# Single-cell transcriptomic atlas of Alzheimer’s disease middle temporal gyrus reveals region, cell type and sex specificity of gene expression with novel genetic risk for MERTK in female

**DOI:** 10.1101/2023.02.18.23286037

**Authors:** Le Zhang, Chuan Hua He, Sarah Coffey, Dominic Yin, I-Uen Hsu, Chang Su, Yixuan Ye, Chi Zhang, Joshua Spurrier, LaShae Nicholson, Carla V. Rothlin, Sourav Ghosh, Pallavi P. Gopal, David A. Hafler, Hongyu Zhao, Stephen M. Strittmatter

**Affiliations:** Department of Neurology, Yale University School of Medicine, New Haven, CT, USA; Department of Neuroscience, Yale University School of Medicine, New Haven, CT, USA; Cellular Neuroscience, Neurodegeneration and Repair Program, Yale University School of Medicine, New Haven, CT, USA; Department of Molecular, Cellular, and Developmental Biology, Yale University, New Haven, CT, USA; Department of Biostatistics, Yale University School of Public Health, New Haven, CT, USA; Department of Immunobiology, Yale University School of Medicine, New Haven, CT, USA; Department of Pharmacology, Yale University School of Medicine, New Haven, CT, USA; Department of Pathology, Yale University School of Medicine, New Haven, CT, USA

## Abstract

Alzheimer’s disease, the most common age-related neurodegenerative disease, is closely associated with both amyloid-ß plaque and neuroinflammation. Two thirds of Alzheimer’s disease patients are females and they have a higher disease risk. Moreover, women with Alzheimer’s disease have more extensive brain histological changes than men along with more severe cognitive symptoms and neurodegeneration. To identify how sex difference induces structural brain changes, we performed unbiased massively parallel single nucleus RNA sequencing on Alzheimer’s disease and control brains focusing on the middle temporal gyrus, a brain region strongly affected by the disease but not previously studied with these methods. We identified a subpopulation of selectively vulnerable layer 2/3 excitatory neurons that that were RORB-negative and CDH9-expressing. This vulnerability differs from that reported for other brain regions, but there was no detectable difference between male and female patterns in middle temporal gyrus samples. Disease-associated, but sex-independent, reactive astrocyte signatures were also present. In clear contrast, the microglia signatures of diseased brains differed between males and females. Combining single cell transcriptomic data with results from genome-wide association studies (GWAS), we identified *MERTK* genetic variation as a risk factor for Alzheimer’s disease selectively in females. Taken together, our single cell dataset revealed a unique cellular-level view of sex-specific transcriptional changes in Alzheimer’s disease, illuminating GWAS identification of sex-specific Alzheimer’s risk genes. These data serve as a rich resource for interrogation of the molecular and cellular basis of Alzheimer’s disease.

## Introduction

Alzheimer’s disease (AD), the most common age-related neurodegenerative disease, is characterized by progressive cognitive decline and dementia. It remains a major unmet medical need with no disease-modifying therapy in clinical practice today^1,2^. Two thirds of AD patients are females, and women have a higher risk of developing AD^3,4^. Women with AD are reported to have more extensive brain histological changes, more severe cognitive symptoms, and more severe neurodegeneration than men with AD^5-9^. However, some studies suggest more rapid progression to death in men^10^, emphasizing our lack of knowledge regarding the differential effect of AD on female and male brains. Thus, research on sex differences in AD is essential to the development of effective interventions and sex-focused therapies. Although sex differences in the risk of AD, vulnerability to genetic load and severity of AD pathology burden have been established^5,11,12^, the underlying mechanisms and molecular pathways that are differentially mediated in female and male AD brains remain poorly understood.

Previous studies have explored certain aspects of genetic variation on sex differences in AD. Apolipoprotein E (*APOE*) has been shown to have a differential impact on age-at-onset between males and females^13,14^. Women who have an *APOE4* allele are more likely to develop AD than are male carriers. This begs the question of whether other known or novel AD risk factors have sex-differential roles in this genetically complex disease. Recent studies suggest that sex differences in genetic risk go well beyond *APOE4*^11,15-17^. Using sex-based Alzheimer’s Disease Genetic Consortium (ADGC) genome-wide association study (GWAS) data and calculation of polygenic hazard scores and polygenic risk scores, it is predicted that effect sizes for *BIN1, MS4A6A, DNAJA2*, and *FERMT2* were higher in women, while *FAM193B, C2orf47*, and *TYW5* variants appeared stronger in men^17^. Using Sex-Specific Family-Based Association Analysis of Whole-Genome Sequence Data, novel loci (*GRID1, RIOK3, MCPH1, ZBTB7C*) were identified showing sex-specific association with AD risk^16^.

It is now clear that inflammation in the central nervous system is an age-dependent process that is associated with impaired memory, cognition, and other brain functions^18-23^, and is a crucial mediator of neurodegenerative diseases^18,19,22^. The important role of neuroinflammation in AD is supported by findings that genes for immune receptors, including *TREM2, CR1* and *CD33* are associated with AD pathology^24,25^. Misfolded and aggregated proteins bind to pattern recognition receptors on microglia and astroglia, triggering an innate immune response characterized by the release of inflammatory mediators that contribute to disease progression and severity^23,26^. Although the importance of myeloid populations in the central nervous system in both health and disease has been increasingly recognized^27-31^, the contribution of brain myeloid cells, microglia in particular^29,32^, to AD-associated neuronal injury remains incomplete. Single cell genomic technologies enable unbiased characterization of cell types and states, transitions from normal to disease, and response to therapies^33-35^. Characterizing the neuroinflammatory networks will be crucial for guiding the development of therapeutic avenues targeting inflammation in AD. Recent research on microglia development identified a microglia-specific gene expression program in mice that was used to create a microglia developmental index. When compared to microglial expression profiles of healthy and AD brain human datasets, differences between males and females^36^ were revealed, suggesting that male microglia are more developmentally mature than female microglia in adult brain. Thus, it is of interest to further explore sex-specific manners of microglia in neuroinflammation and AD.

To define the impact of sex differences on AD risk and neuroinflammation, as well as obtain an accurate and unbiased assessment of sex-specific and cell-type-specific changes associated with AD, we transcriptionally profiled AD brains and age- and sex-matched controls at the single cell level, focusing on the middle temporal gyrus, a brain region strongly affected by AD. We thoroughly characterized our single cell dataset of AD middle temporal gyrus, revealed reactive astrocyte signatures, and identified a subpopulation of excitatory neurons that are selectively vulnerable in Alzheimer’s disease in this brain region. Moreover, this single cell brain atlas of AD brain middle temporal gyrus revealed sex-specific changes in microglia and AD-associated female-specific gene sets. Combining single cell and GWAS data, we analyzed these gene sets and identified *MERTK* as a novel genetic risk factor specific to AD female brains. Taken together, our novel single cell dataset of middle temporal gyrus revealed a unique cellular-level view of sex-specific transcriptional changes in AD, providing a novel tool for analyzing single cell hints using GWAS to identify sex-specific AD risk genes.

## Results

### Single nucleus transcriptomic profiling of human AD brain middle temporal gyrus

To understand the cellular heterogeneity and disease-associated cellular changes in the human AD brain, we performed unbiased massively parallel single nucleus RNA sequencing with postmortem frozen human brain tissues of middle temporal gyrus, a brain cortical region strongly affected by AD. We isolated brain nuclei using sucrose gradient ultracentrifugation, generated single nucleus libraries with 10x Chromium platform (10x Genomics), and sequenced on NovaSeq6000 S4 sequencer (Illumina), from 18 age- and sex-matched individuals with and without AD (Fig. 1a). We integrated our single cell data of human brain middle temporal gyrus from 9 individuals with AD (Braak Stage V/VI) and 9 healthy controls (Braak Stage 0-I/II) by single cell analysis using Seurat^37^ (Supplementary Table 1). After quality control filtering, we profiled and analyzed 104,082 high-quality single nucleus transcriptomes, with 2,153 median genes and 4,460 median UMIs per cell (Supplementary Table 1). We clustered all the cells jointly across the 18 donors that include 8 females and 10 males, and identified and annotated the major cell types of the human brain by interrogating the expression patterns of known gene markers, including neurons (*GRIN1*), excitatory neurons (ExN, *SLC17A7*), inhibitory neurons (InN, *GAD1*), astrocytes (Astro, *AQP4*), microglia (MG, *CSF1R*), oligodendrocytes (Oligo, *MBP*), oligodendrocyte precursor cells (OPC, *PDGFRA*), and endothelial cells (Endo, *CLDN5*) (Fig. 1b, c, Supplementary Fig. 1). We next identified the top marker genes of each major brain cell type, including *ST18, PLP1* and MBP for oligodendrocytes, *SLC17A7, LDB2* and *CLSTN2* for excitatory neurons, *GAD1, GRIP1* and *SYNPR* for inhibitory neurons, *GFAP, RYR3* and *AQP4* for astrocytes, *SPP1, RUNX1* and *DOCK8* for microglia, and *PDGFRA, VCAN* and *XYLT1* for oligodendrocyte precursor cells, which show unique and enriched expression in each cell type (Fig. 1d).

**Fig. 1.**
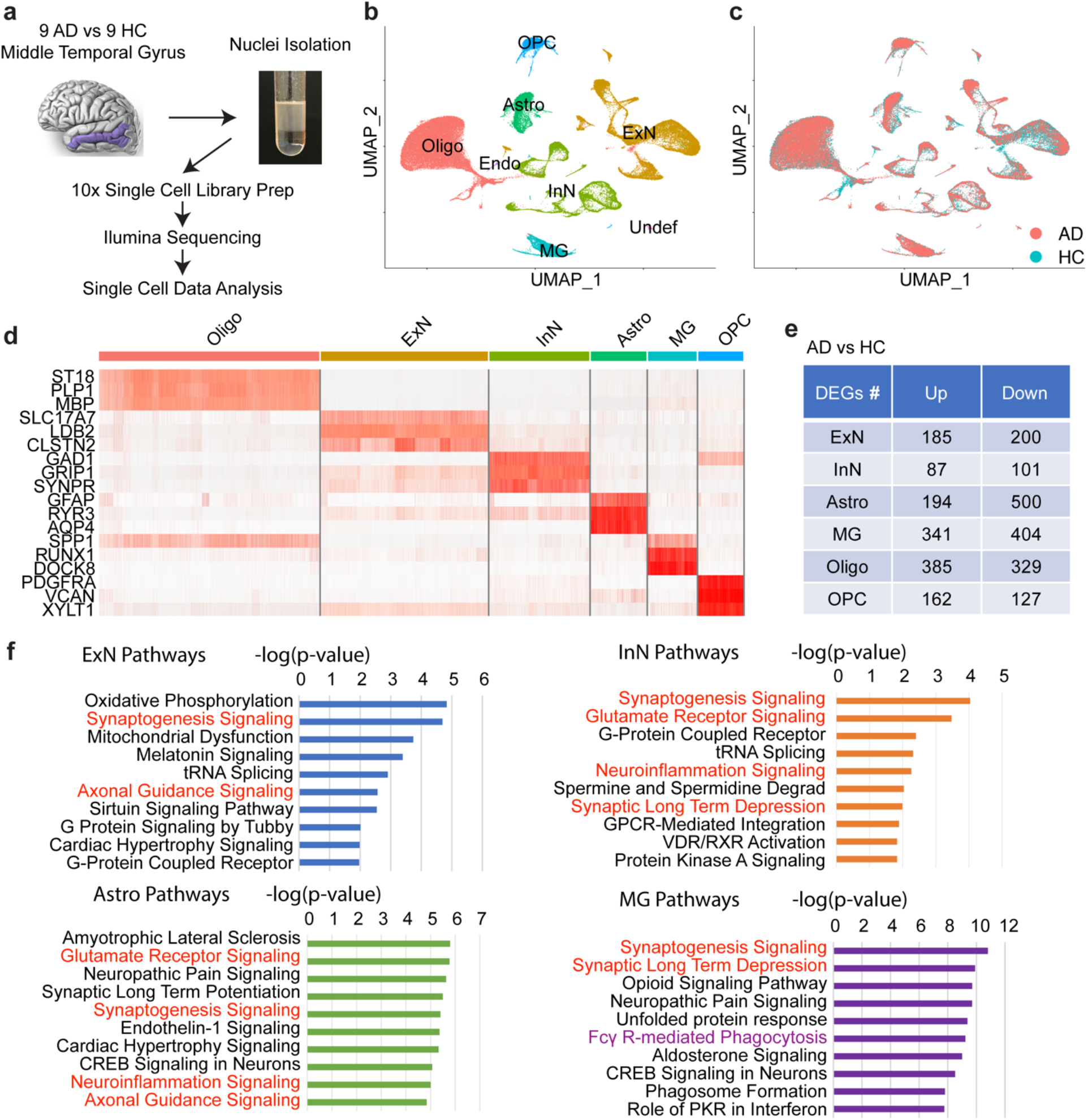
Single nucleus transcriptomic analysis of human brain middle temporal gyrus reveals cell-type-specific changes in AD. **a** Schematic workflow of the sample number, brain region and experimental design. **b-c** UMAP plotting of human brain nuclei (n = 104,082 nuclei), colored by (**b**) major brain cell types: excitatory (ExN) and inhibitory (InN) neurons, oligodendrocytes (Oligo), astrocytes (Astro), microglia (MG), oligodendrocyte precursor cells (OPC), Endothelial cells (Endo) and undefined cells (Undef), and (**c**) disease diagnosis of either Alzheimer’s disease (AD) or healthy controls (HC). **d** Heatmap of top three marker genes in each brain cell type. **e** Differentially expressed genes (DEGs) counts of either upregulated (Up) or downregulated (Down) genes between AD brains and controls for each brain cell type (log2 fold change > 0.25, FDR corrected P-value adjusted < 0.01). **f** Ingenuity Pathway Analysis (IPA) of differentially expressed genes between AD and controls for neuronal (ExN and InN) or glial (Astro and MG) cell types (Top 10 pathways of each cell type were shown; red ink: shared pathways between neuronal and glial cell types; purple ink: unique pathway in microglia).

We performed differential gene expression analysis for each brain cell type between AD and health controls and identified 3,015 significantly differentially expressed genes across all brain cell types that are either upregulated or downregulated in AD brains (Figure 1e, Supplementary Table 2, log2 fold change > 0.25, p value adjusted < 0.01). We validated our single nucleus transcriptomic dataset using the targeted gene expression method, RNAscope *in situ* hybridization on an additional cohort of AD and control brain sections of middle temporal gyrus, focusing on several differentially expressed genes that were identified by single nucleus RNA sequencing, including *AQP4, TREM2, CHRM3* and *HS3ST4* (Supplementary Fig. 2a). We quantified the RNAscope signals at the single RNA molecule level between AD and controls using QuPath (Supplementary Fig. 2b-c), which confirmed differential regulation of these select genes in AD brains compared to controls. We further performed functional analysis for the differentially expressed genes in each major neuronal and glial cell type, such as excitatory neurons, inhibitory neurons, astrocytes and microglia, for the changes in canonical signaling pathways in AD brains compared to healthy controls using Ingenuity Pathway Analysis (IPA) (Fig. 1f, Supplementary Table 3). Interestingly, there were many shared signaling pathways that were affected by the disease across neuronal and glial cell types, including synaptogenesis signaling and synaptic long term depression pathways, which is consistent with the key feature of synapse loss and dysfunction in AD. Neuroinflammation signaling pathway also was altered in both neuronal and glial cell types, consistent with a major role of neuroinflammation in AD pathogenesis. A number of unique pathways were changed in a certain brain cell type in AD, for example, Fc*γ* R-mediated phagocytosis was elevated in AD microglia, which may function as a protective role in the neurodegenerative disease^38,39^.

### Selective vulnerability of excitatory neuron subpopulations in AD middle temporal gyrus

Next, we focused on the neuronal changes in AD brain middle temporal gyrus and unbiasedly re-clustered all neurons into neuronal subpopulations with seven subclusters of excitatory neurons (ExN1, ExN2, …, ExN7) and six subclusters of inhibitory neurons (InN1, InN2, …, InN6) (Figure 2a). We identified a reduction of neurons specifically in excitatory neurons in AD brains, while the cell subtype proportion was similar in inhibitory neuron subpopulations between AD and control brains, which suggests that most selectively vulnerable neurons in AD are excitatory neurons in the brain region of middle temporal gyrus (Fig. 2b, c). Within excitatory neurons, the most significant decline of cell composition and relative abundance in AD was in the subpopulations of ExN1 and ExN6, which showed a severe loss of neurons in AD brains compared to healthy controls in those two subpopulations (Fig. 2c). A similar pattern was observed for male and female samples (Supplementary Fig. 3). We further analyzed the neuronal subpopulations by assessing the top marker genes of each neuronal cluster from either excitatory neurons or inhibitory neurons as shown in the heatmap plot (Fig. 2d, Supplementary Table 4).

**Fig. 2.**
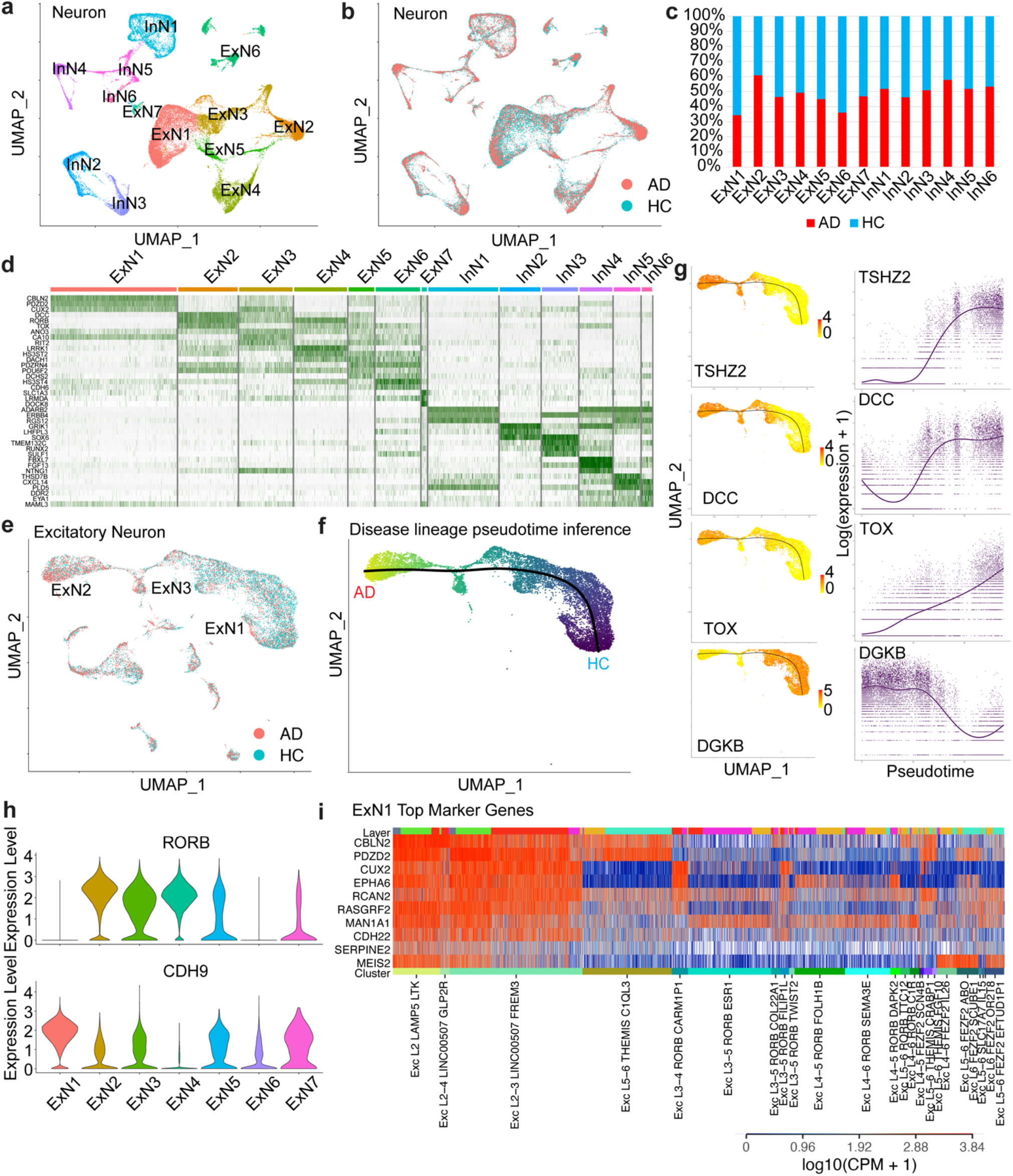
Single cell analysis of neurons reveals selective vulnerability of excitatory neuron subpopulations in human AD brain middle temporal gyrus. **a-b** UMAP plotting of human brain neuronal nuclei (n = 43,197 neurons from 18 individuals with and without AD), colored by neuronal cell subtypes (**a**), such as excitatory neurons (ExN) and inhibitory neurons (InN), or disease diagnosis of either AD or controls (HC) (**b**). **c** Neuronal cell type composition between AD brains or healthy controls. **d** Heatmap of top three marker genes in each excitatory or inhibitory neuronal cell subpopulation in human brain middle temporal gyrus. **e** UMAP plotting of human brain excitatory neurons, colored by disease diagnosis of either AD or controls. **f** UMAP plotting of human brain excitatory neuronal subpopulations ExN1, ExN2 and ExN3, with colors from dark blue to light yellow showing the disease lineage pseudotime inference along the simulation trajectory. **g** Differential expression of significantly altered genes along the disease trajectory (left panels) and their expression levels along the pseudotime (right panels). **h** Violin plotting of the expression of *RORB* and *CDH9* in the subpopulations of excitatory neurons in the middle temporal gyrus. **i** The alignment of highly vulnerable excitatory neuronal subpopulation ExN1 using its top marker genes to Allen Brain single cell atlas of middle temporal gyrus, grouped by brain cortical layers and clusters.

Based on their transcriptional profiles, three major excitatory neuronal subpopulations of ExN1, ExN3 and ExN2 were unbiasedly clustered together in UMAP space and formed a lineage continuum pattern, suggesting a spectrum of cell lineage influencing susceptibility to AD across these major subclusters of excitatory neurons. We subsetted and re-clustered the excitatory neurons (Fig. 2e), performed cellular linage and pseudotime inference using Slingshot^40^ on ExN1, ExN2 and ExN3, which presented a smooth curve as a trajectory for the simulation of the disease lineage from healthy control to AD, and then calculated the differential expression genes along this disease trajectory using TradeSeq^41^ (Fig. 2f, Supplementary Table 5). Along the disease progression trajectory, a set of genes, such as *TSHZ2, DCC* and *TOX*, were upregulated from control to AD, while other genes, such as diacylglycerol kinase *DGKB* for neurite spine formation, were downregulated in the AD excitatory neurons, which may contribute to impaired neuronal function and cognitive decline (Fig. 2g).

A recent study reported that RORB-expressing excitatory neuron subpopulations were selectively vulnerable in AD specifically in the entorhinal cortex, though this pattern was not detected in either superior frontal cortex or prefrontal cortex^42,43^. We further assessed our single cell atlas of middle temporal gyrus, and surprisingly, we identified that the most vulnerable excitatory neurons in AD, such as ExN1 and ExN6 (Fig. 2c), in this distinct brain region are RORB-negative and CDH9-expressing cells (Fig. 2h). Notably, RORB-expressing excitatory neuronal subpopulations of ExN2-ExN5 are abundant in the middle temporal gyrus of AD brains. This demonstrates that RORB-expressing excitatory neurons are affected differently in distinct brain regions during AD pathogenesis. We next performed marker gene alignment for the top marker genes of the most vulnerable excitatory neuron subpopulation ExN1 to an established human middle temporal gyrus brain cell atlas by the Allen Brain Institute, which was generated by Smart-seq2 full length cDNA single cell sequencing. In the middle temporal gyrus, the highly vulnerable RORB-negative and CDH9-expressing excitatory neurons in AD are mostly in the cortical layers 2 and 3 (Fig. 2i).

### Pan reactivation of astrocytes in AD brain middle temporal gyrus

To investigate the role of astrocytes in AD brains, we subsetted and re-clustered astrocytes into 10 subpopulations of AS0, AS1, …, AS9 (Fig. 3a-b), which did not show a notable change in the cell type proportion of astrocytes between AD and healthy controls (Fig. 3c). Neuroinflammation may induce two different types of reactive astrocytes termed as A1 and A2, which were previously identified in mice, whereas A1 astrocytes are neuroinflammatory and A2 are proliferative and anti-inflammatory^44^. We investigated whether those reactive astrocytes are induced in AD using *C3* as the A1 marker gene and *S100A10* as the A2 marker gene. We found that both A1 and A2 astrocytes are highly induced in human AD brain middle temporal gyrus (Fig. 3d), across most of the astrocytes subclusters, with a dramatic increase in the subclusters of AS2, AS3 and AS4 (Fig. 3e). This indicates the presence of both harmful A1 astrocytes and the potentially protective A2 astrocytes in AD pathogenesis. Since reactive astrocytes may be induced by central nervous system disease, including neurogenerative diseases such as AD, we further explored the features of astrocytes in the AD brain middle temporal gyrus using 12 known pan reactive markers and transcripts^44^ for their differential expression analysis (Fig. 3f). We identified a robust induction of reactive astrocytes in AD brain middle temporal gyrus with all 12 marker genes being significantly upregulated in AD compared to healthy controls. These findings show a pan reactivation of astrocytes induced by AD without obvious sex-specific variation (Supplementary Fig. 4).

**Fig. 3.**
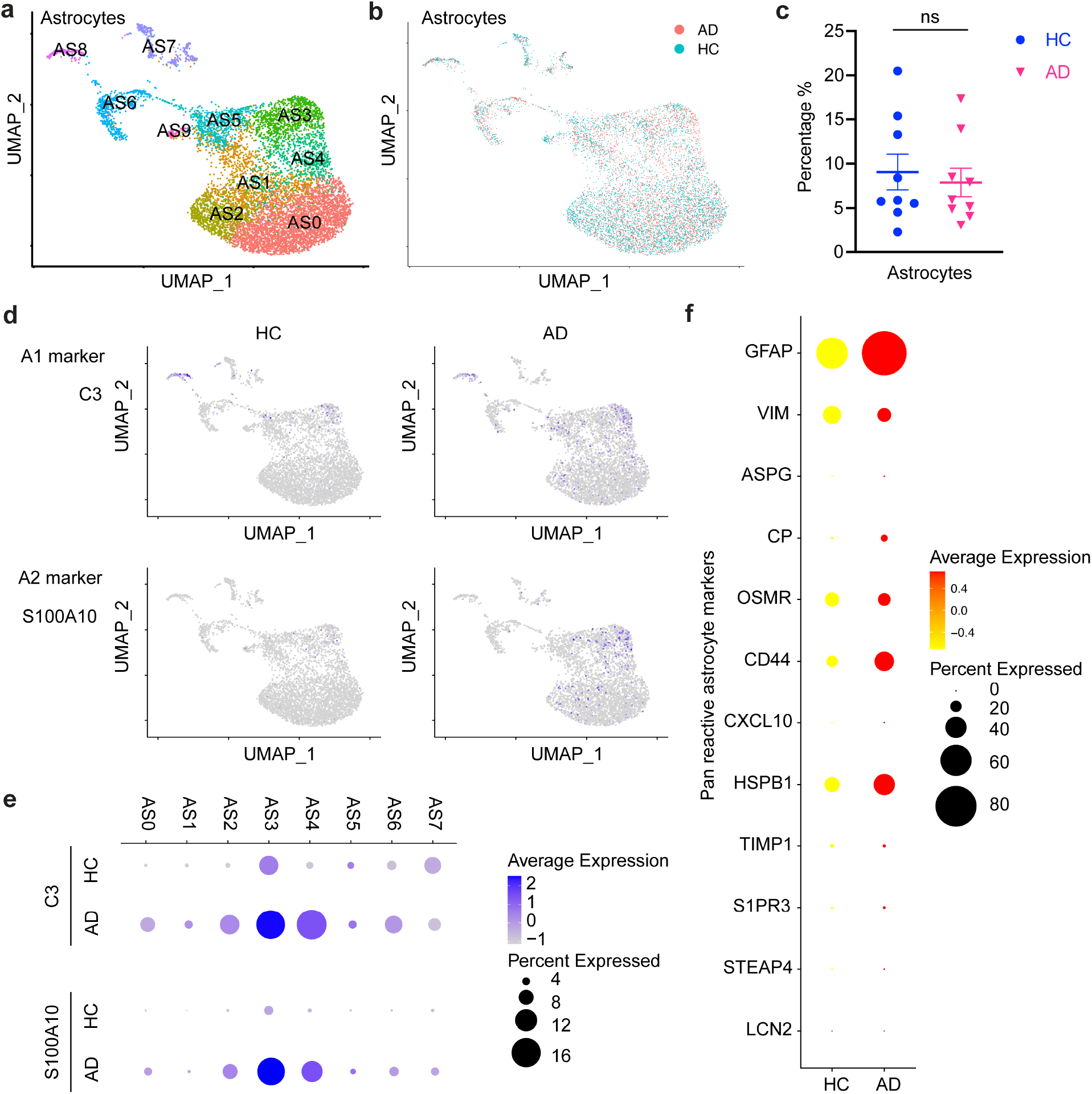
Single cell analysis of astrocytes reveals pan reactivation in human AD brain middle temporal gyrus. **a-b** UMAP plotting of human brain astrocytes (n = 9,000 astrocytes from 18 individuals with and without AD), colored by astrocyte subpopulations (AS0 - AS9) (**a**), or disease diagnosis of either AD or controls (HC) (**b**). **c** The percentage of astrocytes in all brain cells in each individual of AD or HC. ns, no significant difference. **d** UMAP plotting of the expression levels of A1 astrocytes marker gene *C3* or A2 astrocytes marker gene *S100A10* in AD and HC. **e** Dot plotting of the expression levels of A1 astrocytes marker gene *C3* or A2 astrocytes marker gene *S100A10* in each astrocyte subpopulation between AD and HC. **f** Dot plotting of the expression levels of pan reactive astrocytes marker genes as listed between AD and HC.

### AD-associated and female-specific cellular subpopulations and gene sets in microglia

Based on the cellular composition, we observed a specific increase of microglia population in AD brains compared to healthy controls, while there is no change in the percentage of neuronal and other glial cell populations (Fig. 4c). We subsetted and analyzed microglia in the middle temporal gyrus through re-clustering at a finer resolution into 10 subpopulations and identified AD-associated female-specific microglia subclusters of MG0, MG4 and MG7 (Fig. 4a-b, d-e). The marker gene sets of these subclusters provided sex-specific transcriptional features of AD-associated and female-specific microglia subpopulations (Supplementary Table 6). The top marker genes of MG4 showed AD- and female-specific upregulation, such as *CD163, TPRG1* and *MYO1E*. Interestingly, although the complement genes of *C1QA* and *C1QC* showed higher expression in male microglia than in female healthy controls, their expression was increased to a substantially greater degree in only AD female microglia, revealing a female-specific change in AD pathogenesis (Fig. 4f).

**Fig. 4.**
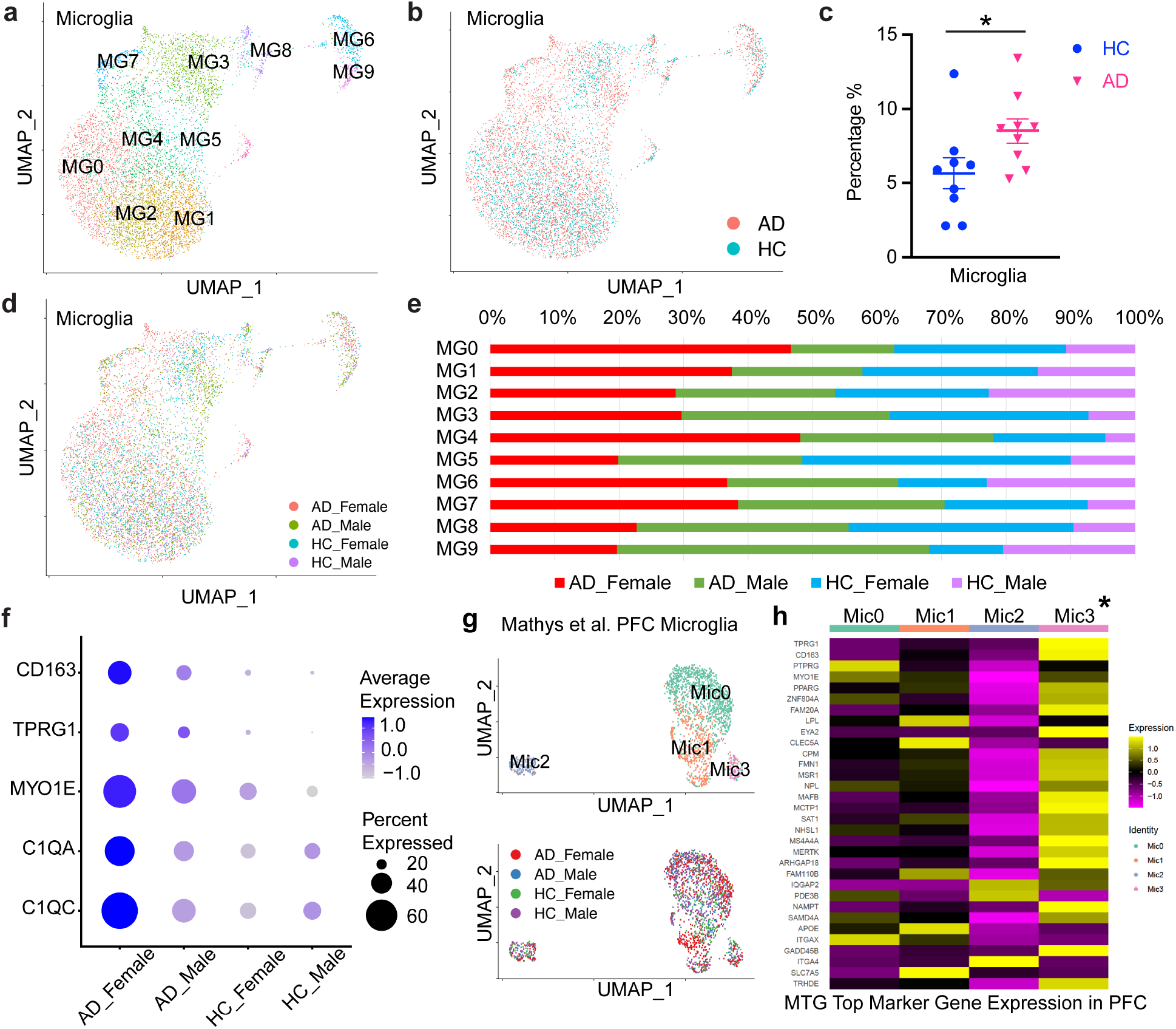
Single cell analysis reveals AD-associated and female-specific microglial subpopulations and gene sets in human AD brain middle temporal gyrus. **a-d** UMAP plotting of human brain microglia (n = 7,874 microglia from 18 individuals with and without AD) in the middle temporal gyrus, colored by microglial subpopulations (MG0 - MG9) (**a**), or disease diagnosis of either AD or controls (HC) (**b**), or sex and disease diagnosis (**d**). **c** The percentage of microglia in all brain cells in each individual of AD or HC. * p-value = 0.0391, wilcoxon test. **e** Microglial cell subtype composition between four groups of AD female (red), AD male (green), healthy control female (blue), and healthy control male (purple) brains. **f** Dot plotting of the expression levels of AD-associated and female-specific marker genes as listed between healthy control males, AD males, healthy control females, and AD females. **g** UMAP plotting of human brain microglia (n = 7,874 microglia from 18 individuals with and without AD) in the prefrontal cortex single cell dataset from Mathys et al., colored by microglial subpopulations (mic0 – mic3, upper panel), or sex and disease diagnosis (lower panel). **h** Gene set enrichment analysis (GSEA) for the comparison of AD-associated and female-specific microglial marker gene sets in the middle temporal gyrus (MTG) and prefrontal cortex (PFC). AD-associated and female-specific MTG microglial subcluster gene set of MG4 is correlated with AD-associated and female-specific PFC subcluser Mic3.

We also compared our microglia dataset from middle temporal gyrus with the microglia from the recently published and first human AD brain prefrontal cortex single cell dataset^45^. We reanalyzed the prefrontal cortex microglia data, and identified four subclusters of Mic0, Mic1, Mic2 and Mic3, of which Mic1 is AD- and female-specific cluster as reported previously^45^ (Fig. 4g). To define the correlation of sex-specific microglial features between these two cortical brain regions, we used gene set enrichment analysis (GSEA) to compare AD-associated and female-specific marker gene sets in both middle temporal gyrus and prefrontal cortex microglia, as well as their subclusters. Strikingly, the AD-associated and female-specific middle temporal gyrus microglia subcluster marker gene set significantly overlapped with that from a prefrontal cortex microglia subcluster (Fig. 4h), which revealed common gene expression features of microglial subpopulation in these two brain regions, indicating that overrepresentation of AD-associated and female-specific microglia subpopulation is robust across multiple brain regions. We also compared both the observed microglia cell proportions in the reported single cell AD prefrontal cortex^45^ data across different sample groups, and inferred microglia cell proportions through deconvoluting bulk RNA-seq data through CIBERSORTx^46^, and observed the same directional changes with statistically significant results (data not shown).

### Identification of novel female-specific AD risk gene *MERTK* from GWAS data

To extend our assessment of AD-associated and female-specific microglial gene sets, we leveraged GWAS data from the Alzheimer’s Disease Genetics Consortium (ADGC) to identify candidate sex-biased and sex-specific AD risk genes in the female AD and microglial specific gene sets that were revealed by single nucleus transcriptomic analysis. We used ADGC data to conduct both overall and sex-specific GWAS analysis using BOLT-LMM. The ADGC dataset was generated from 25,772 participants (Ncase = 13,271), of which 10,468 were male (Ncase = 5,441) and 15304 were female (Ncase = 7,830). We included age and the first 10 genetic principal components as covariates. With the sex-specific GWAS summary statistics, we analyzed three AD-associated and female-specific gene sets (MG0, MG4 and MG7) from microglia using MAGMA^47^, which is a tool for generalized gene-set analysis of GWAS data. We calculated the associations of top marker genes in MG0/MG4/MG7 with AD among the males, females, and the overall population, and identified a set of known AD risk genes of *APOE, APOC1* and *MS4A4A* family in all populations that includes both male and female individuals, which confirms the ability of our pipeline with integrated transcriptomic and genomic tools for identifying AD risk genes in GWAS data (Fig. 5a-b, Supplementary Fig. 5a). Remarkably, we identified the AD risk gene *MERTK* specifically in the female AD population, but not in the entire population nor in the male population after adjusting for multiple comparisons across these marker gene sets (Fig. 5a-b).

**Fig. 5.**
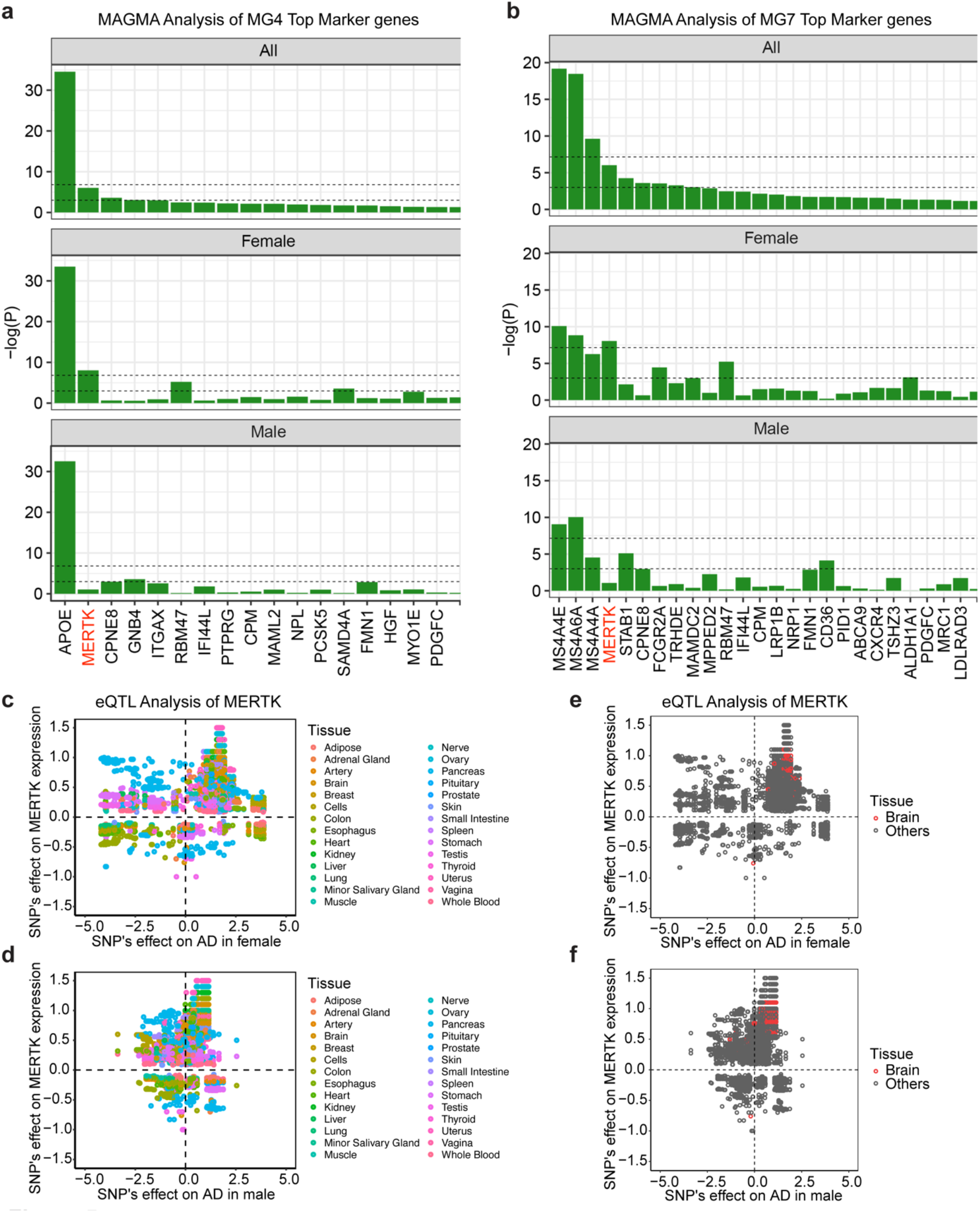
Integrated single cell and GWAS analysis identified novel female-specific AD risk gene *MERTK*. **a-b** Associations between AD and female-specific microglial subcluster MG4 (**a**) or MG7 (**b**) marker gene sets using MAGMA analysis based on ADGC GWAS dataset. The lower dashed line represents a p-value of 0.05 and the upper dashed line indicates a p-value threshold after Bonferroni correction (0.05 divided by the number of genes in the corresponding cluster). Top panel: all population with both male and female; middle panel: female only; bottom panel: male only, in the ADGC dataset. **c-d** *MERTK* eQTL analysis in all tissues in female (**c**) or male (**d**), where each point indicates one SNP in a specific tissue, the x-axis is the SNP’s effect (logOR from GWAS) on AD in female (**c**) or male (**d**), and the y-axis is the SNP’s effect (NES from GTEx) on the expression level of *MERTK* in the specific tissue (points in the same vertical line are usually the same SNP in different tissues). **e-f** MERTK eQTL analysis in brain tissue (red circles) compared to all the other tissues (gray circles) in female (**e**) or male (**f**), where each point indicates one SNP in a specific brain sub-tissue, the x-axis is the SNP’s effect (logOR from GWAS) on AD in female (**e**) or male (**f**), and the y-axis is the SNP’s effect (NES from GTEx) on the expression level of *MERTK*.

We generated LocusZoom plots for *MERTK* as a novel AD risk gene in both female and male GWAS data for direct visual comparison, which revealed several SNPs with r^2^ > 0.8 in the locus of *MERTK* in only the female population and not the males, including rs4264571(Supplementary Fig. 3b). To define the potentially involved human tissues, we then conducted tissue-specific gene-level association between *MERTK* and AD using UTMOST^48^ v8, which showed that the associations were significant in females not in males in most of the tissues including brain sub-tissues specifically (Supplementary Fig. 3c).

We next conducted an expression quantitative trait loci (eQTL) analysis to further explore the role of *MERTK* in terms of AD risk. We extracted *MERTK* significant eQTLs across all tissues from the GTEx v8 database and tested the correlation between these eQTLs’ effects (logOR from AD GWAS) on AD and their effects (normalized effect size, NES from GTEx) on *MERTK*’s expression in either females or males (Fig. 5c-d). We found that in most tissues, these two sets of effect sizes are positively correlated, which suggests from the genetic perspective, higher *MERTK* expression corresponds to a higher risk of AD. Notably, in brain tissues, higher *MERTK* expression corelates with higher AD risk specifically in females not males (Fig. 5e-f), which indicates that *MERTK* may play a risk role in the development of AD pathogenesis specifically in females.

## Discussion

Combining single cell transcriptomics and GWAS data, we analyzed AD brains and sex-specific gene sets, which are unique in microglia, and identified a novel AD genetic risk gene *MERTK* specifically in females. This provides a novel tool for analyzing single cell hints using GWAS data for revealing sex-specific transcriptional changes and identifying sex-specific risk genes in disease. Both TAM receptor tyrosine kinases Mertk (*MERTK*) and Axl (*AXL*) are regulators of microglial physiology^49^ and have been linked to AD^50^, but their roles in the disease may need further investigation. It has been suggested that Axl has a protective role in regulating neuroinflammation and underlying AD, by the longitudinal study of elevated Axl in CSF^51^. Also, in a recent study, higher levels of ApoE and soluble TAM receptors Axl and Tyro3 were detectable even before the onset of cognitive impairment and could serve as reliable indicators of the subsequent development of AD^52^. Intriguingly, our study revealed an upregulation of *AXL* at the transcriptional level in microglia specifically in AD male patients (Supplementary Fig. 6), in contrast to the elevated *MERTK* level in AD females (Fig. 5, Supplementary Fig. 6), which indicates the distinct roles of *AXL* and *MERTK* in regulating neuroinflammation and AD, whereas *AXL* may be protective and expressed more in males, and *MERTK* might be harmful and more in females, resulting sex differences in AD.

Neuroinflammation induced A1 type reactive astrocytes were found abundant in human neurodegenerative diseases, including AD^44,53^. Our data is consistent with previous observation of A1 astrocytes in AD and further showed the increase of A2 reactive astrocytes in the postmortem brains of patients with AD, which may be protective. This indicates the disease progression is complicated with the involvement of both types of reactive astrocytes for neurotoxic A1 and protective A2 astrocytes. Potential blockade of A1 astrocyte conversion was shown to be neuroprotective in models of Parkinson’s disease^54^. It may be helpful to target the reactive astrocytes and further explore the role and application of blocking A1 astrocytes and/or promoting A2 astrocytes in AD.

Selective neuronal vulnerability is a fundamental feature of AD and has been characterized in different brain regions of postmortem AD brains, such as entorhinal cortex, prefrontal cortex and superior frontal gyrus, with diverse findings of the most vulnerable neurons^42,43^. Here we provide the molecular signature of another brain region of middle temporal gyrus that is strongly affected by AD, as well as the selectively vulnerable neurons in this brain region, which are RORB-negative and CDH9-expressing excitatory neurons, different from other brain regions. Our findings demonstrate the brain region specificity of AD pathogenesis and emphasize the importance of potential treatment in targeting certain neuronal populations with a cautious consideration of specific brain regions and disease progression stages.

## Methods

### Human brain tissue samples

The autopsy brain specimens were collected and dissected at Alzheimer’s Disease Research Center (ADRC) at Yale University in New Haven, CT. The samples are fresh frozen brain tissues with post-mortem intervals of less than 24 hours in most of the cases (average 12 hours). Exclusion criteria for tissue specimens included prolonged agonal state, and other confounding medical conditions. Each sample includes a de-identified full autopsy report of neuropathology evaluation and neuro/AD diagnosis based on recommendations of the National Institute on Aging-Alzheimer Association (NIA-AA)^55^, including stains with antibodies to a-synuclein, Aβ and Tau as well as CERAD and Braak staging. A detailed description of the age and sex of the donors, histopathology and clinical diagnosis is listed in Supplementary Table 1.

### Brain nuclei isolation

Nuclei were isolated from post-mortem fresh frozen human brain tissues of middle temporal gyrus. Frozen tissue (50 to 100 mg) was homogenized in 15 ml of ice-cold nuclei homogenization buffer [2 M sucrose, 10 mM Hepes (pH 7.9), 25 mM KCl, 1 mM EDTA (pH 8.0), 10% glycerol, and ribonuclease (RNase) inhibitors freshly added (80 U/ml)] with Dounce tissue grinder (10 strokes with loose pestle and 10 strokes with tight pestle). The homogenate was transferred into an ultracentrifuge tube on top of 10 ml of fresh nuclei homogenization buffer and centrifuged at 25,000 rpm for 60 min at 4°C on ultracentrifuge. The supernatant was removed, and the pellet was resuspended in 1 ml of nuclei resuspension buffer [15 mM Hepes (pH 7.4), 15 mM NaCl, 60 mM KCl, 2 mM MgCl2, 3 mM CaCl2, and RNase inhibitors freshly added (80 U/ml)] and counted on a hemocytometer. The nuclei were centrifuged at 800g for 10 min at 4°C and resuspended at a concentration of 700 to 1200 nuclei/ul for 10x Genomics Chromium loading.

### 10x single nucleus library construction and sequencing

The single nucleus libraries were prepared by the Chromium Single Cell 3′ Reagent Kit v3 chemistry according to the manufacturer’s instructions (10x Genomics) and sequenced using Illumina NovaSeq6000 S4 sequencer, with minimum 300M reads per sample.

### Data processing

For single nucleus RNA sequencing of brain middle temporal gyrus, a custom pre-mRNA human genome reference was generated GRCh38 that included pre-mRNA sequences, and sequencing data were aligned to this GRCh38–pre-mRNA reference to map both unspliced pre-mRNA and mature mRNA using CellRanger version 3.1.0.

### Single cell data analysis

We used Seurat^37^ for single cell data QC, cell clustering, brain cell type annotation (Supplementary Figure 1a), and differential gene expression. Data from different individuals was integrated using *FindIntegrationAnchors* and *IntegrateData* functions. For visualization, the dimensionality of the dataset was reduced by the Uniform Manifold Approximation and Projection (UMAP) based on Principal Component Analysis (PCA). Gene signaling pathway analysis was performed using Ingenuity Pathway Analysis (IPA, QIAGEN). Disease lineage and pseudotime inference for single cell transcriptomics were performed using Slingshot^40^. Trajectory-based differential expression analysis for single cell dataset were performed using using TradeSeq^41^.

### GWAS analysis

MAGMA was used for both gene and gene-set analysis, where the gene analysis is based on a multiple regression model and has been shown to provide robust statistical performance. UTMOST was used for transcriptome-wide association analysis (TWAS), which provides both single-tissue results and cross-tissue results. There are three steps in UTMOST: 1) Gene expression imputation models were firstly trained using GTEx v8 data, where expression levels were normalized and adjusted for possible confounding factors as described in the eQTL analysis in GTEx v8 project; 2) next, the trained cross-tissue gene expression imputation weights were combined with the GWAS summary statistics to identify disease-associated genes in each single tissue; and finally, association test statistics from tissues were combined using the Generalized Berk–Jones test for each gene. Significant disease-associated genes across all tissues and in each single tissue were identified after Bonferroni multiple-testing correction. During our analyses, we mainly used the single-tissue results from UTMOST to prioritize the tissues for AD association.

### RNA *in situ* hybridization, imaging, and quantification

To validate gene expression differences identified by snRNA-seq, RNA *in situ* hybridization was performed using the RNAscope Multiplex Fluorescent v2 Assay (Advanced Cell Diagnostics, Inc., Newark, CA, USA). Human brain tissue from the middle temporal gyrus was flash-frozen at an average post-mortem interval of 12 hours and stored at -80°C. Prior to performing the assay, frozen tissue was embedded in OCT and sectioned at 10*μ*m using a cryostat Leica CM3050S. RNA *in situ* hybridization was performed according to the manufacturer’s protocol. Briefly, fresh frozen sections were fixed in 10% NBF for 1 hour at room temperature, then sequentially dehydrated in 50% EtOH, 70% EtOH, 100% EtOH, and 100% EtOH for 5 minutes each at room temperature. Tissue sections were pretreated with hydrogen peroxide and protease to block endogenous peroxidase activity and optimally permeabilize the sections. Target probes, such as *AQP4, TREM2, CHRM3* and *HS3ST4*, were hybridized to tissue sections, signals were amplified, and fluorescent dyes were applied. Positive control probes for human POLR2S, PPIB, UBC, and HPRT-1 and a negative control probe for *Bacillus subtilis* DapB were used to ensure detection of target RNAs and a lack of non-specific probe binding. All tissue sections were counterstained with DAPI. TrueBlack lipofuscin autofluorescence quencher was used to remove autofluorescent signals. Images were captured using a Leica SP8 motorized staging confocal microscope with a 20x lens (Leica #506517), while confocal imaging settings were kept constant between healthy controls and AD. RNA expression was quantified using QuPath, by detecting the cells using DAPI channel, identifying and measuring probe channel signals, and calculating estimated spot count per cell for the entire cryosection. RNA expression in human brain sections from AD patients was compared to that in healthy control human brains in approximately 10,000 nuclei from each tissue section, and statistical significance was determined using Student’s t-test.

## Data Availability

All postmortem brain single nucleus RNA sequencing data is available through the National Center of Biotechnology Information's Gene Expression Omnibus (GEO) at the accession number GSE188545, including raw sequencing data of fastq files and processed data of gene expression matrix. Additional data related to this paper may be requested from the authors. Code for Seurat, Slingshot, TradeSeq, MAGMA, and UTMOST is available at github.com. Additional code is available upon request.

## Author contributions

L.Z. and S.M.S. conceptualized the study. L.Z. C.H.H., S.C., I.H. and D.Y. performed experiments including nuclei isolation, 10x snRNA-seq, single cell data analysis, pathway analysis, RNAscope and imaging. J.S. and L.N. prepared tissues blocs for molecular studies. P.G. was responsible for pathological diagnosis and curation. C.S., Y.Y., and H.Z. performed bioinformatic and GWAS analyses. C.R. and S.G. contributed to assessment of TAM family proteins. L.Z. and S.M.S. wrote the manuscript with input from all authors.

## Acknowledgements

This work was supported by research funding from NIH NIA R56AG074015 (L.Z., H.Z., C.R., S.G. and S.M.S.), NIA P30AG066508 (L.Z. and S.M.S.), NIA R01AG034924 (S.M.S.), NIA R01AG066165 (S.M.S.), NIA RF1AG070926 (S.M.S.), NIDA DP2DA056169 (L.Z.) and Women’s Health Research Center at Yale (L.Z. and S.M.S.). L.Z. and D.A.H also receive research support from joint efforts of the Michael J. Fox Foundation for Parkinson’s Research (MJFF) and the Aligning Science Across Parkinson’s (ASAP) initiative (ASAP-000529). We thank Philip Coish for reading manuscript and Yale Center for Genome Analysis for 10x Genomics library preparation and Illumina sequencing.

## Ethics statement

All tissue was obtained from pre-existing autopsy tissue without personal identifying information and deemed exempt from human subjects regulation.

