## Supplementary Figures 1-6 for "Single-cell transcriptomic atlas of Alzheimer’s disease middle temporal gyrus reveals region, cell type and sex specificity of gene expression with novel genetic risk for MERTK in female"

Supplementary Tables 1-6 provided as Excel Files, and Supplementary Figures 1-6.

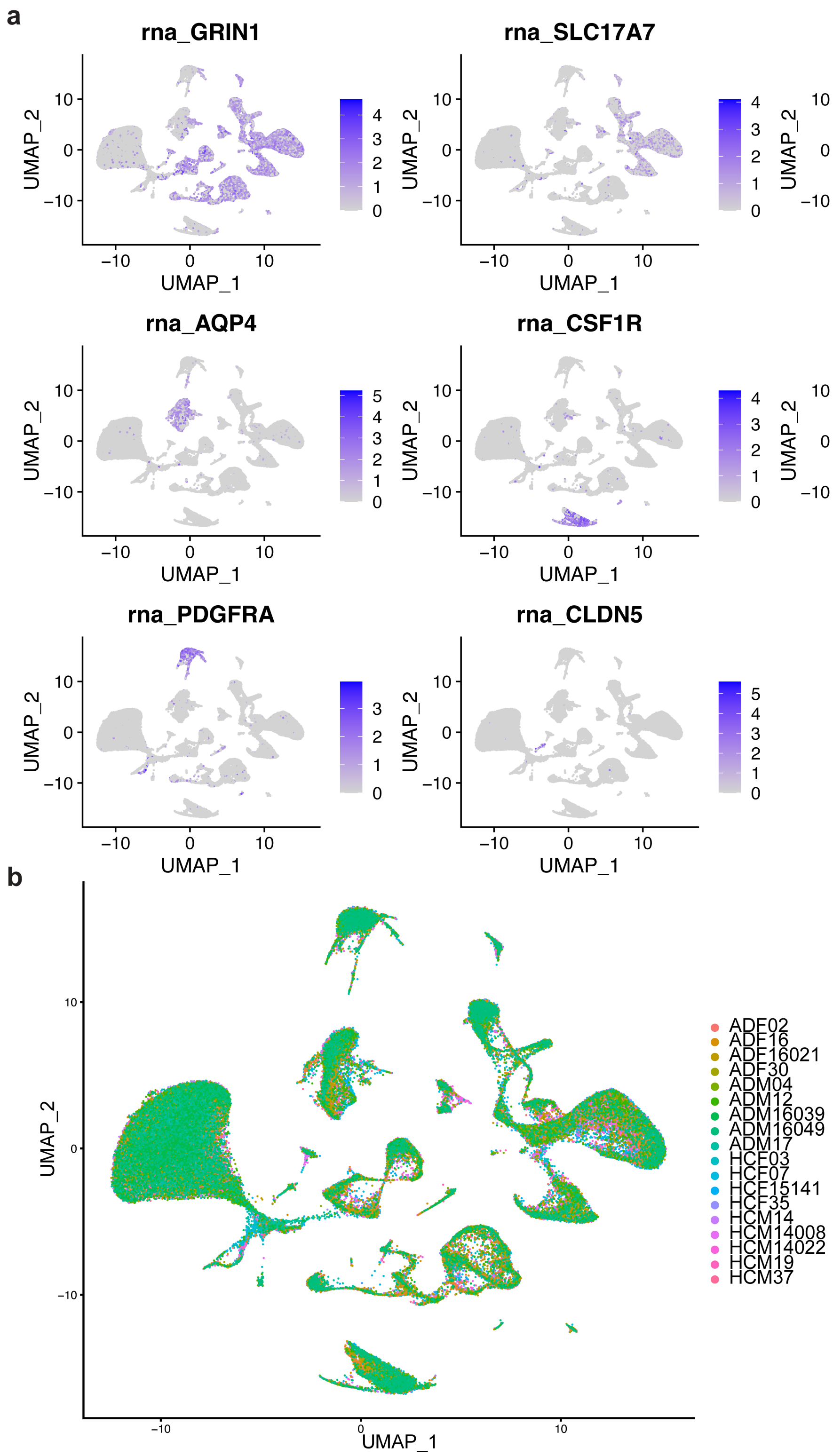


**Supplementary Fig. 1 Single nucleus RNA sequencing QC of human brain middle temporal gyrus. a** UMAP plotting of human brain nuclei (n = 104,082 nuclei) with brain cell type annotations, for neurons (*GRIN1*), excitatory neurons (*SLC17A7*), inhibitory neurons (*GAD1*), astrocytes (*AQP4*), microglia (*CSF1R*), oligodendrocytes (*MBP*), oligodendrocyte precursor cells (*PDGFRA*), and endothelial cells (*CLDN5*). **b** UMAP plotting of human brain nuclei (n = 104,082 nuclei) from 18 individuals with and without AD (ADF: females with Alzheimer's disease, ADM: males with Alzheimer's disease, HCF: healthy control females, HCM: healthy control males).

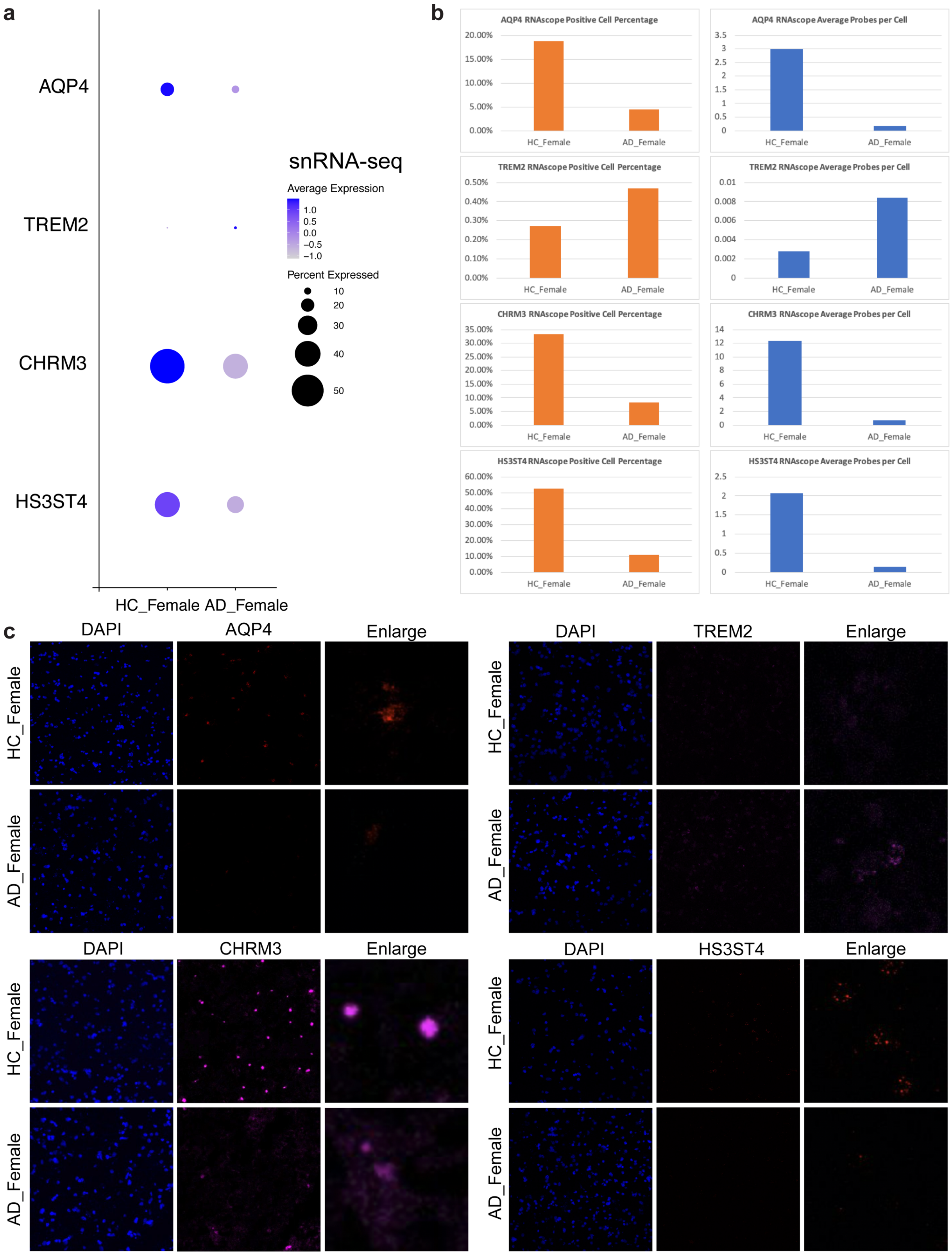


**Supplementary Fig. 2 RNAscope *in situ* hybridization validation of differentially expressed genes in AD and control brain middle temporal gyrus. a** Dot plot of differential expressed genes of *AQP4, TREM2, CHRM3* and *HS3ST4* in single nucleus RNA sequencing dataset (snRNAseq) between female AD (AD_female) brains and female healthy controls (HC_Female). **b** QuPath quantification of RNAscope data for *AQP4, TREM2, CHRM3* and *HS3ST4*. Left panels: RNAscope positive cell percentage in all cells; right panels: RNAscope average probe copies per cell. **c** RNAscope images of *AQP4, TREM2, CHRM3* and *HS3ST4* in female AD (AD_female) or female healthy control (HC_Female) brains. (Blue: DAPI; Red or Magenta: genes as labeled; Enlarge: high magnification images).

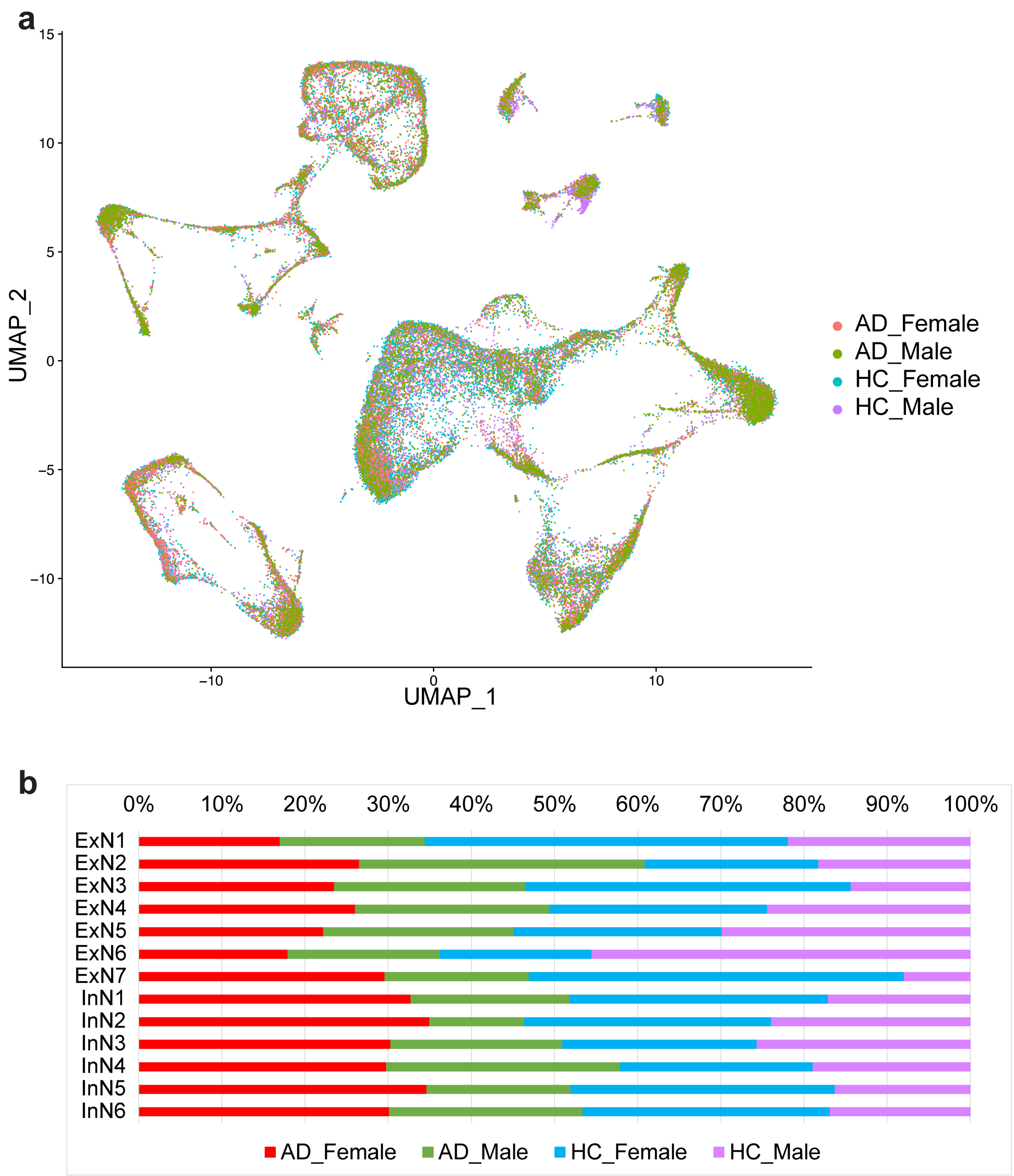


**Supplementary Fig. 3 Single cell analysis of sex difference in the neurons from human AD and control brain middle temporal gyrus. a** UMAP plotting of human brain neuronal nuclei (n = 43,197 neurons from 18 individuals with and without AD), colored by sex and disease diagnosis. **b** Neuronal cell subtype composition of excitatory neuronal subpopulations (ExN) and inhibitory neuronal subpopulations (InN) between four groups of AD female (red), AD male (green), healthy control female (blue), and healthy control male (purple) brains.

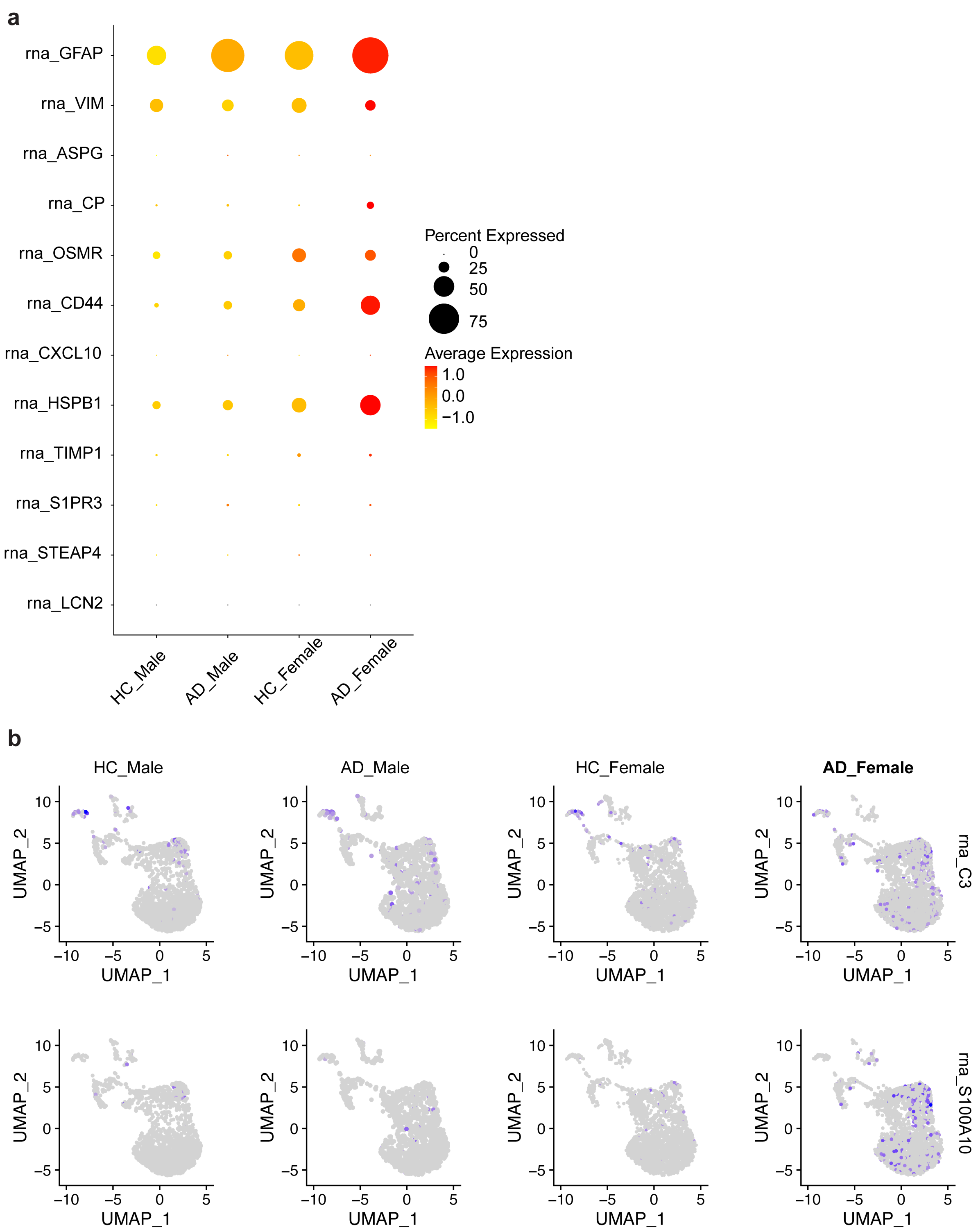


**Supplementary Fig. 4 Single cell analysis of sex difference in astrocytes from human AD brain middle temporal gyrus. a** Dot plotting of the expression levels of pan reactive astrocytes marker genes as listed between healthy control males, AD males, healthy control females, and AD females. **b** UMAP plotting of the expression levels of A1 astrocytes marker gene *C3* or A2 astrocytes marker gene *S100A10* in healthy control males, AD males, healthy control females, and AD females.

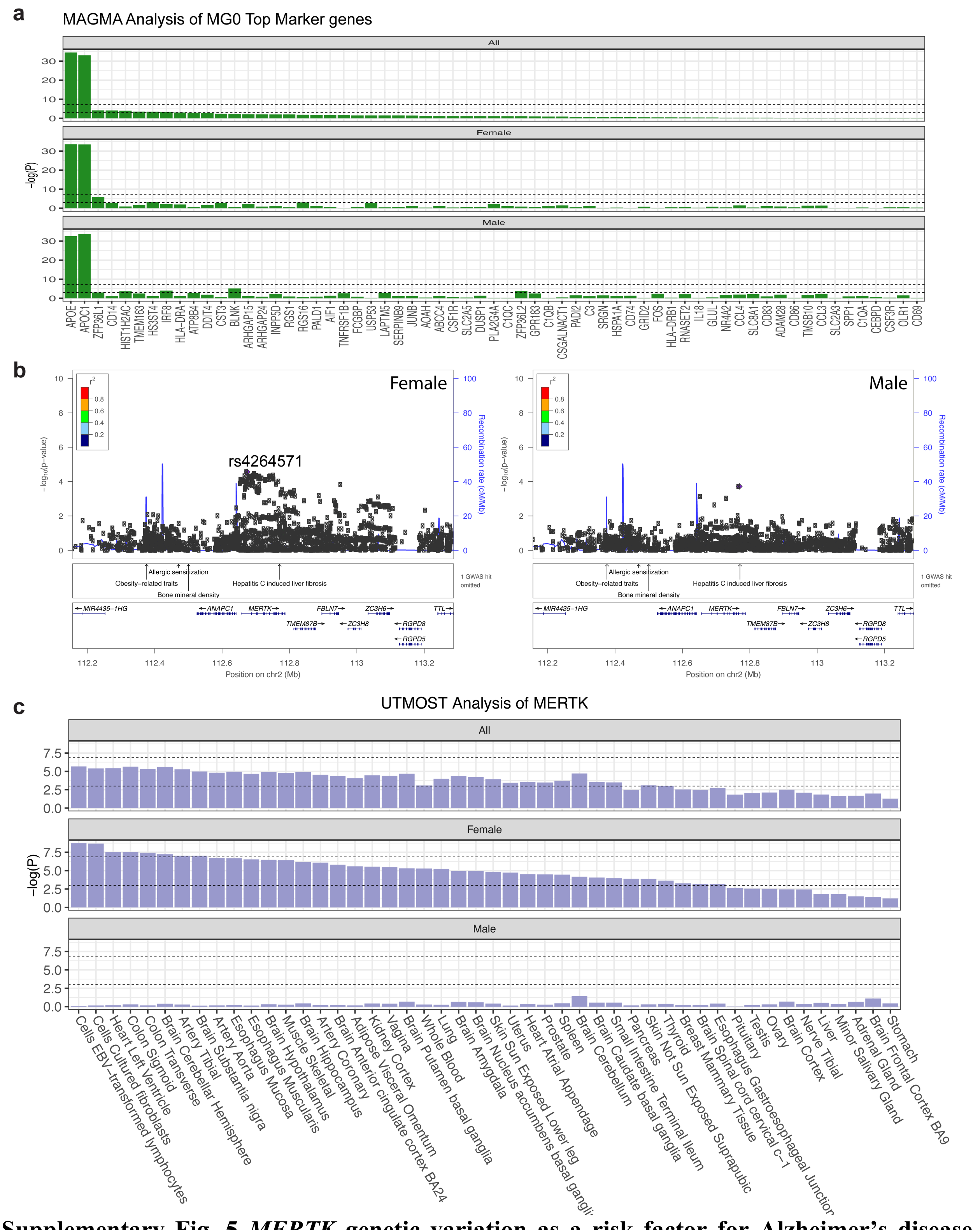


**Supplementary Fig. 5 *MERTK* genetic variation as a risk factor for Alzheimer's disease selectively in females. a** Associations between AD and female-specific microglial subcluster MG0 marker gene sets using MAGMA analysis based on ADGC GWAS dataset. The lower dashed line represents a p-value of 0.05 and the upper dashed line indicates a p-value threshold after

Bonferroni correction. **b** LocusZoom plots of *MERTK* in ADGC for either female (left) or male (right) GWAS dataset. **c** UTMOST tissue-specific associations between *MERTK* and AD in ADGC population using GTEx v8 data (48 tissues in total), the lower dashed line represents a p-value of 0.05 and the upper dashed line indicates a p-value threshold after Bonferroni control.

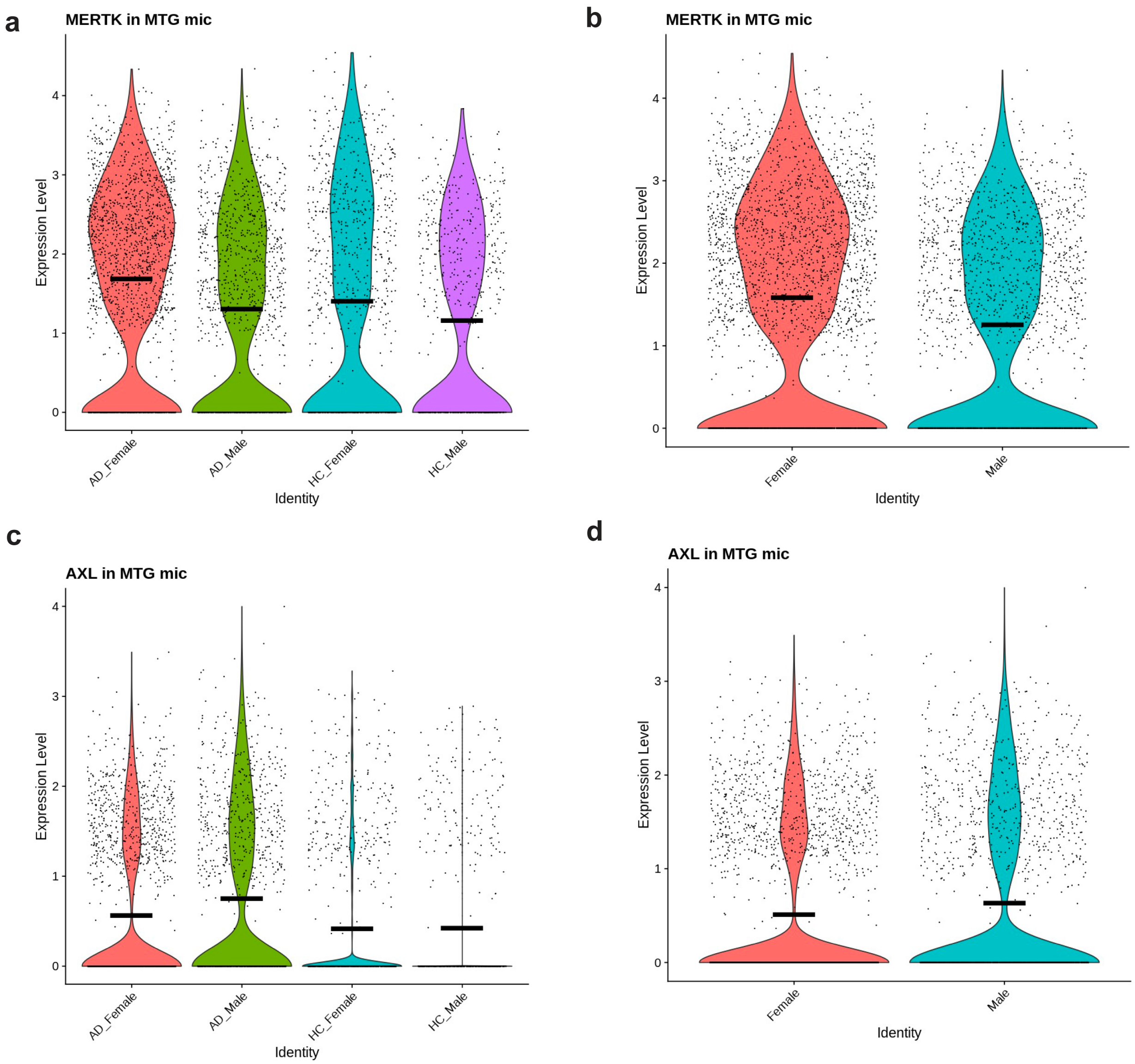


**Supplementary Fig. 6 The expression of *MERTK* and *AXL* in the microglia of human AD brain middle temporal gyrus. a** Violin plot of *MERTK* expression levels in the microglia of AD female, AD male, healthy control female, and healthy control male brains. **b** Violin plot of *MERTK* expression levels in the microglia of either female or male brains. **c** Violin plot of *AXL* expression levels in the microglia of AD female, AD male, healthy control female, and healthy control male brains. **b** Violin plot of *AXL* expression levels in the microglia of either female or male brains.
