## Supplementary Tables 1-6 for "Single-cell transcriptomic atlas of Alzheimer’s disease middle temporal gyrus reveals region, cell type and sex specificity of gene expression with novel genetic risk for MERTK in female"

**Supplementary Table 1. Sample Information and Sequencing QC**

| Group | Age | Sex | PMI | Diagnosis | Braak (Tau) | Cerad | CAA | TDP-43 (hippo-campal) | Median Genes per Cell | Median UMIs per Cell |
|---|---|---|---|---|---|---|---|---|---|---|
| HCF1 | 81-85 | F | 23 | No signif neuropathology | II/III | C0 | Neg | Neg | 1,600 | 3,174 |
| HCF2 | 71-75 | F | 14 | Multifocal demyelination, spinal cord | ND | C0 | Neg | Neg | 3,672 | 9,057 |
| HCF3 | 56-60 | F | 12 | No signif neuropathology | ND | C0 | Neg | Neg | 2,606 | 5,779 |
| HCF4 | 61-65 | F | 26 | Cerebrovascular Disease | 0 | C0 | Neg | Neg | 2,807 | 5,458 |
| ADF1 | 56-60 | F | 11 | AD, CAA | IV+ | C2 | CAA+ | Pos | 458 | 637 |
| ADF2 | 51-55 | F | 3 | AD, Lewy pathology (limbic, brainstem) | IV+ | C2 | Neg | Neg | 2,142 | 4,230 |
| ADF3 | 81-85 | F | 5 | AD, CAA(mild), Cerebrovascular Disease | VI | C3 | Focal | Pos | 2,127 | 3,817 |
| ADF4 | 86-90 | F | 9 | AD, CAA, Cerebrovascular Disease, VBI (multiple) | V/VI | C3 | CAA+ | Pos | 1,506 | 2,496 |
| HCM1 | 41-45 | M | 12 | Pituitary microadenomas (MEN1 syndrome) | ND | C0 | Neg | Neg | 1,969 | 3,764 |
| HCM2 | 46-50 | M | 12 | Cerebrovascular Disease | III | C0 | Neg | Neg | 2,883 | 6,682 |
| HCM3 | 76-80 | M | 16 | Cerebrovascular Disease | 0 | C0 | Neg | Neg | 2,368 | 4,932 |
| HCM4 | 41-45 | M | 16 | Cerebrovascular Disease, focal VBI | 0 | C0 | Neg | Neg | 1,670 | 2,971 |
| HCM5 | 66-70 | M | 35 | no signif neuropathology | 0 | C0 | Neg | Neg | 2,914 | 6,622 |
| ADM1 | 61-65 | M | 6 | AD, CAA(mild) | VI | C3 | Focal | Neg | 2,397 | 5,182 |
| ADM2 | 81-85 | M | 7 | AD, CAA(mild), Cerebrovascular Disease | IV | C3 | Focal | Neg | 3,121 | 7,139 |
| ADM3 | 71-75 | M | 13 | AD, CAA(mild) | V/VI | C3 | Focal | Pos | 1,187 | 1,907 |
| ADM4 | 71-75 | M | 6 | AD, CAA(mild), Cerebrovascular Disease | V/VI | C3 | Focal | ND | 1,661 | 3,074 |
| ADM5 | 81-85 | M | 5 | AD, CAA | V/VI | C3 | CAA+ | Pos | 1,671 | 3,356 |

ND: not determined; block not available

**Supplementary Table 2 Cellular DEGs**

**Astrocyte DEGS**

| | p_val | avg_log2FC | pct.1 | pct.2 | p_val_adj |
|---|---|---|---|---|---|
| HSP90AA1 | 9.31E-93 | 1.71098858 | 0.621 | 0.567 | 3.56E-88 |
| GFAP | 0 | 1.47666913 | 0.872 | 0.607 | 0 |
| NAMPT | 1.44E-16 | 1.42901563 | 0.23 | 0.187 | 5.49E-12 |
| PLCG2 | 8.24E-108 | 1.37662351 | 0.563 | 0.444 | 3.15E-103 |
| NHS | 7.30E-55 | 1.27708236 | 0.333 | 0.229 | 2.79E-50 |
| ELL2 | 9.84E-12 | 1.22666151 | 0.366 | 0.351 | 3.76E-07 |
| MIR4300HG | 3.07E-119 | 1.19931354 | 0.57 | 0.415 | 1.17E-114 |
| SLC39A11 | 4.72E-162 | 1.12457758 | 0.739 | 0.724 | 1.81E-157 |
| CD44 | 4.11E-82 | 1.11240327 | 0.363 | 0.2 | 1.57E-77 |
| HSPB1 | 3.55E-71 | 1.0998379 | 0.399 | 0.277 | 1.36E-66 |
| CRYAB | 9.96E-47 | 1.08628335 | 0.3 | 0.196 | 3.81E-42 |
| NEAT1 | 0 | 1.07032545 | 0.971 | 0.962 | 0 |
| EMP1 | 1.16E-11 | 1.05563019 | 0.199 | 0.161 | 4.42E-07 |
| PLCE1 | 2.27E-113 | 1.05513419 | 0.447 | 0.265 | 8.66E-109 |
| HSPH1 | 1.33E-49 | 1.05426823 | 0.244 | 0.141 | 5.08E-45 |
| MAN1C1 | 2.00E-11 | 1.04892277 | 0.325 | 0.304 | 7.64E-07 |
| HSPA1A | 1.83E-54 | 1.02436623 | 0.356 | 0.248 | 7.00E-50 |
| PFKP | 3.10E-98 | 1.02289514 | 0.589 | 0.506 | 1.19E-93 |
| DGKD | 2.53E-26 | 1.02162342 | 0.309 | 0.247 | 9.66E-22 |
| COLEC12 | 2.03E-91 | 1.01273602 | 0.567 | 0.434 | 7.76E-87 |
| LINC00486 | 3.04E-28 | 1.00102073 | 0.47 | 0.46 | 1.16E-23 |
| RGS6 | 2.30E-28 | 0.99899343 | 0.289 | 0.211 | 8.78E-24 |
| COL21A1 | 1.51E-110 | 0.98838629 | 0.339 | 0.156 | 5.76E-106 |
| ATF3 | 2.00E-77 | 0.98414175 | 0.108 | 0.017 | 7.63E-73 |
| FBXO32 | 5.64E-22 | 0.97384717 | 0.167 | 0.108 | 2.16E-17 |
| AC104574.2 | 1.37E-56 | 0.95665738 | 0.23 | 0.117 | 5.25E-52 |
| TSHZ2 | 5.95E-70 | 0.94878744 | 0.207 | 0.085 | 2.27E-65 |
| CCDC85A | 2.95E-12 | 0.93576082 | 0.108 | 0.072 | 1.13E-07 |
| DNAJB1 | 1.15E-18 | 0.93005456 | 0.248 | 0.2 | 4.41E-14 |
| ITPKB | 8.93E-167 | 0.92905204 | 0.823 | 0.787 | 3.41E-162 |
| GLIS3 | 5.78E-145 | 0.90474514 | 0.892 | 0.896 | 2.21E-140 |
| TTN | 1.95E-29 | 0.89242748 | 0.138 | 0.072 | 7.44E-25 |
| PFKFB3 | 1.07E-47 | 0.8814879 | 0.621 | 0.587 | 4.09E-43 |
| HSPE1 | 1.50E-29 | 0.87451705 | 0.223 | 0.148 | 5.75E-25 |
| ZBTB16 | 7.09E-81 | 0.87364485 | 0.676 | 0.654 | 2.71E-76 |
| DCLK1 | 4.59E-15 | 0.86262257 | 0.277 | 0.231 | 1.75E-10 |
| SORBS1 | 5.40E-238 | 0.84873927 | 0.961 | 0.962 | 2.06E-233 |
| ANGPTL4 | 3.78E-19 | 0.82834661 | 0.451 | 0.423 | 1.45E-14 |
| MRTFA | 3.87E-37 | 0.82426404 | 0.436 | 0.366 | 1.48E-32 |
| ARHGEF3 | 8.94E-52 | 0.81687723 | 0.449 | 0.326 | 3.42E-47 |
| SAMD4A | 9.19E-155 | 0.81067836 | 0.823 | 0.722 | 3.51E-150 |
| SPP1 | 1.94E-49 | 0.80819829 | 0.144 | 0.055 | 7.42E-45 |

| | | | | | |
|---|---|---|---|---|---|
| IRS2 | 1.56E-23 | 0.80325768 | 0.501 | 0.489 | 5.95E-19 |
| RFTN1 | 9.39E-33 | 0.79935365 | 0.202 | 0.122 | 3.59E-28 |
| TPST1 | 3.12E-49 | 0.79477198 | 0.577 | 0.475 | 1.19E-44 |
| MAP1B | 1.28E-14 | 0.78536633 | 0.383 | 0.355 | 4.88E-10 |
| AL390755.1 | 1.10E-82 | 0.78190853 | 0.201 | 0.072 | 4.21E-78 |
| CP | 7.48E-76 | 0.77749996 | 0.112 | 0.02 | 2.86E-71 |
| TESC | 7.97E-37 | 0.77597554 | 0.102 | 0.037 | 3.05E-32 |
| PLEKHA5 | 1.89E-46 | 0.76986248 | 0.592 | 0.535 | 7.23E-42 |
| ST6GAL1 | 2.24E-45 | 0.76868705 | 0.262 | 0.157 | 8.58E-41 |
| SLC5A3 | 1.09E-55 | 0.76662307 | 0.269 | 0.154 | 4.17E-51 |
| UBC | 4.45E-09 | 0.76439186 | 0.372 | 0.379 | 0.00017019 |
| SPATA13 | 2.04E-39 | 0.7613306 | 0.482 | 0.405 | 7.78E-35 |
| ANGPT1 | 4.69E-84 | 0.75340284 | 0.236 | 0.092 | 1.79E-79 |
| ARID5B | 6.46E-11 | 0.75296219 | 0.358 | 0.346 | 2.47E-06 |
| AC119674.1 | 9.51E-62 | 0.75277628 | 0.214 | 0.098 | 3.63E-57 |
| LINC01094 | 3.10E-25 | 0.75014548 | 0.251 | 0.178 | 1.18E-20 |
| AL627171.2 | 1.17E-30 | 0.74323651 | 0.205 | 0.129 | 4.47E-26 |
| VCAN | 4.47E-56 | 0.74296431 | 0.344 | 0.214 | 1.71E-51 |
| AEBP1 | 4.20E-21 | 0.74035762 | 0.257 | 0.196 | 1.61E-16 |
| FKBP5 | 7.47E-47 | 0.73737156 | 0.525 | 0.46 | 2.85E-42 |
| ADAMTS9-AS | 1.06E-19 | 0.73391688 | 0.319 | 0.264 | 4.04E-15 |
| GEM | 2.95E-17 | 0.7179914 | 0.188 | 0.136 | 1.13E-12 |
| MIR29B2CHG | 1.33E-07 | 0.71685884 | 0.413 | 0.439 | 0.00507004 |
| RAMP1 | 4.88E-29 | 0.71418322 | 0.467 | 0.441 | 1.87E-24 |
| CEBPD | 5.56E-11 | 0.7121832 | 0.215 | 0.185 | 2.13E-06 |
| GNA12 | 2.35E-20 | 0.70411549 | 0.477 | 0.468 | 8.96E-16 |
| FOS | 1.40E-15 | 0.7008907 | 0.293 | 0.256 | 5.35E-11 |
| PLP1 | 8.20E-39 | 0.70015841 | 0.267 | 0.162 | 3.14E-34 |
| FAM20C | 3.40E-19 | 0.67414508 | 0.298 | 0.245 | 1.30E-14 |
| TMTC1 | 8.01E-15 | 0.67162519 | 0.48 | 0.48 | 3.06E-10 |
| SLC7A2 | 1.34E-27 | 0.66980165 | 0.467 | 0.413 | 5.13E-23 |
| ADAMTSL3 | 1.37E-64 | 0.66931924 | 0.172 | 0.061 | 5.25E-60 |
| CDH23 | 1.25E-29 | 0.66626572 | 0.281 | 0.202 | 4.78E-25 |
| MYRIP | 3.16E-23 | 0.66581751 | 0.21 | 0.144 | 1.21E-18 |
| SLC38A1 | 7.71E-24 | 0.66222666 | 0.192 | 0.122 | 2.95E-19 |
| LINC01088 | 7.76E-14 | 0.65538675 | 0.152 | 0.105 | 2.97E-09 |
| NAV2 | 2.42E-75 | 0.65424698 | 0.89 | 0.921 | 9.26E-71 |
| SLC20A2 | 7.75E-22 | 0.65153896 | 0.507 | 0.5 | 2.96E-17 |
| FYN | 4.54E-35 | 0.64980629 | 0.873 | 0.92 | 1.74E-30 |
| PRKCA | 6.06E-58 | 0.63921078 | 0.914 | 0.939 | 2.32E-53 |
| HSPB8 | 1.01E-11 | 0.63592354 | 0.241 | 0.203 | 3.86E-07 |
| AHNAK | 2.83E-18 | 0.61894468 | 0.462 | 0.447 | 1.08E-13 |
| GMPR | 2.28E-23 | 0.60475736 | 0.258 | 0.189 | 8.70E-19 |
| RNF19A | 1.43E-11 | 0.60122587 | 0.539 | 0.58 | 5.47E-07 |
| PDE4B | 3.25E-19 | 0.59975445 | 0.777 | 0.859 | 1.24E-14 |

| | | | | | |
|---|---|---|---|---|---|
| PTPRE | 4.16E-22 | 0.59526452 | 0.261 | 0.195 | 1.59E-17 |
| PRRX1 | 7.14E-38 | 0.59449084 | 0.171 | 0.086 | 2.73E-33 |
| MOB3B | 4.48E-30 | 0.59446031 | 0.21 | 0.131 | 1.71E-25 |
| ARHGEF10 | 1.46E-24 | 0.59349438 | 0.512 | 0.476 | 5.60E-20 |
| CNN3 | 1.28E-14 | 0.59248437 | 0.586 | 0.588 | 4.90E-10 |
| DIP2B | 1.47E-09 | 0.58411275 | 0.523 | 0.572 | 5.63E-05 |
| ST8SIA1 | 4.50E-26 | 0.58379954 | 0.326 | 0.255 | 1.72E-21 |
| COTL1 | 2.43E-07 | 0.57878495 | 0.218 | 0.195 | 0.00928192 |
| FAM107A | 3.39E-21 | 0.57799629 | 0.635 | 0.72 | 1.29E-16 |
| COBL | 2.95E-13 | 0.57689316 | 0.476 | 0.49 | 1.13E-08 |
| HILPDA | 1.93E-10 | 0.57387635 | 0.197 | 0.159 | 7.38E-06 |
| HAP1 | 6.74E-25 | 0.57186192 | 0.209 | 0.142 | 2.57E-20 |
| PARD3 | 3.53E-108 | 0.57138563 | 0.883 | 0.908 | 1.35E-103 |
| LHFPL6 | 1.22E-16 | 0.56907302 | 0.617 | 0.666 | 4.68E-12 |
| BAIAP2 | 1.16E-12 | 0.56065952 | 0.208 | 0.164 | 4.45E-08 |
| MAOB | 9.16E-12 | 0.55833123 | 0.448 | 0.435 | 3.50E-07 |
| COL4A2 | 3.93E-09 | 0.55074853 | 0.215 | 0.18 | 0.00015013 |
| SYNM | 1.65E-30 | 0.54731009 | 0.203 | 0.125 | 6.31E-26 |
| SRRM2 | 5.47E-23 | 0.54562197 | 0.555 | 0.592 | 2.09E-18 |
| STAT3 | 5.54E-12 | 0.54423817 | 0.469 | 0.471 | 2.12E-07 |
| TMEM14B | 6.09E-18 | 0.53564201 | 0.416 | 0.385 | 2.33E-13 |
| DGKG | 3.08E-26 | 0.53172222 | 0.685 | 0.769 | 1.18E-21 |
| KAZN | 4.98E-09 | 0.53069094 | 0.409 | 0.404 | 0.00019048 |
| AC005225.2 | 1.52E-21 | 0.53021795 | 0.296 | 0.231 | 5.82E-17 |
| MT-ATP8 | 1.20E-07 | 0.5262893 | 0.259 | 0.236 | 0.00460317 |
| NPNT | 4.82E-89 | 0.52472265 | 0.14 | 0.028 | 1.84E-84 |
| MIR100HG | 2.46E-48 | 0.51825682 | 0.695 | 0.637 | 9.40E-44 |
| SLC7A5 | 7.10E-11 | 0.51212145 | 0.115 | 0.08 | 2.71E-06 |
| AL157886.1 | 4.50E-32 | 0.51049876 | 0.107 | 0.044 | 1.72E-27 |
| DIP2C | 4.75E-11 | 0.50978668 | 0.697 | 0.763 | 1.81E-06 |
| TTLL7 | 2.17E-09 | 0.50882057 | 0.247 | 0.21 | 8.28E-05 |
| PCBP1-AS1 | 2.49E-11 | 0.5068729 | 0.377 | 0.36 | 9.52E-07 |
| SEPTIN9 | 2.52E-08 | 0.50326038 | 0.317 | 0.3 | 0.00096333 |
| ARHGAP39 | 1.16E-11 | 0.50008872 | 0.267 | 0.234 | 4.42E-07 |
| GALNT15 | 1.18E-15 | 0.49898401 | 0.145 | 0.095 | 4.50E-11 |
| LRMDA | 3.24E-20 | 0.49874009 | 0.397 | 0.347 | 1.24E-15 |
| ERBIN | 6.12E-66 | 0.49353189 | 0.77 | 0.796 | 2.34E-61 |
| NR4A2 | 2.60E-08 | 0.48766097 | 0.142 | 0.11 | 0.00099281 |
| TAFA5 | 3.03E-18 | 0.48693015 | 0.452 | 0.42 | 1.16E-13 |
| SLCO3A1 | 1.33E-10 | 0.48612535 | 0.194 | 0.153 | 5.10E-06 |
| LINC00472 | 6.25E-12 | 0.4817606 | 0.521 | 0.537 | 2.39E-07 |
| BAIAP3 | 2.78E-53 | 0.48054032 | 0.16 | 0.062 | 1.06E-48 |
| BMERB1 | 1.40E-07 | 0.47861202 | 0.288 | 0.266 | 0.00536774 |
| CFAP47 | 1.95E-07 | 0.4783964 | 0.195 | 0.165 | 0.0074348 |
| MAML2 | 2.60E-16 | 0.47409169 | 0.85 | 0.913 | 9.95E-12 |

| | | | | | |
|---|---|---|---|---|---|
| CCDC144A | 5.73E-14 | 0.47336708 | 0.347 | 0.311 | 2.19E-09 |
| MALAT1 | 2.21E-58 | 0.47008979 | 1 | 1 | 8.45E-54 |
| HIF3A | 8.93E-16 | 0.46345186 | 0.722 | 0.858 | 3.41E-11 |
| PHACTR3 | 3.89E-22 | 0.4615584 | 0.719 | 0.796 | 1.49E-17 |
| SLC38A2 | 1.30E-13 | 0.45986102 | 0.205 | 0.159 | 4.98E-09 |
| MRPS6 | 2.03E-09 | 0.45985331 | 0.243 | 0.212 | 7.75E-05 |
| AC093865.1 | 5.64E-13 | 0.45450838 | 0.319 | 0.274 | 2.15E-08 |
| PLXDC2 | 5.36E-09 | 0.4520695 | 0.452 | 0.448 | 0.00020478 |
| L3MBTL4 | 1.64E-17 | 0.45028575 | 0.202 | 0.141 | 6.26E-13 |
| KANSL1 | 3.58E-20 | 0.44701373 | 0.643 | 0.716 | 1.37E-15 |
| ZFY | 9.44E-16 | 0.44611602 | 0.132 | 0.084 | 3.61E-11 |
| AOPEP | 9.46E-13 | 0.44430051 | 0.549 | 0.534 | 3.61E-08 |
| TENM4 | 3.62E-14 | 0.44377133 | 0.607 | 0.616 | 1.38E-09 |
| PNISR | 1.51E-24 | 0.44062965 | 0.657 | 0.759 | 5.77E-20 |
| LMNTD1 | 3.90E-19 | 0.43633952 | 0.171 | 0.111 | 1.49E-14 |
| S100A6 | 2.22E-11 | 0.43478323 | 0.136 | 0.099 | 8.47E-07 |
| TMEM91 | 2.12E-08 | 0.43424906 | 0.199 | 0.166 | 0.00081081 |
| POLR2J3 | 2.57E-10 | 0.4294939 | 0.34 | 0.317 | 9.82E-06 |
| TANC1 | 4.72E-09 | 0.42549614 | 0.377 | 0.353 | 0.00018058 |
| MAPK4 | 1.15E-08 | 0.42361679 | 0.682 | 0.757 | 0.00044124 |
| SLC9A9 | 3.90E-15 | 0.41647021 | 0.655 | 0.708 | 1.49E-10 |
| FAM182A | 2.46E-27 | 0.40023927 | 0.223 | 0.143 | 9.41E-23 |
| DAAM1 | 1.41E-09 | 0.39314659 | 0.64 | 0.727 | 5.39E-05 |
| STIP1 | 2.43E-09 | 0.39072034 | 0.128 | 0.094 | 9.28E-05 |
| MAPT | 1.53E-09 | 0.39031655 | 0.593 | 0.657 | 5.85E-05 |
| AC092902.4 | 3.01E-11 | 0.38639492 | 0.224 | 0.182 | 1.15E-06 |
| BCL6 | 3.38E-10 | 0.38598891 | 0.63 | 0.674 | 1.29E-05 |
| ASB3 | 1.18E-17 | 0.38322474 | 0.662 | 0.688 | 4.50E-13 |
| FLCN | 1.47E-19 | 0.37907498 | 0.159 | 0.1 | 5.62E-15 |
| ADCY2 | 1.51E-39 | 0.37901421 | 0.902 | 0.942 | 5.76E-35 |
| PRRT2 | 1.13E-09 | 0.371958 | 0.264 | 0.23 | 4.33E-05 |
| CREB5 | 3.51E-08 | 0.36874385 | 0.177 | 0.142 | 0.0013424 |
| CHD2 | 4.66E-10 | 0.36609566 | 0.65 | 0.72 | 1.78E-05 |
| SYNE2 | 5.71E-16 | 0.36480128 | 0.127 | 0.08 | 2.18E-11 |
| ZNF544 | 1.60E-07 | 0.36332168 | 0.236 | 0.208 | 0.00609769 |
| MAP4K4 | 6.92E-08 | 0.35352057 | 0.734 | 0.816 | 0.00264322 |
| WDR49 | 4.92E-08 | 0.3504496 | 0.261 | 0.223 | 0.00188112 |
| SEMA3D | 3.64E-20 | 0.34992676 | 0.154 | 0.095 | 1.39E-15 |
| TNRC6A | 2.07E-35 | 0.33658515 | 0.853 | 0.922 | 7.91E-31 |
| GALNT8 | 9.48E-12 | 0.33301914 | 0.105 | 0.068 | 3.63E-07 |
| ACTN2 | 1.11E-07 | 0.3325379 | 0.106 | 0.077 | 0.0042297 |
| TFRC | 1.73E-26 | 0.33166365 | 0.232 | 0.371 | 6.62E-22 |
| HVCN1 | 6.78E-09 | 0.32993656 | 0.112 | 0.08 | 0.00025901 |
| NEBL | 9.39E-36 | 0.3287913 | 0.895 | 0.935 | 3.59E-31 |
| PTCHD1-AS | 2.01E-08 | 0.324433 | 0.633 | 0.679 | 0.000768 |

| | | | | | |
|---|---|---|---|---|---|
| ZFAND5 | 1.06E-09 | 0.3154637 | 0.347 | 0.462 | 4.05E-05 |
| AC104389.5 | 3.91E-14 | 0.31425703 | 0.102 | 0.061 | 1.50E-09 |
| DPYSL2 | 4.81E-12 | 0.3127042 | 0.751 | 0.817 | 1.84E-07 |
| CHD1 | 1.78E-07 | 0.30467058 | 0.237 | 0.318 | 0.0067853 |
| WWOX | 4.24E-32 | 0.30457646 | 0.906 | 0.949 | 1.62E-27 |
| AC109927.1 | 5.93E-14 | 0.30133578 | 0.106 | 0.064 | 2.27E-09 |
| NDEL1 | 5.77E-09 | 0.29759662 | 0.176 | 0.247 | 0.00022061 |
| H3F3B | 3.61E-08 | 0.28902354 | 0.233 | 0.319 | 0.00138118 |
| PITPNC1 | 1.66E-19 | 0.28691954 | 0.957 | 0.974 | 6.34E-15 |
| SASH1 | 2.86E-08 | 0.28521124 | 0.8 | 0.879 | 0.00109135 |
| AMD1 | 1.36E-07 | 0.28459342 | 0.161 | 0.222 | 0.00520576 |
| STOM | 3.68E-10 | 0.26690741 | 0.221 | 0.308 | 1.41E-05 |
| C3 | 6.07E-24 | 0.26412778 | 0.101 | 0.047 | 2.32E-19 |
| SAMHD1 | 2.20E-07 | 0.26228482 | 0.116 | 0.087 | 0.00840088 |
| C7orf61 | 1.75E-09 | 0.25802022 | 0.1 | 0.068 | 6.67E-05 |
| OSBPL9 | 6.50E-10 | 0.25658197 | 0.371 | 0.499 | 2.49E-05 |
| ESYT2 | 1.59E-13 | 0.25644899 | 0.305 | 0.428 | 6.08E-09 |
| MYO6 | 2.33E-67 | -0.2500224 | 0.566 | 0.805 | 8.92E-63 |
| NHSL1 | 4.28E-50 | -0.2511541 | 0.625 | 0.837 | 1.63E-45 |
| HHIPL1 | 1.77E-65 | -0.2513637 | 0.138 | 0.297 | 6.77E-61 |
| KCNQ5 | 2.37E-32 | -0.2515719 | 0.136 | 0.244 | 9.07E-28 |
| AC013287.1 | 1.44E-26 | -0.2517186 | 0.043 | 0.103 | 5.51E-22 |
| AKAP7 | 4.30E-71 | -0.251969 | 0.251 | 0.455 | 1.65E-66 |
| AC008957.2 | 5.83E-72 | -0.2520294 | 0.149 | 0.325 | 2.23E-67 |
| ARHGEF12 | 3.18E-47 | -0.2522453 | 0.591 | 0.786 | 1.22E-42 |
| LINC01266 | 9.84E-32 | -0.2530456 | 0.154 | 0.261 | 3.76E-27 |
| TTN-AS1 | 3.16E-49 | -0.2535456 | 0.263 | 0.428 | 1.21E-44 |
| SLC30A10 | 7.64E-50 | -0.2536496 | 0.127 | 0.256 | 2.92E-45 |
| HES4 | 7.11E-55 | -0.2543173 | 0.074 | 0.188 | 2.72E-50 |
| DISP3 | 4.92E-72 | -0.2544765 | 0.066 | 0.203 | 1.88E-67 |
| CA12 | 5.16E-44 | -0.2546253 | 0.055 | 0.146 | 1.97E-39 |
| SNCAIP | 2.56E-54 | -0.2551298 | 0.18 | 0.336 | 9.78E-50 |
| LSM6 | 6.51E-76 | -0.2551636 | 0.086 | 0.235 | 2.49E-71 |
| CPNE8 | 6.31E-30 | -0.2554629 | 0.345 | 0.481 | 2.41E-25 |
| TENM3 | 1.19E-36 | -0.2555321 | 0.118 | 0.225 | 4.55E-32 |
| MYOM1 | 1.31E-55 | -0.2557296 | 0.187 | 0.344 | 5.02E-51 |
| ENTPD1-AS1 | 4.38E-62 | -0.2561258 | 0.382 | 0.614 | 1.68E-57 |
| NAALADL2 | 3.07E-60 | -0.256808 | 0.474 | 0.713 | 1.17E-55 |
| PPM1L | 9.41E-56 | -0.2571984 | 0.377 | 0.58 | 3.60E-51 |
| LRRC8A | 3.13E-80 | -0.2574426 | 0.268 | 0.491 | 1.20E-75 |
| RANBP17 | 2.15E-44 | -0.2576628 | 0.06 | 0.155 | 8.23E-40 |
| CACNB2 | 1.74E-48 | -0.2581491 | 0.736 | 0.894 | 6.64E-44 |
| SLITRK2 | 4.09E-68 | -0.2583298 | 0.133 | 0.293 | 1.56E-63 |
| LRRC37A3 | 2.29E-51 | -0.2584962 | 0.155 | 0.293 | 8.75E-47 |
| GLCCI1 | 6.25E-65 | -0.2585906 | 0.217 | 0.403 | 2.39E-60 |

| | | | | | |
|---|---|---|---|---|---|
| ARHGAP32 | 9.66E-70 | -0.2589181 | 0.519 | 0.768 | 3.69E-65 |
| NRXN3 | 1.79E-41 | -0.2601376 | 0.323 | 0.507 | 6.83E-37 |
| ANKAR | 6.85E-66 | -0.2601687 | 0.212 | 0.395 | 2.62E-61 |
| ARFGEF3 | 4.21E-70 | -0.2602721 | 0.19 | 0.375 | 1.61E-65 |
| FKBP9 | 1.65E-73 | -0.261098 | 0.091 | 0.238 | 6.31E-69 |
| ARID1B | 1.00E-57 | -0.2613382 | 0.643 | 0.849 | 3.84E-53 |
| R3HDM1 | 6.58E-71 | -0.2617893 | 0.361 | 0.594 | 2.51E-66 |
| AL590867.1 | 2.30E-67 | -0.2626346 | 0.039 | 0.15 | 8.81E-63 |
| SLC25A46 | 3.58E-72 | -0.2628055 | 0.137 | 0.305 | 1.37E-67 |
| LINC02427 | 1.34E-72 | -0.2629724 | 0.075 | 0.214 | 5.13E-68 |
| SLC6A11 | 4.36E-76 | -0.2636417 | 0.346 | 0.576 | 1.67E-71 |
| ZNF280D | 2.79E-75 | -0.2641783 | 0.401 | 0.656 | 1.07E-70 |
| NR3C2 | 8.73E-69 | -0.2643761 | 0.467 | 0.713 | 3.34E-64 |
| ACOX1 | 7.81E-82 | -0.2644997 | 0.121 | 0.295 | 2.99E-77 |
| ENHO | 1.04E-90 | -0.2645799 | 0.158 | 0.361 | 3.96E-86 |
| PPP2R2B | 1.70E-40 | -0.2648746 | 0.932 | 0.976 | 6.51E-36 |
| MOXD1 | 6.96E-44 | -0.2648766 | 0.115 | 0.233 | 2.66E-39 |
| PDZD2 | 1.11E-53 | -0.2651827 | 0.189 | 0.348 | 4.25E-49 |
| GRM5 | 8.28E-28 | -0.2654742 | 0.107 | 0.194 | 3.16E-23 |
| SOX2 | 1.24E-73 | -0.265885 | 0.354 | 0.593 | 4.75E-69 |
| MAPK8 | 1.18E-68 | -0.2659133 | 0.343 | 0.57 | 4.49E-64 |
| AL513217.1 | 1.46E-68 | -0.2662379 | 0.054 | 0.177 | 5.56E-64 |
| POU2F1 | 2.72E-69 | -0.266602 | 0.525 | 0.766 | 1.04E-64 |
| GRIA4 | 3.93E-40 | -0.2669037 | 0.147 | 0.269 | 1.50E-35 |
| DLGAP2 | 7.64E-36 | -0.26696 | 0.129 | 0.24 | 2.92E-31 |
| ZC2HC1A | 1.53E-68 | -0.2673498 | 0.226 | 0.419 | 5.84E-64 |
| MALRD1 | 2.28E-43 | -0.2680444 | 0.108 | 0.219 | 8.73E-39 |
| ASPH | 2.36E-52 | -0.2687583 | 0.635 | 0.853 | 9.00E-48 |
| ARHGEF9 | 7.56E-73 | -0.2688274 | 0.188 | 0.374 | 2.89E-68 |
| EFR3B | 1.61E-76 | -0.2692834 | 0.384 | 0.628 | 6.17E-72 |
| CYP4V2 | 3.61E-67 | -0.2694369 | 0.21 | 0.397 | 1.38E-62 |
| LHX2 | 2.05E-68 | -0.2696088 | 0.279 | 0.484 | 7.85E-64 |
| IL33 | 5.43E-72 | -0.269804 | 0.106 | 0.261 | 2.07E-67 |
| CASC2 | 7.05E-76 | -0.2701186 | 0.201 | 0.4 | 2.70E-71 |
| MAST4 | 1.18E-70 | -0.271045 | 0.735 | 0.887 | 4.52E-66 |
| RMST | 1.03E-46 | -0.2714243 | 0.163 | 0.299 | 3.95E-42 |
| MRO | 1.53E-60 | -0.272035 | 0.092 | 0.221 | 5.85E-56 |
| THSD7A | 2.17E-70 | -0.2720852 | 0.376 | 0.607 | 8.29E-66 |
| EXT1 | 2.05E-71 | -0.2723545 | 0.366 | 0.602 | 7.83E-67 |
| MT1X | 8.36E-51 | -0.2731833 | 0.222 | 0.384 | 3.19E-46 |
| AGBL4 | 3.84E-57 | -0.2742682 | 0.462 | 0.687 | 1.47E-52 |
| RBM26 | 1.38E-79 | -0.2744144 | 0.429 | 0.686 | 5.28E-75 |
| AL078602.1 | 6.32E-45 | -0.2750275 | 0.154 | 0.288 | 2.42E-40 |
| EPHA6 | 3.61E-48 | -0.2755117 | 0.384 | 0.582 | 1.38E-43 |
| NMD3 | 4.11E-84 | -0.2755396 | 0.234 | 0.455 | 1.57E-79 |

| | | | | | |
|---|---|---|---|---|---|
| AC013643.2 | 1.87E-68 | -0.2755437 | 0.096 | 0.24 | 7.16E-64 |
| FUT8 | 2.90E-68 | -0.275721 | 0.487 | 0.744 | 1.11E-63 |
| ETFDH | 9.24E-78 | -0.2759303 | 0.189 | 0.383 | 3.53E-73 |
| LINC02060 | 1.35E-75 | -0.2762838 | 0.104 | 0.262 | 5.16E-71 |
| NEK7 | 5.92E-84 | -0.2770409 | 0.257 | 0.482 | 2.26E-79 |
| SLC26A3 | 8.09E-62 | -0.277065 | 0.026 | 0.123 | 3.09E-57 |
| NALCN | 2.81E-80 | -0.2771872 | 0.313 | 0.555 | 1.07E-75 |
| PCCA | 6.96E-78 | -0.2773312 | 0.471 | 0.725 | 2.66E-73 |
| CDH18 | 1.23E-33 | -0.2774812 | 0.084 | 0.174 | 4.72E-29 |
| POU6F2 | 1.62E-54 | -0.2776364 | 0.156 | 0.308 | 6.21E-50 |
| PIK3IP1 | 1.64E-78 | -0.2794585 | 0.096 | 0.253 | 6.28E-74 |
| MED12L | 5.38E-56 | -0.2795114 | 0.645 | 0.852 | 2.06E-51 |
| C11orf65 | 3.12E-66 | -0.2798472 | 0.074 | 0.205 | 1.19E-61 |
| TBC1D4 | 1.21E-54 | -0.2800465 | 0.077 | 0.194 | 4.62E-50 |
| STK33 | 2.76E-80 | -0.2800969 | 0.406 | 0.667 | 1.05E-75 |
| PTPRM | 1.03E-80 | -0.2801642 | 0.461 | 0.724 | 3.96E-76 |
| ATP2C1 | 1.62E-87 | -0.280211 | 0.339 | 0.591 | 6.21E-83 |
| TUBB2A | 1.45E-70 | -0.2803803 | 0.091 | 0.237 | 5.54E-66 |
| CDK14 | 3.78E-89 | -0.2806887 | 0.349 | 0.611 | 1.44E-84 |
| SOBP | 1.17E-79 | -0.2809344 | 0.388 | 0.637 | 4.49E-75 |
| GOPC | 6.76E-64 | -0.2817461 | 0.157 | 0.319 | 2.58E-59 |
| AC079352.1 | 4.45E-37 | -0.2817702 | 0.191 | 0.319 | 1.70E-32 |
| ATL2 | 7.86E-89 | -0.2818558 | 0.218 | 0.436 | 3.01E-84 |
| LCNL1 | 2.90E-71 | -0.2820887 | 0.293 | 0.507 | 1.11E-66 |
| NNT | 4.63E-76 | -0.2823955 | 0.203 | 0.399 | 1.77E-71 |
| ST6GALNAC5 | 9.27E-40 | -0.2828913 | 0.137 | 0.255 | 3.54E-35 |
| NUP58 | 3.63E-94 | -0.2837807 | 0.153 | 0.357 | 1.39E-89 |
| ROBO3 | 4.89E-65 | -0.2838436 | 0.16 | 0.322 | 1.87E-60 |
| UFL1 | 2.94E-75 | -0.2838743 | 0.16 | 0.34 | 1.12E-70 |
| FOXG1 | 1.06E-78 | -0.2842394 | 0.19 | 0.39 | 4.06E-74 |
| PPM1K | 3.57E-70 | -0.2850722 | 0.305 | 0.52 | 1.36E-65 |
| PHKA1 | 1.65E-86 | -0.28558 | 0.181 | 0.383 | 6.32E-82 |
| AC107223.1 | 2.91E-50 | -0.286024 | 0.087 | 0.202 | 1.11E-45 |
| BIRC6 | 3.68E-81 | -0.2862198 | 0.528 | 0.78 | 1.41E-76 |
| AIG1 | 6.29E-87 | -0.2862641 | 0.33 | 0.581 | 2.41E-82 |
| CHRM3 | 9.72E-32 | -0.2863941 | 0.083 | 0.169 | 3.71E-27 |
| WRN | 1.93E-87 | -0.2864493 | 0.263 | 0.492 | 7.39E-83 |
| KCTD16 | 5.53E-43 | -0.287841 | 0.126 | 0.246 | 2.11E-38 |
| CAMSAP2 | 1.42E-62 | -0.2882167 | 0.396 | 0.601 | 5.43E-58 |
| SLF2 | 2.67E-88 | -0.2885643 | 0.209 | 0.424 | 1.02E-83 |
| SLC25A36 | 1.22E-93 | -0.2901004 | 0.225 | 0.454 | 4.67E-89 |
| OLFM1 | 2.92E-79 | -0.2902794 | 0.08 | 0.232 | 1.11E-74 |
| MMP28 | 4.81E-80 | -0.2903002 | 0.163 | 0.35 | 1.84E-75 |
| MBD5 | 5.82E-71 | -0.2903402 | 0.61 | 0.843 | 2.22E-66 |
| PTPRZ1 | 1.06E-22 | -0.2904596 | 0.769 | 0.897 | 4.06E-18 |

| | | | | | |
|---|---|---|---|---|---|
| DACH1 | 1.13E-37 | -0.2918154 | 0.077 | 0.168 | 4.32E-33 |
| PDE3B | 3.58E-58 | -0.2922173 | 0.367 | 0.573 | 1.37E-53 |
| VEGFA | 1.44E-63 | -0.2929138 | 0.178 | 0.348 | 5.52E-59 |
| PIGN | 1.81E-75 | -0.292933 | 0.199 | 0.392 | 6.93E-71 |
| SLC25A48 | 2.81E-73 | -0.2930655 | 0.239 | 0.443 | 1.07E-68 |
| AUTS2 | 1.18E-65 | -0.2932812 | 0.77 | 0.925 | 4.51E-61 |
| AC011287.1 | 1.52E-31 | -0.2941271 | 0.045 | 0.113 | 5.82E-27 |
| GRINA | 7.67E-91 | -0.2947875 | 0.162 | 0.366 | 2.93E-86 |
| PIP5K1B | 2.50E-66 | -0.2953001 | 0.078 | 0.211 | 9.57E-62 |
| MAP2K5 | 3.67E-87 | -0.2960291 | 0.378 | 0.635 | 1.40E-82 |
| EYA1 | 1.62E-74 | -0.2961163 | 0.306 | 0.528 | 6.20E-70 |
| SLC35C2 | 2.13E-92 | -0.2964605 | 0.123 | 0.31 | 8.16E-88 |
| RAP2C-AS1 | 7.50E-64 | -0.2966873 | 0.083 | 0.215 | 2.87E-59 |
| CCDC102B | 6.15E-60 | -0.2969242 | 0.096 | 0.227 | 2.35E-55 |
| CCDC7 | 7.96E-72 | -0.2974318 | 0.289 | 0.496 | 3.04E-67 |
| SETBP1 | 6.55E-69 | -0.2998961 | 0.197 | 0.38 | 2.50E-64 |
| GABRG1 | 8.58E-76 | -0.2999076 | 0.262 | 0.477 | 3.28E-71 |
| CAPZA1 | 1.47E-89 | -0.3008246 | 0.256 | 0.477 | 5.63E-85 |
| SRGAP3 | 7.46E-76 | -0.3016879 | 0.51 | 0.77 | 2.85E-71 |
| FBXL7 | 3.17E-53 | -0.3026315 | 0.821 | 0.924 | 1.21E-48 |
| PDE7B | 1.00E-74 | -0.3026467 | 0.529 | 0.77 | 3.83E-70 |
| HES1 | 1.66E-71 | -0.3027627 | 0.132 | 0.299 | 6.33E-67 |
| DMXL2 | 3.62E-93 | -0.3031276 | 0.212 | 0.437 | 1.38E-88 |
| PRDM5 | 1.90E-76 | -0.3034007 | 0.171 | 0.355 | 7.27E-72 |
| XKR6 | 5.69E-82 | -0.3041773 | 0.293 | 0.524 | 2.17E-77 |
| AP001528.1 | 8.64E-84 | -0.3041965 | 0.067 | 0.216 | 3.30E-79 |
| STON2 | 3.52E-69 | -0.3044469 | 0.582 | 0.809 | 1.34E-64 |
| DNM1L | 3.24E-100 | -0.3045863 | 0.24 | 0.481 | 1.24E-95 |
| TRPM3 | 1.03E-56 | -0.3046329 | 0.719 | 0.867 | 3.93E-52 |
| ALDOC | 8.30E-74 | -0.305051 | 0.197 | 0.386 | 3.17E-69 |
| DMD | 2.36E-62 | -0.3055792 | 0.403 | 0.621 | 9.01E-58 |
| HDAC8 | 2.79E-72 | -0.3057719 | 0.37 | 0.599 | 1.07E-67 |
| TMEM164 | 4.12E-53 | -0.3062365 | 0.349 | 0.538 | 1.57E-48 |
| BAALC-AS1 | 1.08E-17 | -0.3064479 | 0.199 | 0.283 | 4.14E-13 |
| TEX9 | 2.63E-57 | -0.3080472 | 0.134 | 0.277 | 1.01E-52 |
| PLCB1 | 1.90E-68 | -0.3085703 | 0.694 | 0.884 | 7.27E-64 |
| GOLGA8A | 2.82E-79 | -0.3093172 | 0.173 | 0.364 | 1.08E-74 |
| AC113414.1 | 6.60E-64 | -0.309777 | 0.159 | 0.322 | 2.52E-59 |
| BABAM2 | 3.82E-92 | -0.3104389 | 0.404 | 0.671 | 1.46E-87 |
| AL356599.1 | 1.33E-79 | -0.3105678 | 0.149 | 0.33 | 5.10E-75 |
| RBMS3 | 1.51E-35 | -0.3108897 | 0.694 | 0.81 | 5.77E-31 |
| SCML1 | 8.81E-99 | -0.3109171 | 0.154 | 0.364 | 3.37E-94 |
| CLYBL | 1.29E-86 | -0.3111161 | 0.257 | 0.484 | 4.92E-82 |
| CECR2 | 4.51E-86 | -0.3116077 | 0.317 | 0.558 | 1.72E-81 |
| LINC00499 | 1.87E-88 | -0.3119547 | 0.539 | 0.809 | 7.15E-84 |

| | | | | | |
|---|---|---|---|---|---|
| CRY2 | 3.76E-115 | -0.3120107 | 0.216 | 0.467 | 1.44E-110 |
| AC021231.1 | 6.04E-81 | -0.3123562 | 0.166 | 0.357 | 2.31E-76 |
| MID1 | 5.20E-85 | -0.3130373 | 0.261 | 0.486 | 1.99E-80 |
| RAPGEF3 | 2.86E-79 | -0.3134565 | 0.347 | 0.587 | 1.09E-74 |
| AC016831.7 | 1.54E-55 | -0.3140542 | 0.344 | 0.537 | 5.88E-51 |
| FAM85B | 1.45E-68 | -0.3152505 | 0.054 | 0.174 | 5.54E-64 |
| SOX9 | 1.53E-98 | -0.3158355 | 0.322 | 0.575 | 5.84E-94 |
| ASAP2 | 4.12E-82 | -0.3159907 | 0.442 | 0.69 | 1.57E-77 |
| SCFD2 | 3.51E-95 | -0.3160743 | 0.198 | 0.422 | 1.34E-90 |
| GNAQ | 1.37E-70 | -0.3165393 | 0.717 | 0.881 | 5.24E-66 |
| PHACTR1 | 5.60E-76 | -0.3168035 | 0.454 | 0.707 | 2.14E-71 |
| NRP1 | 1.52E-77 | -0.317015 | 0.304 | 0.529 | 5.80E-73 |
| F11-AS1 | 2.36E-89 | -0.3171563 | 0.357 | 0.619 | 9.02E-85 |
| PMP22 | 1.04E-90 | -0.3172951 | 0.287 | 0.522 | 3.97E-86 |
| MLLT3 | 1.28E-78 | -0.3174517 | 0.371 | 0.609 | 4.90E-74 |
| SRPX2 | 1.52E-83 | -0.3174791 | 0.034 | 0.16 | 5.82E-79 |
| LINGO1 | 3.17E-57 | -0.3179261 | 0.039 | 0.137 | 1.21E-52 |
| CHL1 | 1.95E-81 | -0.3189414 | 0.469 | 0.726 | 7.44E-77 |
| SYT1 | 1.04E-36 | -0.3190845 | 0.28 | 0.432 | 3.96E-32 |
| ZRANB3 | 1.32E-82 | -0.3194835 | 0.142 | 0.325 | 5.03E-78 |
| SMARCA2 | 8.24E-83 | -0.3195757 | 0.504 | 0.763 | 3.15E-78 |
| LRRC49 | 4.14E-82 | -0.3213284 | 0.162 | 0.351 | 1.58E-77 |
| ZNF98 | 1.36E-42 | -0.3214847 | 0.538 | 0.714 | 5.20E-38 |
| RNF182 | 3.04E-86 | -0.3224045 | 0.186 | 0.392 | 1.16E-81 |
| LINC02062 | 5.85E-87 | -0.3229673 | 0.261 | 0.488 | 2.23E-82 |
| FGGY | 7.74E-89 | -0.3233841 | 0.332 | 0.577 | 2.96E-84 |
| ATP1A2 | 1.79E-61 | -0.3250825 | 0.668 | 0.895 | 6.86E-57 |
| TTYH3 | 9.42E-86 | -0.3257615 | 0.167 | 0.362 | 3.60E-81 |
| CDC42BPA | 7.47E-75 | -0.3268244 | 0.585 | 0.824 | 2.85E-70 |
| EIF3E | 4.75E-81 | -0.3274043 | 0.29 | 0.517 | 1.82E-76 |
| SEMA4A | 8.48E-73 | -0.3282171 | 0.065 | 0.198 | 3.24E-68 |
| BBS2 | 4.89E-91 | -0.3286167 | 0.394 | 0.665 | 1.87E-86 |
| LINC02328 | 2.04E-68 | -0.3291839 | 0.137 | 0.297 | 7.78E-64 |
| SFPQ | 3.03E-89 | -0.3293241 | 0.413 | 0.67 | 1.16E-84 |
| RABGAP1 | 8.90E-92 | -0.3295908 | 0.397 | 0.657 | 3.40E-87 |
| MTF2 | 9.87E-97 | -0.3312201 | 0.157 | 0.363 | 3.77E-92 |
| CLASP2 | 1.34E-82 | -0.3321018 | 0.551 | 0.811 | 5.10E-78 |
| EFNA5 | 3.60E-87 | -0.3324891 | 0.42 | 0.685 | 1.38E-82 |
| HS3ST5 | 4.50E-62 | -0.3326647 | 0.188 | 0.354 | 1.72E-57 |
| USP24 | 7.55E-100 | -0.332677 | 0.34 | 0.599 | 2.88E-95 |
| BMP2K | 2.28E-88 | -0.3329654 | 0.491 | 0.747 | 8.71E-84 |
| ABCD2 | 1.04E-73 | -0.3330605 | 0.16 | 0.337 | 3.96E-69 |
| AC007402.1 | 3.19E-49 | -0.3351824 | 0.063 | 0.165 | 1.22E-44 |
| RHBDL3 | 9.51E-67 | -0.3355094 | 0.093 | 0.232 | 3.63E-62 |
| DLG2 | 4.54E-66 | -0.3361489 | 0.78 | 0.932 | 1.74E-61 |

| | | | | | |
|---|---|---|---|---|---|
| TXNIP | 5.82E-83 | -0.3365379 | 0.153 | 0.343 | 2.23E-78 |
| ANO5 | 1.58E-72 | -0.3365858 | 0.26 | 0.458 | 6.03E-68 |
| ID2 | 2.17E-84 | -0.3367919 | 0.21 | 0.421 | 8.31E-80 |
| RGS7 | 7.43E-64 | -0.3374516 | 0.368 | 0.584 | 2.84E-59 |
| PLEKHO1 | 2.06E-85 | -0.3377906 | 0.14 | 0.325 | 7.87E-81 |
| ABLIM3 | 4.13E-81 | -0.3384062 | 0.081 | 0.233 | 1.58E-76 |
| DDHD1 | 3.70E-90 | -0.3387226 | 0.208 | 0.424 | 1.42E-85 |
| LINC01184 | 2.35E-94 | -0.3390399 | 0.225 | 0.458 | 9.00E-90 |
| FAM198B-AS | 1.07E-78 | -0.3403949 | 0.252 | 0.467 | 4.11E-74 |
| SLC24A3 | 1.87E-69 | -0.3414113 | 0.291 | 0.495 | 7.15E-65 |
| ATRNL1 | 6.74E-43 | -0.341874 | 0.149 | 0.278 | 2.57E-38 |
| GTDC1 | 2.92E-89 | -0.3419337 | 0.254 | 0.486 | 1.11E-84 |
| MEF2C | 2.86E-76 | -0.3427878 | 0.315 | 0.539 | 1.09E-71 |
| CADM2 | 2.75E-114 | -0.3445182 | 0.858 | 0.97 | 1.05E-109 |
| AC007389.1 | 3.66E-59 | -0.3451199 | 0.337 | 0.523 | 1.40E-54 |
| AGL | 1.15E-83 | -0.3451394 | 0.184 | 0.378 | 4.40E-79 |
| CLDN10 | 1.11E-89 | -0.3458523 | 0.28 | 0.512 | 4.23E-85 |
| METTL15 | 2.36E-91 | -0.3463386 | 0.372 | 0.626 | 9.01E-87 |
| TMTC2 | 4.78E-32 | -0.3471111 | 0.627 | 0.789 | 1.83E-27 |
| GPD2 | 1.15E-89 | -0.3484763 | 0.394 | 0.648 | 4.38E-85 |
| TIMP3 | 3.28E-115 | -0.3494693 | 0.285 | 0.559 | 1.25E-110 |
| DDIT4 | 1.01E-54 | -0.3495876 | 0.163 | 0.309 | 3.87E-50 |
| RBFOX1 | 8.53E-23 | -0.3498148 | 0.444 | 0.604 | 3.26E-18 |
| WDPCP | 1.55E-97 | -0.3499294 | 0.323 | 0.58 | 5.93E-93 |
| AFF3 | 3.80E-63 | -0.3501931 | 0.1 | 0.239 | 1.45E-58 |
| ADGRL3 | 6.27E-91 | -0.3514446 | 0.813 | 0.942 | 2.40E-86 |
| SLC35F1 | 1.68E-81 | -0.3532969 | 0.531 | 0.782 | 6.43E-77 |
| PLAGL1 | 1.36E-84 | -0.3536426 | 0.061 | 0.205 | 5.20E-80 |
| FBXL17 | 1.44E-89 | -0.3542182 | 0.517 | 0.792 | 5.51E-85 |
| ELOVL2 | 5.24E-106 | -0.3543968 | 0.236 | 0.479 | 2.00E-101 |
| DECR1 | 7.66E-98 | -0.3560609 | 0.236 | 0.47 | 2.93E-93 |
| AL359232.1 | 1.91E-68 | -0.3560924 | 0.067 | 0.196 | 7.29E-64 |
| PTCH1 | 2.96E-70 | -0.3564589 | 0.215 | 0.398 | 1.13E-65 |
| WARS | 1.08E-53 | -0.3572884 | 0.084 | 0.204 | 4.13E-49 |
| CYP7B1 | 2.21E-88 | -0.3579746 | 0.179 | 0.384 | 8.44E-84 |
| MAGI2 | 2.01E-98 | -0.3590534 | 0.869 | 0.963 | 7.69E-94 |
| GJB6 | 1.52E-80 | -0.3596883 | 0.137 | 0.312 | 5.80E-76 |
| GPC1 | 1.60E-98 | -0.3601507 | 0.182 | 0.399 | 6.13E-94 |
| LSAMP | 2.45E-177 | -0.3602733 | 0.98 | 0.993 | 9.35E-173 |
| CYP46A1 | 1.63E-105 | -0.3612535 | 0.171 | 0.392 | 6.22E-101 |
| CHRDL1 | 2.19E-90 | -0.3619007 | 0.112 | 0.29 | 8.36E-86 |
| PHLPP1 | 2.03E-86 | -0.3622632 | 0.604 | 0.846 | 7.75E-82 |
| TRPC1 | 2.54E-110 | -0.3623696 | 0.271 | 0.524 | 9.72E-106 |
| AC105383.1 | 6.32E-85 | -0.3643886 | 0.153 | 0.339 | 2.42E-80 |
| PKN2-AS1 | 6.92E-65 | -0.3645229 | 0.091 | 0.226 | 2.65E-60 |

| | | | | | |
|---|---|---|---|---|---|
| NRBP2 | 3.65E-108 | -0.3648652 | 0.301 | 0.568 | 1.39E-103 |
| GFRA1 | 2.79E-38 | -0.3666003 | 0.123 | 0.231 | 1.07E-33 |
| AC025159.1 | 2.42E-62 | -0.3668322 | 0.172 | 0.336 | 9.23E-58 |
| TMEM108 | 8.89E-83 | -0.3675455 | 0.379 | 0.628 | 3.40E-78 |
| C4orf19 | 2.46E-91 | -0.3677995 | 0.258 | 0.485 | 9.41E-87 |
| AC087482.1 | 2.47E-97 | -0.3680681 | 0.087 | 0.263 | 9.44E-93 |
| AC073050.1 | 1.61E-73 | -0.3694519 | 0.548 | 0.792 | 6.17E-69 |
| MYO10 | 1.07E-98 | -0.3704594 | 0.371 | 0.643 | 4.08E-94 |
| LINGO2 | 4.08E-49 | -0.3712269 | 0.067 | 0.173 | 1.56E-44 |
| EPM2A | 2.88E-81 | -0.3714235 | 0.186 | 0.379 | 1.10E-76 |
| AF123462.1 | 3.99E-109 | -0.3721997 | 0.019 | 0.159 | 1.53E-104 |
| ADGRG1 | 8.75E-97 | -0.3729715 | 0.372 | 0.631 | 3.35E-92 |
| CXCL14 | 2.77E-90 | -0.3743697 | 0.149 | 0.342 | 1.06E-85 |
| ITFG1 | 1.35E-106 | -0.3746441 | 0.424 | 0.705 | 5.14E-102 |
| AC073475.1 | 7.67E-59 | -0.3749834 | 0.355 | 0.538 | 2.93E-54 |
| AC115622.1 | 1.11E-69 | -0.3756842 | 0.121 | 0.275 | 4.25E-65 |
| LGI1 | 1.19E-90 | -0.3769604 | 0.444 | 0.703 | 4.55E-86 |
| SESN3 | 9.61E-112 | -0.377566 | 0.282 | 0.551 | 3.67E-107 |
| RAB11FIP2 | 3.65E-109 | -0.3797962 | 0.284 | 0.543 | 1.39E-104 |
| NAV3 | 1.06E-95 | -0.3808947 | 0.785 | 0.951 | 4.04E-91 |
| SLC13A5 | 1.75E-106 | -0.3812037 | 0.077 | 0.256 | 6.70E-102 |
| BDNF-AS | 1.18E-94 | -0.3813291 | 0.186 | 0.397 | 4.51E-90 |
| CNTNAP3B | 9.32E-104 | -0.381716 | 0.291 | 0.548 | 3.56E-99 |
| ATP13A4 | 6.86E-94 | -0.3824677 | 0.558 | 0.834 | 2.62E-89 |
| UBAC2 | 4.49E-75 | -0.3824931 | 0.305 | 0.506 | 1.71E-70 |
| FAT3 | 5.42E-96 | -0.3831245 | 0.419 | 0.686 | 2.07E-91 |
| FAM182B | 2.16E-33 | -0.3832719 | 0.169 | 0.272 | 8.25E-29 |
| ZFP36L2 | 1.25E-85 | -0.3833331 | 0.211 | 0.42 | 4.78E-81 |
| KLF12 | 1.36E-93 | -0.3839522 | 0.377 | 0.635 | 5.20E-89 |
| GJA1 | 8.70E-96 | -0.3840643 | 0.556 | 0.809 | 3.33E-91 |
| ERC2 | 1.18E-61 | -0.3846725 | 0.139 | 0.292 | 4.53E-57 |
| APCDD1 | 2.94E-96 | -0.3848668 | 0.292 | 0.537 | 1.12E-91 |
| FGF14 | 2.44E-62 | -0.3849236 | 0.242 | 0.433 | 9.34E-58 |
| GRID2 | 7.71E-26 | -0.3851912 | 0.133 | 0.226 | 2.95E-21 |
| MCC | 1.55E-95 | -0.3858855 | 0.481 | 0.75 | 5.93E-91 |
| CD38 | 9.64E-105 | -0.3860103 | 0.096 | 0.286 | 3.69E-100 |
| SGIP1 | 5.47E-96 | -0.3864339 | 0.613 | 0.851 | 2.09E-91 |
| NEGR1 | 9.31E-78 | -0.3869044 | 0.291 | 0.511 | 3.56E-73 |
| TMEM241 | 1.27E-99 | -0.3871151 | 0.15 | 0.355 | 4.87E-95 |
| DLC1 | 9.37E-97 | -0.3873897 | 0.411 | 0.684 | 3.58E-92 |
| CADM1 | 8.65E-96 | -0.3891905 | 0.852 | 0.943 | 3.31E-91 |
| HEPN1 | 6.16E-80 | -0.3903766 | 0.199 | 0.395 | 2.35E-75 |
| CDH12 | 2.45E-46 | -0.3906968 | 0.069 | 0.171 | 9.38E-42 |
| LGR4 | 4.93E-95 | -0.3913406 | 0.449 | 0.696 | 1.88E-90 |
| HEPACAM | 5.26E-118 | -0.3920849 | 0.454 | 0.749 | 2.01E-113 |

| | | | | | |
|---|---|---|---|---|---|
| TSPOAP1 | 2.87E-97 | -0.3928364 | 0.198 | 0.417 | 1.10E-92 |
| LUC7L2 | 1.03E-91 | -0.3940789 | 0.348 | 0.574 | 3.93E-87 |
| NTRK2 | 7.93E-86 | -0.3957492 | 0.835 | 0.95 | 3.03E-81 |
| SHISA6 | 1.43E-53 | -0.3976543 | 0.129 | 0.264 | 5.45E-49 |
| CEP85L | 5.89E-105 | -0.3979466 | 0.441 | 0.714 | 2.25E-100 |
| KLHL32 | 7.87E-105 | -0.3986223 | 0.335 | 0.598 | 3.01E-100 |
| AC138627.1 | 5.93E-98 | -0.3992893 | 0.233 | 0.463 | 2.26E-93 |
| CDHR3 | 1.35E-43 | -0.4010631 | 0.526 | 0.705 | 5.16E-39 |
| DZIP3 | 1.22E-106 | -0.4014621 | 0.232 | 0.474 | 4.66E-102 |
| BRINP3 | 2.25E-101 | -0.4041008 | 0.353 | 0.616 | 8.58E-97 |
| FAM66D | 1.07E-120 | -0.405346 | 0.092 | 0.295 | 4.10E-116 |
| UNC5C | 6.33E-72 | -0.4055865 | 0.376 | 0.594 | 2.42E-67 |
| AASS | 1.38E-100 | -0.4060727 | 0.324 | 0.565 | 5.27E-96 |
| OSBPL6 | 1.38E-111 | -0.4067202 | 0.421 | 0.698 | 5.29E-107 |
| ZMAT1 | 3.09E-119 | -0.4070294 | 0.352 | 0.643 | 1.18E-114 |
| FAM189A2 | 1.74E-31 | -0.4072884 | 0.576 | 0.741 | 6.66E-27 |
| TMEM132B | 1.02E-119 | -0.4095625 | 0.352 | 0.646 | 3.91E-115 |
| ITGAV | 8.34E-84 | -0.4118784 | 0.37 | 0.594 | 3.19E-79 |
| GRIK2 | 2.71E-75 | -0.4127455 | 0.212 | 0.414 | 1.04E-70 |
| RAB34 | 8.25E-122 | -0.4133348 | 0.121 | 0.339 | 3.15E-117 |
| GMDS-DT | 2.99E-107 | -0.414079 | 0.435 | 0.717 | 1.14E-102 |
| SUMF1 | 3.22E-108 | -0.4142893 | 0.304 | 0.569 | 1.23E-103 |
| ATP10B | 1.21E-68 | -0.4151169 | 0.185 | 0.359 | 4.63E-64 |
| TPCN1 | 3.10E-87 | -0.41542 | 0.403 | 0.654 | 1.19E-82 |
| SLCO1C1 | 1.37E-110 | -0.4171835 | 0.407 | 0.691 | 5.23E-106 |
| STXBP6 | 7.79E-100 | -0.4178917 | 0.264 | 0.507 | 2.98E-95 |
| CCSER1 | 2.10E-58 | -0.4190567 | 0.119 | 0.261 | 8.03E-54 |
| AC053527.1 | 7.98E-102 | -0.4209417 | 0 | 0.108 | 3.05E-97 |
| PLPP3 | 4.57E-89 | -0.4221844 | 0.574 | 0.803 | 1.75E-84 |
| ROBO2 | 1.70E-49 | -0.4231698 | 0.187 | 0.345 | 6.50E-45 |
| DTNA | 1.37E-67 | -0.4293749 | 0.963 | 0.983 | 5.23E-63 |
| AC079760.1 | 1.23E-104 | -0.4329256 | 0.144 | 0.353 | 4.69E-100 |
| TNS3 | 2.87E-116 | -0.4338179 | 0.34 | 0.621 | 1.10E-111 |
| PCNX2 | 3.20E-111 | -0.4341975 | 0.203 | 0.441 | 1.23E-106 |
| RHOJ | 1.09E-118 | -0.4359428 | 0.307 | 0.582 | 4.16E-114 |
| FAM184A | 2.35E-112 | -0.4371815 | 0.479 | 0.757 | 8.98E-108 |
| NLGN4X | 4.14E-112 | -0.4404584 | 0.303 | 0.568 | 1.58E-107 |
| RFX3-AS1 | 3.15E-110 | -0.4408096 | 0.328 | 0.599 | 1.20E-105 |
| AC116366.3 | 2.11E-103 | -0.4450484 | 0 | 0.109 | 8.08E-99 |
| CSPG5 | 4.53E-93 | -0.4450694 | 0.136 | 0.324 | 1.73E-88 |
| NOL4 | 4.67E-92 | -0.4457679 | 0.357 | 0.606 | 1.78E-87 |
| NDRG2 | 7.50E-108 | -0.4491169 | 0.529 | 0.797 | 2.87E-103 |
| TENT2 | 1.37E-145 | -0.4493061 | 0.324 | 0.62 | 5.25E-141 |
| AIFM3 | 5.06E-110 | -0.4498957 | 0.193 | 0.424 | 1.93E-105 |
| ERBB4 | 1.55E-161 | -0.4532846 | 0.858 | 0.968 | 5.93E-157 |

| | | | | | |
|---|---|---|---|---|---|
| SEPT2 | 6.07E-109 | -0.4560663 | 0 | 0.115 | 2.32E-104 |
| NTM | 1.51E-150 | -0.4564284 | 0.93 | 0.982 | 5.78E-146 |
| NPAS3 | 6.87E-194 | -0.4567116 | 0.975 | 0.987 | 2.62E-189 |
| EPB41L2 | 1.27E-136 | -0.4593272 | 0.404 | 0.706 | 4.87E-132 |
| CSMD1 | 3.84E-55 | -0.459481 | 0.325 | 0.527 | 1.47E-50 |
| GPAM | 7.83E-147 | -0.4606513 | 0.187 | 0.458 | 2.99E-142 |
| ACSL6 | 2.47E-91 | -0.4630271 | 0.369 | 0.594 | 9.43E-87 |
| UNC79 | 2.93E-106 | -0.4632806 | 0.247 | 0.487 | 1.12E-101 |
| WDR17 | 1.04E-120 | -0.4643336 | 0.288 | 0.555 | 3.96E-116 |
| SHROOM3 | 3.50E-116 | -0.4665145 | 0.483 | 0.766 | 1.34E-111 |
| TMEM72-AS | 9.58E-131 | -0.4680384 | 0.124 | 0.353 | 3.66E-126 |
| LRRC4C | 3.62E-117 | -0.4698347 | 0.497 | 0.788 | 1.38E-112 |
| SPATA6 | 2.15E-130 | -0.4702162 | 0.331 | 0.619 | 8.22E-126 |
| CAMK4 | 4.61E-101 | -0.470624 | 0.101 | 0.284 | 1.76E-96 |
| INTU | 2.30E-119 | -0.4706617 | 0.419 | 0.706 | 8.80E-115 |
| FAM19A5 | 3.61E-106 | -0.4713649 | 0 | 0.112 | 1.38E-101 |
| ZFHX4 | 8.55E-128 | -0.4713683 | 0.498 | 0.789 | 3.27E-123 |
| CC2D2B | 3.63E-103 | -0.4714962 | 0.122 | 0.316 | 1.39E-98 |
| MPPED2 | 5.29E-129 | -0.473489 | 0.24 | 0.509 | 2.02E-124 |
| CAMK2G | 5.70E-86 | -0.4737735 | 0.579 | 0.827 | 2.18E-81 |
| PCDH7 | 1.32E-128 | -0.4743959 | 0.44 | 0.74 | 5.03E-124 |
| LINC02340 | 2.20E-146 | -0.4745788 | 0.119 | 0.363 | 8.41E-142 |
| ATP8A1 | 1.87E-126 | -0.4752256 | 0.369 | 0.655 | 7.14E-122 |
| HTRA1 | 1.11E-128 | -0.4776909 | 0.325 | 0.617 | 4.23E-124 |
| LINC01727 | 1.09E-101 | -0.4790612 | 0.129 | 0.329 | 4.17E-97 |
| STXBP5 | 4.53E-127 | -0.4791695 | 0.519 | 0.805 | 1.73E-122 |
| GARNL3 | 1.33E-121 | -0.4794977 | 0.311 | 0.579 | 5.09E-117 |
| AC068308.1 | 6.04E-66 | -0.4813877 | 0.136 | 0.289 | 2.31E-61 |
| ADAMTS17 | 2.24E-119 | -0.4819178 | 0.347 | 0.627 | 8.58E-115 |
| CDH10 | 1.07E-112 | -0.4822942 | 0.262 | 0.512 | 4.09E-108 |
| SEPT11 | 6.07E-109 | -0.4824304 | 0 | 0.115 | 2.32E-104 |
| AGT | 2.50E-145 | -0.4825412 | 0.313 | 0.61 | 9.55E-141 |
| FAM213A | 1.02E-114 | -0.4826743 | 0 | 0.121 | 3.89E-110 |
| UNC80 | 1.47E-123 | -0.4855246 | 0.129 | 0.352 | 5.63E-119 |
| GOLGA8B | 1.13E-144 | -0.4890656 | 0.165 | 0.424 | 4.30E-140 |
| C2orf72 | 8.11E-130 | -0.4905505 | 0.296 | 0.581 | 3.10E-125 |
| RORB | 1.16E-132 | -0.4907803 | 0.448 | 0.747 | 4.42E-128 |
| COL5A3 | 3.24E-90 | -0.4913134 | 0.534 | 0.801 | 1.24E-85 |
| BSG | 2.65E-140 | -0.492639 | 0.284 | 0.569 | 1.01E-135 |
| CKB | 1.07E-115 | -0.4945095 | 0.533 | 0.83 | 4.11E-111 |
| HPSE2 | 1.88E-116 | -0.4962178 | 0.438 | 0.726 | 7.18E-112 |
| LRRC3B | 9.22E-89 | -0.496692 | 0.414 | 0.648 | 3.53E-84 |
| ADGRB3 | 7.74E-162 | -0.4973693 | 0.835 | 0.969 | 2.96E-157 |
| ME1 | 5.33E-131 | -0.4993372 | 0.253 | 0.523 | 2.04E-126 |
| SFXN5 | 9.55E-95 | -0.5066445 | 0.693 | 0.887 | 3.65E-90 |

| | | | | | |
|---|---|---|---|---|---|
| LINC00982 | 3.70E-107 | -0.5080073 | 0 | 0.113 | 1.41E-102 |
| GNA14 | 4.00E-98 | -0.5103634 | 0.609 | 0.817 | 1.53E-93 |
| ZNF518A | 1.56E-149 | -0.5129117 | 0.363 | 0.669 | 5.95E-145 |
| LMO3 | 4.03E-130 | -0.5173883 | 0.376 | 0.68 | 1.54E-125 |
| LINC01376 | 4.25E-135 | -0.5174322 | 0.161 | 0.408 | 1.63E-130 |
| EDNRB | 1.10E-143 | -0.5182668 | 0.162 | 0.419 | 4.21E-139 |
| HSD17B12 | 4.59E-134 | -0.5192748 | 0.136 | 0.372 | 1.76E-129 |
| ADGRB1 | 2.67E-129 | -0.5212796 | 0.144 | 0.38 | 1.02E-124 |
| CD47 | 4.70E-174 | -0.5213099 | 0.258 | 0.575 | 1.80E-169 |
| VAV3 | 1.20E-133 | -0.522638 | 0.324 | 0.615 | 4.58E-129 |
| NDRG4 | 1.56E-203 | -0.5227654 | 0.125 | 0.431 | 5.96E-199 |
| AC098829.1 | 1.84E-95 | -0.5261741 | 0.197 | 0.411 | 7.05E-91 |
| DHFR | 1.21E-67 | -0.5296256 | 0.046 | 0.16 | 4.62E-63 |
| SMURF2 | 7.84E-121 | -0.5320576 | 0.322 | 0.584 | 3.00E-116 |
| TARBP1 | 2.76E-163 | -0.5347868 | 0.187 | 0.474 | 1.06E-158 |
| MCF2L2 | 4.68E-141 | -0.5382386 | 0.143 | 0.391 | 1.79E-136 |
| AC104596.1 | 4.13E-141 | -0.5412223 | 0.318 | 0.609 | 1.58E-136 |
| BCAN | 3.80E-140 | -0.5439869 | 0.22 | 0.49 | 1.45E-135 |
| GLI3 | 2.94E-150 | -0.546329 | 0.469 | 0.778 | 1.12E-145 |
| TSPAN7 | 3.15E-132 | -0.546932 | 0.362 | 0.639 | 1.20E-127 |
| AC097103.2 | 1.60E-118 | -0.5493931 | 0 | 0.125 | 6.10E-114 |
| EPHB1 | 2.49E-155 | -0.5497439 | 0.418 | 0.73 | 9.50E-151 |
| DPH6 | 8.91E-152 | -0.5512669 | 0.297 | 0.6 | 3.40E-147 |
| PPP1R3C | 2.35E-133 | -0.553224 | 0.145 | 0.393 | 8.99E-129 |
| TTYH1 | 1.14E-148 | -0.5534218 | 0.489 | 0.801 | 4.35E-144 |
| ITPR2 | 3.45E-122 | -0.5549245 | 0.75 | 0.922 | 1.32E-117 |
| PDE4D | 4.85E-123 | -0.5589224 | 0.674 | 0.877 | 1.85E-118 |
| DCHS2 | 4.06E-130 | -0.5620565 | 0.162 | 0.405 | 1.55E-125 |
| PTN | 3.36E-167 | -0.563065 | 0.422 | 0.741 | 1.28E-162 |
| GPHN | 1.85E-161 | -0.5635857 | 0.649 | 0.894 | 7.08E-157 |
| RALGAPA2 | 2.70E-116 | -0.5664895 | 0.501 | 0.752 | 1.03E-111 |
| ADGRV1 | 3.31E-226 | -0.5685505 | 0.912 | 0.979 | 1.26E-221 |
| C9orf3 | 3.95E-120 | -0.5718551 | 0 | 0.126 | 1.51E-115 |
| GRIN2C | 3.88E-167 | -0.5741874 | 0.306 | 0.621 | 1.48E-162 |
| SLC6A1 | 1.69E-178 | -0.5751739 | 0.426 | 0.758 | 6.46E-174 |
| FREM2 | 2.44E-148 | -0.5753701 | 0.123 | 0.368 | 9.31E-144 |
| FRMD4A | 1.16E-135 | -0.5803795 | 0.39 | 0.688 | 4.44E-131 |
| KCNQ3 | 2.53E-113 | -0.5825202 | 0.287 | 0.543 | 9.69E-109 |
| SPARCL1 | 2.28E-163 | -0.5831416 | 0.769 | 0.95 | 8.72E-159 |
| SLC1A4 | 9.74E-190 | -0.5843625 | 0.235 | 0.562 | 3.72E-185 |
| OBI1-AS1 | 1.88E-77 | -0.5851815 | 0.6 | 0.715 | 7.19E-73 |
| BRINP2 | 2.10E-168 | -0.5887169 | 0.24 | 0.542 | 8.02E-164 |
| DIO2 | 8.13E-176 | -0.5901209 | 0.104 | 0.367 | 3.11E-171 |
| TUBB2B | 1.48E-139 | -0.590453 | 0.248 | 0.522 | 5.64E-135 |
| TNIK | 6.22E-189 | -0.5935876 | 0.851 | 0.961 | 2.38E-184 |

| | | | | | |
|---|---|---|---|---|---|
| GLUL | 1.01E-159 | -0.5947685 | 0.623 | 0.864 | 3.88E-155 |
| SERPINE2 | 1.59E-166 | -0.5953578 | 0.418 | 0.731 | 6.08E-162 |
| SPON1 | 8.63E-161 | -0.597063 | 0.459 | 0.782 | 3.30E-156 |
| PTPRG | 8.45E-177 | -0.5972845 | 0.555 | 0.848 | 3.23E-172 |
| LRRTM4 | 1.41E-85 | -0.6031759 | 0.276 | 0.503 | 5.40E-81 |
| SGCD | 1.30E-168 | -0.6040821 | 0.523 | 0.822 | 4.96E-164 |
| SCG3 | 2.39E-165 | -0.6081567 | 0.228 | 0.528 | 9.14E-161 |
| GPM6A | 5.79E-255 | -0.6287943 | 0.924 | 0.986 | 2.21E-250 |
| MFGE8 | 3.16E-152 | -0.6335236 | 0.196 | 0.462 | 1.21E-147 |
| GPR37L1 | 3.21E-184 | -0.6403964 | 0.288 | 0.614 | 1.23E-179 |
| OAF | 2.04E-180 | -0.6489034 | 0.253 | 0.568 | 7.79E-176 |
| GRIA2 | 5.83E-184 | -0.6499275 | 0.435 | 0.775 | 2.23E-179 |
| MGAT4C | 7.49E-177 | -0.6501826 | 0.503 | 0.83 | 2.86E-172 |
| ENOX1 | 2.01E-173 | -0.6516545 | 0.309 | 0.631 | 7.68E-169 |
| LINC00854 | 9.97E-120 | -0.6530798 | 0 | 0.126 | 3.81E-115 |
| FAM155A | 1.20E-199 | -0.6543306 | 0.709 | 0.93 | 4.60E-195 |
| LINC02649 | 3.12E-49 | -0.6557658 | 0.35 | 0.484 | 1.19E-44 |
| MMD2 | 1.13E-156 | -0.6717202 | 0.344 | 0.656 | 4.34E-152 |
| AC125613.1 | 2.03E-168 | -0.6729254 | 0.162 | 0.439 | 7.75E-164 |
| MUC20-OT1 | 2.13E-138 | -0.680351 | 0 | 0.144 | 8.15E-134 |
| ENTPD1 | 2.62E-193 | -0.6816033 | 0.164 | 0.464 | 1.00E-188 |
| RERG | 2.35E-161 | -0.6848069 | 0.145 | 0.417 | 8.97E-157 |
| GABRB1 | 4.06E-199 | -0.690133 | 0.694 | 0.915 | 1.55E-194 |
| AL589740.1 | 5.46E-182 | -0.6907625 | 0.618 | 0.883 | 2.09E-177 |
| SPRED1 | 3.14E-188 | -0.7055311 | 0.352 | 0.666 | 1.20E-183 |
| ETNPPL | 1.86E-172 | -0.7090756 | 0.391 | 0.701 | 7.11E-168 |
| NCAN | 2.74E-194 | -0.7145746 | 0.37 | 0.711 | 1.05E-189 |
| AC091826.2 | 9.63E-212 | -0.7162662 | 0.273 | 0.633 | 3.68E-207 |
| LINC00461 | 1.68E-209 | -0.7168836 | 0.511 | 0.838 | 6.42E-205 |
| CIRBP | 7.51E-245 | -0.7233 | 0.333 | 0.714 | 2.87E-240 |
| FGFR3 | 4.73E-225 | -0.7304478 | 0.454 | 0.824 | 1.81E-220 |
| NRXN1 | 6.92E-303 | -0.7308196 | 0.833 | 0.979 | 2.64E-298 |
| ARHGAP24 | 7.54E-198 | -0.7368571 | 0.457 | 0.788 | 2.88E-193 |
| SEPT7 | 5.17E-146 | -0.7371885 | 0 | 0.152 | 1.98E-141 |
| DNAH7 | 1.30E-141 | -0.7403738 | 0.393 | 0.677 | 4.97E-137 |
| GABRA2 | 5.73E-198 | -0.7517073 | 0.436 | 0.755 | 2.19E-193 |
| APOE | 1.98E-168 | -0.7610331 | 0.489 | 0.788 | 7.56E-164 |
| POT1-AS1 | 4.99E-129 | -0.7640878 | 0.251 | 0.506 | 1.91E-124 |
| ESRRG | 7.87E-212 | -0.7651959 | 0.409 | 0.756 | 3.01E-207 |
| FAM19A1 | 1.49E-123 | -0.7975301 | 0 | 0.13 | 5.70E-119 |
| PAMR1 | 1.89E-220 | -0.8005839 | 0.477 | 0.812 | 7.23E-216 |
| C1orf61 | 1.50E-254 | -0.8030192 | 0.546 | 0.876 | 5.72E-250 |
| KCNIP4 | 4.43E-125 | -0.8059806 | 0.392 | 0.691 | 1.69E-120 |
| GLUD1 | 3.68E-242 | -0.8079904 | 0.486 | 0.815 | 1.41E-237 |
| GPC5 | 0 | -0.8084221 | 0.83 | 0.973 | 0 |

| | | | | | |
|---|---|---|---|---|---|
| PREX2 | 8.81E-270 | -0.8222854 | 0.676 | 0.905 | 3.37E-265 |
| F3 | 7.08E-150 | -0.8227284 | 0.407 | 0.696 | 2.71E-145 |
| GPR75-ASB3 | 4.41E-148 | -0.850783 | 0 | 0.154 | 1.69E-143 |
| PIK3C2A | 6.04E-275 | -0.8638603 | 0.353 | 0.728 | 2.31E-270 |
| ACSS1 | 1.82E-212 | -0.876386 | 0.465 | 0.795 | 6.94E-208 |
| HES5 | 5.83E-163 | -0.8927146 | 0.032 | 0.232 | 2.23E-158 |
| NKAIN3 | 1.10E-279 | -0.8958941 | 0.614 | 0.91 | 4.21E-275 |
| SLC1A2 | 0 | -0.901881 | 0.899 | 0.979 | 0 |
| SLC4A4 | 0 | -0.9452928 | 0.632 | 0.923 | 0 |
| CABLES1 | 5.39E-244 | -0.9469279 | 0.479 | 0.811 | 2.06E-239 |
| SLC1A3 | 0 | -0.9528921 | 0.908 | 0.976 | 0 |
| FTH1 | 3.41E-67 | -0.987561 | 0.485 | 0.678 | 1.30E-62 |
| ATP1B2 | 2.82E-291 | -1.0236134 | 0.494 | 0.818 | 1.08E-286 |
| MTSS1L | 3.10E-169 | -1.0619349 | 0 | 0.174 | 1.18E-164 |
| AC002429.2 | 5.87E-266 | -1.095861 | 0.284 | 0.668 | 2.24E-261 |
| ALDH1A1 | 4.22E-274 | -1.1983024 | 0.191 | 0.555 | 1.61E-269 |
| FP236383.1 | 2.60E-180 | -1.1985122 | 0 | 0.184 | 9.94E-176 |
| WIF1 | 0 | -1.3030214 | 0.113 | 0.503 | 0 |
| GRM3 | 0 | -1.3603965 | 0.329 | 0.747 | 0 |
| FP671120.1 | 7.27E-191 | -1.4022333 | 0 | 0.194 | 2.78E-186 |
| AC105402.3 | 6.47E-157 | -1.4619641 | 0.012 | 0.189 | 2.47E-152 |
| RNF219-AS1 | 2.46E-194 | -2.5272325 | 0 | 0.197 | 9.39E-190 |

**Microglial DEGs**

| | p_val | avg_log2FC | pct.1 | pct.2 | p_val_adj |
|---|---|---|---|---|---|
| MT-ND3 | 1.30E-220 | 1.66190287 | 0.752 | 0.492 | 4.97E-216 |
| DPYD | 0 | 1.65876038 | 0.715 | 0.32 | 0 |
| HSP90AA1 | 1.69E-114 | 1.62073811 | 0.537 | 0.298 | 6.46E-110 |
| HSPH1 | 1.84E-86 | 1.61604756 | 0.315 | 0.126 | 7.05E-82 |
| PTPRG | 1.49E-76 | 1.60382729 | 0.304 | 0.136 | 5.70E-72 |
| ACSL1 | 1.62E-252 | 1.5112087 | 0.66 | 0.292 | 6.19E-248 |
| AC131944.1 | 9.39E-92 | 1.49914741 | 0.311 | 0.124 | 3.59E-87 |
| SPP1 | 1.92E-108 | 1.37563653 | 0.735 | 0.588 | 7.34E-104 |
| MT-ATP6 | 5.08E-139 | 1.2829792 | 0.704 | 0.487 | 1.94E-134 |
| TPRG1 | 3.54E-110 | 1.28219344 | 0.245 | 0.052 | 1.35E-105 |
| MT-CYB | 5.68E-145 | 1.27531378 | 0.608 | 0.344 | 2.17E-140 |
| MT-CO2 | 3.81E-142 | 1.21841283 | 0.717 | 0.485 | 1.46E-137 |
| CD163 | 8.90E-91 | 1.20116402 | 0.319 | 0.123 | 3.40E-86 |
| MT-ND4 | 3.68E-107 | 1.12754252 | 0.655 | 0.453 | 1.41E-102 |
| MYO1E | 8.15E-107 | 1.12542203 | 0.475 | 0.246 | 3.11E-102 |
| P4HA1 | 3.52E-89 | 1.10261167 | 0.465 | 0.257 | 1.34E-84 |
| NHSL1 | 1.37E-47 | 1.0948398 | 0.44 | 0.31 | 5.23E-43 |
| HSPD1 | 4.30E-37 | 1.09305965 | 0.225 | 0.115 | 1.64E-32 |
| FMN1 | 4.33E-135 | 1.07566844 | 0.694 | 0.493 | 1.66E-130 |
| HSPA5 | 5.03E-73 | 1.06521723 | 0.273 | 0.109 | 1.92E-68 |
| MT-CO3 | 1.99E-103 | 1.05603546 | 0.65 | 0.444 | 7.61E-99 |
| PLCG2 | 3.29E-36 | 1.05032821 | 0.751 | 0.636 | 1.26E-31 |
| ZBTB16 | 2.22E-122 | 1.04824669 | 0.523 | 0.271 | 8.50E-118 |
| STARD13 | 4.48E-83 | 1.04506406 | 0.45 | 0.254 | 1.71E-78 |
| DNAJB1 | 9.12E-32 | 1.02083062 | 0.126 | 0.047 | 3.49E-27 |
| APOE | 3.30E-96 | 1.01206214 | 0.59 | 0.391 | 1.26E-91 |
| HSPB1 | 1.19E-46 | 0.98123575 | 0.161 | 0.055 | 4.54E-42 |
| MT-ND1 | 1.10E-88 | 0.97597583 | 0.501 | 0.29 | 4.21E-84 |
| HSPE1 | 2.94E-41 | 0.93903308 | 0.157 | 0.057 | 1.12E-36 |
| MTRNR2L12 | 4.35E-99 | 0.9206277 | 0.613 | 0.394 | 1.66E-94 |
| FOXP1 | 1.56E-97 | 0.86664558 | 0.611 | 0.4 | 5.95E-93 |
| MAFB | 3.02E-67 | 0.86549252 | 0.234 | 0.085 | 1.15E-62 |
| HSPA1A | 1.15E-44 | 0.84649402 | 0.476 | 0.338 | 4.39E-40 |
| MT-ND2 | 8.58E-67 | 0.81869069 | 0.499 | 0.315 | 3.28E-62 |
| PVT1 | 1.79E-68 | 0.79842 | 0.284 | 0.119 | 6.84E-64 |
| HSP90AB1 | 7.88E-63 | 0.78287068 | 0.366 | 0.194 | 3.01E-58 |
| AL035446.2 | 9.95E-67 | 0.78258414 | 0.128 | 0.017 | 3.80E-62 |
| CPM | 3.79E-33 | 0.75854473 | 0.365 | 0.25 | 1.45E-28 |
| HSP90B1 | 3.05E-42 | 0.75658098 | 0.346 | 0.218 | 1.17E-37 |
| SCIN | 9.17E-74 | 0.7507026 | 0.549 | 0.358 | 3.50E-69 |
| DOCK5 | 5.07E-73 | 0.73338358 | 0.45 | 0.256 | 1.94E-68 |
| IL15 | 2.23E-72 | 0.72949629 | 0.158 | 0.029 | 8.52E-68 |
| CHORDC1 | 1.64E-50 | 0.72381659 | 0.219 | 0.092 | 6.27E-46 |

| | | | | | |
|---|---|---|---|---|---|
| MS4A6A | 2.84E-84 | 0.7198203 | 0.502 | 0.287 | 1.08E-79 |
| AC007262.2 | 3.26E-43 | 0.71432176 | 0.277 | 0.148 | 1.25E-38 |
| KCNMA1 | 3.11E-46 | 0.71338887 | 0.683 | 0.587 | 1.19E-41 |
| NEAT1 | 1.77E-192 | 0.71093897 | 0.977 | 0.939 | 6.76E-188 |
| SHOC1 | 1.01E-50 | 0.70845177 | 0.173 | 0.059 | 3.87E-46 |
| IQGAP2 | 2.00E-50 | 0.70549818 | 0.176 | 0.06 | 7.65E-46 |
| AP001977.1 | 1.61E-22 | 0.70502312 | 0.114 | 0.052 | 6.15E-18 |
| VSIG4 | 1.49E-60 | 0.69708994 | 0.433 | 0.258 | 5.70E-56 |
| DDIT4 | 5.18E-39 | 0.67210772 | 0.2 | 0.091 | 1.98E-34 |
| CTSD | 7.93E-56 | 0.67086076 | 0.303 | 0.148 | 3.03E-51 |
| FNIP2 | 5.57E-38 | 0.67052275 | 0.497 | 0.371 | 2.13E-33 |
| GK | 7.08E-30 | 0.67021296 | 0.244 | 0.141 | 2.71E-25 |
| FLT1 | 7.89E-75 | 0.66690483 | 0.203 | 0.053 | 3.02E-70 |
| ZNF804A | 6.33E-35 | 0.65741245 | 0.175 | 0.078 | 2.42E-30 |
| UGCG | 9.94E-32 | 0.65397744 | 0.376 | 0.255 | 3.80E-27 |
| HSPA1B | 3.97E-35 | 0.65161277 | 0.308 | 0.188 | 1.52E-30 |
| FCGBP | 2.22E-27 | 0.64610781 | 0.263 | 0.163 | 8.49E-23 |
| HSPA8 | 1.66E-31 | 0.64586385 | 0.201 | 0.103 | 6.36E-27 |
| PRKCE | 7.29E-43 | 0.63926985 | 0.423 | 0.279 | 2.79E-38 |
| BCL2A1 | 5.64E-40 | 0.63598128 | 0.125 | 0.038 | 2.16E-35 |
| BST2 | 4.17E-58 | 0.63407911 | 0.203 | 0.071 | 1.59E-53 |
| ABHD3 | 9.23E-45 | 0.63077948 | 0.329 | 0.189 | 3.53E-40 |
| PTGES3 | 5.61E-40 | 0.61309224 | 0.323 | 0.189 | 2.14E-35 |
| APOC1 | 4.19E-39 | 0.61173665 | 0.239 | 0.121 | 1.60E-34 |
| DNAJB6 | 1.04E-33 | 0.60714489 | 0.369 | 0.242 | 3.97E-29 |
| DNAJC3 | 1.85E-39 | 0.60537044 | 0.333 | 0.203 | 7.08E-35 |
| FKBP5 | 3.70E-116 | 0.59420147 | 0.844 | 0.682 | 1.41E-111 |
| SLC7A5 | 1.87E-30 | 0.58681267 | 0.176 | 0.085 | 7.15E-26 |
| FRMD4B | 6.00E-66 | 0.58468502 | 0.645 | 0.483 | 2.29E-61 |
| MT-ND4L | 3.99E-39 | 0.58361747 | 0.305 | 0.174 | 1.52E-34 |
| FNDC3B | 4.50E-47 | 0.58208166 | 0.55 | 0.404 | 1.72E-42 |
| ANKRD22 | 3.24E-52 | 0.57778869 | 0.222 | 0.089 | 1.24E-47 |
| CD14 | 7.25E-16 | 0.56623165 | 0.336 | 0.262 | 2.77E-11 |
| DNAJA1 | 4.44E-42 | 0.56568538 | 0.186 | 0.076 | 1.70E-37 |
| AC106865.2 | 3.24E-42 | 0.56529634 | 0.198 | 0.086 | 1.24E-37 |
| ALCAM | 2.50E-45 | 0.56496896 | 0.458 | 0.302 | 9.57E-41 |
| GAS7 | 3.42E-42 | 0.56250693 | 0.377 | 0.234 | 1.31E-37 |
| TBC1D8 | 5.84E-42 | 0.55841066 | 0.358 | 0.212 | 2.23E-37 |
| DIRC3 | 1.07E-11 | 0.55480309 | 0.118 | 0.073 | 4.09E-07 |
| CD81 | 2.80E-33 | 0.54573249 | 0.412 | 0.291 | 1.07E-28 |
| PLXNC1 | 3.86E-40 | 0.53959186 | 0.14 | 0.047 | 1.48E-35 |
| LINC02397 | 1.49E-29 | 0.53476879 | 0.12 | 0.046 | 5.69E-25 |
| LITAF | 3.46E-29 | 0.53361836 | 0.213 | 0.116 | 1.32E-24 |
| NEK10 | 3.84E-19 | 0.52809707 | 0.168 | 0.099 | 1.47E-14 |
| PGK1 | 1.12E-22 | 0.5272825 | 0.266 | 0.174 | 4.26E-18 |

| | | | | | |
|---|---|---|---|---|---|
| HERPUD1 | 2.43E-24 | 0.52440803 | 0.336 | 0.235 | 9.31E-20 |
| AL034397.3 | 1.05E-18 | 0.51858861 | 0.35 | 0.266 | 4.00E-14 |
| HLA-DRA | 1.05E-23 | 0.51238419 | 0.437 | 0.338 | 4.02E-19 |
| CEBPB | 4.77E-27 | 0.5095957 | 0.312 | 0.206 | 1.82E-22 |
| SNAP23 | 2.46E-25 | 0.5081394 | 0.275 | 0.176 | 9.41E-21 |
| TRPS1 | 2.64E-32 | 0.50583423 | 0.455 | 0.331 | 1.01E-27 |
| RBM47 | 8.61E-49 | 0.50202861 | 0.73 | 0.626 | 3.29E-44 |
| LNCAROD | 1.86E-40 | 0.50185692 | 0.743 | 0.66 | 7.12E-36 |
| STIP1 | 3.10E-30 | 0.49461094 | 0.142 | 0.06 | 1.18E-25 |
| MT-CO1 | 1.29E-50 | 0.49392858 | 0.607 | 0.447 | 4.94E-46 |
| SH3PXD2B | 7.53E-36 | 0.49302131 | 0.192 | 0.088 | 2.88E-31 |
| PPARG | 9.71E-30 | 0.49283919 | 0.247 | 0.142 | 3.71E-25 |
| NAMPT | 3.62E-22 | 0.48875497 | 0.212 | 0.128 | 1.39E-17 |
| ARID5B | 4.25E-25 | 0.48852503 | 0.342 | 0.237 | 1.63E-20 |
| SERPINA1 | 1.63E-36 | 0.48572088 | 0.157 | 0.063 | 6.25E-32 |
| ELL2 | 3.35E-53 | 0.48551387 | 0.653 | 0.492 | 1.28E-48 |
| CLEC2B | 2.96E-29 | 0.47694424 | 0.103 | 0.035 | 1.13E-24 |
| ATP1B3 | 2.72E-22 | 0.47198821 | 0.224 | 0.136 | 1.04E-17 |
| IQGAP1 | 7.08E-42 | 0.47047816 | 0.237 | 0.112 | 2.71E-37 |
| TLR5 | 1.46E-27 | 0.46746246 | 0.337 | 0.228 | 5.58E-23 |
| MITF | 1.80E-22 | 0.46475273 | 0.339 | 0.24 | 6.87E-18 |
| MAT2A | 5.02E-23 | 0.46456728 | 0.312 | 0.213 | 1.92E-18 |
| FMNL2 | 1.69E-61 | 0.46339153 | 0.821 | 0.717 | 6.47E-57 |
| ANAPC16 | 1.06E-31 | 0.46149103 | 0.331 | 0.209 | 4.06E-27 |
| CALR | 8.05E-23 | 0.46140948 | 0.219 | 0.134 | 3.08E-18 |
| MGAT1 | 8.65E-20 | 0.45952999 | 0.281 | 0.192 | 3.31E-15 |
| SDCBP | 3.22E-18 | 0.45903087 | 0.257 | 0.175 | 1.23E-13 |
| TANC2 | 1.31E-21 | 0.45781984 | 0.543 | 0.463 | 5.02E-17 |
| PADI2 | 1.71E-18 | 0.45764987 | 0.465 | 0.371 | 6.55E-14 |
| PDIA4 | 2.12E-33 | 0.45602542 | 0.13 | 0.048 | 8.10E-29 |
| CALCOCO2 | 9.39E-31 | 0.45364676 | 0.184 | 0.09 | 3.59E-26 |
| GNA13 | 1.10E-14 | 0.44947357 | 0.332 | 0.252 | 4.19E-10 |
| LRRK2 | 3.93E-16 | 0.44840139 | 0.321 | 0.242 | 1.50E-11 |
| IL1B | 4.29E-10 | 0.44391953 | 0.104 | 0.063 | 1.64E-05 |
| FCGR2A | 4.04E-22 | 0.44338406 | 0.322 | 0.222 | 1.54E-17 |
| MT-ND5 | 1.21E-26 | 0.44333322 | 0.286 | 0.179 | 4.61E-22 |
| C1QC | 2.10E-22 | 0.44151754 | 0.419 | 0.323 | 8.02E-18 |
| AC011933.4 | 1.05E-22 | 0.43887275 | 0.15 | 0.077 | 4.02E-18 |
| NRP1 | 1.13E-25 | 0.43705973 | 0.265 | 0.165 | 4.32E-21 |
| RBPJ | 3.28E-24 | 0.43694783 | 0.471 | 0.368 | 1.25E-19 |
| HLA-DRB1 | 9.71E-22 | 0.43468283 | 0.398 | 0.303 | 3.71E-17 |
| CASP1 | 2.34E-27 | 0.43083905 | 0.17 | 0.084 | 8.96E-23 |
| IGF2R | 2.79E-29 | 0.42780198 | 0.332 | 0.217 | 1.07E-24 |
| CDKN1A | 1.73E-11 | 0.42499512 | 0.182 | 0.127 | 6.62E-07 |
| SH2D4A | 5.23E-28 | 0.42407943 | 0.106 | 0.038 | 2.00E-23 |

| | | | | | |
|---|---|---|---|---|---|
| MS4A4A | 6.11E-36 | 0.42339944 | 0.191 | 0.086 | 2.34E-31 |
| PLP1 | 8.27E-24 | 0.42208166 | 0.279 | 0.177 | 3.16E-19 |
| SLC38A2 | 5.22E-25 | 0.42193322 | 0.297 | 0.195 | 1.99E-20 |
| DUSP10 | 3.87E-28 | 0.41973377 | 0.131 | 0.054 | 1.48E-23 |
| THRB | 7.99E-11 | 0.41754263 | 0.173 | 0.122 | 3.05E-06 |
| IFI30 | 4.09E-25 | 0.41686101 | 0.123 | 0.053 | 1.57E-20 |
| SDF2L1 | 2.20E-38 | 0.41618459 | 0.104 | 0.026 | 8.43E-34 |
| BCL6 | 3.93E-22 | 0.41581024 | 0.314 | 0.218 | 1.50E-17 |
| STEAP3 | 6.48E-31 | 0.41493861 | 0.179 | 0.085 | 2.48E-26 |
| SLC5A3 | 1.64E-29 | 0.41468151 | 0.133 | 0.054 | 6.26E-25 |
| RANBP2 | 5.53E-12 | 0.41253375 | 0.342 | 0.271 | 2.12E-07 |
| SLC17A9 | 2.13E-37 | 0.4111868 | 0.121 | 0.037 | 8.15E-33 |
| SOD2 | 2.08E-18 | 0.41063438 | 0.211 | 0.133 | 7.94E-14 |
| DSE | 4.72E-24 | 0.41030961 | 0.377 | 0.27 | 1.80E-19 |
| HSPA6 | 1.21E-21 | 0.40999994 | 0.116 | 0.053 | 4.63E-17 |
| ZFP36 | 5.59E-15 | 0.40909651 | 0.247 | 0.177 | 2.14E-10 |
| LINC01684 | 1.31E-27 | 0.4086234 | 0.294 | 0.183 | 4.99E-23 |
| ARHGAP18 | 4.18E-25 | 0.4083524 | 0.298 | 0.193 | 1.60E-20 |
| KCTD12 | 1.89E-23 | 0.40542522 | 0.34 | 0.238 | 7.21E-19 |
| HMOX1 | 1.44E-11 | 0.40440573 | 0.141 | 0.09 | 5.51E-07 |
| SNHG15 | 6.90E-30 | 0.40361113 | 0.117 | 0.043 | 2.64E-25 |
| ZC3HAV1 | 1.82E-25 | 0.40337502 | 0.335 | 0.225 | 6.95E-21 |
| HLA-B | 1.78E-19 | 0.40291879 | 0.338 | 0.246 | 6.79E-15 |
| ADGRE2 | 1.39E-24 | 0.39959677 | 0.224 | 0.13 | 5.30E-20 |
| FAM107B | 2.62E-16 | 0.39773493 | 0.149 | 0.087 | 1.00E-11 |
| TOM1 | 2.21E-24 | 0.39241571 | 0.151 | 0.074 | 8.46E-20 |
| SND1 | 7.35E-25 | 0.39136624 | 0.57 | 0.47 | 2.81E-20 |
| SEM1 | 4.44E-18 | 0.39009209 | 0.19 | 0.117 | 1.70E-13 |
| DAB2 | 2.34E-25 | 0.38428451 | 0.173 | 0.089 | 8.95E-21 |
| B2M | 7.41E-16 | 0.38336267 | 0.476 | 0.403 | 2.83E-11 |
| CERNA2 | 1.09E-08 | 0.38329683 | 0.108 | 0.07 | 0.00041654 |
| FAM53B | 1.32E-22 | 0.38191678 | 0.408 | 0.3 | 5.05E-18 |
| AP003086.1 | 6.26E-19 | 0.38172817 | 0.225 | 0.145 | 2.39E-14 |
| CLEC5A | 1.55E-24 | 0.37827477 | 0.1 | 0.038 | 5.93E-20 |
| PDIA6 | 1.10E-19 | 0.37622719 | 0.19 | 0.114 | 4.19E-15 |
| SPATS2L | 1.23E-23 | 0.37592823 | 0.261 | 0.163 | 4.70E-19 |
| ELOVL5 | 1.96E-12 | 0.36962961 | 0.246 | 0.179 | 7.51E-08 |
| QDPR | 1.13E-13 | 0.36913888 | 0.145 | 0.09 | 4.32E-09 |
| KANSL1 | 2.36E-32 | 0.36905525 | 0.682 | 0.598 | 9.01E-28 |
| RBM14 | 3.56E-16 | 0.3674579 | 0.176 | 0.109 | 1.36E-11 |
| ERGIC1 | 3.96E-24 | 0.36661186 | 0.267 | 0.166 | 1.51E-19 |
| MT-ATP8 | 4.23E-18 | 0.36646112 | 0.344 | 0.252 | 1.62E-13 |
| ESR1 | 9.87E-20 | 0.36545757 | 0.134 | 0.068 | 3.77E-15 |
| GPATCH2L | 5.86E-20 | 0.36500802 | 0.387 | 0.29 | 2.24E-15 |
| FBXL5 | 9.32E-22 | 0.36334096 | 0.291 | 0.196 | 3.56E-17 |

| | | | | | |
|---|---|---|---|---|---|
| PRKY | 9.33E-20 | 0.36312257 | 0.107 | 0.048 | 3.57E-15 |
| NUFIP2 | 6.97E-21 | 0.36305465 | 0.418 | 0.317 | 2.67E-16 |
| C1orf162 | 8.55E-25 | 0.36275866 | 0.208 | 0.118 | 3.27E-20 |
| LAMP2 | 1.36E-11 | 0.36273776 | 0.238 | 0.178 | 5.19E-07 |
| SIPA1L1 | 5.09E-21 | 0.36206223 | 0.549 | 0.448 | 1.94E-16 |
| GPR65 | 4.47E-23 | 0.36123027 | 0.156 | 0.08 | 1.71E-18 |
| FNDC3A | 1.51E-18 | 0.3608946 | 0.469 | 0.381 | 5.78E-14 |
| KCNJ2 | 9.83E-29 | 0.36049058 | 0.113 | 0.042 | 3.76E-24 |
| MSN | 9.03E-15 | 0.35977867 | 0.382 | 0.298 | 3.45E-10 |
| ACER3 | 8.09E-20 | 0.3585734 | 0.479 | 0.39 | 3.09E-15 |
| EPSTI1 | 3.06E-20 | 0.35745917 | 0.335 | 0.234 | 1.17E-15 |
| MRPS6 | 1.21E-24 | 0.35443209 | 0.136 | 0.062 | 4.62E-20 |
| HCLS1 | 1.90E-16 | 0.35399033 | 0.49 | 0.417 | 7.25E-12 |
| SLA | 8.40E-21 | 0.35239983 | 0.462 | 0.362 | 3.21E-16 |
| FXR1 | 5.75E-17 | 0.35170472 | 0.287 | 0.203 | 2.20E-12 |
| IVNS1ABP | 9.62E-15 | 0.35121101 | 0.23 | 0.159 | 3.68E-10 |
| ASCC3 | 8.55E-20 | 0.3507927 | 0.467 | 0.373 | 3.27E-15 |
| PDGFC | 3.78E-09 | 0.35010949 | 0.301 | 0.245 | 0.00014432 |
| HSD17B4 | 7.88E-19 | 0.34904612 | 0.256 | 0.171 | 3.01E-14 |
| RGS16 | 1.04E-10 | 0.34860607 | 0.103 | 0.061 | 3.97E-06 |
| UBE2J1 | 1.07E-17 | 0.34814152 | 0.186 | 0.114 | 4.09E-13 |
| MS4A7 | 8.83E-18 | 0.34727531 | 0.321 | 0.233 | 3.37E-13 |
| MTHFS | 1.62E-23 | 0.34709118 | 0.163 | 0.084 | 6.20E-19 |
| DNAJB11 | 5.64E-22 | 0.34575101 | 0.123 | 0.057 | 2.16E-17 |
| RAB27A | 1.43E-19 | 0.34542904 | 0.164 | 0.093 | 5.46E-15 |
| CD63 | 3.47E-16 | 0.34505757 | 0.23 | 0.156 | 1.33E-11 |
| UBXN2B | 5.30E-19 | 0.34300413 | 0.305 | 0.215 | 2.03E-14 |
| E2F3 | 2.43E-18 | 0.34283995 | 0.352 | 0.259 | 9.29E-14 |
| MBNL2 | 6.22E-18 | 0.34020398 | 0.469 | 0.38 | 2.38E-13 |
| SESN1 | 2.73E-17 | 0.33484112 | 0.337 | 0.249 | 1.04E-12 |
| MTFR1 | 3.31E-21 | 0.33483554 | 0.153 | 0.081 | 1.27E-16 |
| MIR34AHG | 4.83E-14 | 0.33316759 | 0.251 | 0.179 | 1.85E-09 |
| DOCK1 | 6.10E-18 | 0.33133541 | 0.131 | 0.069 | 2.33E-13 |
| CADM1 | 2.61E-08 | 0.3301041 | 0.206 | 0.158 | 0.00099659 |
| MCL1 | 4.10E-12 | 0.32880192 | 0.166 | 0.11 | 1.57E-07 |
| UBE2D3 | 1.92E-19 | 0.32777223 | 0.469 | 0.374 | 7.32E-15 |
| UTRN | 1.60E-10 | 0.32695771 | 0.455 | 0.397 | 6.10E-06 |
| TMEM51 | 1.77E-14 | 0.32693225 | 0.257 | 0.183 | 6.76E-10 |
| TNFSF13B | 1.71E-19 | 0.32668786 | 0.199 | 0.119 | 6.53E-15 |
| SELENOS | 1.67E-26 | 0.32553356 | 0.118 | 0.047 | 6.39E-22 |
| PAPOLG | 8.35E-09 | 0.32469987 | 0.187 | 0.139 | 0.00031926 |
| GAB1 | 1.10E-08 | 0.32451567 | 0.355 | 0.302 | 0.00042035 |
| HP1BP3 | 2.49E-17 | 0.3244263 | 0.465 | 0.376 | 9.51E-13 |
| TYMP | 4.51E-18 | 0.32440331 | 0.256 | 0.171 | 1.72E-13 |
| CHSY3 | 3.47E-14 | 0.32417703 | 0.388 | 0.306 | 1.33E-09 |

| | | | | | |
|---|---|---|---|---|---|
| ASAP1 | 2.85E-15 | 0.32097613 | 0.426 | 0.341 | 1.09E-10 |
| JUN | 3.18E-09 | 0.32025268 | 0.106 | 0.067 | 0.00012171 |
| LDHA | 1.73E-14 | 0.31959764 | 0.126 | 0.072 | 6.63E-10 |
| CD164 | 1.03E-18 | 0.31754285 | 0.224 | 0.144 | 3.94E-14 |
| IER5L | 5.50E-20 | 0.31677932 | 0.104 | 0.046 | 2.10E-15 |
| GNS | 3.29E-18 | 0.31659999 | 0.182 | 0.109 | 1.26E-13 |
| SARAF | 4.79E-16 | 0.31652528 | 0.288 | 0.208 | 1.83E-11 |
| MCTP1 | 4.79E-10 | 0.31627523 | 0.167 | 0.117 | 1.83E-05 |
| C1QA | 6.32E-14 | 0.31563118 | 0.347 | 0.27 | 2.42E-09 |
| LINC00623 | 4.43E-17 | 0.31514129 | 0.163 | 0.096 | 1.69E-12 |
| CDK6 | 6.58E-21 | 0.31234837 | 0.466 | 0.364 | 2.52E-16 |
| AC008957.2 | 3.17E-19 | 0.31134813 | 0.118 | 0.057 | 1.21E-14 |
| PER1 | 1.05E-13 | 0.31029293 | 0.163 | 0.104 | 4.02E-09 |
| FOXN2 | 8.31E-18 | 0.31011783 | 0.241 | 0.159 | 3.18E-13 |
| USP15 | 2.94E-18 | 0.30939749 | 0.637 | 0.568 | 1.13E-13 |
| SEC24D | 6.03E-13 | 0.30890853 | 0.248 | 0.18 | 2.31E-08 |
| PDIA3 | 3.32E-20 | 0.30772501 | 0.236 | 0.15 | 1.27E-15 |
| RASSF4 | 7.12E-18 | 0.3075371 | 0.427 | 0.334 | 2.72E-13 |
| TUBA1A | 1.21E-13 | 0.30626061 | 0.147 | 0.09 | 4.63E-09 |
| PIK3IP1 | 8.47E-14 | 0.30603778 | 0.146 | 0.09 | 3.24E-09 |
| MYOF | 3.34E-15 | 0.30581849 | 0.386 | 0.301 | 1.28E-10 |
| LCP2 | 1.90E-20 | 0.30547387 | 0.439 | 0.331 | 7.28E-16 |
| GTDC1 | 4.53E-14 | 0.30510453 | 0.43 | 0.35 | 1.73E-09 |
| IDH1 | 2.58E-19 | 0.30463214 | 0.178 | 0.103 | 9.86E-15 |
| CAB39 | 1.01E-19 | 0.30231919 | 0.54 | 0.447 | 3.88E-15 |
| LAT2 | 2.89E-14 | 0.30190254 | 0.39 | 0.309 | 1.11E-09 |
| MAFG | 1.10E-20 | 0.30159365 | 0.177 | 0.099 | 4.19E-16 |
| SNHG5 | 1.34E-09 | 0.30069547 | 0.174 | 0.124 | 5.12E-05 |
| CPNE8 | 6.54E-22 | 0.30040165 | 0.101 | 0.041 | 2.50E-17 |
| ARL17B | 2.46E-24 | 0.29977652 | 0.187 | 0.098 | 9.42E-20 |
| BNIP3L | 3.87E-10 | 0.29674788 | 0.284 | 0.222 | 1.48E-05 |
| ATP6V1B2 | 9.66E-13 | 0.29658722 | 0.309 | 0.233 | 3.69E-08 |
| FPR1 | 3.47E-14 | 0.29549802 | 0.171 | 0.109 | 1.33E-09 |
| VPS9D1 | 1.09E-19 | 0.29263947 | 0.126 | 0.063 | 4.18E-15 |
| HIF1A | 2.14E-10 | 0.29223175 | 0.466 | 0.393 | 8.17E-06 |
| TSPO | 4.46E-19 | 0.29145658 | 0.121 | 0.059 | 1.70E-14 |
| CYTH1 | 1.36E-12 | 0.29109851 | 0.421 | 0.341 | 5.19E-08 |
| MTHFD2 | 8.99E-16 | 0.29081218 | 0.177 | 0.11 | 3.44E-11 |
| MBNL1 | 3.18E-20 | 0.29044073 | 0.806 | 0.77 | 1.22E-15 |
| TMED8 | 9.89E-14 | 0.29026871 | 0.199 | 0.134 | 3.78E-09 |
| ATG7 | 1.16E-14 | 0.29015588 | 0.623 | 0.559 | 4.42E-10 |
| MFSD1 | 2.90E-10 | 0.28854294 | 0.325 | 0.261 | 1.11E-05 |
| FLOT2 | 2.10E-15 | 0.28840561 | 0.201 | 0.131 | 8.02E-11 |
| HLA-DPA1 | 2.27E-09 | 0.28731638 | 0.257 | 0.2 | 8.66E-05 |
| TRAM1 | 2.33E-15 | 0.2869391 | 0.185 | 0.119 | 8.91E-11 |

| | | | | | |
|---|---|---|---|---|---|
| AC015871.1 | 2.87E-15 | 0.28631835 | 0.123 | 0.068 | 1.10E-10 |
| RASA1 | 1.56E-11 | 0.28623248 | 0.462 | 0.395 | 5.95E-07 |
| LCP1 | 3.66E-13 | 0.28583813 | 0.298 | 0.223 | 1.40E-08 |
| LAP3 | 1.41E-14 | 0.2839758 | 0.31 | 0.229 | 5.39E-10 |
| FAM20C | 3.22E-16 | 0.2837138 | 0.18 | 0.111 | 1.23E-11 |
| CHCHD7 | 8.92E-13 | 0.28356454 | 0.171 | 0.113 | 3.41E-08 |
| SPCS3 | 8.13E-14 | 0.28352815 | 0.202 | 0.137 | 3.11E-09 |
| SEC31A | 2.13E-12 | 0.28287814 | 0.307 | 0.235 | 8.13E-08 |
| AC012150.1 | 8.20E-09 | 0.28220429 | 0.333 | 0.273 | 0.00031349 |
| FCER1G | 4.85E-14 | 0.2818887 | 0.161 | 0.102 | 1.85E-09 |
| SKAP2 | 5.65E-20 | 0.28176436 | 0.635 | 0.554 | 2.16E-15 |
| GLRX | 6.45E-16 | 0.2811983 | 0.166 | 0.1 | 2.47E-11 |
| SLC1A3 | 3.97E-65 | 0.28050649 | 0.953 | 0.897 | 1.52E-60 |
| ZNF106 | 2.10E-11 | 0.28042044 | 0.34 | 0.271 | 8.01E-07 |
| BCL2 | 1.31E-11 | 0.27970099 | 0.509 | 0.448 | 5.02E-07 |
| BBX | 2.43E-10 | 0.27967638 | 0.417 | 0.351 | 9.29E-06 |
| TBC1D5 | 1.75E-17 | 0.27834115 | 0.727 | 0.687 | 6.69E-13 |
| TRIM14 | 3.10E-15 | 0.27780153 | 0.193 | 0.124 | 1.18E-10 |
| CCNH | 1.12E-12 | 0.27769362 | 0.499 | 0.422 | 4.27E-08 |
| COPA | 7.65E-11 | 0.27748807 | 0.337 | 0.27 | 2.92E-06 |
| SNHG25 | 7.62E-11 | 0.27637356 | 0.255 | 0.192 | 2.91E-06 |
| EAF2 | 1.35E-18 | 0.27605577 | 0.181 | 0.106 | 5.16E-14 |
| CKLF | 3.52E-12 | 0.27491712 | 0.235 | 0.169 | 1.34E-07 |
| CAPG | 3.75E-14 | 0.27463874 | 0.101 | 0.053 | 1.43E-09 |
| C1QB | 5.66E-12 | 0.27444041 | 0.498 | 0.437 | 2.16E-07 |
| BANP | 2.71E-12 | 0.27296474 | 0.241 | 0.173 | 1.03E-07 |
| SCARB2 | 2.80E-14 | 0.27294798 | 0.271 | 0.195 | 1.07E-09 |
| IRAK3 | 1.40E-11 | 0.27283266 | 0.471 | 0.403 | 5.36E-07 |
| TMEM50B | 2.31E-13 | 0.2726746 | 0.154 | 0.097 | 8.84E-09 |
| YWHAG | 1.61E-13 | 0.27261002 | 0.226 | 0.157 | 6.16E-09 |
| H3F3B | 2.71E-09 | 0.27226061 | 0.225 | 0.17 | 0.0001035 |
| DDX21 | 3.03E-12 | 0.27219406 | 0.208 | 0.146 | 1.16E-07 |
| PHACTR2 | 4.65E-13 | 0.27153054 | 0.305 | 0.23 | 1.78E-08 |
| RUFY3 | 2.27E-14 | 0.27116888 | 0.388 | 0.307 | 8.69E-10 |
| LAIR1 | 9.81E-14 | 0.2693669 | 0.302 | 0.224 | 3.75E-09 |
| TMEM39A | 1.39E-14 | 0.26865185 | 0.137 | 0.08 | 5.33E-10 |
| FNIP1 | 1.06E-12 | 0.26850241 | 0.422 | 0.343 | 4.04E-08 |
| NOP58 | 1.43E-09 | 0.26839443 | 0.179 | 0.128 | 5.48E-05 |
| SAP30 | 2.35E-20 | 0.2682361 | 0.111 | 0.05 | 8.98E-16 |
| ZHX2 | 1.34E-09 | 0.26753503 | 0.238 | 0.182 | 5.13E-05 |
| DISP1 | 4.77E-12 | 0.2657758 | 0.455 | 0.376 | 1.82E-07 |
| MS4A14 | 2.75E-11 | 0.26536238 | 0.144 | 0.094 | 1.05E-06 |
| KDM4B | 1.45E-09 | 0.26477546 | 0.327 | 0.264 | 5.54E-05 |
| THUMPD3-A: | 2.63E-14 | 0.26455103 | 0.214 | 0.145 | 1.00E-09 |
| DTNA | 1.26E-13 | 0.264076 | 0.553 | 0.474 | 4.83E-09 |

| | | | | | |
|---|---|---|---|---|---|
| TRIM27 | 3.86E-10 | 0.26381496 | 0.198 | 0.143 | 1.48E-05 |
| TTYH3 | 1.26E-10 | 0.2630389 | 0.147 | 0.097 | 4.83E-06 |
| FARP1 | 7.84E-14 | 0.26198815 | 0.27 | 0.193 | 3.00E-09 |
| HERC4 | 7.48E-12 | 0.26158128 | 0.518 | 0.453 | 2.86E-07 |
| KLF6 | 1.67E-14 | 0.26129677 | 0.321 | 0.238 | 6.37E-10 |
| ST18 | 1.43E-11 | 0.26121162 | 0.168 | 0.11 | 5.47E-07 |
| ATP6V0B | 3.35E-12 | 0.260706 | 0.237 | 0.169 | 1.28E-07 |
| PNPLA6 | 3.02E-11 | 0.26015462 | 0.174 | 0.118 | 1.16E-06 |
| ABHD15-AS1 | 1.26E-09 | 0.26009648 | 0.179 | 0.128 | 4.81E-05 |
| CHST15 | 1.11E-10 | 0.26007346 | 0.413 | 0.342 | 4.24E-06 |
| LTA4H | 7.07E-15 | 0.25997929 | 0.115 | 0.063 | 2.70E-10 |
| PSME4 | 4.76E-09 | 0.25995883 | 0.39 | 0.326 | 0.00018199 |
| CCR1 | 4.40E-20 | 0.25843904 | 0.15 | 0.079 | 1.68E-15 |
| BAX | 5.31E-14 | 0.2578501 | 0.144 | 0.086 | 2.03E-09 |
| JARID2 | 4.42E-09 | 0.25775294 | 0.389 | 0.326 | 0.00016883 |
| TF | 4.43E-08 | 0.25699582 | 0.119 | 0.08 | 0.00169212 |
| SNHG29 | 6.31E-13 | 0.25478404 | 0.134 | 0.081 | 2.41E-08 |
| PLEKHF2 | 1.20E-14 | 0.25472912 | 0.103 | 0.053 | 4.58E-10 |
| ARFGAP3 | 4.65E-12 | 0.25444538 | 0.171 | 0.113 | 1.78E-07 |
| HLA-DQB1 | 6.44E-14 | 0.2535002 | 0.161 | 0.1 | 2.46E-09 |
| PRKCB | 5.66E-12 | 0.25327896 | 0.539 | 0.472 | 2.16E-07 |
| FRMD3 | 1.62E-07 | 0.25285596 | 0.113 | 0.077 | 0.00621132 |
| SLFN11 | 7.05E-14 | 0.25259261 | 0.13 | 0.076 | 2.69E-09 |
| RASGRP3 | 7.40E-11 | 0.25162369 | 0.152 | 0.101 | 2.83E-06 |
| RAB11A | 7.66E-08 | 0.25062925 | 0.223 | 0.174 | 0.00292977 |
| AL353138.1 | 1.32E-07 | 0.25041924 | 0.128 | 0.09 | 0.00503073 |
| TMED10 | 8.67E-12 | 0.2503964 | 0.209 | 0.147 | 3.31E-07 |
| MLEC | 2.26E-12 | 0.25003673 | 0.171 | 0.113 | 8.63E-08 |
| TPT1-AS1 | 2.72E-08 | -0.2500508 | 0.154 | 0.198 | 0.0010401 |
| GPM6A | 2.13E-11 | -0.2513313 | 0.209 | 0.275 | 8.15E-07 |
| SDK1 | 4.01E-21 | -0.2535295 | 0.489 | 0.59 | 1.53E-16 |
| CLSTN1 | 1.77E-07 | -0.2541635 | 0.161 | 0.202 | 0.0067702 |
| CPLANE1 | 1.36E-07 | -0.2563817 | 0.147 | 0.188 | 0.00520929 |
| FMN2 | 5.47E-12 | -0.2575812 | 0.061 | 0.104 | 2.09E-07 |
| AC046134.2 | 8.45E-08 | -0.2587201 | 0.093 | 0.13 | 0.00322842 |
| C1orf112 | 1.71E-13 | -0.2593414 | 0.076 | 0.125 | 6.52E-09 |
| TPM1 | 2.27E-12 | -0.2598012 | 0.072 | 0.117 | 8.67E-08 |
| CLEC2D | 1.87E-08 | -0.2598137 | 0.207 | 0.253 | 0.00071449 |
| DTNB | 1.08E-11 | -0.2598936 | 0.458 | 0.524 | 4.14E-07 |
| ZSWIM6 | 1.03E-19 | -0.2616469 | 0.752 | 0.8 | 3.92E-15 |
| SPIN1 | 4.57E-08 | -0.2618361 | 0.304 | 0.352 | 0.00174675 |
| SUGP2 | 9.65E-08 | -0.2657557 | 0.234 | 0.278 | 0.00368822 |
| IFNGR1 | 1.46E-09 | -0.2668746 | 0.534 | 0.573 | 5.60E-05 |
| OTULINL | 2.37E-13 | -0.2678098 | 0.287 | 0.357 | 9.06E-09 |
| KHDRBS3 | 7.66E-10 | -0.2678573 | 0.392 | 0.451 | 2.93E-05 |

| | | | | | |
|---|---|---|---|---|---|
| UQCRB | 4.40E-08 | -0.2690657 | 0.074 | 0.108 | 0.00168048 |
| TNFRSF25 | 2.91E-19 | -0.2695426 | 0.054 | 0.108 | 1.11E-14 |
| RASA3 | 2.85E-15 | -0.2695774 | 0.292 | 0.368 | 1.09E-10 |
| SLC8A1 | 6.95E-22 | -0.2698618 | 0.789 | 0.86 | 2.66E-17 |
| ZC3H6 | 2.25E-12 | -0.2700863 | 0.13 | 0.185 | 8.62E-08 |
| SNTG1 | 7.36E-08 | -0.270272 | 0.128 | 0.171 | 0.00281367 |
| SLC23A2 | 1.88E-13 | -0.2724351 | 0.285 | 0.355 | 7.17E-09 |
| DNAJC10 | 1.82E-09 | -0.2732375 | 0.16 | 0.209 | 6.97E-05 |
| PCLO | 6.75E-11 | -0.2743757 | 0.12 | 0.171 | 2.58E-06 |
| QKI | 2.28E-30 | -0.2744089 | 0.88 | 0.909 | 8.71E-26 |
| IQCJ-SCHIP1 | 2.99E-08 | -0.2745272 | 0.177 | 0.227 | 0.00114142 |
| ROCK2 | 1.90E-11 | -0.2749271 | 0.257 | 0.319 | 7.24E-07 |
| CCM2 | 5.28E-10 | -0.2756438 | 0.281 | 0.338 | 2.02E-05 |
| ITGB2-AS1 | 6.63E-15 | -0.2756751 | 0.097 | 0.153 | 2.53E-10 |
| GRM7 | 1.84E-12 | -0.2762195 | 0.079 | 0.128 | 7.04E-08 |
| TMIGD3 | 4.13E-12 | -0.276501 | 0.234 | 0.299 | 1.58E-07 |
| DENND11 | 1.29E-13 | -0.2769305 | 0.211 | 0.278 | 4.92E-09 |
| MICOS10 | 5.43E-11 | -0.2771369 | 0.19 | 0.246 | 2.07E-06 |
| UNC79 | 2.38E-11 | -0.2773118 | 0.078 | 0.122 | 9.11E-07 |
| SLC4A8 | 8.37E-13 | -0.2774256 | 0.105 | 0.159 | 3.20E-08 |
| NEK3 | 1.27E-15 | -0.2774372 | 0.08 | 0.135 | 4.84E-11 |
| ERMAP | 1.74E-12 | -0.2774832 | 0.093 | 0.142 | 6.67E-08 |
| NCKAP1L | 1.39E-15 | -0.2776294 | 0.38 | 0.458 | 5.30E-11 |
| STRBP | 8.01E-09 | -0.2787776 | 0.262 | 0.315 | 0.00030609 |
| TBC1D4 | 2.69E-15 | -0.2791693 | 0.338 | 0.416 | 1.03E-10 |
| ALOX12-AS1 | 1.94E-08 | -0.2795927 | 0.134 | 0.177 | 0.00073998 |
| DENND3 | 1.08E-16 | -0.2804026 | 0.708 | 0.761 | 4.13E-12 |
| KIF3C | 6.94E-09 | -0.2804629 | 0.08 | 0.117 | 0.00026525 |
| SNX29 | 9.65E-23 | -0.28065 | 0.612 | 0.686 | 3.69E-18 |
| TNIK | 1.46E-07 | -0.28112 | 0.129 | 0.168 | 0.00557005 |
| VSIR | 3.07E-16 | -0.2817984 | 0.35 | 0.434 | 1.17E-11 |
| B3GNT5 | 4.11E-13 | -0.2831029 | 0.268 | 0.338 | 1.57E-08 |
| CDKN2B-AS1 | 1.06E-10 | -0.2839638 | 0.1 | 0.146 | 4.05E-06 |
| SPTLC3 | 1.66E-10 | -0.2849495 | 0.112 | 0.161 | 6.35E-06 |
| NPC1 | 1.38E-12 | -0.2852514 | 0.245 | 0.31 | 5.27E-08 |
| CACNB2 | 3.83E-09 | -0.2855009 | 0.082 | 0.121 | 0.00014623 |
| BTBD9 | 4.12E-11 | -0.2863701 | 0.393 | 0.453 | 1.58E-06 |
| DOCK8 | 1.12E-38 | -0.2867446 | 0.836 | 0.88 | 4.27E-34 |
| PACS2 | 2.96E-12 | -0.2871744 | 0.239 | 0.303 | 1.13E-07 |
| CHRM3 | 1.90E-15 | -0.2879302 | 0.073 | 0.127 | 7.26E-11 |
| NRGN | 5.90E-13 | -0.2883012 | 0.057 | 0.1 | 2.26E-08 |
| WWOX | 2.22E-18 | -0.2890415 | 0.564 | 0.65 | 8.50E-14 |
| P2RY13 | 1.26E-10 | -0.2890584 | 0.128 | 0.176 | 4.82E-06 |
| RALYL | 4.40E-10 | -0.2894493 | 0.139 | 0.19 | 1.68E-05 |
| ASIC2 | 1.12E-12 | -0.2894692 | 0.065 | 0.111 | 4.27E-08 |

| | | | | | |
|---|---|---|---|---|---|
| DNM1 | 4.66E-13 | -0.2901041 | 0.091 | 0.143 | 1.78E-08 |
| DOCK4 | 9.29E-40 | -0.2902386 | 0.96 | 0.97 | 3.55E-35 |
| CNTNAP2 | 2.38E-13 | -0.2902551 | 0.329 | 0.414 | 9.09E-09 |
| AC079142.1 | 1.01E-09 | -0.2906853 | 0.104 | 0.149 | 3.86E-05 |
| LDB2 | 5.81E-10 | -0.2912016 | 0.071 | 0.111 | 2.22E-05 |
| PIK3CD | 6.85E-13 | -0.2913615 | 0.255 | 0.32 | 2.62E-08 |
| KIAA1328 | 1.29E-12 | -0.2917336 | 0.23 | 0.297 | 4.93E-08 |
| PTS | 2.82E-11 | -0.2918118 | 0.08 | 0.124 | 1.08E-06 |
| ATP8B4 | 7.00E-25 | -0.2924397 | 0.693 | 0.764 | 2.68E-20 |
| IL6ST | 1.86E-19 | -0.2932231 | 0.502 | 0.587 | 7.12E-15 |
| ADGRL2 | 5.24E-14 | -0.2935686 | 0.056 | 0.101 | 2.00E-09 |
| EMID1 | 7.60E-15 | -0.293762 | 0.08 | 0.133 | 2.90E-10 |
| CCDC92 | 4.24E-12 | -0.2951868 | 0.113 | 0.164 | 1.62E-07 |
| SYNE1 | 2.77E-14 | -0.2953652 | 0.167 | 0.232 | 1.06E-09 |
| NECAB1 | 5.13E-13 | -0.2984861 | 0.059 | 0.102 | 1.96E-08 |
| MARCH3 | 1.16E-12 | -0.2985701 | 0.31 | 0.38 | 4.45E-08 |
| TRIM9 | 1.44E-11 | -0.2995173 | 0.085 | 0.131 | 5.49E-07 |
| GRM5 | 3.56E-13 | -0.299532 | 0.085 | 0.136 | 1.36E-08 |
| CAPZB | 1.61E-19 | -0.2996908 | 0.578 | 0.645 | 6.15E-15 |
| CADM2 | 1.01E-18 | -0.3004235 | 0.366 | 0.47 | 3.85E-14 |
| NUDT3 | 2.28E-13 | -0.3009369 | 0.166 | 0.227 | 8.73E-09 |
| ARHGAP12 | 4.59E-17 | -0.3009734 | 0.514 | 0.589 | 1.76E-12 |
| PCDH9 | 2.56E-25 | -0.3014163 | 0.448 | 0.572 | 9.78E-21 |
| ZNF609 | 1.61E-15 | -0.3015959 | 0.465 | 0.539 | 6.17E-11 |
| ATP9B | 4.43E-14 | -0.3031476 | 0.364 | 0.433 | 1.69E-09 |
| UBAC2 | 2.10E-12 | -0.3032674 | 0.4 | 0.463 | 8.04E-08 |
| NTM | 4.96E-11 | -0.3033779 | 0.151 | 0.208 | 1.90E-06 |
| FCHSD2 | 6.32E-29 | -0.3034997 | 0.698 | 0.786 | 2.42E-24 |
| S100Z | 2.54E-11 | -0.3044487 | 0.136 | 0.191 | 9.71E-07 |
| LINC00882 | 1.25E-11 | -0.3052072 | 0.071 | 0.114 | 4.78E-07 |
| IGSF21 | 7.80E-18 | -0.3065092 | 0.549 | 0.62 | 2.98E-13 |
| DAGLB | 1.45E-14 | -0.3074282 | 0.208 | 0.276 | 5.53E-10 |
| SLC29A3 | 5.37E-16 | -0.3079981 | 0.115 | 0.177 | 2.05E-11 |
| ALPK1 | 2.39E-12 | -0.3082373 | 0.466 | 0.523 | 9.12E-08 |
| KIRREL3 | 4.70E-09 | -0.3094806 | 0.137 | 0.186 | 0.00017974 |
| BCO2 | 4.99E-13 | -0.3098481 | 0.124 | 0.179 | 1.91E-08 |
| PLAGL1 | 4.53E-10 | -0.3105565 | 0.224 | 0.28 | 1.73E-05 |
| LINC01376 | 6.48E-18 | -0.3108846 | 0.05 | 0.1 | 2.48E-13 |
| PIP5K1B | 8.81E-18 | -0.3110478 | 0.158 | 0.232 | 3.37E-13 |
| LST1 | 2.56E-10 | -0.3110929 | 0.182 | 0.232 | 9.78E-06 |
| AL160272.1 | 5.54E-13 | -0.3111321 | 0.061 | 0.105 | 2.12E-08 |
| TTC39C | 4.15E-15 | -0.3113767 | 0.066 | 0.115 | 1.59E-10 |
| CNTN1 | 4.53E-15 | -0.3116673 | 0.093 | 0.15 | 1.73E-10 |
| TMEM72-AS | 4.69E-17 | -0.3119571 | 0.076 | 0.132 | 1.79E-12 |
| PTPRD | 3.68E-16 | -0.3132985 | 0.288 | 0.377 | 1.41E-11 |

| | | | | | |
|---|---|---|---|---|---|
| OSBPL1A | 1.57E-11 | -0.3139246 | 0.307 | 0.37 | 6.02E-07 |
| MAP2 | 1.71E-13 | -0.3141877 | 0.102 | 0.157 | 6.54E-09 |
| BIN2 | 2.69E-12 | -0.3142787 | 0.217 | 0.279 | 1.03E-07 |
| SEMA4D | 3.33E-12 | -0.3153492 | 0.312 | 0.375 | 1.27E-07 |
| C2CD5 | 1.56E-13 | -0.3154049 | 0.261 | 0.33 | 5.98E-09 |
| ARHGAP17 | 1.43E-13 | -0.315548 | 0.269 | 0.336 | 5.48E-09 |
| MAGI2 | 2.54E-13 | -0.3160124 | 0.268 | 0.35 | 9.70E-09 |
| MYO15B | 6.79E-17 | -0.3162464 | 0.234 | 0.312 | 2.60E-12 |
| ARPP21 | 7.96E-13 | -0.317076 | 0.069 | 0.115 | 3.04E-08 |
| C12orf75 | 1.46E-11 | -0.3177851 | 0.154 | 0.208 | 5.58E-07 |
| LRRC7 | 1.42E-12 | -0.3178139 | 0.151 | 0.211 | 5.45E-08 |
| EMB | 6.36E-15 | -0.3179842 | 0.157 | 0.224 | 2.43E-10 |
| TMEM232 | 8.54E-18 | -0.3182818 | 0.062 | 0.115 | 3.26E-13 |
| NXN | 2.95E-21 | -0.3189521 | 0.126 | 0.205 | 1.13E-16 |
| ZNF117 | 9.69E-17 | -0.3217429 | 0.073 | 0.128 | 3.71E-12 |
| BIN1 | 2.10E-10 | -0.3230243 | 0.528 | 0.583 | 8.04E-06 |
| RNF213 | 2.94E-16 | -0.3242262 | 0.419 | 0.493 | 1.12E-11 |
| LRMP | 3.09E-15 | -0.3249175 | 0.305 | 0.38 | 1.18E-10 |
| MAP4K4 | 3.78E-22 | -0.3249365 | 0.663 | 0.718 | 1.45E-17 |
| FRMPD4 | 1.23E-12 | -0.3258316 | 0.088 | 0.139 | 4.71E-08 |
| ENOSF1 | 3.06E-16 | -0.3265619 | 0.085 | 0.141 | 1.17E-11 |
| PIGL | 8.46E-16 | -0.326714 | 0.094 | 0.152 | 3.23E-11 |
| PALD1 | 3.99E-22 | -0.327013 | 0.65 | 0.72 | 1.53E-17 |
| CMKLR1 | 7.27E-17 | -0.3277612 | 0.225 | 0.305 | 2.78E-12 |
| AIF1 | 6.50E-14 | -0.3283769 | 0.132 | 0.191 | 2.48E-09 |
| VASH1 | 1.14E-16 | -0.3284905 | 0.297 | 0.373 | 4.35E-12 |
| SMYD3 | 4.52E-11 | -0.3301208 | 0.502 | 0.557 | 1.73E-06 |
| SLC4A10 | 3.59E-24 | -0.3302733 | 0.058 | 0.123 | 1.37E-19 |
| GRIA4 | 2.17E-11 | -0.3320773 | 0.098 | 0.146 | 8.29E-07 |
| PLXNA2 | 1.14E-20 | -0.3322366 | 0.063 | 0.124 | 4.36E-16 |
| AC005165.1 | 8.08E-19 | -0.3324179 | 0.068 | 0.125 | 3.09E-14 |
| APBA1 | 1.12E-12 | -0.3325707 | 0.27 | 0.336 | 4.27E-08 |
| FAM118A | 4.65E-12 | -0.3329382 | 0.147 | 0.201 | 1.78E-07 |
| CACNA1D | 8.83E-14 | -0.3334545 | 0.327 | 0.398 | 3.38E-09 |
| RIN2 | 2.33E-17 | -0.3335612 | 0.412 | 0.491 | 8.92E-13 |
| WDR70 | 7.03E-19 | -0.3336902 | 0.478 | 0.559 | 2.69E-14 |
| FGF12 | 1.68E-12 | -0.3342734 | 0.116 | 0.172 | 6.43E-08 |
| MAP2K6 | 6.09E-21 | -0.3376408 | 0.066 | 0.127 | 2.33E-16 |
| AC006230.1 | 4.07E-14 | -0.3376886 | 0.148 | 0.211 | 1.55E-09 |
| NLRP3 | 4.01E-17 | -0.3404158 | 0.089 | 0.147 | 1.53E-12 |
| ULK4 | 2.49E-14 | -0.3406444 | 0.204 | 0.275 | 9.53E-10 |
| TERF2IP | 6.27E-10 | -0.3407607 | 0.131 | 0.177 | 2.39E-05 |
| PRKN | 1.24E-23 | -0.3420063 | 0.481 | 0.584 | 4.74E-19 |
| ABCA5 | 1.50E-16 | -0.3423763 | 0.054 | 0.103 | 5.72E-12 |
| LPAR1 | 2.84E-13 | -0.3427432 | 0.251 | 0.318 | 1.08E-08 |

| | | | | | |
|---|---|---|---|---|---|
| PDE1A | 9.01E-15 | -0.344556 | 0.082 | 0.135 | 3.44E-10 |
| NLGN1 | 1.70E-12 | -0.3459467 | 0.201 | 0.268 | 6.52E-08 |
| TLR10 | 1.00E-16 | -0.3473793 | 0.107 | 0.17 | 3.84E-12 |
| WLS | 1.04E-19 | -0.3483756 | 0.082 | 0.145 | 3.99E-15 |
| MED12L | 3.80E-19 | -0.3490726 | 0.078 | 0.139 | 1.45E-14 |
| DPH6 | 4.66E-16 | -0.3509173 | 0.15 | 0.219 | 1.78E-11 |
| FGF14 | 1.84E-21 | -0.3510466 | 0.17 | 0.26 | 7.02E-17 |
| CC2D2B | 1.80E-12 | -0.351518 | 0.092 | 0.141 | 6.90E-08 |
| NRCAM | 4.86E-15 | -0.3515228 | 0.096 | 0.154 | 1.86E-10 |
| ZNRF2 | 6.75E-16 | -0.351794 | 0.373 | 0.446 | 2.58E-11 |
| ADGRL3 | 3.99E-16 | -0.3519598 | 0.183 | 0.259 | 1.53E-11 |
| FAM149A | 9.88E-30 | -0.3520206 | 0.434 | 0.549 | 3.77E-25 |
| MLXIPL | 5.06E-18 | -0.3529965 | 0.307 | 0.392 | 1.93E-13 |
| CTNND2 | 3.34E-18 | -0.3530592 | 0.18 | 0.263 | 1.28E-13 |
| LTC4S | 2.58E-19 | -0.3541989 | 0.102 | 0.169 | 9.88E-15 |
| TMEM178B | 4.90E-22 | -0.3548879 | 0.064 | 0.127 | 1.87E-17 |
| MDGA2 | 1.85E-16 | -0.3549837 | 0.108 | 0.172 | 7.07E-12 |
| ITGA9 | 4.02E-23 | -0.3555428 | 0.308 | 0.409 | 1.54E-18 |
| RPS20 | 1.47E-12 | -0.3559293 | 0.089 | 0.136 | 5.62E-08 |
| CADPS | 5.96E-12 | -0.3565121 | 0.077 | 0.123 | 2.28E-07 |
| CCDC171 | 9.72E-22 | -0.3571948 | 0.069 | 0.132 | 3.71E-17 |
| AC079015.1 | 6.33E-20 | -0.3580357 | 0.179 | 0.26 | 2.42E-15 |
| LINC02232 | 1.22E-24 | -0.358173 | 0.403 | 0.518 | 4.67E-20 |
| GOLGA8A | 4.70E-30 | -0.3596374 | 0.077 | 0.157 | 1.80E-25 |
| SUMF1 | 3.70E-14 | -0.3597758 | 0.415 | 0.481 | 1.41E-09 |
| ARHGAP24 | 8.45E-47 | -0.3600501 | 0.861 | 0.932 | 3.23E-42 |
| LIPC | 1.00E-19 | -0.3600946 | 0.08 | 0.143 | 3.83E-15 |
| DGKB | 1.22E-14 | -0.3602154 | 0.066 | 0.116 | 4.67E-10 |
| DCLK1 | 3.25E-18 | -0.3603117 | 0.064 | 0.12 | 1.24E-13 |
| IGF1 | 3.29E-26 | -0.3604185 | 0.047 | 0.109 | 1.26E-21 |
| LILRB4 | 1.16E-18 | -0.3611108 | 0.119 | 0.186 | 4.44E-14 |
| PDE9A | 6.99E-21 | -0.3612282 | 0.118 | 0.192 | 2.67E-16 |
| MIR31HG | 2.06E-28 | -0.3634089 | 0.054 | 0.124 | 7.87E-24 |
| RPL23 | 5.00E-10 | -0.3635824 | 0.12 | 0.165 | 1.91E-05 |
| APBB1IP | 5.14E-40 | -0.363807 | 0.797 | 0.871 | 1.97E-35 |
| CENPP | 8.84E-15 | -0.3651828 | 0.164 | 0.231 | 3.38E-10 |
| INPP5D | 9.19E-39 | -0.3664338 | 0.679 | 0.772 | 3.51E-34 |
| CKB | 4.28E-17 | -0.3670899 | 0.092 | 0.153 | 1.64E-12 |
| SNHG14 | 1.44E-14 | -0.3671697 | 0.447 | 0.516 | 5.50E-10 |
| SPG7 | 1.92E-21 | -0.3674923 | 0.181 | 0.265 | 7.33E-17 |
| AIG1 | 1.17E-14 | -0.37037 | 0.293 | 0.361 | 4.47E-10 |
| AFF3 | 5.94E-18 | -0.3713034 | 0.061 | 0.116 | 2.27E-13 |
| ARID3A | 6.32E-20 | -0.3717107 | 0.177 | 0.258 | 2.42E-15 |
| RAP1GAP2 | 5.61E-22 | -0.3770951 | 0.218 | 0.308 | 2.14E-17 |
| PCDH7 | 1.59E-20 | -0.3774965 | 0.083 | 0.149 | 6.09E-16 |

| | | | | | |
|---|---|---|---|---|---|
| SP140 | 5.99E-14 | -0.3786173 | 0.126 | 0.183 | 2.29E-09 |
| PTPN14 | 6.00E-27 | -0.3791993 | 0.064 | 0.135 | 2.29E-22 |
| CDH12 | 1.05E-21 | -0.3796683 | 0.058 | 0.118 | 4.00E-17 |
| GASK1B | 2.36E-19 | -0.3813028 | 0.099 | 0.165 | 9.02E-15 |
| NELL2 | 6.39E-22 | -0.3820634 | 0.056 | 0.115 | 2.44E-17 |
| RGS7 | 4.15E-20 | -0.3827666 | 0.097 | 0.166 | 1.59E-15 |
| ENTPD1 | 1.76E-25 | -0.3838146 | 0.469 | 0.569 | 6.73E-21 |
| AZIN1-AS1 | 2.80E-19 | -0.3839557 | 0.146 | 0.221 | 1.07E-14 |
| PELI2 | 2.04E-20 | -0.3849789 | 0.215 | 0.305 | 7.78E-16 |
| TSPAN7 | 3.35E-31 | -0.3861057 | 0.045 | 0.114 | 1.28E-26 |
| TENM2 | 6.26E-17 | -0.3865691 | 0.141 | 0.214 | 2.39E-12 |
| RYR2 | 8.88E-16 | -0.3871418 | 0.13 | 0.196 | 3.39E-11 |
| SLC25A29 | 5.59E-18 | -0.3872029 | 0.182 | 0.257 | 2.14E-13 |
| CAMK1D | 1.57E-31 | -0.3873768 | 0.523 | 0.631 | 6.01E-27 |
| SLC2A5 | 1.03E-10 | -0.3876194 | 0.485 | 0.545 | 3.93E-06 |
| PLCB2 | 9.24E-22 | -0.3877168 | 0.204 | 0.291 | 3.53E-17 |
| MKNK1 | 1.76E-24 | -0.3883916 | 0.504 | 0.594 | 6.73E-20 |
| GAREM1 | 1.44E-07 | -0.388596 | 0.417 | 0.455 | 0.00549521 |
| PLCL2 | 1.40E-23 | -0.3895578 | 0.423 | 0.523 | 5.37E-19 |
| CDH23 | 7.75E-26 | -0.3904643 | 0.114 | 0.197 | 2.96E-21 |
| AKT3 | 1.51E-30 | -0.3916365 | 0.481 | 0.587 | 5.75E-26 |
| CSF1R | 3.34E-35 | -0.3922451 | 0.668 | 0.748 | 1.28E-30 |
| PMEPA1 | 1.62E-20 | -0.3924899 | 0.111 | 0.182 | 6.18E-16 |
| KCNQ5 | 2.64E-21 | -0.3925445 | 0.119 | 0.197 | 1.01E-16 |
| SFMBT2 | 6.56E-35 | -0.3934384 | 0.83 | 0.863 | 2.51E-30 |
| SRGAP3 | 6.37E-19 | -0.3940554 | 0.335 | 0.418 | 2.44E-14 |
| KLF12 | 1.56E-24 | -0.3963713 | 0.077 | 0.148 | 5.97E-20 |
| PARP14 | 3.78E-19 | -0.3966409 | 0.385 | 0.468 | 1.44E-14 |
| LRMDA | 9.96E-87 | -0.3977668 | 0.923 | 0.964 | 3.81E-82 |
| C4orf19 | 6.20E-24 | -0.3988368 | 0.071 | 0.138 | 2.37E-19 |
| PDE3B | 2.60E-25 | -0.3990723 | 0.474 | 0.58 | 9.96E-21 |
| NAALADL2 | 9.71E-19 | -0.3996305 | 0.272 | 0.36 | 3.71E-14 |
| GRIK2 | 5.01E-26 | -0.3996713 | 0.124 | 0.213 | 1.92E-21 |
| AC008014.1 | 7.24E-24 | -0.4020785 | 0.104 | 0.183 | 2.77E-19 |
| ARHGAP32 | 1.12E-22 | -0.402147 | 0.257 | 0.354 | 4.27E-18 |
| LRIG1 | 4.77E-19 | -0.4068519 | 0.099 | 0.164 | 1.82E-14 |
| PPFIA2 | 6.12E-22 | -0.4068739 | 0.102 | 0.175 | 2.34E-17 |
| TTC28 | 1.15E-29 | -0.4070747 | 0.382 | 0.493 | 4.41E-25 |
| TMSB4X | 1.70E-16 | -0.4084272 | 0.529 | 0.603 | 6.50E-12 |
| GRIA2 | 5.11E-28 | -0.4093002 | 0.119 | 0.209 | 1.95E-23 |
| LINC01141 | 6.22E-30 | -0.4110753 | 0.224 | 0.337 | 2.38E-25 |
| ULK2 | 1.54E-23 | -0.4111959 | 0.172 | 0.259 | 5.89E-19 |
| ERC2 | 2.43E-14 | -0.4114413 | 0.298 | 0.372 | 9.28E-10 |
| NEGR1 | 6.25E-25 | -0.4127884 | 0.145 | 0.236 | 2.39E-20 |
| OLR1 | 1.84E-19 | -0.4132948 | 0.312 | 0.404 | 7.02E-15 |

| | | | | | |
|---|---|---|---|---|---|
| DLGAP2 | 6.03E-11 | -0.4137251 | 0.113 | 0.163 | 2.30E-06 |
| TMEM156 | 5.04E-28 | -0.4138108 | 0.328 | 0.437 | 1.93E-23 |
| MEF2A | 1.66E-71 | -0.41416 | 0.872 | 0.923 | 6.34E-67 |
| PSD3 | 7.24E-23 | -0.4142655 | 0.232 | 0.326 | 2.77E-18 |
| HIST1H2AC | 1.63E-08 | -0.4158349 | 0.259 | 0.304 | 0.00062346 |
| CACNA2D3 | 2.52E-15 | -0.4189223 | 0.077 | 0.13 | 9.62E-11 |
| DMD | 5.94E-13 | -0.4192613 | 0.159 | 0.222 | 2.27E-08 |
| CTNNA3 | 1.54E-16 | -0.4238092 | 0.175 | 0.251 | 5.90E-12 |
| HS6ST3 | 7.80E-28 | -0.4259732 | 0.071 | 0.147 | 2.98E-23 |
| UNC5C | 5.29E-10 | -0.4267115 | 0.143 | 0.195 | 2.02E-05 |
| ESRRG | 4.31E-16 | -0.4267554 | 0.08 | 0.135 | 1.65E-11 |
| CTNNA2 | 1.45E-18 | -0.4274054 | 0.176 | 0.256 | 5.55E-14 |
| BCAR3 | 3.46E-22 | -0.4276563 | 0.262 | 0.355 | 1.32E-17 |
| RIMS2 | 3.67E-25 | -0.4297543 | 0.103 | 0.185 | 1.40E-20 |
| OSBPL6 | 1.61E-30 | -0.4304635 | 0.135 | 0.234 | 6.15E-26 |
| FMNL3 | 8.76E-37 | -0.4308285 | 0.445 | 0.568 | 3.35E-32 |
| KCNIP1 | 2.06E-24 | -0.4314532 | 0.355 | 0.461 | 7.86E-20 |
| TARBP1 | 5.73E-29 | -0.4321433 | 0.105 | 0.19 | 2.19E-24 |
| LRRK1 | 1.74E-26 | -0.4339065 | 0.501 | 0.597 | 6.67E-22 |
| PLEKHG5 | 3.57E-38 | -0.4341054 | 0.035 | 0.107 | 1.36E-33 |
| NCKAP5 | 5.40E-44 | -0.4360132 | 0.396 | 0.553 | 2.06E-39 |
| DSCAM | 5.83E-25 | -0.4361281 | 0.496 | 0.583 | 2.23E-20 |
| LMCD1-AS1 | 2.89E-18 | -0.4362837 | 0.083 | 0.143 | 1.11E-13 |
| ANK2 | 2.88E-13 | -0.4365774 | 0.359 | 0.428 | 1.10E-08 |
| FOXP2 | 7.84E-66 | -0.4377649 | 0.427 | 0.631 | 3.00E-61 |
| ADAMTSL2 | 5.21E-25 | -0.4380645 | 0.116 | 0.198 | 1.99E-20 |
| UST | 3.71E-17 | -0.4381615 | 0.221 | 0.3 | 1.42E-12 |
| KCNQ1 | 1.84E-22 | -0.4390411 | 0.319 | 0.405 | 7.04E-18 |
| APPL2 | 5.82E-32 | -0.4390604 | 0.385 | 0.5 | 2.22E-27 |
| CHN1 | 1.78E-27 | -0.4394404 | 0.085 | 0.163 | 6.80E-23 |
| CSMD3 | 1.00E-26 | -0.4402808 | 0.103 | 0.186 | 3.84E-22 |
| TBC1D22A | 4.66E-53 | -0.4434589 | 0.667 | 0.776 | 1.78E-48 |
| MRC2 | 1.08E-30 | -0.4441428 | 0.109 | 0.2 | 4.13E-26 |
| A2M | 1.16E-26 | -0.4446336 | 0.477 | 0.57 | 4.44E-22 |
| LGMN | 3.78E-29 | -0.4453153 | 0.119 | 0.209 | 1.44E-24 |
| KCNMB2-AS1 | 6.72E-20 | -0.4478864 | 0.148 | 0.227 | 2.57E-15 |
| ICA1 | 9.53E-31 | -0.4484632 | 0.089 | 0.175 | 3.64E-26 |
| PLCB1 | 5.53E-26 | -0.4489198 | 0.131 | 0.222 | 2.11E-21 |
| ASTN1 | 1.76E-32 | -0.4545033 | 0.053 | 0.127 | 6.75E-28 |
| EEPD1 | 1.19E-25 | -0.4554707 | 0.085 | 0.159 | 4.56E-21 |
| NRG3 | 5.87E-29 | -0.457927 | 0.248 | 0.363 | 2.24E-24 |
| PID1 | 3.15E-21 | -0.4584629 | 0.095 | 0.164 | 1.20E-16 |
| ARHGAP25 | 1.01E-22 | -0.4585897 | 0.478 | 0.562 | 3.87E-18 |
| P3H2 | 1.45E-35 | -0.463215 | 0.151 | 0.264 | 5.56E-31 |
| SORL1 | 4.36E-55 | -0.4634669 | 0.795 | 0.835 | 1.66E-50 |

| | | | | | |
|---|---|---|---|---|---|
| KALRN | 1.57E-19 | -0.464726 | 0.107 | 0.176 | 6.01E-15 |
| MEF2C | 1.52E-79 | -0.4675685 | 0.84 | 0.918 | 5.82E-75 |
| CFAP74 | 6.66E-48 | -0.4690489 | 0.041 | 0.129 | 2.55E-43 |
| MEG3 | 8.68E-19 | -0.4701117 | 0.25 | 0.33 | 3.32E-14 |
| DLGAP1 | 1.86E-28 | -0.4714293 | 0.187 | 0.293 | 7.10E-24 |
| CIRBP | 2.86E-32 | -0.4744349 | 0.214 | 0.323 | 1.09E-27 |
| ITGB5 | 3.12E-26 | -0.4747347 | 0.233 | 0.329 | 1.19E-21 |
| MAPK10 | 1.13E-20 | -0.4779358 | 0.132 | 0.208 | 4.33E-16 |
| MIR99AHG | 4.96E-34 | -0.478469 | 0.4 | 0.52 | 1.90E-29 |
| ANKRD44 | 5.15E-68 | -0.4791388 | 0.679 | 0.803 | 1.97E-63 |
| PARD3B | 2.58E-25 | -0.4811267 | 0.123 | 0.208 | 9.86E-21 |
| ARHGAP22 | 3.82E-58 | -0.4833106 | 0.649 | 0.766 | 1.46E-53 |
| MIS18BP1 | 1.20E-28 | -0.4837429 | 0.284 | 0.391 | 4.58E-24 |
| LINC01268 | 1.54E-31 | -0.4840083 | 0.134 | 0.23 | 5.87E-27 |
| FAM155A | 2.41E-25 | -0.485828 | 0.18 | 0.277 | 9.21E-21 |
| ANKS1B | 2.24E-23 | -0.4886701 | 0.223 | 0.319 | 8.58E-19 |
| AC087564.1 | 2.55E-22 | -0.4901215 | 0.226 | 0.317 | 9.77E-18 |
| AUTS2 | 1.61E-48 | -0.4909438 | 0.578 | 0.704 | 6.14E-44 |
| AL008633.1 | 4.00E-22 | -0.4912672 | 0.266 | 0.359 | 1.53E-17 |
| NKAIN2 | 6.13E-18 | -0.4918902 | 0.177 | 0.257 | 2.34E-13 |
| PPP6R2 | 3.11E-40 | -0.49465 | 0.382 | 0.509 | 1.19E-35 |
| LSAMP | 1.67E-27 | -0.4978325 | 0.263 | 0.375 | 6.37E-23 |
| MAGI1 | 2.33E-35 | -0.4990113 | 0.219 | 0.341 | 8.89E-31 |
| NOS1AP | 4.32E-42 | -0.4996439 | 0.044 | 0.127 | 1.65E-37 |
| C3 | 3.48E-48 | -0.5010896 | 0.652 | 0.771 | 1.33E-43 |
| SNAP25 | 3.11E-29 | -0.5035605 | 0.104 | 0.191 | 1.19E-24 |
| RGS10 | 8.85E-34 | -0.5042548 | 0.281 | 0.396 | 3.38E-29 |
| AC010378.2 | 6.42E-52 | -0.5049947 | 0.048 | 0.147 | 2.45E-47 |
| SLC22A23 | 5.53E-23 | -0.5069876 | 0.158 | 0.244 | 2.11E-18 |
| LINC01091 | 3.80E-30 | -0.5071676 | 0.142 | 0.243 | 1.45E-25 |
| CLU | 2.02E-20 | -0.5088578 | 0.134 | 0.208 | 7.73E-16 |
| LRP1B | 2.75E-28 | -0.5088605 | 0.291 | 0.405 | 1.05E-23 |
| GLIS3 | 5.20E-32 | -0.5095851 | 0.102 | 0.195 | 1.99E-27 |
| ITPR2 | 6.34E-102 | -0.5109552 | 0.857 | 0.927 | 2.42E-97 |
| SPARCL1 | 2.06E-25 | -0.513394 | 0.096 | 0.173 | 7.89E-21 |
| STXBP5L | 8.10E-34 | -0.5136744 | 0.085 | 0.174 | 3.09E-29 |
| RPS25 | 7.25E-11 | -0.5153578 | 0.167 | 0.216 | 2.77E-06 |
| OPCML | 5.46E-24 | -0.5182974 | 0.203 | 0.298 | 2.09E-19 |
| SETBP1 | 3.18E-34 | -0.5224063 | 0.353 | 0.475 | 1.21E-29 |
| RFX3-AS1 | 2.12E-36 | -0.5233952 | 0.091 | 0.185 | 8.10E-32 |
| AOAH | 3.20E-58 | -0.5240173 | 0.589 | 0.731 | 1.22E-53 |
| NEXMIF | 1.82E-40 | -0.5252515 | 0.034 | 0.108 | 6.96E-36 |
| DLG2 | 1.57E-42 | -0.527448 | 0.399 | 0.55 | 6.00E-38 |
| EML1 | 2.36E-38 | -0.5347903 | 0.127 | 0.234 | 9.03E-34 |
| RTTN | 2.84E-18 | -0.5430044 | 0.333 | 0.41 | 1.08E-13 |

| | | | | | |
|---|---|---|---|---|---|
| ATRNL1 | 7.38E-29 | -0.5430267 | 0.126 | 0.22 | 2.82E-24 |
| PLCXD3 | 6.86E-40 | -0.5446021 | 0.04 | 0.117 | 2.62E-35 |
| RPS11 | 4.49E-16 | -0.5451109 | 0.157 | 0.221 | 1.72E-11 |
| ST6GAL1 | 2.03E-81 | -0.5528852 | 0.832 | 0.887 | 7.75E-77 |
| APBA2 | 3.53E-37 | -0.5574477 | 0.075 | 0.164 | 1.35E-32 |
| AGBL4 | 1.12E-33 | -0.5589916 | 0.084 | 0.173 | 4.27E-29 |
| GMDS-DT | 8.08E-40 | -0.5593816 | 0.34 | 0.47 | 3.09E-35 |
| DPP10 | 6.64E-24 | -0.5596757 | 0.202 | 0.297 | 2.54E-19 |
| TIAM1 | 3.41E-51 | -0.563449 | 0.426 | 0.566 | 1.30E-46 |
| MCF2L | 1.94E-45 | -0.5656096 | 0.422 | 0.552 | 7.41E-41 |
| GAK | 2.15E-39 | -0.5679114 | 0.262 | 0.384 | 8.22E-35 |
| LINC01374 | 2.22E-48 | -0.5705138 | 0.437 | 0.581 | 8.49E-44 |
| PCNX2 | 1.98E-68 | -0.5840293 | 0.49 | 0.656 | 7.56E-64 |
| RPL13A | 2.99E-08 | -0.5841964 | 0.246 | 0.285 | 0.00114337 |
| NRXN3 | 7.98E-25 | -0.5879987 | 0.306 | 0.417 | 3.05E-20 |
| GRID2 | 2.69E-41 | -0.5900885 | 0.427 | 0.57 | 1.03E-36 |
| NLRP1 | 1.18E-48 | -0.5984401 | 0.14 | 0.266 | 4.49E-44 |
| SIPA1L2 | 1.43E-47 | -0.6000207 | 0.42 | 0.554 | 5.47E-43 |
| KCNQ3 | 5.80E-96 | -0.6045213 | 0.788 | 0.898 | 2.22E-91 |
| HIVEP3 | 1.26E-56 | -0.61417 | 0.483 | 0.632 | 4.81E-52 |
| CSMD1 | 9.56E-55 | -0.6256334 | 0.288 | 0.457 | 3.65E-50 |
| SLC1A2 | 1.56E-45 | -0.6292591 | 0.053 | 0.145 | 5.97E-41 |
| SYT1 | 5.48E-50 | -0.6370217 | 0.258 | 0.409 | 2.09E-45 |
| MRC1 | 1.60E-56 | -0.6405451 | 0.126 | 0.267 | 6.11E-52 |
| CYFIP1 | 4.59E-83 | -0.6462654 | 0.64 | 0.773 | 1.76E-78 |
| RBFOX1 | 4.88E-46 | -0.646384 | 0.355 | 0.508 | 1.86E-41 |
| AZIN2 | 7.31E-61 | -0.6468291 | 0.081 | 0.205 | 2.79E-56 |
| CCSER1 | 6.89E-37 | -0.6488862 | 0.091 | 0.189 | 2.63E-32 |
| PSMA1 | 1.81E-53 | -0.6559328 | 0.321 | 0.471 | 6.92E-49 |
| AC008591.1 | 6.44E-49 | -0.6562611 | 0.041 | 0.129 | 2.46E-44 |
| LINC02057 | 1.41E-60 | -0.6580554 | 0.062 | 0.177 | 5.41E-56 |
| PRKCA | 5.51E-65 | -0.6645909 | 0.56 | 0.712 | 2.10E-60 |
| SYNDIG1 | 6.72E-86 | -0.6685591 | 0.542 | 0.724 | 2.57E-81 |
| CPA6 | 1.55E-46 | -0.6736341 | 0.05 | 0.142 | 5.91E-42 |
| ABCC4 | 6.81E-88 | -0.6751575 | 0.56 | 0.719 | 2.60E-83 |
| LINC01266 | 1.65E-44 | -0.6774576 | 0.067 | 0.163 | 6.31E-40 |
| KBTBD12 | 4.65E-56 | -0.6781422 | 0.187 | 0.333 | 1.78E-51 |
| CACNB4 | 2.56E-61 | -0.6816477 | 0.257 | 0.421 | 9.79E-57 |
| ROBO2 | 1.77E-42 | -0.6845446 | 0.163 | 0.292 | 6.77E-38 |
| CX3CR1 | 1.20E-57 | -0.6917341 | 0.29 | 0.445 | 4.59E-53 |
| PRDM11 | 6.52E-47 | -0.6923044 | 0.093 | 0.203 | 2.49E-42 |
| ADAM28 | 1.02E-118 | -0.6969082 | 0.672 | 0.806 | 3.90E-114 |
| DIP2A | 6.57E-60 | -0.7011809 | 0.29 | 0.443 | 2.51E-55 |
| LRRTM4 | 5.89E-68 | -0.706996 | 0.168 | 0.337 | 2.25E-63 |
| NRXN1 | 1.08E-61 | -0.7105825 | 0.233 | 0.402 | 4.13E-57 |

| | | | | | |
|---|---|---|---|---|---|
| CSF2RA | 6.05E-82 | -0.7366921 | 0.384 | 0.569 | 2.31E-77 |
| PPP2R2B | 1.98E-19 | -0.7395295 | 0.206 | 0.293 | 7.58E-15 |
| NAV3 | 7.96E-158 | -0.7400989 | 0.718 | 0.883 | 3.04E-153 |
| CECR2 | 7.53E-81 | -0.7440289 | 0.058 | 0.195 | 2.88E-76 |
| LINC02642 | 1.39E-77 | -0.7474154 | 0.051 | 0.18 | 5.31E-73 |
| AL592429.2 | 2.62E-70 | -0.7554417 | 0.027 | 0.129 | 1.00E-65 |
| HTRA1 | 4.21E-59 | -0.758974 | 0.266 | 0.418 | 1.61E-54 |
| MIR646HG | 2.06E-69 | -0.7631959 | 0.396 | 0.568 | 7.86E-65 |
| LINC02649 | 3.19E-73 | -0.7900645 | 0.134 | 0.294 | 1.22E-68 |
| AC025569.1 | 3.27E-89 | -0.8044566 | 0.122 | 0.3 | 1.25E-84 |
| FRMD4A | 1.76E-251 | -0.8337523 | 0.905 | 0.974 | 6.73E-247 |
| XACT | 1.23E-49 | -0.839673 | 0.116 | 0.241 | 4.69E-45 |
| GPR183 | 3.82E-72 | -0.8538601 | 0.069 | 0.205 | 1.46E-67 |
| TLN2 | 3.47E-119 | -0.8652786 | 0.351 | 0.587 | 1.33E-114 |
| PDE4D | 1.24E-87 | -0.8813514 | 0.24 | 0.443 | 4.73E-83 |
| P2RY12 | 4.21E-119 | -0.8835886 | 0.405 | 0.629 | 1.61E-114 |
| KCNIP4 | 2.00E-107 | -0.997626 | 0.392 | 0.611 | 7.64E-103 |
| ST6GALNAC[illegible] | 3.01E-221 | -1.0683719 | 0.564 | 0.826 | 1.15E-216 |
| NAV2 | 7.20E-106 | -1.0721488 | 0.228 | 0.447 | 2.75E-101 |
| ZFPM2-AS1 | 8.02E-123 | -1.0775272 | 0.053 | 0.232 | 3.07E-118 |
| SOX5 | 5.86E-123 | -1.155299 | 0.1 | 0.303 | 2.24E-118 |
| LINC02712 | 3.26E-150 | -1.1639503 | 0.079 | 0.298 | 1.25E-145 |
| PDK4 | 1.36E-23 | -1.213868 | 0.129 | 0.201 | 5.19E-19 |
| RASGEF1C | 8.47E-285 | -1.5310822 | 0.232 | 0.608 | 3.24E-280 |
| AC008691.1 | 4.90E-221 | -1.8136139 | 0.091 | 0.377 | 1.87E-216 |
| AC008050.1 | 1.62E-236 | -1.8448489 | 0.066 | 0.347 | 6.17E-232 |
| AC011586.2 | 2.62E-174 | -2.4346959 | 0.006 | 0.171 | 1.00E-169 |

**Oligodendrocyte DEGs**

| | p_val | avg_log2FC | pct.1 | pct.2 | p_val_adj |
|---|---|---|---|---|---|
| FKBP5 | 0 | 1.55533455 | 0.61 | 0.195 | 0 |
| AC008571.2 | 0 | 1.49093294 | 0.607 | 0.277 | 0 |
| AC009063.2 | 0 | 1.34920838 | 0.554 | 0.176 | 0 |
| QDPR | 0 | 1.30871309 | 0.806 | 0.505 | 0 |
| PLCG2 | 0 | 1.23763265 | 0.618 | 0.34 | 0 |
| SELENOP | 0 | 1.14797988 | 0.543 | 0.292 | 0 |
| SPP1 | 0 | 1.10857798 | 0.58 | 0.329 | 0 |
| KCNMB4 | 0 | 1.08922687 | 0.675 | 0.384 | 0 |
| SLC38A2 | 0 | 1.07170249 | 0.518 | 0.236 | 0 |
| ANLN | 0 | 1.05855972 | 0.685 | 0.385 | 0 |
| ZBTB16 | 0 | 1.04179058 | 0.727 | 0.401 | 0 |
| ABCA6 | 0 | 1.02188706 | 0.232 | 0.049 | 0 |
| NEAT1 | 0 | 1.0114375 | 0.835 | 0.659 | 0 |
| PTPRM | 0 | 0.99154104 | 0.348 | 0.15 | 0 |
| EFHD1 | 0 | 0.98461916 | 0.569 | 0.317 | 0 |
| SUN2 | 0 | 0.96671704 | 0.591 | 0.355 | 0 |
| MT-ND3 | 0 | 0.93704898 | 0.863 | 0.702 | 0 |
| ZNF652 | 0 | 0.90604048 | 0.567 | 0.338 | 0 |
| AC119674.1 | 0 | 0.88359627 | 0.201 | 0.055 | 0 |
| CDKL1 | 0 | 0.85475897 | 0.351 | 0.144 | 0 |
| LRRC1 | 0 | 0.85223403 | 0.563 | 0.316 | 0 |
| CPQ | 0 | 0.83657313 | 0.639 | 0.403 | 0 |
| BCL2 | 0 | 0.81955268 | 0.404 | 0.221 | 0 |
| RGCC | 0 | 0.79652062 | 0.479 | 0.271 | 0 |
| TMPRSS5 | 0 | 0.79527039 | 0.25 | 0.097 | 0 |
| CRYAB | 2.50E-185 | 0.77268025 | 0.458 | 0.329 | 9.54E-181 |
| NCAM2 | 0 | 0.76934903 | 0.926 | 0.819 | 0 |
| MT3 | 0 | 0.75733026 | 0.276 | 0.115 | 0 |
| EPCAM-DT | 0 | 0.7503684 | 0.336 | 0.155 | 0 |
| LAMP2 | 0 | 0.74366802 | 0.768 | 0.579 | 0 |
| MTRNR2L12 | 0 | 0.73835672 | 0.69 | 0.498 | 0 |
| NDRG1 | 0 | 0.72880531 | 0.594 | 0.374 | 0 |
| ADAMTSL3 | 0 | 0.72373671 | 0.322 | 0.142 | 0 |
| LRP4 | 0 | 0.71074448 | 0.356 | 0.166 | 0 |
| AMER2 | 0 | 0.70619855 | 0.74 | 0.549 | 0 |
| APOLD1 | 1.48E-296 | 0.70409911 | 0.284 | 0.128 | 5.66E-292 |
| MID1IP1 | 0 | 0.69813353 | 0.256 | 0.104 | 0 |
| SEPTIN8 | 0 | 0.69527908 | 0.471 | 0.273 | 0 |
| GPRC5B | 0 | 0.6914032 | 0.703 | 0.514 | 0 |
| SLC39A11 | 0 | 0.6852675 | 0.664 | 0.479 | 0 |
| AC116362.1 | 6.24E-234 | 0.68233532 | 0.195 | 0.076 | 2.39E-229 |
| SGK1 | 0 | 0.67491716 | 0.824 | 0.643 | 0 |
| KIF19 | 0 | 0.67485671 | 0.235 | 0.077 | 0 |

| | | | | | |
|---|---|---|---|---|---|
| ST6GALNAC3 | 0 | 0.67225955 | 0.662 | 0.479 | 0 |
| WWOX | 0 | 0.66867015 | 0.826 | 0.693 | 0 |
| AC093865.1 | 2.28E-278 | 0.66673084 | 0.234 | 0.093 | 8.72E-274 |
| CSRP1 | 1.36E-202 | 0.66274247 | 0.453 | 0.316 | 5.19E-198 |
| PRKX | 0 | 0.6579131 | 0.16 | 0.03 | 0 |
| CCP110 | 0 | 0.65651077 | 0.63 | 0.43 | 0 |
| ERBIN | 0 | 0.64810698 | 0.891 | 0.759 | 0 |
| ADIPOR2 | 0 | 0.6466296 | 0.66 | 0.466 | 0 |
| SORT1 | 0 | 0.6455055 | 0.794 | 0.625 | 0 |
| PRUNE2 | 0 | 0.63083747 | 0.889 | 0.761 | 0 |
| FANCC | 4.77E-275 | 0.62844116 | 0.237 | 0.096 | 1.82E-270 |
| PIEZO2 | 1.53E-233 | 0.6236619 | 0.561 | 0.417 | 5.84E-229 |
| MAN2A1 | 0 | 0.62235555 | 0.89 | 0.761 | 0 |
| ADARB2 | 0 | 0.62191025 | 0.567 | 0.349 | 0 |
| PAQR8 | 7.19E-226 | 0.62076388 | 0.306 | 0.167 | 2.75E-221 |
| FAM53B | 5.44E-278 | 0.62034103 | 0.497 | 0.325 | 2.08E-273 |
| NDRG2 | 1.08E-199 | 0.61054995 | 0.476 | 0.336 | 4.14E-195 |
| METTL7A | 0 | 0.60649087 | 0.209 | 0.064 | 0 |
| HP1BP3 | 0 | 0.6018607 | 0.59 | 0.409 | 0 |
| BOK | 8.25E-187 | 0.60141076 | 0.368 | 0.235 | 3.15E-182 |
| ZDHHC20 | 8.90E-235 | 0.5951877 | 0.549 | 0.397 | 3.40E-230 |
| B2M | 6.42E-180 | 0.59365319 | 0.261 | 0.144 | 2.45E-175 |
| CLMN | 0 | 0.59115388 | 0.844 | 0.696 | 0 |
| HSPA1A | 3.41E-139 | 0.5867888 | 0.261 | 0.154 | 1.30E-134 |
| PDK4 | 1.38E-270 | 0.58358769 | 0.263 | 0.119 | 5.27E-266 |
| PEX5L | 0 | 0.57971982 | 0.887 | 0.774 | 0 |
| IKZF2 | 4.01E-222 | 0.57857954 | 0.401 | 0.249 | 1.53E-217 |
| CA2 | 2.80E-231 | 0.57470476 | 0.392 | 0.239 | 1.07E-226 |
| DCC | 4.22E-131 | 0.57032397 | 0.233 | 0.133 | 1.61E-126 |
| ITPKB | 5.11E-128 | 0.56941564 | 0.367 | 0.256 | 1.95E-123 |
| MT-ATP6 | 3.85E-160 | 0.56297416 | 0.81 | 0.695 | 1.47E-155 |
| PLD5 | 7.09E-225 | 0.5604731 | 0.382 | 0.225 | 2.71E-220 |
| SILC1 | 1.52E-285 | 0.55811025 | 0.555 | 0.38 | 5.81E-281 |
| AC109927.1 | 3.67E-174 | 0.55731294 | 0.203 | 0.097 | 1.40E-169 |
| GIT1 | 1.25E-197 | 0.55555132 | 0.265 | 0.139 | 4.77E-193 |
| LRP2 | 3.97E-247 | 0.5468657 | 0.493 | 0.321 | 1.52E-242 |
| TTYH2 | 2.86E-278 | 0.543028 | 0.585 | 0.423 | 1.09E-273 |
| FRYL | 0 | 0.53451685 | 0.881 | 0.753 | 0 |
| MAPT | 0 | 0.52753343 | 0.696 | 0.535 | 0 |
| VAMP2 | 1.09E-187 | 0.52659337 | 0.443 | 0.305 | 4.18E-183 |
| SEM1 | 1.11E-212 | 0.52613206 | 0.323 | 0.182 | 4.24E-208 |
| DIP2C | 0 | 0.52482963 | 0.843 | 0.718 | 0 |
| SLC44A1 | 0 | 0.51017859 | 0.93 | 0.829 | 0 |
| KANSL1L | 1.60E-212 | 0.50758456 | 0.401 | 0.252 | 6.10E-208 |
| FAM13B | 7.67E-194 | 0.50701237 | 0.521 | 0.383 | 2.93E-189 |

| | | | | | |
|---|---|---|---|---|---|
| BBS9 | 1.31E-193 | 0.50238607 | 0.576 | 0.435 | 5.00E-189 |
| ZSWIM6 | 1.04E-252 | 0.49104498 | 0.777 | 0.655 | 3.96E-248 |
| ANAPC16 | 5.74E-232 | 0.48552343 | 0.347 | 0.196 | 2.19E-227 |
| NFASC | 2.32E-298 | 0.4852097 | 0.812 | 0.685 | 8.89E-294 |
| CHST3 | 7.61E-201 | 0.48493309 | 0.249 | 0.125 | 2.91E-196 |
| PANTR1 | 2.14E-161 | 0.47761001 | 0.344 | 0.219 | 8.19E-157 |
| SORCS2 | 8.40E-154 | 0.47758143 | 0.331 | 0.209 | 3.21E-149 |
| NFKBIA | 5.81E-264 | 0.47667521 | 0.162 | 0.048 | 2.22E-259 |
| CHORDC1 | 2.12E-189 | 0.4764048 | 0.246 | 0.127 | 8.10E-185 |
| DHCR24 | 7.19E-145 | 0.47585507 | 0.18 | 0.089 | 2.75E-140 |
| NDUFAF6 | 9.27E-176 | 0.4750227 | 0.313 | 0.185 | 3.54E-171 |
| ZCCHC24 | 1.51E-196 | 0.47469509 | 0.518 | 0.371 | 5.78E-192 |
| PICK1 | 9.80E-195 | 0.46992601 | 0.205 | 0.092 | 3.75E-190 |
| FDFT1 | 4.89E-194 | 0.46928696 | 0.328 | 0.191 | 1.87E-189 |
| L3MBTL4 | 2.32E-173 | 0.46909281 | 0.569 | 0.435 | 8.89E-169 |
| DBNDD2 | 1.10E-136 | 0.46898564 | 0.44 | 0.322 | 4.20E-132 |
| CREBRF | 6.58E-190 | 0.46638597 | 0.333 | 0.198 | 2.51E-185 |
| ARRDC2 | 1.58E-183 | 0.46531922 | 0.159 | 0.063 | 6.06E-179 |
| LHFPL3 | 5.47E-90 | 0.4650943 | 0.133 | 0.068 | 2.09E-85 |
| SNHG25 | 1.79E-104 | 0.46401679 | 0.283 | 0.19 | 6.85E-100 |
| RHOBTB3 | 4.44E-172 | 0.46376014 | 0.331 | 0.204 | 1.70E-167 |
| OLMALINC | 7.88E-211 | 0.46353224 | 0.345 | 0.198 | 3.01E-206 |
| HTRA1 | 2.82E-202 | 0.46274594 | 0.489 | 0.341 | 1.08E-197 |
| OTUD7A | 8.01E-257 | 0.45444485 | 0.744 | 0.606 | 3.06E-252 |
| CEP128 | 2.15E-144 | 0.45403636 | 0.251 | 0.144 | 8.22E-140 |
| CDKN1C | 1.02E-136 | 0.45254057 | 0.238 | 0.138 | 3.92E-132 |
| BBS2 | 9.09E-196 | 0.44850655 | 0.433 | 0.288 | 3.48E-191 |
| PCA3 | 2.61E-151 | 0.44825501 | 0.145 | 0.061 | 9.99E-147 |
| PLCL1 | 3.94E-230 | 0.44658577 | 0.869 | 0.778 | 1.51E-225 |
| EDA | 7.46E-110 | 0.4460313 | 0.21 | 0.122 | 2.85E-105 |
| CLIP4 | 6.08E-162 | 0.44366747 | 0.501 | 0.374 | 2.32E-157 |
| AZGP1 | 4.11E-220 | 0.44359807 | 0.115 | 0.027 | 1.57E-215 |
| SEMA5A | 5.05E-175 | 0.44224457 | 0.222 | 0.108 | 1.93E-170 |
| ELMO1 | 0 | 0.44100242 | 0.919 | 0.817 | 0 |
| SLC22A23 | 7.02E-165 | 0.44094742 | 0.6 | 0.469 | 2.68E-160 |
| SPOCK1 | 6.41E-284 | 0.4397111 | 0.876 | 0.775 | 2.45E-279 |
| AC016821.1 | 4.44E-191 | 0.43961424 | 0.225 | 0.108 | 1.70E-186 |
| GNAO1 | 3.53E-204 | 0.4367769 | 0.748 | 0.622 | 1.35E-199 |
| ATXN1 | 7.17E-224 | 0.43404258 | 0.785 | 0.67 | 2.74E-219 |
| SLC6A8 | 6.72E-129 | 0.43342044 | 0.281 | 0.177 | 2.57E-124 |
| ZNF365 | 9.73E-123 | 0.43288037 | 0.383 | 0.272 | 3.72E-118 |
| AMD1 | 8.01E-143 | 0.43242255 | 0.505 | 0.381 | 3.06E-138 |
| USP31 | 4.04E-153 | 0.43241704 | 0.311 | 0.193 | 1.54E-148 |
| FAM107B | 5.58E-221 | 0.43137712 | 0.728 | 0.582 | 2.13E-216 |
| DOCK1 | 1.13E-209 | 0.42723275 | 0.745 | 0.616 | 4.32E-205 |

| | | | | | |
|---|---|---|---|---|---|
| AL034405.1 | 1.78E-120 | 0.42421109 | 0.198 | 0.109 | 6.82E-116 |
| SLCO1A2 | 6.51E-205 | 0.42382079 | 0.682 | 0.548 | 2.49E-200 |
| HDAC2-AS2 | 2.56E-128 | 0.42261675 | 0.645 | 0.537 | 9.80E-124 |
| ARRB1 | 1.88E-138 | 0.4216007 | 0.214 | 0.115 | 7.17E-134 |
| HSP90AA1 | 6.65E-275 | 0.4197603 | 0.72 | 0.568 | 2.54E-270 |
| IGF1R | 1.61E-119 | 0.41901897 | 0.469 | 0.355 | 6.17E-115 |
| ATG4B | 9.11E-142 | 0.41816477 | 0.246 | 0.141 | 3.48E-137 |
| SPATA13 | 7.88E-131 | 0.41769935 | 0.495 | 0.375 | 3.01E-126 |
| MT-ND4 | 2.61E-66 | 0.41699338 | 0.78 | 0.688 | 9.96E-62 |
| DOCK5 | 2.00E-256 | 0.41657324 | 0.855 | 0.742 | 7.64E-252 |
| DOCK9 | 2.75E-237 | 0.41580839 | 0.746 | 0.614 | 1.05E-232 |
| PPM1B | 1.30E-167 | 0.41558718 | 0.583 | 0.454 | 4.97E-163 |
| TPPP | 1.74E-127 | 0.41428822 | 0.461 | 0.346 | 6.66E-123 |
| NKX6-2 | 1.37E-127 | 0.41317153 | 0.434 | 0.317 | 5.23E-123 |
| NCAM1 | 0 | 0.41185124 | 0.892 | 0.795 | 0 |
| IRS2 | 6.39E-142 | 0.40975194 | 0.214 | 0.115 | 2.44E-137 |
| LINC00320 | 6.02E-142 | 0.40923052 | 0.592 | 0.473 | 2.30E-137 |
| PDS5B | 5.21E-159 | 0.40848815 | 0.451 | 0.318 | 1.99E-154 |
| JADE2 | 3.81E-171 | 0.40653742 | 0.177 | 0.078 | 1.46E-166 |
| GPM6B | 3.33E-253 | 0.40592003 | 0.867 | 0.755 | 1.27E-248 |
| PHYHIPL | 6.58E-146 | 0.40297525 | 0.555 | 0.428 | 2.52E-141 |
| TACC1 | 7.39E-186 | 0.40173053 | 0.583 | 0.443 | 2.82E-181 |
| SLC6A15 | 7.96E-117 | 0.40104366 | 0.18 | 0.097 | 3.04E-112 |
| TAFA5 | 3.79E-139 | 0.40079022 | 0.245 | 0.14 | 1.45E-134 |
| ARHGAP21 | 6.04E-261 | 0.39918677 | 0.833 | 0.725 | 2.31E-256 |
| AC006148.1 | 2.05E-88 | 0.39650951 | 0.349 | 0.255 | 7.84E-84 |
| NCOA7 | 6.24E-153 | 0.39465353 | 0.624 | 0.505 | 2.38E-148 |
| CDH20 | 4.09E-193 | 0.39339385 | 0.788 | 0.66 | 1.56E-188 |
| NFKBIZ | 4.02E-125 | 0.39213818 | 0.225 | 0.13 | 1.54E-120 |
| KLF15 | 2.31E-196 | 0.39185681 | 0.15 | 0.053 | 8.83E-192 |
| PLP1 | 1.32E-188 | 0.38679249 | 0.893 | 0.775 | 5.04E-184 |
| CTSD | 2.71E-101 | 0.38586103 | 0.242 | 0.155 | 1.04E-96 |
| RFTN2 | 2.56E-169 | 0.38584312 | 0.625 | 0.491 | 9.80E-165 |
| YPEL2 | 8.69E-140 | 0.38398344 | 0.539 | 0.413 | 3.32E-135 |
| SOX10 | 2.43E-122 | 0.38388295 | 0.327 | 0.219 | 9.27E-118 |
| MT-ND1 | 4.42E-163 | 0.38062805 | 0.593 | 0.441 | 1.69E-158 |
| MVB12B | 1.65E-162 | 0.38053832 | 0.658 | 0.516 | 6.30E-158 |
| PIM3 | 4.30E-98 | 0.38052923 | 0.132 | 0.066 | 1.64E-93 |
| NDUFS2 | 5.84E-128 | 0.38005013 | 0.279 | 0.175 | 2.23E-123 |
| SLC5A3 | 4.35E-210 | 0.37940211 | 0.104 | 0.023 | 1.66E-205 |
| RASSF4 | 5.14E-134 | 0.37736968 | 0.281 | 0.173 | 1.96E-129 |
| CDK18 | 6.40E-161 | 0.37690826 | 0.648 | 0.517 | 2.45E-156 |
| MAP4K5 | 9.83E-184 | 0.37645475 | 0.736 | 0.616 | 3.76E-179 |
| PAPOLA | 1.61E-147 | 0.3759558 | 0.374 | 0.252 | 6.15E-143 |
| CADM2 | 4.61E-272 | 0.37478097 | 0.942 | 0.869 | 1.76E-267 |

| | | | | | |
|---|---|---|---|---|---|
| MT-ND2 | 8.42E-101 | 0.37357326 | 0.668 | 0.553 | 3.22E-96 |
| ARHGEF10 | 1.04E-154 | 0.37166869 | 0.436 | 0.302 | 3.97E-150 |
| ALDH3A2 | 7.92E-129 | 0.37139953 | 0.195 | 0.104 | 3.03E-124 |
| DNAJB6 | 2.98E-135 | 0.37129715 | 0.269 | 0.163 | 1.14E-130 |
| PTGES3 | 1.87E-163 | 0.37014603 | 0.312 | 0.188 | 7.14E-159 |
| KANSL1 | 1.97E-193 | 0.36927656 | 0.648 | 0.5 | 7.54E-189 |
| MAT2A | 3.93E-115 | 0.36886722 | 0.441 | 0.33 | 1.50E-110 |
| DPYSL5 | 4.23E-92 | 0.36857218 | 0.324 | 0.229 | 1.62E-87 |
| SEMA4B | 2.02E-166 | 0.36833584 | 0.118 | 0.038 | 7.73E-162 |
| RALGDS | 1.26E-114 | 0.36445051 | 0.399 | 0.291 | 4.81E-110 |
| CHDH | 4.66E-213 | 0.36345834 | 0.105 | 0.023 | 1.78E-208 |
| LCORL | 3.01E-107 | 0.36340698 | 0.388 | 0.281 | 1.15E-102 |
| KLF9 | 1.61E-121 | 0.36268993 | 0.181 | 0.095 | 6.16E-117 |
| ITPK1 | 7.99E-111 | 0.36211264 | 0.35 | 0.244 | 3.05E-106 |
| PRKD1 | 1.39E-91 | 0.36142294 | 0.261 | 0.173 | 5.32E-87 |
| ADAM12 | 1.23E-88 | 0.3613983 | 0.237 | 0.154 | 4.70E-84 |
| LAMP1 | 3.72E-92 | 0.36087519 | 0.312 | 0.222 | 1.42E-87 |
| NACC2 | 1.54E-124 | 0.35990197 | 0.181 | 0.094 | 5.89E-120 |
| FNIP2 | 6.91E-90 | 0.35961095 | 0.484 | 0.389 | 2.64E-85 |
| CORO2B | 1.96E-118 | 0.35944892 | 0.352 | 0.239 | 7.49E-114 |
| FLCN | 2.15E-140 | 0.35899933 | 0.187 | 0.094 | 8.21E-136 |
| RELN | 2.54E-127 | 0.35855982 | 0.136 | 0.06 | 9.71E-123 |
| FXR1 | 2.19E-125 | 0.35700727 | 0.402 | 0.289 | 8.38E-121 |
| PPA1 | 1.54E-116 | 0.35688619 | 0.256 | 0.159 | 5.88E-112 |
| CCNH | 6.01E-137 | 0.35676057 | 0.679 | 0.557 | 2.30E-132 |
| CTSF | 1.29E-117 | 0.35662194 | 0.19 | 0.104 | 4.93E-113 |
| ARHGAP17 | 3.72E-120 | 0.35518222 | 0.32 | 0.213 | 1.42E-115 |
| ELF2 | 6.18E-134 | 0.35408099 | 0.599 | 0.48 | 2.36E-129 |
| SGK3 | 7.53E-128 | 0.35386138 | 0.625 | 0.515 | 2.88E-123 |
| TMC6 | 7.51E-96 | 0.35360656 | 0.256 | 0.168 | 2.87E-91 |
| KANK1 | 3.34E-122 | 0.35350946 | 0.597 | 0.476 | 1.28E-117 |
| APOD | 5.01E-74 | 0.35207825 | 0.376 | 0.29 | 1.91E-69 |
| ZNF397 | 2.99E-111 | 0.35192473 | 0.328 | 0.224 | 1.14E-106 |
| ITPR2 | 5.26E-131 | 0.35116733 | 0.242 | 0.138 | 2.01E-126 |
| CAMKMT | 4.18E-84 | 0.35085425 | 0.373 | 0.277 | 1.60E-79 |
| KIZ | 4.03E-121 | 0.35039352 | 0.442 | 0.325 | 1.54E-116 |
| MT-CYB | 5.24E-124 | 0.35026967 | 0.683 | 0.556 | 2.00E-119 |
| P2RX7 | 1.78E-143 | 0.3495763 | 0.544 | 0.422 | 6.79E-139 |
| LIFR | 1.10E-109 | 0.34860375 | 0.517 | 0.407 | 4.19E-105 |
| TLE4 | 6.73E-121 | 0.34802262 | 0.591 | 0.47 | 2.57E-116 |
| RBM48 | 1.53E-156 | 0.34695274 | 0.165 | 0.073 | 5.86E-152 |
| MAP7 | 1.41E-236 | 0.34367935 | 0.888 | 0.793 | 5.39E-232 |
| DIPK2B | 1.26E-116 | 0.34353027 | 0.15 | 0.073 | 4.81E-112 |
| PDE7A | 7.05E-105 | 0.34295997 | 0.395 | 0.289 | 2.69E-100 |
| STAG1 | 6.12E-134 | 0.34292227 | 0.551 | 0.424 | 2.34E-129 |

| | | | | | |
|---|---|---|---|---|---|
| CADM1 | 1.72E-141 | 0.34235618 | 0.752 | 0.644 | 6.57E-137 |
| RELA | 1.55E-116 | 0.34181174 | 0.106 | 0.042 | 5.93E-112 |
| DIP2B | 2.06E-200 | 0.3417597 | 0.848 | 0.737 | 7.88E-196 |
| BCL2L1 | 3.54E-113 | 0.34090485 | 0.349 | 0.242 | 1.35E-108 |
| TMCC2 | 6.88E-104 | 0.3405063 | 0.27 | 0.175 | 2.63E-99 |
| CNOT2 | 8.67E-114 | 0.34019455 | 0.497 | 0.384 | 3.32E-109 |
| DYNC1LI2 | 5.97E-124 | 0.34010991 | 0.415 | 0.299 | 2.28E-119 |
| PPM1K | 4.18E-119 | 0.33996703 | 0.278 | 0.176 | 1.60E-114 |
| ELMO1-AS1 | 5.00E-131 | 0.33758924 | 0.125 | 0.051 | 1.91E-126 |
| MBNL1 | 2.44E-105 | 0.33641059 | 0.605 | 0.504 | 9.33E-101 |
| RANGAP1 | 1.84E-92 | 0.33633247 | 0.229 | 0.145 | 7.02E-88 |
| PHGDH | 1.43E-133 | 0.33545088 | 0.187 | 0.096 | 5.47E-129 |
| CLCN4 | 3.77E-110 | 0.33530327 | 0.245 | 0.153 | 1.44E-105 |
| AC016597.1 | 3.48E-106 | 0.3339743 | 0.429 | 0.321 | 1.33E-101 |
| LANCL1 | 1.70E-125 | 0.33383152 | 0.614 | 0.493 | 6.49E-121 |
| IRF2BP2 | 3.67E-89 | 0.3335659 | 0.232 | 0.151 | 1.40E-84 |
| YPEL3 | 5.78E-94 | 0.33324176 | 0.252 | 0.165 | 2.21E-89 |
| PLPP4 | 1.99E-122 | 0.33267318 | 0.112 | 0.044 | 7.62E-118 |
| PHLDA3 | 2.91E-86 | 0.33226309 | 0.174 | 0.103 | 1.11E-81 |
| PHACTR3 | 5.99E-117 | 0.33112055 | 0.687 | 0.582 | 2.29E-112 |
| CELSR2 | 3.42E-120 | 0.32964935 | 0.202 | 0.113 | 1.31E-115 |
| AC096677.1 | 3.83E-103 | 0.32958291 | 0.173 | 0.095 | 1.46E-98 |
| PPP1R9A | 1.32E-117 | 0.32854564 | 0.546 | 0.43 | 5.04E-113 |
| RAPGEF3 | 1.68E-64 | 0.32736044 | 0.252 | 0.177 | 6.40E-60 |
| DHCR7 | 6.73E-153 | 0.3272027 | 0.125 | 0.046 | 2.57E-148 |
| ITGB8 | 9.48E-94 | 0.32664736 | 0.448 | 0.347 | 3.62E-89 |
| RPS6KA5 | 1.42E-95 | 0.32559239 | 0.425 | 0.325 | 5.42E-91 |
| SH3RF1 | 6.23E-76 | 0.32522535 | 0.345 | 0.256 | 2.38E-71 |
| TECR | 2.66E-112 | 0.32502083 | 0.258 | 0.162 | 1.02E-107 |
| AGAP1 | 2.79E-229 | 0.32422452 | 0.907 | 0.824 | 1.07E-224 |
| PER1 | 4.85E-118 | 0.32418267 | 0.154 | 0.075 | 1.86E-113 |
| BCL6 | 1.34E-121 | 0.32346732 | 0.189 | 0.1 | 5.14E-117 |
| CLCN7 | 1.30E-83 | 0.32273817 | 0.152 | 0.085 | 4.97E-79 |
| C10orf90 | 2.62E-156 | 0.32109016 | 0.831 | 0.729 | 1.00E-151 |
| MT-CO2 | 8.53E-65 | 0.32053776 | 0.848 | 0.77 | 3.26E-60 |
| CDK19 | 5.27E-131 | 0.32022688 | 0.77 | 0.657 | 2.01E-126 |
| STARD10 | 7.04E-110 | 0.32012303 | 0.102 | 0.041 | 2.69E-105 |
| CTNNA1 | 6.84E-87 | 0.32000606 | 0.447 | 0.347 | 2.61E-82 |
| MSI2 | 3.19E-118 | 0.31828465 | 0.407 | 0.288 | 1.22E-113 |
| PTPRJ | 9.98E-96 | 0.31769925 | 0.571 | 0.462 | 3.81E-91 |
| ZNF488 | 5.60E-129 | 0.31724234 | 0.116 | 0.045 | 2.14E-124 |
| SPOCK3 | 3.71E-165 | 0.31674255 | 0.855 | 0.763 | 1.42E-160 |
| FMNL3 | 3.89E-113 | 0.315751 | 0.145 | 0.07 | 1.49E-108 |
| ARAP2 | 6.86E-155 | 0.3151679 | 0.853 | 0.751 | 2.62E-150 |
| SRRM2 | 5.08E-200 | 0.31491024 | 0.581 | 0.424 | 1.94E-195 |

| | | | | | |
|---|---|---|---|---|---|
| HSPH1 | 2.28E-59 | 0.31419908 | 0.168 | 0.108 | 8.71E-55 |
| RBM14 | 2.61E-85 | 0.31271784 | 0.149 | 0.083 | 9.99E-81 |
| TSC22D4 | 4.94E-59 | 0.31237784 | 0.338 | 0.265 | 1.89E-54 |
| PPP4R2 | 7.82E-101 | 0.31189085 | 0.373 | 0.27 | 2.99E-96 |
| ZNF521 | 1.15E-99 | 0.31180653 | 0.21 | 0.125 | 4.40E-95 |
| FAM221A | 7.74E-153 | 0.31145251 | 0.164 | 0.073 | 2.96E-148 |
| C4orf48 | 1.69E-71 | 0.31022325 | 0.233 | 0.16 | 6.45E-67 |
| KIAA1958 | 1.22E-92 | 0.30985908 | 0.446 | 0.343 | 4.65E-88 |
| BTD | 1.72E-82 | 0.30917984 | 0.246 | 0.164 | 6.58E-78 |
| NCKAP5 | 3.64E-117 | 0.30561895 | 0.848 | 0.767 | 1.39E-112 |
| B3GAT1 | 1.47E-95 | 0.30468137 | 0.38 | 0.278 | 5.63E-91 |
| RASSF2 | 6.64E-75 | 0.30397679 | 0.312 | 0.229 | 2.54E-70 |
| PLOD3 | 1.45E-91 | 0.30325391 | 0.165 | 0.093 | 5.54E-87 |
| PRRG1 | 1.67E-83 | 0.30297383 | 0.41 | 0.315 | 6.38E-79 |
| MAGEC3 | 1.31E-77 | 0.30259844 | 0.11 | 0.055 | 5.00E-73 |
| PAK2 | 3.68E-88 | 0.30237779 | 0.31 | 0.219 | 1.41E-83 |
| EVI2A | 3.57E-73 | 0.30070591 | 0.303 | 0.221 | 1.36E-68 |
| LINC00862 | 4.51E-78 | 0.30065683 | 0.313 | 0.225 | 1.72E-73 |
| TF | 1.32E-169 | 0.30034865 | 0.818 | 0.698 | 5.05E-165 |
| ATG13 | 1.18E-92 | 0.2998281 | 0.352 | 0.255 | 4.50E-88 |
| LINC02199 | 5.93E-92 | 0.29824809 | 0.128 | 0.064 | 2.27E-87 |
| TPD52 | 4.31E-84 | 0.29758488 | 0.431 | 0.333 | 1.65E-79 |
| JAKMIP3 | 2.71E-81 | 0.29755156 | 0.305 | 0.219 | 1.04E-76 |
| HDAC4 | 1.51E-94 | 0.29729992 | 0.441 | 0.338 | 5.77E-90 |
| INSR | 2.86E-96 | 0.29701419 | 0.486 | 0.383 | 1.09E-91 |
| ZNF706 | 9.10E-94 | 0.29623785 | 0.403 | 0.301 | 3.48E-89 |
| TMBIM6 | 4.71E-78 | 0.29471594 | 0.371 | 0.284 | 1.80E-73 |
| TMED10 | 1.52E-86 | 0.29464663 | 0.311 | 0.22 | 5.81E-82 |
| LSS | 5.43E-62 | 0.29404454 | 0.397 | 0.318 | 2.08E-57 |
| DNAJB1 | 1.97E-63 | 0.29360903 | 0.108 | 0.059 | 7.54E-59 |
| XPO1 | 5.22E-104 | 0.29285156 | 0.359 | 0.255 | 2.00E-99 |
| EDIL3 | 2.74E-172 | 0.29277704 | 0.902 | 0.803 | 1.05E-167 |
| CPM | 3.33E-74 | 0.29268894 | 0.388 | 0.296 | 1.27E-69 |
| SLC22A17 | 6.82E-97 | 0.29248345 | 0.275 | 0.183 | 2.61E-92 |
| PWWP3A | 3.18E-89 | 0.29187528 | 0.306 | 0.214 | 1.22E-84 |
| CNTN2 | 3.13E-104 | 0.29143255 | 0.709 | 0.592 | 1.20E-99 |
| PACRG-AS3 | 3.39E-69 | 0.29106268 | 0.197 | 0.128 | 1.30E-64 |
| PNISR | 6.73E-155 | 0.29055103 | 0.566 | 0.431 | 2.57E-150 |
| KLF13 | 1.24E-81 | 0.28881661 | 0.303 | 0.216 | 4.73E-77 |
| ABTB2 | 3.20E-49 | 0.28844705 | 0.185 | 0.128 | 1.22E-44 |
| AGO2 | 5.85E-81 | 0.28844656 | 0.412 | 0.315 | 2.24E-76 |
| C15orf41 | 1.46E-72 | 0.28766806 | 0.244 | 0.167 | 5.59E-68 |
| ITPRID2 | 4.76E-75 | 0.28761065 | 0.222 | 0.147 | 1.82E-70 |
| CTDSP2 | 3.90E-85 | 0.2869182 | 0.209 | 0.131 | 1.49E-80 |
| BANP | 6.26E-94 | 0.28345737 | 0.171 | 0.096 | 2.39E-89 |

| | | | | | |
|---|---|---|---|---|---|
| CXXC4 | 1.69E-82 | 0.28191979 | 0.155 | 0.088 | 6.48E-78 |
| ERBB3 | 1.63E-67 | 0.28179093 | 0.473 | 0.388 | 6.21E-63 |
| SYNGR2 | 1.24E-68 | 0.28098941 | 0.154 | 0.094 | 4.72E-64 |
| TOB2 | 8.11E-91 | 0.27954705 | 0.279 | 0.188 | 3.10E-86 |
| CREBBP | 2.61E-88 | 0.27856784 | 0.399 | 0.301 | 9.99E-84 |
| CLDND1 | 3.76E-81 | 0.27843619 | 0.687 | 0.589 | 1.44E-76 |
| ERMN | 1.05E-57 | 0.27802901 | 0.408 | 0.331 | 4.03E-53 |
| TIAM2 | 1.04E-90 | 0.27780274 | 0.311 | 0.216 | 3.98E-86 |
| SIRT2 | 2.89E-73 | 0.27741835 | 0.368 | 0.281 | 1.11E-68 |
| STARD7 | 2.25E-79 | 0.27735195 | 0.206 | 0.132 | 8.60E-75 |
| FOXO1 | 1.04E-56 | 0.27585473 | 0.344 | 0.267 | 3.96E-52 |
| PIP4K2A | 3.33E-193 | 0.27564439 | 0.927 | 0.823 | 1.27E-188 |
| KDM6A | 1.27E-97 | 0.27427362 | 0.581 | 0.475 | 4.84E-93 |
| RALGAPA2 | 1.12E-71 | 0.27389643 | 0.331 | 0.247 | 4.30E-67 |
| KLHL24 | 1.54E-78 | 0.27381548 | 0.441 | 0.349 | 5.88E-74 |
| PMP22 | 3.41E-61 | 0.27381444 | 0.393 | 0.314 | 1.30E-56 |
| TFEB | 2.37E-64 | 0.2736576 | 0.392 | 0.309 | 9.04E-60 |
| RNF144A | 6.09E-95 | 0.27362712 | 0.599 | 0.493 | 2.33E-90 |
| MAP2K4 | 1.03E-81 | 0.27194117 | 0.377 | 0.284 | 3.96E-77 |
| IRAK2 | 2.10E-81 | 0.27165456 | 0.163 | 0.095 | 8.03E-77 |
| ADAM10 | 6.85E-73 | 0.27050424 | 0.52 | 0.431 | 2.62E-68 |
| ELF1 | 6.84E-61 | 0.27046627 | 0.321 | 0.243 | 2.61E-56 |
| TANC1 | 1.42E-74 | 0.26992167 | 0.38 | 0.288 | 5.41E-70 |
| GPC5 | 5.19E-29 | 0.26872725 | 0.145 | 0.107 | 1.98E-24 |
| TSEN15 | 8.04E-69 | 0.26872081 | 0.259 | 0.183 | 3.07E-64 |
| SOX8 | 6.58E-36 | 0.26738157 | 0.272 | 0.218 | 2.52E-31 |
| IP6K3 | 4.71E-117 | 0.26721322 | 0.118 | 0.049 | 1.80E-112 |
| UTP23 | 9.87E-72 | 0.26711251 | 0.164 | 0.099 | 3.77E-67 |
| CYP51A1 | 3.10E-70 | 0.26689601 | 0.156 | 0.094 | 1.19E-65 |
| AC012494.1 | 1.48E-65 | 0.26633031 | 0.522 | 0.427 | 5.66E-61 |
| CNTNAP2 | 9.96E-83 | 0.26616119 | 0.676 | 0.61 | 3.81E-78 |
| GINM1 | 6.89E-90 | 0.26614283 | 0.204 | 0.124 | 2.63E-85 |
| AC106791.1 | 1.26E-60 | 0.26529222 | 0.364 | 0.284 | 4.81E-56 |
| GLTP | 4.38E-68 | 0.26500934 | 0.321 | 0.24 | 1.67E-63 |
| MIR29B2CHG | 1.87E-92 | 0.26479137 | 0.205 | 0.124 | 7.16E-88 |
| CBR1 | 7.14E-46 | 0.26425347 | 0.264 | 0.204 | 2.73E-41 |
| MICAL3 | 1.14E-83 | 0.26401683 | 0.613 | 0.52 | 4.38E-79 |
| NDE1 | 7.10E-80 | 0.26366138 | 0.358 | 0.266 | 2.71E-75 |
| VPS13C | 3.90E-94 | 0.26324663 | 0.591 | 0.491 | 1.49E-89 |
| ECE1 | 5.36E-73 | 0.26273388 | 0.192 | 0.122 | 2.05E-68 |
| GRB10 | 8.51E-85 | 0.26160678 | 0.33 | 0.235 | 3.25E-80 |
| DPYSL2 | 1.16E-91 | 0.26148501 | 0.701 | 0.599 | 4.45E-87 |
| RDX | 1.75E-69 | 0.26047507 | 0.534 | 0.448 | 6.67E-65 |
| PPFIBP2 | 1.80E-55 | 0.26024519 | 0.299 | 0.228 | 6.88E-51 |
| FUT9 | 7.12E-94 | 0.26005354 | 0.366 | 0.261 | 2.72E-89 |

| | | | | | |
|---|---|---|---|---|---|
| HEPN1 | 1.08E-39 | 0.25991627 | 0.159 | 0.113 | 4.14E-35 |
| NUTM2A-AS | 1.90E-67 | 0.2597921 | 0.498 | 0.41 | 7.25E-63 |
| LGR4 | 1.24E-81 | 0.25925586 | 0.157 | 0.09 | 4.76E-77 |
| CLDN11 | 1.12E-90 | 0.25874721 | 0.673 | 0.548 | 4.30E-86 |
| ZCCHC2 | 6.81E-64 | 0.25787039 | 0.194 | 0.128 | 2.60E-59 |
| SELENOW | 4.14E-62 | 0.25770705 | 0.368 | 0.289 | 1.58E-57 |
| SON | 1.97E-93 | 0.25709335 | 0.451 | 0.348 | 7.51E-89 |
| HLA-A | 1.09E-120 | 0.25680234 | 0.1 | 0.036 | 4.16E-116 |
| LDLRAD3 | 1.38E-38 | 0.25612059 | 0.251 | 0.193 | 5.28E-34 |
| MSMO1 | 5.53E-77 | 0.25571762 | 0.115 | 0.059 | 2.12E-72 |
| ZMYND8 | 3.49E-74 | 0.25555044 | 0.442 | 0.351 | 1.33E-69 |
| CAMK2N1 | 8.63E-70 | 0.25509319 | 0.397 | 0.311 | 3.30E-65 |
| ARHGAP23 | 2.50E-72 | 0.25491097 | 0.475 | 0.384 | 9.55E-68 |
| PACS2 | 2.63E-63 | 0.25483367 | 0.448 | 0.364 | 1.01E-58 |
| FIGN | 7.05E-50 | 0.25445199 | 0.429 | 0.353 | 2.70E-45 |
| HIP1 | 8.95E-87 | 0.25403357 | 0.694 | 0.596 | 3.42E-82 |
| ENOX2 | 2.42E-39 | 0.25342371 | 0.407 | 0.345 | 9.23E-35 |
| ACSL1 | 5.19E-54 | 0.25287032 | 0.284 | 0.216 | 1.99E-49 |
| PTP4A2 | 2.66E-63 | 0.25268011 | 0.399 | 0.317 | 1.02E-58 |
| TTC1 | 3.00E-82 | 0.25240095 | 0.199 | 0.124 | 1.15E-77 |
| LGALS3BP | 3.69E-64 | 0.25217577 | 0.134 | 0.079 | 1.41E-59 |
| SNHG5 | 1.76E-76 | 0.2520811 | 0.201 | 0.128 | 6.72E-72 |
| TMEM14B | 6.38E-80 | 0.25204655 | 0.305 | 0.216 | 2.44E-75 |
| BNIP3L | 9.05E-61 | 0.25174247 | 0.332 | 0.255 | 3.46E-56 |
| STIP1 | 1.22E-93 | 0.25161036 | 0.136 | 0.069 | 4.67E-89 |
| FSCN1 | 2.10E-59 | 0.25154077 | 0.271 | 0.2 | 8.01E-55 |
| CHADL | 6.30E-56 | 0.25003954 | 0.19 | 0.128 | 2.41E-51 |
| SH3GL3 | 8.86E-36 | -0.2510804 | 0.759 | 0.718 | 3.39E-31 |
| SPTBN4 | 7.67E-08 | -0.2512673 | 0.089 | 0.104 | 0.00293125 |
| AC077690.1 | 4.65E-33 | -0.2516095 | 0.115 | 0.157 | 1.78E-28 |
| SPATA6 | 2.86E-23 | -0.2518976 | 0.402 | 0.435 | 1.09E-18 |
| FUT8 | 4.31E-45 | -0.2531397 | 0.76 | 0.733 | 1.65E-40 |
| GFOD1 | 7.43E-35 | -0.2531606 | 0.108 | 0.15 | 2.84E-30 |
| LINC00342 | 1.66E-22 | -0.2532874 | 0.227 | 0.267 | 6.34E-18 |
| LCNL1 | 1.27E-47 | -0.2540923 | 0.082 | 0.128 | 4.84E-43 |
| ZNF189 | 1.41E-36 | -0.2541791 | 0.145 | 0.192 | 5.41E-32 |
| FMN1 | 4.15E-09 | -0.2545184 | 0.101 | 0.119 | 0.00015844 |
| EPB41L3 | 2.93E-17 | -0.2549188 | 0.197 | 0.23 | 1.12E-12 |
| MOG | 1.40E-44 | -0.257218 | 0.594 | 0.613 | 5.34E-40 |
| NTM | 2.83E-51 | -0.2573783 | 0.726 | 0.746 | 1.08E-46 |
| LINC00461 | 4.63E-31 | -0.2582423 | 0.204 | 0.251 | 1.77E-26 |
| PTK2 | 4.56E-83 | -0.2599082 | 0.821 | 0.799 | 1.74E-78 |
| BDNF-AS | 4.26E-46 | -0.2606278 | 0.13 | 0.183 | 1.63E-41 |
| PPM1L | 1.84E-19 | -0.2607953 | 0.14 | 0.172 | 7.02E-15 |
| MYO5A | 2.48E-30 | -0.2617603 | 0.495 | 0.531 | 9.47E-26 |

| | | | | | |
|---|---|---|---|---|---|
| CCDC102B | 2.48E-47 | -0.2622343 | 0.072 | 0.116 | 9.47E-43 |
| PCBP4 | 6.89E-47 | -0.2628541 | 0.195 | 0.252 | 2.63E-42 |
| STMN4 | 9.76E-52 | -0.2635852 | 0.23 | 0.297 | 3.73E-47 |
| DAPK2 | 1.89E-28 | -0.2636571 | 0.2 | 0.243 | 7.22E-24 |
| TRPC1 | 8.01E-32 | -0.2637048 | 0.265 | 0.314 | 3.06E-27 |
| ANO4 | 1.86E-37 | -0.2637093 | 0.687 | 0.681 | 7.12E-33 |
| MEF2A | 3.59E-34 | -0.2639464 | 0.371 | 0.422 | 1.37E-29 |
| ZNF518A | 5.38E-35 | -0.2648333 | 0.28 | 0.332 | 2.06E-30 |
| GIPC1 | 1.26E-28 | -0.2654106 | 0.129 | 0.169 | 4.82E-24 |
| KIAA1324L | 9.64E-29 | -0.2654419 | 0.41 | 0.449 | 3.68E-24 |
| CREB5 | 2.62E-60 | -0.2660065 | 0.763 | 0.736 | 1.00E-55 |
| TTTY14 | 3.23E-46 | -0.2666677 | 0.109 | 0.16 | 1.24E-41 |
| LINC01184 | 5.76E-31 | -0.2680978 | 0.287 | 0.334 | 2.20E-26 |
| ANKS1B | 1.14E-16 | -0.2686665 | 0.811 | 0.792 | 4.37E-12 |
| PKM | 1.27E-07 | -0.2695602 | 0.103 | 0.119 | 0.00485786 |
| SATB2 | 4.69E-20 | -0.2696482 | 0.085 | 0.113 | 1.79E-15 |
| PPP2R3A | 9.68E-37 | -0.2703236 | 0.344 | 0.396 | 3.70E-32 |
| EXOC6B | 2.62E-74 | -0.2706127 | 0.788 | 0.769 | 1.00E-69 |
| AC022182.1 | 5.55E-65 | -0.2710791 | 0.054 | 0.101 | 2.12E-60 |
| SPANXA2-OT | 2.32E-28 | -0.2719639 | 0.091 | 0.126 | 8.86E-24 |
| GRAMD2B | 2.92E-57 | -0.2722844 | 0.225 | 0.294 | 1.12E-52 |
| HS3ST5 | 1.46E-27 | -0.2726634 | 0.468 | 0.498 | 5.57E-23 |
| BTBD9 | 5.39E-39 | -0.2739535 | 0.307 | 0.367 | 2.06E-34 |
| POGZ | 2.10E-49 | -0.2741785 | 0.504 | 0.547 | 8.01E-45 |
| AC108721.2 | 5.84E-31 | -0.2753232 | 0.301 | 0.35 | 2.23E-26 |
| C5orf64 | 2.69E-29 | -0.2775622 | 0.299 | 0.343 | 1.03E-24 |
| PLEKHH1 | 2.51E-76 | -0.2787638 | 0.754 | 0.732 | 9.61E-72 |
| AC115622.1 | 4.11E-53 | -0.2799403 | 0.067 | 0.113 | 1.57E-48 |
| NSF | 5.19E-15 | -0.2826948 | 0.095 | 0.119 | 1.98E-10 |
| TMEFF1 | 2.96E-31 | -0.2827224 | 0.255 | 0.302 | 1.13E-26 |
| GPATCH2 | 2.07E-40 | -0.2828791 | 0.082 | 0.123 | 7.92E-36 |
| CASTOR3 | 5.13E-39 | -0.285238 | 0.345 | 0.396 | 1.96E-34 |
| MAP1LC3A | 2.03E-79 | -0.2852761 | 0.068 | 0.127 | 7.75E-75 |
| LINC01278 | 1.33E-22 | -0.2873891 | 0.145 | 0.18 | 5.08E-18 |
| PROX1-AS1 | 1.85E-58 | -0.2885179 | 0.069 | 0.118 | 7.07E-54 |
| AC138627.1 | 1.61E-44 | -0.2892626 | 0.109 | 0.158 | 6.16E-40 |
| ADAMTS18 | 5.32E-66 | -0.2912697 | 0.122 | 0.187 | 2.03E-61 |
| DMXL2 | 1.20E-34 | -0.2924927 | 0.107 | 0.149 | 4.57E-30 |
| FAR1 | 2.19E-35 | -0.2930787 | 0.44 | 0.476 | 8.37E-31 |
| RABGEF1 | 8.74E-37 | -0.2936573 | 0.401 | 0.447 | 3.34E-32 |
| SLC1A3 | 3.02E-33 | -0.2973236 | 0.421 | 0.459 | 1.15E-28 |
| SCMH1 | 8.93E-16 | -0.2995864 | 0.142 | 0.169 | 3.41E-11 |
| AC026341.1 | 2.05E-42 | -0.3001242 | 0.091 | 0.135 | 7.84E-38 |
| MIR181A1HG | 8.88E-41 | -0.3026454 | 0.613 | 0.618 | 3.39E-36 |
| TNIK | 1.85E-33 | -0.3029473 | 0.354 | 0.403 | 7.06E-29 |

| | | | | | |
|---|---|---|---|---|---|
| AIG1 | 8.13E-39 | -0.303572 | 0.248 | 0.302 | 3.11E-34 |
| MIPEP | 4.17E-49 | -0.3035811 | 0.124 | 0.177 | 1.60E-44 |
| SCFD2 | 5.43E-54 | -0.3061258 | 0.24 | 0.309 | 2.08E-49 |
| TNNI3K | 1.68E-61 | -0.3075637 | 0.118 | 0.178 | 6.42E-57 |
| RBFOX2 | 5.99E-21 | -0.3113615 | 0.259 | 0.295 | 2.29E-16 |
| CCDC88A | 6.05E-99 | -0.3143223 | 0.743 | 0.763 | 2.31E-94 |
| XKR6 | 8.23E-23 | -0.3156581 | 0.108 | 0.14 | 3.15E-18 |
| AL050309.1 | 6.08E-43 | -0.3162998 | 0.117 | 0.166 | 2.33E-38 |
| GOLGA8A | 6.27E-71 | -0.317839 | 0.054 | 0.104 | 2.40E-66 |
| CERCAM | 3.71E-49 | -0.3191666 | 0.661 | 0.655 | 1.42E-44 |
| CACNB2 | 2.26E-12 | -0.3226762 | 0.083 | 0.103 | 8.65E-08 |
| LINC01376 | 1.77E-77 | -0.3233646 | 0.061 | 0.116 | 6.77E-73 |
| R3HDM1 | 2.75E-18 | -0.3240054 | 0.326 | 0.361 | 1.05E-13 |
| BCAT1 | 7.69E-100 | -0.3249052 | 0.051 | 0.111 | 2.94E-95 |
| CPB2-AS1 | 6.83E-60 | -0.3265059 | 0.384 | 0.452 | 2.61E-55 |
| PLS3 | 1.57E-53 | -0.3279916 | 0.162 | 0.223 | 6.01E-49 |
| AC006059.1 | 5.98E-29 | -0.328123 | 0.274 | 0.316 | 2.29E-24 |
| PTK2B | 1.34E-37 | -0.3302392 | 0.072 | 0.109 | 5.12E-33 |
| LINC00609 | 5.69E-47 | -0.3305383 | 0.457 | 0.504 | 2.17E-42 |
| SNAP91 | 2.28E-28 | -0.3325059 | 0.09 | 0.124 | 8.73E-24 |
| PRKCB | 3.71E-16 | -0.332621 | 0.215 | 0.246 | 1.42E-11 |
| FOLH1 | 2.15E-50 | -0.3329095 | 0.451 | 0.493 | 8.22E-46 |
| EXT1 | 5.43E-39 | -0.3343327 | 0.212 | 0.268 | 2.07E-34 |
| KCNH1 | 2.14E-21 | -0.3347087 | 0.174 | 0.211 | 8.18E-17 |
| SH3D19 | 1.93E-87 | -0.3360343 | 0.599 | 0.646 | 7.38E-83 |
| LINC01102 | 7.55E-89 | -0.3361369 | 0.077 | 0.141 | 2.88E-84 |
| BCAS1 | 2.90E-101 | -0.3380317 | 0.647 | 0.674 | 1.11E-96 |
| CCDC7 | 8.36E-54 | -0.3383047 | 0.2 | 0.264 | 3.19E-49 |
| CMTM5 | 1.03E-107 | -0.3390222 | 0.107 | 0.186 | 3.95E-103 |
| CAMK1D | 1.25E-17 | -0.3391003 | 0.082 | 0.107 | 4.77E-13 |
| FAM13C | 1.30E-61 | -0.3410576 | 0.514 | 0.554 | 4.97E-57 |
| ABCA5 | 3.18E-68 | -0.3413137 | 0.136 | 0.203 | 1.21E-63 |
| LINC01877 | 6.62E-68 | -0.341591 | 0.085 | 0.141 | 2.53E-63 |
| ATG10 | 2.15E-65 | -0.3437534 | 0.159 | 0.227 | 8.23E-61 |
| ITFG1 | 1.08E-59 | -0.3443955 | 0.429 | 0.494 | 4.14E-55 |
| SOX5 | 2.58E-15 | -0.3459317 | 0.101 | 0.125 | 9.85E-11 |
| AK5 | 2.12E-79 | -0.3474681 | 0.661 | 0.691 | 8.10E-75 |
| KCNAB1 | 1.26E-35 | -0.3483766 | 0.104 | 0.145 | 4.80E-31 |
| SH3TC2-DT | 1.51E-62 | -0.3489472 | 0.154 | 0.22 | 5.78E-58 |
| FBXL17 | 3.09E-31 | -0.3500612 | 0.31 | 0.358 | 1.18E-26 |
| PPP1R12B | 3.56E-69 | -0.3503747 | 0.531 | 0.576 | 1.36E-64 |
| AC092924.2 | 2.05E-61 | -0.3503892 | 0.183 | 0.25 | 7.82E-57 |
| AC021231.1 | 1.84E-85 | -0.3509786 | 0.058 | 0.115 | 7.04E-81 |
| EVI5 | 1.83E-70 | -0.3518397 | 0.33 | 0.409 | 7.01E-66 |
| ATP2B1 | 2.34E-08 | -0.3519175 | 0.206 | 0.224 | 0.00089446 |

| | | | | | |
|---|---|---|---|---|---|
| LINC01006 | 7.48E-65 | -0.352495 | 0.106 | 0.164 | 2.86E-60 |
| SAMD4B | 2.52E-62 | -0.3528827 | 0.343 | 0.411 | 9.65E-58 |
| DNM1L | 7.97E-69 | -0.3530194 | 0.209 | 0.283 | 3.05E-64 |
| SV2B | 1.58E-12 | -0.353079 | 0.116 | 0.138 | 6.04E-08 |
| LUC7L2 | 4.48E-70 | -0.3534821 | 0.375 | 0.446 | 1.71E-65 |
| PHLDB1 | 2.89E-112 | -0.3555952 | 0.575 | 0.632 | 1.11E-107 |
| INPP4B | 6.67E-49 | -0.3557231 | 0.092 | 0.141 | 2.55E-44 |
| ICA1L | 5.58E-64 | -0.3558878 | 0.081 | 0.134 | 2.13E-59 |
| KCNQ3 | 9.02E-20 | -0.3574537 | 0.089 | 0.116 | 3.45E-15 |
| AC107223.1 | 2.31E-54 | -0.3598436 | 0.063 | 0.108 | 8.83E-50 |
| RBM25 | 4.25E-44 | -0.3622996 | 0.312 | 0.369 | 1.62E-39 |
| JAZF1 | 6.70E-74 | -0.3634204 | 0.308 | 0.39 | 2.56E-69 |
| TSPAN7 | 3.84E-29 | -0.3639618 | 0.07 | 0.102 | 1.47E-24 |
| COL16A1 | 1.27E-46 | -0.3642488 | 0.227 | 0.283 | 4.87E-42 |
| TPM4 | 2.41E-36 | -0.3647244 | 0.143 | 0.189 | 9.20E-32 |
| SIK3 | 2.04E-180 | -0.3652966 | 0.907 | 0.849 | 7.81E-176 |
| RASAL1 | 6.10E-84 | -0.3657311 | 0.135 | 0.209 | 2.33E-79 |
| AGPAT4 | 7.36E-78 | -0.3680195 | 0.458 | 0.531 | 2.81E-73 |
| MRTFB | 1.00E-31 | -0.3682805 | 0.077 | 0.112 | 3.83E-27 |
| POLR2F | 1.20E-88 | -0.3687978 | 0.618 | 0.663 | 4.57E-84 |
| KCTD1 | 5.63E-53 | -0.3689521 | 0.116 | 0.17 | 2.15E-48 |
| AL078590.2 | 3.14E-41 | -0.3706651 | 0.268 | 0.32 | 1.20E-36 |
| ANK2 | 1.49E-197 | -0.3711662 | 0.86 | 0.858 | 5.69E-193 |
| DOCK3 | 1.23E-94 | -0.3712158 | 0.627 | 0.681 | 4.70E-90 |
| CNTN1 | 1.54E-14 | -0.3712227 | 0.137 | 0.164 | 5.88E-10 |
| TMEFF2 | 4.09E-70 | -0.3716875 | 0.7 | 0.702 | 1.56E-65 |
| DMD | 1.28E-56 | -0.3717141 | 0.564 | 0.613 | 4.90E-52 |
| MAP1B | 3.89E-23 | -0.3724658 | 0.384 | 0.417 | 1.49E-18 |
| ENOSF1 | 8.53E-70 | -0.372843 | 0.323 | 0.397 | 3.26E-65 |
| SLC6A1 | 3.30E-46 | -0.3773948 | 0.314 | 0.369 | 1.26E-41 |
| SYT16 | 4.11E-52 | -0.3800909 | 0.058 | 0.1 | 1.57E-47 |
| SAMD12 | 8.74E-79 | -0.3801915 | 0.588 | 0.638 | 3.34E-74 |
| AATK | 1.89E-116 | -0.3817194 | 0.631 | 0.662 | 7.22E-112 |
| LINC01630 | 1.96E-74 | -0.3846299 | 0.387 | 0.462 | 7.50E-70 |
| DANT2 | 3.58E-29 | -0.3856258 | 0.215 | 0.259 | 1.37E-24 |
| LDLRAD4 | 1.41E-70 | -0.3907666 | 0.337 | 0.411 | 5.40E-66 |
| GMDS-DT | 6.39E-71 | -0.3925164 | 0.221 | 0.297 | 2.44E-66 |
| NCALD | 6.73E-36 | -0.3927182 | 0.081 | 0.119 | 2.57E-31 |
| ROR1 | 9.03E-35 | -0.3927853 | 0.169 | 0.216 | 3.45E-30 |
| RNF217-AS1 | 2.25E-67 | -0.3953625 | 0.071 | 0.124 | 8.59E-63 |
| MARCKSL1 | 6.04E-33 | -0.3955976 | 0.293 | 0.333 | 2.31E-28 |
| KCND2 | 5.45E-22 | -0.3960098 | 0.074 | 0.102 | 2.08E-17 |
| FMN2 | 5.33E-33 | -0.3969694 | 0.075 | 0.11 | 2.04E-28 |
| GRIA2 | 1.16E-16 | -0.3984321 | 0.395 | 0.41 | 4.44E-12 |
| TESK2 | 1.96E-89 | -0.4009459 | 0.234 | 0.321 | 7.50E-85 |

| | | | | | |
|---|---|---|---|---|---|
| BABAM2 | 1.47E-82 | -0.4045677 | 0.192 | 0.274 | 5.64E-78 |
| HMGN1 | 9.18E-48 | -0.4077656 | 0.113 | 0.164 | 3.51E-43 |
| NRBP2 | 3.58E-126 | -0.4078885 | 0.139 | 0.233 | 1.37E-121 |
| FOCAD | 2.84E-48 | -0.4095971 | 0.111 | 0.162 | 1.09E-43 |
| MACROD2 | 8.48E-31 | -0.410084 | 0.185 | 0.23 | 3.24E-26 |
| NKAIN2 | 9.03E-133 | -0.4105516 | 0.8 | 0.81 | 3.45E-128 |
| FBXO7 | 1.59E-100 | -0.4108226 | 0.232 | 0.322 | 6.07E-96 |
| PSD3 | 2.07E-67 | -0.4110716 | 0.538 | 0.579 | 7.92E-63 |
| UNC5C | 2.08E-101 | -0.4124916 | 0.845 | 0.807 | 7.93E-97 |
| PAK1 | 9.05E-63 | -0.4130302 | 0.155 | 0.219 | 3.46E-58 |
| LSAMP | 1.64E-167 | -0.4130598 | 0.717 | 0.792 | 6.28E-163 |
| RORA | 3.09E-78 | -0.4163446 | 0.267 | 0.345 | 1.18E-73 |
| ASPA | 1.50E-87 | -0.4195529 | 0.43 | 0.496 | 5.75E-83 |
| AZIN1-AS1 | 1.93E-106 | -0.4239353 | 0.115 | 0.197 | 7.39E-102 |
| SLC22A15 | 2.36E-81 | -0.4256382 | 0.426 | 0.492 | 9.01E-77 |
| ERC2 | 3.86E-21 | -0.4259868 | 0.196 | 0.232 | 1.48E-16 |
| PDE8A | 5.56E-163 | -0.4264215 | 0.758 | 0.748 | 2.12E-158 |
| HMGN2 | 3.23E-28 | -0.4266873 | 0.097 | 0.132 | 1.23E-23 |
| AC104596.1 | 4.87E-66 | -0.4282483 | 0.183 | 0.25 | 1.86E-61 |
| SLC4A8 | 3.05E-80 | -0.4288184 | 0.255 | 0.336 | 1.17E-75 |
| DSCAM | 8.99E-16 | -0.4320714 | 0.143 | 0.17 | 3.44E-11 |
| LURAP1L-AS | 1.20E-75 | -0.4322138 | 0.234 | 0.312 | 4.57E-71 |
| GRIN1 | 6.26E-13 | -0.4335734 | 0.106 | 0.127 | 2.39E-08 |
| PCLO | 6.79E-24 | -0.4373468 | 0.086 | 0.116 | 2.59E-19 |
| ARHGAP26 | 8.95E-19 | -0.4399148 | 0.096 | 0.123 | 3.42E-14 |
| PLPPR1 | 1.27E-40 | -0.4405212 | 0.396 | 0.436 | 4.87E-36 |
| PDE10A | 3.43E-67 | -0.4417431 | 0.082 | 0.137 | 1.31E-62 |
| RASGRF2 | 1.06E-83 | -0.4424728 | 0.292 | 0.375 | 4.03E-79 |
| OPALIN | 2.55E-97 | -0.4432234 | 0.409 | 0.484 | 9.73E-93 |
| RPS28 | 1.94E-14 | -0.4468351 | 0.246 | 0.205 | 7.43E-10 |
| ASTN2 | 5.44E-92 | -0.446905 | 0.468 | 0.54 | 2.08E-87 |
| MAST4 | 1.20E-72 | -0.4478698 | 0.13 | 0.198 | 4.60E-68 |
| TMEM132B | 7.64E-14 | -0.4495925 | 0.092 | 0.114 | 2.92E-09 |
| AL359091.1 | 9.20E-101 | -0.4512078 | 0.498 | 0.564 | 3.52E-96 |
| RIN2 | 1.49E-96 | -0.4522409 | 0.293 | 0.384 | 5.69E-92 |
| TP53TG5 | 1.78E-112 | -0.4534893 | 0.272 | 0.367 | 6.79E-108 |
| DNM1 | 8.23E-27 | -0.4571143 | 0.092 | 0.124 | 3.15E-22 |
| RIMS1 | 6.47E-40 | -0.4590931 | 0.079 | 0.119 | 2.47E-35 |
| CACNA1C | 1.33E-34 | -0.4631846 | 0.11 | 0.152 | 5.09E-30 |
| LINC00882 | 1.61E-121 | -0.4643687 | 0.423 | 0.521 | 6.17E-117 |
| LINC01170 | 2.24E-68 | -0.4650515 | 0.403 | 0.47 | 8.56E-64 |
| SEC14L5 | 1.02E-94 | -0.4691685 | 0.343 | 0.427 | 3.91E-90 |
| H3F3A | 1.32E-08 | -0.4715818 | 0.232 | 0.202 | 0.000503 |
| EFCAB11 | 1.74E-100 | -0.4740436 | 0.128 | 0.209 | 6.66E-96 |
| AFF3 | 8.76E-36 | -0.4770877 | 0.081 | 0.119 | 3.35E-31 |

| | | | | | |
|---|---|---|---|---|---|
| LTBP1 | 1.37E-103 | -0.4777799 | 0.177 | 0.267 | 5.23E-99 |
| MAP2 | 5.89E-32 | -0.4793982 | 0.112 | 0.151 | 2.25E-27 |
| CNTNAP5 | 5.83E-20 | -0.4833077 | 0.076 | 0.101 | 2.23E-15 |
| FNBP1 | 4.84E-211 | -0.4863753 | 0.65 | 0.702 | 1.85E-206 |
| RPS25 | 1.26E-07 | -0.4890297 | 0.134 | 0.15 | 0.00481357 |
| ARPP21 | 9.38E-49 | -0.4910835 | 0.227 | 0.283 | 3.59E-44 |
| TMEM232 | 1.44E-116 | -0.4999615 | 0.082 | 0.159 | 5.52E-112 |
| TARBP1 | 1.63E-125 | -0.5031441 | 0.117 | 0.205 | 6.22E-121 |
| KCTD16 | 1.85E-24 | -0.5032041 | 0.08 | 0.11 | 7.07E-20 |
| KCNH8 | 1.02E-226 | -0.5039539 | 0.721 | 0.735 | 3.91E-222 |
| ZFYVE16 | 5.63E-168 | -0.5063689 | 0.472 | 0.57 | 2.15E-163 |
| SNTG1 | 4.69E-22 | -0.5103569 | 0.119 | 0.151 | 1.79E-17 |
| AC079352.1 | 7.73E-125 | -0.5138368 | 0.31 | 0.42 | 2.96E-120 |
| GRM7 | 1.55E-26 | -0.5162678 | 0.099 | 0.133 | 5.92E-22 |
| ATP8A2 | 5.12E-103 | -0.5206263 | 0.112 | 0.191 | 1.96E-98 |
| CHRM3 | 3.25E-41 | -0.5207203 | 0.076 | 0.116 | 1.24E-36 |
| CNTN5 | 1.53E-27 | -0.5254273 | 0.105 | 0.14 | 5.87E-23 |
| GALNT13 | 1.38E-163 | -0.5272745 | 0.359 | 0.483 | 5.29E-159 |
| CDH12 | 7.42E-59 | -0.5293908 | 0.056 | 0.101 | 2.84E-54 |
| SYT14 | 1.45E-153 | -0.5318992 | 0.147 | 0.253 | 5.53E-149 |
| SEMA3B | 2.69E-156 | -0.5361756 | 0.297 | 0.41 | 1.03E-151 |
| GRIN2A | 1.59E-80 | -0.5362034 | 0.201 | 0.282 | 6.09E-76 |
| CLSTN2 | 2.38E-25 | -0.537884 | 0.075 | 0.105 | 9.12E-21 |
| TMEM178B | 2.72E-77 | -0.5458823 | 0.1 | 0.164 | 1.04E-72 |
| PHACTR1 | 8.60E-38 | -0.5464901 | 0.287 | 0.338 | 3.29E-33 |
| AC233296.1 | 6.03E-127 | -0.54684 | 0.135 | 0.23 | 2.30E-122 |
| NECAB1 | 5.41E-157 | -0.5473652 | 0.405 | 0.513 | 2.07E-152 |
| TAFA2 | 2.37E-38 | -0.5540964 | 0.088 | 0.128 | 9.06E-34 |
| CIRBP | 8.50E-169 | -0.5580129 | 0.324 | 0.444 | 3.25E-164 |
| GPM6A | 6.33E-35 | -0.5613014 | 0.21 | 0.255 | 2.42E-30 |
| CDH11 | 7.78E-155 | -0.5631377 | 0.165 | 0.275 | 2.97E-150 |
| MYT1L | 9.59E-22 | -0.5679604 | 0.075 | 0.102 | 3.66E-17 |
| DPH6 | 4.58E-151 | -0.5690983 | 0.155 | 0.261 | 1.75E-146 |
| AC007364.1 | 3.39E-170 | -0.5727533 | 0.166 | 0.284 | 1.29E-165 |
| MT-CO1 | 7.96E-110 | -0.5728936 | 0.622 | 0.671 | 3.04E-105 |
| DGKB | 6.57E-56 | -0.5731083 | 0.07 | 0.117 | 2.51E-51 |
| ACTB | 2.45E-33 | -0.5785616 | 0.44 | 0.479 | 9.36E-29 |
| CNTN4 | 2.59E-50 | -0.5814353 | 0.067 | 0.11 | 9.90E-46 |
| DCLK1 | 2.68E-63 | -0.5870059 | 0.066 | 0.115 | 1.02E-58 |
| CDH18 | 2.72E-55 | -0.5929743 | 0.07 | 0.116 | 1.04E-50 |
| CADPS | 3.63E-64 | -0.5938843 | 0.074 | 0.126 | 1.39E-59 |
| GNAS | 1.13E-54 | -0.5942725 | 0.226 | 0.288 | 4.32E-50 |
| ASIC2 | 5.14E-56 | -0.5973707 | 0.06 | 0.105 | 1.96E-51 |
| RFX3-AS1 | 2.15E-199 | -0.5977756 | 0.192 | 0.322 | 8.23E-195 |
| PTPRN2 | 8.34E-50 | -0.599028 | 0.07 | 0.114 | 3.19E-45 |

| | | | | | |
|---|---|---|---|---|---|
| DTNA | 1.32E-113 | -0.6008395 | 0.193 | 0.287 | 5.06E-109 |
| KHDRBS2 | 7.26E-39 | -0.6024536 | 0.125 | 0.169 | 2.78E-34 |
| SLC4A10 | 7.31E-76 | -0.6059046 | 0.057 | 0.109 | 2.80E-71 |
| GRIA3 | 2.31E-108 | -0.6072746 | 0.121 | 0.202 | 8.82E-104 |
| PTPRG | 6.20E-70 | -0.6078151 | 0.082 | 0.138 | 2.37E-65 |
| MOBP | 2.89E-187 | -0.612646 | 0.729 | 0.726 | 1.10E-182 |
| IQCJ-SCHIP1 | 2.68E-138 | -0.6183107 | 0.381 | 0.49 | 1.02E-133 |
| LRRTM4 | 6.44E-43 | -0.618401 | 0.296 | 0.355 | 2.46E-38 |
| UNC79 | 2.64E-153 | -0.6243658 | 0.211 | 0.323 | 1.01E-148 |
| GRIK2 | 1.77E-86 | -0.625824 | 0.138 | 0.214 | 6.77E-82 |
| MEF2C | 1.94E-55 | -0.6283698 | 0.087 | 0.137 | 7.42E-51 |
| OXR1 | 9.54E-52 | -0.6320736 | 0.124 | 0.177 | 3.65E-47 |
| CHN1 | 2.49E-63 | -0.6326911 | 0.118 | 0.176 | 9.51E-59 |
| NRCAM | 1.41E-79 | -0.6335955 | 0.081 | 0.141 | 5.37E-75 |
| FGF12 | 1.71E-56 | -0.6387039 | 0.121 | 0.176 | 6.52E-52 |
| NAV3 | 1.97E-73 | -0.638744 | 0.309 | 0.388 | 7.53E-69 |
| LINGO2 | 1.03E-66 | -0.6476981 | 0.063 | 0.113 | 3.94E-62 |
| LDB3 | 6.32E-235 | -0.6506265 | 0.303 | 0.446 | 2.42E-230 |
| GRM5 | 4.62E-63 | -0.655857 | 0.084 | 0.137 | 1.76E-58 |
| ADGRL2 | 2.54E-58 | -0.6577853 | 0.059 | 0.104 | 9.71E-54 |
| CACNA2D3 | 5.82E-67 | -0.6622607 | 0.071 | 0.123 | 2.23E-62 |
| PRKG1 | 7.36E-44 | -0.6639289 | 0.069 | 0.108 | 2.81E-39 |
| CARNS1 | 9.23E-235 | -0.6674672 | 0.377 | 0.504 | 3.53E-230 |
| SLC1A2 | 1.50E-146 | -0.6679506 | 0.052 | 0.127 | 5.74E-142 |
| GLDN | 3.21E-271 | -0.6713998 | 0.44 | 0.572 | 1.23E-266 |
| RPL13 | 3.58E-13 | -0.6715938 | 0.306 | 0.264 | 1.37E-08 |
| CLVS1 | 6.29E-223 | -0.674614 | 0.062 | 0.166 | 2.40E-218 |
| SLC8A1 | 5.16E-64 | -0.6803955 | 0.085 | 0.138 | 1.97E-59 |
| TENM2 | 5.24E-41 | -0.6855769 | 0.129 | 0.175 | 2.00E-36 |
| PWRN1 | 3.07E-149 | -0.6942755 | 0.174 | 0.284 | 1.17E-144 |
| UNC5D | 4.15E-57 | -0.696029 | 0.059 | 0.103 | 1.59E-52 |
| PPFIA2 | 3.19E-261 | -0.6970972 | 0.485 | 0.608 | 1.22E-256 |
| PCDH7 | 1.11E-65 | -0.7048998 | 0.108 | 0.167 | 4.26E-61 |
| GREB1L | 4.04E-265 | -0.7073685 | 0.225 | 0.383 | 1.54E-260 |
| NELL2 | 1.13E-72 | -0.7088537 | 0.053 | 0.103 | 4.31E-68 |
| HS6ST3 | 7.95E-80 | -0.7089573 | 0.065 | 0.121 | 3.04E-75 |
| MEG3 | 1.11E-41 | -0.7112159 | 0.241 | 0.291 | 4.23E-37 |
| AC090164.5 | 0 | -0.728216 | 0.001 | 0.102 | 0 |
| RIMS2 | 2.76E-66 | -0.7290738 | 0.114 | 0.172 | 1.05E-61 |
| NRXN3 | 0 | -0.7317619 | 0.796 | 0.85 | 0 |
| SVEP1 | 1.32E-147 | -0.73448 | 0.195 | 0.3 | 5.03E-143 |
| KALRN | 3.04E-101 | -0.7484511 | 0.152 | 0.233 | 1.16E-96 |
| FRMPD4 | 1.94E-70 | -0.7516087 | 0.075 | 0.129 | 7.43E-66 |
| ATRNL1 | 4.51E-91 | -0.7532616 | 0.118 | 0.191 | 1.72E-86 |
| SNAP25 | 5.47E-95 | -0.7541451 | 0.112 | 0.184 | 2.09E-90 |

| | | | | | |
|---|---|---|---|---|---|
| KCNQ5 | 4.35E-79 | -0.7593498 | 0.103 | 0.167 | 1.66E-74 |
| LMCD1-AS1 | 0 | -0.7650258 | 0.374 | 0.556 | 0 |
| DLGAP2 | 1.01E-68 | -0.7855617 | 0.082 | 0.138 | 3.86E-64 |
| FGF14 | 1.65E-90 | -0.7925671 | 0.156 | 0.233 | 6.29E-86 |
| NRG1 | 3.27E-81 | -0.7935932 | 0.059 | 0.114 | 1.25E-76 |
| RPLP0 | 3.45E-13 | -0.7961564 | 0.1 | 0.121 | 1.32E-08 |
| SPARCL1 | 1.95E-114 | -0.7979742 | 0.09 | 0.168 | 7.47E-110 |
| PLCB1 | 1.86E-74 | -0.8043587 | 0.136 | 0.205 | 7.09E-70 |
| RPL10A | 2.87E-11 | -0.8107966 | 0.086 | 0.104 | 1.10E-06 |
| RPL35A | 5.13E-10 | -0.8115055 | 0.113 | 0.13 | 1.96E-05 |
| RPL9 | 1.34E-12 | -0.818641 | 0.137 | 0.158 | 5.11E-08 |
| NAALADL2 | 0 | -0.8200254 | 0.342 | 0.544 | 0 |
| CCSER1 | 9.38E-121 | -0.8205446 | 0.09 | 0.17 | 3.58E-116 |
| CELF2 | 2.06E-89 | -0.8223086 | 0.139 | 0.216 | 7.88E-85 |
| DLGAP1 | 8.72E-103 | -0.8232015 | 0.175 | 0.26 | 3.33E-98 |
| DPP10 | 3.29E-45 | -0.8243297 | 0.257 | 0.316 | 1.26E-40 |
| RGS7 | 1.31E-150 | -0.8292434 | 0.113 | 0.211 | 5.00E-146 |
| ACTG1 | 1.26E-89 | -0.829838 | 0.177 | 0.258 | 4.81E-85 |
| FAM155A | 1.71E-120 | -0.8640195 | 0.168 | 0.261 | 6.55E-116 |
| KCNMA1 | 1.19E-69 | -0.8667839 | 0.108 | 0.169 | 4.57E-65 |
| PDE4D | 2.10E-194 | -0.8693276 | 0.362 | 0.489 | 8.04E-190 |
| AGBL4 | 7.12E-92 | -0.8760497 | 0.081 | 0.146 | 2.72E-87 |
| RALYL | 1.15E-89 | -0.8770136 | 0.124 | 0.196 | 4.41E-85 |
| RPL37 | 5.66E-14 | -0.87716 | 0.156 | 0.179 | 2.16E-09 |
| PPP2R2B | 0 | -0.906214 | 0.903 | 0.875 | 0 |
| RPS15A | 1.30E-13 | -0.9073332 | 0.105 | 0.126 | 4.96E-09 |
| STXBP5L | 6.10E-114 | -0.9366159 | 0.086 | 0.161 | 2.33E-109 |
| CSMD1 | 9.83E-124 | -0.9795685 | 0.336 | 0.432 | 3.76E-119 |
| NRXN1 | 2.36E-164 | -0.9864117 | 0.25 | 0.366 | 9.02E-160 |
| NRG3 | 3.09E-134 | -1.0060606 | 0.221 | 0.323 | 1.18E-129 |
| RPL15 | 9.18E-14 | -1.0150548 | 0.16 | 0.184 | 3.51E-09 |
| RPL3 | 1.08E-08 | -1.0187099 | 0.161 | 0.177 | 0.00041136 |
| ROBO2 | 1.10E-152 | -1.0220486 | 0.148 | 0.252 | 4.20E-148 |
| OPCML | 8.60E-86 | -1.1094306 | 0.21 | 0.288 | 3.29E-81 |
| PDE1A | 0 | -1.1286462 | 0.407 | 0.606 | 0 |
| RBFOX1 | 9.85E-201 | -1.148152 | 0.323 | 0.453 | 3.76E-196 |
| SYT1 | 8.29E-178 | -1.2052924 | 0.231 | 0.35 | 3.17E-173 |
| RPS12 | 5.58E-29 | -1.219283 | 0.126 | 0.162 | 2.13E-24 |
| RPS18 | 6.09E-16 | -1.2229936 | 0.143 | 0.168 | 2.33E-11 |
| KCNIP4 | 1.67E-201 | -1.2428105 | 0.427 | 0.561 | 6.37E-197 |
| RPS4X | 2.54E-16 | -1.5753697 | 0.094 | 0.116 | 9.72E-12 |

**Oligodendrocyte Precursor DEGs**

| | p_val | avg_log2FC | pct.1 | pct.2 | p_val_adj |
|---|---|---|---|---|---|
| PLCG2 | 9.37E-173 | 1.33225102 | 0.815 | 0.559 | 3.58E-168 |
| MTRNR2L12 | 3.42E-137 | 1.17653339 | 0.735 | 0.538 | 1.31E-132 |
| MT-ND3 | 1.70E-149 | 1.10854252 | 0.821 | 0.664 | 6.51E-145 |
| HSP90AA1 | 1.40E-73 | 0.9061168 | 0.629 | 0.477 | 5.34E-69 |
| MT-ATP6 | 5.96E-89 | 0.87389292 | 0.82 | 0.688 | 2.28E-84 |
| MT-CYB | 1.66E-89 | 0.85624198 | 0.712 | 0.536 | 6.36E-85 |
| AC093865.1 | 1.01E-119 | 0.80390564 | 0.536 | 0.289 | 3.86E-115 |
| EGFR | 5.77E-116 | 0.7787067 | 0.722 | 0.536 | 2.20E-111 |
| MT-CO2 | 5.09E-85 | 0.77312856 | 0.845 | 0.733 | 1.95E-80 |
| MT-ND4 | 4.30E-62 | 0.77136971 | 0.791 | 0.669 | 1.64E-57 |
| FKBP5 | 7.88E-98 | 0.70613458 | 0.695 | 0.511 | 3.01E-93 |
| MT-ND1 | 2.56E-52 | 0.70504248 | 0.544 | 0.391 | 9.77E-48 |
| MT-CO3 | 3.32E-41 | 0.6612224 | 0.727 | 0.627 | 1.27E-36 |
| HSPH1 | 1.53E-27 | 0.63828082 | 0.268 | 0.168 | 5.86E-23 |
| TAFA1 | 5.54E-77 | 0.63387064 | 0.723 | 0.559 | 2.12E-72 |
| MT-ND2 | 7.47E-46 | 0.63162934 | 0.563 | 0.418 | 2.85E-41 |
| HSPA1A | 1.72E-28 | 0.62302655 | 0.226 | 0.13 | 6.56E-24 |
| ZBTB16 | 6.83E-94 | 0.61849755 | 0.763 | 0.618 | 2.61E-89 |
| PDZD2 | 2.41E-140 | 0.59955311 | 0.958 | 0.947 | 9.21E-136 |
| SEM1 | 9.20E-70 | 0.59354889 | 0.527 | 0.35 | 3.52E-65 |
| HIF3A | 1.71E-102 | 0.59303119 | 0.825 | 0.726 | 6.54E-98 |
| KCNMB2 | 2.67E-133 | 0.58850745 | 0.947 | 0.844 | 1.02E-128 |
| PLD5 | 1.38E-88 | 0.58185261 | 0.758 | 0.634 | 5.28E-84 |
| TLE1 | 9.39E-84 | 0.57963652 | 0.417 | 0.215 | 3.59E-79 |
| AL627171.2 | 1.18E-45 | 0.56320312 | 0.249 | 0.125 | 4.50E-41 |
| EEF2K | 1.26E-75 | 0.55853573 | 0.633 | 0.456 | 4.82E-71 |
| HP1BP3 | 8.76E-85 | 0.55578875 | 0.705 | 0.566 | 3.35E-80 |
| KIAA1217 | 6.48E-74 | 0.55280787 | 0.839 | 0.771 | 2.48E-69 |
| DPH6-DT | 4.68E-58 | 0.5429954 | 0.468 | 0.297 | 1.79E-53 |
| MT3 | 5.79E-30 | 0.54155383 | 0.626 | 0.543 | 2.22E-25 |
| SORBS1 | 5.46E-27 | 0.52720624 | 0.784 | 0.711 | 2.09E-22 |
| EEF1A1 | 1.16E-12 | 0.52508942 | 0.452 | 0.388 | 4.42E-08 |
| MGLL | 5.77E-69 | 0.52279401 | 0.63 | 0.463 | 2.20E-64 |
| VCAN | 8.43E-137 | 0.51251288 | 0.973 | 0.97 | 3.22E-132 |
| DOCK5 | 3.85E-46 | 0.4993779 | 0.72 | 0.637 | 1.47E-41 |
| APOLD1 | 8.88E-32 | 0.48569245 | 0.144 | 0.062 | 3.40E-27 |
| DNAJB1 | 1.54E-16 | 0.48383472 | 0.163 | 0.099 | 5.89E-12 |
| FBXO31 | 2.72E-54 | 0.48304488 | 0.359 | 0.204 | 1.04E-49 |
| NEAT1 | 2.69E-19 | 0.46744047 | 0.39 | 0.299 | 1.03E-14 |
| MT-ND4L | 2.09E-24 | 0.46664736 | 0.394 | 0.297 | 7.99E-20 |
| KIF19 | 8.31E-59 | 0.45528101 | 0.313 | 0.157 | 3.18E-54 |
| DGKG | 2.90E-127 | 0.45523772 | 0.947 | 0.93 | 1.11E-122 |
| FAM13C | 6.92E-36 | 0.45201068 | 0.509 | 0.389 | 2.65E-31 |

| | | | | | |
|---|---|---|---|---|---|
| PARD3 | 1.83E-76 | 0.44998746 | 0.904 | 0.88 | 7.01E-72 |
| AC119674.1 | 5.38E-38 | 0.44948268 | 0.301 | 0.177 | 2.06E-33 |
| ITGA8 | 2.14E-48 | 0.4476982 | 0.708 | 0.593 | 8.18E-44 |
| MT2A | 1.13E-19 | 0.44704844 | 0.368 | 0.279 | 4.33E-15 |
| LIFR | 9.38E-50 | 0.44172538 | 0.73 | 0.647 | 3.59E-45 |
| HIP1 | 1.06E-51 | 0.44159068 | 0.859 | 0.837 | 4.06E-47 |
| HDAC4 | 1.72E-53 | 0.44064505 | 0.757 | 0.659 | 6.57E-49 |
| DISP1 | 7.19E-47 | 0.43369665 | 0.557 | 0.406 | 2.75E-42 |
| AC005906.2 | 2.17E-51 | 0.43180056 | 0.418 | 0.259 | 8.31E-47 |
| SLAIN1 | 2.02E-49 | 0.43136423 | 0.799 | 0.744 | 7.72E-45 |
| HSPB1 | 4.58E-38 | 0.42139963 | 0.171 | 0.074 | 1.75E-33 |
| GFAP | 1.41E-11 | 0.41595762 | 0.18 | 0.126 | 5.41E-07 |
| SGO1-AS1 | 1.61E-34 | 0.41488228 | 0.225 | 0.117 | 6.16E-30 |
| BMERB1 | 4.50E-38 | 0.41336417 | 0.704 | 0.584 | 1.72E-33 |
| CCDC144A | 3.20E-50 | 0.40605179 | 0.452 | 0.291 | 1.22E-45 |
| HMGB1 | 1.48E-36 | 0.40406494 | 0.709 | 0.635 | 5.67E-32 |
| SLC5A3 | 2.77E-55 | 0.40080333 | 0.341 | 0.185 | 1.06E-50 |
| LINC01117 | 1.13E-35 | 0.40014132 | 0.304 | 0.18 | 4.32E-31 |
| PANTR1 | 1.76E-45 | 0.39927028 | 0.633 | 0.484 | 6.73E-41 |
| KCNQ3 | 1.19E-24 | 0.39632607 | 0.634 | 0.548 | 4.55E-20 |
| ZMAT3 | 4.80E-40 | 0.39426208 | 0.632 | 0.523 | 1.83E-35 |
| SAMHD1 | 3.13E-37 | 0.39285972 | 0.469 | 0.331 | 1.20E-32 |
| SGK1 | 1.26E-51 | 0.39018395 | 0.861 | 0.844 | 4.81E-47 |
| FLRT2 | 1.28E-31 | 0.3872237 | 0.744 | 0.683 | 4.88E-27 |
| CHST3 | 5.02E-30 | 0.38549496 | 0.364 | 0.246 | 1.92E-25 |
| ZFPM2 | 3.77E-50 | 0.37900498 | 0.918 | 0.928 | 1.44E-45 |
| SPP1 | 5.64E-22 | 0.37741899 | 0.115 | 0.054 | 2.15E-17 |
| CNTN3 | 4.03E-36 | 0.37646008 | 0.741 | 0.681 | 1.54E-31 |
| AJAP1 | 3.04E-40 | 0.37532094 | 0.345 | 0.206 | 1.16E-35 |
| CDH20 | 2.41E-42 | 0.37304231 | 0.893 | 0.905 | 9.20E-38 |
| B2M | 3.23E-23 | 0.36978375 | 0.485 | 0.39 | 1.24E-18 |
| AC114971.1 | 8.20E-25 | 0.36620664 | 0.375 | 0.266 | 3.13E-20 |
| RXRA | 1.20E-30 | 0.36544789 | 0.493 | 0.377 | 4.60E-26 |
| KANSL1L | 1.58E-32 | 0.36311154 | 0.662 | 0.579 | 6.03E-28 |
| CHL1-AS2 | 2.45E-38 | 0.3622246 | 0.271 | 0.149 | 9.38E-34 |
| MT-ND5 | 3.98E-18 | 0.36202954 | 0.383 | 0.292 | 1.52E-13 |
| MT-ATP8 | 3.48E-15 | 0.3604457 | 0.383 | 0.3 | 1.33E-10 |
| RAMP1 | 3.64E-33 | 0.35698737 | 0.606 | 0.5 | 1.39E-28 |
| BCR | 3.78E-25 | 0.35386803 | 0.522 | 0.415 | 1.44E-20 |
| AL110292.1 | 2.51E-34 | 0.35157377 | 0.415 | 0.283 | 9.60E-30 |
| HSPA8 | 4.18E-27 | 0.35004155 | 0.312 | 0.205 | 1.60E-22 |
| LINC00486 | 3.55E-09 | 0.34730385 | 0.566 | 0.506 | 0.00013555 |
| MKLN1 | 2.20E-56 | 0.34585822 | 0.888 | 0.886 | 8.40E-52 |
| ZHX3 | 6.41E-32 | 0.33966505 | 0.677 | 0.595 | 2.45E-27 |
| GAB1 | 4.07E-35 | 0.33799644 | 0.777 | 0.728 | 1.55E-30 |

| | | | | | |
|---|---|---|---|---|---|
| PTPRJ | 5.70E-29 | 0.33739434 | 0.852 | 0.803 | 2.18E-24 |
| HSP90AB1 | 2.89E-13 | 0.33692118 | 0.463 | 0.39 | 1.11E-08 |
| PPM1B | 8.11E-33 | 0.33652758 | 0.682 | 0.616 | 3.10E-28 |
| METTL7A | 2.60E-30 | 0.33582172 | 0.417 | 0.298 | 9.94E-26 |
| SPECC1 | 3.54E-26 | 0.33539042 | 0.481 | 0.371 | 1.35E-21 |
| TAFA5 | 4.18E-38 | 0.33285179 | 0.586 | 0.446 | 1.60E-33 |
| LINC00632 | 6.78E-24 | 0.33283424 | 0.632 | 0.547 | 2.59E-19 |
| RPL28 | 3.31E-12 | 0.32799223 | 0.298 | 0.234 | 1.27E-07 |
| MAPT-AS1 | 3.39E-29 | 0.3272136 | 0.212 | 0.115 | 1.29E-24 |
| LINC01546 | 1.69E-65 | 0.32656049 | 0.188 | 0.058 | 6.45E-61 |
| CDH19 | 1.00E-27 | 0.32468666 | 0.2 | 0.107 | 3.84E-23 |
| PDE8B | 4.40E-43 | 0.32422904 | 0.837 | 0.716 | 1.68E-38 |
| MRPS6 | 9.74E-36 | 0.32144622 | 0.343 | 0.216 | 3.72E-31 |
| ARL6IP1 | 1.34E-07 | 0.31980927 | 0.307 | 0.252 | 0.00510963 |
| CRISPLD2 | 1.16E-26 | 0.31641492 | 0.537 | 0.418 | 4.44E-22 |
| SATB1-AS1 | 3.62E-23 | 0.31632042 | 0.51 | 0.407 | 1.38E-18 |
| GSG1L | 1.42E-25 | 0.31447403 | 0.687 | 0.602 | 5.43E-21 |
| CEBPD | 2.38E-26 | 0.31374492 | 0.223 | 0.132 | 9.11E-22 |
| INHBA | 1.01E-41 | 0.31046907 | 0.175 | 0.072 | 3.87E-37 |
| DST | 1.30E-55 | 0.30571463 | 0.949 | 0.946 | 4.97E-51 |
| HS3ST1 | 8.16E-40 | 0.30544382 | 0.245 | 0.125 | 3.12E-35 |
| ZDHHC14 | 1.64E-39 | 0.30524519 | 0.906 | 0.884 | 6.28E-35 |
| RFTN2 | 7.19E-32 | 0.30463699 | 0.69 | 0.588 | 2.75E-27 |
| UBB | 7.58E-24 | 0.30306607 | 0.296 | 0.197 | 2.90E-19 |
| UTP23 | 3.56E-24 | 0.30187732 | 0.287 | 0.191 | 1.36E-19 |
| PRKX | 3.05E-32 | 0.30005312 | 0.273 | 0.161 | 1.16E-27 |
| MEIKIN | 6.14E-27 | 0.29966187 | 0.215 | 0.122 | 2.35E-22 |
| CALN1 | 2.38E-20 | 0.29806707 | 0.366 | 0.269 | 9.09E-16 |
| GRIA4 | 1.32E-29 | 0.29782639 | 0.87 | 0.865 | 5.06E-25 |
| RAD18 | 4.63E-23 | 0.29688434 | 0.382 | 0.277 | 1.77E-18 |
| KANSL1 | 2.35E-30 | 0.29581345 | 0.789 | 0.734 | 8.99E-26 |
| APOD | 5.22E-19 | 0.29182414 | 0.739 | 0.694 | 1.99E-14 |
| NOVA1 | 6.44E-44 | 0.28754814 | 0.941 | 0.958 | 2.46E-39 |
| GPNMB | 2.01E-12 | 0.28590738 | 0.568 | 0.501 | 7.69E-08 |
| MAPT | 1.04E-23 | 0.28483617 | 0.744 | 0.693 | 3.98E-19 |
| KLHL4 | 7.00E-20 | 0.28406043 | 0.3 | 0.211 | 2.68E-15 |
| VAMP2 | 1.52E-25 | 0.28362093 | 0.48 | 0.367 | 5.80E-21 |
| ZHX2 | 6.48E-25 | 0.28355403 | 0.835 | 0.792 | 2.48E-20 |
| CST3 | 4.49E-08 | 0.28236489 | 0.62 | 0.592 | 0.00171569 |
| CD82 | 1.35E-18 | 0.28217262 | 0.332 | 0.24 | 5.16E-14 |
| SLC38A2 | 1.16E-19 | 0.27977371 | 0.371 | 0.279 | 4.43E-15 |
| MBNL1 | 4.98E-20 | 0.27874811 | 0.732 | 0.683 | 1.90E-15 |
| CHORDC1 | 5.49E-19 | 0.27864631 | 0.285 | 0.2 | 2.10E-14 |
| PYGL | 7.46E-24 | 0.27861941 | 0.384 | 0.273 | 2.85E-19 |
| GPM6B | 9.84E-28 | 0.276811 | 0.86 | 0.852 | 3.76E-23 |

| | | | | | |
|---|---|---|---|---|---|
| UBA6-AS1 | 3.68E-16 | 0.2751365 | 0.577 | 0.5 | 1.41E-11 |
| PEX5L | 1.09E-14 | 0.27374089 | 0.588 | 0.512 | 4.17E-10 |
| FOXO1 | 7.99E-17 | 0.2735931 | 0.465 | 0.375 | 3.05E-12 |
| CALM2 | 3.55E-15 | 0.27353118 | 0.547 | 0.465 | 1.36E-10 |
| KLF15 | 4.37E-30 | 0.27281228 | 0.294 | 0.182 | 1.67E-25 |
| ETV1 | 2.43E-13 | 0.27226415 | 0.608 | 0.561 | 9.29E-09 |
| PAPOLA | 6.91E-16 | 0.26801979 | 0.544 | 0.479 | 2.64E-11 |
| FXR1 | 1.73E-18 | 0.26765252 | 0.526 | 0.44 | 6.59E-14 |
| PTGES3 | 4.95E-17 | 0.26756653 | 0.343 | 0.255 | 1.89E-12 |
| IKZF2 | 2.05E-20 | 0.26521545 | 0.595 | 0.51 | 7.84E-16 |
| CXXC4 | 3.11E-25 | 0.26519429 | 0.368 | 0.255 | 1.19E-20 |
| SRSF6 | 3.54E-21 | 0.26462937 | 0.47 | 0.37 | 1.35E-16 |
| ZCCHC14 | 5.80E-11 | 0.26375133 | 0.428 | 0.364 | 2.22E-06 |
| MAP1LC3B | 2.18E-26 | 0.26276603 | 0.232 | 0.138 | 8.33E-22 |
| ZMIZ1 | 7.93E-12 | 0.26181529 | 0.653 | 0.595 | 3.03E-07 |
| C16orf95 | 6.39E-29 | 0.26152342 | 0.155 | 0.073 | 2.44E-24 |
| SUCLG2-AS1 | 1.42E-13 | 0.25961597 | 0.393 | 0.315 | 5.44E-09 |
| P2RX7 | 2.15E-17 | 0.25864922 | 0.629 | 0.554 | 8.24E-13 |
| AC093523.1 | 7.55E-16 | 0.2585803 | 0.259 | 0.183 | 2.89E-11 |
| AC007106.2 | 9.86E-22 | 0.25835053 | 0.284 | 0.189 | 3.77E-17 |
| ALK | 5.67E-16 | 0.25778431 | 0.767 | 0.731 | 2.17E-11 |
| HIST1H2AC | 5.30E-40 | 0.25763225 | 0.162 | 0.064 | 2.03E-35 |
| NFASC | 3.91E-09 | 0.25611052 | 0.659 | 0.61 | 0.00014936 |
| ANKRD55 | 2.41E-28 | 0.25546945 | 0.228 | 0.129 | 9.22E-24 |
| ARHGAP18 | 6.08E-28 | 0.25348099 | 0.136 | 0.061 | 2.32E-23 |
| ASCL1 | 9.04E-14 | 0.25184 | 0.378 | 0.3 | 3.46E-09 |
| CHST9 | 5.15E-24 | 0.2517516 | 0.61 | 0.49 | 1.97E-19 |
| FMO5 | 2.34E-26 | 0.25113058 | 0.146 | 0.069 | 8.94E-22 |
| B3GAT1 | 1.30E-15 | 0.25078245 | 0.474 | 0.394 | 4.96E-11 |
| DDHD1 | 2.22E-27 | -0.2500168 | 0.401 | 0.507 | 8.47E-23 |
| APOE | 1.54E-33 | -0.2504597 | 0.097 | 0.198 | 5.89E-29 |
| CPNE8 | 1.32E-23 | -0.2512487 | 0.125 | 0.211 | 5.05E-19 |
| CACNB2 | 3.84E-15 | -0.2515052 | 0.342 | 0.426 | 1.47E-10 |
| DLG1 | 1.61E-18 | -0.2523341 | 0.728 | 0.786 | 6.17E-14 |
| ATP1B2 | 1.27E-21 | -0.252825 | 0.442 | 0.533 | 4.84E-17 |
| PCLO | 1.13E-19 | -0.253992 | 0.241 | 0.338 | 4.33E-15 |
| SPRED1 | 1.30E-14 | -0.2540791 | 0.718 | 0.765 | 4.98E-10 |
| LINC00937 | 1.02E-11 | -0.2545135 | 0.33 | 0.387 | 3.90E-07 |
| AC092691.1 | 5.56E-25 | -0.2575127 | 0.923 | 0.944 | 2.12E-20 |
| AL133346.1 | 7.98E-18 | -0.2589638 | 0.402 | 0.494 | 3.05E-13 |
| MTMR1 | 1.42E-21 | -0.2598635 | 0.344 | 0.431 | 5.42E-17 |
| SEMA6D | 4.19E-10 | -0.2606224 | 0.243 | 0.297 | 1.60E-05 |
| GALR1 | 1.42E-16 | -0.2611518 | 0.382 | 0.456 | 5.44E-12 |
| PDE10A | 2.31E-14 | -0.2637313 | 0.248 | 0.319 | 8.83E-10 |
| CENPP | 2.07E-19 | -0.2638134 | 0.179 | 0.265 | 7.90E-15 |

| | | | | | |
|---|---|---|---|---|---|
| ENOSF1 | 1.43E-20 | -0.2641201 | 0.237 | 0.324 | 5.47E-16 |
| GALNT10 | 1.03E-21 | -0.2646015 | 0.505 | 0.587 | 3.95E-17 |
| AC098829.1 | 4.12E-11 | -0.2656251 | 0.151 | 0.206 | 1.58E-06 |
| CACNA1E | 1.48E-19 | -0.2657847 | 0.284 | 0.368 | 5.67E-15 |
| CMSS1 | 8.66E-26 | -0.2664152 | 0.398 | 0.508 | 3.31E-21 |
| PDE8A | 1.26E-15 | -0.2667955 | 0.476 | 0.544 | 4.82E-11 |
| AL359232.1 | 1.56E-19 | -0.2672585 | 0.203 | 0.294 | 5.97E-15 |
| MIAT | 8.79E-25 | -0.2679082 | 0.454 | 0.548 | 3.36E-20 |
| STK33 | 6.92E-17 | -0.2679482 | 0.28 | 0.358 | 2.65E-12 |
| SLC4A10 | 3.32E-16 | -0.2688946 | 0.439 | 0.514 | 1.27E-11 |
| PROM1 | 2.37E-31 | -0.2694938 | 0.192 | 0.307 | 9.08E-27 |
| KLRC2 | 3.29E-18 | -0.2719761 | 0.232 | 0.308 | 1.26E-13 |
| PAM | 6.90E-25 | -0.2722836 | 0.505 | 0.599 | 2.64E-20 |
| PDE3A | 7.14E-31 | -0.272828 | 0.085 | 0.176 | 2.73E-26 |
| ABCA5 | 2.17E-28 | -0.2766285 | 0.32 | 0.432 | 8.28E-24 |
| CNTN4 | 5.43E-20 | -0.2782322 | 0.469 | 0.572 | 2.08E-15 |
| PLPPR1 | 4.57E-30 | -0.2786612 | 0.79 | 0.864 | 1.75E-25 |
| TNK2 | 4.82E-30 | -0.2827962 | 0.744 | 0.831 | 1.84E-25 |
| PTK2 | 8.80E-41 | -0.2846628 | 0.833 | 0.88 | 3.36E-36 |
| GRID1 | 7.92E-42 | -0.2867601 | 0.835 | 0.898 | 3.03E-37 |
| SERINC5 | 3.51E-25 | -0.2868144 | 0.662 | 0.746 | 1.34E-20 |
| PPFIA2 | 6.33E-34 | -0.2893395 | 0.812 | 0.894 | 2.42E-29 |
| CSPG5 | 3.42E-26 | -0.2909079 | 0.569 | 0.664 | 1.31E-21 |
| PPP2R2B | 3.57E-31 | -0.2944836 | 0.907 | 0.947 | 1.37E-26 |
| XYLT1 | 7.35E-43 | -0.2956863 | 0.939 | 0.965 | 2.81E-38 |
| NCALD | 1.22E-21 | -0.2967591 | 0.684 | 0.765 | 4.68E-17 |
| BRINP1 | 1.45E-22 | -0.2970293 | 0.59 | 0.674 | 5.54E-18 |
| SHC3 | 1.88E-39 | -0.2976031 | 0.697 | 0.79 | 7.19E-35 |
| ERC2 | 5.20E-30 | -0.2976876 | 0.713 | 0.808 | 1.99E-25 |
| CABLES1 | 4.81E-16 | -0.2980158 | 0.496 | 0.576 | 1.84E-11 |
| AC023480.1 | 9.42E-17 | -0.2981786 | 0.351 | 0.425 | 3.60E-12 |
| PCDH15 | 6.98E-54 | -0.2989128 | 0.961 | 0.981 | 2.67E-49 |
| GMDS-DT | 5.29E-20 | -0.2993177 | 0.484 | 0.561 | 2.02E-15 |
| GOLGA8A | 5.03E-37 | -0.3001061 | 0.486 | 0.622 | 1.92E-32 |
| PTPRO | 1.07E-18 | -0.3016758 | 0.359 | 0.444 | 4.09E-14 |
| SLC4A4 | 4.30E-24 | -0.3020073 | 0.196 | 0.291 | 1.64E-19 |
| RANBP17 | 7.53E-24 | -0.3033379 | 0.226 | 0.327 | 2.88E-19 |
| XKR4 | 5.66E-39 | -0.3037655 | 0.814 | 0.893 | 2.17E-34 |
| GPR17 | 2.84E-11 | -0.3056241 | 0.36 | 0.417 | 1.09E-06 |
| GALNTL6 | 8.85E-14 | -0.3113876 | 0.145 | 0.208 | 3.38E-09 |
| ADAMTS17 | 4.22E-35 | -0.320719 | 0.727 | 0.814 | 1.61E-30 |
| WSB1 | 9.85E-53 | -0.3220145 | 0.688 | 0.817 | 3.76E-48 |
| SPG7 | 2.76E-43 | -0.3226634 | 0.433 | 0.569 | 1.05E-38 |
| GABRB2 | 4.21E-28 | -0.3278559 | 0.129 | 0.226 | 1.61E-23 |
| SOX6 | 3.88E-57 | -0.327971 | 0.926 | 0.963 | 1.48E-52 |

| | | | | | |
|---|---|---|---|---|---|
| SUMF1 | 4.54E-25 | -0.3282765 | 0.491 | 0.584 | 1.73E-20 |
| SLC16A10 | 1.26E-18 | -0.3299556 | 0.225 | 0.308 | 4.83E-14 |
| LINC02649 | 1.45E-37 | -0.3341872 | 0.078 | 0.176 | 5.55E-33 |
| GLDC | 5.36E-11 | -0.3361299 | 0.309 | 0.369 | 2.05E-06 |
| LINC01184 | 2.56E-25 | -0.3368854 | 0.407 | 0.502 | 9.80E-21 |
| NR6A1 | 2.44E-25 | -0.336913 | 0.348 | 0.451 | 9.32E-21 |
| CIRBP | 2.62E-47 | -0.3383062 | 0.463 | 0.616 | 1.00E-42 |
| SGCZ | 3.99E-39 | -0.3414089 | 0.741 | 0.85 | 1.53E-34 |
| PCDH11X | 7.96E-22 | -0.3417318 | 0.371 | 0.471 | 3.04E-17 |
| AGAP1 | 1.41E-94 | -0.3424448 | 0.959 | 0.978 | 5.37E-90 |
| CFTR | 2.39E-40 | -0.3425038 | 0.115 | 0.228 | 9.13E-36 |
| RAP1GAP | 2.52E-56 | -0.3433919 | 0.219 | 0.376 | 9.65E-52 |
| PTPRT | 2.54E-41 | -0.3471018 | 0.744 | 0.834 | 9.71E-37 |
| EPN2 | 6.15E-68 | -0.3487348 | 0.925 | 0.964 | 2.35E-63 |
| SGCD | 6.06E-48 | -0.3488404 | 0.841 | 0.913 | 2.32E-43 |
| AHI1 | 1.01E-35 | -0.3511326 | 0.561 | 0.662 | 3.84E-31 |
| ASTN2 | 4.82E-74 | -0.3536155 | 0.9 | 0.94 | 1.84E-69 |
| SCFD2 | 2.07E-39 | -0.3545373 | 0.552 | 0.668 | 7.93E-35 |
| HSD17B12 | 9.06E-29 | -0.3565164 | 0.258 | 0.361 | 3.46E-24 |
| GPC5 | 4.55E-10 | -0.3592795 | 0.341 | 0.408 | 1.74E-05 |
| KLHL1 | 3.56E-27 | -0.3609539 | 0.3 | 0.413 | 1.36E-22 |
| TSHZ2 | 1.75E-31 | -0.361652 | 0.164 | 0.276 | 6.68E-27 |
| DOK6 | 5.23E-33 | -0.362589 | 0.424 | 0.543 | 2.00E-28 |
| SPATA6 | 4.74E-43 | -0.3675175 | 0.727 | 0.814 | 1.81E-38 |
| C1orf61 | 1.09E-52 | -0.3702664 | 0.785 | 0.861 | 4.17E-48 |
| DTNA | 2.90E-39 | -0.3794117 | 0.809 | 0.873 | 1.11E-34 |
| LHFPL3 | 1.23E-101 | -0.3810786 | 0.981 | 0.987 | 4.71E-97 |
| ADGRV1 | 4.05E-12 | -0.3848277 | 0.08 | 0.13 | 1.55E-07 |
| ANKS1B | 1.19E-66 | -0.3942258 | 0.929 | 0.966 | 4.55E-62 |
| U91319.1 | 8.05E-50 | -0.3951804 | 0.374 | 0.54 | 3.08E-45 |
| DPH6 | 1.31E-39 | -0.3957014 | 0.382 | 0.524 | 5.01E-35 |
| NRXN3 | 1.80E-37 | -0.3973674 | 0.596 | 0.739 | 6.89E-33 |
| LDLRAD4 | 1.06E-38 | -0.400009 | 0.622 | 0.707 | 4.05E-34 |
| AL359091.1 | 5.45E-28 | -0.4003582 | 0.234 | 0.339 | 2.08E-23 |
| TARBP1 | 1.71E-55 | -0.4004045 | 0.412 | 0.567 | 6.54E-51 |
| AGBL4 | 3.78E-34 | -0.4024333 | 0.397 | 0.525 | 1.44E-29 |
| ENOX1 | 1.03E-35 | -0.4042345 | 0.467 | 0.585 | 3.95E-31 |
| LINC02588 | 1.05E-13 | -0.4123985 | 0.103 | 0.154 | 4.00E-09 |
| RFX3-AS1 | 8.67E-48 | -0.4168104 | 0.313 | 0.462 | 3.31E-43 |
| SHISA9 | 1.38E-60 | -0.423506 | 0.732 | 0.834 | 5.26E-56 |
| HES5 | 4.31E-76 | -0.4259654 | 0.024 | 0.145 | 1.65E-71 |
| ABHD2 | 6.28E-72 | -0.4294305 | 0.851 | 0.91 | 2.40E-67 |
| ROBO2 | 1.49E-32 | -0.4295223 | 0.186 | 0.306 | 5.68E-28 |
| AC061958.1 | 1.08E-11 | -0.430194 | 0.088 | 0.134 | 4.14E-07 |
| GOLGA8B | 1.48E-71 | -0.4304524 | 0.339 | 0.524 | 5.67E-67 |

| | | | | | |
|---|---|---|---|---|---|
| AL392023.2 | 1.40E-41 | -0.4328873 | 0.616 | 0.717 | 5.34E-37 |
| IL1RAP | 2.50E-24 | -0.4345632 | 0.646 | 0.706 | 9.57E-20 |
| VEGFA | 2.26E-31 | -0.4363919 | 0.168 | 0.275 | 8.66E-27 |
| LMCD1-AS1 | 9.96E-45 | -0.4408806 | 0.651 | 0.751 | 3.81E-40 |
| NAV2 | 6.41E-95 | -0.4579999 | 0.926 | 0.943 | 2.45E-90 |
| LRFN5 | 5.81E-42 | -0.4785244 | 0.449 | 0.595 | 2.22E-37 |
| KCNAB1 | 1.49E-15 | -0.4808062 | 0.16 | 0.218 | 5.70E-11 |
| XIST | 1.17E-44 | -0.5045737 | 0.318 | 0.477 | 4.47E-40 |
| RBFOX1 | 1.14E-84 | -0.5233386 | 0.722 | 0.882 | 4.36E-80 |
| KCNQ1OT1 | 7.41E-72 | -0.5371244 | 0.561 | 0.708 | 2.83E-67 |
| FP236383.1 | 2.90E-87 | -0.5578694 | 0 | 0.108 | 1.11E-82 |
| PREX2 | 1.47E-98 | -0.5597102 | 0.605 | 0.768 | 5.64E-94 |
| RPL13 | 2.53E-82 | -0.5615574 | 0.511 | 0.69 | 9.67E-78 |
| CCSER1 | 2.03E-69 | -0.5820749 | 0.54 | 0.707 | 7.75E-65 |
| MTSS1L | 2.53E-90 | -0.584666 | 0 | 0.111 | 9.67E-86 |
| FP671120.1 | 7.40E-92 | -0.6140591 | 0 | 0.113 | 2.83E-87 |
| GPR158 | 2.08E-93 | -0.6588082 | 0.327 | 0.533 | 7.94E-89 |
| AC079352.1 | 3.00E-59 | -0.6614595 | 0.195 | 0.358 | 1.15E-54 |
| AC124254.1 | 4.79E-82 | -0.6810727 | 0 | 0.101 | 1.83E-77 |
| FAM19A2 | 1.61E-85 | -0.7411982 | 0 | 0.106 | 6.14E-81 |
| AC105402.3 | 1.12E-55 | -0.7749742 | 0.043 | 0.15 | 4.27E-51 |

**Excitatory Neuron DEGs**

| | p_val | avg_log2FC | pct.1 | pct.2 | p_val_adj |
|---|---|---|---|---|---|
| MT-ND3 | 0 | 1.40834375 | 0.917 | 0.844 | 0 |
| MTRNR2L12 | 0 | 1.31960236 | 0.82 | 0.695 | 0 |
| MT-CYB | 0 | 1.09565017 | 0.879 | 0.739 | 0 |
| MT-ATP6 | 0 | 1.08150102 | 0.917 | 0.867 | 0 |
| MT-ND4 | 0 | 0.99852582 | 0.907 | 0.83 | 0 |
| PLCG2 | 9.25E-295 | 0.95324861 | 0.723 | 0.613 | 3.53E-290 |
| HSPH1 | 8.03E-129 | 0.93958386 | 0.651 | 0.661 | 3.07E-124 |
| MT-ND1 | 0 | 0.9316361 | 0.862 | 0.751 | 0 |
| MT-ND2 | 0 | 0.92415049 | 0.829 | 0.703 | 0 |
| MT-CO2 | 0 | 0.89834677 | 0.928 | 0.877 | 0 |
| HSPA1A | 6.79E-184 | 0.86569281 | 0.236 | 0.119 | 2.59E-179 |
| MT-CO3 | 8.05E-260 | 0.79358458 | 0.89 | 0.847 | 3.08E-255 |
| AL627171.2 | 0 | 0.76459889 | 0.555 | 0.384 | 0 |
| AC016687.2 | 1.60E-92 | 0.7634629 | 0.299 | 0.203 | 6.13E-88 |
| SLC5A4-AS1 | 0 | 0.72527532 | 0.58 | 0.454 | 0 |
| SATB1-AS1 | 1.54E-271 | 0.69867396 | 0.592 | 0.453 | 5.90E-267 |
| RORA | 0 | 0.66453909 | 0.967 | 0.963 | 0 |
| AP003066.1 | 6.51E-135 | 0.65606551 | 0.221 | 0.118 | 2.49E-130 |
| MT-ND5 | 3.31E-255 | 0.64743513 | 0.64 | 0.511 | 1.27E-250 |
| HSP90AA1 | 7.60E-215 | 0.62902571 | 0.732 | 0.713 | 2.90E-210 |
| TSHZ2 | 1.43E-103 | 0.61039478 | 0.53 | 0.434 | 5.48E-99 |
| NEAT1 | 1.69E-102 | 0.60402945 | 0.358 | 0.269 | 6.47E-98 |
| MT-ND4L | 1.68E-185 | 0.57656163 | 0.586 | 0.482 | 6.41E-181 |
| CDH20 | 2.68E-133 | 0.57387798 | 0.459 | 0.353 | 1.03E-128 |
| DNAJB1 | 1.34E-90 | 0.57360421 | 0.25 | 0.172 | 5.12E-86 |
| RORB | 1.99E-162 | 0.54882354 | 0.567 | 0.429 | 7.61E-158 |
| LINC01322 | 6.47E-98 | 0.53641658 | 0.552 | 0.499 | 2.47E-93 |
| PTPRT | 8.12E-218 | 0.53191683 | 0.821 | 0.77 | 3.11E-213 |
| MT-CO1 | 6.36E-298 | 0.53118121 | 0.919 | 0.847 | 2.43E-293 |
| TAFA2 | 9.31E-272 | 0.53019171 | 0.921 | 0.811 | 3.56E-267 |
| EFNA5 | 0 | 0.51679473 | 0.955 | 0.927 | 0 |
| CNTN5 | 4.51E-186 | 0.50940106 | 0.877 | 0.79 | 1.72E-181 |
| CXXC4 | 8.68E-191 | 0.48787576 | 0.338 | 0.21 | 3.32E-186 |
| SNTG1 | 2.54E-303 | 0.48030664 | 0.97 | 0.949 | 9.70E-299 |
| KANSL1 | 0 | 0.47483201 | 0.886 | 0.866 | 0 |
| PLP1 | 5.66E-153 | 0.47398168 | 0.308 | 0.182 | 2.16E-148 |
| FSTL5 | 1.25E-97 | 0.45855052 | 0.469 | 0.37 | 4.77E-93 |
| CALM1 | 2.05E-156 | 0.44414083 | 0.887 | 0.901 | 7.83E-152 |
| AL356295.1 | 1.14E-27 | 0.44329658 | 0.273 | 0.231 | 4.34E-23 |
| SLCO3A1 | 2.93E-176 | 0.4231648 | 0.765 | 0.768 | 1.12E-171 |
| POU6F2 | 3.88E-124 | 0.4226534 | 0.622 | 0.493 | 1.48E-119 |
| NPTX1 | 5.40E-131 | 0.41635589 | 0.712 | 0.729 | 2.06E-126 |
| AP001977.1 | 2.41E-208 | 0.41384567 | 0.217 | 0.092 | 9.20E-204 |

| | | | | | |
|---|---|---|---|---|---|
| GRM3 | 1.78E-69 | 0.40337251 | 0.597 | 0.527 | 6.80E-65 |
| DNAJA1 | 1.32E-69 | 0.40286603 | 0.356 | 0.303 | 5.03E-65 |
| VWC2L | 4.26E-96 | 0.40136046 | 0.388 | 0.281 | 1.63E-91 |
| BNIP3L | 4.14E-161 | 0.39998739 | 0.649 | 0.633 | 1.58E-156 |
| LHFPL6 | 9.76E-114 | 0.39872629 | 0.617 | 0.529 | 3.73E-109 |
| ADAMTS2 | 4.47E-158 | 0.39591245 | 0.267 | 0.153 | 1.71E-153 |
| ZBTB16 | 8.36E-260 | 0.39401027 | 0.924 | 0.915 | 3.19E-255 |
| VAMP2 | 2.22E-148 | 0.39149948 | 0.688 | 0.637 | 8.50E-144 |
| UBB | 9.64E-104 | 0.39098712 | 0.4 | 0.327 | 3.68E-99 |
| CLSTN2 | 1.26E-208 | 0.38914549 | 0.845 | 0.741 | 4.81E-204 |
| GAB1 | 2.80E-82 | 0.38882963 | 0.714 | 0.721 | 1.07E-77 |
| SERPINB9 | 1.24E-18 | 0.38466525 | 0.149 | 0.201 | 4.74E-14 |
| EDIL3 | 6.90E-111 | 0.3840383 | 0.851 | 0.837 | 2.64E-106 |
| AC103855.2 | 4.32E-113 | 0.3823441 | 0.432 | 0.349 | 1.65E-108 |
| HIVEP3 | 1.07E-165 | 0.37995305 | 0.865 | 0.876 | 4.09E-161 |
| AUXG01000( | 7.44E-82 | 0.3770244 | 0.56 | 0.542 | 2.84E-77 |
| YWHAH | 2.02E-76 | 0.37698202 | 0.604 | 0.612 | 7.73E-72 |
| SPOCK1 | 1.66E-134 | 0.37069922 | 0.791 | 0.75 | 6.35E-130 |
| HSPA8 | 6.30E-77 | 0.36723441 | 0.329 | 0.26 | 2.41E-72 |
| HSP90AB1 | 6.60E-95 | 0.36674439 | 0.731 | 0.756 | 2.52E-90 |
| MLLT3 | 8.47E-135 | 0.3666991 | 0.858 | 0.859 | 3.24E-130 |
| PLCXD3 | 2.59E-72 | 0.36574834 | 0.347 | 0.263 | 9.90E-68 |
| ZNF365 | 9.61E-40 | 0.36461614 | 0.623 | 0.661 | 3.67E-35 |
| SEM1 | 1.20E-78 | 0.36112014 | 0.491 | 0.465 | 4.57E-74 |
| SCN1A-AS1 | 1.55E-22 | 0.35970684 | 0.277 | 0.248 | 5.94E-18 |
| ARL17B | 2.91E-270 | 0.35639292 | 0.469 | 0.28 | 1.11E-265 |
| GNAS | 1.60E-135 | 0.35637108 | 0.812 | 0.842 | 6.13E-131 |
| DCC | 5.54E-69 | 0.353364 | 0.598 | 0.512 | 2.12E-64 |
| ZNF706 | 6.05E-104 | 0.35048035 | 0.669 | 0.68 | 2.31E-99 |
| PLCH1 | 9.42E-09 | 0.34805434 | 0.159 | 0.14 | 0.00036001 |
| SNX31 | 2.18E-163 | 0.34587641 | 0.264 | 0.147 | 8.35E-159 |
| AC120193.1 | 4.48E-12 | 0.34060957 | 0.439 | 0.451 | 1.71E-07 |
| SLC35F1 | 1.39E-113 | 0.34018045 | 0.704 | 0.598 | 5.30E-109 |
| MIR29B2CH( | 7.27E-116 | 0.33579889 | 0.756 | 0.775 | 2.78E-111 |
| PELI2 | 1.73E-105 | 0.3357446 | 0.347 | 0.249 | 6.61E-101 |
| CDHR3 | 1.50E-52 | 0.33509017 | 0.434 | 0.396 | 5.73E-48 |
| ZMAT4 | 1.43E-88 | 0.33486565 | 0.801 | 0.816 | 5.48E-84 |
| RARB | 4.20E-37 | 0.33412199 | 0.481 | 0.457 | 1.61E-32 |
| RBMS3 | 1.07E-57 | 0.33299264 | 0.539 | 0.508 | 4.11E-53 |
| TMTC2 | 1.65E-44 | 0.32940154 | 0.537 | 0.501 | 6.31E-40 |
| RPS6KA5 | 7.95E-112 | 0.32869933 | 0.701 | 0.698 | 3.04E-107 |
| MIR31HG | 3.62E-59 | 0.32764767 | 0.366 | 0.302 | 1.38E-54 |
| HMGCS1 | 5.00E-35 | 0.32334003 | 0.346 | 0.327 | 1.91E-30 |
| HSPA12A | 2.89E-106 | 0.32298016 | 0.883 | 0.906 | 1.11E-101 |
| TAFA5 | 3.54E-81 | 0.32153538 | 0.634 | 0.602 | 1.35E-76 |

| | | | | | |
|---|---|---|---|---|---|
| MYT1L | 0 | 0.31989388 | 0.976 | 0.971 | 0 |
| PTPRK | 1.01E-89 | 0.31830552 | 0.857 | 0.839 | 3.85E-85 |
| CHORDC1 | 1.45E-29 | 0.31642801 | 0.415 | 0.419 | 5.54E-25 |
| PTMA | 1.01E-40 | 0.3157373 | 0.498 | 0.494 | 3.86E-36 |
| KCNJ6 | 1.11E-112 | 0.31540474 | 0.799 | 0.81 | 4.25E-108 |
| ALCAM | 2.12E-16 | 0.31217237 | 0.475 | 0.468 | 8.09E-12 |
| PRICKLE2 | 5.71E-120 | 0.31149835 | 0.815 | 0.844 | 2.18E-115 |
| CACNB4 | 1.52E-131 | 0.30966923 | 0.906 | 0.896 | 5.80E-127 |
| ST18 | 3.62E-28 | 0.30953749 | 0.154 | 0.112 | 1.39E-23 |
| SLC5A3 | 2.55E-88 | 0.30907742 | 0.313 | 0.237 | 9.73E-84 |
| INPP4B | 1.16E-50 | 0.30830692 | 0.74 | 0.739 | 4.43E-46 |
| CALM2 | 7.18E-75 | 0.3070727 | 0.782 | 0.774 | 2.74E-70 |
| HIBCH | 1.37E-49 | 0.3070182 | 0.621 | 0.662 | 5.25E-45 |
| CASC15 | 2.57E-30 | 0.30606164 | 0.616 | 0.632 | 9.81E-26 |
| HMGB1 | 4.45E-88 | 0.30506071 | 0.812 | 0.843 | 1.70E-83 |
| TRHDE | 3.51E-47 | 0.30463559 | 0.645 | 0.636 | 1.34E-42 |
| AC092422.1 | 1.90E-32 | 0.30391647 | 0.303 | 0.255 | 7.26E-28 |
| HP1BP3 | 1.34E-105 | 0.30357576 | 0.697 | 0.717 | 5.12E-101 |
| CNTNAP2 | 8.42E-159 | 0.30205624 | 0.992 | 0.991 | 3.22E-154 |
| ZMYND8 | 1.20E-103 | 0.29981737 | 0.798 | 0.839 | 4.58E-99 |
| AC084816.1 | 9.96E-20 | 0.29941802 | 0.307 | 0.277 | 3.81E-15 |
| NFIC | 1.71E-48 | 0.29933435 | 0.542 | 0.555 | 6.52E-44 |
| ANKH | 8.60E-102 | 0.29910088 | 0.78 | 0.815 | 3.29E-97 |
| CCDC144A | 1.47E-73 | 0.29900107 | 0.717 | 0.687 | 5.61E-69 |
| ELOVL5 | 1.48E-15 | 0.29848539 | 0.284 | 0.271 | 5.66E-11 |
| CFAP161 | 1.59E-70 | 0.29691288 | 0.295 | 0.223 | 6.08E-66 |
| QKI | 7.04E-08 | 0.29660849 | 0.538 | 0.582 | 0.00268991 |
| SSBP3 | 1.56E-108 | 0.29611547 | 0.78 | 0.815 | 5.97E-104 |
| TMEM14B | 2.05E-108 | 0.29569509 | 0.736 | 0.708 | 7.84E-104 |
| PRKAG2 | 1.26E-127 | 0.29534641 | 0.858 | 0.882 | 4.83E-123 |
| RALYL | 9.45E-211 | 0.29529145 | 0.993 | 0.99 | 3.61E-206 |
| TMEM161B-/ | 9.18E-129 | 0.29463598 | 0.896 | 0.908 | 3.51E-124 |
| UBC | 1.21E-31 | 0.29439231 | 0.4 | 0.386 | 4.63E-27 |
| FXYD6 | 4.56E-77 | 0.29417795 | 0.641 | 0.626 | 1.74E-72 |
| FKBP5 | 3.73E-20 | 0.29322772 | 0.372 | 0.362 | 1.42E-15 |
| AC093865.1 | 3.38E-54 | 0.29296033 | 0.264 | 0.204 | 1.29E-49 |
| HSPB1 | 1.78E-81 | 0.29124317 | 0.156 | 0.087 | 6.79E-77 |
| PRNP | 9.40E-69 | 0.29055809 | 0.686 | 0.703 | 3.59E-64 |
| TSHZ3 | 4.06E-42 | 0.2902427 | 0.504 | 0.49 | 1.55E-37 |
| LINC00486 | 5.12E-22 | 0.28998974 | 0.62 | 0.665 | 1.96E-17 |
| KHDRBS2-OT | 3.31E-55 | 0.28928417 | 0.588 | 0.565 | 1.26E-50 |
| KCND3 | 1.12E-75 | 0.28909327 | 0.881 | 0.906 | 4.28E-71 |
| PPP2R2C | 9.71E-143 | 0.28761249 | 0.899 | 0.917 | 3.71E-138 |
| SPP1 | 5.76E-171 | 0.28565284 | 0.139 | 0.044 | 2.20E-166 |
| TMSB10 | 4.94E-10 | 0.2851715 | 0.442 | 0.472 | 1.89E-05 |

| | | | | | |
|---|---|---|---|---|---|
| MAPRE2 | 2.99E-75 | 0.28487102 | 0.783 | 0.825 | 1.14E-70 |
| OIP5-AS1 | 2.43E-58 | 0.28478202 | 0.58 | 0.561 | 9.30E-54 |
| MARCKS | 3.38E-51 | 0.28332058 | 0.395 | 0.353 | 1.29E-46 |
| MSH4 | 2.66E-108 | 0.28223474 | 0.248 | 0.158 | 1.02E-103 |
| ARMCX5-GPI | 1.37E-67 | 0.28199532 | 0.535 | 0.502 | 5.23E-63 |
| LRRC4C | 1.21E-23 | 0.27912831 | 0.888 | 0.916 | 4.63E-19 |
| UTP23 | 1.24E-46 | 0.27860505 | 0.541 | 0.549 | 4.73E-42 |
| GRIP1 | 2.34E-35 | 0.27814817 | 0.71 | 0.718 | 8.93E-31 |
| CXXC5 | 3.36E-42 | 0.27758468 | 0.436 | 0.419 | 1.29E-37 |
| PDZRN3 | 1.92E-66 | 0.27646446 | 0.808 | 0.805 | 7.33E-62 |
| AC109466.1 | 1.96E-45 | 0.27603187 | 0.425 | 0.354 | 7.51E-41 |
| WWOX | 1.28E-42 | 0.27590757 | 0.791 | 0.81 | 4.90E-38 |
| RUNX1 | 5.06E-47 | 0.27554492 | 0.213 | 0.155 | 1.94E-42 |
| HSPA1B | 2.79E-110 | 0.27321822 | 0.125 | 0.053 | 1.07E-105 |
| BANP | 3.91E-53 | 0.27318617 | 0.395 | 0.362 | 1.50E-48 |
| ALDH1A1 | 2.79E-36 | 0.27221588 | 0.156 | 0.109 | 1.06E-31 |
| CCND3 | 1.94E-24 | 0.27219416 | 0.477 | 0.491 | 7.41E-20 |
| UPP2 | 2.59E-48 | 0.27149889 | 0.629 | 0.635 | 9.91E-44 |
| MTRNR2L8 | 3.04E-71 | 0.27133841 | 0.181 | 0.114 | 1.16E-66 |
| CELF4 | 5.62E-139 | 0.27106215 | 0.913 | 0.933 | 2.15E-134 |
| BASP1 | 7.88E-108 | 0.27087251 | 0.856 | 0.88 | 3.01E-103 |
| IGF1R | 2.12E-93 | 0.27025525 | 0.861 | 0.887 | 8.11E-89 |
| PAG1 | 1.52E-25 | 0.26960569 | 0.418 | 0.414 | 5.80E-21 |
| YWHAB | 1.04E-42 | 0.26954833 | 0.641 | 0.678 | 3.99E-38 |
| AC092691.1 | 1.74E-63 | 0.26883006 | 0.857 | 0.879 | 6.63E-59 |
| ELK1 | 5.37E-31 | 0.26665071 | 0.298 | 0.266 | 2.05E-26 |
| KCNQ3 | 1.29E-108 | 0.2664292 | 0.912 | 0.912 | 4.92E-104 |
| ZSCAN31 | 3.37E-58 | 0.26521265 | 0.356 | 0.301 | 1.29E-53 |
| MSMO1 | 6.72E-43 | 0.2651319 | 0.272 | 0.229 | 2.57E-38 |
| DPH6-DT | 3.99E-27 | 0.2644346 | 0.419 | 0.407 | 1.52E-22 |
| DNAJB6 | 1.20E-23 | 0.26402947 | 0.463 | 0.481 | 4.60E-19 |
| YWHAG | 6.73E-31 | 0.26312924 | 0.653 | 0.704 | 2.57E-26 |
| SMARCD3 | 1.61E-47 | 0.2625505 | 0.354 | 0.315 | 6.15E-43 |
| RHBDL3 | 2.54E-37 | 0.26196778 | 0.554 | 0.559 | 9.72E-33 |
| PANTR1 | 1.37E-30 | 0.26063956 | 0.549 | 0.547 | 5.25E-26 |
| GAPDH | 4.66E-30 | 0.26034843 | 0.541 | 0.553 | 1.78E-25 |
| SOX5 | 2.37E-28 | 0.25892022 | 0.716 | 0.723 | 9.06E-24 |
| CEMIP | 6.11E-23 | 0.2581093 | 0.528 | 0.538 | 2.34E-18 |
| CRYAB | 9.51E-143 | 0.25798842 | 0.152 | 0.061 | 3.64E-138 |
| PTGES3 | 8.01E-29 | 0.25751113 | 0.456 | 0.464 | 3.06E-24 |
| SAMD5 | 1.51E-27 | 0.25676032 | 0.354 | 0.315 | 5.76E-23 |
| PEG10 | 5.77E-44 | 0.25501671 | 0.287 | 0.24 | 2.21E-39 |
| PTGDS | 1.85E-23 | 0.25370209 | 0.304 | 0.27 | 7.05E-19 |
| AJAP1 | 8.96E-45 | 0.25340082 | 0.649 | 0.686 | 3.42E-40 |
| DNAJC1 | 7.77E-33 | 0.25179114 | 0.633 | 0.672 | 2.97E-28 |

| | | | | | |
|---|---|---|---|---|---|
| KCNH7 | 1.50E-19 | 0.25147255 | 0.742 | 0.774 | 5.72E-15 |
| NCAM2 | 3.43E-64 | 0.25123859 | 0.953 | 0.946 | 1.31E-59 |
| PAPOLA | 4.10E-48 | 0.25109887 | 0.606 | 0.638 | 1.57E-43 |
| SYN1 | 1.88E-40 | 0.25106034 | 0.574 | 0.589 | 7.18E-36 |
| MAP1A | 9.81E-43 | 0.2507198 | 0.5 | 0.498 | 3.75E-38 |
| SRCIN1 | 3.23E-51 | 0.25061689 | 0.676 | 0.727 | 1.24E-46 |
| CLIP2 | 7.04E-32 | 0.25013246 | 0.616 | 0.66 | 2.69E-27 |
| SIDT1 | 3.59E-176 | -0.250064 | 0.491 | 0.694 | 1.37E-171 |
| MICU3 | 3.33E-181 | -0.2507075 | 0.644 | 0.828 | 1.27E-176 |
| RSRP1 | 1.87E-186 | -0.2514949 | 0.608 | 0.804 | 7.17E-182 |
| INVS | 1.67E-187 | -0.2524559 | 0.549 | 0.763 | 6.37E-183 |
| TENT2 | 2.53E-234 | -0.25277 | 0.501 | 0.733 | 9.66E-230 |
| CBLN2 | 4.02E-140 | -0.2529955 | 0.518 | 0.685 | 1.54E-135 |
| ANO4 | 2.54E-202 | -0.2533065 | 0.363 | 0.584 | 9.70E-198 |
| AC110023.1 | 3.79E-111 | -0.2548511 | 0.368 | 0.518 | 1.45E-106 |
| HTR4 | 1.58E-237 | -0.2549664 | 0.203 | 0.405 | 6.03E-233 |
| ZBBX | 3.02E-239 | -0.2556369 | 0.227 | 0.437 | 1.16E-234 |
| MIR100HG | 3.32E-114 | -0.2563514 | 0.702 | 0.787 | 1.27E-109 |
| GOLT1B | 0 | -0.2564384 | 0.252 | 0.514 | 0 |
| MTR | 2.71E-210 | -0.2564566 | 0.498 | 0.727 | 1.03E-205 |
| CXXC4-AS1 | 1.54E-204 | -0.2596148 | 0.356 | 0.576 | 5.87E-200 |
| PRKN | 2.14E-136 | -0.2598956 | 0.811 | 0.922 | 8.18E-132 |
| SCPEP1 | 1.57E-294 | -0.2601831 | 0.243 | 0.477 | 5.99E-290 |
| AC117453.1 | 1.08E-228 | -0.2608532 | 0.063 | 0.201 | 4.14E-224 |
| LRRTM4 | 3.30E-125 | -0.2613697 | 0.976 | 0.975 | 1.26E-120 |
| AC025887.2 | 1.53E-260 | -0.2623283 | 0.104 | 0.279 | 5.85E-256 |
| AC245060.5 | 9.01E-267 | -0.2623535 | 0.144 | 0.34 | 3.44E-262 |
| AC107223.1 | 2.16E-41 | -0.2629696 | 0.636 | 0.707 | 8.26E-37 |
| ITGA9 | 2.69E-186 | -0.2647975 | 0.384 | 0.585 | 1.03E-181 |
| PWRN1 | 3.03E-102 | -0.2653268 | 0.254 | 0.388 | 1.16E-97 |
| CCDC91 | 1.13E-171 | -0.2662982 | 0.609 | 0.796 | 4.31E-167 |
| CDH11 | 2.98E-200 | -0.2680075 | 0.394 | 0.614 | 1.14E-195 |
| TBC1D32 | 3.28E-201 | -0.2683989 | 0.588 | 0.793 | 1.25E-196 |
| VLDLR-AS1 | 2.60E-239 | -0.2702956 | 0.279 | 0.498 | 9.93E-235 |
| CCPG1 | 6.59E-215 | -0.2704997 | 0.703 | 0.89 | 2.52E-210 |
| AL365295.1 | 3.90E-162 | -0.2708257 | 0.247 | 0.412 | 1.49E-157 |
| SVOP | 5.85E-248 | -0.2714094 | 0.407 | 0.657 | 2.24E-243 |
| LINC00299 | 1.97E-148 | -0.2720639 | 0.267 | 0.432 | 7.52E-144 |
| GNAL | 3.55E-159 | -0.2722686 | 0.324 | 0.501 | 1.36E-154 |
| AL445433.2 | 1.07E-239 | -0.2723664 | 0.242 | 0.454 | 4.07E-235 |
| NOXA1 | 0 | -0.2726983 | 0.26 | 0.511 | 0 |
| RCAN2 | 4.86E-137 | -0.2727436 | 0.733 | 0.861 | 1.86E-132 |
| NIPAL2 | 9.33E-169 | -0.273774 | 0.595 | 0.781 | 3.57E-164 |
| ANKRD30BL | 2.24E-186 | -0.2738985 | 0.615 | 0.809 | 8.57E-182 |
| CAMK2D | 1.70E-104 | -0.2749232 | 0.65 | 0.777 | 6.48E-100 |

| | | | | | |
|---|---|---|---|---|---|
| SORCS3 | 1.53E-129 | -0.2764468 | 0.543 | 0.712 | 5.86E-125 |
| GAREM1 | 3.56E-205 | -0.278397 | 0.389 | 0.61 | 1.36E-200 |
| SNRNP70 | 3.18E-220 | -0.2788707 | 0.637 | 0.836 | 1.22E-215 |
| ANKRD28 | 2.47E-222 | -0.2793923 | 0.579 | 0.783 | 9.42E-218 |
| GRIA4 | 8.93E-109 | -0.2798784 | 0.873 | 0.917 | 3.41E-104 |
| TLK1 | 1.15E-210 | -0.2803651 | 0.547 | 0.76 | 4.40E-206 |
| COL12A1 | 5.81E-167 | -0.2804494 | 0.097 | 0.224 | 2.22E-162 |
| DCLK1 | 1.09E-252 | -0.2810132 | 0.962 | 0.979 | 4.18E-248 |
| NECTIN3-AS1 | 0 | -0.2819035 | 0.087 | 0.288 | 0 |
| ZRANB3 | 8.82E-240 | -0.2824466 | 0.464 | 0.696 | 3.37E-235 |
| LINC02552 | 8.36E-284 | -0.2833591 | 0.036 | 0.177 | 3.19E-279 |
| MPPED2 | 2.97E-229 | -0.283989 | 0.513 | 0.734 | 1.13E-224 |
| AP000829.1 | 8.91E-141 | -0.2847519 | 0.248 | 0.402 | 3.40E-136 |
| MEIS3 | 0 | -0.2870321 | 0.253 | 0.505 | 0 |
| NPAS3 | 3.68E-141 | -0.2878248 | 0.701 | 0.837 | 1.41E-136 |
| CAMK2A | 1.30E-195 | -0.289574 | 0.883 | 0.956 | 4.99E-191 |
| GIPC1 | 6.70E-301 | -0.2899823 | 0.221 | 0.452 | 2.56E-296 |
| MAPK4 | 8.38E-232 | -0.2902496 | 0.404 | 0.633 | 3.20E-227 |
| STAT4 | 7.82E-228 | -0.290797 | 0.408 | 0.635 | 2.99E-223 |
| BDH1 | 3.09E-264 | -0.290809 | 0.435 | 0.69 | 1.18E-259 |
| ST6GAL2 | 1.27E-237 | -0.2919828 | 0.377 | 0.61 | 4.84E-233 |
| MCHR2 | 4.36E-235 | -0.292138 | 0.311 | 0.533 | 1.67E-230 |
| ATRNL1 | 1.78E-228 | -0.2932719 | 0.96 | 0.983 | 6.78E-224 |
| LINC01250 | 5.10E-128 | -0.2949198 | 0.761 | 0.858 | 1.95E-123 |
| AC024901.1 | 1.24E-174 | -0.2959928 | 0.405 | 0.597 | 4.73E-170 |
| HDAC9 | 2.11E-169 | -0.2960371 | 0.708 | 0.849 | 8.06E-165 |
| AC087280.2 | 0 | -0.296158 | 0.161 | 0.414 | 0 |
| TANC2 | 2.78E-200 | -0.2984536 | 0.864 | 0.936 | 1.06E-195 |
| LINC01500 | 2.00E-125 | -0.2984873 | 0.117 | 0.23 | 7.63E-121 |
| CDH12 | 1.23E-104 | -0.3005212 | 0.84 | 0.873 | 4.69E-100 |
| GPC1 | 0 | -0.3012997 | 0.374 | 0.638 | 0 |
| GLIS3 | 9.44E-126 | -0.3021487 | 0.222 | 0.362 | 3.61E-121 |
| TMEM179 | 1.00E-263 | -0.3021513 | 0.213 | 0.426 | 3.83E-259 |
| GRIA3 | 1.79E-214 | -0.3028734 | 0.839 | 0.931 | 6.85E-210 |
| RAPGEF5 | 1.72E-240 | -0.3030821 | 0.897 | 0.952 | 6.58E-236 |
| AC005753.2 | 5.52E-225 | -0.3036784 | 0.179 | 0.359 | 2.11E-220 |
| HGSNAT | 0 | -0.3039036 | 0.385 | 0.657 | 0 |
| CADPS | 4.18E-178 | -0.3055641 | 0.915 | 0.964 | 1.60E-173 |
| CPNE4 | 5.32E-66 | -0.3065774 | 0.579 | 0.719 | 2.03E-61 |
| AL391807.1 | 0 | -0.3069011 | 0.327 | 0.595 | 0 |
| AC006504.5 | 7.40E-224 | -0.307239 | 0.443 | 0.666 | 2.83E-219 |
| PDE4B | 9.29E-218 | -0.3073408 | 0.485 | 0.695 | 3.55E-213 |
| SYTL2 | 1.11E-264 | -0.3074287 | 0.521 | 0.748 | 4.24E-260 |
| DPH6 | 1.66E-262 | -0.3075039 | 0.293 | 0.525 | 6.33E-258 |
| GULP1 | 7.85E-174 | -0.3079809 | 0.308 | 0.487 | 3.00E-169 |

| | | | | | |
|---|---|---|---|---|---|
| SPRED1 | 0 | -0.3119505 | 0.35 | 0.624 | 0 |
| PAK1 | 2.42E-281 | -0.3149305 | 0.694 | 0.867 | 9.26E-277 |
| WSB1 | 6.74E-270 | -0.3169949 | 0.51 | 0.75 | 2.58E-265 |
| CORO6 | 0 | -0.3171952 | 0.301 | 0.564 | 0 |
| SORBS1 | 4.13E-241 | -0.3180745 | 0.882 | 0.951 | 1.58E-236 |
| FBXW7 | 6.35E-272 | -0.3196926 | 0.82 | 0.929 | 2.43E-267 |
| NECTIN3 | 5.50E-209 | -0.3202612 | 0.487 | 0.687 | 2.10E-204 |
| PCED1B | 0 | -0.3204962 | 0.09 | 0.285 | 0 |
| LINC02822 | 3.51E-174 | -0.3215477 | 0.132 | 0.277 | 1.34E-169 |
| DTNA | 4.31E-232 | -0.3218052 | 0.824 | 0.931 | 1.65E-227 |
| RFX3-AS1 | 0 | -0.3230029 | 0.138 | 0.379 | 0 |
| AC108519.1 | 3.16E-230 | -0.3235728 | 0.48 | 0.685 | 1.21E-225 |
| AC092683.1 | 1.08E-270 | -0.3236814 | 0.331 | 0.569 | 4.14E-266 |
| NEK10 | 3.93E-273 | -0.3248542 | 0.548 | 0.775 | 1.50E-268 |
| PRKCA | 1.25E-190 | -0.3255524 | 0.617 | 0.781 | 4.77E-186 |
| GPATCH2 | 2.07E-266 | -0.3263045 | 0.412 | 0.657 | 7.92E-262 |
| GLCE | 0 | -0.3266058 | 0.283 | 0.551 | 0 |
| AC092902.4 | 5.16E-265 | -0.3266637 | 0.212 | 0.42 | 1.97E-260 |
| LMCD1-AS1 | 6.07E-250 | -0.3274936 | 0.171 | 0.365 | 2.32E-245 |
| TMEM241 | 7.05E-284 | -0.3275795 | 0.527 | 0.769 | 2.70E-279 |
| BABAM2 | 1.32E-253 | -0.3286913 | 0.697 | 0.872 | 5.04E-249 |
| LAMB1 | 0 | -0.3287465 | 0.258 | 0.504 | 0 |
| MEG8 | 6.18E-197 | -0.3302223 | 0.792 | 0.913 | 2.36E-192 |
| LINC00326 | 7.58E-147 | -0.3306427 | 0.109 | 0.233 | 2.90E-142 |
| PDE4D | 0 | -0.3351693 | 0.991 | 0.992 | 0 |
| MEG3 | 5.54E-292 | -0.3354952 | 0.979 | 0.968 | 2.12E-287 |
| SLC4A4 | 7.12E-285 | -0.3361745 | 0.248 | 0.474 | 2.72E-280 |
| DCLK2 | 3.49E-264 | -0.336798 | 0.714 | 0.877 | 1.33E-259 |
| ZFPM2 | 2.99E-157 | -0.3370018 | 0.829 | 0.89 | 1.14E-152 |
| TASP1 | 0 | -0.3371266 | 0.481 | 0.732 | 0 |
| DIAPH2 | 2.97E-218 | -0.3372887 | 0.606 | 0.784 | 1.13E-213 |
| TRIM9 | 0 | -0.3379434 | 0.912 | 0.97 | 0 |
| GARNL3 | 5.70E-230 | -0.3381064 | 0.587 | 0.783 | 2.18E-225 |
| LINC01378 | 1.26E-172 | -0.3384542 | 0.426 | 0.615 | 4.80E-168 |
| PID1 | 3.76E-192 | -0.3402655 | 0.403 | 0.598 | 1.44E-187 |
| RFLNA | 0 | -0.3406719 | 0.333 | 0.594 | 0 |
| RGS7 | 0 | -0.345035 | 0.965 | 0.972 | 0 |
| LY86-AS1 | 1.21E-198 | -0.3469466 | 0.658 | 0.826 | 4.64E-194 |
| AC110296.1 | 1.76E-214 | -0.3482658 | 0.173 | 0.347 | 6.74E-210 |
| AC007389.1 | 0 | -0.3513165 | 0.155 | 0.383 | 0 |
| STXBP5L | 0 | -0.3520885 | 0.966 | 0.982 | 0 |
| PAM | 4.06E-227 | -0.3523532 | 0.665 | 0.826 | 1.55E-222 |
| ITFG1 | 1.40E-277 | -0.3531587 | 0.759 | 0.902 | 5.34E-273 |
| ST6GALNAC5 | 1.38E-167 | -0.3532192 | 0.804 | 0.882 | 5.29E-163 |
| GOLGA8A | 0 | -0.3540635 | 0.463 | 0.727 | 0 |

| | | | | | |
|---|---|---|---|---|---|
| LINC02296 | 5.55E-282 | -0.3579632 | 0.139 | 0.339 | 2.12E-277 |
| LUC7L2 | 2.50E-283 | -0.3583076 | 0.583 | 0.79 | 9.55E-279 |
| CCSER1 | 0 | -0.3597481 | 0.953 | 0.976 | 0 |
| CYP1B1-AS1 | 1.83E-234 | -0.3599058 | 0.308 | 0.52 | 7.00E-230 |
| UNC80 | 0 | -0.3620267 | 0.814 | 0.937 | 0 |
| ERC2 | 5.24E-277 | -0.3649039 | 0.958 | 0.976 | 2.00E-272 |
| TTC8 | 0 | -0.366577 | 0.338 | 0.627 | 0 |
| GALNT13 | 2.91E-185 | -0.3669079 | 0.587 | 0.76 | 1.11E-180 |
| SCHLAP1 | 4.01E-169 | -0.3707879 | 0.243 | 0.412 | 1.53E-164 |
| EML6 | 8.06E-305 | -0.37189 | 0.799 | 0.916 | 3.08E-300 |
| RASGRF2 | 3.03E-161 | -0.3746321 | 0.56 | 0.713 | 1.16E-156 |
| CIRBP | 0 | -0.3770708 | 0.351 | 0.648 | 0 |
| MEIS2 | 2.58E-205 | -0.3776076 | 0.366 | 0.564 | 9.86E-201 |
| FRMD6 | 0 | -0.3793501 | 0.157 | 0.401 | 0 |
| LINC01184 | 0 | -0.3835837 | 0.558 | 0.805 | 0 |
| AC002460.2 | 6.89E-197 | -0.3856267 | 0.351 | 0.544 | 2.64E-192 |
| AQP4-AS1 | 1.69E-251 | -0.387233 | 0.59 | 0.798 | 6.46E-247 |
| GMDS-DT | 0 | -0.3903596 | 0.47 | 0.733 | 0 |
| ENC1 | 0 | -0.3921997 | 0.592 | 0.813 | 0 |
| LINC01090 | 4.62E-158 | -0.3952199 | 0.053 | 0.157 | 1.77E-153 |
| HS6ST3 | 4.26E-200 | -0.3984604 | 0.761 | 0.89 | 1.63E-195 |
| SHC3 | 0 | -0.3990632 | 0.571 | 0.825 | 0 |
| KCNN2 | 0 | -0.401673 | 0.386 | 0.654 | 0 |
| DGKB | 4.93E-231 | -0.4096876 | 0.79 | 0.912 | 1.89E-226 |
| LINC00276 | 6.80E-250 | -0.410066 | 0.212 | 0.415 | 2.60E-245 |
| PEX5L | 1.83E-294 | -0.4196692 | 0.839 | 0.93 | 6.99E-290 |
| ZNF804B | 2.32E-141 | -0.4214185 | 0.286 | 0.454 | 8.89E-137 |
| PLCB1 | 0 | -0.4228632 | 0.975 | 0.986 | 0 |
| NDST3 | 0 | -0.4238617 | 0.547 | 0.789 | 0 |
| NELL1 | 0 | -0.4316273 | 0.76 | 0.902 | 0 |
| PTCHD1-AS | 1.06E-300 | -0.4317762 | 0.475 | 0.729 | 4.06E-296 |
| KIAA1217 | 1.21E-220 | -0.4318066 | 0.542 | 0.742 | 4.61E-216 |
| SPRY4-AS1 | 0 | -0.433002 | 0.06 | 0.32 | 0 |
| AC097512.1 | 0 | -0.4333869 | 0.089 | 0.274 | 0 |
| XIST | 6.87E-103 | -0.4436912 | 0.475 | 0.559 | 2.63E-98 |
| AC007368.1 | 2.66E-114 | -0.444801 | 0.392 | 0.507 | 1.02E-109 |
| PDE7B | 1.13E-228 | -0.448358 | 0.414 | 0.605 | 4.34E-224 |
| ICA1 | 0 | -0.4571644 | 0.599 | 0.854 | 0 |
| CA10 | 3.64E-264 | -0.4591861 | 0.51 | 0.729 | 1.39E-259 |
| SGCD | 2.10E-293 | -0.4617056 | 0.574 | 0.798 | 8.04E-289 |
| TESPA1 | 0 | -0.4664151 | 0.416 | 0.645 | 0 |
| ADAMTS3 | 0 | -0.4675975 | 0.279 | 0.528 | 0 |
| ASIC2 | 4.49E-237 | -0.4687936 | 0.864 | 0.953 | 1.72E-232 |
| AGBL1 | 1.43E-221 | -0.4737134 | 0.081 | 0.225 | 5.47E-217 |
| AC073365.1 | 5.08E-288 | -0.4763928 | 0.481 | 0.69 | 1.94E-283 |

| | | | | | |
|---|---|---|---|---|---|
| AC009041.2 | 0 | -0.4926706 | 0.312 | 0.534 | 0 |
| GRID2 | 2.65E-287 | -0.4934601 | 0.63 | 0.808 | 1.01E-282 |
| DOK6 | 0 | -0.4939097 | 0.576 | 0.794 | 0 |
| AC013287.1 | 1.22E-177 | -0.4944727 | 0.597 | 0.721 | 4.68E-173 |
| ROBO1 | 5.32E-270 | -0.497647 | 0.471 | 0.682 | 2.03E-265 |
| NRXN3 | 0 | -0.5130145 | 0.98 | 0.992 | 0 |
| LINC00507 | 0 | -0.5292408 | 0.194 | 0.45 | 0 |
| AJ009632.2 | 3.64E-138 | -0.5342487 | 0.187 | 0.321 | 1.39E-133 |
| ROBO2 | 2.47E-247 | -0.5381262 | 0.721 | 0.887 | 9.45E-243 |
| ADAMTSL1 | 4.54E-24 | -0.5384884 | 0.215 | 0.266 | 1.74E-19 |
| PDZD2 | 1.19E-250 | -0.5414037 | 0.666 | 0.827 | 4.56E-246 |
| LINC01331 | 4.21E-289 | -0.5446337 | 0.246 | 0.457 | 1.61E-284 |
| RPH3A | 0 | -0.5682806 | 0.283 | 0.648 | 0 |
| GOLGA8B | 0 | -0.5688665 | 0.379 | 0.73 | 0 |
| CDH9 | 4.35E-294 | -0.572013 | 0.555 | 0.741 | 1.66E-289 |
| COL24A1 | 0 | -0.5889388 | 0.203 | 0.484 | 0 |
| AC011246.1 | 1.57E-151 | -0.6177613 | 0.285 | 0.435 | 5.99E-147 |
| GPC6 | 1.39E-235 | -0.6347158 | 0.448 | 0.632 | 5.32E-231 |
| TARBP1 | 0 | -0.6470169 | 0.676 | 0.918 | 0 |
| AC011287.1 | 0 | -0.6488429 | 0.587 | 0.801 | 0 |
| AC105402.3 | 1.10E-168 | -0.6811886 | 0.06 | 0.168 | 4.21E-164 |
| MLIP | 0 | -0.7188152 | 0.66 | 0.899 | 0 |
| AC034268.2 | 0 | -0.7277934 | 0.292 | 0.535 | 0 |
| AL157944.1 | 7.37E-285 | -0.7433861 | 0.289 | 0.48 | 2.82E-280 |
| PDE10A | 0 | -0.8816856 | 0.451 | 0.832 | 0 |
| AC093607.1 | 0 | -0.9100365 | 0.159 | 0.405 | 0 |
| LINC02306 | 0 | -1.0056016 | 0.258 | 0.531 | 0 |

**Inhibitory Neuron DEGs**

| | p_val | avg_log2FC | pct.1 | pct.2 | p_val_adj |
|---|---|---|---|---|---|
| HSPH1 | 7.96E-58 | 1.45948368 | 0.454 | 0.433 | 3.04E-53 |
| HSPA1A | 1.33E-85 | 1.34626734 | 0.218 | 0.121 | 5.10E-81 |
| AL627171.2 | 0 | 1.19457821 | 0.559 | 0.332 | 0 |
| PLCG2 | 3.52E-152 | 1.0890464 | 0.681 | 0.592 | 1.35E-147 |
| MT-ND3 | 9.03E-240 | 0.94130324 | 0.898 | 0.824 | 3.45E-235 |
| DNAJB1 | 1.69E-39 | 0.862522 | 0.189 | 0.13 | 6.46E-35 |
| MT-CYB | 1.14E-193 | 0.81985091 | 0.811 | 0.719 | 4.37E-189 |
| NEAT1 | 9.29E-16 | 0.69677222 | 0.504 | 0.524 | 3.55E-11 |
| MT-ND2 | 2.99E-135 | 0.6587271 | 0.769 | 0.7 | 1.14E-130 |
| MT-ND1 | 2.06E-117 | 0.62645629 | 0.786 | 0.72 | 7.86E-113 |
| HSP90AA1 | 1.44E-33 | 0.62314027 | 0.684 | 0.711 | 5.51E-29 |
| MT-ATP6 | 2.88E-95 | 0.61414512 | 0.884 | 0.844 | 1.10E-90 |
| AC132803.1 | 1.58E-23 | 0.6071302 | 0.297 | 0.242 | 6.06E-19 |
| MT-ND4L | 8.76E-82 | 0.59092578 | 0.606 | 0.55 | 3.35E-77 |
| CCDC144A | 4.24E-112 | 0.58919 | 0.627 | 0.558 | 1.62E-107 |
| PLP1 | 6.86E-92 | 0.56654474 | 0.292 | 0.169 | 2.62E-87 |
| HMGB1 | 1.48E-111 | 0.56519682 | 0.738 | 0.765 | 5.66E-107 |
| MT-ND5 | 3.07E-58 | 0.54443262 | 0.593 | 0.555 | 1.17E-53 |
| MTRNR2L12 | 1.04E-94 | 0.53861418 | 0.766 | 0.725 | 3.99E-90 |
| MT-ND4 | 1.27E-87 | 0.53767912 | 0.871 | 0.817 | 4.84E-83 |
| SYN3 | 7.63E-151 | 0.52233726 | 0.826 | 0.793 | 2.92E-146 |
| MT-CO2 | 3.02E-104 | 0.51711505 | 0.919 | 0.876 | 1.16E-99 |
| SPP1 | 9.87E-67 | 0.50095888 | 0.158 | 0.075 | 3.77E-62 |
| SLC5A4-AS1 | 2.60E-46 | 0.47900383 | 0.298 | 0.229 | 9.95E-42 |
| EEF1A1 | 1.56E-26 | 0.45364804 | 0.558 | 0.591 | 5.95E-22 |
| SEM1 | 4.14E-37 | 0.44320211 | 0.495 | 0.499 | 1.58E-32 |
| HSPB1 | 1.88E-51 | 0.44049378 | 0.107 | 0.047 | 7.17E-47 |
| ELOVL5 | 2.35E-07 | 0.43907354 | 0.278 | 0.282 | 0.008966 |
| FLRT2 | 1.10E-30 | 0.43359691 | 0.687 | 0.72 | 4.20E-26 |
| CHORDC1 | 1.73E-17 | 0.43041972 | 0.319 | 0.309 | 6.59E-13 |
| SEZ6L | 5.35E-23 | 0.40645224 | 0.575 | 0.572 | 2.04E-18 |
| KANSL1 | 3.62E-98 | 0.40600077 | 0.761 | 0.801 | 1.38E-93 |
| HMGCS1 | 1.02E-13 | 0.4022477 | 0.293 | 0.283 | 3.89E-09 |
| CRYAB | 3.75E-31 | 0.39923461 | 0.171 | 0.117 | 1.43E-26 |
| MT-CO3 | 4.76E-45 | 0.39758383 | 0.832 | 0.815 | 1.82E-40 |
| ST18 | 1.10E-07 | 0.3933631 | 0.162 | 0.138 | 0.00419026 |
| ARL17B | 1.75E-63 | 0.37761877 | 0.347 | 0.242 | 6.69E-59 |
| MSI2 | 4.08E-47 | 0.36851297 | 0.741 | 0.791 | 1.56E-42 |
| OIP5-AS1 | 5.00E-30 | 0.36829488 | 0.576 | 0.579 | 1.91E-25 |
| SPECC1 | 1.03E-36 | 0.36303084 | 0.66 | 0.705 | 3.96E-32 |
| PTGDS | 3.84E-18 | 0.36175239 | 0.314 | 0.272 | 1.47E-13 |
| RBFOX3 | 1.80E-59 | 0.35612818 | 0.773 | 0.806 | 6.88E-55 |
| HIBCH | 6.99E-15 | 0.3496029 | 0.548 | 0.609 | 2.67E-10 |

| | | | | | |
|---|---|---|---|---|---|
| AC008571.2 | 1.04E-20 | 0.34597728 | 0.137 | 0.095 | 3.98E-16 |
| TENM4 | 1.28E-47 | 0.34218034 | 0.764 | 0.78 | 4.89E-43 |
| SLC5A3 | 3.97E-20 | 0.34192387 | 0.356 | 0.335 | 1.52E-15 |
| EGFR | 2.20E-14 | 0.33816594 | 0.558 | 0.565 | 8.41E-10 |
| LHFPL2 | 6.61E-21 | 0.33214906 | 0.251 | 0.211 | 2.52E-16 |
| MICAL3 | 3.45E-68 | 0.33033005 | 0.822 | 0.872 | 1.32E-63 |
| AC073050.1 | 5.34E-09 | 0.32609704 | 0.688 | 0.752 | 0.0002043 |
| DSCAML1 | 1.94E-53 | 0.32370822 | 0.803 | 0.827 | 7.40E-49 |
| HSPD1 | 4.86E-12 | 0.32368318 | 0.295 | 0.4 | 1.86E-07 |
| DPH6-DT | 1.76E-14 | 0.31844841 | 0.298 | 0.275 | 6.74E-10 |
| TAFA5 | 2.48E-17 | 0.31382882 | 0.583 | 0.604 | 9.47E-13 |
| CFAP161 | 1.65E-09 | 0.31287778 | 0.417 | 0.44 | 6.30E-05 |
| ZBTB16 | 7.97E-38 | 0.3101973 | 0.788 | 0.832 | 3.05E-33 |
| KCNJ6 | 2.70E-13 | 0.30475515 | 0.608 | 0.668 | 1.03E-08 |
| ZMYND8 | 6.06E-28 | 0.30349908 | 0.674 | 0.742 | 2.32E-23 |
| MT-ATP8 | 4.66E-28 | 0.30341634 | 0.611 | 0.602 | 1.78E-23 |
| BNIP3L | 4.94E-09 | 0.30222817 | 0.42 | 0.451 | 0.00018891 |
| CELF4 | 7.59E-64 | 0.30168702 | 0.864 | 0.898 | 2.90E-59 |
| SLCO3A1 | 7.09E-16 | 0.29049888 | 0.656 | 0.721 | 2.71E-11 |
| PWRN4 | 1.10E-12 | 0.28968014 | 0.423 | 0.422 | 4.21E-08 |
| ZDHHC14 | 7.75E-15 | 0.28355243 | 0.612 | 0.664 | 2.96E-10 |
| CXXC4 | 2.20E-09 | 0.28324708 | 0.294 | 0.289 | 8.42E-05 |
| LINC00632 | 2.85E-73 | 0.28185029 | 0.902 | 0.936 | 1.09E-68 |
| SNX31 | 3.85E-54 | 0.28060554 | 0.115 | 0.05 | 1.47E-49 |
| KCND3 | 7.35E-13 | 0.27642021 | 0.686 | 0.751 | 2.81E-08 |
| DPYSL2 | 1.41E-20 | 0.27482041 | 0.698 | 0.778 | 5.40E-16 |
| PARD6G-AS1 | 1.04E-20 | 0.27465105 | 0.123 | 0.084 | 3.99E-16 |
| CMIP | 6.71E-14 | 0.27256601 | 0.671 | 0.765 | 2.56E-09 |
| PAPOLA | 7.33E-09 | 0.26779198 | 0.507 | 0.572 | 0.0002803 |
| HSPA12A | 7.74E-20 | 0.26747427 | 0.709 | 0.79 | 2.96E-15 |
| FOXO1 | 2.02E-07 | 0.26722396 | 0.201 | 0.181 | 0.00773064 |
| INSYN2B | 3.12E-09 | 0.2661538 | 0.213 | 0.189 | 0.00011912 |
| ANLN | 4.25E-21 | 0.26417526 | 0.126 | 0.086 | 1.62E-16 |
| MIR29B2CHG | 1.66E-10 | 0.26377008 | 0.587 | 0.672 | 6.33E-06 |
| SH3RF3 | 2.91E-12 | 0.26344052 | 0.65 | 0.699 | 1.11E-07 |
| RBM14 | 1.15E-11 | 0.26299289 | 0.261 | 0.242 | 4.41E-07 |
| SHANK2 | 1.22E-18 | 0.26062248 | 0.705 | 0.77 | 4.64E-14 |
| IGF1R | 6.41E-22 | 0.26049938 | 0.705 | 0.777 | 2.45E-17 |
| SPECC1L | 2.36E-11 | 0.25670942 | 0.505 | 0.536 | 9.04E-07 |
| PKNOX2 | 4.57E-12 | 0.25442771 | 0.672 | 0.733 | 1.75E-07 |
| DOCK5 | 1.21E-09 | 0.25421085 | 0.105 | 0.08 | 4.62E-05 |
| BMERB1 | 3.24E-20 | 0.25208785 | 0.68 | 0.699 | 1.24E-15 |
| ZNF536 | 1.86E-27 | 0.25167794 | 0.814 | 0.843 | 7.09E-23 |
| MT-CO1 | 1.58E-44 | 0.25014124 | 0.874 | 0.829 | 6.04E-40 |
| KCNMB2-AS1 | 1.71E-107 | -0.2509175 | 0.187 | 0.354 | 6.54E-103 |

| | | | | | |
|---|---|---|---|---|---|
| CNR1 | 2.20E-25 | -0.2519937 | 0.379 | 0.487 | 8.42E-21 |
| CMTM8 | 2.35E-111 | -0.2522894 | 0.116 | 0.26 | 9.00E-107 |
| GALNTL6 | 2.89E-33 | -0.2528344 | 0.714 | 0.789 | 1.10E-28 |
| PCDH8 | 1.70E-154 | -0.2532719 | 0.056 | 0.196 | 6.51E-150 |
| LUC7L2 | 6.45E-95 | -0.2547511 | 0.481 | 0.684 | 2.47E-90 |
| AL359232.1 | 3.37E-129 | -0.255421 | 0.156 | 0.331 | 1.29E-124 |
| AC016766.1 | 8.89E-45 | -0.2565348 | 0.095 | 0.171 | 3.40E-40 |
| DTNA | 2.20E-69 | -0.2577715 | 0.716 | 0.873 | 8.41E-65 |
| AC002460.2 | 1.25E-57 | -0.2592032 | 0.306 | 0.442 | 4.77E-53 |
| XKR4 | 2.55E-46 | -0.2593496 | 0.831 | 0.93 | 9.75E-42 |
| KLF12 | 2.28E-69 | -0.2597043 | 0.544 | 0.709 | 8.71E-65 |
| SORCS3 | 1.68E-71 | -0.2630907 | 0.38 | 0.547 | 6.42E-67 |
| KCNQ1OT1 | 2.45E-109 | -0.2653459 | 0.438 | 0.663 | 9.38E-105 |
| SUMF1 | 1.26E-125 | -0.2668174 | 0.383 | 0.617 | 4.81E-121 |
| ANKRD55 | 2.03E-151 | -0.2675923 | 0.133 | 0.317 | 7.77E-147 |
| OXR1 | 1.37E-73 | -0.2694345 | 0.827 | 0.926 | 5.25E-69 |
| TMEM59L | 2.43E-139 | -0.270269 | 0.313 | 0.546 | 9.29E-135 |
| UNC80 | 8.32E-108 | -0.2706429 | 0.662 | 0.854 | 3.18E-103 |
| IL1RAPL2 | 1.08E-42 | -0.2708152 | 0.186 | 0.284 | 4.13E-38 |
| LINC00461 | 1.50E-74 | -0.2726943 | 0.661 | 0.827 | 5.74E-70 |
| SLC44A5 | 6.68E-75 | -0.2727629 | 0.714 | 0.848 | 2.55E-70 |
| SEZ6L2 | 2.06E-137 | -0.2729806 | 0.353 | 0.589 | 7.89E-133 |
| GAD1 | 2.53E-106 | -0.2752443 | 0.622 | 0.816 | 9.67E-102 |
| CCPG1 | 1.77E-149 | -0.2756481 | 0.454 | 0.721 | 6.77E-145 |
| COL19A1 | 5.32E-69 | -0.2761769 | 0.19 | 0.321 | 2.03E-64 |
| UGGT2 | 1.41E-143 | -0.2773615 | 0.46 | 0.714 | 5.40E-139 |
| COL21A1 | 2.11E-68 | -0.279717 | 0.27 | 0.411 | 8.07E-64 |
| LINC01619 | 1.83E-116 | -0.2812896 | 0.112 | 0.255 | 7.00E-112 |
| ATP1B2 | 1.21E-149 | -0.2826481 | 0.289 | 0.524 | 4.62E-145 |
| ST6GALNAC5 | 1.30E-32 | -0.2837106 | 0.446 | 0.544 | 4.98E-28 |
| TERF2IP | 1.43E-144 | -0.2851345 | 0.395 | 0.648 | 5.45E-140 |
| INVS | 2.00E-150 | -0.287884 | 0.399 | 0.651 | 7.64E-146 |
| KCNT2 | 1.36E-26 | -0.2897813 | 0.496 | 0.569 | 5.22E-22 |
| TTC8 | 7.90E-195 | -0.2902792 | 0.199 | 0.437 | 3.02E-190 |
| SPARCL1 | 1.56E-48 | -0.2926284 | 0.402 | 0.541 | 5.96E-44 |
| SLC22A17 | 1.31E-125 | -0.292907 | 0.421 | 0.647 | 5.01E-121 |
| NELL1 | 1.12E-51 | -0.2932347 | 0.288 | 0.415 | 4.26E-47 |
| AC233296.1 | 2.55E-96 | -0.2937997 | 0.43 | 0.625 | 9.74E-92 |
| AL133346.1 | 1.11E-91 | -0.2954175 | 0.195 | 0.348 | 4.23E-87 |
| HGSNAT | 4.94E-189 | -0.2964484 | 0.29 | 0.56 | 1.89E-184 |
| BACH2 | 7.27E-127 | -0.2965666 | 0.476 | 0.711 | 2.78E-122 |
| ROBO1 | 3.70E-48 | -0.2983328 | 0.736 | 0.84 | 1.42E-43 |
| PDE4B | 9.15E-67 | -0.2989854 | 0.567 | 0.707 | 3.50E-62 |
| PCSK1N | 2.04E-109 | -0.2990668 | 0.363 | 0.572 | 7.79E-105 |
| ANO4 | 7.22E-90 | -0.2994499 | 0.732 | 0.858 | 2.76E-85 |

| | | | | | |
|---|---|---|---|---|---|
| GRIK1 | 3.92E-16 | -0.3010542 | 0.745 | 0.805 | 1.50E-11 |
| RFLNA | 3.06E-158 | -0.3013591 | 0.233 | 0.456 | 1.17E-153 |
| RELN | 1.94E-38 | -0.3026716 | 0.239 | 0.344 | 7.40E-34 |
| NETO2 | 1.48E-99 | -0.3037059 | 0.358 | 0.555 | 5.65E-95 |
| AC008591.1 | 7.08E-97 | -0.3067546 | 0.109 | 0.237 | 2.71E-92 |
| EYS | 1.35E-68 | -0.3070108 | 0.104 | 0.206 | 5.17E-64 |
| BDH1 | 1.32E-190 | -0.3079382 | 0.286 | 0.553 | 5.05E-186 |
| AC098829.1 | 6.21E-94 | -0.3095866 | 0.19 | 0.343 | 2.37E-89 |
| WDPCP | 1.09E-135 | -0.3120192 | 0.532 | 0.77 | 4.17E-131 |
| C8orf34 | 8.73E-64 | -0.3182643 | 0.499 | 0.648 | 3.34E-59 |
| CALY | 6.04E-145 | -0.3190077 | 0.399 | 0.643 | 2.31E-140 |
| GRM7 | 7.60E-68 | -0.3192833 | 0.705 | 0.839 | 2.91E-63 |
| MEF2C-AS2 | 4.56E-140 | -0.3206297 | 0.087 | 0.236 | 1.74E-135 |
| WSB1 | 1.17E-161 | -0.3216574 | 0.468 | 0.729 | 4.48E-157 |
| GMDS-DT | 5.30E-157 | -0.3256092 | 0.369 | 0.62 | 2.03E-152 |
| VGF | 2.92E-202 | -0.3262655 | 0.008 | 0.125 | 1.11E-197 |
| AC092683.1 | 9.44E-142 | -0.3341117 | 0.262 | 0.476 | 3.61E-137 |
| AC025159.1 | 6.13E-120 | -0.3452273 | 0.439 | 0.663 | 2.34E-115 |
| SHC3 | 6.66E-229 | -0.3585415 | 0.3 | 0.59 | 2.55E-224 |
| AL513166.1 | 3.52E-107 | -0.3588856 | 0.317 | 0.499 | 1.34E-102 |
| CDH9 | 1.46E-53 | -0.3602357 | 0.497 | 0.63 | 5.59E-49 |
| DLX6-AS1 | 1.51E-78 | -0.3616744 | 0.536 | 0.7 | 5.76E-74 |
| THSD7A | 1.47E-71 | -0.366171 | 0.629 | 0.757 | 5.62E-67 |
| LINC01184 | 4.90E-196 | -0.3685767 | 0.402 | 0.679 | 1.87E-191 |
| GOLGA8A | 1.59E-264 | -0.3697256 | 0.246 | 0.542 | 6.09E-260 |
| CCDC91 | 5.83E-159 | -0.3711552 | 0.554 | 0.787 | 2.23E-154 |
| COL25A1 | 9.71E-83 | -0.3715144 | 0.242 | 0.392 | 3.71E-78 |
| AC005753.2 | 3.92E-196 | -0.3857436 | 0.076 | 0.25 | 1.50E-191 |
| SST | 8.33E-122 | -0.3928162 | 0.03 | 0.129 | 3.18E-117 |
| LINC02398 | 3.62E-200 | -0.3930109 | 0.153 | 0.37 | 1.38E-195 |
| AC024901.1 | 4.57E-173 | -0.3985374 | 0.251 | 0.481 | 1.75E-168 |
| ATP1B1 | 7.14E-118 | -0.4021278 | 0.708 | 0.851 | 2.73E-113 |
| ICA1 | 3.10E-218 | -0.4067611 | 0.419 | 0.705 | 1.19E-213 |
| RASGRF2 | 3.76E-79 | -0.4079273 | 0.478 | 0.624 | 1.44E-74 |
| TAC3 | 3.31E-69 | -0.4091031 | 0.083 | 0.176 | 1.27E-64 |
| RFX3-AS1 | 1.10E-284 | -0.4181521 | 0.109 | 0.355 | 4.20E-280 |
| PAM | 1.11E-156 | -0.4203141 | 0.676 | 0.862 | 4.24E-152 |
| CIRBP | 1.96E-283 | -0.4204892 | 0.287 | 0.6 | 7.48E-279 |
| DPH6 | 1.91E-251 | -0.435716 | 0.267 | 0.558 | 7.29E-247 |
| ITFG1 | 1.08E-223 | -0.4395679 | 0.6 | 0.849 | 4.12E-219 |
| AL445433.2 | 4.30E-250 | -0.4596088 | 0.19 | 0.453 | 1.64E-245 |
| HTR2C | 7.48E-59 | -0.4598541 | 0.078 | 0.161 | 2.86E-54 |
| MIR4300HG | 8.12E-86 | -0.4746245 | 0.106 | 0.221 | 3.10E-81 |
| VIP | 7.92E-112 | -0.4789948 | 0.079 | 0.201 | 3.03E-107 |
| CNTNAP3B | 2.06E-148 | -0.4818708 | 0.171 | 0.349 | 7.88E-144 |

| | | | | | |
|---|---|---|---|---|---|
| DHFR | 6.96E-90 | -0.5102934 | 0.077 | 0.18 | 2.66E-85 |
| SCG2 | 1.96E-171 | -0.5203695 | 0.258 | 0.48 | 7.49E-167 |
| KCNAB1 | 6.77E-97 | -0.5301896 | 0.166 | 0.309 | 2.59E-92 |
| ROBO2 | 1.34E-119 | -0.5348742 | 0.891 | 0.946 | 5.13E-115 |
| GOLGA8B | 0 | -0.5589941 | 0.212 | 0.583 | 0 |
| CDH12 | 1.41E-87 | -0.561821 | 0.343 | 0.511 | 5.39E-83 |
| TARBP1 | 0 | -0.5786948 | 0.475 | 0.799 | 0 |
| SPRY4-AS1 | 0 | -0.5800834 | 0.065 | 0.325 | 0 |
| PDE10A | 0 | -0.7318049 | 0.567 | 0.845 | 0 |
| AC105402.3 | 4.32E-160 | -1.2270705 | 0.042 | 0.168 | 1.65E-155 |

**Supplementary Table 3. Pathway Analysis by Ingenuity**

**Excitatroy neurons**

| Ingenuity Canonical Pathways | -log(p-valu | Ratio | z-score | Molecules |
|---|---|---|---|---|
| Oxidative Phosphorylation | 4.83 | 0.055 | 2.449 | MT-ATP6,MT-CO3,MT-CYB,MT-ND2,MT-ND3,MT-ND4 |
| Synaptogenesis Signaling Pathway | 4.69 | 0.0288 | -1 | CDH12,CDH20,CDH9,EFNA5,EPHA5,GRIA4,NECTIN3,NRXN3,PLCG2 |
| Mitochondrial Dysfunction | 3.74 | 0.0351 | #NUM! | MT-ATP6,MT-CO3,MT-CYB,MT-ND2,MT-ND3,MT-ND4 |
| Melatonin Signaling | 3.39 | 0.0556 | 1 | PLCB1,PLCG2,RORA,RORB |
| tRNA Splicing | 2.9 | 0.0682 | #NUM! | PDE10A,PDE4D,PDE7B |
| Axonal Guidance Signaling | 2.57 | 0.0162 | #NUM! | ADAMTS3,EFNA5,EPHA5,LINGO1,PLCB1,PLCG2,ROBO1,ROBO2 |
| Sirtuin Signaling Pathway | 2.54 | 0.0206 | -2 | ATG10,MT-ATP6,MT-CYB,MT-ND2,MT-ND3,MT-ND4 |
| D-myo-inositol (1,4,5)-Trisphosphate Biosynthesis | 2.19 | 0.08 | #NUM! | PLCB1,PLCG2 |
| G Protein Signaling Mediated by Tubby | 2.01 | 0.0645 | #NUM! | PLCB1,PLCG2 |
| Cardiac Hypertrophy Signaling (Enhanced) | 1.98 | 0.0141 | -0.378 | HDAC9,PDE10A,PDE4D,PDE7B,PLCB1,PLCG2,RPS6KA5 |
| G-Protein Coupled Receptor Signaling | 1.97 | 0.0182 | #NUM! | PDE10A,PDE4D,PDE7B,PLCB1,RGS7 |
| UVA-Induced MAPK Signaling | 1.92 | 0.0306 | #NUM! | PLCB1,PLCG2,RPS6KA5 |
| Neuropathic Pain Signaling In Dorsal Horn Neurons | 1.89 | 0.0297 | #NUM! | GRIA4,PLCB1,PLCG2 |
| Protein Kinase A Signaling | 1.89 | 0.015 | -1.633 | PDE10A,PDE4D,PDE7B,PLCB1,PLCG2,PTPRT |
| Synaptic Long Term Depression | 1.88 | 0.0212 | -1 | GRIA4,GRID2,PLCB1,PLCG2 |
| Dermatan Sulfate Biosynthesis (Late Stages) | 1.68 | 0.0435 | #NUM! | HS6ST3,NDST3 |
| Ephrin A Signaling | 1.67 | 0.0426 | #NUM! | EFNA5,EPHA5 |
| Chondroitin Sulfate Biosynthesis (Late Stages) | 1.65 | 0.0417 | #NUM! | HS6ST3,NDST3 |
| Sperm Motility | 1.64 | 0.0179 | #NUM! | EPHA5,PDE4D,PLCB1,PLCG2 |
| dTMP De Novo Biosynthesis | 1.62 | 0.2 | #NUM! | DHFR |
| Endocannabinoid Neuronal Synapse Pathway | 1.62 | 0.0234 | #NUM! | GRIA4,PLCB1,PLCG2 |
| Synaptic Long Term Potentiation | 1.61 | 0.0233 | #NUM! | GRIA4,PLCB1,PLCG2 |
| cAMP-mediated signaling | 1.6 | 0.0175 | -1 | PDE10A,PDE4D,PDE7B,RGS7 |
| Cellular Effects of Sildenafil (Viagra) | 1.59 | 0.0229 | #NUM! | PDE4D,PLCB1,PLCG2 |
| Gα12/13 Signaling | 1.59 | 0.0229 | #NUM! | CDH12,CDH20,CDH9 |
| Chondroitin Sulfate Biosynthesis | 1.52 | 0.0357 | #NUM! | HS6ST3,NDST3 |
| Glutamate Receptor Signaling | 1.51 | 0.0351 | #NUM! | GRIA4,GRID2 |
| Cardiac β-adrenergic Signaling | 1.5 | 0.0211 | #NUM! | PDE10A,PDE4D,PDE7B |
| Opioid Signaling Pathway | 1.5 | 0.0162 | 1 | PLCB1,RGS21,RGS7,RPS6KA5 |
| Dermatan Sulfate Biosynthesis | 1.48 | 0.0339 | #NUM! | HS6ST3,NDST3 |
| Wnt/Ca+ pathway | 1.44 | 0.0323 | #NUM! | PLCB1,PLCG2 |
| Relaxin Signaling | 1.44 | 0.0199 | #NUM! | PDE10A,PDE4D,PDE7B |
| Gustation Pathway | 1.41 | 0.0194 | #NUM! | PDE10A,PDE4D,PDE7B |
| Phospholipases | 1.41 | 0.0308 | #NUM! | PLCB1,PLCG2 |
| Gαq Signaling | 1.39 | 0.0191 | #NUM! | PLCB1,PLCG2,RGS7 |
| Growth Hormone Signaling | 1.34 | 0.0282 | #NUM! | PLCG2,RPS6KA5 |
| Heparan Sulfate Biosynthesis (Late Stages) | 1.33 | 0.0278 | #NUM! | HS6ST3,NDST3 |
| GPCR-Mediated Integration of Enteroendocrine Signalir | 1.31 | 0.0274 | #NUM! | PLCB1,PLCG2 |
| Leptin Signaling in Obesity | 1.3 | 0.027 | #NUM! | PLCB1,PLCG2 |
| GDNF Family Ligand-Receptor Interactions | 1.28 | 0.0263 | #NUM! | DOK6,PLCG2 |
| Heparan Sulfate Biosynthesis | 1.25 | 0.0253 | #NUM! | HS6ST3,NDST3 |
| Chemokine Signaling | 1.24 | 0.025 | #NUM! | PLCB1,PLCG2 |
| Oleate Biosynthesis II (Animals) | 1.22 | 0.0769 | #NUM! | PTPRT |
| RhoGDI Signaling | 1.2 | 0.0159 | #NUM! | CDH12,CDH20,CDH9 |
| Xenobiotic Metabolism CAR Signaling Pathway | 1.2 | 0.0159 | #NUM! | HS6ST3,MGST2,NDST3 |
| Ephrin Receptor Signaling | 1.2 | 0.0159 | #NUM! | EFNA5,EPHA5,SORBS1 |
| Leukotriene Biosynthesis | 1.18 | 0.0714 | #NUM! | MGST2 |
| Xenobiotic Metabolism PXR Signaling Pathway | 1.18 | 0.0156 | #NUM! | HS6ST3,MGST2,NDST3 |
| Gap Junction Signaling | 1.15 | 0.0152 | #NUM! | GRIA4,PLCB1,PLCG2 |

| | | | | |
|---|---|---|---|---|
| Mismatch Repair in Eukaryotes | 1.13 | 0.0625 | #NUM! | MLH1 |
| Estrogen Receptor Signaling | 1.13 | 0.0122 | 1 | MT-ATP6,MT-CYB,PLCB1,PLCG2 |
| Melanocyte Development and Pigmentation Signaling | 1.12 | 0.0213 | #NUM! | PLCG2,RPS6KA5 |
| Amyotrophic Lateral Sclerosis Signaling | 1.1 | 0.0206 | #NUM! | GRIA4,GRID2 |
| Apelin Cardiomyocyte Signaling Pathway | 1.08 | 0.0202 | #NUM! | PLCB1,PLCG2 |
| Role of NFAT in Cardiac Hypertrophy | 1.07 | 0.014 | #NUM! | HDAC9,PLCB1,PLCG2 |
| LPS/IL-1 Mediated Inhibition of RXR Function | 1.02 | 0.0133 | #NUM! | HS6ST3,MGST2,NDST3 |
| Endoplasmic Reticulum Stress Pathway | 1.02 | 0.0476 | #NUM! | TAOK3 |
| Antioxidant Action of Vitamin C | 1.01 | 0.0183 | #NUM! | PLCB1,PLCG2 |
| GPCR-Mediated Nutrient Sensing in Enteroendocrine C | 0.991 | 0.0179 | #NUM! | PLCB1,PLCG2 |
| NGF Signaling | 0.979 | 0.0175 | #NUM! | PLCG2,RPS6KA5 |
| Role of Tissue Factor in Cancer | 0.967 | 0.0172 | #NUM! | PLCB1,RPS6KA5 |
| Huntington's Disease Signaling | 0.963 | 0.0126 | #NUM! | HDAC9,PLCB1,RPH3A |
| Sphingosine-1-phosphate Signaling | 0.959 | 0.0171 | #NUM! | PLCB1,PLCG2 |
| Glutathione Redox Reactions I | 0.959 | 0.0417 | #NUM! | MGST2 |
| Insulin Secretion Signaling Pathway | 0.943 | 0.0123 | #NUM! | PLCB1,PLCG2,RPS6KA5 |
| GP6 Signaling Pathway | 0.914 | 0.016 | #NUM! | COL24A1,PLCG2 |
| Signaling by Rho Family GTPases | 0.91 | 0.0119 | #NUM! | CDH12,CDH20,CDH9 |
| 14-3-3-mediated Signaling | 0.903 | 0.0157 | #NUM! | PLCB1,PLCG2 |
| P2Y Purigenic Receptor Signaling Pathway | 0.903 | 0.0157 | #NUM! | PLCB1,PLCG2 |
| p70S6K Signaling | 0.893 | 0.0155 | #NUM! | PLCB1,PLCG2 |
| Phagosome Formation | 0.87 | 0.015 | #NUM! | PLCB1,PLCG2 |
| Phospholipase C Signaling | 0.863 | 0.0113 | #NUM! | HDAC9,PLCB1,PLCG2 |
| PI3K Signaling in B Lymphocytes | 0.845 | 0.0145 | #NUM! | PLCB1,PLCG2 |
| Glutathione-mediated Detoxification | 0.845 | 0.0312 | #NUM! | MGST2 |
| Semaphorin Neuronal Repulsive Signaling Pathway | 0.839 | 0.0144 | #NUM! | PDE4D,PLCG2 |
| Hereditary Breast Cancer Signaling | 0.833 | 0.0143 | #NUM! | HDAC9,MLH1 |
| Aryl Hydrocarbon Receptor Signaling | 0.818 | 0.014 | #NUM! | DHFR,MGST2 |
| Xenobiotic Metabolism Signaling | 0.793 | 0.0105 | #NUM! | HS6ST3,MGST2,NDST3 |
| Factors Promoting Cardiogenesis in Vertebrates | 0.788 | 0.0133 | #NUM! | PLCB1,PLCG2 |
| Epithelial Adherens Junction Signaling | 0.777 | 0.0132 | #NUM! | NECTIN3,SORBS1 |
| D-myo-inositol-5-phosphate Metabolism | 0.757 | 0.0127 | #NUM! | PLCB1,PLCG2 |
| Aldosterone Signaling in Epithelial Cells | 0.752 | 0.0127 | #NUM! | PLCB1,PLCG2 |
| Glucocorticoid Receptor Signaling | 0.735 | 0.00866 | #NUM! | FKBP5,MT-ATP6,MT-CYB,RPS6KA5 |
| Dopamine-DARPP32 Feedback in cAMP Signaling | 0.73 | 0.0123 | #NUM! | PLCB1,PLCG2 |
| Role of IL-17F in Allergic Inflammatory Airway Diseases | 0.726 | 0.0233 | #NUM! | RPS6KA5 |
| Glioblastoma Multiforme Signaling | 0.724 | 0.0121 | #NUM! | PLCB1,PLCG2 |
| IL-23 Signaling Pathway | 0.717 | 0.0227 | #NUM! | RORA |
| Role of Oct4 in Mammalian Embryonic Stem Cell Pluripo | 0.699 | 0.0217 | #NUM! | SPP1 |
| Germ Cell-Sertoli Cell Junction Signaling | 0.699 | 0.0117 | #NUM! | NECTIN3,SORBS1 |
| PFKFB4 Signaling Pathway | 0.699 | 0.0217 | #NUM! | RPS6KA5 |
| Triacylglycerol Degradation | 0.668 | 0.02 | #NUM! | NDST3 |
| Role of NFAT in Regulation of the Immune Response | 0.664 | 0.011 | #NUM! | PLCB1,PLCG2 |
| UVB-Induced MAPK Signaling | 0.654 | 0.0192 | #NUM! | RPS6KA5 |
| Dendritic Cell Maturation | 0.652 | 0.0109 | #NUM! | PLCB1,PLCG2 |
| Endothelin-1 Signaling | 0.638 | 0.0106 | #NUM! | PLCB1,PLCG2 |
| NRF2-mediated Oxidative Stress Response | 0.636 | 0.0106 | #NUM! | FKBP5,MGST2 |
| PPARα/RXRα Activation | 0.629 | 0.0105 | #NUM! | PLCB1,PLCG2 |
| Sertoli Cell-Sertoli Cell Junction Signaling | 0.62 | 0.0103 | #NUM! | NECTIN3,SORBS1 |
| CNTF Signaling | 0.618 | 0.0175 | #NUM! | RPS6KA5 |
| Role of CHK Proteins in Cell Cycle Checkpoint Control | 0.618 | 0.0175 | #NUM! | TLK2 |
| Adrenomedullin signaling pathway | 0.609 | 0.0102 | #NUM! | PLCB1,PLCG2 |

| | | | | |
|---|---|---|---|---|
| Superpathway of Inositol Phosphate Compounds | 0.604 | 0.0101 | #NUM! | PLCB1,PLCG2 |
| Endometrial Cancer Signaling | 0.599 | 0.0167 | #NUM! | MLH1 |
| ERK/MAPK Signaling | 0.593 | 0.0099 | #NUM! | PLCG2,RPS6KA5 |
| Autophagy | 0.592 | 0.0164 | #NUM! | ATG10 |
| Calcium Signaling | 0.582 | 0.00971 | #NUM! | GRIA4,HDAC9 |
| Thrombopoietin Signaling | 0.58 | 0.0159 | #NUM! | PLCG2 |
| Thrombin Signaling | 0.577 | 0.00962 | #NUM! | PLCB1,PLCG2 |
| Role of PI3K/AKT Signaling in the Pathogenesis of Influ | 0.575 | 0.0156 | #NUM! | MLH1 |
| ErbB4 Signaling | 0.558 | 0.0149 | #NUM! | PLCG2 |
| Cell Cycle: G1/S Checkpoint Regulation | 0.558 | 0.0149 | #NUM! | HDAC9 |
| Nur77 Signaling in T Lymphocytes | 0.553 | 0.0147 | #NUM! | HDAC9 |
| ERK5 Signaling | 0.532 | 0.0139 | #NUM! | RPS6KA5 |
| TREM1 Signaling | 0.517 | 0.0133 | #NUM! | PLCG2 |
| FcγRIIB Signaling in B Lymphocytes | 0.517 | 0.0133 | #NUM! | PLCG2 |
| Macropinocytosis Signaling | 0.511 | 0.0132 | #NUM! | PLCG2 |
| VDR/RXR Activation | 0.503 | 0.0128 | #NUM! | SPP1 |
| BEX2 Signaling Pathway | 0.498 | 0.0127 | #NUM! | SPP1 |
| Role of BRCA1 in DNA Damage Response | 0.493 | 0.0125 | #NUM! | MLH1 |
| FLT3 Signaling in Hematopoietic Progenitor Cells | 0.493 | 0.0125 | #NUM! | RPS6KA5 |
| Cardiac Hypertrophy Signaling | 0.492 | 0.00833 | #NUM! | PLCB1,PLCG2 |
| Prolactin Signaling | 0.489 | 0.0123 | #NUM! | PLCG2 |
| CREB Signaling in Neurons | 0.489 | 0.00671 | -1 | GRIA4,GRID2,PLCB1,PLCG2 |
| Cyclins and Cell Cycle Regulation | 0.489 | 0.0123 | #NUM! | HDAC9 |
| Apelin Adipocyte Signaling Pathway | 0.484 | 0.0122 | #NUM! | MGST2 |
| VEGF Family Ligand-Receptor Interactions | 0.476 | 0.0119 | #NUM! | PLCG2 |
| FGF Signaling | 0.476 | 0.0119 | #NUM! | RPS6KA5 |
| HIPPO signaling | 0.472 | 0.0118 | #NUM! | FRMD6 |
| Xenobiotic Metabolism AHR Signaling Pathway | 0.472 | 0.0118 | #NUM! | MGST2 |
| PDGF Signaling | 0.467 | 0.0116 | #NUM! | PLCG2 |
| Altered T Cell and B Cell Signaling in Rheumatoid Arthri | 0.452 | 0.0111 | #NUM! | SPP1 |
| Regulation of IL-2 Expression in Activated and Anergic | 0.452 | 0.0111 | #NUM! | PLCG2 |
| Th17 Activation Pathway | 0.449 | 0.011 | #NUM! | RORA |
| Fcγ Receptor-mediated Phagocytosis in Macrophages a | 0.437 | 0.0106 | #NUM! | DGKB |
| CCR5 Signaling in Macrophages | 0.437 | 0.0106 | #NUM! | PLCG2 |
| ErbB Signaling | 0.437 | 0.0106 | #NUM! | PLCG2 |
| α-Adrenergic Signaling | 0.433 | 0.0105 | #NUM! | PLCG2 |
| ATM Signaling | 0.426 | 0.0103 | #NUM! | TLK2 |
| Bladder Cancer Signaling | 0.426 | 0.0103 | #NUM! | RPS6KA5 |
| p53 Signaling | 0.423 | 0.0102 | #NUM! | HDAC9 |
| VEGF Signaling | 0.419 | 0.0101 | #NUM! | PLCG2 |
| Apoptosis Signaling | 0.416 | 0.01 | #NUM! | PLCG2 |
| Virus Entry via Endocytic Pathways | 0.409 | 0.0098 | #NUM! | PLCG2 |
| Chronic Myeloid Leukemia Signaling | 0.406 | 0.00971 | #NUM! | HDAC9 |
| FAK Signaling | 0.402 | 0.00962 | #NUM! | PLCG2 |
| Neuregulin Signaling | 0.399 | 0.00952 | #NUM! | PLCG2 |
| Telomerase Signaling | 0.394 | 0.00935 | #NUM! | HDAC9 |
| Glioma Signaling | 0.384 | 0.00909 | #NUM! | PLCG2 |
| Apelin Endothelial Signaling Pathway | 0.37 | 0.0087 | #NUM! | PLCB1 |
| fMLP Signaling in Neutrophils | 0.367 | 0.00862 | #NUM! | PLCB1 |
| Fc Epsilon RI Signaling | 0.364 | 0.00855 | #NUM! | PLCG2 |
| Renin-Angiotensin Signaling | 0.362 | 0.00847 | #NUM! | PLCG2 |
| p38 MAPK Signaling | 0.362 | 0.00847 | #NUM! | RPS6KA5 |

| | | | | |
|---|---|---|---|---|
| Cholecystokinin/Gastrin-mediated Signaling | 0.359 | 0.0084 | #NUM! | PLCB1 |
| HGF Signaling | 0.356 | 0.00833 | #NUM! | PLCG2 |
| IL-15 Production | 0.353 | 0.00826 | #NUM! | EPHA5 |
| Reelin Signaling in Neurons | 0.351 | 0.0082 | #NUM! | NECTIN3 |
| G Beta Gamma Signaling | 0.351 | 0.0082 | #NUM! | PLCG2 |
| Role of Macrophages, Fibroblasts and Endothelial Cells | 0.349 | 0.00637 | #NUM! | PLCB1,PLCG2 |
| CCR3 Signaling in Eosinophils | 0.345 | 0.00806 | #NUM! | PLCB1 |
| Gαi Signaling | 0.343 | 0.008 | #NUM! | RGS7 |
| White Adipose Tissue Browning Pathway | 0.333 | 0.00775 | #NUM! | VGF |
| Adipogenesis pathway | 0.321 | 0.00746 | #NUM! | HDAC9 |
| Ovarian Cancer Signaling | 0.31 | 0.00719 | #NUM! | MLH1 |
| Xenobiotic Metabolism General Signaling Pathway | 0.301 | 0.00699 | #NUM! | MGST2 |
| Corticotropin Releasing Hormone Signaling | 0.297 | 0.0069 | #NUM! | PLCG2 |
| Role of Pattern Recognition Receptors in Recognition o | 0.279 | 0.00649 | #NUM! | PLCG2 |
| PKCθ Signaling in T Lymphocytes | 0.277 | 0.00645 | #NUM! | PLCG2 |
| eNOS Signaling | 0.27 | 0.00629 | #NUM! | PLCG2 |
| HOTAIR Regulatory Pathway | 0.268 | 0.00625 | #NUM! | SPP1 |
| Hepatic Fibrosis Signaling Pathway | 0.263 | 0.00529 | #NUM! | PLCG2,SPP1 |
| CXCR4 Signaling | 0.256 | 0.00599 | #NUM! | PLCB1 |
| Tight Junction Signaling | 0.254 | 0.00595 | #NUM! | NECTIN3 |
| GNRH Signaling | 0.246 | 0.00578 | #NUM! | PLCB1 |
| Tec Kinase Signaling | 0.246 | 0.00578 | #NUM! | PLCG2 |
| Wnt/β-catenin Signaling | 0.246 | 0.00578 | #NUM! | CDH12 |
| T Cell Exhaustion Signaling Pathway | 0.243 | 0.00571 | #NUM! | PLCG2 |
| Tumor Microenvironment Pathway | 0.241 | 0.00568 | #NUM! | SPP1 |
| Molecular Mechanisms of Cancer | 0.239 | 0.005 | #NUM! | PLCB1,RASGRF2 |
| NF-κB Signaling | 0.237 | 0.00559 | #NUM! | PLCG2 |
| Hepatic Cholestasis | 0.226 | 0.00538 | #NUM! | SLCO3A1 |
| Hepatic Fibrosis / Hepatic Stellate Cell Activation | 0.226 | 0.00538 | #NUM! | COL24A1 |
| B Cell Receptor Signaling | 0.226 | 0.00538 | #NUM! | PLCG2 |
| Regulation Of The Epithelial Mesenchymal Transition By | 0.223 | 0.00532 | #NUM! | CDH12 |
| Production of Nitric Oxide and Reactive Oxygen Species | 0.222 | 0.00529 | #NUM! | PLCG2 |
| ILK Signaling | 0.22 | 0.00526 | #NUM! | RPS6KA5 |
| Regulation of the Epithelial-Mesenchymal Transition Pat | 0.218 | 0.00521 | #NUM! | CDH12 |
| Leukocyte Extravasation Signaling | 0.216 | 0.00518 | #NUM! | PLCG2 |
| RAR Activation | 0.215 | 0.00515 | #NUM! | ZBTB16 |
| HER-2 Signaling in Breast Cancer | 0.215 | 0.00515 | #NUM! | MT-CO3 |
| Natural Killer Cell Signaling | 0.21 | 0.00508 | #NUM! | PLCG2 |
| HIF1α Signaling | 0.201 | 0.00488 | #NUM! | PLCG2 |
| Colorectal Cancer Metastasis Signaling | 0 | 0.00395 | #NUM! | MLH1 |
| mTOR Signaling | 0 | 0.00476 | #NUM! | RPS6KA5 |
| Systemic Lupus Erythematosus Signaling | 0 | 0.00437 | #NUM! | PLCG2 |
| EIF2 Signaling | 0 | 0.00446 | #NUM! | RPL15 |
| Role of Osteoblasts, Osteoclasts and Chondrocytes in F | 0 | 0.00459 | #NUM! | SPP1 |
| Breast Cancer Regulation by Stathmin1 | 0 | 0.00169 | #NUM! | PLCB1 |
| Integrin Signaling | 0 | 0.00469 | #NUM! | PLCG2 |
| Osteoarthritis Pathway | 0 | 0.00455 | #NUM! | SPP1 |
| Neuroinflammation Signaling Pathway | 0 | 0.00333 | #NUM! | PLCG2 |
| Systemic Lupus Erythematosus In B Cell Signaling Path | 0 | 0.00364 | #NUM! | PLCG2 |
| Senescence Pathway | 0 | 0.00364 | #NUM! | RPS6KA5 |

**Inhibibory Neurons**

| Ingenuity Canonical Pathways | -log(p-valu | Ratio | z-score | Molecules |
|---|---|---|---|---|
| Synaptogenesis Signaling Pathway | 4.03 | 0.0192 | -0.816 | CDH12,GRM7,PLCG2,RELN,SHC3,SYN3 |
| Glutamate Receptor Signaling | 3.47 | 0.0526 | #NUM! | GRM7,SLC1A2,SLC1A3 |
| G-Protein Coupled Receptor Signaling | 2.4 | 0.0146 | #NUM! | GRM7,HTR2C,PDE10A,PDE4B |
| tRNA Splicing | 2.31 | 0.0455 | #NUM! | PDE10A,PDE4B |
| Neuroinflammation Signaling Pathway | 2.26 | 0.0133 | #NUM! | HMGB1,PLCG2,SLC1A2,SLC1A3 |
| Spermine and Spermidine Degradation I | 2.03 | 0.25 | #NUM! | SAT1 |
| Hepatic Fibrosis / Hepatic Stellate Cell Activation | 2.01 | 0.0161 | #NUM! | COL21A1,IGFBP5,IL1RAPL2 |
| Synaptic Long Term Depression | 1.99 | 0.0159 | #NUM! | GRM7,PLCG2,RYR3 |
| GPCR-Mediated Integration of Enteroendocrine Signaling E | 1.89 | 0.0274 | #NUM! | PLCG2,VIP |
| VDR/RXR Activation | 1.83 | 0.0256 | #NUM! | IGFBP5,SPP1 |
| Protein Kinase A Signaling | 1.83 | 0.01 | -1 | PDE10A,PDE4B,PLCG2,RYR3 |
| Osteoarthritis Pathway | 1.82 | 0.0136 | #NUM! | HMGB1,IL1RAPL2,SPP1 |
| cAMP-mediated signaling | 1.77 | 0.0131 | #NUM! | GRM7,PDE10A,PDE4B |
| Neuropathic Pain Signaling In Dorsal Horn Neurons | 1.62 | 0.0198 | #NUM! | GRM7,PLCG2 |
| Oxidative Phosphorylation | 1.56 | 0.0183 | #NUM! | MT-CYB,MT-ND3 |
| Cardiac Hypertrophy Signaling (Enhanced) | 1.52 | 0.00805 | -1 | PDE10A,PDE4B,PLCG2,RYR3 |
| Renin-Angiotensin Signaling | 1.5 | 0.0169 | #NUM! | PLCG2,SHC3 |
| Telomere Extension by Telomerase | 1.46 | 0.0667 | #NUM! | TERF2IP |
| GP6 Signaling Pathway | 1.45 | 0.016 | #NUM! | COL21A1,PLCG2 |
| Synaptic Long Term Potentiation | 1.43 | 0.0155 | #NUM! | GRM7,PLCG2 |
| Cellular Effects of Sildenafil (Viagra) | 1.41 | 0.0153 | #NUM! | PDE4B,PLCG2 |
| Estrogen Receptor Signaling | 1.37 | 0.00915 | #NUM! | MT-CYB,PLCG2,SHC3 |
| Semaphorin Neuronal Repulsive Signaling Pathway | 1.37 | 0.0144 | #NUM! | PDE4B,PLCG2 |
| Cardiac β-adrenergic Signaling | 1.35 | 0.0141 | #NUM! | PDE10A,PDE4B |
| Putrescine Degradation III | 1.31 | 0.0476 | #NUM! | SAT1 |
| Relaxin Signaling | 1.3 | 0.0132 | #NUM! | PDE10A,PDE4B |
| Gustation Pathway | 1.28 | 0.0129 | #NUM! | PDE10A,PDE4B |
| Polyamine Regulation in Colon Cancer | 1.27 | 0.0435 | #NUM! | SAT1 |
| Gαq Signaling | 1.27 | 0.0127 | #NUM! | HTR2C,PLCG2 |
| Aldosterone Signaling in Epithelial Cells | 1.27 | 0.0127 | #NUM! | CRYAB,PLCG2 |
| D-myo-inositol (1,4,5)-Trisphosphate Biosynthesis | 1.24 | 0.04 | #NUM! | PLCG2 |
| Hepatic Fibrosis Signaling Pathway | 1.22 | 0.00794 | #NUM! | IL1RAPL2,PLCG2,SPP1 |
| Mitochondrial Dysfunction | 1.21 | 0.0117 | #NUM! | MT-CYB,MT-ND3 |
| G Protein Signaling Mediated by Tubby | 1.15 | 0.0323 | #NUM! | PLCG2 |
| Endothelin-1 Signaling | 1.14 | 0.0106 | #NUM! | PLCG2,SHC3 |
| PPARα/RXRα Activation | 1.12 | 0.0105 | #NUM! | IL1RAPL2,PLCG2 |
| Circadian Rhythm Signaling | 1.12 | 0.0303 | #NUM! | VIP |
| Adrenomedullin signaling pathway | 1.1 | 0.0102 | #NUM! | PLCG2,SHC3 |
| Gap Junction Signaling | 1.1 | 0.0101 | #NUM! | HTR2C,PLCG2 |
| HIF1α Signaling | 1.07 | 0.00976 | #NUM! | PLCG2,SAT1 |
| nNOS Signaling in Skeletal Muscle Cells | 1.03 | 0.0244 | #NUM! | RYR3 |
| Role of Osteoblasts, Osteoclasts and Chondrocytes in Rhe | 1.03 | 0.00917 | #NUM! | IL1RAPL2,SPP1 |

| | | | | |
|---|---|---|---|---|
| Serotonin Receptor Signaling | 1.01 | 0.0233 | #NUM! | HTR2C |
| Sperm Motility | 1.01 | 0.00897 | #NUM! | PDE4B,PLCG2 |
| Role of Oct4 in Mammalian Embryonic Stem Cell Pluripoten | 0.987 | 0.0217 | #NUM! | SPP1 |
| Axonal Guidance Signaling | 0.951 | 0.00607 | #NUM! | NTNG1,PLCG2,ROBO2 |
| Insulin Secretion Signaling Pathway | 0.943 | 0.0082 | #NUM! | PLCG2,RYR3 |
| Wnt/Ca+ pathway | 0.863 | 0.0161 | #NUM! | PLCG2 |
| Systemic Lupus Erythematosus In B Cell Signaling Pathway | 0.86 | 0.00727 | #NUM! | PLCG2,SHC3 |
| Thrombopoietin Signaling | 0.857 | 0.0159 | #NUM! | PLCG2 |
| Netrin Signaling | 0.845 | 0.0154 | #NUM! | RYR3 |
| Phospholipases | 0.845 | 0.0154 | #NUM! | PLCG2 |
| ErbB4 Signaling | 0.833 | 0.0149 | #NUM! | PLCG2 |
| Sirtuin Signaling Pathway | 0.821 | 0.00687 | #NUM! | MT-CYB,MT-ND3 |
| IL-10 Signaling | 0.815 | 0.0143 | #NUM! | IL1RAPL2 |
| Growth Hormone Signaling | 0.81 | 0.0141 | #NUM! | PLCG2 |
| Melatonin Signaling | 0.804 | 0.0139 | #NUM! | PLCG2 |
| Leptin Signaling in Obesity | 0.793 | 0.0135 | #NUM! | PLCG2 |
| TREM1 Signaling | 0.788 | 0.0133 | #NUM! | PLCG2 |
| FcγRIIB Signaling in B Lymphocytes | 0.788 | 0.0133 | #NUM! | PLCG2 |
| GDNF Family Ligand-Receptor Interactions | 0.783 | 0.0132 | #NUM! | PLCG2 |
| Macropinocytosis Signaling | 0.783 | 0.0132 | #NUM! | PLCG2 |
| CREB Signaling in Neurons | 0.777 | 0.00503 | #NUM! | GRM7,HTR2C,PLCG2 |
| Role of Macrophages, Fibroblasts and Endothelial Cells in I | 0.77 | 0.00637 | #NUM! | IL1RAPL2,PLCG2 |
| BEX2 Signaling Pathway | 0.767 | 0.0127 | #NUM! | SPP1 |
| Chemokine Signaling | 0.762 | 0.0125 | #NUM! | PLCG2 |
| Prolactin Signaling | 0.757 | 0.0123 | #NUM! | PLCG2 |
| VEGF Family Ligand-Receptor Interactions | 0.742 | 0.0119 | #NUM! | PLCG2 |
| PDGF Signaling | 0.733 | 0.0116 | #NUM! | PLCG2 |
| Altered T Cell and B Cell Signaling in Rheumatoid Arthritis | 0.717 | 0.0111 | #NUM! | SPP1 |
| Regulation of IL-2 Expression in Activated and Anergic T Ly | 0.717 | 0.0111 | #NUM! | PLCG2 |
| CCR5 Signaling in Macrophages | 0.699 | 0.0106 | #NUM! | PLCG2 |
| Melanocyte Development and Pigmentation Signaling | 0.699 | 0.0106 | #NUM! | PLCG2 |
| ErbB Signaling | 0.699 | 0.0106 | #NUM! | PLCG2 |
| α-Adrenergic Signaling | 0.695 | 0.0105 | #NUM! | PLCG2 |
| Amyotrophic Lateral Sclerosis Signaling | 0.686 | 0.0103 | #NUM! | SLC1A2 |
| UVA-Induced MAPK Signaling | 0.682 | 0.0102 | #NUM! | PLCG2 |
| VEGF Signaling | 0.68 | 0.0101 | #NUM! | PLCG2 |
| Apelin Cardiomyocyte Signaling Pathway | 0.68 | 0.0101 | #NUM! | PLCG2 |
| Apoptosis Signaling | 0.676 | 0.01 | #NUM! | PLCG2 |
| Virus Entry via Endocytic Pathways | 0.668 | 0.0098 | #NUM! | PLCG2 |
| FAK Signaling | 0.66 | 0.00962 | #NUM! | PLCG2 |
| IGF-1 Signaling | 0.66 | 0.00962 | #NUM! | IGFBP5 |
| Neuregulin Signaling | 0.656 | 0.00952 | #NUM! | PLCG2 |
| PPAR Signaling | 0.656 | 0.00952 | #NUM! | IL1RAPL2 |
| Telomerase Signaling | 0.65 | 0.00935 | #NUM! | TERF2IP |

| | | | | |
|---|---|---|---|---|
| Gαs Signaling | 0.65 | 0.00935 | #NUM! | RYR3 |
| Antioxidant Action of Vitamin C | 0.642 | 0.00917 | #NUM! | PLCG2 |
| Glioma Signaling | 0.638 | 0.00909 | #NUM! | PLCG2 |
| GPCR-Mediated Nutrient Sensing in Enteroendocrine Cells | 0.631 | 0.00893 | #NUM! | PLCG2 |
| NGF Signaling | 0.625 | 0.00877 | #NUM! | PLCG2 |
| Fc Epsilon RI Signaling | 0.614 | 0.00855 | #NUM! | PLCG2 |
| Sphingosine-1-phosphate Signaling | 0.614 | 0.00855 | #NUM! | PLCG2 |
| Role of PKR in Interferon Induction and Antiviral Response | 0.613 | 0.00847 | #NUM! | HMGB1 |
| p38 MAPK Signaling | 0.613 | 0.00847 | #NUM! | IL1RAPL2 |
| HGF Signaling | 0.606 | 0.00833 | #NUM! | PLCG2 |
| LXR/RXR Activation | 0.602 | 0.00826 | #NUM! | IL1RAPL2 |
| Reelin Signaling in Neurons | 0.599 | 0.0082 | #NUM! | RELN |
| G Beta Gamma Signaling | 0.599 | 0.0082 | #NUM! | PLCG2 |
| Gαi Signaling | 0.59 | 0.008 | #NUM! | GRM7 |
| IL-6 Signaling | 0.587 | 0.00794 | #NUM! | IL1RAPL2 |
| Ferroptosis Signaling Pathway | 0.587 | 0.00794 | #NUM! | SAT1 |
| 14-3-3-mediated Signaling | 0.585 | 0.00787 | #NUM! | PLCG2 |
| P2Y Purigenic Receptor Signaling Pathway | 0.585 | 0.00787 | #NUM! | PLCG2 |
| Endocannabinoid Neuronal Synapse Pathway | 0.582 | 0.00781 | #NUM! | PLCG2 |
| p70S6K Signaling | 0.578 | 0.00775 | #NUM! | PLCG2 |
| White Adipose Tissue Browning Pathway | 0.578 | 0.00775 | #NUM! | VGF |
| Gα12/13 Signaling | 0.573 | 0.00763 | #NUM! | CDH12 |
| Phagosome Formation | 0.567 | 0.00752 | #NUM! | PLCG2 |
| PI3K Signaling in B Lymphocytes | 0.553 | 0.00725 | #NUM! | PLCG2 |
| Corticotropin Releasing Hormone Signaling | 0.535 | 0.0069 | #NUM! | PLCG2 |
| Glucocorticoid Receptor Signaling | 0.524 | 0.00433 | #NUM! | HMGB1,MT-CYB |
| Factors Promoting Cardiogenesis in Vertebrates | 0.523 | 0.00667 | #NUM! | PLCG2 |
| Role of Pattern Recognition Receptors in Recognition of Ba | 0.513 | 0.00649 | #NUM! | PLCG2 |
| PKCθ Signaling in T Lymphocytes | 0.511 | 0.00645 | #NUM! | PLCG2 |
| D-myo-inositol-5-phosphate Metabolism | 0.506 | 0.00637 | #NUM! | PLCG2 |
| eNOS Signaling | 0.502 | 0.00629 | #NUM! | PLCG2 |
| HOTAIR Regulatory Pathway | 0.5 | 0.00625 | #NUM! | SPP1 |
| Dopamine-DARPP32 Feedback in cAMP Signaling | 0.493 | 0.00613 | #NUM! | PLCG2 |
| HMGB1 Signaling | 0.488 | 0.00606 | #NUM! | HMGB1 |
| Glioblastoma Multiforme Signaling | 0.488 | 0.00606 | #NUM! | PLCG2 |
| Granulocyte Adhesion and Diapedesis | 0.472 | 0.00578 | #NUM! | IL1RAPL2 |
| Tec Kinase Signaling | 0.472 | 0.00578 | #NUM! | PLCG2 |
| Wnt/β-catenin Signaling | 0.472 | 0.00578 | #NUM! | CDH12 |
| T Cell Exhaustion Signaling Pathway | 0.469 | 0.00571 | #NUM! | PLCG2 |
| Tumor Microenvironment Pathway | 0.466 | 0.00568 | #NUM! | SPP1 |
| NF-κB Signaling | 0.46 | 0.00559 | #NUM! | PLCG2 |
| Role of NFAT in Regulation of the Immune Response | 0.456 | 0.00552 | #NUM! | PLCG2 |
| Dendritic Cell Maturation | 0.45 | 0.00543 | #NUM! | PLCG2 |
| Hepatic Cholestasis | 0.446 | 0.00538 | #NUM! | IL1RAPL2 |

| | | | | |
|---|---|---|---|---|
| B Cell Receptor Signaling | 0.446 | 0.00538 | #NUM! | PLCG2 |
| Regulation Of The Epithelial Mesenchymal Transition By Gr | 0.442 | 0.00532 | #NUM! | CDH12 |
| Production of Nitric Oxide and Reactive Oxygen Species in | 0.441 | 0.00529 | #NUM! | PLCG2 |
| RhoGDI Signaling | 0.441 | 0.00529 | #NUM! | CDH12 |
| Regulation of the Epithelial-Mesenchymal Transition Pathwa | 0.435 | 0.00521 | #NUM! | CDH12 |
| Leukocyte Extravasation Signaling | 0.434 | 0.00518 | #NUM! | PLCG2 |
| Natural Killer Cell Signaling | 0.427 | 0.00508 | #NUM! | PLCG2 |
| Superpathway of Inositol Phosphate Compounds | 0.424 | 0.00503 | #NUM! | PLCG2 |
| ERK/MAPK Signaling | 0.418 | 0.00495 | #NUM! | PLCG2 |
| Calcium Signaling | 0.411 | 0.00485 | #NUM! | RYR3 |
| Thrombin Signaling | 0.409 | 0.00481 | #NUM! | PLCG2 |
| Integrin Signaling | 0.4 | 0.00469 | #NUM! | PLCG2 |
| Role of NFAT in Cardiac Hypertrophy | 0.399 | 0.00467 | #NUM! | PLCG2 |
| Breast Cancer Regulation by Stathmin1 | 0.388 | 0.00339 | #NUM! | GRM7,HTR2C |
| LPS/IL-1 Mediated Inhibition of RXR Function | 0.382 | 0.00444 | #NUM! | IL1RAPL2 |
| Systemic Lupus Erythematosus Signaling | 0.377 | 0.00437 | #NUM! | PLCG2 |
| Cardiac Hypertrophy Signaling | 0.362 | 0.00417 | #NUM! | PLCG2 |
| Opioid Signaling Pathway | 0.352 | 0.00405 | #NUM! | RYR3 |
| Signaling by Rho Family GTPases | 0.344 | 0.00395 | #NUM! | CDH12 |
| Phospholipase C Signaling | 0.328 | 0.00376 | #NUM! | PLCG2 |
| Protein Ubiquitination Pathway | 0.321 | 0.00366 | #NUM! | CRYAB |
| Molecular Mechanisms of Cancer | 0.21 | 0.0025 | #NUM! | RASGRF2 |

**Astrocytes**

| Ingenuity Canonical Pathways | -log(p-valu | Ratio | z-score | Molecules |
|---|---|---|---|---|
| Amyotrophic Lateral Sclerosis Signaling | 5.79 | 0.103 | -0.333 | CAPN2,GLUL,GRIA2,GRID2,GRIN2C,NAIP,PIK3C2A,PPP3CA,RNF19A,SLC1A2 |
| Glutamate Receptor Signaling | 5.76 | 0.14 | -1 | GLUL,GRIA2,GRID2,GRIN2C,GRM3,SLC1A2,SLC1A3,SLC1A4 |
| Neuropathic Pain Signaling In Dorsal Horn Neurons | 5.63 | 0.099 | -1.265 | FOS,GRIA2,GRIN2C,GRM3,ITPR2,KCNQ3,PIK3C2A,PLCB1,PLCE1,PLCG2 |
| Synaptic Long Term Potentiation | 5.49 | 0.0853 | -1.897 | GNA14,GNAQ,GRIA2,GRIN2C,GRM3,ITPR2,PLCB1,PLCE1,PLCG2,PPP3CA,RASD1 |
| Synaptogenesis Signaling Pathway | 5.4 | 0.0545 | -2 | ADCY2,APOE,CADM1,CDH10,CDH23,EFNA5,EPHB1,GRIA2,GRIN2C,GRM3,KALRN,NLGN4X,NRXN1,PIK3C2A,PLCG2,RASD1,SYNGAP1 |
| Endothelin-1 Signaling | 5.36 | 0.0691 | -0.577 | ADCY2,EDNRB,FOS,GNA12,GNA14,GNAQ,ITPR2,JUN,PIK3C2A,PLCB1,PLCE1,PLCG2,RASD1 |
| Cardiac Hypertrophy Signaling (Enhanced) | 5.32 | 0.0443 | -0.943 | ADCY2,EDNRB,FGFR2,FGFR3,GNA12,GNA14,GNAQ,HSPB1,ITGAV,ITGB4,ITPR2,JUN,MPPED2,NFATC4,PDE4D,PDE7B,PIK3C2A,PLCB1,PLCE1,PLCG2,PPP3CA,RASD1 |
| CREB Signaling in Neurons | 5.06 | 0.0403 | -1.964 | ADCY2,ADGRB1,ADGRB3,ADGRV1,BMPR1B,EDNRB,FGFR2,FGFR3,GNA12,GNA14,GNAQ,GPR37L1,GRIA2,GRID2,GRIN2C,GRM3,HRH1,ITPR2,LGR4,PIK3C2A,PLCB1,PLCE1,PLCG2,RASD1 |
| Neuroinflammation Signaling Pathway | 5 | 0.0533 | 0.333 | B2M,FOS,GABRA2,GABRB1,GLUL,GRIN2C,JUN,MFGE8,NAIP,NFATC4,PIK3C2A,PLCG2,PPP3CA,SLC1A2,SLC1A3,SLC6A1 |
| Cardiac Hypertrophy Signaling | 4.88 | 0.0583 | 0 | ADCY2,GNA12,GNA14,GNAQ,HSPB1,JUN,NFATC4,PIK3C2A,PLCB1,PLCE1,PLCG2,PPP3CA,RASD1,RHOJ |
| Axonal Guidance Signaling | 4.84 | 0.0425 | #NUM! | ADAM33,ADAMTS17,EFNA5,EPHB1,GNA12,GNA14,GNAQ,ITGAV,ITGB4,KALRN,LINGO1,LRRC4C,NFATC4,PIK3C2A,PLCB1,PLCE1,PLCG2,PPP3CA,RASD1,SEMA3D,UNC5C |
| Role of NFAT in Regulation of the Immune Respons | 4.81 | 0.0663 | 0 | FOS,GNA12,GNA14,GNAQ,ITPR2,JUN,NFATC4,PIK3C2A,PLCB1,PLCG2,PPP3CA,RASD1 |
| Endocannabinoid Neuronal Synapse Pathway | 4.71 | 0.0781 | -1.897 | ADCY2,GNA14,GNAQ,GRIA2,GRIN2C,PLCB1,PLCE1,PLCG2,PPP3CA,RIMS1 |
| Synaptic Long Term Depression | 4.62 | 0.0635 | -0.905 | GNA12,GNA14,GNAQ,GRIA2,GRID2,GRM3,ITPR2,PLCB1,PLCE1,PLCG2,PPP2R2B,RASD1 |
| Signaling by Rho Family GTPases | 4.62 | 0.0553 | 0 | CDH10,CDH23,FOS,GFAP,GNA12,GNA14,GNAQ,ITGAV,ITGB4,JUN,PARD3,PIK3C2A,PIP5K1B,RHOJ |
| Ephrin Receptor Signaling | 4.62 | 0.0635 | -1.633 | ABI1,ANGPT1,EFNA5,EPHB1,GNA12,GNA14,GNAQ,GRIN2C,ITGAV,ITGB4,KALRN,RASD1 |
| Adrenomedullin signaling pathway | 4.44 | 0.0609 | -0.905 | ADCY2,FOS,GNA14,GNAQ,HIF1A,ITPR2,KCNQ3,PIK3C2A,PLCB1,PLCE1,PLCG2,RASD1 |
| CXCR4 Signaling | 4.43 | 0.0659 | -1.633 | ADCY2,FOS,GNA12,GNA14,GNAQ,ITPR2,JUN,PIK3C2A,PLCB1,RASD1,RHOJ |
| PI3K Signaling in B Lymphocytes | 4.43 | 0.0725 | 0.333 | FOS,ITPR2,JUN,NFATC4,PLCB1,PLCE1,PLCG2,PPP3CA,RASD1,VAV3 |

| | | | | |
|---|---|---|---|---|
| Relaxin Signaling | 4.1 | 0.0662 | 1 | ADCY2,FOS,GNA12,GNA14,GNAQ,JUN,MPPED2,PDE4D,PDE7B,PIK3C2A |
| Ephrin B Signaling | 4.06 | 0.0972 | -1.89 | ABI1,EPHB1,GNA12,GNA14,GNAQ,KALRN,VAV3 |
| Molecular Mechanisms of Cancer | 4.02 | 0.0425 | #NUM! | ADCY2,APC,BMPR1B,FOS,GNA12,GNA14,GNAQ,HIF1A,ITGAV,ITGB4,JUN,NAIP,PIK3C2A,PLCB1,RASD1,RHOJ,SYNGAP1 |
| GPCR-Mediated Integration of Enteroendocrine Sign | 4.02 | 0.0959 | 0.378 | ADCY2,GNA14,GNAQ,ITPR2,PLCB1,PLCE1,PLCG2 |
| UVA-Induced MAPK Signaling | 4.01 | 0.0816 | 0 | FOS,JUN,PIK3C2A,PLCB1,PLCE1,PLCG2,RASD1,TIPARP |
| P2Y Purigenic Receptor Signaling Pathway | 3.97 | 0.0709 | 0.707 | ADCY2,FOS,GNAQ,JUN,PIK3C2A,PLCB1,PLCE1,PLCG2,RASD1 |
| Gαq Signaling | 3.95 | 0.0637 | -1.265 | GNA14,GNAQ,HRH1,ITPR2,NFATC4,PIK3C2A,PLCB1,PLCG2,PPP3CA,RHOJ |
| Aldosterone Signaling in Epithelial Cells | 3.93 | 0.0633 | -0.816 | DUSP1,HSPA1A/HSPA1B,HSPB1,ITPR2,PIK3C2A,PIP5K1B,PLCB1,PLCE1,PLCG2,SACS |
| GDNF Family Ligand-Receptor Interactions | 3.91 | 0.0921 | 0.447 | FOS,GFRA1,ITPR2,JUN,PIK3C2A,PLCG2,RASD1 |
| Phospholipase C Signaling | 3.79 | 0.0489 | 0 | ADCY2,AHNAK,GNAQ,ITGAV,ITGB4,ITPR2,NFATC4,PLCB1,PLCE1,PLCG2,PPP3CA,RASD1,RHOJ |
| G-Protein Coupled Receptor Signaling | 3.66 | 0.0474 | #NUM! | ADCY2,DUSP1,GNA14,GNAQ,GRM3,HRH1,MPPED2,PDE4D,PDE7B,PIK3C2A,PLCB1,RASD1,SYNGAP1 |
| Role of Macrophages, Fibroblasts and Endothelial C | 3.62 | 0.0446 | #NUM! | APC,APC2,CEBPD,FOS,GNAQ,JUN,NFATC4,PIK3C2A,PLCB1,PLCE1,PLCG2,PPP3CA,RASD1,WIF1 |
| Tec Kinase Signaling | 3.61 | 0.0578 | -0.447 | FOS,GNA12,GNA14,GNAQ,ITGAV,ITGB4,PIK3C2A,PLCG2,RHOJ,VAV3 |
| Thrombin Signaling | 3.58 | 0.0529 | -1 | ADCY2,GNA12,GNA14,GNAQ,ITPR2,PIK3C2A,PLCB1,PLCE1,PLCG2,RASD1,RHOJ |
| Wnt/Ca+ pathway | 3.54 | 0.0968 | 0 | NFATC4,PLCB1,PLCE1,PLCG2,PPP3CA,ROR1 |
| Apelin Endothelial Signaling Pathway | 3.53 | 0.0696 | -0.378 | ADCY2,ANGPT1,FOS,HIF1A,JUN,PIK3C2A,PLCB1,RASD1 |
| Protein Kinase A Signaling | 3.53 | 0.04 | -0.775 | ADCY2,DUSP1,GNAQ,H3-3A/H3-3B,ITPR2,MPPED2,NFATC4,PDE4D,PDE7B,PLCB1,PLCE1,PLCG2,PPP3CA,PTPRE,PTPRG,PTPRJ |
| Role of Tissue Factor in Cancer | 3.5 | 0.069 | #NUM! | F3,GNA12,GNA14,GNAQ,ITGAV,PIK3C2A,PLCB1,RASD1 |
| Sphingosine-1-phosphate Signaling | 3.48 | 0.0684 | -0.707 | ADCY2,GNA12,GNAQ,PIK3C2A,PLCB1,PLCE1,PLCG2,RHOJ |
| Renin-Angiotensin Signaling | 3.45 | 0.0678 | 0 | ADCY2,FOS,GNAQ,ITPR2,JUN,PIK3C2A,PLCG2,RASD1 |
| Regulation of IL-2 Expression in Activated and Anerg | 3.45 | 0.0778 | #NUM! | FOS,JUN,NFATC4,PLCG2,PPP3CA,RASD1,VAV3 |
| Cholecystokinin/Gastrin-mediated Signaling | 3.43 | 0.0672 | -0.378 | FOS,GNA12,GNAQ,ITPR2,JUN,PLCB1,RASD1,RHOJ |
| D-myo-inositol (1,4,5)-Trisphosphate Biosynthesis | 3.35 | 0.16 | 0 | PIP5K1B,PLCB1,PLCE1,PLCG2 |
| GABA Receptor Signaling | 3.3 | 0.0737 | #NUM! | ADCY2,GABRA2,GABRB1,GPHN,KCNQ3,SLC6A1,UBC |
| Actin Cytoskeleton Signaling | 3.26 | 0.0485 | -1.134 | APC,APC2,ARHGAP24,GNA12,ITGAV,ITGB4,PIK3C2A,PIP5K1B,RASD1,VAV3,VCL |
| 14-3-3-mediated Signaling | 3.24 | 0.063 | 0.816 | FOS,GFAP,JUN,PIK3C2A,PLCB1,PLCE1,PLCG2,RASD1 |
| p70S6K Signaling | 3.2 | 0.062 | -0.378 | EEF2K,GNAQ,PIK3C2A,PLCB1,PLCE1,PLCG2,PPP2R2B,RASD1 |
| Dopamine-DARPP32 Feedback in cAMP Signaling | 3.16 | 0.0552 | -1 | ADCY2,GNAQ,GRIN2C,ITPR2,PLCB1,PLCE1,PLCG2,PPP2R2B,PPP3CA |
| Gap Junction Signaling | 3.15 | 0.0505 | #NUM! | ADCY2,GNAQ,GRIA2,ITPR2,PIK3C2A,PLCB1,PLCE1,PLCG2,PPP3CA,RASD1 |

| | | | | |
|---|---|---|---|---|
| ERK/MAPK Signaling | 3.08 | 0.0495 | 1.134 | DUSP1,FOS,H3-3A/H3-3B,HSPB1,ITGAV,ITGB4,PIK3C2A,PLCG2,PPP2R2B,RASD1 |
| FAK Signaling | 3.07 | 0.0673 | #NUM! | CAPN2,ITGAV,ITGB4,PIK3C2A,PLCG2,RASD1,VCL |
| Macropinocytosis Signaling | 3.06 | 0.0789 | #NUM! | ABI1,ITGB4,ITGB8,PIK3C2A,PLCG2,RASD1 |
| T Cell Receptor Signaling | 3.02 | 0.066 | #NUM! | FOS,JUN,NFATC4,PIK3C2A,PPP3CA,RASD1,VAV3 |
| Semaphorin Neuronal Repulsive Signaling Pathway | 2.98 | 0.0576 | 0 | CSPG5,ITGAV,ITGB4,NCAN,PDE4D,PIK3C2A,PLCG2,VCAN |
| Renal Cell Carcinoma Signaling | 2.94 | 0.075 | #NUM! | FOS,HIF1A,JUN,PIK3C2A,RASD1,UBC |
| Chemokine Signaling | 2.94 | 0.075 | 0.447 | FOS,GNAQ,JUN,PLCB1,PLCG2,RASD1 |
| Role of NFAT in Cardiac Hypertrophy | 2.89 | 0.0467 | -0.333 | ADCY2,GNAQ,ITPR2,NFATC4,PIK3C2A,PLCB1,PLCE1,PLCG2,PPP3CA,RASD1 |
| GPCR-Mediated Nutrient Sensing in Enteroendocrine | 2.88 | 0.0625 | -0.378 | ADCY2,GNA14,GNAQ,ITPR2,PLCB1,PLCE1,PLCG2 |
| Role of Osteoblasts, Osteoclasts and Chondrocytes | 2.83 | 0.0459 | #NUM! | APC,APC2,FOS,JUN,NAIP,NFATC4,PIK3C2A,PPP3CA,SPP1,WIF1 |
| Sperm Motility | 2.76 | 0.0448 | -0.447 | EPHB1,ERBB4,FGFR2,FGFR3,ITPR2,PDE4D,PLCB1,PLCE1,PLCG2,ROR1 |
| RhoGDI Signaling | 2.71 | 0.0476 | 1 | CDH10,CDH23,GNA12,GNA14,GNAQ,ITGAV,ITGB4,PIP5K1B,RHOJ |
| HGF Signaling | 2.71 | 0.0583 | 1 | FOS,ITGAV,ITGB4,JUN,PIK3C2A,PLCG2,RASD1 |
| ILK Signaling | 2.7 | 0.0474 | 1 | FOS,HIF1A,ITGB4,ITGB8,JUN,PIK3C2A,PPP2R2B,RHOJ,VCL |
| Rac Signaling | 2.69 | 0.0579 | #NUM! | ITGAV,ITGB4,JUN,PARD3,PIK3C2A,PIP5K1B,RASD1 |
| PKCθ Signaling in T Lymphocytes | 2.68 | 0.0516 | 0.378 | FOS,JUN,NFATC4,PIK3C2A,PLCG2,PPP3CA,RASD1,VAV3 |
| Thrombopoietin Signaling | 2.64 | 0.0794 | 1 | FOS,JUN,PIK3C2A,PLCG2,RASD1 |
| IL-1 Signaling | 2.63 | 0.0652 | #NUM! | ADCY2,FOS,GNA12,GNA14,GNAQ,JUN |
| ErbB Signaling | 2.59 | 0.0638 | 0.447 | ERBB4,FOS,JUN,PIK3C2A,PLCG2,RASD1 |
| Superpathway of Inositol Phosphate Compounds | 2.56 | 0.0452 | 0.333 | DUSP1,ITPKB,PIK3C2A,PIP5K1B,PLCB1,PLCE1,PLCG2,PPP3CA,PTPRJ |
| IL-8 Signaling | 2.55 | 0.045 | 0.378 | ANGPT1,FOS,GNA12,ITGAV,JUN,PIK3C2A,RAB11FIP2,RASD1,RHOJ |
| Glioblastoma Multiforme Signaling | 2.51 | 0.0485 | -0.378 | APC,ITPR2,PIK3C2A,PLCB1,PLCE1,PLCG2,RASD1,RHOJ |
| p53 Signaling | 2.49 | 0.0612 | -0.816 | ADGRB1,HIF1A,JUN,PIK3C2A,PLAGL1,SERPINE2 |
| Cellular Effects of Sildenafil (Viagra) | 2.49 | 0.0534 | #NUM! | ADCY2,ITPR2,KCNQ3,PDE4D,PLCB1,PLCE1,PLCG2 |
| Gα12/13 Signaling | 2.49 | 0.0534 | 0 | CDH10,CDH23,GNA12,JUN,PIK3C2A,RASD1,VAV3 |
| Estrogen Receptor Signaling | 2.46 | 0.0366 | 0.577 | ADCY2,FOS,GNA12,GNA14,GNAQ,HIF1A,JUN,PIK3C2A,PLCB1,PLCE1,PLCG2,RASD1 |
| Phagosome Formation | 2.46 | 0.0526 | #NUM! | ITGAV,ITGB4,PIK3C2A,PLCB1,PLCE1,PLCG2,RHOJ |
| Virus Entry via Endocytic Pathways | 2.41 | 0.0588 | #NUM! | B2M,ITGB4,ITGB8,PIK3C2A,PLCG2,RASD1 |
| PTEN Signaling | 2.4 | 0.0515 | 2 | BMPR1B,FGFR2,FGFR3,ITGAV,ITGB4,PREX2,RASD1 |
| GNRH Signaling | 2.39 | 0.0462 | -0.378 | ADCY2,FOS,GNA14,GNAQ,ITPR2,JUN,PLCB1,RASD1 |
| Melatonin Signaling | 2.39 | 0.0694 | -0.447 | GNAQ,PLCB1,PLCE1,PLCG2,RORB |
| Integrin Signaling | 2.37 | 0.0423 | 0 | CAPN2,ITGAV,ITGB4,ITGB8,PIK3C2A,PLCG2,RASD1,RHOJ,VCL |
| T Cell Exhaustion Signaling Pathway | 2.36 | 0.0457 | -0.816 | FOS,JUN,MGAT5,NFATC4,PIK3C2A,PLCG2,PPP2R2B,RASD1 |
| Neuregulin Signaling | 2.35 | 0.0571 | #NUM! | ERBB4,ERBIN,ITGAV,ITGB4,PLCG2,RASD1 |
| Cdc42 Signaling | 2.35 | 0.0455 | 1 | APC,APC2,B2M,FOS,ITGAV,ITGB4,JUN,PARD3 |
| Leptin Signaling in Obesity | 2.34 | 0.0676 | #NUM! | ADCY2,PIK3C2A,PLCB1,PLCE1,PLCG2 |
| Regulation of Cellular Mechanics by Calpain Proteas | 2.34 | 0.0676 | #NUM! | CAPN2,ITGAV,ITGB4,RASD1,VCL |
| Estrogen-Dependent Breast Cancer Signaling | 2.31 | 0.0667 | #NUM! | FOS,HSD17B12,JUN,PIK3C2A,RASD1 |

| | | | | |
|---|---|---|---|---|
| nNOS Signaling in Neurons | 2.31 | 0.0851 | #NUM! | CAPN2,GRIN2C,PPP3CA,RASD1 |
| Paxillin Signaling | 2.29 | 0.0556 | #NUM! | ITGAV,ITGB4,ITGB8,PIK3C2A,RASD1,VCL |
| IL-3 Signaling | 2.22 | 0.0633 | 0 | FOS,JUN,PIK3C2A,PPP3CA,RASD1 |
| JAK/Stat Signaling | 2.19 | 0.0625 | 0 | FOS,GNAQ,JUN,PIK3C2A,RASD1 |
| Prolactin Signaling | 2.17 | 0.0617 | 1 | FOS,JUN,PIK3C2A,PLCG2,RASD1 |
| Actin Nucleation by ARP-WASP Complex | 2.17 | 0.0617 | #NUM! | GNA12,ITGAV,ITGB4,RASD1,RHOJ |
| UVB-Induced MAPK Signaling | 2.15 | 0.0769 | #NUM! | FOS,H3-3A/H3-3B,JUN,PIK3C2A |
| fMLP Signaling in Neutrophils | 2.14 | 0.0517 | -1.342 | ITPR2,NFATC4,PIK3C2A,PLCB1,PPP3CA,RASD1 |
| PPARα/RXRα Activation | 2.14 | 0.0419 | #NUM! | ADCY2,GNA14,GNAQ,JUN,PLCB1,PLCE1,PLCG2,RASD1 |
| EGF Signaling | 2.07 | 0.0727 | 0 | FOS,ITPR2,JUN,PIK3C2A |
| PDGF Signaling | 2.06 | 0.0581 | 1 | FOS,JUN,PIK3C2A,PLCG2,RASD1 |
| CD28 Signaling in T Helper Cells | 2.05 | 0.0496 | 0 | FOS,ITPR2,JUN,NFATC4,PIK3C2A,PPP3CA |
| Huntington's Disease Signaling | 2.05 | 0.0377 | 0 | CAPN2,GNA14,GNAQ,HAP1,HSPA1A/HSPA1B,JUN,PIK3C2A,PLCB1,UBC |
| G Beta Gamma Signaling | 2.04 | 0.0492 | 0.447 | ADCY2,GNA12,GNA14,GNAQ,PLCG2,RASD1 |
| TNFR2 Signaling | 2.03 | 0.1 | #NUM! | FOS,JUN,NAIP |
| Ceramide Signaling | 2.02 | 0.0568 | 1 | FOS,JUN,PIK3C2A,PPP2R2B,RASD1 |
| G Protein Signaling Mediated by Tubby | 1.99 | 0.0968 | #NUM! | GNAQ,PLCB1,PLCG2 |
| IL-2 Signaling | 1.91 | 0.0656 | #NUM! | FOS,JUN,PIK3C2A,RASD1 |
| α-Adrenergic Signaling | 1.89 | 0.0526 | #NUM! | ADCY2,GNAQ,ITPR2,PLCG2,RASD1 |
| TGF-β Signaling | 1.87 | 0.0521 | 1 | BMPR1B,FOS,JUN,RASD1,SMURF2 |
| Wnt/β-catenin Signaling | 1.85 | 0.0405 | 0.816 | APC,APC2,GNAQ,JUN,PPP2R2B,UBC,WIF1 |
| MSP-RON Signaling In Cancer Cells Pathway | 1.85 | 0.0448 | 0 | FOS,HIF1A,ITGB4,JUN,PIK3C2A,RASD1 |
| Glutamine Biosynthesis I | 1.84 | 1 | #NUM! | GLUL |
| VEGF Signaling | 1.82 | 0.0505 | 0 | HIF1A,PIK3C2A,PLCG2,RASD1,VCL |
| Apelin Cardiomyocyte Signaling Pathway | 1.82 | 0.0505 | -0.447 | HIF1A,PIK3C2A,PLCB1,PLCE1,PLCG2 |
| Tumor Microenvironment Pathway | 1.82 | 0.0398 | 0.378 | FOS,HIF1A,JUN,PIK3C2A,RASD1,SLC1A4,SPP1 |
| IL-17A Signaling in Fibroblasts | 1.81 | 0.0833 | #NUM! | CEBPD,FOS,JUN |
| ErbB4 Signaling | 1.77 | 0.0597 | #NUM! | ERBB4,PIK3C2A,PLCG2,RASD1 |
| B Cell Receptor Signaling | 1.7 | 0.0376 | 0 | JUN,NFATC4,PIK3C2A,PLCG2,PPP3CA,RASD1,VAV3 |
| Corticotropin Releasing Hormone Signaling | 1.7 | 0.0414 | 1 | ADCY2,FOS,GNAQ,ITPR2,JUN,PLCG2 |
| April Mediated Signaling | 1.69 | 0.075 | #NUM! | FOS,JUN,NFATC4 |
| Regulation Of The Epithelial Mesenchymal Transition | 1.68 | 0.0372 | -0.378 | FGFR2,FGFR3,FOS,JUN,PIK3C2A,RASD1,WWTR1 |
| ERK5 Signaling | 1.67 | 0.0556 | #NUM! | FOS,GNA12,GNAQ,RASD1 |
| CDK5 Signaling | 1.67 | 0.0463 | 0 | ADCY2,CABLES1,LAMA5,PPP2R2B,RASD1 |
| NRF2-mediated Oxidative Stress Response | 1.67 | 0.037 | 0 | FKBP5,FOS,FTH1,JUN,JUNB,PIK3C2A,RASD1 |
| Production of Nitric Oxide and Reactive Oxygen Spe | 1.67 | 0.037 | 0.378 | APOE,FOS,JUN,PIK3C2A,PLCG2,PPP2R2B,RHOJ |
| B Cell Activating Factor Signaling | 1.66 | 0.0732 | #NUM! | FOS,JUN,NFATC4 |
| Caveolar-mediated Endocytosis Signaling | 1.65 | 0.0548 | #NUM! | B2M,ITGAV,ITGB4,ITGB8 |
| Glioma Invasiveness Signaling | 1.65 | 0.0548 | #NUM! | ITGAV,PIK3C2A,RASD1,RHOJ |
| Hepatic Fibrosis Signaling Pathway | 1.62 | 0.0291 | 0.333 | FOS,FTH1,HIF1A,ITGAV,ITGB4,JUN,PIK3C2A,PLCG2,RASD1,RHOJ,SPP1 |

| | | | | |
|---|---|---|---|---|
| HER-2 Signaling in Breast Cancer | 1.61 | 0.0361 | 1.89 | FOS,ITGB4,ITGB8,JUN,PARD3,PIK3C2A,RASD1 |
| Oncostatin M Signaling | 1.6 | 0.0698 | #NUM! | CHI3L1,EPAS1,RASD1 |
| Neurotrophin/TRK Signaling | 1.6 | 0.0526 | #NUM! | FOS,JUN,PIK3C2A,RASD1 |
| Natural Killer Cell Signaling | 1.58 | 0.0355 | 0.378 | B2M,HSPA1A/HSPA1B,NFATC4,PIK3C2A,PLCG2,RASD1,VAV3 |
| tRNA Splicing | 1.58 | 0.0682 | #NUM! | MPPED2,PDE4D,PDE7B |
| Insulin Secretion Signaling Pathway | 1.56 | 0.0328 | -0.707 | ADCY2,GNA14,GNAQ,ITPR2,PIK3C2A,PLCB1,PLCE1,PLCG2 |
| Breast Cancer Regulation by Stathmin1 | 1.55 | 0.0254 | -2.324 | ADGRB1,ADGRB3,ADGRV1,EDNRB,GNAQ,GPR37L1,GRM3,HIF1A,HRH1,JUN,LGR4,PIK3C2A,PLCB1,PPP2R2B,RASD1 |
| D-myo-inositol-5-phosphate Metabolism | 1.55 | 0.0382 | 0.816 | DUSP1,PLCB1,PLCE1,PLCG2,PPP3CA,PTPRJ |
| Thyroid Cancer Signaling | 1.54 | 0.0506 | 1 | FOS,JUN,PIK3C2A,RASD1 |
| BEX2 Signaling Pathway | 1.54 | 0.0506 | 1 | HIF1A,JUN,PPP2R2B,SPP1 |
| Asparagine Degradation I | 1.54 | 0.5 | #NUM! | ASRGL1 |
| Glutamate Biosynthesis II | 1.54 | 0.5 | #NUM! | GLUD1 |
| Glutamate Degradation X | 1.54 | 0.5 | #NUM! | GLUD1 |
| Opioid Signaling Pathway | 1.53 | 0.0324 | -1.134 | ADCY2,FOS,GRIN2C,ITPR2,PLCB1,PPP3CA,RASD1,RGS6 |
| eNOS Signaling | 1.52 | 0.0377 | -0.447 | ADCY2,GNAQ,HSPA1A/HSPA1B,ITPR2,PIK3C2A,PLCG2 |
| Oxidative Ethanol Degradation III | 1.51 | 0.105 | #NUM! | ACSS1,ALDH1A1 |
| Ephrin A Signaling | 1.5 | 0.0638 | #NUM! | EFNA5,PIK3C2A,VAV3 |
| HIF1α Signaling | 1.5 | 0.0341 | -0.378 | HIF1A,HSPA1A/HSPA1B,JUN,PIK3C2A,PLCG2,PPP3CA,RASD1 |
| IL-15 Production | 1.48 | 0.0413 | -2.236 | EPHB1,ERBB4,FGFR2,FGFR3,ROR1 |
| LPS-stimulated MAPK Signaling | 1.47 | 0.0482 | #NUM! | FOS,JUN,PIK3C2A,RASD1 |
| TR/RXR Activation | 1.46 | 0.0476 | #NUM! | DIO2,HIF1A,ME1,PIK3C2A |
| VEGF Family Ligand-Receptor Interactions | 1.46 | 0.0476 | #NUM! | FOS,PIK3C2A,PLCG2,RASD1 |
| TNFR1 Signaling | 1.43 | 0.06 | #NUM! | FOS,JUN,NAIP |
| GP6 Signaling Pathway | 1.43 | 0.04 | 1.342 | COL21A1,COL27A1,LAMA5,PIK3C2A,PLCG2 |
| IL-6 Signaling | 1.42 | 0.0397 | 1 | FOS,HSPB1,JUN,PIK3C2A,RASD1 |
| Ferroptosis Signaling Pathway | 1.42 | 0.0397 | -0.447 | ANGPTL4,FTH1,HSPB1,RASD1,WWTR1 |
| UVC-Induced MAPK Signaling | 1.41 | 0.0588 | #NUM! | FOS,JUN,RASD1 |
| RANK Signaling in Osteoclasts | 1.38 | 0.0449 | 0 | FOS,JUN,PIK3C2A,PPP3CA |
| Erythropoietin Signaling Pathway | 1.37 | 0.0347 | 0 | FOS,HIF1A,ITPR2,JUN,PIK3C2A,RASD1 |
| L-carnitine Biosynthesis | 1.36 | 0.333 | #NUM! | BBOX1 |
| Thyronamine and Iodothyronamine Metabolism | 1.36 | 0.333 | #NUM! | DIO2 |
| Thyroid Hormone Metabolism I (via Deiodination) | 1.36 | 0.333 | #NUM! | DIO2 |
| Ethanol Degradation IV | 1.35 | 0.087 | #NUM! | ACSS1,ALDH1A1 |
| Adipogenesis pathway | 1.32 | 0.0373 | #NUM! | CEBPD,FGFR2,FGFR3,HIF1A,NFATC4 |
| NF-κB Signaling | 1.31 | 0.0335 | -1.342 | BMPR1B,FGFR2,FGFR3,PIK3C2A,PLCG2,RASD1 |
| Melanocyte Development and Pigmentation Signalin | 1.31 | 0.0426 | #NUM! | ADCY2,PIK3C2A,PLCG2,RASD1 |
| Senescence Pathway | 1.3 | 0.0291 | 0 | CAPN2,ITPR2,JUN,NFATC4,PIK3C2A,PPP2R2B,PPP3CA,RASD1 |
| Iron homeostasis signaling pathway | 1.29 | 0.0365 | #NUM! | BMPR1B,EPAS1,FTH1,HIF1A,HIF3A |
| cAMP-mediated signaling | 1.28 | 0.0306 | -1.633 | ADCY2,DUSP1,GRM3,MPPED2,PDE4D,PDE7B,PPP3CA |
| ATM Signaling | 1.27 | 0.0412 | #NUM! | HP1BP3,JUN,PPP2R2B,RAD9A |
| IL-17A Signaling in Gastric Cells | 1.26 | 0.0769 | #NUM! | FOS,JUN |

| | | | | |
|---|---|---|---|---|
| Phenylalanine Degradation I (Aerobic) | 1.24 | 0.25 | #NUM! | QDPR |
| Molybdenum Cofactor Biosynthesis | 1.24 | 0.25 | #NUM! | GPHN |
| Acetate Conversion to Acetyl-CoA | 1.24 | 0.25 | #NUM! | ACSS1 |
| Endometrial Cancer Signaling | 1.24 | 0.05 | #NUM! | APC2,PIK3C2A,RASD1 |
| PCP pathway | 1.24 | 0.05 | #NUM! | JUN,JUNB,LGR4 |
| Cardiac β-adrenergic Signaling | 1.24 | 0.0352 | 0 | ADCY2,MPPED2,PDE4D,PDE7B,PPP2R2B |
| Systemic Lupus Erythematosus In T Cell Signaling P | 1.23 | 0.0269 | -1.667 | B2M,FOS,GNA12,JUN,PIK3C2A,PPP2R2B,PPP3CA,RASD1,RHOJ |
| Apoptosis Signaling | 1.23 | 0.04 | #NUM! | CAPN2,NAIP,PLCG2,RASD1 |
| Aryl Hydrocarbon Receptor Signaling | 1.23 | 0.035 | #NUM! | ALDH1A1,DHFR,FOS,HSPB1,JUN |
| SAPK/JNK Signaling | 1.2 | 0.0392 | #NUM! | GNA12,JUN,PIK3C2A,RASD1 |
| Regulation of the Epithelial-Mesenchymal Transition | 1.2 | 0.0312 | #NUM! | APC,FGFR2,FGFR3,HIF1A,PIK3C2A,RASD1 |
| Regulation of Actin-based Motility by Rho | 1.19 | 0.0388 | #NUM! | ITGAV,ITGB4,PIP5K1B,RHOJ |
| Clathrin-mediated Endocytosis Signaling | 1.19 | 0.0311 | #NUM! | APOE,ITGB4,ITGB8,PIK3C2A,PPP3CA,UBC |
| RAR Activation | 1.18 | 0.0309 | #NUM! | ADCY2,ALDH1A1,DUSP1,FOS,JUN,ZBTB16 |
| IGF-1 Signaling | 1.18 | 0.0385 | #NUM! | FOS,JUN,PIK3C2A,RASD1 |
| CD40 Signaling | 1.15 | 0.0462 | #NUM! | FOS,JUN,PIK3C2A |
| Factors Promoting Cardiogenesis in Vertebrates | 1.15 | 0.0333 | 0.447 | APC,BMPR1B,PLCB1,PLCE1,PLCG2 |
| ErbB2-ErbB3 Signaling | 1.15 | 0.0462 | #NUM! | JUN,PIK3C2A,RASD1 |
| Netrin Signaling | 1.15 | 0.0462 | #NUM! | NFATC4,PPP3CA,UNC5C |
| Phospholipases | 1.15 | 0.0462 | #NUM! | PLCB1,PLCE1,PLCG2 |
| PAK Signaling | 1.15 | 0.0377 | #NUM! | ITGAV,ITGB4,PIK3C2A,RASD1 |
| Serine Biosynthesis | 1.15 | 0.2 | #NUM! | UBAC2 |
| dTMP De Novo Biosynthesis | 1.15 | 0.2 | #NUM! | DHFR |
| Calcium-induced T Lymphocyte Apoptosis | 1.14 | 0.0455 | #NUM! | CAPN2,ITPR2,PPP3CA |
| Gustation Pathway | 1.11 | 0.0323 | #NUM! | ADCY2,ITPR2,MPPED2,PDE4D,PDE7B |
| Colorectal Cancer Metastasis Signaling | 1.1 | 0.0277 | 0.816 | ADCY2,APC,FOS,JUN,PIK3C2A,RASD1,RHOJ |
| Ethanol Degradation II | 1.1 | 0.0625 | #NUM! | ACSS1,ALDH1A1 |
| iCOS-iCOSL Signaling in T Helper Cells | 1.09 | 0.036 | #NUM! | ITPR2,NFATC4,PIK3C2A,PPP3CA |
| Necroptosis Signaling Pathway | 1.09 | 0.0318 | -0.447 | CAPN2,GLUD1,GLUL,PPP3CA,UBC |
| Agrin Interactions at Neuromuscular Junction | 1.08 | 0.0429 | #NUM! | ERBB4,JUN,RASD1 |
| GM-CSF Signaling | 1.08 | 0.0429 | #NUM! | PIK3C2A,PPP3CA,RASD1 |
| Circadian Rhythm Signaling | 1.08 | 0.0606 | #NUM! | CRY2,GRIN2C |
| MSP-RON Signaling In Macrophages Pathway | 1.07 | 0.0354 | 1 | FOS,JUN,PIK3C2A,RASD1 |
| Arginine Biosynthesis IV | 1.07 | 0.167 | #NUM! | GLUD1 |
| Growth Hormone Signaling | 1.07 | 0.0423 | #NUM! | FOS,PIK3C2A,PLCG2 |
| Inhibition of Angiogenesis by TSP1 | 1.05 | 0.0588 | #NUM! | CD47,JUN |
| Non-Small Cell Lung Cancer Signaling | 1.04 | 0.0411 | #NUM! | ITPR2,PIK3C2A,RASD1 |
| Fc Epsilon RI Signaling | 1.03 | 0.0342 | #NUM! | PIK3C2A,PLCG2,RASD1,VAV3 |
| Role of PKR in Interferon Induction and Antiviral Res | 1.02 | 0.0339 | #NUM! | COLEC12,FOS,HSPA1A/HSPA1B,JUN |
| p38 MAPK Signaling | 1.02 | 0.0339 | 0 | DUSP1,EEF2K,H3-3A/H3-3B,HSPB1 |
| HMGB1 Signaling | 1.02 | 0.0303 | 0 | FOS,JUN,PIK3C2A,RASD1,RHOJ |
| Regulation of eIF4 and p70S6K Signaling | 1.01 | 0.0301 | #NUM! | ITGAV,ITGB4,PIK3C2A,PPP2R2B,RASD1 |

| | | | | |
|---|---|---|---|---|
| 3-phosphoinositide Biosynthesis | 1.01 | 0.0301 | -0.447 | DUSP1,PIK3C2A,PIP5K1B,PPP3CA,PTPRJ |
| FcγRIIB Signaling in B Lymphocytes | 1.01 | 0.04 | #NUM! | PIK3C2A,PLCG2,RASD1 |
| Angiopoietin Signaling | 1.01 | 0.04 | #NUM! | ANGPT1,PIK3C2A,RASD1 |
| Role of NANOG in Mammalian Embryonic Stem Cell I | 1.01 | 0.0336 | #NUM! | APC,BMPR1B,PIK3C2A,RASD1 |
| Superpathway of Serine and Glycine Biosynthesis I | 1.01 | 0.143 | #NUM! | UBAC2 |
| NF-κB Activation by Viruses | 1 | 0.0395 | #NUM! | ITGAV,PIK3C2A,RASD1 |
| Toll-like Receptor Signaling | 1 | 0.0395 | #NUM! | FOS,JUN,UBC |
| Dopamine Receptor Signaling | 0.987 | 0.039 | #NUM! | ADCY2,PPP2R2B,QDPR |
| Osteoarthritis Pathway | 0.979 | 0.0273 | 1 | EPAS1,FGFR3,HIF1A,ITGAV,ITGB4,SPP1 |
| Protein Ubiquitination Pathway | 0.971 | 0.0256 | #NUM! | B2M,HSPA1A/HSPA1B,HSPB1,SACS,SMURF2,UBC,USP24 |
| CCR3 Signaling in Eosinophils | 0.963 | 0.0323 | #NUM! | ITPR2,PIK3C2A,PLCB1,RASD1 |
| Systemic Lupus Erythematosus In B Cell Signaling P | 0.955 | 0.0255 | 1.134 | FOS,JUN,NFATC4,PIK3C2A,PLCG2,PPP3CA,RASD1 |
| Systemic Lupus Erythematosus Signaling | 0.917 | 0.0262 | #NUM! | FOS,JUN,NFATC4,PIK3C2A,PLCG2,RASD1 |
| White Adipose Tissue Browning Pathway | 0.917 | 0.031 | -1 | ADCY2,DIO2,FGFR2,FGFR3 |
| FGF Signaling | 0.903 | 0.0357 | #NUM! | FGFR2,FGFR3,PIK3C2A |
| MIF Regulation of Innate Immunity | 0.9 | 0.0476 | #NUM! | FOS,JUN |
| HIPPO signaling | 0.889 | 0.0353 | #NUM! | PARD3,PPP2R2B,WWTR1 |
| IL-4 Signaling | 0.889 | 0.0353 | #NUM! | NFATC4,PIK3C2A,RASD1 |
| BMP signaling pathway | 0.889 | 0.0353 | #NUM! | BMPR1B,JUN,RASD1 |
| Serotonin Receptor Signaling | 0.883 | 0.0465 | #NUM! | ADCY2,QDPR |
| IL-12 Signaling and Production in Macrophages | 0.879 | 0.0301 | #NUM! | APOE,FOS,JUN,PIK3C2A |
| Dendritic Cell Maturation | 0.873 | 0.0272 | 0.447 | B2M,PIK3C2A,PLCB1,PLCE1,PLCG2 |
| IL-23 Signaling Pathway | 0.866 | 0.0455 | #NUM! | HIF1A,PIK3C2A |
| Human Embryonic Stem Cell Pluripotency | 0.863 | 0.0296 | #NUM! | APC,FGFR2,FGFR3,PIK3C2A |
| STAT3 Pathway | 0.863 | 0.0296 | #NUM! | BMPR1B,FGFR2,FGFR3,RASD1 |
| Sirtuin Signaling Pathway | 0.863 | 0.0241 | -0.816 | EPAS1,GLUD1,H3-3A/H3-3B,HIF1A,JUN,MT-ND3,NDRG1 |
| Hepatic Fibrosis / Hepatic Stellate Cell Activation | 0.86 | 0.0269 | #NUM! | COL21A1,COL27A1,EDNRB,FGFR2,LHX2 |
| Androgen Signaling | 0.857 | 0.0294 | #NUM! | GNA12,GNA14,GNAQ,JUN |
| iNOS Signaling | 0.851 | 0.0444 | #NUM! | FOS,JUN |
| CTLA4 Signaling in Cytotoxic T Lymphocytes | 0.848 | 0.0337 | #NUM! | B2M,PIK3C2A,PPP2R2B |
| Dermatan Sulfate Biosynthesis (Late Stages) | 0.836 | 0.0435 | #NUM! | HS3ST5,UST |
| Death Receptor Signaling | 0.818 | 0.0326 | #NUM! | HSPB1,NAIP,TIPARP |
| Sertoli Cell-Sertoli Cell Junction Signaling | 0.807 | 0.0258 | #NUM! | ITGAV,ITGB4,JUN,RASD1,VCL |
| Chondroitin Sulfate Biosynthesis (Late Stages) | 0.807 | 0.0417 | #NUM! | HS3ST5,UST |
| Endocannabinoid Cancer Inhibition Pathway | 0.801 | 0.028 | 0 | ADCY2,GNAQ,HIF1A,PIK3C2A |
| CCR5 Signaling in Macrophages | 0.799 | 0.0319 | #NUM! | FOS,JUN,PLCG2 |
| Melanoma Signaling | 0.78 | 0.04 | #NUM! | PIK3C2A,RASD1 |
| CD27 Signaling in Lymphocytes | 0.74 | 0.0377 | #NUM! | FOS,JUN |
| Epithelial Adherens Junction Signaling | 0.735 | 0.0263 | #NUM! | APC,PARD3,RASD1,VCL |
| Calcium Signaling | 0.733 | 0.0243 | -1.342 | GRIA2,GRIN2C,ITPR2,NFATC4,PPP3CA |
| Mouse Embryonic Stem Cell Pluripotency | 0.717 | 0.0291 | #NUM! | APC,PIK3C2A,RASD1 |
| Sumoylation Pathway | 0.717 | 0.0291 | #NUM! | FOS,JUN,RHOJ |

| | | | | |
|---|---|---|---|---|
| mTOR Signaling | 0.71 | 0.0238 | -1 | HIF1A,PIK3C2A,PPP2R2B,RASD1,RHOJ |
| Chondroitin Sulfate Biosynthesis | 0.703 | 0.0357 | #NUM! | HS3ST5,UST |
| Unfolded protein response | 0.703 | 0.0357 | #NUM! | CEBPD,HSPA1A/HSPA1B |
| PPAR Signaling | 0.701 | 0.0286 | #NUM! | FOS,JUN,RASD1 |
| CNTF Signaling | 0.693 | 0.0351 | #NUM! | PIK3C2A,RASD1 |
| Role of CHK Proteins in Cell Cycle Checkpoint Contr | 0.693 | 0.0351 | #NUM! | PPP2R2B,RAD9A |
| Telomerase Signaling | 0.684 | 0.028 | #NUM! | PIK3C2A,PPP2R2B,RASD1 |
| HOTAIR Regulatory Pathway | 0.682 | 0.025 | #NUM! | H3-3A/H3-3B,PIK3C2A,SPP1,WIF1 |
| Extrinsic Prothrombin Activation Pathway | 0.678 | 0.0625 | #NUM! | F3 |
| Dermatan Sulfate Biosynthesis | 0.67 | 0.0339 | #NUM! | HS3ST5,UST |
| Antioxidant Action of Vitamin C | 0.67 | 0.0275 | #NUM! | PLCB1,PLCE1,PLCG2 |
| Glioma Signaling | 0.662 | 0.0273 | #NUM! | PIK3C2A,PLCG2,RASD1 |
| Histamine Degradation | 0.654 | 0.0588 | #NUM! | ALDH1A1 |
| Tight Junction Signaling | 0.635 | 0.0238 | #NUM! | FOS,JUN,PPP2R2B,VCL |
| 1D-myo-inositol Hexakisphosphate Biosynthesis II (M | 0.633 | 0.0556 | #NUM! | ITPKB |
| D-myo-inositol (1,3,4)-trisphosphate Biosynthesis | 0.633 | 0.0556 | #NUM! | ITPKB |
| NGF Signaling | 0.631 | 0.0263 | #NUM! | PIK3C2A,PLCG2,RASD1 |
| LPS/IL-1 Mediated Inhibition of RXR Function | 0.629 | 0.0222 | #NUM! | ALDH1A1,APOE,HS3ST5,JUN,UST |
| Glucocorticoid Receptor Signaling | 0.623 | 0.0195 | #NUM! | DUSP1,FKBP5,FOS,HSPA1A/HSPA1B,JUN,NFATC4,PIK3C2A,PPP3CA,RASD1 |
| Endocannabinoid Developing Neuron Pathway | 0.623 | 0.0261 | #NUM! | ADCY2,PIK3C2A,RASD1 |
| Germ Cell-Sertoli Cell Junction Signaling | 0.616 | 0.0234 | #NUM! | PIK3C2A,RASD1,RHOJ,VCL |
| DNA damage-induced 14-3-3σ Signaling | 0.613 | 0.0526 | #NUM! | RAD9A |
| Xenobiotic Metabolism Signaling | 0.611 | 0.0209 | #NUM! | ALDH1A1,HS3ST5,PIK3C2A,PPP2R2B,RASD1,UST |
| Cardiomyocyte Differentiation via BMP Receptors | 0.592 | 0.05 | #NUM! | BMPR1B |
| Fatty Acid α-oxidation | 0.592 | 0.05 | #NUM! | ALDH1A1 |
| Inflammasome pathway | 0.592 | 0.05 | #NUM! | NAIP |
| Remodeling of Epithelial Adherens Junctions | 0.58 | 0.0294 | #NUM! | APC,VCL |
| Reelin Signaling in Neurons | 0.577 | 0.0246 | #NUM! | APOE,GRIN2C,PIK3C2A |
| Inhibition of ARE-Mediated mRNA Degradation Path | 0.577 | 0.0246 | #NUM! | PPP2R2B,ZFP36,ZFP36L2 |
| Putrescine Degradation III | 0.575 | 0.0476 | #NUM! | ALDH1A1 |
| Role of JAK1 and JAK3 in γc Cytokine Signaling | 0.572 | 0.029 | #NUM! | PIK3C2A,RASD1 |
| SPINK1 General Cancer Pathway | 0.572 | 0.029 | #NUM! | PIK3C2A,RASD1 |
| RhoA Signaling | 0.57 | 0.0244 | #NUM! | GNA12,PIP5K1B,RHPN1 |
| IL-10 Signaling | 0.564 | 0.0286 | #NUM! | FOS,JUN |
| CDP-diacylglycerol Biosynthesis I | 0.558 | 0.0455 | #NUM! | GPAM |
| Methionine Degradation I (to Homocysteine) | 0.558 | 0.0455 | #NUM! | AHCYL1 |
| Gαi Signaling | 0.558 | 0.024 | #NUM! | ADCY2,GRM3,RASD1 |
| Basal Cell Carcinoma Signaling | 0.547 | 0.0278 | #NUM! | APC,APC2 |
| Heparan Sulfate Biosynthesis (Late Stages) | 0.547 | 0.0278 | #NUM! | HS3ST5,UST |
| PI3K/AKT Signaling | 0.547 | 0.0217 | #NUM! | ITGAV,ITGB4,PPP2R2B,RASD1 |
| Polyamine Regulation in Colon Cancer | 0.541 | 0.0435 | #NUM! | APC |

| | | | | |
|---|---|---|---|---|
| Superpathway of D-myo-inositol (1,4,5)-trisphosphate | 0.541 | 0.0435 | #NUM! | ITPKB |
| IL-17 Signaling | 0.533 | 0.0214 | 1 | FOS,JUN,PIK3C2A,RASD1 |
| Hypoxia Signaling in the Cardiovascular System | 0.53 | 0.027 | #NUM! | HIF1A,JUN |
| Phosphatidylglycerol Biosynthesis II (Non-plastidic) | 0.526 | 0.0417 | #NUM! | GPAM |
| Cysteine Biosynthesis III (mammalia) | 0.526 | 0.0417 | #NUM! | AHCYL1 |
| IL-15 Signaling | 0.523 | 0.0267 | #NUM! | PIK3C2A,RASD1 |
| Xenobiotic Metabolism CAR Signaling Pathway | 0.523 | 0.0212 | -2 | ALDH1A1,HS3ST5,PPP2R2B,UST |
| Tryptophan Degradation X (Mammalian, via Tryptami | 0.511 | 0.04 | #NUM! | ALDH1A1 |
| Antiproliferative Role of Somatostatin Receptor 2 | 0.507 | 0.026 | #NUM! | PIK3C2A,RASD1 |
| Leukocyte Extravasation Signaling | 0.504 | 0.0207 | #NUM! | PIK3C2A,PLCG2,VAV3,VCL |
| Role of Wnt/GSK-3β Signaling in the Pathogenesis c | 0.499 | 0.0256 | #NUM! | APC,APC2 |
| IL-7 Signaling Pathway | 0.499 | 0.0256 | #NUM! | JUN,PIK3C2A |
| Lipid Antigen Presentation by CD1 | 0.496 | 0.0385 | #NUM! | B2M |
| Gluconeogenesis I | 0.496 | 0.0385 | #NUM! | ME1 |
| Heparan Sulfate Biosynthesis | 0.492 | 0.0253 | #NUM! | HS3ST5,UST |
| FLT3 Signaling in Hematopoietic Progenitor Cells | 0.485 | 0.025 | #NUM! | PIK3C2A,RASD1 |
| Ovarian Cancer Signaling | 0.478 | 0.0216 | #NUM! | APC,PIK3C2A,RASD1 |
| Hereditary Breast Cancer Signaling | 0.472 | 0.0214 | #NUM! | PIK3C2A,RASD1,UBC |
| PEDF Signaling | 0.471 | 0.0244 | #NUM! | PIK3C2A,RASD1 |
| Apelin Adipocyte Signaling Pathway | 0.471 | 0.0244 | #NUM! | ADCY2,HIF1A |
| D-myo-inositol (1,4,5,6)-Tetrakisphosphate Biosynthe | 0.462 | 0.0211 | #NUM! | DUSP1,PPP3CA,PTPRJ |
| D-myo-inositol (3,4,5,6)-tetrakisphosphate Biosynthe: | 0.462 | 0.0211 | #NUM! | DUSP1,PPP3CA,PTPRJ |
| Role of p14/p19ARF in Tumor Suppression | 0.458 | 0.0345 | #NUM! | PIK3C2A |
| Regulation Of The Epithelial Mesenchymal Transition | 0.457 | 0.0238 | #NUM! | APC,HIF1A |
| Dopamine Degradation | 0.446 | 0.0333 | #NUM! | ALDH1A1 |
| Acute Myeloid Leukemia Signaling | 0.426 | 0.0225 | #NUM! | PIK3C2A,RASD1 |
| 4-1BB Signaling in T Lymphocytes | 0.425 | 0.0312 | #NUM! | JUN |
| AMPK Signaling | 0.421 | 0.0188 | #NUM! | EEF2K,PHLPP1,PIK3C2A,PPP2R2B |
| OX40 Signaling Pathway | 0.42 | 0.0222 | #NUM! | B2M,JUN |
| IL-9 Signaling | 0.413 | 0.0303 | #NUM! | PIK3C2A |
| Prostate Cancer Signaling | 0.413 | 0.022 | #NUM! | PIK3C2A,RASD1 |
| Pyrimidine Ribonucleotides Interconversion | 0.413 | 0.0303 | #NUM! | ENTPD1 |
| Th17 Activation Pathway | 0.413 | 0.022 | #NUM! | HIF1A,NFATC4 |
| Cytotoxic T Lymphocyte-mediated Apoptosis of Targe | 0.403 | 0.0294 | #NUM! | B2M |
| Retinoate Biosynthesis I | 0.403 | 0.0294 | #NUM! | ALDH1A1 |
| 3-phosphoinositide Degradation | 0.398 | 0.0192 | #NUM! | DUSP1,PPP3CA,PTPRJ |
| Coagulation System | 0.395 | 0.0286 | #NUM! | F3 |
| DNA Methylation and Transcriptional Repression Sigr | 0.395 | 0.0286 | #NUM! | H3-3A/H3-3B |
| TWEAK Signaling | 0.395 | 0.0286 | #NUM! | NAIP |
| Pyrimidine Ribonucleotides De Novo Biosynthesis | 0.395 | 0.0286 | #NUM! | ENTPD1 |
| Noradrenaline and Adrenaline Degradation | 0.395 | 0.0286 | #NUM! | ALDH1A1 |
| Bladder Cancer Signaling | 0.381 | 0.0206 | #NUM! | FGFR3,RASD1 |

| | | | | |
|---|---|---|---|---|
| Cell Cycle Regulation by BTG Family Proteins | 0.376 | 0.027 | #NUM! | PPP2R2B |
| Salvage Pathways of Pyrimidine Ribonucleotides | 0.376 | 0.0204 | #NUM! | PRKX,UPP2 |
| Superpathway of Methionine Degradation | 0.376 | 0.027 | #NUM! | AHCYL1 |
| Notch Signaling | 0.376 | 0.027 | #NUM! | HES5 |
| Nitric Oxide Signaling in the Cardiovascular System | 0.371 | 0.0202 | #NUM! | ITPR2,PIK3C2A |
| Docosahexaenoic Acid (DHA) Signaling | 0.368 | 0.0263 | #NUM! | PIK3C2A |
| Thyroid Hormone Metabolism II (via Conjugation and | 0.368 | 0.0263 | #NUM! | DIO2 |
| tRNA Charging | 0.359 | 0.0256 | #NUM! | WARS1 |
| Antigen Presentation Pathway | 0.359 | 0.0256 | #NUM! | B2M |
| Chronic Myeloid Leukemia Signaling | 0.351 | 0.0194 | #NUM! | PIK3C2A,RASD1 |
| Th1 and Th2 Activation Pathway | 0.339 | 0.0175 | #NUM! | JUN,NFATC4,PIK3C2A |
| PD-1, PD-L1 cancer immunotherapy pathway | 0.336 | 0.0189 | #NUM! | B2M,PIK3C2A |
| Estrogen Biosynthesis | 0.335 | 0.0238 | #NUM! | HSD17B12 |
| BAG2 Signaling Pathway | 0.328 | 0.0233 | #NUM! | HSPA1A/HSPA1B |
| Triacylglycerol Biosynthesis | 0.321 | 0.0227 | #NUM! | GPAM |
| Apelin Pancreas Signaling Pathway | 0.321 | 0.0227 | #NUM! | PIK3C2A |
| Role of MAPK Signaling in Promoting the Pathogene | 0.319 | 0.0182 | #NUM! | JUN,RASD1 |
| Acute Phase Response Signaling | 0.308 | 0.0167 | #NUM! | FOS,JUN,RASD1 |
| Role of Oct4 in Mammalian Embryonic Stem Cell Plur | 0.307 | 0.0217 | #NUM! | SPP1 |
| Spliceosomal Cycle | 0.294 | 0.0208 | #NUM! | DDX46 |
| Hepatic Cholestasis | 0.289 | 0.0161 | #NUM! | ADCY2,JUN,SLCO3A1 |
| Amyloid Processing | 0.282 | 0.02 | #NUM! | CAPN2 |
| Th1 Pathway | 0.275 | 0.0165 | #NUM! | NFATC4,PIK3C2A |
| Xenobiotic Metabolism PXR Signaling Pathway | 0.272 | 0.0156 | #NUM! | ALDH1A1,HS3ST5,UST |
| Lymphotoxin β Receptor Signaling | 0.265 | 0.0189 | #NUM! | PIK3C2A |
| Transcriptional Regulatory Network in Embryonic Ster | 0.26 | 0.0185 | #NUM! | H3-3A/H3-3B |
| Role of IL-17A in Arthritis | 0.255 | 0.0182 | #NUM! | PIK3C2A |
| Atherosclerosis Signaling | 0.254 | 0.0157 | #NUM! | APOE,F3 |
| MSP-RON Signaling Pathway | 0.24 | 0.0172 | #NUM! | PIK3C2A |
| Cancer Drug Resistance By Drug Efflux | 0.24 | 0.0172 | #NUM! | RASD1 |
| Semaphorin Signaling in Neurons | 0.231 | 0.0167 | #NUM! | RHOJ |
| Maturity Onset Diabetes of Young (MODY) Signaling | 0.231 | 0.0167 | #NUM! | APOE |
| Retinoic acid Mediated Apoptosis Signaling | 0.231 | 0.0167 | #NUM! | TIPARP |
| Induction of Apoptosis by HIV1 | 0.226 | 0.0164 | #NUM! | NAIP |
| Th2 Pathway | 0.226 | 0.0147 | #NUM! | JUN,PIK3C2A |
| Activation of IRF by Cytosolic Pattern Recognition Re | 0.218 | 0.0159 | #NUM! | JUN |
| Role of PI3K/AKT Signaling in the Pathogenesis of I | 0.214 | 0.0156 | #NUM! | PIK3C2A |
| PXR/RXR Activation | 0.21 | 0.0154 | #NUM! | ALDH1A1 |
| IL-17A Signaling in Airway Cells | 0.21 | 0.0154 | #NUM! | PIK3C2A |
| Mitotic Roles of Polo-Like Kinase | 0.206 | 0.0152 | #NUM! | PPP2R2B |
| Pyridoxal 5'-phosphate Salvage Pathway | 0.206 | 0.0152 | #NUM! | PRKX |
| Serotonin Degradation | 0.202 | 0.0149 | #NUM! | ALDH1A1 |

| | | | | |
|---|---|---|---|---|
| Nur77 Signaling in T Lymphocytes | 0.198 | 0.0147 | #NUM! | PPP3CA |
| LXR/RXR Activation | 0 | 0.00826 | #NUM! | APOE |
| Mitochondrial Dysfunction | 0 | 0.00585 | #NUM! | MT-ND3 |
| VDR/RXR Activation | 0 | 0.0128 | #NUM! | SPP1 |
| FXR/RXR Activation | 0 | 0.00794 | #NUM! | APOE |
| Fcγ Receptor-mediated Phagocytosis in Macrophage | 0 | 0.0106 | #NUM! | VAV3 |
| Role of Pattern Recognition Receptors in Recognitio | 0 | 0.013 | #NUM! | PIK3C2A,PLCG2 |
| TREM1 Signaling | 0 | 0.0133 | #NUM! | PLCG2 |
| Small Cell Lung Cancer Signaling | 0 | 0.0141 | #NUM! | PIK3C2A |
| Allograft Rejection Signaling | 0 | 0.0116 | #NUM! | B2M |
| Type II Diabetes Mellitus Signaling | 0 | 0.00704 | #NUM! | PIK3C2A |
| Pancreatic Adenocarcinoma Signaling | 0 | 0.00917 | #NUM! | PIK3C2A |
| Communication between Innate and Adaptive Immur | 0 | 0.0104 | #NUM! | B2M |
| EIF2 Signaling | 0 | 0.0134 | #NUM! | PIK3C2A,RASD1,WARS1 |
| Altered T Cell and B Cell Signaling in Rheumatoid Ar | 0 | 0.0111 | #NUM! | SPP1 |
| Role of MAPK Signaling in the Pathogenesis of Influ | 0 | 0.0125 | #NUM! | RASD1 |
| Cyclins and Cell Cycle Regulation | 0 | 0.0123 | #NUM! | PPP2R2B |
| Gαs Signaling | 0 | 0.00935 | #NUM! | ADCY2 |
| Agranulocyte Adhesion and Diapedesis | 0 | 0.0104 | #NUM! | CXCL14,HRH1 |
| Granulocyte Adhesion and Diapedesis | 0 | 0.0116 | #NUM! | CXCL14,HRH1 |
| Oxidative Phosphorylation | 0 | 0.00917 | #NUM! | MT-ND3 |
| Insulin Receptor Signaling | 0 | 0.0144 | #NUM! | PIK3C2A,RASD1 |
| Phagosome Maturation | 0 | 0.00662 | #NUM! | B2M |
| NER Pathway | 0 | 0.00971 | #NUM! | H3-3A/H3-3B |
| Xenobiotic Metabolism General Signaling Pathway | 0 | 0.014 | #NUM! | PIK3C2A,RASD1 |
| Xenobiotic Metabolism AHR Signaling Pathway | 0 | 0.0118 | #NUM! | ALDH1A1 |
| Coronavirus Pathogenesis Pathway | 0 | 0.0133 | #NUM! | FOS,JUN |
| Role of MAPK Signaling in Inhibiting the Pathogenes | 0 | 0.0133 | #NUM! | JUN |

**Microglia**

| Ingenuity Canonical Pathways | -log(p-valu | Ratio | z-score | Molecules |
|---|---|---|---|---|
| Synaptogenesis Signaling Pathway | 10.8 | 0.0994 | -3.9 | ADCY2,AKT3,APOE,CACNB4,CDH12,CDH18,CDH23,CHN1,CNTNAP2,DAB1,EPHA6,FGR,FYN,GRIA2,GRIA3,GRIA4,GRIN2A,GRM5,GRM7,HSPA8,KALRN,NLGN1,NLGN4Y,NRXN1,NRXN3,NTRK2,PRKCE,RAPGEF1,SNAP25,SYT1,TIAM1 |
| Synaptic Long Term Depression | 9.9 | 0.122 | -2.711 | GRM7,IGF1,ITPR2,PDIA3,PLCB1,PLCB2,PPP2R2B,PRKCA,PRKCB,PRKCE,PRKG1,RYR2,RYR3 |
| Opioid Signaling Pathway | 9.7 | 0.105 | -1.961 | ADCY2,AKT3,CACNA1A,CACNA1C,CACNA2D3,CACNB2,CACNB4,CAMK1D,FGR,FOS,FYN,GRIN2A,ITPR2,KCNJ5,MAP2K6,PDE1A,PLCB1,PRKCA,PRKCB,PRKCE,RGS16,RGS6,RGS7,RPS6KA5,RYR2,RYR3 |
| Neuropathic Pain Signaling In Dorsal Horn Neurons | 9.69 | 0.168 | -2.183 | CAMK1D,FOS,GRIA2,GRIA3,GRIA4,GRIN2A,GRM5,GRM7,ITPR2,KCNQ3,NTRK2,PDIA3,PLCB1,PLCB2,PRKCA,PRKCB,PRKCE |
| Unfolded protein response | 9.37 | 0.232 | 2.887 | CALR,CEBPB,CEBPD,DNAJC3,HSP90B1,HSPA1A/HSPA1B,HSPA5,HSPA6,HSPA8,HSPH1,PDIA6,PPARG,XBP1 |
| Fcγ Receptor-mediated Phagocytosis in Macrophages a | 9.23 | 0.17 | 1.5 | AKT3,DGKB,FCGR1A,FCGR2A,FCGR3A/FCGR3B,FGR,FYN,HMOX1,INPP5D,LCP2,NCK2,PRKCA,PRKCB,PRKCE,RAB11A,TLN2 |
| Aldosterone Signaling in Epithelial Cells | 9 | 0.127 | -0.707 | ASIC2,DNAJA1,DNAJC3,HSP90AA1,HSP90AB1,HSP90B1,HSPA1A/HSPA1B,HSPA5,HSPA6,HSPA8,HSPB1,HSPD1,HSPH1,ITPR2,PDIA3,PLCB1,PLCB2,PRKCA,PRKCB,PRKCE |
| CREB Signaling in Neurons | 8.5 | 0.0671 | -2.53 | ADCY2,ADGRB3,ADGRE2,ADGRG1,ADGRL3,ADGRV1,AKT3,C3AR1,CACNA1A,CACNA1C,CACNA2D3,CACNB2,CACNB4,CCR1,CHRM3,CMKLR1,CX3CR1,FPR1,GPR183,GPR34,GRIA2,GRIA3,GRIA4,GRID2,GRIK2,GRIN2A,GRM5,GRM7,IGF1,IGF2R,ITPR2,NTRK2,NTRK3,P2RY12,PDIA3,PLCB1,PLCB2,PRKCA,PRKCB,PRKCE |
| Phagosome Formation | 7.8 | 0.128 | #NUM! | C3AR1,FCER1G,FCGR1A,FCGR2A,FCGR3A/FCGR3B,ITGAX,ITGB5,MRC2,MSR1,PDIA3,PLCB1,PLCB2,PRKCA,PRKCB,PRKCE,TLR2,TLR5 |
| Role of PKR in Interferon Induction and Antiviral Respor | 7.75 | 0.136 | -1.291 | DNAJC3,FCGR1A,FOS,HSP90AA1,HSP90AB1,HSP90B1,HSPA1A/HSPA1B,HSPA5,HSPA6,HSPA8,IL1B,JUN,MAP2K6,MAPK10,MSR1,TNFRSF1A |
| Cardiac Hypertrophy Signaling (Enhanced) | 7.51 | 0.0684 | -2.121 | ADCY2,AKT3,CACNA1A,CACNA1C,FGF12,FGF14,HDAC4,HSPB1,IGF1,IL15,IL17RA,IL1B,ITGB5,ITPR2,JUN,MAP2K6,MAPK10,MEF2C,PDE1A,PDE3B,PDE4D,PDE9A,PDIA3,PLCB1,PLCB2,PRKCA,PRKCB,PRKCE,PRKG1,ROCK1,RPS6KA5,RYR2,RYR3,TNFRSF1A |
| Netrin Signaling | 7.49 | 0.185 | -1.732 | ABLIM1,CACNA1A,CACNA1C,CACNA2D3,CACNB2,CACNB4,NCK2,PRKG1,RYR2,RYR3,UNC5C,UNC5D |
| Role of NFAT in Cardiac Hypertrophy | 7.41 | 0.0981 | -2.683 | ADCY2,AKT3,CACNA1A,CACNA1C,CACNA2D3,CACNB2,CACNB4,CAMK1D,HDAC4,IGF1,ITPR2,MAP2K6,MAPK10,MEF2C,PDIA3,PLCB1,PLCB2,PRKCA,PRKCB,PRKCE,SLC8A1 |
| Endocannabinoid Neuronal Synapse Pathway | 7.24 | 0.125 | -2 | ADCY2,CACNA1A,CACNA1C,CACNA2D3,CACNB2,CACNB4,GRIA2,GRIA3,GRIA4,GRIN2A,GRM5,KCNJ5,MAPK10,PDIA3,PLCB1,PLCB2 |
| Dendritic Cell Maturation | 7.12 | 0.103 | 2.183 | AKT3,B2M,FCER1G,FCGR1A,FCGR2A,FCGR3A/FCGR3B,HLA-B,HLA-DRA,HLA-DRB1,HLA-DRB5,IL15,IL1B,IRF8,MAPK10,PDIA3,PLCB1,PLCB2,TLR2,TNFRSF1A |
| Nitric Oxide Signaling in the Cardiovascular System | 7.1 | 0.141 | -0.277 | AKT3,CACNA1A,CACNA1C,HSP90AA1,HSP90AB1,HSP90B1,ITPR2,PDE1A,PDE3B,PRKCA,PRKCB,PRKCE,PRKG1,RYR2 |
| HIF1α Signaling | 7.05 | 0.0976 | -0.447 | AKT3,CAMK1D,CDKN1A,HK2,HMOX1,HSP90AA1,HSPA1A/HSPA1B,HSPA5,HSPA6,HSPA8,IGF1,JUN,LDHA,MAP2K6,MMP16,PRKCA,PRKCB,PRKCE,SAT1,SLC2A3 |
| Role of Macrophages, Fibroblasts and Endothelial Cells | 6.32 | 0.0764 | #NUM! | AKT3,CEBPB,CEBPD,DAAM1,FCGR1A,FCGR3A/FCGR3B,FOS,IL15,IL17RA,IL1B,IL1RAPL1,IRAK3,JUN,MAP2K6,PDIA3,PLCB1,PLCB2,PRKCA,PRKCB,PRKCE,ROCK1,TLR2,TLR5,TNFRSF1A |

| | | | | |
|---|---|---|---|---|
| IL-12 Signaling and Production in Macrophages | 6.24 | 0.113 | #NUM! | AKT3,APOC1,APOE,CEBPB,CLU,FOS,IRF8,JUN,MAPK10,PPARG,PRKCA,PRKCB,PRKCE,SERPINA1,TLR2 |
| GNRH Signaling | 6.13 | 0.0983 | -1.155 | ADCY2,CACNA1A,CACNA1C,CACNA2D3,CACNB2,CACNB4,FOS,ITPR2,JUN,MAP2K6,MAPK10,PAK3,PLCB1,PLCB2,PRKCA,PRKCB,PRKCE |
| Glutamate Receptor Signaling | 6.12 | 0.175 | -2.646 | GRIA2,GRIA3,GRIA4,GRID2,GRIK2,GRIN2A,GRIP1,GRM5,GRM7,SLC1A2 |
| Cholecystokinin/Gastrin-mediated Signaling | 6.09 | 0.118 | -0.535 | CREM,FOS,IL1B,ITPR2,JUN,MAP2K6,MAPK10,MEF2C,PLCB1,PLCB2,PRKCA,PRKCB,PRKCE,ROCK1 |
| Role of NFAT in Regulation of the Immune Response | 5.85 | 0.0939 | 1.5 | AKT3,FCER1G,FCGR1A,FCGR2A,FCGR3A/FCGR3B,FOS,FYN,HLA-B,HLA-DRA,HLA-DRB1,HLA-DRB5,ITPR2,JUN,LCP2,MEF2C,PLCB1,PLCB2 |
| Insulin Secretion Signaling Pathway | 5.84 | 0.082 | -1.342 | ADCY2,CACNA1A,CACNA1C,CHRM3,CPE,EIF1,FGR,FYN,ITPR2,PDIA3,PLCB1,PLCB2,PRKCA,PRKCB,PRKCE,RPS6KA5,RYR2,RYR3,SLC2A3,SNAP25 |
| CCR5 Signaling in Macrophages | 5.68 | 0.128 | 0.816 | CACNA1A,CACNA1C,CACNA2D3,CACNB2,CACNB4,FCER1G,FOS,JUN,MAPK10,PRKCA,PRKCB,PRKCE |
| Synaptic Long Term Potentiation | 5.66 | 0.109 | -1.941 | CACNA1C,GRIA2,GRIA3,GRIA4,GRIN2A,GRM5,GRM7,ITPR2,PDIA3,PLCB1,PLCB2,PRKCA,PRKCB,PRKCE |
| GPCR-Mediated Nutrient Sensing in Enteroendocrine C | 5.63 | 0.116 | -1.387 | ADCY2,CACNA1A,CACNA1C,CACNA2D3,CACNB2,CACNB4,ITPR2,PDIA3,PLCB1,PLCB2,PRKCA,PRKCB,PRKCE |
| Antigen Presentation Pathway | 5.55 | 0.205 | #NUM! | B2M,CALR,HLA-B,HLA-DPA1,HLA-DRA,HLA-DRB1,HLA-DRB5,PDIA3 |
| nNOS Signaling in Skeletal Muscle Cells | 5.37 | 0.195 | #NUM! | CACNA1A,CACNA1C,CACNA2D3,CACNB2,CACNB4,DMD,RYR2,RYR3 |
| Cdc42 Signaling | 5.36 | 0.0909 | 0.707 | B2M,CDC42BPA,FCER1G,FOS,HLA-B,HLA-DPA1,HLA-DRA,HLA-DRB1,HLA-DRB5,IQGAP1,IQGAP2,ITGB5,JUN,MAPK10,PAK3,PARD3 |
| eNOS Signaling | 5.26 | 0.0943 | -2.111 | ADCY2,AKT3,CHRM3,HSP90AA1,HSP90AB1,HSP90B1,HSPA1A/HSPA1B,HSPA5,HSPA6,HSPA8,ITPR2,PRKCA,PRKCB,PRKCE,PRKG1 |
| Sperm Motility | 5.22 | 0.0807 | -1.265 | CSF2RA,EPHA6,ERBB4,FGR,FYN,ITPR2,MAP2K6,NTRK2,NTRK3,PDE1A,PDE4D,PDIA3,PLCB1,PLCB2,PRKCA,PRKCB,PRKCE,PRKG1 |
| Type II Diabetes Mellitus Signaling | 5.17 | 0.0986 | -0.707 | ACSL1,AKT3,CACNA1A,CACNA1C,CACNA2D3,CACNB2,CACNB4,CEBPB,MAPK10,PPARG,PRKCA,PRKCB,PRKCE,TNFRSF1A |
| Axonal Guidance Signaling | 5.16 | 0.0587 | #NUM! | ABLIM1,ADAM28,AKT3,EPHA6,FYN,IGF1,ITGB5,KALRN,LRRC4C,MMP16,NCK2,NRP1,NTRK2,NTRK3,PAK3,PDIA3,PLCB1,PLCB2,PLXNA2,PLXNC1,PRKCA,PRKCB,PRKCE,ROBO2,ROCK1,TUBA1A,TUBA1B,UNC5C,UNC5D |
| Dopamine-DARPP32 Feedback in cAMP Signaling | 5.13 | 0.092 | -1.387 | ADCY2,CACNA1A,CACNA1C,CREM,GRIN2A,ITPR2,KCNJ5,PDIA3,PLCB1,PLCB2,PPP2R2B,PRKCA,PRKCB,PRKCE,PRKG1 |
| Calcium Signaling | 5.09 | 0.0825 | -3.357 | CACNA1A,CACNA1C,CACNA2D3,CACNB2,CACNB4,CALR,CAMK1D,GRIA2,GRIA3,GRIA4,GRIN2A,HDAC4,ITPR2,MEF2C,RYR2,RYR3,SLC8A1 |
| 14-3-3-mediated Signaling | 5.02 | 0.102 | 0.302 | AKT3,FOS,JUN,MAPK10,PDIA3,PLCB1,PLCB2,PRKCA,PRKCB,PRKCE,TNFRSF1A,TUBA1A,TUBA1B |
| Neuroinflammation Signaling Pathway | 5.02 | 0.07 | 2.357 | AKT3,B2M,CX3CR1,FOS,GABRB1,GABRB3,GRIN2A,HLA-B,HLA-DRA,HLA-DRB1,HLA-DRB5,HMOX1,IL1B,IRAK3,JUN,KCNJ5,MAPK10,SLC1A2,TLR2,TLR5,TNFRSF1A |
| Estrogen Receptor Signaling | 4.93 | 0.0671 | -0.229 | ,IGF2R,JUN,MMP16,MT-ATP6,MT-CYB,PDIA3,PLCB1,PLCB2,PRKCA,PRKCB,PRKCE,ROCK1 |
| Type I Diabetes Mellitus Signaling | 4.92 | 0.108 | 0.447 | CPE,FCER1G,HLA-B,HLA-DRA,HLA-DRB1,HLA-DRB5,HSPD1,ICA1,IL1B,MAP2K6,MAPK10,TNFRSF1A |
| ErbB Signaling | 4.89 | 0.117 | -0.302 | ERBB4,FOS,JUN,MAP2K6,MAPK10,NCK2,NRG3,PAK3,PRKCA,PRKCB,PRKCE |

| | | | | |
|---|---|---|---|---|
| Cellular Effects of Sildenafil (Viagra) | 4.88 | 0.0992 | #NUM! | ADCY2,CACNA1A,CACNA1C,ITPR2,KCNQ3,PDE1A,PDE3B,PDE4D,PDIA3,PLCB1,PLCB2,PRKG1,SLC4A10 |
| GABA Receptor Signaling | 4.84 | 0.116 | #NUM! | ADCY2,CACNA1A,CACNA1C,CACNA2D3,CACNB2,CACNB4,DNM1,GABRB1,GABRB3,KCNQ3,UBC |
| Apelin Endothelial Signaling Pathway | 4.76 | 0.104 | -0.577 | ADCY2,AKT3,FOS,HDAC4,JUN,MAPK10,MEF2C,PLCB1,PLCB2,PRKCA,PRKCB,PRKCE |
| Amyotrophic Lateral Sclerosis Signaling | 4.76 | 0.113 | -1.414 | AKT3,CACNA1A,CACNA1C,GRIA2,GRIA3,GRIA4,GRID2,GRIK2,GRIN2A,IGF1,SLC1A2 |
| Natural Killer Cell Signaling | 4.74 | 0.0812 | 1 | AKT3,B2M,FCER1G,FCGR2A,FCGR3A/FCGR3B,FYN,HLA-B,HSPA1A/HSPA1B,HSPA5,HSPA6,HSPA8,IL15,LAIR1,LCP2,PAK3,ROCK1 |
| PKCθ Signaling in T Lymphocytes | 4.74 | 0.0903 | 2.333 | CACNA1A,CACNA1C,CACNA2D3,CACNB2,CACNB4,FCER1G,FOS,FYN,HLA-B,HLA-DRA,HLA-DRB1,HLA-DRB5,JUN,LCP2 |
| Gustation Pathway | 4.74 | 0.0903 | #NUM! | ADCY2,ASIC2,CACNA1A,CACNA1C,CACNA2D3,CACNB2,CACNB4,ITPR2,P2RY12,PDE1A,PDE3B,PDE4D,PDE9A,PLCB2 |
| Gap Junction Signaling | 4.71 | 0.0808 | #NUM! | ADCY2,AKT3,GRIA2,GRIA3,GRIA4,GRIK2,ITPR2,PDIA3,PLCB1,PLCB2,PRKCA,PRKCB,PRKCE,PRKG1,TUBA1A,TUBA1B |
| Calcium-induced T Lymphocyte Apoptosis | 4.64 | 0.136 | 1.414 | FCER1G,HLA-B,HLA-DRA,HLA-DRB1,HLA-DRB5,ITPR2,PRKCA,PRKCB,PRKCE |
| UVB-Induced MAPK Signaling | 4.57 | 0.154 | 0.378 | FOS,H3-3A/H3-3B,JUN,MAPK10,PRKCA,PRKCB,PRKCE,RPS6KA5 |
| CD28 Signaling in T Helper Cells | 4.54 | 0.0992 | -0.378 | DRB5,ITPR2,JUN,LCP2,MAPK10 |
| G-Protein Coupled Receptor Signaling | 4.54 | 0.0693 | #NUM! | ADCY2,AKT3,CHRM3,FPR1,FYN,GRM5,GRM7,P2RY12,PDE1A,PDE3B,PDE4D,PDE9A,PLCB1,PLCB2,PRKCA,PRKCB,PRKCE,RGS16,RGS7 |
| Aryl Hydrocarbon Receptor Signaling | 4.47 | 0.0909 | -0.302 | CDK6,CDKN1A,DHFR,FOS,GSTK1,HSP90AA1,HSP90AB1,HSP90B1,HSPB1,IL1B,JUN,NFIB,PTGES3 |
| IL-10 Signaling | 4.43 | 0.129 | #NUM! | CCR1,CD14,FCGR2A,FOS,HMOX1,IL1B,IL1RAPL1,JUN,MAP2K6 |
| Corticotropin Releasing Hormone Signaling | 4.4 | 0.0897 | 0.905 | ADCY2,CACNA1A,CACNA1C,CACNA2D3,CACNB2,CACNB4,FOS,ITPR2,JUN,MEF2C,PRKCA,PRKCB,PRKCE |
| NRF2-mediated Oxidative Stress Response | 4.37 | 0.0794 | 0.905 | ABCC4,DNAJA1,DNAJC3,FKBP5,FOS,GSTK1,HERPUD1,HMOX1,JUN,JUNB,MAFG,MAP2K6,PRKCA,PRKCB,PRKCE |
| Production of Nitric Oxide and Reactive Oxygen Species | 4.37 | 0.0794 | 1.807 | AKT3,APOC1,APOE,CLU,FOS,IRF8,JUN,MAPK10,PPP2R2B,PRKCA,PRKCB,PRKCE,SERPINA1,TLR2,TNFRSF1A |
| PPARα/RXRα Activation | 4.31 | 0.0785 | -1.732 | ADCY2,HSP90AA1,HSP90AB1,HSP90B1,IL1B,IL1RAPL1,ITGB5,JUN,MAP2K6,MEF2C,PDIA3,PLCB1,PLCB2,PRKCA,PRKCB |
| Altered T Cell and B Cell Signaling in Rheumatoid Arthrit | 4.3 | 0.111 | #NUM! | FCER1G,HLA-B,HLA-DRA,HLA-DRB1,HLA-DRB5,IL15,IL1B,SPP1,TLR2,TLR5 |
| Huntington's Disease Signaling | 4.26 | 0.0711 | -0.632 | AKT3,DNM1,GRM5,HDAC4,HSPA1A/HSPA1B,HSPA5,HSPA6,HSPA8,IGF1,JUN,PLCB1,PLCB2,PRKCA,PRKCB,PRKCE,SNAP25,UBC |
| Toll-like Receptor Signaling | 4.14 | 0.118 | 1.89 | CD14,FOS,IL1B,IRAK3,JUN,MAP2K6,TLR2,TLR5,UBC |
| Protein Kinase A Signaling | 4.08 | 0.0575 | -1.279 | ADCY2,AKAP6,CREM,EYA2,H3-3A/H3-3B,ITPR2,PDE1A,PDE3B,PDE4D,PDE9A,PDIA3,PLCB1,PLCB2,PRKCA,PRKCB,PRKCE,PTPRD,PTPRG,PTPRZ1,PYGL,ROCK1,RYR2,RYR3 |
| Communication between Innate and Adaptive Immune ( | 4.06 | 0.104 | #NUM! | B2M,FCER1G,HLA-B,HLA-DRA,HLA-DRB1,HLA-DRB5,IL15,IL1B,TLR2,TLR5 |
| Protein Ubiquitination Pathway | 4.05 | 0.0659 | #NUM! | B2M,DNAJA1,DNAJC3,HLA-B,HSP90AA1,HSP90AB1,HSP90B1,HSPA1A/HSPA1B,HSPA5,HSPA6,HSPA8,HSPB1,HSPD1,HSPH1,PSMA1,UBC,UBE2J1,USP15 |
| Gαq Signaling | 4.05 | 0.0828 | -0.832 | AKT3,CHRM3,GRM5,HMOX1,ITPR2,PLCB1,PLCB2,PRKCA,PRKCB,PRKCE,RGS16,RGS7,ROCK1 |
| Endoplasmic Reticulum Stress Pathway | 3.98 | 0.238 | 1.342 | CALR,DNAJC3,HSP90B1,HSPA5,XBP1 |

| | | | | |
|---|---|---|---|---|
| Glucocorticoid Receptor Signaling | 3.97 | 0.0541 | #NUM! | AKT3,CD163,CDKN1A,FCGR1A,FKBP5,FOS,HSP90AA1,HSP90AB1,HSP90B1,HSPA1A/HSPA1B,HSPA5,HSPA6,HSPA8,IGF1,IL17RA,IL1B,JUN,MAPK10,MT-ATP6,MT-CYB,PDK4,PPARG,PTGES3,RPS6KA5,TLR2 |
| G Beta Gamma Signaling | 3.84 | 0.0902 | -0.905 | ADCY2,AKT3,CACNA1A,CACNA1C,CACNA2D3,CACNB2,CACNB4,KCNJ5,PRKCA,PRKCB,PRKCE |
| CXCR4 Signaling | 3.78 | 0.0778 | -0.577 | CK1 |
| Breast Cancer Regulation by Stathmin1 | 3.78 | 0.0492 | -0.928 | ADGRB3,ADGRE2,ADGRG1,ADGRL3,ADGRV1,C3AR1,CAMK1D,CCR1,CDK6,CHRM3,CMKLR1,CX3CR1,E2F3,FPR1,GPR183,GPR34,GRM5,GRM7,IGF1,JUN,P2RY12,PLCB1,PLCB2,PPP2R2B,PRKCA,PRKCB,PRKCE,TUBA1A,TUBA1B |
| Xenobiotic Metabolism Signaling | 3.78 | 0.0627 | #NUM! | CAMK1D,CHST15,GRIP1,GSTK1,HDAC4,HMOX1,HS6ST3,HSP90AA1,HSP90AB1,HSP90B1,IL1B,MAP2K6,PPP2R2B,PRKCA,PRKCB,PRKCE,PTGES3,UST |
| Neuregulin Signaling | 3.74 | 0.0952 | -0.816 | AKT3,ERBB4,HSP90AA1,HSP90AB1,HSP90B1,ITGB5,NRG3,PRKCA,PRKCB,PRKCE |
| IL-6 Signaling | 3.71 | 0.0873 | 0.632 | AKT3,CD14,CEBPB,FOS,HSPB1,IL1B,IL1RAPL1,JUN,MAP2K6,MAPK10,TNFRSF1A |
| Clathrin-mediated Endocytosis Signaling | 3.7 | 0.0725 | #NUM! | APOC1,APOE,CLU,DNM1,FGF12,FGF14,GAK,HSPA8,IGF1,ITGB5,MYO1E,RAB11A,SERPINA1,UBC |
| P2Y Purigenic Receptor Signaling Pathway | 3.68 | 0.0866 | 0.302 | ADCY2,AKT3,FOS,JUN,P2RY12,PDIA3,PLCB1,PLCB2,PRKCA,PRKCB,PRKCE |
| HER-2 Signaling in Breast Cancer | 3.68 | 0.0722 | 0 | AKT3,CDKN1A,FCER1G,FGR,FOS,FYN,ITGB5,JUN,MT-CO2,MT-CO3,PARD3,PRKCA,PRKCB,PRKCE |
| White Adipose Tissue Browning Pathway | 3.62 | 0.0853 | -0.302 | ADCY2,CACNA1A,CACNA1C,CACNA2D3,CACNB2,CACNB4,CEBPB,FCER1G,LDHA,PPARG,PRKG1 |
| Adrenomedullin signaling pathway | 3.61 | 0.0711 | -1.069 | ADCY2,AKT3,CEBPB,FOS,IL1B,ITPR2,KCNQ3,MAP2K6,MAPK10,PDIA3,PLCB1,PLCB2,PPARG,PRKG1 |
| T Cell Exhaustion Signaling Pathway | 3.58 | 0.0743 | 0 | AKT3,BCL6,FCER1G,FOS,FOXP1,HLA-B,HLA-DPA1,HLA-DRA,HLA-DRB1,HLA-DRB5,JUN,MAPK10,PPP2R2B |
| OX40 Signaling Pathway | 3.58 | 0.1 | #NUM! | B2M,FCER1G,HLA-B,HLA-DPA1,HLA-DRA,HLA-DRB1,HLA-DRB5,JUN,MAPK10 |
| Glioma Signaling | 3.57 | 0.0909 | -0.378 | AKT3,CAMK1D,CDK6,CDKN1A,E2F3,IGF1,IGF2R,PRKCA,PRKCB,PRKCE |
| Tumor Microenvironment Pathway | 3.55 | 0.0739 | -0.277 | B,IGF1,IL1B,JUN,MMP16,SLC2A3,SPP1,TIAM1,TNFRSF1A |
| Melatonin Signaling | 3.55 | 0.111 | -0.707 | MAP2K6,PDIA3,PLCB1,PLCB2,PRKCA,PRKCB,PRKCE,RORA |
| Role of Pattern Recognition Receptors in Recognition of | 3.53 | 0.0779 | 1.897 | C1QA,C1QB,C1QC,C3AR1,IL15,IL1B,MAPK10,PRKCA,PRKCB,PRKCE,TLR2,TLR5 |
| ERK/MAPK Signaling | 3.5 | 0.0693 | 0 | 3B,HSPB1,ITGB5,PAK3,PPARG,PPP2R2B,PRKCA,PRKCB,PRKCE,RAPGEF1,RPS6KA5,TLN2 |
| Androgen Signaling | 3.42 | 0.0809 | -0.632 | CACNA1A,CACNA1C,CACNA2D3,CACNB2,CACNB4,CALR,HSP90AA1,JUN,PRKCA,PRKCB,PRKCE |
| PI3K Signaling in B Lymphocytes | 3.37 | 0.0797 | 0 | AKT3,CD81,FOS,FYN,INPP5D,ITPR2,JUN,PDIA3,PLCB1,PLCB2,PRKCB |
| Fc Epsilon RI Signaling | 3.35 | 0.0855 | 0 | AKT3,FCER1G,FYN,INPP5D,LCP2,MAP2K6,MAPK10,PRKCA,PRKCB,PRKCE |
| BAG2 Signaling Pathway | 3.32 | 0.14 | 2.449 | CDKN1A,HSP90AA1,HSPA1A/HSPA1B,HSPA5,HSPA6,HSPA8 |
| IL-17 Signaling | 3.3 | 0.0695 | 1.941 | AKT3,CEBPB,FOS,HSP90AA1,HSP90AB1,HSP90B1,IL15,IL17RA,IL1B,JUN,MAP2K6,MAPK10,RGS16 |
| IL-3 Signaling | 3.27 | 0.101 | 0.707 | AKT3,FOS,INPP5D,JUN,PRKCA,PRKCB,PRKCE,RAPGEF1 |
| BEX2 Signaling Pathway | 3.27 | 0.101 | 0 | AKT3,CDKN1A,JUN,MAP2,MAPK10,PLXNC1,PPP2R2B,SPP1 |
| Apelin Cardiomyocyte Signaling Pathway | 3.27 | 0.0909 | -1 | AKT3,MAPK10,PDIA3,PLCB1,PLCB2,PRKCA,PRKCB,PRKCE,SLC8A1 |
| HGF Signaling | 3.27 | 0.0833 | -0.333 | AKT3,CDKN1A,FOS,ITGB5,JUN,MAPK10,PRKCA,PRKCB,PRKCE,RAPGEF1 |
| Cardiac β-adrenergic Signaling | 3.27 | 0.0775 | -1.414 | C8A1 |

| | | | | |
|---|---|---|---|---|
| Xenobiotic Metabolism CAR Signaling Pathway | 3.26 | 0.0688 | 0.277 | CHST15,GRIP1,GSTK1,HS6ST3,HSP90AA1,HSP90AB1,HSP90B1,MAP2K6,PPP2R2B,PRKCA,PRKCB,PRKCE,UST |
| LXR/RXR Activation | 3.24 | 0.0826 | -0.333 | APOC1,APOE,CD14,CLU,IL1B,IL1RAPL1,MSR1,PLTP,SERPINA1,TNFRSF1A |
| Reelin Signaling in Neurons | 3.21 | 0.082 | -1.667 | AKT3,APOE,DAB1,FGR,FYN,GRIN2A,MAP2K6,MAPK10,PDK4,RAPGEF1 |
| Virus Entry via Endocytic Pathways | 3.17 | 0.0882 | #NUM! | B2M,CXADR,DNM1,FYN,HLA-B,ITGB5,PRKCA,PRKCB,PRKCE |
| Hepatic Fibrosis Signaling Pathway | 3.17 | 0.0529 | 1.147 | AKT3,CACNA1A,CACNA1C,CEBPB,FOS,IL17RA,IL1B,IL1RAPL1,IRAK3,ITGB5,JUN,MAP2K6,MAPK10,PPARG,PRKCA,PRKCB,PRKCE,ROCK1,SPP1,TNFRSF1A |
| LPS-stimulated MAPK Signaling | 3.13 | 0.0964 | 0.707 | CD14,FOS,JUN,MAP2K6,MAPK10,PRKCA,PRKCB,PRKCE |
| Osteoarthritis Pathway | 3.13 | 0.0636 | 0 | CEBPB,CTNNA2,DDIT4,HDAC4,HTRA1,IL1B,IL1RAPL1,ITGB5,MEF2C,PPARG,RBPJ,SPP1,TLR2,TNFRSF1A |
| nNOS Signaling in Neurons | 3.11 | 0.128 | #NUM! | DLG2,GRIN2A,NOS1AP,PRKCA,PRKCB,PRKCE |
| PPAR Signaling | 3.08 | 0.0857 | -1.667 | FOS,HSP90AA1,HSP90AB1,HSP90B1,IL1B,IL1RAPL1,JUN,PPARG,TNFRSF1A |
| Graft-versus-Host Disease Signaling | 3.06 | 0.125 | #NUM! | FCER1G,HLA-B,HLA-DRA,HLA-DRB1,HLA-DRB5,IL1B |
| PD-1, PD-L1 cancer immunotherapy pathway | 3.05 | 0.0849 | -1.414 | AKT3,B2M,HLA-B,HLA-DPA1,HLA-DRA,HLA-DRB1,HLA-DRB5,LCP2,TNFRSF1A |
| LPS/IL-1 Mediated Inhibition of RXR Function | 3.03 | 0.0622 | 1.414 | ABCC4,ACSL1,APOC1,APOE,CD14,CHST15,GSTK1,HS6ST3,IL1B,IL1RAPL1,JUN,PLTP,TNFRSF1A,UST |
| IL-8 Signaling | 3.03 | 0.065 | 0.832 | AKT3,FOS,HMOX1,IQGAP1,IRAK3,ITGAX,JUN,MAPK10,PLCB2,PRKCA,PRKCB,PRKCE,ROCK1 |
| NF-κB Signaling | 2.95 | 0.067 | 0.577 | A |
| UVC-Induced MAPK Signaling | 2.92 | 0.118 | 0.816 | FOS,JUN,MAPK10,PRKCA,PRKCB,PRKCE |
| iCOS-iCOSL Signaling in T Helper Cells | 2.91 | 0.0811 | 1.89 | AKT3,FCER1G,HLA-B,HLA-DRA,HLA-DRB1,HLA-DRB5,INPP5D,ITPR2,LCP2 |
| Molecular Mechanisms of Cancer | 2.87 | 0.05 | #NUM! | ADCY2,AKT3,CDK6,CDKN1A,CTNNA2,E2F3,FOS,FYN,ITGB5,JUN,MAP2K6,MAPK10,PAK3,PLCB1,PLCB2,PRKCA,PRKCB,PRKCE,RAPGEF1,RBPJ |
| PTEN Signaling | 2.84 | 0.0735 | 1.667 | AKT3,CDKN1A,IGF2R,INPP5D,ITGB5,MAGI1,MAGI2,NTRK2,NTRK3,PREX2 |
| IL-17A Signaling in Fibroblasts | 2.84 | 0.139 | #NUM! | CEBPB,CEBPD,FOS,IL17RA,JUN |
| Hepatic Cholestasis | 2.81 | 0.0645 | #NUM! | 1A |
| T Helper Cell Differentiation | 2.79 | 0.0959 | #NUM! | BCL6,FCER1G,HLA-B,HLA-DRA,HLA-DRB1,HLA-DRB5,TNFRSF1A |
| Complement System | 2.79 | 0.135 | #NUM! | C1QA,C1QB,C1QC,C3AR1,ITGAX |
| Phospholipase C Signaling | 2.78 | 0.0564 | 0 | ADCY2,FCER1G,FCGR2A,FYN,HDAC4,HMOX1,ITGB5,ITPR2,LCP2,MEF2C,PLCB1,PLCB2,PRKCA,PRKCB,PRKCE |
| Semaphorin Neuronal Repulsive Signaling Pathway | 2.77 | 0.0719 | 0 | AKT3,FYN,ITGB5,MAP2K6,NRP1,PAK3,PDE4D,PLXNA2,PRKG1,ROCK1 |
| Endothelin-1 Signaling | 2.77 | 0.0638 | 0.577 | ADCY2,FOS,HMOX1,ITPR2,JUN,MAPK10,PDIA3,PLCB1,PLCB2,PRKCA,PRKCB,PRKCE |
| Renin-Angiotensin Signaling | 2.72 | 0.0763 | 0 | ADCY2,FOS,ITPR2,JUN,MAPK10,PAK3,PRKCA,PRKCB,PRKCE |
| p38 MAPK Signaling | 2.72 | 0.0763 | 0.333 | H3-3A/H3-3B,HSPB1,IL1B,IL1RAPL1,IRAK3,MAP2K6,MEF2C,RPS6KA5,TNFRSF1A |
| FcγRIIB Signaling in B Lymphocytes | 2.72 | 0.0933 | #NUM! | CACNA1A,CACNA1C,CACNA2D3,CACNB2,CACNB4,INPP5D,MAPK10 |
| UVA-Induced MAPK Signaling | 2.66 | 0.0816 | -1.342 | FOS,JUN,MAPK10,PDIA3,PLCB1,PLCB2,PRKCA,RPS6KA5 |
| IL-15 Production | 2.65 | 0.0744 | -1.667 | CSF2RA,EPHA6,ERBB4,FGR,FYN,IL15,MAP2K6,NTRK2,NTRK3 |
| VDR/RXR Activation | 2.62 | 0.0897 | 0 | CD14,CDKN1A,CEBPB,PRKCA,PRKCB,PRKCE,SPP1 |
| Chemokine Signaling | 2.56 | 0.0875 | -0.378 | CAMK1D,FOS,JUN,PLCB1,PLCB2,PRKCA,PRKCB |
| Actin Cytoskeleton Signaling | 2.54 | 0.0573 | -1.89 | ARHGAP24,CD14,CYFIP1,FGF12,FGF14,GSN,IQGAP1,IQGAP2,ITGB5,PAK3,ROCK1,TIAM1,TLN2 |
| Prolactin Signaling | 2.53 | 0.0864 | 0.816 | CEBPB,FOS,FYN,JUN,PRKCA,PRKCB,PRKCE |
| FXR/RXR Activation | 2.53 | 0.0714 | #NUM! | AKT3,APOC1,APOE,CLU,IL1B,MAPK10,PLTP,PPARG,SERPINA1 |
| Lipid Antigen Presentation by CD1 | 2.52 | 0.154 | #NUM! | B2M,CALR,FCER1G,PDIA3 |

| | | | | |
|---|---|---|---|---|
| IL-17A Signaling in Gastric Cells | 2.52 | 0.154 | 1 | FOS,IL17RA,JUN,MAPK10 |
| Phagosome Maturation | 2.51 | 0.0662 | #NUM! | B2M,CALR,CTSC,HLA-B,HLA-DRA,HLA-DRB1,HLA-DRB5,SNAP25,TUBA1A,TUBA1B |
| cAMP-mediated signaling | 2.51 | 0.0568 | -1.732 | ADCY2,AKAP6,CAMK1D,CHRM3,CREM,FPR1,GRM7,P2RY12,PDE1A,PDE3B,PDE4D,PDE9A,RGS7 |
| Epithelial Adherens Junction Signaling | 2.49 | 0.0658 | #NUM! | AKT3,CTNNA2,IQGAP1,MAGI1,MAGI2,PARD3,RAPGEF1,SORBS1,TUBA1A,TUBA1B |
| Telomerase Signaling | 2.42 | 0.0748 | 1.342 | AKT3,CDKN1A,HDAC4,HSP90AA1,HSP90AB1,HSP90B1,PPP2R2B,PTGES3 |
| Xenobiotic Metabolism AHR Signaling Pathway | 2.41 | 0.0824 | -1.134 | GSTK1,HDAC4,HSP90AA1,HSP90AB1,HSP90B1,IL1B,PTGES3 |
| Allograft Rejection Signaling | 2.38 | 0.0814 | #NUM! | B2M,FCER1G,HLA-B,HLA-DPA1,HLA-DRA,HLA-DRB1,HLA-DRB5 |
| Antioxidant Action of Vitamin C | 2.38 | 0.0734 | 0.816 | CSF2RA,HMOX1,MAPK10,NXN,PDIA3,PLCB1,PLCB2,SLC2A3 |
| Oxidative Phosphorylation | 2.38 | 0.0734 | 2.828 | MT-ATP6,MT-CO2,MT-CO3,MT-CYB,MT-ND1,MT-ND3,MT-ND4,VPS9D1 |
| Cardiac Hypertrophy Signaling | 2.33 | 0.0542 | -0.832 | ADCY2,CACNA1A,CACNA1C,HSPB1,IGF1,JUN,MAP2K6,MAPK10,MEF2C,PDIA3,PLCB1,PLCB2,ROCK1 |
| CTLA4 Signaling in Cytotoxic T Lymphocytes | 2.3 | 0.0787 | #NUM! | AKT3,B2M,FCER1G,FYN,HLA-B,LCP2,PPP2R2B |
| RANK Signaling in Osteoclasts | 2.3 | 0.0787 | 0.378 | AKT3,FOS,GSN,JUN,MAP2K6,MAPK10,MITF |
| Chondroitin Sulfate Biosynthesis (Late Stages) | 2.28 | 0.104 | 0.447 | CHST15,CHSY3,CSGALNACT1,HS6ST3,UST |
| Ephrin Receptor Signaling | 2.28 | 0.0582 | -1.667 | AKT3,EPHA6,FYN,GRIN2A,ITGB5,KALRN,NCK2,PAK3,RAPGEF1,ROCK1,SORBS1 |
| Th17 Activation Pathway | 2.25 | 0.0769 | -0.378 | FCER1G,HSP90AA1,HSP90AB1,HSP90B1,IL1B,IRAK3,RORA |
| Autoimmune Thyroid Disease Signaling | 2.24 | 0.102 | #NUM! | FCER1G,HLA-B,HLA-DRA,HLA-DRB1,HLA-DRB5 |
| Senescence Pathway | 2.24 | 0.0509 | -0.535 | AKT3,CACNA1A,CACNA1C,CDK6,CDKN1A,CEBPB,E2F3,ITPR2,JUN,MAP2K6,PDK4,PPP2R2B,RPS6KA5,TLR2 |
| Xenobiotic Metabolism PXR Signaling Pathway | 2.23 | 0.0573 | -1.508 | CHST15,GRIP1,GSTK1,HS6ST3,HSP90AA1,HSP90AB1,HSP90B1,PRKCA,PRKCB,PRKCE,UST |
| Role of Tissue Factor in Cancer | 2.21 | 0.069 | #NUM! | AKT3,FGR,FYN,IL1B,PDIA6,PLCB1,PRKCA,RPS6KA5 |
| Sertoli Cell-Sertoli Cell Junction Signaling | 2.2 | 0.0567 | #NUM! | B |
| Growth Hormone Signaling | 2.19 | 0.0845 | 0.447 | FOS,IGF1,PRKCA,PRKCB,PRKCE,RPS6KA5 |
| α-Adrenergic Signaling | 2.15 | 0.0737 | 0 | ADCY2,ITPR2,PRKCA,PRKCB,PRKCE,PYGL,SLC8A1 |
| Mitochondrial Dysfunction | 2.13 | 0.0585 | #NUM! | LRRK2,MAPK10,MT-ATP6,MT-CO2,MT-CO3,MT-CYB,MT-ND1,MT-ND3,MT-ND4,VPS9D1 |
| Germ Cell-Sertoli Cell Junction Signaling | 2.13 | 0.0585 | #NUM! | CTNNA2,GSN,IQGAP1,MAP2K6,MAPK10,PAK3,SORBS1,TNFRSF1A,TUBA1A,TUBA1B |
| Caveolar-mediated Endocytosis Signaling | 2.13 | 0.0822 | #NUM! | B2M,FYN,HLA-B,ITGAX,ITGB5,PRKCA |
| Chondroitin and Dermatan Biosynthesis | 2.11 | 0.333 | #NUM! | CHSY3,CSGALNACT1 |
| Ceramide Biosynthesis | 2.11 | 0.333 | #NUM! | SPTLC2,SPTLC3 |
| Rac Signaling | 2.11 | 0.0661 | 0 | CYFIP1,IQGAP1,IQGAP2,ITGB5,JUN,PAK3,PARD3,TIAM1 |
| Leptin Signaling in Obesity | 2.1 | 0.0811 | #NUM! | ADCY2,AKT3,PDE3B,PDIA3,PLCB1,PLCB2 |
| Hypoxia Signaling in the Cardiovascular System | 2.1 | 0.0811 | #NUM! | HSP90AA1,HSP90AB1,HSP90B1,JUN,LDHA,UBE2J1 |
| Tec Kinase Signaling | 2.1 | 0.0578 | 0.333 | FCER1G,FGR,FOS,FYN,ITGB5,MAPK10,PAK3,PRKCA,PRKCB,PRKCE |
| Inhibition of Angiogenesis by TSP1 | 2.1 | 0.118 | -1 | AKT3,FYN,JUN,MAPK10 |
| TREM1 Signaling | 2.08 | 0.08 | 0.816 | AKT3,IL1B,ITGAX,LAT2,TLR2,TLR5 |
| CCR3 Signaling in Eosinophils | 2.04 | 0.0645 | -0.816 | ITPR2,PAK3,PLCB1,PLCB2,PRKCA,PRKCB,PRKCE,ROCK1 |
| Factors Promoting Cardiogenesis in Vertebrates | 2.04 | 0.06 | -0.333 | MAPK10,MEF2C,PDIA3,PLCB1,PLCB2,PRKCA,PRKCB,PRKCE,ROCK1 |
| EGF Signaling | 2.03 | 0.0909 | -0.447 | AKT3,FOS,ITPR2,JUN,PRKCA |
| B Cell Development | 2.01 | 0.111 | #NUM! | HLA-B,HLA-DRA,HLA-DRB1,HLA-DRB5 |
| Chondroitin Sulfate Biosynthesis | 2 | 0.0893 | 0.447 | CHST15,CHSY3,CSGALNACT1,HS6ST3,UST |
| Thrombin Signaling | 1.98 | 0.0529 | -0.302 | ADCY2,AKT3,CAMK1D,ITPR2,PDIA3,PLCB1,PLCB2,PRKCA,PRKCB,PRKCE,ROCK1 |
| Thyroid Cancer Signaling | 1.97 | 0.0759 | -0.816 | AKT3,FOS,IGF1,JUN,NTRK2,NTRK3 |

| | | | | |
|---|---|---|---|---|
| Cell Cycle Regulation by BTG Family Proteins | 1.97 | 0.108 | #NUM! | BTG1,BTG2,E2F3,PPP2R2B |
| p70S6K Signaling | 1.95 | 0.062 | 0 | AKT3,PDIA3,PLCB1,PLCB2,PPP2R2B,PRKCA,PRKCB,PRKCE |
| Renal Cell Carcinoma Signaling | 1.94 | 0.075 | -0.447 | AKT3,FOS,JUN,PAK3,RAPGEF1,UBC |
| Gα12/13 Signaling | 1.91 | 0.0611 | 0.707 | AKT3,CDH12,CDH18,CDH23,JUN,MAPK10,MEF2C,ROCK1 |
| Integrin Signaling | 1.91 | 0.0516 | -1.897 | AKT3,BCAR3,FYN,GSN,ITGAX,ITGB5,NCK2,PAK3,RAPGEF1,ROCK1,TLN2 |
| Dermatan Sulfate Biosynthesis | 1.91 | 0.0847 | 0.447 | CHST15,CHSY3,CSGALNACT1,HS6ST3,UST |
| Systemic Lupus Erythematosus In T Cell Signaling Path | 1.88 | 0.0449 | 1.291 | AKT3,B2M,BCL6,CREM,FCER1G,FOS,HLA-B,HLA-DPA1,HLA-DRA,HLA-DRB1,HLA-DRB5,JUN,MAP2K6,PPP2R2B,ROCK1 |
| PCP pathway | 1.88 | 0.0833 | 0.447 | DAAM1,JUN,JUNB,MAPK10,ROCK1 |
| Regulation Of The Epithelial Mesenchymal Transition By | 1.86 | 0.0532 | -0.707 | AKT3,CDH12,FGF12,FGF14,FOS,ID2,JUN,MAP2K6,MAPK10,TNFRSF1A |
| VEGF Family Ligand-Receptor Interactions | 1.85 | 0.0714 | 0.447 | AKT3,FOS,NRP1,PRKCA,PRKCB,PRKCE |
| FGF Signaling | 1.85 | 0.0714 | -2.449 | AKT3,FGF12,FGF14,MAP2K6,PRKCA,RPS6KA5 |
| Role of Osteoblasts, Osteoclasts and Chondrocytes in R | 1.84 | 0.0505 | #NUM! | AKT3,FOS,GSN,IGF1,IL1B,IL1RAPL1,JUN,MAP2K6,MAPK10,SPP1,TNFRSF1A |
| STAT3 Pathway | 1.84 | 0.0593 | -0.816 | CDKN1A,IGF1,IGF2R,IL17RA,IL1B,MAPK10,NTRK2,NTRK3 |
| ILK Signaling | 1.83 | 0.0526 | 0 | AKT3,FOS,ITGB5,JUN,MAP2K6,MAPK10,NCK2,PPP2R2B,RPS6KA5,TNFRSF1A |
| IL-4 Signaling | 1.82 | 0.0706 | #NUM! | AKT3,HLA-B,HLA-DRA,HLA-DRB1,HLA-DRB5,INPP5D |
| Th2 Pathway | 1.82 | 0.0588 | #NUM! | BHLHE41,CCR1,HLA-B,HLA-DPA1,HLA-DRA,HLA-DRB1,HLA-DRB5,JUN |
| Thrombopoietin Signaling | 1.79 | 0.0794 | 1.342 | FOS,JUN,PRKCA,PRKCB,PRKCE |
| Glioblastoma Multiforme Signaling | 1.79 | 0.0545 | -1.89 | AKT3,CDK6,CDKN1A,E2F3,IGF1,ITPR2,PDIA3,PLCB1,PLCB2 |
| MIF Regulation of Innate Immunity | 1.78 | 0.0952 | 1 | CD14,FOS,JUN,MAPK10 |
| Signaling by Rho Family GTPases | 1.77 | 0.0474 | -0.632 | CDH12,CDH18,CDH23,CYFIP1,FOS,IQGAP1,ITGB5,JUN,MAPK10,PAK3,PARD3,ROCK1 |
| Role of IL-17F in Allergic Inflammatory Airway Diseases | 1.74 | 0.093 | #NUM! | IGF1,IL17RA,IL1B,RPS6KA5 |
| Crosstalk between Dendritic Cells and Natural Killer Cells | 1.74 | 0.0674 | 2 | HLA-B,HLA-DRA,HLA-DRB1,HLA-DRB5,IL15,TLN2 |
| tRNA Splicing | 1.71 | 0.0909 | -2 | PDE1A,PDE3B,PDE4D,PDE9A |
| fMLP Signaling in Neutrophils | 1.7 | 0.0603 | -0.816 | FPR1,ITPR2,PLCB1,PLCB2,PRKCA,PRKCB,PRKCE |
| Systemic Lupus Erythematosus Signaling | 1.7 | 0.048 | #NUM! | AKT3,CREM,FCER1G,FCGR1A,FCGR2A,FCGR3A/FCGR3B,FOS,HLA-B,IL1B,INPP5D,JUN |
| Th1 and Th2 Activation Pathway | 1.7 | 0.0526 | #NUM! | BHLHE41,CCR1,HLA-B,HLA-DPA1,HLA-DRA,HLA-DRB1,HLA-DRB5,JUN,MAP2K6 |
| ErbB4 Signaling | 1.69 | 0.0746 | -0.447 | ERBB4,NRG3,PRKCA,PRKCB,PRKCE |
| Sirtuin Signaling Pathway | 1.68 | 0.0447 | -0.707 | H3-3A/H3-3B,JUN,LDHA,MT-ATP6,MT-CYB,MT-ND1,MT-ND3,MT-ND4,PGK1,PPARG,TSPO,TUBA1A,TUBA1B |
| iNOS Signaling | 1.68 | 0.0889 | 2 | CD14,FOS,IRAK3,JUN |
| IL-1 Signaling | 1.67 | 0.0652 | 0.447 | ADCY2,FOS,IRAK3,JUN,MAP2K6,MAPK10 |
| Erythropoietin Signaling Pathway | 1.67 | 0.052 | -0.333 | AKT3,FOS,IL15,IL1B,ITPR2,JUN,PRKCA,PRKCB,PRKCE |
| Remodeling of Epithelial Adherens Junctions | 1.66 | 0.0735 | #NUM! | CTNNA2,DNM1,IQGAP1,TUBA1A,TUBA1B |
| Agrin Interactions at Neuromuscular Junction | 1.61 | 0.0714 | -1.342 | ERBB4,JUN,MAPK10,NRG3,PAK3 |
| Relaxin Signaling | 1.58 | 0.053 | 0 | ADCY2,AKT3,FOS,JUN,PDE1A,PDE3B,PDE4D,PDE9A |
| Acute Phase Response Signaling | 1.57 | 0.05 | 1.414 | AKT3,CEBPB,FOS,HMOX1,IL1B,JUN,MAP2K6,SERPINA1,TNFRSF1A |
| GP6 Signaling Pathway | 1.55 | 0.056 | 0.378 | AKT3,FCER1G,FYN,LCP2,PRKCA,PRKCB,PRKCE |
| GPCR-Mediated Integration of Enteroendocrine Signalin | 1.54 | 0.0685 | -0.447 | ADCY2,ITPR2,PDIA3,PLCB1,PLCB2 |
| TNFR1 Signaling | 1.53 | 0.08 | 1 | FOS,JUN,PAK3,TNFRSF1A |
| Systemic Lupus Erythematosus In B Cell Signaling Path | 1.52 | 0.0436 | 1.155 | AKT3,FCGR2A,FGR,FOS,FYN,IL15,IL1B,INPP5D,JUN,PRKCA,PRKCB,PRKCE |
| PI3K/AKT Signaling | 1.52 | 0.0489 | 1.134 | AKT3,CDKN1A,HSP90AA1,HSP90AB1,HSP90B1,IL17RA,INPP5D,ITGB5,PPP2R2B |
| Atherosclerosis Signaling | 1.52 | 0.0551 | #NUM! | ALOX15B,APOC1,APOE,CLU,IL1B,MSR1,SERPINA1 |
| Glycogen Degradation II | 1.51 | 0.167 | #NUM! | PYGL,TYMP |

| | | | | |
|---|---|---|---|---|
| B Cell Receptor Signaling | 1.49 | 0.0484 | 0.707 | AKT3,BCL2A1,BCL6,FCGR2A,INPP5D,JUN,MAP2K6,MEF2C,PRKCB |
| Neurotrophin/TRK Signaling | 1.48 | 0.0658 | #NUM! | FOS,JUN,MAP2K6,NTRK2,NTRK3 |
| Macropinocytosis Signaling | 1.48 | 0.0658 | #NUM! | CD14,ITGB5,PRKCA,PRKCB,PRKCE |
| CD27 Signaling in Lymphocytes | 1.45 | 0.0755 | #NUM! | FOS,JUN,MAP2K6,MAPK10 |
| JAK/Stat Signaling | 1.4 | 0.0625 | 0.447 | AKT3,CDKN1A,CEBPB,FOS,JUN |
| RAR Activation | 1.39 | 0.0464 | #NUM! | ADCY2,AKT3,FOS,JUN,MAPK10,PRKCA,PRKCB,PRKCE,ZBTB16 |
| HMGB1 Signaling | 1.38 | 0.0485 | 0.378 | AKT3,FOS,IL15,IL1B,JUN,MAP2K6,MAPK10,TNFRSF1A |
| Glycogen Degradation III | 1.38 | 0.143 | #NUM! | PYGL,TYMP |
| Cyclins and Cell Cycle Regulation | 1.38 | 0.0617 | 1 | CDK6,CDKN1A,E2F3,HDAC4,PPP2R2B |
| CDK5 Signaling | 1.38 | 0.0556 | -0.816 | ADCY2,CABLES1,CACNA1A,MAPK10,NTRK2,PPP2R2B |
| Paxillin Signaling | 1.38 | 0.0556 | -1 | ITGAX,ITGB5,MAPK10,NCK2,PAK3,TLN2 |
| Cytotoxic T Lymphocyte-mediated Apoptosis of Target C | 1.35 | 0.0882 | #NUM! | B2M,FCER1G,HLA-B |
| Tight Junction Signaling | 1.34 | 0.0476 | #NUM! | AKT3,FOS,JUN,MAGI2,PPP2R2B,SNAP25,TIAM1,TNFRSF1A |
| Putrescine Biosynthesis III | 1.33 | 0.5 | #NUM! | AZIN2 |
| MSP-RON Signaling In Macrophages Pathway | 1.3 | 0.0531 | -0.816 | FOS,HLA-DPA1,HLA-DRA,HLA-DRB1,HLA-DRB5,JUN |
| PDGF Signaling | 1.28 | 0.0581 | 0.447 | FOS,INPP5D,JUN,PRKCA,PRKCB |
| Xenobiotic Metabolism General Signaling Pathway | 1.28 | 0.049 | 0.378 | AKT3,GSTK1,HMOX1,MAP2K6,PRKCA,PRKCB,PRKCE |
| Semaphorin Signaling in Neurons | 1.28 | 0.0667 | #NUM! | FYN,NRP1,PAK3,ROCK1 |
| Maturity Onset Diabetes of Young (MODY) Signaling | 1.28 | 0.0667 | #NUM! | APOC1,APOE,CACNA1A,CACNA1C |
| Ceramide Signaling | 1.25 | 0.0568 | 1.342 | AKT3,FOS,JUN,PPP2R2B,TNFRSF1A |
| Wnt/Ca+ pathway | 1.24 | 0.0645 | -1 | PDIA3,PLCB1,PLCB2,PRKCA |
| Sphingosine-1-phosphate Signaling | 1.24 | 0.0513 | -0.447 | ACER3,ADCY2,AKT3,PDIA3,PLCB1,PLCB2 |
| Docosahexaenoic Acid (DHA) Signaling | 1.23 | 0.0789 | #NUM! | AKT3,BCL2A1,IL1B |
| Prostate Cancer Signaling | 1.2 | 0.0549 | #NUM! | AKT3,CDKN1A,HSP90AA1,HSP90AB1,HSP90B1 |
| Th1 Pathway | 1.18 | 0.0496 | 1.342 | HLA-B,HLA-DPA1,HLA-DRA,HLA-DRB1,HLA-DRB5,MAP2K6 |
| CD40 Signaling | 1.18 | 0.0615 | 0 | FOS,JUN,MAP2K6,MAPK10 |
| Phospholipases | 1.18 | 0.0615 | 0 | HMOX1,PDIA3,PLCB1,PLCB2 |
| April Mediated Signaling | 1.17 | 0.075 | #NUM! | FOS,JUN,MAPK10 |
| Uracil Degradation II (Reductive) | 1.16 | 0.333 | #NUM! | DPYD |
| Diphthamide Biosynthesis | 1.16 | 0.333 | #NUM! | DPH6 |
| Thymine Degradation | 1.16 | 0.333 | #NUM! | DPYD |
| Mitotic Roles of Polo-Like Kinase | 1.16 | 0.0606 | #NUM! | HSP90AA1,HSP90AB1,HSP90B1,PPP2R2B |
| Pyridoxal 5'-phosphate Salvage Pathway | 1.16 | 0.0606 | 0 | CDK6,MAP2K6,PAK3,PRKCE |
| B Cell Activating Factor Signaling | 1.15 | 0.0732 | #NUM! | FOS,JUN,MAPK10 |
| Mechanisms of Viral Exit from Host Cells | 1.15 | 0.0732 | #NUM! | PRKCA,PRKCB,PRKCE |
| Cell Cycle: G1/S Checkpoint Regulation | 1.14 | 0.0597 | -1 | CDK6,CDKN1A,E2F3,HDAC4 |
| Nur77 Signaling in T Lymphocytes | 1.12 | 0.0588 | 1 | FCER1G,PRKCA,PRKCB,PRKCE |
| Ferroptosis Signaling Pathway | 1.11 | 0.0476 | 0.816 | ALOX15B,CDKN1A,HMOX1,HSPB1,SAT1,STEAP3 |
| RhoGDI Signaling | 1.11 | 0.0423 | 1.633 | CDH12,CDH18,CDH23,GRIP1,ITGB5,PAK3,PRKCA,ROCK1 |
| Bladder Cancer Signaling | 1.11 | 0.0515 | #NUM! | CDKN1A,FGF12,FGF14,MMP16,RPS6KA5 |
| Inflammasome pathway | 1.1 | 0.1 | #NUM! | IL1B,NLRP1 |
| GM-CSF Signaling | 1.08 | 0.0571 | #NUM! | AKT3,BCL2A1,CSF2RA,PRKCB |
| VEGF Signaling | 1.08 | 0.0505 | 0 | AKT3,EIF1,PRKCA,PRKCB,ROCK1 |
| IL-23 Signaling Pathway | 1.07 | 0.0682 | #NUM! | AKT3,IL1B,RORA |

| | | | | |
|---|---|---|---|---|
| Leukocyte Extravasation Signaling | 1.06 | 0.0415 | -0.707 | CTNNA2,CTNNA3,MAPK10,MMP16,PRKCA,PRKCB,PRKCE,ROCK1 |
| Apoptosis Signaling | 1.06 | 0.05 | 0.447 | BCL2A1,PRKCA,PRKCE,ROCK1,TNFRSF1A |
| Heme Degradation | 1.04 | 0.25 | #NUM! | HMOX1 |
| Spermine and Spermidine Degradation I | 1.04 | 0.25 | #NUM! | SAT1 |
| Glutathione Redox Reactions II | 1.04 | 0.25 | #NUM! | PDIA3 |
| Acetate Conversion to Acetyl-CoA | 1.04 | 0.25 | #NUM! | ACSL1 |
| Non-Small Cell Lung Cancer Signaling | 1.03 | 0.0548 | #NUM! | AKT3,CDK6,ITPR2,PRKCA |
| Dermatan Sulfate Biosynthesis (Late Stages) | 1.03 | 0.0652 | #NUM! | CHST15,HS6ST3,UST |
| PFKFB4 Signaling Pathway | 1.03 | 0.0652 | #NUM! | HK2,MAP2K6,RPS6KA5 |
| Chronic Myeloid Leukemia Signaling | 1.02 | 0.0485 | #NUM! | AKT3,CDK6,CDKN1A,E2F3,HDAC4 |
| Adipogenesis pathway | 1.02 | 0.0448 | #NUM! | CEBPB,CEBPD,HDAC4,PPARG,TNFRSF1A,XBP1 |
| Ephrin A Signaling | 1.01 | 0.0638 | #NUM! | EPHA6,FYN,ROCK1 |
| FAK Signaling | 1.01 | 0.0481 | #NUM! | AKT3,FYN,ITGB5,PAK3,TLN2 |
| Estrogen-Dependent Breast Cancer Signaling | 1 | 0.0533 | 0 | AKT3,FOS,IGF1,JUN |
| Role of MAPK Signaling in Inhibiting the Pathogenesis c | 1 | 0.0533 | 0 | IL1B,JUN,MAP2K6,MAPK10 |
| Polyamine Regulation in Colon Cancer | 0.996 | 0.087 | #NUM! | PPARG,SAT1 |
| NF-κB Activation by Viruses | 0.983 | 0.0526 | 0 | AKT3,PRKCA,PRKCB,PRKCE |
| GDNF Family Ligand-Receptor Interactions | 0.983 | 0.0526 | #NUM! | FOS,ITPR2,JUN,MAPK10 |
| PAK Signaling | 0.979 | 0.0472 | -1 | DSCAM,ITGB5,MAPK10,NCK2,PAK3 |
| IL-22 Signaling | 0.967 | 0.0833 | #NUM! | AKT3,MAPK10 |
| Vitamin-C Transport | 0.967 | 0.0833 | #NUM! | NXN,SLC2A3 |
| Insulin Receptor Signaling | 0.959 | 0.0432 | -1.633 | AKT3,ASIC2,FYN,INPP5D,PDE3B,RAPGEF1 |
| IL-7 Signaling Pathway | 0.955 | 0.0513 | #NUM! | AKT3,BCL6,FYN,JUN |
| Creatine-phosphate Biosynthesis | 0.951 | 0.2 | #NUM! | CKB |
| Tetrahydrofolate Salvage from 5,10-methenyltetrahydrof | 0.951 | 0.2 | #NUM! | MTHFD2 |
| dTMP De Novo Biosynthesis | 0.951 | 0.2 | #NUM! | DHFR |
| Melanoma Signaling | 0.947 | 0.06 | #NUM! | AKT3,CDKN1A,MITF |
| Pancreatic Adenocarcinoma Signaling | 0.943 | 0.0459 | 0 | AKT3,CDKN1A,E2F3,HMOX1,MAPK10 |
| D-myo-inositol (1,4,5)-Trisphosphate Biosynthesis | 0.936 | 0.08 | #NUM! | PLCB1,PLCB2 |
| Role of MAPK Signaling in the Pathogenesis of Influenz | 0.924 | 0.05 | #NUM! | AKT3,MAP2K6,MAPK10,PRKCA |
| Endocannabinoid Cancer Inhibition Pathway | 0.917 | 0.042 | 0.816 | ADCY2,AKT3,CDKN1A,MAP2K6,ROCK1,SPTLC2 |
| mTOR Signaling | 0.91 | 0.0381 | 0.378 | AKT3,DDIT4,HMOX1,PPP2R2B,PRKCA,PRKCB,PRKCE,RPS6KA5 |
| Estrogen-mediated S-phase Entry | 0.907 | 0.0769 | #NUM! | CDKN1A,E2F3 |
| Pyruvate Fermentation to Lactate | 0.876 | 0.167 | #NUM! | LDHA |
| Ceramide Degradation | 0.876 | 0.167 | #NUM! | ACER3 |
| Role of IL-17A in Arthritis | 0.857 | 0.0545 | #NUM! | IL17RA,MAP2K6,MAPK10 |
| Colorectal Cancer Metastasis Signaling | 0.845 | 0.0356 | 0.333 | ADCY2,AKT3,FOS,JUN,MAPK10,MMP16,TLR2,TLR5,TNFRSF1A |
| Role of CHK Proteins in Cell Cycle Checkpoint Control | 0.824 | 0.0526 | #NUM! | CDKN1A,E2F3,PPP2R2B |
| Trehalose Degradation II (Trehalase) | 0.815 | 0.143 | #NUM! | HK2 |
| Thioredoxin Pathway | 0.815 | 0.143 | #NUM! | NXN |
| TNFR2 Signaling | 0.807 | 0.0667 | #NUM! | FOS,JUN |
| Inhibition of ARE-Mediated mRNA Degradation Pathway | 0.796 | 0.041 | -0.447 | AKT3,PABPN1,PPP2R2B,TNFRSF1A,ZFP36 |
| Regulation of IL-2 Expression in Activated and Anergic | 0.79 | 0.0444 | #NUM! | FOS,FYN,JUN,MAPK10 |
| G Protein Signaling Mediated by Tubby | 0.785 | 0.0645 | #NUM! | PLCB1,PLCB2 |

| | | | | |
|---|---|---|---|---|
| Regulation of the Epithelial-Mesenchymal Transition Pat | 0.777 | 0.0365 | #NUM! | AKT3,CDH12,FGF12,FGF14,ID2,MAP2K6,RBPJ |
| SPINK1 Pancreatic Cancer Pathway | 0.777 | 0.05 | #NUM! | CPA6,CPE,CPM |
| Gαi Signaling | 0.764 | 0.04 | 0.447 | ADCY2,FPR1,GRM7,P2RY12,RGS7 |
| 4-1BB Signaling in T Lymphocytes | 0.762 | 0.0625 | #NUM! | JUN,MAPK10 |
| Sphingosine and Sphingosine-1-phosphate Metabolism | 0.762 | 0.125 | #NUM! | ACER3 |
| Histidine Degradation III | 0.762 | 0.125 | #NUM! | MTHFD2 |
| Salvage Pathways of Pyrimidine Deoxyribonucleotides | 0.762 | 0.125 | #NUM! | TYMP |
| IL-2 Signaling | 0.762 | 0.0492 | #NUM! | AKT3,FOS,JUN |
| HOTAIR Regulatory Pathway | 0.754 | 0.0375 | -0.447 | AKT3,CDKN1A,H3-3A/H3-3B,MMP16,ROCK1,SPP1 |
| Circadian Rhythm Signaling | 0.742 | 0.0606 | #NUM! | BHLHE41,GRIN2A |
| Pathogenesis of Multiple Sclerosis | 0.714 | 0.111 | #NUM! | CCR1 |
| Folate Transformations I | 0.714 | 0.111 | #NUM! | MTHFD2 |
| ATM Signaling | 0.712 | 0.0412 | 1 | CDKN1A,JUN,MAPK10,PPP2R2B |
| IL-17A Signaling in Airway Cells | 0.708 | 0.0462 | #NUM! | AKT3,IL17RA,MAPK10 |
| Salvage Pathways of Pyrimidine Ribonucleotides | 0.701 | 0.0408 | 0 | CDK6,MAP2K6,PAK3,PRKCE |
| Prostanoid Biosynthesis | 0.674 | 0.1 | #NUM! | PTGES3 |
| Notch Signaling | 0.666 | 0.0541 | #NUM! | CNTN1,RBPJ |
| Iron homeostasis signaling pathway | 0.656 | 0.0365 | #NUM! | CD163,HMOX1,SLC11A1,SLC25A37,STEAP3 |
| Wnt/β-catenin Signaling | 0.652 | 0.0347 | 1 | AKT3,CDH12,JUN,PPP2R2B,SOX5,UBC |
| Regulation of Actin-based Motility by Rho | 0.65 | 0.0388 | #NUM! | GSN,ITGB5,PAK3,ROCK1 |
| IGF-1 Signaling | 0.64 | 0.0385 | 0 | AKT3,FOS,IGF1,JUN |
| GDP-glucose Biosynthesis | 0.638 | 0.0909 | #NUM! | HK2 |
| Hereditary Breast Cancer Signaling | 0.633 | 0.0357 | #NUM! | AKT3,CDK6,CDKN1A,HDAC4,UBC |
| AMPK Signaling | 0.627 | 0.0329 | 0.816 | AKT3,CAB39,CDKN1A,CHRM3,PPP2R2B,RAB11A,RAB27A |
| ERK5 Signaling | 0.622 | 0.0417 | #NUM! | FOS,MEF2C,RPS6KA5 |
| Ephrin B Signaling | 0.622 | 0.0417 | #NUM! | KALRN,NCK2,ROCK1 |
| Heparan Sulfate Biosynthesis (Late Stages) | 0.622 | 0.0417 | #NUM! | CHST15,HS6ST3,UST |
| T Cell Receptor Signaling | 0.622 | 0.0377 | #NUM! | FOS,FYN,JUN,LCP2 |
| Gαs Signaling | 0.613 | 0.0374 | -2 | ADCY2,CHRM3,RYR2,RYR3 |
| Cleavage and Polyadenylation of Pre-mRNA | 0.606 | 0.0833 | #NUM! | PABPN1 |
| Hematopoiesis from Multipotent Stem Cells | 0.606 | 0.0833 | #NUM! | IL15 |
| Glucose and Glucose-1-phosphate Degradation | 0.606 | 0.0833 | #NUM! | HK2 |
| Regulation of Cellular Mechanics by Calpain Protease | 0.599 | 0.0405 | #NUM! | CDK6,ITGB5,TLN2 |
| Role of MAPK Signaling in Promoting the Pathogenesis | 0.587 | 0.0364 | -1 | JUN,MAP2K6,MAPK10,PRKCA |
| Fatty Acid Activation | 0.575 | 0.0769 | #NUM! | ACSL1 |
| UDP-N-acetyl-D-galactosamine Biosynthesis II | 0.575 | 0.0769 | #NUM! | HK2 |
| Hepatic Fibrosis / Hepatic Stellate Cell Activation | 0.564 | 0.0323 | #NUM! | CD14,IGF1,IL1B,IL1RAPL1,KLF6,TNFRSF1A |
| Apelin Pancreas Signaling Pathway | 0.558 | 0.0455 | #NUM! | MAPK10,PDE3B |
| Coronavirus Pathogenesis Pathway | 0.556 | 0.0333 | 0.447 | E2F3,FOS,IL1B,JUN,MAPK10 |
| NGF Signaling | 0.553 | 0.0351 | #NUM! | AKT3,MAPK10,ROCK1,RPS6KA5 |
| Role of IL-17A in Psoriasis | 0.548 | 0.0714 | #NUM! | IL17RA |
| Heparan Sulfate Biosynthesis | 0.547 | 0.038 | #NUM! | CHST15,HS6ST3,UST |
| Endocannabinoid Developing Neuron Pathway | 0.545 | 0.0348 | -1 | ADCY2,AKT3,MAP2K6,MAPK10 |
| Coronavirus Replication Pathway | 0.545 | 0.0444 | #NUM! | TUBA1A,TUBA1B |

| | | | | |
|---|---|---|---|---|
| FLT3 Signaling in Hematopoietic Progenitor Cells | 0.538 | 0.0375 | #NUM! | AKT3,INPP5D,RPS6KA5 |
| FAT10 Cancer Signaling Pathway | 0.532 | 0.0435 | #NUM! | AKT3,TNFRSF1A |
| Actin Nucleation by ARP-WASP Complex | 0.527 | 0.037 | #NUM! | ITGB5,NCK2,ROCK1 |
| Choline Biosynthesis III | 0.523 | 0.0667 | #NUM! | HMOX1 |
| Airway Pathology in Chronic Obstructive Pulmonary Dise | 0.521 | 0.0339 | #NUM! | FGF12,FGF14,IL15,IL1B |
| PEDF Signaling | 0.519 | 0.0366 | #NUM! | AKT3,PPARG,ROCK1 |
| Apelin Adipocyte Signaling Pathway | 0.519 | 0.0366 | #NUM! | ADCY2,GSTK1,MAPK10 |
| TR/RXR Activation | 0.5 | 0.0357 | #NUM! | AKT3,PDE3B,STRBP |
| HIPPO signaling | 0.492 | 0.0353 | #NUM! | DLG2,PARD3,PPP2R2B |
| Amyloid Processing | 0.483 | 0.04 | #NUM! | AKT3,PRKCE |
| γ-linolenate Biosynthesis II (Animals) | 0.478 | 0.0588 | #NUM! | ACSL1 |
| Mitochondrial L-carnitine Shuttle Pathway | 0.478 | 0.0588 | #NUM! | ACSL1 |
| D-myo-inositol (1,4,5)-trisphosphate Degradation | 0.478 | 0.0588 | #NUM! | INPP5D |
| Acute Myeloid Leukemia Signaling | 0.458 | 0.0337 | #NUM! | AKT3,CSF2RA,MAP2K6 |
| Differential Regulation of Cytokine Production in Macrop | 0.458 | 0.0556 | #NUM! | IL1B |
| 1D-myo-inositol Hexakisphosphate Biosynthesis II (Mam | 0.458 | 0.0556 | #NUM! | INPP5D |
| D-myo-inositol (1,3,4)-trisphosphate Biosynthesis | 0.458 | 0.0556 | #NUM! | INPP5D |
| Role of Cytokines in Mediating Communication between | 0.44 | 0.037 | #NUM! | IL15,IL1B |
| GADD45 Signaling | 0.439 | 0.0526 | #NUM! | CDKN1A |
| Oxidative Ethanol Degradation III | 0.439 | 0.0526 | #NUM! | ACSL1 |
| DNA damage-induced 14-3-3σ Signaling | 0.439 | 0.0526 | #NUM! | AKT3 |
| Apelin Muscle Signaling Pathway | 0.439 | 0.0526 | #NUM! | AKT3 |
| Death Receptor Signaling | 0.435 | 0.0326 | #NUM! | HSPB1,ROCK1,TNFRSF1A |
| Cardiomyocyte Differentiation via BMP Receptors | 0.421 | 0.05 | #NUM! | MEF2C |
| Melanocyte Development and Pigmentation Signaling | 0.42 | 0.0319 | #NUM! | ADCY2,MITF,RPS6KA5 |
| Granulocyte Adhesion and Diapedesis | 0.416 | 0.0289 | #NUM! | FPR1,IL1B,IL1RAPL1,MMP16,TNFRSF1A |
| Putrescine Degradation III | 0.406 | 0.0476 | #NUM! | SAT1 |
| TGF-β Signaling | 0.406 | 0.0312 | #NUM! | FOS,JUN,MAP2K6 |
| p53 Signaling | 0.391 | 0.0306 | #NUM! | AKT3,CDKN1A,JUN |
| Endometrial Cancer Signaling | 0.384 | 0.0333 | #NUM! | AKT3,CTNNA2 |
| Induction of Apoptosis by HIV1 | 0.376 | 0.0328 | #NUM! | MAPK10,TNFRSF1A |
| Differential Regulation of Cytokine Production in Intestin | 0.376 | 0.0435 | #NUM! | IL1B |
| Superpathway of D-myo-inositol (1,4,5)-trisphosphate M | 0.376 | 0.0435 | #NUM! | INPP5D |
| Ethanol Degradation IV | 0.376 | 0.0435 | #NUM! | ACSL1 |
| Apelin Cardiac Fibroblast Signaling Pathway | 0.376 | 0.0435 | #NUM! | AKT3 |
| SAPK/JNK Signaling | 0.366 | 0.0294 | #NUM! | FCER1G,JUN,MAPK10 |
| Tumoricidal Function of Hepatic Natural Killer Cells | 0.362 | 0.0417 | #NUM! | SERPINB9 |
| Glutathione Redox Reactions I | 0.362 | 0.0417 | #NUM! | GSTK1 |
| Sumoylation Pathway | 0.36 | 0.0291 | #NUM! | FOS,JUN,MAPK10 |
| Activation of IRF by Cytosolic Pattern Recognition Rece | 0.359 | 0.0317 | #NUM! | JUN,MAPK10 |
| Role of JAK family kinases in IL-6-type Cytokine Signalir | 0.349 | 0.04 | #NUM! | MAPK10 |
| ErbB2-ErbB3 Signaling | 0.344 | 0.0308 | #NUM! | JUN,NRG3 |
| Glycolysis I | 0.336 | 0.0385 | #NUM! | PGK1 |
| Gluconeogenesis I | 0.336 | 0.0385 | #NUM! | PGK1 |

| | | | | |
|---|---|---|---|---|
| Apelin Liver Signaling Pathway | 0.336 | 0.0385 | #NUM! | MAPK10 |
| Eicosanoid Signaling | 0.336 | 0.0303 | #NUM! | ALOX15B,ALOX5AP |
| Small Cell Lung Cancer Signaling | 0.301 | 0.0282 | #NUM! | AKT3,CDK6 |
| Necroptosis Signaling Pathway | 0.296 | 0.0255 | 2 | PYGL,TNFRSF1A,TSPO,UBC |
| IL-15 Signaling | 0.277 | 0.0267 | #NUM! | AKT3,IL15 |
| Angiopoietin Signaling | 0.277 | 0.0267 | #NUM! | AKT3,PAK3 |
| Glutathione-mediated Detoxification | 0.273 | 0.0312 | #NUM! | GSTK1 |
| Ethanol Degradation II | 0.273 | 0.0312 | #NUM! | ACSL1 |
| Dopamine Receptor Signaling | 0.265 | 0.026 | #NUM! | ADCY2,PPP2R2B |
| Fatty Acid β-oxidation I | 0.264 | 0.0303 | #NUM! | ACSL1 |
| Regulation of eIF4 and p70S6K Signaling | 0.26 | 0.0241 | #NUM! | AKT3,EIF1,ITGB5,PPP2R2B |
| RhoA Signaling | 0.256 | 0.0244 | #NUM! | IGF1,PLEKHG5,ROCK1 |
| MIF-mediated Glucocorticoid Regulation | 0.256 | 0.0294 | #NUM! | CD14 |
| Role of BRCA1 in DNA Damage Response | 0.249 | 0.025 | #NUM! | CDKN1A,E2F3 |
| Coagulation System | 0.248 | 0.0286 | #NUM! | SERPINA1 |
| DNA Methylation and Transcriptional Repression Signali | 0.248 | 0.0286 | #NUM! | H3-3A/H3-3B |
| Thyroid Hormone Metabolism II (via Conjugation and/or | 0.225 | 0.0263 | #NUM! | CSGALNACT1 |
| BMP signaling pathway | 0.224 | 0.0235 | #NUM! | JUN,MAPK10 |
| Antiproliferative Role of TOB in T Cell Signaling | 0.218 | 0.0256 | #NUM! | FCER1G |
| Inhibition of Matrix Metalloproteases | 0.218 | 0.0256 | #NUM! | MMP16 |
| PXR/RXR Activation | 0 | 0.0154 | #NUM! | AKT3 |
| Lymphotoxin β Receptor Signaling | 0 | 0.0189 | #NUM! | AKT3 |
| CNTF Signaling | 0 | 0.0175 | #NUM! | RPS6KA5 |
| Human Embryonic Stem Cell Pluripotency | 0 | 0.0222 | #NUM! | AKT3,NTRK2,NTRK3 |
| Antiproliferative Role of Somatostatin Receptor 2 | 0 | 0.013 | #NUM! | CDKN1A |
| Role of Oct4 in Mammalian Embryonic Stem Cell Pluripo | 0 | 0.0217 | #NUM! | SPP1 |
| Role of NANOG in Mammalian Embryonic Stem Cell Plur | 0 | 0.0084 | #NUM! | AKT3 |
| Myc Mediated Apoptosis Signaling | 0 | 0.02 | #NUM! | TNFRSF1A |
| EIF2 Signaling | 0 | 0.0179 | #NUM! | AKT3,EIF1,HSPA5,RPL13A |
| Ovarian Cancer Signaling | 0 | 0.00719 | #NUM! | AKT3 |
| Neuroprotective Role of THOP1 in Alzheimer's Disease | 0 | 0.0172 | #NUM! | HLA-B,HTRA1 |
| Role of PI3K/AKT Signaling in the Pathogenesis of Influ | 0 | 0.0156 | #NUM! | AKT3 |
| Role of Hypercytokinemia/hyperchemokinemia in the Pa | 0 | 0.0116 | #NUM! | IL1B |
| MSP-RON Signaling Pathway | 0 | 0.0172 | #NUM! | TLR2 |
| Cell Cycle Control of Chromosomal Replication | 0 | 0.0179 | #NUM! | CDK6 |
| Spliceosomal Cycle | 0 | 0.0208 | #NUM! | HSPA8 |
| Role of JAK1 and JAK3 in γc Cytokine Signaling | 0 | 0.0145 | #NUM! | IL15 |
| Mouse Embryonic Stem Cell Pluripotency | 0 | 0.0194 | #NUM! | AKT3,ID2 |
| Hematopoiesis from Pluripotent Stem Cells | 0 | 0.0204 | #NUM! | FCER1G |
| Transcriptional Regulatory Network in Embryonic Stem C | 0 | 0.0185 | #NUM! | H3-3A/H3-3B |
| Nicotine Degradation III | 0 | 0.0175 | #NUM! | CSGALNACT1 |
| D-myo-inositol-5-phosphate Metabolism | 0 | 0.0191 | #NUM! | PLCB1,PLCB2,PPFIA2 |
| Melatonin Degradation I | 0 | 0.0167 | #NUM! | CSGALNACT1 |
| D-myo-inositol (1,4,5,6)-Tetrakisphosphate Biosynthesis | 0 | 0.00704 | #NUM! | PPFIA2 |

| | | | | |
|---|---|---|---|---|
| Nicotine Degradation II | 0 | 0.0154 | #NUM! | CSGALNACT1 |
| Superpathway of Inositol Phosphate Compounds | 0 | 0.0201 | -2 | INPP5D,PLCB1,PLCB2,PPFIA2 |
| Serotonin Degradation | 0 | 0.0149 | #NUM! | CSGALNACT1 |
| D-myo-inositol (3,4,5,6)-tetrakisphosphate Biosynthesis | 0 | 0.00704 | #NUM! | PPFIA2 |
| 3-phosphoinositide Degradation | 0 | 0.0128 | #NUM! | INPP5D,PPFIA2 |
| 3-phosphoinositide Biosynthesis | 0 | 0.00602 | #NUM! | PPFIA2 |
| Superpathway of Melatonin Degradation | 0 | 0.0154 | #NUM! | CSGALNACT1 |
| Stearate Biosynthesis I (Animals) | 0 | 0.0204 | #NUM! | ACSL1 |
| Agranulocyte Adhesion and Diapedesis | 0 | 0.0155 | #NUM! | IL1B,MMP16,TNFRSF1A |
| Cell Cycle: G2/M DNA Damage Checkpoint Regulation | 0 | 0.0204 | #NUM! | CDKN1A |
| Serotonin Receptor Signaling | 0 | 0.0233 | #NUM! | ADCY2 |
| Autophagy | 0 | 0.0164 | #NUM! | CTSC |
| Cancer Drug Resistance By Drug Efflux | 0 | 0.0172 | #NUM! | AKT3 |
| NER Pathway | 0 | 0.00971 | #NUM! | H3-3A/H3-3B |
| SPINK1 General Cancer Pathway | 0 | 0.0145 | #NUM! | AKT3 |
| Regulation Of The Epithelial Mesenchymal Transition In | 0 | 0.0119 | #NUM! | RBPJ |
| MSP-RON Signaling In Cancer Cells Pathway | 0 | 0.0224 | #NUM! | AKT3,FOS,JUN |

**Supplementary Table 4 Neuronal Marker Genes**

| | p_val | avg_log2FC | pct.1 | pct.2 | p_val_adj | cluster | gene |
|---|---|---|---|---|---|---|---|
| CBLN2 | 0 | 2.51986753 | 0.973 | 0.286 | 0 | ExN1 | CBLN2 |
| LINC02306 | 0 | 2.17302819 | 0.733 | 0.14 | 0 | ExN1 | LINC02306 |
| PDZD2 | 0 | 2.13617429 | 0.985 | 0.571 | 0 | ExN1 | PDZD2 |
| AC011287.1 | 0 | 1.99463208 | 0.923 | 0.353 | 0 | ExN1 | AC011287.1 |
| LINC01378 | 0 | 1.77777407 | 0.848 | 0.216 | 0 | ExN1 | LINC01378 |
| MEIS2 | 0 | 1.63419183 | 0.878 | 0.173 | 0 | ExN1 | MEIS2 |
| HS6ST3 | 0 | 1.57325141 | 0.992 | 0.683 | 0 | ExN1 | HS6ST3 |
| DGKB | 0 | 1.56556774 | 0.988 | 0.76 | 0 | ExN1 | DGKB |
| ENC1 | 0 | 1.53050935 | 0.903 | 0.433 | 0 | ExN1 | ENC1 |
| AC093607.1 | 0 | 1.51922348 | 0.552 | 0.095 | 0 | ExN1 | AC093607.1 |
| CUX2 | 0 | 1.44612041 | 0.926 | 0.254 | 0 | ExN1 | CUX2 |
| AC021613.1 | 0 | 1.41360309 | 0.726 | 0.165 | 0 | ExN1 | AC021613.1 |
| LINC00507 | 0 | 1.39040191 | 0.717 | 0.079 | 0 | ExN1 | LINC00507 |
| TESPA1 | 0 | 1.34883201 | 0.832 | 0.218 | 0 | ExN1 | TESPA1 |
| LINC00276 | 0 | 1.31446264 | 0.59 | 0.138 | 0 | ExN1 | LINC00276 |
| AC013287.1 | 0 | 1.31168905 | 0.759 | 0.369 | 0 | ExN1 | AC013287.1 |
| CYP1B1-AS1 | 0 | 1.30671021 | 0.765 | 0.171 | 0 | ExN1 | CYP1B1-AS1 |
| AC008574.1 | 0 | 1.28731536 | 0.655 | 0.111 | 0 | ExN1 | AC008574.1 |
| LINC01331 | 0 | 1.28072256 | 0.625 | 0.131 | 0 | ExN1 | LINC01331 |
| AC110023.1 | 0 | 1.26394459 | 0.695 | 0.208 | 0 | ExN1 | AC110023.1 |
| LINC01250 | 0 | 1.26026179 | 0.932 | 0.569 | 0 | ExN1 | LINC01250 |
| CDH12 | 0 | 1.23121924 | 0.981 | 0.617 | 0 | ExN1 | CDH12 |
| LINC02263 | 0 | 1.23088714 | 0.531 | 0.121 | 0 | ExN1 | LINC02263 |
| GREB1L | 0 | 1.22562763 | 0.709 | 0.21 | 0 | ExN1 | GREB1L |
| SLIT3 | 0 | 1.21583256 | 0.926 | 0.347 | 0 | ExN1 | SLIT3 |
| PTK2B | 0 | 1.18684735 | 0.893 | 0.423 | 0 | ExN1 | PTK2B |
| GRIA4 | 0 | 1.17370891 | 0.988 | 0.807 | 0 | ExN1 | GRIA4 |
| SERPINE2 | 0 | 1.16264312 | 0.679 | 0.111 | 0 | ExN1 | SERPINE2 |
| AC091078.1 | 0 | 1.15208492 | 0.677 | 0.209 | 0 | ExN1 | AC091078.1 |
| TAFA1 | 0 | 1.14834989 | 0.875 | 0.424 | 0 | ExN1 | TAFA1 |
| LRRTM4 | 0 | 1.12958864 | 0.997 | 0.901 | 0 | ExN1 | LRRTM4 |
| CHN1 | 0 | 1.11870975 | 0.969 | 0.694 | 0 | ExN1 | CHN1 |
| RASGRF2 | 0 | 1.11723727 | 0.933 | 0.521 | 0 | ExN1 | RASGRF2 |
| NDST3 | 0 | 1.11698217 | 0.892 | 0.471 | 0 | ExN1 | NDST3 |
| LINC01122 | 0 | 1.11324571 | 0.954 | 0.622 | 0 | ExN1 | LINC01122 |
| ZFPM2 | 0 | 1.11093231 | 0.958 | 0.631 | 0 | ExN1 | ZFPM2 |
| DIAPH2 | 0 | 1.10029621 | 0.9 | 0.535 | 0 | ExN1 | DIAPH2 |
| KIAA1211L | 0 | 1.0997974 | 0.922 | 0.472 | 0 | ExN1 | KIAA1211L |
| AC011246.1 | 0 | 1.08277571 | 0.575 | 0.156 | 0 | ExN1 | AC011246.1 |
| MLIP | 0 | 1.06363943 | 0.868 | 0.422 | 0 | ExN1 | MLIP |
| LY86-AS1 | 0 | 1.05499662 | 0.825 | 0.397 | 0 | ExN1 | LY86-AS1 |
| MAN1A1 | 0 | 1.05111258 | 0.818 | 0.402 | 0 | ExN1 | MAN1A1 |
| HHAT | 0 | 1.05104621 | 0.555 | 0.197 | 0 | ExN1 | HHAT |
| ADAMTS3 | 0 | 1.03576293 | 0.657 | 0.165 | 0 | ExN1 | ADAMTS3 |
| CDH9 | 0 | 1.03267836 | 0.927 | 0.539 | 0 | ExN1 | CDH9 |
| TTC6 | 0 | 1.02675255 | 0.536 | 0.083 | 0 | ExN1 | TTC6 |
| AL117329.1 | 0 | 1.00776942 | 0.829 | 0.379 | 0 | ExN1 | AL117329.1 |
| CDH22 | 0 | 1.00432014 | 0.79 | 0.303 | 0 | ExN1 | CDH22 |
| CDH18 | 0 | 0.99948255 | 0.986 | 0.841 | 0 | ExN1 | CDH18 |
| NLK | 0 | 0.99179569 | 0.932 | 0.637 | 0 | ExN1 | NLK |
| SYN3 | 0 | 0.98841468 | 0.99 | 0.833 | 0 | ExN1 | SYN3 |
| PDGFD | 0 | 0.98689411 | 0.587 | 0.089 | 0 | ExN1 | PDGFD |
| EPHA6 | 0 | 0.97569398 | 0.923 | 0.592 | 0 | ExN1 | EPHA6 |
| KIF13A | 0 | 0.96894235 | 0.844 | 0.499 | 0 | ExN1 | KIF13A |
| DGKG | 0 | 0.96747579 | 0.895 | 0.44 | 0 | ExN1 | DGKG |
| RCAN2 | 0 | 0.96383564 | 0.934 | 0.729 | 0 | ExN1 | RCAN2 |
| CA10 | 0 | 0.95268272 | 0.831 | 0.337 | 0 | ExN1 | CA10 |
| PPFIA2 | 0 | 0.95203125 | 0.988 | 0.908 | 0 | ExN1 | PPFIA2 |
| KCTD1 | 0 | 0.94345781 | 0.926 | 0.606 | 0 | ExN1 | KCTD1 |
| RAPGEF5 | 0 | 0.94103071 | 0.967 | 0.695 | 0 | ExN1 | RAPGEF5 |
| DPYD | 0 | 0.93990284 | 0.924 | 0.599 | 0 | ExN1 | DPYD |

| | | | | | | | |
|---|---|---|---|---|---|---|---|
| SORBS2 | 0 | 0.93660304 | 0.976 | 0.809 | 0 | ExN1 | SORBS2 |
| HOPX | 0 | 0.92595956 | 0.67 | 0.203 | 0 | ExN1 | HOPX |
| AL591501.1 | 0 | 0.92571221 | 0.854 | 0.498 | 0 | ExN1 | AL591501.1 |
| LINC01500 | 0 | 0.91981307 | 0.385 | 0.065 | 0 | ExN1 | LINC01500 |
| AP000829.1 | 0 | 0.91828184 | 0.554 | 0.122 | 0 | ExN1 | AP000829.1 |
| PDE8B | 0 | 0.90949312 | 0.917 | 0.596 | 0 | ExN1 | PDE8B |
| KCNIP4 | 0 | 0.90881705 | 1 | 0.922 | 0 | ExN1 | KCNIP4 |
| SMAD3 | 0 | 0.90696342 | 0.753 | 0.282 | 0 | ExN1 | SMAD3 |
| ATP2B1 | 0 | 0.90511641 | 0.973 | 0.816 | 0 | ExN1 | ATP2B1 |
| RFX3 | 0 | 0.89204879 | 0.937 | 0.765 | 0 | ExN1 | RFX3 |
| SIPA1L1 | 0 | 0.89146629 | 0.988 | 0.898 | 0 | ExN1 | SIPA1L1 |
| HPCAL1 | 0 | 0.88622551 | 0.675 | 0.194 | 0 | ExN1 | HPCAL1 |
| BTBD9 | 0 | 0.88320668 | 0.964 | 0.803 | 0 | ExN1 | BTBD9 |
| ST6GALNAC5 | 0 | 0.87761674 | 0.96 | 0.648 | 0 | ExN1 | ST6GALNAC5 |
| PDE7B | 0 | 0.87701088 | 0.817 | 0.394 | 0 | ExN1 | PDE7B |
| CAMK2A | 0 | 0.87397713 | 0.959 | 0.753 | 0 | ExN1 | CAMK2A |
| PVT1 | 0 | 0.86623547 | 0.792 | 0.35 | 0 | ExN1 | PVT1 |
| EPB41L2 | 0 | 0.8624195 | 0.927 | 0.656 | 0 | ExN1 | EPB41L2 |
| ITPR1 | 0 | 0.85375267 | 0.955 | 0.685 | 0 | ExN1 | ITPR1 |
| R3HDM1 | 0 | 0.85370325 | 0.987 | 0.869 | 0 | ExN1 | R3HDM1 |
| AL049828.1 | 0 | 0.85076962 | 0.81 | 0.602 | 0 | ExN1 | AL049828.1 |
| PTPRR | 0 | 0.84026171 | 0.901 | 0.641 | 0 | ExN1 | PTPRR |
| AC005162.3 | 0 | 0.83908192 | 0.592 | 0.143 | 0 | ExN1 | AC005162.3 |
| NGEF | 0 | 0.83733852 | 0.79 | 0.384 | 0 | ExN1 | NGEF |
| CAMK2N1 | 0 | 0.83522366 | 0.873 | 0.598 | 0 | ExN1 | CAMK2N1 |
| AL353604.1 | 0 | 0.83113283 | 0.507 | 0.074 | 0 | ExN1 | AL353604.1 |
| NTRK3 | 0 | 0.82862951 | 0.983 | 0.835 | 0 | ExN1 | NTRK3 |
| P2RX7 | 0 | 0.82049798 | 0.76 | 0.372 | 0 | ExN1 | P2RX7 |
| PSD3 | 0 | 0.81925035 | 0.935 | 0.685 | 0 | ExN1 | PSD3 |
| SLC24A4 | 0 | 0.81685297 | 0.755 | 0.243 | 0 | ExN1 | SLC24A4 |
| TANC2 | 0 | 0.8112738 | 0.966 | 0.835 | 0 | ExN1 | TANC2 |
| LAMP5 | 0 | 0.81066044 | 0.535 | 0.08 | 0 | ExN1 | LAMP5 |
| LINC00326 | 0 | 0.80708251 | 0.323 | 0.06 | 0 | ExN1 | LINC00326 |
| NECTIN3 | 0 | 0.80583915 | 0.76 | 0.408 | 0 | ExN1 | NECTIN3 |
| SV2B | 0 | 0.80537747 | 0.942 | 0.521 | 0 | ExN1 | SV2B |
| LDB2 | 0 | 0.80308763 | 0.985 | 0.707 | 0 | ExN1 | LDB2 |
| KCNK1 | 0 | 0.80259618 | 0.811 | 0.447 | 0 | ExN1 | KCNK1 |
| PTCHD1-AS | 0 | 0.79955386 | 0.734 | 0.378 | 0 | ExN1 | PTCHD1-AS |
| GNAL | 0 | 0.79096695 | 0.818 | 0.388 | 0 | ExN1 | GNAL |
| AL391840.1 | 0 | 0.78518211 | 0.755 | 0.45 | 0 | ExN1 | AL391840.1 |
| RGS6 | 0 | 0.78490583 | 0.981 | 0.63 | 0 | ExN1 | RGS6 |
| AQP4-AS1 | 0 | 0.78484825 | 0.796 | 0.426 | 0 | ExN1 | AQP4-AS1 |
| KCND2 | 0 | 0.78472454 | 0.981 | 0.844 | 0 | ExN1 | KCND2 |
| AC110614.1 | 0 | 0.78056598 | 0.372 | 0.137 | 0 | ExN1 | AC110614.1 |
| SATB2 | 0 | 0.77854021 | 0.923 | 0.487 | 0 | ExN1 | SATB2 |
| AC097512.1 | 0 | 0.77838174 | 0.382 | 0.052 | 0 | ExN1 | AC097512.1 |
| RFTN1 | 0 | 0.77756402 | 0.788 | 0.4 | 0 | ExN1 | RFTN1 |
| MAP3K5 | 0 | 0.77386422 | 0.911 | 0.57 | 0 | ExN1 | MAP3K5 |
| ARPP19 | 0 | 0.76845887 | 0.771 | 0.461 | 0 | ExN1 | ARPP19 |
| NRGN | 0 | 0.76665686 | 0.882 | 0.494 | 0 | ExN1 | NRGN |
| NECAB1 | 0 | 0.76358498 | 0.93 | 0.57 | 0 | ExN1 | NECAB1 |
| USP25 | 0 | 0.75854318 | 0.868 | 0.614 | 0 | ExN1 | USP25 |
| FLRT2 | 0 | 0.75596696 | 0.969 | 0.801 | 0 | ExN1 | FLRT2 |
| KCTD16 | 0 | 0.75539749 | 0.985 | 0.836 | 0 | ExN1 | KCTD16 |
| AC009041.2 | 0 | 0.75107568 | 0.595 | 0.251 | 0 | ExN1 | AC009041.2 |
| ADGRL3 | 0 | 0.74916788 | 0.994 | 0.955 | 0 | ExN1 | ADGRL3 |
| ART3 | 0 | 0.7440811 | 0.551 | 0.143 | 0 | ExN1 | ART3 |
| AL157944.1 | 0 | 0.74354389 | 0.446 | 0.235 | 0 | ExN1 | AL157944.1 |
| LMO3 | 0 | 0.74013328 | 0.862 | 0.526 | 0 | ExN1 | LMO3 |
| IQCJ-SCHIP1 | 0 | 0.73457779 | 0.99 | 0.794 | 0 | ExN1 | IQCJ-SCHIP1 |
| LAMB1 | 0 | 0.73224181 | 0.606 | 0.198 | 0 | ExN1 | LAMB1 |
| DLEU2 | 0 | 0.72272712 | 0.796 | 0.497 | 0 | ExN1 | DLEU2 |
| VSNL1 | 0 | 0.72146141 | 0.938 | 0.737 | 0 | ExN1 | VSNL1 |

| | | | | | | | |
|---|---|---|---|---|---|---|---|
| LNX1 | 0 | 0.72106965 | 0.843 | 0.539 | 0 | ExN1 | LNX1 |
| NIPAL2 | 0 | 0.71941448 | 0.789 | 0.463 | 0 | ExN1 | NIPAL2 |
| CACNA2D1 | 0 | 0.71912339 | 0.972 | 0.722 | 0 | ExN1 | CACNA2D1 |
| CELF2 | 0 | 0.71706961 | 0.996 | 0.961 | 0 | ExN1 | CELF2 |
| FBXW7 | 0 | 0.71569901 | 0.925 | 0.713 | 0 | ExN1 | FBXW7 |
| COL24A1 | 0 | 0.71552871 | 0.553 | 0.16 | 0 | ExN1 | COL24A1 |
| MPPED2 | 0 | 0.7038848 | 0.791 | 0.411 | 0 | ExN1 | MPPED2 |
| AL008633.1 | 0 | 0.70357475 | 0.84 | 0.684 | 0 | ExN1 | AL008633.1 |
| LIN7A | 0 | 0.70060795 | 0.802 | 0.476 | 0 | ExN1 | LIN7A |
| STXBP6 | 0 | 0.69533235 | 0.747 | 0.291 | 0 | ExN1 | STXBP6 |
| GRM1 | 0 | 0.68928811 | 0.907 | 0.57 | 0 | ExN1 | GRM1 |
| PHYHIP | 0 | 0.68878656 | 0.734 | 0.418 | 0 | ExN1 | PHYHIP |
| AL136441.1 | 0 | 0.68664115 | 0.365 | 0.094 | 0 | ExN1 | AL136441.1 |
| KLHL2 | 0 | 0.67975235 | 0.756 | 0.434 | 0 | ExN1 | KLHL2 |
| CAMKK1 | 0 | 0.67926837 | 0.725 | 0.382 | 0 | ExN1 | CAMKK1 |
| PHACTR1 | 0 | 0.67859916 | 0.993 | 0.942 | 0 | ExN1 | PHACTR1 |
| AC007563.2 | 0 | 0.67802006 | 0.862 | 0.544 | 0 | ExN1 | AC007563.2 |
| RPS6KC1 | 0 | 0.67755182 | 0.925 | 0.73 | 0 | ExN1 | RPS6KC1 |
| SYNJ2 | 0 | 0.67508498 | 0.697 | 0.313 | 0 | ExN1 | SYNJ2 |
| PTPRO | 0 | 0.67264921 | 0.899 | 0.623 | 0 | ExN1 | PTPRO |
| BAIAP2 | 0 | 0.67133454 | 0.776 | 0.435 | 0 | ExN1 | BAIAP2 |
| MBNL2 | 0 | 0.66606786 | 0.952 | 0.717 | 0 | ExN1 | MBNL2 |
| ATP2B4 | 0 | 0.66457912 | 0.806 | 0.442 | 0 | ExN1 | ATP2B4 |
| PTH2R | 0 | 0.66270045 | 0.53 | 0.193 | 0 | ExN1 | PTH2R |
| AC069133.1 | 0 | 0.65830824 | 0.461 | 0.119 | 0 | ExN1 | AC069133.1 |
| WASF1 | 0 | 0.65322229 | 0.851 | 0.588 | 0 | ExN1 | WASF1 |
| TYRO3 | 0 | 0.65257192 | 0.651 | 0.277 | 0 | ExN1 | TYRO3 |
| NELL1 | 0 | 0.65160946 | 0.892 | 0.591 | 0 | ExN1 | NELL1 |
| PTK2 | 0 | 0.65010437 | 0.975 | 0.879 | 0 | ExN1 | PTK2 |
| TMEM108 | 0 | 0.64668388 | 0.958 | 0.784 | 0 | ExN1 | TMEM108 |
| CEMIP2 | 0 | 0.64638831 | 0.654 | 0.273 | 0 | ExN1 | CEMIP2 |
| ACVR1C | 0 | 0.64458754 | 0.396 | 0.06 | 0 | ExN1 | ACVR1C |
| SORBS1 | 0 | 0.64126295 | 0.968 | 0.87 | 0 | ExN1 | SORBS1 |
| SNTB2 | 0 | 0.63999886 | 0.709 | 0.378 | 0 | ExN1 | SNTB2 |
| ACTN1 | 0 | 0.6395763 | 0.818 | 0.529 | 0 | ExN1 | ACTN1 |
| GRIA2 | 0 | 0.6395196 | 0.988 | 0.941 | 0 | ExN1 | GRIA2 |
| DGCR9 | 0 | 0.63854099 | 0.796 | 0.578 | 0 | ExN1 | DGCR9 |
| FBXO32 | 0 | 0.6378041 | 0.703 | 0.381 | 0 | ExN1 | FBXO32 |
| UPP2 | 0 | 0.63246796 | 0.759 | 0.479 | 0 | ExN1 | UPP2 |
| ITGA9 | 0 | 0.63029127 | 0.746 | 0.351 | 0 | ExN1 | ITGA9 |
| FAM153CP | 0 | 0.62857572 | 0.785 | 0.594 | 0 | ExN1 | FAM153CP |
| NHSL2 | 0 | 0.62670073 | 0.581 | 0.215 | 0 | ExN1 | NHSL2 |
| KLHL4 | 0 | 0.62284041 | 0.56 | 0.22 | 0 | ExN1 | KLHL4 |
| CHRM3 | 0 | 0.62085901 | 0.985 | 0.849 | 0 | ExN1 | CHRM3 |
| MED12L | 0 | 0.61863788 | 0.893 | 0.663 | 0 | ExN1 | MED12L |
| PRAG1 | 0 | 0.6181854 | 0.52 | 0.141 | 0 | ExN1 | PRAG1 |
| ITPKA | 0 | 0.61627021 | 0.502 | 0.117 | 0 | ExN1 | ITPKA |
| FSTL4 | 0 | 0.61540493 | 0.934 | 0.575 | 0 | ExN1 | FSTL4 |
| PTPN13 | 0 | 0.61494906 | 0.606 | 0.343 | 0 | ExN1 | PTPN13 |
| GLRA3 | 0 | 0.61231366 | 0.59 | 0.229 | 0 | ExN1 | GLRA3 |
| PPP4R4 | 0 | 0.60983372 | 0.676 | 0.332 | 0 | ExN1 | PPP4R4 |
| MIR137HG | 0 | 0.60837961 | 0.888 | 0.603 | 0 | ExN1 | MIR137HG |
| BRINP2 | 0 | 0.60731634 | 0.845 | 0.595 | 0 | ExN1 | BRINP2 |
| ZNF365 | 0 | 0.60727707 | 0.71 | 0.405 | 0 | ExN1 | ZNF365 |
| ARHGAP10 | 0 | 0.60501768 | 0.578 | 0.233 | 0 | ExN1 | ARHGAP10 |
| MAST3 | 0 | 0.60225458 | 0.684 | 0.353 | 0 | ExN1 | MAST3 |
| GABRA2 | 0 | 0.59668981 | 0.833 | 0.587 | 0 | ExN1 | GABRA2 |
| ANKRD33B | 0 | 0.59502539 | 0.712 | 0.364 | 0 | ExN1 | ANKRD33B |
| NWD2 | 0 | 0.59461399 | 0.732 | 0.399 | 0 | ExN1 | NWD2 |
| TMOD1 | 0 | 0.59407694 | 0.745 | 0.416 | 0 | ExN1 | TMOD1 |
| ELL2 | 0 | 0.58761972 | 0.78 | 0.486 | 0 | ExN1 | ELL2 |
| ATRNL1 | 0 | 0.58704685 | 0.99 | 0.915 | 0 | ExN1 | ATRNL1 |
| EML1 | 0 | 0.58661868 | 0.629 | 0.303 | 0 | ExN1 | EML1 |

| | | | | | | | |
|---|---|---|---|---|---|---|---|
| TMEM59L | 0 | 0.58601762 | 0.771 | 0.522 | 0 | ExN1 | TMEM59L |
| PLEKHA5 | 0 | 0.58555816 | 0.97 | 0.906 | 0 | ExN1 | PLEKHA5 |
| NIN | 0 | 0.58294256 | 0.904 | 0.661 | 0 | ExN1 | NIN |
| AK5 | 0 | 0.58100119 | 0.949 | 0.738 | 0 | ExN1 | AK5 |
| ZBTB7C | 0 | 0.58070342 | 0.551 | 0.217 | 0 | ExN1 | ZBTB7C |
| LZTS1 | 0 | 0.57730787 | 0.516 | 0.149 | 0 | ExN1 | LZTS1 |
| KCNIP2 | 0 | 0.57614005 | 0.638 | 0.28 | 0 | ExN1 | KCNIP2 |
| CNTNAP5 | 0 | 0.57576075 | 0.976 | 0.881 | 0 | ExN1 | CNTNAP5 |
| PCDH7 | 0 | 0.57193258 | 0.984 | 0.785 | 0 | ExN1 | PCDH7 |
| MCHR2 | 0 | 0.57099766 | 0.533 | 0.208 | 0 | ExN1 | MCHR2 |
| AC007091.1 | 0 | 0.56968828 | 0.283 | 0.061 | 0 | ExN1 | AC007091.1 |
| ARNT2 | 0 | 0.567433 | 0.942 | 0.811 | 0 | ExN1 | ARNT2 |
| COL27A1 | 0 | 0.56463763 | 0.441 | 0.088 | 0 | ExN1 | COL27A1 |
| ARFGEF3 | 0 | 0.56250382 | 0.91 | 0.744 | 0 | ExN1 | ARFGEF3 |
| CKB | 0 | 0.56173981 | 0.778 | 0.576 | 0 | ExN1 | CKB |
| AC013652.1 | 0 | 0.5541641 | 0.53 | 0.287 | 0 | ExN1 | AC013652.1 |
| SH3RF2 | 0 | 0.55070372 | 0.396 | 0.059 | 0 | ExN1 | SH3RF2 |
| PPP3CA | 0 | 0.54993102 | 0.984 | 0.898 | 0 | ExN1 | PPP3CA |
| RTN4 | 0 | 0.54907006 | 0.914 | 0.795 | 0 | ExN1 | RTN4 |
| TLN2 | 0 | 0.54790128 | 0.902 | 0.693 | 0 | ExN1 | TLN2 |
| ADGRL2 | 0 | 0.54789533 | 0.974 | 0.825 | 0 | ExN1 | ADGRL2 |
| CDC40 | 0 | 0.54744674 | 0.699 | 0.418 | 0 | ExN1 | CDC40 |
| MYOM2 | 0 | 0.54638926 | 0.459 | 0.142 | 0 | ExN1 | MYOM2 |
| CAMKK2 | 0 | 0.54282125 | 0.706 | 0.397 | 0 | ExN1 | CAMKK2 |
| MFSD4A | 0 | 0.54180268 | 0.592 | 0.279 | 0 | ExN1 | MFSD4A |
| DDAH1 | 0 | 0.54022806 | 0.674 | 0.359 | 0 | ExN1 | DDAH1 |
| CTDSPL | 0 | 0.5398142 | 0.869 | 0.666 | 0 | ExN1 | CTDSPL |
| OLFM1 | 0 | 0.53952159 | 0.827 | 0.563 | 0 | ExN1 | OLFM1 |
| MIR222HG | 0 | 0.53510755 | 0.566 | 0.25 | 0 | ExN1 | MIR222HG |
| KIAA2012 | 0 | 0.53440601 | 0.451 | 0.111 | 0 | ExN1 | KIAA2012 |
| TMEM132D | 0 | 0.53372254 | 0.963 | 0.735 | 0 | ExN1 | TMEM132D |
| ARL4A | 0 | 0.53313836 | 0.418 | 0.1 | 0 | ExN1 | ARL4A |
| PKIG | 0 | 0.53118291 | 0.732 | 0.457 | 0 | ExN1 | PKIG |
| MEG3 | 0 | 0.5311511 | 0.971 | 0.957 | 0 | ExN1 | MEG3 |
| VWA3A | 0 | 0.52827634 | 0.498 | 0.182 | 0 | ExN1 | VWA3A |
| FAM163A | 0 | 0.52800624 | 0.32 | 0.03 | 0 | ExN1 | FAM163A |
| SEMA5B | 0 | 0.52790128 | 0.576 | 0.223 | 0 | ExN1 | SEMA5B |
| STXBP5L | 0 | 0.52737891 | 0.986 | 0.938 | 0 | ExN1 | STXBP5L |
| SLC12A8 | 0 | 0.52731396 | 0.548 | 0.228 | 0 | ExN1 | SLC12A8 |
| NPTX1 | 0 | 0.52436363 | 0.738 | 0.42 | 0 | ExN1 | NPTX1 |
| ATP6V1H | 0 | 0.52326961 | 0.813 | 0.617 | 0 | ExN1 | ATP6V1H |
| AC106795.3 | 0 | 0.52110009 | 0.491 | 0.149 | 0 | ExN1 | AC106795.3 |
| SGK3 | 0 | 0.51947022 | 0.8 | 0.563 | 0 | ExN1 | SGK3 |
| ICAM5 | 0 | 0.51927072 | 0.516 | 0.203 | 0 | ExN1 | ICAM5 |
| ADAMTS16 | 0 | 0.518396 | 0.437 | 0.141 | 0 | ExN1 | ADAMTS16 |
| SNED1 | 0 | 0.51810481 | 0.688 | 0.381 | 0 | ExN1 | SNED1 |
| SGCD | 0 | 0.51803107 | 0.779 | 0.543 | 0 | ExN1 | SGCD |
| CCDC88C | 0 | 0.51783783 | 0.361 | 0.034 | 0 | ExN1 | CCDC88C |
| PRKAR2B | 0 | 0.51537138 | 0.821 | 0.57 | 0 | ExN1 | PRKAR2B |
| SYBU | 0 | 0.51413769 | 0.878 | 0.672 | 0 | ExN1 | SYBU |
| HRH2 | 0 | 0.51367314 | 0.627 | 0.33 | 0 | ExN1 | HRH2 |
| BRINP1 | 0 | 0.5125206 | 0.904 | 0.703 | 0 | ExN1 | BRINP1 |
| ARHGAP32 | 0 | 0.51211392 | 0.951 | 0.821 | 0 | ExN1 | ARHGAP32 |
| KALRN | 0 | 0.51131357 | 0.994 | 0.948 | 0 | ExN1 | KALRN |
| EDIL3 | 0 | 0.51091206 | 0.91 | 0.684 | 0 | ExN1 | EDIL3 |
| FAT3 | 0 | 0.51089845 | 0.924 | 0.659 | 0 | ExN1 | FAT3 |
| MBNL1 | 0 | 0.51027816 | 0.905 | 0.746 | 0 | ExN1 | MBNL1 |
| TBC1D1 | 0 | 0.51001233 | 0.787 | 0.519 | 0 | ExN1 | TBC1D1 |
| CNKSR2 | 0 | 0.50883541 | 0.958 | 0.785 | 0 | ExN1 | CNKSR2 |
| PDE1A | 0 | 0.50695515 | 0.95 | 0.611 | 0 | ExN1 | PDE1A |
| GALNT13 | 0 | 0.50673576 | 0.892 | 0.63 | 0 | ExN1 | GALNT13 |
| CDK19 | 0 | 0.50631153 | 0.776 | 0.506 | 0 | ExN1 | CDK19 |
| ZDHHC14 | 0 | 0.50587381 | 0.917 | 0.74 | 0 | ExN1 | ZDHHC14 |

| | | | | | | | |
|---|---|---|---|---|---|---|---|
| ST8SIA3 | 0 | 0.50507752 | 0.751 | 0.503 | 0 | ExN1 | ST8SIA3 |
| EML6 | 0 | 0.50039637 | 0.92 | 0.745 | 0 | ExN1 | EML6 |
| CTNNAL1 | 0 | 0.4993283 | 0.674 | 0.381 | 0 | ExN1 | CTNNAL1 |
| LINC01484 | 0 | 0.49929853 | 0.326 | 0.02 | 0 | ExN1 | LINC01484 |
| GSG1L | 0 | 0.49919501 | 0.584 | 0.235 | 0 | ExN1 | GSG1L |
| AC068286.2 | 0 | 0.49818344 | 0.319 | 0.082 | 0 | ExN1 | AC068286.2 |
| DLGAP2 | 0 | 0.49691093 | 0.992 | 0.915 | 0 | ExN1 | DLGAP2 |
| LONRF2 | 0 | 0.49645949 | 0.816 | 0.61 | 0 | ExN1 | LONRF2 |
| SESN1 | 0 | 0.49624136 | 0.82 | 0.603 | 0 | ExN1 | SESN1 |
| CIT | 0 | 0.49592616 | 0.909 | 0.711 | 0 | ExN1 | CIT |
| NOVA1 | 0 | 0.49524331 | 0.924 | 0.783 | 0 | ExN1 | NOVA1 |
| SNAP25 | 0 | 0.49378759 | 0.977 | 0.907 | 0 | ExN1 | SNAP25 |
| GALNT18 | 0 | 0.49354819 | 0.909 | 0.702 | 0 | ExN1 | GALNT18 |
| ARMC9 | 0 | 0.49331542 | 0.805 | 0.565 | 0 | ExN1 | ARMC9 |
| HSPA12A | 0 | 0.4933154 | 0.924 | 0.818 | 0 | ExN1 | HSPA12A |
| LRATD1 | 0 | 0.49145218 | 0.433 | 0.138 | 0 | ExN1 | LRATD1 |
| MAL2 | 0 | 0.49134837 | 0.532 | 0.235 | 0 | ExN1 | MAL2 |
| FGFR1OP2 | 0 | 0.49092301 | 0.655 | 0.365 | 0 | ExN1 | FGFR1OP2 |
| MYH9 | 0 | 0.48959291 | 0.546 | 0.239 | 0 | ExN1 | MYH9 |
| CACNA1E | 0 | 0.48955765 | 0.942 | 0.774 | 0 | ExN1 | CACNA1E |
| AC011995.2 | 0 | 0.48865482 | 0.619 | 0.357 | 0 | ExN1 | AC011995.2 |
| SLC4A4 | 0 | 0.48776636 | 0.594 | 0.265 | 0 | ExN1 | SLC4A4 |
| TMEM178B | 0 | 0.48548491 | 0.978 | 0.905 | 0 | ExN1 | TMEM178B |
| PRKCB | 0 | 0.48477218 | 0.977 | 0.84 | 0 | ExN1 | PRKCB |
| FAM153B | 0 | 0.48426631 | 0.609 | 0.37 | 0 | ExN1 | FAM153B |
| ACTB | 0 | 0.48355351 | 0.78 | 0.603 | 0 | ExN1 | ACTB |
| FAM160A1 | 0 | 0.48133286 | 0.321 | 0.072 | 0 | ExN1 | FAM160A1 |
| SEMA3C | 0 | 0.48010495 | 0.378 | 0.077 | 0 | ExN1 | SEMA3C |
| FBXO34 | 0 | 0.4800424 | 0.792 | 0.578 | 0 | ExN1 | FBXO34 |
| MOXD1 | 0 | 0.47799405 | 0.349 | 0.094 | 0 | ExN1 | MOXD1 |
| CACNG3 | 0 | 0.47504716 | 0.847 | 0.604 | 0 | ExN1 | CACNG3 |
| ADAM23 | 0 | 0.4742423 | 0.914 | 0.732 | 0 | ExN1 | ADAM23 |
| AC034268.2 | 0 | 0.47330032 | 0.471 | 0.244 | 0 | ExN1 | AC034268.2 |
| AC092902.4 | 0 | 0.47205522 | 0.447 | 0.17 | 0 | ExN1 | AC092902.4 |
| COL26A1 | 0 | 0.47122884 | 0.701 | 0.423 | 0 | ExN1 | COL26A1 |
| EPB41L3 | 0 | 0.4700297 | 0.908 | 0.749 | 0 | ExN1 | EPB41L3 |
| LRFN5 | 0 | 0.46729374 | 0.978 | 0.87 | 0 | ExN1 | LRFN5 |
| DCAF6 | 0 | 0.46719528 | 0.789 | 0.582 | 0 | ExN1 | DCAF6 |
| KCNH3 | 0 | 0.46716504 | 0.613 | 0.345 | 0 | ExN1 | KCNH3 |
| PLEKHA2 | 0 | 0.46640593 | 0.487 | 0.186 | 0 | ExN1 | PLEKHA2 |
| BMPR1B | 0 | 0.46581647 | 0.284 | 0.055 | 0 | ExN1 | BMPR1B |
| SEC14L5 | 0 | 0.46579203 | 0.682 | 0.391 | 0 | ExN1 | SEC14L5 |
| KCNIP4-IT1 | 0 | 0.46573854 | 0.702 | 0.451 | 0 | ExN1 | KCNIP4-IT1 |
| NFAT5 | 0 | 0.46503084 | 0.887 | 0.743 | 0 | ExN1 | NFAT5 |
| COX7A2L | 0 | 0.46454324 | 0.61 | 0.344 | 0 | ExN1 | COX7A2L |
| ITGB8 | 0 | 0.46431888 | 0.565 | 0.215 | 0 | ExN1 | ITGB8 |
| CAMK4 | 0 | 0.46229256 | 0.883 | 0.678 | 0 | ExN1 | CAMK4 |
| SH3BGRL2 | 0 | 0.46217509 | 0.732 | 0.486 | 0 | ExN1 | SH3BGRL2 |
| DLGAP1-AS4 | 0 | 0.46172764 | 0.571 | 0.299 | 0 | ExN1 | DLGAP1-AS4 |
| CALN1 | 0 | 0.46145455 | 0.902 | 0.597 | 0 | ExN1 | CALN1 |
| CABP1 | 0 | 0.45972447 | 0.903 | 0.653 | 0 | ExN1 | CABP1 |
| PRXL2A | 0 | 0.45951984 | 0.505 | 0.291 | 0 | ExN1 | PRXL2A |
| IPO5 | 0 | 0.45889982 | 0.633 | 0.372 | 0 | ExN1 | IPO5 |
| ARHGAP26 | 0 | 0.45883926 | 0.966 | 0.847 | 0 | ExN1 | ARHGAP26 |
| RNF115 | 0 | 0.45813077 | 0.767 | 0.546 | 0 | ExN1 | RNF115 |
| EPHX4 | 0 | 0.45727832 | 0.516 | 0.237 | 0 | ExN1 | EPHX4 |
| CNGB1 | 0 | 0.45691411 | 0.343 | 0.069 | 0 | ExN1 | CNGB1 |
| CYP46A1 | 0 | 0.45668107 | 0.612 | 0.373 | 0 | ExN1 | CYP46A1 |
| ANXA11 | 0 | 0.45654273 | 0.605 | 0.321 | 0 | ExN1 | ANXA11 |
| ACAP3 | 0 | 0.45491577 | 0.769 | 0.563 | 0 | ExN1 | ACAP3 |
| ROS1 | 0 | 0.45062117 | 0.293 | 0.083 | 0 | ExN1 | ROS1 |
| SLC17A7 | 0 | 0.44989944 | 0.683 | 0.375 | 0 | ExN1 | SLC17A7 |
| SLF1 | 0 | 0.44972883 | 0.679 | 0.437 | 0 | ExN1 | SLF1 |

| | | | | | | | |
|---|---|---|---|---|---|---|---|
| ACTN4 | 0 | 0.447277 | 0.743 | 0.48 | 0 | ExN1 | ACTN4 |
| GUCY1A2 | 0 | 0.44679846 | 0.904 | 0.686 | 0 | ExN1 | GUCY1A2 |
| GNG2 | 0 | 0.44282748 | 0.7 | 0.378 | 0 | ExN1 | GNG2 |
| NCDN | 0 | 0.44062306 | 0.685 | 0.481 | 0 | ExN1 | NCDN |
| CDK6 | 0 | 0.44034919 | 0.654 | 0.349 | 0 | ExN1 | CDK6 |
| RAPGEF2 | 0 | 0.43939222 | 0.954 | 0.832 | 0 | ExN1 | RAPGEF2 |
| SRPK1 | 0 | 0.43870659 | 0.667 | 0.433 | 0 | ExN1 | SRPK1 |
| GFOD1 | 0 | 0.43863985 | 0.895 | 0.719 | 0 | ExN1 | GFOD1 |
| AC002377.1 | 0 | 0.43838916 | 0.423 | 0.189 | 0 | ExN1 | AC002377.1 |
| UNC5A | 0 | 0.43528965 | 0.659 | 0.405 | 0 | ExN1 | UNC5A |
| AC025887.2 | 0 | 0.43274102 | 0.328 | 0.104 | 0 | ExN1 | AC025887.2 |
| PTPRB | 0 | 0.4313738 | 0.63 | 0.365 | 0 | ExN1 | PTPRB |
| FAM106A | 0 | 0.43133509 | 0.549 | 0.341 | 0 | ExN1 | FAM106A |
| PLXNA2 | 0 | 0.43046977 | 0.86 | 0.624 | 0 | ExN1 | PLXNA2 |
| SEZ6L | 0 | 0.42972911 | 0.903 | 0.681 | 0 | ExN1 | SEZ6L |
| PI4KA | 0 | 0.4293844 | 0.92 | 0.79 | 0 | ExN1 | PI4KA |
| DENND3 | 0 | 0.42773021 | 0.571 | 0.288 | 0 | ExN1 | DENND3 |
| GABRB3 | 0 | 0.42632249 | 0.971 | 0.883 | 0 | ExN1 | GABRB3 |
| ZNF831 | 0 | 0.4251076 | 0.447 | 0.172 | 0 | ExN1 | ZNF831 |
| NDFIP2 | 0 | 0.42493541 | 0.715 | 0.461 | 0 | ExN1 | NDFIP2 |
| MAPK4 | 0 | 0.42319783 | 0.717 | 0.416 | 0 | ExN1 | MAPK4 |
| MAN1C1 | 0 | 0.42241983 | 0.8 | 0.57 | 0 | ExN1 | MAN1C1 |
| DTNB | 0 | 0.42213993 | 0.906 | 0.778 | 0 | ExN1 | DTNB |
| TJP1 | 0 | 0.42054871 | 0.936 | 0.807 | 0 | ExN1 | TJP1 |
| CSMD1 | 0 | 0.41993087 | 0.999 | 0.977 | 0 | ExN1 | CSMD1 |
| TENM2 | 0 | 0.41872163 | 0.992 | 0.919 | 0 | ExN1 | TENM2 |
| NPTX2 | 0 | 0.41805609 | 0.32 | 0.059 | 0 | ExN1 | NPTX2 |
| IL12A-AS1 | 0 | 0.41777376 | 0.379 | 0.133 | 0 | ExN1 | IL12A-AS1 |
| CAMK1D | 0 | 0.41616686 | 0.977 | 0.83 | 0 | ExN1 | CAMK1D |
| FKBP5 | 0 | 0.41305779 | 0.472 | 0.208 | 0 | ExN1 | FKBP5 |
| STXBP5 | 0 | 0.41264837 | 0.922 | 0.804 | 0 | ExN1 | STXBP5 |
| GOLGA8B | 0 | 0.4123546 | 0.664 | 0.464 | 0 | ExN1 | GOLGA8B |
| HTR4 | 0 | 0.41161645 | 0.498 | 0.19 | 0 | ExN1 | HTR4 |
| KCNQ5 | 0 | 0.41156196 | 0.994 | 0.881 | 0 | ExN1 | KCNQ5 |
| WWC1 | 0 | 0.41130985 | 0.572 | 0.3 | 0 | ExN1 | WWC1 |
| DCLK2 | 0 | 0.40865183 | 0.862 | 0.721 | 0 | ExN1 | DCLK2 |
| FANCL | 0 | 0.40798233 | 0.65 | 0.425 | 0 | ExN1 | FANCL |
| ADGRB2 | 0 | 0.40727777 | 0.548 | 0.301 | 0 | ExN1 | ADGRB2 |
| TNFRSF25 | 0 | 0.40636466 | 0.6 | 0.364 | 0 | ExN1 | TNFRSF25 |
| MIR4435-2H | 0 | 0.40612342 | 0.538 | 0.328 | 0 | ExN1 | MIR4435-2H |
| TLE1 | 0 | 0.40606445 | 0.522 | 0.257 | 0 | ExN1 | TLE1 |
| BICC1 | 0 | 0.4032403 | 0.671 | 0.368 | 0 | ExN1 | BICC1 |
| HYDIN | 0 | 0.40253656 | 0.61 | 0.373 | 0 | ExN1 | HYDIN |
| WDR17 | 0 | 0.40175756 | 0.823 | 0.659 | 0 | ExN1 | WDR17 |
| CAMK2B | 0 | 0.40003429 | 0.91 | 0.783 | 0 | ExN1 | CAMK2B |
| ERC2 | 0 | 0.39935306 | 0.967 | 0.922 | 0 | ExN1 | ERC2 |
| PRKCI | 0 | 0.39784291 | 0.673 | 0.454 | 0 | ExN1 | PRKCI |
| VGLL4 | 0 | 0.39747971 | 0.823 | 0.642 | 0 | ExN1 | VGLL4 |
| GPM6B | 0 | 0.39704317 | 0.931 | 0.77 | 0 | ExN1 | GPM6B |
| ABLIM2 | 0 | 0.39647219 | 0.834 | 0.663 | 0 | ExN1 | ABLIM2 |
| PEAK1 | 0 | 0.39575809 | 0.791 | 0.581 | 0 | ExN1 | PEAK1 |
| STYK1 | 0 | 0.39538413 | 0.386 | 0.121 | 0 | ExN1 | STYK1 |
| RASA3 | 0 | 0.39532099 | 0.541 | 0.301 | 0 | ExN1 | RASA3 |
| JAG1 | 0 | 0.39281684 | 0.334 | 0.064 | 0 | ExN1 | JAG1 |
| ARAP2 | 0 | 0.39267997 | 0.841 | 0.542 | 0 | ExN1 | ARAP2 |
| UNC13A | 0 | 0.39145786 | 0.757 | 0.553 | 0 | ExN1 | UNC13A |
| AC073365.1 | 0 | 0.38952273 | 0.625 | 0.32 | 0 | ExN1 | AC073365.1 |
| CSGALNACT1 | 0 | 0.38910955 | 0.842 | 0.552 | 0 | ExN1 | CSGALNACT1 |
| ECHDC2 | 0 | 0.38866966 | 0.515 | 0.284 | 0 | ExN1 | ECHDC2 |
| KCNH1 | 0 | 0.38821612 | 0.949 | 0.801 | 0 | ExN1 | KCNH1 |
| LRRK2 | 0 | 0.38812077 | 0.624 | 0.389 | 0 | ExN1 | LRRK2 |
| RAPH1 | 0 | 0.38802931 | 0.688 | 0.453 | 0 | ExN1 | RAPH1 |
| FAM153A | 0 | 0.38800249 | 0.541 | 0.313 | 0 | ExN1 | FAM153A |

| | | | | | | | |
|---|---|---|---|---|---|---|---|
| PRICKLE1 | 0 | 0.38790729 | 0.856 | 0.65 | 0 | ExN1 | PRICKLE1 |
| AP001993.1 | 0 | 0.38677006 | 0.305 | 0.067 | 0 | ExN1 | AP001993.1 |
| EPB41L4B | 0 | 0.38603072 | 0.759 | 0.552 | 0 | ExN1 | EPB41L4B |
| SLITRK5 | 0 | 0.38533218 | 0.718 | 0.498 | 0 | ExN1 | SLITRK5 |
| KITLG | 0 | 0.38508488 | 0.498 | 0.239 | 0 | ExN1 | KITLG |
| MEF2A | 0 | 0.38274968 | 0.873 | 0.718 | 0 | ExN1 | MEF2A |
| AKAP5 | 0 | 0.38191055 | 0.374 | 0.124 | 0 | ExN1 | AKAP5 |
| GULP1 | 0 | 0.38171307 | 0.624 | 0.282 | 0 | ExN1 | GULP1 |
| CACNB2 | 0 | 0.38169156 | 0.955 | 0.854 | 0 | ExN1 | CACNB2 |
| GAREM1 | 0 | 0.37949786 | 0.669 | 0.418 | 0 | ExN1 | GAREM1 |
| ZNRF3 | 0 | 0.37935826 | 0.615 | 0.361 | 0 | ExN1 | ZNRF3 |
| SLC37A1 | 0 | 0.3791551 | 0.488 | 0.253 | 0 | ExN1 | SLC37A1 |
| PPM1L | 0 | 0.37769358 | 0.92 | 0.781 | 0 | ExN1 | PPM1L |
| MYO5C | 0 | 0.37503802 | 0.483 | 0.227 | 0 | ExN1 | MYO5C |
| SLC39A10 | 0 | 0.3748806 | 0.79 | 0.608 | 0 | ExN1 | SLC39A10 |
| E2F3 | 0 | 0.37348653 | 0.526 | 0.29 | 0 | ExN1 | E2F3 |
| KIRREL3 | 0 | 0.37203931 | 0.98 | 0.743 | 0 | ExN1 | KIRREL3 |
| SLC38A11 | 0 | 0.37189441 | 0.429 | 0.209 | 0 | ExN1 | SLC38A11 |
| PDE4D | 0 | 0.37097576 | 0.994 | 0.953 | 0 | ExN1 | PDE4D |
| EPHA4 | 0 | 0.37000059 | 0.881 | 0.625 | 0 | ExN1 | EPHA4 |
| MIS18BP1 | 0 | 0.36916977 | 0.402 | 0.166 | 0 | ExN1 | MIS18BP1 |
| PALMD | 0 | 0.36884214 | 0.262 | 0.038 | 0 | ExN1 | PALMD |
| DKK3 | 0 | 0.36876868 | 0.789 | 0.594 | 0 | ExN1 | DKK3 |
| AMPH | 0 | 0.36739189 | 0.918 | 0.785 | 0 | ExN1 | AMPH |
| MPP7 | 0 | 0.36718371 | 0.603 | 0.352 | 0 | ExN1 | MPP7 |
| MIAT | 0 | 0.36678813 | 0.966 | 0.86 | 0 | ExN1 | MIAT |
| CNST | 0 | 0.36639536 | 0.645 | 0.443 | 0 | ExN1 | CNST |
| TYW1B | 0 | 0.36582186 | 0.561 | 0.317 | 0 | ExN1 | TYW1B |
| JPH1 | 0 | 0.36568234 | 0.636 | 0.403 | 0 | ExN1 | JPH1 |
| LZTS3 | 0 | 0.36499948 | 0.607 | 0.388 | 0 | ExN1 | LZTS3 |
| ADCY9 | 0 | 0.36424373 | 0.809 | 0.613 | 0 | ExN1 | ADCY9 |
| BCL2 | 0 | 0.36266732 | 0.711 | 0.478 | 0 | ExN1 | BCL2 |
| KCNMA1 | 0 | 0.36208524 | 0.991 | 0.951 | 0 | ExN1 | KCNMA1 |
| NAGPA | 0 | 0.36144609 | 0.478 | 0.254 | 0 | ExN1 | NAGPA |
| MAPK1 | 0 | 0.36074341 | 0.794 | 0.64 | 0 | ExN1 | MAPK1 |
| ATP11B | 0 | 0.36073951 | 0.67 | 0.469 | 0 | ExN1 | ATP11B |
| PDE2A | 0 | 0.36031587 | 0.734 | 0.541 | 0 | ExN1 | PDE2A |
| TMEM241 | 0 | 0.36025122 | 0.677 | 0.454 | 0 | ExN1 | TMEM241 |
| MAP2K1 | 0 | 0.3599525 | 0.663 | 0.447 | 0 | ExN1 | MAP2K1 |
| SLC4A10 | 0 | 0.3596675 | 0.974 | 0.909 | 0 | ExN1 | SLC4A10 |
| ANKRD62 | 0 | 0.35948284 | 0.284 | 0.051 | 0 | ExN1 | ANKRD62 |
| NETO1 | 0 | 0.35742321 | 0.9 | 0.686 | 0 | ExN1 | NETO1 |
| NPTN | 0 | 0.35736256 | 0.831 | 0.678 | 0 | ExN1 | NPTN |
| CACNA1C | 0 | 0.35698567 | 0.99 | 0.955 | 0 | ExN1 | CACNA1C |
| ATP9B | 0 | 0.35682619 | 0.845 | 0.707 | 0 | ExN1 | ATP9B |
| DAB1 | 0 | 0.35603941 | 0.982 | 0.852 | 0 | ExN1 | DAB1 |
| RTN4RL1 | 0 | 0.35582176 | 0.614 | 0.375 | 0 | ExN1 | RTN4RL1 |
| GOLGA8A | 0 | 0.35443636 | 0.679 | 0.487 | 0 | ExN1 | GOLGA8A |
| AC093817.2 | 0 | 0.3541874 | 0.261 | 0.072 | 0 | ExN1 | AC093817.2 |
| NBAS | 0 | 0.35416569 | 0.879 | 0.753 | 0 | ExN1 | NBAS |
| CPLX2 | 0 | 0.35397858 | 0.744 | 0.528 | 0 | ExN1 | CPLX2 |
| HS6ST1 | 0 | 0.35258205 | 0.551 | 0.324 | 0 | ExN1 | HS6ST1 |
| CRLF1 | 0 | 0.35246206 | 0.311 | 0.063 | 0 | ExN1 | CRLF1 |
| ETV5 | 0 | 0.35223128 | 0.335 | 0.14 | 0 | ExN1 | ETV5 |
| TNPO1 | 0 | 0.35215613 | 0.786 | 0.607 | 0 | ExN1 | TNPO1 |
| KCNN3 | 0 | 0.35211978 | 0.532 | 0.265 | 0 | ExN1 | KCNN3 |
| TEK | 0 | 0.35171142 | 0.342 | 0.079 | 0 | ExN1 | TEK |
| PCSK2 | 0 | 0.35144 | 0.933 | 0.769 | 0 | ExN1 | PCSK2 |
| RAB40B | 0 | 0.35058014 | 0.53 | 0.295 | 0 | ExN1 | RAB40B |
| MYO1D | 0 | 0.35028844 | 0.737 | 0.462 | 0 | ExN1 | MYO1D |
| COP1 | 0 | 0.34841956 | 0.859 | 0.728 | 0 | ExN1 | COP1 |
| LRFN2 | 0 | 0.34793448 | 0.855 | 0.644 | 0 | ExN1 | LRFN2 |
| C11orf87 | 0 | 0.34759217 | 0.523 | 0.283 | 0 | ExN1 | C11orf87 |

| | | | | | | | |
|---|---|---|---|---|---|---|---|
| ADCY1 | 0 | 0.34712834 | 0.771 | 0.556 | 0 | ExN1 | ADCY1 |
| OPN3 | 0 | 0.34696567 | 0.424 | 0.196 | 0 | ExN1 | OPN3 |
| STEAP2 | 0 | 0.34627831 | 0.563 | 0.342 | 0 | ExN1 | STEAP2 |
| TMTC1 | 0 | 0.34600429 | 0.752 | 0.513 | 0 | ExN1 | TMTC1 |
| ROCK2 | 0 | 0.34563231 | 0.879 | 0.746 | 0 | ExN1 | ROCK2 |
| CREG2 | 0 | 0.34561502 | 0.684 | 0.471 | 0 | ExN1 | CREG2 |
| CHL1 | 0 | 0.3453969 | 0.937 | 0.771 | 0 | ExN1 | CHL1 |
| PAK1 | 0 | 0.34487878 | 0.82 | 0.659 | 0 | ExN1 | PAK1 |
| KCNMB2 | 0 | 0.34428191 | 0.812 | 0.514 | 0 | ExN1 | KCNMB2 |
| SLC22A10 | 0 | 0.34405512 | 0.722 | 0.398 | 0 | ExN1 | SLC22A10 |
| ANKRD24 | 0 | 0.34345977 | 0.585 | 0.357 | 0 | ExN1 | ANKRD24 |
| AL583827.1 | 0 | 0.34288903 | 0.266 | 0.067 | 0 | ExN1 | AL583827.1 |
| CDKL1 | 0 | 0.34277983 | 0.631 | 0.422 | 0 | ExN1 | CDKL1 |
| CCDC144NL | 0 | 0.34198258 | 0.335 | 0.095 | 0 | ExN1 | CCDC144NL |
| SLC44A1 | 0 | 0.3381798 | 0.871 | 0.68 | 0 | ExN1 | SLC44A1 |
| AC004158.1 | 0 | 0.3380912 | 0.375 | 0.175 | 0 | ExN1 | AC004158.1 |
| DGKA | 0 | 0.33743807 | 0.414 | 0.171 | 0 | ExN1 | DGKA |
| ATP11A | 0 | 0.33626596 | 0.618 | 0.401 | 0 | ExN1 | ATP11A |
| BSN | 0 | 0.3360333 | 0.759 | 0.566 | 0 | ExN1 | BSN |
| HTR3B | 0 | 0.33597495 | 0.3 | 0.079 | 0 | ExN1 | HTR3B |
| DTWD2 | 0 | 0.33566155 | 0.559 | 0.346 | 0 | ExN1 | DTWD2 |
| UBE2E2 | 0 | 0.33534207 | 0.963 | 0.855 | 0 | ExN1 | UBE2E2 |
| DOCK4 | 0 | 0.33236578 | 0.938 | 0.798 | 0 | ExN1 | DOCK4 |
| NAV3 | 0 | 0.33193957 | 0.993 | 0.939 | 0 | ExN1 | NAV3 |
| KCNAB2 | 0 | 0.3319342 | 0.693 | 0.489 | 0 | ExN1 | KCNAB2 |
| GPM6A | 0 | 0.33180661 | 0.985 | 0.933 | 0 | ExN1 | GPM6A |
| KCNN2 | 0 | 0.33157476 | 0.672 | 0.448 | 0 | ExN1 | KCNN2 |
| SYNPO | 0 | 0.33145754 | 0.478 | 0.255 | 0 | ExN1 | SYNPO |
| HOMER1 | 0 | 0.33074753 | 0.768 | 0.585 | 0 | ExN1 | HOMER1 |
| MIR4500HG | 0 | 0.33073258 | 0.469 | 0.261 | 0 | ExN1 | MIR4500HG |
| KCNG3 | 0 | 0.33033141 | 0.311 | 0.083 | 0 | ExN1 | KCNG3 |
| LINC01476 | 0 | 0.33032372 | 0.316 | 0.103 | 0 | ExN1 | LINC01476 |
| MTA3 | 0 | 0.32981716 | 0.798 | 0.619 | 0 | ExN1 | MTA3 |
| ABLIM1 | 0 | 0.3290512 | 0.938 | 0.806 | 0 | ExN1 | ABLIM1 |
| KBTBD11 | 0 | 0.32879783 | 0.397 | 0.189 | 0 | ExN1 | KBTBD11 |
| CACNA2D3 | 0 | 0.32861777 | 0.982 | 0.838 | 0 | ExN1 | CACNA2D3 |
| DOC2A | 0 | 0.32744629 | 0.486 | 0.279 | 0 | ExN1 | DOC2A |
| RAPGEFL1 | 0 | 0.32665883 | 0.596 | 0.4 | 0 | ExN1 | RAPGEFL1 |
| ABI2 | 0 | 0.32657491 | 0.879 | 0.742 | 0 | ExN1 | ABI2 |
| TMEM196 | 0 | 0.32586292 | 0.403 | 0.166 | 0 | ExN1 | TMEM196 |
| GLS | 0 | 0.32539426 | 0.919 | 0.803 | 0 | ExN1 | GLS |
| DNM1 | 0 | 0.32475567 | 0.929 | 0.815 | 0 | ExN1 | DNM1 |
| INKA2 | 0 | 0.32299275 | 0.397 | 0.194 | 0 | ExN1 | INKA2 |
| FHL2 | 0 | 0.32192141 | 0.449 | 0.222 | 0 | ExN1 | FHL2 |
| SYTL2 | 0 | 0.32070463 | 0.665 | 0.437 | 0 | ExN1 | SYTL2 |
| POU2F2 | 0 | 0.31981408 | 0.517 | 0.294 | 0 | ExN1 | POU2F2 |
| MOB1B | 0 | 0.31800625 | 0.534 | 0.322 | 0 | ExN1 | MOB1B |
| CA4 | 0 | 0.31567348 | 0.276 | 0.07 | 0 | ExN1 | CA4 |
| PITPNM2 | 0 | 0.31547528 | 0.718 | 0.516 | 0 | ExN1 | PITPNM2 |
| PTPRQ | 0 | 0.31464804 | 0.275 | 0.078 | 0 | ExN1 | PTPRQ |
| CA11 | 0 | 0.31408683 | 0.595 | 0.38 | 0 | ExN1 | CA11 |
| FSIP1 | 0 | 0.31270658 | 0.431 | 0.222 | 0 | ExN1 | FSIP1 |
| IQSEC2 | 0 | 0.31266866 | 0.646 | 0.448 | 0 | ExN1 | IQSEC2 |
| KIAA1257 | 0 | 0.3101321 | 0.32 | 0.103 | 0 | ExN1 | KIAA1257 |
| INSYN2B | 0 | 0.30914611 | 0.419 | 0.179 | 0 | ExN1 | INSYN2B |
| PTCHD1 | 0 | 0.30764038 | 0.414 | 0.208 | 0 | ExN1 | PTCHD1 |
| LRRIQ1 | 0 | 0.30577529 | 0.32 | 0.129 | 0 | ExN1 | LRRIQ1 |
| WIPF3 | 0 | 0.30546598 | 0.586 | 0.363 | 0 | ExN1 | WIPF3 |
| ST3GAL1 | 0 | 0.30470021 | 0.498 | 0.251 | 0 | ExN1 | ST3GAL1 |
| CUX1 | 0 | 0.30412228 | 0.87 | 0.693 | 0 | ExN1 | CUX1 |
| CRIP2 | 0 | 0.30372026 | 0.356 | 0.149 | 0 | ExN1 | CRIP2 |
| CTXN1 | 0 | 0.30340138 | 0.382 | 0.182 | 0 | ExN1 | CTXN1 |
| MSRA | 0 | 0.30257635 | 0.975 | 0.891 | 0 | ExN1 | MSRA |

| | | | | | | | |
|---|---|---|---|---|---|---|---|
| RFX2 | 0 | 0.30173112 | 0.431 | 0.199 | 0 | ExN1 | RFX2 |
| RASAL1 | 0 | 0.30049145 | 0.348 | 0.148 | 0 | ExN1 | RASAL1 |
| CYTOR | 0 | 0.29937248 | 0.39 | 0.187 | 0 | ExN1 | CYTOR |
| MTHFD1 | 0 | 0.2993171 | 0.428 | 0.218 | 0 | ExN1 | MTHFD1 |
| TTC3 | 0 | 0.29914398 | 0.956 | 0.898 | 0 | ExN1 | TTC3 |
| SLC17A6 | 0 | 0.29765633 | 0.304 | 0.102 | 0 | ExN1 | SLC17A6 |
| FXYD7 | 0 | 0.29710517 | 0.573 | 0.358 | 0 | ExN1 | FXYD7 |
| LINC01793 | 0 | 0.29431576 | 0.269 | 0.09 | 0 | ExN1 | LINC01793 |
| HTR2A | 0 | 0.29415297 | 0.51 | 0.291 | 0 | ExN1 | HTR2A |
| KCNH4 | 0 | 0.29331917 | 0.295 | 0.096 | 0 | ExN1 | KCNH4 |
| MGAT5B | 0 | 0.29296656 | 0.666 | 0.43 | 0 | ExN1 | MGAT5B |
| LINC00937 | 0 | 0.29230811 | 0.449 | 0.246 | 0 | ExN1 | LINC00937 |
| LGI4 | 0 | 0.28936018 | 0.47 | 0.267 | 0 | ExN1 | LGI4 |
| NRN1 | 0 | 0.2892669 | 0.375 | 0.185 | 0 | ExN1 | NRN1 |
| AP1S1 | 0 | 0.28895437 | 0.321 | 0.132 | 0 | ExN1 | AP1S1 |
| CFAP54 | 0 | 0.288029 | 0.382 | 0.187 | 0 | ExN1 | CFAP54 |
| BEGAIN | 0 | 0.28778288 | 0.425 | 0.214 | 0 | ExN1 | BEGAIN |
| PRSS3 | 0 | 0.28665376 | 0.352 | 0.159 | 0 | ExN1 | PRSS3 |
| CAT | 0 | 0.2860226 | 0.262 | 0.046 | 0 | ExN1 | CAT |
| MAP7 | 0 | 0.28393879 | 0.946 | 0.863 | 0 | ExN1 | MAP7 |
| AL078600.1 | 0 | 0.28247598 | 0.337 | 0.161 | 0 | ExN1 | AL078600.1 |
| LHX2 | 0 | 0.28188269 | 0.356 | 0.155 | 0 | ExN1 | LHX2 |
| NPEPL1 | 0 | 0.27918833 | 0.404 | 0.211 | 0 | ExN1 | NPEPL1 |
| PCSK6 | 0 | 0.27897296 | 0.391 | 0.173 | 0 | ExN1 | PCSK6 |
| PRKCA | 0 | 0.27665533 | 0.917 | 0.688 | 0 | ExN1 | PRKCA |
| SLC2A12 | 0 | 0.27654043 | 0.32 | 0.123 | 0 | ExN1 | SLC2A12 |
| RBFOX1 | 0 | 0.27635661 | 1 | 0.98 | 0 | ExN1 | RBFOX1 |
| KCNV1 | 0 | 0.2735909 | 0.301 | 0.116 | 0 | ExN1 | KCNV1 |
| SLC6A7 | 0 | 0.2734704 | 0.491 | 0.28 | 0 | ExN1 | SLC6A7 |
| DCBLD1 | 0 | 0.27295861 | 0.275 | 0.089 | 0 | ExN1 | DCBLD1 |
| CRHR1 | 0 | 0.27271507 | 0.352 | 0.17 | 0 | ExN1 | CRHR1 |
| RHPN1 | 0 | 0.27249584 | 0.396 | 0.211 | 0 | ExN1 | RHPN1 |
| DLG2 | 0 | 0.26438073 | 0.997 | 0.993 | 0 | ExN1 | DLG2 |
| GRIN2B | 0 | 0.26020903 | 0.98 | 0.938 | 0 | ExN1 | GRIN2B |
| NBPF11 | 0 | 0.25982771 | 0.322 | 0.128 | 0 | ExN1 | NBPF11 |
| PLEKHG5 | 0 | 0.25954596 | 0.426 | 0.227 | 0 | ExN1 | PLEKHG5 |
| ZNF184 | 0 | 0.25807046 | 0.279 | 0.094 | 0 | ExN1 | ZNF184 |
| VGF | 0 | 0.25334069 | 0.259 | 0.092 | 0 | ExN1 | VGF |
| PNCK | 2.22E-305 | 0.31970144 | 0.524 | 0.325 | 8.47E-301 | ExN1 | PNCK |
| TTLL11 | 7.54E-302 | 0.32845257 | 0.771 | 0.589 | 2.88E-297 | ExN1 | TTLL11 |
| SYT7 | 9.52E-302 | 0.30933976 | 0.714 | 0.488 | 3.64E-297 | ExN1 | SYT7 |
| DNAH7 | 1.33E-301 | 0.30932356 | 0.559 | 0.351 | 5.07E-297 | ExN1 | DNAH7 |
| RMDN2 | 4.32E-301 | 0.27593122 | 0.44 | 0.248 | 1.65E-296 | ExN1 | RMDN2 |
| PPFIA4 | 2.88E-300 | 0.31171984 | 0.708 | 0.512 | 1.10E-295 | ExN1 | PPFIA4 |
| PAM | 1.53E-299 | 0.25225368 | 0.893 | 0.723 | 5.83E-295 | ExN1 | PAM |
| CDYL2 | 3.20E-298 | 0.30496531 | 0.572 | 0.363 | 1.22E-293 | ExN1 | CDYL2 |
| CLASP1 | 9.30E-298 | 0.29537363 | 0.939 | 0.85 | 3.56E-293 | ExN1 | CLASP1 |
| TRERF1 | 5.03E-296 | 0.28880535 | 0.808 | 0.617 | 1.92E-291 | ExN1 | TRERF1 |
| SSX2IP | 6.22E-296 | 0.32063555 | 0.723 | 0.527 | 2.38E-291 | ExN1 | SSX2IP |
| ACVR1 | 1.50E-294 | 0.2510475 | 0.391 | 0.208 | 5.74E-290 | ExN1 | ACVR1 |
| EXOC6 | 1.12E-293 | 0.33136129 | 0.78 | 0.606 | 4.29E-289 | ExN1 | EXOC6 |
| CCPG1 | 1.65E-293 | 0.33024261 | 0.842 | 0.691 | 6.32E-289 | ExN1 | CCPG1 |
| SLC8A2 | 3.73E-293 | 0.29859954 | 0.626 | 0.424 | 1.43E-288 | ExN1 | SLC8A2 |
| AGAP1 | 1.68E-291 | 0.2781426 | 0.973 | 0.893 | 6.41E-287 | ExN1 | AGAP1 |
| SHC3 | 6.29E-291 | 0.33562827 | 0.767 | 0.569 | 2.40E-286 | ExN1 | SHC3 |
| NEBL | 7.22E-291 | 0.27913046 | 0.952 | 0.856 | 2.76E-286 | ExN1 | NEBL |
| SH3GL2 | 1.28E-289 | 0.2963545 | 0.921 | 0.701 | 4.87E-285 | ExN1 | SH3GL2 |
| LRRC8B | 2.99E-289 | 0.30581458 | 0.812 | 0.642 | 1.14E-284 | ExN1 | LRRC8B |
| SORT1 | 3.99E-289 | 0.30475029 | 0.824 | 0.644 | 1.52E-284 | ExN1 | SORT1 |
| TLK1 | 2.69E-288 | 0.30189069 | 0.772 | 0.59 | 1.03E-283 | ExN1 | TLK1 |
| MEIS3 | 6.37E-288 | 0.2959864 | 0.452 | 0.265 | 2.43E-283 | ExN1 | MEIS3 |
| ZFR | 1.13E-287 | 0.32310894 | 0.773 | 0.612 | 4.31E-283 | ExN1 | ZFR |
| GRIA3 | 3.64E-287 | 0.28059519 | 0.933 | 0.778 | 1.39E-282 | ExN1 | GRIA3 |

| | | | | | | | |
|---|---|---|---|---|---|---|---|
| TTL | 1.54E-284 | 0.26502358 | 0.489 | 0.299 | 5.89E-280 | ExN1 | TTL |
| AC006504.5 | 2.20E-284 | 0.33684916 | 0.587 | 0.392 | 8.41E-280 | ExN1 | AC006504.5 |
| PTPDC1 | 2.48E-283 | 0.28020534 | 0.465 | 0.279 | 9.48E-279 | ExN1 | PTPDC1 |
| DMD | 5.13E-283 | 0.27228214 | 0.981 | 0.906 | 1.96E-278 | ExN1 | DMD |
| FKBP1A | 1.97E-282 | 0.3145155 | 0.44 | 0.263 | 7.52E-278 | ExN1 | FKBP1A |
| TARBP1 | 6.92E-282 | 0.34117873 | 0.841 | 0.718 | 2.64E-277 | ExN1 | TARBP1 |
| CELF5 | 7.80E-282 | 0.32991559 | 0.827 | 0.687 | 2.98E-277 | ExN1 | CELF5 |
| MAPK6 | 7.98E-281 | 0.28101009 | 0.493 | 0.302 | 3.05E-276 | ExN1 | MAPK6 |
| EML5 | 1.85E-279 | 0.29256601 | 0.715 | 0.524 | 7.09E-275 | ExN1 | EML5 |
| PTPRF | 1.27E-277 | 0.27845962 | 0.503 | 0.313 | 4.85E-273 | ExN1 | PTPRF |
| PPFIBP1 | 5.82E-277 | 0.29447619 | 0.835 | 0.617 | 2.23E-272 | ExN1 | PPFIBP1 |
| KIAA1549L | 2.89E-276 | 0.30748307 | 0.925 | 0.813 | 1.11E-271 | ExN1 | KIAA1549L |
| YWHAZ | 6.00E-276 | 0.31291238 | 0.814 | 0.658 | 2.29E-271 | ExN1 | YWHAZ |
| PEA15 | 1.52E-275 | 0.30007726 | 0.489 | 0.306 | 5.79E-271 | ExN1 | PEA15 |
| CNTN1 | 3.13E-275 | 0.27869277 | 0.982 | 0.941 | 1.20E-270 | ExN1 | CNTN1 |
| KCNB1 | 3.43E-275 | 0.31115064 | 0.911 | 0.787 | 1.31E-270 | ExN1 | KCNB1 |
| CPEB4 | 7.19E-275 | 0.29944919 | 0.861 | 0.729 | 2.75E-270 | ExN1 | CPEB4 |
| MRTFB | 1.14E-274 | 0.34086348 | 0.849 | 0.739 | 4.38E-270 | ExN1 | MRTFB |
| ARNTL2 | 2.00E-274 | 0.26425829 | 0.539 | 0.338 | 7.65E-270 | ExN1 | ARNTL2 |
| ATF7IP2 | 9.81E-272 | 0.30934268 | 0.602 | 0.41 | 3.75E-267 | ExN1 | ATF7IP2 |
| ACAP2 | 1.45E-271 | 0.31488449 | 0.778 | 0.631 | 5.54E-267 | ExN1 | ACAP2 |
| DAPK1 | 9.94E-271 | 0.26044519 | 0.878 | 0.722 | 3.80E-266 | ExN1 | DAPK1 |
| AC239809.3 | 1.23E-267 | 0.29882157 | 0.518 | 0.335 | 4.70E-263 | ExN1 | AC239809.3 |
| PPP1R16B | 1.92E-267 | 0.28111567 | 0.811 | 0.605 | 7.33E-263 | ExN1 | PPP1R16B |
| ASXL3 | 8.49E-266 | 0.3141994 | 0.767 | 0.587 | 3.25E-261 | ExN1 | ASXL3 |
| DDX10 | 1.12E-265 | 0.29605177 | 0.678 | 0.497 | 4.29E-261 | ExN1 | DDX10 |
| APLP1 | 3.92E-265 | 0.31991547 | 0.582 | 0.399 | 1.50E-260 | ExN1 | APLP1 |
| C14orf132 | 6.14E-265 | 0.25413052 | 0.5 | 0.3 | 2.35E-260 | ExN1 | C14orf132 |
| NCKAP1 | 7.23E-264 | 0.30974078 | 0.852 | 0.712 | 2.76E-259 | ExN1 | NCKAP1 |
| PTPN5 | 9.73E-264 | 0.2979663 | 0.674 | 0.488 | 3.72E-259 | ExN1 | PTPN5 |
| MTURN | 2.11E-263 | 0.30308931 | 0.682 | 0.495 | 8.08E-259 | ExN1 | MTURN |
| NUP93 | 2.11E-261 | 0.25216668 | 0.635 | 0.437 | 8.06E-257 | ExN1 | NUP93 |
| TMEM181 | 8.25E-260 | 0.27603384 | 0.662 | 0.476 | 3.15E-255 | ExN1 | TMEM181 |
| TUT7 | 1.48E-259 | 0.26138647 | 0.498 | 0.311 | 5.64E-255 | ExN1 | TUT7 |
| ZFAND3 | 3.59E-259 | 0.28771181 | 0.925 | 0.83 | 1.37E-254 | ExN1 | ZFAND3 |
| AMD1 | 3.73E-258 | 0.30151447 | 0.626 | 0.437 | 1.42E-253 | ExN1 | AMD1 |
| AC010210.1 | 2.85E-256 | 0.2803628 | 0.464 | 0.285 | 1.09E-251 | ExN1 | AC010210.1 |
| PCNT | 4.44E-256 | 0.29350754 | 0.652 | 0.471 | 1.70E-251 | ExN1 | PCNT |
| OGA | 1.46E-254 | 0.29318417 | 0.82 | 0.683 | 5.60E-250 | ExN1 | OGA |
| ZNF609 | 2.09E-253 | 0.30166048 | 0.836 | 0.693 | 8.01E-249 | ExN1 | ZNF609 |
| CAMKV | 7.74E-252 | 0.28795172 | 0.557 | 0.371 | 2.96E-247 | ExN1 | CAMKV |
| LINC01197 | 3.32E-251 | 0.25090913 | 0.512 | 0.311 | 1.27E-246 | ExN1 | LINC01197 |
| DTNA | 3.70E-251 | 0.27420086 | 0.948 | 0.823 | 1.41E-246 | ExN1 | DTNA |
| TRIM9 | 9.57E-251 | 0.26846987 | 0.959 | 0.871 | 3.66E-246 | ExN1 | TRIM9 |
| ZC3H14 | 4.69E-250 | 0.28372389 | 0.704 | 0.529 | 1.79E-245 | ExN1 | ZC3H14 |
| PRKDC | 2.47E-248 | 0.29087057 | 0.756 | 0.607 | 9.44E-244 | ExN1 | PRKDC |
| TSPAN3 | 2.49E-248 | 0.2798028 | 0.623 | 0.441 | 9.52E-244 | ExN1 | TSPAN3 |
| TYW1 | 2.70E-248 | 0.27672392 | 0.612 | 0.431 | 1.03E-243 | ExN1 | TYW1 |
| MTMR12 | 2.25E-246 | 0.27179794 | 0.634 | 0.452 | 8.59E-242 | ExN1 | MTMR12 |
| DLG1 | 6.41E-246 | 0.27123983 | 0.904 | 0.742 | 2.45E-241 | ExN1 | DLG1 |
| MPP6 | 1.02E-245 | 0.25396571 | 0.73 | 0.546 | 3.88E-241 | ExN1 | MPP6 |
| FLCN | 1.48E-245 | 0.30856164 | 0.476 | 0.31 | 5.64E-241 | ExN1 | FLCN |
| RBFOX3 | 3.09E-245 | 0.28243141 | 0.959 | 0.865 | 1.18E-240 | ExN1 | RBFOX3 |
| CELF1 | 1.76E-244 | 0.28001299 | 0.876 | 0.753 | 6.73E-240 | ExN1 | CELF1 |
| BZW2 | 3.49E-243 | 0.26475351 | 0.528 | 0.347 | 1.33E-238 | ExN1 | BZW2 |
| RAB40C | 3.04E-241 | 0.25415949 | 0.601 | 0.414 | 1.16E-236 | ExN1 | RAB40C |
| GPC5 | 3.86E-241 | 0.38181294 | 0.576 | 0.383 | 1.48E-236 | ExN1 | GPC5 |
| YWHAH | 6.12E-241 | 0.33162527 | 0.651 | 0.463 | 2.34E-236 | ExN1 | YWHAH |
| PPP2R2C | 4.02E-239 | 0.28997053 | 0.915 | 0.771 | 1.54E-234 | ExN1 | PPP2R2C |
| DLGAP3 | 1.31E-238 | 0.26501633 | 0.617 | 0.434 | 5.01E-234 | ExN1 | DLGAP3 |
| GLIS3 | 1.08E-237 | 0.41902807 | 0.41 | 0.241 | 4.12E-233 | ExN1 | GLIS3 |
| ERICH3 | 2.35E-237 | 0.28909509 | 0.585 | 0.407 | 8.97E-233 | ExN1 | ERICH3 |
| ARMC2 | 5.30E-237 | 0.2600493 | 0.68 | 0.504 | 2.03E-232 | ExN1 | ARMC2 |

| | | | | | | | |
|---|---|---|---|---|---|---|---|
| SPECC1 | 1.70E-236 | 0.25316944 | 0.861 | 0.703 | 6.51E-232 | ExN1 | SPECC1 |
| RSRP1 | 1.43E-235 | 0.30190226 | 0.754 | 0.61 | 5.48E-231 | ExN1 | RSRP1 |
| STAT4 | 1.85E-235 | 0.27197959 | 0.592 | 0.401 | 7.07E-231 | ExN1 | STAT4 |
| TTC8 | 1.45E-233 | 0.2768669 | 0.565 | 0.394 | 5.55E-229 | ExN1 | TTC8 |
| PHYHIPL | 1.74E-233 | 0.302239 | 0.727 | 0.558 | 6.64E-229 | ExN1 | PHYHIPL |
| ARHGEF7 | 2.17E-233 | 0.26696106 | 0.904 | 0.792 | 8.29E-229 | ExN1 | ARHGEF7 |
| TCERG1L | 2.35E-231 | 0.35750137 | 0.48 | 0.312 | 8.97E-227 | ExN1 | TCERG1L |
| LRRC28 | 6.09E-230 | 0.28626841 | 0.758 | 0.602 | 2.33E-225 | ExN1 | LRRC28 |
| AC124852.1 | 1.18E-229 | 0.3022095 | 0.633 | 0.44 | 4.49E-225 | ExN1 | AC124852.1 |
| NELL2 | 5.85E-227 | 0.26589933 | 0.96 | 0.746 | 2.24E-222 | ExN1 | NELL2 |
| FBXO33 | 9.48E-225 | 0.26040405 | 0.548 | 0.375 | 3.62E-220 | ExN1 | FBXO33 |
| BEX1 | 1.74E-224 | 0.34827207 | 0.517 | 0.341 | 6.65E-220 | ExN1 | BEX1 |
| MALAT1 | 3.75E-224 | 0.38433855 | 1 | 1 | 1.43E-219 | ExN1 | MALAT1 |
| PUM3 | 1.40E-219 | 0.28499149 | 0.73 | 0.575 | 5.34E-215 | ExN1 | PUM3 |
| AFF3 | 5.10E-219 | 0.26169847 | 0.961 | 0.887 | 1.95E-214 | ExN1 | AFF3 |
| GPC6 | 2.88E-218 | 0.36019573 | 0.622 | 0.459 | 1.10E-213 | ExN1 | GPC6 |
| GRM5 | 3.07E-218 | 0.26053317 | 0.985 | 0.877 | 1.17E-213 | ExN1 | GRM5 |
| SPRED1 | 1.81E-216 | 0.26089655 | 0.56 | 0.385 | 6.91E-212 | ExN1 | SPRED1 |
| OPA1 | 1.86E-216 | 0.2796291 | 0.731 | 0.59 | 7.12E-212 | ExN1 | OPA1 |
| SIRPA | 1.58E-214 | 0.28060036 | 0.734 | 0.559 | 6.03E-210 | ExN1 | SIRPA |
| LYST | 2.24E-214 | 0.26705744 | 0.834 | 0.695 | 8.55E-210 | ExN1 | LYST |
| NEDD4L | 7.98E-214 | 0.25565092 | 0.908 | 0.79 | 3.05E-209 | ExN1 | NEDD4L |
| CCDC30 | 2.04E-212 | 0.27211262 | 0.71 | 0.537 | 7.78E-208 | ExN1 | CCDC30 |
| KCNJ6 | 2.93E-212 | 0.27707676 | 0.858 | 0.71 | 1.12E-207 | ExN1 | KCNJ6 |
| THY1 | 3.46E-211 | 0.34684132 | 0.632 | 0.472 | 1.32E-206 | ExN1 | THY1 |
| INTS10 | 9.27E-211 | 0.2577183 | 0.611 | 0.449 | 3.54E-206 | ExN1 | INTS10 |
| DNMT3A | 4.62E-210 | 0.26249974 | 0.759 | 0.595 | 1.77E-205 | ExN1 | DNMT3A |
| NPTXR | 7.05E-210 | 0.26583002 | 0.678 | 0.496 | 2.69E-205 | ExN1 | NPTXR |
| DZIP3 | 1.50E-208 | 0.26514954 | 0.744 | 0.597 | 5.74E-204 | ExN1 | DZIP3 |
| UGGT2 | 7.86E-208 | 0.26816086 | 0.824 | 0.682 | 3.00E-203 | ExN1 | UGGT2 |
| PPP1R12A | 7.44E-206 | 0.26102923 | 0.841 | 0.703 | 2.84E-201 | ExN1 | PPP1R12A |
| RBM26 | 1.23E-204 | 0.25778446 | 0.868 | 0.75 | 4.71E-200 | ExN1 | RBM26 |
| CALM2 | 1.49E-203 | 0.40774411 | 0.786 | 0.68 | 5.69E-199 | ExN1 | CALM2 |
| TIAM2 | 5.18E-203 | 0.26048936 | 0.852 | 0.706 | 1.98E-198 | ExN1 | TIAM2 |
| HTT | 1.77E-201 | 0.25306187 | 0.79 | 0.651 | 6.77E-197 | ExN1 | HTT |
| AC069228.1 | 9.88E-200 | 0.28944639 | 0.31 | 0.175 | 3.78E-195 | ExN1 | AC069228.1 |
| DIRC3 | 1.47E-198 | 0.26740117 | 0.703 | 0.505 | 5.62E-194 | ExN1 | DIRC3 |
| BSG | 2.56E-198 | 0.25925434 | 0.565 | 0.4 | 9.78E-194 | ExN1 | BSG |
| LINC02693 | 7.51E-197 | 0.28527174 | 0.602 | 0.45 | 2.87E-192 | ExN1 | LINC02693 |
| SCN3B | 4.54E-194 | 0.25487739 | 0.651 | 0.481 | 1.73E-189 | ExN1 | SCN3B |
| AC107223.1 | 5.87E-190 | 0.53068009 | 0.659 | 0.539 | 2.24E-185 | ExN1 | AC107223.1 |
| GDAP1 | 8.03E-184 | 0.27162518 | 0.881 | 0.764 | 3.07E-179 | ExN1 | GDAP1 |
| DYNLL1 | 9.46E-178 | 0.25754913 | 0.56 | 0.399 | 3.62E-173 | ExN1 | DYNLL1 |
| AC110296.1 | 8.57E-176 | 0.32242235 | 0.291 | 0.167 | 3.28E-171 | ExN1 | AC110296.1 |
| CCDC144A | 6.95E-173 | 0.29373966 | 0.74 | 0.639 | 2.66E-168 | ExN1 | CCDC144A |
| TOMM20 | 1.01E-172 | 0.28706055 | 0.59 | 0.44 | 3.86E-168 | ExN1 | TOMM20 |
| MKLN1 | 6.70E-172 | 0.25008151 | 0.911 | 0.81 | 2.56E-167 | ExN1 | MKLN1 |
| CALM1 | 4.54E-170 | 0.32396577 | 0.925 | 0.841 | 1.73E-165 | ExN1 | CALM1 |
| XIST | 3.05E-164 | 0.4824146 | 0.593 | 0.532 | 1.17E-159 | ExN1 | XIST |
| SERINC3 | 8.39E-164 | 0.26403797 | 0.604 | 0.458 | 3.21E-159 | ExN1 | SERINC3 |
| AC002460.2 | 9.02E-148 | 0.25189846 | 0.548 | 0.393 | 3.45E-143 | ExN1 | AC002460.2 |
| PDE4DIP | 2.73E-82 | 0.25134421 | 0.771 | 0.679 | 1.04E-77 | ExN1 | PDE4DIP |
| TSHZ2 | 0 | 2.554354 | 0.908 | 0.347 | 0 | ExN2 | TSHZ2 |
| AL356295.1 | 0 | 2.28635537 | 0.754 | 0.109 | 0 | ExN2 | AL356295.1 |
| DCC | 0 | 2.18932517 | 0.937 | 0.504 | 0 | ExN2 | DCC |
| AC016687.2 | 0 | 2.05799048 | 0.697 | 0.098 | 0 | ExN2 | AC016687.2 |
| RORB | 0 | 1.95749013 | 0.894 | 0.273 | 0 | ExN2 | RORB |
| IL1RAPL2 | 0 | 1.95699415 | 0.863 | 0.266 | 0 | ExN2 | IL1RAPL2 |
| LINC01725 | 0 | 1.79720721 | 0.796 | 0.311 | 0 | ExN2 | LINC01725 |
| TOX | 0 | 1.66931006 | 0.89 | 0.396 | 0 | ExN2 | TOX |
| SLC22A101 | 0 | 1.58092292 | 0.856 | 0.424 | 0 | ExN2 | SLC22A10 |
| COBLL1 | 0 | 1.54058277 | 0.736 | 0.108 | 0 | ExN2 | COBLL1 |
| POU6F2 | 0 | 1.49489783 | 0.869 | 0.355 | 0 | ExN2 | POU6F2 |

| | | | | | | | |
|---|---|---|---|---|---|---|---|
| FOXP2 | 0 | 1.49163199 | 0.837 | 0.26 | 0 | ExN2 | FOXP2 |
| LINC02232 | 0 | 1.4701298 | 0.678 | 0.139 | 0 | ExN2 | LINC02232 |
| SCHLAP1 | 0 | 1.46888737 | 0.603 | 0.174 | 0 | ExN2 | SCHLAP1 |
| FSHR | 0 | 1.44584744 | 0.327 | 0.016 | 0 | ExN2 | FSHR |
| AC073091.3 | 0 | 1.43265423 | 0.754 | 0.27 | 0 | ExN2 | AC073091.3 |
| VWC2L | 0 | 1.41083053 | 0.804 | 0.296 | 0 | ExN2 | VWC2L |
| AC060765.2 | 0 | 1.39294298 | 0.401 | 0.174 | 0 | ExN2 | AC060765.2 |
| ALDH1A1 | 0 | 1.3914538 | 0.57 | 0.03 | 0 | ExN2 | ALDH1A1 |
| AC084816.1 | 0 | 1.32409221 | 0.607 | 0.153 | 0 | ExN2 | AC084816.1 |
| KCNH5 | 0 | 1.28995833 | 0.832 | 0.375 | 0 | ExN2 | KCNH5 |
| AL450352.1 | 0 | 1.22718362 | 0.54 | 0.029 | 0 | ExN2 | AL450352.1 |
| RARB | 0 | 1.20485405 | 0.803 | 0.391 | 0 | ExN2 | RARB |
| TAFA2 | 0 | 1.19704994 | 0.938 | 0.776 | 0 | ExN2 | TAFA2 |
| ALCAM | 0 | 1.17643606 | 0.883 | 0.427 | 0 | ExN2 | ALCAM |
| GRM3 | 0 | 1.14715652 | 0.861 | 0.351 | 0 | ExN2 | GRM3 |
| KHDRBS2 | 0 | 1.13401443 | 0.985 | 0.891 | 0 | ExN2 | KHDRBS2 |
| SLC38A111 | 0 | 1.1321294 | 0.688 | 0.208 | 0 | ExN2 | SLC38A11 |
| PTPRK | 0 | 1.12016363 | 0.964 | 0.57 | 0 | ExN2 | PTPRK |
| AL117329.11 | 0 | 1.11155679 | 0.903 | 0.428 | 0 | ExN2 | AL117329.1 |
| FSTL41 | 0 | 1.10319577 | 0.954 | 0.619 | 0 | ExN2 | FSTL4 |
| CASC15 | 0 | 1.10061401 | 0.782 | 0.496 | 0 | ExN2 | CASC15 |
| LINC01414 | 0 | 1.09958274 | 0.735 | 0.43 | 0 | ExN2 | LINC01414 |
| PLCXD3 | 0 | 1.07603705 | 0.744 | 0.266 | 0 | ExN2 | PLCXD3 |
| AC073365.11 | 0 | 1.06182229 | 0.75 | 0.345 | 0 | ExN2 | AC073365.1 |
| AC019211.1 | 0 | 1.0585814 | 0.764 | 0.31 | 0 | ExN2 | AC019211.1 |
| GABRG1 | 0 | 1.04388121 | 0.565 | 0.089 | 0 | ExN2 | GABRG1 |
| RORA | 0 | 1.04265156 | 0.987 | 0.961 | 0 | ExN2 | RORA |
| MIR31HG | 0 | 1.02757174 | 0.605 | 0.168 | 0 | ExN2 | MIR31HG |
| AP003066.1 | 0 | 1.02259223 | 0.369 | 0.077 | 0 | ExN2 | AP003066.1 |
| AC092422.1 | 0 | 1.01584067 | 0.695 | 0.243 | 0 | ExN2 | AC092422.1 |
| SLC35F3 | 0 | 0.98550934 | 0.94 | 0.752 | 0 | ExN2 | SLC35F3 |
| GAB1 | 0 | 0.98258849 | 0.866 | 0.516 | 0 | ExN2 | GAB1 |
| TRHDE | 0 | 0.98032836 | 0.869 | 0.484 | 0 | ExN2 | TRHDE |
| LHFPL6 | 0 | 0.97741479 | 0.819 | 0.382 | 0 | ExN2 | LHFPL6 |
| AC067956.1 | 0 | 0.95731461 | 0.709 | 0.286 | 0 | ExN2 | AC067956.1 |
| EDIL31 | 0 | 0.93404473 | 0.944 | 0.709 | 0 | ExN2 | EDIL3 |
| SCN1A-AS1 | 0 | 0.93240226 | 0.591 | 0.234 | 0 | ExN2 | SCN1A-AS1 |
| MTHFD2L | 0 | 0.88776587 | 0.65 | 0.334 | 0 | ExN2 | MTHFD2L |
| AC246817.1 | 0 | 0.88741848 | 0.447 | 0.109 | 0 | ExN2 | AC246817.1 |
| LINC01202 | 0 | 0.88357461 | 0.516 | 0.123 | 0 | ExN2 | LINC01202 |
| EFNA5 | 0 | 0.88168875 | 0.965 | 0.781 | 0 | ExN2 | EFNA5 |
| SATB1-AS1 | 0 | 0.87401156 | 0.741 | 0.407 | 0 | ExN2 | SATB1-AS1 |
| KHDRBS3 | 0 | 0.87227686 | 0.963 | 0.78 | 0 | ExN2 | KHDRBS3 |
| CTNNA2 | 0 | 0.87153507 | 0.994 | 0.954 | 0 | ExN2 | CTNNA2 |
| FAM13A | 0 | 0.86043827 | 0.847 | 0.578 | 0 | ExN2 | FAM13A |
| SLC5A4-AS1 | 0 | 0.85718309 | 0.742 | 0.381 | 0 | ExN2 | SLC5A4-AS1 |
| OLFM3 | 0 | 0.85374043 | 0.94 | 0.692 | 0 | ExN2 | OLFM3 |
| KCND3 | 0 | 0.84871285 | 0.941 | 0.816 | 0 | ExN2 | KCND3 |
| AC109466.1 | 0 | 0.81891336 | 0.56 | 0.263 | 0 | ExN2 | AC109466.1 |
| IQCJ-SCHIP11 | 0 | 0.81643386 | 0.993 | 0.819 | 0 | ExN2 | IQCJ-SCHIP1 |
| NRGN1 | 0 | 0.81376253 | 0.83 | 0.549 | 0 | ExN2 | NRGN |
| LINC01721 | 0 | 0.8090192 | 0.351 | 0.029 | 0 | ExN2 | LINC01721 |
| RALYL | 0 | 0.78174177 | 0.991 | 0.801 | 0 | ExN2 | RALYL |
| KIF16B | 0 | 0.77940769 | 0.776 | 0.516 | 0 | ExN2 | KIF16B |
| FOXP1 | 0 | 0.7757103 | 0.956 | 0.806 | 0 | ExN2 | FOXP1 |
| CARMIL1 | 0 | 0.76217466 | 0.862 | 0.597 | 0 | ExN2 | CARMIL1 |
| CAMK1D1 | 0 | 0.75991851 | 0.974 | 0.849 | 0 | ExN2 | CAMK1D |
| BMPER | 0 | 0.74949283 | 0.845 | 0.594 | 0 | ExN2 | BMPER |
| AL445250.1 | 0 | 0.74723971 | 0.615 | 0.315 | 0 | ExN2 | AL445250.1 |
| LMO4 | 0 | 0.74544844 | 0.787 | 0.504 | 0 | ExN2 | LMO4 |
| AC073578.2 | 0 | 0.74363007 | 0.4 | 0.046 | 0 | ExN2 | AC073578.2 |
| LINC01821 | 0 | 0.7419755 | 0.496 | 0.186 | 0 | ExN2 | LINC01821 |
| SATB21 | 0 | 0.74117999 | 0.904 | 0.544 | 0 | ExN2 | SATB2 |

| | | | | | | | |
|---|---|---|---|---|---|---|---|
| EPHA41 | 0 | 0.7362049 | 0.889 | 0.657 | 0 | ExN2 | EPHA4 |
| PTPRD | 0 | 0.72436151 | 0.997 | 0.961 | 0 | ExN2 | PTPRD |
| UBE2E3 | 0 | 0.72168424 | 0.761 | 0.53 | 0 | ExN2 | UBE2E3 |
| LINC02389 | 0 | 0.71982724 | 0.72 | 0.449 | 0 | ExN2 | LINC02389 |
| SPATS2L | 0 | 0.71955785 | 0.861 | 0.636 | 0 | ExN2 | SPATS2L |
| SOX5 | 0 | 0.7074669 | 0.921 | 0.645 | 0 | ExN2 | SOX5 |
| AC107223.11 | 0 | 0.69992701 | 0.762 | 0.542 | 0 | ExN2 | AC107223.1 |
| SEMA3A | 0 | 0.69446307 | 0.543 | 0.177 | 0 | ExN2 | SEMA3A |
| DPY19L1 | 0 | 0.69291996 | 0.655 | 0.323 | 0 | ExN2 | DPY19L1 |
| HECW1 | 0 | 0.68964915 | 0.959 | 0.82 | 0 | ExN2 | HECW1 |
| SPOCK1 | 0 | 0.68530542 | 0.962 | 0.728 | 0 | ExN2 | SPOCK1 |
| LDB21 | 0 | 0.68110735 | 0.98 | 0.743 | 0 | ExN2 | LDB2 |
| TSHZ3 | 0 | 0.68099509 | 0.715 | 0.419 | 0 | ExN2 | TSHZ3 |
| AC113383.1 | 0 | 0.68029845 | 0.583 | 0.295 | 0 | ExN2 | AC113383.1 |
| CCDC3 | 0 | 0.67295995 | 0.841 | 0.629 | 0 | ExN2 | CCDC3 |
| EPHA3 | 0 | 0.67211762 | 0.694 | 0.395 | 0 | ExN2 | EPHA3 |
| PTK7 | 0 | 0.67069333 | 0.467 | 0.102 | 0 | ExN2 | PTK7 |
| AC087883.2 | 0 | 0.6702455 | 0.341 | 0.049 | 0 | ExN2 | AC087883.2 |
| MSC-AS1 | 0 | 0.66992206 | 0.504 | 0.202 | 0 | ExN2 | MSC-AS1 |
| SLIT1 | 0 | 0.66741703 | 0.819 | 0.571 | 0 | ExN2 | SLIT1 |
| CFAP221 | 0 | 0.66087307 | 0.652 | 0.39 | 0 | ExN2 | CFAP221 |
| CNTN5 | 0 | 0.65265615 | 0.964 | 0.801 | 0 | ExN2 | CNTN5 |
| VAV3 | 0 | 0.64441204 | 0.376 | 0.125 | 0 | ExN2 | VAV3 |
| PPP3CA1 | 0 | 0.64435988 | 0.982 | 0.909 | 0 | ExN2 | PPP3CA |
| TDO2 | 0 | 0.63849702 | 0.436 | 0.104 | 0 | ExN2 | TDO2 |
| BRINP3 | 0 | 0.63683018 | 0.793 | 0.465 | 0 | ExN2 | BRINP3 |
| PAPSS2 | 0 | 0.63104984 | 0.443 | 0.112 | 0 | ExN2 | PAPSS2 |
| ARAP21 | 0 | 0.62329682 | 0.822 | 0.582 | 0 | ExN2 | ARAP2 |
| CHSY3 | 0 | 0.61753047 | 0.953 | 0.824 | 0 | ExN2 | CHSY3 |
| RFTN11 | 0 | 0.61708552 | 0.756 | 0.453 | 0 | ExN2 | RFTN1 |
| DPF3 | 0 | 0.6148935 | 0.609 | 0.251 | 0 | ExN2 | DPF3 |
| LY86-AS11 | 0 | 0.61214998 | 0.791 | 0.455 | 0 | ExN2 | LY86-AS1 |
| PPP2R2B | 0 | 0.60838809 | 0.989 | 0.953 | 0 | ExN2 | PPP2R2B |
| NEO1 | 0 | 0.6032236 | 0.777 | 0.576 | 0 | ExN2 | NEO1 |
| LAMA4 | 0 | 0.59884282 | 0.418 | 0.094 | 0 | ExN2 | LAMA4 |
| NUAK1 | 0 | 0.59571966 | 0.778 | 0.578 | 0 | ExN2 | NUAK1 |
| SLC24A2 | 0 | 0.59561236 | 0.968 | 0.821 | 0 | ExN2 | SLC24A2 |
| FAP | 0 | 0.59549141 | 0.401 | 0.096 | 0 | ExN2 | FAP |
| MLLT3 | 0 | 0.59408338 | 0.914 | 0.81 | 0 | ExN2 | MLLT3 |
| MARCH1 | 0 | 0.59328673 | 0.955 | 0.739 | 0 | ExN2 | 1-Mar |
| SND1 | 0 | 0.59218058 | 0.835 | 0.684 | 0 | ExN2 | SND1 |
| MROH7 | 0 | 0.5912464 | 0.481 | 0.187 | 0 | ExN2 | MROH7 |
| NELL21 | 0 | 0.57763334 | 0.952 | 0.774 | 0 | ExN2 | NELL2 |
| AC093843.1 | 0 | 0.57577038 | 0.475 | 0.194 | 0 | ExN2 | AC093843.1 |
| OPCML | 0 | 0.57478616 | 0.997 | 0.987 | 0 | ExN2 | OPCML |
| MEGF10 | 0 | 0.5741023 | 0.366 | 0.071 | 0 | ExN2 | MEGF10 |
| MBNL21 | 0 | 0.57408127 | 0.924 | 0.75 | 0 | ExN2 | MBNL2 |
| NKAIN2 | 0 | 0.57216929 | 0.987 | 0.824 | 0 | ExN2 | NKAIN2 |
| SCN1B | 0 | 0.57014797 | 0.503 | 0.242 | 0 | ExN2 | SCN1B |
| CEMIP | 0 | 0.56799263 | 0.665 | 0.384 | 0 | ExN2 | CEMIP |
| LINC02296 | 0 | 0.5582652 | 0.371 | 0.145 | 0 | ExN2 | LINC02296 |
| RHOBTB2 | 0 | 0.55701765 | 0.536 | 0.295 | 0 | ExN2 | RHOBTB2 |
| TLE4 | 0 | 0.55609678 | 0.708 | 0.391 | 0 | ExN2 | TLE4 |
| COL6A1 | 0 | 0.55246256 | 0.496 | 0.221 | 0 | ExN2 | COL6A1 |
| ANO3 | 0 | 0.55241478 | 0.685 | 0.37 | 0 | ExN2 | ANO3 |
| DLG11 | 0 | 0.54480181 | 0.9 | 0.763 | 0 | ExN2 | DLG1 |
| ACOT11 | 0 | 0.54368152 | 0.491 | 0.198 | 0 | ExN2 | ACOT11 |
| LINGO2 | 0 | 0.54063308 | 0.972 | 0.81 | 0 | ExN2 | LINGO2 |
| ITPK1 | 0 | 0.54055623 | 0.685 | 0.444 | 0 | ExN2 | ITPK1 |
| GRM7 | 0 | 0.53777563 | 0.974 | 0.852 | 0 | ExN2 | GRM7 |
| MIR181A1HG | 0 | 0.53678715 | 0.687 | 0.421 | 0 | ExN2 | MIR181A1HG |
| ATP8B1 | 0 | 0.52616439 | 0.512 | 0.238 | 0 | ExN2 | ATP8B1 |
| CXXC4 | 0 | 0.52556102 | 0.515 | 0.249 | 0 | ExN2 | CXXC4 |

| | | | | | | | |
|---|---|---|---|---|---|---|---|
| MARCKS | 0 | 0.52522736 | 0.504 | 0.265 | 0 | ExN2 | MARCKS |
| CACNB4 | 0 | 0.52232534 | 0.942 | 0.845 | 0 | ExN2 | CACNB4 |
| DOCK41 | 0 | 0.51588455 | 0.936 | 0.816 | 0 | ExN2 | DOCK4 |
| SNTG1 | 0 | 0.51317009 | 0.982 | 0.938 | 0 | ExN2 | SNTG1 |
| VIPR2 | 0 | 0.50896995 | 0.361 | 0.057 | 0 | ExN2 | VIPR2 |
| INPP4B | 0 | 0.50762056 | 0.923 | 0.737 | 0 | ExN2 | INPP4B |
| PTPN14 | 0 | 0.50517266 | 0.475 | 0.211 | 0 | ExN2 | PTPN14 |
| MET | 0 | 0.50344176 | 0.345 | 0.075 | 0 | ExN2 | MET |
| LRRK1 | 0 | 0.50322472 | 0.639 | 0.3 | 0 | ExN2 | LRRK1 |
| HS3ST2 | 0 | 0.49316342 | 0.562 | 0.235 | 0 | ExN2 | HS3ST2 |
| CDH121 | 0 | 0.48932417 | 0.962 | 0.666 | 0 | ExN2 | CDH12 |
| BCAR3 | 0 | 0.48739607 | 0.476 | 0.22 | 0 | ExN2 | BCAR3 |
| PLEKHG1 | 0 | 0.48663558 | 0.397 | 0.132 | 0 | ExN2 | PLEKHG1 |
| GSG1L1 | 0 | 0.48501023 | 0.555 | 0.282 | 0 | ExN2 | GSG1L |
| AC007614.1 | 0 | 0.4822139 | 0.299 | 0.052 | 0 | ExN2 | AC007614.1 |
| TNNT2 | 0 | 0.47725534 | 0.377 | 0.102 | 0 | ExN2 | TNNT2 |
| ESR1 | 0 | 0.47675561 | 0.304 | 0.085 | 0 | ExN2 | ESR1 |
| TANC1 | 0 | 0.46462017 | 0.298 | 0.099 | 0 | ExN2 | TANC1 |
| SGCG | 0 | 0.4623363 | 0.285 | 0.044 | 0 | ExN2 | SGCG |
| ATRNL11 | 0 | 0.45744485 | 0.984 | 0.925 | 0 | ExN2 | ATRNL1 |
| AL109930.1 | 0 | 0.43851793 | 0.327 | 0.068 | 0 | ExN2 | AL109930.1 |
| NRXN1 | 0 | 0.43174134 | 0.996 | 0.986 | 0 | ExN2 | NRXN1 |
| UNC5D | 0 | 0.42820049 | 0.964 | 0.817 | 0 | ExN2 | UNC5D |
| LOXHD1 | 0 | 0.42583454 | 0.258 | 0.05 | 0 | ExN2 | LOXHD1 |
| GOLIM4 | 0 | 0.42108186 | 0.427 | 0.138 | 0 | ExN2 | GOLIM4 |
| AC104689.2 | 0 | 0.41011993 | 0.258 | 0.045 | 0 | ExN2 | AC104689.2 |
| TPBG | 0 | 0.40899451 | 0.263 | 0.02 | 0 | ExN2 | TPBG |
| LRMP | 0 | 0.40633982 | 0.375 | 0.129 | 0 | ExN2 | LRMP |
| CNTNAP4 | 0 | 0.3969783 | 0.844 | 0.557 | 0 | ExN2 | CNTNAP4 |
| PTGFRN | 0 | 0.3933647 | 0.37 | 0.147 | 0 | ExN2 | PTGFRN |
| MAGI2 | 0 | 0.39006581 | 0.994 | 0.986 | 0 | ExN2 | MAGI2 |
| CCDC39 | 0 | 0.38389431 | 0.293 | 0.098 | 0 | ExN2 | CCDC39 |
| SLC22A9 | 0 | 0.35087983 | 0.305 | 0.107 | 0 | ExN2 | SLC22A9 |
| RORB-AS1 | 0 | 0.34731631 | 0.27 | 0.056 | 0 | ExN2 | RORB-AS1 |
| BARD1 | 0 | 0.34233336 | 0.275 | 0.066 | 0 | ExN2 | BARD1 |
| SLCO2A1 | 0 | 0.32388773 | 0.255 | 0.071 | 0 | ExN2 | SLCO2A1 |
| AC068051.1 | 0 | 0.32115207 | 0.253 | 0.077 | 0 | ExN2 | AC068051.1 |
| POU3F2 | 0 | 0.31448289 | 0.274 | 0.069 | 0 | ExN2 | POU3F2 |
| MERTK | 0 | 0.2991418 | 0.279 | 0.092 | 0 | ExN2 | MERTK |
| COL5A1 | 0 | 0.28179446 | 0.278 | 0.091 | 0 | ExN2 | COL5A1 |
| SCARA3 | 1.86E-307 | 0.29122543 | 0.261 | 0.082 | 7.11E-303 | ExN2 | SCARA3 |
| COL5A2 | 4.26E-307 | 0.31504088 | 0.335 | 0.121 | 1.63E-302 | ExN2 | COL5A2 |
| STXBP1 | 5.76E-303 | 0.50856659 | 0.899 | 0.843 | 2.20E-298 | ExN2 | STXBP1 |
| PPFIBP11 | 1.09E-300 | 0.51949566 | 0.843 | 0.644 | 4.16E-296 | ExN2 | PPFIBP1 |
| NOVA11 | 1.45E-298 | 0.48456258 | 0.897 | 0.804 | 5.54E-294 | ExN2 | NOVA1 |
| NPTX11 | 3.96E-298 | 0.54101922 | 0.721 | 0.462 | 1.51E-293 | ExN2 | NPTX1 |
| LINGO1 | 4.57E-298 | 0.5600666 | 0.709 | 0.484 | 1.75E-293 | ExN2 | LINGO1 |
| IPCEF1 | 1.66E-296 | 0.49545209 | 0.636 | 0.379 | 6.36E-292 | ExN2 | IPCEF1 |
| NAPEPLD | 1.41E-295 | 0.51792974 | 0.691 | 0.475 | 5.40E-291 | ExN2 | NAPEPLD |
| KHDRBS2-OT | 3.17E-295 | 0.6507375 | 0.657 | 0.433 | 1.21E-290 | ExN2 | KHDRBS2-OT |
| EPB41L4A | 2.12E-286 | 0.34673174 | 0.357 | 0.144 | 8.11E-282 | ExN2 | EPB41L4A |
| RNF144A | 3.38E-285 | 0.4965418 | 0.875 | 0.728 | 1.29E-280 | ExN2 | RNF144A |
| RFPL2 | 6.91E-284 | 0.36075557 | 0.34 | 0.138 | 2.64E-279 | ExN2 | RFPL2 |
| MIAT1 | 1.09E-282 | 0.46280823 | 0.957 | 0.875 | 4.16E-278 | ExN2 | MIAT |
| STXBP5-AS1 | 3.65E-282 | 0.55109187 | 0.803 | 0.567 | 1.40E-277 | ExN2 | STXBP5-AS1 |
| KCNH11 | 6.89E-282 | 0.52778776 | 0.943 | 0.821 | 2.63E-277 | ExN2 | KCNH1 |
| IL1RAP | 1.50E-281 | 0.44458929 | 0.554 | 0.303 | 5.74E-277 | ExN2 | IL1RAP |
| LINC023061 | 1.06E-277 | 0.42876454 | 0.509 | 0.241 | 4.05E-273 | ExN2 | LINC02306 |
| IGDCC3 | 2.96E-277 | 0.29441901 | 0.296 | 0.108 | 1.13E-272 | ExN2 | IGDCC3 |
| KLHL32 | 6.99E-276 | 0.52342174 | 0.837 | 0.672 | 2.67E-271 | ExN2 | KLHL32 |
| UBE2E21 | 6.76E-275 | 0.43785591 | 0.953 | 0.87 | 2.59E-270 | ExN2 | UBE2E2 |
| RASSF4 | 1.78E-273 | 0.36044015 | 0.436 | 0.209 | 6.79E-269 | ExN2 | RASSF4 |
| AGBL4 | 1.02E-272 | 0.39930561 | 0.978 | 0.929 | 3.91E-268 | ExN2 | AGBL4 |

| | | | | | | | |
|---|---|---|---|---|---|---|---|
| TESK2 | 4.46E-270 | 0.37551212 | 0.346 | 0.144 | 1.70E-265 | ExN2 | TESK2 |
| LRFN51 | 7.13E-269 | 0.44036287 | 0.963 | 0.885 | 2.72E-264 | ExN2 | LRFN5 |
| FRMD5 | 2.19E-268 | 0.45421743 | 0.96 | 0.855 | 8.37E-264 | ExN2 | FRMD5 |
| CNKSR21 | 3.57E-268 | 0.46108789 | 0.941 | 0.809 | 1.36E-263 | ExN2 | CNKSR2 |
| OLFM11 | 2.93E-266 | 0.52068951 | 0.777 | 0.602 | 1.12E-261 | ExN2 | OLFM1 |
| SPAG16 | 2.10E-265 | 0.61394414 | 0.815 | 0.673 | 8.04E-261 | ExN2 | SPAG16 |
| PAG1 | 2.77E-264 | 0.48642346 | 0.589 | 0.352 | 1.06E-259 | ExN2 | PAG1 |
| TGFBR1 | 1.63E-262 | 0.25370584 | 0.262 | 0.092 | 6.25E-258 | ExN2 | TGFBR1 |
| GLIS1 | 3.74E-261 | 0.53371504 | 0.672 | 0.459 | 1.43E-256 | ExN2 | GLIS1 |
| AC008415.1 | 6.00E-261 | 0.66939736 | 0.408 | 0.182 | 2.29E-256 | ExN2 | AC008415.1 |
| TRPC5 | 4.11E-260 | 0.41410998 | 0.564 | 0.306 | 1.57E-255 | ExN2 | TRPC5 |
| ST3GAL6 | 1.60E-259 | 0.49375776 | 0.637 | 0.414 | 6.11E-255 | ExN2 | ST3GAL6 |
| PLXDC2 | 9.54E-259 | 0.48584476 | 0.843 | 0.588 | 3.65E-254 | ExN2 | PLXDC2 |
| KCNIP41 | 5.14E-258 | 0.46811982 | 0.999 | 0.932 | 1.96E-253 | ExN2 | KCNIP4 |
| NWD21 | 4.85E-257 | 0.46373266 | 0.714 | 0.444 | 1.85E-252 | ExN2 | NWD2 |
| TBL1X | 9.50E-257 | 0.31460096 | 0.33 | 0.133 | 3.63E-252 | ExN2 | TBL1X |
| ENOX1 | 2.18E-255 | 0.37699081 | 0.95 | 0.841 | 8.34E-251 | ExN2 | ENOX1 |
| SNED11 | 1.36E-254 | 0.45295617 | 0.676 | 0.421 | 5.18E-250 | ExN2 | SNED1 |
| SLC17A71 | 3.93E-253 | 0.48552848 | 0.658 | 0.417 | 1.50E-248 | ExN2 | SLC17A7 |
| FAM171A1 | 3.55E-252 | 0.50153396 | 0.822 | 0.664 | 1.36E-247 | ExN2 | FAM171A1 |
| CPNE9 | 4.80E-252 | 0.3599444 | 0.381 | 0.178 | 1.84E-247 | ExN2 | CPNE9 |
| AC025809.1 | 1.08E-251 | 0.44014211 | 0.527 | 0.3 | 4.13E-247 | ExN2 | AC025809.1 |
| PDE1A1 | 1.26E-251 | 0.42552063 | 0.956 | 0.654 | 4.80E-247 | ExN2 | PDE1A |
| AC005906.2 | 1.53E-251 | 0.53877421 | 0.495 | 0.276 | 5.85E-247 | ExN2 | AC005906.2 |
| EPB41 | 1.92E-250 | 0.4825655 | 0.671 | 0.435 | 7.33E-246 | ExN2 | EPB41 |
| SCN1A | 1.21E-249 | 0.45395904 | 0.906 | 0.837 | 4.62E-245 | ExN2 | SCN1A |
| R3HDM11 | 4.23E-249 | 0.39655622 | 0.968 | 0.886 | 1.62E-244 | ExN2 | R3HDM1 |
| CDK18 | 1.53E-248 | 0.39463536 | 0.377 | 0.171 | 5.85E-244 | ExN2 | CDK18 |
| MLIP1 | 1.97E-248 | 0.52562711 | 0.758 | 0.492 | 7.51E-244 | ExN2 | MLIP |
| PTPRT | 2.78E-246 | 0.30929482 | 0.922 | 0.661 | 1.06E-241 | ExN2 | PTPRT |
| CTNNA3 | 7.62E-246 | 0.91866787 | 0.64 | 0.437 | 2.91E-241 | ExN2 | CTNNA3 |
| ZBTB7C1 | 2.67E-243 | 0.50110124 | 0.494 | 0.266 | 1.02E-238 | ExN2 | ZBTB7C |
| LRRN3 | 5.66E-242 | 0.45041603 | 0.539 | 0.317 | 2.16E-237 | ExN2 | LRRN3 |
| RAPGEF21 | 1.52E-241 | 0.40156833 | 0.929 | 0.851 | 5.81E-237 | ExN2 | RAPGEF2 |
| CPNE5 | 7.90E-241 | 0.47552836 | 0.63 | 0.41 | 3.02E-236 | ExN2 | CPNE5 |
| TMEM72-AS1 | 1.93E-239 | 0.48162471 | 0.542 | 0.33 | 7.37E-235 | ExN2 | TMEM72-AS1 |
| PDZRN4 | 7.67E-239 | 0.28149039 | 0.66 | 0.372 | 2.93E-234 | ExN2 | PDZRN4 |
| CDHR3 | 1.15E-238 | 0.41286043 | 0.606 | 0.357 | 4.41E-234 | ExN2 | CDHR3 |
| MAN2A1 | 1.76E-236 | 0.58771076 | 0.769 | 0.573 | 6.72E-232 | ExN2 | MAN2A1 |
| RPS6KA5 | 6.68E-235 | 0.48108902 | 0.771 | 0.582 | 2.55E-230 | ExN2 | RPS6KA5 |
| PELI2 | 1.19E-234 | 0.39880428 | 0.499 | 0.264 | 4.57E-230 | ExN2 | PELI2 |
| MGAT5B1 | 2.69E-233 | 0.45086049 | 0.681 | 0.458 | 1.03E-228 | ExN2 | MGAT5B |
| KCNQ3 | 1.18E-232 | 0.41784237 | 0.959 | 0.896 | 4.50E-228 | ExN2 | KCNQ3 |
| MRPS30-DT | 2.60E-232 | 0.50234848 | 0.648 | 0.444 | 9.96E-228 | ExN2 | MRPS30-DT |
| ADGRB3 | 2.57E-231 | 0.30929379 | 0.997 | 0.992 | 9.82E-227 | ExN2 | ADGRB3 |
| PLXND1 | 5.05E-230 | 0.2983768 | 0.305 | 0.13 | 1.93E-225 | ExN2 | PLXND1 |
| ATG7 | 2.21E-229 | 0.4535417 | 0.871 | 0.768 | 8.45E-225 | ExN2 | ATG7 |
| DNAJC1 | 2.68E-228 | 0.48112662 | 0.729 | 0.544 | 1.02E-223 | ExN2 | DNAJC1 |
| EPB41L21 | 6.38E-228 | 0.42095443 | 0.874 | 0.697 | 2.44E-223 | ExN2 | EPB41L2 |
| KIAA0319 | 3.17E-226 | 0.48943908 | 0.72 | 0.546 | 1.21E-221 | ExN2 | KIAA0319 |
| SLC8A1 | 8.24E-226 | 0.4077896 | 0.978 | 0.85 | 3.15E-221 | ExN2 | SLC8A1 |
| AL133166.1 | 6.65E-225 | 0.4700962 | 0.4 | 0.205 | 2.54E-220 | ExN2 | AL133166.1 |
| GUCY1A21 | 2.46E-224 | 0.44224159 | 0.862 | 0.719 | 9.40E-220 | ExN2 | GUCY1A2 |
| DGKH | 1.38E-223 | 0.48668607 | 0.572 | 0.364 | 5.26E-219 | ExN2 | DGKH |
| MYO5C1 | 2.34E-223 | 0.38752986 | 0.473 | 0.261 | 8.95E-219 | ExN2 | MYO5C |
| PTPN3 | 7.20E-223 | 0.38815974 | 0.426 | 0.224 | 2.75E-218 | ExN2 | PTPN3 |
| TRHDE-AS1 | 5.29E-222 | 0.32390225 | 0.375 | 0.179 | 2.02E-217 | ExN2 | TRHDE-AS1 |
| CTNNB1 | 6.82E-222 | 0.48883838 | 0.657 | 0.483 | 2.61E-217 | ExN2 | CTNNB1 |
| MICAL2 | 6.97E-221 | 0.45009968 | 0.854 | 0.722 | 2.67E-216 | ExN2 | MICAL2 |
| UBE3D | 7.62E-221 | 0.50453506 | 0.719 | 0.55 | 2.91E-216 | ExN2 | UBE3D |
| GREM2 | 1.73E-217 | 0.34703712 | 0.342 | 0.154 | 6.60E-213 | ExN2 | GREM2 |
| SH3GL21 | 2.85E-217 | 0.44949022 | 0.912 | 0.73 | 1.09E-212 | ExN2 | SH3GL2 |
| ZMYND8 | 4.97E-217 | 0.43414242 | 0.863 | 0.769 | 1.90E-212 | ExN2 | ZMYND8 |

| | | | | | | | |
|---|---|---|---|---|---|---|---|
| DUXAP8 | 3.55E-216 | 0.26762361 | 0.259 | 0.101 | 1.36E-211 | ExN2 | DUXAP8 |
| USP43 | 4.18E-215 | 0.39235142 | 0.444 | 0.242 | 1.60E-210 | ExN2 | USP43 |
| ALS2 | 2.73E-214 | 0.47357357 | 0.671 | 0.508 | 1.04E-209 | ExN2 | ALS2 |
| ADGRL21 | 2.39E-213 | 0.39248373 | 0.953 | 0.847 | 9.14E-209 | ExN2 | ADGRL2 |
| AC090578.1 | 1.98E-211 | 0.5744964 | 0.741 | 0.545 | 7.58E-207 | ExN2 | AC090578.1 |
| AC103855.2 | 3.79E-211 | 0.43296109 | 0.47 | 0.266 | 1.45E-206 | ExN2 | AC103855.2 |
| RSPO2 | 6.88E-206 | 0.36663461 | 0.623 | 0.383 | 2.63E-201 | ExN2 | RSPO2 |
| NEK11 | 2.58E-204 | 0.46938081 | 0.611 | 0.421 | 9.88E-200 | ExN2 | NEK11 |
| LYPD6B | 4.69E-204 | 0.29098648 | 0.358 | 0.163 | 1.79E-199 | ExN2 | LYPD6B |
| GLDN | 1.28E-201 | 0.35249265 | 0.419 | 0.218 | 4.89E-197 | ExN2 | GLDN |
| SORCS1 | 1.87E-201 | 0.34087146 | 0.907 | 0.654 | 7.16E-197 | ExN2 | SORCS1 |
| CXXC5 | 7.38E-200 | 0.39055931 | 0.541 | 0.337 | 2.82E-195 | ExN2 | CXXC5 |
| COBL | 1.01E-199 | 0.38949113 | 0.899 | 0.735 | 3.86E-195 | ExN2 | COBL |
| GPATCH8 | 1.18E-198 | 0.38663099 | 0.896 | 0.83 | 4.52E-194 | ExN2 | GPATCH8 |
| RAP1GAP2 | 1.28E-196 | 0.45509382 | 0.804 | 0.622 | 4.88E-192 | ExN2 | RAP1GAP2 |
| THRB | 1.90E-196 | 0.36633536 | 0.962 | 0.912 | 7.27E-192 | ExN2 | THRB |
| OSTF1 | 6.48E-196 | 0.34563729 | 0.421 | 0.229 | 2.48E-191 | ExN2 | OSTF1 |
| PCLO | 2.60E-195 | 0.3444284 | 0.96 | 0.92 | 9.95E-191 | ExN2 | PCLO |
| KIAA1958 | 3.07E-195 | 0.40835692 | 0.628 | 0.412 | 1.17E-190 | ExN2 | KIAA1958 |
| ST8SIA4 | 6.37E-195 | 0.28706837 | 0.352 | 0.167 | 2.43E-190 | ExN2 | ST8SIA4 |
| RHBDL3 | 2.76E-192 | 0.44155512 | 0.594 | 0.378 | 1.05E-187 | ExN2 | RHBDL3 |
| TSC22D3 | 7.18E-188 | 0.31347873 | 0.35 | 0.175 | 2.74E-183 | ExN2 | TSC22D3 |
| STAMBPL1 | 8.11E-188 | 0.44581721 | 0.673 | 0.5 | 3.10E-183 | ExN2 | STAMBPL1 |
| VPS41 | 1.05E-187 | 0.43228007 | 0.723 | 0.579 | 4.00E-183 | ExN2 | VPS41 |
| DGKI | 3.10E-186 | 0.33276566 | 0.952 | 0.887 | 1.18E-181 | ExN2 | DGKI |
| TSBP1-AS1 | 1.42E-185 | 0.40552206 | 0.476 | 0.275 | 5.42E-181 | ExN2 | TSBP1-AS1 |
| GABRB31 | 7.48E-185 | 0.33913812 | 0.953 | 0.896 | 2.86E-180 | ExN2 | GABRB3 |
| FOCAD | 1.84E-183 | 0.39599772 | 0.883 | 0.795 | 7.02E-179 | ExN2 | FOCAD |
| LINC00923 | 8.59E-183 | 0.50507399 | 0.433 | 0.238 | 3.29E-178 | ExN2 | LINC00923 |
| PTH2R1 | 4.27E-182 | 0.32785922 | 0.451 | 0.244 | 1.63E-177 | ExN2 | PTH2R |
| CACNA1G | 1.11E-180 | 0.30301801 | 0.388 | 0.205 | 4.26E-176 | ExN2 | CACNA1G |
| SATB1 | 1.38E-180 | 0.37623266 | 0.513 | 0.323 | 5.29E-176 | ExN2 | SATB1 |
| MEF2C | 3.42E-179 | 0.31429794 | 0.971 | 0.935 | 1.31E-174 | ExN2 | MEF2C |
| SPIDR | 2.36E-178 | 0.35420583 | 0.917 | 0.857 | 9.04E-174 | ExN2 | SPIDR |
| CREG21 | 2.29E-177 | 0.42299877 | 0.665 | 0.5 | 8.77E-173 | ExN2 | CREG2 |
| AFF1 | 3.68E-174 | 0.37297244 | 0.599 | 0.396 | 1.41E-169 | ExN2 | AFF1 |
| BACH2 | 6.14E-174 | 0.38597355 | 0.764 | 0.597 | 2.35E-169 | ExN2 | BACH2 |
| SGPL1 | 8.24E-174 | 0.33703264 | 0.47 | 0.288 | 3.15E-169 | ExN2 | SGPL1 |
| ANKS1B | 1.04E-172 | 0.2610173 | 0.995 | 0.986 | 3.98E-168 | ExN2 | ANKS1B |
| AC093523.1 | 1.23E-171 | 0.45102916 | 0.598 | 0.402 | 4.70E-167 | ExN2 | AC093523.1 |
| CAMK2A1 | 2.62E-171 | 0.33473022 | 0.907 | 0.785 | 1.00E-166 | ExN2 | CAMK2A |
| LINC02609 | 1.28E-169 | 0.29672607 | 0.3 | 0.143 | 4.88E-165 | ExN2 | LINC02609 |
| RTN1 | 1.71E-169 | 0.33044894 | 0.93 | 0.884 | 6.53E-165 | ExN2 | RTN1 |
| ZNF3651 | 2.76E-169 | 0.45722364 | 0.645 | 0.452 | 1.05E-164 | ExN2 | ZNF365 |
| EPHA5 | 6.02E-167 | 0.43515275 | 0.751 | 0.585 | 2.30E-162 | ExN2 | EPHA5 |
| CABP11 | 1.24E-165 | 0.36154683 | 0.868 | 0.689 | 4.72E-161 | ExN2 | CABP1 |
| AL355472.2 | 2.49E-165 | 0.39344797 | 0.435 | 0.26 | 9.52E-161 | ExN2 | AL355472.2 |
| SYTL21 | 2.58E-165 | 0.38551334 | 0.653 | 0.467 | 9.86E-161 | ExN2 | SYTL2 |
| ZNF385B | 2.99E-164 | 0.34304168 | 0.937 | 0.809 | 1.14E-159 | ExN2 | ZNF385B |
| PPP1R16B1 | 3.42E-164 | 0.38776247 | 0.794 | 0.633 | 1.31E-159 | ExN2 | PPP1R16B |
| SLC44A5 | 6.53E-164 | 0.35268154 | 0.835 | 0.681 | 2.49E-159 | ExN2 | SLC44A5 |
| CSMD2 | 7.64E-164 | 0.38859689 | 0.875 | 0.761 | 2.92E-159 | ExN2 | CSMD2 |
| PHYHIP1 | 3.15E-163 | 0.41747196 | 0.645 | 0.468 | 1.21E-158 | ExN2 | PHYHIP |
| SERPINI1 | 3.69E-163 | 0.36876794 | 0.799 | 0.667 | 1.41E-158 | ExN2 | SERPINI1 |
| CPNE8 | 3.68E-162 | 0.33707925 | 0.684 | 0.458 | 1.41E-157 | ExN2 | CPNE8 |
| CA111 | 3.92E-162 | 0.38728471 | 0.58 | 0.409 | 1.50E-157 | ExN2 | CA11 |
| FXYD71 | 9.88E-160 | 0.38284106 | 0.562 | 0.386 | 3.78E-155 | ExN2 | FXYD7 |
| SSBP3 | 5.81E-158 | 0.39605043 | 0.791 | 0.667 | 2.22E-153 | ExN2 | SSBP3 |
| TSPAN5 | 6.55E-157 | 0.3613739 | 0.927 | 0.835 | 2.50E-152 | ExN2 | TSPAN5 |
| LINC00882 | 1.21E-156 | 0.42974125 | 0.929 | 0.871 | 4.64E-152 | ExN2 | LINC00882 |
| RYR2 | 1.64E-155 | 0.27896205 | 0.984 | 0.923 | 6.26E-151 | ExN2 | RYR2 |
| SPARCL1 | 1.91E-155 | 0.35755955 | 0.803 | 0.653 | 7.30E-151 | ExN2 | SPARCL1 |
| LPGAT1 | 2.38E-155 | 0.40913464 | 0.675 | 0.533 | 9.10E-151 | ExN2 | LPGAT1 |

| | | | | | | | |
|---|---|---|---|---|---|---|---|
| CCDC175 | 2.82E-154 | 0.27310459 | 0.303 | 0.152 | 1.08E-149 | ExN2 | CCDC175 |
| CAMK2N11 | 8.05E-154 | 0.38489731 | 0.782 | 0.644 | 3.08E-149 | ExN2 | CAMK2N1 |
| SLC35F1 | 5.05E-153 | 0.37351945 | 0.865 | 0.694 | 1.93E-148 | ExN2 | SLC35F1 |
| RETREG1 | 6.45E-152 | 0.37449822 | 0.725 | 0.565 | 2.46E-147 | ExN2 | RETREG1 |
| ADORA1 | 1.98E-151 | 0.30277153 | 0.351 | 0.192 | 7.58E-147 | ExN2 | ADORA1 |
| ATP8A1 | 1.99E-150 | 0.32714729 | 0.909 | 0.846 | 7.62E-146 | ExN2 | ATP8A1 |
| JCAD | 4.54E-148 | 0.2633339 | 0.361 | 0.2 | 1.74E-143 | ExN2 | JCAD |
| MPPED21 | 8.74E-148 | 0.33456143 | 0.682 | 0.472 | 3.34E-143 | ExN2 | MPPED2 |
| DPY19L3 | 9.14E-146 | 0.29562913 | 0.434 | 0.263 | 3.49E-141 | ExN2 | DPY19L3 |
| TPK1 | 4.77E-145 | 0.42401302 | 0.592 | 0.429 | 1.82E-140 | ExN2 | TPK1 |
| FARP1 | 2.39E-144 | 0.35096178 | 0.785 | 0.646 | 9.14E-140 | ExN2 | FARP1 |
| KBTBD12 | 1.02E-143 | 0.31524164 | 0.417 | 0.248 | 3.90E-139 | ExN2 | KBTBD12 |
| PPFIA41 | 1.12E-143 | 0.39514518 | 0.686 | 0.539 | 4.27E-139 | ExN2 | PPFIA4 |
| HRH1 | 1.69E-143 | 0.29278537 | 0.38 | 0.217 | 6.44E-139 | ExN2 | HRH1 |
| SUN2 | 2.79E-143 | 0.27152518 | 0.345 | 0.189 | 1.07E-138 | ExN2 | SUN2 |
| KIAA0930 | 4.55E-142 | 0.4090914 | 0.73 | 0.603 | 1.74E-137 | ExN2 | KIAA0930 |
| PDZRN3 | 1.19E-139 | 0.3294356 | 0.895 | 0.75 | 4.54E-135 | ExN2 | PDZRN3 |
| C1orf115 | 5.43E-139 | 0.25777712 | 0.335 | 0.184 | 2.08E-134 | ExN2 | C1orf115 |
| DLG5 | 2.47E-138 | 0.30014651 | 0.469 | 0.298 | 9.43E-134 | ExN2 | DLG5 |
| MB21D2 | 1.10E-137 | 0.36399188 | 0.55 | 0.381 | 4.20E-133 | ExN2 | MB21D2 |
| TTYH1 | 1.55E-137 | 0.27878301 | 0.364 | 0.207 | 5.91E-133 | ExN2 | TTYH1 |
| CCNG2 | 1.15E-136 | 0.3589276 | 0.418 | 0.252 | 4.38E-132 | ExN2 | CCNG2 |
| DIRC31 | 1.61E-136 | 0.36651532 | 0.71 | 0.529 | 6.14E-132 | ExN2 | DIRC3 |
| TBR1 | 2.22E-136 | 0.29127753 | 0.384 | 0.225 | 8.50E-132 | ExN2 | TBR1 |
| AHR | 2.24E-136 | 0.29878852 | 0.406 | 0.247 | 8.56E-132 | ExN2 | AHR |
| SLC6A71 | 2.26E-136 | 0.31220873 | 0.481 | 0.309 | 8.62E-132 | ExN2 | SLC6A7 |
| AC103855.3 | 3.77E-135 | 0.28265518 | 0.293 | 0.151 | 1.44E-130 | ExN2 | AC103855.3 |
| BASP1-AS1 | 7.45E-135 | 0.34509911 | 0.816 | 0.644 | 2.85E-130 | ExN2 | BASP1-AS1 |
| PFN2 | 6.79E-134 | 0.33951338 | 0.437 | 0.282 | 2.60E-129 | ExN2 | PFN2 |
| MMP16 | 7.82E-133 | 0.30960668 | 0.919 | 0.788 | 2.99E-128 | ExN2 | MMP16 |
| PCDH9 | 8.26E-130 | 0.29268803 | 0.997 | 0.964 | 3.16E-125 | ExN2 | PCDH9 |
| CCDC102B | 1.04E-129 | 0.37249983 | 0.518 | 0.357 | 3.99E-125 | ExN2 | CCDC102B |
| KCNIP4-IT11 | 1.42E-129 | 0.3463654 | 0.684 | 0.485 | 5.44E-125 | ExN2 | KCNIP4-IT1 |
| CREB5 | 8.91E-129 | 0.43263136 | 0.415 | 0.258 | 3.40E-124 | ExN2 | CREB5 |
| C5orf30 | 1.79E-128 | 0.31478392 | 0.45 | 0.294 | 6.83E-124 | ExN2 | C5orf30 |
| MAP4K4 | 1.30E-127 | 0.36566426 | 0.807 | 0.715 | 4.97E-123 | ExN2 | MAP4K4 |
| MAGI3 | 1.31E-127 | 0.30165375 | 0.802 | 0.677 | 5.00E-123 | ExN2 | MAGI3 |
| KCTD8 | 4.07E-127 | 0.30812713 | 0.816 | 0.644 | 1.55E-122 | ExN2 | KCTD8 |
| NMT2 | 1.04E-126 | 0.34693645 | 0.633 | 0.482 | 3.97E-122 | ExN2 | NMT2 |
| AC110614.11 | 3.68E-126 | 0.30907245 | 0.321 | 0.173 | 1.41E-121 | ExN2 | AC110614.1 |
| TCF7L2 | 7.75E-126 | 0.34031124 | 0.422 | 0.265 | 2.96E-121 | ExN2 | TCF7L2 |
| STOX2 | 2.72E-124 | 0.31980155 | 0.859 | 0.791 | 1.04E-119 | ExN2 | STOX2 |
| MCHR21 | 2.26E-123 | 0.25231005 | 0.431 | 0.261 | 8.62E-119 | ExN2 | MCHR2 |
| TLCD4 | 5.16E-123 | 0.30417267 | 0.418 | 0.269 | 1.97E-118 | ExN2 | TLCD4 |
| SPEF2 | 1.35E-121 | 0.32989912 | 0.565 | 0.41 | 5.16E-117 | ExN2 | SPEF2 |
| MINDY4 | 2.63E-121 | 0.33369283 | 0.44 | 0.286 | 1.00E-116 | ExN2 | MINDY4 |
| ADAMTS19 | 5.90E-121 | 0.27822843 | 0.536 | 0.352 | 2.25E-116 | ExN2 | ADAMTS19 |
| AQP4-AS11 | 5.95E-121 | 0.32525554 | 0.687 | 0.485 | 2.27E-116 | ExN2 | AQP4-AS1 |
| MSRB3 | 1.01E-120 | 0.32099159 | 0.481 | 0.32 | 3.86E-116 | ExN2 | MSRB3 |
| RCL1 | 1.59E-119 | 0.30471586 | 0.46 | 0.304 | 6.08E-115 | ExN2 | RCL1 |
| SORL1 | 2.29E-119 | 0.31753993 | 0.785 | 0.669 | 8.76E-115 | ExN2 | SORL1 |
| STX1A | 4.36E-119 | 0.30890677 | 0.517 | 0.363 | 1.67E-114 | ExN2 | STX1A |
| CEP112 | 1.80E-118 | 0.34439027 | 0.723 | 0.581 | 6.89E-114 | ExN2 | CEP112 |
| JAKMIP2 | 2.94E-118 | 0.30896665 | 0.892 | 0.83 | 1.13E-113 | ExN2 | JAKMIP2 |
| NME9 | 3.39E-118 | 0.34667536 | 0.478 | 0.323 | 1.30E-113 | ExN2 | NME9 |
| LRRN1 | 4.20E-118 | 0.30456607 | 0.506 | 0.344 | 1.60E-113 | ExN2 | LRRN1 |
| CHGA | 2.17E-117 | 0.32521083 | 0.507 | 0.358 | 8.28E-113 | ExN2 | CHGA |
| FHL21 | 2.39E-117 | 0.26501961 | 0.407 | 0.255 | 9.13E-113 | ExN2 | FHL2 |
| SEC14L51 | 5.27E-117 | 0.29632761 | 0.611 | 0.436 | 2.02E-112 | ExN2 | SEC14L5 |
| AC124852.11 | 5.51E-117 | 0.34820932 | 0.628 | 0.465 | 2.11E-112 | ExN2 | AC124852.1 |
| ELMO1 | 6.28E-117 | 0.46728639 | 0.791 | 0.643 | 2.40E-112 | ExN2 | ELMO1 |
| AC010280.1 | 8.29E-117 | 0.33559627 | 0.403 | 0.253 | 3.17E-112 | ExN2 | AC010280.1 |
| PALM2-AKAF | 4.12E-116 | 0.36278818 | 0.795 | 0.694 | 1.57E-111 | ExN2 | PALM2-AKAP |

| | | | | | | | | |
|---|---|---|---|---|---|---|---|---|
| AC013652.11 | 1.13E-114 | 0.37695631 | 0.479 | 0.324 | 4.32E-110 | ExN2 | | AC013652.1 |
| VPS13D | 1.16E-114 | 0.30208851 | 0.833 | 0.744 | 4.44E-110 | ExN2 | | VPS13D |
| KCNA2 | 7.90E-114 | 0.3017485 | 0.479 | 0.329 | 3.02E-109 | ExN2 | | KCNA2 |
| SEC16B | 1.17E-112 | 0.2677638 | 0.327 | 0.195 | 4.48E-108 | ExN2 | | SEC16B |
| PRKCB1 | 1.18E-112 | 0.25871277 | 0.958 | 0.86 | 4.50E-108 | ExN2 | | PRKCB |
| DIPK2A | 3.60E-112 | 0.30797603 | 0.505 | 0.347 | 1.37E-107 | ExN2 | | DIPK2A |
| GTDC1 | 4.48E-112 | 0.32151458 | 0.866 | 0.785 | 1.71E-107 | ExN2 | | GTDC1 |
| KIAA1549 | 1.08E-111 | 0.32610264 | 0.646 | 0.514 | 4.13E-107 | ExN2 | | KIAA1549 |
| MT-ND1 | 1.65E-111 | 0.53456759 | 0.864 | 0.773 | 6.31E-107 | ExN2 | | MT-ND1 |
| CARNMT1 | 3.28E-111 | 0.28594512 | 0.466 | 0.321 | 1.26E-106 | ExN2 | | CARNMT1 |
| NPTXR1 | 5.41E-111 | 0.31056721 | 0.663 | 0.521 | 2.07E-106 | ExN2 | | NPTXR |
| UNC13B | 6.37E-111 | 0.26531113 | 0.53 | 0.369 | 2.43E-106 | ExN2 | | UNC13B |
| SIK3 | 9.09E-110 | 0.35908933 | 0.903 | 0.856 | 3.47E-105 | ExN2 | | SIK3 |
| GAP43 | 1.55E-109 | 0.289157 | 0.687 | 0.525 | 5.92E-105 | ExN2 | | GAP43 |
| TACC1 | 1.76E-109 | 0.31445255 | 0.735 | 0.629 | 6.73E-105 | ExN2 | | TACC1 |
| MAMLD1 | 2.56E-109 | 0.28704037 | 0.447 | 0.295 | 9.80E-105 | ExN2 | | MAMLD1 |
| CCND3 | 2.63E-109 | 0.34649135 | 0.528 | 0.376 | 1.00E-104 | ExN2 | | CCND3 |
| AC124854.1 | 6.34E-109 | 0.37140452 | 0.482 | 0.333 | 2.42E-104 | ExN2 | | AC124854.1 |
| TM7SF3 | 1.67E-108 | 0.31191919 | 0.537 | 0.408 | 6.40E-104 | ExN2 | | TM7SF3 |
| SYT13 | 1.86E-108 | 0.28828966 | 0.494 | 0.35 | 7.13E-104 | ExN2 | | SYT13 |
| RIMS3 | 5.84E-108 | 0.30514521 | 0.54 | 0.396 | 2.23E-103 | ExN2 | | RIMS3 |
| BRINP11 | 3.60E-107 | 0.28346381 | 0.836 | 0.736 | 1.38E-102 | ExN2 | | BRINP1 |
| PREP | 4.44E-107 | 0.30342323 | 0.574 | 0.425 | 1.70E-102 | ExN2 | | PREP |
| AC007038.1 | 5.23E-107 | 0.2545324 | 0.318 | 0.187 | 2.00E-102 | ExN2 | | AC007038.1 |
| FLRT21 | 8.57E-107 | 0.27135373 | 0.94 | 0.825 | 3.28E-102 | ExN2 | | FLRT2 |
| MAGI2-AS3 | 1.28E-106 | 0.29729835 | 0.498 | 0.356 | 4.89E-102 | ExN2 | | MAGI2-AS3 |
| MRTFB1 | 6.64E-106 | 0.28688605 | 0.863 | 0.752 | 2.54E-101 | ExN2 | | MRTFB |
| JAK2 | 1.98E-103 | 0.32297629 | 0.712 | 0.601 | 7.56E-99 | ExN2 | | JAK2 |
| B4GALNT3 | 3.78E-103 | 0.30474773 | 0.447 | 0.303 | 1.45E-98 | ExN2 | | B4GALNT3 |
| ARPP21 | 1.22E-102 | 0.27709624 | 0.959 | 0.748 | 4.68E-98 | ExN2 | | ARPP21 |
| PPP2R2C1 | 7.72E-102 | 0.28871192 | 0.889 | 0.793 | 2.95E-97 | ExN2 | | PPP2R2C |
| SLC16A7 | 9.09E-102 | 0.33232654 | 0.711 | 0.596 | 3.47E-97 | ExN2 | | SLC16A7 |
| AMPH1 | 7.21E-101 | 0.26119792 | 0.873 | 0.807 | 2.75E-96 | ExN2 | | AMPH |
| MSRA1 | 9.84E-101 | 0.25508793 | 0.961 | 0.903 | 3.76E-96 | ExN2 | | MSRA |
| LNX11 | 1.49E-100 | 0.27798682 | 0.736 | 0.59 | 5.70E-96 | ExN2 | | LNX1 |
| IFT122 | 4.20E-100 | 0.28994718 | 0.535 | 0.399 | 1.60E-95 | ExN2 | | IFT122 |
| OPRD1 | 4.29E-100 | 0.25001803 | 0.408 | 0.262 | 1.64E-95 | ExN2 | | OPRD1 |
| FUT9 | 1.16E-99 | 0.27465769 | 0.885 | 0.81 | 4.45E-95 | ExN2 | | FUT9 |
| SH3GL3 | 3.04E-99 | 0.27936817 | 0.878 | 0.816 | 1.16E-94 | ExN2 | | SH3GL3 |
| SASH1 | 3.68E-99 | 0.28808002 | 0.5 | 0.342 | 1.41E-94 | ExN2 | | SASH1 |
| HTR2A1 | 4.64E-99 | 0.26610924 | 0.477 | 0.322 | 1.77E-94 | ExN2 | | HTR2A |
| KIAA1549L1 | 7.59E-99 | 0.2657306 | 0.902 | 0.83 | 2.90E-94 | ExN2 | | KIAA1549L |
| SNX30 | 8.13E-99 | 0.29018484 | 0.509 | 0.37 | 3.11E-94 | ExN2 | | SNX30 |
| SHROOM3 | 1.15E-98 | 0.25796031 | 0.441 | 0.285 | 4.38E-94 | ExN2 | | SHROOM3 |
| SMARCC1 | 1.02E-97 | 0.27519441 | 0.614 | 0.488 | 3.89E-93 | ExN2 | | SMARCC1 |
| STX8 | 1.37E-97 | 0.32052489 | 0.673 | 0.546 | 5.24E-93 | ExN2 | | STX8 |
| AL513166.1 | 1.85E-97 | 0.37661757 | 0.741 | 0.613 | 7.08E-93 | ExN2 | | AL513166.1 |
| SEPTIN9 | 8.91E-97 | 0.30956588 | 0.483 | 0.351 | 3.41E-92 | ExN2 | | SEPTIN9 |
| IQCA1 | 1.04E-96 | 0.26526663 | 0.489 | 0.344 | 3.99E-92 | ExN2 | | IQCA1 |
| NDFIP21 | 2.04E-96 | 0.28066753 | 0.651 | 0.501 | 7.80E-92 | ExN2 | | NDFIP2 |
| ZEB1 | 4.26E-96 | 0.29129865 | 0.855 | 0.755 | 1.63E-91 | ExN2 | | ZEB1 |
| SESN3 | 4.89E-96 | 0.31792597 | 0.721 | 0.623 | 1.87E-91 | ExN2 | | SESN3 |
| AC104806.2 | 4.90E-96 | 0.30768592 | 0.603 | 0.472 | 1.87E-91 | ExN2 | | AC104806.2 |
| LRRTM3 | 2.05E-95 | 0.28580083 | 0.916 | 0.824 | 7.85E-91 | ExN2 | | LRRTM3 |
| MT-CO1 | 1.46E-94 | 0.47684313 | 0.919 | 0.863 | 5.58E-90 | ExN2 | | MT-CO1 |
| CTNND1 | 3.29E-94 | 0.29513778 | 0.591 | 0.463 | 1.26E-89 | ExN2 | | CTNND1 |
| FHIT | 5.86E-94 | 0.37195595 | 0.828 | 0.736 | 2.24E-89 | ExN2 | | FHIT |
| ST8SIA1 | 6.41E-94 | 0.28047073 | 0.691 | 0.547 | 2.45E-89 | ExN2 | | ST8SIA1 |
| DLGAP1-AS4 | 1.15E-93 | 0.25687614 | 0.492 | 0.342 | 4.41E-89 | ExN2 | | DLGAP1-AS4 |
| ASPH | 3.18E-93 | 0.30668645 | 0.697 | 0.591 | 1.22E-88 | ExN2 | | ASPH |
| STMN1 | 2.07E-92 | 0.36169991 | 0.587 | 0.466 | 7.90E-88 | ExN2 | | STMN1 |
| LZTS31 | 1.79E-91 | 0.28106809 | 0.554 | 0.422 | 6.85E-87 | ExN2 | | LZTS3 |
| ST6GAL2 | 3.91E-91 | 0.25727438 | 0.527 | 0.379 | 1.49E-86 | ExN2 | | ST6GAL2 |

| | | | | | | | |
|---|---|---|---|---|---|---|---|
| LINC01252 | 2.76E-90 | 0.2844337 | 0.434 | 0.304 | 1.05E-85 | ExN2 | LINC01252 |
| FANCC | 1.48E-89 | 0.28677275 | 0.336 | 0.213 | 5.68E-85 | ExN2 | FANCC |
| ST7 | 2.97E-89 | 0.30480823 | 0.718 | 0.619 | 1.13E-84 | ExN2 | ST7 |
| PDGFC | 2.36E-88 | 0.25477966 | 0.41 | 0.27 | 9.03E-84 | ExN2 | PDGFC |
| NCDN1 | 2.17E-87 | 0.3153677 | 0.625 | 0.514 | 8.30E-83 | ExN2 | NCDN |
| MT-CO3 | 4.21E-87 | 0.39568383 | 0.905 | 0.844 | 1.61E-82 | ExN2 | MT-CO3 |
| TNR | 4.93E-87 | 0.25633944 | 0.893 | 0.793 | 1.89E-82 | ExN2 | TNR |
| COL26A11 | 6.84E-87 | 0.25318405 | 0.622 | 0.467 | 2.61E-82 | ExN2 | COL26A1 |
| NFIC | 7.30E-86 | 0.27581308 | 0.627 | 0.501 | 2.79E-81 | ExN2 | NFIC |
| LYST1 | 9.25E-86 | 0.25458441 | 0.793 | 0.718 | 3.54E-81 | ExN2 | LYST |
| MMP17 | 6.00E-85 | 0.26781897 | 0.547 | 0.417 | 2.29E-80 | ExN2 | MMP17 |
| FNIP2 | 7.61E-85 | 0.25665875 | 0.721 | 0.605 | 2.91E-80 | ExN2 | FNIP2 |
| CENPC | 1.15E-84 | 0.28413905 | 0.672 | 0.564 | 4.38E-80 | ExN2 | CENPC |
| NCEH1 | 2.62E-84 | 0.27711718 | 0.62 | 0.501 | 1.00E-79 | ExN2 | NCEH1 |
| AGAP3 | 7.99E-84 | 0.27706885 | 0.67 | 0.556 | 3.06E-79 | ExN2 | AGAP3 |
| PRB2 | 9.66E-84 | 0.26955247 | 0.362 | 0.241 | 3.69E-79 | ExN2 | PRB2 |
| PDP1 | 5.23E-83 | 0.25825498 | 0.441 | 0.316 | 2.00E-78 | ExN2 | PDP1 |
| PDE10A | 6.73E-83 | 0.37293091 | 0.743 | 0.671 | 2.57E-78 | ExN2 | PDE10A |
| CDYL21 | 6.91E-83 | 0.25168257 | 0.533 | 0.394 | 2.64E-78 | ExN2 | CDYL2 |
| SMAP1 | 9.08E-83 | 0.30233569 | 0.72 | 0.619 | 3.47E-78 | ExN2 | SMAP1 |
| RNF220 | 1.10E-82 | 0.69441071 | 0.358 | 0.242 | 4.19E-78 | ExN2 | RNF220 |
| BASP1 | 1.19E-81 | 0.25660407 | 0.841 | 0.765 | 4.56E-77 | ExN2 | BASP1 |
| AKT2 | 2.55E-80 | 0.25917765 | 0.552 | 0.432 | 9.74E-76 | ExN2 | AKT2 |
| LINC00970 | 2.89E-80 | 0.2689854 | 0.531 | 0.394 | 1.11E-75 | ExN2 | LINC00970 |
| IGSF9B | 3.53E-79 | 0.26744059 | 0.626 | 0.507 | 1.35E-74 | ExN2 | IGSF9B |
| MT-ATP6 | 8.85E-79 | 0.44745842 | 0.925 | 0.874 | 3.38E-74 | ExN2 | MT-ATP6 |
| PLCL1 | 1.15E-77 | 0.31653756 | 0.786 | 0.636 | 4.39E-73 | ExN2 | PLCL1 |
| MT-CYB | 1.97E-77 | 0.35976447 | 0.869 | 0.778 | 7.53E-73 | ExN2 | MT-CYB |
| ARMC91 | 1.22E-76 | 0.26843626 | 0.731 | 0.604 | 4.67E-72 | ExN2 | ARMC9 |
| RNLS | 1.93E-76 | 0.30772452 | 0.584 | 0.461 | 7.38E-72 | ExN2 | RNLS |
| TMEM161B-/ | 2.52E-76 | 0.25881739 | 0.916 | 0.861 | 9.64E-72 | ExN2 | TMEM161B-/ |
| ELK1 | 4.71E-76 | 0.2572928 | 0.328 | 0.214 | 1.80E-71 | ExN2 | ELK1 |
| SDCCAG8 | 4.86E-75 | 0.25594546 | 0.792 | 0.707 | 1.86E-70 | ExN2 | SDCCAG8 |
| PTMA | 7.73E-75 | 0.25051045 | 0.578 | 0.456 | 2.95E-70 | ExN2 | PTMA |
| APBB2 | 8.51E-75 | 0.27133859 | 0.769 | 0.683 | 3.25E-70 | ExN2 | APBB2 |
| TSPYL1 | 1.27E-73 | 0.27962134 | 0.604 | 0.497 | 4.87E-69 | ExN2 | TSPYL1 |
| SLCO3A1 | 1.87E-73 | 0.26650613 | 0.816 | 0.728 | 7.14E-69 | ExN2 | SLCO3A1 |
| PPP3R1 | 2.05E-73 | 0.28234864 | 0.717 | 0.617 | 7.84E-69 | ExN2 | PPP3R1 |
| VWA8 | 5.59E-73 | 0.25015565 | 0.8 | 0.735 | 2.14E-68 | ExN2 | VWA8 |
| BSN1 | 7.42E-73 | 0.25901875 | 0.698 | 0.597 | 2.84E-68 | ExN2 | BSN |
| TPD52 | 1.69E-72 | 0.26130539 | 0.639 | 0.529 | 6.44E-68 | ExN2 | TPD52 |
| CATSPER2 | 2.46E-72 | 0.26537484 | 0.712 | 0.62 | 9.39E-68 | ExN2 | CATSPER2 |
| SYBU1 | 2.49E-72 | 0.25044744 | 0.801 | 0.707 | 9.51E-68 | ExN2 | SYBU |
| MTA31 | 3.34E-72 | 0.25517875 | 0.745 | 0.648 | 1.28E-67 | ExN2 | MTA3 |
| LYRM4 | 3.77E-72 | 0.27146564 | 0.59 | 0.476 | 1.44E-67 | ExN2 | LYRM4 |
| CMSS1 | 1.07E-71 | 0.25687012 | 0.805 | 0.72 | 4.09E-67 | ExN2 | CMSS1 |
| CLU | 3.22E-71 | 0.26969663 | 0.781 | 0.711 | 1.23E-66 | ExN2 | CLU |
| PKN2-AS1 | 4.14E-71 | 0.33431721 | 0.428 | 0.305 | 1.58E-66 | ExN2 | PKN2-AS1 |
| PLP1 | 4.98E-70 | 0.98014496 | 0.327 | 0.225 | 1.90E-65 | ExN2 | PLP1 |
| AC079382.2 | 1.01E-69 | 0.30377766 | 0.33 | 0.223 | 3.87E-65 | ExN2 | AC079382.2 |
| LINC01811 | 1.17E-69 | 0.30541464 | 0.421 | 0.299 | 4.47E-65 | ExN2 | LINC01811 |
| CLIP2 | 3.37E-69 | 0.29745266 | 0.68 | 0.567 | 1.29E-64 | ExN2 | CLIP2 |
| GPM6B1 | 1.12E-68 | 0.27943059 | 0.875 | 0.797 | 4.26E-64 | ExN2 | GPM6B |
| KCNQ2 | 1.11E-66 | 0.26394553 | 0.733 | 0.649 | 4.23E-62 | ExN2 | KCNQ2 |
| TUBA1B | 7.19E-65 | 0.30287937 | 0.64 | 0.561 | 2.75E-60 | ExN2 | TUBA1B |
| SEPTIN7 | 1.06E-63 | 0.30140328 | 0.741 | 0.689 | 4.04E-59 | ExN2 | SEPTIN7 |
| AC008591.1 | 4.18E-63 | 0.305098 | 0.373 | 0.264 | 1.60E-58 | ExN2 | AC008591.1 |
| MT-ND2 | 3.46E-61 | 0.3675975 | 0.825 | 0.741 | 1.32E-56 | ExN2 | MT-ND2 |
| TF | 1.03E-60 | 0.37423058 | 0.278 | 0.186 | 3.92E-56 | ExN2 | TF |
| DUBR | 1.70E-60 | 0.26039986 | 0.665 | 0.567 | 6.52E-56 | ExN2 | DUBR |
| TNNI3K | 6.65E-60 | 0.26870355 | 0.472 | 0.366 | 2.54E-55 | ExN2 | TNNI3K |
| CXXC4-AS1 | 1.51E-57 | 0.29838006 | 0.485 | 0.379 | 5.79E-53 | ExN2 | CXXC4-AS1 |
| TTLL7 | 2.35E-55 | 0.26726271 | 0.861 | 0.821 | 8.98E-51 | ExN2 | TTLL7 |

| | | | | | | | |
|---|---|---|---|---|---|---|---|
| MTRNR2L12 | 2.03E-50 | 0.34759477 | 0.823 | 0.74 | 7.78E-46 | ExN2 | MTRNR2L12 |
| MT-ND3 | 3.17E-50 | 0.35457921 | 0.914 | 0.866 | 1.21E-45 | ExN2 | MT-ND3 |
| ABCA2 | 6.53E-40 | 0.29837199 | 0.451 | 0.373 | 2.49E-35 | ExN2 | ABCA2 |
| PIP4K2A | 4.20E-36 | 0.58560783 | 0.485 | 0.422 | 1.60E-31 | ExN2 | PIP4K2A |
| AC106793.1 | 4.78E-33 | 0.27026914 | 0.446 | 0.37 | 1.83E-28 | ExN2 | AC106793.1 |
| PTGDS | 1.30E-29 | 0.33231599 | 0.351 | 0.281 | 4.95E-25 | ExN2 | PTGDS |
| QDPR | 1.92E-28 | 0.26190776 | 0.461 | 0.395 | 7.34E-24 | ExN2 | QDPR |
| PLEKHH1 | 6.14E-23 | 0.26559283 | 0.297 | 0.242 | 2.35E-18 | ExN2 | PLEKHH1 |
| SLC44A11 | 2.62E-19 | 0.38803511 | 0.77 | 0.715 | 1.00E-14 | ExN2 | SLC44A1 |
| TMEM165 | 5.10E-17 | 0.25056104 | 0.457 | 0.417 | 1.95E-12 | ExN2 | TMEM165 |
| PHLPP1 | 4.85E-05 | 0.34134804 | 0.564 | 0.556 | 1 | ExN2 | PHLPP1 |
| AJ009632.2 | 0 | 2.06134853 | 0.72 | 0.118 | 0 | ExN3 | AJ009632.2 |
| RMST | 0 | 1.76456582 | 0.642 | 0.126 | 0 | ExN3 | RMST |
| TAFA11 | 0 | 1.44300908 | 0.935 | 0.48 | 0 | ExN3 | TAFA1 |
| GLIS31 | 0 | 1.38957926 | 0.641 | 0.241 | 0 | ExN3 | GLIS3 |
| CA101 | 0 | 1.34448392 | 0.905 | 0.398 | 0 | ExN3 | CA10 |
| COL5A21 | 0 | 1.34432391 | 0.603 | 0.096 | 0 | ExN3 | COL5A2 |
| AC034268.21 | 0 | 1.21063307 | 0.629 | 0.259 | 0 | ExN3 | AC034268.2 |
| CUX21 | 0 | 1.13906037 | 0.788 | 0.36 | 0 | ExN3 | CUX2 |
| RIT2 | 0 | 1.09626605 | 0.713 | 0.288 | 0 | ExN3 | RIT2 |
| ANO31 | 0 | 1.04665225 | 0.825 | 0.36 | 0 | ExN3 | ANO3 |
| AL157944.11 | 0 | 1.02130539 | 0.583 | 0.25 | 0 | ExN3 | AL157944.1 |
| NTNG1 | 0 | 1.0202859 | 0.776 | 0.241 | 0 | ExN3 | NTNG1 |
| PTPRD1 | 0 | 0.9952657 | 0.995 | 0.962 | 0 | ExN3 | PTPRD |
| LAMA2 | 0 | 0.98972486 | 0.39 | 0.131 | 0 | ExN3 | LAMA2 |
| PTPRK1 | 0 | 0.96962975 | 0.945 | 0.576 | 0 | ExN3 | PTPRK |
| RORB1 | 0 | 0.95641103 | 0.804 | 0.289 | 0 | ExN3 | RORB |
| AC073365.12 | 0 | 0.92719417 | 0.78 | 0.346 | 0 | ExN3 | AC073365.1 |
| IQCJ-SCHIP12 | 0 | 0.90940207 | 0.984 | 0.821 | 0 | ExN3 | IQCJ-SCHIP1 |
| ST6GALNAC5 | 0 | 0.88508832 | 0.953 | 0.691 | 0 | ExN3 | ST6GALNAC5 |
| LINC022961 | 0 | 0.88431155 | 0.481 | 0.136 | 0 | ExN3 | LINC02296 |
| LHFPL61 | 0 | 0.87549998 | 0.84 | 0.385 | 0 | ExN3 | LHFPL6 |
| OTOGL | 0 | 0.86221371 | 0.454 | 0.171 | 0 | ExN3 | OTOGL |
| POU6F21 | 0 | 0.85471076 | 0.735 | 0.374 | 0 | ExN3 | POU6F2 |
| CPNE4 | 0 | 0.84168948 | 0.837 | 0.437 | 0 | ExN3 | CPNE4 |
| LRRC4C | 0 | 0.84076835 | 0.966 | 0.883 | 0 | ExN3 | LRRC4C |
| NECAB11 | 0 | 0.83855519 | 0.95 | 0.617 | 0 | ExN3 | NECAB1 |
| AP000829.11 | 0 | 0.83570939 | 0.604 | 0.176 | 0 | ExN3 | AP000829.1 |
| TCERG1L1 | 0 | 0.82366663 | 0.746 | 0.308 | 0 | ExN3 | TCERG1L |
| AC019211.11 | 0 | 0.80753632 | 0.782 | 0.314 | 0 | ExN3 | AC019211.1 |
| AC067956.11 | 0 | 0.80507883 | 0.746 | 0.287 | 0 | ExN3 | AC067956.1 |
| AC120193.1 | 0 | 0.77936426 | 0.641 | 0.299 | 0 | ExN3 | AC120193.1 |
| LINC018211 | 0 | 0.76995993 | 0.508 | 0.189 | 0 | ExN3 | LINC01821 |
| RFX31 | 0 | 0.75800742 | 0.923 | 0.79 | 0 | ExN3 | RFX3 |
| SGCD1 | 0 | 0.75522546 | 0.8 | 0.573 | 0 | ExN3 | SGCD |
| MSC-AS11 | 0 | 0.7538599 | 0.586 | 0.197 | 0 | ExN3 | MSC-AS1 |
| DCC1 | 0 | 0.74827084 | 0.815 | 0.521 | 0 | ExN3 | DCC |
| SCHLAP11 | 0 | 0.74761687 | 0.517 | 0.188 | 0 | ExN3 | SCHLAP1 |
| PKN2-AS11 | 0 | 0.70833598 | 0.618 | 0.287 | 0 | ExN3 | PKN2-AS1 |
| MARCH11 | 0 | 0.70295997 | 0.959 | 0.741 | 0 | ExN3 | 1-Mar |
| GRM71 | 0 | 0.69352136 | 0.975 | 0.853 | 0 | ExN3 | GRM7 |
| PTPRT1 | 0 | 0.68967298 | 0.933 | 0.662 | 0 | ExN3 | PTPRT |
| SHROOM31 | 0 | 0.689038 | 0.632 | 0.267 | 0 | ExN3 | SHROOM3 |
| OLFM31 | 0 | 0.6799534 | 0.952 | 0.694 | 0 | ExN3 | OLFM3 |
| LDB22 | 0 | 0.67648803 | 0.979 | 0.746 | 0 | ExN3 | LDB2 |
| FAT31 | 0 | 0.65738853 | 0.939 | 0.694 | 0 | ExN3 | FAT3 |
| HS6ST31 | 0 | 0.6569427 | 0.965 | 0.729 | 0 | ExN3 | HS6ST3 |
| R3HDM12 | 0 | 0.65461753 | 0.983 | 0.885 | 0 | ExN3 | R3HDM1 |
| KCNB2 | 0 | 0.6490232 | 0.91 | 0.667 | 0 | ExN3 | KCNB2 |
| KCNIP42 | 0 | 0.64781645 | 1 | 0.932 | 0 | ExN3 | KCNIP4 |
| MED12L1 | 0 | 0.64428238 | 0.897 | 0.694 | 0 | ExN3 | MED12L |
| AC007368.1 | 0 | 0.63516968 | 0.718 | 0.417 | 0 | ExN3 | AC007368.1 |
| CNGB11 | 0 | 0.62565386 | 0.443 | 0.097 | 0 | ExN3 | CNGB1 |

| | | | | | | | |
|---|---|---|---|---|---|---|---|
| CARMIL11 | 0 | 0.62443193 | 0.896 | 0.597 | 0 | ExN3 | CARMIL1 |
| TRHDE1 | 0 | 0.61105304 | 0.833 | 0.492 | 0 | ExN3 | TRHDE |
| GAP431 | 0 | 0.60700656 | 0.823 | 0.513 | 0 | ExN3 | GAP43 |
| AC246817.11 | 0 | 0.60045724 | 0.403 | 0.117 | 0 | ExN3 | AC246817.1 |
| EPHA42 | 0 | 0.59739822 | 0.912 | 0.657 | 0 | ExN3 | EPHA4 |
| RAPGEF51 | 0 | 0.5973864 | 0.947 | 0.735 | 0 | ExN3 | RAPGEF5 |
| FMN1 | 0 | 0.59530899 | 0.938 | 0.744 | 0 | ExN3 | FMN1 |
| COL26A12 | 0 | 0.58974762 | 0.757 | 0.455 | 0 | ExN3 | COL26A1 |
| PCDH91 | 0 | 0.58203353 | 0.996 | 0.965 | 0 | ExN3 | PCDH9 |
| LMO41 | 0 | 0.58112759 | 0.834 | 0.503 | 0 | ExN3 | LMO4 |
| DLGAP21 | 0 | 0.57822261 | 0.984 | 0.926 | 0 | ExN3 | DLGAP2 |
| NRGN2 | 0 | 0.57583849 | 0.888 | 0.547 | 0 | ExN3 | NRGN |
| SNED12 | 0 | 0.57412975 | 0.749 | 0.417 | 0 | ExN3 | SNED1 |
| DPYD1 | 0 | 0.57318576 | 0.913 | 0.645 | 0 | ExN3 | DPYD |
| KHDRBS31 | 0 | 0.57102215 | 0.957 | 0.783 | 0 | ExN3 | KHDRBS3 |
| AC008574.11 | 0 | 0.5658492 | 0.587 | 0.192 | 0 | ExN3 | AC008574.1 |
| AC018697.1 | 0 | 0.5633169 | 0.394 | 0.105 | 0 | ExN3 | AC018697.1 |
| UNC5D1 | 0 | 0.56315161 | 0.965 | 0.819 | 0 | ExN3 | UNC5D |
| FAM13A1 | 0 | 0.56274309 | 0.834 | 0.582 | 0 | ExN3 | FAM13A |
| RHOBTB1 | 0 | 0.56129162 | 0.592 | 0.263 | 0 | ExN3 | RHOBTB1 |
| ADAMTS31 | 0 | 0.55486729 | 0.558 | 0.242 | 0 | ExN3 | ADAMTS3 |
| SATB22 | 0 | 0.55131599 | 0.91 | 0.548 | 0 | ExN3 | SATB2 |
| PLCL11 | 0 | 0.54664311 | 0.899 | 0.626 | 0 | ExN3 | PLCL1 |
| MRTFB2 | 0 | 0.544506 | 0.914 | 0.748 | 0 | ExN3 | MRTFB |
| KHDRBS21 | 0 | 0.54381517 | 0.978 | 0.893 | 0 | ExN3 | KHDRBS2 |
| KCNH12 | 0 | 0.5425387 | 0.96 | 0.82 | 0 | ExN3 | KCNH1 |
| GRM31 | 0 | 0.53832798 | 0.764 | 0.366 | 0 | ExN3 | GRM3 |
| USP431 | 0 | 0.53781655 | 0.588 | 0.23 | 0 | ExN3 | USP43 |
| PTK2B1 | 0 | 0.53713972 | 0.836 | 0.493 | 0 | ExN3 | PTK2B |
| AL162493.1 | 0 | 0.5370742 | 0.312 | 0.087 | 0 | ExN3 | AL162493.1 |
| AL392023.2 | 0 | 0.53688522 | 0.409 | 0.131 | 0 | ExN3 | AL392023.2 |
| HECW11 | 0 | 0.53396918 | 0.961 | 0.822 | 0 | ExN3 | HECW1 |
| CDH221 | 0 | 0.53171633 | 0.728 | 0.376 | 0 | ExN3 | CDH22 |
| KIAA1211L1 | 0 | 0.53132619 | 0.867 | 0.539 | 0 | ExN3 | KIAA1211L |
| AC124290.1 | 0 | 0.5302809 | 0.3 | 0.102 | 0 | ExN3 | AC124290.1 |
| CFAP299 | 0 | 0.52804378 | 0.471 | 0.178 | 0 | ExN3 | CFAP299 |
| KCNH51 | 0 | 0.5230223 | 0.758 | 0.388 | 0 | ExN3 | KCNH5 |
| NKAIN21 | 0 | 0.51849237 | 0.983 | 0.827 | 0 | ExN3 | NKAIN2 |
| MYO1D1 | 0 | 0.5109928 | 0.796 | 0.494 | 0 | ExN3 | MYO1D |
| GAB11 | 0 | 0.51079177 | 0.829 | 0.524 | 0 | ExN3 | GAB1 |
| AC093843.11 | 0 | 0.50640196 | 0.536 | 0.191 | 0 | ExN3 | AC093843.1 |
| TNNT21 | 0 | 0.49824765 | 0.43 | 0.1 | 0 | ExN3 | TNNT2 |
| ROS11 | 0 | 0.49819429 | 0.379 | 0.104 | 0 | ExN3 | ROS1 |
| MIR99AHG | 0 | 0.4969544 | 0.97 | 0.888 | 0 | ExN3 | MIR99AHG |
| MYLK | 0 | 0.4961564 | 0.281 | 0.061 | 0 | ExN3 | MYLK |
| OPRM1 | 0 | 0.49501508 | 0.538 | 0.204 | 0 | ExN3 | OPRM1 |
| ZBTB7C2 | 0 | 0.49338547 | 0.571 | 0.26 | 0 | ExN3 | ZBTB7C |
| ZBBX | 0 | 0.4869581 | 0.487 | 0.209 | 0 | ExN3 | ZBBX |
| AC117453.1 | 0 | 0.4854996 | 0.288 | 0.076 | 0 | ExN3 | AC117453.1 |
| COL6A11 | 0 | 0.48512009 | 0.533 | 0.22 | 0 | ExN3 | COL6A1 |
| MIAT2 | 0 | 0.48279853 | 0.977 | 0.874 | 0 | ExN3 | MIAT |
| LRRK11 | 0 | 0.47910526 | 0.694 | 0.298 | 0 | ExN3 | LRRK1 |
| CELF21 | 0 | 0.47458774 | 0.989 | 0.966 | 0 | ExN3 | CELF2 |
| PCED1B | 0 | 0.46937333 | 0.365 | 0.112 | 0 | ExN3 | PCED1B |
| BAIAP3 | 0 | 0.46324367 | 0.56 | 0.25 | 0 | ExN3 | BAIAP3 |
| HTR2A2 | 0 | 0.45481604 | 0.612 | 0.31 | 0 | ExN3 | HTR2A |
| MET1 | 0 | 0.45355468 | 0.367 | 0.076 | 0 | ExN3 | MET |
| LINC02217 | 0 | 0.45310584 | 0.514 | 0.203 | 0 | ExN3 | LINC02217 |
| ADGRB31 | 0 | 0.42996673 | 0.996 | 0.992 | 0 | ExN3 | ADGRB3 |
| ANTXR2 | 0 | 0.42365702 | 0.328 | 0.069 | 0 | ExN3 | ANTXR2 |
| SLC38A112 | 0 | 0.4040737 | 0.528 | 0.229 | 0 | ExN3 | SLC38A11 |
| PRDM8 | 0 | 0.38273114 | 0.386 | 0.121 | 0 | ExN3 | PRDM8 |
| NRP1 | 0 | 0.36768568 | 0.466 | 0.182 | 0 | ExN3 | NRP1 |

| | | | | | | | |
|---|---|---|---|---|---|---|---|
| LINC012021 | 0 | 0.3620188 | 0.382 | 0.141 | 0 | ExN3 | LINC01202 |
| GRB14 | 0 | 0.33381291 | 0.319 | 0.086 | 0 | ExN3 | GRB14 |
| PRSS31 | 0 | 0.32933957 | 0.434 | 0.177 | 0 | ExN3 | PRSS3 |
| MIR3976HG | 0 | 0.26232543 | 0.252 | 0.054 | 0 | ExN3 | MIR3976HG |
| MLIP2 | 2.37E-306 | 0.51431882 | 0.836 | 0.487 | 9.05E-302 | ExN3 | MLIP |
| ARHGEF28 | 4.17E-306 | 0.37822679 | 0.591 | 0.286 | 1.59E-301 | ExN3 | ARHGEF28 |
| SLC30A3 | 6.92E-303 | 0.32584703 | 0.426 | 0.178 | 2.65E-298 | ExN3 | SLC30A3 |
| MAGI21 | 1.46E-301 | 0.40759607 | 0.995 | 0.986 | 5.56E-297 | ExN3 | MAGI2 |
| GULP11 | 5.63E-296 | 0.50129688 | 0.621 | 0.329 | 2.15E-291 | ExN3 | GULP1 |
| IL12A-AS11 | 3.67E-295 | 0.36100514 | 0.403 | 0.165 | 1.40E-290 | ExN3 | IL12A-AS1 |
| HDAC9 | 1.00E-294 | 0.51599557 | 0.919 | 0.766 | 3.84E-290 | ExN3 | HDAC9 |
| TRIM54 | 6.75E-294 | 0.27422462 | 0.316 | 0.111 | 2.58E-289 | ExN3 | TRIM54 |
| FLRT22 | 2.23E-293 | 0.52023257 | 0.958 | 0.825 | 8.52E-289 | ExN3 | FLRT2 |
| CDH8 | 2.51E-293 | 0.5299765 | 0.904 | 0.639 | 9.60E-289 | ExN3 | CDH8 |
| MGAT5B2 | 1.18E-291 | 0.45331737 | 0.751 | 0.454 | 4.49E-287 | ExN3 | MGAT5B |
| AC011995.21 | 1.43E-290 | 0.4365252 | 0.69 | 0.386 | 5.48E-286 | ExN3 | AC011995.2 |
| AL133166.11 | 8.26E-290 | 0.42293797 | 0.453 | 0.202 | 3.16E-285 | ExN3 | AL133166.1 |
| TMEM132D1 | 1.62E-285 | 0.51726192 | 0.948 | 0.768 | 6.20E-281 | ExN3 | TMEM132D |
| FOXP11 | 5.64E-285 | 0.51557388 | 0.948 | 0.809 | 2.15E-280 | ExN3 | FOXP1 |
| CRYM | 2.15E-284 | 0.32972242 | 0.52 | 0.247 | 8.21E-280 | ExN3 | CRYM |
| AC013287.11 | 5.31E-284 | 0.81813336 | 0.692 | 0.429 | 2.03E-279 | ExN3 | AC013287.1 |
| RGS7 | 7.19E-279 | 0.43660741 | 0.981 | 0.955 | 2.75E-274 | ExN3 | RGS7 |
| SH3GL22 | 2.94E-277 | 0.50627637 | 0.938 | 0.729 | 1.12E-272 | ExN3 | SH3GL2 |
| LINC012501 | 4.71E-276 | 0.47683845 | 0.883 | 0.624 | 1.80E-271 | ExN3 | LINC01250 |
| AC016598.1 | 6.82E-275 | 0.30049819 | 0.319 | 0.116 | 2.61E-270 | ExN3 | AC016598.1 |
| SOX51 | 9.18E-275 | 0.45815104 | 0.951 | 0.645 | 3.51E-270 | ExN3 | SOX5 |
| CTNND11 | 3.21E-273 | 0.42974042 | 0.716 | 0.452 | 1.23E-268 | ExN3 | CTNND1 |
| CACNA2D31 | 1.67E-269 | 0.49460133 | 0.966 | 0.859 | 6.37E-265 | ExN3 | CACNA2D3 |
| STXBP5L1 | 5.37E-269 | 0.52800616 | 0.972 | 0.946 | 2.05E-264 | ExN3 | STXBP5L |
| XYLT1 | 3.55E-268 | 0.45956275 | 0.815 | 0.537 | 1.36E-263 | ExN3 | XYLT1 |
| PCBP3 | 1.31E-267 | 0.52310254 | 0.388 | 0.165 | 5.00E-263 | ExN3 | PCBP3 |
| CNKSR22 | 8.01E-265 | 0.4968963 | 0.948 | 0.81 | 3.06E-260 | ExN3 | CNKSR2 |
| CADPS2 | 3.25E-263 | 0.50991245 | 0.838 | 0.567 | 1.24E-258 | ExN3 | CADPS2 |
| AKAP13 | 3.67E-263 | 0.38614541 | 0.624 | 0.344 | 1.40E-258 | ExN3 | AKAP13 |
| KCNH7 | 1.64E-262 | 0.50404156 | 0.936 | 0.712 | 6.28E-258 | ExN3 | KCNH7 |
| NEGR1 | 1.06E-260 | 0.45125872 | 0.99 | 0.953 | 4.06E-256 | ExN3 | NEGR1 |
| IQCA11 | 2.27E-258 | 0.38730589 | 0.612 | 0.333 | 8.67E-254 | ExN3 | IQCA1 |
| UBE2E22 | 1.04E-257 | 0.41855662 | 0.953 | 0.871 | 3.99E-253 | ExN3 | UBE2E2 |
| SEMA6D | 1.53E-257 | 0.37149629 | 0.852 | 0.559 | 5.86E-253 | ExN3 | SEMA6D |
| FAM155A | 1.90E-257 | 0.38133605 | 0.995 | 0.985 | 7.28E-253 | ExN3 | FAM155A |
| KIAA20121 | 2.80E-257 | 0.33603606 | 0.386 | 0.164 | 1.07E-252 | ExN3 | KIAA2012 |
| ZBTB46 | 6.35E-257 | 0.35567042 | 0.535 | 0.272 | 2.43E-252 | ExN3 | ZBTB46 |
| PLCH1 | 1.15E-256 | 1.12209766 | 0.403 | 0.195 | 4.40E-252 | ExN3 | PLCH1 |
| CABP12 | 1.97E-247 | 0.48777923 | 0.894 | 0.688 | 7.55E-243 | ExN3 | CABP1 |
| PART1 | 1.31E-246 | 0.37261403 | 0.534 | 0.274 | 5.02E-242 | ExN3 | PART1 |
| FRMD6 | 5.21E-246 | 0.30877597 | 0.445 | 0.207 | 1.99E-241 | ExN3 | FRMD6 |
| LRFN52 | 1.11E-245 | 0.42978147 | 0.971 | 0.885 | 4.26E-241 | ExN3 | LRFN5 |
| DENND31 | 1.47E-244 | 0.31367598 | 0.601 | 0.324 | 5.63E-240 | ExN3 | DENND3 |
| AL807742.1 | 2.45E-241 | 0.31479368 | 0.317 | 0.123 | 9.38E-237 | ExN3 | AL807742.1 |
| UBE2E31 | 1.81E-238 | 0.52202166 | 0.765 | 0.532 | 6.94E-234 | ExN3 | UBE2E3 |
| CCDC31 | 1.53E-236 | 0.47113968 | 0.861 | 0.629 | 5.84E-232 | ExN3 | CCDC3 |
| PAG11 | 2.95E-236 | 0.33365327 | 0.625 | 0.352 | 1.13E-231 | ExN3 | PAG1 |
| LY86-AS12 | 1.30E-235 | 0.47126213 | 0.766 | 0.461 | 4.96E-231 | ExN3 | LY86-AS1 |
| LINC003261 | 1.46E-234 | 0.47609155 | 0.276 | 0.101 | 5.59E-230 | ExN3 | LINC00326 |
| AGBL41 | 2.16E-231 | 0.43304142 | 0.982 | 0.929 | 8.27E-227 | ExN3 | AGBL4 |
| DLGAP1-AS4 | 4.96E-231 | 0.38933157 | 0.598 | 0.333 | 1.90E-226 | ExN3 | DLGAP1-AS4 |
| STYK11 | 1.80E-230 | 0.27673944 | 0.367 | 0.159 | 6.89E-226 | ExN3 | STYK1 |
| EFNA51 | 4.90E-229 | 0.50148377 | 0.958 | 0.784 | 1.87E-224 | ExN3 | EFNA5 |
| MAPK10 | 2.08E-228 | 0.33377985 | 0.979 | 0.95 | 7.95E-224 | ExN3 | MAPK10 |
| NELL22 | 5.48E-226 | 0.44856206 | 0.957 | 0.776 | 2.09E-221 | ExN3 | NELL2 |
| MIR137HG1 | 2.15E-224 | 0.45007105 | 0.899 | 0.641 | 8.23E-220 | ExN3 | MIR137HG |
| GUCY1A22 | 4.40E-224 | 0.49618986 | 0.91 | 0.716 | 1.68E-219 | ExN3 | GUCY1A2 |
| MPPED22 | 1.98E-223 | 0.39968903 | 0.753 | 0.467 | 7.56E-219 | ExN3 | MPPED2 |

| | | | | | | | |
|---|---|---|---|---|---|---|---|
| ST6GAL21 | 3.16E-223 | 0.45764381 | 0.629 | 0.37 | 1.21E-218 | ExN3 | ST6GAL2 |
| AC002460.21 | 4.59E-223 | 0.59659099 | 0.639 | 0.405 | 1.75E-218 | ExN3 | AC002460.2 |
| AC124852.12 | 1.16E-221 | 0.41120622 | 0.728 | 0.457 | 4.42E-217 | ExN3 | AC124852.1 |
| LINC013311 | 4.59E-220 | 0.45937668 | 0.446 | 0.217 | 1.76E-215 | ExN3 | LINC01331 |
| SLC17A72 | 7.43E-220 | 0.35463615 | 0.706 | 0.415 | 2.84E-215 | ExN3 | SLC17A7 |
| SYNPO2 | 1.53E-219 | 0.32753018 | 0.691 | 0.405 | 5.85E-215 | ExN3 | SYNPO2 |
| FNDC3B | 1.58E-219 | 0.33231349 | 0.581 | 0.324 | 6.04E-215 | ExN3 | FNDC3B |
| PRAG11 | 4.44E-219 | 0.29409629 | 0.426 | 0.202 | 1.70E-214 | ExN3 | PRAG1 |
| ATRNL12 | 1.44E-218 | 0.44982484 | 0.983 | 0.926 | 5.50E-214 | ExN3 | ATRNL1 |
| AC107223.12 | 2.74E-217 | 0.70708647 | 0.73 | 0.548 | 1.05E-212 | ExN3 | AC107223.1 |
| DNAH71 | 3.22E-217 | 0.36200041 | 0.631 | 0.373 | 1.23E-212 | ExN3 | DNAH7 |
| AL024495.1 | 9.88E-217 | 0.3543702 | 0.535 | 0.283 | 3.77E-212 | ExN3 | AL024495.1 |
| ILDR2 | 7.25E-215 | 0.30351545 | 0.585 | 0.322 | 2.77E-210 | ExN3 | ILDR2 |
| RYR21 | 8.26E-214 | 0.38813382 | 0.986 | 0.923 | 3.16E-209 | ExN3 | RYR2 |
| LINC02254 | 8.57E-213 | 0.29638226 | 0.347 | 0.15 | 3.27E-208 | ExN3 | LINC02254 |
| LINC02613 | 2.08E-212 | 0.25803202 | 0.332 | 0.141 | 7.95E-208 | ExN3 | LINC02613 |
| GABRB32 | 3.22E-211 | 0.35648385 | 0.968 | 0.895 | 1.23E-206 | ExN3 | GABRB3 |
| BASP1-AS11 | 3.47E-210 | 0.40730604 | 0.883 | 0.639 | 1.33E-205 | ExN3 | BASP1-AS1 |
| SV2B1 | 4.45E-210 | 0.41153399 | 0.9 | 0.583 | 1.70E-205 | ExN3 | SV2B |
| ST6GAL1 | 1.15E-208 | 0.26489562 | 0.494 | 0.255 | 4.39E-204 | ExN3 | ST6GAL1 |
| LGI41 | 1.15E-208 | 0.29942855 | 0.532 | 0.289 | 4.40E-204 | ExN3 | LGI4 |
| AL391840.11 | 9.05E-208 | 0.4400513 | 0.762 | 0.491 | 3.46E-203 | ExN3 | AL391840.1 |
| CSMD11 | 9.57E-208 | 0.33232597 | 0.997 | 0.98 | 3.66E-203 | ExN3 | CSMD1 |
| TESPA11 | 1.17E-205 | 0.34849534 | 0.596 | 0.326 | 4.46E-201 | ExN3 | TESPA1 |
| MAOB | 3.94E-205 | 0.30581221 | 0.525 | 0.283 | 1.51E-200 | ExN3 | MAOB |
| ZNF827 | 4.62E-205 | 0.37315441 | 0.713 | 0.464 | 1.76E-200 | ExN3 | ZNF827 |
| ANKRD18B | 9.25E-205 | 0.26371244 | 0.445 | 0.223 | 3.54E-200 | ExN3 | ANKRD18B |
| AC011287.11 | 1.02E-204 | 0.45532139 | 0.736 | 0.45 | 3.92E-200 | ExN3 | AC011287.1 |
| KCNIP4-IT12 | 6.98E-204 | 0.36854542 | 0.76 | 0.479 | 2.67E-199 | ExN3 | KCNIP4-IT1 |
| AC006504.51 | 7.21E-204 | 0.42518134 | 0.644 | 0.413 | 2.76E-199 | ExN3 | AC006504.5 |
| LRRTM41 | 1.29E-201 | 0.42084327 | 0.995 | 0.914 | 4.93E-197 | ExN3 | LRRTM4 |
| ARHGAP22 | 2.32E-199 | 0.27351262 | 0.531 | 0.288 | 8.88E-195 | ExN3 | ARHGAP22 |
| ZNF521 | 1.16E-197 | 0.27219118 | 0.352 | 0.158 | 4.43E-193 | ExN3 | ZNF521 |
| TSBP1-AS11 | 1.20E-197 | 0.36980827 | 0.509 | 0.274 | 4.58E-193 | ExN3 | TSBP1-AS1 |
| NWD22 | 7.67E-197 | 0.31051112 | 0.739 | 0.444 | 2.93E-192 | ExN3 | NWD2 |
| SMAD31 | 4.39E-196 | 0.331917 | 0.622 | 0.36 | 1.68E-191 | ExN3 | SMAD3 |
| KALRN1 | 1.67E-195 | 0.33129852 | 0.987 | 0.955 | 6.38E-191 | ExN3 | KALRN |
| SYT71 | 7.09E-195 | 0.34914066 | 0.763 | 0.514 | 2.71E-190 | ExN3 | SYT7 |
| LDLRAD3 | 5.37E-194 | 0.28082711 | 0.436 | 0.216 | 2.05E-189 | ExN3 | LDLRAD3 |
| PTPRF1 | 1.36E-193 | 0.29665544 | 0.574 | 0.332 | 5.22E-189 | ExN3 | PTPRF |
| PRKG1 | 3.03E-193 | 0.51489137 | 0.87 | 0.584 | 1.16E-188 | ExN3 | PRKG1 |
| NIN1 | 3.03E-191 | 0.38083668 | 0.89 | 0.696 | 1.16E-186 | ExN3 | NIN |
| SNTG11 | 4.11E-191 | 0.36962405 | 0.983 | 0.938 | 1.57E-186 | ExN3 | SNTG1 |
| RASAL2 | 2.36E-190 | 0.36152108 | 0.952 | 0.871 | 9.03E-186 | ExN3 | RASAL2 |
| GLRA31 | 2.99E-188 | 0.30371454 | 0.519 | 0.286 | 1.14E-183 | ExN3 | GLRA3 |
| LINC02224 | 1.18E-187 | 0.28691599 | 0.318 | 0.14 | 4.49E-183 | ExN3 | LINC02224 |
| PRKCB2 | 1.52E-187 | 0.37053186 | 0.968 | 0.86 | 5.81E-183 | ExN3 | PRKCB |
| LAMB11 | 9.29E-187 | 0.27524202 | 0.496 | 0.266 | 3.55E-182 | ExN3 | LAMB1 |
| ADAMTS191 | 1.42E-186 | 0.35267029 | 0.6 | 0.347 | 5.42E-182 | ExN3 | ADAMTS19 |
| C11orf871 | 7.27E-185 | 0.29473319 | 0.548 | 0.314 | 2.78E-180 | ExN3 | C11orf87 |
| MIR2052HG | 2.09E-184 | 0.44395746 | 0.529 | 0.301 | 7.99E-180 | ExN3 | MIR2052HG |
| TMEM150C | 1.22E-183 | 0.27522569 | 0.511 | 0.286 | 4.65E-179 | ExN3 | TMEM150C |
| PPP4R41 | 1.28E-183 | 0.31281043 | 0.636 | 0.383 | 4.90E-179 | ExN3 | PPP4R4 |
| NOL4 | 2.70E-183 | 0.39733502 | 0.874 | 0.726 | 1.03E-178 | ExN3 | NOL4 |
| CLSTN2 | 5.94E-183 | 0.30254391 | 0.881 | 0.602 | 2.27E-178 | ExN3 | CLSTN2 |
| NDFIP22 | 2.39E-182 | 0.36417023 | 0.737 | 0.494 | 9.12E-178 | ExN3 | NDFIP2 |
| MRPS30-DT1 | 6.35E-182 | 0.34894843 | 0.68 | 0.443 | 2.43E-177 | ExN3 | MRPS30-DT |
| SLC6A72 | 2.05E-181 | 0.25765383 | 0.541 | 0.304 | 7.84E-177 | ExN3 | SLC6A7 |
| HOPX1 | 2.66E-181 | 0.27482794 | 0.518 | 0.282 | 1.02E-176 | ExN3 | HOPX |
| PRUNE2 | 2.88E-181 | 0.33751893 | 0.79 | 0.583 | 1.10E-176 | ExN3 | PRUNE2 |
| FOCAD1 | 9.32E-181 | 0.38509378 | 0.909 | 0.793 | 3.56E-176 | ExN3 | FOCAD |
| JAZF1-AS1 | 1.84E-180 | 0.26449387 | 0.409 | 0.209 | 7.01E-176 | ExN3 | JAZF1-AS1 |
| TMEM163 | 6.24E-178 | 0.25149469 | 0.5 | 0.272 | 2.39E-173 | ExN3 | TMEM163 |

| | | | | | | | |
|---|---|---|---|---|---|---|---|
| LDLRAD4 | 1.20E-177 | 0.37732963 | 0.926 | 0.768 | 4.59E-173 | ExN3 | LDLRAD4 |
| CACNA1A | 6.17E-177 | 0.3260078 | 0.972 | 0.904 | 2.36E-172 | ExN3 | CACNA1A |
| TOM1L1 | 1.32E-176 | 0.2512688 | 0.444 | 0.238 | 5.06E-172 | ExN3 | TOM1L1 |
| AC007389.1 | 2.54E-175 | 0.35243413 | 0.404 | 0.207 | 9.70E-171 | ExN3 | AC007389.1 |
| PTK21 | 3.82E-175 | 0.31122625 | 0.967 | 0.893 | 1.46E-170 | ExN3 | PTK2 |
| ARHGAP321 | 7.46E-174 | 0.3554438 | 0.951 | 0.839 | 2.85E-169 | ExN3 | ARHGAP32 |
| ACTN11 | 4.22E-173 | 0.34903744 | 0.797 | 0.57 | 1.61E-168 | ExN3 | ACTN1 |
| DNAH14 | 5.10E-173 | 0.29098148 | 0.51 | 0.28 | 1.95E-168 | ExN3 | DNAH14 |
| CAMK2A2 | 9.30E-173 | 0.36152688 | 0.938 | 0.784 | 3.56E-168 | ExN3 | CAMK2A |
| CNTN3 | 1.15E-172 | 0.34050066 | 0.886 | 0.644 | 4.40E-168 | ExN3 | CNTN3 |
| KIAA15491 | 5.65E-170 | 0.34190634 | 0.728 | 0.507 | 2.16E-165 | ExN3 | KIAA1549 |
| AL096794.1 | 6.12E-170 | 0.30259105 | 0.464 | 0.255 | 2.34E-165 | ExN3 | AL096794.1 |
| TSPAN51 | 3.56E-169 | 0.36431867 | 0.938 | 0.835 | 1.36E-164 | ExN3 | TSPAN5 |
| LINC02822 | 6.16E-168 | 0.45875943 | 0.3 | 0.136 | 2.36E-163 | ExN3 | LINC02822 |
| STX1A1 | 1.72E-166 | 0.32053208 | 0.581 | 0.359 | 6.56E-162 | ExN3 | STX1A |
| PIK3R1 | 2.04E-166 | 0.35202899 | 0.864 | 0.676 | 7.79E-162 | ExN3 | PIK3R1 |
| OXR1 | 6.16E-166 | 0.30633529 | 0.974 | 0.917 | 2.36E-161 | ExN3 | OXR1 |
| PRKAG2 | 4.76E-165 | 0.3411789 | 0.905 | 0.713 | 1.82E-160 | ExN3 | PRKAG2 |
| TTN | 2.03E-164 | 0.30046782 | 0.299 | 0.139 | 7.75E-160 | ExN3 | TTN |
| PPP2R2C2 | 1.34E-163 | 0.33598759 | 0.926 | 0.79 | 5.12E-159 | ExN3 | PPP2R2C |
| SORL11 | 8.06E-162 | 0.32847424 | 0.856 | 0.663 | 3.08E-157 | ExN3 | SORL1 |
| AC090578.11 | 1.46E-160 | 0.66095376 | 0.77 | 0.544 | 5.58E-156 | ExN3 | AC090578.1 |
| AC009041.21 | 2.37E-160 | 0.34981278 | 0.52 | 0.306 | 9.06E-156 | ExN3 | AC009041.2 |
| CFAP2211 | 2.41E-160 | 0.30631139 | 0.619 | 0.397 | 9.23E-156 | ExN3 | CFAP221 |
| LINGO21 | 6.96E-160 | 0.38910302 | 0.953 | 0.814 | 2.66E-155 | ExN3 | LINGO2 |
| MPP71 | 8.26E-160 | 0.25250967 | 0.624 | 0.384 | 3.16E-155 | ExN3 | MPP7 |
| PPP1R21 | 7.99E-157 | 0.30923891 | 0.676 | 0.464 | 3.05E-152 | ExN3 | PPP1R21 |
| AL117329.12 | 1.16E-156 | 0.566057 | 0.671 | 0.456 | 4.43E-152 | ExN3 | AL117329.1 |
| GRM8 | 2.89E-156 | 0.26996273 | 0.58 | 0.334 | 1.10E-151 | ExN3 | GRM8 |
| NRCAM | 3.59E-156 | 0.29931668 | 0.975 | 0.916 | 1.37E-151 | ExN3 | NRCAM |
| DCLK1 | 4.47E-156 | 0.29530431 | 0.978 | 0.938 | 1.71E-151 | ExN3 | DCLK1 |
| RNF144A1 | 1.93E-155 | 0.35479862 | 0.895 | 0.728 | 7.38E-151 | ExN3 | RNF144A |
| TPD521 | 3.32E-154 | 0.30877532 | 0.738 | 0.52 | 1.27E-149 | ExN3 | TPD52 |
| ZEB11 | 6.91E-154 | 0.36004325 | 0.901 | 0.752 | 2.64E-149 | ExN3 | ZEB1 |
| KHDRBS2-OT | 5.06E-153 | 0.31563478 | 0.664 | 0.435 | 1.94E-148 | ExN3 | KHDRBS2-OT |
| ARL15 | 6.78E-151 | 0.40642383 | 0.905 | 0.8 | 2.59E-146 | ExN3 | ARL15 |
| AQP4-AS12 | 7.94E-151 | 0.4385005 | 0.696 | 0.486 | 3.04E-146 | ExN3 | AQP4-AS1 |
| AL513166.11 | 3.52E-150 | 0.38807679 | 0.821 | 0.606 | 1.34E-145 | ExN3 | AL513166.1 |
| DCAF61 | 1.39E-149 | 0.32472631 | 0.807 | 0.609 | 5.30E-145 | ExN3 | DCAF6 |
| CDYL22 | 1.66E-149 | 0.28496304 | 0.607 | 0.388 | 6.33E-145 | ExN3 | CDYL2 |
| AC068587.4 | 2.46E-149 | 0.26730465 | 0.696 | 0.498 | 9.40E-145 | ExN3 | AC068587.4 |
| AL049828.11 | 2.38E-148 | 0.36940862 | 0.829 | 0.629 | 9.09E-144 | ExN3 | AL049828.1 |
| MAST31 | 6.91E-148 | 0.25550729 | 0.641 | 0.403 | 2.64E-143 | ExN3 | MAST3 |
| ARHGAP261 | 1.27E-147 | 0.41624469 | 0.936 | 0.867 | 4.86E-143 | ExN3 | ARHGAP26 |
| RALYL1 | 5.96E-147 | 0.35400668 | 0.991 | 0.803 | 2.28E-142 | ExN3 | RALYL |
| SLC2A13 | 1.48E-146 | 0.2924457 | 0.933 | 0.849 | 5.67E-142 | ExN3 | SLC2A13 |
| FAM81A | 8.49E-145 | 0.28813133 | 0.689 | 0.472 | 3.25E-140 | ExN3 | FAM81A |
| KIF13A1 | 1.01E-144 | 0.26413322 | 0.786 | 0.552 | 3.88E-140 | ExN3 | KIF13A |
| PARM1 | 2.24E-143 | 0.25718007 | 0.612 | 0.379 | 8.55E-139 | ExN3 | PARM1 |
| ADAMTS161 | 5.89E-141 | 0.41650853 | 0.35 | 0.19 | 2.25E-136 | ExN3 | ADAMTS16 |
| AKT3 | 7.43E-141 | 0.29656796 | 0.97 | 0.923 | 2.84E-136 | ExN3 | AKT3 |
| DYNC1I1 | 4.42E-139 | 0.28987166 | 0.943 | 0.866 | 1.69E-134 | ExN3 | DYNC1I1 |
| MLLT31 | 1.23E-138 | 0.34009428 | 0.921 | 0.811 | 4.70E-134 | ExN3 | MLLT3 |
| AC109466.11 | 1.92E-138 | 0.60271787 | 0.468 | 0.276 | 7.36E-134 | ExN3 | AC109466.1 |
| CHN11 | 3.60E-137 | 0.29160499 | 0.94 | 0.735 | 1.38E-132 | ExN3 | CHN1 |
| CEP126 | 2.11E-136 | 0.31745634 | 0.727 | 0.54 | 8.08E-132 | ExN3 | CEP126 |
| ARPP211 | 6.77E-136 | 0.30453149 | 0.966 | 0.75 | 2.59E-131 | ExN3 | ARPP21 |
| HOMER11 | 1.40E-134 | 0.31674204 | 0.791 | 0.607 | 5.35E-130 | ExN3 | HOMER1 |
| SYNDIG1 | 5.19E-134 | 0.27363756 | 0.801 | 0.58 | 1.98E-129 | ExN3 | SYNDIG1 |
| RAP1GAP21 | 1.39E-133 | 0.28401906 | 0.842 | 0.62 | 5.31E-129 | ExN3 | RAP1GAP2 |
| SYTL22 | 5.12E-133 | 0.25714321 | 0.699 | 0.465 | 1.96E-128 | ExN3 | SYTL2 |
| STMN2 | 4.96E-132 | 0.30111487 | 0.675 | 0.47 | 1.90E-127 | ExN3 | STMN2 |
| CAMK1D2 | 6.96E-132 | 0.29370371 | 0.97 | 0.851 | 2.66E-127 | ExN3 | CAMK1D |

| | | | | | | | |
|---|---|---|---|---|---|---|---|
| RHBDL31 | 7.60E-132 | 0.26128978 | 0.599 | 0.38 | 2.90E-127 | ExN3 | RHBDL3 |
| CUX11 | 2.00E-131 | 0.27525321 | 0.881 | 0.716 | 7.64E-127 | ExN3 | CUX1 |
| KIAA09301 | 3.42E-131 | 0.29986013 | 0.788 | 0.598 | 1.31E-126 | ExN3 | KIAA0930 |
| NLK1 | 5.51E-131 | 0.34047916 | 0.86 | 0.685 | 2.11E-126 | ExN3 | NLK |
| GRM51 | 5.67E-131 | 0.29743634 | 0.981 | 0.892 | 2.17E-126 | ExN3 | GRM5 |
| ALS21 | 2.86E-130 | 0.26950841 | 0.704 | 0.506 | 1.09E-125 | ExN3 | ALS2 |
| MAST4 | 3.12E-130 | 0.33388359 | 0.902 | 0.811 | 1.19E-125 | ExN3 | MAST4 |
| RGS61 | 1.65E-129 | 0.33655591 | 0.894 | 0.687 | 6.29E-125 | ExN3 | RGS6 |
| TMEM1081 | 8.00E-129 | 0.35202784 | 0.919 | 0.811 | 3.06E-124 | ExN3 | TMEM108 |
| MYH10 | 3.08E-128 | 0.29694902 | 0.871 | 0.714 | 1.18E-123 | ExN3 | MYH10 |
| FBXO341 | 9.15E-128 | 0.32782052 | 0.797 | 0.607 | 3.50E-123 | ExN3 | FBXO34 |
| AACS | 3.03E-125 | 0.25847965 | 0.653 | 0.451 | 1.16E-120 | ExN3 | AACS |
| FSTL42 | 1.28E-124 | 0.3063402 | 0.887 | 0.629 | 4.91E-120 | ExN3 | FSTL4 |
| AC073091.31 | 3.52E-123 | 0.31858707 | 0.502 | 0.301 | 1.35E-118 | ExN3 | AC073091.3 |
| TMEM2411 | 6.54E-123 | 0.28583428 | 0.687 | 0.484 | 2.50E-118 | ExN3 | TMEM241 |
| MSRA2 | 7.27E-123 | 0.27814163 | 0.966 | 0.903 | 2.78E-118 | ExN3 | MSRA |
| DUBR1 | 1.56E-122 | 0.26654404 | 0.758 | 0.558 | 5.95E-118 | ExN3 | DUBR |
| HCN1 | 2.08E-122 | 0.29637976 | 0.933 | 0.853 | 7.96E-118 | ExN3 | HCN1 |
| FBXW71 | 4.64E-122 | 0.28183256 | 0.897 | 0.745 | 1.78E-117 | ExN3 | FBXW7 |
| SPATS2 | 7.38E-122 | 0.26971871 | 0.76 | 0.566 | 2.82E-117 | ExN3 | SPATS2 |
| ZRANB3 | 1.28E-121 | 0.28683138 | 0.676 | 0.482 | 4.89E-117 | ExN3 | ZRANB3 |
| MEG31 | 6.97E-121 | 0.48253972 | 0.977 | 0.959 | 2.66E-116 | ExN3 | MEG3 |
| LINGO11 | 1.66E-120 | 0.25456558 | 0.709 | 0.487 | 6.35E-116 | ExN3 | LINGO1 |
| AC008415.11 | 5.83E-117 | 0.48689264 | 0.348 | 0.191 | 2.23E-112 | ExN3 | AC008415.1 |
| PDK3 | 6.68E-117 | 0.26832259 | 0.649 | 0.459 | 2.56E-112 | ExN3 | PDK3 |
| PDE4D1 | 9.53E-116 | 0.29812057 | 0.986 | 0.959 | 3.64E-111 | ExN3 | PDE4D |
| PPP1R12A1 | 1.28E-115 | 0.27756313 | 0.859 | 0.72 | 4.90E-111 | ExN3 | PPP1R12A |
| SNCA | 4.32E-115 | 0.27633119 | 0.817 | 0.594 | 1.65E-110 | ExN3 | SNCA |
| SEMA3A1 | 1.45E-113 | 0.32915377 | 0.358 | 0.2 | 5.55E-109 | ExN3 | SEMA3A |
| AL591501.11 | 2.01E-113 | 0.27385502 | 0.778 | 0.554 | 7.69E-109 | ExN3 | AL591501.1 |
| CDKL5 | 2.81E-112 | 0.29572794 | 0.865 | 0.72 | 1.07E-107 | ExN3 | CDKL5 |
| KCTD161 | 4.91E-112 | 0.27429319 | 0.962 | 0.859 | 1.88E-107 | ExN3 | KCTD16 |
| PPFIA21 | 9.49E-112 | 0.2655889 | 0.973 | 0.92 | 3.63E-107 | ExN3 | PPFIA2 |
| ABHD2 | 7.84E-110 | 0.25376591 | 0.809 | 0.638 | 3.00E-105 | ExN3 | ABHD2 |
| CDKL11 | 2.10E-109 | 0.26803738 | 0.643 | 0.45 | 8.04E-105 | ExN3 | CDKL1 |
| AC007563.21 | 1.40E-106 | 0.26960473 | 0.806 | 0.593 | 5.36E-102 | ExN3 | AC007563.2 |
| PUM31 | 3.33E-106 | 0.270766 | 0.768 | 0.593 | 1.27E-101 | ExN3 | PUM3 |
| TTC39C | 2.34E-105 | 0.26651499 | 0.768 | 0.587 | 8.93E-101 | ExN3 | TTC39C |
| MAN1C11 | 1.58E-104 | 0.25027574 | 0.783 | 0.603 | 6.03E-100 | ExN3 | MAN1C1 |
| KIFC2 | 1.83E-104 | 0.25648983 | 0.677 | 0.496 | 6.98E-100 | ExN3 | KIFC2 |
| AK51 | 8.57E-103 | 0.25482604 | 0.911 | 0.77 | 3.28E-98 | ExN3 | AK5 |
| SLC39A101 | 6.62E-101 | 0.2836132 | 0.8 | 0.632 | 2.53E-96 | ExN3 | SLC39A10 |
| MBNL22 | 3.91E-100 | 0.27112185 | 0.925 | 0.752 | 1.49E-95 | ExN3 | MBNL2 |
| AL008633.11 | 1.16E-98 | 0.32727573 | 0.866 | 0.703 | 4.44E-94 | ExN3 | AL008633.1 |
| AC024901.1 | 2.33E-98 | 0.33637081 | 0.604 | 0.442 | 8.91E-94 | ExN3 | AC024901.1 |
| LRRC281 | 3.29E-98 | 0.27737149 | 0.789 | 0.62 | 1.26E-93 | ExN3 | LRRC28 |
| EVI5 | 1.03E-97 | 0.28397646 | 0.786 | 0.651 | 3.95E-93 | ExN3 | EVI5 |
| TJP11 | 4.35E-95 | 0.250081 | 0.928 | 0.825 | 1.66E-90 | ExN3 | TJP1 |
| AC084816.11 | 3.68E-93 | 0.29651462 | 0.325 | 0.187 | 1.41E-88 | ExN3 | AC084816.1 |
| AC016831.7 | 3.74E-93 | 0.27461308 | 0.825 | 0.673 | 1.43E-88 | ExN3 | AC016831.7 |
| PI4KA1 | 3.30E-91 | 0.2528588 | 0.917 | 0.808 | 1.26E-86 | ExN3 | PI4KA |
| CAMK41 | 3.98E-88 | 0.27727347 | 0.861 | 0.709 | 1.52E-83 | ExN3 | CAMK4 |
| DMD1 | 2.33E-86 | 0.26046954 | 0.97 | 0.917 | 8.92E-82 | ExN3 | DMD |
| KCTD11 | 6.24E-85 | 0.28834833 | 0.803 | 0.663 | 2.38E-80 | ExN3 | KCTD1 |
| PDZRN41 | 9.88E-85 | 0.45974904 | 0.554 | 0.386 | 3.78E-80 | ExN3 | PDZRN4 |
| SLC35F31 | 1.49E-80 | 0.29141738 | 0.89 | 0.759 | 5.70E-76 | ExN3 | SLC35F3 |
| ZNF804B | 5.47E-78 | 0.27652488 | 0.512 | 0.346 | 2.09E-73 | ExN3 | ZNF804B |
| PCSK21 | 7.35E-77 | 0.27281668 | 0.912 | 0.793 | 2.81E-72 | ExN3 | PCSK2 |
| GRIA31 | 3.87E-73 | 0.26226554 | 0.885 | 0.804 | 1.48E-68 | ExN3 | GRIA3 |
| ASXL31 | 3.44E-71 | 0.25626487 | 0.769 | 0.612 | 1.32E-66 | ExN3 | ASXL3 |
| DGKB1 | 2.75E-52 | 0.25968104 | 0.794 | 0.811 | 1.05E-47 | ExN3 | DGKB |
| SYN31 | 5.15E-25 | 0.2507052 | 0.885 | 0.865 | 1.97E-20 | ExN3 | SYN3 |
| AC109466.12 | 0 | 2.50130631 | 0.859 | 0.237 | 0 | ExN4 | AC109466.1 |

| | | | | | | | |
|---|---|---|---|---|---|---|---|
| AC008415.12 | 0 | 2.12060151 | 0.769 | 0.149 | 0 | ExN4 | AC008415.1 |
| AC060765.21 | 0 | 2.11312693 | 0.738 | 0.143 | 0 | ExN4 | AC060765.2 |
| CASC151 | 0 | 2.06169783 | 0.94 | 0.484 | 0 | ExN4 | CASC15 |
| AC073091.32 | 0 | 1.92794411 | 0.839 | 0.267 | 0 | ExN4 | AC073091.3 |
| AC016687.21 | 0 | 1.71582503 | 0.558 | 0.119 | 0 | ExN4 | AC016687.2 |
| LRRK12 | 0 | 1.69632209 | 0.88 | 0.28 | 0 | ExN4 | LRRK1 |
| RORB2 | 0 | 1.64309999 | 0.958 | 0.274 | 0 | ExN4 | RORB |
| CADPS21 | 0 | 1.6293176 | 0.978 | 0.553 | 0 | ExN4 | CADPS2 |
| NRG1 | 0 | 1.59140114 | 0.95 | 0.511 | 0 | ExN4 | NRG1 |
| CLSTN21 | 0 | 1.58660915 | 0.975 | 0.592 | 0 | ExN4 | CLSTN2 |
| AP003066.11 | 0 | 1.50832498 | 0.571 | 0.06 | 0 | ExN4 | AP003066.1 |
| CPNE41 | 0 | 1.46966563 | 0.908 | 0.43 | 0 | ExN4 | CPNE4 |
| TLL1 | 0 | 1.44560839 | 0.734 | 0.076 | 0 | ExN4 | TLL1 |
| HS3ST21 | 0 | 1.30095468 | 0.806 | 0.215 | 0 | ExN4 | HS3ST2 |
| DPP10 | 0 | 1.25509111 | 0.995 | 0.835 | 0 | ExN4 | DPP10 |
| NKAIN22 | 0 | 1.2513576 | 0.993 | 0.826 | 0 | ExN4 | NKAIN2 |
| PRKG11 | 0 | 1.23598745 | 0.921 | 0.579 | 0 | ExN4 | PRKG1 |
| KCNH71 | 0 | 1.22986937 | 0.967 | 0.709 | 0 | ExN4 | KCNH7 |
| LHFPL62 | 0 | 1.21210171 | 0.873 | 0.382 | 0 | ExN4 | LHFPL6 |
| PDE1C | 0 | 1.17992891 | 0.917 | 0.457 | 0 | ExN4 | PDE1C |
| IL1RAPL21 | 0 | 1.17262727 | 0.771 | 0.283 | 0 | ExN4 | IL1RAPL2 |
| DACH1 | 0 | 1.15910428 | 0.671 | 0.162 | 0 | ExN4 | DACH1 |
| FOXP21 | 0 | 1.12015655 | 0.894 | 0.261 | 0 | ExN4 | FOXP2 |
| RXFP1 | 0 | 1.09912955 | 0.703 | 0.184 | 0 | ExN4 | RXFP1 |
| CDH20 | 0 | 1.09117053 | 0.797 | 0.345 | 0 | ExN4 | CDH20 |
| BRINP31 | 0 | 1.08909512 | 0.89 | 0.459 | 0 | ExN4 | BRINP3 |
| LINC018212 | 0 | 1.08175801 | 0.637 | 0.176 | 0 | ExN4 | LINC01821 |
| PTPRT2 | 0 | 1.07984985 | 0.977 | 0.658 | 0 | ExN4 | PTPRT |
| GRM32 | 0 | 1.06830654 | 0.858 | 0.357 | 0 | ExN4 | GRM3 |
| CAMK2D | 0 | 1.06281855 | 0.918 | 0.647 | 0 | ExN4 | CAMK2D |
| LRP1B | 0 | 1.0396715 | 0.995 | 0.946 | 0 | ExN4 | LRP1B |
| FSTL43 | 0 | 1.03499808 | 0.954 | 0.623 | 0 | ExN4 | FSTL4 |
| LINC018111 | 0 | 1.0282382 | 0.701 | 0.273 | 0 | ExN4 | LINC01811 |
| PDE1A2 | 0 | 1.02330993 | 0.975 | 0.656 | 0 | ExN4 | PDE1A |
| FRMPD4 | 0 | 1.02229228 | 0.986 | 0.934 | 0 | ExN4 | FRMPD4 |
| AC099517.1 | 0 | 0.98361511 | 0.318 | 0.041 | 0 | ExN4 | AC099517.1 |
| PRUNE21 | 0 | 0.97610179 | 0.831 | 0.579 | 0 | ExN4 | PRUNE2 |
| LDLRAD31 | 0 | 0.95426556 | 0.663 | 0.194 | 0 | ExN4 | LDLRAD3 |
| DNAH6 | 0 | 0.95426492 | 0.848 | 0.505 | 0 | ExN4 | DNAH6 |
| TAFA12 | 0 | 0.94409047 | 0.828 | 0.491 | 0 | ExN4 | TAFA1 |
| PHACTR11 | 0 | 0.94389512 | 0.994 | 0.949 | 0 | ExN4 | PHACTR1 |
| GRM81 | 0 | 0.94048537 | 0.765 | 0.316 | 0 | ExN4 | GRM8 |
| CCDC85A | 0 | 0.90634993 | 0.918 | 0.633 | 0 | ExN4 | CCDC85A |
| DPY19L11 | 0 | 0.90486747 | 0.768 | 0.316 | 0 | ExN4 | DPY19L1 |
| EPHA31 | 0 | 0.8988112 | 0.806 | 0.387 | 0 | ExN4 | EPHA3 |
| PCDH71 | 0 | 0.89398955 | 0.983 | 0.813 | 0 | ExN4 | PCDH7 |
| IPCEF11 | 0 | 0.89193215 | 0.781 | 0.368 | 0 | ExN4 | IPCEF1 |
| ST6GALNAC3 | 0 | 0.89089248 | 0.804 | 0.479 | 0 | ExN4 | ST6GALNAC3 |
| TENM4 | 0 | 0.88822064 | 0.967 | 0.849 | 0 | ExN4 | TENM4 |
| CTNNA21 | 0 | 0.8751756 | 0.993 | 0.955 | 0 | ExN4 | CTNNA2 |
| RAB27B | 0 | 0.87431239 | 0.783 | 0.427 | 0 | ExN4 | RAB27B |
| CNTN31 | 0 | 0.87334162 | 0.935 | 0.639 | 0 | ExN4 | CNTN3 |
| HS3ST4 | 0 | 0.86623035 | 0.876 | 0.386 | 0 | ExN4 | HS3ST4 |
| CDH10 | 0 | 0.8657117 | 0.92 | 0.679 | 0 | ExN4 | CDH10 |
| ARPP212 | 0 | 0.84822213 | 0.98 | 0.749 | 0 | ExN4 | ARPP21 |
| PTPRD2 | 0 | 0.84784928 | 0.997 | 0.961 | 0 | ExN4 | PTPRD |
| STXBP5-AS11 | 0 | 0.8442337 | 0.879 | 0.562 | 0 | ExN4 | STXBP5-AS1 |
| SORCS11 | 0 | 0.84404612 | 0.923 | 0.655 | 0 | ExN4 | SORCS1 |
| GHR | 0 | 0.84201392 | 0.749 | 0.35 | 0 | ExN4 | GHR |
| POU6F22 | 0 | 0.81786442 | 0.89 | 0.359 | 0 | ExN4 | POU6F2 |
| FMN11 | 0 | 0.81145513 | 0.948 | 0.743 | 0 | ExN4 | FMN1 |
| RALYL2 | 0 | 0.81099383 | 0.995 | 0.803 | 0 | ExN4 | RALYL |
| ARSJ | 0 | 0.80336138 | 0.491 | 0.054 | 0 | ExN4 | ARSJ |

| | | | | | | | |
|---|---|---|---|---|---|---|---|
| FAT32 | 0 | 0.80313642 | 0.95 | 0.693 | 0 | ExN4 | FAT3 |
| RHBDL32 | 0 | 0.80264612 | 0.76 | 0.364 | 0 | ExN4 | RHBDL3 |
| AC013287.12 | 0 | 0.79508074 | 0.795 | 0.419 | 0 | ExN4 | AC013287.1 |
| MDGA2 | 0 | 0.79468107 | 0.988 | 0.934 | 0 | ExN4 | MDGA2 |
| HECW12 | 0 | 0.79061771 | 0.97 | 0.821 | 0 | ExN4 | HECW1 |
| COL11A1 | 0 | 0.78689098 | 0.676 | 0.221 | 0 | ExN4 | COL11A1 |
| RYR22 | 0 | 0.77028082 | 0.992 | 0.923 | 0 | ExN4 | RYR2 |
| RORA1 | 0 | 0.76138218 | 0.988 | 0.961 | 0 | ExN4 | RORA |
| MIR3681HG | 0 | 0.75650325 | 0.439 | 0.058 | 0 | ExN4 | MIR3681HG |
| MIR99AHG1 | 0 | 0.75252608 | 0.969 | 0.888 | 0 | ExN4 | MIR99AHG |
| TMEM117 | 0 | 0.74772747 | 0.843 | 0.629 | 0 | ExN4 | TMEM117 |
| TOX1 | 0 | 0.73836882 | 0.864 | 0.405 | 0 | ExN4 | TOX |
| OPCML1 | 0 | 0.73206471 | 0.996 | 0.987 | 0 | ExN4 | OPCML |
| SLC22A102 | 0 | 0.73178887 | 0.824 | 0.433 | 0 | ExN4 | SLC22A10 |
| KHDRBS32 | 0 | 0.72343088 | 0.964 | 0.782 | 0 | ExN4 | KHDRBS3 |
| ELAVL4 | 0 | 0.72325131 | 0.838 | 0.59 | 0 | ExN4 | ELAVL4 |
| PDGFC1 | 0 | 0.72088842 | 0.637 | 0.249 | 0 | ExN4 | PDGFC |
| AC008127.1 | 0 | 0.71878327 | 0.43 | 0.047 | 0 | ExN4 | AC008127.1 |
| KCNB21 | 0 | 0.71664296 | 0.88 | 0.67 | 0 | ExN4 | KCNB2 |
| STMN21 | 0 | 0.69949824 | 0.753 | 0.462 | 0 | ExN4 | STMN2 |
| BMPER1 | 0 | 0.69121475 | 0.873 | 0.594 | 0 | ExN4 | BMPER |
| MKX | 0 | 0.68634091 | 0.511 | 0.125 | 0 | ExN4 | MKX |
| AL138927.1 | 0 | 0.68556312 | 0.29 | 0.015 | 0 | ExN4 | AL138927.1 |
| AGBL42 | 0 | 0.68146063 | 0.984 | 0.929 | 0 | ExN4 | AGBL4 |
| LINC022321 | 0 | 0.66836264 | 0.484 | 0.165 | 0 | ExN4 | LINC02232 |
| KCNQ51 | 0 | 0.6671137 | 0.99 | 0.897 | 0 | ExN4 | KCNQ5 |
| SLC24A21 | 0 | 0.66641303 | 0.977 | 0.821 | 0 | ExN4 | SLC24A2 |
| PRKAG21 | 0 | 0.66497341 | 0.927 | 0.711 | 0 | ExN4 | PRKAG2 |
| AC009899.1 | 0 | 0.66486476 | 0.753 | 0.474 | 0 | ExN4 | AC009899.1 |
| SYNDIG11 | 0 | 0.66374716 | 0.845 | 0.576 | 0 | ExN4 | SYNDIG1 |
| KLHL321 | 0 | 0.65934179 | 0.888 | 0.669 | 0 | ExN4 | KLHL32 |
| NAV31 | 0 | 0.65880995 | 0.993 | 0.946 | 0 | ExN4 | NAV3 |
| ZNF385B1 | 0 | 0.65355097 | 0.966 | 0.808 | 0 | ExN4 | ZNF385B |
| AC021134.1 | 0 | 0.64611844 | 0.273 | 0.025 | 0 | ExN4 | AC021134.1 |
| GALNT17 | 0 | 0.644239 | 0.978 | 0.894 | 0 | ExN4 | GALNT17 |
| OCA2 | 0 | 0.63714235 | 0.601 | 0.246 | 0 | ExN4 | OCA2 |
| AP000943.2 | 0 | 0.6367862 | 0.535 | 0.182 | 0 | ExN4 | AP000943.2 |
| CADM2 | 0 | 0.63274956 | 0.997 | 0.992 | 0 | ExN4 | CADM2 |
| NEGR11 | 0 | 0.63026532 | 0.995 | 0.952 | 0 | ExN4 | NEGR1 |
| DNAJC11 | 0 | 0.62814349 | 0.803 | 0.539 | 0 | ExN4 | DNAJC1 |
| CRIM1 | 0 | 0.62569214 | 0.774 | 0.463 | 0 | ExN4 | CRIM1 |
| PCLO1 | 0 | 0.6239856 | 0.978 | 0.918 | 0 | ExN4 | PCLO |
| CDH7 | 0 | 0.62280089 | 0.635 | 0.242 | 0 | ExN4 | CDH7 |
| EPHA51 | 0 | 0.61737368 | 0.841 | 0.578 | 0 | ExN4 | EPHA5 |
| NMNAT3 | 0 | 0.6139717 | 0.579 | 0.221 | 0 | ExN4 | NMNAT3 |
| AL033539.2 | 0 | 0.6125907 | 0.423 | 0.043 | 0 | ExN4 | AL033539.2 |
| KCNK10 | 0 | 0.61118955 | 0.642 | 0.3 | 0 | ExN4 | KCNK10 |
| SH3GL23 | 0 | 0.60944656 | 0.943 | 0.729 | 0 | ExN4 | SH3GL2 |
| MRTFB3 | 0 | 0.60793966 | 0.921 | 0.747 | 0 | ExN4 | MRTFB |
| PFKP | 0 | 0.60468461 | 0.681 | 0.381 | 0 | ExN4 | PFKP |
| RMST1 | 0 | 0.6007016 | 0.425 | 0.148 | 0 | ExN4 | RMST |
| PDE3B | 0 | 0.60054348 | 0.758 | 0.497 | 0 | ExN4 | PDE3B |
| SNCA1 | 0 | 0.59793801 | 0.848 | 0.591 | 0 | ExN4 | SNCA |
| LINC022171 | 0 | 0.59789038 | 0.557 | 0.199 | 0 | ExN4 | LINC02217 |
| CEMIP1 | 0 | 0.58548236 | 0.682 | 0.385 | 0 | ExN4 | CEMIP |
| PDE4B | 0 | 0.58321399 | 0.899 | 0.586 | 0 | ExN4 | PDE4B |
| TRPM3 | 0 | 0.57645158 | 0.762 | 0.378 | 0 | ExN4 | TRPM3 |
| ST8SIA11 | 0 | 0.57618543 | 0.799 | 0.538 | 0 | ExN4 | ST8SIA1 |
| AL391117.1 | 0 | 0.56976023 | 0.307 | 0.046 | 0 | ExN4 | AL391117.1 |
| TRABD2A | 0 | 0.5688858 | 0.396 | 0.033 | 0 | ExN4 | TRABD2A |
| CELF4 | 0 | 0.56314596 | 0.968 | 0.902 | 0 | ExN4 | CELF4 |
| PTPRG-AS1 | 0 | 0.5626439 | 0.433 | 0.133 | 0 | ExN4 | PTPRG-AS1 |
| NCALD | 0 | 0.56253962 | 0.922 | 0.757 | 0 | ExN4 | NCALD |

| | | | | | | | |
|---|---|---|---|---|---|---|---|
| TMEFF2 | 0 | 0.55966121 | 0.918 | 0.75 | 0 | ExN4 | TMEFF2 |
| KIAA03191 | 0 | 0.55931498 | 0.798 | 0.54 | 0 | ExN4 | KIAA0319 |
| PPP3CA2 | 0 | 0.55494335 | 0.983 | 0.91 | 0 | ExN4 | PPP3CA |
| LINC01902 | 0 | 0.55402227 | 0.476 | 0.133 | 0 | ExN4 | LINC01902 |
| RSPO3 | 0 | 0.55186095 | 0.376 | 0.049 | 0 | ExN4 | RSPO3 |
| NYAP2 | 0 | 0.55073505 | 0.805 | 0.482 | 0 | ExN4 | NYAP2 |
| CDK14 | 0 | 0.55060196 | 0.951 | 0.856 | 0 | ExN4 | CDK14 |
| KCNQ31 | 0 | 0.54746044 | 0.976 | 0.895 | 0 | ExN4 | KCNQ3 |
| SMYD2 | 0 | 0.54447275 | 0.556 | 0.247 | 0 | ExN4 | SMYD2 |
| NOS1AP | 0 | 0.54420761 | 0.87 | 0.645 | 0 | ExN4 | NOS1AP |
| NCK2 | 0 | 0.53867727 | 0.706 | 0.421 | 0 | ExN4 | NCK2 |
| ALDH1A2 | 0 | 0.53789697 | 0.478 | 0.174 | 0 | ExN4 | ALDH1A2 |
| LRMP1 | 0 | 0.53772209 | 0.493 | 0.12 | 0 | ExN4 | LRMP |
| SYT1 | 0 | 0.53724829 | 0.997 | 0.984 | 0 | ExN4 | SYT1 |
| OPRM11 | 0 | 0.52273896 | 0.499 | 0.208 | 0 | ExN4 | OPRM1 |
| PDE4D2 | 0 | 0.51965008 | 0.992 | 0.959 | 0 | ExN4 | PDE4D |
| SORCS2 | 0 | 0.51421062 | 0.608 | 0.298 | 0 | ExN4 | SORCS2 |
| CALD1 | 0 | 0.51188585 | 0.599 | 0.296 | 0 | ExN4 | CALD1 |
| SATB1-AS11 | 0 | 0.51154687 | 0.757 | 0.409 | 0 | ExN4 | SATB1-AS1 |
| PID1 | 0 | 0.49559698 | 0.749 | 0.476 | 0 | ExN4 | PID1 |
| HTR2A3 | 0 | 0.48371928 | 0.608 | 0.311 | 0 | ExN4 | HTR2A |
| LINC022631 | 0 | 0.48284273 | 0.509 | 0.18 | 0 | ExN4 | LINC02263 |
| ATP8B11 | 0 | 0.4790211 | 0.551 | 0.237 | 0 | ExN4 | ATP8B1 |
| FER1L6-AS2 | 0 | 0.46474797 | 0.293 | 0.063 | 0 | ExN4 | FER1L6-AS2 |
| PLD1 | 0 | 0.46071545 | 0.405 | 0.104 | 0 | ExN4 | PLD1 |
| RNF182 | 0 | 0.44461194 | 0.325 | 0.022 | 0 | ExN4 | RNF182 |
| WHRN | 0 | 0.44015114 | 0.329 | 0.059 | 0 | ExN4 | WHRN |
| LNX2 | 0 | 0.43804361 | 0.402 | 0.13 | 0 | ExN4 | LNX2 |
| SYT9 | 0 | 0.43493128 | 0.404 | 0.122 | 0 | ExN4 | SYT9 |
| SEMA3E | 0 | 0.42769303 | 0.329 | 0.11 | 0 | ExN4 | SEMA3E |
| KCNK2 | 0 | 0.42228835 | 0.461 | 0.145 | 0 | ExN4 | KCNK2 |
| ARHGAP29 | 0 | 0.41689489 | 0.454 | 0.17 | 0 | ExN4 | ARHGAP29 |
| AL589693.1 | 0 | 0.41001578 | 0.337 | 0.116 | 0 | ExN4 | AL589693.1 |
| MDFIC | 0 | 0.40499355 | 0.333 | 0.058 | 0 | ExN4 | MDFIC |
| AL138701.2 | 0 | 0.40058861 | 0.295 | 0.092 | 0 | ExN4 | AL138701.2 |
| RBM20 | 0 | 0.39467138 | 0.28 | 0.049 | 0 | ExN4 | RBM20 |
| PLOD2 | 0 | 0.39171367 | 0.463 | 0.198 | 0 | ExN4 | PLOD2 |
| ANK2 | 0 | 0.38807283 | 0.991 | 0.976 | 0 | ExN4 | ANK2 |
| CMTM8 | 0 | 0.38201144 | 0.341 | 0.087 | 0 | ExN4 | CMTM8 |
| THEMIS | 0 | 0.36756279 | 0.315 | 0.068 | 0 | ExN4 | THEMIS |
| TDO21 | 0 | 0.34927228 | 0.354 | 0.116 | 0 | ExN4 | TDO2 |
| TMTC4 | 0 | 0.34479637 | 0.384 | 0.149 | 0 | ExN4 | TMTC4 |
| DAPK2 | 0 | 0.31767963 | 0.299 | 0.075 | 0 | ExN4 | DAPK2 |
| Z68323.1 | 0 | 0.30512141 | 0.254 | 0.033 | 0 | ExN4 | Z68323.1 |
| AC017050.1 | 0 | 0.28859899 | 0.268 | 0.069 | 0 | ExN4 | AC017050.1 |
| AC015712.2 | 0 | 0.26328388 | 0.262 | 0.061 | 0 | ExN4 | AC015712.2 |
| FRMD3 | 4.24E-304 | 0.40351742 | 0.619 | 0.319 | 1.62E-299 | ExN4 | FRMD3 |
| GPC51 | 6.50E-302 | 0.59005014 | 0.688 | 0.399 | 2.48E-297 | ExN4 | GPC5 |
| FAM13A2 | 1.03E-300 | 0.54019846 | 0.836 | 0.582 | 3.92E-296 | ExN4 | FAM13A |
| FOXP12 | 4.92E-300 | 0.48731632 | 0.955 | 0.808 | 1.88E-295 | ExN4 | FOXP1 |
| FNBP1L | 9.97E-300 | 0.42773994 | 0.675 | 0.364 | 3.81E-295 | ExN4 | FNBP1L |
| RIMS1 | 1.58E-298 | 0.47130599 | 0.95 | 0.888 | 6.03E-294 | ExN4 | RIMS1 |
| AL512662.2 | 5.17E-298 | 0.31951232 | 0.387 | 0.154 | 1.98E-293 | ExN4 | AL512662.2 |
| PTPRK2 | 1.49E-297 | 0.51556011 | 0.886 | 0.583 | 5.69E-293 | ExN4 | PTPRK |
| FAM189A1 | 2.50E-297 | 0.50205384 | 0.856 | 0.614 | 9.55E-293 | ExN4 | FAM189A1 |
| ANKRD33B1 | 2.61E-295 | 0.47155682 | 0.707 | 0.413 | 9.99E-291 | ExN4 | ANKRD33B |
| CNIH3 | 4.27E-295 | 0.54029715 | 0.663 | 0.394 | 1.63E-290 | ExN4 | CNIH3 |
| BAALC | 3.14E-294 | 0.5580662 | 0.712 | 0.456 | 1.20E-289 | ExN4 | BAALC |
| TAFA21 | 8.42E-293 | 0.59796997 | 0.948 | 0.777 | 3.22E-288 | ExN4 | TAFA2 |
| ADD2 | 6.05E-292 | 0.51970529 | 0.847 | 0.663 | 2.31E-287 | ExN4 | ADD2 |
| VPS13A | 4.44E-291 | 0.49383861 | 0.877 | 0.722 | 1.70E-286 | ExN4 | VPS13A |
| NTN4 | 2.73E-288 | 0.35351947 | 0.473 | 0.208 | 1.04E-283 | ExN4 | NTN4 |
| GCNT1 | 1.82E-284 | 0.28793805 | 0.309 | 0.109 | 6.97E-280 | ExN4 | GCNT1 |

| | | | | | | | |
|---|---|---|---|---|---|---|---|
| CDH81 | 2.38E-284 | 0.51368297 | 0.886 | 0.641 | 9.10E-280 | ExN4 | CDH8 |
| PART11 | 1.14E-283 | 0.42551539 | 0.542 | 0.274 | 4.37E-279 | ExN4 | PART1 |
| EPB411 | 2.49E-282 | 0.50091974 | 0.715 | 0.433 | 9.51E-278 | ExN4 | EPB41 |
| FMNL2 | 8.39E-282 | 0.46825501 | 0.96 | 0.866 | 3.21E-277 | ExN4 | FMNL2 |
| PARVA | 1.86E-280 | 0.37885586 | 0.477 | 0.225 | 7.12E-276 | ExN4 | PARVA |
| COL12A1 | 1.05E-277 | 0.37706057 | 0.423 | 0.179 | 4.01E-273 | ExN4 | COL12A1 |
| DIRAS2 | 1.44E-277 | 0.429951 | 0.563 | 0.296 | 5.51E-273 | ExN4 | DIRAS2 |
| CPNE81 | 4.88E-277 | 0.53797865 | 0.731 | 0.456 | 1.86E-272 | ExN4 | CPNE8 |
| CHGA1 | 9.46E-277 | 0.52346672 | 0.601 | 0.35 | 3.62E-272 | ExN4 | CHGA |
| BCL11A | 6.02E-276 | 0.457731 | 0.833 | 0.579 | 2.30E-271 | ExN4 | BCL11A |
| ADAMTS2 | 2.48E-274 | 0.48209298 | 0.333 | 0.13 | 9.48E-270 | ExN4 | ADAMTS2 |
| SSTR2 | 2.19E-273 | 0.3420136 | 0.397 | 0.168 | 8.39E-269 | ExN4 | SSTR2 |
| CCDC144NL-/ | 4.48E-271 | 0.33030898 | 0.342 | 0.134 | 1.71E-266 | ExN4 | CCDC144NL-/ |
| GALNT7 | 6.60E-271 | 0.49951363 | 0.736 | 0.494 | 2.52E-266 | ExN4 | GALNT7 |
| SORCS3 | 4.03E-270 | 0.54592487 | 0.81 | 0.548 | 1.54E-265 | ExN4 | SORCS3 |
| IQCJ-SCHIP13 | 2.38E-269 | 0.49746349 | 0.99 | 0.821 | 9.10E-265 | ExN4 | IQCJ-SCHIP1 |
| CRYM1 | 3.08E-269 | 0.36142371 | 0.51 | 0.248 | 1.18E-264 | ExN4 | CRYM |
| KCNT2 | 8.23E-269 | 0.36658543 | 0.816 | 0.522 | 3.15E-264 | ExN4 | KCNT2 |
| LINC00299 | 2.15E-268 | 0.47975331 | 0.516 | 0.262 | 8.23E-264 | ExN4 | LINC00299 |
| STAMBPL11 | 3.92E-268 | 0.48494286 | 0.747 | 0.495 | 1.50E-263 | ExN4 | STAMBPL1 |
| SYT72 | 6.96E-268 | 0.46708941 | 0.772 | 0.513 | 2.66E-263 | ExN4 | SYT7 |
| LINC026091 | 7.23E-268 | 0.29737857 | 0.356 | 0.139 | 2.76E-263 | ExN4 | LINC02609 |
| TMSB10 | 6.58E-267 | 0.74474077 | 0.561 | 0.315 | 2.51E-262 | ExN4 | TMSB10 |
| RIT21 | 1.59E-265 | 0.43331839 | 0.596 | 0.301 | 6.07E-261 | ExN4 | RIT2 |
| STXBP5L2 | 1.90E-265 | 0.43024882 | 0.981 | 0.945 | 7.26E-261 | ExN4 | STXBP5L |
| ANO32 | 1.21E-263 | 0.51033524 | 0.662 | 0.376 | 4.63E-259 | ExN4 | ANO3 |
| NRCAM1 | 1.49E-263 | 0.39727242 | 0.97 | 0.916 | 5.70E-259 | ExN4 | NRCAM |
| SSBP31 | 6.63E-263 | 0.47835698 | 0.866 | 0.661 | 2.53E-258 | ExN4 | SSBP3 |
| MROH71 | 3.38E-262 | 0.33834441 | 0.439 | 0.195 | 1.29E-257 | ExN4 | MROH7 |
| SEC24D | 6.59E-262 | 0.35843918 | 0.517 | 0.256 | 2.52E-257 | ExN4 | SEC24D |
| GRM72 | 2.88E-261 | 0.5384855 | 0.973 | 0.854 | 1.10E-256 | ExN4 | GRM7 |
| ARL151 | 4.86E-259 | 0.6275104 | 0.903 | 0.801 | 1.86E-254 | ExN4 | ARL15 |
| ITPK11 | 4.38E-256 | 0.44811268 | 0.699 | 0.446 | 1.67E-251 | ExN4 | ITPK1 |
| FGD6 | 4.63E-256 | 0.31464519 | 0.347 | 0.141 | 1.77E-251 | ExN4 | FGD6 |
| MAN2A11 | 4.73E-256 | 0.43080963 | 0.806 | 0.571 | 1.81E-251 | ExN4 | MAN2A1 |
| ST3GAL61 | 6.78E-255 | 0.46783043 | 0.673 | 0.414 | 2.59E-250 | ExN4 | ST3GAL6 |
| SLC35F11 | 1.86E-252 | 0.46732861 | 0.937 | 0.689 | 7.10E-248 | ExN4 | SLC35F1 |
| ANKRD34C-A | 4.74E-252 | 0.46995786 | 0.736 | 0.475 | 1.81E-247 | ExN4 | ANKRD34C-A |
| NR2F1-AS1 | 7.06E-252 | 0.47544503 | 0.917 | 0.776 | 2.70E-247 | ExN4 | NR2F1-AS1 |
| SLC39A11 | 7.58E-251 | 0.54016101 | 0.647 | 0.4 | 2.90E-246 | ExN4 | SLC39A11 |
| CTBP2 | 2.46E-250 | 0.40201667 | 0.591 | 0.332 | 9.39E-246 | ExN4 | CTBP2 |
| MID1 | 5.18E-250 | 0.2725548 | 0.326 | 0.123 | 1.98E-245 | ExN4 | MID1 |
| ATXN1 | 6.86E-250 | 0.39846304 | 0.966 | 0.924 | 2.62E-245 | ExN4 | ATXN1 |
| RBFOX31 | 1.75E-249 | 0.46323087 | 0.956 | 0.878 | 6.68E-245 | ExN4 | RBFOX3 |
| AC103855.21 | 8.61E-249 | 0.41474883 | 0.511 | 0.264 | 3.29E-244 | ExN4 | AC103855.2 |
| SLIT31 | 7.75E-244 | 0.65248649 | 0.707 | 0.449 | 2.96E-239 | ExN4 | SLIT3 |
| EFHD2 | 4.58E-243 | 0.36810475 | 0.489 | 0.251 | 1.75E-238 | ExN4 | EFHD2 |
| AL731769.2 | 6.66E-242 | 0.37365994 | 0.356 | 0.152 | 2.55E-237 | ExN4 | AL731769.2 |
| CABP13 | 5.90E-241 | 0.45999617 | 0.893 | 0.688 | 2.25E-236 | ExN4 | CABP1 |
| LINC01435 | 1.86E-238 | 0.32120902 | 0.3 | 0.115 | 7.11E-234 | ExN4 | LINC01435 |
| MAPK101 | 2.28E-238 | 0.35018982 | 0.981 | 0.95 | 8.70E-234 | ExN4 | MAPK10 |
| MEF2C1 | 1.01E-236 | 0.38508352 | 0.978 | 0.935 | 3.88E-232 | ExN4 | MEF2C |
| RETREG11 | 2.26E-235 | 0.43763219 | 0.797 | 0.56 | 8.66E-231 | ExN4 | RETREG1 |
| NAPEPLD1 | 8.12E-235 | 0.42199049 | 0.713 | 0.475 | 3.10E-230 | ExN4 | NAPEPLD |
| TTC39C1 | 8.43E-235 | 0.45319647 | 0.795 | 0.585 | 3.22E-230 | ExN4 | TTC39C |
| ST6GAL11 | 1.96E-234 | 0.32206422 | 0.497 | 0.255 | 7.49E-230 | ExN4 | ST6GAL1 |
| NEK111 | 1.37E-233 | 0.46234454 | 0.652 | 0.419 | 5.24E-229 | ExN4 | NEK11 |
| SYNPO21 | 1.70E-233 | 0.42685086 | 0.664 | 0.408 | 6.49E-229 | ExN4 | SYNPO2 |
| IL12RB2 | 4.10E-233 | 0.27705517 | 0.419 | 0.19 | 1.57E-228 | ExN4 | IL12RB2 |
| NAALAD2 | 2.36E-232 | 0.40622329 | 0.686 | 0.456 | 9.02E-228 | ExN4 | NAALAD2 |
| DYNC1I11 | 3.81E-232 | 0.41693966 | 0.942 | 0.866 | 1.46E-227 | ExN4 | DYNC1I1 |
| BASP1-AS12 | 5.43E-231 | 0.4755679 | 0.872 | 0.64 | 2.08E-226 | ExN4 | BASP1-AS1 |
| CCSER1 | 4.26E-230 | 0.37647775 | 0.983 | 0.963 | 1.63E-225 | ExN4 | CCSER1 |

| | | | | | | | |
|---|---|---|---|---|---|---|---|
| STX1A2 | 1.16E-229 | 0.41275374 | 0.594 | 0.358 | 4.42E-225 | ExN4 | STX1A |
| XYLT11 | 2.02E-228 | 0.5532138 | 0.764 | 0.542 | 7.71E-224 | ExN4 | XYLT1 |
| COL23A1 | 6.72E-228 | 0.33396315 | 0.405 | 0.189 | 2.57E-223 | ExN4 | COL23A1 |
| LHX21 | 4.10E-227 | 0.30613314 | 0.391 | 0.179 | 1.57E-222 | ExN4 | LHX2 |
| MIR31HG1 | 8.43E-227 | 0.35830216 | 0.418 | 0.192 | 3.22E-222 | ExN4 | MIR31HG |
| ATP2B41 | 1.23E-226 | 0.41705923 | 0.776 | 0.495 | 4.68E-222 | ExN4 | ATP2B4 |
| SV2B2 | 2.02E-225 | 0.45811821 | 0.883 | 0.585 | 7.72E-221 | ExN4 | SV2B |
| NUDT4 | 9.56E-225 | 0.37789041 | 0.502 | 0.274 | 3.65E-220 | ExN4 | NUDT4 |
| KBTBD121 | 2.53E-224 | 0.35373939 | 0.477 | 0.244 | 9.68E-220 | ExN4 | KBTBD12 |
| FAM3C | 7.03E-222 | 0.46550152 | 0.621 | 0.396 | 2.69E-217 | ExN4 | FAM3C |
| MAP1B | 7.33E-222 | 0.42542459 | 0.961 | 0.893 | 2.80E-217 | ExN4 | MAP1B |
| AC120193.11 | 3.36E-220 | 0.56448767 | 0.542 | 0.309 | 1.28E-215 | ExN4 | AC120193.1 |
| PEX5L | 4.28E-220 | 0.41908884 | 0.951 | 0.818 | 1.64E-215 | ExN4 | PEX5L |
| AC092691.1 | 1.10E-219 | 0.37185231 | 0.97 | 0.86 | 4.22E-215 | ExN4 | AC092691.1 |
| PAK11 | 2.61E-219 | 0.4575672 | 0.837 | 0.679 | 9.97E-215 | ExN4 | PAK1 |
| PPP2R2B1 | 2.75E-219 | 0.35527 | 0.985 | 0.954 | 1.05E-214 | ExN4 | PPP2R2B |
| ACOT111 | 2.51E-218 | 0.28146946 | 0.437 | 0.207 | 9.61E-214 | ExN4 | ACOT11 |
| SYTL23 | 4.93E-215 | 0.43062816 | 0.7 | 0.465 | 1.89E-210 | ExN4 | SYTL2 |
| CNKSR23 | 3.48E-214 | 0.44880503 | 0.943 | 0.81 | 1.33E-209 | ExN4 | CNKSR2 |
| EFNB2 | 4.76E-212 | 0.33368465 | 0.49 | 0.265 | 1.82E-207 | ExN4 | EFNB2 |
| RHOBTB11 | 2.62E-208 | 0.32312821 | 0.505 | 0.272 | 1.00E-203 | ExN4 | RHOBTB1 |
| SASH11 | 8.74E-208 | 0.40307241 | 0.575 | 0.336 | 3.34E-203 | ExN4 | SASH1 |
| NPTX12 | 1.62E-207 | 0.3671582 | 0.732 | 0.464 | 6.21E-203 | ExN4 | NPTX1 |
| NUAK11 | 4.85E-207 | 0.42266115 | 0.789 | 0.58 | 1.85E-202 | ExN4 | NUAK1 |
| PLAGL1 | 5.03E-207 | 0.35497348 | 0.436 | 0.219 | 1.92E-202 | ExN4 | PLAGL1 |
| NELL23 | 2.75E-206 | 0.40719423 | 0.966 | 0.775 | 1.05E-201 | ExN4 | NELL2 |
| PDZRN31 | 2.97E-206 | 0.4354424 | 0.921 | 0.749 | 1.13E-201 | ExN4 | PDZRN3 |
| CDHR31 | 1.81E-202 | 0.34946207 | 0.612 | 0.359 | 6.93E-198 | ExN4 | CDHR3 |
| CNTN51 | 1.46E-201 | 0.3065942 | 0.98 | 0.801 | 5.57E-197 | ExN4 | CNTN5 |
| ANKS1B1 | 2.30E-201 | 0.29480574 | 0.994 | 0.986 | 8.81E-197 | ExN4 | ANKS1B |
| THRB1 | 6.67E-199 | 0.37986088 | 0.963 | 0.913 | 2.55E-194 | ExN4 | THRB |
| AL158077.2 | 9.69E-198 | 0.33834947 | 0.484 | 0.259 | 3.70E-193 | ExN4 | AL158077.2 |
| LIN7A1 | 1.32E-197 | 0.4102797 | 0.763 | 0.525 | 5.03E-193 | ExN4 | LIN7A |
| SRRM4 | 2.90E-197 | 0.38841679 | 0.938 | 0.847 | 1.11E-192 | ExN4 | SRRM4 |
| KLHL41 | 3.41E-195 | 0.3384199 | 0.502 | 0.273 | 1.30E-190 | ExN4 | KLHL4 |
| UNC13B1 | 3.86E-195 | 0.33913466 | 0.602 | 0.363 | 1.47E-190 | ExN4 | UNC13B |
| SAMD5 | 3.16E-194 | 0.28543741 | 0.552 | 0.296 | 1.21E-189 | ExN4 | SAMD5 |
| R3HDM13 | 7.89E-194 | 0.35241893 | 0.977 | 0.886 | 3.01E-189 | ExN4 | R3HDM1 |
| CAPS2 | 1.81E-193 | 0.43036058 | 0.62 | 0.415 | 6.93E-189 | ExN4 | CAPS2 |
| CELF22 | 4.38E-191 | 0.3314661 | 0.995 | 0.966 | 1.68E-186 | ExN4 | CELF2 |
| PPP1R16B2 | 3.28E-188 | 0.39875844 | 0.833 | 0.632 | 1.25E-183 | ExN4 | PPP1R16B |
| SNAP251 | 3.40E-188 | 0.34277678 | 0.968 | 0.917 | 1.30E-183 | ExN4 | SNAP25 |
| DNAH141 | 4.76E-187 | 0.3239826 | 0.51 | 0.28 | 1.82E-182 | ExN4 | DNAH14 |
| TEAD1 | 6.01E-187 | 0.42511193 | 0.753 | 0.547 | 2.30E-182 | ExN4 | TEAD1 |
| BASP11 | 6.96E-187 | 0.4075757 | 0.897 | 0.76 | 2.66E-182 | ExN4 | BASP1 |
| AC019211.12 | 8.17E-186 | 0.34469023 | 0.601 | 0.332 | 3.12E-181 | ExN4 | AC019211.1 |
| AL513166.12 | 6.35E-182 | 0.46264851 | 0.815 | 0.607 | 2.43E-177 | ExN4 | AL513166.1 |
| AC093523.11 | 4.65E-181 | 0.42033248 | 0.626 | 0.401 | 1.78E-176 | ExN4 | AC093523.1 |
| CACNB41 | 6.74E-180 | 0.35436428 | 0.954 | 0.845 | 2.58E-175 | ExN4 | CACNB4 |
| ST6GALNAC5 | 3.92E-179 | 0.47384437 | 0.895 | 0.697 | 1.50E-174 | ExN4 | ST6GALNAC5 |
| DCAF62 | 2.32E-178 | 0.39924411 | 0.798 | 0.61 | 8.86E-174 | ExN4 | DCAF6 |
| PCDH15 | 1.88E-177 | 0.28709544 | 0.54 | 0.308 | 7.18E-173 | ExN4 | PCDH15 |
| PHACTR2 | 2.60E-177 | 0.45917561 | 0.617 | 0.391 | 9.93E-173 | ExN4 | PHACTR2 |
| PLCL2 | 1.39E-176 | 0.37944792 | 0.83 | 0.685 | 5.31E-172 | ExN4 | PLCL2 |
| AC124854.11 | 1.59E-174 | 0.37881128 | 0.549 | 0.328 | 6.08E-170 | ExN4 | AC124854.1 |
| GABRG3 | 6.34E-174 | 0.32686419 | 0.938 | 0.789 | 2.42E-169 | ExN4 | GABRG3 |
| AFF11 | 2.70E-173 | 0.34640771 | 0.63 | 0.395 | 1.03E-168 | ExN4 | AFF1 |
| COBL1 | 2.19E-171 | 0.34230764 | 0.901 | 0.737 | 8.39E-167 | ExN4 | COBL |
| LARGE1 | 4.68E-171 | 0.3548879 | 0.959 | 0.907 | 1.79E-166 | ExN4 | LARGE1 |
| MMP161 | 1.34E-170 | 0.37338507 | 0.936 | 0.788 | 5.12E-166 | ExN4 | MMP16 |
| STARD13 | 2.23E-170 | 0.32517399 | 0.755 | 0.521 | 8.53E-166 | ExN4 | STARD13 |
| ARHGAP12 | 3.76E-170 | 0.26901982 | 0.55 | 0.318 | 1.44E-165 | ExN4 | ARHGAP12 |
| LINC013781 | 1.95E-168 | 0.31435949 | 0.586 | 0.329 | 7.46E-164 | ExN4 | LINC01378 |

| | | | | | | | |
|---|---|---|---|---|---|---|---|
| HIVEP1 | 9.69E-167 | 0.33714287 | 0.65 | 0.446 | 3.70E-162 | ExN4 | HIVEP1 |
| DLG12 | 4.67E-164 | 0.36249949 | 0.909 | 0.764 | 1.78E-159 | ExN4 | DLG1 |
| ACVR2A | 1.96E-163 | 0.36334735 | 0.635 | 0.444 | 7.48E-159 | ExN4 | ACVR2A |
| MBP | 3.32E-162 | 0.26142801 | 0.744 | 0.513 | 1.27E-157 | ExN4 | MBP |
| TLN21 | 7.60E-162 | 0.34545889 | 0.89 | 0.723 | 2.90E-157 | ExN4 | TLN2 |
| GLRB | 7.28E-161 | 0.36250847 | 0.742 | 0.559 | 2.78E-156 | ExN4 | GLRB |
| HIVEP3 | 1.06E-159 | 0.38132448 | 0.937 | 0.855 | 4.06E-155 | ExN4 | HIVEP3 |
| SLC6A73 | 1.39E-159 | 0.28665842 | 0.516 | 0.307 | 5.31E-155 | ExN4 | SLC6A7 |
| LINC00632 | 3.95E-159 | 0.31043245 | 0.97 | 0.944 | 1.51E-154 | ExN4 | LINC00632 |
| IQSEC1 | 1.24E-158 | 0.32838882 | 0.893 | 0.772 | 4.74E-154 | ExN4 | IQSEC1 |
| AC091078.11 | 1.08E-156 | 0.26832281 | 0.516 | 0.29 | 4.14E-152 | ExN4 | AC091078.1 |
| SNTG2 | 3.64E-155 | 0.3274512 | 0.303 | 0.141 | 1.39E-150 | ExN4 | SNTG2 |
| EPHA43 | 6.63E-155 | 0.35120762 | 0.884 | 0.66 | 2.53E-150 | ExN4 | EPHA4 |
| SLC17A73 | 8.09E-155 | 0.32362311 | 0.652 | 0.421 | 3.09E-150 | ExN4 | SLC17A7 |
| GPR158 | 3.05E-154 | 0.31613886 | 0.874 | 0.74 | 1.17E-149 | ExN4 | GPR158 |
| AC067956.12 | 1.31E-153 | 0.28132725 | 0.551 | 0.307 | 4.99E-149 | ExN4 | AC067956.1 |
| LINC01322 | 1.86E-153 | 0.37090717 | 0.744 | 0.526 | 7.09E-149 | ExN4 | LINC01322 |
| CXXC41 | 2.37E-152 | 0.29033225 | 0.461 | 0.257 | 9.06E-148 | ExN4 | CXXC4 |
| MGLL | 2.85E-151 | 0.26652323 | 0.644 | 0.421 | 1.09E-146 | ExN4 | MGLL |
| RAPGEF22 | 3.39E-151 | 0.35004081 | 0.943 | 0.85 | 1.29E-146 | ExN4 | RAPGEF2 |
| DMD2 | 1.98E-149 | 0.38731121 | 0.971 | 0.917 | 7.57E-145 | ExN4 | DMD |
| MCTP1 | 3.89E-149 | 0.34662022 | 0.9 | 0.757 | 1.49E-144 | ExN4 | MCTP1 |
| KLF12 | 3.98E-149 | 0.30541845 | 0.812 | 0.594 | 1.52E-144 | ExN4 | KLF12 |
| SYN2 | 9.60E-149 | 0.28389549 | 0.953 | 0.895 | 3.67E-144 | ExN4 | SYN2 |
| TCF7L21 | 3.53E-148 | 0.35738854 | 0.451 | 0.264 | 1.35E-143 | ExN4 | TCF7L2 |
| LINC011221 | 1.49E-147 | 0.27226116 | 0.903 | 0.673 | 5.70E-143 | ExN4 | LINC01122 |
| LINC014141 | 1.72E-147 | 0.3383051 | 0.662 | 0.441 | 6.58E-143 | ExN4 | LINC01414 |
| RAP1GAP22 | 5.99E-147 | 0.3523796 | 0.804 | 0.624 | 2.29E-142 | ExN4 | RAP1GAP2 |
| PCED1B1 | 6.85E-147 | 0.26838619 | 0.268 | 0.122 | 2.62E-142 | ExN4 | PCED1B |
| PLXDC21 | 1.41E-146 | 0.33726208 | 0.816 | 0.594 | 5.39E-142 | ExN4 | PLXDC2 |
| AC073365.13 | 7.25E-146 | 0.38029856 | 0.586 | 0.366 | 2.77E-141 | ExN4 | AC073365.1 |
| CACNA1A1 | 6.87E-145 | 0.29419477 | 0.97 | 0.904 | 2.62E-140 | ExN4 | CACNA1A |
| AGAP11 | 1.17E-142 | 0.28634775 | 0.969 | 0.905 | 4.45E-138 | ExN4 | AGAP1 |
| PRKCB3 | 1.48E-142 | 0.31258808 | 0.974 | 0.859 | 5.67E-138 | ExN4 | PRKCB |
| NREP | 1.87E-142 | 0.27620897 | 0.546 | 0.344 | 7.14E-138 | ExN4 | NREP |
| CHN12 | 1.30E-141 | 0.29785021 | 0.935 | 0.736 | 4.95E-137 | ExN4 | CHN1 |
| KCNIP3 | 1.15E-140 | 0.25952573 | 0.463 | 0.276 | 4.39E-136 | ExN4 | KCNIP3 |
| FAM13C | 3.55E-140 | 0.29536462 | 0.691 | 0.496 | 1.36E-135 | ExN4 | FAM13C |
| AC010280.11 | 1.42E-139 | 0.28110339 | 0.441 | 0.251 | 5.43E-135 | ExN4 | AC010280.1 |
| TUBA1B1 | 3.18E-139 | 0.4446082 | 0.715 | 0.554 | 1.22E-134 | ExN4 | TUBA1B |
| LINGO12 | 7.65E-139 | 0.31690328 | 0.698 | 0.488 | 2.93E-134 | ExN4 | LINGO1 |
| PCDH10 | 1.44E-138 | 0.33700358 | 0.64 | 0.435 | 5.50E-134 | ExN4 | PCDH10 |
| TMEM72-AS1 | 1.71E-138 | 0.26828513 | 0.533 | 0.333 | 6.54E-134 | ExN4 | TMEM72-AS1 |
| RGS62 | 1.97E-138 | 0.49185851 | 0.8 | 0.696 | 7.52E-134 | ExN4 | RGS6 |
| VPS411 | 4.98E-138 | 0.32796718 | 0.748 | 0.579 | 1.90E-133 | ExN4 | VPS41 |
| SLIT11 | 8.78E-138 | 0.28207509 | 0.792 | 0.577 | 3.35E-133 | ExN4 | SLIT1 |
| HERC1 | 1.56E-137 | 0.28611316 | 0.927 | 0.875 | 5.96E-133 | ExN4 | HERC1 |
| MIR137HG2 | 2.25E-137 | 0.33963873 | 0.868 | 0.644 | 8.61E-133 | ExN4 | MIR137HG |
| SNTG12 | 1.54E-136 | 0.26718416 | 0.986 | 0.938 | 5.89E-132 | ExN4 | SNTG1 |
| ZBTB16 | 2.63E-134 | 0.32037826 | 0.947 | 0.871 | 1.00E-129 | ExN4 | ZBTB16 |
| KCNMB4 | 2.91E-134 | 0.31712603 | 0.647 | 0.453 | 1.11E-129 | ExN4 | KCNMB4 |
| EPS15 | 3.29E-134 | 0.29597104 | 0.658 | 0.472 | 1.26E-129 | ExN4 | EPS15 |
| AL807742.11 | 3.35E-134 | 0.43004161 | 0.266 | 0.128 | 1.28E-129 | ExN4 | AL807742.1 |
| OXR11 | 1.18E-131 | 0.27105125 | 0.972 | 0.917 | 4.51E-127 | ExN4 | OXR1 |
| MYRIP | 1.15E-129 | 0.25378673 | 0.911 | 0.797 | 4.38E-125 | ExN4 | MYRIP |
| MSC-AS12 | 1.35E-129 | 0.38871623 | 0.385 | 0.217 | 5.15E-125 | ExN4 | MSC-AS1 |
| FAM155A1 | 3.66E-129 | 0.25119528 | 0.995 | 0.985 | 1.40E-124 | ExN4 | FAM155A |
| MYO5A | 5.28E-129 | 0.31310226 | 0.929 | 0.843 | 2.02E-124 | ExN4 | MYO5A |
| DIRC32 | 9.28E-129 | 0.34598328 | 0.728 | 0.529 | 3.55E-124 | ExN4 | DIRC3 |
| KALRN2 | 1.00E-128 | 0.27698488 | 0.992 | 0.954 | 3.84E-124 | ExN4 | KALRN |
| CHSY31 | 6.35E-127 | 0.31445006 | 0.94 | 0.827 | 2.43E-122 | ExN4 | CHSY3 |
| CAMK2G | 2.55E-126 | 0.30744044 | 0.741 | 0.573 | 9.74E-122 | ExN4 | CAMK2G |
| ARHGAP221 | 2.20E-125 | 0.25135089 | 0.476 | 0.294 | 8.42E-121 | ExN4 | ARHGAP22 |

| | | | | | | | |
|---|---|---|---|---|---|---|---|
| HDAC4 | 2.46E-124 | 0.30709266 | 0.811 | 0.653 | 9.39E-120 | ExN4 | HDAC4 |
| KCNIP4-IT13 | 6.80E-123 | 0.25115531 | 0.724 | 0.483 | 2.60E-118 | ExN4 | KCNIP4-IT1 |
| HLF | 1.93E-122 | 0.36331607 | 0.68 | 0.499 | 7.38E-118 | ExN4 | HLF |
| NUMB | 2.15E-121 | 0.29183545 | 0.709 | 0.543 | 8.21E-117 | ExN4 | NUMB |
| PIK3R11 | 7.71E-120 | 0.3030656 | 0.834 | 0.679 | 2.95E-115 | ExN4 | PIK3R1 |
| FAM135B | 1.00E-119 | 0.32099843 | 0.778 | 0.62 | 3.84E-115 | ExN4 | FAM135B |
| HPCA | 2.76E-119 | 0.25537684 | 0.406 | 0.239 | 1.06E-114 | ExN4 | HPCA |
| KCNIP43 | 8.34E-118 | 0.31448659 | 0.999 | 0.933 | 3.19E-113 | ExN4 | KCNIP4 |
| DSCAM | 2.56E-117 | 0.27519224 | 0.984 | 0.898 | 9.80E-113 | ExN4 | DSCAM |
| LRFN53 | 4.23E-117 | 0.28753232 | 0.969 | 0.886 | 1.62E-112 | ExN4 | LRFN5 |
| SIDT1 | 1.05E-116 | 0.2821791 | 0.692 | 0.504 | 4.01E-112 | ExN4 | SIDT1 |
| RALGPS2 | 8.37E-116 | 0.29467705 | 0.698 | 0.526 | 3.20E-111 | ExN4 | RALGPS2 |
| STRBP | 3.06E-115 | 0.30286791 | 0.865 | 0.751 | 1.17E-110 | ExN4 | STRBP |
| EFNA52 | 3.82E-115 | 0.30525179 | 0.945 | 0.785 | 1.46E-110 | ExN4 | EFNA5 |
| RALGAPA2 | 2.04E-114 | 0.32299478 | 0.748 | 0.577 | 7.79E-110 | ExN4 | RALGAPA2 |
| EPHB6 | 4.53E-113 | 0.27382017 | 0.592 | 0.411 | 1.73E-108 | ExN4 | EPHB6 |
| RAPGEF52 | 1.10E-112 | 0.29579465 | 0.932 | 0.736 | 4.20E-108 | ExN4 | RAPGEF5 |
| RAPGEF4 | 7.93E-112 | 0.27066646 | 0.932 | 0.87 | 3.03E-107 | ExN4 | RAPGEF4 |
| LDB23 | 8.26E-110 | 0.31222737 | 0.971 | 0.747 | 3.16E-105 | ExN4 | LDB2 |
| AL391840.12 | 1.58E-109 | 0.26388358 | 0.723 | 0.495 | 6.06E-105 | ExN4 | AL391840.1 |
| TRIM44 | 1.27E-108 | 0.29964706 | 0.824 | 0.699 | 4.86E-104 | ExN4 | TRIM44 |
| FUT8 | 1.80E-108 | 0.30278946 | 0.799 | 0.688 | 6.87E-104 | ExN4 | FUT8 |
| MRPS30-DT2 | 4.01E-108 | 0.27371846 | 0.632 | 0.448 | 1.53E-103 | ExN4 | MRPS30-DT |
| STK39 | 1.20E-107 | 0.28835353 | 0.743 | 0.589 | 4.60E-103 | ExN4 | STK39 |
| CXXC4-AS11 | 1.64E-106 | 0.32569497 | 0.545 | 0.374 | 6.26E-102 | ExN4 | CXXC4-AS1 |
| TMEM232 | 1.82E-106 | 0.32686862 | 0.894 | 0.763 | 6.94E-102 | ExN4 | TMEM232 |
| DDAH11 | 5.96E-105 | 0.27066107 | 0.597 | 0.41 | 2.28E-100 | ExN4 | DDAH1 |
| LINC022541 | 8.52E-105 | 0.25296855 | 0.291 | 0.155 | 3.26E-100 | ExN4 | LINC02254 |
| AL008633.12 | 8.60E-100 | 0.30391062 | 0.872 | 0.703 | 3.29E-95 | ExN4 | AL008633.1 |
| SHISA9 | 1.69E-99 | 0.3027652 | 0.956 | 0.811 | 6.44E-95 | ExN4 | SHISA9 |
| SLCO3A11 | 3.22E-98 | 0.27805415 | 0.858 | 0.725 | 1.23E-93 | ExN4 | SLCO3A1 |
| ZFYVE28 | 1.72E-97 | 0.27052675 | 0.81 | 0.683 | 6.58E-93 | ExN4 | ZFYVE28 |
| GCC2 | 2.05E-97 | 0.26529184 | 0.693 | 0.539 | 7.85E-93 | ExN4 | GCC2 |
| ASAP2 | 5.79E-96 | 0.2533335 | 0.558 | 0.389 | 2.21E-91 | ExN4 | ASAP2 |
| ZEB12 | 2.77E-94 | 0.28533668 | 0.878 | 0.754 | 1.06E-89 | ExN4 | ZEB1 |
| SLC1A2 | 2.04E-91 | 0.30670456 | 0.62 | 0.448 | 7.80E-87 | ExN4 | SLC1A2 |
| AJ009632.21 | 4.55E-91 | 0.3293026 | 0.292 | 0.161 | 1.74E-86 | ExN4 | AJ009632.2 |
| PPM1L1 | 3.10E-90 | 0.27233401 | 0.909 | 0.802 | 1.19E-85 | ExN4 | PPM1L |
| NEFL | 4.34E-90 | 0.25905696 | 0.449 | 0.3 | 1.66E-85 | ExN4 | NEFL |
| PPP2R2C3 | 5.20E-89 | 0.25204048 | 0.906 | 0.792 | 1.99E-84 | ExN4 | PPP2R2C |
| ARNTL | 2.22E-85 | 0.25897508 | 0.612 | 0.453 | 8.49E-81 | ExN4 | ARNTL |
| HCN11 | 6.71E-85 | 0.2581633 | 0.926 | 0.854 | 2.57E-80 | ExN4 | HCN1 |
| PRICKLE11 | 1.30E-83 | 0.25453416 | 0.831 | 0.681 | 4.95E-79 | ExN4 | PRICKLE1 |
| SNAP91 | 1.52E-82 | 0.25058924 | 0.912 | 0.842 | 5.83E-78 | ExN4 | SNAP91 |
| ELL21 | 4.00E-81 | 0.25128376 | 0.722 | 0.533 | 1.53E-76 | ExN4 | ELL2 |
| FARS2 | 3.31E-79 | 0.25014377 | 0.872 | 0.754 | 1.27E-74 | ExN4 | FARS2 |
| RNLS1 | 4.65E-77 | 0.25725387 | 0.609 | 0.46 | 1.78E-72 | ExN4 | RNLS |
| STMN11 | 1.56E-73 | 0.28492951 | 0.605 | 0.466 | 5.97E-69 | ExN4 | STMN1 |
| SLC5A4-AS11 | 6.71E-72 | 0.30535471 | 0.552 | 0.404 | 2.57E-67 | ExN4 | SLC5A4-AS1 |
| SLC23A2 | 1.85E-70 | 0.25345078 | 0.794 | 0.672 | 7.09E-66 | ExN4 | SLC23A2 |
| PTPRK3 | 0 | 1.66614173 | 0.98 | 0.593 | 0 | ExN5 | PTPRK |
| CTNNA22 | 0 | 1.66363768 | 0.995 | 0.957 | 0 | ExN5 | CTNNA2 |
| LINC02196 | 0 | 1.61719232 | 0.384 | 0.059 | 0 | ExN5 | LINC02196 |
| PDZRN42 | 0 | 1.56404321 | 0.919 | 0.378 | 0 | ExN5 | PDZRN4 |
| AC019211.13 | 0 | 1.46605487 | 0.914 | 0.331 | 0 | ExN5 | AC019211.1 |
| AC067956.13 | 0 | 1.31519747 | 0.864 | 0.305 | 0 | ExN5 | AC067956.1 |
| LRP1B1 | 0 | 1.28955093 | 0.995 | 0.948 | 0 | ExN5 | LRP1B |
| CLSTN22 | 0 | 1.25633696 | 0.965 | 0.612 | 0 | ExN5 | CLSTN2 |
| ADGRL4 | 0 | 1.18662446 | 0.294 | 0.048 | 0 | ExN5 | ADGRL4 |
| AC099517.11 | 0 | 1.11265249 | 0.435 | 0.049 | 0 | ExN5 | AC099517.1 |
| DACH11 | 0 | 1.10433449 | 0.678 | 0.187 | 0 | ExN5 | DACH1 |
| HS3ST22 | 0 | 1.07630289 | 0.741 | 0.247 | 0 | ExN5 | HS3ST2 |
| POU6F23 | 0 | 1.06294003 | 0.901 | 0.384 | 0 | ExN5 | POU6F2 |

| | | | | | | | |
|---|---|---|---|---|---|---|---|
| PLXDC22 | 0 | 1.03768791 | 0.936 | 0.599 | 0 | ExN5 | PLXDC2 |
| RXFP11 | 0 | 1.02934873 | 0.757 | 0.207 | 0 | ExN5 | RXFP1 |
| THEMIS1 | 0 | 1.02697998 | 0.583 | 0.067 | 0 | ExN5 | THEMIS |
| RIT22 | 0 | 0.94667233 | 0.792 | 0.306 | 0 | ExN5 | RIT2 |
| KHDRBS33 | 0 | 0.92924074 | 0.975 | 0.791 | 0 | ExN5 | KHDRBS3 |
| SLC39A111 | 0 | 0.88057009 | 0.793 | 0.405 | 0 | ExN5 | SLC39A11 |
| SEMA3A2 | 0 | 0.86819418 | 0.587 | 0.197 | 0 | ExN5 | SEMA3A |
| AL138701.21 | 0 | 0.86474259 | 0.563 | 0.09 | 0 | ExN5 | AL138701.2 |
| AC093843.12 | 0 | 0.85252628 | 0.715 | 0.2 | 0 | ExN5 | AC093843.1 |
| NELL24 | 0 | 0.84526669 | 0.973 | 0.784 | 0 | ExN5 | NELL2 |
| ARHGEF281 | 0 | 0.82387653 | 0.71 | 0.295 | 0 | ExN5 | ARHGEF28 |
| PDE4D3 | 0 | 0.80975555 | 0.995 | 0.96 | 0 | ExN5 | PDE4D |
| MMP162 | 0 | 0.80483016 | 0.959 | 0.794 | 0 | ExN5 | MMP16 |
| RALYL3 | 0 | 0.79239589 | 0.995 | 0.812 | 0 | ExN5 | RALYL |
| LINC00343 | 0 | 0.77777941 | 0.429 | 0.047 | 0 | ExN5 | LINC00343 |
| LINC022542 | 0 | 0.74111046 | 0.566 | 0.149 | 0 | ExN5 | LINC02254 |
| DCHS2 | 0 | 0.72169564 | 0.523 | 0.11 | 0 | ExN5 | DCHS2 |
| HS3ST5 | 0 | 0.71820392 | 0.746 | 0.299 | 0 | ExN5 | HS3ST5 |
| PDGFC2 | 0 | 0.64332758 | 0.661 | 0.267 | 0 | ExN5 | PDGFC |
| AL136456.1 | 0 | 0.61407971 | 0.523 | 0.132 | 0 | ExN5 | AL136456.1 |
| AC084816.12 | 0 | 0.60593374 | 0.587 | 0.182 | 0 | ExN5 | AC084816.1 |
| OPCML2 | 0 | 0.60155946 | 0.993 | 0.987 | 0 | ExN5 | OPCML |
| MGST1 | 0 | 0.572157 | 0.432 | 0.103 | 0 | ExN5 | MGST1 |
| TEX41 | 0 | 0.45007752 | 0.363 | 0.078 | 0 | ExN5 | TEX41 |
| AC020704.1 | 0 | 0.4447778 | 0.361 | 0.062 | 0 | ExN5 | AC020704.1 |
| BACE2 | 0 | 0.41144053 | 0.466 | 0.133 | 0 | ExN5 | BACE2 |
| AC010931.3 | 0 | 0.39226213 | 0.287 | 0.045 | 0 | ExN5 | AC010931.3 |
| AC008127.11 | 0 | 0.31501104 | 0.332 | 0.07 | 0 | ExN5 | AC008127.1 |
| SEC24D1 | 1.41E-293 | 0.54579315 | 0.641 | 0.263 | 5.38E-289 | ExN5 | SEC24D |
| FOXP22 | 8.19E-288 | 0.9943897 | 0.695 | 0.301 | 3.13E-283 | ExN5 | FOXP2 |
| SV2B3 | 5.24E-287 | 0.76949877 | 0.937 | 0.597 | 2.00E-282 | ExN5 | SV2B |
| GAP432 | 1.68E-283 | 0.74039485 | 0.859 | 0.527 | 6.43E-279 | ExN5 | GAP43 |
| AC090578.12 | 9.86E-281 | 0.8653287 | 0.879 | 0.55 | 3.77E-276 | ExN5 | AC090578.1 |
| NAV32 | 2.14E-279 | 0.62194429 | 0.991 | 0.949 | 8.18E-275 | ExN5 | NAV3 |
| SLC35F32 | 2.51E-279 | 0.92877519 | 0.947 | 0.763 | 9.58E-275 | ExN5 | SLC35F3 |
| FMNL21 | 1.15E-278 | 0.67058179 | 0.967 | 0.87 | 4.39E-274 | ExN5 | FMNL2 |
| NECAB12 | 7.11E-276 | 0.74180135 | 0.932 | 0.635 | 2.72E-271 | ExN5 | NECAB1 |
| TNNT22 | 1.66E-274 | 0.50729823 | 0.392 | 0.118 | 6.35E-270 | ExN5 | TNNT2 |
| OPRM12 | 1.13E-272 | 0.60659462 | 0.568 | 0.219 | 4.31E-268 | ExN5 | OPRM1 |
| LINC00320 | 6.95E-272 | 0.34948428 | 0.381 | 0.109 | 2.66E-267 | ExN5 | LINC00320 |
| EFNA53 | 5.35E-270 | 0.76473034 | 0.966 | 0.792 | 2.04E-265 | ExN5 | EFNA5 |
| CACHD1 | 6.35E-261 | 0.45549011 | 0.603 | 0.244 | 2.43E-256 | ExN5 | CACHD1 |
| LINC014351 | 1.05E-259 | 0.45163372 | 0.393 | 0.119 | 4.03E-255 | ExN5 | LINC01435 |
| DNAH142 | 1.42E-259 | 0.51331492 | 0.667 | 0.284 | 5.44E-255 | ExN5 | DNAH14 |
| SORCS12 | 9.47E-258 | 0.78670714 | 0.954 | 0.667 | 3.62E-253 | ExN5 | SORCS1 |
| NRCAM2 | 1.20E-257 | 0.52870122 | 0.98 | 0.918 | 4.60E-253 | ExN5 | NRCAM |
| LINC028221 | 4.38E-257 | 0.7180101 | 0.422 | 0.139 | 1.67E-252 | ExN5 | LINC02822 |
| CSGALNACT1 | 6.75E-253 | 0.71894493 | 0.886 | 0.602 | 2.58E-248 | ExN5 | CSGALNACT1 |
| CCDC32 | 3.45E-251 | 0.7796011 | 0.914 | 0.638 | 1.32E-246 | ExN5 | CCDC3 |
| AC007614.11 | 3.16E-249 | 0.4397131 | 0.277 | 0.068 | 1.21E-244 | ExN5 | AC007614.1 |
| MCTP11 | 4.44E-249 | 1.0292601 | 0.924 | 0.763 | 1.70E-244 | ExN5 | MCTP1 |
| FOXP13 | 2.14E-246 | 0.62434885 | 0.963 | 0.815 | 8.19E-242 | ExN5 | FOXP1 |
| MYO1D2 | 2.87E-246 | 0.78098733 | 0.811 | 0.508 | 1.10E-241 | ExN5 | MYO1D |
| SOX52 | 6.79E-246 | 0.77117659 | 0.942 | 0.661 | 2.60E-241 | ExN5 | SOX5 |
| IL12RB21 | 4.47E-244 | 0.3835985 | 0.528 | 0.196 | 1.71E-239 | ExN5 | IL12RB2 |
| FRMD61 | 1.08E-240 | 0.58307007 | 0.529 | 0.215 | 4.13E-236 | ExN5 | FRMD6 |
| GRM11 | 3.03E-239 | 0.65166205 | 0.936 | 0.63 | 1.16E-234 | ExN5 | GRM1 |
| TENM41 | 3.69E-238 | 0.61729698 | 0.967 | 0.855 | 1.41E-233 | ExN5 | TENM4 |
| CPNE42 | 2.44E-237 | 0.90491118 | 0.802 | 0.458 | 9.31E-233 | ExN5 | CPNE4 |
| RYR23 | 3.69E-234 | 0.57159727 | 0.986 | 0.926 | 1.41E-229 | ExN5 | RYR2 |
| PEX5L1 | 1.36E-232 | 0.67359288 | 0.961 | 0.824 | 5.19E-228 | ExN5 | PEX5L |
| PKN2-AS12 | 3.56E-232 | 0.63671797 | 0.65 | 0.302 | 1.36E-227 | ExN5 | PKN2-AS1 |
| LINC022322 | 4.58E-231 | 0.82257717 | 0.472 | 0.181 | 1.75E-226 | ExN5 | LINC02232 |

| | | | | | | | |
|---|---|---|---|---|---|---|---|
| MSC-AS13 | 1.99E-230 | 0.57728032 | 0.546 | 0.218 | 7.61E-226 | ExN5 | MSC-AS1 |
| ADAMTS32 | 3.24E-225 | 0.55462241 | 0.601 | 0.256 | 1.24E-220 | ExN5 | ADAMTS3 |
| AC117834.1 | 8.99E-223 | 0.27558518 | 0.255 | 0.062 | 3.44E-218 | ExN5 | AC117834.1 |
| SAMD51 | 7.18E-222 | 0.42619503 | 0.683 | 0.303 | 2.74E-217 | ExN5 | SAMD5 |
| NKAIN23 | 5.76E-218 | 0.59740996 | 0.987 | 0.834 | 2.20E-213 | ExN5 | NKAIN2 |
| IL1RAPL22 | 1.22E-214 | 0.94594404 | 0.654 | 0.312 | 4.66E-210 | ExN5 | IL1RAPL2 |
| LINC01748 | 3.11E-213 | 0.28588944 | 0.365 | 0.116 | 1.19E-208 | ExN5 | LINC01748 |
| SYT11 | 3.20E-211 | 0.43111504 | 0.997 | 0.985 | 1.22E-206 | ExN5 | SYT1 |
| CRIM11 | 3.54E-211 | 0.57234679 | 0.809 | 0.477 | 1.35E-206 | ExN5 | CRIM1 |
| LMO42 | 1.44E-209 | 0.55284015 | 0.856 | 0.518 | 5.51E-205 | ExN5 | LMO4 |
| PART12 | 1.18E-206 | 0.45337254 | 0.616 | 0.283 | 4.53E-202 | ExN5 | PART1 |
| PTK22 | 2.78E-206 | 0.56975839 | 0.971 | 0.897 | 1.06E-201 | ExN5 | PTK2 |
| CABP14 | 1.62E-203 | 0.61600985 | 0.932 | 0.696 | 6.19E-199 | ExN5 | CABP1 |
| SSTR21 | 7.75E-203 | 0.30254528 | 0.467 | 0.176 | 2.96E-198 | ExN5 | SSTR2 |
| ZNF804B1 | 1.47E-201 | 0.70783165 | 0.667 | 0.347 | 5.61E-197 | ExN5 | ZNF804B |
| SLIT12 | 3.79E-201 | 0.56664368 | 0.893 | 0.583 | 1.45E-196 | ExN5 | SLIT1 |
| SPOCK11 | 4.77E-200 | 0.6007625 | 0.95 | 0.743 | 1.82E-195 | ExN5 | SPOCK1 |
| ANO33 | 1.69E-198 | 0.49556964 | 0.766 | 0.386 | 6.47E-194 | ExN5 | ANO3 |
| TBR11 | 3.65E-196 | 0.35534262 | 0.536 | 0.228 | 1.40E-191 | ExN5 | TBR1 |
| KCNQ52 | 7.06E-196 | 0.53073833 | 0.988 | 0.902 | 2.70E-191 | ExN5 | KCNQ5 |
| AL033504.1 | 3.25E-195 | 0.40669251 | 0.583 | 0.251 | 1.24E-190 | ExN5 | AL033504.1 |
| SNTG21 | 1.09E-194 | 0.5176695 | 0.398 | 0.145 | 4.16E-190 | ExN5 | SNTG2 |
| FAP1 | 2.96E-193 | 0.33418047 | 0.352 | 0.116 | 1.13E-188 | ExN5 | FAP |
| AL356295.11 | 7.38E-190 | 0.90042038 | 0.418 | 0.164 | 2.82E-185 | ExN5 | AL356295.1 |
| NRG11 | 1.70E-189 | 0.75981214 | 0.84 | 0.537 | 6.52E-185 | ExN5 | NRG1 |
| ATP8B12 | 2.52E-186 | 0.39868779 | 0.565 | 0.252 | 9.64E-182 | ExN5 | ATP8B1 |
| ADGRL22 | 4.88E-186 | 0.53677195 | 0.969 | 0.852 | 1.87E-181 | ExN5 | ADGRL2 |
| RASGRP1 | 1.05E-184 | 0.29448183 | 0.468 | 0.182 | 4.02E-180 | ExN5 | RASGRP1 |
| SNCA2 | 2.50E-184 | 0.55447202 | 0.888 | 0.602 | 9.57E-180 | ExN5 | SNCA |
| SH3GL24 | 1.34E-182 | 0.56840125 | 0.945 | 0.739 | 5.14E-178 | ExN5 | SH3GL2 |
| LY86-AS13 | 2.62E-181 | 0.5181888 | 0.856 | 0.473 | 1.00E-176 | ExN5 | LY86-AS1 |
| DPP101 | 2.49E-177 | 0.34791918 | 0.989 | 0.843 | 9.53E-173 | ExN5 | DPP10 |
| MRTFB4 | 5.96E-177 | 0.53473733 | 0.924 | 0.756 | 2.28E-172 | ExN5 | MRTFB |
| CDH6 | 3.58E-175 | 0.39273932 | 0.444 | 0.18 | 1.37E-170 | ExN5 | CDH6 |
| RPS6KA51 | 6.03E-175 | 0.55288378 | 0.85 | 0.59 | 2.31E-170 | ExN5 | RPS6KA5 |
| PTPRD3 | 1.41E-172 | 0.48326469 | 0.995 | 0.963 | 5.37E-168 | ExN5 | PTPRD |
| CDYL23 | 2.06E-172 | 0.49680303 | 0.703 | 0.394 | 7.89E-168 | ExN5 | CDYL2 |
| AC011995.22 | 5.27E-172 | 0.43422081 | 0.744 | 0.398 | 2.01E-167 | ExN5 | AC011995.2 |
| PPP3CA3 | 6.01E-172 | 0.48322989 | 0.983 | 0.914 | 2.30E-167 | ExN5 | PPP3CA |
| COL19A1 | 2.82E-171 | 0.32465782 | 0.648 | 0.317 | 1.08E-166 | ExN5 | COL19A1 |
| MAN2A12 | 1.44E-169 | 0.46889004 | 0.863 | 0.58 | 5.51E-165 | ExN5 | MAN2A1 |
| CPNE91 | 1.66E-167 | 0.29299021 | 0.451 | 0.187 | 6.33E-163 | ExN5 | CPNE9 |
| DENND32 | 9.06E-167 | 0.38022829 | 0.648 | 0.335 | 3.46E-162 | ExN5 | DENND3 |
| CAMK1D3 | 6.44E-166 | 0.52007814 | 0.978 | 0.857 | 2.46E-161 | ExN5 | CAMK1D |
| B3GLCT | 1.55E-164 | 0.31736759 | 0.602 | 0.287 | 5.93E-160 | ExN5 | B3GLCT |
| DPYD2 | 1.94E-164 | 0.54048592 | 0.932 | 0.657 | 7.42E-160 | ExN5 | DPYD |
| RAB27B1 | 7.21E-164 | 0.46782221 | 0.768 | 0.445 | 2.76E-159 | ExN5 | RAB27B |
| BMPER2 | 9.85E-163 | 0.55803303 | 0.879 | 0.608 | 3.77E-158 | ExN5 | BMPER |
| KALRN3 | 9.98E-162 | 0.41445135 | 0.989 | 0.956 | 3.82E-157 | ExN5 | KALRN |
| KIAA1211L2 | 2.17E-160 | 0.49979595 | 0.886 | 0.555 | 8.31E-156 | ExN5 | KIAA1211L |
| SNED13 | 2.90E-160 | 0.47152877 | 0.756 | 0.433 | 1.11E-155 | ExN5 | SNED1 |
| RETREG12 | 5.97E-160 | 0.48470059 | 0.836 | 0.569 | 2.28E-155 | ExN5 | RETREG1 |
| USP432 | 6.73E-160 | 0.30038923 | 0.552 | 0.249 | 2.57E-155 | ExN5 | USP43 |
| SHC31 | 2.54E-159 | 0.51263733 | 0.849 | 0.601 | 9.70E-155 | ExN5 | SHC3 |
| PCDH92 | 2.62E-159 | 0.47853477 | 0.998 | 0.966 | 1.00E-154 | ExN5 | PCDH9 |
| SATB23 | 1.23E-157 | 0.4900836 | 0.914 | 0.566 | 4.69E-153 | ExN5 | SATB2 |
| LINC012022 | 9.78E-157 | 0.36034026 | 0.389 | 0.153 | 3.74E-152 | ExN5 | LINC01202 |
| TSHZ1 | 5.35E-156 | 0.38883384 | 0.681 | 0.371 | 2.05E-151 | ExN5 | TSHZ1 |
| ADAMTS162 | 1.30E-155 | 0.34743548 | 0.457 | 0.193 | 4.95E-151 | ExN5 | ADAMTS16 |
| MYOM21 | 2.97E-155 | 0.33995589 | 0.457 | 0.2 | 1.14E-150 | ExN5 | MYOM2 |
| VPS412 | 1.69E-154 | 0.4586399 | 0.839 | 0.583 | 6.45E-150 | ExN5 | VPS41 |
| FMN12 | 8.86E-154 | 0.53654184 | 0.927 | 0.754 | 3.39E-149 | ExN5 | FMN1 |
| CCDC144NL-/ | 3.09E-152 | 0.28756984 | 0.37 | 0.143 | 1.18E-147 | ExN5 | CCDC144NL-/ |

| | | | | | | | |
|---|---|---|---|---|---|---|---|
| AC060765.22 | 3.34E-152 | 0.69434086 | 0.437 | 0.186 | 1.27E-147 | ExN5 | AC060765.2 |
| B4GALNT31 | 3.58E-152 | 0.3421463 | 0.606 | 0.305 | 1.37E-147 | ExN5 | B4GALNT3 |
| AC015943.1 | 6.13E-152 | 0.32500746 | 0.311 | 0.11 | 2.34E-147 | ExN5 | AC015943.1 |
| PPFIBP12 | 3.04E-151 | 0.49778517 | 0.906 | 0.653 | 1.16E-146 | ExN5 | PPFIBP1 |
| NAB1 | 4.40E-151 | 0.3284311 | 0.602 | 0.305 | 1.68E-146 | ExN5 | NAB1 |
| LRRC7 | 7.12E-151 | 0.38292526 | 0.989 | 0.965 | 2.72E-146 | ExN5 | LRRC7 |
| SYNPO22 | 1.71E-150 | 0.43527207 | 0.716 | 0.418 | 6.54E-146 | ExN5 | SYNPO2 |
| PTPN141 | 5.08E-150 | 0.32409415 | 0.494 | 0.226 | 1.94E-145 | ExN5 | PTPN14 |
| GPM6A1 | 5.66E-150 | 0.40055937 | 0.977 | 0.943 | 2.17E-145 | ExN5 | GPM6A |
| CADM21 | 5.80E-149 | 0.34943921 | 0.996 | 0.993 | 2.22E-144 | ExN5 | CADM2 |
| GABRB33 | 1.07E-148 | 0.43082552 | 0.969 | 0.899 | 4.09E-144 | ExN5 | GABRB3 |
| ACTN2 | 1.44E-148 | 0.27591288 | 0.533 | 0.252 | 5.50E-144 | ExN5 | ACTN2 |
| NCAM2 | 1.45E-148 | 0.49748968 | 0.969 | 0.874 | 5.52E-144 | ExN5 | NCAM2 |
| ITGB81 | 2.35E-148 | 0.30128321 | 0.575 | 0.277 | 8.98E-144 | ExN5 | ITGB8 |
| ATRNL13 | 5.07E-148 | 0.5142883 | 0.986 | 0.929 | 1.94E-143 | ExN5 | ATRNL1 |
| AKAP131 | 9.16E-148 | 0.37566326 | 0.662 | 0.356 | 3.50E-143 | ExN5 | AKAP13 |
| MIR137HG3 | 1.07E-147 | 0.49403738 | 0.906 | 0.653 | 4.10E-143 | ExN5 | MIR137HG |
| PPP1R14C | 8.60E-147 | 0.29856737 | 0.493 | 0.229 | 3.29E-142 | ExN5 | PPP1R14C |
| SLC17A74 | 1.20E-146 | 0.42447437 | 0.751 | 0.428 | 4.58E-142 | ExN5 | SLC17A7 |
| GPATCH2 | 1.53E-146 | 0.47598618 | 0.721 | 0.443 | 5.86E-142 | ExN5 | GPATCH2 |
| TOX2 | 4.80E-146 | 1.11574999 | 0.691 | 0.435 | 1.83E-141 | ExN5 | TOX |
| PAPSS1 | 6.23E-144 | 0.44352664 | 0.775 | 0.513 | 2.38E-139 | ExN5 | PAPSS1 |
| AC120193.12 | 2.03E-143 | 0.51803076 | 0.6 | 0.318 | 7.77E-139 | ExN5 | AC120193.1 |
| NPTX13 | 2.06E-143 | 0.40279795 | 0.802 | 0.474 | 7.87E-139 | ExN5 | NPTX1 |
| NFIA | 5.33E-143 | 0.94647704 | 0.619 | 0.415 | 2.04E-138 | ExN5 | NFIA |
| AL024495.11 | 6.85E-143 | 0.36033364 | 0.589 | 0.293 | 2.62E-138 | ExN5 | AL024495.1 |
| DIPK2A1 | 2.89E-142 | 0.40926928 | 0.645 | 0.35 | 1.10E-137 | ExN5 | DIPK2A |
| EPB41L4A1 | 8.16E-142 | 0.27704338 | 0.385 | 0.156 | 3.12E-137 | ExN5 | EPB41L4A |
| PCSK22 | 1.29E-141 | 0.46237108 | 0.937 | 0.798 | 4.94E-137 | ExN5 | PCSK2 |
| KHDRBS22 | 6.74E-141 | 0.45664354 | 0.983 | 0.897 | 2.57E-136 | ExN5 | KHDRBS2 |
| FIGN | 8.77E-141 | 0.26079262 | 0.67 | 0.345 | 3.35E-136 | ExN5 | FIGN |
| CADPS22 | 9.78E-140 | 0.44816291 | 0.914 | 0.577 | 3.74E-135 | ExN5 | CADPS2 |
| GALNT14 | 2.76E-138 | 0.28706909 | 0.626 | 0.321 | 1.06E-133 | ExN5 | GALNT14 |
| LINC022172 | 1.70E-136 | 0.26336448 | 0.489 | 0.219 | 6.52E-132 | ExN5 | LINC02217 |
| KCNMB41 | 1.73E-136 | 0.43514946 | 0.737 | 0.458 | 6.60E-132 | ExN5 | KCNMB4 |
| AL391840.13 | 9.18E-136 | 0.46167972 | 0.82 | 0.502 | 3.51E-131 | ExN5 | AL391840.1 |
| SEC16B1 | 1.02E-135 | 0.37011501 | 0.43 | 0.198 | 3.88E-131 | ExN5 | SEC16B |
| VAV31 | 2.80E-135 | 0.44514453 | 0.349 | 0.141 | 1.07E-130 | ExN5 | VAV3 |
| OTOGL1 | 1.84E-134 | 0.34359862 | 0.422 | 0.187 | 7.02E-130 | ExN5 | OTOGL |
| ANKS1B2 | 3.08E-134 | 0.33722413 | 0.992 | 0.986 | 1.18E-129 | ExN5 | ANKS1B |
| GABRA5 | 3.80E-134 | 0.27718329 | 0.492 | 0.231 | 1.45E-129 | ExN5 | GABRA5 |
| IPCEF12 | 5.33E-134 | 0.39874653 | 0.692 | 0.392 | 2.04E-129 | ExN5 | IPCEF1 |
| ITPK12 | 7.13E-134 | 0.36838361 | 0.752 | 0.456 | 2.72E-129 | ExN5 | ITPK1 |
| IGSF21 | 8.31E-133 | 0.49402645 | 0.783 | 0.524 | 3.18E-128 | ExN5 | IGSF21 |
| PRKD1 | 1.01E-131 | 0.44415862 | 0.773 | 0.502 | 3.84E-127 | ExN5 | PRKD1 |
| RASA2 | 1.51E-130 | 0.43178287 | 0.849 | 0.65 | 5.76E-126 | ExN5 | RASA2 |
| RUNX1T1 | 1.70E-130 | 0.41724192 | 0.907 | 0.74 | 6.51E-126 | ExN5 | RUNX1T1 |
| KCNQ32 | 2.44E-130 | 0.48352789 | 0.974 | 0.899 | 9.32E-126 | ExN5 | KCNQ3 |
| LINC02223 | 2.94E-129 | 0.38485744 | 0.558 | 0.287 | 1.13E-124 | ExN5 | LINC02223 |
| MLIP3 | 4.13E-129 | 0.50900947 | 0.815 | 0.505 | 1.58E-124 | ExN5 | MLIP |
| MCHR22 | 3.41E-128 | 0.30697164 | 0.543 | 0.266 | 1.30E-123 | ExN5 | MCHR2 |
| AC034268.22 | 2.34E-127 | 0.63836355 | 0.544 | 0.281 | 8.96E-123 | ExN5 | AC034268.2 |
| NMNAT31 | 1.53E-126 | 0.27938332 | 0.504 | 0.242 | 5.84E-122 | ExN5 | NMNAT3 |
| CDH122 | 3.76E-126 | 0.4593967 | 0.961 | 0.684 | 1.44E-121 | ExN5 | CDH12 |
| AC124852.13 | 9.51E-126 | 0.42318822 | 0.767 | 0.468 | 3.64E-121 | ExN5 | AC124852.1 |
| CDH101 | 9.96E-126 | 0.49096286 | 0.899 | 0.692 | 3.81E-121 | ExN5 | CDH10 |
| CREG22 | 1.42E-124 | 0.43148561 | 0.77 | 0.505 | 5.42E-120 | ExN5 | CREG2 |
| FSTL44 | 1.74E-124 | 0.46652171 | 0.932 | 0.64 | 6.65E-120 | ExN5 | FSTL4 |
| ESRRG | 1.48E-123 | 0.5009925 | 0.88 | 0.677 | 5.66E-119 | ExN5 | ESRRG |
| MAP1B1 | 4.34E-122 | 0.4103013 | 0.959 | 0.896 | 1.66E-117 | ExN5 | MAP1B |
| DCC2 | 1.81E-121 | 0.79004624 | 0.742 | 0.539 | 6.91E-117 | ExN5 | DCC |
| LRRN31 | 4.62E-121 | 0.30572158 | 0.608 | 0.327 | 1.76E-116 | ExN5 | LRRN3 |
| AQP4-AS13 | 5.56E-121 | 0.46575251 | 0.786 | 0.493 | 2.13E-116 | ExN5 | AQP4-AS1 |

| | | | | | | | |
|---|---|---|---|---|---|---|---|
| AC005906.21 | 1.68E-120 | 0.26904108 | 0.568 | 0.286 | 6.42E-116 | ExN5 | AC005906.2 |
| ARNTL21 | 1.83E-119 | 0.30970817 | 0.657 | 0.369 | 6.98E-115 | ExN5 | ARNTL2 |
| ARPP213 | 2.68E-119 | 0.44485551 | 0.979 | 0.76 | 1.02E-114 | ExN5 | ARPP21 |
| GRIK4 | 3.54E-119 | 0.39784581 | 0.716 | 0.435 | 1.35E-114 | ExN5 | GRIK4 |
| AL133166.12 | 4.58E-118 | 0.28748447 | 0.452 | 0.214 | 1.75E-113 | ExN5 | AL133166.1 |
| AL355472.21 | 5.02E-118 | 0.28396201 | 0.522 | 0.266 | 1.92E-113 | ExN5 | AL355472.2 |
| SLC35F12 | 5.24E-118 | 0.42574269 | 0.933 | 0.701 | 2.00E-113 | ExN5 | SLC35F1 |
| GPM6B2 | 2.99E-117 | 0.39287468 | 0.935 | 0.799 | 1.14E-112 | ExN5 | GPM6B |
| SLC24A22 | 1.93E-116 | 0.39272987 | 0.952 | 0.83 | 7.37E-112 | ExN5 | SLC24A2 |
| RGS4 | 4.64E-116 | 0.2919619 | 0.527 | 0.275 | 1.77E-111 | ExN5 | RGS4 |
| DKK2 | 1.12E-115 | 0.25202815 | 0.284 | 0.107 | 4.28E-111 | ExN5 | DKK2 |
| WDPCP | 1.52E-115 | 0.42154828 | 0.912 | 0.758 | 5.79E-111 | ExN5 | WDPCP |
| SNTB21 | 2.16E-115 | 0.43485315 | 0.7 | 0.438 | 8.27E-111 | ExN5 | SNTB2 |
| MAGI31 | 4.64E-115 | 0.38843266 | 0.883 | 0.681 | 1.77E-110 | ExN5 | MAGI3 |
| AL158077.21 | 1.11E-114 | 0.29872651 | 0.527 | 0.268 | 4.25E-110 | ExN5 | AL158077.2 |
| ATP2B11 | 2.42E-114 | 0.36921743 | 0.955 | 0.845 | 9.27E-110 | ExN5 | ATP2B1 |
| FAM81A1 | 5.22E-114 | 0.39039335 | 0.748 | 0.48 | 2.00E-109 | ExN5 | FAM81A |
| AC007368.11 | 5.29E-114 | 0.70163547 | 0.652 | 0.435 | 2.02E-109 | ExN5 | AC007368.1 |
| KCNK101 | 6.49E-114 | 0.31451164 | 0.586 | 0.319 | 2.48E-109 | ExN5 | KCNK10 |
| COBL2 | 1.28E-113 | 0.39702759 | 0.923 | 0.744 | 4.88E-109 | ExN5 | COBL |
| UGP2 | 3.88E-112 | 0.330361 | 0.662 | 0.4 | 1.48E-107 | ExN5 | UGP2 |
| DNAJC12 | 6.99E-112 | 0.42157212 | 0.797 | 0.552 | 2.67E-107 | ExN5 | DNAJC1 |
| TM7SF31 | 3.84E-110 | 0.3478626 | 0.666 | 0.41 | 1.47E-105 | ExN5 | TM7SF3 |
| SYNJ21 | 1.01E-109 | 0.29349431 | 0.671 | 0.384 | 3.85E-105 | ExN5 | SYNJ2 |
| PRKAG22 | 2.21E-109 | 0.40993412 | 0.91 | 0.722 | 8.45E-105 | ExN5 | PRKAG2 |
| KCNIP44 | 1.26E-108 | 0.43299687 | 0.999 | 0.936 | 4.81E-104 | ExN5 | KCNIP4 |
| PLOD21 | 2.91E-108 | 0.33314164 | 0.434 | 0.212 | 1.11E-103 | ExN5 | PLOD2 |
| LMO31 | 2.19E-107 | 0.38675862 | 0.847 | 0.587 | 8.39E-103 | ExN5 | LMO3 |
| ADD21 | 3.21E-107 | 0.39955446 | 0.866 | 0.671 | 1.23E-102 | ExN5 | ADD2 |
| NWD23 | 3.65E-107 | 0.34359122 | 0.757 | 0.458 | 1.39E-102 | ExN5 | NWD2 |
| NRGN3 | 4.44E-107 | 0.35640329 | 0.874 | 0.564 | 1.70E-102 | ExN5 | NRGN |
| DIRAS21 | 6.44E-107 | 0.25767663 | 0.572 | 0.308 | 2.46E-102 | ExN5 | DIRAS2 |
| SLC39A102 | 6.63E-107 | 0.41158185 | 0.842 | 0.638 | 2.53E-102 | ExN5 | SLC39A10 |
| MIR4500HG1 | 1.03E-106 | 0.34689176 | 0.541 | 0.295 | 3.93E-102 | ExN5 | MIR4500HG |
| NEO11 | 5.61E-106 | 0.40476839 | 0.824 | 0.586 | 2.14E-101 | ExN5 | NEO1 |
| ANKRD33B2 | 6.53E-106 | 0.29826026 | 0.715 | 0.427 | 2.50E-101 | ExN5 | ANKRD33B |
| TMEFF21 | 7.86E-106 | 0.43741344 | 0.893 | 0.76 | 3.00E-101 | ExN5 | TMEFF2 |
| DGKI1 | 9.49E-106 | 0.3745895 | 0.957 | 0.891 | 3.63E-101 | ExN5 | DGKI |
| NLK2 | 1.71E-105 | 0.40343242 | 0.906 | 0.691 | 6.54E-101 | ExN5 | NLK |
| DMD3 | 1.77E-105 | 0.52539379 | 0.971 | 0.92 | 6.76E-101 | ExN5 | DMD |
| AC006504.52 | 3.85E-105 | 0.39864985 | 0.678 | 0.423 | 1.47E-100 | ExN5 | AC006504.5 |
| AC011474.1 | 1.90E-103 | 0.27081473 | 0.54 | 0.285 | 7.26E-99 | ExN5 | AC011474.1 |
| DNAH61 | 2.29E-103 | 0.33873357 | 0.809 | 0.524 | 8.74E-99 | ExN5 | DNAH6 |
| AK52 | 6.24E-103 | 0.53938327 | 0.892 | 0.778 | 2.39E-98 | ExN5 | AK5 |
| DLGAP1-AS4 | 2.43E-102 | 0.25278527 | 0.62 | 0.346 | 9.28E-98 | ExN5 | DLGAP1-AS4 |
| RASAL21 | 4.56E-102 | 0.35618882 | 0.949 | 0.875 | 1.74E-97 | ExN5 | RASAL2 |
| ST6GAL22 | 5.60E-102 | 0.26732251 | 0.656 | 0.382 | 2.14E-97 | ExN5 | ST6GAL2 |
| GAB12 | 1.46E-101 | 0.3156048 | 0.821 | 0.539 | 5.58E-97 | ExN5 | GAB1 |
| FOCAD2 | 4.45E-101 | 0.37507385 | 0.925 | 0.798 | 1.70E-96 | ExN5 | FOCAD |
| SMAP11 | 7.43E-101 | 0.3930082 | 0.826 | 0.62 | 2.84E-96 | ExN5 | SMAP1 |
| CCDC85A1 | 1.93E-100 | 0.41540515 | 0.88 | 0.649 | 7.38E-96 | ExN5 | CCDC85A |
| NELL11 | 4.12E-100 | 0.44218965 | 0.86 | 0.647 | 1.57E-95 | ExN5 | NELL1 |
| RAP1GAP23 | 4.52E-100 | 0.35700936 | 0.865 | 0.63 | 1.73E-95 | ExN5 | RAP1GAP2 |
| LINC00894 | 1.29E-99 | 0.3492459 | 0.647 | 0.394 | 4.95E-95 | ExN5 | LINC00894 |
| VPS13A1 | 2.69E-99 | 0.35541784 | 0.893 | 0.729 | 1.03E-94 | ExN5 | VPS13A |
| ATP2B42 | 4.47E-99 | 0.32173644 | 0.796 | 0.508 | 1.71E-94 | ExN5 | ATP2B4 |
| MEIS31 | 3.83E-98 | 0.28825915 | 0.537 | 0.295 | 1.46E-93 | ExN5 | MEIS3 |
| SH3BGRL21 | 7.78E-98 | 0.35267456 | 0.779 | 0.528 | 2.97E-93 | ExN5 | SH3BGRL2 |
| AL157944.12 | 1.31E-97 | 0.42693226 | 0.513 | 0.27 | 5.02E-93 | ExN5 | AL157944.1 |
| MAPK102 | 2.17E-97 | 0.28840757 | 0.977 | 0.952 | 8.28E-93 | ExN5 | MAPK10 |
| SEC14L52 | 2.33E-97 | 0.2777424 | 0.722 | 0.442 | 8.90E-93 | ExN5 | SEC14L5 |
| CARMIL12 | 3.43E-97 | 0.36287363 | 0.861 | 0.613 | 1.31E-92 | ExN5 | CARMIL1 |
| DCLK11 | 1.39E-96 | 0.30697857 | 0.976 | 0.94 | 5.33E-92 | ExN5 | DCLK1 |

| | | | | | | | |
|---|---|---|---|---|---|---|---|
| THRB2 | 2.21E-96 | 0.36003755 | 0.968 | 0.915 | 8.45E-92 | ExN5 | THRB |
| BASP1-AS13 | 2.96E-96 | 0.38043124 | 0.884 | 0.651 | 1.13E-91 | ExN5 | BASP1-AS1 |
| ZBBX1 | 5.78E-96 | 0.36522018 | 0.431 | 0.225 | 2.21E-91 | ExN5 | ZBBX |
| DDAH12 | 3.46E-95 | 0.3527303 | 0.668 | 0.416 | 1.32E-90 | ExN5 | DDAH1 |
| OXR12 | 3.66E-95 | 0.38664936 | 0.974 | 0.92 | 1.40E-90 | ExN5 | OXR1 |
| KCNIP4-IT14 | 3.96E-95 | 0.29965662 | 0.788 | 0.492 | 1.52E-90 | ExN5 | KCNIP4-IT1 |
| PGBD5 | 4.33E-95 | 0.36418482 | 0.867 | 0.681 | 1.66E-90 | ExN5 | PGBD5 |
| CNTN11 | 4.69E-95 | 0.32328746 | 0.973 | 0.949 | 1.79E-90 | ExN5 | CNTN1 |
| SYN32 | 6.78E-95 | 0.45190661 | 0.965 | 0.862 | 2.59E-90 | ExN5 | SYN3 |
| CHN13 | 1.51E-94 | 0.35533212 | 0.943 | 0.745 | 5.78E-90 | ExN5 | CHN1 |
| LDB24 | 4.07E-94 | 0.40048259 | 0.974 | 0.758 | 1.55E-89 | ExN5 | LDB2 |
| SORL12 | 6.02E-94 | 0.34178503 | 0.879 | 0.672 | 2.30E-89 | ExN5 | SORL1 |
| CCND31 | 6.07E-93 | 0.27258941 | 0.637 | 0.38 | 2.32E-88 | ExN5 | CCND3 |
| CACNA2D32 | 7.85E-93 | 0.38692178 | 0.972 | 0.864 | 3.00E-88 | ExN5 | CACNA2D3 |
| ANKMY2 | 8.71E-93 | 0.28528994 | 0.628 | 0.385 | 3.33E-88 | ExN5 | ANKMY2 |
| SULF2 | 1.20E-92 | 0.27475815 | 0.292 | 0.128 | 4.57E-88 | ExN5 | SULF2 |
| HOMER12 | 4.93E-92 | 0.36343885 | 0.831 | 0.615 | 1.89E-87 | ExN5 | HOMER1 |
| CASC152 | 9.84E-92 | 0.73810381 | 0.736 | 0.515 | 3.76E-87 | ExN5 | CASC15 |
| LGI42 | 6.77E-91 | 0.25361948 | 0.545 | 0.3 | 2.59E-86 | ExN5 | LGI4 |
| GTDC11 | 6.87E-91 | 0.37892331 | 0.924 | 0.788 | 2.63E-86 | ExN5 | GTDC1 |
| GFOD11 | 2.26E-90 | 0.34083503 | 0.908 | 0.75 | 8.65E-86 | ExN5 | GFOD1 |
| PDP11 | 6.20E-90 | 0.2589509 | 0.556 | 0.318 | 2.37E-85 | ExN5 | PDP1 |
| RAPGEF23 | 1.02E-89 | 0.34379723 | 0.942 | 0.855 | 3.91E-85 | ExN5 | RAPGEF2 |
| NUAK12 | 1.23E-89 | 0.33778512 | 0.823 | 0.588 | 4.71E-85 | ExN5 | NUAK1 |
| GPATCH81 | 1.70E-88 | 0.33331299 | 0.942 | 0.832 | 6.49E-84 | ExN5 | GPATCH8 |
| UBE2E23 | 2.04E-88 | 0.33872249 | 0.955 | 0.875 | 7.81E-84 | ExN5 | UBE2E2 |
| CNIH31 | 2.93E-88 | 0.27692311 | 0.661 | 0.407 | 1.12E-83 | ExN5 | CNIH3 |
| GBE1 | 3.73E-88 | 0.28729202 | 0.443 | 0.236 | 1.42E-83 | ExN5 | GBE1 |
| FBXW72 | 6.84E-88 | 0.36103006 | 0.911 | 0.752 | 2.61E-83 | ExN5 | FBXW7 |
| CPE | 1.35E-87 | 0.33296134 | 0.862 | 0.673 | 5.17E-83 | ExN5 | CPE |
| ME1 | 2.61E-87 | 0.3185287 | 0.751 | 0.516 | 9.97E-83 | ExN5 | ME1 |
| C4orf50 | 7.94E-86 | 0.25995157 | 0.593 | 0.357 | 3.04E-81 | ExN5 | C4orf50 |
| AC016687.22 | 8.90E-86 | 0.64150789 | 0.322 | 0.151 | 3.40E-81 | ExN5 | AC016687.2 |
| CEMIP21 | 7.23E-85 | 0.27312096 | 0.582 | 0.345 | 2.77E-80 | ExN5 | CEMIP2 |
| TLL11 | 5.04E-84 | 0.28557988 | 0.291 | 0.128 | 1.93E-79 | ExN5 | TLL1 |
| TMEM2412 | 6.35E-84 | 0.29220361 | 0.756 | 0.491 | 2.43E-79 | ExN5 | TMEM241 |
| KHDRBS2-OT | 2.29E-83 | 0.30787706 | 0.7 | 0.444 | 8.76E-79 | ExN5 | KHDRBS2-OT |
| SYN21 | 3.33E-83 | 0.28117916 | 0.965 | 0.897 | 1.27E-78 | ExN5 | SYN2 |
| SLC4A101 | 1.02E-82 | 0.29412524 | 0.966 | 0.921 | 3.89E-78 | ExN5 | SLC4A10 |
| NOS1AP1 | 2.57E-82 | 0.3461282 | 0.844 | 0.657 | 9.81E-78 | ExN5 | NOS1AP |
| PCDH101 | 2.73E-82 | 0.32720869 | 0.687 | 0.443 | 1.04E-77 | ExN5 | PCDH10 |
| RORB3 | 3.40E-82 | 0.36502381 | 0.575 | 0.326 | 1.30E-77 | ExN5 | RORB |
| AC093523.12 | 9.54E-82 | 0.32027257 | 0.653 | 0.411 | 3.65E-77 | ExN5 | AC093523.1 |
| HTR1E | 3.39E-81 | 0.32666487 | 0.809 | 0.57 | 1.30E-76 | ExN5 | HTR1E |
| GLS1 | 3.52E-81 | 0.31841789 | 0.92 | 0.824 | 1.35E-76 | ExN5 | GLS |
| AMPH2 | 2.12E-80 | 0.35183788 | 0.921 | 0.809 | 8.09E-76 | ExN5 | AMPH |
| MBNL23 | 3.21E-80 | 0.31059511 | 0.939 | 0.76 | 1.23E-75 | ExN5 | MBNL2 |
| NETO11 | 1.31E-79 | 0.32326965 | 0.896 | 0.725 | 5.01E-75 | ExN5 | NETO1 |
| LINC017251 | 2.51E-79 | 0.42262717 | 0.575 | 0.35 | 9.59E-75 | ExN5 | LINC01725 |
| TMSB101 | 1.85E-78 | 0.31826249 | 0.549 | 0.327 | 7.08E-74 | ExN5 | TMSB10 |
| RFTN12 | 1.67E-77 | 0.25754298 | 0.749 | 0.472 | 6.40E-73 | ExN5 | RFTN1 |
| KCNK11 | 1.91E-77 | 0.27170572 | 0.769 | 0.515 | 7.28E-73 | ExN5 | KCNK1 |
| AL591501.12 | 3.05E-77 | 0.32945714 | 0.813 | 0.564 | 1.16E-72 | ExN5 | AL591501.1 |
| SLC38A6 | 1.01E-76 | 0.30442177 | 0.754 | 0.528 | 3.87E-72 | ExN5 | SLC38A6 |
| LINGO13 | 2.06E-76 | 0.25197002 | 0.755 | 0.496 | 7.87E-72 | ExN5 | LINGO1 |
| CAMK2A3 | 2.17E-76 | 0.29180523 | 0.955 | 0.791 | 8.31E-72 | ExN5 | CAMK2A |
| ZNF385B2 | 2.96E-76 | 0.33053796 | 0.943 | 0.817 | 1.13E-71 | ExN5 | ZNF385B |
| HECW13 | 5.76E-76 | 0.31811399 | 0.949 | 0.829 | 2.20E-71 | ExN5 | HECW1 |
| AL008633.13 | 1.81E-75 | 0.40858592 | 0.894 | 0.71 | 6.91E-71 | ExN5 | AL008633.1 |
| DSCAM1 | 4.29E-75 | 0.29246473 | 0.984 | 0.902 | 1.64E-70 | ExN5 | DSCAM |
| COBLL11 | 2.49E-74 | 0.35636832 | 0.328 | 0.165 | 9.51E-70 | ExN5 | COBLL1 |
| SYT73 | 1.74E-73 | 0.26621554 | 0.786 | 0.525 | 6.64E-69 | ExN5 | SYT7 |
| MICAL21 | 5.29E-73 | 0.3023043 | 0.9 | 0.728 | 2.02E-68 | ExN5 | MICAL2 |

| | | | | | | | |
|---|---|---|---|---|---|---|---|
| MSRA3 | 6.25E-73 | 0.28625932 | 0.968 | 0.906 | 2.39E-68 | ExN5 | MSRA |
| STX1A3 | 1.05E-72 | 0.25871489 | 0.595 | 0.369 | 3.99E-68 | ExN5 | STX1A |
| PACRG | 1.14E-72 | 0.33404967 | 0.882 | 0.702 | 4.35E-68 | ExN5 | PACRG |
| KCNH13 | 1.79E-72 | 0.36472519 | 0.951 | 0.828 | 6.84E-68 | ExN5 | KCNH1 |
| PPP1R12A2 | 7.16E-72 | 0.32894361 | 0.88 | 0.726 | 2.74E-67 | ExN5 | PPP1R12A |
| AC092691.11 | 1.75E-71 | 0.35488073 | 0.945 | 0.866 | 6.69E-67 | ExN5 | AC092691.1 |
| NIN2 | 2.71E-71 | 0.3102323 | 0.886 | 0.705 | 1.04E-66 | ExN5 | NIN |
| DLGAP22 | 1.02E-70 | 0.2830854 | 0.984 | 0.929 | 3.92E-66 | ExN5 | DLGAP2 |
| ARHGAP262 | 1.12E-70 | 0.34301029 | 0.951 | 0.87 | 4.27E-66 | ExN5 | ARHGAP26 |
| SEPTIN11 | 2.56E-70 | 0.31057866 | 0.856 | 0.673 | 9.80E-66 | ExN5 | SEPTIN11 |
| AL117329.13 | 1.23E-69 | 0.4228739 | 0.691 | 0.466 | 4.69E-65 | ExN5 | AL117329.1 |
| KIAA09302 | 3.56E-69 | 0.29039444 | 0.82 | 0.606 | 1.36E-64 | ExN5 | KIAA0930 |
| CXXC4-AS12 | 3.95E-69 | 0.31981956 | 0.597 | 0.38 | 1.51E-64 | ExN5 | CXXC4-AS1 |
| KCNMA11 | 4.57E-69 | 0.27540954 | 0.984 | 0.958 | 1.75E-64 | ExN5 | KCNMA1 |
| ANKRD10 | 1.21E-68 | 0.25753678 | 0.722 | 0.506 | 4.61E-64 | ExN5 | ANKRD10 |
| MNAT1 | 2.91E-68 | 0.29834084 | 0.904 | 0.786 | 1.11E-63 | ExN5 | MNAT1 |
| PTK2B2 | 5.29E-68 | 0.33545932 | 0.731 | 0.515 | 2.02E-63 | ExN5 | PTK2B |
| AC007563.22 | 5.69E-68 | 0.32932891 | 0.829 | 0.603 | 2.17E-63 | ExN5 | AC007563.2 |
| STXBP5L3 | 1.32E-67 | 0.28006508 | 0.976 | 0.947 | 5.05E-63 | ExN5 | STXBP5L |
| APBA2 | 2.26E-67 | 0.29952825 | 0.887 | 0.739 | 8.63E-63 | ExN5 | APBA2 |
| CCDC6 | 2.68E-67 | 0.27997831 | 0.771 | 0.559 | 1.02E-62 | ExN5 | CCDC6 |
| FNIP21 | 4.26E-67 | 0.28362498 | 0.808 | 0.608 | 1.63E-62 | ExN5 | FNIP2 |
| KCTD12 | 8.79E-67 | 0.29932151 | 0.883 | 0.666 | 3.36E-62 | ExN5 | KCTD1 |
| LINC002991 | 9.51E-67 | 0.26066501 | 0.467 | 0.277 | 3.64E-62 | ExN5 | LINC00299 |
| DNAH72 | 2.48E-66 | 0.25094669 | 0.609 | 0.387 | 9.47E-62 | ExN5 | DNAH7 |
| AC021613.11 | 6.25E-66 | 0.29662696 | 0.471 | 0.278 | 2.39E-61 | ExN5 | AC021613.1 |
| LINC023891 | 2.16E-65 | 0.2775574 | 0.691 | 0.467 | 8.28E-61 | ExN5 | LINC02389 |
| AGBL43 | 3.95E-65 | 0.28816821 | 0.98 | 0.932 | 1.51E-60 | ExN5 | AGBL4 |
| FMN2 | 9.05E-65 | 0.27020113 | 0.957 | 0.899 | 3.46E-60 | ExN5 | FMN2 |
| PRKCB4 | 3.04E-64 | 0.2956248 | 0.97 | 0.865 | 1.16E-59 | ExN5 | PRKCB |
| WASF11 | 5.10E-64 | 0.27474679 | 0.834 | 0.636 | 1.95E-59 | ExN5 | WASF1 |
| PI4KA2 | 5.43E-64 | 0.29191744 | 0.924 | 0.813 | 2.08E-59 | ExN5 | PI4KA |
| EXOC61 | 8.44E-64 | 0.28840559 | 0.824 | 0.635 | 3.22E-59 | ExN5 | EXOC6 |
| ADAM231 | 3.24E-63 | 0.27250992 | 0.909 | 0.765 | 1.24E-58 | ExN5 | ADAM23 |
| MYH101 | 5.65E-63 | 0.28357076 | 0.88 | 0.721 | 2.16E-58 | ExN5 | MYH10 |
| AC073091.33 | 6.93E-63 | 0.39638759 | 0.503 | 0.311 | 2.65E-58 | ExN5 | AC073091.3 |
| KCNB22 | 7.62E-63 | 0.29362032 | 0.879 | 0.681 | 2.91E-58 | ExN5 | KCNB2 |
| LINC014142 | 7.73E-63 | 0.2646944 | 0.681 | 0.451 | 2.96E-58 | ExN5 | LINC01414 |
| TBC1D11 | 1.81E-62 | 0.25620701 | 0.784 | 0.567 | 6.92E-58 | ExN5 | TBC1D1 |
| MEG32 | 2.54E-62 | 0.3459454 | 0.984 | 0.959 | 9.71E-58 | ExN5 | MEG3 |
| PARP8 | 4.30E-62 | 0.30649557 | 0.823 | 0.647 | 1.64E-57 | ExN5 | PARP8 |
| AC106793.11 | 1.69E-61 | 0.2871987 | 0.575 | 0.369 | 6.47E-57 | ExN5 | AC106793.1 |
| ATP8A11 | 1.70E-61 | 0.26368467 | 0.931 | 0.849 | 6.48E-57 | ExN5 | ATP8A1 |
| AL513166.13 | 3.88E-60 | 0.25848686 | 0.843 | 0.616 | 1.48E-55 | ExN5 | AL513166.1 |
| DIRC33 | 1.17E-59 | 0.29778241 | 0.744 | 0.538 | 4.48E-55 | ExN5 | DIRC3 |
| VPS13D1 | 5.38E-59 | 0.3000402 | 0.875 | 0.748 | 2.06E-54 | ExN5 | VPS13D |
| LRFN54 | 1.09E-58 | 0.27630436 | 0.966 | 0.89 | 4.18E-54 | ExN5 | LRFN5 |
| CNKSR24 | 1.10E-58 | 0.29631788 | 0.94 | 0.817 | 4.21E-54 | ExN5 | CNKSR2 |
| ARHGEF3 | 1.10E-58 | 0.26149332 | 0.8 | 0.601 | 4.21E-54 | ExN5 | ARHGEF3 |
| LINC012502 | 2.57E-58 | 0.27797922 | 0.84 | 0.639 | 9.82E-54 | ExN5 | LINC01250 |
| PRKN | 4.91E-58 | 0.28392747 | 0.933 | 0.86 | 1.88E-53 | ExN5 | PRKN |
| SSBP32 | 1.08E-57 | 0.25435644 | 0.86 | 0.671 | 4.12E-53 | ExN5 | SSBP3 |
| CDH2 | 3.01E-57 | 0.31270998 | 0.879 | 0.752 | 1.15E-52 | ExN5 | CDH2 |
| PTCHD1-AS1 | 3.81E-57 | 0.2604818 | 0.668 | 0.445 | 1.46E-52 | ExN5 | PTCHD1-AS |
| ZEB13 | 2.43E-56 | 0.272895 | 0.891 | 0.76 | 9.30E-52 | ExN5 | ZEB1 |
| SLIT32 | 9.32E-55 | 0.56330426 | 0.582 | 0.468 | 3.56E-50 | ExN5 | SLIT3 |
| MIR646HG | 3.92E-54 | 0.26930392 | 0.448 | 0.272 | 1.50E-49 | ExN5 | MIR646HG |
| TMEM132D2 | 4.28E-54 | 0.31377862 | 0.902 | 0.779 | 1.63E-49 | ExN5 | TMEM132D |
| GLDN1 | 5.45E-54 | 0.29502215 | 0.382 | 0.232 | 2.08E-49 | ExN5 | GLDN |
| PDE1A3 | 2.33E-53 | 0.25763306 | 0.924 | 0.674 | 8.90E-49 | ExN5 | PDE1A |
| FARS21 | 2.40E-53 | 0.26582635 | 0.894 | 0.758 | 9.17E-49 | ExN5 | FARS2 |
| DYNC1I12 | 5.13E-53 | 0.25224507 | 0.943 | 0.87 | 1.96E-48 | ExN5 | DYNC1I1 |
| CELF23 | 7.38E-53 | 0.25569449 | 0.991 | 0.967 | 2.82E-48 | ExN5 | CELF2 |

| | | | | | | | |
|---|---|---|---|---|---|---|---|
| KCNH72 | 7.42E-53 | 0.25249114 | 0.894 | 0.725 | 2.84E-48 | ExN5 | KCNH7 |
| CNTN4 | 1.58E-52 | 0.26701275 | 0.939 | 0.848 | 6.06E-48 | ExN5 | CNTN4 |
| CA102 | 1.92E-51 | 0.30545595 | 0.625 | 0.436 | 7.35E-47 | ExN5 | CA10 |
| RAPGEF53 | 6.37E-49 | 0.28537159 | 0.921 | 0.746 | 2.43E-44 | ExN5 | RAPGEF5 |
| AC024901.11 | 1.07E-46 | 0.26258577 | 0.645 | 0.448 | 4.11E-42 | ExN5 | AC024901.1 |
| AFF31 | 6.60E-45 | 0.25087655 | 0.956 | 0.9 | 2.52E-40 | ExN5 | AFF3 |
| ERC21 | 3.85E-44 | 0.30763957 | 0.967 | 0.93 | 1.47E-39 | ExN5 | ERC2 |
| AC011287.12 | 2.27E-43 | 0.34918873 | 0.639 | 0.469 | 8.69E-39 | ExN5 | AC011287.1 |
| DGKG1 | 1.26E-36 | 0.35499449 | 0.64 | 0.534 | 4.81E-32 | ExN5 | DGKG |
| USP9Y | 2.10E-31 | 0.2873771 | 0.476 | 0.349 | 8.02E-27 | ExN5 | USP9Y |
| NLGN4Y | 5.75E-25 | 0.25936928 | 0.481 | 0.367 | 2.20E-20 | ExN5 | NLGN4Y |
| KCNH52 | 3.09E-20 | 0.4422419 | 0.476 | 0.419 | 1.18E-15 | ExN5 | KCNH5 |
| EPHA32 | 6.62E-12 | 0.33434724 | 0.484 | 0.423 | 2.53E-07 | ExN5 | EPHA3 |
| ADAMTSL1 | 0 | 2.50894094 | 0.621 | 0.151 | 0 | ExN6 | ADAMTSL1 |
| HS3ST41 | 0 | 2.24365312 | 0.951 | 0.388 | 0 | ExN6 | HS3ST4 |
| HTR2C | 0 | 2.13102373 | 0.315 | 0.062 | 0 | ExN6 | HTR2C |
| DLC1 | 0 | 2.0437488 | 0.783 | 0.33 | 0 | ExN6 | DLC1 |
| SEMA3E1 | 0 | 1.87163067 | 0.633 | 0.089 | 0 | ExN6 | SEMA3E |
| PDZRN43 | 0 | 1.71293367 | 0.866 | 0.363 | 0 | ExN6 | PDZRN4 |
| TLE41 | 0 | 1.6225348 | 0.86 | 0.387 | 0 | ExN6 | TLE4 |
| CLSTN23 | 0 | 1.59677914 | 0.975 | 0.599 | 0 | ExN6 | CLSTN2 |
| AC068587.41 | 0 | 1.45366748 | 0.793 | 0.493 | 0 | ExN6 | AC068587.4 |
| KIAA1217 | 0 | 1.34655966 | 0.885 | 0.642 | 0 | ExN6 | KIAA1217 |
| ASIC2 | 0 | 1.33443 | 0.953 | 0.84 | 0 | ExN6 | ASIC2 |
| NRG12 | 0 | 1.28219648 | 0.833 | 0.527 | 0 | ExN6 | NRG1 |
| FOXP23 | 0 | 1.27241218 | 0.705 | 0.287 | 0 | ExN6 | FOXP2 |
| CADPS23 | 0 | 1.20698005 | 0.798 | 0.575 | 0 | ExN6 | CADPS2 |
| FRAS1 | 0 | 1.20106263 | 0.846 | 0.436 | 0 | ExN6 | FRAS1 |
| DPP102 | 0 | 1.14940951 | 0.995 | 0.838 | 0 | ExN6 | DPP10 |
| CDH61 | 0 | 1.13334407 | 0.664 | 0.153 | 0 | ExN6 | CDH6 |
| GHR1 | 0 | 1.13056637 | 0.637 | 0.366 | 0 | ExN6 | GHR |
| AC090578.13 | 0 | 1.11655655 | 0.855 | 0.541 | 0 | ExN6 | AC090578.1 |
| LRP1B2 | 0 | 1.09485363 | 0.991 | 0.947 | 0 | ExN6 | LRP1B |
| GRM33 | 0 | 1.07824636 | 0.642 | 0.383 | 0 | ExN6 | GRM3 |
| ZFHX3 | 0 | 1.07786368 | 0.567 | 0.273 | 0 | ExN6 | ZFHX3 |
| ERC22 | 0 | 1.07285689 | 0.982 | 0.927 | 0 | ExN6 | ERC2 |
| MDGA21 | 0 | 1.05683616 | 0.965 | 0.937 | 0 | ExN6 | MDGA2 |
| AK53 | 0 | 1.02600264 | 0.952 | 0.769 | 0 | ExN6 | AK5 |
| MEIS21 | 0 | 0.99011644 | 0.64 | 0.3 | 0 | ExN6 | MEIS2 |
| SORBS21 | 0 | 0.94721594 | 0.957 | 0.836 | 0 | ExN6 | SORBS2 |
| TLL12 | 0 | 0.9196802 | 0.367 | 0.116 | 0 | ExN6 | TLL1 |
| CDH11 | 0 | 0.91712496 | 0.721 | 0.434 | 0 | ExN6 | CDH11 |
| CADPS | 0 | 0.9074388 | 0.985 | 0.935 | 0 | ExN6 | CADPS |
| PDE1A4 | 0 | 0.90468254 | 0.839 | 0.672 | 0 | ExN6 | PDE1A |
| SGCD2 | 0 | 0.89382075 | 0.874 | 0.571 | 0 | ExN6 | SGCD |
| BMPER3 | 0 | 0.88088106 | 0.875 | 0.599 | 0 | ExN6 | BMPER |
| ZNF385B3 | 0 | 0.87362046 | 0.926 | 0.814 | 0 | ExN6 | ZNF385B |
| DOCK42 | 0 | 0.86307454 | 0.951 | 0.818 | 0 | ExN6 | DOCK4 |
| SOX53 | 0 | 0.84544181 | 0.927 | 0.652 | 0 | ExN6 | SOX5 |
| MMP163 | 0 | 0.82472288 | 0.935 | 0.79 | 0 | ExN6 | MMP16 |
| DIRAS22 | 0 | 0.8066156 | 0.601 | 0.297 | 0 | ExN6 | DIRAS2 |
| DPY19L12 | 0 | 0.80363871 | 0.648 | 0.333 | 0 | ExN6 | DPY19L1 |
| MCTP2 | 0 | 0.8000109 | 0.311 | 0.036 | 0 | ExN6 | MCTP2 |
| DACH12 | 0 | 0.79061127 | 0.519 | 0.183 | 0 | ExN6 | DACH1 |
| CHRM2 | 0 | 0.7848932 | 0.468 | 0.199 | 0 | ExN6 | CHRM2 |
| STARD131 | 0 | 0.78364398 | 0.82 | 0.52 | 0 | ExN6 | STARD13 |
| SV2B4 | 0 | 0.75922589 | 0.91 | 0.587 | 0 | ExN6 | SV2B |
| KLHL1 | 0 | 0.75844496 | 0.373 | 0.084 | 0 | ExN6 | KLHL1 |
| SEMA3A3 | 0 | 0.74998695 | 0.507 | 0.19 | 0 | ExN6 | SEMA3A |
| RASGRP11 | 0 | 0.74645959 | 0.564 | 0.164 | 0 | ExN6 | RASGRP1 |
| NAV33 | 0 | 0.74149633 | 0.991 | 0.947 | 0 | ExN6 | NAV3 |
| AC092902.41 | 0 | 0.71605792 | 0.574 | 0.201 | 0 | ExN6 | AC092902.4 |
| B3GLCT1 | 0 | 0.71517125 | 0.621 | 0.274 | 0 | ExN6 | B3GLCT |

| | | | | | | | |
|---|---|---|---|---|---|---|---|
| TMEM1631 | 0 | 0.69019243 | 0.587 | 0.268 | 0 | ExN6 | TMEM163 |
| LINC022173 | 0 | 0.68534031 | 0.51 | 0.208 | 0 | ExN6 | LINC02217 |
| ARPP214 | 0 | 0.68263041 | 0.97 | 0.753 | 0 | ExN6 | ARPP21 |
| LUZP2 | 0 | 0.67818687 | 0.658 | 0.285 | 0 | ExN6 | LUZP2 |
| SLC8A11 | 0 | 0.66176891 | 0.981 | 0.854 | 0 | ExN6 | SLC8A1 |
| RXFP12 | 0 | 0.65743719 | 0.529 | 0.207 | 0 | ExN6 | RXFP1 |
| SH3GL25 | 0 | 0.6567466 | 0.927 | 0.734 | 0 | ExN6 | SH3GL2 |
| RAPGEF6 | 0 | 0.63541748 | 0.875 | 0.681 | 0 | ExN6 | RAPGEF6 |
| PCDH93 | 0 | 0.63433798 | 0.998 | 0.965 | 0 | ExN6 | PCDH9 |
| HS3ST23 | 0 | 0.63430457 | 0.565 | 0.244 | 0 | ExN6 | HS3ST2 |
| DSCAML1 | 0 | 0.63361456 | 0.931 | 0.746 | 0 | ExN6 | DSCAML1 |
| DGKI2 | 0 | 0.61787427 | 0.945 | 0.889 | 0 | ExN6 | DGKI |
| SAMD3 | 0 | 0.61422795 | 0.279 | 0.063 | 0 | ExN6 | SAMD3 |
| ATP2C2 | 0 | 0.60877662 | 0.346 | 0.032 | 0 | ExN6 | ATP2C2 |
| GFRA1 | 0 | 0.59929397 | 0.313 | 0.063 | 0 | ExN6 | GFRA1 |
| NELL25 | 0 | 0.59876955 | 0.963 | 0.778 | 0 | ExN6 | NELL2 |
| PCDH17 | 0 | 0.59502184 | 0.615 | 0.31 | 0 | ExN6 | PCDH17 |
| ATP8B13 | 0 | 0.59205855 | 0.546 | 0.242 | 0 | ExN6 | ATP8B1 |
| TEX2 | 0 | 0.57988463 | 0.651 | 0.331 | 0 | ExN6 | TEX2 |
| AL138701.22 | 0 | 0.56441989 | 0.379 | 0.089 | 0 | ExN6 | AL138701.2 |
| CALCRL | 0 | 0.56161807 | 0.323 | 0.084 | 0 | ExN6 | CALCRL |
| DYSF | 0 | 0.56152231 | 0.539 | 0.195 | 0 | ExN6 | DYSF |
| PTPRG-AS11 | 0 | 0.54382114 | 0.414 | 0.14 | 0 | ExN6 | PTPRG-AS1 |
| DCLK12 | 0 | 0.54155342 | 0.976 | 0.938 | 0 | ExN6 | DCLK1 |
| BCL11B | 0 | 0.5229842 | 0.58 | 0.234 | 0 | ExN6 | BCL11B |
| DCHS21 | 0 | 0.52171368 | 0.385 | 0.107 | 0 | ExN6 | DCHS2 |
| CNTNAP3 | 0 | 0.51174992 | 0.486 | 0.125 | 0 | ExN6 | CNTNAP3 |
| NMNAT32 | 0 | 0.5018075 | 0.555 | 0.228 | 0 | ExN6 | NMNAT3 |
| GNB4 | 0 | 0.49912551 | 0.358 | 0.099 | 0 | ExN6 | GNB4 |
| ETV1 | 0 | 0.49863015 | 0.345 | 0.094 | 0 | ExN6 | ETV1 |
| MIR31HG2 | 0 | 0.49500074 | 0.481 | 0.191 | 0 | ExN6 | MIR31HG |
| CNTNAP3B | 0 | 0.48519736 | 0.508 | 0.219 | 0 | ExN6 | CNTNAP3B |
| B3GAT2 | 0 | 0.45431813 | 0.366 | 0.109 | 0 | ExN6 | B3GAT2 |
| LRMP2 | 0 | 0.42369135 | 0.398 | 0.134 | 0 | ExN6 | LRMP |
| LINC017481 | 0 | 0.4218816 | 0.365 | 0.108 | 0 | ExN6 | LINC01748 |
| STUM | 0 | 0.41420076 | 0.445 | 0.172 | 0 | ExN6 | STUM |
| SULF1 | 0 | 0.41166171 | 0.311 | 0.061 | 0 | ExN6 | SULF1 |
| RNF152 | 0 | 0.40304042 | 0.293 | 0.052 | 0 | ExN6 | RNF152 |
| ABCC9 | 0 | 0.3982821 | 0.286 | 0.054 | 0 | ExN6 | ABCC9 |
| TRIM22 | 0 | 0.38504243 | 0.282 | 0.041 | 0 | ExN6 | TRIM22 |
| AC107072.2 | 0 | 0.3647488 | 0.279 | 0.037 | 0 | ExN6 | AC107072.2 |
| SHB | 0 | 0.33086127 | 0.306 | 0.069 | 0 | ExN6 | SHB |
| ADTRP | 0 | 0.32971913 | 0.263 | 0.042 | 0 | ExN6 | ADTRP |
| UNC13B2 | 8.46E-307 | 0.55551392 | 0.666 | 0.362 | 3.23E-302 | ExN6 | UNC13B |
| RYR24 | 4.25E-303 | 0.61068896 | 0.983 | 0.924 | 1.62E-298 | ExN6 | RYR2 |
| AFDN | 1.59E-302 | 0.60266765 | 0.85 | 0.636 | 6.07E-298 | ExN6 | AFDN |
| ADAM232 | 4.44E-301 | 0.6640075 | 0.91 | 0.76 | 1.70E-296 | ExN6 | ADAM23 |
| ANKRD18A | 5.07E-301 | 0.33633051 | 0.348 | 0.118 | 1.94E-296 | ExN6 | ANKRD18A |
| RIT23 | 1.07E-299 | 0.85377113 | 0.601 | 0.305 | 4.09E-295 | ExN6 | RIT2 |
| RALYL4 | 1.54E-298 | 0.69337296 | 0.988 | 0.806 | 5.87E-294 | ExN6 | RALYL |
| MCTP12 | 1.84E-297 | 0.94516684 | 0.872 | 0.762 | 7.03E-293 | ExN6 | MCTP1 |
| AFF12 | 9.14E-297 | 0.53973727 | 0.699 | 0.393 | 3.49E-292 | ExN6 | AFF1 |
| TMEFF22 | 6.35E-293 | 0.95382403 | 0.89 | 0.755 | 2.43E-288 | ExN6 | TMEFF2 |
| ZNF804B2 | 1.72E-286 | 1.37681616 | 0.592 | 0.342 | 6.57E-282 | ExN6 | ZNF804B |
| HDAC41 | 2.99E-282 | 0.56374146 | 0.856 | 0.652 | 1.14E-277 | ExN6 | HDAC4 |
| LRMDA | 8.31E-282 | 1.62250146 | 0.393 | 0.176 | 3.17E-277 | ExN6 | LRMDA |
| SLC24A23 | 4.07E-281 | 0.5754458 | 0.957 | 0.826 | 1.56E-276 | ExN6 | SLC24A2 |
| RYR3 | 8.04E-281 | 1.09578729 | 0.603 | 0.375 | 3.07E-276 | ExN6 | RYR3 |
| DGKG2 | 2.48E-280 | 0.88186404 | 0.778 | 0.519 | 9.49E-276 | ExN6 | DGKG |
| AC024610.2 | 4.43E-280 | 0.44719008 | 0.423 | 0.173 | 1.69E-275 | ExN6 | AC024610.2 |
| RMST2 | 2.25E-277 | 0.42069746 | 0.408 | 0.154 | 8.58E-273 | ExN6 | RMST |
| PCSK5 | 3.78E-277 | 0.77763504 | 0.436 | 0.184 | 1.45E-272 | ExN6 | PCSK5 |
| SGCZ | 4.52E-277 | 0.40733256 | 0.814 | 0.492 | 1.73E-272 | ExN6 | SGCZ |

| | | | | | | | |
|---|---|---|---|---|---|---|---|
| RASGEF1C | 7.95E-273 | 0.38670711 | 0.369 | 0.141 | 3.04E-268 | ExN6 | RASGEF1C |
| UST | 2.65E-272 | 0.58181108 | 0.35 | 0.131 | 1.01E-267 | ExN6 | UST |
| CARMIL13 | 9.87E-267 | 0.59026669 | 0.845 | 0.606 | 3.77E-262 | ExN6 | CARMIL1 |
| LINC018112 | 4.82E-265 | 0.79670768 | 0.569 | 0.291 | 1.84E-260 | ExN6 | LINC01811 |
| MAMDC2 | 8.48E-264 | 0.8204334 | 0.376 | 0.157 | 3.24E-259 | ExN6 | MAMDC2 |
| VWA3A1 | 2.23E-263 | 0.49026475 | 0.492 | 0.23 | 8.54E-259 | ExN6 | VWA3A |
| KHDRBS34 | 3.91E-262 | 0.60866994 | 0.946 | 0.786 | 1.49E-257 | ExN6 | KHDRBS3 |
| SLIT13 | 1.10E-260 | 0.59298484 | 0.816 | 0.579 | 4.20E-256 | ExN6 | SLIT1 |
| ILDR21 | 2.59E-259 | 0.54797825 | 0.584 | 0.326 | 9.90E-255 | ExN6 | ILDR2 |
| TBR12 | 7.91E-257 | 0.3579792 | 0.486 | 0.221 | 3.02E-252 | ExN6 | TBR1 |
| GRM4 | 8.25E-256 | 0.45868849 | 0.407 | 0.169 | 3.15E-251 | ExN6 | GRM4 |
| FILIP1 | 6.72E-255 | 0.29624357 | 0.278 | 0.09 | 2.57E-250 | ExN6 | FILIP1 |
| DIP2A | 1.63E-254 | 0.50379341 | 0.703 | 0.444 | 6.24E-250 | ExN6 | DIP2A |
| OLFM32 | 3.76E-254 | 0.67906132 | 0.91 | 0.702 | 1.44E-249 | ExN6 | OLFM3 |
| PTPRU | 6.74E-254 | 0.38037429 | 0.428 | 0.184 | 2.57E-249 | ExN6 | PTPRU |
| SEC24D2 | 9.52E-254 | 0.65407358 | 0.496 | 0.262 | 3.64E-249 | ExN6 | SEC24D |
| CELF24 | 4.57E-253 | 0.46552702 | 0.991 | 0.967 | 1.75E-248 | ExN6 | CELF2 |
| GPM6A2 | 3.12E-252 | 0.46085654 | 0.975 | 0.942 | 1.19E-247 | ExN6 | GPM6A |
| LINC02646 | 5.88E-251 | 0.33633111 | 0.341 | 0.126 | 2.25E-246 | ExN6 | LINC02646 |
| GRIK3 | 3.06E-243 | 0.40326474 | 0.57 | 0.282 | 1.17E-238 | ExN6 | GRIK3 |
| GALNT141 | 1.97E-240 | 0.60212442 | 0.574 | 0.315 | 7.52E-236 | ExN6 | GALNT14 |
| SEMA5A | 2.67E-238 | 0.55094765 | 0.389 | 0.16 | 1.02E-233 | ExN6 | SEMA5A |
| NKAIN24 | 1.48E-237 | 0.6741233 | 0.96 | 0.831 | 5.65E-233 | ExN6 | NKAIN2 |
| TRMT9B | 6.46E-237 | 0.72134818 | 0.625 | 0.404 | 2.47E-232 | ExN6 | TRMT9B |
| KALRN4 | 1.00E-234 | 0.43282905 | 0.983 | 0.956 | 3.82E-230 | ExN6 | KALRN |
| CPNE43 | 1.06E-231 | 0.89279527 | 0.692 | 0.456 | 4.04E-227 | ExN6 | CPNE4 |
| SDK2 | 1.16E-228 | 0.36083393 | 0.47 | 0.217 | 4.42E-224 | ExN6 | SDK2 |
| NPTX14 | 1.08E-225 | 0.50329267 | 0.742 | 0.467 | 4.12E-221 | ExN6 | NPTX1 |
| SEPTIN111 | 4.86E-225 | 0.51417074 | 0.856 | 0.667 | 1.86E-220 | ExN6 | SEPTIN11 |
| NOS1AP2 | 9.31E-225 | 0.64614771 | 0.809 | 0.654 | 3.56E-220 | ExN6 | NOS1AP |
| LMCD1-AS1 | 2.32E-224 | 0.48642775 | 0.535 | 0.273 | 8.87E-220 | ExN6 | LMCD1-AS1 |
| CPE1 | 2.18E-223 | 0.54715952 | 0.841 | 0.668 | 8.35E-219 | ExN6 | CPE |
| KCNMB42 | 3.06E-223 | 0.4826354 | 0.706 | 0.451 | 1.17E-218 | ExN6 | KCNMB4 |
| FRMPD41 | 4.95E-223 | 0.79028756 | 0.956 | 0.937 | 1.89E-218 | ExN6 | FRMPD4 |
| SDK1 | 6.39E-223 | 0.58108678 | 0.724 | 0.463 | 2.44E-218 | ExN6 | SDK1 |
| ASAP21 | 1.11E-222 | 0.53561016 | 0.627 | 0.386 | 4.26E-218 | ExN6 | ASAP2 |
| FAM102B | 3.61E-222 | 0.4263898 | 0.59 | 0.33 | 1.38E-217 | ExN6 | FAM102B |
| CACHD11 | 1.68E-221 | 0.73613027 | 0.466 | 0.243 | 6.44E-217 | ExN6 | CACHD1 |
| SRGAP1 | 1.69E-221 | 0.43886565 | 0.487 | 0.237 | 6.45E-217 | ExN6 | SRGAP1 |
| MIR137HG4 | 1.69E-221 | 0.54019274 | 0.867 | 0.648 | 6.46E-217 | ExN6 | MIR137HG |
| DENND11 | 1.73E-219 | 0.39132679 | 0.562 | 0.301 | 6.62E-215 | ExN6 | DENND11 |
| AFF32 | 5.62E-219 | 0.54288565 | 0.95 | 0.899 | 2.15E-214 | ExN6 | AFF3 |
| AL158077.22 | 7.13E-219 | 0.46218583 | 0.514 | 0.26 | 2.73E-214 | ExN6 | AL158077.2 |
| SSTR22 | 5.79E-218 | 0.28877981 | 0.397 | 0.172 | 2.21E-213 | ExN6 | SSTR2 |
| SYNPO23 | 4.71E-217 | 0.73961071 | 0.623 | 0.415 | 1.80E-212 | ExN6 | SYNPO2 |
| LMO32 | 7.03E-217 | 0.59040886 | 0.803 | 0.582 | 2.69E-212 | ExN6 | LMO3 |
| KLHL21 | 8.95E-217 | 0.46996715 | 0.743 | 0.484 | 3.42E-212 | ExN6 | KLHL2 |
| RAB3B | 1.86E-216 | 0.26705981 | 0.325 | 0.122 | 7.13E-212 | ExN6 | RAB3B |
| TANC21 | 5.09E-216 | 0.41447621 | 0.938 | 0.857 | 1.94E-211 | ExN6 | TANC2 |
| RAP1GAP24 | 1.56E-215 | 0.50877607 | 0.849 | 0.623 | 5.97E-211 | ExN6 | RAP1GAP2 |
| SLC35F13 | 2.17E-215 | 0.57336506 | 0.907 | 0.695 | 8.29E-211 | ExN6 | SLC35F1 |
| DPYS | 2.45E-213 | 0.28170416 | 0.265 | 0.092 | 9.37E-209 | ExN6 | DPYS |
| GRM82 | 4.79E-213 | 0.9977319 | 0.544 | 0.341 | 1.83E-208 | ExN6 | GRM8 |
| CEMIP22 | 5.05E-213 | 0.50782409 | 0.581 | 0.337 | 1.93E-208 | ExN6 | CEMIP2 |
| RETREG13 | 6.84E-212 | 0.49198516 | 0.787 | 0.564 | 2.61E-207 | ExN6 | RETREG1 |
| SPHKAP | 2.85E-209 | 0.3353713 | 0.368 | 0.151 | 1.09E-204 | ExN6 | SPHKAP |
| NRCAM3 | 3.47E-209 | 0.38483227 | 0.962 | 0.918 | 1.33E-204 | ExN6 | NRCAM |
| LMO7 | 6.90E-208 | 0.50316009 | 0.796 | 0.6 | 2.64E-203 | ExN6 | LMO7 |
| SSBP2 | 5.44E-207 | 0.50983084 | 0.903 | 0.818 | 2.08E-202 | ExN6 | SSBP2 |
| PTPRN2 | 1.01E-206 | 0.39390702 | 0.976 | 0.948 | 3.86E-202 | ExN6 | PTPRN2 |
| AL133346.1 | 1.52E-204 | 0.36897407 | 0.344 | 0.137 | 5.80E-200 | ExN6 | AL133346.1 |
| CA103 | 7.54E-204 | 0.6017426 | 0.715 | 0.422 | 2.88E-199 | ExN6 | CA10 |
| SH3GL31 | 1.18E-203 | 0.41026536 | 0.915 | 0.814 | 4.51E-199 | ExN6 | SH3GL3 |

| | | | | | | | |
|---|---|---|---|---|---|---|---|
| NDFIP23 | 1.02E-202 | 0.44694493 | 0.739 | 0.498 | 3.91E-198 | ExN6 | NDFIP2 |
| MMD | 8.57E-202 | 0.25420102 | 0.362 | 0.156 | 3.27E-197 | ExN6 | MMD |
| CRTAC1 | 6.48E-201 | 0.38759034 | 0.464 | 0.223 | 2.48E-196 | ExN6 | CRTAC1 |
| FAM171B | 1.04E-200 | 0.48160652 | 0.805 | 0.64 | 3.99E-196 | ExN6 | FAM171B |
| MAP2K11 | 1.23E-199 | 0.50469642 | 0.702 | 0.477 | 4.72E-195 | ExN6 | MAP2K1 |
| NLGN4Y1 | 4.07E-199 | 0.77205863 | 0.576 | 0.355 | 1.56E-194 | ExN6 | NLGN4Y |
| SORCS31 | 3.31E-197 | 0.65166865 | 0.731 | 0.559 | 1.27E-192 | ExN6 | SORCS3 |
| ZNF704 | 6.21E-196 | 0.55774552 | 0.824 | 0.661 | 2.37E-191 | ExN6 | ZNF704 |
| OCA21 | 8.23E-196 | 0.44270274 | 0.497 | 0.26 | 3.15E-191 | ExN6 | OCA2 |
| AC011246.11 | 1.78E-195 | 1.32383867 | 0.427 | 0.232 | 6.80E-191 | ExN6 | AC011246.1 |
| FILIP1L | 3.20E-195 | 0.36689989 | 0.419 | 0.203 | 1.22E-190 | ExN6 | FILIP1L |
| PRKAG23 | 1.19E-194 | 0.58762893 | 0.87 | 0.719 | 4.56E-190 | ExN6 | PRKAG2 |
| AKAP132 | 2.70E-194 | 0.46082141 | 0.595 | 0.351 | 1.03E-189 | ExN6 | AKAP13 |
| PLXDC23 | 1.39E-193 | 0.63713229 | 0.781 | 0.6 | 5.29E-189 | ExN6 | PLXDC2 |
| MEGF11 | 5.33E-192 | 0.34608063 | 0.324 | 0.133 | 2.04E-187 | ExN6 | MEGF11 |
| MBOAT2 | 5.15E-191 | 0.49590321 | 0.782 | 0.575 | 1.97E-186 | ExN6 | MBOAT2 |
| TESC | 7.46E-188 | 0.37639342 | 0.354 | 0.163 | 2.85E-183 | ExN6 | TESC |
| PAK5 | 6.49E-187 | 0.48124396 | 0.879 | 0.751 | 2.48E-182 | ExN6 | PAK5 |
| AC092969.1 | 1.94E-184 | 0.49705098 | 0.34 | 0.151 | 7.42E-180 | ExN6 | AC092969.1 |
| HMGCLL1 | 3.12E-183 | 0.4596016 | 0.72 | 0.503 | 1.19E-178 | ExN6 | HMGCLL1 |
| NIN3 | 5.46E-180 | 0.48164323 | 0.868 | 0.701 | 2.09E-175 | ExN6 | NIN |
| PDE4D4 | 1.10E-177 | 0.51197753 | 0.991 | 0.959 | 4.22E-173 | ExN6 | PDE4D |
| SNTG22 | 3.78E-176 | 0.46594753 | 0.323 | 0.142 | 1.45E-171 | ExN6 | SNTG2 |
| SYT12 | 1.75E-175 | 0.31981302 | 0.995 | 0.985 | 6.70E-171 | ExN6 | SYT1 |
| EDA | 5.42E-175 | 0.28790192 | 0.31 | 0.129 | 2.07E-170 | ExN6 | EDA |
| IGSF211 | 3.89E-174 | 0.54319812 | 0.713 | 0.521 | 1.49E-169 | ExN6 | IGSF21 |
| HLF1 | 3.14E-173 | 0.44070771 | 0.718 | 0.499 | 1.20E-168 | ExN6 | HLF |
| BASP1-AS14 | 3.60E-171 | 0.4672862 | 0.858 | 0.645 | 1.37E-166 | ExN6 | BASP1-AS1 |
| NRP11 | 8.96E-171 | 0.47780409 | 0.384 | 0.193 | 3.42E-166 | ExN6 | NRP1 |
| ST6GAL23 | 4.55E-170 | 0.49925649 | 0.596 | 0.377 | 1.74E-165 | ExN6 | ST6GAL2 |
| EPHA52 | 5.47E-170 | 0.54922492 | 0.767 | 0.588 | 2.09E-165 | ExN6 | EPHA5 |
| PWRN1 | 6.31E-170 | 0.55910347 | 0.673 | 0.457 | 2.41E-165 | ExN6 | PWRN1 |
| DENND33 | 1.34E-169 | 0.34778059 | 0.576 | 0.33 | 5.14E-165 | ExN6 | DENND3 |
| RIMS2 | 4.09E-169 | 0.3319019 | 0.98 | 0.968 | 1.56E-164 | ExN6 | RIMS2 |
| GAS7 | 1.36E-168 | 0.46978171 | 0.897 | 0.772 | 5.21E-164 | ExN6 | GAS7 |
| TRPM31 | 3.62E-168 | 1.36030121 | 0.542 | 0.402 | 1.38E-163 | ExN6 | TRPM3 |
| ATP2B12 | 1.29E-167 | 0.39185634 | 0.943 | 0.842 | 4.95E-163 | ExN6 | ATP2B1 |
| GLCE | 5.14E-167 | 0.40149341 | 0.578 | 0.349 | 1.97E-162 | ExN6 | GLCE |
| CEP70 | 4.42E-166 | 0.39305079 | 0.646 | 0.421 | 1.69E-161 | ExN6 | CEP70 |
| GCNT4 | 6.61E-166 | 0.39314585 | 0.364 | 0.177 | 2.53E-161 | ExN6 | GCNT4 |
| AC090709.1 | 2.08E-165 | 0.30469209 | 0.341 | 0.155 | 7.94E-161 | ExN6 | AC090709.1 |
| CRYBG3 | 5.86E-165 | 0.27351133 | 0.271 | 0.108 | 2.24E-160 | ExN6 | CRYBG3 |
| MAPK103 | 2.53E-164 | 0.35009566 | 0.977 | 0.951 | 9.67E-160 | ExN6 | MAPK10 |
| GNAO1 | 3.66E-164 | 0.58390272 | 0.889 | 0.805 | 1.40E-159 | ExN6 | GNAO1 |
| MEG33 | 1.11E-163 | 0.3576771 | 0.988 | 0.958 | 4.24E-159 | ExN6 | MEG3 |
| EFHD21 | 3.82E-163 | 0.32062197 | 0.47 | 0.256 | 1.46E-158 | ExN6 | EFHD2 |
| LINC022231 | 1.48E-162 | 0.41101618 | 0.503 | 0.282 | 5.67E-158 | ExN6 | LINC02223 |
| AL589843.1 | 3.88E-162 | 0.35104773 | 0.621 | 0.387 | 1.48E-157 | ExN6 | AL589843.1 |
| CHN14 | 8.93E-162 | 0.35894016 | 0.928 | 0.739 | 3.41E-157 | ExN6 | CHN1 |
| SLC36A1 | 3.23E-161 | 0.31373236 | 0.52 | 0.295 | 1.23E-156 | ExN6 | SLC36A1 |
| KIAA1671 | 8.01E-161 | 0.49050511 | 0.626 | 0.419 | 3.06E-156 | ExN6 | KIAA1671 |
| ST8SIA5 | 2.06E-160 | 0.45876306 | 0.794 | 0.611 | 7.87E-156 | ExN6 | ST8SIA5 |
| NFIA1 | 2.15E-160 | 0.68954949 | 0.602 | 0.409 | 8.20E-156 | ExN6 | NFIA |
| HIVEP11 | 2.78E-160 | 0.44472749 | 0.656 | 0.449 | 1.06E-155 | ExN6 | HIVEP1 |
| SYT74 | 4.61E-160 | 0.37903898 | 0.763 | 0.518 | 1.76E-155 | ExN6 | SYT7 |
| HIVEP2 | 6.10E-160 | 0.42181737 | 0.886 | 0.794 | 2.33E-155 | ExN6 | HIVEP2 |
| CACNA1E1 | 9.89E-159 | 0.47244326 | 0.881 | 0.804 | 3.78E-154 | ExN6 | CACNA1E |
| AL591519.1 | 2.14E-157 | 0.42269313 | 0.508 | 0.286 | 8.17E-153 | ExN6 | AL591519.1 |
| CTNNA31 | 1.30E-155 | 0.85364242 | 0.645 | 0.443 | 4.96E-151 | ExN6 | CTNNA3 |
| AL136456.11 | 1.75E-155 | 0.31961761 | 0.31 | 0.135 | 6.71E-151 | ExN6 | AL136456.1 |
| ARHGAP44 | 2.78E-155 | 0.37663258 | 0.874 | 0.755 | 1.06E-150 | ExN6 | ARHGAP44 |
| SETBP1 | 3.62E-155 | 0.42526082 | 0.941 | 0.885 | 1.39E-150 | ExN6 | SETBP1 |
| DSCAM2 | 3.77E-155 | 0.35947653 | 0.98 | 0.9 | 1.44E-150 | ExN6 | DSCAM |

| | | | | | | | |
|---|---|---|---|---|---|---|---|
| RAP1GAP | 2.45E-154 | 0.41368166 | 0.716 | 0.527 | 9.37E-150 | ExN6 | RAP1GAP |
| PCLO2 | 4.67E-154 | 0.3719567 | 0.957 | 0.921 | 1.79E-149 | ExN6 | PCLO |
| PTPRD4 | 1.09E-153 | 0.43525516 | 0.993 | 0.962 | 4.17E-149 | ExN6 | PTPRD |
| BAIAP21 | 3.10E-152 | 0.3683815 | 0.73 | 0.49 | 1.18E-147 | ExN6 | BAIAP2 |
| MCC | 3.99E-150 | 0.40086818 | 0.459 | 0.252 | 1.53E-145 | ExN6 | MCC |
| AC040168.1 | 1.20E-149 | 0.36342366 | 0.532 | 0.312 | 4.58E-145 | ExN6 | AC040168.1 |
| DMD4 | 3.33E-149 | 0.43321889 | 0.969 | 0.918 | 1.27E-144 | ExN6 | DMD |
| GUCY1A23 | 8.37E-149 | 0.4175577 | 0.869 | 0.722 | 3.20E-144 | ExN6 | GUCY1A2 |
| RPE | 3.56E-148 | 0.35994246 | 0.589 | 0.369 | 1.36E-143 | ExN6 | RPE |
| SH3RF3 | 3.71E-148 | 0.46210009 | 0.842 | 0.67 | 1.42E-143 | ExN6 | SH3RF3 |
| CAMK2D1 | 1.98E-147 | 0.52828589 | 0.831 | 0.659 | 7.56E-143 | ExN6 | CAMK2D |
| ANKS1B3 | 1.25E-146 | 0.337925 | 0.992 | 0.986 | 4.76E-142 | ExN6 | ANKS1B |
| SPOCK12 | 1.45E-146 | 0.71271209 | 0.845 | 0.744 | 5.56E-142 | ExN6 | SPOCK1 |
| PTPRR1 | 3.10E-146 | 0.44647068 | 0.841 | 0.685 | 1.19E-141 | ExN6 | PTPRR |
| SGIP1 | 1.45E-145 | 0.3627264 | 0.928 | 0.871 | 5.56E-141 | ExN6 | SGIP1 |
| GRID1 | 2.74E-143 | 0.38412499 | 0.929 | 0.847 | 1.05E-138 | ExN6 | GRID1 |
| MAOB1 | 2.57E-140 | 0.35385786 | 0.492 | 0.29 | 9.84E-136 | ExN6 | MAOB |
| MNAT11 | 3.62E-140 | 0.3739493 | 0.884 | 0.784 | 1.38E-135 | ExN6 | MNAT1 |
| GABRG31 | 1.15E-137 | 0.34066501 | 0.926 | 0.792 | 4.41E-133 | ExN6 | GABRG3 |
| ANTXR1 | 2.16E-137 | 0.40168037 | 0.604 | 0.398 | 8.25E-133 | ExN6 | ANTXR1 |
| USP9Y1 | 1.00E-136 | 0.4855064 | 0.544 | 0.339 | 3.83E-132 | ExN6 | USP9Y |
| NTNG2 | 1.74E-136 | 0.66721292 | 0.265 | 0.128 | 6.66E-132 | ExN6 | NTNG2 |
| FUT91 | 2.22E-136 | 0.45489356 | 0.891 | 0.811 | 8.48E-132 | ExN6 | FUT9 |
| ITGA8 | 9.84E-136 | 0.59572786 | 0.267 | 0.118 | 3.76E-131 | ExN6 | ITGA8 |
| ST3GAL62 | 4.00E-135 | 0.36158592 | 0.638 | 0.421 | 1.53E-130 | ExN6 | ST3GAL6 |
| AL513166.14 | 7.43E-135 | 0.45480182 | 0.795 | 0.612 | 2.84E-130 | ExN6 | AL513166.1 |
| SPARCL11 | 4.04E-134 | 0.29437961 | 0.855 | 0.652 | 1.54E-129 | ExN6 | SPARCL1 |
| ZNF8271 | 6.11E-134 | 0.35079341 | 0.684 | 0.47 | 2.33E-129 | ExN6 | ZNF827 |
| AC120193.13 | 7.63E-134 | 0.38156276 | 0.529 | 0.314 | 2.92E-129 | ExN6 | AC120193.1 |
| ADAMTS192 | 1.85E-133 | 0.41835718 | 0.564 | 0.355 | 7.06E-129 | ExN6 | ADAMTS19 |
| FMN21 | 5.47E-133 | 0.31620635 | 0.948 | 0.898 | 2.09E-128 | ExN6 | FMN2 |
| AL133166.13 | 7.50E-133 | 0.26302775 | 0.401 | 0.21 | 2.87E-128 | ExN6 | AL133166.1 |
| UTY | 2.67E-132 | 0.43068945 | 0.507 | 0.306 | 1.02E-127 | ExN6 | UTY |
| AC005906.22 | 8.69E-132 | 0.30597519 | 0.488 | 0.282 | 3.32E-127 | ExN6 | AC005906.2 |
| STK391 | 1.40E-131 | 0.36956013 | 0.778 | 0.589 | 5.35E-127 | ExN6 | STK39 |
| AC007038.11 | 1.51E-130 | 0.2628414 | 0.365 | 0.187 | 5.77E-126 | ExN6 | AC007038.1 |
| CRYM2 | 5.88E-130 | 0.57874423 | 0.432 | 0.259 | 2.25E-125 | ExN6 | CRYM |
| TNFRSF21 | 1.28E-129 | 0.2758691 | 0.466 | 0.271 | 4.89E-125 | ExN6 | TNFRSF21 |
| GLIS11 | 5.01E-129 | 0.31375882 | 0.682 | 0.465 | 1.92E-124 | ExN6 | GLIS1 |
| DPYD3 | 6.15E-129 | 0.37095125 | 0.873 | 0.653 | 2.35E-124 | ExN6 | DPYD |
| GAREM11 | 7.71E-128 | 0.44895274 | 0.643 | 0.459 | 2.95E-123 | ExN6 | GAREM1 |
| ADCY11 | 1.70E-127 | 0.41741519 | 0.757 | 0.59 | 6.50E-123 | ExN6 | ADCY1 |
| FGFR1 | 1.51E-126 | 0.38542414 | 0.319 | 0.163 | 5.77E-122 | ExN6 | FGFR1 |
| CACNA2D33 | 3.14E-126 | 0.4447435 | 0.95 | 0.862 | 1.20E-121 | ExN6 | CACNA2D3 |
| EFR3A | 1.92E-125 | 0.37807603 | 0.762 | 0.605 | 7.34E-121 | ExN6 | EFR3A |
| KLF7 | 2.80E-125 | 0.30998901 | 0.53 | 0.329 | 1.07E-120 | ExN6 | KLF7 |
| SVOP | 8.39E-125 | 0.37732032 | 0.691 | 0.494 | 3.21E-120 | ExN6 | SVOP |
| KLHL5 | 1.88E-124 | 0.27560824 | 0.515 | 0.306 | 7.18E-120 | ExN6 | KLHL5 |
| VIPR1 | 1.03E-123 | 0.26276955 | 0.425 | 0.233 | 3.94E-119 | ExN6 | VIPR1 |
| GABRA3 | 1.18E-123 | 0.34270182 | 0.526 | 0.331 | 4.50E-119 | ExN6 | GABRA3 |
| TNIK | 1.35E-123 | 0.2977513 | 0.905 | 0.797 | 5.16E-119 | ExN6 | TNIK |
| PCDH11Y | 7.25E-123 | 0.44703202 | 0.393 | 0.215 | 2.77E-118 | ExN6 | PCDH11Y |
| CDH21 | 7.40E-123 | 0.44630746 | 0.836 | 0.751 | 2.83E-118 | ExN6 | CDH2 |
| KCNQ33 | 8.44E-123 | 0.38737919 | 0.959 | 0.898 | 3.23E-118 | ExN6 | KCNQ3 |
| DYNC1I13 | 8.98E-123 | 0.31846623 | 0.93 | 0.869 | 3.43E-118 | ExN6 | DYNC1I1 |
| ARHGAP222 | 1.03E-122 | 0.32742346 | 0.492 | 0.296 | 3.93E-118 | ExN6 | ARHGAP22 |
| PHKB | 2.19E-122 | 0.33935587 | 0.746 | 0.562 | 8.38E-118 | ExN6 | PHKB |
| MAGI32 | 9.11E-121 | 0.50049504 | 0.822 | 0.679 | 3.48E-116 | ExN6 | MAGI3 |
| GNPTAB | 1.63E-120 | 0.34271556 | 0.712 | 0.525 | 6.23E-116 | ExN6 | GNPTAB |
| JPH11 | 1.44E-119 | 0.31541845 | 0.642 | 0.438 | 5.52E-115 | ExN6 | JPH1 |
| ARHGEF282 | 1.99E-119 | 0.32814722 | 0.495 | 0.299 | 7.59E-115 | ExN6 | ARHGEF28 |
| AC069437.1 | 6.83E-119 | 0.28495549 | 0.349 | 0.183 | 2.61E-114 | ExN6 | AC069437.1 |
| ABLIM3 | 9.43E-119 | 0.29679961 | 0.578 | 0.37 | 3.61E-114 | ExN6 | ABLIM3 |

| | | | | | | | |
|---|---|---|---|---|---|---|---|
| FARS22 | 1.67E-118 | 0.35292744 | 0.874 | 0.755 | 6.40E-114 | ExN6 | FARS2 |
| SCPEP1 | 8.37E-118 | 0.30009026 | 0.471 | 0.288 | 3.20E-113 | ExN6 | SCPEP1 |
| SIDT11 | 9.23E-118 | 0.37236885 | 0.699 | 0.506 | 3.53E-113 | ExN6 | SIDT1 |
| AC091078.12 | 2.07E-117 | 0.7551941 | 0.471 | 0.297 | 7.93E-113 | ExN6 | AC091078.1 |
| ZC2HC1A | 2.94E-116 | 0.29806159 | 0.624 | 0.426 | 1.13E-111 | ExN6 | ZC2HC1A |
| EPHA7 | 6.05E-116 | 0.30166064 | 0.737 | 0.531 | 2.31E-111 | ExN6 | EPHA7 |
| VPS13A2 | 1.68E-115 | 0.3179474 | 0.853 | 0.727 | 6.42E-111 | ExN6 | VPS13A |
| CDH181 | 6.06E-115 | 0.53596111 | 0.906 | 0.869 | 2.32E-110 | ExN6 | CDH18 |
| KHDRBS23 | 3.77E-114 | 0.31410074 | 0.976 | 0.895 | 1.44E-109 | ExN6 | KHDRBS2 |
| SMAP12 | 6.96E-114 | 0.41527227 | 0.776 | 0.617 | 2.66E-109 | ExN6 | SMAP1 |
| SATB24 | 4.07E-113 | 0.57952345 | 0.792 | 0.564 | 1.56E-108 | ExN6 | SATB2 |
| GFPT2 | 8.55E-113 | 0.25749417 | 0.449 | 0.264 | 3.27E-108 | ExN6 | GFPT2 |
| SFXN5 | 1.77E-112 | 0.32733984 | 0.715 | 0.517 | 6.78E-108 | ExN6 | SFXN5 |
| LRRN2 | 2.74E-112 | 0.2684445 | 0.711 | 0.505 | 1.05E-107 | ExN6 | LRRN2 |
| ANO6 | 5.08E-112 | 0.35993938 | 0.459 | 0.285 | 1.94E-107 | ExN6 | ANO6 |
| TRPM2 | 1.38E-111 | 0.30940242 | 0.599 | 0.402 | 5.29E-107 | ExN6 | TRPM2 |
| CTNNAL11 | 3.35E-111 | 0.33395323 | 0.622 | 0.43 | 1.28E-106 | ExN6 | CTNNAL1 |
| KATNAL1 | 6.55E-111 | 0.26983141 | 0.609 | 0.402 | 2.50E-106 | ExN6 | KATNAL1 |
| TTTY14 | 1.06E-110 | 0.43281429 | 0.553 | 0.355 | 4.04E-106 | ExN6 | TTTY14 |
| DGKH1 | 1.17E-110 | 0.34871098 | 0.556 | 0.371 | 4.45E-106 | ExN6 | DGKH |
| LSAMP | 1.79E-109 | 0.48533587 | 0.99 | 0.973 | 6.85E-105 | ExN6 | LSAMP |
| FARP11 | 4.63E-109 | 0.42118342 | 0.759 | 0.652 | 1.77E-104 | ExN6 | FARP1 |
| PRICKLE2 | 1.45E-108 | 0.31708467 | 0.882 | 0.788 | 5.54E-104 | ExN6 | PRICKLE2 |
| PPP1R12B | 1.94E-108 | 0.32668057 | 0.855 | 0.752 | 7.40E-104 | ExN6 | PPP1R12B |
| UNC5C | 1.50E-106 | 0.8644066 | 0.846 | 0.781 | 5.75E-102 | ExN6 | UNC5C |
| C4orf501 | 2.14E-106 | 0.28226632 | 0.548 | 0.353 | 8.17E-102 | ExN6 | C4orf50 |
| ASAP1 | 7.68E-106 | 0.33049907 | 0.889 | 0.796 | 2.93E-101 | ExN6 | ASAP1 |
| POU6F24 | 2.50E-105 | 0.36153326 | 0.611 | 0.39 | 9.54E-101 | ExN6 | POU6F2 |
| ELL22 | 8.69E-105 | 0.32516924 | 0.736 | 0.534 | 3.32E-100 | ExN6 | ELL2 |
| GPD2 | 1.13E-103 | 0.32047438 | 0.493 | 0.318 | 4.31E-99 | ExN6 | GPD2 |
| GPR63 | 1.25E-103 | 0.3443493 | 0.402 | 0.243 | 4.77E-99 | ExN6 | GPR63 |
| IQSEC11 | 1.71E-103 | 0.33811302 | 0.887 | 0.774 | 6.54E-99 | ExN6 | IQSEC1 |
| DONSON | 2.20E-103 | 0.32825215 | 0.308 | 0.16 | 8.42E-99 | ExN6 | DONSON |
| B4GALNT32 | 5.10E-103 | 0.26522884 | 0.49 | 0.304 | 1.95E-98 | ExN6 | B4GALNT3 |
| EPS151 | 5.46E-103 | 0.27467417 | 0.667 | 0.474 | 2.09E-98 | ExN6 | EPS15 |
| FMNL22 | 1.36E-102 | 0.34722975 | 0.923 | 0.87 | 5.21E-98 | ExN6 | FMNL2 |
| JAZF1 | 2.20E-102 | 0.27376159 | 0.905 | 0.815 | 8.41E-98 | ExN6 | JAZF1 |
| AFF2 | 4.41E-102 | 0.52835107 | 0.553 | 0.38 | 1.69E-97 | ExN6 | AFF2 |
| NEGR12 | 5.98E-102 | 0.33397955 | 0.983 | 0.954 | 2.29E-97 | ExN6 | NEGR1 |
| PTCHD1-AS2 | 2.62E-101 | 0.37647884 | 0.642 | 0.439 | 1.00E-96 | ExN6 | PTCHD1-AS |
| LHFPL5 | 1.03E-100 | 0.25748667 | 0.445 | 0.268 | 3.95E-96 | ExN6 | LHFPL5 |
| SEMA5B1 | 1.30E-100 | 0.39873919 | 0.448 | 0.287 | 4.98E-96 | ExN6 | SEMA5B |
| ATP8B4 | 1.57E-100 | 0.40744912 | 0.378 | 0.223 | 5.99E-96 | ExN6 | ATP8B4 |
| CAP2 | 3.07E-100 | 0.34104578 | 0.792 | 0.648 | 1.17E-95 | ExN6 | CAP2 |
| SGMS1 | 3.92E-99 | 0.35310372 | 0.786 | 0.66 | 1.50E-94 | ExN6 | SGMS1 |
| SCD5 | 6.89E-99 | 0.2791286 | 0.773 | 0.596 | 2.64E-94 | ExN6 | SCD5 |
| STK38L | 7.78E-99 | 0.31200405 | 0.525 | 0.35 | 2.97E-94 | ExN6 | STK38L |
| C1GALT1 | 8.00E-99 | 0.36097638 | 0.479 | 0.312 | 3.06E-94 | ExN6 | C1GALT1 |
| AACS1 | 8.74E-99 | 0.29178506 | 0.645 | 0.455 | 3.34E-94 | ExN6 | AACS |
| VPS413 | 3.79E-98 | 0.30305515 | 0.754 | 0.581 | 1.45E-93 | ExN6 | VPS41 |
| SLC16A1-AS1 | 8.46E-98 | 0.33398087 | 0.773 | 0.597 | 3.23E-93 | ExN6 | SLC16A1-AS1 |
| TMEM178A | 1.92E-97 | 0.33140041 | 0.741 | 0.581 | 7.32E-93 | ExN6 | TMEM178A |
| NPY1R | 2.11E-97 | 0.29521927 | 0.523 | 0.347 | 8.08E-93 | ExN6 | NPY1R |
| ARNTL22 | 4.14E-97 | 0.4558598 | 0.515 | 0.37 | 1.58E-92 | ExN6 | ARNTL2 |
| EFNA54 | 4.92E-97 | 0.25537267 | 0.928 | 0.789 | 1.88E-92 | ExN6 | EFNA5 |
| AC092691.12 | 7.23E-97 | 0.66200806 | 0.836 | 0.872 | 2.77E-92 | ExN6 | AC092691.1 |
| SNCA3 | 4.12E-96 | 0.28571006 | 0.797 | 0.599 | 1.57E-91 | ExN6 | SNCA |
| KSR2 | 5.86E-96 | 0.32290231 | 0.933 | 0.889 | 2.24E-91 | ExN6 | KSR2 |
| TMEM131 | 1.29E-95 | 0.29004388 | 0.779 | 0.638 | 4.94E-91 | ExN6 | TMEM131 |
| GNG7 | 3.81E-95 | 0.33953866 | 0.575 | 0.397 | 1.46E-90 | ExN6 | GNG7 |
| FRMD31 | 5.09E-95 | 0.3875342 | 0.504 | 0.333 | 1.95E-90 | ExN6 | FRMD3 |
| CDKL51 | 1.29E-94 | 0.35808376 | 0.846 | 0.724 | 4.94E-90 | ExN6 | CDKL5 |
| PAPSS11 | 2.61E-94 | 0.28378832 | 0.682 | 0.511 | 9.98E-90 | ExN6 | PAPSS1 |

| | | | | | | | |
|---|---|---|---|---|---|---|---|
| INPP4B1 | 4.14E-94 | 0.35927804 | 0.89 | 0.745 | 1.58E-89 | ExN6 | INPP4B |
| NFASC | 4.48E-94 | 0.27565266 | 0.894 | 0.805 | 1.71E-89 | ExN6 | NFASC |
| GDA | 1.35E-92 | 0.38352875 | 0.537 | 0.389 | 5.17E-88 | ExN6 | GDA |
| COL11A11 | 1.71E-92 | 0.37796998 | 0.41 | 0.251 | 6.52E-88 | ExN6 | COL11A1 |
| RAPGEF41 | 2.51E-92 | 0.39467037 | 0.922 | 0.872 | 9.60E-88 | ExN6 | RAPGEF4 |
| ZNF385D | 2.52E-92 | 0.84070389 | 0.735 | 0.5 | 9.65E-88 | ExN6 | ZNF385D |
| ADCY2 | 3.15E-91 | 0.36250708 | 0.915 | 0.85 | 1.20E-86 | ExN6 | ADCY2 |
| AC006504.53 | 1.57E-90 | 0.26171295 | 0.616 | 0.419 | 6.00E-86 | ExN6 | AC006504.5 |
| SLC38A61 | 2.94E-90 | 0.28326685 | 0.699 | 0.525 | 1.13E-85 | ExN6 | SLC38A6 |
| RPTOR | 1.68E-89 | 0.30382924 | 0.77 | 0.619 | 6.43E-85 | ExN6 | RPTOR |
| ST3GAL11 | 4.34E-89 | 0.28890047 | 0.463 | 0.291 | 1.66E-84 | ExN6 | ST3GAL1 |
| SSBP33 | 5.15E-89 | 0.30592765 | 0.812 | 0.668 | 1.97E-84 | ExN6 | SSBP3 |
| GRIK41 | 6.19E-89 | 0.53088534 | 0.566 | 0.438 | 2.37E-84 | ExN6 | GRIK4 |
| SIK2 | 8.66E-89 | 0.30542015 | 0.76 | 0.615 | 3.31E-84 | ExN6 | SIK2 |
| STXBP5-AS12 | 1.21E-87 | 0.3235508 | 0.746 | 0.578 | 4.64E-83 | ExN6 | STXBP5-AS1 |
| MPP72 | 2.14E-87 | 0.48556288 | 0.529 | 0.396 | 8.16E-83 | ExN6 | MPP7 |
| WDPCP1 | 5.59E-87 | 0.30989816 | 0.865 | 0.756 | 2.14E-82 | ExN6 | WDPCP |
| MIR100HG | 6.67E-86 | 0.29311224 | 0.821 | 0.696 | 2.55E-81 | ExN6 | MIR100HG |
| PRR16 | 7.66E-86 | 0.45801682 | 0.685 | 0.542 | 2.93E-81 | ExN6 | PRR16 |
| ITGB82 | 1.14E-85 | 0.66437507 | 0.405 | 0.281 | 4.37E-81 | ExN6 | ITGB8 |
| STRBP1 | 1.39E-85 | 0.30255039 | 0.86 | 0.754 | 5.31E-81 | ExN6 | STRBP |
| PPP2R2C4 | 1.24E-84 | 0.27860673 | 0.899 | 0.794 | 4.73E-80 | ExN6 | PPP2R2C |
| SGPP2 | 1.25E-84 | 0.29052179 | 0.472 | 0.315 | 4.80E-80 | ExN6 | SGPP2 |
| AL008633.14 | 4.74E-84 | 0.3114864 | 0.872 | 0.705 | 1.81E-79 | ExN6 | AL008633.1 |
| MAP4K3-DT | 1.12E-83 | 0.30365747 | 0.81 | 0.651 | 4.28E-79 | ExN6 | MAP4K3-DT |
| TOX3 | 1.51E-83 | 0.41043196 | 0.588 | 0.435 | 5.78E-79 | ExN6 | TOX |
| TUSC3 | 2.72E-83 | 0.27234591 | 0.777 | 0.65 | 1.04E-78 | ExN6 | TUSC3 |
| AC110023.11 | 1.16E-82 | 0.72620964 | 0.443 | 0.303 | 4.42E-78 | ExN6 | AC110023.1 |
| SHC32 | 1.43E-82 | 0.33284105 | 0.753 | 0.6 | 5.45E-78 | ExN6 | SHC3 |
| RALGPS21 | 1.87E-82 | 0.30930021 | 0.688 | 0.53 | 7.16E-78 | ExN6 | RALGPS2 |
| FKBP51 | 4.00E-82 | 0.27720111 | 0.412 | 0.253 | 1.53E-77 | ExN6 | FKBP5 |
| BBS9 | 5.35E-82 | 0.27298808 | 0.829 | 0.717 | 2.05E-77 | ExN6 | BBS9 |
| BASP12 | 3.78E-81 | 0.27120641 | 0.869 | 0.765 | 1.45E-76 | ExN6 | BASP1 |
| CPNE51 | 4.14E-81 | 0.34411104 | 0.579 | 0.421 | 1.58E-76 | ExN6 | CPNE5 |
| HOMER13 | 5.24E-81 | 0.34086796 | 0.756 | 0.613 | 2.00E-76 | ExN6 | HOMER1 |
| ADK | 1.42E-80 | 0.33361232 | 0.864 | 0.779 | 5.42E-76 | ExN6 | ADK |
| KCNH53 | 3.11E-80 | 0.42569456 | 0.56 | 0.41 | 1.19E-75 | ExN6 | KCNH5 |
| DDAH13 | 1.57E-79 | 0.32668756 | 0.575 | 0.415 | 6.02E-75 | ExN6 | DDAH1 |
| LINC018213 | 1.68E-79 | 0.31529441 | 0.349 | 0.207 | 6.43E-75 | ExN6 | LINC01821 |
| PRICKLE12 | 1.69E-79 | 0.33436108 | 0.808 | 0.685 | 6.44E-75 | ExN6 | PRICKLE1 |
| PEX5L2 | 9.40E-79 | 0.47982863 | 0.85 | 0.828 | 3.59E-74 | ExN6 | PEX5L |
| AC008269.1 | 1.30E-78 | 0.27276364 | 0.761 | 0.583 | 4.97E-74 | ExN6 | AC008269.1 |
| SKAP2 | 1.93E-78 | 0.25538656 | 0.544 | 0.37 | 7.37E-74 | ExN6 | SKAP2 |
| CCNY | 1.91E-76 | 0.27315603 | 0.814 | 0.688 | 7.31E-72 | ExN6 | CCNY |
| FOXP14 | 2.20E-76 | 0.25388221 | 0.907 | 0.815 | 8.40E-72 | ExN6 | FOXP1 |
| BCL11A1 | 1.18E-75 | 0.32678469 | 0.728 | 0.591 | 4.53E-71 | ExN6 | BCL11A |
| CHSY32 | 2.13E-75 | 0.28728635 | 0.925 | 0.83 | 8.16E-71 | ExN6 | CHSY3 |
| WIPF31 | 1.52E-74 | 0.32375148 | 0.557 | 0.399 | 5.80E-70 | ExN6 | WIPF3 |
| OSBPL10 | 3.64E-74 | 0.28610463 | 0.632 | 0.475 | 1.39E-69 | ExN6 | OSBPL10 |
| AGAP12 | 7.03E-74 | 0.28944049 | 0.949 | 0.907 | 2.69E-69 | ExN6 | AGAP1 |
| ARGLU1 | 2.01E-73 | 0.29130845 | 0.815 | 0.7 | 7.68E-69 | ExN6 | ARGLU1 |
| DOP1B | 6.65E-73 | 0.25447919 | 0.552 | 0.382 | 2.54E-68 | ExN6 | DOP1B |
| STPG2 | 8.07E-73 | 0.28919033 | 0.301 | 0.174 | 3.08E-68 | ExN6 | STPG2 |
| LINC011222 | 1.23E-72 | 0.29019395 | 0.845 | 0.681 | 4.72E-68 | ExN6 | LINC01122 |
| GSG1L2 | 1.28E-72 | 0.33400194 | 0.441 | 0.299 | 4.91E-68 | ExN6 | GSG1L |
| NR3C1 | 2.41E-72 | 0.25686855 | 0.811 | 0.696 | 9.21E-68 | ExN6 | NR3C1 |
| AC245060.5 | 3.04E-72 | 0.30811152 | 0.344 | 0.21 | 1.16E-67 | ExN6 | AC245060.5 |
| MICAL22 | 9.81E-72 | 0.26959133 | 0.858 | 0.725 | 3.75E-67 | ExN6 | MICAL2 |
| PPP1R9A | 1.31E-71 | 0.26387189 | 0.862 | 0.786 | 5.02E-67 | ExN6 | PPP1R9A |
| C11orf80 | 1.57E-71 | 0.26752382 | 0.756 | 0.608 | 5.99E-67 | ExN6 | C11orf80 |
| TRIM2 | 2.37E-71 | 0.25213478 | 0.882 | 0.786 | 9.07E-67 | ExN6 | TRIM2 |
| CALN11 | 1.52E-70 | 0.27076716 | 0.794 | 0.653 | 5.83E-66 | ExN6 | CALN1 |
| CABP15 | 1.69E-70 | 0.28809106 | 0.851 | 0.695 | 6.44E-66 | ExN6 | CABP1 |

| | | | | | | | |
|---|---|---|---|---|---|---|---|
| CCDC61 | 3.32E-70 | 0.26527932 | 0.712 | 0.556 | 1.27E-65 | ExN6 | CCDC6 |
| RAP1GDS1 | 3.09E-69 | 0.25835651 | 0.757 | 0.611 | 1.18E-64 | ExN6 | RAP1GDS1 |
| SEL1L3 | 5.82E-68 | 0.27620648 | 0.425 | 0.282 | 2.22E-63 | ExN6 | SEL1L3 |
| TNPO11 | 3.71E-67 | 0.25185575 | 0.776 | 0.635 | 1.42E-62 | ExN6 | TNPO1 |
| COL24A11 | 5.85E-67 | 0.52947133 | 0.352 | 0.236 | 2.23E-62 | ExN6 | COL24A1 |
| LRRTM31 | 1.08E-66 | 0.27949402 | 0.901 | 0.828 | 4.13E-62 | ExN6 | LRRTM3 |
| HIVEP31 | 6.56E-66 | 0.28988266 | 0.899 | 0.86 | 2.51E-61 | ExN6 | HIVEP3 |
| AC019211.14 | 6.83E-66 | 0.41763662 | 0.499 | 0.345 | 2.61E-61 | ExN6 | AC019211.1 |
| SLIT33 | 9.62E-65 | 0.349489 | 0.622 | 0.46 | 3.68E-60 | ExN6 | SLIT3 |
| PDE7A | 3.36E-63 | 0.30726946 | 0.62 | 0.481 | 1.28E-58 | ExN6 | PDE7A |
| KIAA09303 | 1.14E-62 | 0.27432441 | 0.74 | 0.605 | 4.36E-58 | ExN6 | KIAA0930 |
| HULC | 1.41E-62 | 0.25711427 | 0.688 | 0.525 | 5.40E-58 | ExN6 | HULC |
| KCNH73 | 1.89E-62 | 0.3146579 | 0.842 | 0.723 | 7.24E-58 | ExN6 | KCNH7 |
| LHFPL3 | 9.95E-62 | 0.4820094 | 0.389 | 0.269 | 3.80E-57 | ExN6 | LHFPL3 |
| NEK112 | 4.28E-58 | 0.28103023 | 0.578 | 0.429 | 1.64E-53 | ExN6 | NEK11 |
| ADD3 | 4.89E-58 | 0.3077402 | 0.691 | 0.574 | 1.87E-53 | ExN6 | ADD3 |
| MARCH12 | 7.34E-58 | 0.3374025 | 0.883 | 0.751 | 2.81E-53 | ExN6 | 1-Mar |
| GMDS-DT | 1.92E-56 | 0.26358783 | 0.7 | 0.56 | 7.34E-52 | ExN6 | GMDS-DT |
| WASF12 | 4.34E-56 | 0.27109929 | 0.784 | 0.634 | 1.66E-51 | ExN6 | WASF1 |
| GARNL3 | 5.26E-56 | 0.2878291 | 0.817 | 0.719 | 2.01E-51 | ExN6 | GARNL3 |
| BACH21 | 1.39E-55 | 0.26522815 | 0.733 | 0.604 | 5.30E-51 | ExN6 | BACH2 |
| ARHGAP263 | 9.80E-55 | 0.25530937 | 0.929 | 0.869 | 3.75E-50 | ExN6 | ARHGAP26 |
| ROBO2 | 1.16E-53 | 0.74152703 | 0.836 | 0.855 | 4.43E-49 | ExN6 | ROBO2 |
| AC110296.11 | 1.75E-53 | 0.29109712 | 0.292 | 0.186 | 6.70E-49 | ExN6 | AC110296.1 |
| STXBP51 | 7.40E-53 | 0.27785126 | 0.877 | 0.825 | 2.83E-48 | ExN6 | STXBP5 |
| GPATCH21 | 7.66E-51 | 0.27561413 | 0.573 | 0.445 | 2.93E-46 | ExN6 | GPATCH2 |
| USP33 | 1.63E-50 | 0.28244989 | 0.789 | 0.693 | 6.23E-46 | ExN6 | USP33 |
| MARK1 | 8.85E-49 | 0.25674332 | 0.776 | 0.674 | 3.38E-44 | ExN6 | MARK1 |
| WSB1 | 3.07E-48 | 0.25397936 | 0.727 | 0.618 | 1.17E-43 | ExN6 | WSB1 |
| TMEM2321 | 2.31E-47 | 0.27622961 | 0.851 | 0.769 | 8.84E-43 | ExN6 | TMEM232 |
| DLGAP1-AS4 | 8.70E-46 | 0.27701896 | 0.466 | 0.349 | 3.32E-41 | ExN6 | DLGAP1-AS4 |
| STXBP5L4 | 3.85E-45 | 0.26993385 | 0.97 | 0.946 | 1.47E-40 | ExN6 | STXBP5L |
| COL12A11 | 8.32E-45 | 0.31060591 | 0.293 | 0.194 | 3.18E-40 | ExN6 | COL12A1 |
| COL19A11 | 1.38E-44 | 0.26332631 | 0.434 | 0.323 | 5.26E-40 | ExN6 | COL19A1 |
| MGAT5 | 3.53E-43 | 0.38941939 | 0.744 | 0.656 | 1.35E-38 | ExN6 | MGAT5 |
| CLVS2 | 1.15E-42 | 0.34720763 | 0.682 | 0.593 | 4.41E-38 | ExN6 | CLVS2 |
| AC067956.14 | 1.09E-41 | 0.28198012 | 0.439 | 0.32 | 4.16E-37 | ExN6 | AC067956.1 |
| FMN13 | 1.90E-40 | 0.28708993 | 0.868 | 0.753 | 7.27E-36 | ExN6 | FMN1 |
| ITPR2 | 8.19E-38 | 0.41415542 | 0.456 | 0.358 | 3.13E-33 | ExN6 | ITPR2 |
| LRRC4C1 | 2.69E-35 | 0.27952212 | 0.929 | 0.887 | 1.03E-30 | ExN6 | LRRC4C |
| RNF144A2 | 1.04E-33 | 0.29838605 | 0.797 | 0.739 | 3.96E-29 | ExN6 | RNF144A |
| DYNLL11 | 1.62E-33 | 0.30628124 | 0.543 | 0.425 | 6.18E-29 | ExN6 | DYNLL1 |
| AQP4-AS14 | 3.39E-32 | 0.29361825 | 0.597 | 0.498 | 1.30E-27 | ExN6 | AQP4-AS1 |
| TENM42 | 1.45E-31 | 0.40346928 | 0.914 | 0.856 | 5.55E-27 | ExN6 | TENM4 |
| TSHZ21 | 1.37E-26 | 0.49664426 | 0.496 | 0.396 | 5.24E-22 | ExN6 | TSHZ2 |
| CNTN6 | 3.58E-26 | 0.28977552 | 0.301 | 0.23 | 1.37E-21 | ExN6 | CNTN6 |
| VPS13D2 | 7.30E-25 | 0.32986601 | 0.806 | 0.749 | 2.79E-20 | ExN6 | VPS13D |
| GPC52 | 1.45E-24 | 0.39864632 | 0.496 | 0.419 | 5.52E-20 | ExN6 | GPC5 |
| CSGALNACT1 | 1.82E-24 | 0.30539951 | 0.666 | 0.61 | 6.96E-20 | ExN6 | CSGALNACT1 |
| TRHDE2 | 3.70E-22 | 0.45702795 | 0.518 | 0.523 | 1.42E-17 | ExN6 | TRHDE |
| SORCS21 | 2.47E-19 | 0.59457095 | 0.367 | 0.323 | 9.44E-15 | ExN6 | SORCS2 |
| NWD24 | 8.28E-17 | 0.32533157 | 0.337 | 0.482 | 3.17E-12 | ExN6 | NWD2 |
| CDH82 | 6.62E-12 | 0.25467186 | 0.67 | 0.663 | 2.53E-07 | ExN6 | CDH8 |
| CUX12 | 1.44E-11 | 0.31739571 | 0.655 | 0.738 | 5.50E-07 | ExN6 | CUX1 |
| CPNE82 | 0.00027899 | 0.25127977 | 0.462 | 0.483 | 1 | ExN6 | CPNE8 |
| SLC1A3 | 0 | 2.66912104 | 0.896 | 0.137 | 0 | ExN7 | SLC1A3 |
| LRMDA1 | 0 | 2.65754986 | 0.925 | 0.186 | 0 | ExN7 | LRMDA |
| ARHGAP24 | 0 | 2.43769254 | 0.863 | 0.147 | 0 | ExN7 | ARHGAP24 |
| DOCK8 | 0 | 2.19425947 | 0.839 | 0.022 | 0 | ExN7 | DOCK8 |
| APBB1IP | 0 | 2.06101785 | 0.834 | 0.023 | 0 | ExN7 | APBB1IP |
| RUNX1 | 0 | 2.00686112 | 0.821 | 0.138 | 0 | ExN7 | RUNX1 |
| TBXAS1 | 0 | 1.82297165 | 0.798 | 0.028 | 0 | ExN7 | TBXAS1 |
| FYB1 | 0 | 1.82251326 | 0.736 | 0.013 | 0 | ExN7 | FYB1 |

| | | | | | | |
|---|---|---|---|---|---|---|
| LNCAROD | 0 | 1.80756707 | 0.679 | 0.012 | 0 | ExN7 | LNCAROD |
| ARHGAP15 | 0 | 1.66997236 | 0.705 | 0.066 | 0 | ExN7 | ARHGAP15 |
| DISC1 | 0 | 1.62408202 | 0.764 | 0.135 | 0 | ExN7 | DISC1 |
| DOCK2 | 0 | 1.58366265 | 0.756 | 0.049 | 0 | ExN7 | DOCK2 |
| ADAM28 | 0 | 1.56609731 | 0.699 | 0.018 | 0 | ExN7 | ADAM28 |
| SLCO2B1 | 0 | 1.56292404 | 0.723 | 0.027 | 0 | ExN7 | SLCO2B1 |
| INPP5D | 0 | 1.51375754 | 0.687 | 0.043 | 0 | ExN7 | INPP5D |
| ZFP36L1 | 0 | 1.46829301 | 0.63 | 0.039 | 0 | ExN7 | ZFP36L1 |
| PTPRC | 0 | 1.38773222 | 0.697 | 0.011 | 0 | ExN7 | PTPRC |
| C3 | 0 | 1.37924047 | 0.648 | 0.019 | 0 | ExN7 | C3 |
| CD74 | 0 | 1.33224289 | 0.609 | 0.025 | 0 | ExN7 | CD74 |
| RBM47 | 0 | 1.33111837 | 0.642 | 0.027 | 0 | ExN7 | RBM47 |
| PALD1 | 0 | 1.30859883 | 0.648 | 0.065 | 0 | ExN7 | PALD1 |
| CSF1R | 0 | 1.28543125 | 0.65 | 0.01 | 0 | ExN7 | CSF1R |
| SYK | 0 | 1.25369167 | 0.632 | 0.008 | 0 | ExN7 | SYK |
| FLI1 | 0 | 1.21664221 | 0.63 | 0.009 | 0 | ExN7 | FLI1 |
| IKZF1 | 0 | 1.21421848 | 0.609 | 0.007 | 0 | ExN7 | IKZF1 |
| RHBDF2 | 0 | 1.16937262 | 0.614 | 0.015 | 0 | ExN7 | RHBDF2 |
| LYN | 0 | 1.14068188 | 0.567 | 0.019 | 0 | ExN7 | LYN |
| LINC01374 | 0 | 1.0731139 | 0.492 | 0.029 | 0 | ExN7 | LINC01374 |
| LPCAT2 | 0 | 1.06240961 | 0.554 | 0.058 | 0 | ExN7 | LPCAT2 |
| BNC2 | 0 | 1.04251809 | 0.5 | 0.02 | 0 | ExN7 | BNC2 |
| P2RY12 | 0 | 1.02063418 | 0.523 | 0.021 | 0 | ExN7 | P2RY12 |
| BLNK | 0 | 1.01647357 | 0.508 | 0.009 | 0 | ExN7 | BLNK |
| TLR2 | 0 | 1.01092058 | 0.521 | 0.007 | 0 | ExN7 | TLR2 |
| A2M | 0 | 0.96492205 | 0.544 | 0.033 | 0 | ExN7 | A2M |
| ZFP36L2 | 0 | 0.95201849 | 0.417 | 0.02 | 0 | ExN7 | ZFP36L2 |
| TGFBR2 | 0 | 0.94958379 | 0.5 | 0.01 | 0 | ExN7 | TGFBR2 |
| SP100 | 0 | 0.9479557 | 0.526 | 0.016 | 0 | ExN7 | SP100 |
| SCIN | 0 | 0.93687723 | 0.422 | 0.006 | 0 | ExN7 | SCIN |
| CPED1 | 0 | 0.93392284 | 0.487 | 0.029 | 0 | ExN7 | CPED1 |
| LPAR6 | 0 | 0.92917209 | 0.5 | 0.018 | 0 | ExN7 | LPAR6 |
| SAMSN1 | 0 | 0.92915596 | 0.49 | 0.016 | 0 | ExN7 | SAMSN1 |
| LY86 | 0 | 0.92759159 | 0.557 | 0.033 | 0 | ExN7 | LY86 |
| NFATC2 | 0 | 0.90280369 | 0.497 | 0.015 | 0 | ExN7 | NFATC2 |
| ARHGAP25 | 0 | 0.89358696 | 0.541 | 0.062 | 0 | ExN7 | ARHGAP25 |
| SLC2A5 | 0 | 0.88611614 | 0.477 | 0.012 | 0 | ExN7 | SLC2A5 |
| MSR1 | 0 | 0.88347081 | 0.409 | 0.021 | 0 | ExN7 | MSR1 |
| IFI16 | 0 | 0.87470993 | 0.534 | 0.059 | 0 | ExN7 | IFI16 |
| PIK3AP1 | 0 | 0.87368426 | 0.482 | 0.013 | 0 | ExN7 | PIK3AP1 |
| AL357522.1 | 0 | 0.86891451 | 0.448 | 0.016 | 0 | ExN7 | AL357522.1 |
| ENTPD1 | 0 | 0.86123218 | 0.495 | 0.025 | 0 | ExN7 | ENTPD1 |
| RREB1 | 0 | 0.85605908 | 0.484 | 0.014 | 0 | ExN7 | RREB1 |
| C1QB | 0 | 0.85085361 | 0.43 | 0.009 | 0 | ExN7 | C1QB |
| WDFY4 | 0 | 0.84774736 | 0.497 | 0.01 | 0 | ExN7 | WDFY4 |
| MYO1F | 0 | 0.80298981 | 0.508 | 0.023 | 0 | ExN7 | MYO1F |
| LAPTM5 | 0 | 0.78070717 | 0.469 | 0.011 | 0 | ExN7 | LAPTM5 |
| ALPK1 | 0 | 0.76068781 | 0.443 | 0.042 | 0 | ExN7 | ALPK1 |
| CSF2RA | 0 | 0.75711479 | 0.446 | 0.015 | 0 | ExN7 | CSF2RA |
| CIITA | 0 | 0.75702589 | 0.415 | 0.015 | 0 | ExN7 | CIITA |
| CD53 | 0 | 0.75650941 | 0.5 | 0.052 | 0 | ExN7 | CD53 |
| MS4A6A | 0 | 0.74717731 | 0.368 | 0.003 | 0 | ExN7 | MS4A6A |
| IRAK3 | 0 | 0.74008249 | 0.427 | 0.038 | 0 | ExN7 | IRAK3 |
| SLC11A1 | 0 | 0.73760044 | 0.391 | 0.017 | 0 | ExN7 | SLC11A1 |
| PIK3R5 | 0 | 0.73361251 | 0.409 | 0.005 | 0 | ExN7 | PIK3R5 |
| VSIG4 | 0 | 0.71308174 | 0.326 | 0.003 | 0 | ExN7 | VSIG4 |
| AL392172.2 | 0 | 0.7112662 | 0.35 | 0.003 | 0 | ExN7 | AL392172.2 |
| C1QC | 0 | 0.7053793 | 0.347 | 0.005 | 0 | ExN7 | C1QC |
| ALOX5 | 0 | 0.70452323 | 0.422 | 0.018 | 0 | ExN7 | ALOX5 |
| CX3CR1 | 0 | 0.69917188 | 0.378 | 0.004 | 0 | ExN7 | CX3CR1 |
| HCLS1 | 0 | 0.69911772 | 0.415 | 0.016 | 0 | ExN7 | HCLS1 |
| PARP14 | 0 | 0.6985809 | 0.412 | 0.016 | 0 | ExN7 | PARP14 |
| CD86 | 0 | 0.69818889 | 0.381 | 0.003 | 0 | ExN7 | CD86 |

| | | | | | | | |
|---|---|---|---|---|---|---|---|
| OLR1 | 0 | 0.69750217 | 0.381 | 0.021 | 0 | ExN7 | OLR1 |
| IL18 | 0 | 0.69693337 | 0.404 | 0.014 | 0 | ExN7 | IL18 |
| FGD2 | 0 | 0.68266452 | 0.404 | 0.005 | 0 | ExN7 | FGD2 |
| CYBB | 0 | 0.68113246 | 0.399 | 0.025 | 0 | ExN7 | CYBB |
| TNFRSF1B | 0 | 0.68045668 | 0.396 | 0.006 | 0 | ExN7 | TNFRSF1B |
| HLA-DRA | 0 | 0.67346048 | 0.347 | 0.011 | 0 | ExN7 | HLA-DRA |
| NIBAN1 | 0 | 0.67338814 | 0.324 | 0.013 | 0 | ExN7 | NIBAN1 |
| ITGAX | 0 | 0.65318694 | 0.399 | 0.004 | 0 | ExN7 | ITGAX |
| CSF3R | 0 | 0.64432321 | 0.358 | 0.004 | 0 | ExN7 | CSF3R |
| IRF8 | 0 | 0.62759117 | 0.319 | 0.004 | 0 | ExN7 | IRF8 |
| IL13RA1 | 0 | 0.62476447 | 0.394 | 0.009 | 0 | ExN7 | IL13RA1 |
| NCKAP1L | 0 | 0.62084268 | 0.381 | 0.012 | 0 | ExN7 | NCKAP1L |
| VSIR | 0 | 0.61058911 | 0.386 | 0.009 | 0 | ExN7 | VSIR |
| SH3TC1 | 0 | 0.60243402 | 0.378 | 0.004 | 0 | ExN7 | SH3TC1 |
| TMEM156 | 0 | 0.60236762 | 0.355 | 0.009 | 0 | ExN7 | TMEM156 |
| STAB1 | 0 | 0.59858114 | 0.337 | 0.009 | 0 | ExN7 | STAB1 |
| RGS10 | 0 | 0.59683493 | 0.347 | 0.007 | 0 | ExN7 | RGS10 |
| MYOF | 0 | 0.59457933 | 0.337 | 0.01 | 0 | ExN7 | MYOF |
| SRGN | 0 | 0.57672786 | 0.316 | 0.007 | 0 | ExN7 | SRGN |
| KCNQ1 | 0 | 0.57308484 | 0.36 | 0.023 | 0 | ExN7 | KCNQ1 |
| HLA-DRB1 | 0 | 0.5596246 | 0.311 | 0.013 | 0 | ExN7 | HLA-DRB1 |
| HLA-E | 0 | 0.55784632 | 0.342 | 0.009 | 0 | ExN7 | HLA-E |
| TLR1 | 0 | 0.55362741 | 0.35 | 0.013 | 0 | ExN7 | TLR1 |
| CYBA | 0 | 0.54037583 | 0.329 | 0.008 | 0 | ExN7 | CYBA |
| AL034397.3 | 0 | 0.50303087 | 0.259 | 0.002 | 0 | ExN7 | AL034397.3 |
| ITGAM | 0 | 0.50291247 | 0.337 | 0.005 | 0 | ExN7 | ITGAM |
| C1QA | 0 | 0.49359914 | 0.28 | 0.006 | 0 | ExN7 | C1QA |
| TFEC | 0 | 0.49066409 | 0.339 | 0.016 | 0 | ExN7 | TFEC |
| IL4R | 0 | 0.48920878 | 0.313 | 0.012 | 0 | ExN7 | IL4R |
| PARVG | 0 | 0.46819922 | 0.303 | 0.015 | 0 | ExN7 | PARVG |
| VAV1 | 0 | 0.46765365 | 0.308 | 0.011 | 0 | ExN7 | VAV1 |
| LCP1 | 0 | 0.46690885 | 0.264 | 0.012 | 0 | ExN7 | LCP1 |
| CMTM7 | 0 | 0.466034 | 0.324 | 0.019 | 0 | ExN7 | CMTM7 |
| GAB3 | 0 | 0.45851647 | 0.285 | 0.005 | 0 | ExN7 | GAB3 |
| AC012150.1 | 0 | 0.45719451 | 0.264 | 0.011 | 0 | ExN7 | AC012150.1 |
| CYTH4 | 0 | 0.45090116 | 0.254 | 0.005 | 0 | ExN7 | CYTH4 |
| PGM5 | 0 | 0.45046452 | 0.264 | 0.015 | 0 | ExN7 | PGM5 |
| CD84 | 0 | 0.43532947 | 0.269 | 0.004 | 0 | ExN7 | CD84 |
| LINC01094 | 0 | 0.42481365 | 0.269 | 0.007 | 0 | ExN7 | LINC01094 |
| BIN2 | 0 | 0.38666582 | 0.256 | 0.005 | 0 | ExN7 | BIN2 |
| ITGB2 | 0 | 0.37936986 | 0.254 | 0.009 | 0 | ExN7 | ITGB2 |
| HCK | 2.08E-304 | 0.57962687 | 0.347 | 0.026 | 7.96E-300 | ExN7 | HCK |
| TLR5 | 1.16E-296 | 0.39353462 | 0.259 | 0.015 | 4.42E-292 | ExN7 | TLR5 |
| RCSD1 | 1.02E-284 | 0.85286871 | 0.484 | 0.056 | 3.88E-280 | ExN7 | RCSD1 |
| SPP1 | 1.99E-277 | 2.49630685 | 0.604 | 0.093 | 7.59E-273 | ExN7 | SPP1 |
| AL163541.1 | 7.26E-275 | 1.66407285 | 0.472 | 0.056 | 2.77E-270 | ExN7 | AL163541.1 |
| SLA | 5.62E-272 | 0.6955482 | 0.386 | 0.037 | 2.15E-267 | ExN7 | SLA |
| AOAH | 6.45E-272 | 1.19510293 | 0.65 | 0.11 | 2.47E-267 | ExN7 | AOAH |
| TGFBR11 | 1.47E-260 | 1.15701871 | 0.624 | 0.104 | 5.62E-256 | ExN7 | TGFBR1 |
| PADI2 | 4.28E-258 | 0.94113273 | 0.427 | 0.048 | 1.64E-253 | ExN7 | PADI2 |
| HLA-DPB1 | 5.39E-258 | 0.41794524 | 0.272 | 0.019 | 2.06E-253 | ExN7 | HLA-DPB1 |
| CARD11 | 7.66E-241 | 0.45489091 | 0.272 | 0.02 | 2.93E-236 | ExN7 | CARD11 |
| MSN | 6.78E-240 | 0.53104421 | 0.36 | 0.036 | 2.59E-235 | ExN7 | MSN |
| ST6GAL12 | 3.13E-238 | 2.05165963 | 0.863 | 0.272 | 1.20E-233 | ExN7 | ST6GAL1 |
| RIN3 | 1.40E-236 | 0.8817012 | 0.552 | 0.086 | 5.34E-232 | ExN7 | RIN3 |
| SUCLG2 | 3.26E-235 | 0.72924289 | 0.474 | 0.064 | 1.25E-230 | ExN7 | SUCLG2 |
| ABCC4 | 8.69E-229 | 1.21156425 | 0.637 | 0.125 | 3.32E-224 | ExN7 | ABCC4 |
| SP110 | 5.64E-227 | 0.42216954 | 0.262 | 0.02 | 2.16E-222 | ExN7 | SP110 |
| MERTK1 | 1.35E-225 | 1.09400962 | 0.596 | 0.106 | 5.16E-221 | ExN7 | MERTK |
| CCDC26 | 2.98E-225 | 1.10517439 | 0.376 | 0.043 | 1.14E-220 | ExN7 | CCDC26 |
| LAT2 | 1.63E-219 | 0.50611664 | 0.355 | 0.038 | 6.25E-215 | ExN7 | LAT2 |
| MAML2 | 2.65E-219 | 1.75011183 | 0.837 | 0.231 | 1.01E-214 | ExN7 | MAML2 |
| LCP2 | 1.26E-212 | 0.51791321 | 0.347 | 0.037 | 4.82E-208 | ExN7 | LCP2 |

| | | | | | | | |
|---|---|---|---|---|---|---|---|
| TNS3 | 2.84E-211 | 0.93797045 | 0.596 | 0.108 | 1.09E-206 | ExN7 | TNS3 |
| DLEU1 | 2.98E-204 | 1.60195026 | 0.65 | 0.147 | 1.14E-199 | ExN7 | DLEU1 |
| CTSC | 6.03E-194 | 0.51289059 | 0.332 | 0.038 | 2.31E-189 | ExN7 | CTSC |
| MYO15B | 7.10E-194 | 0.38291327 | 0.301 | 0.03 | 2.71E-189 | ExN7 | MYO15B |
| PARP4 | 4.83E-193 | 0.42104492 | 0.29 | 0.029 | 1.85E-188 | ExN7 | PARP4 |
| NEAT1 | 8.10E-192 | 1.95880602 | 0.943 | 0.38 | 3.10E-187 | ExN7 | NEAT1 |
| WIPF1 | 6.21E-187 | 0.65828739 | 0.443 | 0.067 | 2.37E-182 | ExN7 | WIPF1 |
| PLXDC24 | 1.08E-184 | 2.14231352 | 0.977 | 0.611 | 4.12E-180 | ExN7 | PLXDC2 |
| PRKCH | 7.19E-180 | 0.58975865 | 0.391 | 0.055 | 2.75E-175 | ExN7 | PRKCH |
| CPVL | 1.13E-177 | 0.57943618 | 0.332 | 0.041 | 4.33E-173 | ExN7 | CPVL |
| APOE | 9.39E-174 | 0.91333313 | 0.456 | 0.077 | 3.59E-169 | ExN7 | APOE |
| NHSL1 | 2.75E-171 | 0.91045989 | 0.352 | 0.048 | 1.05E-166 | ExN7 | NHSL1 |
| SFMBT2 | 1.41E-166 | 1.74544582 | 0.904 | 0.382 | 5.39E-162 | ExN7 | SFMBT2 |
| RNASET2 | 1.48E-165 | 1.29210694 | 0.775 | 0.264 | 5.66E-161 | ExN7 | RNASET2 |
| PRAM1 | 9.32E-164 | 0.38026653 | 0.264 | 0.028 | 3.56E-159 | ExN7 | PRAM1 |
| TMCC3 | 4.59E-162 | 0.46974409 | 0.313 | 0.039 | 1.75E-157 | ExN7 | TMCC3 |
| PREX1 | 6.32E-162 | 1.12911234 | 0.699 | 0.2 | 2.42E-157 | ExN7 | PREX1 |
| ITPR21 | 3.46E-161 | 1.64621723 | 0.896 | 0.361 | 1.32E-156 | ExN7 | ITPR2 |
| CPM | 1.30E-160 | 0.56947524 | 0.301 | 0.036 | 4.97E-156 | ExN7 | CPM |
| LHFPL2 | 9.99E-157 | 1.45343676 | 0.749 | 0.244 | 3.82E-152 | ExN7 | LHFPL2 |
| NPL | 3.41E-156 | 0.79985377 | 0.51 | 0.106 | 1.30E-151 | ExN7 | NPL |
| ATP8B41 | 1.70E-155 | 1.2100217 | 0.731 | 0.23 | 6.48E-151 | ExN7 | ATP8B4 |
| SRGAP2 | 1.77E-150 | 1.48716981 | 0.902 | 0.483 | 6.77E-146 | ExN7 | SRGAP2 |
| FKBP52 | 9.64E-148 | 1.3761184 | 0.78 | 0.261 | 3.68E-143 | ExN7 | FKBP5 |
| BMP2K | 5.62E-146 | 1.4284699 | 0.788 | 0.31 | 2.15E-141 | ExN7 | BMP2K |
| CHST11 | 3.20E-141 | 1.457001 | 0.953 | 0.513 | 1.22E-136 | ExN7 | CHST11 |
| ARHGAP31 | 7.80E-140 | 0.46268935 | 0.332 | 0.048 | 2.98E-135 | ExN7 | ARHGAP31 |
| CHD7 | 1.17E-137 | 0.73334635 | 0.526 | 0.113 | 4.48E-133 | ExN7 | CHD7 |
| ELF1 | 2.79E-136 | 0.63694996 | 0.479 | 0.1 | 1.07E-131 | ExN7 | ELF1 |
| HLA-B | 5.62E-136 | 0.53215961 | 0.345 | 0.054 | 2.15E-131 | ExN7 | HLA-B |
| ADAP2 | 1.29E-133 | 0.41168209 | 0.316 | 0.047 | 4.95E-129 | ExN7 | ADAP2 |
| HLA-DMA | 2.85E-132 | 0.39380721 | 0.277 | 0.037 | 1.09E-127 | ExN7 | HLA-DMA |
| RAB31 | 1.34E-128 | 0.791968 | 0.539 | 0.131 | 5.12E-124 | ExN7 | RAB31 |
| ADCY7 | 1.82E-128 | 0.48005806 | 0.342 | 0.057 | 6.97E-124 | ExN7 | ADCY7 |
| SPATA13 | 4.45E-126 | 0.77164462 | 0.56 | 0.143 | 1.70E-121 | ExN7 | SPATA13 |
| SLC9A9 | 8.70E-126 | 1.25224668 | 0.837 | 0.351 | 3.32E-121 | ExN7 | SLC9A9 |
| B3GNT5 | 3.62E-125 | 0.39653124 | 0.256 | 0.034 | 1.38E-120 | ExN7 | B3GNT5 |
| FGD4 | 1.22E-124 | 1.14295722 | 0.811 | 0.332 | 4.66E-120 | ExN7 | FGD4 |
| ARHGAP4 | 6.22E-120 | 0.32700841 | 0.251 | 0.033 | 2.38E-115 | ExN7 | ARHGAP4 |
| B2M | 8.35E-120 | 0.72014091 | 0.518 | 0.125 | 3.19E-115 | ExN7 | B2M |
| PRKD3 | 6.93E-117 | 0.43947659 | 0.324 | 0.055 | 2.65E-112 | ExN7 | PRKD3 |
| KCNK13 | 7.09E-117 | 0.60295799 | 0.368 | 0.069 | 2.71E-112 | ExN7 | KCNK13 |
| ZFHX31 | 3.02E-116 | 1.01422883 | 0.769 | 0.291 | 1.15E-111 | ExN7 | ZFHX3 |
| SRGAP2B | 1.16E-115 | 0.93495005 | 0.684 | 0.249 | 4.43E-111 | ExN7 | SRGAP2B |
| IL17RA | 2.27E-115 | 0.64113644 | 0.459 | 0.107 | 8.69E-111 | ExN7 | IL17RA |
| CARD8 | 1.01E-113 | 0.32782552 | 0.282 | 0.043 | 3.85E-109 | ExN7 | CARD8 |
| ARPC1B | 6.30E-113 | 0.37846291 | 0.282 | 0.044 | 2.41E-108 | ExN7 | ARPC1B |
| FAM149A | 8.46E-113 | 0.80032582 | 0.518 | 0.143 | 3.24E-108 | ExN7 | FAM149A |
| RPS6KA1 | 5.44E-112 | 0.35190908 | 0.256 | 0.037 | 2.08E-107 | ExN7 | RPS6KA1 |
| DOCK43 | 1.60E-110 | 1.33313 | 0.987 | 0.827 | 6.12E-106 | ExN7 | DOCK4 |
| EPS8 | 9.07E-110 | 0.65929146 | 0.497 | 0.12 | 3.47E-105 | ExN7 | EPS8 |
| ADGRG1 | 9.70E-110 | 0.62523181 | 0.415 | 0.092 | 3.71E-105 | ExN7 | ADGRG1 |
| SOCS6 | 2.97E-108 | 0.81234506 | 0.557 | 0.167 | 1.13E-103 | ExN7 | SOCS6 |
| DENND34 | 9.88E-108 | 1.12510033 | 0.764 | 0.345 | 3.78E-103 | ExN7 | DENND3 |
| FRMD4B | 1.85E-107 | 0.74873188 | 0.596 | 0.168 | 7.09E-103 | ExN7 | FRMD4B |
| ZNF710 | 1.78E-106 | 0.70006895 | 0.536 | 0.152 | 6.79E-102 | ExN7 | ZNF710 |
| IFNGR1 | 5.58E-106 | 0.8832449 | 0.624 | 0.219 | 2.13E-101 | ExN7 | IFNGR1 |
| ELMO11 | 1.22E-104 | 1.26692403 | 0.93 | 0.655 | 4.66E-100 | ExN7 | ELMO1 |
| BHLHE41 | 1.08E-102 | 0.69872479 | 0.505 | 0.14 | 4.13E-98 | ExN7 | BHLHE41 |
| LPAR1 | 1.96E-102 | 0.43063062 | 0.345 | 0.065 | 7.51E-98 | ExN7 | LPAR1 |
| LINC01141 | 1.85E-101 | 0.45209628 | 0.329 | 0.063 | 7.06E-97 | ExN7 | LINC01141 |
| TXNIP | 5.93E-100 | 0.46552275 | 0.277 | 0.046 | 2.27E-95 | ExN7 | TXNIP |
| ARHGAP223 | 8.97E-100 | 1.13792471 | 0.705 | 0.307 | 3.43E-95 | ExN7 | ARHGAP22 |

| | | | | | | | |
|---|---|---|---|---|---|---|---|
| MAML3 | 9.57E-99 | 1.08823392 | 0.813 | 0.361 | 3.66E-94 | ExN7 | MAML3 |
| RUBCNL | 3.94E-98 | 0.49290721 | 0.373 | 0.082 | 1.51E-93 | ExN7 | RUBCNL |
| MEF2A1 | 8.24E-98 | 1.09178119 | 0.93 | 0.75 | 3.15E-93 | ExN7 | MEF2A |
| USP6NL | 3.19E-96 | 0.69364843 | 0.513 | 0.151 | 1.22E-91 | ExN7 | USP6NL |
| ETV6 | 5.66E-96 | 1.08357149 | 0.855 | 0.466 | 2.16E-91 | ExN7 | ETV6 |
| RNF144B | 2.22E-95 | 0.46376899 | 0.298 | 0.054 | 8.47E-91 | ExN7 | RNF144B |
| GNB41 | 1.02E-93 | 0.51970697 | 0.448 | 0.115 | 3.89E-89 | ExN7 | GNB4 |
| SRGAP11 | 7.11E-93 | 0.99222429 | 0.668 | 0.253 | 2.72E-88 | ExN7 | SRGAP1 |
| RUNX2 | 2.41E-92 | 0.59336394 | 0.56 | 0.162 | 9.21E-88 | ExN7 | RUNX2 |
| GCNT2 | 3.43E-92 | 0.44969851 | 0.355 | 0.075 | 1.31E-87 | ExN7 | GCNT2 |
| ANOS1 | 3.64E-91 | 0.40717599 | 0.339 | 0.07 | 1.39E-86 | ExN7 | ANOS1 |
| SAT1 | 2.03E-89 | 1.19033373 | 0.754 | 0.366 | 7.77E-85 | ExN7 | SAT1 |
| AKAP133 | 2.83E-89 | 0.97619183 | 0.756 | 0.366 | 1.08E-84 | ExN7 | AKAP13 |
| MYO1E | 7.14E-89 | 0.7896414 | 0.352 | 0.079 | 2.73E-84 | ExN7 | MYO1E |
| ANKRD44 | 8.30E-88 | 1.04725616 | 0.816 | 0.427 | 3.17E-83 | ExN7 | ANKRD44 |
| MGAT4A | 1.18E-87 | 0.95884061 | 0.837 | 0.468 | 4.50E-83 | ExN7 | MGAT4A |
| BACH1 | 3.85E-86 | 0.89384828 | 0.632 | 0.233 | 1.47E-81 | ExN7 | BACH1 |
| PLA2G4A | 3.61E-84 | 0.48955663 | 0.37 | 0.089 | 1.38E-79 | ExN7 | PLA2G4A |
| PICALM | 6.74E-84 | 0.98271351 | 0.909 | 0.674 | 2.58E-79 | ExN7 | PICALM |
| TRIM221 | 2.24E-83 | 0.36743599 | 0.29 | 0.057 | 8.56E-79 | ExN7 | TRIM22 |
| SUSD6 | 9.95E-83 | 0.75675265 | 0.64 | 0.264 | 3.80E-78 | ExN7 | SUSD6 |
| FOXN3 | 1.07E-82 | 1.07836369 | 0.935 | 0.707 | 4.10E-78 | ExN7 | FOXN3 |
| ARHGAP6 | 4.48E-82 | 0.72719032 | 0.541 | 0.174 | 1.71E-77 | ExN7 | ARHGAP6 |
| RASGEF1C1 | 9.67E-82 | 0.76436516 | 0.487 | 0.156 | 3.70E-77 | ExN7 | RASGEF1C |
| IPCEF13 | 1.67E-81 | 1.05657144 | 0.785 | 0.402 | 6.36E-77 | ExN7 | IPCEF1 |
| CTSS | 5.22E-81 | 0.41367535 | 0.334 | 0.077 | 1.99E-76 | ExN7 | CTSS |
| ZC3HAV1 | 3.96E-80 | 0.38547652 | 0.262 | 0.049 | 1.51E-75 | ExN7 | ZC3HAV1 |
| RASSF8 | 3.77E-77 | 0.73528897 | 0.523 | 0.177 | 1.44E-72 | ExN7 | RASSF8 |
| CYFIP1 | 8.61E-76 | 0.91571922 | 0.801 | 0.46 | 3.29E-71 | ExN7 | CYFIP1 |
| SORL13 | 1.42E-75 | 1.05957934 | 0.912 | 0.679 | 5.42E-71 | ExN7 | SORL1 |
| IL6R | 7.27E-74 | 0.38078388 | 0.329 | 0.078 | 2.78E-69 | ExN7 | IL6R |
| TGFB1 | 1.99E-73 | 0.38155622 | 0.288 | 0.063 | 7.61E-69 | ExN7 | TGFB1 |
| DOCK11 | 6.23E-72 | 0.62348754 | 0.585 | 0.212 | 2.38E-67 | ExN7 | DOCK11 |
| AC025569.1 | 6.60E-72 | 0.36421489 | 0.251 | 0.051 | 2.52E-67 | ExN7 | AC025569.1 |
| PLSCR1 | 1.96E-71 | 0.49524827 | 0.394 | 0.113 | 7.50E-67 | ExN7 | PLSCR1 |
| GSN | 4.70E-70 | 0.54982533 | 0.484 | 0.164 | 1.80E-65 | ExN7 | GSN |
| LPP | 3.35E-69 | 0.8222259 | 0.832 | 0.474 | 1.28E-64 | ExN7 | LPP |
| AKNA | 3.82E-69 | 0.38079509 | 0.339 | 0.087 | 1.46E-64 | ExN7 | AKNA |
| OTULINL | 1.01E-68 | 0.42569943 | 0.378 | 0.108 | 3.88E-64 | ExN7 | OTULINL |
| RAD51B | 4.24E-68 | 0.82835587 | 0.64 | 0.282 | 1.62E-63 | ExN7 | RAD51B |
| COTL1 | 8.51E-68 | 0.44571168 | 0.365 | 0.1 | 3.25E-63 | ExN7 | COTL1 |
| PTPRJ | 1.31E-67 | 0.97532338 | 0.863 | 0.625 | 5.02E-63 | ExN7 | PTPRJ |
| SPTLC2 | 3.27E-67 | 0.77406851 | 0.63 | 0.291 | 1.25E-62 | ExN7 | SPTLC2 |
| DOCK5 | 3.47E-67 | 0.50991298 | 0.352 | 0.092 | 1.32E-62 | ExN7 | DOCK5 |
| FMNL3 | 4.09E-67 | 0.71098914 | 0.56 | 0.228 | 1.56E-62 | ExN7 | FMNL3 |
| CTSB | 2.13E-66 | 0.8352599 | 0.689 | 0.352 | 8.14E-62 | ExN7 | CTSB |
| HSPA1B | 2.43E-66 | 0.44569233 | 0.311 | 0.076 | 9.27E-62 | ExN7 | HSPA1B |
| PELI1 | 1.59E-64 | 0.72962377 | 0.591 | 0.254 | 6.09E-60 | ExN7 | PELI1 |
| RYR1 | 1.90E-64 | 0.59005752 | 0.575 | 0.235 | 7.28E-60 | ExN7 | RYR1 |
| RNF213 | 9.93E-64 | 0.55822779 | 0.513 | 0.198 | 3.80E-59 | ExN7 | RNF213 |
| HLA-C | 1.55E-63 | 0.37251848 | 0.282 | 0.067 | 5.92E-59 | ExN7 | HLA-C |
| SWAP70 | 2.39E-63 | 0.45331254 | 0.407 | 0.126 | 9.12E-59 | ExN7 | SWAP70 |
| TBC1D22A | 4.04E-63 | 0.77198453 | 0.881 | 0.62 | 1.54E-58 | ExN7 | TBC1D22A |
| LINC02798 | 6.81E-63 | 0.76231702 | 0.544 | 0.212 | 2.60E-58 | ExN7 | LINC02798 |
| NAIP | 2.96E-61 | 0.50855038 | 0.451 | 0.158 | 1.13E-56 | ExN7 | NAIP |
| PAG12 | 2.02E-60 | 0.81660663 | 0.702 | 0.374 | 7.72E-56 | ExN7 | PAG1 |
| MAF | 2.19E-60 | 0.51432217 | 0.531 | 0.186 | 8.35E-56 | ExN7 | MAF |
| EPB41L22 | 2.51E-60 | 0.81726744 | 0.933 | 0.713 | 9.59E-56 | ExN7 | EPB41L2 |
| EHBP1L1 | 2.70E-60 | 0.33413133 | 0.332 | 0.091 | 1.03E-55 | ExN7 | EHBP1L1 |
| LDLRAD41 | 3.63E-60 | 0.81530357 | 0.956 | 0.781 | 1.39E-55 | ExN7 | LDLRAD4 |
| STK10 | 3.40E-58 | 0.43283534 | 0.324 | 0.091 | 1.30E-53 | ExN7 | STK10 |
| NCK21 | 6.53E-58 | 0.90827244 | 0.744 | 0.444 | 2.50E-53 | ExN7 | NCK2 |
| PYGL | 8.91E-58 | 0.30400076 | 0.259 | 0.062 | 3.41E-53 | ExN7 | PYGL |

| | | | | | | | |
|---|---|---|---|---|---|---|---|
| HSPA1A | 2.29E-57 | 0.71946557 | 0.464 | 0.168 | 8.77E-53 | ExN7 | HSPA1A |
| USP53 | 3.81E-57 | 0.67187511 | 0.585 | 0.271 | 1.46E-52 | ExN7 | USP53 |
| PLD11 | 1.27E-56 | 0.47074334 | 0.402 | 0.129 | 4.85E-52 | ExN7 | PLD1 |
| SLC2A9 | 5.80E-56 | 0.36186319 | 0.303 | 0.081 | 2.22E-51 | ExN7 | SLC2A9 |
| KCTD121 | 1.36E-55 | 0.37236391 | 0.293 | 0.079 | 5.21E-51 | ExN7 | KCTD12 |
| PRKX | 3.43E-55 | 0.27624131 | 0.264 | 0.064 | 1.31E-50 | ExN7 | PRKX |
| ABR | 3.95E-55 | 0.75115709 | 0.915 | 0.717 | 1.51E-50 | ExN7 | ABR |
| DOCK10 | 4.32E-55 | 0.64784405 | 0.811 | 0.405 | 1.65E-50 | ExN7 | DOCK10 |
| ACSL1 | 5.56E-55 | 0.93390254 | 0.503 | 0.213 | 2.13E-50 | ExN7 | ACSL1 |
| ITGB5 | 1.18E-54 | 0.28125114 | 0.254 | 0.06 | 4.50E-50 | ExN7 | ITGB5 |
| QKI | 1.87E-54 | 0.79383995 | 0.933 | 0.673 | 7.14E-50 | ExN7 | QKI |
| ST8SIA41 | 2.43E-54 | 0.53941744 | 0.487 | 0.183 | 9.27E-50 | ExN7 | ST8SIA4 |
| GAB2 | 2.44E-54 | 0.74571731 | 0.811 | 0.505 | 9.34E-50 | ExN7 | GAB2 |
| SAMHD1 | 3.98E-54 | 0.46227026 | 0.42 | 0.15 | 1.52E-49 | ExN7 | SAMHD1 |
| GRB2 | 1.66E-53 | 0.67050194 | 0.728 | 0.417 | 6.33E-49 | ExN7 | GRB2 |
| SSH2 | 1.80E-53 | 0.70859223 | 0.959 | 0.821 | 6.88E-49 | ExN7 | SSH2 |
| SMAP2 | 6.01E-53 | 0.7398554 | 0.775 | 0.501 | 2.30E-48 | ExN7 | SMAP2 |
| SKAP21 | 1.16E-52 | 0.72823933 | 0.674 | 0.381 | 4.44E-48 | ExN7 | SKAP2 |
| TBC1D16 | 3.13E-52 | 0.39053781 | 0.35 | 0.111 | 1.20E-47 | ExN7 | TBC1D16 |
| WASF2 | 4.73E-51 | 0.56911927 | 0.565 | 0.264 | 1.81E-46 | ExN7 | WASF2 |
| RAPGEF1 | 5.30E-51 | 0.69510404 | 0.767 | 0.48 | 2.03E-46 | ExN7 | RAPGEF1 |
| SNX29 | 4.41E-50 | 0.66194942 | 0.837 | 0.581 | 1.68E-45 | ExN7 | SNX29 |
| GLDN2 | 1.10E-49 | 0.71128329 | 0.534 | 0.236 | 4.20E-45 | ExN7 | GLDN |
| STK3 | 8.44E-49 | 0.61379754 | 0.622 | 0.316 | 3.23E-44 | ExN7 | STK3 |
| STK32B | 1.34E-48 | 0.34965814 | 0.262 | 0.069 | 5.13E-44 | ExN7 | STK32B |
| SH3RF31 | 1.82E-48 | 0.76119015 | 0.894 | 0.681 | 6.94E-44 | ExN7 | SH3RF3 |
| SRGAP2C | 1.83E-48 | 0.58873835 | 0.565 | 0.274 | 6.99E-44 | ExN7 | SRGAP2C |
| MIR646HG1 | 2.19E-48 | 0.61766117 | 0.619 | 0.277 | 8.38E-44 | ExN7 | MIR646HG |
| LAP3 | 3.86E-48 | 0.39916925 | 0.275 | 0.079 | 1.48E-43 | ExN7 | LAP3 |
| ARHGAP27 | 4.53E-48 | 0.27896779 | 0.254 | 0.067 | 1.73E-43 | ExN7 | ARHGAP27 |
| LRRK13 | 9.75E-48 | 0.55940398 | 0.676 | 0.331 | 3.73E-43 | ExN7 | LRRK1 |
| MTHFD1L | 1.23E-47 | 0.66482662 | 0.803 | 0.531 | 4.69E-43 | ExN7 | MTHFD1L |
| HAVCR2 | 1.58E-47 | 0.41360961 | 0.396 | 0.145 | 6.03E-43 | ExN7 | HAVCR2 |
| NCKAP5 | 1.87E-47 | 0.46245366 | 0.518 | 0.208 | 7.14E-43 | ExN7 | NCKAP5 |
| MBNL11 | 2.29E-47 | 0.72075537 | 0.935 | 0.779 | 8.77E-43 | ExN7 | MBNL1 |
| NOTCH2 | 2.70E-47 | 0.28864877 | 0.28 | 0.079 | 1.03E-42 | ExN7 | NOTCH2 |
| CTBP21 | 1.65E-46 | 0.54126767 | 0.674 | 0.353 | 6.30E-42 | ExN7 | CTBP2 |
| ATP11C | 2.87E-46 | 0.32315869 | 0.275 | 0.075 | 1.10E-41 | ExN7 | ATP11C |
| SPATA6 | 4.50E-46 | 0.54244906 | 0.472 | 0.202 | 1.72E-41 | ExN7 | SPATA6 |
| PIK3CD | 1.18E-45 | 0.37822734 | 0.321 | 0.103 | 4.50E-41 | ExN7 | PIK3CD |
| ST6GALNAC3 | 3.41E-45 | 0.96822609 | 0.762 | 0.507 | 1.30E-40 | ExN7 | ST6GALNAC3 |
| RIN2 | 6.76E-45 | 0.4028401 | 0.526 | 0.211 | 2.58E-40 | ExN7 | RIN2 |
| TANC11 | 7.99E-45 | 0.39802563 | 0.352 | 0.117 | 3.05E-40 | ExN7 | TANC1 |
| MKNK1 | 1.91E-44 | 0.59888287 | 0.744 | 0.462 | 7.29E-40 | ExN7 | MKNK1 |
| ZSWIM6 | 9.50E-44 | 0.6303414 | 0.917 | 0.73 | 3.63E-39 | ExN7 | ZSWIM6 |
| BAZ1A | 1.41E-43 | 0.30253497 | 0.308 | 0.099 | 5.38E-39 | ExN7 | BAZ1A |
| DISP1 | 1.46E-43 | 0.56545764 | 0.472 | 0.206 | 5.57E-39 | ExN7 | DISP1 |
| REL | 8.13E-43 | 0.43610885 | 0.376 | 0.141 | 3.11E-38 | ExN7 | REL |
| CFLAR | 1.02E-42 | 0.42234823 | 0.459 | 0.194 | 3.89E-38 | ExN7 | CFLAR |
| ARHGAP264 | 1.59E-42 | 0.56199986 | 0.979 | 0.872 | 6.09E-38 | ExN7 | ARHGAP26 |
| USP3 | 1.67E-42 | 0.37751147 | 0.446 | 0.176 | 6.39E-38 | ExN7 | USP3 |
| FTL | 2.03E-42 | 0.96824145 | 0.666 | 0.379 | 7.76E-38 | ExN7 | FTL |
| FCHSD2 | 1.47E-41 | 0.68777782 | 0.855 | 0.614 | 5.63E-37 | ExN7 | FCHSD2 |
| RASSF3 | 1.69E-41 | 0.28395763 | 0.293 | 0.091 | 6.45E-37 | ExN7 | RASSF3 |
| IL6ST | 1.94E-41 | 0.53795634 | 0.617 | 0.327 | 7.42E-37 | ExN7 | IL6ST |
| PTPRE | 3.54E-41 | 0.61189125 | 0.852 | 0.633 | 1.35E-36 | ExN7 | PTPRE |
| ATM | 4.19E-41 | 0.62057555 | 0.788 | 0.552 | 1.60E-36 | ExN7 | ATM |
| SERPINB9 | 6.06E-41 | 0.44817442 | 0.43 | 0.181 | 2.31E-36 | ExN7 | SERPINB9 |
| EML4 | 1.61E-40 | 0.57329815 | 0.661 | 0.382 | 6.17E-36 | ExN7 | EML4 |
| DIAPH21 | 2.05E-40 | 0.62432761 | 0.873 | 0.611 | 7.85E-36 | ExN7 | DIAPH2 |
| CREB3L2 | 3.23E-40 | 0.29821001 | 0.311 | 0.105 | 1.23E-35 | ExN7 | CREB3L2 |
| CSGALNACT1 | 3.43E-40 | 0.71269954 | 0.863 | 0.612 | 1.31E-35 | ExN7 | CSGALNACT1 |
| TTC7A | 5.36E-40 | 0.54101726 | 0.549 | 0.279 | 2.05E-35 | ExN7 | TTC7A |

| | | | | | | | |
|---|---|---|---|---|---|---|---|
| ANKS1A | 7.50E-40 | 0.61527513 | 0.837 | 0.596 | 2.87E-35 | ExN7 | ANKS1A |
| VASH1 | 8.64E-40 | 0.43857297 | 0.435 | 0.189 | 3.30E-35 | ExN7 | VASH1 |
| FMNL1 | 1.97E-39 | 0.30646025 | 0.342 | 0.126 | 7.51E-35 | ExN7 | FMNL1 |
| TCF12 | 4.81E-39 | 0.61543969 | 0.896 | 0.705 | 1.84E-34 | ExN7 | TCF12 |
| B4GALT1 | 5.89E-39 | 0.26134421 | 0.29 | 0.095 | 2.25E-34 | ExN7 | B4GALT1 |
| KLF6 | 6.38E-39 | 0.38839527 | 0.329 | 0.121 | 2.44E-34 | ExN7 | KLF6 |
| UTRN | 1.09E-38 | 0.46696817 | 0.549 | 0.255 | 4.17E-34 | ExN7 | UTRN |
| TBC1D14 | 1.39E-38 | 0.60472391 | 0.588 | 0.333 | 5.30E-34 | ExN7 | TBC1D14 |
| ETV51 | 6.25E-38 | 0.4204437 | 0.425 | 0.18 | 2.39E-33 | ExN7 | ETV5 |
| TAB2 | 1.02E-37 | 0.64267064 | 0.772 | 0.538 | 3.91E-33 | ExN7 | TAB2 |
| LRMP3 | 1.43E-37 | 0.3837091 | 0.389 | 0.152 | 5.48E-33 | ExN7 | LRMP |
| PFKFB3 | 1.92E-37 | 0.56958216 | 0.598 | 0.34 | 7.35E-33 | ExN7 | PFKFB3 |
| TSPAN14 | 2.86E-37 | 0.47749221 | 0.518 | 0.26 | 1.09E-32 | ExN7 | TSPAN14 |
| GALNT10 | 3.45E-37 | 0.33197611 | 0.373 | 0.145 | 1.32E-32 | ExN7 | GALNT10 |
| PIP4K2A1 | 4.32E-37 | 0.58310011 | 0.705 | 0.426 | 1.65E-32 | ExN7 | PIP4K2A |
| ETS2 | 5.65E-37 | 0.43956219 | 0.44 | 0.198 | 2.16E-32 | ExN7 | ETS2 |
| GALNT2 | 7.17E-37 | 0.57767079 | 0.733 | 0.481 | 2.74E-32 | ExN7 | GALNT2 |
| FNDC3B1 | 7.83E-37 | 0.47137323 | 0.627 | 0.345 | 2.99E-32 | ExN7 | FNDC3B |
| LIMS1 | 1.48E-36 | 0.58119634 | 0.601 | 0.333 | 5.66E-32 | ExN7 | LIMS1 |
| PPARD | 1.58E-36 | 0.45717884 | 0.531 | 0.27 | 6.02E-32 | ExN7 | PPARD |
| SCAMP2 | 4.53E-36 | 0.26075849 | 0.311 | 0.112 | 1.73E-31 | ExN7 | SCAMP2 |
| ZFAND6 | 6.90E-36 | 0.36943728 | 0.469 | 0.217 | 2.64E-31 | ExN7 | ZFAND6 |
| PACSIN2 | 7.73E-36 | 0.48099701 | 0.627 | 0.349 | 2.95E-31 | ExN7 | PACSIN2 |
| TMSB4X | 9.89E-36 | 0.7212291 | 0.759 | 0.513 | 3.78E-31 | ExN7 | TMSB4X |
| ITPKB | 1.24E-35 | 0.31313536 | 0.269 | 0.087 | 4.74E-31 | ExN7 | ITPKB |
| RPS19 | 1.80E-35 | 0.54353523 | 0.562 | 0.291 | 6.88E-31 | ExN7 | RPS19 |
| RAB1A | 2.69E-35 | 0.46851456 | 0.679 | 0.424 | 1.03E-30 | ExN7 | RAB1A |
| SPRED11 | 1.13E-34 | 0.62573192 | 0.668 | 0.421 | 4.33E-30 | ExN7 | SPRED1 |
| SOAT1 | 1.40E-34 | 0.33486573 | 0.334 | 0.13 | 5.37E-30 | ExN7 | SOAT1 |
| HLA-A | 1.48E-34 | 0.33335473 | 0.282 | 0.098 | 5.66E-30 | ExN7 | HLA-A |
| AC087564.1 | 3.88E-34 | 0.29018509 | 0.376 | 0.142 | 1.48E-29 | ExN7 | AC087564.1 |
| NCOA3 | 5.70E-34 | 0.47771388 | 0.658 | 0.396 | 2.18E-29 | ExN7 | NCOA3 |
| ARHGEF6 | 9.14E-34 | 0.36040839 | 0.404 | 0.177 | 3.49E-29 | ExN7 | ARHGEF6 |
| RNF166 | 9.23E-34 | 0.27251391 | 0.259 | 0.088 | 3.53E-29 | ExN7 | RNF166 |
| RCBTB2 | 9.38E-34 | 0.31566635 | 0.324 | 0.126 | 3.59E-29 | ExN7 | RCBTB2 |
| MIS18BP11 | 1.34E-33 | 0.38872545 | 0.453 | 0.215 | 5.12E-29 | ExN7 | MIS18BP1 |
| CEBPB | 1.39E-33 | 0.39843056 | 0.311 | 0.119 | 5.33E-29 | ExN7 | CEBPB |
| SYNDIG12 | 1.83E-33 | 0.7242936 | 0.79 | 0.598 | 6.99E-29 | ExN7 | SYNDIG1 |
| HTRA1 | 2.00E-33 | 0.36938428 | 0.402 | 0.166 | 7.65E-29 | ExN7 | HTRA1 |
| RB1 | 2.37E-33 | 0.50686206 | 0.764 | 0.52 | 9.08E-29 | ExN7 | RB1 |
| MED13L | 3.22E-33 | 0.54098737 | 0.943 | 0.835 | 1.23E-28 | ExN7 | MED13L |
| FAM49B | 4.79E-33 | 0.52914893 | 0.933 | 0.807 | 1.83E-28 | ExN7 | FAM49B |
| BCL6 | 5.19E-33 | 0.33320745 | 0.303 | 0.113 | 1.98E-28 | ExN7 | BCL6 |
| RAP1A | 7.10E-33 | 0.56074308 | 0.715 | 0.486 | 2.72E-28 | ExN7 | RAP1A |
| MAP4K41 | 1.05E-32 | 0.53792242 | 0.878 | 0.723 | 3.99E-28 | ExN7 | MAP4K4 |
| HS3ST42 | 8.84E-32 | 0.41474373 | 0.736 | 0.428 | 3.38E-27 | ExN7 | HS3ST4 |
| SLC25A37 | 8.88E-32 | 0.48770585 | 0.65 | 0.393 | 3.39E-27 | ExN7 | SLC25A37 |
| SLC25A13 | 1.21E-31 | 0.40981089 | 0.513 | 0.263 | 4.62E-27 | ExN7 | SLC25A13 |
| IGSF212 | 1.38E-31 | 0.58193031 | 0.759 | 0.534 | 5.28E-27 | ExN7 | IGSF21 |
| ARAP1 | 1.75E-31 | 0.34248703 | 0.352 | 0.148 | 6.70E-27 | ExN7 | ARAP1 |
| UST1 | 2.60E-31 | 0.31177825 | 0.36 | 0.146 | 9.93E-27 | ExN7 | UST |
| WASHC4 | 4.52E-31 | 0.45891026 | 0.565 | 0.329 | 1.73E-26 | ExN7 | WASHC4 |
| POU2F21 | 4.56E-31 | 0.50567705 | 0.585 | 0.34 | 1.74E-26 | ExN7 | POU2F2 |
| CPQ | 5.39E-31 | 0.44747894 | 0.456 | 0.219 | 2.06E-26 | ExN7 | CPQ |
| BIN1 | 1.30E-30 | 0.56815863 | 0.788 | 0.583 | 4.95E-26 | ExN7 | BIN1 |
| GPRIN3 | 1.33E-30 | 0.4158541 | 0.412 | 0.179 | 5.08E-26 | ExN7 | GPRIN3 |
| CEBPD | 1.46E-30 | 0.4940629 | 0.324 | 0.137 | 5.59E-26 | ExN7 | CEBPD |
| MYO9B | 1.96E-30 | 0.50445031 | 0.689 | 0.446 | 7.50E-26 | ExN7 | MYO9B |
| SLC4A7 | 2.93E-30 | 0.50972278 | 0.674 | 0.447 | 1.12E-25 | ExN7 | SLC4A7 |
| ATG16L2 | 2.97E-30 | 0.33318449 | 0.358 | 0.157 | 1.14E-25 | ExN7 | ATG16L2 |
| PLEKHA21 | 5.52E-30 | 0.43543195 | 0.477 | 0.249 | 2.11E-25 | ExN7 | PLEKHA2 |
| ARHGAP121 | 5.72E-30 | 0.46883933 | 0.588 | 0.337 | 2.19E-25 | ExN7 | ARHGAP12 |
| CTTNBP2NL | 6.14E-30 | 0.45279234 | 0.534 | 0.301 | 2.35E-25 | ExN7 | CTTNBP2NL |

| | | | | | | | |
|---|---|---|---|---|---|---|---|
| USP15 | 7.83E-30 | 0.54807651 | 0.793 | 0.6 | 2.99E-25 | ExN7 | USP15 |
| LINC022323 | 1.89E-29 | 0.35834499 | 0.44 | 0.192 | 7.24E-25 | ExN7 | LINC02232 |
| AP1B1 | 2.06E-29 | 0.43103997 | 0.505 | 0.277 | 7.86E-25 | ExN7 | AP1B1 |
| RTTN | 2.30E-29 | 0.66676266 | 0.523 | 0.302 | 8.78E-25 | ExN7 | RTTN |
| DLEU21 | 3.04E-29 | 0.48267191 | 0.79 | 0.56 | 1.16E-24 | ExN7 | DLEU2 |
| ZNRF2 | 3.39E-29 | 0.47599818 | 0.539 | 0.313 | 1.29E-24 | ExN7 | ZNRF2 |
| MAP3K1 | 5.39E-29 | 0.28699384 | 0.376 | 0.164 | 2.06E-24 | ExN7 | MAP3K1 |
| HIF1A | 7.51E-29 | 0.50018529 | 0.544 | 0.317 | 2.87E-24 | ExN7 | HIF1A |
| LRCH1 | 1.10E-28 | 0.49751447 | 0.756 | 0.542 | 4.20E-24 | ExN7 | LRCH1 |
| CTNND12 | 1.23E-28 | 0.46185263 | 0.699 | 0.474 | 4.70E-24 | ExN7 | CTNND1 |
| SH2B3 | 4.76E-28 | 0.32702091 | 0.378 | 0.176 | 1.82E-23 | ExN7 | SH2B3 |
| MANBA | 8.21E-28 | 0.47442178 | 0.622 | 0.382 | 3.14E-23 | ExN7 | MANBA |
| CD81 | 1.08E-27 | 0.43144656 | 0.469 | 0.243 | 4.13E-23 | ExN7 | CD81 |
| RASSF41 | 2.33E-27 | 0.37180048 | 0.453 | 0.23 | 8.89E-23 | ExN7 | RASSF4 |
| PSMA1 | 2.99E-27 | 0.39986503 | 0.583 | 0.346 | 1.14E-22 | ExN7 | PSMA1 |
| MGAT1 | 3.40E-27 | 0.33522045 | 0.36 | 0.163 | 1.30E-22 | ExN7 | MGAT1 |
| CST3 | 3.96E-27 | 0.26627758 | 0.259 | 0.097 | 1.51E-22 | ExN7 | CST3 |
| RNF149 | 4.67E-27 | 0.47485956 | 0.754 | 0.558 | 1.79E-22 | ExN7 | RNF149 |
| NEK6 | 4.82E-27 | 0.28158503 | 0.293 | 0.118 | 1.84E-22 | ExN7 | NEK6 |
| ITGAV | 4.95E-27 | 0.43319017 | 0.482 | 0.26 | 1.89E-22 | ExN7 | ITGAV |
| ARRB2 | 5.99E-27 | 0.39024877 | 0.42 | 0.213 | 2.29E-22 | ExN7 | ARRB2 |
| CBFB | 1.78E-26 | 0.2704471 | 0.365 | 0.165 | 6.80E-22 | ExN7 | CBFB |
| PSAP | 2.04E-26 | 0.44381936 | 0.855 | 0.639 | 7.81E-22 | ExN7 | PSAP |
| AFF13 | 4.11E-26 | 0.46418131 | 0.655 | 0.414 | 1.57E-21 | ExN7 | AFF1 |
| RPL13 | 6.51E-26 | 0.60056985 | 0.614 | 0.4 | 2.49E-21 | ExN7 | RPL13 |
| PCED1B2 | 1.21E-25 | 0.33359445 | 0.311 | 0.133 | 4.62E-21 | ExN7 | PCED1B |
| ZBTB20 | 1.51E-25 | 0.28614433 | 0.78 | 0.55 | 5.77E-21 | ExN7 | ZBTB20 |
| STAT3 | 1.86E-25 | 0.42267339 | 0.513 | 0.295 | 7.10E-21 | ExN7 | STAT3 |
| CORO1A | 2.80E-25 | 0.25077644 | 0.301 | 0.128 | 1.07E-20 | ExN7 | CORO1A |
| GPCPD1 | 3.15E-25 | 0.37331942 | 0.44 | 0.232 | 1.20E-20 | ExN7 | GPCPD1 |
| AP3B1 | 4.51E-25 | 0.42237204 | 0.611 | 0.386 | 1.73E-20 | ExN7 | AP3B1 |
| ASAH1 | 5.77E-25 | 0.41114114 | 0.56 | 0.342 | 2.20E-20 | ExN7 | ASAH1 |
| GRAMD1A | 9.06E-25 | 0.36519979 | 0.334 | 0.155 | 3.46E-20 | ExN7 | GRAMD1A |
| H2AFY | 9.55E-25 | 0.38199291 | 0.699 | 0.49 | 3.65E-20 | ExN7 | H2AFY |
| SERPINB6 | 1.47E-24 | 0.34075401 | 0.402 | 0.204 | 5.62E-20 | ExN7 | SERPINB6 |
| CHSY1 | 2.41E-24 | 0.407034 | 0.547 | 0.337 | 9.23E-20 | ExN7 | CHSY1 |
| CCDC18-AS1 | 3.04E-24 | 0.41828521 | 0.632 | 0.414 | 1.16E-19 | ExN7 | CCDC18-AS1 |
| WNT2B | 3.12E-24 | 0.2880846 | 0.277 | 0.116 | 1.19E-19 | ExN7 | WNT2B |
| SCAF11 | 3.41E-24 | 0.41472523 | 0.697 | 0.494 | 1.30E-19 | ExN7 | SCAF11 |
| CDK61 | 5.06E-24 | 0.40279726 | 0.653 | 0.413 | 1.93E-19 | ExN7 | CDK6 |
| IFNGR2 | 5.23E-24 | 0.35513562 | 0.484 | 0.275 | 2.00E-19 | ExN7 | IFNGR2 |
| HPS3 | 6.02E-24 | 0.29119404 | 0.345 | 0.161 | 2.30E-19 | ExN7 | HPS3 |
| SH3BGRL | 6.64E-24 | 0.4002147 | 0.694 | 0.475 | 2.54E-19 | ExN7 | SH3BGRL |
| ARHGEF1 | 8.97E-24 | 0.25842866 | 0.321 | 0.145 | 3.43E-19 | ExN7 | ARHGEF1 |
| NRIP1 | 9.23E-24 | 0.45592444 | 0.725 | 0.522 | 3.53E-19 | ExN7 | NRIP1 |
| RASA31 | 1.16E-23 | 0.42138681 | 0.57 | 0.351 | 4.42E-19 | ExN7 | RASA3 |
| TBC1D12 | 1.43E-23 | 0.44402552 | 0.513 | 0.308 | 5.47E-19 | ExN7 | TBC1D12 |
| LRRC8C | 3.25E-23 | 0.37004793 | 0.531 | 0.32 | 1.24E-18 | ExN7 | LRRC8C |
| SLC15A2 | 3.41E-23 | 0.27620605 | 0.326 | 0.151 | 1.31E-18 | ExN7 | SLC15A2 |
| NFE2L2 | 4.82E-23 | 0.35558125 | 0.469 | 0.262 | 1.84E-18 | ExN7 | NFE2L2 |
| EDA1 | 6.83E-23 | 0.29130763 | 0.316 | 0.141 | 2.61E-18 | ExN7 | EDA |
| RHOQ | 6.84E-23 | 0.3073039 | 0.415 | 0.215 | 2.61E-18 | ExN7 | RHOQ |
| CKLF | 7.95E-23 | 0.28411423 | 0.339 | 0.162 | 3.04E-18 | ExN7 | CKLF |
| DIP2B | 1.03E-22 | 0.42235781 | 0.829 | 0.691 | 3.94E-18 | ExN7 | DIP2B |
| CCNY1 | 2.28E-22 | 0.40016465 | 0.86 | 0.696 | 8.72E-18 | ExN7 | CCNY |
| JAZF11 | 2.66E-22 | 0.39121002 | 0.935 | 0.82 | 1.02E-17 | ExN7 | JAZF1 |
| ARL5A | 4.94E-22 | 0.40416824 | 0.497 | 0.299 | 1.89E-17 | ExN7 | ARL5A |
| MBP1 | 6.11E-22 | 0.33802339 | 0.749 | 0.532 | 2.34E-17 | ExN7 | MBP |
| MARCH3 | 6.49E-22 | 0.27298589 | 0.329 | 0.148 | 2.48E-17 | ExN7 | 3-Mar |
| RPL41 | 6.58E-22 | 0.37805759 | 0.565 | 0.342 | 2.52E-17 | ExN7 | RPL41 |
| PBX3 | 6.66E-22 | 0.25745605 | 0.334 | 0.155 | 2.55E-17 | ExN7 | PBX3 |
| ADPGK | 7.35E-22 | 0.26823352 | 0.319 | 0.154 | 2.81E-17 | ExN7 | ADPGK |
| STK4 | 7.35E-22 | 0.39275806 | 0.578 | 0.371 | 2.81E-17 | ExN7 | STK4 |

| | | | | | | | |
|---|---|---|---|---|---|---|---|
| CCM2 | 9.83E-22 | 0.34818996 | 0.466 | 0.275 | 3.76E-17 | ExN7 | CCM2 |
| CCND32 | 1.09E-21 | 0.41797211 | 0.598 | 0.389 | 4.17E-17 | ExN7 | CCND3 |
| FHIT1 | 1.10E-21 | 0.45866903 | 0.912 | 0.744 | 4.22E-17 | ExN7 | FHIT |
| GNAQ | 1.11E-21 | 0.41074581 | 0.94 | 0.858 | 4.23E-17 | ExN7 | GNAQ |
| APMAP | 1.71E-21 | 0.33255772 | 0.446 | 0.254 | 6.52E-17 | ExN7 | APMAP |
| HERPUD1 | 1.98E-21 | 0.31320933 | 0.386 | 0.204 | 7.57E-17 | ExN7 | HERPUD1 |
| CAB39 | 2.00E-21 | 0.40437078 | 0.715 | 0.52 | 7.64E-17 | ExN7 | CAB39 |
| UGCG | 2.46E-21 | 0.3811157 | 0.503 | 0.304 | 9.40E-17 | ExN7 | UGCG |
| SYT17 | 2.56E-21 | 0.34059401 | 0.518 | 0.3 | 9.79E-17 | ExN7 | SYT17 |
| ZFPM2-AS1 | 2.71E-21 | 0.27383506 | 0.259 | 0.111 | 1.04E-16 | ExN7 | ZFPM2-AS1 |
| WDFY2 | 2.88E-21 | 0.41745595 | 0.645 | 0.446 | 1.10E-16 | ExN7 | WDFY2 |
| ARPC2 | 2.92E-21 | 0.34163412 | 0.611 | 0.405 | 1.12E-16 | ExN7 | ARPC2 |
| APAF1 | 2.94E-21 | 0.29897128 | 0.352 | 0.178 | 1.12E-16 | ExN7 | APAF1 |
| GNG71 | 2.99E-21 | 0.36634628 | 0.627 | 0.409 | 1.14E-16 | ExN7 | GNG7 |
| MPZL1 | 3.16E-21 | 0.33960657 | 0.469 | 0.273 | 1.21E-16 | ExN7 | MPZL1 |
| GATAD2A | 3.62E-21 | 0.29917069 | 0.347 | 0.169 | 1.38E-16 | ExN7 | GATAD2A |
| ZFAND31 | 4.10E-21 | 0.35043331 | 0.953 | 0.849 | 1.57E-16 | ExN7 | ZFAND3 |
| SNX2 | 5.07E-21 | 0.28850232 | 0.541 | 0.335 | 1.94E-16 | ExN7 | SNX2 |
| MAP3K14 | 7.69E-21 | 0.25973156 | 0.326 | 0.158 | 2.94E-16 | ExN7 | MAP3K14 |
| ACER3 | 1.04E-20 | 0.34975341 | 0.614 | 0.401 | 3.99E-16 | ExN7 | ACER3 |
| VOPP1 | 1.91E-20 | 0.35611798 | 0.676 | 0.473 | 7.31E-16 | ExN7 | VOPP1 |
| CAPZB | 2.73E-20 | 0.38376864 | 0.785 | 0.636 | 1.04E-15 | ExN7 | CAPZB |
| APPL2 | 3.05E-20 | 0.39223295 | 0.663 | 0.466 | 1.17E-15 | ExN7 | APPL2 |
| CREBL2 | 3.06E-20 | 0.26639726 | 0.358 | 0.183 | 1.17E-15 | ExN7 | CREBL2 |
| RAP1B | 3.15E-20 | 0.34573154 | 0.534 | 0.334 | 1.20E-15 | ExN7 | RAP1B |
| RPS9 | 3.23E-20 | 0.31435494 | 0.422 | 0.235 | 1.24E-15 | ExN7 | RPS9 |
| SIPA1L2 | 4.88E-20 | 0.34678286 | 0.64 | 0.428 | 1.87E-15 | ExN7 | SIPA1L2 |
| TRPM21 | 4.98E-20 | 0.37723503 | 0.609 | 0.416 | 1.90E-15 | ExN7 | TRPM2 |
| TRMT1 | 7.56E-20 | 0.25904844 | 0.396 | 0.214 | 2.89E-15 | ExN7 | TRMT1 |
| RPL13A | 8.21E-20 | 0.39795144 | 0.495 | 0.298 | 3.14E-15 | ExN7 | RPL13A |
| ELL23 | 8.45E-20 | 0.41894538 | 0.749 | 0.548 | 3.23E-15 | ExN7 | ELL2 |
| RPS18 | 1.04E-19 | 0.38472217 | 0.482 | 0.288 | 3.99E-15 | ExN7 | RPS18 |
| RPS27A | 1.07E-19 | 0.41499695 | 0.477 | 0.282 | 4.10E-15 | ExN7 | RPS27A |
| GLUL | 1.22E-19 | 0.33081986 | 0.301 | 0.141 | 4.66E-15 | ExN7 | GLUL |
| C6orf62 | 1.27E-19 | 0.35154406 | 0.557 | 0.364 | 4.85E-15 | ExN7 | C6orf62 |
| SH3KBP1 | 1.54E-19 | 0.44602262 | 0.767 | 0.606 | 5.88E-15 | ExN7 | SH3KBP1 |
| STX7 | 1.63E-19 | 0.35817262 | 0.684 | 0.499 | 6.24E-15 | ExN7 | STX7 |
| UBE2E1 | 1.64E-19 | 0.35368645 | 0.578 | 0.378 | 6.28E-15 | ExN7 | UBE2E1 |
| PKN2 | 2.42E-19 | 0.41092626 | 0.777 | 0.633 | 9.26E-15 | ExN7 | PKN2 |
| SMURF2 | 3.25E-19 | 0.3208684 | 0.573 | 0.374 | 1.24E-14 | ExN7 | SMURF2 |
| SDCCAG81 | 3.73E-19 | 0.41008416 | 0.85 | 0.714 | 1.43E-14 | ExN7 | SDCCAG8 |
| RPS3 | 4.88E-19 | 0.38780736 | 0.541 | 0.343 | 1.87E-14 | ExN7 | RPS3 |
| ITM2B | 5.02E-19 | 0.38616319 | 0.782 | 0.62 | 1.92E-14 | ExN7 | ITM2B |
| MTUS1 | 6.20E-19 | 0.31789445 | 0.51 | 0.299 | 2.37E-14 | ExN7 | MTUS1 |
| P4HA1 | 8.41E-19 | 0.4318479 | 0.518 | 0.339 | 3.22E-14 | ExN7 | P4HA1 |
| SETX | 8.71E-19 | 0.36607486 | 0.725 | 0.555 | 3.33E-14 | ExN7 | SETX |
| RPS8 | 9.15E-19 | 0.37778016 | 0.534 | 0.336 | 3.50E-14 | ExN7 | RPS8 |
| TBC1D13 | 1.07E-18 | 0.39517508 | 0.751 | 0.575 | 4.11E-14 | ExN7 | TBC1D1 |
| PDE3B1 | 1.36E-18 | 0.39977709 | 0.712 | 0.519 | 5.21E-14 | ExN7 | PDE3B |
| ANP32A | 1.38E-18 | 0.29005378 | 0.495 | 0.306 | 5.29E-14 | ExN7 | ANP32A |
| LPIN2 | 1.41E-18 | 0.3306278 | 0.562 | 0.364 | 5.38E-14 | ExN7 | LPIN2 |
| ACTR3 | 1.69E-18 | 0.32369092 | 0.518 | 0.337 | 6.46E-14 | ExN7 | ACTR3 |
| PLCG2 | 1.85E-18 | 0.46000156 | 0.852 | 0.651 | 7.08E-14 | ExN7 | PLCG2 |
| ERBIN | 1.98E-18 | 0.36712973 | 0.731 | 0.539 | 7.57E-14 | ExN7 | ERBIN |
| ATG71 | 2.06E-18 | 0.39629876 | 0.912 | 0.777 | 7.89E-14 | ExN7 | ATG7 |
| JAK21 | 2.12E-18 | 0.37381091 | 0.793 | 0.611 | 8.09E-14 | ExN7 | JAK2 |
| ADAM9 | 2.53E-18 | 0.27457995 | 0.593 | 0.392 | 9.67E-14 | ExN7 | ADAM9 |
| MITF | 2.80E-18 | 0.30767155 | 0.383 | 0.199 | 1.07E-13 | ExN7 | MITF |
| PPP6R3 | 3.72E-18 | 0.31977193 | 0.837 | 0.698 | 1.42E-13 | ExN7 | PPP6R3 |
| FTH1 | 3.88E-18 | 0.55668564 | 0.741 | 0.566 | 1.48E-13 | ExN7 | FTH1 |
| PIAS1 | 4.56E-18 | 0.35719213 | 0.837 | 0.697 | 1.74E-13 | ExN7 | PIAS1 |
| KLHL51 | 4.85E-18 | 0.30116142 | 0.531 | 0.32 | 1.85E-13 | ExN7 | KLHL5 |
| GNA13 | 5.49E-18 | 0.27606175 | 0.383 | 0.211 | 2.10E-13 | ExN7 | GNA13 |

| | | | | | | | |
|---|---|---|---|---|---|---|---|
| ROCK1 | 5.89E-18 | 0.35254116 | 0.741 | 0.577 | 2.25E-13 | ExN7 | ROCK1 |
| GAS6 | 7.57E-18 | 0.36080052 | 0.459 | 0.292 | 2.89E-13 | ExN7 | GAS6 |
| RPL36A | 1.01E-17 | 0.29114967 | 0.311 | 0.156 | 3.87E-13 | ExN7 | RPL36A |
| TRAK1 | 1.10E-17 | 0.29833395 | 0.733 | 0.542 | 4.22E-13 | ExN7 | TRAK1 |
| NUMB1 | 1.64E-17 | 0.32832155 | 0.731 | 0.557 | 6.26E-13 | ExN7 | NUMB |
| PABPC1 | 1.68E-17 | 0.35861646 | 0.653 | 0.467 | 6.42E-13 | ExN7 | PABPC1 |
| ZEB2 | 1.69E-17 | 0.3064316 | 0.943 | 0.822 | 6.47E-13 | ExN7 | ZEB2 |
| NIPBL | 1.87E-17 | 0.35978567 | 0.87 | 0.729 | 7.16E-13 | ExN7 | NIPBL |
| MLXIPL | 2.02E-17 | 0.29375107 | 0.557 | 0.357 | 7.74E-13 | ExN7 | MLXIPL |
| RPS28 | 2.40E-17 | 0.38166368 | 0.552 | 0.359 | 9.16E-13 | ExN7 | RPS28 |
| ATP10D | 2.79E-17 | 0.30645916 | 0.404 | 0.233 | 1.07E-12 | ExN7 | ATP10D |
| ABHD12 | 2.99E-17 | 0.29785846 | 0.622 | 0.441 | 1.14E-12 | ExN7 | ABHD12 |
| RPL10 | 3.07E-17 | 0.34213223 | 0.539 | 0.344 | 1.18E-12 | ExN7 | RPL10 |
| RPS11 | 3.09E-17 | 0.29340275 | 0.339 | 0.18 | 1.18E-12 | ExN7 | RPS11 |
| NFIA2 | 3.18E-17 | 0.2891618 | 0.653 | 0.422 | 1.22E-12 | ExN7 | NFIA |
| TET2 | 3.40E-17 | 0.31022966 | 0.627 | 0.438 | 1.30E-12 | ExN7 | TET2 |
| E2F31 | 3.88E-17 | 0.32077604 | 0.528 | 0.339 | 1.48E-12 | ExN7 | E2F3 |
| HNRNPH1 | 3.99E-17 | 0.32613204 | 0.663 | 0.484 | 1.52E-12 | ExN7 | HNRNPH1 |
| ENOX2 | 3.99E-17 | 0.30053442 | 0.342 | 0.18 | 1.53E-12 | ExN7 | ENOX2 |
| BCL21 | 4.05E-17 | 0.45638882 | 0.72 | 0.527 | 1.55E-12 | ExN7 | BCL2 |
| RPL3 | 5.08E-17 | 0.42726914 | 0.484 | 0.31 | 1.94E-12 | ExN7 | RPL3 |
| SLC38A10 | 5.12E-17 | 0.25597932 | 0.355 | 0.194 | 1.96E-12 | ExN7 | SLC38A10 |
| MGST2 | 5.29E-17 | 0.26927038 | 0.368 | 0.202 | 2.02E-12 | ExN7 | MGST2 |
| FMNL23 | 5.67E-17 | 0.36924318 | 0.951 | 0.874 | 2.17E-12 | ExN7 | FMNL2 |
| GSAP | 6.79E-17 | 0.29864702 | 0.425 | 0.256 | 2.59E-12 | ExN7 | GSAP |
| PHYKPL | 7.38E-17 | 0.25963416 | 0.43 | 0.258 | 2.82E-12 | ExN7 | PHYKPL |
| RPL11 | 8.39E-17 | 0.38453629 | 0.477 | 0.301 | 3.21E-12 | ExN7 | RPL11 |
| TMEM1632 | 8.90E-17 | 0.64382307 | 0.456 | 0.291 | 3.40E-12 | ExN7 | TMEM163 |
| ERICH1 | 8.90E-17 | 0.30545898 | 0.482 | 0.305 | 3.40E-12 | ExN7 | ERICH1 |
| FAM53B | 9.25E-17 | 0.34134687 | 0.585 | 0.392 | 3.54E-12 | ExN7 | FAM53B |
| MAP3K51 | 9.59E-17 | 0.30396243 | 0.845 | 0.642 | 3.67E-12 | ExN7 | MAP3K5 |
| DNAJC3 | 1.10E-16 | 0.31745319 | 0.446 | 0.278 | 4.20E-12 | ExN7 | DNAJC3 |
| SKIL | 1.38E-16 | 0.29016776 | 0.44 | 0.268 | 5.29E-12 | ExN7 | SKIL |
| ATP11A1 | 1.61E-16 | 0.3061995 | 0.63 | 0.446 | 6.14E-12 | ExN7 | ATP11A |
| UVRAG | 2.04E-16 | 0.34948473 | 0.801 | 0.671 | 7.80E-12 | ExN7 | UVRAG |
| ST3GAL63 | 2.14E-16 | 0.30000964 | 0.63 | 0.435 | 8.20E-12 | ExN7 | ST3GAL6 |
| SHTN1 | 2.21E-16 | 0.32195653 | 0.813 | 0.675 | 8.45E-12 | ExN7 | SHTN1 |
| STAG1 | 2.71E-16 | 0.31679426 | 0.902 | 0.788 | 1.03E-11 | ExN7 | STAG1 |
| ITGA91 | 2.71E-16 | 0.31990368 | 0.645 | 0.434 | 1.04E-11 | ExN7 | ITGA9 |
| RPL28 | 3.09E-16 | 0.37662488 | 0.552 | 0.368 | 1.18E-11 | ExN7 | RPL28 |
| HNRNPM | 3.53E-16 | 0.30988453 | 0.611 | 0.438 | 1.35E-11 | ExN7 | HNRNPM |
| DTNBP1 | 3.64E-16 | 0.25310111 | 0.492 | 0.307 | 1.39E-11 | ExN7 | DTNBP1 |
| OGFRL1 | 4.28E-16 | 0.35561163 | 0.484 | 0.318 | 1.63E-11 | ExN7 | OGFRL1 |
| MCF2L | 5.12E-16 | 0.34166601 | 0.795 | 0.634 | 1.96E-11 | ExN7 | MCF2L |
| PTBP3 | 6.35E-16 | 0.29468067 | 0.544 | 0.366 | 2.43E-11 | ExN7 | PTBP3 |
| MYH91 | 8.03E-16 | 0.28007408 | 0.482 | 0.304 | 3.07E-11 | ExN7 | MYH9 |
| BAZ2B | 9.74E-16 | 0.3175662 | 0.837 | 0.71 | 3.72E-11 | ExN7 | BAZ2B |
| PHC2 | 1.29E-15 | 0.33535085 | 0.653 | 0.487 | 4.91E-11 | ExN7 | PHC2 |
| LYST2 | 1.33E-15 | 0.30138022 | 0.863 | 0.724 | 5.09E-11 | ExN7 | LYST |
| ATF6 | 1.34E-15 | 0.32029162 | 0.715 | 0.537 | 5.14E-11 | ExN7 | ATF6 |
| MTAP | 2.14E-15 | 0.28541785 | 0.402 | 0.239 | 8.17E-11 | ExN7 | MTAP |
| RPS2 | 2.44E-15 | 0.29985503 | 0.415 | 0.252 | 9.31E-11 | ExN7 | RPS2 |
| SLC11A2 | 2.70E-15 | 0.2949257 | 0.528 | 0.365 | 1.03E-10 | ExN7 | SLC11A2 |
| TRPS1 | 2.84E-15 | 0.39969014 | 0.635 | 0.439 | 1.08E-10 | ExN7 | TRPS1 |
| EEF1A1 | 3.22E-15 | 0.33511573 | 0.751 | 0.576 | 1.23E-10 | ExN7 | EEF1A1 |
| FAM133B | 3.29E-15 | 0.26662836 | 0.575 | 0.396 | 1.26E-10 | ExN7 | FAM133B |
| PTPN2 | 3.39E-15 | 0.32888316 | 0.655 | 0.477 | 1.30E-10 | ExN7 | PTPN2 |
| MOB1B1 | 5.88E-15 | 0.28740574 | 0.544 | 0.366 | 2.25E-10 | ExN7 | MOB1B |
| PRMT2 | 6.05E-15 | 0.33020697 | 0.534 | 0.37 | 2.31E-10 | ExN7 | PRMT2 |
| TAOK3 | 8.47E-15 | 0.29225612 | 0.847 | 0.709 | 3.24E-10 | ExN7 | TAOK3 |
| RPLP1 | 8.66E-15 | 0.32000415 | 0.477 | 0.309 | 3.31E-10 | ExN7 | RPLP1 |
| PHF20 | 8.81E-15 | 0.29111382 | 0.632 | 0.459 | 3.37E-10 | ExN7 | PHF20 |
| TBC1D2B | 9.79E-15 | 0.27584917 | 0.482 | 0.317 | 3.74E-10 | ExN7 | TBC1D2B |

| | | | | | | | |
|---|---|---|---|---|---|---|---|
| IKBKB | 1.02E-14 | 0.26785422 | 0.373 | 0.221 | 3.91E-10 | ExN7 | IKBKB |
| SLTM | 1.09E-14 | 0.26365256 | 0.632 | 0.462 | 4.18E-10 | ExN7 | SLTM |
| DEPTOR | 1.11E-14 | 0.28087023 | 0.44 | 0.266 | 4.25E-10 | ExN7 | DEPTOR |
| OSTF11 | 1.45E-14 | 0.25134763 | 0.412 | 0.247 | 5.54E-10 | ExN7 | OSTF1 |
| ADAM17 | 1.47E-14 | 0.25640704 | 0.606 | 0.42 | 5.61E-10 | ExN7 | ADAM17 |
| CCNL1 | 2.53E-14 | 0.28433744 | 0.482 | 0.319 | 9.65E-10 | ExN7 | CCNL1 |
| RAB12 | 2.69E-14 | 0.29040019 | 0.578 | 0.408 | 1.03E-09 | ExN7 | RAB12 |
| SKI | 2.79E-14 | 0.29557082 | 0.622 | 0.464 | 1.07E-09 | ExN7 | SKI |
| STK24 | 2.86E-14 | 0.2659608 | 0.544 | 0.375 | 1.09E-09 | ExN7 | STK24 |
| GAREM12 | 3.78E-14 | 0.27860017 | 0.679 | 0.471 | 1.45E-09 | ExN7 | GAREM1 |
| XIST1 | 4.08E-14 | 0.39960333 | 0.749 | 0.543 | 1.56E-09 | ExN7 | XIST |
| TMEM1651 | 4.40E-14 | 0.30873149 | 0.575 | 0.42 | 1.68E-09 | ExN7 | TMEM165 |
| SSH1 | 4.76E-14 | 0.31384874 | 0.601 | 0.432 | 1.82E-09 | ExN7 | SSH1 |
| SLC39A112 | 4.88E-14 | 0.29641058 | 0.611 | 0.42 | 1.87E-09 | ExN7 | SLC39A11 |
| ACAP21 | 4.93E-14 | 0.30758877 | 0.806 | 0.661 | 1.88E-09 | ExN7 | ACAP2 |
| RPS12 | 5.58E-14 | 0.2539655 | 0.394 | 0.235 | 2.13E-09 | ExN7 | RPS12 |
| NEK7 | 6.08E-14 | 0.26385745 | 0.378 | 0.222 | 2.32E-09 | ExN7 | NEK7 |
| CHKA | 6.83E-14 | 0.27231434 | 0.601 | 0.445 | 2.61E-09 | ExN7 | CHKA |
| XRN2 | 9.15E-14 | 0.28115346 | 0.585 | 0.427 | 3.50E-09 | ExN7 | XRN2 |
| CHST15 | 1.49E-13 | 0.283472 | 0.601 | 0.42 | 5.68E-09 | ExN7 | CHST15 |
| JARID2 | 1.51E-13 | 0.33008361 | 0.775 | 0.631 | 5.77E-09 | ExN7 | JARID2 |
| RAB7A | 1.73E-13 | 0.27390944 | 0.681 | 0.517 | 6.62E-09 | ExN7 | RAB7A |
| TPT1 | 1.85E-13 | 0.31228334 | 0.495 | 0.334 | 7.09E-09 | ExN7 | TPT1 |
| EIF4A1 | 1.89E-13 | 0.2660201 | 0.479 | 0.326 | 7.23E-09 | ExN7 | EIF4A1 |
| KIF13B | 2.84E-13 | 0.27157284 | 0.469 | 0.312 | 1.09E-08 | ExN7 | KIF13B |
| KMT2C | 2.91E-13 | 0.28479206 | 0.878 | 0.774 | 1.11E-08 | ExN7 | KMT2C |
| CDYL | 3.12E-13 | 0.28899045 | 0.58 | 0.421 | 1.19E-08 | ExN7 | CDYL |
| RPL26 | 3.28E-13 | 0.27718338 | 0.435 | 0.271 | 1.25E-08 | ExN7 | RPL26 |
| COMMD10 | 3.57E-13 | 0.31828077 | 0.658 | 0.499 | 1.36E-08 | ExN7 | COMMD10 |
| MGAT51 | 3.63E-13 | 0.3030704 | 0.772 | 0.661 | 1.39E-08 | ExN7 | MGAT5 |
| RPL35 | 4.43E-13 | 0.26308937 | 0.422 | 0.264 | 1.69E-08 | ExN7 | RPL35 |
| CD63 | 4.60E-13 | 0.26961344 | 0.288 | 0.158 | 1.76E-08 | ExN7 | CD63 |
| SMAD32 | 4.82E-13 | 0.2660537 | 0.56 | 0.382 | 1.84E-08 | ExN7 | SMAD3 |
| TMEM65 | 5.23E-13 | 0.31680393 | 0.573 | 0.399 | 2.00E-08 | ExN7 | TMEM65 |
| SF3B1 | 5.61E-13 | 0.26607783 | 0.705 | 0.542 | 2.14E-08 | ExN7 | SF3B1 |
| AC008691.1 | 5.76E-13 | 0.25590369 | 0.319 | 0.177 | 2.20E-08 | ExN7 | AC008691.1 |
| EPS152 | 6.08E-13 | 0.32504473 | 0.64 | 0.488 | 2.33E-08 | ExN7 | EPS15 |
| RAB10 | 1.13E-12 | 0.2907063 | 0.712 | 0.575 | 4.32E-08 | ExN7 | RAB10 |
| ATF7IP | 1.21E-12 | 0.28422504 | 0.63 | 0.492 | 4.62E-08 | ExN7 | ATF7IP |
| FKBP15 | 1.36E-12 | 0.27228393 | 0.42 | 0.277 | 5.22E-08 | ExN7 | FKBP15 |
| RPS6 | 1.43E-12 | 0.34032429 | 0.365 | 0.226 | 5.46E-08 | ExN7 | RPS6 |
| TPK11 | 1.50E-12 | 0.31744987 | 0.598 | 0.444 | 5.72E-08 | ExN7 | TPK1 |
| KDM6A | 1.89E-12 | 0.27600174 | 0.744 | 0.582 | 7.21E-08 | ExN7 | KDM6A |
| PRIM2 | 1.92E-12 | 0.2859032 | 0.427 | 0.282 | 7.35E-08 | ExN7 | PRIM2 |
| SRRM2 | 2.13E-12 | 0.25932276 | 0.845 | 0.721 | 8.14E-08 | ExN7 | SRRM2 |
| DNAJB6 | 2.26E-12 | 0.27623584 | 0.604 | 0.445 | 8.62E-08 | ExN7 | DNAJB6 |
| LRRK21 | 2.80E-12 | 0.29759455 | 0.596 | 0.439 | 1.07E-07 | ExN7 | LRRK2 |
| RPLP2 | 4.45E-12 | 0.25225173 | 0.368 | 0.226 | 1.70E-07 | ExN7 | RPLP2 |
| FNIP22 | 4.87E-12 | 0.34011164 | 0.754 | 0.616 | 1.86E-07 | ExN7 | FNIP2 |
| ZFX | 5.07E-12 | 0.25407503 | 0.487 | 0.329 | 1.94E-07 | ExN7 | ZFX |
| ELF2 | 5.30E-12 | 0.26329463 | 0.712 | 0.558 | 2.03E-07 | ExN7 | ELF2 |
| MTSS1 | 5.37E-12 | 0.26980393 | 0.733 | 0.571 | 2.05E-07 | ExN7 | MTSS1 |
| RPL7A | 5.83E-12 | 0.2814073 | 0.446 | 0.294 | 2.23E-07 | ExN7 | RPL7A |
| PRRC2B | 6.01E-12 | 0.26494647 | 0.798 | 0.665 | 2.30E-07 | ExN7 | PRRC2B |
| CYTH1 | 7.28E-12 | 0.25337635 | 0.596 | 0.441 | 2.78E-07 | ExN7 | CYTH1 |
| PRKCA1 | 7.98E-12 | 0.2767209 | 0.876 | 0.736 | 3.05E-07 | ExN7 | PRKCA |
| PIK3R12 | 8.74E-12 | 0.30283334 | 0.834 | 0.692 | 3.34E-07 | ExN7 | PIK3R1 |
| RPS25 | 1.02E-11 | 0.26467561 | 0.438 | 0.292 | 3.89E-07 | ExN7 | RPS25 |
| RPS7 | 1.29E-11 | 0.25761675 | 0.407 | 0.265 | 4.92E-07 | ExN7 | RPS7 |
| RPS15 | 1.36E-11 | 0.32336503 | 0.433 | 0.289 | 5.22E-07 | ExN7 | RPS15 |
| RPS23 | 1.44E-11 | 0.2527112 | 0.451 | 0.296 | 5.51E-07 | ExN7 | RPS23 |
| SLC2A3 | 1.58E-11 | 0.33185674 | 0.339 | 0.211 | 6.05E-07 | ExN7 | SLC2A3 |
| ZMAT3 | 1.97E-11 | 0.2845435 | 0.484 | 0.342 | 7.52E-07 | ExN7 | ZMAT3 |

| | | | | | | | |
|---|---|---|---|---|---|---|---|
| SNX13 | 2.56E-11 | 0.29667884 | 0.712 | 0.578 | 9.78E-07 | ExN7 | SNX13 |
| DENND4C | 3.04E-11 | 0.26383984 | 0.637 | 0.495 | 1.16E-06 | ExN7 | DENND4C |
| DDX5 | 3.06E-11 | 0.26621858 | 0.832 | 0.716 | 1.17E-06 | ExN7 | DDX5 |
| RPL19 | 3.20E-11 | 0.33089605 | 0.422 | 0.28 | 1.22E-06 | ExN7 | RPL19 |
| HDAC91 | 3.34E-11 | 0.33812348 | 0.883 | 0.779 | 1.28E-06 | ExN7 | HDAC9 |
| EIF3H | 3.64E-11 | 0.25477365 | 0.508 | 0.363 | 1.39E-06 | ExN7 | EIF3H |
| DCAF63 | 3.83E-11 | 0.27798649 | 0.785 | 0.626 | 1.47E-06 | ExN7 | DCAF6 |
| CORO1C | 4.03E-11 | 0.28982542 | 0.492 | 0.35 | 1.54E-06 | ExN7 | CORO1C |
| ACTB1 | 4.46E-11 | 0.32649842 | 0.795 | 0.64 | 1.70E-06 | ExN7 | ACTB |
| RPS14 | 4.50E-11 | 0.26781028 | 0.451 | 0.311 | 1.72E-06 | ExN7 | RPS14 |
| HSPA5 | 5.21E-11 | 0.28192209 | 0.355 | 0.223 | 1.99E-06 | ExN7 | HSPA5 |
| DNAJC13 | 5.40E-11 | 0.2529528 | 0.557 | 0.416 | 2.07E-06 | ExN7 | DNAJC13 |
| RPS24 | 5.94E-11 | 0.2712293 | 0.588 | 0.427 | 2.27E-06 | ExN7 | RPS24 |
| CAMKMT | 6.74E-11 | 0.27110639 | 0.544 | 0.382 | 2.58E-06 | ExN7 | CAMKMT |
| RNF13 | 7.45E-11 | 0.29112675 | 0.71 | 0.577 | 2.85E-06 | ExN7 | RNF13 |
| NCOR2 | 8.34E-11 | 0.28493586 | 0.736 | 0.595 | 3.19E-06 | ExN7 | NCOR2 |
| RNF130 | 8.44E-11 | 0.25819303 | 0.826 | 0.729 | 3.23E-06 | ExN7 | RNF130 |
| ACTR2 | 1.06E-10 | 0.26750802 | 0.687 | 0.529 | 4.03E-06 | ExN7 | ACTR2 |
| TBC1D5 | 1.32E-10 | 0.28338232 | 0.92 | 0.858 | 5.06E-06 | ExN7 | TBC1D5 |
| WDFY3 | 1.96E-10 | 0.25932618 | 0.915 | 0.81 | 7.48E-06 | ExN7 | WDFY3 |
| ZHX2 | 2.51E-10 | 0.26402006 | 0.355 | 0.224 | 9.58E-06 | ExN7 | ZHX2 |
| RPL18 | 3.43E-10 | 0.25016073 | 0.459 | 0.316 | 1.31E-05 | ExN7 | RPL18 |
| KCNQ34 | 6.78E-10 | 0.25572873 | 0.969 | 0.902 | 2.59E-05 | ExN7 | KCNQ3 |
| DPYD4 | 8.42E-10 | 0.31387693 | 0.834 | 0.668 | 3.22E-05 | ExN7 | DPYD |
| RIC1 | 9.10E-10 | 0.25053678 | 0.536 | 0.407 | 3.48E-05 | ExN7 | RIC1 |
| LRP1 | 9.72E-10 | 0.26062391 | 0.617 | 0.486 | 3.72E-05 | ExN7 | LRP1 |
| MVB12B | 9.94E-10 | 0.26956333 | 0.674 | 0.555 | 3.80E-05 | ExN7 | MVB12B |
| NSMCE2 | 1.01E-09 | 0.2556157 | 0.811 | 0.672 | 3.84E-05 | ExN7 | NSMCE2 |
| NACA | 1.07E-09 | 0.25343284 | 0.539 | 0.391 | 4.10E-05 | ExN7 | NACA |
| IRS2 | 1.11E-09 | 0.25105603 | 0.552 | 0.42 | 4.23E-05 | ExN7 | IRS2 |
| KIF16B1 | 1.49E-09 | 0.25386468 | 0.671 | 0.541 | 5.70E-05 | ExN7 | KIF16B |
| SEMA4D | 1.95E-09 | 0.25067775 | 0.573 | 0.44 | 7.44E-05 | ExN7 | SEMA4D |
| RPS27 | 2.25E-09 | 0.36384981 | 0.438 | 0.308 | 8.60E-05 | ExN7 | RPS27 |
| SUMF1 | 7.22E-09 | 0.25641567 | 0.705 | 0.591 | 0.00027586 | ExN7 | SUMF1 |
| RPS4X | 1.92E-08 | 0.27121162 | 0.355 | 0.245 | 0.00073533 | ExN7 | RPS4X |
| NAMPT | 3.18E-08 | 0.27831593 | 0.324 | 0.218 | 0.00121462 | ExN7 | NAMPT |
| RBPJ | 6.91E-08 | 0.28298937 | 0.674 | 0.581 | 0.00264057 | ExN7 | RBPJ |
| HHAT1 | 1.46E-05 | 0.25368248 | 0.363 | 0.273 | 0.55989897 | ExN7 | HHAT |
| HSP90AA1 | 6.07E-05 | 0.34831123 | 0.793 | 0.712 | 1 | ExN7 | HSP90AA1 |
| ADARB2 | 0 | 2.318592 | 0.993 | 0.29 | 0 | InN1 | ADARB2 |
| ERBB4 | 0 | 2.22004728 | 0.992 | 0.523 | 0 | InN1 | ERBB4 |
| RGS12 | 0 | 2.09118671 | 0.941 | 0.463 | 0 | InN1 | RGS12 |
| SYNPR | 0 | 2.03183131 | 0.958 | 0.485 | 0 | InN1 | SYNPR |
| L3MBTL4 | 0 | 1.96069398 | 0.883 | 0.339 | 0 | InN1 | L3MBTL4 |
| SLC24A3 | 0 | 1.92479284 | 0.866 | 0.241 | 0 | InN1 | SLC24A3 |
| CALB2 | 0 | 1.90529752 | 0.676 | 0.051 | 0 | InN1 | CALB2 |
| AL589740.1 | 0 | 1.88006098 | 0.716 | 0.403 | 0 | InN1 | AL589740.1 |
| THSD7A | 0 | 1.8554226 | 0.858 | 0.465 | 0 | InN1 | THSD7A |
| BTBD11 | 0 | 1.75222646 | 0.823 | 0.203 | 0 | InN1 | BTBD11 |
| PLD5 | 0 | 1.74383279 | 0.865 | 0.254 | 0 | InN1 | PLD5 |
| GALNTL6 | 0 | 1.65737729 | 0.96 | 0.716 | 0 | InN1 | GALNTL6 |
| LAMA3 | 0 | 1.602751 | 0.574 | 0.075 | 0 | InN1 | LAMA3 |
| IGFBP5 | 0 | 1.59025526 | 0.489 | 0.144 | 0 | InN1 | IGFBP5 |
| PROX1 | 0 | 1.57842054 | 0.718 | 0.102 | 0 | InN1 | PROX1 |
| AC087564.11 | 0 | 1.57690761 | 0.6 | 0.082 | 0 | InN1 | AC087564.1 |
| RYR31 | 0 | 1.5391368 | 0.56 | 0.37 | 0 | InN1 | RYR3 |
| AC233296.1 | 0 | 1.53546109 | 0.708 | 0.213 | 0 | InN1 | AC233296.1 |
| PWRN11 | 0 | 1.51800151 | 0.87 | 0.419 | 0 | InN1 | PWRN1 |
| PLCE1 | 0 | 1.50126374 | 0.562 | 0.09 | 0 | InN1 | PLCE1 |
| DLX6-AS1 | 0 | 1.48936985 | 0.724 | 0.179 | 0 | InN1 | DLX6-AS1 |
| NR2F2-AS1 | 0 | 1.48703434 | 0.64 | 0.155 | 0 | InN1 | NR2F2-AS1 |
| C8orf34 | 0 | 1.43715874 | 0.71 | 0.352 | 0 | InN1 | C8orf34 |
| NPAS3 | 0 | 1.42707948 | 0.918 | 0.806 | 0 | InN1 | NPAS3 |

| | | | | | | | |
|---|---|---|---|---|---|---|---|
| NEAT11 | 0 | 1.40575623 | 0.679 | 0.345 | 0 | InN1 | NEAT1 |
| ZBTB201 | 0 | 1.40139786 | 0.689 | 0.534 | 0 | InN1 | ZBTB20 |
| PDE3A | 0 | 1.3965516 | 0.509 | 0.088 | 0 | InN1 | PDE3A |
| PROX1-AS1 | 0 | 1.38531796 | 0.54 | 0.059 | 0 | InN1 | PROX1-AS1 |
| KCNT21 | 0 | 1.37381592 | 0.807 | 0.514 | 0 | InN1 | KCNT2 |
| GALNT131 | 0 | 1.36307892 | 0.896 | 0.658 | 0 | InN1 | GALNT13 |
| LRRC3B | 0 | 1.35490322 | 0.64 | 0.205 | 0 | InN1 | LRRC3B |
| CCNH | 0 | 1.32967405 | 0.863 | 0.756 | 0 | InN1 | CCNH |
| FIGN1 | 0 | 1.32813543 | 0.671 | 0.317 | 0 | InN1 | FIGN |
| PTPRE1 | 0 | 1.32640146 | 0.848 | 0.606 | 0 | InN1 | PTPRE |
| EGFR | 0 | 1.31277402 | 0.729 | 0.18 | 0 | InN1 | EGFR |
| ANO4 | 0 | 1.29244456 | 0.865 | 0.566 | 0 | InN1 | ANO4 |
| SHISA6 | 0 | 1.27525068 | 0.64 | 0.308 | 0 | InN1 | SHISA6 |
| ADAMTS6 | 0 | 1.26605004 | 0.514 | 0.08 | 0 | InN1 | ADAMTS6 |
| AC013265.1 | 0 | 1.25645135 | 0.448 | 0.025 | 0 | InN1 | AC013265.1 |
| PDE4B1 | 0 | 1.2554735 | 0.869 | 0.58 | 0 | InN1 | PDE4B |
| AL589693.11 | 0 | 1.24478344 | 0.347 | 0.108 | 0 | InN1 | AL589693.1 |
| GPR149 | 0 | 1.23361247 | 0.484 | 0.035 | 0 | InN1 | GPR149 |
| SLC22A3 | 0 | 1.23047673 | 0.434 | 0.008 | 0 | InN1 | SLC22A3 |
| LINC01931 | 0 | 1.22176648 | 0.273 | 0.038 | 0 | InN1 | LINC01931 |
| TAC3 | 0 | 1.21581719 | 0.316 | 0.02 | 0 | InN1 | TAC3 |
| ROBO1 | 0 | 1.21566042 | 0.922 | 0.628 | 0 | InN1 | ROBO1 |
| SEZ6 | 0 | 1.20821668 | 0.57 | 0.192 | 0 | InN1 | SEZ6 |
| SHISA8 | 0 | 1.20713933 | 0.45 | 0.025 | 0 | InN1 | SHISA8 |
| AL133346.11 | 0 | 1.20665124 | 0.522 | 0.103 | 0 | InN1 | AL133346.1 |
| DOCK101 | 0 | 1.17394983 | 0.83 | 0.351 | 0 | InN1 | DOCK10 |
| VIP | 0 | 1.16851243 | 0.365 | 0.014 | 0 | InN1 | VIP |
| MCC1 | 0 | 1.16699146 | 0.641 | 0.217 | 0 | InN1 | MCC |
| ITGA81 | 0 | 1.15759265 | 0.468 | 0.083 | 0 | InN1 | ITGA8 |
| SDK11 | 0 | 1.14524422 | 0.779 | 0.443 | 0 | InN1 | SDK1 |
| ZNF536 | 0 | 1.13724428 | 0.89 | 0.309 | 0 | InN1 | ZNF536 |
| ARHGAP28 | 0 | 1.13624203 | 0.544 | 0.166 | 0 | InN1 | ARHGAP28 |
| SVEP1 | 0 | 1.12765865 | 0.494 | 0.165 | 0 | InN1 | SVEP1 |
| COL21A1 | 0 | 1.12069888 | 0.559 | 0.15 | 0 | InN1 | COL21A1 |
| LINC013221 | 0 | 1.09813278 | 0.729 | 0.521 | 0 | InN1 | LINC01322 |
| ADRA1B | 0 | 1.0958215 | 0.526 | 0.183 | 0 | InN1 | ADRA1B |
| B4GALT6 | 0 | 1.09044152 | 0.659 | 0.385 | 0 | InN1 | B4GALT6 |
| SEMA5A1 | 0 | 1.0769308 | 0.46 | 0.139 | 0 | InN1 | SEMA5A |
| NKAIN3 | 0 | 1.06812537 | 0.589 | 0.239 | 0 | InN1 | NKAIN3 |
| PITPNC1 | 0 | 1.05802713 | 0.789 | 0.457 | 0 | InN1 | PITPNC1 |
| KIAA1211 | 0 | 1.05191375 | 0.771 | 0.354 | 0 | InN1 | KIAA1211 |
| NCKAP51 | 0 | 1.04770026 | 0.448 | 0.179 | 0 | InN1 | NCKAP5 |
| AL136114.1 | 0 | 1.04162639 | 0.328 | 0.079 | 0 | InN1 | AL136114.1 |
| CHD71 | 0 | 1.03150704 | 0.452 | 0.071 | 0 | InN1 | CHD7 |
| PLXNA4 | 0 | 1.01805399 | 0.913 | 0.784 | 0 | InN1 | PLXNA4 |
| CALN12 | 0 | 1.01787941 | 0.794 | 0.645 | 0 | InN1 | CALN1 |
| AC016766.1 | 0 | 1.01655956 | 0.295 | 0.025 | 0 | InN1 | AC016766.1 |
| PARD3 | 0 | 0.99964936 | 0.637 | 0.282 | 0 | InN1 | PARD3 |
| AC092422.11 | 0 | 0.9992193 | 0.516 | 0.258 | 0 | InN1 | AC092422.1 |
| VWC2L1 | 0 | 0.99455092 | 0.665 | 0.304 | 0 | InN1 | VWC2L |
| LYPD6B1 | 0 | 0.98776279 | 0.488 | 0.141 | 0 | InN1 | LYPD6B |
| DPF31 | 0 | 0.97493379 | 0.566 | 0.249 | 0 | InN1 | DPF3 |
| GLRA2 | 0 | 0.96960399 | 0.542 | 0.17 | 0 | InN1 | GLRA2 |
| AL356108.1 | 0 | 0.96842815 | 0.317 | 0.033 | 0 | InN1 | AL356108.1 |
| SLC35F4 | 0 | 0.96692464 | 0.552 | 0.195 | 0 | InN1 | SLC35F4 |
| RERG | 0 | 0.96673369 | 0.527 | 0.238 | 0 | InN1 | RERG |
| CCNG21 | 0 | 0.96518266 | 0.507 | 0.236 | 0 | InN1 | CCNG2 |
| MAPK41 | 0 | 0.95652404 | 0.705 | 0.451 | 0 | InN1 | MAPK4 |
| TRMT9B1 | 0 | 0.95264398 | 0.71 | 0.382 | 0 | InN1 | TRMT9B |
| GALNT171 | 0 | 0.94723803 | 0.893 | 0.903 | 0 | InN1 | GALNT17 |
| CLVS1 | 0 | 0.94230924 | 0.793 | 0.641 | 0 | InN1 | CLVS1 |
| ARL4C | 0 | 0.94228853 | 0.557 | 0.235 | 0 | InN1 | ARL4C |
| ASIC4 | 0 | 0.93705393 | 0.452 | 0.035 | 0 | InN1 | ASIC4 |

| | | | | | | | |
|---|---|---|---|---|---|---|---|
| ANKRD29 | 0 | 0.93467695 | 0.499 | 0.161 | 0 | InN1 | ANKRD29 |
| KCNMB21 | 0 | 0.9330378 | 0.801 | 0.548 | 0 | InN1 | KCNMB2 |
| ZNF804A | 0 | 0.92747835 | 0.768 | 0.516 | 0 | InN1 | ZNF804A |
| CNR1 | 0 | 0.92334071 | 0.565 | 0.362 | 0 | InN1 | CNR1 |
| GAD1 | 0 | 0.92272689 | 0.677 | 0.239 | 0 | InN1 | GAD1 |
| REV3L | 0 | 0.91725334 | 0.864 | 0.767 | 0 | InN1 | REV3L |
| PTPRZ1 | 0 | 0.91300306 | 0.589 | 0.267 | 0 | InN1 | PTPRZ1 |
| FHOD3 | 0 | 0.91017002 | 0.78 | 0.623 | 0 | InN1 | FHOD3 |
| ADRA1A | 0 | 0.90641698 | 0.556 | 0.17 | 0 | InN1 | ADRA1A |
| ENTPD3 | 0 | 0.90329215 | 0.587 | 0.259 | 0 | InN1 | ENTPD3 |
| NLGN1 | 0 | 0.89460549 | 0.979 | 0.958 | 0 | InN1 | NLGN1 |
| HRH3 | 0 | 0.89419098 | 0.427 | 0.116 | 0 | InN1 | HRH3 |
| CNTNAP41 | 0 | 0.89220231 | 0.769 | 0.562 | 0 | InN1 | CNTNAP4 |
| IQGAP2 | 0 | 0.8804182 | 0.257 | 0.028 | 0 | InN1 | IQGAP2 |
| LYPD6 | 0 | 0.87870912 | 0.403 | 0.089 | 0 | InN1 | LYPD6 |
| CNTNAP2 | 0 | 0.87690416 | 0.995 | 0.993 | 0 | InN1 | CNTNAP2 |
| PREX2 | 0 | 0.87675917 | 0.5 | 0.137 | 0 | InN1 | PREX2 |
| OSBPL3 | 0 | 0.87592091 | 0.648 | 0.411 | 0 | InN1 | OSBPL3 |
| HIPK2 | 0 | 0.87025569 | 0.607 | 0.334 | 0 | InN1 | HIPK2 |
| CHRNA2 | 0 | 0.85944518 | 0.383 | 0.01 | 0 | InN1 | CHRNA2 |
| NHS | 0 | 0.85226852 | 0.718 | 0.417 | 0 | InN1 | NHS |
| SLC44A51 | 0 | 0.8449089 | 0.843 | 0.677 | 0 | InN1 | SLC44A5 |
| NR3C2 | 0 | 0.84401844 | 0.881 | 0.796 | 0 | InN1 | NR3C2 |
| AP1S2 | 0 | 0.84264094 | 0.489 | 0.167 | 0 | InN1 | AP1S2 |
| ADAMTS17 | 0 | 0.83835137 | 0.454 | 0.201 | 0 | InN1 | ADAMTS17 |
| SLC4A71 | 0 | 0.83347657 | 0.634 | 0.424 | 0 | InN1 | SLC4A7 |
| RIN21 | 0 | 0.8255659 | 0.501 | 0.175 | 0 | InN1 | RIN2 |
| KANK1 | 0 | 0.81521846 | 0.358 | 0.121 | 0 | InN1 | KANK1 |
| CNTN61 | 0 | 0.81306738 | 0.418 | 0.211 | 0 | InN1 | CNTN6 |
| TCF4 | 0 | 0.80777291 | 0.962 | 0.957 | 0 | InN1 | TCF4 |
| AC022523.1 | 0 | 0.80703648 | 0.305 | 0.028 | 0 | InN1 | AC022523.1 |
| NRXN3 | 0 | 0.79992739 | 0.996 | 0.99 | 0 | InN1 | NRXN3 |
| LCNL1 | 0 | 0.79841096 | 0.313 | 0.118 | 0 | InN1 | LCNL1 |
| CRB1 | 0 | 0.79707919 | 0.536 | 0.287 | 0 | InN1 | CRB1 |
| ANK1 | 0 | 0.78827021 | 0.624 | 0.281 | 0 | InN1 | ANK1 |
| ANGPT1 | 0 | 0.78739799 | 0.271 | 0.059 | 0 | InN1 | ANGPT1 |
| KCNIP1 | 0 | 0.78483535 | 0.644 | 0.264 | 0 | InN1 | KCNIP1 |
| SCG2 | 0 | 0.78476628 | 0.454 | 0.239 | 0 | InN1 | SCG2 |
| ROBO21 | 0 | 0.78373659 | 0.985 | 0.836 | 0 | InN1 | ROBO2 |
| AFAP1 | 0 | 0.77428309 | 0.613 | 0.371 | 0 | InN1 | AFAP1 |
| SLC6A1 | 0 | 0.77240907 | 0.75 | 0.306 | 0 | InN1 | SLC6A1 |
| ZEB21 | 0 | 0.77052132 | 0.877 | 0.816 | 0 | InN1 | ZEB2 |
| STARD132 | 0 | 0.76991188 | 0.701 | 0.521 | 0 | InN1 | STARD13 |
| SMARCA2 | 0 | 0.76736513 | 0.861 | 0.801 | 0 | InN1 | SMARCA2 |
| DNER | 0 | 0.76661026 | 0.761 | 0.5 | 0 | InN1 | DNER |
| NR2F2 | 0 | 0.7658951 | 0.407 | 0.077 | 0 | InN1 | NR2F2 |
| DSCAM3 | 0 | 0.76585781 | 0.979 | 0.896 | 0 | InN1 | DSCAM |
| PDE9A | 0 | 0.7595977 | 0.324 | 0.065 | 0 | InN1 | PDE9A |
| RXRA | 0 | 0.75004659 | 0.447 | 0.179 | 0 | InN1 | RXRA |
| COL11A12 | 0 | 0.7482112 | 0.459 | 0.236 | 0 | InN1 | COL11A1 |
| AFDN1 | 0 | 0.74068502 | 0.748 | 0.639 | 0 | InN1 | AFDN |
| CPNE2 | 0 | 0.73700171 | 0.358 | 0.05 | 0 | InN1 | CPNE2 |
| HTRA11 | 0 | 0.73192271 | 0.41 | 0.135 | 0 | InN1 | HTRA1 |
| AL391832.4 | 0 | 0.73109192 | 0.324 | 0.035 | 0 | InN1 | AL391832.4 |
| PLPPR1 | 0 | 0.73068839 | 0.569 | 0.263 | 0 | InN1 | PLPPR1 |
| SNTG13 | 0 | 0.72830885 | 0.977 | 0.938 | 0 | InN1 | SNTG1 |
| ENAH | 0 | 0.72121754 | 0.657 | 0.528 | 0 | InN1 | ENAH |
| PWRN4 | 0 | 0.71469442 | 0.459 | 0.197 | 0 | InN1 | PWRN4 |
| SMOC1 | 0 | 0.71420556 | 0.283 | 0.066 | 0 | InN1 | SMOC1 |
| CNTN41 | 0 | 0.70810195 | 0.916 | 0.844 | 0 | InN1 | CNTN4 |
| LRP1B3 | 0 | 0.70764584 | 0.986 | 0.946 | 0 | InN1 | LRP1B |
| FOXN31 | 0 | 0.70681105 | 0.805 | 0.696 | 0 | InN1 | FOXN3 |
| VOPP11 | 0 | 0.69706835 | 0.633 | 0.453 | 0 | InN1 | VOPP1 |

| | | | | | | | |
|---|---|---|---|---|---|---|---|
| ARHGAP241 | 0 | 0.69291799 | 0.35 | 0.126 | 0 | InN1 | ARHGAP24 |
| NPR3 | 0 | 0.69126585 | 0.296 | 0.011 | 0 | InN1 | NPR3 |
| CACNG4 | 0 | 0.69027031 | 0.417 | 0.128 | 0 | InN1 | CACNG4 |
| LRRC4C2 | 0 | 0.68908539 | 0.943 | 0.883 | 0 | InN1 | LRRC4C |
| NCOA1 | 0 | 0.68866068 | 0.892 | 0.874 | 0 | InN1 | NCOA1 |
| OSBPL1A | 0 | 0.68738185 | 0.818 | 0.769 | 0 | InN1 | OSBPL1A |
| LPP1 | 0 | 0.68718671 | 0.64 | 0.455 | 0 | InN1 | LPP |
| ZNF608 | 0 | 0.67640771 | 0.586 | 0.33 | 0 | InN1 | ZNF608 |
| PPP1R1C | 0 | 0.67233693 | 0.265 | 0.016 | 0 | InN1 | PPP1R1C |
| CHRNA7 | 0 | 0.67182025 | 0.362 | 0.158 | 0 | InN1 | CHRNA7 |
| PPM1H | 0 | 0.67018252 | 0.792 | 0.711 | 0 | InN1 | PPM1H |
| VWC2 | 0 | 0.66939485 | 0.587 | 0.329 | 0 | InN1 | VWC2 |
| AC009093.2 | 0 | 0.66637488 | 0.344 | 0.109 | 0 | InN1 | AC009093.2 |
| RIPOR2 | 0 | 0.66009108 | 0.656 | 0.445 | 0 | InN1 | RIPOR2 |
| TGFB2 | 0 | 0.65950133 | 0.368 | 0.133 | 0 | InN1 | TGFB2 |
| FAT4 | 0 | 0.65189741 | 0.37 | 0.136 | 0 | InN1 | FAT4 |
| TTC28 | 0 | 0.65185502 | 0.845 | 0.782 | 0 | InN1 | TTC28 |
| HIF3A | 0 | 0.64775609 | 0.281 | 0.047 | 0 | InN1 | HIF3A |
| LINC01102 | 0 | 0.64671132 | 0.53 | 0.301 | 0 | InN1 | LINC01102 |
| ASIC1 | 0 | 0.64160395 | 0.404 | 0.162 | 0 | InN1 | ASIC1 |
| ADCYAP1R1 | 0 | 0.63783564 | 0.517 | 0.276 | 0 | InN1 | ADCYAP1R1 |
| MTUS11 | 0 | 0.6362016 | 0.528 | 0.271 | 0 | InN1 | MTUS1 |
| TIAM1 | 0 | 0.63357895 | 0.854 | 0.743 | 0 | InN1 | TIAM1 |
| GRM73 | 0 | 0.62805628 | 0.941 | 0.854 | 0 | InN1 | GRM7 |
| CACNA1D | 0 | 0.62694901 | 0.897 | 0.887 | 0 | InN1 | CACNA1D |
| LARGE11 | 0 | 0.62562738 | 0.931 | 0.909 | 0 | InN1 | LARGE1 |
| QKI1 | 0 | 0.61912403 | 0.828 | 0.654 | 0 | InN1 | QKI |
| TNIK1 | 0 | 0.61844169 | 0.853 | 0.799 | 0 | InN1 | TNIK |
| LRRC39 | 0 | 0.6177865 | 0.374 | 0.138 | 0 | InN1 | LRRC39 |
| FGD5 | 0 | 0.61702351 | 0.307 | 0.046 | 0 | InN1 | FGD5 |
| INHBA-AS1 | 0 | 0.6154444 | 0.383 | 0.123 | 0 | InN1 | INHBA-AS1 |
| ATP1B2 | 0 | 0.61427109 | 0.458 | 0.205 | 0 | InN1 | ATP1B2 |
| TNS31 | 0 | 0.61344265 | 0.265 | 0.091 | 0 | InN1 | TNS3 |
| GRIN2D | 0 | 0.61057051 | 0.395 | 0.113 | 0 | InN1 | GRIN2D |
| MTSS11 | 0 | 0.60977623 | 0.739 | 0.549 | 0 | InN1 | MTSS1 |
| TIMP2 | 0 | 0.60907747 | 0.508 | 0.279 | 0 | InN1 | TIMP2 |
| HIP1 | 0 | 0.60339888 | 0.381 | 0.165 | 0 | InN1 | HIP1 |
| KLHL13 | 0 | 0.59841997 | 0.323 | 0.098 | 0 | InN1 | KLHL13 |
| RGS8 | 0 | 0.59218506 | 0.312 | 0.047 | 0 | InN1 | RGS8 |
| SOBP | 0 | 0.5869459 | 0.848 | 0.816 | 0 | InN1 | SOBP |
| IGF1 | 0 | 0.58663538 | 0.381 | 0.124 | 0 | InN1 | IGF1 |
| DGKD | 0 | 0.58500387 | 0.561 | 0.282 | 0 | InN1 | DGKD |
| NINL | 0 | 0.58258334 | 0.407 | 0.161 | 0 | InN1 | NINL |
| NFIB | 0 | 0.57807808 | 0.829 | 0.628 | 0 | InN1 | NFIB |
| DPYSL3 | 0 | 0.57612285 | 0.379 | 0.174 | 0 | InN1 | DPYSL3 |
| GCNT21 | 0 | 0.57580048 | 0.305 | 0.046 | 0 | InN1 | GCNT2 |
| SNRPN | 0 | 0.57384721 | 0.855 | 0.751 | 0 | InN1 | SNRPN |
| EMID1 | 0 | 0.57265655 | 0.286 | 0.076 | 0 | InN1 | EMID1 |
| PXDN | 0 | 0.5671947 | 0.314 | 0.095 | 0 | InN1 | PXDN |
| USP31 | 0 | 0.56718599 | 0.387 | 0.149 | 0 | InN1 | USP3 |
| SMAD9 | 0 | 0.56573657 | 0.297 | 0.105 | 0 | InN1 | SMAD9 |
| SILC1 | 0 | 0.5657036 | 0.451 | 0.191 | 0 | InN1 | SILC1 |
| EPS81 | 0 | 0.56312729 | 0.302 | 0.1 | 0 | InN1 | EPS8 |
| INSYN2A | 0 | 0.56009165 | 0.278 | 0.076 | 0 | InN1 | INSYN2A |
| AC092691.13 | 0 | 0.55844413 | 0.927 | 0.862 | 0 | InN1 | AC092691.1 |
| ELFN1 | 0 | 0.54799011 | 0.309 | 0.057 | 0 | InN1 | ELFN1 |
| SALL1 | 0 | 0.54382822 | 0.28 | 0.022 | 0 | InN1 | SALL1 |
| FRMD4A | 0 | 0.54329595 | 0.986 | 0.988 | 0 | InN1 | FRMD4A |
| NCAM1 | 0 | 0.54051722 | 0.941 | 0.937 | 0 | InN1 | NCAM1 |
| SLC25A48 | 0 | 0.5356701 | 0.33 | 0.098 | 0 | InN1 | SLC25A48 |
| ADAM33 | 0 | 0.53178005 | 0.257 | 0.027 | 0 | InN1 | ADAM33 |
| SLC47A1 | 0 | 0.5258528 | 0.306 | 0.085 | 0 | InN1 | SLC47A1 |
| PKP2 | 0 | 0.51907075 | 0.328 | 0.081 | 0 | InN1 | PKP2 |

| | | | | | | | |
|---|---|---|---|---|---|---|---|
| ANO1 | 0 | 0.50582266 | 0.25 | 0.047 | 0 | InN1 | ANO1 |
| MIR99AHG2 | 0 | 0.49767874 | 0.927 | 0.891 | 0 | InN1 | MIR99AHG |
| SLC5A8 | 0 | 0.48440852 | 0.263 | 0.053 | 0 | InN1 | SLC5A8 |
| TMEM200C | 0 | 0.43923922 | 0.262 | 0.061 | 0 | InN1 | TMEM200C |
| BCL61 | 0 | 0.43345454 | 0.271 | 0.093 | 0 | InN1 | BCL6 |
| MARCKSL1 | 0 | 0.40449249 | 0.277 | 0.1 | 0 | InN1 | MARCKSL1 |
| DLX1 | 0 | 0.40177445 | 0.277 | 0.078 | 0 | InN1 | DLX1 |
| FRMPD3 | 0 | 0.3861395 | 0.271 | 0.09 | 0 | InN1 | FRMPD3 |
| CXCL14 | 0 | 0.27976868 | 0.445 | 0.121 | 0 | InN1 | CXCL14 |
| ZNF385D1 | 0 | 0.26769526 | 0.84 | 0.474 | 0 | InN1 | ZNF385D |
| KCNJ3 | 8.41E-308 | 0.57212889 | 0.874 | 0.869 | 3.21E-303 | InN1 | KCNJ3 |
| GNAS | 1.14E-307 | 0.66591566 | 0.835 | 0.824 | 4.36E-303 | InN1 | GNAS |
| GPRIN31 | 9.18E-305 | 0.4481685 | 0.364 | 0.156 | 3.51E-300 | InN1 | GPRIN3 |
| PARM11 | 8.98E-304 | 0.71821478 | 0.574 | 0.377 | 3.43E-299 | InN1 | PARM1 |
| PLS3 | 5.33E-303 | 0.72447072 | 0.373 | 0.181 | 2.04E-298 | InN1 | PLS3 |
| NFIX | 4.46E-300 | 0.62262772 | 0.742 | 0.619 | 1.71E-295 | InN1 | NFIX |
| DOCK111 | 6.47E-294 | 0.50862974 | 0.404 | 0.19 | 2.47E-289 | InN1 | DOCK11 |
| COL12A12 | 4.15E-293 | 0.55646259 | 0.38 | 0.177 | 1.59E-288 | InN1 | COL12A1 |
| B3GAT1 | 3.01E-288 | 0.66329486 | 0.631 | 0.473 | 1.15E-283 | InN1 | B3GAT1 |
| SNRK | 1.37E-286 | 0.61977126 | 0.5 | 0.318 | 5.25E-282 | InN1 | SNRK |
| NEDD4L1 | 2.80E-283 | 0.51776095 | 0.868 | 0.808 | 1.07E-278 | InN1 | NEDD4L |
| PPM1E | 3.57E-281 | 0.61679939 | 0.815 | 0.755 | 1.36E-276 | InN1 | PPM1E |
| PCDH8 | 1.02E-280 | 0.42740644 | 0.264 | 0.101 | 3.89E-276 | InN1 | PCDH8 |
| STIM1 | 1.28E-280 | 0.68454128 | 0.637 | 0.496 | 4.89E-276 | InN1 | STIM1 |
| TBL1X1 | 1.74E-278 | 0.60968634 | 0.306 | 0.132 | 6.64E-274 | InN1 | TBL1X |
| GAB21 | 3.34E-277 | 0.64578899 | 0.652 | 0.488 | 1.28E-272 | InN1 | GAB2 |
| UBASH3B | 6.50E-277 | 0.4750319 | 0.55 | 0.316 | 2.48E-272 | InN1 | UBASH3B |
| CA8 | 1.87E-275 | 0.5697372 | 0.26 | 0.1 | 7.16E-271 | InN1 | CA8 |
| CIT1 | 3.27E-275 | 0.58274532 | 0.82 | 0.745 | 1.25E-270 | InN1 | CIT |
| SEZ6L1 | 5.25E-275 | 0.62595464 | 0.813 | 0.718 | 2.01E-270 | InN1 | SEZ6L |
| AL590705.1 | 7.38E-273 | 0.63132952 | 0.416 | 0.235 | 2.82E-268 | InN1 | AL590705.1 |
| JAK1 | 8.13E-272 | 0.66733097 | 0.787 | 0.739 | 3.11E-267 | InN1 | JAK1 |
| PLCB1 | 2.71E-271 | 0.42903811 | 0.969 | 0.98 | 1.04E-266 | InN1 | PLCB1 |
| CCNI | 1.58E-268 | 0.75000433 | 0.62 | 0.502 | 6.06E-264 | InN1 | CCNI |
| MAF1 | 3.32E-268 | 0.3390571 | 0.373 | 0.164 | 1.27E-263 | InN1 | MAF |
| LCORL | 8.38E-267 | 0.74084628 | 0.625 | 0.481 | 3.20E-262 | InN1 | LCORL |
| KCNQ1OT1 | 8.46E-265 | 0.77262142 | 0.632 | 0.493 | 3.23E-260 | InN1 | KCNQ1OT1 |
| PODXL2 | 4.48E-263 | 0.57598073 | 0.552 | 0.373 | 1.71E-258 | InN1 | PODXL2 |
| DAB11 | 6.98E-262 | 0.38886699 | 0.956 | 0.87 | 2.67E-257 | InN1 | DAB1 |
| LSAMP1 | 8.54E-258 | 0.38993745 | 0.982 | 0.974 | 3.26E-253 | InN1 | LSAMP |
| RUNX1T11 | 1.23E-256 | 0.52078105 | 0.808 | 0.739 | 4.72E-252 | InN1 | RUNX1T1 |
| LRFN21 | 1.34E-254 | 0.70772494 | 0.767 | 0.679 | 5.12E-250 | InN1 | LRFN2 |
| ANKH | 4.00E-252 | 0.55891031 | 0.803 | 0.786 | 1.53E-247 | InN1 | ANKH |
| MIR4300HG | 2.12E-251 | 0.90055641 | 0.274 | 0.118 | 8.11E-247 | InN1 | MIR4300HG |
| CHRM31 | 2.13E-250 | 0.49239377 | 0.942 | 0.87 | 8.14E-246 | InN1 | CHRM3 |
| LARP1 | 3.29E-250 | 0.60380655 | 0.728 | 0.663 | 1.26E-245 | InN1 | LARP1 |
| LINC00461 | 1.23E-249 | 0.59332041 | 0.808 | 0.783 | 4.70E-245 | InN1 | LINC00461 |
| SHTN11 | 9.67E-245 | 0.58560867 | 0.749 | 0.667 | 3.70E-240 | InN1 | SHTN1 |
| DSEL | 1.45E-242 | 0.46764632 | 0.266 | 0.113 | 5.52E-238 | InN1 | DSEL |
| TRPC6 | 1.49E-239 | 0.64321447 | 0.274 | 0.118 | 5.70E-235 | InN1 | TRPC6 |
| FXYD6 | 1.70E-239 | 0.69941986 | 0.678 | 0.613 | 6.51E-235 | InN1 | FXYD6 |
| N4BP2 | 4.40E-239 | 0.61734605 | 0.516 | 0.348 | 1.68E-234 | InN1 | N4BP2 |
| GRID2 | 3.45E-238 | 0.65800547 | 0.847 | 0.722 | 1.32E-233 | InN1 | GRID2 |
| KAZN | 9.45E-237 | 0.29251627 | 0.987 | 0.949 | 3.61E-232 | InN1 | KAZN |
| PANTR1 | 4.40E-236 | 0.62809048 | 0.657 | 0.529 | 1.68E-231 | InN1 | PANTR1 |
| CHST151 | 6.55E-235 | 0.84557219 | 0.54 | 0.405 | 2.50E-230 | InN1 | CHST15 |
| TP53I11 | 7.32E-235 | 0.38989344 | 0.279 | 0.123 | 2.80E-230 | InN1 | TP53I11 |
| IRS1 | 1.11E-228 | 0.49193116 | 0.378 | 0.207 | 4.26E-224 | InN1 | IRS1 |
| ARHGAP5 | 1.17E-227 | 0.6042266 | 0.666 | 0.583 | 4.49E-223 | InN1 | ARHGAP5 |
| SYT14 | 1.72E-226 | 0.52688155 | 0.797 | 0.78 | 6.56E-222 | InN1 | SYT14 |
| LINC009231 | 1.12E-224 | 0.62197851 | 0.418 | 0.236 | 4.29E-220 | InN1 | LINC00923 |
| SGCZ1 | 5.46E-223 | 0.68871721 | 0.658 | 0.497 | 2.09E-218 | InN1 | SGCZ |
| ARHGEF26-A | 4.32E-222 | 0.53510488 | 0.27 | 0.12 | 1.65E-217 | InN1 | ARHGEF26-A |

| | | | | | | | |
|---|---|---|---|---|---|---|---|
| FRMD51 | 2.42E-221 | 0.56926585 | 0.897 | 0.861 | 9.25E-217 | InN1 | FRMD5 |
| ATP1B1 | 1.35E-220 | 0.65614405 | 0.775 | 0.781 | 5.18E-216 | InN1 | ATP1B1 |
| TRAF5 | 7.44E-220 | 0.3827645 | 0.272 | 0.123 | 2.84E-215 | InN1 | TRAF5 |
| NAALADL2 | 1.77E-219 | 0.70745763 | 0.635 | 0.508 | 6.75E-215 | InN1 | NAALADL2 |
| AC105285.1 | 2.25E-219 | 0.42838238 | 0.253 | 0.108 | 8.61E-215 | InN1 | AC105285.1 |
| NBL1 | 8.26E-219 | 0.47239767 | 0.348 | 0.186 | 3.16E-214 | InN1 | NBL1 |
| BACH11 | 4.93E-218 | 0.43852011 | 0.399 | 0.215 | 1.88E-213 | InN1 | BACH1 |
| CHST111 | 5.01E-216 | 0.58850527 | 0.649 | 0.499 | 1.91E-211 | InN1 | CHST11 |
| AC008691.11 | 3.53E-215 | 0.79361565 | 0.317 | 0.16 | 1.35E-210 | InN1 | AC008691.1 |
| AL445250.11 | 1.74E-211 | 0.74406679 | 0.491 | 0.326 | 6.65E-207 | InN1 | AL445250.1 |
| TENM3 | 3.38E-211 | 0.50365274 | 0.779 | 0.662 | 1.29E-206 | InN1 | TENM3 |
| ST8SIA51 | 1.19E-209 | 0.60842263 | 0.703 | 0.614 | 4.57E-205 | InN1 | ST8SIA5 |
| PROS1 | 9.50E-208 | 0.53700058 | 0.302 | 0.153 | 3.63E-203 | InN1 | PROS1 |
| MT-CO2 | 1.55E-206 | 0.58437627 | 0.922 | 0.896 | 5.92E-202 | InN1 | MT-CO2 |
| ZNF322 | 1.16E-203 | 0.53913829 | 0.328 | 0.177 | 4.44E-199 | InN1 | ZNF322 |
| AUTS2 | 1.85E-202 | 0.33491205 | 0.963 | 0.975 | 7.08E-198 | InN1 | AUTS2 |
| ZNF292 | 1.49E-201 | 0.49437762 | 0.818 | 0.814 | 5.70E-197 | InN1 | ZNF292 |
| ASIC21 | 4.77E-201 | 0.48744821 | 0.877 | 0.845 | 1.82E-196 | InN1 | ASIC2 |
| ARHGAP122 | 9.66E-201 | 0.50024164 | 0.485 | 0.319 | 3.69E-196 | InN1 | ARHGAP12 |
| GTF2IRD1 | 7.38E-199 | 0.60397826 | 0.576 | 0.441 | 2.82E-194 | InN1 | GTF2IRD1 |
| USP13 | 2.72E-198 | 0.52990687 | 0.501 | 0.349 | 1.04E-193 | InN1 | USP13 |
| RANBP17 | 4.14E-198 | 0.51088321 | 0.624 | 0.454 | 1.58E-193 | InN1 | RANBP17 |
| ECE2 | 6.55E-197 | 0.45016021 | 0.38 | 0.213 | 2.51E-192 | InN1 | ECE2 |
| AC092640.1 | 2.45E-196 | 0.42569107 | 0.25 | 0.109 | 9.37E-192 | InN1 | AC092640.1 |
| RPS6KA2 | 4.53E-196 | 0.33000948 | 0.914 | 0.85 | 1.73E-191 | InN1 | RPS6KA2 |
| FCHSD21 | 6.49E-196 | 0.48160976 | 0.712 | 0.603 | 2.48E-191 | InN1 | FCHSD2 |
| WWOX | 3.79E-195 | 0.52125466 | 0.847 | 0.815 | 1.45E-190 | InN1 | WWOX |
| LINC011971 | 4.39E-194 | 0.5695393 | 0.503 | 0.334 | 1.68E-189 | InN1 | LINC01197 |
| ZNF641 | 6.53E-194 | 0.39714387 | 0.294 | 0.149 | 2.50E-189 | InN1 | ZNF641 |
| TNRC6A | 4.91E-193 | 0.33645018 | 0.953 | 0.959 | 1.88E-188 | InN1 | TNRC6A |
| MYT1L | 4.84E-190 | 0.30630439 | 0.964 | 0.972 | 1.85E-185 | InN1 | MYT1L |
| MARCH31 | 1.45E-189 | 0.43397706 | 0.28 | 0.132 | 5.55E-185 | InN1 | 3-Mar |
| ANKFN1 | 7.18E-189 | 0.81121707 | 0.35 | 0.204 | 2.74E-184 | InN1 | ANKFN1 |
| PCSK51 | 3.32E-184 | 0.31198439 | 0.358 | 0.183 | 1.27E-179 | InN1 | PCSK5 |
| SOX2-OT | 6.22E-183 | 0.90275977 | 0.611 | 0.478 | 2.38E-178 | InN1 | SOX2-OT |
| KCNK21 | 3.71E-181 | 0.39605516 | 0.306 | 0.155 | 1.42E-176 | InN1 | KCNK2 |
| LANCL1 | 9.52E-181 | 0.67081508 | 0.579 | 0.493 | 3.64E-176 | InN1 | LANCL1 |
| GALNT71 | 3.10E-180 | 0.55570175 | 0.617 | 0.502 | 1.19E-175 | InN1 | GALNT7 |
| PTGDS1 | 9.66E-180 | 0.73133082 | 0.416 | 0.271 | 3.69E-175 | InN1 | PTGDS |
| IGLON5 | 6.93E-179 | 0.33038742 | 0.252 | 0.118 | 2.65E-174 | InN1 | IGLON5 |
| CPQ1 | 3.76E-176 | 0.47677873 | 0.351 | 0.203 | 1.44E-171 | InN1 | CPQ |
| MAN1A2 | 1.36E-175 | 0.5036864 | 0.761 | 0.752 | 5.21E-171 | InN1 | MAN1A2 |
| CTTNBP2 | 1.44E-174 | 0.55573331 | 0.71 | 0.675 | 5.50E-170 | InN1 | CTTNBP2 |
| SLC6A11 | 6.88E-174 | 0.50812906 | 0.384 | 0.241 | 2.63E-169 | InN1 | SLC6A11 |
| POT1-AS1 | 6.40E-173 | 0.64221004 | 0.522 | 0.381 | 2.45E-168 | InN1 | POT1-AS1 |
| AC079296.1 | 1.47E-170 | 0.35627559 | 0.322 | 0.169 | 5.60E-166 | InN1 | AC079296.1 |
| AMPD2 | 1.40E-169 | 0.40171209 | 0.33 | 0.189 | 5.36E-165 | InN1 | AMPD2 |
| OLFM2 | 2.42E-169 | 0.47757171 | 0.436 | 0.281 | 9.25E-165 | InN1 | OLFM2 |
| GRIP2 | 1.32E-168 | 0.29008229 | 0.341 | 0.173 | 5.04E-164 | InN1 | GRIP2 |
| ARNT21 | 4.56E-168 | 0.39005503 | 0.847 | 0.838 | 1.74E-163 | InN1 | ARNT2 |
| GNAL1 | 2.79E-167 | 0.43390868 | 0.658 | 0.457 | 1.07E-162 | InN1 | GNAL |
| PDE1C1 | 3.21E-166 | 0.63670654 | 0.618 | 0.483 | 1.23E-161 | InN1 | PDE1C |
| SPECC11 | 7.85E-165 | 0.49649705 | 0.776 | 0.732 | 3.00E-160 | InN1 | SPECC1 |
| C1orf21 | 6.83E-162 | 0.46779349 | 0.742 | 0.717 | 2.61E-157 | InN1 | C1orf21 |
| NRIP3 | 3.15E-160 | 0.26412533 | 0.364 | 0.202 | 1.20E-155 | InN1 | NRIP3 |
| FGD41 | 1.35E-159 | 0.49268526 | 0.462 | 0.319 | 5.17E-155 | InN1 | FGD4 |
| ANKRD30BL | 2.48E-159 | 0.48717867 | 0.785 | 0.699 | 9.48E-155 | InN1 | ANKRD30BL |
| CRY1 | 3.08E-159 | 0.45161445 | 0.306 | 0.17 | 1.18E-154 | InN1 | CRY1 |
| CSMD12 | 4.51E-159 | 0.41725024 | 0.992 | 0.98 | 1.72E-154 | InN1 | CSMD1 |
| NFIA3 | 9.65E-158 | 0.38981345 | 0.571 | 0.404 | 3.69E-153 | InN1 | NFIA |
| MT-ND4 | 2.18E-155 | 0.51212625 | 0.879 | 0.854 | 8.33E-151 | InN1 | MT-ND4 |
| TAFA5 | 1.16E-154 | 0.56842086 | 0.664 | 0.6 | 4.45E-150 | InN1 | TAFA5 |
| GPHN | 1.25E-152 | 0.35212316 | 0.928 | 0.95 | 4.77E-148 | InN1 | GPHN |

| | | | | | | | |
|---|---|---|---|---|---|---|---|
| SPOCK2 | 1.28E-152 | 0.45915959 | 0.537 | 0.424 | 4.88E-148 | InN1 | SPOCK2 |
| ZBTB161 | 1.69E-152 | 0.35945113 | 0.881 | 0.878 | 6.45E-148 | InN1 | ZBTB16 |
| INPP4B2 | 7.43E-152 | 0.40742109 | 0.821 | 0.748 | 2.84E-147 | InN1 | INPP4B |
| SMIM17 | 2.30E-151 | 0.40363263 | 0.337 | 0.201 | 8.78E-147 | InN1 | SMIM17 |
| TUSC31 | 1.59E-150 | 0.51630062 | 0.691 | 0.655 | 6.09E-146 | InN1 | TUSC3 |
| GNG72 | 5.00E-150 | 0.52947196 | 0.522 | 0.396 | 1.91E-145 | InN1 | GNG7 |
| TRAF3IP2-AS | 2.71E-148 | 0.44839531 | 0.763 | 0.748 | 1.03E-143 | InN1 | TRAF3IP2-AS |
| ELFN2 | 7.37E-146 | 0.36914436 | 0.321 | 0.189 | 2.82E-141 | InN1 | ELFN2 |
| CAMTA1 | 1.00E-145 | 0.31790867 | 0.902 | 0.929 | 3.83E-141 | InN1 | CAMTA1 |
| TNKS | 6.19E-145 | 0.41945383 | 0.805 | 0.818 | 2.37E-140 | InN1 | TNKS |
| PCDH11X | 6.44E-144 | 0.54834455 | 0.631 | 0.528 | 2.46E-139 | InN1 | PCDH11X |
| MEG8 | 5.09E-143 | 0.39189534 | 0.865 | 0.846 | 1.95E-138 | InN1 | MEG8 |
| KIF5C | 2.27E-142 | 0.38469038 | 0.841 | 0.859 | 8.67E-138 | InN1 | KIF5C |
| RARB1 | 2.59E-142 | 0.67341781 | 0.534 | 0.419 | 9.90E-138 | InN1 | RARB |
| CNTLN | 2.27E-141 | 0.44938131 | 0.355 | 0.222 | 8.68E-137 | InN1 | CNTLN |
| MACROD2 | 2.49E-141 | 0.27295437 | 0.951 | 0.951 | 9.53E-137 | InN1 | MACROD2 |
| SUSD4 | 9.49E-139 | 0.52328726 | 0.66 | 0.606 | 3.63E-134 | InN1 | SUSD4 |
| SYNE1 | 1.56E-136 | 0.32373977 | 0.945 | 0.953 | 5.95E-132 | InN1 | SYNE1 |
| HDAC92 | 5.55E-136 | 0.41557343 | 0.81 | 0.776 | 2.12E-131 | InN1 | HDAC9 |
| ASTN2 | 7.67E-136 | 0.34178936 | 0.81 | 0.729 | 2.93E-131 | InN1 | ASTN2 |
| KAT2B | 1.03E-134 | 0.48393356 | 0.427 | 0.3 | 3.94E-130 | InN1 | KAT2B |
| CDH102 | 5.44E-132 | 0.37614916 | 0.785 | 0.69 | 2.08E-127 | InN1 | CDH10 |
| SH3D19 | 3.39E-131 | 0.55192582 | 0.548 | 0.45 | 1.29E-126 | InN1 | SH3D19 |
| KLF3-AS1 | 4.27E-130 | 0.45454393 | 0.386 | 0.254 | 1.63E-125 | InN1 | KLF3-AS1 |
| OLFM33 | 3.23E-129 | 0.48168535 | 0.783 | 0.709 | 1.24E-124 | InN1 | OLFM3 |
| SYT171 | 2.99E-128 | 0.44280078 | 0.414 | 0.287 | 1.14E-123 | InN1 | SYT17 |
| BMERB1 | 1.28E-127 | 0.480548 | 0.734 | 0.724 | 4.90E-123 | InN1 | BMERB1 |
| MSI2 | 1.29E-127 | 0.36940752 | 0.719 | 0.654 | 4.93E-123 | InN1 | MSI2 |
| ERO1B | 6.20E-126 | 0.466679 | 0.508 | 0.407 | 2.37E-121 | InN1 | ERO1B |
| DENND2A | 1.76E-125 | 0.45487844 | 0.39 | 0.254 | 6.72E-121 | InN1 | DENND2A |
| DCX | 1.35E-124 | 0.43215645 | 0.504 | 0.385 | 5.18E-120 | InN1 | DCX |
| IL12RB22 | 2.68E-124 | 0.47458539 | 0.316 | 0.196 | 1.02E-119 | InN1 | IL12RB2 |
| ZFHX2-AS1 | 1.39E-123 | 0.43610049 | 0.377 | 0.256 | 5.32E-119 | InN1 | ZFHX2-AS1 |
| USP6NL1 | 2.44E-123 | 0.34580712 | 0.254 | 0.14 | 9.34E-119 | InN1 | USP6NL |
| HACD3 | 2.98E-123 | 0.47167096 | 0.567 | 0.503 | 1.14E-118 | InN1 | HACD3 |
| MT-ND31 | 1.85E-122 | 0.37400466 | 0.891 | 0.868 | 7.09E-118 | InN1 | MT-ND3 |
| CYTH11 | 1.00E-121 | 0.45138709 | 0.53 | 0.431 | 3.83E-117 | InN1 | CYTH1 |
| C3orf70 | 2.77E-121 | 0.41024208 | 0.326 | 0.208 | 1.06E-116 | InN1 | C3orf70 |
| SERPINB61 | 1.39E-119 | 0.34611341 | 0.31 | 0.191 | 5.31E-115 | InN1 | SERPINB6 |
| SLC25A131 | 3.60E-118 | 0.40141521 | 0.37 | 0.251 | 1.37E-113 | InN1 | SLC25A13 |
| HFM1 | 1.03E-117 | 0.35639349 | 0.299 | 0.18 | 3.92E-113 | InN1 | HFM1 |
| ZFHX32 | 1.60E-117 | 0.4049879 | 0.409 | 0.28 | 6.12E-113 | InN1 | ZFHX3 |
| INPP5F | 5.86E-117 | 0.42763176 | 0.626 | 0.578 | 2.24E-112 | InN1 | INPP5F |
| VAV2 | 1.16E-114 | 0.31028571 | 0.3 | 0.172 | 4.45E-110 | InN1 | VAV2 |
| ANKRD12 | 1.52E-114 | 0.38140098 | 0.78 | 0.78 | 5.80E-110 | InN1 | ANKRD12 |
| MT-ATP61 | 7.53E-114 | 0.39742311 | 0.898 | 0.877 | 2.88E-109 | InN1 | MT-ATP6 |
| POU3F3 | 3.86E-113 | 0.29339344 | 0.278 | 0.16 | 1.47E-108 | InN1 | POU3F3 |
| DENND1B | 1.92E-112 | 0.47604512 | 0.565 | 0.499 | 7.34E-108 | InN1 | DENND1B |
| WWP1 | 8.50E-112 | 0.45908609 | 0.555 | 0.478 | 3.25E-107 | InN1 | WWP1 |
| ATP1A3 | 9.88E-110 | 0.53221384 | 0.571 | 0.535 | 3.78E-105 | InN1 | ATP1A3 |
| MT-ND4L | 4.04E-107 | 0.51054893 | 0.624 | 0.536 | 1.55E-102 | InN1 | MT-ND4L |
| NGLY1 | 4.10E-107 | 0.42962528 | 0.487 | 0.405 | 1.57E-102 | InN1 | NGLY1 |
| PRKD11 | 6.11E-107 | 0.50561928 | 0.587 | 0.504 | 2.33E-102 | InN1 | PRKD1 |
| MT-CO11 | 3.27E-106 | 0.37999009 | 0.887 | 0.866 | 1.25E-101 | InN1 | MT-CO1 |
| GTF2I | 3.80E-106 | 0.32756116 | 0.811 | 0.838 | 1.45E-101 | InN1 | GTF2I |
| EEF1A11 | 6.54E-106 | 0.51540302 | 0.631 | 0.57 | 2.50E-101 | InN1 | EEF1A1 |
| TXNDC16 | 2.73E-105 | 0.44003878 | 0.593 | 0.54 | 1.04E-100 | InN1 | TXNDC16 |
| TCF121 | 4.89E-104 | 0.35264093 | 0.732 | 0.703 | 1.87E-99 | InN1 | TCF12 |
| GREB1 | 1.29E-102 | 0.40835917 | 0.437 | 0.33 | 4.95E-98 | InN1 | GREB1 |
| KIAA19581 | 6.23E-102 | 0.38986628 | 0.529 | 0.421 | 2.38E-97 | InN1 | KIAA1958 |
| SAMD4A | 3.98E-101 | 0.43557481 | 0.551 | 0.461 | 1.52E-96 | InN1 | SAMD4A |
| PDE10A1 | 1.30E-100 | 0.51715378 | 0.714 | 0.674 | 4.96E-96 | InN1 | PDE10A |
| TGFBR3 | 3.48E-100 | 0.41273895 | 0.288 | 0.18 | 1.33E-95 | InN1 | TGFBR3 |

| | | | | | | | |
|---|---|---|---|---|---|---|---|
| KLHL52 | 8.50E-99 | 0.40745364 | 0.429 | 0.307 | 3.25E-94 | InN1 | KLHL5 |
| KLHL42 | 1.13E-98 | 0.40102947 | 0.499 | 0.423 | 4.33E-94 | InN1 | KLHL42 |
| GNG21 | 1.28E-98 | 0.38995364 | 0.541 | 0.435 | 4.91E-94 | InN1 | GNG2 |
| TLE2 | 2.33E-98 | 0.34335633 | 0.299 | 0.189 | 8.90E-94 | InN1 | TLE2 |
| CCDC91 | 1.83E-97 | 0.46589794 | 0.697 | 0.697 | 6.99E-93 | InN1 | CCDC91 |
| FNBP1L1 | 3.43E-97 | 0.36380088 | 0.5 | 0.378 | 1.31E-92 | InN1 | FNBP1L |
| DIP2C | 7.00E-97 | 0.29751103 | 0.872 | 0.892 | 2.68E-92 | InN1 | DIP2C |
| MTRNR2L12 | 9.47E-97 | 0.35801631 | 0.796 | 0.742 | 3.62E-92 | InN1 | MTRNR2L12 |
| MT-CO31 | 1.51E-96 | 0.4380775 | 0.867 | 0.848 | 5.78E-92 | InN1 | MT-CO3 |
| SVIL | 6.05E-96 | 0.39391973 | 0.269 | 0.165 | 2.31E-91 | InN1 | SVIL |
| NSG2 | 1.36E-94 | 0.39061331 | 0.599 | 0.553 | 5.20E-90 | InN1 | NSG2 |
| BID | 3.11E-93 | 0.40026041 | 0.292 | 0.189 | 1.19E-88 | InN1 | BID |
| PKD1L1 | 4.09E-93 | 0.40723661 | 0.366 | 0.256 | 1.56E-88 | InN1 | PKD1L1 |
| AC106795.31 | 2.08E-92 | 0.46455523 | 0.312 | 0.211 | 7.97E-88 | InN1 | AC106795.3 |
| GSE1 | 2.33E-92 | 0.44911654 | 0.569 | 0.495 | 8.90E-88 | InN1 | GSE1 |
| SORCS32 | 7.32E-92 | 0.41084424 | 0.653 | 0.561 | 2.80E-87 | InN1 | SORCS3 |
| PDE7B1 | 7.65E-92 | 0.52688053 | 0.556 | 0.476 | 2.92E-87 | InN1 | PDE7B |
| WSCD2 | 1.02E-91 | 0.46994164 | 0.489 | 0.408 | 3.88E-87 | InN1 | WSCD2 |
| HIVEP32 | 2.15E-91 | 0.29563355 | 0.854 | 0.864 | 8.21E-87 | InN1 | HIVEP3 |
| RGS63 | 2.45E-91 | 0.61317776 | 0.77 | 0.697 | 9.37E-87 | InN1 | RGS6 |
| APP | 9.98E-89 | 0.299943 | 0.884 | 0.916 | 3.81E-84 | InN1 | APP |
| CCK | 1.89E-88 | 1.02205019 | 0.497 | 0.452 | 7.21E-84 | InN1 | CCK |
| ALDOC | 2.02E-88 | 0.33480989 | 0.269 | 0.171 | 7.72E-84 | InN1 | ALDOC |
| PDIA2 | 3.21E-88 | 0.6072487 | 0.446 | 0.366 | 1.23E-83 | InN1 | PDIA2 |
| RALGPS1 | 4.87E-87 | 0.3527015 | 0.714 | 0.708 | 1.86E-82 | InN1 | RALGPS1 |
| THSD7B | 1.18E-86 | 0.65651646 | 0.25 | 0.153 | 4.51E-82 | InN1 | THSD7B |
| HSP90AB1 | 1.52E-86 | 0.3892541 | 0.726 | 0.728 | 5.83E-82 | InN1 | HSP90AB1 |
| SDK21 | 4.44E-84 | 0.42590519 | 0.326 | 0.224 | 1.70E-79 | InN1 | SDK2 |
| PTPRJ1 | 6.41E-84 | 0.39600768 | 0.649 | 0.625 | 2.45E-79 | InN1 | PTPRJ |
| ATP6V0E2 | 7.65E-84 | 0.41524758 | 0.43 | 0.351 | 2.92E-79 | InN1 | ATP6V0E2 |
| NMNAT2 | 7.65E-84 | 0.28416492 | 0.834 | 0.856 | 2.93E-79 | InN1 | NMNAT2 |
| MAST41 | 1.24E-82 | 0.29510562 | 0.826 | 0.818 | 4.74E-78 | InN1 | MAST4 |
| FBXO31 | 6.31E-82 | 0.44984666 | 0.393 | 0.312 | 2.41E-77 | InN1 | FBXO31 |
| ARHGAP18 | 3.81E-81 | 0.56241276 | 0.295 | 0.207 | 1.46E-76 | InN1 | ARHGAP18 |
| SLC22A17 | 5.59E-81 | 0.45614242 | 0.587 | 0.571 | 2.14E-76 | InN1 | SLC22A17 |
| AC004083.1 | 9.19E-81 | 0.48594163 | 0.385 | 0.293 | 3.51E-76 | InN1 | AC004083.1 |
| AC068587.42 | 9.54E-81 | 0.33397187 | 0.569 | 0.508 | 3.65E-76 | InN1 | AC068587.4 |
| TUT4 | 3.38E-80 | 0.36988085 | 0.705 | 0.702 | 1.29E-75 | InN1 | TUT4 |
| ZNF483 | 4.78E-80 | 0.35343778 | 0.737 | 0.752 | 1.83E-75 | InN1 | ZNF483 |
| L3MBTL1 | 5.70E-80 | 0.3622107 | 0.446 | 0.362 | 2.18E-75 | InN1 | L3MBTL1 |
| NSG1 | 1.31E-78 | 0.26562894 | 0.265 | 0.172 | 5.01E-74 | InN1 | NSG1 |
| SLC4A3 | 1.50E-78 | 0.29770397 | 0.291 | 0.198 | 5.72E-74 | InN1 | SLC4A3 |
| CDC42EP3 | 3.59E-78 | 0.27240656 | 0.268 | 0.173 | 1.37E-73 | InN1 | CDC42EP3 |
| GSAP1 | 1.00E-77 | 0.34825059 | 0.339 | 0.247 | 3.83E-73 | InN1 | GSAP |
| GABRA1 | 1.09E-77 | 0.39200372 | 0.646 | 0.644 | 4.16E-73 | InN1 | GABRA1 |
| CCDC85A2 | 3.15E-77 | 0.37570512 | 0.697 | 0.653 | 1.20E-72 | InN1 | CCDC85A |
| TFDP2 | 4.08E-77 | 0.38294421 | 0.733 | 0.741 | 1.56E-72 | InN1 | TFDP2 |
| ANO10 | 2.76E-76 | 0.40334891 | 0.579 | 0.547 | 1.06E-71 | InN1 | ANO10 |
| SMARCD3 | 7.51E-76 | 0.37847499 | 0.435 | 0.344 | 2.87E-71 | InN1 | SMARCD3 |
| ATRX | 2.12E-75 | 0.28378292 | 0.814 | 0.839 | 8.10E-71 | InN1 | ATRX |
| GPR137C | 2.69E-75 | 0.3572004 | 0.633 | 0.605 | 1.03E-70 | InN1 | GPR137C |
| POLB | 9.05E-74 | 0.33938777 | 0.292 | 0.205 | 3.46E-69 | InN1 | POLB |
| SIPA1L3 | 4.48E-73 | 0.40557657 | 0.599 | 0.556 | 1.71E-68 | InN1 | SIPA1L3 |
| KCNH74 | 6.30E-73 | 0.25250484 | 0.815 | 0.721 | 2.41E-68 | InN1 | KCNH7 |
| LINC01551 | 8.88E-73 | 0.40648348 | 0.441 | 0.374 | 3.39E-68 | InN1 | LINC01551 |
| CDH201 | 9.45E-73 | 0.57400359 | 0.459 | 0.376 | 3.61E-68 | InN1 | CDH20 |
| CYFIP2 | 9.89E-73 | 0.35082818 | 0.696 | 0.713 | 3.78E-68 | InN1 | CYFIP2 |
| ARHGAP21 | 2.78E-72 | 0.3215735 | 0.819 | 0.843 | 1.06E-67 | InN1 | ARHGAP21 |
| OSBPL2 | 9.10E-72 | 0.30400082 | 0.294 | 0.205 | 3.48E-67 | InN1 | OSBPL2 |
| MAP71 | 1.27E-70 | 0.25888615 | 0.866 | 0.883 | 4.87E-66 | InN1 | MAP7 |
| KCNMB2-AS1 | 2.82E-70 | 0.29056269 | 0.316 | 0.221 | 1.08E-65 | InN1 | KCNMB2-AS1 |
| SCMH1 | 2.85E-70 | 0.32664778 | 0.718 | 0.728 | 1.09E-65 | InN1 | SCMH1 |
| PLCB4 | 3.05E-70 | 0.33213696 | 0.615 | 0.558 | 1.16E-65 | InN1 | PLCB4 |

| | | | | | | | |
|---|---|---|---|---|---|---|---|
| EPHB1 | 1.56E-69 | 0.48976983 | 0.534 | 0.494 | 5.95E-65 | InN1 | EPHB1 |
| GPR155 | 1.65E-69 | 0.36817158 | 0.614 | 0.6 | 6.32E-65 | InN1 | GPR155 |
| BCL9 | 6.35E-69 | 0.29087437 | 0.301 | 0.213 | 2.43E-64 | InN1 | BCL9 |
| KLHL23 | 1.39E-68 | 0.38957867 | 0.505 | 0.457 | 5.32E-64 | InN1 | KLHL23 |
| PACSIN21 | 1.57E-68 | 0.37338834 | 0.42 | 0.343 | 5.99E-64 | InN1 | PACSIN2 |
| CDK5RAP2 | 4.24E-68 | 0.41630791 | 0.511 | 0.465 | 1.62E-63 | InN1 | CDK5RAP2 |
| EXT1 | 8.12E-68 | 0.34850528 | 0.783 | 0.818 | 3.10E-63 | InN1 | EXT1 |
| LDLRAD42 | 6.28E-67 | 0.34173427 | 0.786 | 0.782 | 2.40E-62 | InN1 | LDLRAD4 |
| ZHX21 | 1.15E-66 | 0.36175456 | 0.305 | 0.215 | 4.41E-62 | InN1 | ZHX2 |
| HSP90AA11 | 1.20E-66 | 0.41809543 | 0.707 | 0.713 | 4.57E-62 | InN1 | HSP90AA1 |
| NAAA | 1.94E-66 | 0.30423291 | 0.289 | 0.203 | 7.41E-62 | InN1 | NAAA |
| MYRIP1 | 2.05E-66 | 0.3995885 | 0.8 | 0.808 | 7.84E-62 | InN1 | MYRIP |
| BICD1 | 3.78E-66 | 0.26215086 | 0.79 | 0.795 | 1.44E-61 | InN1 | BICD1 |
| LRCH2 | 3.96E-66 | 0.36522722 | 0.404 | 0.334 | 1.51E-61 | InN1 | LRCH2 |
| MAP1B2 | 6.48E-66 | 0.28110836 | 0.876 | 0.902 | 2.48E-61 | InN1 | MAP1B |
| ARFGEF31 | 8.40E-66 | 0.27560072 | 0.775 | 0.781 | 3.21E-61 | InN1 | ARFGEF3 |
| TPP2 | 1.27E-65 | 0.38940499 | 0.529 | 0.495 | 4.86E-61 | InN1 | TPP2 |
| KLF121 | 1.32E-65 | 0.46144633 | 0.628 | 0.612 | 5.06E-61 | InN1 | KLF12 |
| NAP1L3 | 1.70E-65 | 0.3185686 | 0.379 | 0.298 | 6.50E-61 | InN1 | NAP1L3 |
| SRRM41 | 1.91E-65 | 0.2696432 | 0.844 | 0.857 | 7.29E-61 | InN1 | SRRM4 |
| ACACA | 3.91E-65 | 0.31159984 | 0.777 | 0.792 | 1.50E-60 | InN1 | ACACA |
| RHOQ1 | 1.31E-64 | 0.28558233 | 0.292 | 0.207 | 5.00E-60 | InN1 | RHOQ |
| AC002070.1 | 2.22E-64 | 0.39646327 | 0.265 | 0.184 | 8.47E-60 | InN1 | AC002070.1 |
| HECTD2 | 6.86E-64 | 0.38957331 | 0.522 | 0.478 | 2.62E-59 | InN1 | HECTD2 |
| PXDNL | 1.12E-63 | 0.33772056 | 0.304 | 0.221 | 4.28E-59 | InN1 | PXDNL |
| SEM1 | 2.37E-63 | 0.41756692 | 0.524 | 0.478 | 9.06E-59 | InN1 | SEM1 |
| DLGAP4 | 3.61E-63 | 0.35841769 | 0.689 | 0.681 | 1.38E-58 | InN1 | DLGAP4 |
| TMEM145 | 4.70E-63 | 0.34044909 | 0.413 | 0.34 | 1.80E-58 | InN1 | TMEM145 |
| TEX21 | 8.64E-63 | 0.3603982 | 0.417 | 0.347 | 3.30E-58 | InN1 | TEX2 |
| CNTRL | 1.25E-62 | 0.39833292 | 0.445 | 0.385 | 4.78E-58 | InN1 | CNTRL |
| EPS15L1 | 1.42E-62 | 0.377025 | 0.575 | 0.546 | 5.41E-58 | InN1 | EPS15L1 |
| INSYN2B1 | 2.81E-62 | 0.32878393 | 0.309 | 0.221 | 1.07E-57 | InN1 | INSYN2B |
| TPST1 | 1.29E-61 | 0.36411207 | 0.684 | 0.684 | 4.93E-57 | InN1 | TPST1 |
| NEK10 | 1.89E-61 | 0.39837438 | 0.638 | 0.632 | 7.24E-57 | InN1 | NEK10 |
| RPL181 | 2.22E-61 | 0.37692673 | 0.381 | 0.308 | 8.48E-57 | InN1 | RPL18 |
| SENP7 | 4.43E-61 | 0.3570651 | 0.545 | 0.502 | 1.69E-56 | InN1 | SENP7 |
| IL1RAP1 | 7.18E-61 | 0.32411913 | 0.403 | 0.319 | 2.75E-56 | InN1 | IL1RAP |
| KIF1A | 8.88E-61 | 0.36234527 | 0.633 | 0.638 | 3.40E-56 | InN1 | KIF1A |
| RPS271 | 1.45E-60 | 0.41978902 | 0.373 | 0.3 | 5.54E-56 | InN1 | RPS27 |
| RPL131 | 2.96E-60 | 0.44558168 | 0.452 | 0.395 | 1.13E-55 | InN1 | RPL13 |
| CSPG5 | 5.39E-60 | 0.27762527 | 0.295 | 0.216 | 2.06E-55 | InN1 | CSPG5 |
| FOXG1 | 6.81E-60 | 0.35255662 | 0.355 | 0.281 | 2.60E-55 | InN1 | FOXG1 |
| KCNN31 | 1.18E-59 | 0.41724831 | 0.386 | 0.314 | 4.50E-55 | InN1 | KCNN3 |
| STAT5B | 3.96E-59 | 0.3052458 | 0.421 | 0.345 | 1.52E-54 | InN1 | STAT5B |
| TACC2 | 5.04E-59 | 0.38610874 | 0.463 | 0.408 | 1.92E-54 | InN1 | TACC2 |
| RNF180 | 2.32E-58 | 0.37780176 | 0.591 | 0.58 | 8.88E-54 | InN1 | RNF180 |
| GULP12 | 2.82E-58 | 0.53695819 | 0.415 | 0.348 | 1.08E-53 | InN1 | GULP1 |
| OSBPL9 | 3.91E-58 | 0.34193173 | 0.58 | 0.558 | 1.49E-53 | InN1 | OSBPL9 |
| KIF2A | 8.61E-58 | 0.36964618 | 0.528 | 0.499 | 3.29E-53 | InN1 | KIF2A |
| LRRC2 | 2.00E-57 | 0.280276 | 0.261 | 0.182 | 7.64E-53 | InN1 | LRRC2 |
| CTPS2 | 2.28E-57 | 0.263449 | 0.254 | 0.175 | 8.73E-53 | InN1 | CTPS2 |
| ABLIM11 | 9.36E-57 | 0.26518829 | 0.829 | 0.835 | 3.58E-52 | InN1 | ABLIM1 |
| OBSL1 | 1.87E-56 | 0.30872179 | 0.348 | 0.278 | 7.14E-52 | InN1 | OBSL1 |
| REEP1 | 5.10E-56 | 0.34413789 | 0.625 | 0.627 | 1.95E-51 | InN1 | REEP1 |
| RPL18A | 1.36E-55 | 0.28460594 | 0.258 | 0.181 | 5.18E-51 | InN1 | RPL18A |
| DIP2A1 | 4.40E-55 | 0.30957676 | 0.513 | 0.457 | 1.68E-50 | InN1 | DIP2A |
| ATCAY | 1.29E-54 | 0.34637484 | 0.605 | 0.598 | 4.95E-50 | InN1 | ATCAY |
| MT-ND21 | 1.85E-54 | 0.31515343 | 0.773 | 0.747 | 7.06E-50 | InN1 | MT-ND2 |
| CRADD | 3.10E-54 | 0.37822174 | 0.486 | 0.429 | 1.19E-49 | InN1 | CRADD |
| ALCAM1 | 1.14E-53 | 0.72316369 | 0.502 | 0.47 | 4.37E-49 | InN1 | ALCAM |
| SLC25A12 | 1.25E-53 | 0.26125046 | 0.788 | 0.823 | 4.77E-49 | InN1 | SLC25A12 |
| SRGAP21 | 7.33E-53 | 0.30170679 | 0.523 | 0.481 | 2.80E-48 | InN1 | SRGAP2 |
| CTIF | 1.35E-52 | 0.29926794 | 0.744 | 0.762 | 5.17E-48 | InN1 | CTIF |

| | | | | | | | |
|---|---|---|---|---|---|---|---|
| ALDH2 | 2.83E-52 | 0.2782456 | 0.289 | 0.214 | 1.08E-47 | InN1 | ALDH2 |
| LINC00599 | 5.88E-52 | 0.30364765 | 0.712 | 0.736 | 2.25E-47 | InN1 | LINC00599 |
| MT-CYB1 | 7.74E-52 | 0.30127448 | 0.808 | 0.785 | 2.96E-47 | InN1 | MT-CYB |
| OSTM1 | 1.15E-51 | 0.29202811 | 0.327 | 0.257 | 4.40E-47 | InN1 | OSTM1 |
| AC025159.1 | 1.98E-51 | 0.52234453 | 0.6 | 0.592 | 7.56E-47 | InN1 | AC025159.1 |
| USP11 | 3.59E-51 | 0.3421544 | 0.556 | 0.543 | 1.37E-46 | InN1 | USP11 |
| SCAPER | 4.16E-51 | 0.25092366 | 0.793 | 0.807 | 1.59E-46 | InN1 | SCAPER |
| LRIG1 | 4.28E-51 | 0.30475196 | 0.285 | 0.21 | 1.64E-46 | InN1 | LRIG1 |
| GPD21 | 5.14E-51 | 0.31174189 | 0.39 | 0.323 | 1.97E-46 | InN1 | GPD2 |
| RGS17 | 2.45E-50 | 0.36315892 | 0.411 | 0.357 | 9.37E-46 | InN1 | RGS17 |
| C3orf67-AS1 | 4.70E-50 | 0.48186103 | 0.339 | 0.278 | 1.80E-45 | InN1 | C3orf67-AS1 |
| FNDC3A | 5.56E-50 | 0.30233022 | 0.67 | 0.681 | 2.12E-45 | InN1 | FNDC3A |
| SEL1L31 | 7.59E-50 | 0.46060041 | 0.351 | 0.286 | 2.90E-45 | InN1 | SEL1L3 |
| SYT111 | 8.33E-50 | 0.32035819 | 0.634 | 0.649 | 3.18E-45 | InN1 | SYT11 |
| POU2F1 | 1.24E-49 | 0.27390399 | 0.732 | 0.761 | 4.75E-45 | InN1 | POU2F1 |
| SERINC1 | 1.28E-49 | 0.36471171 | 0.603 | 0.624 | 4.89E-45 | InN1 | SERINC1 |
| SEMA4B | 2.38E-49 | 0.32757456 | 0.403 | 0.348 | 9.11E-45 | InN1 | SEMA4B |
| ANKRD6 | 2.61E-49 | 0.31502094 | 0.477 | 0.413 | 9.96E-45 | InN1 | ANKRD6 |
| MT-ND5 | 5.28E-49 | 0.36424585 | 0.607 | 0.565 | 2.02E-44 | InN1 | MT-ND5 |
| DNAJB2 | 5.35E-49 | 0.32500104 | 0.471 | 0.426 | 2.04E-44 | InN1 | DNAJB2 |
| MAST2 | 8.09E-49 | 0.33772686 | 0.454 | 0.404 | 3.09E-44 | InN1 | MAST2 |
| RPL281 | 8.81E-49 | 0.3451208 | 0.421 | 0.363 | 3.37E-44 | InN1 | RPL28 |
| LINC009701 | 9.09E-49 | 0.47668429 | 0.445 | 0.403 | 3.47E-44 | InN1 | LINC00970 |
| PCSK1N | 1.28E-48 | 0.45340512 | 0.489 | 0.456 | 4.91E-44 | InN1 | PCSK1N |
| RHBDD1 | 1.66E-48 | 0.31898773 | 0.333 | 0.27 | 6.36E-44 | InN1 | RHBDD1 |
| PRUNE22 | 2.13E-48 | 0.25256355 | 0.634 | 0.598 | 8.15E-44 | InN1 | PRUNE2 |
| GALNT142 | 5.90E-48 | 0.35036735 | 0.393 | 0.327 | 2.25E-43 | InN1 | GALNT14 |
| OSBPL8 | 7.39E-48 | 0.34993138 | 0.722 | 0.757 | 2.82E-43 | InN1 | OSBPL8 |
| RPS281 | 8.09E-48 | 0.37464702 | 0.408 | 0.354 | 3.09E-43 | InN1 | RPS28 |
| PWWP2A | 1.29E-47 | 0.32696049 | 0.426 | 0.371 | 4.92E-43 | InN1 | PWWP2A |
| MXD4 | 1.64E-47 | 0.29652673 | 0.317 | 0.251 | 6.26E-43 | InN1 | MXD4 |
| URI1 | 1.92E-47 | 0.30490463 | 0.507 | 0.465 | 7.35E-43 | InN1 | URI1 |
| AC007364.1 | 3.33E-47 | 0.35027994 | 0.346 | 0.276 | 1.27E-42 | InN1 | AC007364.1 |
| NRXN2 | 7.88E-47 | 0.29723455 | 0.639 | 0.637 | 3.01E-42 | InN1 | NRXN2 |
| ZSWIM5 | 1.66E-46 | 0.33624478 | 0.561 | 0.545 | 6.34E-42 | InN1 | ZSWIM5 |
| CALY | 1.83E-45 | 0.3960677 | 0.549 | 0.55 | 7.00E-41 | InN1 | CALY |
| PCGF5 | 2.50E-45 | 0.28636855 | 0.628 | 0.632 | 9.54E-41 | InN1 | PCGF5 |
| ARL6IP1 | 5.00E-45 | 0.37836779 | 0.466 | 0.445 | 1.91E-40 | InN1 | ARL6IP1 |
| APC | 6.82E-45 | 0.26539666 | 0.717 | 0.745 | 2.61E-40 | InN1 | APC |
| CAMKMT1 | 8.99E-45 | 0.40323859 | 0.43 | 0.377 | 3.44E-40 | InN1 | CAMKMT |
| CHRM21 | 9.81E-44 | 0.27792719 | 0.288 | 0.211 | 3.75E-39 | InN1 | CHRM2 |
| RDX | 1.18E-43 | 0.31184836 | 0.455 | 0.4 | 4.50E-39 | InN1 | RDX |
| ABCC1 | 1.42E-43 | 0.34697608 | 0.403 | 0.352 | 5.45E-39 | InN1 | ABCC1 |
| NR3C11 | 2.78E-43 | 0.31817555 | 0.68 | 0.709 | 1.06E-38 | InN1 | NR3C1 |
| RAVER2 | 7.68E-43 | 0.27267086 | 0.349 | 0.287 | 2.94E-38 | InN1 | RAVER2 |
| LINC01138 | 1.10E-42 | 0.34830795 | 0.463 | 0.429 | 4.20E-38 | InN1 | LINC01138 |
| ANKRD36C | 1.48E-42 | 0.27835969 | 0.796 | 0.82 | 5.65E-38 | InN1 | ANKRD36C |
| MT-ATP8 | 2.29E-42 | 0.46417144 | 0.617 | 0.59 | 8.75E-38 | InN1 | MT-ATP8 |
| DNAH143 | 2.70E-42 | 0.37781106 | 0.358 | 0.293 | 1.03E-37 | InN1 | DNAH14 |
| AL024495.12 | 3.07E-42 | 0.53618175 | 0.347 | 0.301 | 1.17E-37 | InN1 | AL024495.1 |
| PPP2CB | 3.29E-42 | 0.29391665 | 0.419 | 0.374 | 1.26E-37 | InN1 | PPP2CB |
| PDE8A | 1.19E-41 | 0.30148251 | 0.484 | 0.436 | 4.54E-37 | InN1 | PDE8A |
| CASTOR3 | 1.31E-41 | 0.32148829 | 0.614 | 0.608 | 5.00E-37 | InN1 | CASTOR3 |
| OIP5-AS1 | 1.90E-41 | 0.32748064 | 0.572 | 0.572 | 7.24E-37 | InN1 | OIP5-AS1 |
| TSPAN7 | 6.03E-41 | 0.27302011 | 0.713 | 0.758 | 2.30E-36 | InN1 | TSPAN7 |
| FNBP1 | 2.05E-40 | 0.29367921 | 0.632 | 0.638 | 7.84E-36 | InN1 | FNBP1 |
| SLC8A3 | 2.29E-40 | 0.30424337 | 0.524 | 0.484 | 8.77E-36 | InN1 | SLC8A3 |
| CD811 | 2.34E-40 | 0.25944685 | 0.3 | 0.237 | 8.96E-36 | InN1 | CD81 |
| CNTN32 | 2.50E-40 | 0.34777376 | 0.686 | 0.663 | 9.55E-36 | InN1 | CNTN3 |
| BTBD3 | 3.21E-40 | 0.37582354 | 0.405 | 0.369 | 1.23E-35 | InN1 | BTBD3 |
| ERBIN1 | 5.61E-40 | 0.32920379 | 0.557 | 0.538 | 2.14E-35 | InN1 | ERBIN |
| PPP1R13B | 6.01E-40 | 0.32019127 | 0.628 | 0.647 | 2.30E-35 | InN1 | PPP1R13B |
| DCLK13 | 6.47E-40 | 0.27777812 | 0.921 | 0.944 | 2.47E-35 | InN1 | DCLK1 |

| | | | | | | | |
|---|---|---|---|---|---|---|---|
| LRRTM2 | 2.01E-39 | 0.26272236 | 0.353 | 0.295 | 7.70E-35 | InN1 | LRRTM2 |
| BCAT1 | 2.54E-39 | 0.268414 | 0.402 | 0.346 | 9.71E-35 | InN1 | BCAT1 |
| AMER2 | 3.34E-39 | 0.25532231 | 0.306 | 0.242 | 1.28E-34 | InN1 | AMER2 |
| ROCK21 | 3.49E-39 | 0.26694438 | 0.754 | 0.778 | 1.33E-34 | InN1 | ROCK2 |
| CXXC51 | 3.74E-39 | 0.28293489 | 0.403 | 0.351 | 1.43E-34 | InN1 | CXXC5 |
| ADCY5 | 5.43E-39 | 0.31738731 | 0.49 | 0.46 | 2.08E-34 | InN1 | ADCY5 |
| WDR6 | 8.44E-39 | 0.27681589 | 0.299 | 0.238 | 3.23E-34 | InN1 | WDR6 |
| PIK3C2A | 1.74E-38 | 0.31048283 | 0.494 | 0.468 | 6.65E-34 | InN1 | PIK3C2A |
| LMCD1-AS11 | 2.29E-38 | 0.40007622 | 0.344 | 0.286 | 8.74E-34 | InN1 | LMCD1-AS1 |
| DZIP1 | 2.46E-38 | 0.30218854 | 0.609 | 0.617 | 9.41E-34 | InN1 | DZIP1 |
| TSPYL2 | 4.57E-38 | 0.29388016 | 0.618 | 0.632 | 1.75E-33 | InN1 | TSPYL2 |
| CANX | 5.51E-38 | 0.29735979 | 0.506 | 0.492 | 2.11E-33 | InN1 | CANX |
| TBC1D8 | 9.16E-38 | 0.32790914 | 0.459 | 0.422 | 3.50E-33 | InN1 | TBC1D8 |
| TOP2B | 9.73E-38 | 0.32317829 | 0.568 | 0.576 | 3.72E-33 | InN1 | TOP2B |
| ITGAV1 | 1.87E-37 | 0.2725221 | 0.314 | 0.255 | 7.13E-33 | InN1 | ITGAV |
| BICC11 | 3.20E-37 | 0.6482854 | 0.449 | 0.431 | 1.22E-32 | InN1 | BICC1 |
| SEZ6L2 | 5.38E-37 | 0.30829833 | 0.485 | 0.464 | 2.05E-32 | InN1 | SEZ6L2 |
| TDRP | 5.55E-37 | 0.29928164 | 0.349 | 0.299 | 2.12E-32 | InN1 | TDRP |
| WWP2 | 6.08E-37 | 0.33017702 | 0.353 | 0.308 | 2.33E-32 | InN1 | WWP2 |
| RPS231 | 1.11E-36 | 0.33009911 | 0.349 | 0.291 | 4.26E-32 | InN1 | RPS23 |
| ALDH7A1 | 1.73E-36 | 0.25382581 | 0.272 | 0.214 | 6.61E-32 | InN1 | ALDH7A1 |
| SLC16A10 | 2.38E-36 | 0.31530823 | 0.423 | 0.376 | 9.09E-32 | InN1 | SLC16A10 |
| ATAD2B | 2.64E-36 | 0.31385492 | 0.464 | 0.437 | 1.01E-31 | InN1 | ATAD2B |
| AL356124.1 | 4.01E-36 | 0.51833003 | 0.289 | 0.239 | 1.53E-31 | InN1 | AL356124.1 |
| SLC5A3 | 6.07E-36 | 0.26919138 | 0.348 | 0.292 | 2.32E-31 | InN1 | SLC5A3 |
| RPS191 | 1.04E-35 | 0.28692544 | 0.341 | 0.287 | 3.98E-31 | InN1 | RPS19 |
| PACS2 | 1.36E-35 | 0.3190165 | 0.461 | 0.431 | 5.19E-31 | InN1 | PACS2 |
| TRIM24 | 2.43E-35 | 0.31676104 | 0.421 | 0.386 | 9.29E-31 | InN1 | TRIM24 |
| AC002460.22 | 4.35E-35 | 0.41018693 | 0.466 | 0.421 | 1.66E-30 | InN1 | AC002460.2 |
| WDR11-AS1 | 5.99E-35 | 0.28510413 | 0.341 | 0.289 | 2.29E-30 | InN1 | WDR11-AS1 |
| GIPC2 | 7.40E-35 | 0.29636646 | 0.305 | 0.252 | 2.83E-30 | InN1 | GIPC2 |
| RASA1 | 8.05E-35 | 0.2697472 | 0.682 | 0.715 | 3.08E-30 | InN1 | RASA1 |
| POR | 1.20E-34 | 0.26449642 | 0.301 | 0.243 | 4.57E-30 | InN1 | POR |
| MAP4 | 3.15E-34 | 0.32833162 | 0.561 | 0.561 | 1.20E-29 | InN1 | MAP4 |
| MBOAT21 | 3.79E-34 | 0.27848532 | 0.602 | 0.589 | 1.45E-29 | InN1 | MBOAT2 |
| SMYD21 | 4.26E-34 | 0.26761736 | 0.327 | 0.268 | 1.63E-29 | InN1 | SMYD2 |
| SREBF2 | 6.99E-34 | 0.31099836 | 0.578 | 0.584 | 2.67E-29 | InN1 | SREBF2 |
| ZNF83 | 1.40E-33 | 0.3140257 | 0.372 | 0.327 | 5.37E-29 | InN1 | ZNF83 |
| AC090115.1 | 2.97E-33 | 0.284191 | 0.273 | 0.22 | 1.14E-28 | InN1 | AC090115.1 |
| LPIN21 | 6.25E-33 | 0.27777736 | 0.405 | 0.361 | 2.39E-28 | InN1 | LPIN2 |
| UPF3A | 6.80E-33 | 0.3023905 | 0.55 | 0.547 | 2.60E-28 | InN1 | UPF3A |
| UPP1 | 6.97E-33 | 0.30204381 | 0.381 | 0.338 | 2.66E-28 | InN1 | UPP1 |
| MIRLET7BHG | 1.11E-32 | 0.28377118 | 0.458 | 0.423 | 4.26E-28 | InN1 | MIRLET7BHG |
| CHST1 | 1.37E-32 | 0.28142006 | 0.338 | 0.291 | 5.24E-28 | InN1 | CHST1 |
| C5orf56 | 4.24E-32 | 0.27586142 | 0.329 | 0.279 | 1.62E-27 | InN1 | C5orf56 |
| HSPA4L | 4.86E-32 | 0.28454778 | 0.498 | 0.478 | 1.86E-27 | InN1 | HSPA4L |
| CFAP161 | 4.92E-32 | 0.2506971 | 0.372 | 0.312 | 1.88E-27 | InN1 | CFAP161 |
| HSPA8 | 5.55E-32 | 0.30085776 | 0.341 | 0.287 | 2.12E-27 | InN1 | HSPA8 |
| RAMP1 | 2.29E-31 | 0.27250386 | 0.337 | 0.287 | 8.74E-27 | InN1 | RAMP1 |
| ASPH1 | 3.90E-31 | 0.27070909 | 0.596 | 0.602 | 1.49E-26 | InN1 | ASPH |
| AC245297.3 | 4.27E-31 | 0.26023702 | 0.31 | 0.261 | 1.63E-26 | InN1 | AC245297.3 |
| ARMH4 | 5.02E-31 | 0.31176398 | 0.446 | 0.421 | 1.92E-26 | InN1 | ARMH4 |
| PCBP4 | 5.99E-31 | 0.25238223 | 0.333 | 0.287 | 2.29E-26 | InN1 | PCBP4 |
| FAM214A | 1.89E-30 | 0.30269103 | 0.544 | 0.542 | 7.24E-26 | InN1 | FAM214A |
| RPS27A1 | 4.09E-30 | 0.32753577 | 0.329 | 0.277 | 1.56E-25 | InN1 | RPS27A |
| RPL14 | 6.63E-30 | 0.32291294 | 0.411 | 0.377 | 2.53E-25 | InN1 | RPL14 |
| CAPZB1 | 1.36E-29 | 0.28184494 | 0.615 | 0.64 | 5.19E-25 | InN1 | CAPZB |
| SMCHD1 | 9.75E-29 | 0.29742706 | 0.632 | 0.659 | 3.73E-24 | InN1 | SMCHD1 |
| SNRNP70 | 1.01E-28 | 0.2795209 | 0.695 | 0.726 | 3.86E-24 | InN1 | SNRNP70 |
| HDAC2-AS2 | 1.24E-28 | 0.29304336 | 0.312 | 0.266 | 4.73E-24 | InN1 | HDAC2-AS2 |
| ANKRD26 | 2.85E-28 | 0.27926847 | 0.721 | 0.758 | 1.09E-23 | InN1 | ANKRD26 |
| RPL13A1 | 3.30E-28 | 0.26089399 | 0.34 | 0.294 | 1.26E-23 | InN1 | RPL13A |
| AC007368.12 | 4.06E-28 | 0.30852615 | 0.491 | 0.438 | 1.55E-23 | InN1 | AC007368.1 |

| | | | | | | | |
|---|---|---|---|---|---|---|---|
| DIXDC1 | 1.43E-27 | 0.26165143 | 0.493 | 0.463 | 5.48E-23 | InN1 | DIXDC1 |
| SFMBT1 | 2.05E-27 | 0.26291887 | 0.368 | 0.327 | 7.83E-23 | InN1 | SFMBT1 |
| NPY1R1 | 4.30E-27 | 0.28347216 | 0.391 | 0.356 | 1.64E-22 | InN1 | NPY1R |
| LRP11 | 5.29E-27 | 0.29936053 | 0.488 | 0.487 | 2.02E-22 | InN1 | LRP1 |
| HNRNPA2B1 | 6.29E-27 | 0.2605596 | 0.657 | 0.688 | 2.40E-22 | InN1 | HNRNPA2B1 |
| PRRC2B1 | 1.98E-26 | 0.25193226 | 0.637 | 0.67 | 7.57E-22 | InN1 | PRRC2B |
| INTU | 3.26E-26 | 0.30649274 | 0.477 | 0.471 | 1.25E-21 | InN1 | INTU |
| DHX36 | 5.37E-26 | 0.32295397 | 0.507 | 0.513 | 2.05E-21 | InN1 | DHX36 |
| NAP1L4 | 7.01E-26 | 0.26411874 | 0.486 | 0.475 | 2.68E-21 | InN1 | NAP1L4 |
| EPN2 | 1.21E-25 | 0.27782944 | 0.552 | 0.557 | 4.62E-21 | InN1 | EPN2 |
| RPL101 | 5.32E-25 | 0.31950605 | 0.373 | 0.342 | 2.03E-20 | InN1 | RPL10 |
| SV2A | 6.54E-25 | 0.26227124 | 0.568 | 0.596 | 2.50E-20 | InN1 | SV2A |
| GAPDH | 6.85E-25 | 0.28671914 | 0.531 | 0.529 | 2.62E-20 | InN1 | GAPDH |
| RPS151 | 1.10E-24 | 0.25154897 | 0.328 | 0.285 | 4.21E-20 | InN1 | RPS15 |
| TSTD3 | 1.57E-24 | 0.26808414 | 0.315 | 0.278 | 6.00E-20 | InN1 | TSTD3 |
| SH3KBP11 | 2.88E-24 | 0.27789792 | 0.595 | 0.609 | 1.10E-19 | InN1 | SH3KBP1 |
| ZNF680 | 6.48E-24 | 0.27048409 | 0.366 | 0.332 | 2.48E-19 | InN1 | ZNF680 |
| CCDC180 | 7.71E-24 | 0.29406385 | 0.35 | 0.317 | 2.95E-19 | InN1 | CCDC180 |
| SEMA6D1 | 1.84E-23 | 0.61688198 | 0.55 | 0.591 | 7.03E-19 | InN1 | SEMA6D |
| TP53BP1 | 4.44E-23 | 0.28940532 | 0.555 | 0.57 | 1.70E-18 | InN1 | TP53BP1 |
| NETO2 | 6.62E-23 | 0.37091567 | 0.427 | 0.411 | 2.53E-18 | InN1 | NETO2 |
| WDR19 | 1.83E-22 | 0.30932242 | 0.522 | 0.532 | 6.99E-18 | InN1 | WDR19 |
| PLCG21 | 2.05E-22 | 0.55494669 | 0.654 | 0.652 | 7.84E-18 | InN1 | PLCG2 |
| SLC24A41 | 2.19E-22 | 0.30213361 | 0.39 | 0.349 | 8.36E-18 | InN1 | SLC24A4 |
| PSAP1 | 2.55E-22 | 0.25408561 | 0.607 | 0.646 | 9.76E-18 | InN1 | PSAP |
| TNK2 | 4.51E-22 | 0.27773422 | 0.487 | 0.49 | 1.72E-17 | InN1 | TNK2 |
| EVA1C | 5.16E-22 | 0.25616483 | 0.384 | 0.354 | 1.97E-17 | InN1 | EVA1C |
| RPS181 | 6.08E-22 | 0.27856349 | 0.323 | 0.285 | 2.33E-17 | InN1 | RPS18 |
| RNF24 | 9.55E-22 | 0.28166293 | 0.572 | 0.597 | 3.65E-17 | InN1 | RNF24 |
| GLB1 | 3.07E-21 | 0.29744951 | 0.406 | 0.392 | 1.17E-16 | InN1 | GLB1 |
| UBXN2A | 3.91E-21 | 0.27754586 | 0.351 | 0.32 | 1.49E-16 | InN1 | UBXN2A |
| MICU1 | 7.04E-21 | 0.27395876 | 0.69 | 0.731 | 2.69E-16 | InN1 | MICU1 |
| ATF7IP1 | 7.15E-21 | 0.25167671 | 0.496 | 0.493 | 2.73E-16 | InN1 | ATF7IP |
| HIBCH | 9.60E-21 | 0.28556984 | 0.599 | 0.622 | 3.67E-16 | InN1 | HIBCH |
| EFCAB2 | 9.79E-21 | 0.27464376 | 0.521 | 0.518 | 3.74E-16 | InN1 | EFCAB2 |
| CDK8 | 1.12E-20 | 0.31070628 | 0.546 | 0.567 | 4.27E-16 | InN1 | CDK8 |
| CTBP22 | 3.05E-20 | 0.27613681 | 0.382 | 0.352 | 1.17E-15 | InN1 | CTBP2 |
| MRPL33 | 8.93E-19 | 0.28649125 | 0.405 | 0.392 | 3.41E-14 | InN1 | MRPL33 |
| GP1BB | 1.03E-18 | 0.26323176 | 0.289 | 0.258 | 3.95E-14 | InN1 | GP1BB |
| RPL31 | 1.52E-18 | 0.26533199 | 0.339 | 0.308 | 5.83E-14 | InN1 | RPL3 |
| ATP6V0C | 5.41E-18 | 0.29996547 | 0.452 | 0.46 | 2.07E-13 | InN1 | ATP6V0C |
| LIPA | 1.40E-17 | 0.28912074 | 0.479 | 0.485 | 5.36E-13 | InN1 | LIPA |
| SLC4A8 | 1.79E-17 | 0.26103983 | 0.593 | 0.628 | 6.83E-13 | InN1 | SLC4A8 |
| PPP1CB | 4.57E-17 | 0.25636197 | 0.484 | 0.493 | 1.75E-12 | InN1 | PPP1CB |
| EDIL32 | 4.67E-17 | 0.26832312 | 0.692 | 0.739 | 1.78E-12 | InN1 | EDIL3 |
| TRPC4 | 8.28E-17 | 0.2660269 | 0.31 | 0.275 | 3.16E-12 | InN1 | TRPC4 |
| BCR | 1.49E-16 | 0.25915139 | 0.423 | 0.416 | 5.68E-12 | InN1 | BCR |
| CDRT4 | 1.91E-16 | 0.25212596 | 0.409 | 0.398 | 7.29E-12 | InN1 | CDRT4 |
| EML51 | 2.63E-15 | 0.35876963 | 0.528 | 0.57 | 1.01E-10 | InN1 | EML5 |
| CRIM12 | 7.35E-15 | 0.26413677 | 0.485 | 0.493 | 2.81E-10 | InN1 | CRIM1 |
| RPL6 | 2.31E-14 | 0.25270534 | 0.342 | 0.32 | 8.82E-10 | InN1 | RPL6 |
| ANKS6 | 2.56E-14 | 0.25817395 | 0.406 | 0.396 | 9.78E-10 | InN1 | ANKS6 |
| MIR3936HG | 2.83E-14 | 0.26926321 | 0.476 | 0.492 | 1.08E-09 | InN1 | MIR3936HG |
| RPL32 | 5.42E-14 | 0.2904657 | 0.384 | 0.367 | 2.07E-09 | InN1 | RPL32 |
| DPH6 | 9.24E-14 | 0.25428419 | 0.423 | 0.417 | 3.53E-09 | InN1 | DPH6 |
| RPS81 | 2.30E-12 | 0.25661574 | 0.35 | 0.336 | 8.77E-08 | InN1 | RPS8 |
| TCF25 | 2.52E-12 | 0.26479296 | 0.587 | 0.627 | 9.65E-08 | InN1 | TCF25 |
| HYDIN1 | 4.36E-12 | 0.2685674 | 0.423 | 0.424 | 1.67E-07 | InN1 | HYDIN |
| SCG3 | 1.14E-11 | 0.25695083 | 0.428 | 0.434 | 4.37E-07 | InN1 | SCG3 |
| RPL34 | 1.71E-11 | 0.29331425 | 0.389 | 0.384 | 6.52E-07 | InN1 | RPL34 |
| RYR11 | 1.36E-09 | 0.25575153 | 0.253 | 0.236 | 5.18E-05 | InN1 | RYR1 |
| AL627171.2 | 2.20E-09 | 0.52251749 | 0.443 | 0.456 | 8.42E-05 | InN1 | AL627171.2 |
| MB21D21 | 4.28E-08 | 0.25187703 | 0.393 | 0.399 | 0.00163488 | InN1 | MB21D2 |

| | | | | | | | |
|---|---|---|---|---|---|---|---|
| HTR1E1 | 1.46E-07 | 0.34813482 | 0.541 | 0.586 | 0.00559227 | InN1 | HTR1E |
| ZMAT4 | 7.86E-06 | 0.32052001 | 0.617 | 0.791 | 0.30061021 | InN1 | ZMAT4 |
| RPS241 | 3.33E-05 | 0.25733445 | 0.415 | 0.431 | 1 | InN1 | RPS24 |
| ANKRD30B | 0.00039265 | 0.2604703 | 0.275 | 0.282 | 1 | InN1 | ANKRD30B |
| GRIK1 | 0 | 3.31187761 | 0.984 | 0.352 | 0 | InN2 | GRIK1 |
| SOX6 | 0 | 2.66947477 | 0.859 | 0.177 | 0 | InN2 | SOX6 |
| NXPH1 | 0 | 2.40130764 | 0.976 | 0.221 | 0 | InN2 | NXPH1 |
| SPOCK3 | 0 | 2.33475231 | 0.977 | 0.527 | 0 | InN2 | SPOCK3 |
| KIF26B | 0 | 2.32258867 | 0.887 | 0.401 | 0 | InN2 | KIF26B |
| LHFPL31 | 0 | 2.08622964 | 0.774 | 0.241 | 0 | InN2 | LHFPL3 |
| GRIK31 | 0 | 2.0365511 | 0.864 | 0.262 | 0 | InN2 | GRIK3 |
| KIAA12171 | 0 | 2.02815139 | 0.984 | 0.636 | 0 | InN2 | KIAA1217 |
| PCDH151 | 0 | 2.01994478 | 0.698 | 0.302 | 0 | InN2 | PCDH15 |
| PLCH11 | 0 | 1.90257527 | 0.776 | 0.172 | 0 | InN2 | PLCH1 |
| ASTN21 | 0 | 1.83713483 | 0.955 | 0.723 | 0 | InN2 | ASTN2 |
| SYNPR1 | 0 | 1.8154131 | 0.959 | 0.51 | 0 | InN2 | SYNPR |
| XKR4 | 0 | 1.80797871 | 0.981 | 0.853 | 0 | InN2 | XKR4 |
| ROBO22 | 0 | 1.80644449 | 0.992 | 0.843 | 0 | InN2 | ROBO2 |
| GRID21 | 0 | 1.80606351 | 0.945 | 0.721 | 0 | InN2 | GRID2 |
| STXBP61 | 0 | 1.79750392 | 0.845 | 0.356 | 0 | InN2 | STXBP6 |
| TRHDE3 | 0 | 1.7887791 | 0.745 | 0.506 | 0 | InN2 | TRHDE |
| ROBO11 | 0 | 1.72238862 | 0.96 | 0.641 | 0 | InN2 | ROBO1 |
| GPC61 | 0 | 1.7156307 | 0.816 | 0.47 | 0 | InN2 | GPC6 |
| WLS | 0 | 1.62214652 | 0.748 | 0.245 | 0 | InN2 | WLS |
| COL25A1 | 0 | 1.62073355 | 0.573 | 0.192 | 0 | InN2 | COL25A1 |
| NPAS31 | 0 | 1.60383096 | 0.938 | 0.811 | 0 | InN2 | NPAS3 |
| SHISA61 | 0 | 1.57345771 | 0.748 | 0.318 | 0 | InN2 | SHISA6 |
| GRIN3A | 0 | 1.55560435 | 0.669 | 0.209 | 0 | InN2 | GRIN3A |
| TENM1 | 0 | 1.55138069 | 0.748 | 0.357 | 0 | InN2 | TENM1 |
| ADCY8 | 0 | 1.47433089 | 0.708 | 0.26 | 0 | InN2 | ADCY8 |
| GRIK2 | 0 | 1.46541526 | 0.991 | 0.946 | 0 | InN2 | GRIK2 |
| PCDH11X1 | 0 | 1.45972176 | 0.777 | 0.522 | 0 | InN2 | PCDH11X |
| TENM31 | 0 | 1.41750576 | 0.879 | 0.66 | 0 | InN2 | TENM3 |
| PTPRM | 0 | 1.41321489 | 0.874 | 0.565 | 0 | InN2 | PTPRM |
| RBMS3 | 0 | 1.38389116 | 0.916 | 0.622 | 0 | InN2 | RBMS3 |
| PAWR | 0 | 1.38093015 | 0.528 | 0.058 | 0 | InN2 | PAWR |
| MAF2 | 0 | 1.36809462 | 0.59 | 0.159 | 0 | InN2 | MAF |
| KCNMB22 | 0 | 1.32655157 | 0.781 | 0.563 | 0 | InN2 | KCNMB2 |
| PCSK52 | 0 | 1.30593123 | 0.488 | 0.182 | 0 | InN2 | PCSK5 |
| SLC24A31 | 0 | 1.29868663 | 0.719 | 0.285 | 0 | InN2 | SLC24A3 |
| CDH13 | 0 | 1.28866042 | 0.794 | 0.636 | 0 | InN2 | CDH13 |
| FBN2 | 0 | 1.28391544 | 0.291 | 0.035 | 0 | InN2 | FBN2 |
| RUNX1T12 | 0 | 1.27163193 | 0.887 | 0.737 | 0 | InN2 | RUNX1T1 |
| SATB1-AS12 | 0 | 1.24960829 | 0.713 | 0.42 | 0 | InN2 | SATB1-AS1 |
| KCNIP11 | 0 | 1.23574965 | 0.756 | 0.276 | 0 | InN2 | KCNIP1 |
| GAD11 | 0 | 1.2160219 | 0.745 | 0.258 | 0 | InN2 | GAD1 |
| MGAT4C | 0 | 1.20248928 | 0.821 | 0.564 | 0 | InN2 | MGAT4C |
| TMTC2 | 0 | 1.17919154 | 0.778 | 0.525 | 0 | InN2 | TMTC2 |
| NETO12 | 0 | 1.17841339 | 0.865 | 0.723 | 0 | InN2 | NETO1 |
| ELAVL2 | 0 | 1.16415694 | 0.77 | 0.506 | 0 | InN2 | ELAVL2 |
| LINC013222 | 0 | 1.14776373 | 0.779 | 0.529 | 0 | InN2 | LINC01322 |
| CDH91 | 0 | 1.14416218 | 0.835 | 0.607 | 0 | InN2 | CDH9 |
| GALNT143 | 0 | 1.14201153 | 0.591 | 0.315 | 0 | InN2 | GALNT14 |
| LINC00535 | 0 | 1.13756896 | 0.529 | 0.285 | 0 | InN2 | LINC00535 |
| NRXN31 | 0 | 1.12065958 | 0.999 | 0.99 | 0 | InN2 | NRXN3 |
| SLC27A6 | 0 | 1.11840615 | 0.301 | 0.054 | 0 | InN2 | SLC27A6 |
| TIAM11 | 0 | 1.10895509 | 0.893 | 0.746 | 0 | InN2 | TIAM1 |
| SEMA6A | 0 | 1.08639642 | 0.525 | 0.198 | 0 | InN2 | SEMA6A |
| RAB3C | 0 | 1.07787431 | 0.841 | 0.626 | 0 | InN2 | RAB3C |
| FLT3 | 0 | 1.07562244 | 0.443 | 0.051 | 0 | InN2 | FLT3 |
| TSHZ31 | 0 | 1.07461974 | 0.704 | 0.43 | 0 | InN2 | TSHZ3 |
| SPARCL12 | 0 | 1.07438925 | 0.812 | 0.657 | 0 | InN2 | SPARCL1 |
| ZNF385D2 | 0 | 1.06895464 | 0.773 | 0.499 | 0 | InN2 | ZNF385D |

| | | | | | | | |
|---|---|---|---|---|---|---|---|
| GRIP1 | 0 | 1.06061337 | 0.953 | 0.79 | 0 | InN2 | GRIP1 |
| SST | 0 | 1.03754166 | 0.282 | 0.015 | 0 | InN2 | SST |
| CRB11 | 0 | 1.01811997 | 0.595 | 0.296 | 0 | InN2 | CRB1 |
| RPS6KA21 | 0 | 1.01488175 | 0.941 | 0.852 | 0 | InN2 | RPS6KA2 |
| DAPK11 | 0 | 1.00749604 | 0.868 | 0.748 | 0 | InN2 | DAPK1 |
| MITF1 | 0 | 1.00488703 | 0.443 | 0.182 | 0 | InN2 | MITF |
| LHX6 | 0 | 0.99123486 | 0.447 | 0.049 | 0 | InN2 | LHX6 |
| FRMD32 | 0 | 0.98097963 | 0.598 | 0.327 | 0 | InN2 | FRMD3 |
| AC027288.3 | 0 | 0.97174402 | 0.488 | 0.234 | 0 | InN2 | AC027288.3 |
| PAM1 | 0 | 0.95775791 | 0.838 | 0.754 | 0 | InN2 | PAM |
| GRIA32 | 0 | 0.93186383 | 0.921 | 0.803 | 0 | InN2 | GRIA3 |
| TANC12 | 0 | 0.92889898 | 0.388 | 0.099 | 0 | InN2 | TANC1 |
| KIAA12111 | 0 | 0.92289515 | 0.732 | 0.379 | 0 | InN2 | KIAA1211 |
| QKI2 | 0 | 0.88215805 | 0.864 | 0.661 | 0 | InN2 | QKI |
| SNRPN1 | 0 | 0.8613379 | 0.883 | 0.754 | 0 | InN2 | SNRPN |
| ANK11 | 0 | 0.84575519 | 0.644 | 0.298 | 0 | InN2 | ANK1 |
| CNTNAP21 | 0 | 0.83704884 | 0.997 | 0.993 | 0 | InN2 | CNTNAP2 |
| VAV21 | 0 | 0.819133 | 0.472 | 0.166 | 0 | InN2 | VAV2 |
| ARX | 0 | 0.80824478 | 0.397 | 0.087 | 0 | InN2 | ARX |
| ELFN11 | 0 | 0.78541238 | 0.37 | 0.066 | 0 | InN2 | ELFN1 |
| SLC8A12 | 0 | 0.77726928 | 0.964 | 0.856 | 0 | InN2 | SLC8A1 |
| DSCAML11 | 0 | 0.76835942 | 0.89 | 0.751 | 0 | InN2 | DSCAML1 |
| TNIK2 | 0 | 0.76151842 | 0.878 | 0.8 | 0 | InN2 | TNIK |
| RAB3B1 | 0 | 0.76140402 | 0.382 | 0.119 | 0 | InN2 | RAB3B |
| DOCK112 | 0 | 0.75731409 | 0.474 | 0.196 | 0 | InN2 | DOCK11 |
| RBFOX11 | 0 | 0.74727622 | 0.999 | 0.983 | 0 | InN2 | RBFOX1 |
| AUTS21 | 0 | 0.74717225 | 0.981 | 0.973 | 0 | InN2 | AUTS2 |
| SLC24A24 | 0 | 0.74436601 | 0.924 | 0.829 | 0 | InN2 | SLC24A2 |
| AC076968.2 | 0 | 0.74427828 | 0.289 | 0.053 | 0 | InN2 | AC076968.2 |
| LIMCH1 | 0 | 0.74222358 | 0.941 | 0.917 | 0 | InN2 | LIMCH1 |
| OXR13 | 0 | 0.73334186 | 0.954 | 0.92 | 0 | InN2 | OXR1 |
| SLC9A2 | 0 | 0.72895843 | 0.278 | 0.055 | 0 | InN2 | SLC9A2 |
| CADPS1 | 0 | 0.72303922 | 0.968 | 0.937 | 0 | InN2 | CADPS |
| AAK1 | 0 | 0.72051005 | 0.896 | 0.878 | 0 | InN2 | AAK1 |
| DNM3 | 0 | 0.71846615 | 0.921 | 0.89 | 0 | InN2 | DNM3 |
| GAD2 | 0 | 0.66631719 | 0.601 | 0.238 | 0 | InN2 | GAD2 |
| KLF5 | 0 | 0.65232972 | 0.266 | 0.034 | 0 | InN2 | KLF5 |
| PRDM1 | 0 | 0.64972873 | 0.281 | 0.049 | 0 | InN2 | PRDM1 |
| SLC6A12 | 0 | 0.63828959 | 0.753 | 0.329 | 0 | InN2 | SLC6A1 |
| MAFB | 0 | 0.61597783 | 0.28 | 0.066 | 0 | InN2 | MAFB |
| MCUB | 0 | 0.6124793 | 0.254 | 0.066 | 0 | InN2 | MCUB |
| TRIM67 | 0 | 0.59592121 | 0.262 | 0.045 | 0 | InN2 | TRIM67 |
| DLGAP1 | 0 | 0.58844724 | 0.996 | 0.989 | 0 | InN2 | DLGAP1 |
| NTRK2 | 0 | 0.58009505 | 0.947 | 0.944 | 0 | InN2 | NTRK2 |
| RELN | 0 | 0.4252783 | 0.387 | 0.121 | 0 | InN2 | RELN |
| TENM21 | 2.69E-307 | 0.69135473 | 0.975 | 0.931 | 1.03E-302 | InN2 | TENM2 |
| NOS1 | 2.08E-305 | 0.87617196 | 0.317 | 0.104 | 7.94E-301 | InN2 | NOS1 |
| KLHL53 | 7.06E-299 | 0.8157288 | 0.567 | 0.303 | 2.70E-294 | InN2 | KLHL5 |
| CACNA2D34 | 5.93E-295 | 0.76571941 | 0.91 | 0.866 | 2.27E-290 | InN2 | CACNA2D3 |
| RASGRF21 | 5.70E-293 | 0.84017498 | 0.81 | 0.595 | 2.18E-288 | InN2 | RASGRF2 |
| TOX31 | 1.11E-289 | 0.6651837 | 0.438 | 0.174 | 4.26E-285 | InN2 | TOX3 |
| UBASH3B1 | 1.25E-287 | 0.784978 | 0.6 | 0.325 | 4.79E-283 | InN2 | UBASH3B |
| MARCH111 | 1.99E-282 | 0.95207068 | 0.556 | 0.328 | 7.61E-278 | InN2 | 11-Mar |
| NAV2 | 1.29E-281 | 0.65616375 | 0.926 | 0.921 | 4.92E-277 | InN2 | NAV2 |
| AC092969.11 | 1.04E-277 | 0.88583624 | 0.373 | 0.15 | 3.97E-273 | InN2 | AC092969.1 |
| AL445623.2 | 3.35E-277 | 0.91246156 | 0.593 | 0.364 | 1.28E-272 | InN2 | AL445623.2 |
| L3MBTL41 | 4.10E-275 | 0.93777208 | 0.642 | 0.386 | 1.57E-270 | InN2 | L3MBTL4 |
| PCLO3 | 1.84E-273 | 0.58893799 | 0.943 | 0.922 | 7.02E-269 | InN2 | PCLO |
| COL19A12 | 6.35E-269 | 1.19198048 | 0.529 | 0.317 | 2.43E-264 | InN2 | COL19A1 |
| MYO5B | 3.83E-263 | 0.59162653 | 0.256 | 0.073 | 1.46E-258 | InN2 | MYO5B |
| ANKRD441 | 1.93E-262 | 0.88879139 | 0.624 | 0.416 | 7.38E-258 | InN2 | ANKRD44 |
| NXPH2 | 2.91E-262 | 0.79195469 | 0.302 | 0.101 | 1.11E-257 | InN2 | NXPH2 |
| FRMD4B1 | 7.26E-262 | 0.72223905 | 0.389 | 0.156 | 2.77E-257 | InN2 | FRMD4B |

| | | | | | | | |
|---|---|---|---|---|---|---|---|
| SILC11 | 2.29E-258 | 0.67644883 | 0.456 | 0.205 | 8.77E-254 | InN2 | SILC1 |
| GPRIN32 | 5.83E-256 | 0.64046206 | 0.399 | 0.165 | 2.23E-251 | InN2 | GPRIN3 |
| CDH83 | 1.38E-255 | 0.8474523 | 0.786 | 0.654 | 5.27E-251 | InN2 | CDH8 |
| ERC1 | 5.41E-248 | 0.60647656 | 0.892 | 0.866 | 2.07E-243 | InN2 | ERC1 |
| FAM110B | 7.22E-248 | 0.91090855 | 0.622 | 0.421 | 2.76E-243 | InN2 | FAM110B |
| PTCHD4 | 7.09E-246 | 0.73982226 | 0.642 | 0.386 | 2.71E-241 | InN2 | PTCHD4 |
| ENOX21 | 8.51E-244 | 0.88664824 | 0.372 | 0.168 | 3.25E-239 | InN2 | ENOX2 |
| GRIP21 | 3.29E-242 | 0.54440722 | 0.419 | 0.176 | 1.26E-237 | InN2 | GRIP2 |
| NCOA2 | 2.67E-240 | 0.73091846 | 0.798 | 0.74 | 1.02E-235 | InN2 | NCOA2 |
| TMTC11 | 1.97E-239 | 0.88708368 | 0.702 | 0.555 | 7.54E-235 | InN2 | TMTC1 |
| MID11 | 4.32E-237 | 0.62514728 | 0.33 | 0.128 | 1.65E-232 | InN2 | MID1 |
| MIR181A1HG | 6.17E-236 | 0.83378246 | 0.636 | 0.434 | 2.36E-231 | InN2 | MIR181A1HG |
| SMYD3 | 4.42E-234 | 0.61778197 | 0.935 | 0.932 | 1.69E-229 | InN2 | SMYD3 |
| AL592156.2 | 1.36E-230 | 0.90913215 | 0.297 | 0.113 | 5.21E-226 | InN2 | AL592156.2 |
| AL365361.1 | 8.31E-230 | 0.52998307 | 0.299 | 0.114 | 3.18E-225 | InN2 | AL365361.1 |
| GARNL31 | 6.50E-227 | 0.66491554 | 0.808 | 0.72 | 2.48E-222 | InN2 | GARNL3 |
| CACNA2D2 | 5.26E-225 | 0.58497059 | 0.378 | 0.168 | 2.01E-220 | InN2 | CACNA2D2 |
| ZNF6081 | 4.17E-224 | 0.71604221 | 0.57 | 0.345 | 1.60E-219 | InN2 | ZNF608 |
| PACRG1 | 3.85E-222 | 0.82939531 | 0.783 | 0.704 | 1.47E-217 | InN2 | PACRG |
| DLX11 | 9.00E-222 | 0.44215964 | 0.267 | 0.089 | 3.44E-217 | InN2 | DLX1 |
| BCL11A2 | 1.07E-220 | 0.88800782 | 0.719 | 0.593 | 4.10E-216 | InN2 | BCL11A |
| ADAMTSL3 | 3.62E-217 | 0.94637431 | 0.486 | 0.28 | 1.38E-212 | InN2 | ADAMTSL3 |
| TOX21 | 1.69E-215 | 0.74060132 | 0.406 | 0.194 | 6.46E-211 | InN2 | TOX2 |
| NBEA | 4.86E-215 | 0.44302812 | 0.965 | 0.96 | 1.86E-210 | InN2 | NBEA |
| RAB311 | 1.99E-212 | 0.67829061 | 0.305 | 0.122 | 7.62E-208 | InN2 | RAB31 |
| CTNND2 | 5.04E-210 | 0.51571846 | 0.98 | 0.97 | 1.93E-205 | InN2 | CTNND2 |
| TCF41 | 7.90E-209 | 0.44214888 | 0.956 | 0.958 | 3.02E-204 | InN2 | TCF4 |
| GRIA1 | 2.63E-208 | 0.54230438 | 0.933 | 0.922 | 1.00E-203 | InN2 | GRIA1 |
| RIPOR21 | 2.81E-208 | 0.75582748 | 0.635 | 0.458 | 1.07E-203 | InN2 | RIPOR2 |
| PTPRN21 | 1.62E-207 | 0.48433826 | 0.951 | 0.95 | 6.20E-203 | InN2 | PTPRN2 |
| GNG4 | 4.47E-203 | 0.54376261 | 0.329 | 0.135 | 1.71E-198 | InN2 | GNG4 |
| DKK21 | 3.76E-200 | 0.63652718 | 0.272 | 0.103 | 1.44E-195 | InN2 | DKK2 |
| ZNF462 | 2.70E-199 | 0.65997002 | 0.485 | 0.276 | 1.03E-194 | InN2 | ZNF462 |
| SFMBT21 | 7.65E-199 | 0.87636334 | 0.562 | 0.374 | 2.92E-194 | InN2 | SFMBT2 |
| NTN41 | 2.06E-195 | 0.62346592 | 0.425 | 0.218 | 7.88E-191 | InN2 | NTN4 |
| TRPC61 | 3.69E-193 | 0.67747981 | 0.304 | 0.124 | 1.41E-188 | InN2 | TRPC6 |
| SHTN12 | 5.59E-190 | 0.69800546 | 0.755 | 0.671 | 2.14E-185 | InN2 | SHTN1 |
| NPAS1 | 1.20E-186 | 0.58297641 | 0.316 | 0.137 | 4.60E-182 | InN2 | NPAS1 |
| UST2 | 3.52E-186 | 0.60582204 | 0.314 | 0.135 | 1.35E-181 | InN2 | UST |
| PHACTR21 | 3.89E-185 | 0.97954669 | 0.559 | 0.401 | 1.49E-180 | InN2 | PHACTR2 |
| AC025809.11 | 6.91E-178 | 0.75527961 | 0.492 | 0.31 | 2.64E-173 | InN2 | AC025809.1 |
| SATB11 | 4.64E-177 | 0.80501243 | 0.495 | 0.331 | 1.77E-172 | InN2 | SATB1 |
| KAZN1 | 3.79E-176 | 0.40874146 | 0.986 | 0.951 | 1.45E-171 | InN2 | KAZN |
| ADCYAP1R11 | 5.17E-172 | 0.57915209 | 0.493 | 0.291 | 1.98E-167 | InN2 | ADCYAP1R1 |
| BCL11B1 | 1.08E-167 | 0.52608789 | 0.462 | 0.245 | 4.15E-163 | InN2 | BCL11B |
| CLMP | 8.62E-160 | 0.63724956 | 0.3 | 0.135 | 3.30E-155 | InN2 | CLMP |
| TMEM47 | 9.52E-158 | 0.43107981 | 0.256 | 0.106 | 3.64E-153 | InN2 | TMEM47 |
| KIZ | 6.68E-157 | 0.70592635 | 0.637 | 0.545 | 2.55E-152 | InN2 | KIZ |
| DAB12 | 8.08E-156 | 0.61083282 | 0.905 | 0.878 | 3.09E-151 | InN2 | DAB1 |
| PIP5K1B | 5.31E-153 | 0.71971171 | 0.614 | 0.473 | 2.03E-148 | InN2 | PIP5K1B |
| FAM135B1 | 4.44E-152 | 0.69629283 | 0.73 | 0.627 | 1.70E-147 | InN2 | FAM135B |
| PLPPR11 | 1.15E-151 | 0.71788333 | 0.474 | 0.286 | 4.39E-147 | InN2 | PLPPR1 |
| PDE4B2 | 6.79E-150 | 1.00194804 | 0.698 | 0.609 | 2.60E-145 | InN2 | PDE4B |
| AC006115.2 | 4.42E-149 | 0.79696595 | 0.52 | 0.379 | 1.69E-144 | InN2 | AC006115.2 |
| FRMD4A1 | 3.32E-148 | 0.31353152 | 0.991 | 0.988 | 1.27E-143 | InN2 | FRMD4A |
| MYT1L1 | 4.66E-148 | 0.35264305 | 0.97 | 0.971 | 1.78E-143 | InN2 | MYT1L |
| SETBP11 | 1.75E-147 | 0.46510287 | 0.893 | 0.889 | 6.70E-143 | InN2 | SETBP1 |
| AMOTL1 | 4.69E-147 | 0.49896264 | 0.374 | 0.195 | 1.79E-142 | InN2 | AMOTL1 |
| PITPNC11 | 7.66E-147 | 0.57770896 | 0.658 | 0.485 | 2.93E-142 | InN2 | PITPNC1 |
| ZNF7041 | 1.29E-146 | 0.65049212 | 0.733 | 0.669 | 4.95E-142 | InN2 | ZNF704 |
| RIN22 | 2.36E-146 | 0.60299556 | 0.382 | 0.201 | 9.03E-142 | InN2 | RIN2 |
| DTNA1 | 5.07E-142 | 0.45960143 | 0.884 | 0.847 | 1.94E-137 | InN2 | DTNA |
| PPP2R2B2 | 5.86E-140 | 0.40984695 | 0.958 | 0.956 | 2.24E-135 | InN2 | PPP2R2B |

| | | | | | | | |
|---|---|---|---|---|---|---|---|
| B3GAT21 | 4.07E-138 | 0.60497575 | 0.262 | 0.119 | 1.55E-133 | InN2 | B3GAT2 |
| SNHG14 | 4.85E-138 | 0.25089797 | 0.943 | 0.948 | 1.86E-133 | InN2 | SNHG14 |
| NETO21 | 1.61E-136 | 0.70491754 | 0.54 | 0.404 | 6.15E-132 | InN2 | NETO2 |
| ANKRD36 | 4.09E-136 | 0.52754189 | 0.831 | 0.842 | 1.56E-131 | InN2 | ANKRD36 |
| AC073332.1 | 5.31E-133 | 0.97289144 | 0.468 | 0.32 | 2.03E-128 | InN2 | AC073332.1 |
| PALM2-AKAF | 6.35E-133 | 0.6829936 | 0.733 | 0.702 | 2.43E-128 | InN2 | PALM2-AKAP |
| CACNG41 | 3.02E-129 | 0.47029619 | 0.309 | 0.151 | 1.15E-124 | InN2 | CACNG4 |
| IGF11 | 6.55E-129 | 0.41757998 | 0.305 | 0.143 | 2.50E-124 | InN2 | IGF1 |
| TIMP21 | 7.19E-129 | 0.56056138 | 0.462 | 0.295 | 2.75E-124 | InN2 | TIMP2 |
| EPB41L4A2 | 1.41E-127 | 0.89303627 | 0.297 | 0.156 | 5.39E-123 | InN2 | EPB41L4A |
| RNASET21 | 3.25E-126 | 0.58438984 | 0.409 | 0.258 | 1.24E-121 | InN2 | RNASET2 |
| IQSEC12 | 7.91E-126 | 0.55194576 | 0.798 | 0.781 | 3.02E-121 | InN2 | IQSEC1 |
| TMEFF23 | 2.37E-124 | 0.61668884 | 0.814 | 0.762 | 9.05E-120 | InN2 | TMEFF2 |
| BACH12 | 1.91E-123 | 0.46693325 | 0.394 | 0.225 | 7.29E-119 | InN2 | BACH1 |
| RIMS21 | 5.93E-123 | 0.34650121 | 0.97 | 0.969 | 2.27E-118 | InN2 | RIMS2 |
| CDH71 | 1.83E-121 | 0.71398396 | 0.418 | 0.267 | 6.99E-117 | InN2 | CDH7 |
| LRP8 | 6.51E-121 | 0.66691327 | 0.438 | 0.304 | 2.49E-116 | InN2 | LRP8 |
| ST5 | 4.19E-120 | 0.62293296 | 0.481 | 0.337 | 1.60E-115 | InN2 | ST5 |
| AFAP11 | 1.11E-119 | 0.56262761 | 0.536 | 0.39 | 4.25E-115 | InN2 | AFAP1 |
| C3orf67-AS1 | 1.93E-118 | 0.65989772 | 0.426 | 0.275 | 7.39E-114 | InN2 | C3orf67-AS1 |
| OSBPL81 | 4.21E-118 | 0.51377521 | 0.766 | 0.752 | 1.61E-113 | InN2 | OSBPL8 |
| PEG3 | 5.96E-118 | 0.64209228 | 0.645 | 0.58 | 2.28E-113 | InN2 | PEG3 |
| RSPO21 | 1.69E-116 | 0.71756979 | 0.538 | 0.398 | 6.48E-112 | InN2 | RSPO2 |
| SIAH3 | 5.86E-114 | 0.5111152 | 0.378 | 0.226 | 2.24E-109 | InN2 | SIAH3 |
| SAMD52 | 8.75E-114 | 0.93186136 | 0.438 | 0.31 | 3.34E-109 | InN2 | SAMD5 |
| MPPED1 | 2.47E-113 | 0.57074967 | 0.562 | 0.447 | 9.45E-109 | InN2 | MPPED1 |
| KCNC2 | 1.05E-111 | 0.40032068 | 0.86 | 0.835 | 4.02E-107 | InN2 | KCNC2 |
| ANO41 | 3.32E-111 | 0.51830961 | 0.705 | 0.594 | 1.27E-106 | InN2 | ANO4 |
| NRIP31 | 2.22E-110 | 0.40275472 | 0.371 | 0.21 | 8.48E-106 | InN2 | NRIP3 |
| FARP12 | 5.45E-110 | 0.53087073 | 0.71 | 0.656 | 2.09E-105 | InN2 | FARP1 |
| CTTNBP21 | 2.37E-109 | 0.56334554 | 0.699 | 0.677 | 9.06E-105 | InN2 | CTTNBP2 |
| ARHGAP61 | 2.94E-109 | 0.81068545 | 0.301 | 0.168 | 1.12E-104 | InN2 | ARHGAP6 |
| CCDC136 | 2.67E-107 | 0.56290765 | 0.505 | 0.387 | 1.02E-102 | InN2 | CCDC136 |
| DPP6 | 7.42E-107 | 0.49619154 | 0.887 | 0.921 | 2.83E-102 | InN2 | DPP6 |
| SLC44A52 | 1.23E-105 | 0.50078898 | 0.776 | 0.691 | 4.69E-101 | InN2 | SLC44A5 |
| SH3PXD2A | 2.01E-105 | 0.54073769 | 0.715 | 0.704 | 7.67E-101 | InN2 | SH3PXD2A |
| SPHKAP1 | 1.73E-103 | 0.91181998 | 0.292 | 0.158 | 6.63E-99 | InN2 | SPHKAP |
| BICD11 | 1.02E-102 | 0.47830976 | 0.791 | 0.795 | 3.91E-98 | InN2 | BICD1 |
| TMSB4X1 | 2.32E-102 | 0.66123436 | 0.594 | 0.509 | 8.86E-98 | InN2 | TMSB4X |
| VPS13C | 7.61E-102 | 0.44284514 | 0.793 | 0.808 | 2.91E-97 | InN2 | VPS13C |
| ANKRD36C1 | 7.80E-102 | 0.46603613 | 0.799 | 0.818 | 2.98E-97 | InN2 | ANKRD36C |
| CELF41 | 7.44E-100 | 0.43416752 | 0.895 | 0.909 | 2.84E-95 | InN2 | CELF4 |
| GRIN2D1 | 6.04E-99 | 0.35821949 | 0.274 | 0.137 | 2.31E-94 | InN2 | GRIN2D |
| GRM12 | 1.31E-98 | 0.65398536 | 0.699 | 0.639 | 5.02E-94 | InN2 | GRM1 |
| TLN22 | 1.60E-97 | 0.55732074 | 0.758 | 0.736 | 6.10E-93 | InN2 | TLN2 |
| PRKG2 | 4.65E-97 | 0.57016364 | 0.296 | 0.172 | 1.78E-92 | InN2 | PRKG2 |
| KCNK22 | 4.67E-97 | 0.47461675 | 0.297 | 0.164 | 1.79E-92 | InN2 | KCNK2 |
| LINC011021 | 1.47E-96 | 0.53996157 | 0.457 | 0.319 | 5.61E-92 | InN2 | LINC01102 |
| SLC8A31 | 1.58E-96 | 0.57418647 | 0.583 | 0.482 | 6.05E-92 | InN2 | SLC8A3 |
| RAP1GDS11 | 1.06E-95 | 0.55664437 | 0.652 | 0.62 | 4.05E-91 | InN2 | RAP1GDS1 |
| INSYN2B2 | 2.21E-95 | 0.62918142 | 0.351 | 0.222 | 8.43E-91 | InN2 | INSYN2B |
| FRMD52 | 2.53E-95 | 0.37985469 | 0.898 | 0.863 | 9.65E-91 | InN2 | FRMD5 |
| HTRA12 | 1.74E-94 | 0.40896805 | 0.294 | 0.159 | 6.67E-90 | InN2 | HTRA1 |
| PLAGL11 | 1.21E-93 | 0.54404498 | 0.359 | 0.23 | 4.63E-89 | InN2 | PLAGL1 |
| REV3L1 | 5.78E-93 | 0.42652748 | 0.781 | 0.778 | 2.21E-88 | InN2 | REV3L |
| MAP3K52 | 7.22E-93 | 0.69559765 | 0.661 | 0.642 | 2.76E-88 | InN2 | MAP3K5 |
| MAGI22 | 1.30E-92 | 0.25648997 | 0.987 | 0.987 | 4.99E-88 | InN2 | MAGI2 |
| CDH23 | 1.64E-91 | 0.56194609 | 0.757 | 0.758 | 6.25E-87 | InN2 | CDH2 |
| SPRY4-AS1 | 2.21E-91 | 0.59224912 | 0.317 | 0.192 | 8.46E-87 | InN2 | SPRY4-AS1 |
| ADCY21 | 2.69E-91 | 0.53196955 | 0.784 | 0.86 | 1.03E-86 | InN2 | ADCY2 |
| EPB41L23 | 6.34E-90 | 0.44964086 | 0.791 | 0.709 | 2.42E-85 | InN2 | EPB41L2 |
| AP000943.21 | 1.55E-89 | 0.51966037 | 0.341 | 0.205 | 5.91E-85 | InN2 | AP000943.2 |
| NTM | 3.57E-89 | 0.55011958 | 0.813 | 0.855 | 1.37E-84 | InN2 | NTM |

| | | | | | | | |
|---|---|---|---|---|---|---|---|
| OSBPL101 | 4.18E-88 | 0.52361237 | 0.577 | 0.48 | 1.60E-83 | InN2 | OSBPL10 |
| FSTL5 | 6.32E-87 | 0.43940902 | 0.642 | 0.494 | 2.41E-82 | InN2 | FSTL5 |
| GRIN2B1 | 1.74E-86 | 0.28004394 | 0.93 | 0.948 | 6.66E-82 | InN2 | GRIN2B |
| ABAT | 1.55E-85 | 0.55668524 | 0.568 | 0.5 | 5.92E-81 | InN2 | ABAT |
| EVA1C1 | 1.70E-85 | 0.54063224 | 0.461 | 0.349 | 6.50E-81 | InN2 | EVA1C |
| UTRN1 | 2.01E-81 | 0.7650866 | 0.366 | 0.25 | 7.68E-77 | InN2 | UTRN |
| NF1 | 5.22E-81 | 0.40015207 | 0.797 | 0.818 | 1.99E-76 | InN2 | NF1 |
| RBMS1 | 5.17E-80 | 0.57408815 | 0.527 | 0.447 | 1.97E-75 | InN2 | RBMS1 |
| AC079296.11 | 8.24E-80 | 0.37130333 | 0.305 | 0.178 | 3.15E-75 | InN2 | AC079296.1 |
| ATP1B21 | 1.63E-79 | 0.39232236 | 0.363 | 0.225 | 6.24E-75 | InN2 | ATP1B2 |
| RPH3A | 5.75E-79 | 0.46795812 | 0.597 | 0.505 | 2.20E-74 | InN2 | RPH3A |
| NRSN1 | 6.35E-78 | 0.50726571 | 0.553 | 0.487 | 2.43E-73 | InN2 | NRSN1 |
| ANTXR11 | 7.24E-78 | 0.63841878 | 0.49 | 0.408 | 2.77E-73 | InN2 | ANTXR1 |
| SCN9A | 3.44E-77 | 0.52388363 | 0.308 | 0.196 | 1.31E-72 | InN2 | SCN9A |
| ADAMTS9-AS | 1.01E-76 | 0.81883767 | 0.458 | 0.341 | 3.87E-72 | InN2 | ADAMTS9-AS |
| AL033504.11 | 3.27E-76 | 0.74454191 | 0.368 | 0.258 | 1.25E-71 | InN2 | AL033504.1 |
| PTPRG | 2.32E-74 | 0.39818582 | 0.898 | 0.91 | 8.87E-70 | InN2 | PTPRG |
| ADGRL31 | 6.43E-74 | 0.25626008 | 0.972 | 0.963 | 2.46E-69 | InN2 | ADGRL3 |
| ARHGAP20 | 6.84E-74 | 0.56841022 | 0.522 | 0.464 | 2.62E-69 | InN2 | ARHGAP20 |
| PAN3 | 2.18E-72 | 0.47394691 | 0.677 | 0.683 | 8.34E-68 | InN2 | PAN3 |
| LGMN | 6.06E-72 | 0.46657061 | 0.369 | 0.262 | 2.32E-67 | InN2 | LGMN |
| PLOD22 | 6.60E-72 | 0.46479633 | 0.329 | 0.214 | 2.52E-67 | InN2 | PLOD2 |
| GNAS1 | 1.23E-71 | 0.43777726 | 0.806 | 0.827 | 4.68E-67 | InN2 | GNAS |
| STARD9 | 1.57E-71 | 0.5624178 | 0.44 | 0.345 | 6.02E-67 | InN2 | STARD9 |
| ARHGAP281 | 1.79E-71 | 0.42836522 | 0.323 | 0.203 | 6.85E-67 | InN2 | ARHGAP28 |
| TNRC6A1 | 1.86E-71 | 0.26394099 | 0.945 | 0.96 | 7.11E-67 | InN2 | TNRC6A |
| PWRN12 | 1.27E-70 | 0.37540357 | 0.604 | 0.464 | 4.84E-66 | InN2 | PWRN1 |
| PPM1K | 1.45E-70 | 0.51537477 | 0.607 | 0.589 | 5.53E-66 | InN2 | PPM1K |
| ADAMTS193 | 1.03E-69 | 0.66503701 | 0.456 | 0.364 | 3.94E-65 | InN2 | ADAMTS19 |
| MACF1 | 2.14E-69 | 0.32753901 | 0.897 | 0.924 | 8.18E-65 | InN2 | MACF1 |
| TRIO | 4.45E-68 | 0.29105488 | 0.909 | 0.933 | 1.70E-63 | InN2 | TRIO |
| TRPC41 | 5.92E-68 | 0.45588696 | 0.39 | 0.271 | 2.26E-63 | InN2 | TRPC4 |
| AGPAT4 | 1.06E-67 | 0.44980895 | 0.702 | 0.706 | 4.06E-63 | InN2 | AGPAT4 |
| LINC011223 | 5.63E-67 | 0.41573603 | 0.767 | 0.688 | 2.15E-62 | InN2 | LINC01122 |
| NFASC1 | 1.27E-66 | 0.43572583 | 0.78 | 0.814 | 4.85E-62 | InN2 | NFASC |
| ENOX11 | 3.31E-66 | 0.39542545 | 0.831 | 0.854 | 1.27E-61 | InN2 | ENOX1 |
| ITGA92 | 3.97E-66 | 0.70229361 | 0.496 | 0.432 | 1.52E-61 | InN2 | ITGA9 |
| KLF122 | 4.12E-66 | 0.43323407 | 0.665 | 0.61 | 1.58E-61 | InN2 | KLF12 |
| FAM184B | 9.08E-66 | 0.39925249 | 0.252 | 0.151 | 3.47E-61 | InN2 | FAM184B |
| NHS1 | 9.43E-66 | 0.39043168 | 0.57 | 0.445 | 3.60E-61 | InN2 | NHS |
| PWRN41 | 2.79E-65 | 0.38480192 | 0.345 | 0.22 | 1.07E-60 | InN2 | PWRN4 |
| CCSER11 | 3.93E-65 | 0.28495618 | 0.961 | 0.965 | 1.50E-60 | InN2 | CCSER1 |
| PALLD | 1.05E-64 | 0.46552323 | 0.501 | 0.413 | 4.01E-60 | InN2 | PALLD |
| ANKRD121 | 1.49E-64 | 0.45752897 | 0.759 | 0.782 | 5.71E-60 | InN2 | ANKRD12 |
| GALNT8 | 8.50E-64 | 0.4503639 | 0.396 | 0.297 | 3.25E-59 | InN2 | GALNT8 |
| TMEM178B1 | 6.01E-62 | 0.26639029 | 0.912 | 0.922 | 2.30E-57 | InN2 | TMEM178B |
| LRP1B4 | 1.83E-61 | 0.28782021 | 0.954 | 0.95 | 7.00E-57 | InN2 | LRP1B |
| WASF3 | 1.60E-60 | 0.44543261 | 0.463 | 0.38 | 6.10E-56 | InN2 | WASF3 |
| EPHA71 | 2.05E-60 | 0.53389966 | 0.592 | 0.543 | 7.86E-56 | InN2 | EPHA7 |
| TRHDE-AS11 | 5.48E-60 | 0.45768179 | 0.289 | 0.192 | 2.09E-55 | InN2 | TRHDE-AS1 |
| ZEB22 | 1.23E-58 | 0.31228582 | 0.816 | 0.823 | 4.72E-54 | InN2 | ZEB2 |
| GREB11 | 3.87E-58 | 0.5117925 | 0.422 | 0.337 | 1.48E-53 | InN2 | GREB1 |
| PDE1C2 | 7.82E-57 | 0.41679467 | 0.596 | 0.492 | 2.99E-52 | InN2 | PDE1C |
| RAB8B | 1.53E-56 | 0.41460086 | 0.38 | 0.287 | 5.85E-52 | InN2 | RAB8B |
| MPDZ | 1.77E-56 | 0.39715443 | 0.74 | 0.773 | 6.76E-52 | InN2 | MPDZ |
| RANBP171 | 2.16E-56 | 0.40761716 | 0.556 | 0.468 | 8.24E-52 | InN2 | RANBP17 |
| SLC9A91 | 1.36E-55 | 0.34025309 | 0.451 | 0.349 | 5.19E-51 | InN2 | SLC9A9 |
| NLGN4X | 1.08E-54 | 0.41123061 | 0.709 | 0.732 | 4.14E-50 | InN2 | NLGN4X |
| MARCH4 | 1.51E-54 | 0.49455571 | 0.473 | 0.404 | 5.76E-50 | InN2 | 4-Mar |
| FAM49B1 | 2.34E-53 | 0.3476098 | 0.775 | 0.81 | 8.94E-49 | InN2 | FAM49B |
| CDH4 | 3.77E-53 | 0.49309479 | 0.502 | 0.424 | 1.44E-48 | InN2 | CDH4 |
| MYRIP2 | 1.00E-52 | 0.34809946 | 0.812 | 0.807 | 3.83E-48 | InN2 | MYRIP |
| DIXDC11 | 1.01E-52 | 0.4343349 | 0.523 | 0.463 | 3.88E-48 | InN2 | DIXDC1 |

| | | | | | | | |
|---|---|---|---|---|---|---|---|
| AKAP7 | 1.16E-51 | 0.41994682 | 0.453 | 0.386 | 4.42E-47 | InN2 | AKAP7 |
| PTPRU1 | 1.30E-51 | 0.40053363 | 0.286 | 0.196 | 4.98E-47 | InN2 | PTPRU |
| SIDT12 | 2.29E-51 | 0.48321616 | 0.564 | 0.517 | 8.77E-47 | InN2 | SIDT1 |
| VAT1L | 3.15E-51 | 0.4273055 | 0.333 | 0.242 | 1.20E-46 | InN2 | VAT1L |
| MTSS12 | 4.41E-51 | 0.26929709 | 0.651 | 0.566 | 1.69E-46 | InN2 | MTSS1 |
| JAZF12 | 4.84E-51 | 0.33428177 | 0.792 | 0.824 | 1.85E-46 | InN2 | JAZF1 |
| EPHB61 | 4.85E-51 | 0.47818111 | 0.481 | 0.424 | 1.85E-46 | InN2 | EPHB6 |
| UMAD1 | 9.84E-51 | 0.45085571 | 0.65 | 0.651 | 3.76E-46 | InN2 | UMAD1 |
| SAMD4A1 | 3.10E-50 | 0.46549528 | 0.53 | 0.467 | 1.18E-45 | InN2 | SAMD4A |
| CACNB42 | 9.64E-50 | 0.32690847 | 0.843 | 0.855 | 3.68E-45 | InN2 | CACNB4 |
| ABCA5 | 1.40E-49 | 0.37032566 | 0.733 | 0.763 | 5.34E-45 | InN2 | ABCA5 |
| CHRM22 | 1.97E-49 | 0.48862217 | 0.312 | 0.213 | 7.53E-45 | InN2 | CHRM2 |
| PPP4R42 | 2.52E-49 | 0.44037838 | 0.474 | 0.401 | 9.62E-45 | InN2 | PPP4R4 |
| PARD31 | 3.48E-49 | 0.27856178 | 0.441 | 0.315 | 1.33E-44 | InN2 | PARD3 |
| INPP5F1 | 5.04E-49 | 0.42993979 | 0.591 | 0.583 | 1.93E-44 | InN2 | INPP5F |
| ILDR22 | 5.47E-49 | 0.46408552 | 0.417 | 0.34 | 2.09E-44 | InN2 | ILDR2 |
| POU3F31 | 1.52E-48 | 0.32487838 | 0.258 | 0.167 | 5.81E-44 | InN2 | POU3F3 |
| CABLES1 | 1.66E-48 | 0.54695177 | 0.424 | 0.361 | 6.33E-44 | InN2 | CABLES1 |
| GRID11 | 1.21E-47 | 0.2960316 | 0.836 | 0.854 | 4.61E-43 | InN2 | GRID1 |
| OSBPL91 | 1.70E-47 | 0.41272208 | 0.58 | 0.559 | 6.49E-43 | InN2 | OSBPL9 |
| RAB3IP | 9.43E-46 | 0.30640516 | 0.369 | 0.271 | 3.60E-41 | InN2 | RAB3IP |
| CCDC88A | 7.02E-45 | 0.31754311 | 0.831 | 0.847 | 2.68E-40 | InN2 | CCDC88A |
| TET21 | 1.55E-44 | 0.43797971 | 0.485 | 0.437 | 5.94E-40 | InN2 | TET2 |
| HUNK | 1.58E-44 | 0.44233433 | 0.386 | 0.307 | 6.04E-40 | InN2 | HUNK |
| REPS2 | 4.22E-44 | 0.37762439 | 0.687 | 0.709 | 1.61E-39 | InN2 | REPS2 |
| ST6GALNAC5 | 7.07E-44 | 0.34113156 | 0.776 | 0.711 | 2.70E-39 | InN2 | ST6GALNAC5 |
| PLA2R1 | 2.65E-43 | 0.38000666 | 0.271 | 0.189 | 1.01E-38 | InN2 | PLA2R1 |
| PDE3B2 | 2.76E-43 | 0.44248531 | 0.556 | 0.518 | 1.05E-38 | InN2 | PDE3B |
| EPHA61 | 2.92E-43 | 0.46226322 | 0.688 | 0.661 | 1.12E-38 | InN2 | EPHA6 |
| AHR1 | 1.02E-42 | 0.46053385 | 0.332 | 0.258 | 3.90E-38 | InN2 | AHR |
| C11orf58 | 1.73E-42 | 0.52258612 | 0.535 | 0.527 | 6.61E-38 | InN2 | C11orf58 |
| ANXA4 | 1.30E-41 | 0.40831544 | 0.252 | 0.175 | 4.98E-37 | InN2 | ANXA4 |
| TRAF3IP2-AS | 3.32E-41 | 0.3411687 | 0.723 | 0.752 | 1.27E-36 | InN2 | TRAF3IP2-AS |
| WWC2 | 6.21E-41 | 0.41411815 | 0.507 | 0.469 | 2.37E-36 | InN2 | WWC2 |
| OSBPL31 | 7.13E-40 | 0.34479062 | 0.503 | 0.435 | 2.73E-35 | InN2 | OSBPL3 |
| ST6GALNAC3 | 1.14E-38 | 0.47045893 | 0.548 | 0.506 | 4.35E-34 | InN2 | ST6GALNAC3 |
| FCHSD22 | 1.79E-38 | 0.30545204 | 0.643 | 0.614 | 6.86E-34 | InN2 | FCHSD2 |
| DAAM1 | 2.75E-38 | 0.33014121 | 0.725 | 0.754 | 1.05E-33 | InN2 | DAAM1 |
| SLITRK3 | 1.55E-37 | 0.34010366 | 0.276 | 0.201 | 5.91E-33 | InN2 | SLITRK3 |
| DGKE | 1.69E-37 | 0.39438056 | 0.468 | 0.43 | 6.47E-33 | InN2 | DGKE |
| DMD5 | 1.50E-36 | 0.28937672 | 0.878 | 0.925 | 5.71E-32 | InN2 | DMD |
| PHKB1 | 2.08E-36 | 0.38219967 | 0.573 | 0.576 | 7.96E-32 | InN2 | PHKB |
| RNF157 | 1.13E-35 | 0.39094284 | 0.541 | 0.529 | 4.33E-31 | InN2 | RNF157 |
| PRKN1 | 1.82E-35 | 0.27946064 | 0.834 | 0.866 | 6.96E-31 | InN2 | PRKN |
| RAPGEF61 | 2.29E-35 | 0.34469563 | 0.673 | 0.697 | 8.77E-31 | InN2 | RAPGEF6 |
| BACH22 | 3.01E-35 | 0.44740196 | 0.613 | 0.614 | 1.15E-30 | InN2 | BACH2 |
| TCF122 | 3.21E-35 | 0.33268637 | 0.691 | 0.708 | 1.23E-30 | InN2 | TCF12 |
| SEPTIN6 | 3.75E-35 | 0.36570244 | 0.357 | 0.288 | 1.43E-30 | InN2 | SEPTIN6 |
| CYTH3 | 1.15E-34 | 0.42164848 | 0.528 | 0.518 | 4.39E-30 | InN2 | CYTH3 |
| NREP1 | 1.81E-34 | 0.42344061 | 0.415 | 0.358 | 6.94E-30 | InN2 | NREP |
| SRGAP12 | 2.29E-34 | 0.35746114 | 0.326 | 0.251 | 8.75E-30 | InN2 | SRGAP1 |
| ARL4C1 | 3.46E-34 | 0.25680493 | 0.355 | 0.268 | 1.32E-29 | InN2 | ARL4C |
| POLA1 | 4.48E-34 | 0.42217331 | 0.447 | 0.4 | 1.71E-29 | InN2 | POLA1 |
| KCNB23 | 5.28E-34 | 0.41021534 | 0.71 | 0.688 | 2.02E-29 | InN2 | KCNB2 |
| FHIT2 | 1.54E-33 | 0.32643938 | 0.733 | 0.746 | 5.88E-29 | InN2 | FHIT |
| NAALAD21 | 2.20E-33 | 0.44090109 | 0.497 | 0.475 | 8.41E-29 | InN2 | NAALAD2 |
| AF117829.1 | 2.78E-33 | 0.40860471 | 0.543 | 0.533 | 1.06E-28 | InN2 | AF117829.1 |
| IL6ST1 | 5.60E-33 | 0.36470819 | 0.388 | 0.325 | 2.14E-28 | InN2 | IL6ST |
| ABLIM12 | 5.97E-33 | 0.25906857 | 0.819 | 0.835 | 2.28E-28 | InN2 | ABLIM1 |
| WWOX1 | 8.25E-33 | 0.36180687 | 0.782 | 0.822 | 3.15E-28 | InN2 | WWOX |
| CRTAC11 | 9.42E-33 | 0.30796476 | 0.314 | 0.236 | 3.60E-28 | InN2 | CRTAC1 |
| SPATS2L1 | 1.22E-32 | 0.35564545 | 0.658 | 0.659 | 4.65E-28 | InN2 | SPATS2L |
| VSTM2L | 1.14E-31 | 0.34533089 | 0.358 | 0.297 | 4.37E-27 | InN2 | VSTM2L |

| | | | | | | | |
|---|---|---|---|---|---|---|---|
| PRKCG | 1.53E-31 | 0.39222347 | 0.355 | 0.297 | 5.83E-27 | InN2 | PRKCG |
| AC091138.1 | 2.56E-31 | 0.37872135 | 0.339 | 0.284 | 9.80E-27 | InN2 | AC091138.1 |
| OSBPL6 | 2.91E-31 | 0.25969937 | 0.764 | 0.807 | 1.11E-26 | InN2 | OSBPL6 |
| GPI | 2.93E-31 | 0.4574054 | 0.615 | 0.663 | 1.12E-26 | InN2 | GPI |
| LRRFIP1 | 5.63E-31 | 0.30961904 | 0.707 | 0.753 | 2.15E-26 | InN2 | LRRFIP1 |
| FRAS11 | 1.57E-30 | 0.57255542 | 0.485 | 0.466 | 6.02E-26 | InN2 | FRAS1 |
| OLFM21 | 3.75E-30 | 0.31618259 | 0.366 | 0.295 | 1.43E-25 | InN2 | OLFM2 |
| CFAP1611 | 4.78E-30 | 0.33308781 | 0.382 | 0.315 | 1.83E-25 | InN2 | CFAP161 |
| SCD51 | 5.86E-30 | 0.35719293 | 0.603 | 0.61 | 2.24E-25 | InN2 | SCD5 |
| AC073050.1 | 1.36E-29 | 0.25518349 | 0.757 | 0.776 | 5.19E-25 | InN2 | AC073050.1 |
| KLHL322 | 2.07E-29 | 0.29556808 | 0.687 | 0.689 | 7.91E-25 | InN2 | KLHL32 |
| SH3GL32 | 2.75E-29 | 0.33246407 | 0.771 | 0.826 | 1.05E-24 | InN2 | SH3GL3 |
| PKD1L11 | 1.72E-28 | 0.34818385 | 0.33 | 0.265 | 6.59E-24 | InN2 | PKD1L1 |
| LINC023892 | 2.11E-28 | 0.40069544 | 0.507 | 0.474 | 8.07E-24 | InN2 | LINC02389 |
| DCX1 | 2.56E-28 | 0.30610279 | 0.448 | 0.396 | 9.79E-24 | InN2 | DCX |
| TRANK1 | 2.75E-28 | 0.40408919 | 0.579 | 0.598 | 1.05E-23 | InN2 | TRANK1 |
| CYTH12 | 3.31E-28 | 0.38235654 | 0.472 | 0.44 | 1.26E-23 | InN2 | CYTH1 |
| TMEM651 | 4.39E-28 | 0.38663971 | 0.429 | 0.399 | 1.68E-23 | InN2 | TMEM65 |
| ZNF618 | 5.76E-28 | 0.3680668 | 0.328 | 0.273 | 2.20E-23 | InN2 | ZNF618 |
| KSR1 | 6.02E-28 | 0.35280103 | 0.413 | 0.366 | 2.30E-23 | InN2 | KSR1 |
| AL158077.23 | 2.82E-27 | 0.38561027 | 0.337 | 0.275 | 1.08E-22 | InN2 | AL158077.2 |
| VSTM2B | 2.83E-27 | 0.41452135 | 0.38 | 0.339 | 1.08E-22 | InN2 | VSTM2B |
| PODXL21 | 5.19E-27 | 0.34119441 | 0.438 | 0.391 | 1.98E-22 | InN2 | PODXL2 |
| LINC027981 | 6.49E-27 | 0.34542534 | 0.274 | 0.21 | 2.48E-22 | InN2 | LINC02798 |
| PDE10A2 | 1.05E-26 | 0.33868883 | 0.691 | 0.678 | 4.00E-22 | InN2 | PDE10A |
| PRKG12 | 1.11E-26 | 0.35061184 | 0.647 | 0.607 | 4.24E-22 | InN2 | PRKG1 |
| SSBP21 | 1.52E-26 | 0.30804029 | 0.764 | 0.829 | 5.81E-22 | InN2 | SSBP2 |
| LINC011381 | 1.67E-26 | 0.39012932 | 0.455 | 0.431 | 6.38E-22 | InN2 | LINC01138 |
| NAV1 | 1.71E-26 | 0.30507838 | 0.651 | 0.678 | 6.53E-22 | InN2 | NAV1 |
| PTPRT3 | 4.65E-26 | 0.50330093 | 0.668 | 0.689 | 1.78E-21 | InN2 | PTPRT |
| PDZRN32 | 7.66E-26 | 0.26064552 | 0.765 | 0.765 | 2.93E-21 | InN2 | PDZRN3 |
| THSD7A1 | 1.30E-25 | 0.63191562 | 0.519 | 0.511 | 4.97E-21 | InN2 | THSD7A |
| STAT5B1 | 3.58E-25 | 0.36667769 | 0.39 | 0.352 | 1.37E-20 | InN2 | STAT5B |
| MSI21 | 7.12E-25 | 0.25000214 | 0.657 | 0.662 | 2.72E-20 | InN2 | MSI2 |
| PKD2 | 1.23E-24 | 0.35427289 | 0.597 | 0.625 | 4.68E-20 | InN2 | PKD2 |
| KIF21A | 1.61E-24 | 0.27930099 | 0.672 | 0.724 | 6.15E-20 | InN2 | KIF21A |
| TGFBR31 | 2.07E-24 | 0.27946236 | 0.25 | 0.189 | 7.91E-20 | InN2 | TGFBR3 |
| NIPAL3 | 2.86E-24 | 0.42156454 | 0.332 | 0.292 | 1.09E-19 | InN2 | NIPAL3 |
| SLC9A7 | 4.00E-24 | 0.32904093 | 0.425 | 0.39 | 1.53E-19 | InN2 | SLC9A7 |
| ENAH1 | 6.02E-24 | 0.3209128 | 0.544 | 0.543 | 2.30E-19 | InN2 | ENAH |
| GPR137C1 | 6.58E-24 | 0.32117847 | 0.602 | 0.609 | 2.51E-19 | InN2 | GPR137C |
| SERINC5 | 8.40E-24 | 0.36572225 | 0.451 | 0.424 | 3.21E-19 | InN2 | SERINC5 |
| AKAP9 | 1.60E-23 | 0.26368757 | 0.731 | 0.781 | 6.10E-19 | InN2 | AKAP9 |
| ACSL6 | 1.84E-23 | 0.32567505 | 0.476 | 0.457 | 7.05E-19 | InN2 | ACSL6 |
| ANKRD50 | 2.60E-23 | 0.26031632 | 0.255 | 0.197 | 9.94E-19 | InN2 | ANKRD50 |
| TRMT11 | 3.19E-23 | 0.28965921 | 0.747 | 0.807 | 1.22E-18 | InN2 | TRMT11 |
| HSPH1 | 4.34E-23 | 0.52613018 | 0.428 | 0.588 | 1.66E-18 | InN2 | HSPH1 |
| HMGB1 | 6.55E-23 | 0.29220018 | 0.752 | 0.803 | 2.50E-18 | InN2 | HMGB1 |
| TACC21 | 1.77E-22 | 0.32541392 | 0.438 | 0.413 | 6.78E-18 | InN2 | TACC2 |
| C2orf27A | 2.59E-22 | 0.3511717 | 0.345 | 0.303 | 9.89E-18 | InN2 | C2orf27A |
| CPSF6 | 2.66E-22 | 0.32090003 | 0.6 | 0.635 | 1.02E-17 | InN2 | CPSF6 |
| AC011474.11 | 3.43E-22 | 0.43617264 | 0.336 | 0.294 | 1.31E-17 | InN2 | AC011474.1 |
| GALNT21 | 5.81E-22 | 0.3440089 | 0.492 | 0.482 | 2.22E-17 | InN2 | GALNT2 |
| CHST152 | 1.14E-21 | 0.33204639 | 0.447 | 0.419 | 4.34E-17 | InN2 | CHST15 |
| ANKRD36B | 1.34E-21 | 0.35160567 | 0.523 | 0.543 | 5.11E-17 | InN2 | ANKRD36B |
| GLG1 | 1.44E-21 | 0.27525107 | 0.691 | 0.754 | 5.52E-17 | InN2 | GLG1 |
| ITFG1 | 1.67E-21 | 0.28893402 | 0.737 | 0.799 | 6.37E-17 | InN2 | ITFG1 |
| CORO2B | 2.28E-21 | 0.34355076 | 0.571 | 0.592 | 8.73E-17 | InN2 | CORO2B |
| FREM1 | 2.35E-21 | 0.38276596 | 0.275 | 0.221 | 8.98E-17 | InN2 | FREM1 |
| BCAT11 | 5.17E-21 | 0.28617932 | 0.388 | 0.35 | 1.97E-16 | InN2 | BCAT1 |
| DLEU7 | 5.64E-21 | 0.40017354 | 0.35 | 0.313 | 2.15E-16 | InN2 | DLEU7 |
| PLA2G4C | 5.64E-21 | 0.39687348 | 0.431 | 0.414 | 2.16E-16 | InN2 | PLA2G4C |
| APC1 | 8.59E-21 | 0.26644508 | 0.695 | 0.745 | 3.28E-16 | InN2 | APC |

| | | | | | | | |
|---|---|---|---|---|---|---|---|
| KCNMB2-AS1 | 1.19E-20 | 0.31712211 | 0.282 | 0.229 | 4.53E-16 | InN2 | KCNMB2-AS1 |
| ABCA10 | 1.41E-20 | 0.37963427 | 0.516 | 0.527 | 5.39E-16 | InN2 | ABCA10 |
| SRGAP22 | 2.77E-20 | 0.29264942 | 0.49 | 0.486 | 1.06E-15 | InN2 | SRGAP2 |
| TASP1 | 4.37E-20 | 0.33761381 | 0.533 | 0.549 | 1.67E-15 | InN2 | TASP1 |
| GNG22 | 5.75E-20 | 0.29495986 | 0.476 | 0.446 | 2.20E-15 | InN2 | GNG2 |
| PTPRJ2 | 1.14E-19 | 0.272939 | 0.602 | 0.629 | 4.36E-15 | InN2 | PTPRJ |
| PNMA2 | 2.65E-19 | 0.34513053 | 0.518 | 0.532 | 1.01E-14 | InN2 | PNMA2 |
| TMEFF1 | 3.74E-19 | 0.33613287 | 0.41 | 0.388 | 1.43E-14 | InN2 | TMEFF1 |
| PHF20L1 | 5.53E-19 | 0.34098983 | 0.588 | 0.639 | 2.12E-14 | InN2 | PHF20L1 |
| LINC011972 | 6.01E-19 | 0.3026059 | 0.399 | 0.351 | 2.30E-14 | InN2 | LINC01197 |
| MIR100HG1 | 6.29E-19 | 0.29946776 | 0.662 | 0.709 | 2.40E-14 | InN2 | MIR100HG |
| IGSF3 | 7.20E-19 | 0.27233862 | 0.254 | 0.207 | 2.75E-14 | InN2 | IGSF3 |
| MGLL1 | 1.05E-18 | 0.32106848 | 0.46 | 0.44 | 4.01E-14 | InN2 | MGLL |
| UNC13C | 1.49E-18 | 0.32427241 | 0.567 | 0.584 | 5.69E-14 | InN2 | UNC13C |
| RALGPS22 | 1.82E-18 | 0.33286302 | 0.531 | 0.543 | 6.95E-14 | InN2 | RALGPS2 |
| EEF1A12 | 2.16E-18 | 0.36160026 | 0.569 | 0.578 | 8.26E-14 | InN2 | EEF1A1 |
| ZFYVE9 | 2.36E-18 | 0.2895733 | 0.64 | 0.693 | 9.02E-14 | InN2 | ZFYVE9 |
| MAN1A11 | 6.84E-18 | 0.27790722 | 0.521 | 0.49 | 2.62E-13 | InN2 | MAN1A1 |
| LINC00342 | 9.05E-18 | 0.37056737 | 0.617 | 0.639 | 3.46E-13 | InN2 | LINC00342 |
| ETV61 | 2.74E-17 | 0.38408882 | 0.473 | 0.469 | 1.05E-12 | InN2 | ETV6 |
| MT-ATP81 | 4.65E-17 | 0.3261145 | 0.59 | 0.593 | 1.78E-12 | InN2 | MT-ATP8 |
| ZFHX2-AS11 | 7.09E-17 | 0.26412161 | 0.311 | 0.267 | 2.71E-12 | InN2 | ZFHX2-AS1 |
| ZC3H12C | 9.24E-17 | 0.329416 | 0.32 | 0.288 | 3.53E-12 | InN2 | ZC3H12C |
| XYLT12 | 9.87E-17 | 0.35319989 | 0.569 | 0.562 | 3.77E-12 | InN2 | XYLT1 |
| FUT81 | 1.01E-16 | 0.2866593 | 0.657 | 0.701 | 3.87E-12 | InN2 | FUT8 |
| ZFYVE16 | 1.39E-16 | 0.30670631 | 0.563 | 0.592 | 5.33E-12 | InN2 | ZFYVE16 |
| PPARG | 2.63E-16 | 0.31816143 | 0.314 | 0.274 | 1.00E-11 | InN2 | PPARG |
| FXYD61 | 2.86E-16 | 0.29005477 | 0.593 | 0.623 | 1.09E-11 | InN2 | FXYD6 |
| CACNG31 | 2.96E-16 | 0.26735185 | 0.652 | 0.657 | 1.13E-11 | InN2 | CACNG3 |
| RIMKLB | 5.86E-16 | 0.3036816 | 0.364 | 0.342 | 2.24E-11 | InN2 | RIMKLB |
| PCDH19 | 6.48E-16 | 0.31439094 | 0.367 | 0.337 | 2.48E-11 | InN2 | PCDH19 |
| FAM189A11 | 7.61E-16 | 0.30981177 | 0.612 | 0.638 | 2.91E-11 | InN2 | FAM189A1 |
| NRIP11 | 1.04E-15 | 0.37676218 | 0.506 | 0.525 | 3.98E-11 | InN2 | NRIP1 |
| PCDH11Y1 | 1.41E-15 | 0.66766296 | 0.267 | 0.226 | 5.39E-11 | InN2 | PCDH11Y |
| SASH12 | 1.84E-15 | 0.46542728 | 0.368 | 0.357 | 7.03E-11 | InN2 | SASH1 |
| AK3 | 2.91E-15 | 0.29552908 | 0.314 | 0.281 | 1.11E-10 | InN2 | AK3 |
| DENND2A1 | 1.43E-14 | 0.28155944 | 0.312 | 0.267 | 5.47E-10 | InN2 | DENND2A |
| ALCAM2 | 1.87E-14 | 0.39159745 | 0.482 | 0.473 | 7.14E-10 | InN2 | ALCAM |
| CALD11 | 1.88E-14 | 0.35067547 | 0.347 | 0.322 | 7.20E-10 | InN2 | CALD1 |
| AC092683.1 | 2.44E-14 | 0.42051655 | 0.434 | 0.427 | 9.33E-10 | InN2 | AC092683.1 |
| LINC00486 | 3.44E-14 | 0.35181813 | 0.595 | 0.627 | 1.31E-09 | InN2 | LINC00486 |
| KCNC1 | 4.51E-14 | 0.31091545 | 0.541 | 0.566 | 1.72E-09 | InN2 | KCNC1 |
| HS2ST1 | 7.61E-14 | 0.32147185 | 0.562 | 0.6 | 2.91E-09 | InN2 | HS2ST1 |
| CACNG2 | 8.35E-14 | 0.27694395 | 0.616 | 0.661 | 3.19E-09 | InN2 | CACNG2 |
| DENND5A | 8.62E-14 | 0.31512823 | 0.464 | 0.471 | 3.30E-09 | InN2 | DENND5A |
| GAP433 | 1.00E-13 | 0.26429781 | 0.553 | 0.541 | 3.84E-09 | InN2 | GAP43 |
| RNF1801 | 1.43E-13 | 0.27745229 | 0.547 | 0.584 | 5.48E-09 | InN2 | RNF180 |
| PEG10 | 2.31E-13 | 0.26388622 | 0.302 | 0.267 | 8.84E-09 | InN2 | PEG10 |
| DYNC1I2 | 4.06E-13 | 0.31197549 | 0.575 | 0.627 | 1.55E-08 | InN2 | DYNC1I2 |
| SLC36A11 | 5.07E-13 | 0.30676182 | 0.33 | 0.311 | 1.94E-08 | InN2 | SLC36A1 |
| KANSL1L | 7.58E-13 | 0.32369511 | 0.455 | 0.463 | 2.90E-08 | InN2 | KANSL1L |
| ANKRD28 | 9.55E-13 | 0.2740171 | 0.612 | 0.658 | 3.65E-08 | InN2 | ANKRD28 |
| EFHD22 | 1.82E-12 | 0.31023783 | 0.298 | 0.27 | 6.97E-08 | InN2 | EFHD2 |
| VSTM2A | 3.74E-12 | 0.26248817 | 0.411 | 0.395 | 1.43E-07 | InN2 | VSTM2A |
| AL627171.21 | 4.48E-12 | 0.34246371 | 0.457 | 0.455 | 1.71E-07 | InN2 | AL627171.2 |
| SNX25 | 4.95E-12 | 0.27074816 | 0.391 | 0.377 | 1.89E-07 | InN2 | SNX25 |
| GIT2 | 6.74E-12 | 0.29267688 | 0.501 | 0.531 | 2.57E-07 | InN2 | GIT2 |
| RBPJ1 | 7.39E-12 | 0.29818733 | 0.54 | 0.585 | 2.82E-07 | InN2 | RBPJ |
| SH3KBP12 | 8.06E-12 | 0.26743398 | 0.57 | 0.61 | 3.08E-07 | InN2 | SH3KBP1 |
| PCGF51 | 2.33E-11 | 0.25186782 | 0.58 | 0.636 | 8.91E-07 | InN2 | PCGF5 |
| ERO1B1 | 2.47E-11 | 0.25551905 | 0.42 | 0.419 | 9.46E-07 | InN2 | ERO1B |
| NCALD1 | 2.53E-11 | 0.32551607 | 0.688 | 0.778 | 9.67E-07 | InN2 | NCALD |
| POU6F1 | 2.97E-11 | 0.28337444 | 0.325 | 0.306 | 1.14E-06 | InN2 | POU6F1 |

| | | | | | | | |
|---|---|---|---|---|---|---|---|
| GMCL1 | 3.01E-11 | 0.289294 | 0.418 | 0.427 | 1.15E-06 | InN2 | GMCL1 |
| ADAM10 | 6.75E-11 | 0.29569552 | 0.56 | 0.612 | 2.58E-06 | InN2 | ADAM10 |
| LMBRD1 | 8.16E-11 | 0.27257205 | 0.572 | 0.631 | 3.12E-06 | InN2 | LMBRD1 |
| MGAT4A1 | 1.28E-10 | 0.30076367 | 0.457 | 0.472 | 4.89E-06 | InN2 | MGAT4A |
| AC046134.2 | 2.85E-10 | 0.30657571 | 0.524 | 0.563 | 1.09E-05 | InN2 | AC046134.2 |
| NUP931 | 3.09E-10 | 0.28529593 | 0.47 | 0.481 | 1.18E-05 | InN2 | NUP93 |
| SMARCA21 | 6.66E-10 | 0.32595517 | 0.72 | 0.814 | 2.55E-05 | InN2 | SMARCA2 |
| JARID21 | 8.32E-10 | 0.25235185 | 0.584 | 0.636 | 3.18E-05 | InN2 | JARID2 |
| DPYSL5 | 1.56E-09 | 0.30290107 | 0.358 | 0.35 | 5.95E-05 | InN2 | DPYSL5 |
| MIR646HG2 | 1.69E-09 | 0.43917708 | 0.301 | 0.278 | 6.47E-05 | InN2 | MIR646HG |
| TBC1D4 | 2.04E-09 | 0.28931366 | 0.459 | 0.468 | 7.81E-05 | InN2 | TBC1D4 |
| DIP2A2 | 2.21E-09 | 0.25409111 | 0.454 | 0.464 | 8.46E-05 | InN2 | DIP2A |
| CPEB2 | 4.35E-09 | 0.30868301 | 0.427 | 0.443 | 0.00016633 | InN2 | CPEB2 |
| NEXMIF | 4.83E-09 | 0.29199887 | 0.541 | 0.586 | 0.00018472 | InN2 | NEXMIF |
| ZBTB202 | 8.56E-09 | 0.36618526 | 0.524 | 0.555 | 0.00032706 | InN2 | ZBTB20 |
| SEL1L32 | 9.44E-09 | 0.30446001 | 0.31 | 0.292 | 0.00036088 | InN2 | SEL1L3 |
| CKMT2-AS1 | 1.12E-08 | 0.31041982 | 0.373 | 0.376 | 0.00042991 | InN2 | CKMT2-AS1 |
| N4BP21 | 1.74E-08 | 0.25796003 | 0.375 | 0.367 | 0.00066655 | InN2 | N4BP2 |
| CC2D2A | 2.07E-08 | 0.28505095 | 0.285 | 0.269 | 0.00079232 | InN2 | CC2D2A |
| KCNT1 | 4.75E-08 | 0.29915033 | 0.528 | 0.568 | 0.00181455 | InN2 | KCNT1 |
| SIPA1L31 | 7.47E-08 | 0.25269085 | 0.525 | 0.564 | 0.00285362 | InN2 | SIPA1L3 |
| KCNJ61 | 9.66E-08 | 0.25102195 | 0.671 | 0.747 | 0.00369226 | InN2 | KCNJ6 |
| MAP2K6 | 1.14E-07 | 0.28987356 | 0.35 | 0.349 | 0.00436894 | InN2 | MAP2K6 |
| MTCL1 | 1.32E-07 | 0.26104039 | 0.558 | 0.615 | 0.00505499 | InN2 | MTCL1 |
| AC034195.1 | 1.92E-06 | 0.27992591 | 0.269 | 0.252 | 0.07343572 | InN2 | AC034195.1 |
| LRRC8B1 | 2.20E-06 | 0.27460432 | 0.606 | 0.684 | 0.08407914 | InN2 | LRRC8B |
| EPB41L5 | 3.27E-06 | 0.25221016 | 0.431 | 0.455 | 0.1249568 | InN2 | EPB41L5 |
| DTD1 | 3.69E-06 | 0.25357568 | 0.423 | 0.449 | 0.14094938 | InN2 | DTD1 |
| GALNTL61 | 4.23E-06 | 0.26987203 | 0.593 | 0.756 | 0.16175649 | InN2 | GALNTL6 |
| MRPL41 | 8.04E-06 | 0.3703763 | 0.421 | 0.448 | 0.30736892 | InN2 | MRPL41 |
| LRRN21 | 6.29E-05 | 0.30673744 | 0.473 | 0.525 | 1 | InN2 | LRRN2 |
| HSPA4L1 | 7.80E-05 | 0.25659238 | 0.448 | 0.483 | 1 | InN2 | HSPA4L |
| ERICH31 | 0.0001139 | 0.25626687 | 0.419 | 0.448 | 1 | InN2 | ERICH3 |
| SGCZ2 | 0.000267 | 1.11020621 | 0.475 | 0.52 | 1 | InN2 | SGCZ |
| CTNNA1 | 0.00033946 | 0.26903652 | 0.33 | 0.34 | 1 | InN2 | CTNNA1 |
| SLC7A8 | 0.00035425 | 0.26829583 | 0.359 | 0.375 | 1 | InN2 | SLC7A8 |
| CDH111 | 0.00040637 | 0.27815381 | 0.431 | 0.457 | 1 | InN2 | CDH11 |
| RAPH11 | 0.00140467 | 0.28376584 | 0.456 | 0.507 | 1 | InN2 | RAPH1 |
| JAM3 | 0.00404352 | 0.26729678 | 0.464 | 0.517 | 1 | InN2 | JAM3 |
| SOX61 | 0 | 2.64375991 | 0.95 | 0.176 | 0 | InN3 | SOX6 |
| NXPH11 | 0 | 2.54121499 | 0.984 | 0.227 | 0 | InN3 | NXPH1 |
| DPP103 | 0 | 2.44508874 | 0.915 | 0.845 | 0 | InN3 | DPP10 |
| ERBB41 | 0 | 2.35437627 | 0.999 | 0.552 | 0 | InN3 | ERBB4 |
| TMEM132C | 0 | 2.26706461 | 0.87 | 0.227 | 0 | InN3 | TMEM132C |
| KIAA12172 | 0 | 2.18448045 | 0.99 | 0.639 | 0 | InN3 | KIAA1217 |
| SLC9A92 | 0 | 2.12897213 | 0.828 | 0.324 | 0 | InN3 | SLC9A9 |
| SLIT2 | 0 | 2.0874352 | 0.972 | 0.581 | 0 | InN3 | SLIT2 |
| MYO5B1 | 0 | 1.99631381 | 0.795 | 0.039 | 0 | InN3 | MYO5B |
| RUNX21 | 0 | 1.97117896 | 0.81 | 0.123 | 0 | InN3 | RUNX2 |
| MYO16 | 0 | 1.90636622 | 0.912 | 0.25 | 0 | InN3 | MYO16 |
| ADAMTS171 | 0 | 1.88679157 | 0.816 | 0.193 | 0 | InN3 | ADAMTS17 |
| PTPRM1 | 0 | 1.8867575 | 0.972 | 0.561 | 0 | InN3 | PTPRM |
| BTBD111 | 0 | 1.88483841 | 0.929 | 0.235 | 0 | InN3 | BTBD11 |
| SCN1A-AS11 | 0 | 1.88309647 | 0.828 | 0.234 | 0 | InN3 | SCN1A-AS1 |
| ZNF804A1 | 0 | 1.86349316 | 0.924 | 0.521 | 0 | InN3 | ZNF804A |
| ASTN22 | 0 | 1.69197448 | 0.98 | 0.723 | 0 | InN3 | ASTN2 |
| KCNC21 | 0 | 1.66984543 | 0.968 | 0.828 | 0 | InN3 | KCNC2 |
| TENM11 | 0 | 1.59770061 | 0.875 | 0.352 | 0 | InN3 | TENM1 |
| ADAMTS9-AS | 0 | 1.58872623 | 0.799 | 0.32 | 0 | InN3 | ADAMTS9-AS |
| SULF11 | 0 | 1.56964767 | 0.579 | 0.047 | 0 | InN3 | SULF1 |
| MEPE | 0 | 1.55587329 | 0.311 | 0.006 | 0 | InN3 | MEPE |
| ZNF385D3 | 0 | 1.51764766 | 0.969 | 0.488 | 0 | InN3 | ZNF385D |
| ZNF5361 | 0 | 1.50047237 | 0.954 | 0.34 | 0 | InN3 | ZNF536 |

| | | | | | | | |
|---|---|---|---|---|---|---|---|
| ADCY81 | 0 | 1.49413044 | 0.809 | 0.257 | 0 | InN3 | ADCY8 |
| ROR1 | 0 | 1.48635399 | 0.614 | 0.102 | 0 | InN3 | ROR1 |
| VWC21 | 0 | 1.48366482 | 0.848 | 0.327 | 0 | InN3 | VWC2 |
| NEAT12 | 0 | 1.47460781 | 0.725 | 0.362 | 0 | InN3 | NEAT1 |
| TRPC42 | 0 | 1.46557184 | 0.798 | 0.245 | 0 | InN3 | TRPC4 |
| EPHA62 | 0 | 1.46490012 | 0.903 | 0.647 | 0 | InN3 | EPHA6 |
| SPARCL13 | 0 | 1.42963737 | 0.92 | 0.651 | 0 | InN3 | SPARCL1 |
| ANK12 | 0 | 1.42782394 | 0.844 | 0.288 | 0 | InN3 | ANK1 |
| ANO42 | 0 | 1.41145655 | 0.932 | 0.58 | 0 | InN3 | ANO4 |
| SPOCK31 | 0 | 1.40095394 | 0.959 | 0.532 | 0 | InN3 | SPOCK3 |
| GAD21 | 0 | 1.36052885 | 0.846 | 0.224 | 0 | InN3 | GAD2 |
| LUZP21 | 0 | 1.33189165 | 0.646 | 0.292 | 0 | InN3 | LUZP2 |
| GAD12 | 0 | 1.32593619 | 0.837 | 0.256 | 0 | InN3 | GAD1 |
| PRKG13 | 0 | 1.30897088 | 0.947 | 0.588 | 0 | InN3 | PRKG1 |
| TENM32 | 0 | 1.30179022 | 0.922 | 0.659 | 0 | InN3 | TENM3 |
| CNTNAP3B1 | 0 | 1.29738008 | 0.66 | 0.214 | 0 | InN3 | CNTNAP3B |
| SGCZ3 | 0 | 1.28508704 | 0.868 | 0.493 | 0 | InN3 | SGCZ |
| RBMS31 | 0 | 1.28310784 | 0.945 | 0.623 | 0 | InN3 | RBMS3 |
| GRIK32 | 0 | 1.27379008 | 0.838 | 0.269 | 0 | InN3 | GRIK3 |
| KIF26B1 | 0 | 1.27248993 | 0.864 | 0.407 | 0 | InN3 | KIF26B |
| GPC62 | 0 | 1.2610483 | 0.742 | 0.478 | 0 | InN3 | GPC6 |
| KLHL54 | 0 | 1.20059748 | 0.766 | 0.292 | 0 | InN3 | KLHL5 |
| RPS6KA22 | 0 | 1.19427662 | 0.969 | 0.85 | 0 | InN3 | RPS6KA2 |
| OSBPL32 | 0 | 1.18151892 | 0.822 | 0.414 | 0 | InN3 | OSBPL3 |
| GABRG32 | 0 | 1.18041101 | 0.954 | 0.792 | 0 | InN3 | GABRG3 |
| FGF12 | 0 | 1.16515823 | 0.993 | 0.97 | 0 | InN3 | FGF12 |
| KCNMB23 | 0 | 1.16323581 | 0.933 | 0.555 | 0 | InN3 | KCNMB2 |
| PTCHD41 | 0 | 1.16092857 | 0.848 | 0.375 | 0 | InN3 | PTCHD4 |
| CNTNAP31 | 0 | 1.14893848 | 0.62 | 0.122 | 0 | InN3 | CNTNAP3 |
| KLF123 | 0 | 1.1404843 | 0.885 | 0.596 | 0 | InN3 | KLF12 |
| KAZN2 | 0 | 1.13544237 | 0.997 | 0.951 | 0 | InN3 | KAZN |
| LRRC4C3 | 0 | 1.12798771 | 0.973 | 0.885 | 0 | InN3 | LRRC4C |
| SUPT3H | 0 | 1.12284763 | 0.929 | 0.789 | 0 | InN3 | SUPT3H |
| PLCE11 | 0 | 1.11554807 | 0.554 | 0.12 | 0 | InN3 | PLCE1 |
| GRIP11 | 0 | 1.11298335 | 0.981 | 0.789 | 0 | InN3 | GRIP1 |
| TAFA4 | 0 | 1.09680011 | 0.484 | 0.041 | 0 | InN3 | TAFA4 |
| NHS2 | 0 | 1.08942657 | 0.842 | 0.428 | 0 | InN3 | NHS |
| PPARGC1A | 0 | 1.07222566 | 0.745 | 0.38 | 0 | InN3 | PPARGC1A |
| PAM2 | 0 | 1.06078836 | 0.919 | 0.749 | 0 | InN3 | PAM |
| ELAVL21 | 0 | 1.0565806 | 0.815 | 0.505 | 0 | InN3 | ELAVL2 |
| DOCK113 | 0 | 1.04828411 | 0.66 | 0.186 | 0 | InN3 | DOCK11 |
| FAR2 | 0 | 1.04789429 | 0.797 | 0.515 | 0 | InN3 | FAR2 |
| KCNS3 | 0 | 1.03519552 | 0.52 | 0.043 | 0 | InN3 | KCNS3 |
| SAMD53 | 0 | 1.03186159 | 0.686 | 0.295 | 0 | InN3 | SAMD5 |
| CNTNAP22 | 0 | 1.02988217 | 0.999 | 0.993 | 0 | InN3 | CNTNAP2 |
| SAT11 | 0 | 1.02755746 | 0.661 | 0.35 | 0 | InN3 | SAT1 |
| MYO1B | 0 | 1.01655954 | 0.595 | 0.235 | 0 | InN3 | MYO1B |
| TRPC51 | 0 | 1.01647808 | 0.705 | 0.307 | 0 | InN3 | TRPC5 |
| ETV62 | 0 | 1.00935248 | 0.778 | 0.449 | 0 | InN3 | ETV6 |
| ST8SIA42 | 0 | 1.00673431 | 0.576 | 0.16 | 0 | InN3 | ST8SIA4 |
| AL445623.21 | 0 | 1.00481835 | 0.733 | 0.356 | 0 | InN3 | AL445623.2 |
| RSPO22 | 0 | 1.00364555 | 0.731 | 0.386 | 0 | InN3 | RSPO2 |
| NCALD2 | 0 | 0.99249957 | 0.91 | 0.763 | 0 | InN3 | NCALD |
| SLC44A53 | 0 | 0.99193904 | 0.915 | 0.683 | 0 | InN3 | SLC44A5 |
| TOX32 | 0 | 0.99019339 | 0.629 | 0.163 | 0 | InN3 | TOX3 |
| PRDM11 | 0 | 0.98192681 | 0.499 | 0.036 | 0 | InN3 | PRDM1 |
| ESRRG1 | 0 | 0.98124574 | 0.86 | 0.674 | 0 | InN3 | ESRRG |
| PLCH12 | 0 | 0.97493377 | 0.688 | 0.183 | 0 | InN3 | PLCH1 |
| SNRPN2 | 0 | 0.96272238 | 0.938 | 0.752 | 0 | InN3 | SNRPN |
| FAM78B | 0 | 0.95605568 | 0.715 | 0.432 | 0 | InN3 | FAM78B |
| RGS5 | 0 | 0.95435931 | 0.474 | 0.066 | 0 | InN3 | RGS5 |
| OSBPL102 | 0 | 0.9532836 | 0.78 | 0.467 | 0 | InN3 | OSBPL10 |
| MAGI1 | 0 | 0.95182305 | 0.971 | 0.921 | 0 | InN3 | MAGI1 |

| | | | | | | | |
|---|---|---|---|---|---|---|---|
| XKR41 | 0 | 0.95102885 | 0.961 | 0.855 | 0 | InN3 | XKR4 |
| AP000943.22 | 0 | 0.95028348 | 0.575 | 0.191 | 0 | InN3 | AP000943.2 |
| SLC35F41 | 0 | 0.95024961 | 0.672 | 0.209 | 0 | InN3 | SLC35F4 |
| AL033504.12 | 0 | 0.94981267 | 0.614 | 0.242 | 0 | InN3 | AL033504.1 |
| KCND21 | 0 | 0.94352575 | 0.971 | 0.867 | 0 | InN3 | KCND2 |
| TENM22 | 0 | 0.93640326 | 0.99 | 0.931 | 0 | InN3 | TENM2 |
| ZMAT41 | 0 | 0.91410091 | 0.932 | 0.759 | 0 | InN3 | ZMAT4 |
| CPLX1 | 0 | 0.90049761 | 0.673 | 0.33 | 0 | InN3 | CPLX1 |
| PLCXD31 | 0 | 0.88447364 | 0.703 | 0.289 | 0 | InN3 | PLCXD3 |
| SFMBT22 | 0 | 0.88313661 | 0.651 | 0.37 | 0 | InN3 | SFMBT2 |
| ENOX12 | 0 | 0.88246945 | 0.96 | 0.845 | 0 | InN3 | ENOX1 |
| DTNA2 | 0 | 0.87615614 | 0.949 | 0.843 | 0 | InN3 | DTNA |
| CACNB43 | 0 | 0.8722878 | 0.956 | 0.848 | 0 | InN3 | CACNB4 |
| CLEC16A | 0 | 0.8617719 | 0.831 | 0.649 | 0 | InN3 | CLEC16A |
| LANCL11 | 0 | 0.86126832 | 0.729 | 0.489 | 0 | InN3 | LANCL1 |
| FRMD53 | 0 | 0.8563272 | 0.97 | 0.859 | 0 | InN3 | FRMD5 |
| SH3RF32 | 0 | 0.8546156 | 0.887 | 0.669 | 0 | InN3 | SH3RF3 |
| FHIT3 | 0 | 0.8541203 | 0.888 | 0.736 | 0 | InN3 | FHIT |
| SOX54 | 0 | 0.85398746 | 0.926 | 0.656 | 0 | InN3 | SOX5 |
| TAFA22 | 0 | 0.85156228 | 0.94 | 0.782 | 0 | InN3 | TAFA2 |
| TCF123 | 0 | 0.84900854 | 0.878 | 0.695 | 0 | InN3 | TCF12 |
| SERPINI11 | 0 | 0.84715492 | 0.841 | 0.669 | 0 | InN3 | SERPINI1 |
| PTPRZ11 | 0 | 0.84043627 | 0.623 | 0.285 | 0 | InN3 | PTPRZ1 |
| SLC26A4 | 0 | 0.83543443 | 0.59 | 0.247 | 0 | InN3 | SLC26A4 |
| SLC4A41 | 0 | 0.83109767 | 0.665 | 0.314 | 0 | InN3 | SLC4A4 |
| LINC023893 | 0 | 0.83105015 | 0.75 | 0.459 | 0 | InN3 | LINC02389 |
| TSPAN9 | 0 | 0.82441176 | 0.627 | 0.242 | 0 | InN3 | TSPAN9 |
| PLXNA41 | 0 | 0.82028322 | 0.92 | 0.792 | 0 | InN3 | PLXNA4 |
| MITF2 | 0 | 0.82009988 | 0.585 | 0.175 | 0 | InN3 | MITF |
| GRIP22 | 0 | 0.81691953 | 0.603 | 0.166 | 0 | InN3 | GRIP2 |
| CACNA2D21 | 0 | 0.81591641 | 0.567 | 0.157 | 0 | InN3 | CACNA2D2 |
| SLC6A13 | 0 | 0.81557463 | 0.87 | 0.325 | 0 | InN3 | SLC6A1 |
| LPP2 | 0 | 0.80929819 | 0.748 | 0.459 | 0 | InN3 | LPP |
| ARHGAP42 | 0 | 0.80308335 | 0.42 | 0.071 | 0 | InN3 | ARHGAP42 |
| ADCY12 | 0 | 0.80137191 | 0.809 | 0.589 | 0 | InN3 | ADCY1 |
| IL1RAPL1 | 0 | 0.8006568 | 0.993 | 0.974 | 0 | InN3 | IL1RAPL1 |
| PPARG1 | 0 | 0.79785063 | 0.596 | 0.256 | 0 | InN3 | PPARG |
| SILC12 | 0 | 0.79279465 | 0.606 | 0.197 | 0 | InN3 | SILC1 |
| COL12A13 | 0 | 0.79271389 | 0.527 | 0.18 | 0 | InN3 | COL12A1 |
| MTSS13 | 0 | 0.7897186 | 0.846 | 0.554 | 0 | InN3 | MTSS1 |
| PLD51 | 0 | 0.78491423 | 0.762 | 0.299 | 0 | InN3 | PLD5 |
| SCN1A1 | 0 | 0.78331859 | 0.934 | 0.838 | 0 | InN3 | SCN1A |
| HS6ST2 | 0 | 0.77743038 | 0.499 | 0.202 | 0 | InN3 | HS6ST2 |
| LHX61 | 0 | 0.77713775 | 0.508 | 0.049 | 0 | InN3 | LHX6 |
| DLGAP11 | 0 | 0.77610421 | 0.995 | 0.989 | 0 | InN3 | DLGAP1 |
| SLC24A25 | 0 | 0.77603957 | 0.963 | 0.827 | 0 | InN3 | SLC24A2 |
| TAC1 | 0 | 0.7688755 | 0.398 | 0.018 | 0 | InN3 | TAC1 |
| RNF144B1 | 0 | 0.76841953 | 0.438 | 0.031 | 0 | InN3 | RNF144B |
| CRTAC12 | 0 | 0.76104195 | 0.55 | 0.221 | 0 | InN3 | CRTAC1 |
| TBC1D41 | 0 | 0.7578852 | 0.716 | 0.451 | 0 | InN3 | TBC1D4 |
| LRRFIP11 | 0 | 0.75571094 | 0.869 | 0.742 | 0 | InN3 | LRRFIP1 |
| SETBP12 | 0 | 0.74282928 | 0.948 | 0.885 | 0 | InN3 | SETBP1 |
| NCOA21 | 0 | 0.74047659 | 0.866 | 0.736 | 0 | InN3 | NCOA2 |
| ALK | 0 | 0.73886657 | 0.661 | 0.224 | 0 | InN3 | ALK |
| CFAP1612 | 0 | 0.73729366 | 0.611 | 0.3 | 0 | InN3 | CFAP161 |
| PRKCA2 | 0 | 0.7357539 | 0.9 | 0.727 | 0 | InN3 | PRKCA |
| HCN12 | 0 | 0.73037285 | 0.936 | 0.855 | 0 | InN3 | HCN1 |
| KCNAB3 | 0 | 0.72672577 | 0.495 | 0.105 | 0 | InN3 | KCNAB3 |
| RHOBTB3 | 0 | 0.72401035 | 0.573 | 0.237 | 0 | InN3 | RHOBTB3 |
| NTRK21 | 0 | 0.70907489 | 0.969 | 0.943 | 0 | InN3 | NTRK2 |
| DGKD1 | 0 | 0.7070716 | 0.667 | 0.292 | 0 | InN3 | DGKD |
| COL4A2 | 0 | 0.70083576 | 0.561 | 0.229 | 0 | InN3 | COL4A2 |
| SIAH31 | 0 | 0.69228174 | 0.554 | 0.216 | 0 | InN3 | SIAH3 |

| | | | | | | | |
|---|---|---|---|---|---|---|---|
| WIF1 | 0 | 0.68553035 | 0.404 | 0.036 | 0 | InN3 | WIF1 |
| RAB3IP1 | 0 | 0.6829989 | 0.607 | 0.256 | 0 | InN3 | RAB3IP |
| TNIK3 | 0 | 0.68276683 | 0.915 | 0.798 | 0 | InN3 | TNIK |
| CADPS3 | 0 | 0.67840779 | 0.978 | 0.936 | 0 | InN3 | CADPS |
| AC087457.1 | 0 | 0.67828481 | 0.478 | 0.103 | 0 | InN3 | AC087457.1 |
| ADCYAP1R12 | 0 | 0.67794274 | 0.654 | 0.282 | 0 | InN3 | ADCYAP1R1 |
| CLMP1 | 0 | 0.66645604 | 0.449 | 0.126 | 0 | InN3 | CLMP |
| BCL11B2 | 0 | 0.66639401 | 0.583 | 0.239 | 0 | InN3 | BCL11B |
| MIR34AHG | 0 | 0.66604677 | 0.439 | 0.117 | 0 | InN3 | MIR34AHG |
| HGF | 0 | 0.66350082 | 0.351 | 0.025 | 0 | InN3 | HGF |
| MTUS12 | 0 | 0.65381294 | 0.608 | 0.281 | 0 | InN3 | MTUS1 |
| VAV22 | 0 | 0.63210961 | 0.522 | 0.165 | 0 | InN3 | VAV2 |
| DNM31 | 0 | 0.62760913 | 0.944 | 0.889 | 0 | InN3 | DNM3 |
| CAMTA11 | 0 | 0.61955221 | 0.957 | 0.923 | 0 | InN3 | CAMTA1 |
| ARX1 | 0 | 0.61423712 | 0.462 | 0.086 | 0 | InN3 | ARX |
| DLC11 | 0 | 0.6121057 | 0.723 | 0.341 | 0 | InN3 | DLC1 |
| MAF3 | 0 | 0.6091379 | 0.546 | 0.166 | 0 | InN3 | MAF |
| AMOTL11 | 0 | 0.58922462 | 0.498 | 0.188 | 0 | InN3 | AMOTL1 |
| GLRA21 | 0 | 0.58230706 | 0.515 | 0.194 | 0 | InN3 | GLRA2 |
| CCNJL | 0 | 0.58210435 | 0.422 | 0.113 | 0 | InN3 | CCNJL |
| PVALB | 0 | 0.57319563 | 0.251 | 0.004 | 0 | InN3 | PVALB |
| MYBPC1 | 0 | 0.55893718 | 0.258 | 0.05 | 0 | InN3 | MYBPC1 |
| RASSF81 | 0 | 0.55126075 | 0.458 | 0.162 | 0 | InN3 | RASSF8 |
| SGO1-AS1 | 0 | 0.50900348 | 0.41 | 0.121 | 0 | InN3 | SGO1-AS1 |
| C1QL1 | 0 | 0.50663897 | 0.286 | 0.026 | 0 | InN3 | C1QL1 |
| LYPD61 | 0 | 0.50506606 | 0.405 | 0.108 | 0 | InN3 | LYPD6 |
| GOLIM41 | 0 | 0.49908825 | 0.434 | 0.15 | 0 | InN3 | GOLIM4 |
| POSTN | 0 | 0.49344797 | 0.262 | 0.02 | 0 | InN3 | POSTN |
| MAGI23 | 0 | 0.49083548 | 0.994 | 0.987 | 0 | InN3 | MAGI2 |
| TENM3-AS1 | 0 | 0.49077154 | 0.307 | 0.063 | 0 | InN3 | TENM3-AS1 |
| FLT31 | 0 | 0.48839037 | 0.348 | 0.061 | 0 | InN3 | FLT3 |
| ACADL | 0 | 0.4829293 | 0.307 | 0.03 | 0 | InN3 | ACADL |
| NRXN32 | 0 | 0.46027523 | 0.997 | 0.99 | 0 | InN3 | NRXN3 |
| HHATL | 0 | 0.44292368 | 0.322 | 0.075 | 0 | InN3 | HHATL |
| GRIN2D2 | 0 | 0.44261967 | 0.417 | 0.129 | 0 | InN3 | GRIN2D |
| TACR1 | 0 | 0.43924027 | 0.345 | 0.081 | 0 | InN3 | TACR1 |
| MYO3B | 0 | 0.43133682 | 0.27 | 0.044 | 0 | InN3 | MYO3B |
| DNAI1 | 0 | 0.42885611 | 0.298 | 0.074 | 0 | InN3 | DNAI1 |
| NACC2 | 0 | 0.42853145 | 0.339 | 0.101 | 0 | InN3 | NACC2 |
| DUSP10 | 0 | 0.42682513 | 0.297 | 0.052 | 0 | InN3 | DUSP10 |
| RASSF31 | 0 | 0.41782514 | 0.306 | 0.079 | 0 | InN3 | RASSF3 |
| LINC00886 | 0 | 0.41631565 | 0.26 | 0.038 | 0 | InN3 | LINC00886 |
| RPH3AL | 0 | 0.41589571 | 0.261 | 0.062 | 0 | InN3 | RPH3AL |
| NDST4 | 0 | 0.40402151 | 0.278 | 0.058 | 0 | InN3 | NDST4 |
| SNHG141 | 0 | 0.39448414 | 0.97 | 0.946 | 0 | InN3 | SNHG14 |
| PAWR1 | 0 | 0.37215883 | 0.302 | 0.077 | 0 | InN3 | PAWR |
| PPARA | 0 | 0.36718911 | 0.29 | 0.062 | 0 | InN3 | PPARA |
| TIAM12 | 1.69E-305 | 0.7153442 | 0.89 | 0.747 | 6.45E-301 | InN3 | TIAM1 |
| ANKRD18A1 | 2.98E-305 | 0.52937513 | 0.367 | 0.12 | 1.14E-300 | InN3 | ANKRD18A |
| PLOD23 | 2.57E-304 | 0.57546524 | 0.497 | 0.204 | 9.83E-300 | InN3 | PLOD2 |
| KCTD81 | 1.68E-303 | 0.78087593 | 0.868 | 0.648 | 6.43E-299 | InN3 | KCTD8 |
| AL596087.2 | 1.14E-302 | 0.81598013 | 0.459 | 0.192 | 4.34E-298 | InN3 | AL596087.2 |
| GPR1491 | 2.01E-301 | 0.49257783 | 0.29 | 0.076 | 7.69E-297 | InN3 | GPR149 |
| ANKRD34C-A | 1.21E-298 | 0.7798879 | 0.739 | 0.483 | 4.61E-294 | InN3 | ANKRD34C-A |
| SNCG | 1.78E-297 | 0.37940254 | 0.301 | 0.086 | 6.80E-293 | InN3 | SNCG |
| ST8SIA12 | 3.62E-296 | 0.72850283 | 0.777 | 0.548 | 1.38E-291 | InN3 | ST8SIA1 |
| SDK12 | 1.99E-294 | 0.98364127 | 0.747 | 0.466 | 7.62E-290 | InN3 | SDK1 |
| EYS | 2.36E-292 | 0.69426584 | 0.328 | 0.102 | 9.03E-288 | InN3 | EYS |
| NEFH | 4.71E-287 | 0.32950811 | 0.266 | 0.07 | 1.80E-282 | InN3 | NEFH |
| FBLN7 | 1.64E-284 | 0.50632862 | 0.446 | 0.176 | 6.27E-280 | InN3 | FBLN7 |
| TMEM131L | 8.65E-284 | 0.68004122 | 0.607 | 0.324 | 3.31E-279 | InN3 | TMEM131L |
| ABCA1 | 2.24E-283 | 0.45517362 | 0.359 | 0.117 | 8.55E-279 | InN3 | ABCA1 |
| OPRD11 | 1.18E-277 | 0.71621765 | 0.533 | 0.26 | 4.50E-273 | InN3 | OPRD1 |

| | | | | | | | |
|---|---|---|---|---|---|---|---|
| GDPD5 | 1.63E-275 | 0.35260288 | 0.304 | 0.091 | 6.23E-271 | InN3 | GDPD5 |
| FGFR2 | 3.52E-275 | 0.6299087 | 0.451 | 0.187 | 1.34E-270 | InN3 | FGFR2 |
| OXR14 | 4.50E-274 | 0.56062278 | 0.97 | 0.919 | 1.72E-269 | InN3 | OXR1 |
| FRMD4B2 | 1.82E-272 | 0.54053169 | 0.423 | 0.155 | 6.94E-268 | InN3 | FRMD4B |
| AFAP12 | 1.49E-270 | 0.65443389 | 0.663 | 0.383 | 5.70E-266 | InN3 | AFAP1 |
| HUNK1 | 5.55E-269 | 0.65488455 | 0.574 | 0.295 | 2.12E-264 | InN3 | HUNK |
| FGF13 | 4.72E-268 | 0.43049138 | 0.675 | 0.404 | 1.80E-263 | InN3 | FGF13 |
| GLDC | 1.10E-266 | 0.44502337 | 0.329 | 0.108 | 4.21E-262 | InN3 | GLDC |
| MEF2C2 | 3.53E-266 | 0.57201551 | 0.969 | 0.937 | 1.35E-261 | InN3 | MEF2C |
| INPP5J | 2.16E-265 | 0.28853164 | 0.25 | 0.066 | 8.26E-261 | InN3 | INPP5J |
| KCNAB1 | 1.04E-262 | 0.56305041 | 0.377 | 0.133 | 3.96E-258 | InN3 | KCNAB1 |
| GNAL2 | 1.05E-262 | 0.61061388 | 0.784 | 0.461 | 4.01E-258 | InN3 | GNAL |
| RAB3C1 | 3.87E-262 | 0.58462994 | 0.841 | 0.627 | 1.48E-257 | InN3 | RAB3C |
| TMEM178A1 | 1.30E-260 | 0.7134316 | 0.775 | 0.582 | 4.95E-256 | InN3 | TMEM178A |
| AC092969.12 | 5.63E-259 | 0.49013482 | 0.405 | 0.149 | 2.15E-254 | InN3 | AC092969.1 |
| MARCH112 | 1.59E-258 | 0.63008738 | 0.603 | 0.327 | 6.07E-254 | InN3 | 11-Mar |
| GALNT72 | 7.17E-258 | 0.68860653 | 0.726 | 0.502 | 2.74E-253 | InN3 | GALNT7 |
| SLC8A32 | 8.86E-257 | 0.68024377 | 0.725 | 0.473 | 3.39E-252 | InN3 | SLC8A3 |
| FHOD31 | 2.28E-253 | 0.69283922 | 0.815 | 0.63 | 8.71E-249 | InN3 | FHOD3 |
| GRK3 | 1.08E-250 | 0.63715969 | 0.851 | 0.7 | 4.15E-246 | InN3 | GRK3 |
| PLCL12 | 1.74E-250 | 0.84505929 | 0.828 | 0.639 | 6.64E-246 | InN3 | PLCL1 |
| NDST31 | 1.97E-250 | 0.75129084 | 0.791 | 0.547 | 7.53E-246 | InN3 | NDST3 |
| SCN7A | 4.88E-248 | 0.42790308 | 0.305 | 0.097 | 1.87E-243 | InN3 | SCN7A |
| AFF21 | 1.02E-247 | 0.64454189 | 0.654 | 0.376 | 3.89E-243 | InN3 | AFF2 |
| AC006148.1 | 5.20E-246 | 0.74925699 | 0.853 | 0.7 | 1.99E-241 | InN3 | AC006148.1 |
| DNER1 | 1.52E-245 | 0.4919868 | 0.8 | 0.514 | 5.80E-241 | InN3 | DNER |
| SATB1-AS13 | 2.23E-245 | 0.66471944 | 0.721 | 0.422 | 8.52E-241 | InN3 | SATB1-AS1 |
| ERC11 | 1.51E-244 | 0.54654196 | 0.922 | 0.864 | 5.78E-240 | InN3 | ERC1 |
| KIAA12112 | 1.62E-244 | 0.47688543 | 0.73 | 0.382 | 6.20E-240 | InN3 | KIAA1211 |
| NAV21 | 2.78E-242 | 0.45944607 | 0.962 | 0.919 | 1.06E-237 | InN3 | NAV2 |
| SYTL5 | 6.63E-242 | 0.40053921 | 0.308 | 0.098 | 2.53E-237 | InN3 | SYTL5 |
| FMN14 | 3.04E-241 | 0.61194484 | 0.908 | 0.752 | 1.16E-236 | InN3 | FMN1 |
| AC008105.3 | 3.05E-241 | 0.54673243 | 0.471 | 0.219 | 1.17E-236 | InN3 | AC008105.3 |
| DCX2 | 5.15E-241 | 0.60291858 | 0.644 | 0.383 | 1.97E-236 | InN3 | DCX |
| UPP11 | 1.12E-237 | 0.59811529 | 0.58 | 0.327 | 4.29E-233 | InN3 | UPP1 |
| AKNA1 | 1.79E-233 | 0.30711575 | 0.261 | 0.078 | 6.84E-229 | InN3 | AKNA |
| NAV11 | 9.35E-233 | 0.63846655 | 0.81 | 0.667 | 3.57E-228 | InN3 | NAV1 |
| KLHL131 | 3.47E-232 | 0.41369138 | 0.325 | 0.111 | 1.32E-227 | InN3 | KLHL13 |
| CNTN42 | 7.17E-232 | 0.65414862 | 0.925 | 0.848 | 2.74E-227 | InN3 | CNTN4 |
| MSI22 | 4.46E-231 | 0.57511078 | 0.83 | 0.651 | 1.70E-226 | InN3 | MSI2 |
| ASIC22 | 2.05E-230 | 0.57104282 | 0.965 | 0.841 | 7.85E-226 | InN3 | ASIC2 |
| LYPD6B2 | 2.26E-230 | 0.41582533 | 0.425 | 0.167 | 8.63E-226 | InN3 | LYPD6B |
| SPTB | 2.78E-230 | 0.56673951 | 0.571 | 0.318 | 1.06E-225 | InN3 | SPTB |
| SEPTIN4 | 1.46E-229 | 0.44499586 | 0.402 | 0.165 | 5.60E-225 | InN3 | SEPTIN4 |
| NXPH21 | 4.15E-227 | 0.32509408 | 0.316 | 0.102 | 1.59E-222 | InN3 | NXPH2 |
| ZEB23 | 3.47E-222 | 0.57171943 | 0.92 | 0.816 | 1.33E-217 | InN3 | ZEB2 |
| CMTM81 | 3.47E-221 | 0.41420537 | 0.296 | 0.098 | 1.33E-216 | InN3 | CMTM8 |
| INPP4B3 | 1.32E-220 | 0.46532245 | 0.921 | 0.745 | 5.03E-216 | InN3 | INPP4B |
| IGF12 | 6.98E-219 | 0.33602216 | 0.38 | 0.139 | 2.67E-214 | InN3 | IGF1 |
| FCHSD23 | 7.65E-219 | 0.57083807 | 0.8 | 0.604 | 2.93E-214 | InN3 | FCHSD2 |
| PCDH152 | 4.97E-217 | 0.95733064 | 0.567 | 0.314 | 1.90E-212 | InN3 | PCDH15 |
| SHTN13 | 4.09E-215 | 0.60153596 | 0.812 | 0.668 | 1.56E-210 | InN3 | SHTN1 |
| ARHGAP123 | 1.53E-213 | 0.51861451 | 0.583 | 0.323 | 5.85E-209 | InN3 | ARHGAP12 |
| STXBP62 | 8.93E-213 | 0.47886587 | 0.677 | 0.371 | 3.41E-208 | InN3 | STXBP6 |
| PDE3B3 | 1.37E-212 | 0.67700758 | 0.726 | 0.507 | 5.23E-208 | InN3 | PDE3B |
| UBASH3B2 | 1.73E-210 | 0.45068054 | 0.617 | 0.326 | 6.61E-206 | InN3 | UBASH3B |
| CYTH31 | 1.02E-209 | 0.64681125 | 0.703 | 0.506 | 3.88E-205 | InN3 | CYTH3 |
| FAM135B2 | 1.76E-209 | 0.61909031 | 0.816 | 0.622 | 6.71E-205 | InN3 | FAM135B |
| KANSL1L1 | 3.25E-209 | 0.60937819 | 0.668 | 0.448 | 1.24E-204 | InN3 | KANSL1L |
| QKI3 | 2.94E-207 | 0.47971737 | 0.876 | 0.662 | 1.12E-202 | InN3 | QKI |
| SLITRK2 | 4.75E-207 | 0.38314947 | 0.345 | 0.136 | 1.82E-202 | InN3 | SLITRK2 |
| PALLD1 | 8.27E-206 | 0.57663749 | 0.646 | 0.404 | 3.16E-201 | InN3 | PALLD |
| NF11 | 1.89E-205 | 0.52153655 | 0.873 | 0.813 | 7.22E-201 | InN3 | NF1 |

| | | | | | | | |
|---|---|---|---|---|---|---|---|
| ANKS1A1 | 1.03E-204 | 0.62953657 | 0.744 | 0.589 | 3.95E-200 | InN3 | ANKS1A |
| MCC2 | 5.14E-204 | 0.46040478 | 0.525 | 0.25 | 1.96E-199 | InN3 | MCC |
| SLC35F14 | 1.52E-203 | 0.51910971 | 0.918 | 0.698 | 5.80E-199 | InN3 | SLC35F1 |
| ACSL61 | 6.36E-202 | 0.58279557 | 0.656 | 0.445 | 2.43E-197 | InN3 | ACSL6 |
| ADARB1 | 7.61E-202 | 0.62193844 | 0.674 | 0.479 | 2.91E-197 | InN3 | ADARB1 |
| CLMN | 1.62E-200 | 0.48614856 | 0.388 | 0.163 | 6.18E-196 | InN3 | CLMN |
| VAV32 | 2.57E-200 | 0.46587209 | 0.35 | 0.137 | 9.84E-196 | InN3 | VAV3 |
| GREB12 | 1.33E-199 | 0.54272746 | 0.566 | 0.328 | 5.08E-195 | InN3 | GREB1 |
| ME3 | 6.04E-198 | 0.57321168 | 0.829 | 0.701 | 2.31E-193 | InN3 | ME3 |
| TSHZ32 | 9.69E-197 | 0.58825266 | 0.674 | 0.434 | 3.70E-192 | InN3 | TSHZ3 |
| DIXDC12 | 4.57E-196 | 0.59009454 | 0.67 | 0.454 | 1.75E-191 | InN3 | DIXDC1 |
| XYLT13 | 6.14E-196 | 0.70894598 | 0.76 | 0.549 | 2.35E-191 | InN3 | XYLT1 |
| BACH13 | 8.42E-196 | 0.42177385 | 0.473 | 0.221 | 3.22E-191 | InN3 | BACH1 |
| UBA6-AS1 | 3.82E-195 | 0.68508658 | 0.762 | 0.596 | 1.46E-190 | InN3 | UBA6-AS1 |
| RASA11 | 1.32E-194 | 0.60431158 | 0.8 | 0.705 | 5.06E-190 | InN3 | RASA1 |
| ABAT1 | 2.27E-193 | 0.55215432 | 0.695 | 0.492 | 8.68E-189 | InN3 | ABAT |
| SYN22 | 1.52E-192 | 0.46750802 | 0.94 | 0.898 | 5.82E-188 | InN3 | SYN2 |
| PLCH2 | 2.44E-187 | 0.41183383 | 0.401 | 0.182 | 9.34E-183 | InN3 | PLCH2 |
| AC007405.1 | 4.79E-187 | 0.34002498 | 0.303 | 0.112 | 1.83E-182 | InN3 | AC007405.1 |
| UNC5D2 | 6.10E-187 | 0.64124781 | 0.945 | 0.825 | 2.33E-182 | InN3 | UNC5D |
| GHR2 | 3.05E-185 | 0.57646082 | 0.62 | 0.371 | 1.17E-180 | InN3 | GHR |
| GRIA41 | 1.60E-179 | 0.43643357 | 0.968 | 0.838 | 6.10E-175 | InN3 | GRIA4 |
| NINL1 | 3.26E-179 | 0.3433486 | 0.403 | 0.177 | 1.24E-174 | InN3 | NINL |
| AC025809.12 | 3.79E-176 | 0.48836889 | 0.542 | 0.308 | 1.45E-171 | InN3 | AC025809.1 |
| PTPRN22 | 4.02E-174 | 0.4006237 | 0.968 | 0.949 | 1.54E-169 | InN3 | PTPRN2 |
| GABRB2 | 2.39E-173 | 0.42858665 | 0.949 | 0.921 | 9.12E-169 | InN3 | GABRB2 |
| SHISA91 | 2.71E-173 | 0.48830641 | 0.936 | 0.817 | 1.04E-168 | InN3 | SHISA9 |
| RNASET22 | 5.88E-173 | 0.42214768 | 0.48 | 0.254 | 2.25E-168 | InN3 | RNASET2 |
| PITPNC12 | 7.93E-173 | 0.40737983 | 0.743 | 0.481 | 3.03E-168 | InN3 | PITPNC1 |
| RASGRF22 | 1.67E-172 | 0.48668375 | 0.839 | 0.594 | 6.39E-168 | InN3 | RASGRF2 |
| RALGPS11 | 2.09E-172 | 0.50566213 | 0.817 | 0.701 | 7.99E-168 | InN3 | RALGPS1 |
| PLPPR12 | 3.62E-170 | 0.52576007 | 0.53 | 0.284 | 1.38E-165 | InN3 | PLPPR1 |
| CEMIP3 | 4.58E-170 | 0.74823675 | 0.61 | 0.399 | 1.75E-165 | InN3 | CEMIP |
| TRERF11 | 1.30E-168 | 0.57112669 | 0.788 | 0.649 | 4.98E-164 | InN3 | TRERF1 |
| SPAG161 | 6.43E-167 | 0.55996743 | 0.806 | 0.68 | 2.46E-162 | InN3 | SPAG16 |
| SLC4A72 | 9.01E-166 | 0.51176603 | 0.639 | 0.437 | 3.45E-161 | InN3 | SLC4A7 |
| GAS2 | 2.60E-165 | 0.55944281 | 0.595 | 0.4 | 9.93E-161 | InN3 | GAS2 |
| DECR1 | 1.77E-164 | 0.42572178 | 0.459 | 0.248 | 6.78E-160 | InN3 | DECR1 |
| DEPTOR1 | 2.93E-163 | 0.46094126 | 0.474 | 0.254 | 1.12E-158 | InN3 | DEPTOR |
| PLEKHA7 | 7.14E-163 | 0.29469381 | 0.258 | 0.095 | 2.73E-158 | InN3 | PLEKHA7 |
| NCOA31 | 2.75E-161 | 0.54599687 | 0.592 | 0.385 | 1.05E-156 | InN3 | NCOA3 |
| PRPSAP1 | 1.70E-160 | 0.46772354 | 0.515 | 0.305 | 6.48E-156 | InN3 | PRPSAP1 |
| CLEC2L | 1.76E-160 | 0.44259415 | 0.429 | 0.224 | 6.73E-156 | InN3 | CLEC2L |
| PDE8A1 | 8.92E-160 | 0.50760679 | 0.636 | 0.429 | 3.41E-155 | InN3 | PDE8A |
| TIMP22 | 3.42E-159 | 0.40084198 | 0.537 | 0.292 | 1.31E-154 | InN3 | TIMP2 |
| MANBA1 | 1.59E-158 | 0.51754513 | 0.575 | 0.372 | 6.08E-154 | InN3 | MANBA |
| TOX22 | 3.61E-158 | 0.40228111 | 0.409 | 0.195 | 1.38E-153 | InN3 | TOX2 |
| FILIP11 | 2.88E-157 | 0.40447607 | 0.25 | 0.094 | 1.10E-152 | InN3 | FILIP1 |
| TGFBR32 | 4.41E-156 | 0.42278662 | 0.381 | 0.181 | 1.68E-151 | InN3 | TGFBR3 |
| PTPN131 | 7.50E-156 | 0.51801724 | 0.617 | 0.385 | 2.87E-151 | InN3 | PTPN13 |
| TRPS11 | 4.55E-155 | 0.57360955 | 0.636 | 0.428 | 1.74E-150 | InN3 | TRPS1 |
| AAK11 | 1.68E-154 | 0.39588056 | 0.907 | 0.878 | 6.41E-150 | InN3 | AAK1 |
| AC233296.11 | 1.24E-153 | 0.44592645 | 0.498 | 0.257 | 4.75E-149 | InN3 | AC233296.1 |
| GPR176 | 2.28E-153 | 0.53164525 | 0.523 | 0.323 | 8.73E-149 | InN3 | GPR176 |
| CLIP4 | 2.55E-153 | 0.51242521 | 0.685 | 0.535 | 9.76E-149 | InN3 | CLIP4 |
| EFR3B | 3.44E-152 | 0.56390713 | 0.681 | 0.495 | 1.32E-147 | InN3 | EFR3B |
| AL158077.24 | 6.61E-152 | 0.479355 | 0.479 | 0.266 | 2.53E-147 | InN3 | AL158077.2 |
| COL4A1 | 1.69E-151 | 0.35731064 | 0.345 | 0.157 | 6.44E-147 | InN3 | COL4A1 |
| PCLO4 | 2.21E-150 | 0.38326816 | 0.962 | 0.921 | 8.45E-146 | InN3 | PCLO |
| ATP8A2 | 4.32E-150 | 0.30411011 | 0.973 | 0.951 | 1.65E-145 | InN3 | ATP8A2 |
| CBLB | 1.59E-149 | 0.49190111 | 0.599 | 0.401 | 6.07E-145 | InN3 | CBLB |
| WLS1 | 2.37E-149 | 0.38040325 | 0.501 | 0.265 | 9.08E-145 | InN3 | WLS |
| KATNAL2 | 4.08E-149 | 0.45217594 | 0.44 | 0.239 | 1.56E-144 | InN3 | KATNAL2 |

| | | | | | | | |
|---|---|---|---|---|---|---|---|
| ZNF124 | 5.45E-149 | 0.37892277 | 0.34 | 0.158 | 2.08E-144 | InN3 | ZNF124 |
| ATRNL14 | 1.84E-148 | 0.42868786 | 0.981 | 0.928 | 7.02E-144 | InN3 | ATRNL1 |
| GRIN2B2 | 8.47E-148 | 0.34301575 | 0.962 | 0.946 | 3.24E-143 | InN3 | GRIN2B |
| GAB22 | 1.22E-147 | 0.5161435 | 0.682 | 0.496 | 4.66E-143 | InN3 | GAB2 |
| SGK1 | 2.64E-147 | 0.58886615 | 0.681 | 0.499 | 1.01E-142 | InN3 | SGK1 |
| CACNA1A2 | 5.14E-145 | 0.4410836 | 0.959 | 0.907 | 1.96E-140 | InN3 | CACNA1A |
| SRRM42 | 2.41E-144 | 0.46247616 | 0.9 | 0.852 | 9.20E-140 | InN3 | SRRM4 |
| LUZP1 | 2.17E-143 | 0.51391581 | 0.67 | 0.509 | 8.28E-139 | InN3 | LUZP1 |
| OSBPL61 | 1.03E-142 | 0.44337464 | 0.853 | 0.801 | 3.95E-138 | InN3 | OSBPL6 |
| PPP2R2B3 | 2.16E-142 | 0.36739328 | 0.976 | 0.955 | 8.25E-138 | InN3 | PPP2R2B |
| LRRC391 | 6.44E-142 | 0.27164778 | 0.344 | 0.155 | 2.46E-137 | InN3 | LRRC39 |
| EPB412 | 1.12E-141 | 0.52812616 | 0.641 | 0.447 | 4.27E-137 | InN3 | EPB41 |
| AC115622.1 | 1.21E-141 | 0.31653066 | 0.253 | 0.099 | 4.64E-137 | InN3 | AC115622.1 |
| TRIO1 | 3.18E-141 | 0.33694826 | 0.947 | 0.93 | 1.22E-136 | InN3 | TRIO |
| DOK6 | 1.44E-140 | 0.60400165 | 0.825 | 0.7 | 5.52E-136 | InN3 | DOK6 |
| LRP81 | 2.56E-140 | 0.46023622 | 0.497 | 0.301 | 9.78E-136 | InN3 | LRP8 |
| CHRM23 | 3.23E-140 | 0.8392063 | 0.387 | 0.209 | 1.23E-135 | InN3 | CHRM2 |
| KCNC11 | 4.32E-140 | 0.5055853 | 0.71 | 0.554 | 1.65E-135 | InN3 | KCNC1 |
| LDLRAD43 | 7.24E-140 | 0.47670322 | 0.888 | 0.775 | 2.77E-135 | InN3 | LDLRAD4 |
| VPS13D3 | 6.99E-139 | 0.47410669 | 0.839 | 0.748 | 2.67E-134 | InN3 | VPS13D |
| UTRN2 | 5.76E-138 | 0.51187693 | 0.446 | 0.245 | 2.20E-133 | InN3 | UTRN |
| LINC027982 | 5.50E-137 | 0.39315099 | 0.396 | 0.203 | 2.10E-132 | InN3 | LINC02798 |
| RBFOX12 | 7.38E-137 | 0.29937742 | 0.999 | 0.983 | 2.82E-132 | InN3 | RBFOX1 |
| EEF2K | 1.70E-135 | 0.45537635 | 0.505 | 0.31 | 6.49E-131 | InN3 | EEF2K |
| TASOR2 | 1.07E-134 | 0.5004609 | 0.677 | 0.523 | 4.10E-130 | InN3 | TASOR2 |
| TBC1D30 | 1.58E-134 | 0.39189806 | 0.487 | 0.29 | 6.05E-130 | InN3 | TBC1D30 |
| RYR12 | 1.60E-133 | 0.47639276 | 0.409 | 0.227 | 6.13E-129 | InN3 | RYR1 |
| KLF3-AS11 | 6.69E-133 | 0.60197523 | 0.441 | 0.258 | 2.56E-128 | InN3 | KLF3-AS1 |
| DENND1B1 | 1.07E-132 | 0.45427882 | 0.661 | 0.496 | 4.07E-128 | InN3 | DENND1B |
| APC2 | 1.83E-132 | 0.45049844 | 0.811 | 0.737 | 7.01E-128 | InN3 | APC |
| NTN42 | 4.52E-132 | 0.34262176 | 0.421 | 0.22 | 1.73E-127 | InN3 | NTN4 |
| KSR11 | 7.32E-130 | 0.42525094 | 0.555 | 0.357 | 2.80E-125 | InN3 | KSR1 |
| ABLIM13 | 1.14E-129 | 0.402493 | 0.903 | 0.83 | 4.35E-125 | InN3 | ABLIM1 |
| FNBP1L2 | 3.67E-129 | 0.44306077 | 0.594 | 0.379 | 1.40E-124 | InN3 | FNBP1L |
| PRKCE | 2.19E-128 | 0.35282312 | 0.942 | 0.912 | 8.37E-124 | InN3 | PRKCE |
| NQO2 | 6.34E-128 | 0.50631237 | 0.689 | 0.554 | 2.42E-123 | InN3 | NQO2 |
| ARHGAP201 | 7.03E-128 | 0.49068757 | 0.616 | 0.459 | 2.69E-123 | InN3 | ARHGAP20 |
| CNTN52 | 1.75E-126 | 0.46157941 | 0.909 | 0.811 | 6.69E-122 | InN3 | CNTN5 |
| HS3ST1 | 2.44E-126 | 0.37533802 | 0.327 | 0.16 | 9.31E-122 | InN3 | HS3ST1 |
| CNTNAP51 | 8.52E-126 | 0.51984585 | 0.926 | 0.9 | 3.26E-121 | InN3 | CNTNAP5 |
| WWOX2 | 1.38E-125 | 0.47192643 | 0.901 | 0.813 | 5.28E-121 | InN3 | WWOX |
| ZFYVE161 | 3.64E-125 | 0.50712904 | 0.716 | 0.582 | 1.39E-120 | InN3 | ZFYVE16 |
| TCF42 | 1.10E-123 | 0.28135522 | 0.971 | 0.957 | 4.21E-119 | InN3 | TCF4 |
| ATP10D1 | 3.55E-123 | 0.34336213 | 0.406 | 0.223 | 1.36E-118 | InN3 | ATP10D |
| AHI1 | 5.44E-122 | 0.36883655 | 0.943 | 0.919 | 2.08E-117 | InN3 | AHI1 |
| ENTPD31 | 6.30E-122 | 0.30501129 | 0.499 | 0.285 | 2.41E-117 | InN3 | ENTPD3 |
| GPSM2 | 7.68E-122 | 0.25971043 | 0.258 | 0.109 | 2.93E-117 | InN3 | GPSM2 |
| PHACTR22 | 1.35E-120 | 0.43262579 | 0.599 | 0.4 | 5.17E-116 | InN3 | PHACTR2 |
| SH3PXD2A1 | 1.79E-120 | 0.43831601 | 0.796 | 0.699 | 6.83E-116 | InN3 | SH3PXD2A |
| KCNMB2-AS: | 2.25E-120 | 0.40658541 | 0.408 | 0.221 | 8.59E-116 | InN3 | KCNMB2-AS1 |
| JARID22 | 4.43E-120 | 0.4551727 | 0.738 | 0.625 | 1.69E-115 | InN3 | JARID2 |
| GPRC5B | 6.61E-120 | 0.28004794 | 0.344 | 0.163 | 2.53E-115 | InN3 | GPRC5B |
| TEAD11 | 1.93E-119 | 0.47909863 | 0.705 | 0.556 | 7.37E-115 | InN3 | TEAD1 |
| CCDC911 | 2.16E-119 | 0.41369692 | 0.792 | 0.69 | 8.25E-115 | InN3 | CCDC91 |
| ADGRL32 | 8.77E-119 | 0.3107743 | 0.982 | 0.963 | 3.35E-114 | InN3 | ADGRL3 |
| RAB3B2 | 1.12E-118 | 0.30383013 | 0.284 | 0.128 | 4.29E-114 | InN3 | RAB3B |
| PKD21 | 2.81E-118 | 0.45980291 | 0.721 | 0.617 | 1.07E-113 | InN3 | PKD2 |
| TCP11L2 | 3.22E-118 | 0.27647995 | 0.282 | 0.128 | 1.23E-113 | InN3 | TCP11L2 |
| FAM189A12 | 4.94E-118 | 0.48543098 | 0.751 | 0.628 | 1.89E-113 | InN3 | FAM189A1 |
| GRM83 | 1.13E-117 | 0.72048392 | 0.517 | 0.346 | 4.31E-113 | InN3 | GRM8 |
| AK31 | 1.96E-117 | 0.36091441 | 0.461 | 0.271 | 7.50E-113 | InN3 | AK3 |
| DAAM11 | 1.43E-116 | 0.4074921 | 0.818 | 0.747 | 5.46E-112 | InN3 | DAAM1 |
| MB21D22 | 2.13E-116 | 0.44771277 | 0.572 | 0.387 | 8.14E-112 | InN3 | MB21D2 |

| | | | | | | | |
|---|---|---|---|---|---|---|---|
| TMEM1082 | 2.89E-116 | 0.39315988 | 0.906 | 0.816 | 1.10E-111 | InN3 | TMEM108 |
| PKNOX2 | 3.88E-116 | 0.48197172 | 0.835 | 0.753 | 1.48E-111 | InN3 | PKNOX2 |
| SASH13 | 1.07E-115 | 0.47403344 | 0.531 | 0.346 | 4.08E-111 | InN3 | SASH1 |
| GRID12 | 1.66E-114 | 0.39230023 | 0.89 | 0.85 | 6.36E-110 | InN3 | GRID1 |
| TNRC6A2 | 6.20E-114 | 0.298327 | 0.962 | 0.958 | 2.37E-109 | InN3 | TNRC6A |
| ATP1B11 | 5.90E-113 | 0.45264754 | 0.84 | 0.776 | 2.25E-108 | InN3 | ATP1B1 |
| GNG73 | 5.11E-112 | 0.42715037 | 0.584 | 0.399 | 1.95E-107 | InN3 | GNG7 |
| FAM49B2 | 1.46E-111 | 0.37304351 | 0.856 | 0.805 | 5.57E-107 | InN3 | FAM49B |
| ZNF4621 | 1.54E-110 | 0.32822636 | 0.474 | 0.279 | 5.88E-106 | InN3 | ZNF462 |
| SLC7A81 | 3.84E-110 | 0.39766529 | 0.535 | 0.363 | 1.47E-105 | InN3 | SLC7A8 |
| KIAA1324L | 4.26E-110 | 0.42432768 | 0.65 | 0.491 | 1.63E-105 | InN3 | KIAA1324L |
| NOS11 | 4.75E-110 | 0.26323337 | 0.253 | 0.11 | 1.81E-105 | InN3 | NOS1 |
| PLAGL12 | 7.49E-109 | 0.30591454 | 0.409 | 0.227 | 2.86E-104 | InN3 | PLAGL1 |
| FGD42 | 7.20E-108 | 0.43957241 | 0.5 | 0.325 | 2.75E-103 | InN3 | FGD4 |
| ADRA1A1 | 1.69E-107 | 0.25352454 | 0.402 | 0.204 | 6.44E-103 | InN3 | ADRA1A |
| MIR181A1HG | 1.82E-107 | 0.45064454 | 0.617 | 0.437 | 6.94E-103 | InN3 | MIR181A1HG |
| SERINC51 | 7.30E-107 | 0.41435792 | 0.594 | 0.415 | 2.79E-102 | InN3 | SERINC5 |
| COG5 | 1.68E-106 | 0.40809426 | 0.784 | 0.706 | 6.43E-102 | InN3 | COG5 |
| GABRG3-AS1 | 3.82E-106 | 0.28505354 | 0.259 | 0.118 | 1.46E-101 | InN3 | GABRG3-AS1 |
| BACH23 | 5.05E-106 | 0.46143682 | 0.741 | 0.605 | 1.93E-101 | InN3 | BACH2 |
| TCAIM | 6.23E-106 | 0.41516661 | 0.546 | 0.387 | 2.38E-101 | InN3 | TCAIM |
| LRCH11 | 1.16E-105 | 0.40836895 | 0.677 | 0.535 | 4.44E-101 | InN3 | LRCH1 |
| FLRT3 | 5.90E-105 | 0.35053884 | 0.37 | 0.207 | 2.26E-100 | InN3 | FLRT3 |
| USP2-AS1 | 6.02E-105 | 0.34614685 | 0.372 | 0.207 | 2.30E-100 | InN3 | USP2-AS1 |
| RAPGEF42 | 8.50E-105 | 0.31600108 | 0.907 | 0.874 | 3.25E-100 | InN3 | RAPGEF4 |
| ZC3H12C1 | 1.91E-104 | 0.37925136 | 0.45 | 0.28 | 7.31E-100 | InN3 | ZC3H12C |
| GRIA11 | 2.63E-104 | 0.36741465 | 0.948 | 0.921 | 1.01E-99 | InN3 | GRIA1 |
| STS | 5.32E-104 | 0.50522194 | 0.574 | 0.409 | 2.03E-99 | InN3 | STS |
| STARD91 | 1.49E-103 | 0.44648014 | 0.508 | 0.342 | 5.70E-99 | InN3 | STARD9 |
| PCTP | 1.50E-103 | 0.29750678 | 0.3 | 0.15 | 5.73E-99 | InN3 | PCTP |
| KCNIP21 | 2.41E-103 | 0.35753902 | 0.536 | 0.346 | 9.22E-99 | InN3 | KCNIP2 |
| PRKN2 | 3.46E-103 | 0.37905709 | 0.9 | 0.861 | 1.32E-98 | InN3 | PRKN |
| DIP2C1 | 4.91E-103 | 0.33043072 | 0.91 | 0.888 | 1.88E-98 | InN3 | DIP2C |
| ITSN1 | 3.80E-101 | 0.39772486 | 0.806 | 0.748 | 1.45E-96 | InN3 | ITSN1 |
| LIMA1 | 3.82E-101 | 0.30286079 | 0.361 | 0.196 | 1.46E-96 | InN3 | LIMA1 |
| PACRG2 | 7.04E-100 | 0.45620633 | 0.799 | 0.704 | 2.69E-95 | InN3 | PACRG |
| TTBK2 | 1.13E-99 | 0.39469623 | 0.77 | 0.68 | 4.31E-95 | InN3 | TTBK2 |
| MACF11 | 2.03E-99 | 0.3133004 | 0.946 | 0.92 | 7.78E-95 | InN3 | MACF1 |
| MEIKIN | 2.61E-99 | 0.35996646 | 0.334 | 0.179 | 9.98E-95 | InN3 | MEIKIN |
| PPIP5K2 | 4.07E-99 | 0.43199903 | 0.714 | 0.623 | 1.56E-94 | InN3 | PPIP5K2 |
| AIG1 | 4.97E-99 | 0.39726584 | 0.835 | 0.77 | 1.90E-94 | InN3 | AIG1 |
| ZNF718 | 1.58E-97 | 0.39817973 | 0.524 | 0.369 | 6.05E-93 | InN3 | ZNF718 |
| TDRD3 | 2.02E-97 | 0.4378245 | 0.641 | 0.509 | 7.72E-93 | InN3 | TDRD3 |
| CDC42EP31 | 8.99E-97 | 0.29221985 | 0.33 | 0.175 | 3.43E-92 | InN3 | CDC42EP3 |
| ARHGAP291 | 1.82E-96 | 0.31174151 | 0.343 | 0.186 | 6.97E-92 | InN3 | ARHGAP29 |
| AFF14 | 2.35E-96 | 0.38331573 | 0.585 | 0.405 | 8.98E-92 | InN3 | AFF1 |
| TAOK31 | 2.50E-96 | 0.42217622 | 0.773 | 0.706 | 9.56E-92 | InN3 | TAOK3 |
| ATP2B2 | 3.43E-96 | 0.32412737 | 0.921 | 0.877 | 1.31E-91 | InN3 | ATP2B2 |
| GSAP2 | 4.58E-96 | 0.30318473 | 0.416 | 0.247 | 1.75E-91 | InN3 | GSAP |
| SACS | 8.91E-96 | 0.41740023 | 0.747 | 0.637 | 3.41E-91 | InN3 | SACS |
| BEND6 | 1.63E-95 | 0.4136213 | 0.589 | 0.443 | 6.24E-91 | InN3 | BEND6 |
| AC073050.11 | 2.07E-95 | 0.39750093 | 0.862 | 0.769 | 7.92E-91 | InN3 | AC073050.1 |
| CUX13 | 3.22E-95 | 0.35690304 | 0.845 | 0.724 | 1.23E-90 | InN3 | CUX1 |
| DSCAML12 | 3.88E-95 | 0.35492253 | 0.842 | 0.755 | 1.48E-90 | InN3 | DSCAML1 |
| AF117829.11 | 1.13E-94 | 0.429849 | 0.656 | 0.526 | 4.33E-90 | InN3 | AF117829.1 |
| EPRS | 3.93E-94 | 0.40663542 | 0.567 | 0.418 | 1.50E-89 | InN3 | EPRS |
| PTPRG1 | 1.88E-93 | 0.39578516 | 0.927 | 0.908 | 7.19E-89 | InN3 | PTPRG |
| WASF31 | 3.38E-93 | 0.34593474 | 0.537 | 0.376 | 1.29E-88 | InN3 | WASF3 |
| NEK1 | 3.63E-93 | 0.37975507 | 0.717 | 0.611 | 1.39E-88 | InN3 | NEK1 |
| SPIN1 | 7.03E-92 | 0.3542173 | 0.777 | 0.717 | 2.69E-87 | InN3 | SPIN1 |
| SEPTIN61 | 1.31E-91 | 0.31336643 | 0.457 | 0.282 | 5.01E-87 | InN3 | SEPTIN6 |
| SETD7 | 6.64E-91 | 0.31214136 | 0.393 | 0.232 | 2.54E-86 | InN3 | SETD7 |
| FAM110B1 | 1.04E-90 | 0.311295 | 0.599 | 0.424 | 3.99E-86 | InN3 | FAM110B |

| | | | | | | | |
|---|---|---|---|---|---|---|---|
| DLGAP23 | 1.17E-90 | 0.31001335 | 0.975 | 0.929 | 4.47E-86 | InN3 | DLGAP2 |
| UMAD11 | 6.24E-89 | 0.39104959 | 0.741 | 0.645 | 2.39E-84 | InN3 | UMAD1 |
| AC011346.1 | 1.57E-88 | 0.45670832 | 0.483 | 0.324 | 5.99E-84 | InN3 | AC011346.1 |
| ARSG | 3.60E-88 | 0.34563583 | 0.413 | 0.256 | 1.37E-83 | InN3 | ARSG |
| CCDC1361 | 5.37E-88 | 0.34843736 | 0.549 | 0.385 | 2.05E-83 | InN3 | CCDC136 |
| RASAL22 | 2.09E-87 | 0.30526771 | 0.913 | 0.876 | 8.00E-83 | InN3 | RASAL2 |
| ZNF487 | 2.48E-87 | 0.31905352 | 0.333 | 0.187 | 9.46E-83 | InN3 | ZNF487 |
| LGMN1 | 5.74E-87 | 0.34348358 | 0.416 | 0.26 | 2.19E-82 | InN3 | LGMN |
| CYP4V2 | 7.20E-87 | 0.30150808 | 0.384 | 0.234 | 2.75E-82 | InN3 | CYP4V2 |
| KCNT11 | 2.16E-86 | 0.42337361 | 0.666 | 0.558 | 8.24E-82 | InN3 | KCNT1 |
| MEG81 | 2.92E-85 | 0.32473475 | 0.894 | 0.845 | 1.12E-80 | InN3 | MEG8 |
| CORO6 | 3.00E-85 | 0.43923277 | 0.557 | 0.429 | 1.15E-80 | InN3 | CORO6 |
| PCSK61 | 6.39E-85 | 0.28494739 | 0.372 | 0.21 | 2.44E-80 | InN3 | PCSK6 |
| AL645568.1 | 6.43E-85 | 0.35981206 | 0.588 | 0.445 | 2.46E-80 | InN3 | AL645568.1 |
| CHCHD3 | 8.48E-85 | 0.37139453 | 0.76 | 0.686 | 3.24E-80 | InN3 | CHCHD3 |
| AGBL44 | 9.83E-85 | 0.26256388 | 0.973 | 0.931 | 3.76E-80 | InN3 | AGBL4 |
| WWC21 | 2.04E-84 | 0.36428756 | 0.608 | 0.462 | 7.79E-80 | InN3 | WWC2 |
| DST | 2.16E-84 | 0.26399874 | 0.964 | 0.953 | 8.25E-80 | InN3 | DST |
| RASSF42 | 6.22E-84 | 0.25932859 | 0.379 | 0.222 | 2.38E-79 | InN3 | RASSF4 |
| HTR41 | 2.58E-83 | 0.44654002 | 0.392 | 0.248 | 9.84E-79 | InN3 | HTR4 |
| NEGR13 | 5.14E-83 | 0.33254858 | 0.979 | 0.954 | 1.96E-78 | InN3 | NEGR1 |
| DLEU71 | 1.85E-82 | 0.35183666 | 0.463 | 0.306 | 7.06E-78 | InN3 | DLEU7 |
| ST6GALNAC3 | 1.24E-81 | 0.40446057 | 0.648 | 0.5 | 4.75E-77 | InN3 | ST6GALNAC3 |
| ZNF516 | 1.31E-81 | 0.33383281 | 0.491 | 0.322 | 5.02E-77 | InN3 | ZNF516 |
| GABRD | 1.44E-81 | 0.35818658 | 0.401 | 0.256 | 5.51E-77 | InN3 | GABRD |
| EFCAB21 | 5.71E-81 | 0.41272281 | 0.638 | 0.511 | 2.18E-76 | InN3 | EFCAB2 |
| SLC7A14 | 7.90E-81 | 0.37914435 | 0.643 | 0.508 | 3.02E-76 | InN3 | SLC7A14 |
| SMARCA22 | 2.01E-80 | 0.35226978 | 0.84 | 0.806 | 7.68E-76 | InN3 | SMARCA2 |
| SND11 | 2.59E-79 | 0.39695793 | 0.765 | 0.695 | 9.92E-75 | InN3 | SND1 |
| VPS13C1 | 5.22E-79 | 0.34117813 | 0.835 | 0.805 | 2.00E-74 | InN3 | VPS13C |
| VSTM2L1 | 4.73E-78 | 0.32046135 | 0.446 | 0.292 | 1.81E-73 | InN3 | VSTM2L |
| SIPA1L32 | 5.13E-78 | 0.3820626 | 0.675 | 0.554 | 1.96E-73 | InN3 | SIPA1L3 |
| ANKRD442 | 1.25E-77 | 0.30306539 | 0.585 | 0.421 | 4.77E-73 | InN3 | ANKRD44 |
| IDH3A | 1.43E-77 | 0.31116193 | 0.422 | 0.278 | 5.48E-73 | InN3 | IDH3A |
| LINC008941 | 2.75E-77 | 0.41269941 | 0.541 | 0.396 | 1.05E-72 | InN3 | LINC00894 |
| CALN13 | 3.16E-77 | 0.39586315 | 0.766 | 0.657 | 1.21E-72 | InN3 | CALN1 |
| C2orf27A1 | 4.27E-77 | 0.32647064 | 0.447 | 0.297 | 1.63E-72 | InN3 | C2orf27A |
| GRM52 | 8.49E-77 | 0.27767063 | 0.97 | 0.896 | 3.25E-72 | InN3 | GRM5 |
| NDUFS4 | 2.73E-76 | 0.3390224 | 0.544 | 0.412 | 1.04E-71 | InN3 | NDUFS4 |
| ARHGEF10L | 5.69E-76 | 0.28140963 | 0.376 | 0.232 | 2.17E-71 | InN3 | ARHGEF10L |
| TRPM32 | 7.93E-76 | 0.97516328 | 0.552 | 0.404 | 3.03E-71 | InN3 | TRPM3 |
| MROH1 | 1.23E-75 | 0.38004306 | 0.668 | 0.558 | 4.69E-71 | InN3 | MROH1 |
| SNCAIP | 1.24E-75 | 0.27070662 | 0.29 | 0.156 | 4.73E-71 | InN3 | SNCAIP |
| VWA81 | 1.30E-75 | 0.34541961 | 0.787 | 0.738 | 4.97E-71 | InN3 | VWA8 |
| SIK31 | 1.83E-75 | 0.30005991 | 0.887 | 0.859 | 7.00E-71 | InN3 | SIK3 |
| ATP1A31 | 2.14E-75 | 0.35349113 | 0.659 | 0.531 | 8.17E-71 | InN3 | ATP1A3 |
| GOT1 | 5.19E-75 | 0.32361802 | 0.441 | 0.3 | 1.99E-70 | InN3 | GOT1 |
| AOPEP | 5.89E-75 | 0.32731043 | 0.805 | 0.741 | 2.25E-70 | InN3 | AOPEP |
| GRK4 | 9.08E-75 | 0.36452944 | 0.566 | 0.432 | 3.47E-70 | InN3 | GRK4 |
| GALNT81 | 1.37E-74 | 0.29006751 | 0.45 | 0.294 | 5.24E-70 | InN3 | GALNT8 |
| MAN1A21 | 1.60E-74 | 0.38148372 | 0.8 | 0.75 | 6.12E-70 | InN3 | MAN1A2 |
| PCCA | 2.51E-74 | 0.34884891 | 0.76 | 0.693 | 9.59E-70 | InN3 | PCCA |
| PPM1E1 | 3.24E-74 | 0.36565614 | 0.816 | 0.758 | 1.24E-69 | InN3 | PPM1E |
| LINC01619 | 5.58E-74 | 0.25445948 | 0.273 | 0.144 | 2.13E-69 | InN3 | LINC01619 |
| IL1RAP2 | 8.56E-74 | 0.35489923 | 0.473 | 0.319 | 3.27E-69 | InN3 | IL1RAP |
| RNF1571 | 9.03E-74 | 0.33544797 | 0.643 | 0.523 | 3.45E-69 | InN3 | RNF157 |
| NLGN4X1 | 2.75E-73 | 0.37139492 | 0.788 | 0.726 | 1.05E-68 | InN3 | NLGN4X |
| PXDNL1 | 2.85E-73 | 0.34734095 | 0.361 | 0.222 | 1.09E-68 | InN3 | PXDNL |
| MFHAS1 | 5.91E-73 | 0.34946238 | 0.524 | 0.385 | 2.26E-68 | InN3 | MFHAS1 |
| SLC38A1 | 1.18E-72 | 0.32999612 | 0.697 | 0.593 | 4.51E-68 | InN3 | SLC38A1 |
| AUH | 2.70E-72 | 0.36421756 | 0.692 | 0.607 | 1.03E-67 | InN3 | AUH |
| TTC39B | 9.40E-72 | 0.3470719 | 0.582 | 0.455 | 3.59E-67 | InN3 | TTC39B |
| KDM4C | 1.05E-71 | 0.33071044 | 0.792 | 0.735 | 4.00E-67 | InN3 | KDM4C |

| | | | | | | | |
|---|---|---|---|---|---|---|---|
| MYO19 | 1.90E-71 | 0.29376566 | 0.389 | 0.251 | 7.26E-67 | InN3 | MYO19 |
| GARNL32 | 3.49E-71 | 0.2897381 | 0.828 | 0.72 | 1.33E-66 | InN3 | GARNL3 |
| BCAS4 | 3.49E-71 | 0.30126613 | 0.363 | 0.225 | 1.33E-66 | InN3 | BCAS4 |
| NMNAT21 | 4.07E-71 | 0.31455801 | 0.883 | 0.852 | 1.56E-66 | InN3 | NMNAT2 |
| P3H2 | 7.26E-71 | 0.36233698 | 0.484 | 0.343 | 2.77E-66 | InN3 | P3H2 |
| DNMBP | 1.08E-70 | 0.25115675 | 0.332 | 0.197 | 4.13E-66 | InN3 | DNMBP |
| HERC6 | 1.12E-70 | 0.28230432 | 0.376 | 0.234 | 4.26E-66 | InN3 | HERC6 |
| APBB21 | 1.15E-70 | 0.33733612 | 0.762 | 0.687 | 4.41E-66 | InN3 | APBB2 |
| CDS1 | 2.04E-69 | 0.26240265 | 0.394 | 0.253 | 7.79E-65 | InN3 | CDS1 |
| ATG5 | 3.10E-69 | 0.3433596 | 0.615 | 0.495 | 1.19E-64 | InN3 | ATG5 |
| KIF9-AS1 | 4.11E-69 | 0.40353376 | 0.697 | 0.594 | 1.57E-64 | InN3 | KIF9-AS1 |
| PPM1K1 | 4.66E-69 | 0.34834798 | 0.677 | 0.584 | 1.78E-64 | InN3 | PPM1K |
| LINC01473 | 5.40E-68 | 0.38188196 | 0.65 | 0.545 | 2.07E-63 | InN3 | LINC01473 |
| CACNG21 | 1.41E-67 | 0.3521489 | 0.733 | 0.653 | 5.39E-63 | InN3 | CACNG2 |
| AKAP1 | 2.06E-67 | 0.2524199 | 0.398 | 0.26 | 7.89E-63 | InN3 | AKAP1 |
| ETFDH | 7.74E-67 | 0.29962071 | 0.481 | 0.348 | 2.96E-62 | InN3 | ETFDH |
| SATB12 | 1.79E-66 | 0.26258452 | 0.479 | 0.333 | 6.84E-62 | InN3 | SATB1 |
| CHD6 | 2.47E-66 | 0.3022928 | 0.775 | 0.725 | 9.43E-62 | InN3 | CHD6 |
| PDGFC3 | 2.56E-66 | 0.33329017 | 0.414 | 0.276 | 9.78E-62 | InN3 | PDGFC |
| FAM20C | 5.52E-66 | 0.25128194 | 0.296 | 0.171 | 2.11E-61 | InN3 | FAM20C |
| RDX1 | 1.17E-65 | 0.28875041 | 0.54 | 0.398 | 4.47E-61 | InN3 | RDX |
| CDH72 | 3.07E-65 | 0.30289864 | 0.415 | 0.269 | 1.17E-60 | InN3 | CDH7 |
| FBN1 | 4.81E-65 | 0.35805305 | 0.721 | 0.642 | 1.84E-60 | InN3 | FBN1 |
| TPST2 | 4.89E-65 | 0.2956216 | 0.354 | 0.224 | 1.87E-60 | InN3 | TPST2 |
| PPP2R3A | 5.29E-65 | 0.3196384 | 0.442 | 0.309 | 2.02E-60 | InN3 | PPP2R3A |
| MGAT52 | 9.65E-65 | 0.3361911 | 0.732 | 0.658 | 3.69E-60 | InN3 | MGAT5 |
| PTPRS | 1.09E-64 | 0.32960984 | 0.772 | 0.711 | 4.16E-60 | InN3 | PTPRS |
| SFXN4 | 1.28E-64 | 0.26355039 | 0.325 | 0.198 | 4.91E-60 | InN3 | SFXN4 |
| CCDC146 | 1.43E-64 | 0.34939493 | 0.465 | 0.33 | 5.45E-60 | InN3 | CCDC146 |
| DENND5A1 | 1.70E-64 | 0.32432624 | 0.589 | 0.462 | 6.50E-60 | InN3 | DENND5A |
| ABCA101 | 4.82E-64 | 0.33590237 | 0.63 | 0.52 | 1.84E-59 | InN3 | ABCA10 |
| MRRF | 9.47E-64 | 0.25544923 | 0.357 | 0.229 | 3.62E-59 | InN3 | MRRF |
| NRG2 | 1.03E-63 | 0.31913046 | 0.705 | 0.614 | 3.94E-59 | InN3 | NRG2 |
| LGR4 | 1.62E-63 | 0.32405668 | 0.423 | 0.292 | 6.21E-59 | InN3 | LGR4 |
| OSBPL82 | 2.77E-63 | 0.28235093 | 0.809 | 0.749 | 1.06E-58 | InN3 | OSBPL8 |
| HSPA4L2 | 1.71E-62 | 0.30517776 | 0.594 | 0.473 | 6.52E-58 | InN3 | HSPA4L |
| HIVEP21 | 4.16E-62 | 0.30327173 | 0.837 | 0.798 | 1.59E-57 | InN3 | HIVEP2 |
| CDK62 | 4.42E-62 | 0.35339406 | 0.544 | 0.407 | 1.69E-57 | InN3 | CDK6 |
| DHX32 | 8.31E-62 | 0.28754169 | 0.408 | 0.276 | 3.18E-57 | InN3 | DHX32 |
| PIP5K1B1 | 9.55E-62 | 0.25239258 | 0.622 | 0.474 | 3.65E-57 | InN3 | PIP5K1B |
| FASTKD1 | 1.08E-61 | 0.32174254 | 0.557 | 0.443 | 4.13E-57 | InN3 | FASTKD1 |
| UBL3 | 1.64E-61 | 0.33279601 | 0.575 | 0.457 | 6.26E-57 | InN3 | UBL3 |
| ARFGEF2 | 4.11E-61 | 0.33086295 | 0.694 | 0.608 | 1.57E-56 | InN3 | ARFGEF2 |
| STK392 | 4.14E-61 | 0.34088639 | 0.686 | 0.597 | 1.58E-56 | InN3 | STK39 |
| TMEM652 | 1.43E-60 | 0.30887368 | 0.525 | 0.392 | 5.47E-56 | InN3 | TMEM65 |
| ABCA9 | 2.71E-60 | 0.29010073 | 0.409 | 0.279 | 1.04E-55 | InN3 | ABCA9 |
| FAM3C1 | 1.37E-59 | 0.3207292 | 0.533 | 0.408 | 5.22E-55 | InN3 | FAM3C |
| UBE2O | 1.40E-59 | 0.3474418 | 0.618 | 0.522 | 5.34E-55 | InN3 | UBE2O |
| FRYL | 1.98E-59 | 0.32166734 | 0.769 | 0.701 | 7.58E-55 | InN3 | FRYL |
| EPM2A | 2.00E-59 | 0.2828004 | 0.371 | 0.244 | 7.65E-55 | InN3 | EPM2A |
| CPEB3 | 2.39E-59 | 0.27860496 | 0.883 | 0.845 | 9.13E-55 | InN3 | CPEB3 |
| TRPM22 | 2.93E-59 | 0.30280073 | 0.53 | 0.41 | 1.12E-54 | InN3 | TRPM2 |
| PARVA1 | 3.62E-59 | 0.25563667 | 0.371 | 0.24 | 1.39E-54 | InN3 | PARVA |
| STAU2 | 4.78E-59 | 0.29385291 | 0.832 | 0.806 | 1.83E-54 | InN3 | STAU2 |
| VPS13B | 8.34E-59 | 0.29267626 | 0.848 | 0.816 | 3.19E-54 | InN3 | VPS13B |
| SRGAP23 | 1.05E-58 | 0.27280268 | 0.591 | 0.479 | 4.02E-54 | InN3 | SRGAP2 |
| LMBR1 | 1.40E-58 | 0.33665237 | 0.664 | 0.577 | 5.36E-54 | InN3 | LMBR1 |
| GTF2IRD11 | 1.83E-58 | 0.31073915 | 0.582 | 0.449 | 6.98E-54 | InN3 | GTF2IRD1 |
| TRAF3IP2-AS | 2.11E-58 | 0.29065759 | 0.804 | 0.747 | 8.08E-54 | InN3 | TRAF3IP2-AS |
| GAS61 | 5.49E-58 | 0.26805581 | 0.413 | 0.286 | 2.10E-53 | InN3 | GAS6 |
| ANXA6 | 6.37E-58 | 0.29705505 | 0.512 | 0.386 | 2.43E-53 | InN3 | ANXA6 |
| SIDT13 | 7.71E-58 | 0.30769861 | 0.645 | 0.513 | 2.95E-53 | InN3 | SIDT1 |
| GUCY1A1 | 1.01E-57 | 0.31902929 | 0.451 | 0.322 | 3.87E-53 | InN3 | GUCY1A1 |

| | | | | | | | |
|---|---|---|---|---|---|---|---|
| CC2D2A1 | 2.14E-57 | 0.25932457 | 0.389 | 0.263 | 8.18E-53 | InN3 | CC2D2A |
| JDP2 | 3.11E-57 | 0.26652485 | 0.369 | 0.244 | 1.19E-52 | InN3 | JDP2 |
| CCDC88A1 | 1.01E-56 | 0.26741493 | 0.882 | 0.844 | 3.87E-52 | InN3 | CCDC88A |
| CRADD1 | 1.75E-56 | 0.33212937 | 0.561 | 0.428 | 6.70E-52 | InN3 | CRADD |
| SYT141 | 2.90E-56 | 0.26869958 | 0.814 | 0.78 | 1.11E-51 | InN3 | SYT14 |
| PIK3R3 | 8.37E-56 | 0.26112019 | 0.449 | 0.312 | 3.20E-51 | InN3 | PIK3R3 |
| FARP13 | 2.25E-55 | 0.28396616 | 0.754 | 0.653 | 8.58E-51 | InN3 | FARP1 |
| COX10 | 1.89E-54 | 0.26949916 | 0.383 | 0.263 | 7.22E-50 | InN3 | COX10 |
| ZBTB1 | 1.93E-54 | 0.2797757 | 0.493 | 0.369 | 7.39E-50 | InN3 | ZBTB1 |
| NCK22 | 2.17E-54 | 0.28300048 | 0.564 | 0.439 | 8.29E-50 | InN3 | NCK2 |
| AC084198.2 | 2.48E-54 | 0.25816102 | 0.421 | 0.294 | 9.47E-50 | InN3 | AC084198.2 |
| EPHA72 | 3.04E-54 | 0.30330864 | 0.658 | 0.539 | 1.16E-49 | InN3 | EPHA7 |
| AC120193.14 | 3.85E-54 | 0.58269555 | 0.439 | 0.323 | 1.47E-49 | InN3 | AC120193.1 |
| WIPF32 | 3.08E-53 | 0.29665358 | 0.526 | 0.404 | 1.18E-48 | InN3 | WIPF3 |
| TRANK11 | 6.34E-53 | 0.31544012 | 0.676 | 0.592 | 2.42E-48 | InN3 | TRANK1 |
| CDH112 | 7.85E-53 | 0.44038565 | 0.548 | 0.449 | 3.00E-48 | InN3 | CDH11 |
| EML2 | 2.79E-52 | 0.29175847 | 0.507 | 0.389 | 1.07E-47 | InN3 | EML2 |
| DGKE1 | 4.51E-52 | 0.29638408 | 0.537 | 0.426 | 1.72E-47 | InN3 | DGKE |
| SFXN51 | 5.21E-52 | 0.30578874 | 0.624 | 0.526 | 1.99E-47 | InN3 | SFXN5 |
| HSPD1 | 6.84E-52 | 0.33180939 | 0.454 | 0.33 | 2.62E-47 | InN3 | HSPD1 |
| MPC1 | 1.12E-51 | 0.26070618 | 0.407 | 0.293 | 4.27E-47 | InN3 | MPC1 |
| SLC25A42 | 1.13E-51 | 0.25381839 | 0.393 | 0.27 | 4.33E-47 | InN3 | SLC25A42 |
| SLC25A121 | 1.23E-51 | 0.27042644 | 0.842 | 0.817 | 4.68E-47 | InN3 | SLC25A12 |
| CCDC148 | 1.29E-51 | 0.30887671 | 0.464 | 0.347 | 4.94E-47 | InN3 | CCDC148 |
| SENP71 | 2.01E-51 | 0.27716962 | 0.606 | 0.501 | 7.69E-47 | InN3 | SENP7 |
| NUBPL | 2.24E-51 | 0.30594751 | 0.564 | 0.452 | 8.58E-47 | InN3 | NUBPL |
| URI11 | 2.51E-51 | 0.27412815 | 0.574 | 0.463 | 9.60E-47 | InN3 | URI1 |
| UHRF1BP1 | 6.96E-51 | 0.27265454 | 0.436 | 0.315 | 2.66E-46 | InN3 | UHRF1BP1 |
| ERBIN2 | 7.08E-51 | 0.28885164 | 0.64 | 0.534 | 2.70E-46 | InN3 | ERBIN |
| SESTD1 | 1.08E-49 | 0.30110835 | 0.687 | 0.62 | 4.13E-45 | InN3 | SESTD1 |
| TUT41 | 1.26E-49 | 0.27882827 | 0.753 | 0.699 | 4.81E-45 | InN3 | TUT4 |
| DNAJC21 | 1.93E-49 | 0.26222006 | 0.4 | 0.284 | 7.40E-45 | InN3 | DNAJC21 |
| SPECC12 | 3.35E-49 | 0.27062286 | 0.796 | 0.733 | 1.28E-44 | InN3 | SPECC1 |
| COA1 | 8.57E-49 | 0.3071303 | 0.629 | 0.547 | 3.28E-44 | InN3 | COA1 |
| OSBPL92 | 7.09E-48 | 0.25241581 | 0.644 | 0.555 | 2.71E-43 | InN3 | OSBPL9 |
| RARB2 | 8.54E-48 | 0.29506652 | 0.555 | 0.425 | 3.26E-43 | InN3 | RARB |
| LPCAT4 | 1.13E-47 | 0.28113266 | 0.471 | 0.364 | 4.31E-43 | InN3 | LPCAT4 |
| CLVS21 | 1.31E-47 | 0.28738204 | 0.67 | 0.595 | 5.01E-43 | InN3 | CLVS2 |
| HERC3 | 1.89E-47 | 0.27717754 | 0.633 | 0.542 | 7.24E-43 | InN3 | HERC3 |
| PTPRJ3 | 2.26E-47 | 0.27170119 | 0.697 | 0.623 | 8.63E-43 | InN3 | PTPRJ |
| BTD | 3.54E-47 | 0.26908325 | 0.423 | 0.304 | 1.35E-42 | InN3 | BTD |
| INSR | 4.32E-47 | 0.288945 | 0.729 | 0.66 | 1.65E-42 | InN3 | INSR |
| MAGED1 | 1.37E-46 | 0.2820029 | 0.657 | 0.572 | 5.22E-42 | InN3 | MAGED1 |
| TUBGCP5 | 1.69E-46 | 0.25183511 | 0.409 | 0.294 | 6.47E-42 | InN3 | TUBGCP5 |
| SLC45A4 | 6.27E-45 | 0.26860051 | 0.43 | 0.318 | 2.40E-40 | InN3 | SLC45A4 |
| MTR | 1.07E-44 | 0.27494343 | 0.662 | 0.593 | 4.11E-40 | InN3 | MTR |
| PIP4K2A2 | 1.14E-44 | 0.31314508 | 0.531 | 0.422 | 4.34E-40 | InN3 | PIP4K2A |
| WDR60 | 1.36E-44 | 0.28482359 | 0.699 | 0.63 | 5.20E-40 | InN3 | WDR60 |
| AC007364.11 | 1.61E-44 | 0.28752644 | 0.391 | 0.277 | 6.14E-40 | InN3 | AC007364.1 |
| NT5DC1 | 7.43E-44 | 0.28777873 | 0.467 | 0.359 | 2.84E-39 | InN3 | NT5DC1 |
| C11orf581 | 1.38E-43 | 0.33187749 | 0.597 | 0.523 | 5.26E-39 | InN3 | C11orf58 |
| TBC1D81 | 1.99E-43 | 0.25059452 | 0.53 | 0.419 | 7.62E-39 | InN3 | TBC1D8 |
| CMC1 | 3.77E-43 | 0.26542987 | 0.454 | 0.348 | 1.44E-38 | InN3 | CMC1 |
| NRSN11 | 4.67E-43 | 0.29841445 | 0.584 | 0.485 | 1.78E-38 | InN3 | NRSN1 |
| PCGF52 | 5.58E-43 | 0.25865787 | 0.692 | 0.628 | 2.13E-38 | InN3 | PCGF5 |
| AC105383.1 | 7.73E-43 | 0.285691 | 0.424 | 0.319 | 2.95E-38 | InN3 | AC105383.1 |
| SH3KBP13 | 3.46E-42 | 0.27501086 | 0.683 | 0.603 | 1.32E-37 | InN3 | SH3KBP1 |
| PUS7L | 7.17E-42 | 0.26376226 | 0.556 | 0.462 | 2.74E-37 | InN3 | PUS7L |
| TMTC12 | 9.30E-42 | 0.25119657 | 0.679 | 0.557 | 3.56E-37 | InN3 | TMTC1 |
| AC046134.21 | 2.38E-41 | 0.30537274 | 0.633 | 0.555 | 9.08E-37 | InN3 | AC046134.2 |
| ACYP2 | 2.91E-41 | 0.2864627 | 0.764 | 0.72 | 1.11E-36 | InN3 | ACYP2 |
| SRRM3 | 3.19E-41 | 0.25307271 | 0.834 | 0.808 | 1.22E-36 | InN3 | SRRM3 |
| ST3GAL3 | 3.69E-41 | 0.26591058 | 0.628 | 0.548 | 1.41E-36 | InN3 | ST3GAL3 |

| | | | | | | | |
|---|---|---|---|---|---|---|---|
| BMPR1A | 2.61E-40 | 0.26350607 | 0.604 | 0.518 | 9.98E-36 | InN3 | BMPR1A |
| PNKD | 3.93E-40 | 0.26966994 | 0.449 | 0.35 | 1.50E-35 | InN3 | PNKD |
| USP45 | 4.49E-40 | 0.26818156 | 0.442 | 0.341 | 1.71E-35 | InN3 | USP45 |
| CHRD | 4.61E-40 | 0.26866486 | 0.386 | 0.288 | 1.76E-35 | InN3 | CHRD |
| MEF2C-AS1 | 5.64E-40 | 0.28550521 | 0.801 | 0.741 | 2.16E-35 | InN3 | MEF2C-AS1 |
| RALGAPA21 | 1.06E-39 | 0.25310871 | 0.66 | 0.588 | 4.06E-35 | InN3 | RALGAPA2 |
| TARSL2 | 1.43E-39 | 0.27597285 | 0.562 | 0.478 | 5.46E-35 | InN3 | TARSL2 |
| TAFA51 | 8.33E-39 | 0.30906995 | 0.672 | 0.603 | 3.18E-34 | InN3 | TAFA5 |
| ANKRD261 | 1.10E-38 | 0.27654369 | 0.787 | 0.752 | 4.19E-34 | InN3 | ANKRD26 |
| STX18-AS1 | 1.22E-38 | 0.29416224 | 0.667 | 0.591 | 4.67E-34 | InN3 | STX18-AS1 |
| LINC01505 | 1.38E-37 | 0.31047012 | 0.313 | 0.214 | 5.29E-33 | InN3 | LINC01505 |
| TTC21B | 3.27E-37 | 0.25995575 | 0.658 | 0.599 | 1.25E-32 | InN3 | TTC21B |
| SERPINB91 | 2.97E-36 | 0.44481209 | 0.271 | 0.177 | 1.14E-31 | InN3 | SERPINB9 |
| U91319.1 | 1.00E-35 | 0.25324537 | 0.537 | 0.432 | 3.84E-31 | InN3 | U91319.1 |
| GDPD1 | 1.06E-35 | 0.26559971 | 0.528 | 0.439 | 4.03E-31 | InN3 | GDPD1 |
| DZIP11 | 1.83E-35 | 0.26289943 | 0.673 | 0.613 | 6.98E-31 | InN3 | DZIP1 |
| ARHGAP62 | 2.27E-35 | 0.36767991 | 0.255 | 0.172 | 8.66E-31 | InN3 | ARHGAP6 |
| CDC14B | 3.78E-35 | 0.25155616 | 0.575 | 0.489 | 1.45E-30 | InN3 | CDC14B |
| CNNM2 | 9.43E-35 | 0.25050203 | 0.732 | 0.692 | 3.60E-30 | InN3 | CNNM2 |
| ZSWIM51 | 3.55E-33 | 0.25680833 | 0.617 | 0.542 | 1.36E-28 | InN3 | ZSWIM5 |
| SVIL1 | 3.62E-33 | 0.39362733 | 0.25 | 0.172 | 1.38E-28 | InN3 | SVIL |
| AC073332.11 | 7.77E-33 | 0.3081209 | 0.423 | 0.324 | 2.97E-28 | InN3 | AC073332.1 |
| DENND2A2 | 1.18E-32 | 0.26578513 | 0.362 | 0.264 | 4.50E-28 | InN3 | DENND2A |
| MAP1A | 1.39E-32 | 0.26565462 | 0.542 | 0.461 | 5.33E-28 | InN3 | MAP1A |
| KCNA21 | 4.45E-28 | 0.2938395 | 0.431 | 0.339 | 1.70E-23 | InN3 | KCNA2 |
| KCNN21 | 1.17E-27 | 0.25043774 | 0.572 | 0.492 | 4.47E-23 | InN3 | KCNN2 |
| ZNF804B3 | 4.13E-24 | 0.41497198 | 0.432 | 0.356 | 1.58E-19 | InN3 | ZNF804B |
| PLCG22 | 1.01E-19 | 0.35050063 | 0.704 | 0.649 | 3.85E-15 | InN3 | PLCG2 |
| KLHL29 | 2.27E-12 | 0.29385405 | 0.694 | 0.702 | 8.68E-08 | InN3 | KLHL29 |
| PCDH11X2 | 9.06E-11 | 0.25772381 | 0.541 | 0.54 | 3.46E-06 | InN3 | PCDH11X |
| GPC53 | 9.94E-09 | 0.7506003 | 0.441 | 0.424 | 0.00037978 | InN3 | GPC5 |
| GALNTL62 | 2.55E-06 | 0.26321584 | 0.692 | 0.748 | 0.09748931 | InN3 | GALNTL6 |
| MT-CO21 | 0.00051013 | 0.30757033 | 0.884 | 0.9 | 1 | InN3 | MT-CO2 |
| FBXL7 | 0 | 3.56951733 | 0.957 | 0.13 | 0 | InN4 | FBXL7 |
| FGF131 | 0 | 3.3795477 | 0.968 | 0.388 | 0 | InN4 | FGF13 |
| AC132803.1 | 0 | 3.19666093 | 0.842 | 0.143 | 0 | InN4 | AC132803.1 |
| PRELID2 | 0 | 3.1664132 | 0.919 | 0.385 | 0 | InN4 | PRELID2 |
| NTNG11 | 0 | 2.73084873 | 0.917 | 0.253 | 0 | InN4 | NTNG1 |
| ALK1 | 0 | 2.66681684 | 0.933 | 0.211 | 0 | InN4 | ALK |
| EYA4 | 0 | 2.54684887 | 0.834 | 0.063 | 0 | InN4 | EYA4 |
| KIT | 0 | 2.52283133 | 0.841 | 0.06 | 0 | InN4 | KIT |
| ADARB21 | 0 | 2.49755819 | 0.984 | 0.338 | 0 | InN4 | ADARB2 |
| SGCZ4 | 0 | 2.41536822 | 0.984 | 0.489 | 0 | InN4 | SGCZ |
| MYO161 | 0 | 2.38195366 | 0.924 | 0.253 | 0 | InN4 | MYO16 |
| SLC6A14 | 0 | 2.35939229 | 0.947 | 0.324 | 0 | InN4 | SLC6A1 |
| SOX2-OT1 | 0 | 2.25438065 | 0.898 | 0.47 | 0 | InN4 | SOX2-OT |
| PTCHD42 | 0 | 2.24172303 | 0.914 | 0.374 | 0 | InN4 | PTCHD4 |
| KCNIP12 | 0 | 2.20329458 | 0.931 | 0.273 | 0 | InN4 | KCNIP1 |
| RBMS32 | 0 | 2.14276195 | 0.976 | 0.623 | 0 | InN4 | RBMS3 |
| GRIK11 | 0 | 2.05964659 | 0.969 | 0.362 | 0 | InN4 | GRIK1 |
| UNC13C1 | 0 | 2.04528574 | 0.937 | 0.562 | 0 | InN4 | UNC13C |
| ZNF5362 | 0 | 2.02778916 | 0.958 | 0.344 | 0 | InN4 | ZNF536 |
| NXPH12 | 0 | 2.0028902 | 0.96 | 0.233 | 0 | InN4 | NXPH1 |
| LUZP22 | 0 | 1.94901134 | 0.869 | 0.281 | 0 | InN4 | LUZP2 |
| FSTL51 | 0 | 1.92384536 | 0.947 | 0.478 | 0 | InN4 | FSTL5 |
| GRIP12 | 0 | 1.91481579 | 0.99 | 0.79 | 0 | InN4 | GRIP1 |
| DNER2 | 0 | 1.89384259 | 0.928 | 0.508 | 0 | InN4 | DNER |
| GAD22 | 0 | 1.87647337 | 0.853 | 0.228 | 0 | InN4 | GAD2 |
| UBASH3B3 | 0 | 1.8390328 | 0.852 | 0.314 | 0 | InN4 | UBASH3B |
| EGFR1 | 0 | 1.82685279 | 0.891 | 0.208 | 0 | InN4 | EGFR |
| SV2C | 0 | 1.7784791 | 0.701 | 0.052 | 0 | InN4 | SV2C |
| PTPRT4 | 0 | 1.77361166 | 0.976 | 0.67 | 0 | InN4 | PTPRT |
| NKAIN31 | 0 | 1.75653193 | 0.704 | 0.255 | 0 | InN4 | NKAIN3 |

| | | | | | | | |
|---|---|---|---|---|---|---|---|
| AC137770.1 | 0 | 1.75518131 | 0.656 | 0.082 | 0 | InN4 | AC137770.1 |
| SPHKAP2 | 0 | 1.71892547 | 0.729 | 0.134 | 0 | InN4 | SPHKAP |
| PID11 | 0 | 1.70450796 | 0.856 | 0.48 | 0 | InN4 | PID1 |
| CDH131 | 0 | 1.70448198 | 0.866 | 0.634 | 0 | InN4 | CDH13 |
| RAB3C2 | 0 | 1.69841606 | 0.91 | 0.625 | 0 | InN4 | RAB3C |
| PDGFD1 | 0 | 1.69439852 | 0.54 | 0.176 | 0 | InN4 | PDGFD |
| GRIN3A1 | 0 | 1.67106259 | 0.789 | 0.208 | 0 | InN4 | GRIN3A |
| CHST9 | 0 | 1.6596597 | 0.321 | 0.054 | 0 | InN4 | CHST9 |
| NFIB1 | 0 | 1.64689746 | 0.952 | 0.634 | 0 | InN4 | NFIB |
| ADAMTSL31 | 0 | 1.61765174 | 0.749 | 0.267 | 0 | InN4 | ADAMTSL3 |
| TOX23 | 0 | 1.59821329 | 0.772 | 0.175 | 0 | InN4 | TOX2 |
| WWP11 | 0 | 1.58443578 | 0.834 | 0.467 | 0 | InN4 | WWP1 |
| ADAMTS9-AS | 0 | 1.58043868 | 0.872 | 0.318 | 0 | InN4 | ADAMTS9-AS |
| TACR11 | 0 | 1.57232497 | 0.641 | 0.065 | 0 | InN4 | TACR1 |
| KAZN3 | 0 | 1.55554216 | 0.997 | 0.951 | 0 | InN4 | KAZN |
| TMTC21 | 0 | 1.53559215 | 0.884 | 0.523 | 0 | InN4 | TMTC2 |
| PKP21 | 0 | 1.52289053 | 0.66 | 0.078 | 0 | InN4 | PKP2 |
| KIAA12113 | 0 | 1.51653163 | 0.871 | 0.376 | 0 | InN4 | KIAA1211 |
| LINC00298 | 0 | 1.51649041 | 0.639 | 0.069 | 0 | InN4 | LINC00298 |
| MGAT4C1 | 0 | 1.50770285 | 0.934 | 0.561 | 0 | InN4 | MGAT4C |
| DGKD2 | 0 | 1.50237432 | 0.771 | 0.289 | 0 | InN4 | DGKD |
| DOCK102 | 0 | 1.48528568 | 0.881 | 0.381 | 0 | InN4 | DOCK10 |
| NXPH22 | 0 | 1.44793525 | 0.621 | 0.085 | 0 | InN4 | NXPH2 |
| PMEPA1 | 0 | 1.44599586 | 0.646 | 0.144 | 0 | InN4 | PMEPA1 |
| BCL11B3 | 0 | 1.40896511 | 0.768 | 0.23 | 0 | InN4 | BCL11B |
| PTPRM2 | 0 | 1.40323171 | 0.924 | 0.566 | 0 | InN4 | PTPRM |
| GRIN2A | 0 | 1.39647878 | 0.983 | 0.922 | 0 | InN4 | GRIN2A |
| SGK11 | 0 | 1.38937915 | 0.838 | 0.49 | 0 | InN4 | SGK1 |
| RAB3IP2 | 0 | 1.37093024 | 0.722 | 0.252 | 0 | InN4 | RAB3IP |
| NRIP32 | 0 | 1.37023106 | 0.687 | 0.194 | 0 | InN4 | NRIP3 |
| CDHR32 | 0 | 1.35442322 | 0.752 | 0.36 | 0 | InN4 | CDHR3 |
| UNC5D3 | 0 | 1.34981073 | 0.959 | 0.825 | 0 | InN4 | UNC5D |
| DSCAML13 | 0 | 1.34979473 | 0.957 | 0.749 | 0 | InN4 | DSCAML1 |
| TMEM132D3 | 0 | 1.34962624 | 0.976 | 0.773 | 0 | InN4 | TMEM132D |
| UNC5C1 | 0 | 1.34597945 | 0.95 | 0.776 | 0 | InN4 | UNC5C |
| IGSF11 | 0 | 1.33351167 | 0.793 | 0.498 | 0 | InN4 | IGSF11 |
| NFIA4 | 0 | 1.32523616 | 0.759 | 0.404 | 0 | InN4 | NFIA |
| MTSS14 | 0 | 1.32469955 | 0.886 | 0.553 | 0 | InN4 | MTSS1 |
| IL1RAPL11 | 0 | 1.31572811 | 0.996 | 0.974 | 0 | InN4 | IL1RAPL1 |
| PIP5K1B2 | 0 | 1.30521786 | 0.815 | 0.463 | 0 | InN4 | PIP5K1B |
| QKI4 | 0 | 1.30347045 | 0.918 | 0.661 | 0 | InN4 | QKI |
| CHST112 | 0 | 1.30195757 | 0.848 | 0.497 | 0 | InN4 | CHST11 |
| SLC35F42 | 0 | 1.3000503 | 0.686 | 0.211 | 0 | InN4 | SLC35F4 |
| CTXND1 | 0 | 1.29805725 | 0.549 | 0.044 | 0 | InN4 | CTXND1 |
| GRK31 | 0 | 1.29422672 | 0.907 | 0.697 | 0 | InN4 | GRK3 |
| RPH3A1 | 0 | 1.29105967 | 0.837 | 0.492 | 0 | InN4 | RPH3A |
| SOX1-OT | 0 | 1.29059141 | 0.619 | 0.093 | 0 | InN4 | SOX1-OT |
| FAM110B2 | 0 | 1.28926229 | 0.781 | 0.415 | 0 | InN4 | FAM110B |
| NYAP21 | 0 | 1.28268967 | 0.816 | 0.493 | 0 | InN4 | NYAP2 |
| MSI23 | 0 | 1.27249747 | 0.884 | 0.649 | 0 | InN4 | MSI2 |
| CNTNAP42 | 0 | 1.27121311 | 0.906 | 0.567 | 0 | InN4 | CNTNAP4 |
| ROR2 | 0 | 1.26439028 | 0.506 | 0.038 | 0 | InN4 | ROR2 |
| EEPD1 | 0 | 1.26136383 | 0.758 | 0.365 | 0 | InN4 | EEPD1 |
| LTBP1 | 0 | 1.23921964 | 0.518 | 0.104 | 0 | InN4 | LTBP1 |
| ATP11C1 | 0 | 1.23401725 | 0.577 | 0.048 | 0 | InN4 | ATP11C |
| MACROD21 | 0 | 1.20899936 | 0.983 | 0.949 | 0 | InN4 | MACROD2 |
| ATP8A21 | 0 | 1.19638772 | 0.983 | 0.95 | 0 | InN4 | ATP8A2 |
| ARHGAP311 | 0 | 1.19220108 | 0.499 | 0.024 | 0 | InN4 | ARHGAP31 |
| LINC00472 | 0 | 1.17225153 | 0.537 | 0.129 | 0 | InN4 | LINC00472 |
| LINC01344 | 0 | 1.17116703 | 0.375 | 0.015 | 0 | InN4 | LINC01344 |
| GRIA42 | 0 | 1.16427171 | 0.965 | 0.839 | 0 | InN4 | GRIA4 |
| EXT11 | 0 | 1.16195665 | 0.933 | 0.806 | 0 | InN4 | EXT1 |
| GOLIM42 | 0 | 1.16154072 | 0.603 | 0.141 | 0 | InN4 | GOLIM4 |

| | | | | | | | |
|---|---|---|---|---|---|---|---|
| KIRREL1 | 0 | 1.15913564 | 0.571 | 0.064 | 0 | InN4 | KIRREL1 |
| CACNA2D11 | 0 | 1.15597458 | 0.933 | 0.767 | 0 | InN4 | CACNA2D1 |
| SH3RF33 | 0 | 1.13749623 | 0.879 | 0.671 | 0 | InN4 | SH3RF3 |
| RERG1 | 0 | 1.12423163 | 0.698 | 0.247 | 0 | InN4 | RERG |
| ZBTB203 | 0 | 1.1179554 | 0.807 | 0.537 | 0 | InN4 | ZBTB20 |
| HPSE2 | 0 | 1.09662909 | 0.347 | 0.099 | 0 | InN4 | HPSE2 |
| SPOCK13 | 0 | 1.09300537 | 0.946 | 0.74 | 0 | InN4 | SPOCK1 |
| TOX33 | 0 | 1.0898281 | 0.606 | 0.168 | 0 | InN4 | TOX3 |
| COL4A21 | 0 | 1.08903736 | 0.609 | 0.228 | 0 | InN4 | COL4A2 |
| AP001347.1 | 0 | 1.08607075 | 0.789 | 0.591 | 0 | InN4 | AP001347.1 |
| TIAM13 | 0 | 1.08585081 | 0.922 | 0.746 | 0 | InN4 | TIAM1 |
| GRIP23 | 0 | 1.08246308 | 0.633 | 0.167 | 0 | InN4 | GRIP2 |
| KIRREL31 | 0 | 1.07265323 | 0.965 | 0.784 | 0 | InN4 | KIRREL3 |
| KCNC22 | 0 | 1.0699595 | 0.942 | 0.83 | 0 | InN4 | KCNC2 |
| SLC35F15 | 0 | 1.06765574 | 0.938 | 0.698 | 0 | InN4 | SLC35F1 |
| FGF14 | 0 | 1.06742311 | 0.994 | 0.989 | 0 | InN4 | FGF14 |
| TOX4 | 0 | 1.0644621 | 0.786 | 0.426 | 0 | InN4 | TOX |
| TRPC3 | 0 | 1.06196646 | 0.463 | 0.052 | 0 | InN4 | TRPC3 |
| LSAMP2 | 0 | 1.05892211 | 0.996 | 0.973 | 0 | InN4 | LSAMP |
| VWC22 | 0 | 1.05239987 | 0.739 | 0.337 | 0 | InN4 | VWC2 |
| PREX21 | 0 | 1.05189261 | 0.52 | 0.161 | 0 | InN4 | PREX2 |
| FREM11 | 0 | 1.05032149 | 0.505 | 0.208 | 0 | InN4 | FREM1 |
| ZEB24 | 0 | 1.04145447 | 0.933 | 0.816 | 0 | InN4 | ZEB2 |
| RIN23 | 0 | 1.02713691 | 0.619 | 0.19 | 0 | InN4 | RIN2 |
| COL25A11 | 0 | 1.02594842 | 0.495 | 0.202 | 0 | InN4 | COL25A1 |
| CDK141 | 0 | 1.01801902 | 0.936 | 0.861 | 0 | InN4 | CDK14 |
| MTUS2 | 0 | 1.00756955 | 0.979 | 0.923 | 0 | InN4 | MTUS2 |
| DOCK9 | 0 | 0.99896745 | 0.889 | 0.745 | 0 | InN4 | DOCK9 |
| ADRA1A2 | 0 | 0.9988358 | 0.655 | 0.19 | 0 | InN4 | ADRA1A |
| GRIK42 | 0 | 0.99120961 | 0.742 | 0.43 | 0 | InN4 | GRIK4 |
| MPPED11 | 0 | 0.98354681 | 0.736 | 0.438 | 0 | InN4 | MPPED1 |
| GNG41 | 0 | 0.98168525 | 0.534 | 0.126 | 0 | InN4 | GNG4 |
| HOMER2 | 0 | 0.97904626 | 0.635 | 0.329 | 0 | InN4 | HOMER2 |
| FRY | 0 | 0.97282861 | 0.927 | 0.848 | 0 | InN4 | FRY |
| UTRN3 | 0 | 0.97090467 | 0.587 | 0.238 | 0 | InN4 | UTRN |
| AC079296.12 | 0 | 0.96179684 | 0.544 | 0.166 | 0 | InN4 | AC079296.1 |
| KIAA1324L1 | 0 | 0.95690572 | 0.715 | 0.488 | 0 | InN4 | KIAA1324L |
| PDGFRA | 0 | 0.95343571 | 0.42 | 0.026 | 0 | InN4 | PDGFRA |
| PRKCA3 | 0 | 0.94989954 | 0.91 | 0.727 | 0 | InN4 | PRKCA |
| CA2 | 0 | 0.93688383 | 0.434 | 0.036 | 0 | InN4 | CA2 |
| LYPD62 | 0 | 0.92978784 | 0.509 | 0.104 | 0 | InN4 | LYPD6 |
| PITPNC13 | 0 | 0.92659707 | 0.798 | 0.479 | 0 | InN4 | PITPNC1 |
| ERBB42 | 0 | 0.92010368 | 0.992 | 0.555 | 0 | InN4 | ERBB4 |
| SYN23 | 0 | 0.91982045 | 0.945 | 0.898 | 0 | InN4 | SYN2 |
| DLX6-AS11 | 0 | 0.91857024 | 0.676 | 0.219 | 0 | InN4 | DLX6-AS1 |
| FOXO1 | 0 | 0.91558809 | 0.464 | 0.111 | 0 | InN4 | FOXO1 |
| SOX13 | 0 | 0.91196393 | 0.442 | 0.029 | 0 | InN4 | SOX13 |
| ITGB51 | 0 | 0.90936105 | 0.435 | 0.04 | 0 | InN4 | ITGB5 |
| ZNF4622 | 0 | 0.90709372 | 0.613 | 0.272 | 0 | InN4 | ZNF462 |
| ZNF385D4 | 0 | 0.90652921 | 0.853 | 0.498 | 0 | InN4 | ZNF385D |
| ENTPD32 | 0 | 0.89038034 | 0.611 | 0.28 | 0 | InN4 | ENTPD3 |
| WWOX3 | 0 | 0.8894019 | 0.914 | 0.813 | 0 | InN4 | WWOX |
| SLC25A481 | 0 | 0.88495807 | 0.414 | 0.109 | 0 | InN4 | SLC25A48 |
| CRTAC13 | 0 | 0.88015635 | 0.583 | 0.221 | 0 | InN4 | CRTAC1 |
| MAF4 | 0 | 0.87865576 | 0.583 | 0.166 | 0 | InN4 | MAF |
| AP1S21 | 0 | 0.87548582 | 0.553 | 0.185 | 0 | InN4 | AP1S2 |
| HAPLN1 | 0 | 0.87002464 | 0.405 | 0.025 | 0 | InN4 | HAPLN1 |
| COBL3 | 0 | 0.85218621 | 0.898 | 0.743 | 0 | InN4 | COBL |
| PRKCE1 | 0 | 0.84612413 | 0.96 | 0.911 | 0 | InN4 | PRKCE |
| PDE8B1 | 0 | 0.82943473 | 0.876 | 0.653 | 0 | InN4 | PDE8B |
| NAV22 | 0 | 0.82815489 | 0.962 | 0.919 | 0 | InN4 | NAV2 |
| MARCH32 | 0 | 0.82450809 | 0.422 | 0.133 | 0 | InN4 | 3-Mar |
| SCN1A2 | 0 | 0.81545759 | 0.916 | 0.84 | 0 | InN4 | SCN1A |

| | | | | | | | |
|---|---|---|---|---|---|---|---|
| NTM1 | 0 | 0.80252228 | 0.942 | 0.846 | 0 | InN4 | NTM |
| LINC00354 | 0 | 0.78589169 | 0.389 | 0.043 | 0 | InN4 | LINC00354 |
| NTRK22 | 0 | 0.78174013 | 0.971 | 0.943 | 0 | InN4 | NTRK2 |
| TMEM132B | 0 | 0.78172581 | 0.962 | 0.918 | 0 | InN4 | TMEM132B |
| LAMP51 | 0 | 0.76713127 | 0.47 | 0.161 | 0 | InN4 | LAMP5 |
| CA81 | 0 | 0.76634239 | 0.401 | 0.102 | 0 | InN4 | CA8 |
| LINC01237 | 0 | 0.76187274 | 0.456 | 0.148 | 0 | InN4 | LINC01237 |
| DPP61 | 0 | 0.75990743 | 0.982 | 0.915 | 0 | InN4 | DPP6 |
| DAB13 | 0 | 0.758111 | 0.976 | 0.874 | 0 | InN4 | DAB1 |
| PCBP31 | 0 | 0.75630479 | 0.463 | 0.169 | 0 | InN4 | PCBP3 |
| PARD32 | 0 | 0.75054793 | 0.694 | 0.302 | 0 | InN4 | PARD3 |
| GAD13 | 0 | 0.74601262 | 0.675 | 0.269 | 0 | InN4 | GAD1 |
| TENM43 | 0 | 0.73934728 | 0.957 | 0.854 | 0 | InN4 | TENM4 |
| GRIK21 | 0 | 0.72062953 | 0.978 | 0.947 | 0 | InN4 | GRIK2 |
| GPRC5B1 | 0 | 0.71833134 | 0.476 | 0.156 | 0 | InN4 | GPRC5B |
| TPD52L1 | 0 | 0.71039652 | 0.428 | 0.135 | 0 | InN4 | TPD52L1 |
| PRSS12 | 0 | 0.70837837 | 0.376 | 0.067 | 0 | InN4 | PRSS12 |
| AC023095.1 | 0 | 0.70680959 | 0.32 | 0.062 | 0 | InN4 | AC023095.1 |
| GRIA12 | 0 | 0.70186437 | 0.954 | 0.921 | 0 | InN4 | GRIA1 |
| WLS2 | 0 | 0.69605456 | 0.602 | 0.26 | 0 | InN4 | WLS |
| IGF13 | 0 | 0.68258959 | 0.47 | 0.136 | 0 | InN4 | IGF1 |
| TCF43 | 0 | 0.67650738 | 0.974 | 0.957 | 0 | InN4 | TCF4 |
| CHRDL1 | 0 | 0.66853579 | 0.346 | 0.055 | 0 | InN4 | CHRDL1 |
| RUNX22 | 0 | 0.65955153 | 0.49 | 0.146 | 0 | InN4 | RUNX2 |
| FAT1 | 0 | 0.65765918 | 0.388 | 0.074 | 0 | InN4 | FAT1 |
| ARID1B | 0 | 0.62990185 | 0.94 | 0.918 | 0 | InN4 | ARID1B |
| RGS71 | 0 | 0.61189297 | 0.982 | 0.956 | 0 | InN4 | RGS7 |
| TNFAIP8L3 | 0 | 0.58523266 | 0.336 | 0.07 | 0 | InN4 | TNFAIP8L3 |
| FRMPD1 | 0 | 0.58068274 | 0.326 | 0.056 | 0 | InN4 | FRMPD1 |
| MYRFL | 0 | 0.56980248 | 0.26 | 0.046 | 0 | InN4 | MYRFL |
| CPLX3 | 0 | 0.55615632 | 0.253 | 0.017 | 0 | InN4 | CPLX3 |
| COL18A1 | 0 | 0.54553813 | 0.34 | 0.101 | 0 | InN4 | COL18A1 |
| HEG1 | 0 | 0.53679582 | 0.323 | 0.061 | 0 | InN4 | HEG1 |
| CLIC4 | 0 | 0.53521851 | 0.284 | 0.042 | 0 | InN4 | CLIC4 |
| MDGA1 | 0 | 0.5178441 | 0.287 | 0.065 | 0 | InN4 | MDGA1 |
| SFRP1 | 0 | 0.51502289 | 0.266 | 0.029 | 0 | InN4 | SFRP1 |
| SLC7A2 | 0 | 0.50652633 | 0.298 | 0.072 | 0 | InN4 | SLC7A2 |
| SOX1 | 0 | 0.48411927 | 0.276 | 0.034 | 0 | InN4 | SOX1 |
| SOX2 | 0 | 0.44400836 | 0.307 | 0.08 | 0 | InN4 | SOX2 |
| ANO11 | 0 | 0.4137076 | 0.263 | 0.06 | 0 | InN4 | ANO1 |
| PLCB11 | 2.00E-307 | 0.68125865 | 0.983 | 0.978 | 7.63E-303 | InN4 | PLCB1 |
| GABRB21 | 4.32E-299 | 0.71806957 | 0.947 | 0.921 | 1.65E-294 | InN4 | GABRB2 |
| ASTN23 | 2.39E-297 | 0.68017811 | 0.91 | 0.729 | 9.13E-293 | InN4 | ASTN2 |
| SEMA6D2 | 6.79E-289 | 1.13871224 | 0.796 | 0.574 | 2.60E-284 | InN4 | SEMA6D |
| COL4A11 | 1.66E-287 | 0.67598129 | 0.41 | 0.154 | 6.35E-283 | InN4 | COL4A1 |
| CA3 | 2.62E-285 | 1.04280711 | 0.453 | 0.196 | 1.00E-280 | InN4 | CA3 |
| PDZRN33 | 3.39E-284 | 0.82783311 | 0.904 | 0.756 | 1.30E-279 | InN4 | PDZRN3 |
| B3GALT2 | 1.38E-282 | 0.60528218 | 0.379 | 0.134 | 5.27E-278 | InN4 | B3GALT2 |
| JAM2 | 6.89E-282 | 0.67569581 | 0.413 | 0.156 | 2.64E-277 | InN4 | JAM2 |
| SH3GL33 | 9.30E-282 | 0.72779847 | 0.884 | 0.818 | 3.55E-277 | InN4 | SH3GL3 |
| RNF150 | 5.24E-281 | 0.85238932 | 0.805 | 0.653 | 2.00E-276 | InN4 | RNF150 |
| RORA2 | 1.79E-280 | 0.57876561 | 0.989 | 0.962 | 6.83E-276 | InN4 | RORA |
| ZNF6082 | 3.30E-279 | 0.75876545 | 0.644 | 0.344 | 1.26E-274 | InN4 | ZNF608 |
| DDR2 | 1.84E-275 | 0.48203988 | 0.281 | 0.075 | 7.05E-271 | InN4 | DDR2 |
| RIMBP2 | 2.55E-274 | 0.78765407 | 0.843 | 0.755 | 9.73E-270 | InN4 | RIMBP2 |
| PLCXD32 | 7.08E-274 | 0.98379644 | 0.599 | 0.298 | 2.71E-269 | InN4 | PLCXD3 |
| CCSER12 | 5.61E-273 | 0.55447878 | 0.977 | 0.964 | 2.15E-268 | InN4 | CCSER1 |
| GNG12 | 4.87E-269 | 0.42146544 | 0.26 | 0.065 | 1.86E-264 | InN4 | GNG12 |
| CXCL141 | 5.36E-267 | 0.45310956 | 0.415 | 0.145 | 2.05E-262 | InN4 | CXCL14 |
| LRRC71 | 2.71E-265 | 0.54838756 | 0.978 | 0.965 | 1.03E-260 | InN4 | LRRC7 |
| TMEM132C1 | 1.54E-264 | 0.58727916 | 0.565 | 0.249 | 5.88E-260 | InN4 | TMEM132C |
| AC092691.14 | 1.08E-263 | 0.67820558 | 0.952 | 0.865 | 4.13E-259 | InN4 | AC092691.1 |
| TCF124 | 5.37E-263 | 0.86901449 | 0.826 | 0.7 | 2.05E-258 | InN4 | TCF12 |

| | | | | | | | |
|---|---|---|---|---|---|---|---|
| PROX11 | 1.00E-262 | 0.46705994 | 0.454 | 0.16 | 3.83E-258 | InN4 | PROX1 |
| ANKH1 | 8.56E-262 | 0.74713559 | 0.862 | 0.784 | 3.27E-257 | InN4 | ANKH |
| XKR42 | 2.32E-260 | 0.61400947 | 0.934 | 0.858 | 8.85E-256 | InN4 | XKR4 |
| GRID13 | 2.05E-258 | 0.69148889 | 0.911 | 0.849 | 7.83E-254 | InN4 | GRID1 |
| ADD31 | 1.74E-254 | 0.79458415 | 0.75 | 0.573 | 6.67E-250 | InN4 | ADD3 |
| C8orf341 | 1.92E-254 | 0.96706455 | 0.665 | 0.378 | 7.35E-250 | InN4 | C8orf34 |
| AC005009.2 | 2.18E-254 | 0.52970168 | 0.347 | 0.12 | 8.35E-250 | InN4 | AC005009.2 |
| VAV23 | 5.91E-254 | 0.53065285 | 0.444 | 0.172 | 2.26E-249 | InN4 | VAV2 |
| ANKRD55 | 7.25E-253 | 0.56232842 | 0.335 | 0.111 | 2.77E-248 | InN4 | ANKRD55 |
| ANKRD443 | 4.61E-250 | 0.72724461 | 0.679 | 0.416 | 1.76E-245 | InN4 | ANKRD44 |
| RPS6KA23 | 6.15E-248 | 0.58427418 | 0.937 | 0.853 | 2.35E-243 | InN4 | RPS6KA2 |
| BACH14 | 9.86E-243 | 0.67424664 | 0.492 | 0.222 | 3.77E-238 | InN4 | BACH1 |
| RIMS11 | 1.69E-241 | 0.60845053 | 0.927 | 0.891 | 6.48E-237 | InN4 | RIMS1 |
| GPR1581 | 1.98E-240 | 0.7055082 | 0.85 | 0.746 | 7.55E-236 | InN4 | GPR158 |
| ABI1 | 2.85E-240 | 0.85251574 | 0.721 | 0.57 | 1.09E-235 | InN4 | ABI1 |
| PARD3B | 3.63E-240 | 0.72624452 | 0.325 | 0.11 | 1.39E-235 | InN4 | PARD3B |
| VLDLR-AS1 | 4.39E-240 | 0.79286764 | 0.596 | 0.343 | 1.68E-235 | InN4 | VLDLR-AS1 |
| AKAP6 | 1.16E-238 | 0.52316963 | 0.952 | 0.944 | 4.42E-234 | InN4 | AKAP6 |
| RASSF82 | 1.32E-237 | 0.62442057 | 0.413 | 0.166 | 5.04E-233 | InN4 | RASSF8 |
| LRRC3B1 | 1.15E-236 | 0.64243971 | 0.521 | 0.242 | 4.38E-232 | InN4 | LRRC3B |
| ARL4C2 | 8.21E-236 | 0.68451713 | 0.53 | 0.258 | 3.14E-231 | InN4 | ARL4C |
| CELF42 | 2.98E-235 | 0.57304944 | 0.947 | 0.905 | 1.14E-230 | InN4 | CELF4 |
| LRIG11 | 6.46E-233 | 0.60144093 | 0.458 | 0.205 | 2.47E-228 | InN4 | LRIG1 |
| SLC24A32 | 8.68E-233 | 0.413999 | 0.647 | 0.296 | 3.32E-228 | InN4 | SLC24A3 |
| PROX1-AS11 | 8.45E-232 | 0.43899931 | 0.328 | 0.104 | 3.23E-227 | InN4 | PROX1-AS1 |
| MAP3K53 | 4.61E-231 | 0.77908377 | 0.817 | 0.633 | 1.76E-226 | InN4 | MAP3K5 |
| UNC5B | 2.82E-230 | 0.53098269 | 0.37 | 0.141 | 1.08E-225 | InN4 | UNC5B |
| AL136456.12 | 5.12E-230 | 1.55350626 | 0.349 | 0.137 | 1.96E-225 | InN4 | AL136456.1 |
| KLF3-AS12 | 2.26E-227 | 0.69420395 | 0.518 | 0.255 | 8.65E-223 | InN4 | KLF3-AS1 |
| HS3ST51 | 9.43E-227 | 0.76340619 | 0.579 | 0.303 | 3.61E-222 | InN4 | HS3ST5 |
| PTPRB1 | 3.66E-225 | 0.85404291 | 0.625 | 0.41 | 1.40E-220 | InN4 | PTPRB |
| ATXN11 | 7.25E-225 | 0.53235864 | 0.948 | 0.926 | 2.77E-220 | InN4 | ATXN1 |
| TLE21 | 2.12E-221 | 0.58366604 | 0.431 | 0.189 | 8.08E-217 | InN4 | TLE2 |
| DLGAP12 | 2.12E-220 | 0.46920325 | 0.994 | 0.989 | 8.10E-216 | InN4 | DLGAP1 |
| CCNH1 | 3.90E-220 | 0.79072102 | 0.853 | 0.764 | 1.49E-215 | InN4 | CCNH |
| SLCO5A1 | 1.01E-219 | 0.7113601 | 0.307 | 0.107 | 3.87E-215 | InN4 | SLCO5A1 |
| SLC2A131 | 1.12E-219 | 0.70799637 | 0.896 | 0.855 | 4.27E-215 | InN4 | SLC2A13 |
| PBX1 | 1.43E-219 | 0.66986585 | 0.933 | 0.887 | 5.45E-215 | InN4 | PBX1 |
| PAWR2 | 6.39E-219 | 0.40293308 | 0.272 | 0.08 | 2.44E-214 | InN4 | PAWR |
| PKNOX21 | 4.17E-218 | 0.72253623 | 0.857 | 0.752 | 1.60E-213 | InN4 | PKNOX2 |
| PTPRG2 | 1.78E-216 | 0.59096923 | 0.941 | 0.908 | 6.82E-212 | InN4 | PTPRG |
| KCNAB11 | 5.14E-216 | 0.69677213 | 0.358 | 0.135 | 1.97E-211 | InN4 | KCNAB1 |
| PLEKHO1 | 1.51E-215 | 0.62562814 | 0.463 | 0.23 | 5.76E-211 | InN4 | PLEKHO1 |
| HPCAL11 | 1.22E-214 | 0.68896847 | 0.545 | 0.283 | 4.68E-210 | InN4 | HPCAL1 |
| DNM32 | 1.58E-213 | 0.5480977 | 0.919 | 0.891 | 6.04E-209 | InN4 | DNM3 |
| PCP4L1 | 9.03E-213 | 0.3660898 | 0.258 | 0.077 | 3.45E-208 | InN4 | PCP4L1 |
| AF279873.3 | 1.44E-212 | 0.66418501 | 0.36 | 0.139 | 5.52E-208 | InN4 | AF279873.3 |
| SERTM1 | 2.62E-212 | 0.43279957 | 0.284 | 0.095 | 1.00E-207 | InN4 | SERTM1 |
| SYT15 | 4.13E-210 | 0.46192705 | 0.995 | 0.985 | 1.58E-205 | InN4 | SYT1 |
| LINC002992 | 2.52E-209 | 0.69785313 | 0.528 | 0.271 | 9.63E-205 | InN4 | LINC00299 |
| AC073050.12 | 3.73E-208 | 0.78939944 | 0.863 | 0.77 | 1.43E-203 | InN4 | AC073050.1 |
| ANK3 | 3.77E-207 | 0.39689999 | 0.98 | 0.976 | 1.44E-202 | InN4 | ANK3 |
| NHS3 | 8.74E-205 | 0.67294996 | 0.707 | 0.438 | 3.34E-200 | InN4 | NHS |
| SAMD12 | 1.04E-204 | 0.51259472 | 0.941 | 0.921 | 3.97E-200 | InN4 | SAMD12 |
| EPHA33 | 5.08E-203 | 0.75723855 | 0.658 | 0.411 | 1.94E-198 | InN4 | EPHA3 |
| RXRA1 | 5.85E-202 | 0.5416822 | 0.435 | 0.198 | 2.24E-197 | InN4 | RXRA |
| DOK5 | 1.43E-201 | 0.71452915 | 0.54 | 0.306 | 5.46E-197 | InN4 | DOK5 |
| NFIX1 | 7.15E-201 | 0.70987867 | 0.778 | 0.626 | 2.73E-196 | InN4 | NFIX |
| USP32 | 9.43E-201 | 0.47093856 | 0.396 | 0.165 | 3.60E-196 | InN4 | USP3 |
| ANK13 | 1.70E-197 | 0.51358451 | 0.606 | 0.306 | 6.49E-193 | InN4 | ANK1 |
| NXN | 3.41E-195 | 0.5630738 | 0.355 | 0.141 | 1.31E-190 | InN4 | NXN |
| SPOCK21 | 8.92E-194 | 0.75550454 | 0.623 | 0.426 | 3.41E-189 | InN4 | SPOCK2 |
| PIK3R31 | 1.26E-191 | 0.6716941 | 0.542 | 0.307 | 4.82E-187 | InN4 | PIK3R3 |

| | | | | | | | |
|---|---|---|---|---|---|---|---|
| WASF32 | 2.41E-191 | 0.64388807 | 0.589 | 0.374 | 9.23E-187 | InN4 | WASF3 |
| NRG13 | 2.82E-191 | 0.92997976 | 0.761 | 0.538 | 1.08E-186 | InN4 | NRG1 |
| KCNQ35 | 4.00E-191 | 0.5320631 | 0.963 | 0.899 | 1.53E-186 | InN4 | KCNQ3 |
| GALNT181 | 1.47E-190 | 0.62811454 | 0.874 | 0.739 | 5.63E-186 | InN4 | GALNT18 |
| SLIT21 | 1.90E-190 | 0.96140194 | 0.771 | 0.596 | 7.26E-186 | InN4 | SLIT2 |
| SOX55 | 4.76E-189 | 0.67612927 | 0.889 | 0.66 | 1.82E-184 | InN4 | SOX5 |
| ARX2 | 4.99E-186 | 0.34724836 | 0.29 | 0.098 | 1.91E-181 | InN4 | ARX |
| AC003991.1 | 2.56E-185 | 0.54031763 | 0.284 | 0.104 | 9.77E-181 | InN4 | AC003991.1 |
| CDH41 | 7.11E-184 | 0.71715894 | 0.634 | 0.417 | 2.72E-179 | InN4 | CDH4 |
| TRPS12 | 1.93E-183 | 0.8344469 | 0.637 | 0.429 | 7.38E-179 | InN4 | TRPS1 |
| ETV11 | 1.09E-182 | 0.46058244 | 0.283 | 0.103 | 4.18E-178 | InN4 | ETV1 |
| DOK61 | 1.05E-181 | 0.78303153 | 0.824 | 0.701 | 4.02E-177 | InN4 | DOK6 |
| PDSS1 | 3.64E-180 | 0.58226438 | 0.411 | 0.201 | 1.39E-175 | InN4 | PDSS1 |
| ARID5B | 1.11E-179 | 0.65699653 | 0.808 | 0.726 | 4.24E-175 | InN4 | ARID5B |
| ST6GAL24 | 8.27E-179 | 0.69450459 | 0.595 | 0.382 | 3.16E-174 | InN4 | ST6GAL2 |
| AMMECR1 | 1.70E-178 | 0.75978566 | 0.717 | 0.572 | 6.51E-174 | InN4 | AMMECR1 |
| RASGRF1 | 2.15E-178 | 0.67406214 | 0.677 | 0.505 | 8.23E-174 | InN4 | RASGRF1 |
| NOS12 | 5.87E-178 | 0.69599983 | 0.289 | 0.109 | 2.24E-173 | InN4 | NOS1 |
| CA13 | 1.58E-176 | 0.64779212 | 0.349 | 0.157 | 6.04E-172 | InN4 | CA13 |
| FRMD4B3 | 1.19E-175 | 0.5972558 | 0.376 | 0.16 | 4.54E-171 | InN4 | FRMD4B |
| CSMD3 | 1.35E-175 | 0.44820622 | 0.97 | 0.94 | 5.15E-171 | InN4 | CSMD3 |
| ITGA82 | 2.72E-175 | 0.78671449 | 0.306 | 0.119 | 1.04E-170 | InN4 | ITGA8 |
| SBF2 | 2.49E-173 | 0.55621462 | 0.841 | 0.799 | 9.53E-169 | InN4 | SBF2 |
| FOXN32 | 3.16E-172 | 0.64348906 | 0.815 | 0.703 | 1.21E-167 | InN4 | FOXN3 |
| SLCO3A12 | 4.17E-170 | 0.6010199 | 0.826 | 0.732 | 1.60E-165 | InN4 | SLCO3A1 |
| BICD12 | 6.45E-170 | 0.56238931 | 0.85 | 0.792 | 2.47E-165 | InN4 | BICD1 |
| FAM13C1 | 2.58E-168 | 0.70949538 | 0.665 | 0.505 | 9.86E-164 | InN4 | FAM13C |
| KCNH54 | 1.27E-167 | 0.6645474 | 0.648 | 0.408 | 4.87E-163 | InN4 | KCNH5 |
| OSBPL33 | 1.49E-167 | 0.66862485 | 0.647 | 0.427 | 5.68E-163 | InN4 | OSBPL3 |
| LRRFIP2 | 1.77E-167 | 0.67815684 | 0.769 | 0.689 | 6.78E-163 | InN4 | LRRFIP2 |
| MYRIP3 | 9.30E-167 | 0.52572754 | 0.886 | 0.803 | 3.55E-162 | InN4 | MYRIP |
| AJAP1 | 5.16E-165 | 0.64760088 | 0.725 | 0.575 | 1.97E-160 | InN4 | AJAP1 |
| SLC8A13 | 1.07E-164 | 0.50666032 | 0.969 | 0.857 | 4.08E-160 | InN4 | SLC8A1 |
| MTR1 | 1.10E-163 | 0.64451043 | 0.708 | 0.591 | 4.21E-159 | InN4 | MTR |
| VPS13C2 | 8.89E-163 | 0.5644497 | 0.844 | 0.805 | 3.40E-158 | InN4 | VPS13C |
| MBP2 | 1.16E-162 | 0.63097562 | 0.708 | 0.524 | 4.45E-158 | InN4 | MBP |
| PRICKLE21 | 2.42E-162 | 0.61543792 | 0.844 | 0.792 | 9.25E-158 | InN4 | PRICKLE2 |
| ABCC11 | 6.52E-162 | 0.6503827 | 0.549 | 0.347 | 2.49E-157 | InN4 | ABCC1 |
| GALNT22 | 2.51E-161 | 0.65696021 | 0.637 | 0.474 | 9.60E-157 | InN4 | GALNT2 |
| AC087457.11 | 3.35E-160 | 0.35182756 | 0.301 | 0.116 | 1.28E-155 | InN4 | AC087457.1 |
| ERC12 | 8.38E-160 | 0.47411326 | 0.894 | 0.866 | 3.20E-155 | InN4 | ERC1 |
| CNTN53 | 8.03E-159 | 0.56266665 | 0.947 | 0.81 | 3.07E-154 | InN4 | CNTN5 |
| MMD2 | 2.09E-158 | 0.55584239 | 0.333 | 0.143 | 7.99E-154 | InN4 | MMD2 |
| GRIN2B3 | 2.72E-158 | 0.42915775 | 0.959 | 0.946 | 1.04E-153 | InN4 | GRIN2B |
| REV3L2 | 1.84E-157 | 0.52438258 | 0.836 | 0.775 | 7.01E-153 | InN4 | REV3L |
| PCDH153 | 7.39E-157 | 0.44784463 | 0.575 | 0.315 | 2.82E-152 | InN4 | PCDH15 |
| ANKRD61 | 2.96E-156 | 0.63588664 | 0.61 | 0.409 | 1.13E-151 | InN4 | ANKRD6 |
| GRM41 | 5.73E-155 | 0.4413183 | 0.379 | 0.176 | 2.19E-150 | InN4 | GRM4 |
| OSBPL62 | 6.26E-155 | 0.5294416 | 0.844 | 0.802 | 2.39E-150 | InN4 | OSBPL6 |
| CDH202 | 1.64E-153 | 0.64435827 | 0.611 | 0.372 | 6.27E-149 | InN4 | CDH20 |
| NR2F1-AS11 | 2.24E-153 | 0.56528397 | 0.855 | 0.785 | 8.56E-149 | InN4 | NR2F1-AS1 |
| KIAA1109 | 3.81E-153 | 0.55619949 | 0.838 | 0.801 | 1.46E-148 | InN4 | KIAA1109 |
| LPIN22 | 1.50E-152 | 0.61569859 | 0.554 | 0.355 | 5.75E-148 | InN4 | LPIN2 |
| MCTP13 | 8.26E-151 | 0.46426823 | 0.883 | 0.763 | 3.16E-146 | InN4 | MCTP1 |
| RERE | 2.33E-150 | 0.40524828 | 0.929 | 0.925 | 8.89E-146 | InN4 | RERE |
| NINL2 | 5.52E-150 | 0.39528392 | 0.386 | 0.179 | 2.11E-145 | InN4 | NINL |
| IL6ST2 | 9.16E-150 | 0.6124202 | 0.515 | 0.319 | 3.50E-145 | InN4 | IL6ST |
| TLK11 | 1.88E-148 | 0.62219472 | 0.734 | 0.623 | 7.20E-144 | InN4 | TLK1 |
| SERINC52 | 1.11E-146 | 0.60769628 | 0.601 | 0.415 | 4.26E-142 | InN4 | SERINC5 |
| OPHN1 | 3.79E-146 | 0.66989086 | 0.71 | 0.607 | 1.45E-141 | InN4 | OPHN1 |
| AMOTL12 | 1.66E-143 | 0.41701059 | 0.405 | 0.195 | 6.35E-139 | InN4 | AMOTL1 |
| FARP14 | 1.43E-142 | 0.55424483 | 0.787 | 0.652 | 5.46E-138 | InN4 | FARP1 |
| ADAM22 | 4.07E-141 | 0.54255887 | 0.846 | 0.825 | 1.56E-136 | InN4 | ADAM22 |

| | | | | | | | |
|---|---|---|---|---|---|---|---|
| MAML31 | 1.29E-140 | 0.68209884 | 0.559 | 0.353 | 4.92E-136 | InN4 | MAML3 |
| LRRTM32 | 8.65E-140 | 0.66266544 | 0.891 | 0.83 | 3.31E-135 | InN4 | LRRTM3 |
| MTUS13 | 1.21E-139 | 0.47932183 | 0.513 | 0.289 | 4.61E-135 | InN4 | MTUS1 |
| ARHGEF26-A | 1.46E-139 | 0.40981475 | 0.304 | 0.128 | 5.58E-135 | InN4 | ARHGEF26-A |
| SWAP701 | 4.45E-139 | 0.36221259 | 0.286 | 0.119 | 1.70E-134 | InN4 | SWAP70 |
| KCNQ4 | 1.19E-138 | 0.34284945 | 0.255 | 0.099 | 4.54E-134 | InN4 | KCNQ4 |
| MGLL2 | 2.31E-138 | 0.57161845 | 0.616 | 0.431 | 8.83E-134 | InN4 | MGLL |
| HECW2 | 1.44E-134 | 0.4542005 | 0.89 | 0.866 | 5.51E-130 | InN4 | HECW2 |
| PHYHIPL1 | 2.85E-132 | 0.62331776 | 0.702 | 0.588 | 1.09E-127 | InN4 | PHYHIPL |
| LYPD6B3 | 3.46E-132 | 0.38568705 | 0.372 | 0.171 | 1.32E-127 | InN4 | LYPD6B |
| SEC14L1 | 2.11E-131 | 0.57606801 | 0.553 | 0.381 | 8.08E-127 | InN4 | SEC14L1 |
| FCHSD24 | 1.90E-129 | 0.51464608 | 0.744 | 0.608 | 7.28E-125 | InN4 | FCHSD2 |
| MIRLET7BHG | 4.05E-128 | 0.59491832 | 0.587 | 0.418 | 1.55E-123 | InN4 | MIRLET7BHG |
| ATP1B22 | 8.66E-128 | 0.45341167 | 0.423 | 0.224 | 3.31E-123 | InN4 | ATP1B2 |
| MIR181A1HG | 8.13E-123 | 0.59483746 | 0.612 | 0.439 | 3.11E-118 | InN4 | MIR181A1HG |
| ADCYAP1R13 | 1.71E-122 | 0.43919924 | 0.504 | 0.293 | 6.54E-118 | InN4 | ADCYAP1R1 |
| KCTD162 | 3.26E-122 | 0.46306902 | 0.938 | 0.864 | 1.25E-117 | InN4 | KCTD16 |
| SGMS11 | 3.91E-122 | 0.57854423 | 0.74 | 0.665 | 1.50E-117 | InN4 | SGMS1 |
| BCKDHB | 1.45E-119 | 0.59922204 | 0.609 | 0.469 | 5.55E-115 | InN4 | BCKDHB |
| PKP4 | 5.32E-118 | 0.46403448 | 0.867 | 0.821 | 2.03E-113 | InN4 | PKP4 |
| CACNA1A3 | 8.88E-117 | 0.39750956 | 0.946 | 0.908 | 3.40E-112 | InN4 | CACNA1A |
| ANO61 | 7.45E-116 | 0.53017888 | 0.463 | 0.289 | 2.85E-111 | InN4 | ANO6 |
| FXYD62 | 4.21E-114 | 0.56077849 | 0.708 | 0.615 | 1.61E-109 | InN4 | FXYD6 |
| MEF2C-AS11 | 8.00E-113 | 0.55098255 | 0.809 | 0.741 | 3.06E-108 | InN4 | MEF2C-AS1 |
| ATXN7L1 | 1.18E-112 | 0.53124557 | 0.77 | 0.696 | 4.50E-108 | InN4 | ATXN7L1 |
| RASGRP12 | 5.31E-112 | 0.43304433 | 0.352 | 0.186 | 2.03E-107 | InN4 | RASGRP1 |
| ZMAT42 | 3.34E-111 | 0.45418671 | 0.867 | 0.764 | 1.28E-106 | InN4 | ZMAT4 |
| DOCK3 | 4.10E-111 | 0.31850017 | 0.962 | 0.965 | 1.57E-106 | InN4 | DOCK3 |
| RBMS11 | 1.02E-109 | 0.53724241 | 0.595 | 0.444 | 3.89E-105 | InN4 | RBMS1 |
| FAM135B3 | 1.04E-109 | 0.5321377 | 0.758 | 0.627 | 3.97E-105 | InN4 | FAM135B |
| HUNK2 | 5.65E-109 | 0.52807904 | 0.478 | 0.303 | 2.16E-104 | InN4 | HUNK |
| SSH21 | 6.23E-109 | 0.44439133 | 0.853 | 0.82 | 2.38E-104 | InN4 | SSH2 |
| MYT1 | 2.60E-108 | 0.29537821 | 0.25 | 0.107 | 9.94E-104 | InN4 | MYT1 |
| CCDC85A3 | 1.20E-106 | 0.54513286 | 0.77 | 0.652 | 4.57E-102 | InN4 | CCDC85A |
| CFAP1613 | 1.86E-106 | 0.53623954 | 0.485 | 0.31 | 7.12E-102 | InN4 | CFAP161 |
| KCNN22 | 6.32E-104 | 0.54003479 | 0.63 | 0.489 | 2.42E-99 | InN4 | KCNN2 |
| TMEM653 | 1.66E-103 | 0.52044214 | 0.544 | 0.392 | 6.36E-99 | InN4 | TMEM65 |
| ID2 | 1.77E-103 | 0.45074285 | 0.341 | 0.186 | 6.75E-99 | InN4 | ID2 |
| AC007405.11 | 6.37E-103 | 0.33748521 | 0.259 | 0.116 | 2.44E-98 | InN4 | AC007405.1 |
| CPEB31 | 8.35E-103 | 0.41795486 | 0.857 | 0.847 | 3.19E-98 | InN4 | CPEB3 |
| CYFIP21 | 2.12E-102 | 0.46695929 | 0.759 | 0.709 | 8.12E-98 | InN4 | CYFIP2 |
| KCNJ31 | 2.75E-102 | 0.41184135 | 0.886 | 0.869 | 1.05E-97 | InN4 | KCNJ3 |
| MPDZ1 | 6.79E-102 | 0.50757891 | 0.795 | 0.769 | 2.59E-97 | InN4 | MPDZ |
| NECAB2 | 2.83E-101 | 0.37231459 | 0.328 | 0.171 | 1.08E-96 | InN4 | NECAB2 |
| EPHB2 | 1.70E-99 | 0.53471625 | 0.53 | 0.375 | 6.52E-95 | InN4 | EPHB2 |
| CDC42BPA | 3.15E-99 | 0.36538954 | 0.91 | 0.905 | 1.20E-94 | InN4 | CDC42BPA |
| TGFB21 | 8.68E-99 | 0.35670872 | 0.308 | 0.153 | 3.32E-94 | InN4 | TGFB2 |
| RALGPS12 | 5.55E-98 | 0.46871091 | 0.76 | 0.705 | 2.12E-93 | InN4 | RALGPS1 |
| CNTNAP52 | 8.91E-98 | 0.50769224 | 0.931 | 0.9 | 3.41E-93 | InN4 | CNTNAP5 |
| SLC20A2 | 1.31E-96 | 0.52789627 | 0.717 | 0.634 | 5.01E-92 | InN4 | SLC20A2 |
| PAK51 | 5.59E-96 | 0.44303796 | 0.812 | 0.758 | 2.14E-91 | InN4 | PAK5 |
| KANSL1L2 | 2.37E-94 | 0.50213233 | 0.584 | 0.455 | 9.06E-90 | InN4 | KANSL1L |
| VAT1L1 | 2.76E-94 | 0.47845602 | 0.399 | 0.239 | 1.05E-89 | InN4 | VAT1L |
| MYH14 | 5.17E-94 | 0.33768345 | 0.289 | 0.144 | 1.98E-89 | InN4 | MYH14 |
| BTBD8 | 7.73E-94 | 0.52481832 | 0.692 | 0.622 | 2.96E-89 | InN4 | BTBD8 |
| MAST42 | 7.82E-94 | 0.41247734 | 0.865 | 0.816 | 2.99E-89 | InN4 | MAST4 |
| SSBP22 | 1.80E-93 | 0.44923296 | 0.837 | 0.824 | 6.89E-89 | InN4 | SSBP2 |
| LCORL1 | 2.00E-92 | 0.48432544 | 0.617 | 0.491 | 7.66E-88 | InN4 | LCORL |
| SVIL2 | 5.29E-92 | 0.41224503 | 0.319 | 0.169 | 2.02E-87 | InN4 | SVIL |
| SDK13 | 1.26E-91 | 0.60233811 | 0.636 | 0.474 | 4.80E-87 | InN4 | SDK1 |
| AC087564.12 | 2.00E-91 | 0.36135126 | 0.287 | 0.136 | 7.65E-87 | InN4 | AC087564.1 |
| CSMD21 | 4.95E-91 | 0.44344362 | 0.811 | 0.771 | 1.89E-86 | InN4 | CSMD2 |
| PHF14 | 6.63E-91 | 0.44537233 | 0.796 | 0.78 | 2.54E-86 | InN4 | PHF14 |

| | | | | | | | |
|---|---|---|---|---|---|---|---|
| NCOA22 | 1.43E-90 | 0.40479723 | 0.793 | 0.741 | 5.47E-86 | InN4 | NCOA2 |
| HDAC93 | 2.26E-90 | 0.4097395 | 0.847 | 0.776 | 8.65E-86 | InN4 | HDAC9 |
| LINC027983 | 1.34E-89 | 0.44267941 | 0.357 | 0.206 | 5.11E-85 | InN4 | LINC02798 |
| FUT82 | 3.29E-88 | 0.45564863 | 0.749 | 0.695 | 1.26E-83 | InN4 | FUT8 |
| TENM12 | 4.29E-88 | 0.33307689 | 0.564 | 0.374 | 1.64E-83 | InN4 | TENM1 |
| TSPAN91 | 8.77E-88 | 0.41632171 | 0.418 | 0.257 | 3.35E-83 | InN4 | TSPAN9 |
| LINC005991 | 4.47E-87 | 0.45593713 | 0.764 | 0.731 | 1.71E-82 | InN4 | LINC00599 |
| TMEM164 | 3.10E-86 | 0.42381221 | 0.389 | 0.241 | 1.18E-81 | InN4 | TMEM164 |
| MARCH41 | 3.17E-86 | 0.49420582 | 0.538 | 0.401 | 1.21E-81 | InN4 | 4-Mar |
| SSTR23 | 1.15E-85 | 0.387438 | 0.323 | 0.181 | 4.41E-81 | InN4 | SSTR2 |
| ADAMTS21 | 3.40E-85 | 0.36002824 | 0.278 | 0.14 | 1.30E-80 | InN4 | ADAMTS2 |
| APBA1 | 5.47E-85 | 0.43747931 | 0.789 | 0.754 | 2.09E-80 | InN4 | APBA1 |
| MAP3K11 | 8.18E-85 | 0.32074909 | 0.301 | 0.158 | 3.13E-80 | InN4 | MAP3K1 |
| PPFIBP13 | 1.35E-84 | 0.44944707 | 0.76 | 0.659 | 5.14E-80 | InN4 | PPFIBP1 |
| RARS2 | 2.20E-84 | 0.4567176 | 0.526 | 0.399 | 8.42E-80 | InN4 | RARS2 |
| UBTD2 | 6.93E-84 | 0.3367982 | 0.295 | 0.157 | 2.65E-79 | InN4 | UBTD2 |
| HRH11 | 8.83E-84 | 0.47425402 | 0.368 | 0.225 | 3.37E-79 | InN4 | HRH1 |
| MTMR7 | 3.14E-83 | 0.46150723 | 0.678 | 0.613 | 1.20E-78 | InN4 | MTMR7 |
| PRKG21 | 4.22E-83 | 0.40057921 | 0.311 | 0.172 | 1.61E-78 | InN4 | PRKG2 |
| CACNG32 | 8.65E-83 | 0.48487101 | 0.743 | 0.652 | 3.31E-78 | InN4 | CACNG3 |
| MXD41 | 1.01E-82 | 0.41010113 | 0.399 | 0.25 | 3.87E-78 | InN4 | MXD4 |
| ST51 | 3.53E-82 | 0.53341415 | 0.475 | 0.339 | 1.35E-77 | InN4 | ST5 |
| URI12 | 5.23E-82 | 0.4509255 | 0.574 | 0.464 | 2.00E-77 | InN4 | URI1 |
| SNCAIP1 | 1.00E-81 | 0.37123304 | 0.296 | 0.157 | 3.84E-77 | InN4 | SNCAIP |
| PODXL22 | 1.35E-81 | 0.43255773 | 0.535 | 0.386 | 5.15E-77 | InN4 | PODXL2 |
| CACNA1I | 1.10E-80 | 0.40634858 | 0.442 | 0.292 | 4.22E-76 | InN4 | CACNA1I |
| TRERF12 | 1.30E-80 | 0.43144801 | 0.734 | 0.653 | 4.97E-76 | InN4 | TRERF1 |
| FTO | 3.53E-79 | 0.35655252 | 0.852 | 0.855 | 1.35E-74 | InN4 | FTO |
| PDE10A3 | 9.87E-79 | 0.4166215 | 0.785 | 0.672 | 3.77E-74 | InN4 | PDE10A |
| ITM2C | 1.23E-78 | 0.4977049 | 0.392 | 0.258 | 4.71E-74 | InN4 | ITM2C |
| PLAGL13 | 1.78E-78 | 0.37243195 | 0.378 | 0.23 | 6.79E-74 | InN4 | PLAGL1 |
| CAP21 | 5.97E-78 | 0.45584206 | 0.729 | 0.654 | 2.28E-73 | InN4 | CAP2 |
| BRINP21 | 1.26E-77 | 0.46722605 | 0.746 | 0.644 | 4.82E-73 | InN4 | BRINP2 |
| CASK | 2.73E-77 | 0.4361164 | 0.688 | 0.631 | 1.04E-72 | InN4 | CASK |
| ZDHHC141 | 5.47E-76 | 0.38088157 | 0.837 | 0.775 | 2.09E-71 | InN4 | ZDHHC14 |
| ADGRL23 | 5.75E-76 | 0.40835194 | 0.907 | 0.855 | 2.20E-71 | InN4 | ADGRL2 |
| TMEFF24 | 6.21E-76 | 0.47665333 | 0.815 | 0.763 | 2.37E-71 | InN4 | TMEFF2 |
| VAV33 | 2.36E-75 | 0.59340273 | 0.266 | 0.143 | 9.02E-71 | InN4 | VAV3 |
| GABBR2 | 3.47E-75 | 0.31201366 | 0.924 | 0.918 | 1.33E-70 | InN4 | GABBR2 |
| POLA11 | 1.07E-74 | 0.48173667 | 0.519 | 0.397 | 4.11E-70 | InN4 | POLA1 |
| ST8SIA43 | 1.26E-74 | 0.3817187 | 0.315 | 0.179 | 4.83E-70 | InN4 | ST8SIA4 |
| CCNJL1 | 2.56E-74 | 0.27592112 | 0.251 | 0.125 | 9.80E-70 | InN4 | CCNJL |
| SNRPN3 | 7.25E-74 | 0.37765501 | 0.816 | 0.76 | 2.77E-69 | InN4 | SNRPN |
| PDE8A2 | 2.37E-73 | 0.4370557 | 0.557 | 0.435 | 9.04E-69 | InN4 | PDE8A |
| ANKRD30BL1 | 4.74E-73 | 0.42788758 | 0.784 | 0.705 | 1.81E-68 | InN4 | ANKRD30BL |
| LMCD1-AS12 | 7.16E-73 | 0.62981383 | 0.417 | 0.285 | 2.74E-68 | InN4 | LMCD1-AS1 |
| KIAA19582 | 2.23E-72 | 0.42310066 | 0.559 | 0.426 | 8.53E-68 | InN4 | KIAA1958 |
| GPR1551 | 2.38E-72 | 0.445736 | 0.664 | 0.598 | 9.09E-68 | InN4 | GPR155 |
| SH3PXD2B | 2.49E-72 | 0.29021467 | 0.258 | 0.134 | 9.53E-68 | InN4 | SH3PXD2B |
| TRAF3IP2-AS | 2.15E-71 | 0.39683507 | 0.775 | 0.749 | 8.23E-67 | InN4 | TRAF3IP2-AS |
| ST3GAL12 | 2.31E-71 | 0.45104032 | 0.435 | 0.297 | 8.84E-67 | InN4 | ST3GAL1 |
| MBOAT22 | 5.71E-71 | 0.41900515 | 0.679 | 0.586 | 2.18E-66 | InN4 | MBOAT2 |
| GRIN2D3 | 5.90E-71 | 0.2575821 | 0.272 | 0.139 | 2.26E-66 | InN4 | GRIN2D |
| DCX3 | 2.07E-70 | 0.39108006 | 0.53 | 0.392 | 7.89E-66 | InN4 | DCX |
| LRRC8B2 | 4.32E-70 | 0.43887711 | 0.723 | 0.676 | 1.65E-65 | InN4 | LRRC8B |
| ATP13A3 | 4.32E-70 | 0.46442975 | 0.535 | 0.437 | 1.65E-65 | InN4 | ATP13A3 |
| TBC1D42 | 4.67E-70 | 0.44595416 | 0.574 | 0.461 | 1.79E-65 | InN4 | TBC1D4 |
| SH3KBP14 | 5.59E-70 | 0.43680752 | 0.675 | 0.604 | 2.14E-65 | InN4 | SH3KBP1 |
| TNR1 | 6.08E-70 | 0.40472236 | 0.862 | 0.799 | 2.32E-65 | InN4 | TNR |
| SYNE11 | 9.21E-70 | 0.30672112 | 0.962 | 0.952 | 3.52E-65 | InN4 | SYNE1 |
| CCK1 | 1.64E-69 | 0.36178698 | 0.58 | 0.45 | 6.27E-65 | InN4 | CCK |
| LINC01630 | 2.41E-69 | 0.35661683 | 0.251 | 0.131 | 9.22E-65 | InN4 | LINC01630 |
| ASPH2 | 1.36E-68 | 0.45043942 | 0.661 | 0.598 | 5.19E-64 | InN4 | ASPH |

| | | | | | | | |
|---|---|---|---|---|---|---|---|
| TTC39B1 | 7.14E-68 | 0.45412854 | 0.558 | 0.457 | 2.73E-63 | InN4 | TTC39B |
| KIF21A1 | 9.32E-68 | 0.41613176 | 0.738 | 0.719 | 3.56E-63 | InN4 | KIF21A |
| GRM53 | 4.59E-67 | 0.30202074 | 0.96 | 0.897 | 1.75E-62 | InN4 | GRM5 |
| TTC39C2 | 2.89E-66 | 0.42745886 | 0.683 | 0.599 | 1.11E-61 | InN4 | TTC39C |
| NF2 | 3.04E-66 | 0.42761841 | 0.418 | 0.294 | 1.16E-61 | InN4 | NF2 |
| IL1RAP3 | 6.37E-66 | 0.57062849 | 0.438 | 0.322 | 2.44E-61 | InN4 | IL1RAP |
| MPP61 | 1.32E-65 | 0.46827119 | 0.652 | 0.582 | 5.05E-61 | InN4 | MPP6 |
| EPN21 | 1.44E-65 | 0.4402404 | 0.634 | 0.552 | 5.49E-61 | InN4 | EPN2 |
| NETO13 | 3.84E-65 | 0.37467208 | 0.798 | 0.729 | 1.47E-60 | InN4 | NETO1 |
| GALNT101 | 4.08E-65 | 0.37181495 | 0.254 | 0.141 | 1.56E-60 | InN4 | GALNT10 |
| KLF3 | 6.31E-65 | 0.26207735 | 0.256 | 0.138 | 2.41E-60 | InN4 | KLF3 |
| ARG2 | 8.68E-65 | 0.25076479 | 0.257 | 0.139 | 3.32E-60 | InN4 | ARG2 |
| LIFR | 1.23E-64 | 0.46359854 | 0.432 | 0.311 | 4.70E-60 | InN4 | LIFR |
| CYTH13 | 3.94E-64 | 0.40357434 | 0.546 | 0.437 | 1.51E-59 | InN4 | CYTH1 |
| CLIP41 | 1.21E-63 | 0.42664996 | 0.606 | 0.541 | 4.64E-59 | InN4 | CLIP4 |
| KITLG1 | 1.52E-63 | 0.42759409 | 0.416 | 0.288 | 5.81E-59 | InN4 | KITLG |
| AIG11 | 2.69E-63 | 0.36995904 | 0.788 | 0.773 | 1.03E-58 | InN4 | AIG1 |
| PLPPR13 | 8.03E-63 | 0.26743971 | 0.454 | 0.29 | 3.07E-58 | InN4 | PLPPR1 |
| SRGAP3 | 1.59E-62 | 0.32010847 | 0.883 | 0.873 | 6.06E-58 | InN4 | SRGAP3 |
| PWRN13 | 1.79E-62 | 0.34670917 | 0.631 | 0.464 | 6.83E-58 | InN4 | PWRN1 |
| RBFOX32 | 1.01E-61 | 0.31130389 | 0.912 | 0.883 | 3.85E-57 | InN4 | RBFOX3 |
| DYNC1I14 | 1.24E-61 | 0.31219733 | 0.883 | 0.873 | 4.75E-57 | InN4 | DYNC1I1 |
| NAV12 | 1.37E-61 | 0.39738817 | 0.719 | 0.674 | 5.22E-57 | InN4 | NAV1 |
| JAK11 | 8.98E-61 | 0.38160893 | 0.757 | 0.744 | 3.43E-56 | InN4 | JAK1 |
| LINGO22 | 1.56E-60 | 0.31812789 | 0.953 | 0.819 | 5.98E-56 | InN4 | LINGO2 |
| CALD12 | 7.85E-60 | 0.37111835 | 0.442 | 0.317 | 3.00E-55 | InN4 | CALD1 |
| RIPOR22 | 1.10E-59 | 0.35468348 | 0.584 | 0.464 | 4.20E-55 | InN4 | RIPOR2 |
| MOB3B | 1.30E-59 | 0.46456693 | 0.502 | 0.388 | 4.97E-55 | InN4 | MOB3B |
| VSTM2L2 | 5.26E-59 | 0.41066408 | 0.417 | 0.295 | 2.01E-54 | InN4 | VSTM2L |
| ECE21 | 1.55E-58 | 0.30062205 | 0.357 | 0.226 | 5.92E-54 | InN4 | ECE2 |
| ABLIM14 | 1.62E-58 | 0.27687527 | 0.866 | 0.832 | 6.21E-54 | InN4 | ABLIM1 |
| PIK3C2B | 5.86E-58 | 0.33356869 | 0.356 | 0.235 | 2.24E-53 | InN4 | PIK3C2B |
| PACSIN22 | 7.94E-58 | 0.39665841 | 0.461 | 0.345 | 3.04E-53 | InN4 | PACSIN2 |
| CHD3 | 1.72E-57 | 0.39994977 | 0.47 | 0.362 | 6.57E-53 | InN4 | CHD3 |
| C5orf561 | 2.44E-57 | 0.34604782 | 0.404 | 0.278 | 9.33E-53 | InN4 | C5orf56 |
| TTBK21 | 5.57E-57 | 0.41166779 | 0.709 | 0.684 | 2.13E-52 | InN4 | TTBK2 |
| SLC7A82 | 5.94E-57 | 0.39577827 | 0.478 | 0.368 | 2.27E-52 | InN4 | SLC7A8 |
| ANKRD30B1 | 8.08E-57 | 0.43019712 | 0.391 | 0.275 | 3.09E-52 | InN4 | ANKRD30B |
| FNBP1L3 | 1.47E-56 | 0.32237496 | 0.528 | 0.384 | 5.63E-52 | InN4 | FNBP1L |
| TCF20 | 1.84E-56 | 0.39085984 | 0.677 | 0.63 | 7.03E-52 | InN4 | TCF20 |
| CDK81 | 3.46E-56 | 0.4061436 | 0.624 | 0.561 | 1.32E-51 | InN4 | CDK8 |
| DST1 | 3.90E-56 | 0.25178534 | 0.949 | 0.954 | 1.49E-51 | InN4 | DST |
| BACH24 | 4.52E-56 | 0.38602949 | 0.681 | 0.61 | 1.73E-51 | InN4 | BACH2 |
| PLCH21 | 9.20E-56 | 0.28275773 | 0.307 | 0.189 | 3.52E-51 | InN4 | PLCH2 |
| PLXDC1 | 6.32E-55 | 0.33264808 | 0.292 | 0.18 | 2.42E-50 | InN4 | PLXDC1 |
| TMEM74B | 8.54E-55 | 0.27286999 | 0.265 | 0.158 | 3.26E-50 | InN4 | TMEM74B |
| CCBE1 | 1.48E-53 | 0.48806907 | 0.325 | 0.212 | 5.64E-49 | InN4 | CCBE1 |
| CHL11 | 2.75E-53 | 0.34362834 | 0.846 | 0.805 | 1.05E-48 | InN4 | CHL1 |
| GPR1761 | 6.26E-53 | 0.38884034 | 0.442 | 0.329 | 2.39E-48 | InN4 | GPR176 |
| RGS7BP | 9.51E-53 | 0.43097486 | 0.678 | 0.634 | 3.63E-48 | InN4 | RGS7BP |
| BCAT12 | 1.40E-52 | 0.33509538 | 0.461 | 0.346 | 5.36E-48 | InN4 | BCAT1 |
| TRPC52 | 8.72E-52 | 0.35818944 | 0.451 | 0.325 | 3.33E-47 | InN4 | TRPC5 |
| PNPLA7 | 9.17E-52 | 0.42568776 | 0.499 | 0.411 | 3.50E-47 | InN4 | PNPLA7 |
| GABRB1 | 1.42E-51 | 0.28695045 | 0.917 | 0.917 | 5.42E-47 | InN4 | GABRB1 |
| PWRN42 | 1.75E-51 | 0.26890096 | 0.355 | 0.221 | 6.68E-47 | InN4 | PWRN4 |
| EXOC2 | 3.86E-51 | 0.38912844 | 0.643 | 0.59 | 1.47E-46 | InN4 | EXOC2 |
| PALLD2 | 4.64E-51 | 0.34395614 | 0.527 | 0.413 | 1.77E-46 | InN4 | PALLD |
| DIP2B1 | 2.68E-50 | 0.35020806 | 0.713 | 0.691 | 1.02E-45 | InN4 | DIP2B |
| RAPGEF43 | 1.42E-49 | 0.26153469 | 0.882 | 0.875 | 5.43E-45 | InN4 | RAPGEF4 |
| EFR3B1 | 8.79E-49 | 0.37689672 | 0.586 | 0.502 | 3.36E-44 | InN4 | EFR3B |
| CRADD2 | 1.18E-48 | 0.47899647 | 0.525 | 0.431 | 4.50E-44 | InN4 | CRADD |
| GARNL33 | 1.64E-48 | 0.29790837 | 0.778 | 0.724 | 6.29E-44 | InN4 | GARNL3 |
| PXK | 1.72E-48 | 0.39932005 | 0.555 | 0.468 | 6.56E-44 | InN4 | PXK |

| | | | | | | | |
|---|---|---|---|---|---|---|---|
| TIAM21 | 2.37E-48 | 0.3791207 | 0.76 | 0.736 | 9.05E-44 | InN4 | TIAM2 |
| NRG21 | 2.92E-48 | 0.34559447 | 0.666 | 0.616 | 1.12E-43 | InN4 | NRG2 |
| GLDN3 | 3.16E-48 | 0.28275196 | 0.351 | 0.232 | 1.21E-43 | InN4 | GLDN |
| CADM1 | 3.34E-47 | 0.312328 | 0.865 | 0.861 | 1.28E-42 | InN4 | CADM1 |
| RNF217-AS1 | 3.97E-47 | 0.33734052 | 0.259 | 0.159 | 1.52E-42 | InN4 | RNF217-AS1 |
| KSR12 | 4.75E-47 | 0.35623122 | 0.468 | 0.364 | 1.82E-42 | InN4 | KSR1 |
| CNNM21 | 6.60E-47 | 0.36140483 | 0.724 | 0.692 | 2.52E-42 | InN4 | CNNM2 |
| ADCY22 | 1.17E-46 | 0.29481729 | 0.863 | 0.854 | 4.46E-42 | InN4 | ADCY2 |
| KIDINS220 | 2.17E-46 | 0.3369178 | 0.739 | 0.739 | 8.29E-42 | InN4 | KIDINS220 |
| CFLAR1 | 2.82E-46 | 0.2983911 | 0.295 | 0.19 | 1.08E-41 | InN4 | CFLAR |
| CDC73 | 4.36E-46 | 0.42974011 | 0.549 | 0.479 | 1.67E-41 | InN4 | CDC73 |
| VSTM2A1 | 9.77E-46 | 0.33634307 | 0.495 | 0.39 | 3.74E-41 | InN4 | VSTM2A |
| GSE11 | 1.05E-45 | 0.38239517 | 0.589 | 0.499 | 4.03E-41 | InN4 | GSE1 |
| NEBL1 | 2.07E-45 | 0.31872684 | 0.88 | 0.876 | 7.90E-41 | InN4 | NEBL |
| SAT12 | 2.63E-45 | 0.39324349 | 0.461 | 0.364 | 1.00E-40 | InN4 | SAT1 |
| CPEB21 | 2.92E-45 | 0.40971215 | 0.511 | 0.438 | 1.12E-40 | InN4 | CPEB2 |
| FMNL24 | 6.17E-45 | 0.29049858 | 0.892 | 0.873 | 2.36E-40 | InN4 | FMNL2 |
| KCNN32 | 6.27E-45 | 0.36342518 | 0.423 | 0.317 | 2.40E-40 | InN4 | KCNN3 |
| PLCL21 | 9.14E-45 | 0.35925148 | 0.72 | 0.696 | 3.49E-40 | InN4 | PLCL2 |
| NDRG1 | 1.06E-44 | 0.28325316 | 0.268 | 0.168 | 4.05E-40 | InN4 | NDRG1 |
| KLHL291 | 2.61E-44 | 0.4131669 | 0.711 | 0.701 | 9.98E-40 | InN4 | KLHL29 |
| NMNAT33 | 2.67E-44 | 0.33390331 | 0.354 | 0.247 | 1.02E-39 | InN4 | NMNAT3 |
| GABRA51 | 6.10E-44 | 0.3757206 | 0.337 | 0.236 | 2.33E-39 | InN4 | GABRA5 |
| OIP5-AS11 | 9.62E-44 | 0.43726335 | 0.622 | 0.569 | 3.68E-39 | InN4 | OIP5-AS1 |
| C1orf211 | 1.01E-43 | 0.30623757 | 0.735 | 0.719 | 3.87E-39 | InN4 | C1orf21 |
| CC2D2A2 | 1.64E-43 | 0.35975243 | 0.363 | 0.265 | 6.25E-39 | InN4 | CC2D2A |
| GNAL3 | 3.04E-43 | 0.33707504 | 0.629 | 0.473 | 1.16E-38 | InN4 | GNAL |
| LINC015051 | 2.31E-42 | 0.38379977 | 0.317 | 0.214 | 8.83E-38 | InN4 | LINC01505 |
| PGAP1 | 9.34E-42 | 0.36759559 | 0.523 | 0.454 | 3.57E-37 | InN4 | PGAP1 |
| BCL22 | 9.35E-42 | 0.3916524 | 0.603 | 0.524 | 3.57E-37 | InN4 | BCL2 |
| AC004083.11 | 1.48E-41 | 0.37375891 | 0.397 | 0.298 | 5.64E-37 | InN4 | AC004083.1 |
| PRKN3 | 3.42E-41 | 0.25944152 | 0.854 | 0.864 | 1.31E-36 | InN4 | PRKN |
| NEDD4L2 | 1.14E-40 | 0.27012229 | 0.851 | 0.813 | 4.35E-36 | InN4 | NEDD4L |
| DIAPH22 | 1.59E-40 | 0.28858265 | 0.705 | 0.608 | 6.06E-36 | InN4 | DIAPH2 |
| CRIM13 | 1.85E-40 | 0.37942734 | 0.573 | 0.487 | 7.08E-36 | InN4 | CRIM1 |
| DAAM12 | 2.38E-40 | 0.29952373 | 0.757 | 0.751 | 9.11E-36 | InN4 | DAAM1 |
| GTF2I1 | 2.59E-40 | 0.25156651 | 0.822 | 0.835 | 9.88E-36 | InN4 | GTF2I |
| PELI21 | 5.94E-40 | 0.30876709 | 0.393 | 0.281 | 2.27E-35 | InN4 | PELI2 |
| APC3 | 1.34E-39 | 0.30443055 | 0.743 | 0.742 | 5.14E-35 | InN4 | APC |
| B3GLCT2 | 1.95E-39 | 0.61857513 | 0.374 | 0.296 | 7.44E-35 | InN4 | B3GLCT |
| AC007368.13 | 4.53E-39 | 0.36180685 | 0.55 | 0.438 | 1.73E-34 | InN4 | AC007368.1 |
| SH3BGRL1 | 4.64E-39 | 0.33931008 | 0.542 | 0.474 | 1.77E-34 | InN4 | SH3BGRL |
| CRB12 | 7.84E-39 | 0.29318929 | 0.417 | 0.31 | 3.00E-34 | InN4 | CRB1 |
| GREB13 | 1.62E-38 | 0.30275258 | 0.432 | 0.337 | 6.21E-34 | InN4 | GREB1 |
| ST3GAL5 | 2.69E-38 | 0.36602722 | 0.5 | 0.43 | 1.03E-33 | InN4 | ST3GAL5 |
| CDR2 | 3.94E-38 | 0.32525727 | 0.618 | 0.554 | 1.51E-33 | InN4 | CDR2 |
| PHACTR23 | 5.90E-38 | 0.28389226 | 0.508 | 0.406 | 2.25E-33 | InN4 | PHACTR2 |
| USP311 | 8.38E-38 | 0.39690318 | 0.49 | 0.422 | 3.20E-33 | InN4 | USP31 |
| SEM11 | 1.02E-37 | 0.38806657 | 0.545 | 0.48 | 3.89E-33 | InN4 | SEM1 |
| RFC1 | 1.06E-37 | 0.33059352 | 0.386 | 0.298 | 4.04E-33 | InN4 | RFC1 |
| FAM168A | 2.48E-37 | 0.29816534 | 0.766 | 0.764 | 9.47E-33 | InN4 | FAM168A |
| AFF22 | 3.34E-37 | 0.30918276 | 0.485 | 0.387 | 1.28E-32 | InN4 | AFF2 |
| C1orf61 | 3.99E-37 | 0.3312133 | 0.413 | 0.326 | 1.53E-32 | InN4 | C1orf61 |
| CEP164 | 4.84E-37 | 0.35992192 | 0.453 | 0.374 | 1.85E-32 | InN4 | CEP164 |
| ZNF8272 | 4.85E-37 | 0.36158348 | 0.549 | 0.483 | 1.85E-32 | InN4 | ZNF827 |
| MXRA7 | 6.81E-37 | 0.29348181 | 0.346 | 0.253 | 2.60E-32 | InN4 | MXRA7 |
| PTPRS1 | 2.19E-36 | 0.3104007 | 0.736 | 0.714 | 8.36E-32 | InN4 | PTPRS |
| BIRC6 | 4.97E-36 | 0.28734382 | 0.805 | 0.817 | 1.90E-31 | InN4 | BIRC6 |
| NR3C21 | 8.59E-36 | 0.27735761 | 0.817 | 0.805 | 3.28E-31 | InN4 | NR3C2 |
| AC098829.1 | 1.34E-35 | 0.46789449 | 0.327 | 0.238 | 5.12E-31 | InN4 | AC098829.1 |
| FAM49A | 1.40E-35 | 0.33548511 | 0.724 | 0.716 | 5.36E-31 | InN4 | FAM49A |
| ATP1B12 | 1.93E-35 | 0.28417418 | 0.779 | 0.78 | 7.39E-31 | InN4 | ATP1B1 |
| ZSWIM52 | 2.22E-35 | 0.32773376 | 0.597 | 0.544 | 8.49E-31 | InN4 | ZSWIM5 |

| | | | | | | | |
|---|---|---|---|---|---|---|---|
| SSH11 | 2.46E-35 | 0.36377048 | 0.494 | 0.43 | 9.42E-31 | InN4 | SSH1 |
| SREBF21 | 3.99E-35 | 0.35093884 | 0.615 | 0.581 | 1.53E-30 | InN4 | SREBF2 |
| PLPP1 | 4.36E-35 | 0.37083071 | 0.577 | 0.534 | 1.67E-30 | InN4 | PLPP1 |
| EFCAB22 | 9.22E-35 | 0.34304433 | 0.576 | 0.515 | 3.52E-30 | InN4 | EFCAB2 |
| ZSWIM61 | 2.52E-34 | 0.33901392 | 0.745 | 0.73 | 9.63E-30 | InN4 | ZSWIM6 |
| SPOPL | 8.74E-34 | 0.33558739 | 0.42 | 0.345 | 3.34E-29 | InN4 | SPOPL |
| L3MBTL11 | 1.04E-33 | 0.31174916 | 0.448 | 0.367 | 3.99E-29 | InN4 | L3MBTL1 |
| LIMA11 | 6.48E-33 | 0.25846521 | 0.288 | 0.201 | 2.48E-28 | InN4 | LIMA1 |
| WSB11 | 2.03E-32 | 0.3305616 | 0.629 | 0.626 | 7.76E-28 | InN4 | WSB1 |
| DENND5A2 | 3.31E-32 | 0.3220855 | 0.53 | 0.467 | 1.26E-27 | InN4 | DENND5A |
| ANKRD262 | 9.52E-32 | 0.32082865 | 0.752 | 0.754 | 3.64E-27 | InN4 | ANKRD26 |
| ARHGAP51 | 3.79E-31 | 0.30735506 | 0.623 | 0.591 | 1.45E-26 | InN4 | ARHGAP5 |
| NAAA1 | 3.82E-31 | 0.26191704 | 0.293 | 0.209 | 1.46E-26 | InN4 | NAAA |
| FUT92 | 3.89E-31 | 0.27848504 | 0.819 | 0.817 | 1.49E-26 | InN4 | FUT9 |
| ELAVL3 | 4.64E-31 | 0.2984984 | 0.774 | 0.776 | 1.77E-26 | InN4 | ELAVL3 |
| INSR1 | 4.88E-31 | 0.32097837 | 0.685 | 0.663 | 1.86E-26 | InN4 | INSR |
| TACC22 | 7.39E-31 | 0.29546968 | 0.479 | 0.411 | 2.83E-26 | InN4 | TACC2 |
| MT-ND32 | 2.24E-30 | 0.49862598 | 0.866 | 0.871 | 8.55E-26 | InN4 | MT-ND3 |
| SRGAP24 | 2.83E-30 | 0.27762109 | 0.54 | 0.483 | 1.08E-25 | InN4 | SRGAP2 |
| FYN | 6.02E-30 | 0.27734494 | 0.746 | 0.749 | 2.30E-25 | InN4 | FYN |
| CHD9 | 7.25E-30 | 0.25212507 | 0.825 | 0.839 | 2.77E-25 | InN4 | CHD9 |
| GABRD1 | 7.92E-30 | 0.31699426 | 0.335 | 0.26 | 3.03E-25 | InN4 | GABRD |
| CCDC144A1 | 1.79E-29 | 0.32658463 | 0.704 | 0.658 | 6.86E-25 | InN4 | CCDC144A |
| ITSN11 | 2.30E-29 | 0.29020731 | 0.743 | 0.752 | 8.78E-25 | InN4 | ITSN1 |
| PPP1R2 | 3.93E-29 | 0.29162979 | 0.347 | 0.275 | 1.50E-24 | InN4 | PPP1R2 |
| TRAF3 | 8.02E-29 | 0.31892668 | 0.572 | 0.526 | 3.06E-24 | InN4 | TRAF3 |
| AC073332.12 | 1.02E-28 | 0.4224328 | 0.403 | 0.326 | 3.89E-24 | InN4 | AC073332.1 |
| AL357054.2 | 1.38E-28 | 0.27887125 | 0.294 | 0.216 | 5.27E-24 | InN4 | AL357054.2 |
| ABAT2 | 1.64E-28 | 0.29800614 | 0.549 | 0.502 | 6.26E-24 | InN4 | ABAT |
| ADCY51 | 2.76E-28 | 0.28336083 | 0.52 | 0.46 | 1.05E-23 | InN4 | ADCY5 |
| APBB22 | 5.74E-28 | 0.27527459 | 0.701 | 0.691 | 2.19E-23 | InN4 | APBB2 |
| PDCD4 | 6.75E-28 | 0.2927623 | 0.419 | 0.351 | 2.58E-23 | InN4 | PDCD4 |
| NAALADL21 | 2.16E-27 | 0.34675286 | 0.572 | 0.521 | 8.26E-23 | InN4 | NAALADL2 |
| ANKMY1 | 2.29E-27 | 0.25809394 | 0.287 | 0.212 | 8.74E-23 | InN4 | ANKMY1 |
| REPS1 | 2.48E-27 | 0.29788107 | 0.635 | 0.642 | 9.46E-23 | InN4 | REPS1 |
| ZNF804B4 | 7.43E-27 | 0.45363212 | 0.441 | 0.356 | 2.84E-22 | InN4 | ZNF804B |
| GTF2IRD12 | 1.33E-26 | 0.31364495 | 0.517 | 0.454 | 5.10E-22 | InN4 | GTF2IRD1 |
| TUT42 | 1.50E-26 | 0.26722909 | 0.691 | 0.703 | 5.73E-22 | InN4 | TUT4 |
| GPR631 | 2.33E-26 | 0.28152878 | 0.322 | 0.251 | 8.89E-22 | InN4 | GPR63 |
| SLC4A31 | 4.38E-26 | 0.25374925 | 0.28 | 0.205 | 1.67E-21 | InN4 | SLC4A3 |
| AC009899.11 | 1.26E-25 | 0.31604502 | 0.549 | 0.497 | 4.83E-21 | InN4 | AC009899.1 |
| PKD1L12 | 1.29E-25 | 0.25377246 | 0.349 | 0.264 | 4.93E-21 | InN4 | PKD1L1 |
| SLC43A2 | 2.68E-25 | 0.3137063 | 0.363 | 0.293 | 1.02E-20 | InN4 | SLC43A2 |
| KIAA0895 | 4.99E-25 | 0.26723701 | 0.321 | 0.251 | 1.91E-20 | InN4 | KIAA0895 |
| RALGAPA22 | 5.62E-25 | 0.27566393 | 0.621 | 0.591 | 2.15E-20 | InN4 | RALGAPA2 |
| ATAD2B1 | 7.92E-25 | 0.29877156 | 0.489 | 0.437 | 3.03E-20 | InN4 | ATAD2B |
| ANKS1A2 | 8.28E-25 | 0.30126838 | 0.618 | 0.597 | 3.16E-20 | InN4 | ANKS1A |
| PER3 | 8.31E-25 | 0.295137 | 0.538 | 0.491 | 3.18E-20 | InN4 | PER3 |
| BAZ2B1 | 1.58E-24 | 0.28033163 | 0.713 | 0.711 | 6.05E-20 | InN4 | BAZ2B |
| RNF217 | 2.35E-24 | 0.29800984 | 0.452 | 0.395 | 8.98E-20 | InN4 | RNF217 |
| GNG74 | 2.98E-24 | 0.29645612 | 0.477 | 0.407 | 1.14E-19 | InN4 | GNG7 |
| SEPTIN62 | 3.62E-24 | 0.25189517 | 0.367 | 0.289 | 1.38E-19 | InN4 | SEPTIN6 |
| SLC5A31 | 3.94E-24 | 0.25644262 | 0.368 | 0.294 | 1.51E-19 | InN4 | SLC5A3 |
| VPS45 | 4.78E-24 | 0.30945928 | 0.589 | 0.575 | 1.83E-19 | InN4 | VPS45 |
| NFIC1 | 5.14E-24 | 0.30104777 | 0.556 | 0.512 | 1.96E-19 | InN4 | NFIC |
| UBAC2 | 5.57E-24 | 0.28224053 | 0.621 | 0.618 | 2.13E-19 | InN4 | UBAC2 |
| HSP90AA12 | 7.62E-24 | 0.48213127 | 0.722 | 0.712 | 2.91E-19 | InN4 | HSP90AA1 |
| NR6A1 | 9.36E-24 | 0.32789281 | 0.591 | 0.567 | 3.58E-19 | InN4 | NR6A1 |
| SLC1A1 | 3.81E-23 | 0.27671227 | 0.451 | 0.392 | 1.46E-18 | InN4 | SLC1A1 |
| NME91 | 1.11E-22 | 0.35655446 | 0.392 | 0.336 | 4.25E-18 | InN4 | NME9 |
| TMEM50B | 1.43E-22 | 0.26955441 | 0.363 | 0.301 | 5.46E-18 | InN4 | TMEM50B |
| ELOVL5 | 2.55E-22 | 0.38108579 | 0.341 | 0.274 | 9.74E-18 | InN4 | ELOVL5 |
| PCDH11Y2 | 2.81E-22 | 0.40147608 | 0.294 | 0.225 | 1.07E-17 | InN4 | PCDH11Y |

| | | | | | | | |
|---|---|---|---|---|---|---|---|
| SUSD41 | 2.87E-22 | 0.28967903 | 0.633 | 0.611 | 1.10E-17 | InN4 | SUSD4 |
| CHCHD6 | 4.89E-22 | 0.29538055 | 0.509 | 0.479 | 1.87E-17 | InN4 | CHCHD6 |
| ZBTB11 | 6.00E-22 | 0.27878843 | 0.428 | 0.374 | 2.29E-17 | InN4 | ZBTB1 |
| SEPTIN71 | 6.08E-22 | 0.25324349 | 0.687 | 0.695 | 2.32E-17 | InN4 | SEPTIN7 |
| MAMLD11 | 2.40E-21 | 0.30116884 | 0.369 | 0.307 | 9.19E-17 | InN4 | MAMLD1 |
| DPYSL51 | 2.91E-21 | 0.2641551 | 0.408 | 0.347 | 1.11E-16 | InN4 | DPYSL5 |
| DOCK7 | 4.59E-21 | 0.26543686 | 0.559 | 0.538 | 1.75E-16 | InN4 | DOCK7 |
| SYNDIG13 | 4.76E-21 | 0.34071608 | 0.615 | 0.599 | 1.82E-16 | InN4 | SYNDIG1 |
| MGAT53 | 4.79E-21 | 0.25546183 | 0.676 | 0.661 | 1.83E-16 | InN4 | MGAT5 |
| GRK5 | 5.71E-21 | 0.31712285 | 0.342 | 0.28 | 2.18E-16 | InN4 | GRK5 |
| CD99L2 | 6.08E-21 | 0.29953461 | 0.465 | 0.424 | 2.32E-16 | InN4 | CD99L2 |
| TRAPPC9 | 6.43E-21 | 0.28842663 | 0.641 | 0.639 | 2.46E-16 | InN4 | TRAPPC9 |
| RNF1572 | 7.47E-21 | 0.26598284 | 0.555 | 0.529 | 2.85E-16 | InN4 | RNF157 |
| AGPS | 8.47E-21 | 0.27303701 | 0.453 | 0.414 | 3.24E-16 | InN4 | AGPS |
| PDK31 | 9.69E-21 | 0.28202573 | 0.517 | 0.474 | 3.70E-16 | InN4 | PDK3 |
| MT-ND41 | 1.61E-20 | 0.4482222 | 0.843 | 0.858 | 6.15E-16 | InN4 | MT-ND4 |
| LRRN22 | 1.87E-20 | 0.2821757 | 0.553 | 0.519 | 7.17E-16 | InN4 | LRRN2 |
| GABPB2 | 2.16E-20 | 0.29479139 | 0.377 | 0.317 | 8.26E-16 | InN4 | GABPB2 |
| FAM214A1 | 2.65E-20 | 0.2805984 | 0.563 | 0.541 | 1.01E-15 | InN4 | FAM214A |
| XPO1 | 8.69E-20 | 0.33154614 | 0.623 | 0.618 | 3.32E-15 | InN4 | XPO1 |
| RRAS2 | 1.03E-19 | 0.25517403 | 0.354 | 0.294 | 3.96E-15 | InN4 | RRAS2 |
| PIP4K2A3 | 1.12E-19 | 0.2984167 | 0.476 | 0.426 | 4.29E-15 | InN4 | PIP4K2A |
| EPHA53 | 1.31E-19 | 0.27169706 | 0.638 | 0.599 | 5.02E-15 | InN4 | EPHA5 |
| LGI1 | 1.41E-19 | 0.31018795 | 0.477 | 0.441 | 5.41E-15 | InN4 | LGI1 |
| MRPS6 | 2.04E-19 | 0.2532196 | 0.361 | 0.3 | 7.79E-15 | InN4 | MRPS6 |
| PRMT8 | 2.05E-19 | 0.30368007 | 0.468 | 0.423 | 7.82E-15 | InN4 | PRMT8 |
| EXOSC10 | 2.63E-19 | 0.28314889 | 0.368 | 0.315 | 1.00E-14 | InN4 | EXOSC10 |
| SVOP1 | 3.21E-19 | 0.29041868 | 0.537 | 0.508 | 1.23E-14 | InN4 | SVOP |
| NR2F1 | 4.10E-19 | 0.30049022 | 0.451 | 0.404 | 1.57E-14 | InN4 | NR2F1 |
| OLFM22 | 6.08E-19 | 0.25363374 | 0.368 | 0.296 | 2.33E-14 | InN4 | OLFM2 |
| ZNF832 | 1.33E-18 | 0.25917324 | 0.382 | 0.329 | 5.07E-14 | InN4 | ZNF83 |
| SH3PXD2A2 | 1.70E-18 | 0.25860975 | 0.696 | 0.705 | 6.48E-14 | InN4 | SH3PXD2A |
| MT-ND11 | 2.03E-18 | 0.35490809 | 0.789 | 0.782 | 7.76E-14 | InN4 | MT-ND1 |
| DNM2 | 2.18E-18 | 0.3027293 | 0.482 | 0.447 | 8.33E-14 | InN4 | DNM2 |
| ALOX12-AS1 | 6.05E-18 | 0.2958929 | 0.454 | 0.414 | 2.31E-13 | InN4 | ALOX12-AS1 |
| TMEM131L1 | 9.21E-18 | 0.25164465 | 0.396 | 0.338 | 3.52E-13 | InN4 | TMEM131L |
| UBA6-AS11 | 9.97E-18 | 0.26320671 | 0.625 | 0.605 | 3.81E-13 | InN4 | UBA6-AS1 |
| APLP2 | 1.05E-17 | 0.29750837 | 0.591 | 0.597 | 4.01E-13 | InN4 | APLP2 |
| AP2A2 | 1.09E-17 | 0.26439756 | 0.577 | 0.568 | 4.18E-13 | InN4 | AP2A2 |
| FLNB | 3.70E-17 | 0.26898241 | 0.487 | 0.454 | 1.41E-12 | InN4 | FLNB |
| PITPNM21 | 3.88E-17 | 0.26425176 | 0.577 | 0.559 | 1.48E-12 | InN4 | PITPNM2 |
| HLF2 | 5.15E-17 | 0.29058086 | 0.534 | 0.514 | 1.97E-12 | InN4 | HLF |
| LSS | 5.85E-17 | 0.26253201 | 0.331 | 0.278 | 2.24E-12 | InN4 | LSS |
| GABRA4 | 9.50E-17 | 0.26181569 | 0.619 | 0.623 | 3.63E-12 | InN4 | GABRA4 |
| FDFT1 | 1.38E-16 | 0.27006629 | 0.345 | 0.297 | 5.27E-12 | InN4 | FDFT1 |
| PTPRO1 | 2.24E-16 | 0.2615442 | 0.739 | 0.679 | 8.58E-12 | InN4 | PTPRO |
| RYBP | 3.28E-16 | 0.25182775 | 0.477 | 0.437 | 1.25E-11 | InN4 | RYBP |
| MT-ND4L1 | 3.72E-16 | 0.37796696 | 0.575 | 0.545 | 1.42E-11 | InN4 | MT-ND4L |
| FAIM2 | 6.05E-16 | 0.25655299 | 0.664 | 0.677 | 2.31E-11 | InN4 | FAIM2 |
| RAD21 | 7.48E-16 | 0.25239631 | 0.411 | 0.373 | 2.86E-11 | InN4 | RAD21 |
| CAMKMT2 | 7.56E-16 | 0.28598707 | 0.425 | 0.381 | 2.89E-11 | InN4 | CAMKMT |
| AC027288.31 | 8.03E-16 | 0.28514828 | 0.304 | 0.249 | 3.07E-11 | InN4 | AC027288.3 |
| MTRNR2L12 | 1.42E-15 | 0.2542449 | 0.752 | 0.748 | 5.43E-11 | InN4 | MTRNR2L12 |
| MT-ND22 | 3.59E-15 | 0.30899475 | 0.749 | 0.75 | 1.37E-10 | InN4 | MT-ND2 |
| SKI1 | 4.81E-15 | 0.252051 | 0.489 | 0.464 | 1.84E-10 | InN4 | SKI |
| HMGCS1 | 4.90E-15 | 0.35578219 | 0.358 | 0.315 | 1.87E-10 | InN4 | HMGCS1 |
| PLCG23 | 5.27E-15 | 0.3924991 | 0.679 | 0.651 | 2.01E-10 | InN4 | PLCG2 |
| LINC009371 | 2.13E-14 | 0.25178097 | 0.337 | 0.287 | 8.13E-10 | InN4 | LINC00937 |
| TET22 | 2.42E-14 | 0.25249045 | 0.471 | 0.438 | 9.26E-10 | InN4 | TET2 |
| DPH61 | 4.30E-14 | 0.25712634 | 0.451 | 0.416 | 1.64E-09 | InN4 | DPH6 |
| NCS1 | 4.70E-14 | 0.25128301 | 0.507 | 0.486 | 1.80E-09 | InN4 | NCS1 |
| AMN1 | 7.58E-14 | 0.27198403 | 0.439 | 0.408 | 2.90E-09 | InN4 | AMN1 |
| COLQ | 1.83E-13 | 0.25415875 | 0.443 | 0.414 | 6.99E-09 | InN4 | COLQ |

| | | | | | | | |
|---|---|---|---|---|---|---|---|
| IQCK | 4.54E-13 | 0.26964408 | 0.345 | 0.306 | 1.73E-08 | InN4 | IQCK |
| SPANXA2-OT | 7.89E-13 | 0.3559112 | 0.308 | 0.264 | 3.02E-08 | InN4 | SPANXA2-OT |
| AC098588.3 | 9.87E-13 | 0.28938219 | 0.565 | 0.547 | 3.77E-08 | InN4 | AC098588.3 |
| PRKD12 | 1.01E-12 | 0.42381824 | 0.517 | 0.514 | 3.86E-08 | InN4 | PRKD1 |
| MT-CO22 | 6.82E-12 | 0.26991665 | 0.891 | 0.899 | 2.61E-07 | InN4 | MT-CO2 |
| MT-ND51 | 7.78E-12 | 0.34588927 | 0.579 | 0.57 | 2.97E-07 | InN4 | MT-ND5 |
| AL627171.22 | 1.20E-11 | 0.43331638 | 0.482 | 0.453 | 4.60E-07 | InN4 | AL627171.2 |
| LINC02245 | 2.08E-11 | 0.27991862 | 0.508 | 0.495 | 7.96E-07 | InN4 | LINC02245 |
| MT-CYB2 | 1.35E-10 | 0.33020936 | 0.777 | 0.788 | 5.14E-06 | InN4 | MT-CYB |
| MT-ATP62 | 1.38E-10 | 0.34290671 | 0.862 | 0.881 | 5.28E-06 | InN4 | MT-ATP6 |
| AC119673.2 | 2.85E-10 | 0.29419171 | 0.37 | 0.349 | 1.09E-05 | InN4 | AC119673.2 |
| NECTIN31 | 7.65E-08 | 0.25475702 | 0.496 | 0.484 | 0.00292312 | InN4 | NECTIN3 |
| MRPL411 | 9.47E-08 | 0.25294521 | 0.45 | 0.446 | 0.00362126 | InN4 | MRPL41 |
| CXCL142 | 0 | 3.39736596 | 0.885 | 0.125 | 0 | InN5 | CXCL14 |
| RELN1 | 0 | 3.0564758 | 0.514 | 0.121 | 0 | InN5 | RELN |
| THSD7B1 | 0 | 2.81762737 | 0.754 | 0.137 | 0 | InN5 | THSD7B |
| SGO1-AS11 | 0 | 2.55056752 | 0.551 | 0.119 | 0 | InN5 | SGO1-AS1 |
| ADARB22 | 0 | 2.26530898 | 0.999 | 0.345 | 0 | InN5 | ADARB2 |
| FSTL52 | 0 | 2.19864844 | 0.91 | 0.485 | 0 | InN5 | FSTL5 |
| PLD52 | 0 | 2.10603747 | 0.91 | 0.3 | 0 | InN5 | PLD5 |
| INPP4B4 | 0 | 1.9464761 | 0.935 | 0.748 | 0 | InN5 | INPP4B |
| DLX6-AS12 | 0 | 1.90532147 | 0.753 | 0.22 | 0 | InN5 | DLX6-AS1 |
| HS3ST52 | 0 | 1.84536106 | 0.693 | 0.301 | 0 | InN5 | HS3ST5 |
| ERBB43 | 0 | 1.82912263 | 0.997 | 0.56 | 0 | InN5 | ERBB4 |
| SDK14 | 0 | 1.815611 | 0.882 | 0.464 | 0 | InN5 | SDK1 |
| EGFR2 | 0 | 1.81166387 | 0.83 | 0.218 | 0 | InN5 | EGFR |
| ADRA1A3 | 0 | 1.802236 | 0.82 | 0.188 | 0 | InN5 | ADRA1A |
| PRELID21 | 0 | 1.71055304 | 0.82 | 0.395 | 0 | InN5 | PRELID2 |
| CNTNAP43 | 0 | 1.69165668 | 0.896 | 0.572 | 0 | InN5 | CNTNAP4 |
| RGS121 | 0 | 1.67828498 | 0.949 | 0.5 | 0 | InN5 | RGS12 |
| NR2F2-AS11 | 0 | 1.67461401 | 0.796 | 0.186 | 0 | InN5 | NR2F2-AS1 |
| CNTN54 | 0 | 1.64159024 | 0.97 | 0.81 | 0 | InN5 | CNTN5 |
| AC132803.11 | 0 | 1.63340635 | 0.654 | 0.16 | 0 | InN5 | AC132803.1 |
| GRIK22 | 0 | 1.63045721 | 0.991 | 0.947 | 0 | InN5 | GRIK2 |
| DOCK103 | 0 | 1.62990614 | 0.906 | 0.385 | 0 | InN5 | DOCK10 |
| SLC25A482 | 0 | 1.62091333 | 0.706 | 0.098 | 0 | InN5 | SLC25A48 |
| C8orf342 | 0 | 1.59432544 | 0.831 | 0.374 | 0 | InN5 | C8orf34 |
| ZNF385D5 | 0 | 1.57443311 | 0.872 | 0.501 | 0 | InN5 | ZNF385D |
| RPH3A2 | 0 | 1.54251255 | 0.88 | 0.494 | 0 | InN5 | RPH3A |
| NCAM21 | 0 | 1.48035756 | 0.986 | 0.873 | 0 | InN5 | NCAM2 |
| GALNTL63 | 0 | 1.46331276 | 0.949 | 0.735 | 0 | InN5 | GALNTL6 |
| CHST113 | 0 | 1.4345049 | 0.855 | 0.501 | 0 | InN5 | CHST11 |
| PROX12 | 0 | 1.42229868 | 0.716 | 0.15 | 0 | InN5 | PROX1 |
| CNR11 | 0 | 1.39661592 | 0.71 | 0.371 | 0 | InN5 | CNR1 |
| COL21A11 | 0 | 1.39418445 | 0.639 | 0.178 | 0 | InN5 | COL21A1 |
| CHRNA71 | 0 | 1.39291534 | 0.627 | 0.161 | 0 | InN5 | CHRNA7 |
| AL136114.11 | 0 | 1.3782123 | 0.53 | 0.088 | 0 | InN5 | AL136114.1 |
| NPAS32 | 0 | 1.37527363 | 0.956 | 0.813 | 0 | InN5 | NPAS3 |
| NR2F21 | 0 | 1.33773716 | 0.608 | 0.093 | 0 | InN5 | NR2F2 |
| MEGF111 | 0 | 1.32142672 | 0.555 | 0.129 | 0 | InN5 | MEGF11 |
| DNER3 | 0 | 1.31454587 | 0.85 | 0.516 | 0 | InN5 | DNER |
| TOX34 | 0 | 1.30886354 | 0.602 | 0.173 | 0 | InN5 | TOX3 |
| ZNF5363 | 0 | 1.301698 | 0.927 | 0.352 | 0 | InN5 | ZNF536 |
| HPSE21 | 0 | 1.28762054 | 0.42 | 0.099 | 0 | InN5 | HPSE2 |
| GAD23 | 0 | 1.26615565 | 0.729 | 0.241 | 0 | InN5 | GAD2 |
| SCG21 | 0 | 1.2632471 | 0.59 | 0.249 | 0 | InN5 | SCG2 |
| GRIK12 | 0 | 1.21440236 | 0.793 | 0.377 | 0 | InN5 | GRIK1 |
| CCNH2 | 0 | 1.20745165 | 0.893 | 0.763 | 0 | InN5 | CCNH |
| BTBD112 | 0 | 1.16151559 | 0.796 | 0.253 | 0 | InN5 | BTBD11 |
| DAB14 | 0 | 1.15613902 | 0.952 | 0.877 | 0 | InN5 | DAB1 |
| PTPRM3 | 0 | 1.1457759 | 0.848 | 0.574 | 0 | InN5 | PTPRM |
| PLPPR14 | 0 | 1.13887358 | 0.643 | 0.283 | 0 | InN5 | PLPPR1 |
| AL161629.1 | 0 | 1.12688482 | 0.426 | 0.022 | 0 | InN5 | AL161629.1 |

| | | | | | | | |
|---|---|---|---|---|---|---|---|
| RERG2 | 0 | 1.12539845 | 0.66 | 0.254 | 0 | InN5 | RERG |
| FAT41 | 0 | 1.11503186 | 0.551 | 0.145 | 0 | InN5 | FAT4 |
| HDAC94 | 0 | 1.10933181 | 0.911 | 0.774 | 0 | InN5 | HDAC9 |
| SPOCK32 | 0 | 1.09230619 | 0.916 | 0.541 | 0 | InN5 | SPOCK3 |
| PEX5L3 | 0 | 1.08271532 | 0.938 | 0.825 | 0 | InN5 | PEX5L |
| RAB3C3 | 0 | 1.06332567 | 0.857 | 0.63 | 0 | InN5 | RAB3C |
| AC233296.12 | 0 | 1.05958055 | 0.625 | 0.255 | 0 | InN5 | AC233296.1 |
| GNG121 | 0 | 1.01350755 | 0.39 | 0.061 | 0 | InN5 | GNG12 |
| PWRN14 | 0 | 0.99924767 | 0.816 | 0.457 | 0 | InN5 | PWRN1 |
| CALB21 | 0 | 0.98349538 | 0.481 | 0.109 | 0 | InN5 | CALB2 |
| DISC11 | 0 | 0.95612531 | 0.432 | 0.127 | 0 | InN5 | DISC1 |
| GAD14 | 0 | 0.95165737 | 0.688 | 0.273 | 0 | InN5 | GAD1 |
| PREX22 | 0 | 0.94419305 | 0.511 | 0.165 | 0 | InN5 | PREX2 |
| ARHGAP421 | 0 | 0.91630046 | 0.401 | 0.078 | 0 | InN5 | ARHGAP42 |
| TAFA41 | 0 | 0.90603146 | 0.273 | 0.059 | 0 | InN5 | TAFA4 |
| BMP6 | 0 | 0.89573754 | 0.327 | 0.038 | 0 | InN5 | BMP6 |
| AL391832.41 | 0 | 0.89468336 | 0.393 | 0.054 | 0 | InN5 | AL391832.4 |
| PWRN43 | 0 | 0.89367507 | 0.552 | 0.213 | 0 | InN5 | PWRN4 |
| AC137770.11 | 0 | 0.88507951 | 0.415 | 0.1 | 0 | InN5 | AC137770.1 |
| EPS82 | 0 | 0.86715244 | 0.416 | 0.11 | 0 | InN5 | EPS8 |
| NPAS11 | 0 | 0.84801875 | 0.464 | 0.134 | 0 | InN5 | NPAS1 |
| KAZN4 | 0 | 0.84125242 | 0.994 | 0.952 | 0 | InN5 | KAZN |
| SNCAIP2 | 0 | 0.83992174 | 0.446 | 0.151 | 0 | InN5 | SNCAIP |
| CNTNAP23 | 0 | 0.83167534 | 0.996 | 0.993 | 0 | InN5 | CNTNAP2 |
| SMOC11 | 0 | 0.80755425 | 0.333 | 0.081 | 0 | InN5 | SMOC1 |
| IGF14 | 0 | 0.74797064 | 0.446 | 0.141 | 0 | InN5 | IGF1 |
| NDNF | 0 | 0.73851474 | 0.264 | 0.007 | 0 | InN5 | NDNF |
| TCF44 | 0 | 0.73309568 | 0.98 | 0.957 | 0 | InN5 | TCF4 |
| AC009093.21 | 0 | 0.73287997 | 0.407 | 0.124 | 0 | InN5 | AC009093.2 |
| PROX1-AS12 | 0 | 0.71267571 | 0.387 | 0.104 | 0 | InN5 | PROX1-AS1 |
| RXRG | 0 | 0.64032124 | 0.25 | 0.024 | 0 | InN5 | RXRG |
| MN1 | 0 | 0.61074753 | 0.343 | 0.082 | 0 | InN5 | MN1 |
| DLX12 | 0 | 0.60155027 | 0.346 | 0.09 | 0 | InN5 | DLX1 |
| KMO | 0 | 0.58803413 | 0.269 | 0.055 | 0 | InN5 | KMO |
| PNOC | 0 | 0.5638335 | 0.274 | 0.033 | 0 | InN5 | PNOC |
| GCNT22 | 0 | 0.53198506 | 0.308 | 0.066 | 0 | InN5 | GCNT2 |
| NR2E1 | 0 | 0.51612051 | 0.277 | 0.035 | 0 | InN5 | NR2E1 |
| SDK22 | 1.28E-307 | 0.94391122 | 0.544 | 0.221 | 4.89E-303 | InN5 | SDK2 |
| PRKCA4 | 8.06E-303 | 0.95182245 | 0.903 | 0.73 | 3.08E-298 | InN5 | PRKCA |
| PARD33 | 1.13E-301 | 0.85740183 | 0.686 | 0.307 | 4.33E-297 | InN5 | PARD3 |
| PLS31 | 1.51E-290 | 0.82009684 | 0.493 | 0.19 | 5.76E-286 | InN5 | PLS3 |
| SLC6A15 | 1.07E-286 | 0.9624163 | 0.721 | 0.342 | 4.10E-282 | InN5 | SLC6A1 |
| LINC004721 | 2.55E-286 | 0.73976488 | 0.42 | 0.139 | 9.76E-282 | InN5 | LINC00472 |
| L3MBTL42 | 1.06E-278 | 1.03222827 | 0.711 | 0.389 | 4.05E-274 | InN5 | L3MBTL4 |
| AFF23 | 1.26E-277 | 0.97929981 | 0.675 | 0.379 | 4.82E-273 | InN5 | AFF2 |
| ARHGEF26-A | 2.95E-271 | 0.71953365 | 0.39 | 0.126 | 1.13E-266 | InN5 | ARHGEF26-A |
| ADAMTS9-AS | 1.55E-267 | 1.57563183 | 0.639 | 0.335 | 5.91E-263 | InN5 | ADAMTS9-AS |
| KCNIP13 | 3.81E-264 | 0.69510605 | 0.673 | 0.292 | 1.46E-259 | InN5 | KCNIP1 |
| PRR161 | 2.72E-263 | 1.1069939 | 0.761 | 0.543 | 1.04E-258 | InN5 | PRR16 |
| AC087564.13 | 1.30E-257 | 0.66735095 | 0.41 | 0.131 | 4.96E-253 | InN5 | AC087564.1 |
| NINL3 | 2.79E-255 | 0.67080495 | 0.466 | 0.177 | 1.06E-250 | InN5 | NINL |
| SEMA3C1 | 5.38E-254 | 0.83574949 | 0.38 | 0.131 | 2.06E-249 | InN5 | SEMA3C |
| PCSK53 | 3.08E-253 | 0.67946518 | 0.496 | 0.19 | 1.18E-248 | InN5 | PCSK5 |
| SLC44A54 | 6.14E-253 | 0.89652489 | 0.877 | 0.688 | 2.35E-248 | InN5 | SLC44A5 |
| PITPNC14 | 1.11E-249 | 0.87465083 | 0.774 | 0.484 | 4.24E-245 | InN5 | PITPNC1 |
| THSD7A2 | 1.40E-243 | 1.22109779 | 0.729 | 0.501 | 5.37E-239 | InN5 | THSD7A |
| LUZP23 | 2.49E-242 | 0.99121791 | 0.617 | 0.299 | 9.50E-238 | InN5 | LUZP2 |
| GRIN2D4 | 1.87E-240 | 0.58438018 | 0.396 | 0.135 | 7.16E-236 | InN5 | GRIN2D |
| GNG12-AS1 | 3.74E-239 | 0.87074509 | 0.302 | 0.088 | 1.43E-234 | InN5 | GNG12-AS1 |
| AMOTL13 | 8.30E-237 | 0.65806874 | 0.479 | 0.194 | 3.17E-232 | InN5 | AMOTL1 |
| NDST41 | 2.27E-236 | 0.61863573 | 0.255 | 0.063 | 8.67E-232 | InN5 | NDST4 |
| GRIP13 | 6.67E-235 | 0.80537266 | 0.931 | 0.795 | 2.55E-230 | InN5 | GRIP1 |
| PDE1C3 | 8.01E-232 | 1.04878656 | 0.749 | 0.487 | 3.06E-227 | InN5 | PDE1C |

| | | | | | | | |
|---|---|---|---|---|---|---|---|
| DDR21 | 7.86E-230 | 0.56419887 | 0.283 | 0.078 | 3.00E-225 | InN5 | DDR2 |
| CHSY33 | 4.21E-229 | 0.87619573 | 0.924 | 0.833 | 1.61E-224 | InN5 | CHSY3 |
| KCNC23 | 2.13E-228 | 0.71127374 | 0.914 | 0.833 | 8.13E-224 | InN5 | KCNC2 |
| MARCH33 | 2.98E-228 | 0.70634243 | 0.388 | 0.138 | 1.14E-223 | InN5 | 3-Mar |
| NCKAP52 | 1.13E-226 | 1.08220344 | 0.469 | 0.199 | 4.33E-222 | InN5 | NCKAP5 |
| SLCO5A11 | 2.73E-224 | 0.72590321 | 0.333 | 0.108 | 1.05E-219 | InN5 | SLCO5A1 |
| ROBO12 | 1.55E-223 | 1.02283963 | 0.878 | 0.653 | 5.94E-219 | InN5 | ROBO1 |
| GRIK43 | 6.28E-221 | 0.91465038 | 0.695 | 0.436 | 2.40E-216 | InN5 | GRIK4 |
| GRIN3A2 | 2.77E-220 | 0.69697778 | 0.534 | 0.227 | 1.06E-215 | InN5 | GRIN3A |
| ANKFN11 | 5.83E-219 | 0.91034873 | 0.473 | 0.209 | 2.23E-214 | InN5 | ANKFN1 |
| AUTS22 | 1.81E-217 | 0.52156895 | 0.98 | 0.974 | 6.90E-213 | InN5 | AUTS2 |
| TTC281 | 3.32E-217 | 0.75609699 | 0.867 | 0.786 | 1.27E-212 | InN5 | TTC28 |
| ZNF6083 | 2.55E-216 | 0.78524912 | 0.629 | 0.348 | 9.74E-212 | InN5 | ZNF608 |
| PCP4L11 | 8.37E-216 | 0.46378222 | 0.277 | 0.078 | 3.20E-211 | InN5 | PCP4L1 |
| NLGN11 | 2.98E-214 | 0.64611347 | 0.984 | 0.96 | 1.14E-209 | InN5 | NLGN1 |
| EXT12 | 1.36E-213 | 0.88142916 | 0.876 | 0.81 | 5.19E-209 | InN5 | EXT1 |
| MTSS15 | 2.19E-213 | 0.80127731 | 0.779 | 0.562 | 8.38E-209 | InN5 | MTSS1 |
| NYAP22 | 5.35E-213 | 0.87493545 | 0.729 | 0.501 | 2.04E-208 | InN5 | NYAP2 |
| PANTR11 | 7.50E-209 | 0.91762948 | 0.723 | 0.536 | 2.87E-204 | InN5 | PANTR1 |
| NHS4 | 1.74E-208 | 0.79911061 | 0.721 | 0.441 | 6.64E-204 | InN5 | NHS |
| BCL11B4 | 5.69E-208 | 0.71362974 | 0.534 | 0.247 | 2.18E-203 | InN5 | BCL11B |
| LINC011022 | 2.11E-206 | 0.76973221 | 0.58 | 0.316 | 8.08E-202 | InN5 | LINC01102 |
| NECAB21 | 1.94E-205 | 0.65542879 | 0.409 | 0.168 | 7.41E-201 | InN5 | NECAB2 |
| NEBL2 | 2.92E-205 | 0.6726383 | 0.919 | 0.874 | 1.12E-200 | InN5 | NEBL |
| SLC24A33 | 5.19E-203 | 0.9025903 | 0.613 | 0.301 | 1.98E-198 | InN5 | SLC24A3 |
| RIPOR23 | 2.93E-202 | 0.80889139 | 0.695 | 0.46 | 1.12E-197 | InN5 | RIPOR2 |
| KIAA12114 | 4.11E-200 | 0.59511549 | 0.727 | 0.388 | 1.57E-195 | InN5 | KIAA1211 |
| MTUS21 | 9.68E-195 | 0.58564655 | 0.957 | 0.924 | 3.70E-190 | InN5 | MTUS2 |
| NFIX2 | 1.57E-194 | 0.7876438 | 0.783 | 0.627 | 5.99E-190 | InN5 | NFIX |
| NRIP33 | 5.80E-194 | 0.59599622 | 0.474 | 0.209 | 2.22E-189 | InN5 | NRIP3 |
| C1orf212 | 2.30E-191 | 0.80333715 | 0.808 | 0.716 | 8.78E-187 | InN5 | C1orf21 |
| ANKRD63 | 4.42E-190 | 0.78812676 | 0.638 | 0.41 | 1.69E-185 | InN5 | ANKRD6 |
| SRGAP13 | 1.04E-188 | 0.88128444 | 0.487 | 0.245 | 3.98E-184 | InN5 | SRGAP1 |
| ARL4C3 | 4.20E-187 | 0.64952032 | 0.522 | 0.262 | 1.61E-182 | InN5 | ARL4C |
| ADAMTS61 | 4.31E-185 | 0.64888702 | 0.343 | 0.122 | 1.65E-180 | InN5 | ADAMTS6 |
| GRIP24 | 7.91E-185 | 0.5126405 | 0.444 | 0.181 | 3.02E-180 | InN5 | GRIP2 |
| NFIB2 | 1.51E-183 | 0.66946097 | 0.851 | 0.643 | 5.76E-179 | InN5 | NFIB |
| EPHA73 | 8.09E-182 | 0.91116354 | 0.719 | 0.539 | 3.09E-177 | InN5 | EPHA7 |
| FRMPD31 | 2.34E-179 | 0.45584595 | 0.303 | 0.103 | 8.93E-175 | InN5 | FRMPD3 |
| LINGO23 | 4.71E-179 | 0.6595611 | 0.952 | 0.82 | 1.80E-174 | InN5 | LINGO2 |
| ADCY23 | 1.19E-177 | 0.65545864 | 0.906 | 0.852 | 4.55E-173 | InN5 | ADCY2 |
| LSAMP3 | 1.98E-177 | 0.55662415 | 0.993 | 0.974 | 7.58E-173 | InN5 | LSAMP |
| FAT11 | 2.83E-176 | 0.5932432 | 0.263 | 0.084 | 1.08E-171 | InN5 | FAT1 |
| FOXN33 | 5.03E-176 | 0.68498441 | 0.822 | 0.704 | 1.92E-171 | InN5 | FOXN3 |
| MIR4300HG1 | 1.66E-175 | 1.16699624 | 0.329 | 0.127 | 6.36E-171 | InN5 | MIR4300HG |
| PTPRO2 | 8.92E-175 | 0.7642945 | 0.831 | 0.676 | 3.41E-170 | InN5 | PTPRO |
| SOX21 | 1.75E-173 | 0.42100592 | 0.269 | 0.085 | 6.69E-169 | InN5 | SOX2 |
| GNAL4 | 7.82E-173 | 0.67419384 | 0.741 | 0.469 | 2.99E-168 | InN5 | GNAL |
| MGAT4C2 | 2.01E-171 | 0.88398236 | 0.793 | 0.572 | 7.70E-167 | InN5 | MGAT4C |
| PKNOX22 | 3.69E-171 | 0.6803788 | 0.854 | 0.754 | 1.41E-166 | InN5 | PKNOX2 |
| SLC7A21 | 5.52E-170 | 0.43358912 | 0.251 | 0.077 | 2.11E-165 | InN5 | SLC7A2 |
| BICD13 | 2.87E-166 | 0.66604224 | 0.864 | 0.792 | 1.10E-161 | InN5 | BICD1 |
| ATP1B23 | 2.01E-165 | 0.57245193 | 0.467 | 0.224 | 7.67E-161 | InN5 | ATP1B2 |
| ARHGAP181 | 6.68E-165 | 0.95378035 | 0.419 | 0.208 | 2.55E-160 | InN5 | ARHGAP18 |
| SOX2-OT2 | 8.06E-165 | 0.99711093 | 0.687 | 0.485 | 3.08E-160 | InN5 | SOX2-OT |
| AL589740.11 | 6.14E-164 | 1.07446467 | 0.635 | 0.431 | 2.35E-159 | InN5 | AL589740.1 |
| MIR99AHG3 | 6.93E-163 | 0.64791525 | 0.933 | 0.894 | 2.65E-158 | InN5 | MIR99AHG |
| TCF125 | 1.74E-162 | 0.69876637 | 0.812 | 0.702 | 6.65E-158 | InN5 | TCF12 |
| SLC1A21 | 3.53E-162 | 0.7228063 | 0.645 | 0.455 | 1.35E-157 | InN5 | SLC1A2 |
| AP1S22 | 5.28E-162 | 0.54958043 | 0.431 | 0.195 | 2.02E-157 | InN5 | AP1S2 |
| SCLT1 | 1.48E-161 | 0.82845891 | 0.714 | 0.568 | 5.67E-157 | InN5 | SCLT1 |
| XKR43 | 1.69E-160 | 0.61723994 | 0.918 | 0.859 | 6.46E-156 | InN5 | XKR4 |
| MAML32 | 5.37E-160 | 1.00967977 | 0.569 | 0.355 | 2.05E-155 | InN5 | MAML3 |

| | | | | | | | |
|---|---|---|---|---|---|---|---|
| LINC002981 | 1.77E-159 | 0.66996947 | 0.27 | 0.093 | 6.75E-155 | InN5 | LINC00298 |
| CDR21 | 2.89E-158 | 0.78930091 | 0.706 | 0.55 | 1.10E-153 | InN5 | CDR2 |
| QKI5 | 3.13E-155 | 0.6291707 | 0.835 | 0.667 | 1.20E-150 | InN5 | QKI |
| EPHA63 | 3.49E-155 | 0.65398893 | 0.869 | 0.654 | 1.34E-150 | InN5 | EPHA6 |
| LINC016301 | 1.59E-151 | 0.66158093 | 0.323 | 0.129 | 6.08E-147 | InN5 | LINC01630 |
| PKP22 | 2.45E-150 | 0.41221078 | 0.291 | 0.102 | 9.36E-146 | InN5 | PKP2 |
| POU3F32 | 1.61E-149 | 0.51358813 | 0.372 | 0.164 | 6.17E-145 | InN5 | POU3F3 |
| ANO43 | 8.59E-149 | 0.63863276 | 0.788 | 0.593 | 3.28E-144 | InN5 | ANO4 |
| MTUS14 | 1.44E-147 | 0.65512881 | 0.52 | 0.291 | 5.49E-143 | InN5 | MTUS1 |
| ADRA1B1 | 1.44E-147 | 0.73009053 | 0.428 | 0.214 | 5.51E-143 | InN5 | ADRA1B |
| DSCAML14 | 5.93E-146 | 0.58597301 | 0.885 | 0.754 | 2.27E-141 | InN5 | DSCAML1 |
| SEMA6A1 | 1.00E-144 | 0.61804604 | 0.431 | 0.211 | 3.82E-140 | InN5 | SEMA6A |
| PID12 | 2.02E-143 | 0.75391145 | 0.681 | 0.493 | 7.70E-139 | InN5 | PID1 |
| MAGI24 | 1.41E-142 | 0.5540039 | 0.99 | 0.987 | 5.40E-138 | InN5 | MAGI2 |
| SORCS33 | 3.96E-142 | 0.63110805 | 0.762 | 0.563 | 1.51E-137 | InN5 | SORCS3 |
| SIPA1L21 | 4.94E-142 | 0.96443764 | 0.578 | 0.422 | 1.89E-137 | InN5 | SIPA1L2 |
| SNRPN4 | 1.53E-141 | 0.57021236 | 0.864 | 0.758 | 5.85E-137 | InN5 | SNRPN |
| SLC35F43 | 2.02E-141 | 0.58602505 | 0.477 | 0.227 | 7.72E-137 | InN5 | SLC35F4 |
| ERC13 | 1.56E-140 | 0.5250457 | 0.894 | 0.866 | 5.94E-136 | InN5 | ERC1 |
| GLRA22 | 3.17E-137 | 0.57394844 | 0.422 | 0.204 | 1.21E-132 | InN5 | GLRA2 |
| ANKRD501 | 1.56E-136 | 0.57026259 | 0.392 | 0.192 | 5.97E-132 | InN5 | ANKRD50 |
| PODXL23 | 1.05E-135 | 0.63175776 | 0.581 | 0.385 | 4.00E-131 | InN5 | PODXL2 |
| SILC13 | 2.93E-134 | 0.52593504 | 0.439 | 0.212 | 1.12E-129 | InN5 | SILC1 |
| ZBTB162 | 5.02E-134 | 0.52015226 | 0.91 | 0.876 | 1.92E-129 | InN5 | ZBTB16 |
| MCC3 | 1.98E-133 | 0.59849951 | 0.491 | 0.257 | 7.55E-129 | InN5 | MCC |
| ARHGAP282 | 2.63E-133 | 0.61375863 | 0.413 | 0.202 | 1.00E-128 | InN5 | ARHGAP28 |
| CHRM32 | 1.29E-132 | 0.55117748 | 0.951 | 0.875 | 4.92E-128 | InN5 | CHRM3 |
| ESRRG2 | 2.56E-132 | 0.7803226 | 0.805 | 0.68 | 9.77E-128 | InN5 | ESRRG |
| FNBP1L4 | 6.96E-131 | 0.65081683 | 0.59 | 0.383 | 2.66E-126 | InN5 | FNBP1L |
| POU2F11 | 1.42E-129 | 0.63693226 | 0.796 | 0.756 | 5.43E-125 | InN5 | POU2F1 |
| EMID11 | 8.30E-128 | 0.36736918 | 0.257 | 0.094 | 3.17E-123 | InN5 | EMID1 |
| LINC011973 | 8.84E-127 | 0.65496821 | 0.559 | 0.344 | 3.38E-122 | InN5 | LINC01197 |
| FREM12 | 9.75E-127 | 1.03060947 | 0.402 | 0.216 | 3.73E-122 | InN5 | FREM1 |
| PEG101 | 1.26E-126 | 0.83208471 | 0.452 | 0.26 | 4.83E-122 | InN5 | PEG10 |
| MSI24 | 4.88E-126 | 0.69887887 | 0.764 | 0.657 | 1.87E-121 | InN5 | MSI2 |
| AC079296.13 | 1.32E-125 | 0.47293005 | 0.379 | 0.178 | 5.04E-121 | InN5 | AC079296.1 |
| FAM110B3 | 5.02E-125 | 0.6241398 | 0.62 | 0.427 | 1.92E-120 | InN5 | FAM110B |
| PTPRS2 | 4.18E-123 | 0.63194179 | 0.786 | 0.711 | 1.60E-118 | InN5 | PTPRS |
| BACH15 | 5.91E-123 | 0.56815819 | 0.433 | 0.227 | 2.26E-118 | InN5 | BACH1 |
| RANBP172 | 9.32E-123 | 0.60124009 | 0.665 | 0.465 | 3.56E-118 | InN5 | RANBP17 |
| COL25A12 | 9.55E-122 | 0.57092698 | 0.411 | 0.21 | 3.65E-117 | InN5 | COL25A1 |
| TMTC22 | 6.69E-121 | 0.78282092 | 0.714 | 0.535 | 2.56E-116 | InN5 | TMTC2 |
| SYNPR2 | 9.66E-120 | 0.62781211 | 0.723 | 0.533 | 3.69E-115 | InN5 | SYNPR |
| TIMP23 | 2.05E-119 | 0.6205936 | 0.5 | 0.298 | 7.84E-115 | InN5 | TIMP2 |
| CNTN43 | 8.28E-119 | 0.72020355 | 0.895 | 0.85 | 3.17E-114 | InN5 | CNTN4 |
| AKAP61 | 6.70E-118 | 0.41757042 | 0.953 | 0.944 | 2.56E-113 | InN5 | AKAP6 |
| MACROD22 | 3.09E-117 | 0.52040015 | 0.96 | 0.951 | 1.18E-112 | InN5 | MACROD2 |
| PCSK23 | 3.49E-116 | 0.57327681 | 0.86 | 0.801 | 1.33E-111 | InN5 | PCSK2 |
| KIAA19583 | 1.85E-115 | 0.6300715 | 0.612 | 0.425 | 7.06E-111 | InN5 | KIAA1958 |
| MAML21 | 1.01E-114 | 0.78432841 | 0.42 | 0.228 | 3.87E-110 | InN5 | MAML2 |
| PRICKLE22 | 2.20E-114 | 0.7889163 | 0.822 | 0.794 | 8.41E-110 | InN5 | PRICKLE2 |
| CACNB21 | 2.35E-114 | 0.49360898 | 0.928 | 0.874 | 8.99E-110 | InN5 | CACNB2 |
| AC092691.15 | 1.14E-113 | 0.4988579 | 0.939 | 0.866 | 4.37E-109 | InN5 | AC092691.1 |
| CEP1121 | 1.29E-113 | 0.7200912 | 0.706 | 0.59 | 4.92E-109 | InN5 | CEP112 |
| GRB141 | 3.42E-112 | 0.42589149 | 0.253 | 0.101 | 1.31E-107 | InN5 | GRB14 |
| NTN43 | 6.28E-111 | 0.68390675 | 0.407 | 0.224 | 2.40E-106 | InN5 | NTN4 |
| AFAP13 | 2.11E-109 | 0.54401244 | 0.576 | 0.392 | 8.07E-105 | InN5 | AFAP1 |
| CSMD31 | 2.89E-109 | 0.51382973 | 0.959 | 0.941 | 1.11E-104 | InN5 | CSMD3 |
| FGF141 | 2.85E-108 | 0.38331077 | 0.989 | 0.989 | 1.09E-103 | InN5 | FGF14 |
| FIGN2 | 4.94E-108 | 0.58831561 | 0.549 | 0.35 | 1.89E-103 | InN5 | FIGN |
| KCNT22 | 2.99E-107 | 0.53926512 | 0.724 | 0.541 | 1.14E-102 | InN5 | KCNT2 |
| DISP3 | 3.69E-107 | 0.48127434 | 0.342 | 0.17 | 1.41E-102 | InN5 | DISP3 |
| SNTG14 | 1.26E-106 | 0.47024005 | 0.97 | 0.941 | 4.82E-102 | InN5 | SNTG1 |

| | | | | | | | |
|---|---|---|---|---|---|---|---|
| SGK12 | 5.57E-105 | 0.58347162 | 0.656 | 0.503 | 2.13E-100 | InN5 | SGK1 |
| ANKRD444 | 7.56E-103 | 0.54333413 | 0.601 | 0.423 | 2.89E-98 | InN5 | ANKRD44 |
| MT-ND33 | 3.28E-101 | 0.65457562 | 0.905 | 0.869 | 1.26E-96 | InN5 | MT-ND3 |
| ZBTB204 | 6.76E-101 | 1.1829297 | 0.661 | 0.547 | 2.59E-96 | InN5 | ZBTB20 |
| NAP1L31 | 1.23E-99 | 0.64046186 | 0.461 | 0.3 | 4.68E-95 | InN5 | NAP1L3 |
| LHFPL32 | 5.04E-99 | 0.55552105 | 0.47 | 0.269 | 1.93E-94 | InN5 | LHFPL3 |
| IL1RAPL23 | 6.53E-98 | 0.52067147 | 0.55 | 0.316 | 2.49E-93 | InN5 | IL1RAPL2 |
| PEG31 | 6.90E-98 | 0.67699616 | 0.654 | 0.582 | 2.64E-93 | InN5 | PEG3 |
| MT-ND42 | 1.15E-97 | 0.59113611 | 0.91 | 0.854 | 4.39E-93 | InN5 | MT-ND4 |
| KIDINS2201 | 3.56E-97 | 0.52413817 | 0.769 | 0.738 | 1.36E-92 | InN5 | KIDINS220 |
| KCNJ32 | 1.87E-96 | 0.53091457 | 0.891 | 0.869 | 7.17E-92 | InN5 | KCNJ3 |
| CACNG42 | 2.41E-96 | 0.38432915 | 0.325 | 0.155 | 9.23E-92 | InN5 | CACNG4 |
| MYT1L2 | 6.22E-96 | 0.32583158 | 0.974 | 0.971 | 2.38E-91 | InN5 | MYT1L |
| GABBR21 | 3.27E-95 | 0.40474825 | 0.927 | 0.918 | 1.25E-90 | InN5 | GABBR2 |
| PRKCE2 | 5.17E-94 | 0.3869806 | 0.93 | 0.913 | 1.97E-89 | InN5 | PRKCE |
| KLHL132 | 2.19E-92 | 0.50673873 | 0.262 | 0.118 | 8.37E-88 | InN5 | KLHL13 |
| NRXN11 | 6.02E-92 | 0.3667415 | 0.994 | 0.986 | 2.30E-87 | InN5 | NRXN1 |
| SHISA92 | 4.60E-91 | 0.51110829 | 0.898 | 0.821 | 1.76E-86 | InN5 | SHISA9 |
| ABLIM15 | 2.79E-90 | 0.458564 | 0.868 | 0.833 | 1.07E-85 | InN5 | ABLIM1 |
| APBA11 | 1.18E-89 | 0.51891978 | 0.785 | 0.755 | 4.53E-85 | InN5 | APBA1 |
| JAM21 | 1.44E-89 | 0.48236762 | 0.318 | 0.163 | 5.50E-85 | InN5 | JAM2 |
| NMNAT22 | 1.53E-89 | 0.44614944 | 0.857 | 0.854 | 5.86E-85 | InN5 | NMNAT2 |
| SYT172 | 1.88E-89 | 0.63310842 | 0.454 | 0.295 | 7.18E-85 | InN5 | SYT17 |
| DNAH144 | 3.14E-89 | 0.56714481 | 0.465 | 0.293 | 1.20E-84 | InN5 | DNAH14 |
| NETO22 | 9.63E-89 | 0.69060236 | 0.541 | 0.407 | 3.68E-84 | InN5 | NETO2 |
| PDE3A1 | 1.04E-88 | 0.59632756 | 0.284 | 0.131 | 3.99E-84 | InN5 | PDE3A |
| UBASH3B4 | 2.17E-88 | 0.37627037 | 0.536 | 0.335 | 8.31E-84 | InN5 | UBASH3B |
| AF279873.31 | 4.80E-88 | 0.51455457 | 0.297 | 0.144 | 1.84E-83 | InN5 | AF279873.3 |
| HIP11 | 4.92E-87 | 0.42354678 | 0.352 | 0.183 | 1.88E-82 | InN5 | HIP1 |
| TLE3 | 6.34E-87 | 0.41022464 | 0.268 | 0.126 | 2.42E-82 | InN5 | TLE3 |
| ADCYAP1R14 | 9.24E-87 | 0.43872373 | 0.48 | 0.297 | 3.53E-82 | InN5 | ADCYAP1R1 |
| FOXO11 | 9.45E-87 | 0.61385626 | 0.266 | 0.125 | 3.61E-82 | InN5 | FOXO1 |
| MARCH13 | 1.32E-86 | 0.62769434 | 0.846 | 0.757 | 5.06E-82 | InN5 | 1-Mar |
| MT-CO23 | 1.49E-86 | 0.55506981 | 0.931 | 0.897 | 5.68E-82 | InN5 | MT-CO2 |
| NRXN21 | 5.62E-85 | 0.53523336 | 0.709 | 0.634 | 2.15E-80 | InN5 | NRXN2 |
| LONRF3 | 1.12E-84 | 0.37397007 | 0.277 | 0.131 | 4.30E-80 | InN5 | LONRF3 |
| VSTM2A2 | 1.79E-84 | 0.57383962 | 0.531 | 0.39 | 6.83E-80 | InN5 | VSTM2A |
| CDH92 | 3.47E-84 | 0.67008048 | 0.764 | 0.616 | 1.32E-79 | InN5 | CDH9 |
| GPRIN33 | 7.29E-84 | 0.3670442 | 0.339 | 0.174 | 2.79E-79 | InN5 | GPRIN3 |
| ELAVL22 | 8.66E-84 | 0.58641339 | 0.662 | 0.518 | 3.31E-79 | InN5 | ELAVL2 |
| NCAM11 | 1.64E-83 | 0.35250185 | 0.95 | 0.937 | 6.27E-79 | InN5 | NCAM1 |
| LRRC4C4 | 7.08E-82 | 0.29460454 | 0.943 | 0.888 | 2.70E-77 | InN5 | LRRC4C |
| AP001347.11 | 3.00E-81 | 0.56999822 | 0.691 | 0.598 | 1.15E-76 | InN5 | AP001347.1 |
| GPR137C2 | 3.86E-81 | 0.57884961 | 0.684 | 0.605 | 1.48E-76 | InN5 | GPR137C |
| MAGI33 | 8.28E-81 | 0.55890528 | 0.752 | 0.687 | 3.16E-76 | InN5 | MAGI3 |
| MARCKSL11 | 7.45E-80 | 0.33106744 | 0.251 | 0.115 | 2.85E-75 | InN5 | MARCKSL1 |
| AC019197.1 | 3.24E-78 | 1.03567778 | 0.258 | 0.132 | 1.24E-73 | InN5 | AC019197.1 |
| LPIN23 | 9.22E-78 | 0.54212836 | 0.499 | 0.36 | 3.53E-73 | InN5 | LPIN2 |
| RIMBP21 | 2.66E-77 | 0.57384431 | 0.771 | 0.76 | 1.02E-72 | InN5 | RIMBP2 |
| HIVEP33 | 1.08E-76 | 0.41404159 | 0.879 | 0.862 | 4.12E-72 | InN5 | HIVEP3 |
| SH3RF34 | 2.69E-76 | 0.5027437 | 0.75 | 0.68 | 1.03E-71 | InN5 | SH3RF3 |
| ZMAT43 | 4.17E-75 | 0.51712502 | 0.766 | 0.77 | 1.60E-70 | InN5 | ZMAT4 |
| OSTM11 | 5.68E-75 | 0.48676998 | 0.399 | 0.259 | 2.17E-70 | InN5 | OSTM1 |
| RBMS33 | 1.73E-74 | 0.66190975 | 0.73 | 0.638 | 6.63E-70 | InN5 | RBMS3 |
| RASSF83 | 7.33E-74 | 0.51364496 | 0.316 | 0.174 | 2.80E-69 | InN5 | RASSF8 |
| HS6ST32 | 1.69E-73 | 0.34846502 | 0.925 | 0.742 | 6.47E-69 | InN5 | HS6ST3 |
| OSBPL93 | 1.73E-73 | 0.49686434 | 0.636 | 0.557 | 6.62E-69 | InN5 | OSBPL9 |
| SLC7A141 | 1.14E-72 | 0.59274717 | 0.606 | 0.512 | 4.36E-68 | InN5 | SLC7A14 |
| NBEA1 | 3.25E-72 | 0.39114146 | 0.963 | 0.96 | 1.24E-67 | InN5 | NBEA |
| MYO1B1 | 1.07E-71 | 0.54697171 | 0.396 | 0.251 | 4.10E-67 | InN5 | MYO1B |
| AL133346.12 | 1.94E-71 | 0.42478749 | 0.292 | 0.147 | 7.43E-67 | InN5 | AL133346.1 |
| CHD31 | 3.54E-71 | 0.54162318 | 0.488 | 0.362 | 1.35E-66 | InN5 | CHD3 |
| NIPAL31 | 1.13E-70 | 0.47929189 | 0.424 | 0.288 | 4.31E-66 | InN5 | NIPAL3 |

| | | | | | | | |
|---|---|---|---|---|---|---|---|
| IL6ST3 | 1.16E-70 | 0.51988964 | 0.459 | 0.323 | 4.42E-66 | InN5 | IL6ST |
| ANKRD30BL2 | 3.66E-70 | 0.5173279 | 0.786 | 0.706 | 1.40E-65 | InN5 | ANKRD30BL |
| RGMB | 1.06E-69 | 0.50018066 | 0.386 | 0.252 | 4.04E-65 | InN5 | RGMB |
| GABRB22 | 2.52E-69 | 0.3895668 | 0.91 | 0.923 | 9.63E-65 | InN5 | GABRB2 |
| SLC35F16 | 3.07E-69 | 0.45176826 | 0.831 | 0.706 | 1.17E-64 | InN5 | SLC35F1 |
| LRRN32 | 3.38E-69 | 0.50776598 | 0.469 | 0.333 | 1.29E-64 | InN5 | LRRN3 |
| TPD52L11 | 6.28E-69 | 0.39112814 | 0.281 | 0.146 | 2.40E-64 | InN5 | TPD52L1 |
| DACH2 | 1.13E-68 | 1.12055565 | 0.344 | 0.227 | 4.34E-64 | InN5 | DACH2 |
| PTPRE2 | 2.14E-68 | 0.51725525 | 0.695 | 0.632 | 8.18E-64 | InN5 | PTPRE |
| B3GLCT3 | 3.27E-68 | 0.59839869 | 0.43 | 0.295 | 1.25E-63 | InN5 | B3GLCT |
| CACNA1D1 | 3.36E-68 | 0.38762453 | 0.886 | 0.888 | 1.28E-63 | InN5 | CACNA1D |
| GRID22 | 9.06E-68 | 0.63832929 | 0.809 | 0.734 | 3.46E-63 | InN5 | GRID2 |
| COL5A22 | 1.89E-67 | 0.67466898 | 0.261 | 0.137 | 7.22E-63 | InN5 | COL5A2 |
| PLPPR4 | 2.13E-67 | 0.49047578 | 0.641 | 0.585 | 8.14E-63 | InN5 | PLPPR4 |
| OLFM23 | 1.35E-66 | 0.40587264 | 0.443 | 0.293 | 5.16E-62 | InN5 | OLFM2 |
| ZHX22 | 1.21E-65 | 0.47791573 | 0.361 | 0.219 | 4.62E-61 | InN5 | ZHX2 |
| POT1-AS11 | 1.39E-65 | 0.57165836 | 0.521 | 0.392 | 5.31E-61 | InN5 | POT1-AS1 |
| ANK31 | 5.92E-65 | 0.25694541 | 0.979 | 0.976 | 2.26E-60 | InN5 | ANK3 |
| MT-ATP63 | 6.50E-65 | 0.44106814 | 0.909 | 0.878 | 2.49E-60 | InN5 | MT-ATP6 |
| GNAS2 | 1.02E-64 | 0.52054323 | 0.818 | 0.825 | 3.91E-60 | InN5 | GNAS |
| GABRB11 | 1.23E-64 | 0.34208584 | 0.919 | 0.916 | 4.72E-60 | InN5 | GABRB1 |
| MT-CO32 | 2.44E-63 | 0.56719702 | 0.881 | 0.848 | 9.31E-59 | InN5 | MT-CO3 |
| USP131 | 9.03E-63 | 0.44420568 | 0.487 | 0.361 | 3.45E-58 | InN5 | USP13 |
| DGKD3 | 1.98E-61 | 0.36819131 | 0.467 | 0.309 | 7.55E-57 | InN5 | DGKD |
| GRK32 | 1.35E-60 | 0.50295428 | 0.713 | 0.709 | 5.17E-56 | InN5 | GRK3 |
| LRRC392 | 1.46E-60 | 0.37322958 | 0.287 | 0.161 | 5.59E-56 | InN5 | LRRC39 |
| ADCY82 | 1.11E-59 | 0.31555179 | 0.446 | 0.284 | 4.23E-55 | InN5 | ADCY8 |
| GSE12 | 2.41E-59 | 0.45010744 | 0.601 | 0.5 | 9.20E-55 | InN5 | GSE1 |
| PCDH154 | 5.12E-59 | 0.66066295 | 0.46 | 0.323 | 1.96E-54 | InN5 | PCDH15 |
| NCOA23 | 5.20E-59 | 0.40285814 | 0.774 | 0.743 | 1.99E-54 | InN5 | NCOA2 |
| TXNDC161 | 6.54E-59 | 0.47918093 | 0.606 | 0.543 | 2.50E-54 | InN5 | TXNDC16 |
| LINC012371 | 2.53E-58 | 0.4264605 | 0.282 | 0.159 | 9.68E-54 | InN5 | LINC01237 |
| DAAM13 | 2.56E-58 | 0.39452789 | 0.765 | 0.751 | 9.77E-54 | InN5 | DAAM1 |
| PDE7B2 | 6.69E-58 | 0.59141695 | 0.609 | 0.48 | 2.56E-53 | InN5 | PDE7B |
| TNR2 | 1.11E-56 | 0.46402653 | 0.807 | 0.803 | 4.23E-52 | InN5 | TNR |
| ANK14 | 1.79E-56 | 0.32470417 | 0.487 | 0.315 | 6.84E-52 | InN5 | ANK1 |
| ATP1B13 | 2.41E-56 | 0.53965701 | 0.771 | 0.781 | 9.20E-52 | InN5 | ATP1B1 |
| MT-CO12 | 1.18E-55 | 0.42399125 | 0.902 | 0.867 | 4.53E-51 | InN5 | MT-CO1 |
| RTN3 | 2.21E-55 | 0.32666822 | 0.885 | 0.903 | 8.45E-51 | InN5 | RTN3 |
| HSP90AA13 | 2.96E-55 | 0.53488149 | 0.743 | 0.711 | 1.13E-50 | InN5 | HSP90AA1 |
| SRM | 3.25E-55 | 0.33534281 | 0.287 | 0.17 | 1.24E-50 | InN5 | SRM |
| PARM12 | 3.88E-55 | 0.52349395 | 0.506 | 0.396 | 1.48E-50 | InN5 | PARM1 |
| SLCO3A13 | 1.90E-54 | 0.49935474 | 0.748 | 0.737 | 7.27E-50 | InN5 | SLCO3A1 |
| REV3L3 | 1.72E-53 | 0.33980324 | 0.794 | 0.778 | 6.58E-49 | InN5 | REV3L |
| ABI11 | 1.91E-53 | 0.47846254 | 0.631 | 0.576 | 7.31E-49 | InN5 | ABI1 |
| HSP90AB11 | 2.22E-53 | 0.49227358 | 0.735 | 0.728 | 8.50E-49 | InN5 | HSP90AB1 |
| PCDH102 | 2.54E-53 | 0.51362353 | 0.542 | 0.449 | 9.72E-49 | InN5 | PCDH10 |
| VOPP12 | 2.70E-53 | 0.44445598 | 0.57 | 0.47 | 1.03E-48 | InN5 | VOPP1 |
| CACNA1E2 | 6.32E-53 | 0.43350788 | 0.802 | 0.811 | 2.42E-48 | InN5 | CACNA1E |
| LINC004611 | 8.32E-53 | 0.39096928 | 0.789 | 0.786 | 3.18E-48 | InN5 | LINC00461 |
| GAS71 | 8.50E-53 | 0.41310167 | 0.793 | 0.78 | 3.25E-48 | InN5 | GAS7 |
| MTRNR2L12 | 1.44E-52 | 0.61955262 | 0.799 | 0.746 | 5.50E-48 | InN5 | MTRNR2L12 |
| NPTXR2 | 2.08E-52 | 0.45710974 | 0.603 | 0.532 | 7.95E-48 | InN5 | NPTXR |
| SERINC53 | 2.14E-52 | 0.46849572 | 0.521 | 0.421 | 8.18E-48 | InN5 | SERINC5 |
| THRB3 | 4.19E-52 | 0.33621418 | 0.919 | 0.917 | 1.60E-47 | InN5 | THRB |
| REEP11 | 6.13E-52 | 0.44256333 | 0.655 | 0.625 | 2.34E-47 | InN5 | REEP1 |
| N4BP22 | 6.25E-52 | 0.41288824 | 0.477 | 0.363 | 2.39E-47 | InN5 | N4BP2 |
| PDSS11 | 9.87E-52 | 0.38819533 | 0.324 | 0.207 | 3.77E-47 | InN5 | PDSS1 |
| SOBP1 | 1.14E-51 | 0.34922526 | 0.827 | 0.819 | 4.35E-47 | InN5 | SOBP |
| GTF2I2 | 2.12E-50 | 0.33558382 | 0.835 | 0.835 | 8.09E-46 | InN5 | GTF2I |
| NSG21 | 2.41E-50 | 0.4692581 | 0.609 | 0.556 | 9.20E-46 | InN5 | NSG2 |
| CAMK42 | 2.73E-50 | 0.39359098 | 0.774 | 0.72 | 1.04E-45 | InN5 | CAMK4 |
| TMEM2322 | 3.57E-50 | 0.45216259 | 0.808 | 0.773 | 1.36E-45 | InN5 | TMEM232 |

| | | | | | | | |
|---|---|---|---|---|---|---|---|
| MT-ND4L2 | 6.01E-50 | 0.45452814 | 0.635 | 0.542 | 2.30E-45 | InN5 | MT-ND4L |
| LPP3 | 6.42E-49 | 0.42803696 | 0.58 | 0.472 | 2.45E-44 | InN5 | LPP |
| ERBIN3 | 8.71E-49 | 0.43617592 | 0.612 | 0.537 | 3.33E-44 | InN5 | ERBIN |
| CHD91 | 1.17E-48 | 0.33776366 | 0.837 | 0.839 | 4.48E-44 | InN5 | CHD9 |
| LCORL2 | 6.05E-48 | 0.44209534 | 0.575 | 0.494 | 2.31E-43 | InN5 | LCORL |
| MYRIP4 | 1.15E-47 | 0.42121861 | 0.816 | 0.807 | 4.41E-43 | InN5 | MYRIP |
| IRF2BP2 | 1.59E-47 | 0.35708293 | 0.276 | 0.168 | 6.07E-43 | InN5 | IRF2BP2 |
| CHL12 | 1.77E-47 | 0.47976111 | 0.817 | 0.807 | 6.78E-43 | InN5 | CHL1 |
| TNRC6A3 | 4.93E-47 | 0.25306265 | 0.946 | 0.959 | 1.88E-42 | InN5 | TNRC6A |
| GNAI1 | 1.07E-46 | 0.46624157 | 0.585 | 0.526 | 4.08E-42 | InN5 | GNAI1 |
| MAST21 | 1.43E-46 | 0.42531186 | 0.494 | 0.406 | 5.45E-42 | InN5 | MAST2 |
| SGCZ5 | 1.98E-46 | 0.40488591 | 0.61 | 0.512 | 7.57E-42 | InN5 | SGCZ |
| SEZ6L21 | 4.52E-46 | 0.48637927 | 0.531 | 0.463 | 1.73E-41 | InN5 | SEZ6L2 |
| PXDNL2 | 1.15E-45 | 0.57730114 | 0.331 | 0.226 | 4.39E-41 | InN5 | PXDNL |
| SBF21 | 3.82E-45 | 0.32916693 | 0.804 | 0.802 | 1.46E-40 | InN5 | SBF2 |
| PLCB41 | 2.56E-44 | 0.40446597 | 0.638 | 0.562 | 9.77E-40 | InN5 | PLCB4 |
| SHANK2 | 2.66E-44 | 0.3283366 | 0.82 | 0.816 | 1.02E-39 | InN5 | SHANK2 |
| INHBA-AS11 | 3.80E-44 | 0.31887143 | 0.255 | 0.15 | 1.45E-39 | InN5 | INHBA-AS1 |
| ARHGAP52 | 8.37E-44 | 0.43970342 | 0.638 | 0.591 | 3.20E-39 | InN5 | ARHGAP5 |
| ZNF5161 | 8.68E-44 | 0.37911547 | 0.439 | 0.327 | 3.32E-39 | InN5 | ZNF516 |
| TUBB2A | 1.33E-43 | 0.46842909 | 0.387 | 0.286 | 5.07E-39 | InN5 | TUBB2A |
| COLGALT2 | 2.46E-43 | 0.4283586 | 0.522 | 0.441 | 9.39E-39 | InN5 | COLGALT2 |
| SCN3A | 2.53E-43 | 0.35218962 | 0.793 | 0.786 | 9.67E-39 | InN5 | SCN3A |
| BCL23 | 4.89E-43 | 0.45416649 | 0.604 | 0.525 | 1.87E-38 | InN5 | BCL2 |
| ATRX1 | 6.94E-43 | 0.32119814 | 0.821 | 0.837 | 2.65E-38 | InN5 | ATRX |
| ARNT22 | 7.85E-43 | 0.30207151 | 0.854 | 0.838 | 3.00E-38 | InN5 | ARNT2 |
| JAK12 | 9.44E-43 | 0.39960091 | 0.752 | 0.745 | 3.61E-38 | InN5 | JAK1 |
| SLC41A2 | 1.71E-42 | 0.41552125 | 0.623 | 0.581 | 6.55E-38 | InN5 | SLC41A2 |
| MSRA4 | 2.46E-42 | 0.26465043 | 0.939 | 0.908 | 9.40E-38 | InN5 | MSRA |
| DOK62 | 3.02E-42 | 0.5256549 | 0.743 | 0.706 | 1.15E-37 | InN5 | DOK6 |
| ENTPD33 | 5.61E-42 | 0.31338333 | 0.414 | 0.293 | 2.14E-37 | InN5 | ENTPD3 |
| HIPK21 | 1.25E-41 | 0.5112083 | 0.458 | 0.363 | 4.78E-37 | InN5 | HIPK2 |
| ATXN12 | 1.45E-41 | 0.26233535 | 0.927 | 0.928 | 5.52E-37 | InN5 | ATXN1 |
| OCA22 | 4.06E-41 | 0.54038546 | 0.373 | 0.273 | 1.55E-36 | InN5 | OCA2 |
| TMEM1451 | 4.32E-41 | 0.36669449 | 0.438 | 0.345 | 1.65E-36 | InN5 | TMEM145 |
| AC008591.11 | 6.33E-41 | 0.64970443 | 0.371 | 0.27 | 2.42E-36 | InN5 | AC008591.1 |
| USP34 | 1.67E-40 | 0.26131051 | 0.283 | 0.173 | 6.37E-36 | InN5 | USP3 |
| TMEM1083 | 1.76E-40 | 0.40568715 | 0.852 | 0.82 | 6.72E-36 | InN5 | TMEM108 |
| MT-ND52 | 3.36E-40 | 0.45088292 | 0.638 | 0.567 | 1.28E-35 | InN5 | MT-ND5 |
| CDK82 | 7.02E-40 | 0.47951257 | 0.599 | 0.562 | 2.68E-35 | InN5 | CDK8 |
| TMEFF25 | 1.96E-39 | 0.3184732 | 0.797 | 0.764 | 7.47E-35 | InN5 | TMEFF2 |
| AC008691.12 | 2.73E-39 | 0.4429348 | 0.275 | 0.174 | 1.05E-34 | InN5 | AC008691.1 |
| NT5DC3 | 2.99E-39 | 0.38452663 | 0.399 | 0.302 | 1.14E-34 | InN5 | NT5DC3 |
| CLIP3 | 2.05E-38 | 0.43876219 | 0.469 | 0.397 | 7.82E-34 | InN5 | CLIP3 |
| COL16A1 | 3.85E-38 | 0.55843463 | 0.27 | 0.184 | 1.47E-33 | InN5 | COL16A1 |
| PIP5K1B3 | 6.98E-38 | 0.42210756 | 0.557 | 0.48 | 2.67E-33 | InN5 | PIP5K1B |
| GREM21 | 9.49E-38 | 0.32868136 | 0.271 | 0.169 | 3.63E-33 | InN5 | GREM2 |
| EEPD11 | 2.75E-37 | 0.36505502 | 0.483 | 0.383 | 1.05E-32 | InN5 | EEPD1 |
| TRMT9B2 | 4.14E-37 | 0.27638426 | 0.536 | 0.415 | 1.58E-32 | InN5 | TRMT9B |
| SEMA6D3 | 5.40E-37 | 0.44267406 | 0.649 | 0.583 | 2.06E-32 | InN5 | SEMA6D |
| SEMA5A2 | 9.67E-37 | 0.4253938 | 0.275 | 0.173 | 3.70E-32 | InN5 | SEMA5A |
| STARD133 | 2.42E-36 | 0.5169578 | 0.591 | 0.54 | 9.24E-32 | InN5 | STARD13 |
| KIAA1549L2 | 3.18E-36 | 0.35765385 | 0.83 | 0.838 | 1.21E-31 | InN5 | KIAA1549L |
| MT-CYB3 | 3.87E-36 | 0.37153728 | 0.819 | 0.786 | 1.48E-31 | InN5 | MT-CYB |
| B3GAT11 | 4.10E-36 | 0.38920073 | 0.551 | 0.489 | 1.57E-31 | InN5 | B3GAT1 |
| CBLB1 | 4.15E-36 | 0.5456373 | 0.473 | 0.41 | 1.59E-31 | InN5 | CBLB |
| FCHSD25 | 1.11E-35 | 0.31341234 | 0.67 | 0.614 | 4.24E-31 | InN5 | FCHSD2 |
| RIN24 | 1.33E-35 | 0.27280366 | 0.322 | 0.209 | 5.07E-31 | InN5 | RIN2 |
| DCX4 | 1.89E-35 | 0.33843494 | 0.49 | 0.395 | 7.22E-31 | InN5 | DCX |
| ASIC11 | 4.19E-35 | 0.26614297 | 0.284 | 0.186 | 1.60E-30 | InN5 | ASIC1 |
| AL356124.11 | 8.52E-35 | 0.57413804 | 0.326 | 0.241 | 3.26E-30 | InN5 | AL356124.1 |
| CELF43 | 9.65E-35 | 0.25266331 | 0.898 | 0.908 | 3.69E-30 | InN5 | CELF4 |
| MT-ND12 | 1.18E-34 | 0.33881632 | 0.817 | 0.781 | 4.51E-30 | InN5 | MT-ND1 |

| | | | | | | | |
|---|---|---|---|---|---|---|---|
| FRMPD42 | 4.33E-34 | 0.2996368 | 0.945 | 0.938 | 1.66E-29 | InN5 | FRMPD4 |
| WASF33 | 5.00E-34 | 0.35489644 | 0.46 | 0.382 | 1.91E-29 | InN5 | WASF3 |
| AGPAT41 | 1.01E-33 | 0.35935919 | 0.717 | 0.705 | 3.87E-29 | InN5 | AGPAT4 |
| ARL6IP11 | 1.17E-33 | 0.49023572 | 0.491 | 0.446 | 4.47E-29 | InN5 | ARL6IP1 |
| PGBD51 | 1.30E-33 | 0.34374521 | 0.705 | 0.689 | 4.97E-29 | InN5 | PGBD5 |
| CSRNP3 | 2.95E-33 | 0.30949651 | 0.819 | 0.851 | 1.13E-28 | InN5 | CSRNP3 |
| ANKRD263 | 3.45E-33 | 0.34807004 | 0.744 | 0.754 | 1.32E-28 | InN5 | ANKRD26 |
| PRNP | 3.91E-33 | 0.41010874 | 0.67 | 0.661 | 1.50E-28 | InN5 | PRNP |
| MT-ND23 | 4.77E-33 | 0.31545506 | 0.791 | 0.748 | 1.82E-28 | InN5 | MT-ND2 |
| ZEB25 | 7.98E-33 | 0.27007394 | 0.831 | 0.822 | 3.05E-28 | InN5 | ZEB2 |
| CYFIP22 | 8.50E-33 | 0.31514298 | 0.707 | 0.712 | 3.25E-28 | InN5 | CYFIP2 |
| KDM6A1 | 1.24E-32 | 0.41170913 | 0.61 | 0.582 | 4.73E-28 | InN5 | KDM6A |
| ADAM221 | 1.52E-32 | 0.3029219 | 0.809 | 0.827 | 5.81E-28 | InN5 | ADAM22 |
| NR2F11 | 1.61E-32 | 0.4001138 | 0.471 | 0.404 | 6.16E-28 | InN5 | NR2F1 |
| HDAC2-AS21 | 1.95E-32 | 0.35297876 | 0.363 | 0.267 | 7.44E-28 | InN5 | HDAC2-AS2 |
| TUT43 | 2.87E-32 | 0.32367876 | 0.696 | 0.702 | 1.10E-27 | InN5 | TUT4 |
| LYPD6B4 | 4.74E-32 | 0.27886402 | 0.278 | 0.178 | 1.81E-27 | InN5 | LYPD6B |
| PELI22 | 5.34E-32 | 0.38180278 | 0.377 | 0.283 | 2.04E-27 | InN5 | PELI2 |
| LURAP1L | 5.88E-32 | 0.29699194 | 0.268 | 0.181 | 2.25E-27 | InN5 | LURAP1L |
| DOCK71 | 1.02E-31 | 0.3679499 | 0.571 | 0.538 | 3.90E-27 | InN5 | DOCK7 |
| GREB1L1 | 1.06E-31 | 0.32877148 | 0.421 | 0.313 | 4.04E-27 | InN5 | GREB1L |
| SIPA1L33 | 1.33E-31 | 0.36917329 | 0.6 | 0.56 | 5.07E-27 | InN5 | SIPA1L3 |
| MGLL3 | 2.74E-31 | 0.37495437 | 0.514 | 0.438 | 1.05E-26 | InN5 | MGLL |
| SEPTIN63 | 8.16E-31 | 0.3337869 | 0.378 | 0.289 | 3.12E-26 | InN5 | SEPTIN6 |
| TMEM1171 | 3.45E-30 | 0.34626708 | 0.689 | 0.646 | 1.32E-25 | InN5 | TMEM117 |
| FNBP11 | 4.07E-30 | 0.339154 | 0.651 | 0.636 | 1.56E-25 | InN5 | FNBP1 |
| SYT112 | 5.73E-30 | 0.34575504 | 0.654 | 0.647 | 2.19E-25 | InN5 | SYT11 |
| INPP5F2 | 6.52E-30 | 0.34308033 | 0.614 | 0.583 | 2.49E-25 | InN5 | INPP5F |
| PIK3R32 | 1.68E-29 | 0.34426182 | 0.4 | 0.317 | 6.42E-25 | InN5 | PIK3R3 |
| CHST153 | 2.60E-29 | 0.47456449 | 0.473 | 0.419 | 9.94E-25 | InN5 | CHST15 |
| CANX1 | 3.49E-29 | 0.39935666 | 0.529 | 0.492 | 1.33E-24 | InN5 | CANX |
| USP111 | 9.80E-29 | 0.37510733 | 0.572 | 0.543 | 3.74E-24 | InN5 | USP11 |
| MITF3 | 9.80E-29 | 0.28816133 | 0.289 | 0.196 | 3.75E-24 | InN5 | MITF |
| ANKRD122 | 1.03E-28 | 0.30267071 | 0.768 | 0.781 | 3.93E-24 | InN5 | ANKRD12 |
| SYNGR3 | 1.04E-28 | 0.32488055 | 0.333 | 0.254 | 3.98E-24 | InN5 | SYNGR3 |
| ZNF2921 | 1.07E-28 | 0.26837582 | 0.785 | 0.816 | 4.10E-24 | InN5 | ZNF292 |
| TSPYL21 | 1.08E-28 | 0.37463592 | 0.635 | 0.63 | 4.14E-24 | InN5 | TSPYL2 |
| MAPK42 | 1.23E-28 | 0.31699773 | 0.553 | 0.478 | 4.71E-24 | InN5 | MAPK4 |
| GNG23 | 2.48E-28 | 0.34973994 | 0.525 | 0.444 | 9.47E-24 | InN5 | GNG2 |
| FGF132 | 2.62E-28 | 0.40323663 | 0.47 | 0.418 | 1.00E-23 | InN5 | FGF13 |
| ERO1B2 | 2.91E-28 | 0.30739205 | 0.478 | 0.416 | 1.11E-23 | InN5 | ERO1B |
| RPL282 | 4.11E-28 | 0.42343095 | 0.433 | 0.367 | 1.57E-23 | InN5 | RPL28 |
| ECE22 | 5.63E-28 | 0.28778653 | 0.319 | 0.229 | 2.15E-23 | InN5 | ECE2 |
| WSCD21 | 7.77E-28 | 0.36480054 | 0.482 | 0.415 | 2.97E-23 | InN5 | WSCD2 |
| RIF1 | 1.74E-27 | 0.35151317 | 0.592 | 0.568 | 6.64E-23 | InN5 | RIF1 |
| MXD42 | 5.81E-27 | 0.29849633 | 0.337 | 0.255 | 2.22E-22 | InN5 | MXD4 |
| GPD22 | 6.11E-27 | 0.2966608 | 0.406 | 0.328 | 2.34E-22 | InN5 | GPD2 |
| SRRM43 | 7.90E-27 | 0.25202759 | 0.845 | 0.856 | 3.02E-22 | InN5 | SRRM4 |
| AC007364.12 | 8.32E-27 | 0.43457169 | 0.359 | 0.281 | 3.18E-22 | InN5 | AC007364.1 |
| SERINC11 | 1.82E-26 | 0.40505085 | 0.609 | 0.622 | 6.97E-22 | InN5 | SERINC1 |
| INTU1 | 3.56E-26 | 0.37527975 | 0.501 | 0.471 | 1.36E-21 | InN5 | INTU |
| CAMKMT3 | 6.00E-26 | 0.4225095 | 0.448 | 0.38 | 2.29E-21 | InN5 | CAMKMT |
| SLC22A23 | 6.86E-26 | 0.34744451 | 0.603 | 0.59 | 2.62E-21 | InN5 | SLC22A23 |
| APC4 | 1.61E-25 | 0.27472343 | 0.726 | 0.742 | 6.14E-21 | InN5 | APC |
| ADGRB1 | 2.12E-25 | 0.44952122 | 0.494 | 0.466 | 8.08E-21 | InN5 | ADGRB1 |
| SCAPER1 | 2.18E-25 | 0.28031423 | 0.795 | 0.806 | 8.33E-21 | InN5 | SCAPER |
| RYR32 | 2.35E-25 | 0.39634655 | 0.459 | 0.389 | 8.99E-21 | InN5 | RYR3 |
| RXRA2 | 2.54E-25 | 0.25245065 | 0.292 | 0.208 | 9.72E-21 | InN5 | RXRA |
| GRM74 | 4.70E-25 | 0.40151198 | 0.891 | 0.863 | 1.80E-20 | InN5 | GRM7 |
| FOXG11 | 5.98E-25 | 0.33214196 | 0.355 | 0.287 | 2.28E-20 | InN5 | FOXG1 |
| RDX2 | 1.03E-24 | 0.35181541 | 0.463 | 0.404 | 3.94E-20 | InN5 | RDX |
| KLHL421 | 1.83E-24 | 0.34043767 | 0.477 | 0.43 | 6.98E-20 | InN5 | KLHL42 |
| MBOAT23 | 2.52E-24 | 0.30839149 | 0.621 | 0.589 | 9.63E-20 | InN5 | MBOAT2 |

| | | | | | | | |
|---|---|---|---|---|---|---|---|
| SLC16A2 | 2.53E-24 | 0.27884527 | 0.28 | 0.203 | 9.68E-20 | InN5 | SLC16A2 |
| SV2A1 | 2.81E-24 | 0.35053114 | 0.599 | 0.592 | 1.07E-19 | InN5 | SV2A |
| AC006148.11 | 2.89E-24 | 0.36394291 | 0.717 | 0.71 | 1.10E-19 | InN5 | AC006148.1 |
| TRIM36 | 5.90E-24 | 0.29239054 | 0.288 | 0.213 | 2.25E-19 | InN5 | TRIM36 |
| AL445250.12 | 6.80E-24 | 0.47379828 | 0.409 | 0.343 | 2.60E-19 | InN5 | AL445250.1 |
| HACD31 | 6.94E-24 | 0.31717604 | 0.538 | 0.509 | 2.65E-19 | InN5 | HACD3 |
| EEF1A13 | 9.02E-24 | 0.36900436 | 0.601 | 0.577 | 3.45E-19 | InN5 | EEF1A1 |
| GABRA11 | 1.11E-23 | 0.33164477 | 0.641 | 0.644 | 4.24E-19 | InN5 | GABRA1 |
| PNMA21 | 1.32E-23 | 0.37677585 | 0.545 | 0.53 | 5.03E-19 | InN5 | PNMA2 |
| NPAS2 | 2.34E-23 | 0.37435905 | 0.625 | 0.606 | 8.95E-19 | InN5 | NPAS2 |
| BMERB11 | 2.89E-23 | 0.29887958 | 0.706 | 0.726 | 1.10E-18 | InN5 | BMERB1 |
| GALNT132 | 4.52E-23 | 0.47536517 | 0.701 | 0.686 | 1.73E-18 | InN5 | GALNT13 |
| FRYL1 | 6.21E-23 | 0.27142824 | 0.697 | 0.706 | 2.37E-18 | InN5 | FRYL |
| HIVEP12 | 8.73E-23 | 0.35371426 | 0.508 | 0.462 | 3.34E-18 | InN5 | HIVEP1 |
| MT-ATP82 | 9.41E-23 | 0.43816055 | 0.619 | 0.592 | 3.60E-18 | InN5 | MT-ATP8 |
| PALLD3 | 1.32E-22 | 0.31896042 | 0.485 | 0.416 | 5.06E-18 | InN5 | PALLD |
| TNRC18 | 3.49E-22 | 0.30477031 | 0.599 | 0.589 | 1.34E-17 | InN5 | TNRC18 |
| PGRMC1 | 3.53E-22 | 0.35848968 | 0.391 | 0.339 | 1.35E-17 | InN5 | PGRMC1 |
| ALK2 | 6.37E-22 | 0.44431746 | 0.326 | 0.248 | 2.43E-17 | InN5 | ALK |
| ANKRD30B2 | 1.16E-21 | 0.58826341 | 0.334 | 0.279 | 4.44E-17 | InN5 | ANKRD30B |
| KCNH75 | 9.70E-21 | 0.30725157 | 0.781 | 0.73 | 3.71E-16 | InN5 | KCNH7 |
| PNMA8A | 9.86E-21 | 0.28181394 | 0.312 | 0.246 | 3.77E-16 | InN5 | PNMA8A |
| EIF1AX | 4.15E-20 | 0.3380759 | 0.379 | 0.326 | 1.59E-15 | InN5 | EIF1AX |
| AMMECR11 | 4.20E-20 | 0.31969668 | 0.596 | 0.579 | 1.61E-15 | InN5 | AMMECR1 |
| BCAT13 | 4.21E-20 | 0.29959363 | 0.407 | 0.35 | 1.61E-15 | InN5 | BCAT1 |
| PLPPR5 | 4.78E-20 | 0.31016806 | 0.525 | 0.504 | 1.83E-15 | InN5 | PLPPR5 |
| PLXNC1 | 4.94E-20 | 0.39832837 | 0.444 | 0.412 | 1.89E-15 | InN5 | PLXNC1 |
| SEL1L33 | 8.65E-20 | 0.29523159 | 0.357 | 0.29 | 3.31E-15 | InN5 | SEL1L3 |
| LINC011224 | 1.22E-19 | 0.26020125 | 0.724 | 0.692 | 4.66E-15 | InN5 | LINC01122 |
| SEM12 | 1.32E-19 | 0.32161815 | 0.523 | 0.482 | 5.05E-15 | InN5 | SEM1 |
| ABHD21 | 1.34E-19 | 0.26288199 | 0.654 | 0.654 | 5.10E-15 | InN5 | ABHD2 |
| SAMD4A2 | 2.65E-19 | 0.33049146 | 0.513 | 0.47 | 1.01E-14 | InN5 | SAMD4A |
| TUBB | 4.17E-19 | 0.27089921 | 0.342 | 0.283 | 1.59E-14 | InN5 | TUBB |
| PLCL22 | 5.27E-19 | 0.27959594 | 0.682 | 0.698 | 2.01E-14 | InN5 | PLCL2 |
| CSGALNACT1 | 6.10E-19 | 0.5022315 | 0.574 | 0.616 | 2.33E-14 | InN5 | CSGALNACT1 |
| SYT4 | 6.33E-19 | 0.29419471 | 0.406 | 0.354 | 2.42E-14 | InN5 | SYT4 |
| CALN14 | 8.15E-19 | 0.26817508 | 0.702 | 0.661 | 3.11E-14 | InN5 | CALN1 |
| SUSD42 | 1.23E-18 | 0.3958594 | 0.613 | 0.612 | 4.69E-14 | InN5 | SUSD4 |
| EPHA54 | 1.68E-18 | 0.25451744 | 0.643 | 0.6 | 6.43E-14 | InN5 | EPHA5 |
| PHYHIPL2 | 2.42E-18 | 0.26030394 | 0.609 | 0.594 | 9.24E-14 | InN5 | PHYHIPL |
| TENM33 | 2.80E-18 | 0.39488053 | 0.685 | 0.675 | 1.07E-13 | InN5 | TENM3 |
| HIPK1 | 4.34E-18 | 0.26842851 | 0.34 | 0.282 | 1.66E-13 | InN5 | HIPK1 |
| RCN2 | 4.41E-18 | 0.28857288 | 0.41 | 0.366 | 1.69E-13 | InN5 | RCN2 |
| SLC9A71 | 5.56E-18 | 0.26159743 | 0.437 | 0.39 | 2.13E-13 | InN5 | SLC9A7 |
| CDH42 | 7.83E-18 | 0.61887941 | 0.449 | 0.428 | 2.99E-13 | InN5 | CDH4 |
| TACC23 | 9.42E-18 | 0.28009083 | 0.453 | 0.413 | 3.60E-13 | InN5 | TACC2 |
| LINC005992 | 1.27E-17 | 0.27386134 | 0.706 | 0.734 | 4.85E-13 | InN5 | LINC00599 |
| FXYD63 | 1.33E-17 | 0.37990138 | 0.587 | 0.622 | 5.09E-13 | InN5 | FXYD6 |
| TCF201 | 1.72E-17 | 0.27141464 | 0.635 | 0.633 | 6.59E-13 | InN5 | TCF20 |
| SRP14 | 2.48E-17 | 0.35038586 | 0.355 | 0.3 | 9.50E-13 | InN5 | SRP14 |
| ATP6V0C1 | 2.67E-17 | 0.38900199 | 0.479 | 0.459 | 1.02E-12 | InN5 | ATP6V0C |
| CD812 | 4.08E-17 | 0.26256775 | 0.303 | 0.242 | 1.56E-12 | InN5 | CD81 |
| GALNT82 | 4.77E-17 | 0.31288088 | 0.356 | 0.301 | 1.82E-12 | InN5 | GALNT8 |
| ATP1A1 | 5.62E-17 | 0.34558965 | 0.598 | 0.603 | 2.15E-12 | InN5 | ATP1A1 |
| MACROD1 | 7.72E-17 | 0.25218463 | 0.278 | 0.217 | 2.95E-12 | InN5 | MACROD1 |
| DIP2A3 | 1.47E-16 | 0.25875929 | 0.494 | 0.462 | 5.62E-12 | InN5 | DIP2A |
| PITPNM3 | 1.56E-16 | 0.28653723 | 0.338 | 0.288 | 5.96E-12 | InN5 | PITPNM3 |
| DKK31 | 1.57E-16 | 0.29886089 | 0.632 | 0.636 | 6.00E-12 | InN5 | DKK3 |
| HMBOX1 | 2.30E-16 | 0.28479195 | 0.586 | 0.593 | 8.79E-12 | InN5 | HMBOX1 |
| SPOCK22 | 2.82E-16 | 0.25692301 | 0.475 | 0.436 | 1.08E-11 | InN5 | SPOCK2 |
| CAPN2 | 3.78E-16 | 0.26305006 | 0.284 | 0.228 | 1.45E-11 | InN5 | CAPN2 |
| ATP1A32 | 4.82E-16 | 0.38550411 | 0.535 | 0.539 | 1.84E-11 | InN5 | ATP1A3 |
| GRM84 | 5.03E-16 | 0.47573751 | 0.405 | 0.354 | 1.92E-11 | InN5 | GRM8 |

| | | | | | | | |
|---|---|---|---|---|---|---|---|
| XPO11 | 5.30E-16 | 0.25363958 | 0.604 | 0.619 | 2.03E-11 | InN5 | XPO1 |
| L3MBTL12 | 8.08E-16 | 0.27662467 | 0.415 | 0.37 | 3.09E-11 | InN5 | L3MBTL1 |
| SCAMP1 | 1.14E-15 | 0.27353307 | 0.52 | 0.514 | 4.36E-11 | InN5 | SCAMP1 |
| HPCAL4 | 1.15E-15 | 0.27266923 | 0.388 | 0.345 | 4.38E-11 | InN5 | HPCAL4 |
| TSPYL4 | 1.43E-15 | 0.29227864 | 0.351 | 0.303 | 5.45E-11 | InN5 | TSPYL4 |
| KCTD82 | 2.82E-15 | 0.46606846 | 0.637 | 0.663 | 1.08E-10 | InN5 | KCTD8 |
| WWP12 | 3.34E-15 | 0.32225212 | 0.513 | 0.486 | 1.28E-10 | InN5 | WWP1 |
| SLC1A11 | 3.43E-15 | 0.25741256 | 0.432 | 0.393 | 1.31E-10 | InN5 | SLC1A1 |
| SLC38A12 | 3.60E-15 | 0.27136652 | 0.589 | 0.6 | 1.38E-10 | InN5 | SLC38A1 |
| ZNF7042 | 5.20E-15 | 0.28365433 | 0.666 | 0.674 | 1.99E-10 | InN5 | ZNF704 |
| ACTG1 | 5.61E-15 | 0.3336429 | 0.413 | 0.377 | 2.14E-10 | InN5 | ACTG1 |
| PTPN21 | 1.28E-14 | 0.30396784 | 0.492 | 0.478 | 4.89E-10 | InN5 | PTPN2 |
| PLD3 | 1.34E-14 | 0.26390012 | 0.654 | 0.686 | 5.12E-10 | InN5 | PLD3 |
| HSPA4L3 | 1.37E-14 | 0.27640941 | 0.5 | 0.479 | 5.25E-10 | InN5 | HSPA4L |
| HSPA51 | 1.39E-14 | 0.28156034 | 0.273 | 0.222 | 5.31E-10 | InN5 | HSPA5 |
| TSHZ11 | 1.47E-14 | 0.32039246 | 0.416 | 0.383 | 5.61E-10 | InN5 | TSHZ1 |
| PTMA1 | 2.58E-14 | 0.3491197 | 0.486 | 0.467 | 9.85E-10 | InN5 | PTMA |
| ZSWIM53 | 2.62E-14 | 0.28775542 | 0.549 | 0.547 | 1.00E-09 | InN5 | ZSWIM5 |
| PGM2L1 | 3.13E-14 | 0.28436316 | 0.516 | 0.506 | 1.20E-09 | InN5 | PGM2L1 |
| UBB | 3.59E-14 | 0.34666457 | 0.386 | 0.338 | 1.37E-09 | InN5 | UBB |
| SHISA62 | 8.34E-14 | 0.39847135 | 0.398 | 0.346 | 3.19E-09 | InN5 | SHISA6 |
| OIP5-AS12 | 1.12E-13 | 0.30534821 | 0.564 | 0.573 | 4.27E-09 | InN5 | OIP5-AS1 |
| ABCC12 | 1.17E-13 | 0.31021093 | 0.394 | 0.357 | 4.48E-09 | InN5 | ABCC1 |
| KIF1A1 | 1.76E-13 | 0.25941803 | 0.605 | 0.639 | 6.74E-09 | InN5 | KIF1A |
| STMN12 | 1.77E-13 | 0.27950708 | 0.501 | 0.477 | 6.77E-09 | InN5 | STMN1 |
| ADAMTS194 | 2.25E-13 | 0.28638207 | 0.423 | 0.368 | 8.61E-09 | InN5 | ADAMTS19 |
| PGAP11 | 3.35E-13 | 0.26313376 | 0.47 | 0.457 | 1.28E-08 | InN5 | PGAP1 |
| RPS27A2 | 4.16E-13 | 0.31484433 | 0.334 | 0.281 | 1.59E-08 | InN5 | RPS27A |
| PRMT81 | 4.54E-13 | 0.30973815 | 0.449 | 0.424 | 1.73E-08 | InN5 | PRMT8 |
| TRIM241 | 4.90E-13 | 0.27721127 | 0.415 | 0.389 | 1.87E-08 | InN5 | TRIM24 |
| LRP12 | 6.70E-13 | 0.29989172 | 0.445 | 0.428 | 2.56E-08 | InN5 | LRP12 |
| TET23 | 1.22E-12 | 0.26723409 | 0.461 | 0.439 | 4.67E-08 | InN5 | TET2 |
| SLC5A32 | 1.84E-12 | 0.27894251 | 0.338 | 0.297 | 7.05E-08 | InN5 | SLC5A3 |
| TMX4 | 2.85E-12 | 0.31297848 | 0.446 | 0.44 | 1.09E-07 | InN5 | TMX4 |
| PCSK1N1 | 2.85E-12 | 0.31883429 | 0.472 | 0.46 | 1.09E-07 | InN5 | PCSK1N |
| AL445623.22 | 3.90E-12 | 0.28339299 | 0.423 | 0.378 | 1.49E-07 | InN5 | AL445623.2 |
| CLVS11 | 5.39E-12 | 0.26051337 | 0.635 | 0.66 | 2.06E-07 | InN5 | CLVS1 |
| TERF2IP | 7.63E-12 | 0.27058171 | 0.558 | 0.566 | 2.92E-07 | InN5 | TERF2IP |
| KLC1 | 8.03E-12 | 0.26511343 | 0.633 | 0.661 | 3.07E-07 | InN5 | KLC1 |
| SARAF | 8.07E-12 | 0.31972147 | 0.557 | 0.577 | 3.08E-07 | InN5 | SARAF |
| RPL13A2 | 8.13E-12 | 0.31077325 | 0.337 | 0.298 | 3.11E-07 | InN5 | RPL13A |
| PUS7L1 | 9.91E-12 | 0.2784521 | 0.475 | 0.468 | 3.79E-07 | InN5 | PUS7L |
| LRRTM21 | 1.03E-11 | 0.25488737 | 0.337 | 0.3 | 3.92E-07 | InN5 | LRRTM2 |
| H3F3B | 1.04E-11 | 0.277713 | 0.331 | 0.29 | 3.97E-07 | InN5 | H3F3B |
| UBC | 1.18E-11 | 0.30760912 | 0.407 | 0.374 | 4.50E-07 | InN5 | UBC |
| REPS21 | 1.42E-11 | 0.26005517 | 0.661 | 0.71 | 5.41E-07 | InN5 | REPS2 |
| SEZ6L3 | 3.27E-11 | 0.2554687 | 0.72 | 0.73 | 1.25E-06 | InN5 | SEZ6L |
| PDE4A | 3.41E-11 | 0.25986289 | 0.451 | 0.432 | 1.30E-06 | InN5 | PDE4A |
| SLMAP | 5.75E-11 | 0.27425349 | 0.574 | 0.593 | 2.20E-06 | InN5 | SLMAP |
| CCBE11 | 6.15E-11 | 0.31463985 | 0.263 | 0.216 | 2.35E-06 | InN5 | CCBE1 |
| TUBA1A | 9.68E-11 | 0.30117997 | 0.355 | 0.323 | 3.70E-06 | InN5 | TUBA1A |
| RGS7BP1 | 1.12E-10 | 0.26266277 | 0.609 | 0.637 | 4.30E-06 | InN5 | RGS7BP |
| CALY1 | 1.15E-10 | 0.29085533 | 0.533 | 0.551 | 4.38E-06 | InN5 | CALY |
| PPP2R1A | 5.66E-10 | 0.25296399 | 0.43 | 0.421 | 2.16E-05 | InN5 | PPP2R1A |
| LYPLA1 | 5.88E-10 | 0.26993276 | 0.271 | 0.234 | 2.25E-05 | InN5 | LYPLA1 |
| FEZ1 | 8.29E-10 | 0.28728069 | 0.412 | 0.399 | 3.17E-05 | InN5 | FEZ1 |
| TMEM179 | 8.73E-10 | 0.26531933 | 0.324 | 0.293 | 3.34E-05 | InN5 | TMEM179 |
| SLITRK4 | 1.17E-09 | 0.25695832 | 0.396 | 0.372 | 4.49E-05 | InN5 | SLITRK4 |
| ATAD2B2 | 1.24E-09 | 0.25804263 | 0.448 | 0.44 | 4.76E-05 | InN5 | ATAD2B |
| MARCH42 | 1.33E-09 | 0.25716474 | 0.426 | 0.408 | 5.07E-05 | InN5 | 4-Mar |
| SPINT2 | 2.44E-09 | 0.26512196 | 0.306 | 0.279 | 9.32E-05 | InN5 | SPINT2 |
| LRRC4B | 2.62E-09 | 0.25065838 | 0.517 | 0.518 | 1.00E-04 | InN5 | LRRC4B |
| RPS282 | 2.66E-09 | 0.31075578 | 0.384 | 0.359 | 0.00010155 | InN5 | RPS28 |

| | | | | | | | |
|---|---|---|---|---|---|---|---|
| CDHR33 | 4.13E-09 | 0.30163931 | 0.407 | 0.381 | 0.00015776 | InN5 | CDHR3 |
| RNF1573 | 4.24E-09 | 0.25848902 | 0.521 | 0.53 | 0.00016196 | InN5 | RNF157 |
| MORF4L1 | 5.17E-09 | 0.26207657 | 0.52 | 0.533 | 0.00019747 | InN5 | MORF4L1 |
| CCSAP | 7.66E-09 | 0.25006859 | 0.473 | 0.481 | 0.00029285 | InN5 | CCSAP |
| ADCY3 | 8.68E-09 | 0.2680493 | 0.4 | 0.387 | 0.00033183 | InN5 | ADCY3 |
| AC011346.11 | 9.32E-09 | 0.28367169 | 0.36 | 0.332 | 0.00035627 | InN5 | AC011346.1 |
| H3F3A | 1.46E-08 | 0.29175855 | 0.292 | 0.263 | 0.00055919 | InN5 | H3F3A |
| PALM2-AKAF | 2.32E-08 | 0.28185167 | 0.655 | 0.707 | 0.00088567 | InN5 | PALM2-AKAP |
| GAPDH1 | 2.78E-08 | 0.3040799 | 0.526 | 0.53 | 0.00106441 | InN5 | GAPDH |
| RPS6KA3 | 7.30E-08 | 0.26897402 | 0.562 | 0.597 | 0.00279021 | InN5 | RPS6KA3 |
| RPS272 | 7.92E-08 | 0.28124391 | 0.334 | 0.308 | 0.00302856 | InN5 | RPS27 |
| RPL321 | 1.79E-06 | 0.29193623 | 0.384 | 0.368 | 0.06844201 | InN5 | RPL32 |
| ATP2C1 | 2.28E-06 | 0.25505516 | 0.63 | 0.678 | 0.0873076 | InN5 | ATP2C1 |
| HSP90B1 | 2.35E-06 | 0.25117438 | 0.446 | 0.453 | 0.08993138 | InN5 | HSP90B1 |
| PXK1 | 6.33E-06 | 0.26499857 | 0.464 | 0.473 | 0.24183417 | InN5 | PXK |
| HOMER21 | 6.74E-06 | 0.25136853 | 0.354 | 0.346 | 0.25755293 | InN5 | HOMER2 |
| NKAIN32 | 8.94E-06 | 0.48161863 | 0.294 | 0.28 | 0.34188746 | InN5 | NKAIN3 |
| RPL411 | 1.13E-05 | 0.26450264 | 0.358 | 0.343 | 0.4303933 | InN5 | RPL41 |
| ALCAM3 | 1.30E-05 | 0.27949019 | 0.476 | 0.473 | 0.49704645 | InN5 | ALCAM |
| RPL191 | 1.94E-05 | 0.30326561 | 0.297 | 0.28 | 0.74326949 | InN5 | RPL19 |
| RPS152 | 2.82E-05 | 0.26621119 | 0.31 | 0.29 | 1 | InN5 | RPS15 |
| CREB51 | 3.89E-05 | 0.45755138 | 0.282 | 0.274 | 1 | InN5 | CREB5 |
| WWC11 | 4.30E-05 | 0.28677561 | 0.354 | 0.359 | 1 | InN5 | WWC1 |
| AC005225.2 | 0.00017457 | 0.25360032 | 0.4 | 0.404 | 1 | InN5 | AC005225.2 |
| RPL15 | 0.00041683 | 0.2522672 | 0.362 | 0.363 | 1 | InN5 | RPL15 |
| RPL61 | 0.00085951 | 0.26609236 | 0.324 | 0.323 | 1 | InN5 | RPL6 |
| MPP62 | 0.00086548 | 0.33857884 | 0.53 | 0.589 | 1 | InN5 | MPP6 |
| RPS251 | 0.00250236 | 0.25749574 | 0.298 | 0.293 | 1 | InN5 | RPS25 |
| AC006115.21 | 0.00376154 | 0.26423017 | 0.37 | 0.39 | 1 | InN5 | AC006115.2 |
| MPPED12 | 0.00682069 | 0.35101051 | 0.405 | 0.457 | 1 | InN5 | MPPED1 |
| RPL261 | 0.00750977 | 0.2556995 | 0.278 | 0.272 | 1 | InN5 | RPL26 |
| RELN2 | 0 | 3.29692493 | 0.725 | 0.129 | 0 | InN6 | RELN |
| CXCL143 | 0 | 2.51824417 | 0.873 | 0.147 | 0 | InN6 | CXCL14 |
| CNR12 | 0 | 2.44727658 | 0.838 | 0.378 | 0 | InN6 | CNR1 |
| ADARB23 | 0 | 2.38785636 | 0.997 | 0.363 | 0 | InN6 | ADARB2 |
| ZNF385D6 | 0 | 2.36400803 | 0.97 | 0.51 | 0 | InN6 | ZNF385D |
| CDH43 | 0 | 2.13711328 | 0.904 | 0.421 | 0 | InN6 | CDH4 |
| SORCS34 | 0 | 2.07450099 | 0.951 | 0.565 | 0 | InN6 | SORCS3 |
| RBMS34 | 0 | 2.065819 | 0.989 | 0.636 | 0 | InN6 | RBMS3 |
| AC006305.1 | 0 | 1.97082612 | 0.573 | 0.009 | 0 | InN6 | AC006305.1 |
| MAML33 | 0 | 1.92817844 | 0.882 | 0.356 | 0 | InN6 | MAML3 |
| SLC35F44 | 0 | 1.88921937 | 0.873 | 0.227 | 0 | InN6 | SLC35F4 |
| DDR22 | 0 | 1.87716355 | 0.608 | 0.078 | 0 | InN6 | DDR2 |
| SDK23 | 0 | 1.77313747 | 0.8 | 0.226 | 0 | InN6 | SDK2 |
| MYO162 | 0 | 1.75839681 | 0.844 | 0.281 | 0 | InN6 | MYO16 |
| NR2F2-AS12 | 0 | 1.73065901 | 0.82 | 0.202 | 0 | InN6 | NR2F2-AS1 |
| CHRNA72 | 0 | 1.70230049 | 0.753 | 0.172 | 0 | InN6 | CHRNA7 |
| EYA1 | 0 | 1.47930454 | 0.371 | 0.038 | 0 | InN6 | EYA1 |
| PLS32 | 0 | 1.46457562 | 0.756 | 0.194 | 0 | InN6 | PLS3 |
| LINC01416 | 0 | 1.36464014 | 0.419 | 0.012 | 0 | InN6 | LINC01416 |
| SLC25A483 | 0 | 1.25347619 | 0.687 | 0.116 | 0 | InN6 | SLC25A48 |
| FBXL71 | 0 | 1.15613927 | 0.733 | 0.167 | 0 | InN6 | FBXL7 |
| WIF11 | 0 | 1.14065598 | 0.532 | 0.051 | 0 | InN6 | WIF1 |
| CDH231 | 0 | 0.88991188 | 0.314 | 0.025 | 0 | InN6 | CDH23 |
| ST8SIA6 | 0 | 0.68120295 | 0.276 | 0.025 | 0 | InN6 | ST8SIA6 |
| PAX6 | 0 | 0.67252715 | 0.336 | 0.011 | 0 | InN6 | PAX6 |
| CPLX31 | 0 | 0.66313027 | 0.293 | 0.026 | 0 | InN6 | CPLX3 |
| SP8 | 0 | 0.47080308 | 0.272 | 0.015 | 0 | InN6 | SP8 |
| NRXN33 | 4.33E-303 | 1.14741418 | 0.999 | 0.99 | 1.65E-298 | InN6 | NRXN3 |
| KCNIP14 | 9.82E-303 | 1.31679927 | 0.89 | 0.299 | 3.76E-298 | InN6 | KCNIP1 |
| ARHGAP422 | 2.34E-299 | 0.91178872 | 0.469 | 0.086 | 8.95E-295 | InN6 | ARHGAP42 |
| GRIP14 | 1.39E-296 | 1.66977422 | 0.995 | 0.798 | 5.33E-292 | InN6 | GRIP1 |
| PLD53 | 3.52E-293 | 1.76345302 | 0.846 | 0.318 | 1.34E-288 | InN6 | PLD5 |

| | | | | | | | |
|---|---|---|---|---|---|---|---|
| ROR11 | 1.11E-290 | 1.13472183 | 0.572 | 0.126 | 4.24E-286 | InN6 | ROR1 |
| IL1RAPL24 | 8.52E-290 | 1.7947869 | 0.861 | 0.317 | 3.26E-285 | InN6 | IL1RAPL2 |
| FRMD4B4 | 3.16E-287 | 1.14929596 | 0.64 | 0.164 | 1.21E-282 | InN6 | FRMD4B |
| NPAS33 | 3.89E-286 | 1.76122672 | 0.988 | 0.817 | 1.49E-281 | InN6 | NPAS3 |
| NR2F22 | 1.03E-279 | 0.89949198 | 0.522 | 0.109 | 3.93E-275 | InN6 | NR2F2 |
| RGS122 | 1.20E-271 | 1.51036898 | 0.947 | 0.512 | 4.58E-267 | InN6 | RGS12 |
| ANGPT11 | 4.37E-271 | 1.02271588 | 0.427 | 0.078 | 1.67E-266 | InN6 | ANGPT1 |
| ZNF5364 | 4.67E-268 | 1.34299435 | 0.942 | 0.368 | 1.78E-263 | InN6 | ZNF536 |
| SYT6 | 2.02E-267 | 0.5158378 | 0.276 | 0.034 | 7.71E-263 | InN6 | SYT6 |
| SHISA63 | 3.23E-267 | 1.44679946 | 0.846 | 0.339 | 1.24E-262 | InN6 | SHISA6 |
| AC064875.1 | 4.11E-266 | 0.76354474 | 0.318 | 0.045 | 1.57E-261 | InN6 | AC064875.1 |
| AC073050.13 | 1.18E-265 | 1.86290329 | 0.954 | 0.772 | 4.53E-261 | InN6 | AC073050.1 |
| WLS3 | 9.15E-264 | 1.28797096 | 0.762 | 0.271 | 3.50E-259 | InN6 | WLS |
| L3MBTL43 | 3.06E-261 | 1.52534849 | 0.88 | 0.395 | 1.17E-256 | InN6 | L3MBTL4 |
| SLC7A22 | 9.08E-259 | 0.72683574 | 0.42 | 0.078 | 3.47E-254 | InN6 | SLC7A2 |
| GRIK23 | 3.17E-257 | 1.31076725 | 0.991 | 0.948 | 1.21E-252 | InN6 | GRIK2 |
| HS3ST53 | 1.01E-255 | 1.36219598 | 0.794 | 0.31 | 3.86E-251 | InN6 | HS3ST5 |
| AP1S23 | 1.42E-251 | 1.12916802 | 0.645 | 0.198 | 5.41E-247 | InN6 | AP1S2 |
| ALK3 | 3.05E-247 | 1.7415105 | 0.702 | 0.243 | 1.17E-242 | InN6 | ALK |
| KIAA12115 | 9.19E-247 | 1.32365352 | 0.883 | 0.395 | 3.51E-242 | InN6 | KIAA1211 |
| MAML22 | 2.35E-243 | 1.54189668 | 0.668 | 0.229 | 8.97E-239 | InN6 | MAML2 |
| HMBOX11 | 1.40E-234 | 1.43493247 | 0.865 | 0.588 | 5.37E-230 | InN6 | HMBOX1 |
| RASSF32 | 6.68E-232 | 0.72351081 | 0.422 | 0.087 | 2.55E-227 | InN6 | RASSF3 |
| PRKCA5 | 5.20E-231 | 1.23838278 | 0.966 | 0.733 | 1.99E-226 | InN6 | PRKCA |
| DLX6-AS13 | 1.33E-229 | 1.05982024 | 0.732 | 0.236 | 5.07E-225 | InN6 | DLX6-AS1 |
| ATP11C2 | 4.73E-226 | 0.63979525 | 0.385 | 0.072 | 1.81E-221 | InN6 | ATP11C |
| GALNTL64 | 2.66E-222 | 1.59633154 | 0.957 | 0.741 | 1.02E-217 | InN6 | GALNTL6 |
| WWP13 | 2.12E-221 | 1.3546075 | 0.838 | 0.481 | 8.11E-217 | InN6 | WWP1 |
| TNFAIP8L31 | 1.95E-215 | 0.68928776 | 0.389 | 0.079 | 7.45E-211 | InN6 | TNFAIP8L3 |
| NFIB3 | 4.79E-215 | 1.268382 | 0.957 | 0.647 | 1.83E-210 | InN6 | NFIB |
| LINC004722 | 4.04E-213 | 0.89891571 | 0.531 | 0.145 | 1.54E-208 | InN6 | LINC00472 |
| DOCK104 | 2.98E-212 | 1.134633 | 0.876 | 0.4 | 1.14E-207 | InN6 | DOCK10 |
| NXPH13 | 4.27E-204 | 1.53833359 | 0.741 | 0.266 | 1.63E-199 | InN6 | NXPH1 |
| PITPNC15 | 3.72E-203 | 1.18682001 | 0.892 | 0.49 | 1.42E-198 | InN6 | PITPNC1 |
| CNTNAP24 | 1.31E-201 | 1.180007 | 1 | 0.993 | 5.02E-197 | InN6 | CNTNAP2 |
| PID13 | 1.00E-198 | 1.37227001 | 0.855 | 0.495 | 3.83E-194 | InN6 | PID1 |
| PBX31 | 2.73E-194 | 1.307872 | 0.507 | 0.15 | 1.04E-189 | InN6 | PBX3 |
| ROBO13 | 4.68E-193 | 1.52860577 | 0.949 | 0.658 | 1.79E-188 | InN6 | ROBO1 |
| ENOX13 | 1.14E-191 | 1.27174195 | 0.962 | 0.85 | 4.37E-187 | InN6 | ENOX1 |
| SHISA93 | 5.37E-190 | 1.09425247 | 0.97 | 0.822 | 2.05E-185 | InN6 | SHISA9 |
| NPAS12 | 2.39E-188 | 0.82989423 | 0.501 | 0.143 | 9.13E-184 | InN6 | NPAS1 |
| SGCD3 | 3.53E-187 | 1.60738395 | 0.894 | 0.589 | 1.35E-182 | InN6 | SGCD |
| FSTL53 | 1.53E-185 | 1.25619911 | 0.898 | 0.497 | 5.84E-181 | InN6 | FSTL5 |
| FAM110B4 | 8.23E-183 | 1.15552357 | 0.795 | 0.429 | 3.15E-178 | InN6 | FAM110B |
| KAZN5 | 1.98E-178 | 0.86873319 | 0.999 | 0.953 | 7.57E-174 | InN6 | KAZN |
| SEMA6D4 | 4.36E-176 | 1.28694672 | 0.894 | 0.581 | 1.67E-171 | InN6 | SEMA6D |
| MTSS16 | 3.62E-174 | 1.0436946 | 0.886 | 0.566 | 1.38E-169 | InN6 | MTSS1 |
| SPOCK33 | 9.15E-174 | 1.25749922 | 0.928 | 0.552 | 3.50E-169 | InN6 | SPOCK3 |
| NRIP34 | 3.15E-173 | 0.83416516 | 0.611 | 0.214 | 1.20E-168 | InN6 | NRIP3 |
| RAB3C4 | 5.37E-172 | 1.06847211 | 0.901 | 0.636 | 2.05E-167 | InN6 | RAB3C |
| EPS83 | 5.42E-172 | 0.82149902 | 0.442 | 0.118 | 2.07E-167 | InN6 | EPS8 |
| PNOC1 | 8.99E-172 | 0.44328622 | 0.25 | 0.04 | 3.44E-167 | InN6 | PNOC |
| MCC4 | 6.67E-171 | 0.95736 | 0.681 | 0.26 | 2.55E-166 | InN6 | MCC |
| CNTN55 | 1.30E-164 | 1.34440266 | 0.959 | 0.815 | 4.97E-160 | InN6 | CNTN5 |
| TMEM200C1 | 2.12E-164 | 0.5465176 | 0.353 | 0.08 | 8.09E-160 | InN6 | TMEM200C |
| SLC6A16 | 2.88E-164 | 0.86191582 | 0.824 | 0.351 | 1.10E-159 | InN6 | SLC6A1 |
| PTPRM4 | 1.18E-163 | 1.1907021 | 0.901 | 0.58 | 4.50E-159 | InN6 | PTPRM |
| AC068722.1 | 5.22E-163 | 0.71500474 | 0.343 | 0.077 | 2.00E-158 | InN6 | AC068722.1 |
| PARD34 | 1.46E-160 | 1.02658058 | 0.745 | 0.317 | 5.58E-156 | InN6 | PARD3 |
| EGFR3 | 4.50E-159 | 0.79810836 | 0.682 | 0.238 | 1.72E-154 | InN6 | EGFR |
| CDHR34 | 6.92E-159 | 1.13190431 | 0.749 | 0.376 | 2.64E-154 | InN6 | CDHR3 |
| ROBO23 | 1.52E-157 | 1.08816123 | 0.999 | 0.851 | 5.79E-153 | InN6 | ROBO2 |
| QKI6 | 6.91E-156 | 0.97684505 | 0.922 | 0.671 | 2.64E-151 | InN6 | QKI |

| | | | | | | | |
|---|---|---|---|---|---|---|---|
| SCN7A1 | 1.15E-154 | 0.75657615 | 0.396 | 0.105 | 4.40E-150 | InN6 | SCN7A |
| PRR162 | 1.13E-152 | 1.31570006 | 0.834 | 0.548 | 4.31E-148 | InN6 | PRR16 |
| MSI25 | 8.30E-151 | 0.97343087 | 0.895 | 0.658 | 3.17E-146 | InN6 | MSI2 |
| SYTL51 | 1.31E-150 | 0.70054285 | 0.401 | 0.106 | 5.01E-146 | InN6 | SYTL5 |
| FGD51 | 1.55E-150 | 0.52616514 | 0.325 | 0.073 | 5.91E-146 | InN6 | FGD5 |
| PDLIM5 | 4.47E-148 | 0.9688057 | 0.359 | 0.093 | 1.71E-143 | InN6 | PDLIM5 |
| DAB15 | 2.47E-147 | 0.86257226 | 0.979 | 0.878 | 9.44E-143 | InN6 | DAB1 |
| CCDC85A4 | 1.14E-146 | 1.21007616 | 0.862 | 0.655 | 4.36E-142 | InN6 | CCDC85A |
| PWRN44 | 3.90E-146 | 0.84728111 | 0.597 | 0.222 | 1.49E-141 | InN6 | PWRN4 |
| ABTB2 | 8.33E-146 | 0.52218402 | 0.281 | 0.057 | 3.19E-141 | InN6 | ABTB2 |
| ENTPD34 | 6.46E-144 | 0.89613251 | 0.65 | 0.292 | 2.47E-139 | InN6 | ENTPD3 |
| CHST114 | 8.33E-143 | 1.06684737 | 0.833 | 0.511 | 3.19E-138 | InN6 | CHST11 |
| NRXN12 | 4.79E-142 | 0.63748036 | 1 | 0.986 | 1.83E-137 | InN6 | NRXN1 |
| TNS32 | 2.33E-141 | 0.67907222 | 0.388 | 0.107 | 8.91E-137 | InN6 | TNS3 |
| SILC14 | 1.12E-139 | 0.77151896 | 0.583 | 0.216 | 4.30E-135 | InN6 | SILC1 |
| MN11 | 2.37E-139 | 0.58977045 | 0.348 | 0.089 | 9.07E-135 | InN6 | MN1 |
| HIF3A1 | 2.94E-138 | 0.52500983 | 0.31 | 0.071 | 1.12E-133 | InN6 | HIF3A |
| DCHS22 | 1.06E-137 | 0.67910479 | 0.415 | 0.123 | 4.06E-133 | InN6 | DCHS2 |
| CLMP2 | 2.97E-136 | 0.85727202 | 0.443 | 0.141 | 1.14E-131 | InN6 | CLMP |
| PLCXD33 | 7.14E-136 | 1.22840089 | 0.664 | 0.309 | 2.73E-131 | InN6 | PLCXD3 |
| FBN21 | 1.01E-135 | 0.65744923 | 0.252 | 0.05 | 3.85E-131 | InN6 | FBN2 |
| AC233296.13 | 1.29E-133 | 0.89086989 | 0.649 | 0.266 | 4.94E-129 | InN6 | AC233296.1 |
| PWRN15 | 5.44E-133 | 0.90289903 | 0.848 | 0.467 | 2.08E-128 | InN6 | PWRN1 |
| GRIK13 | 2.39E-131 | 0.59036753 | 0.824 | 0.389 | 9.13E-127 | InN6 | GRIK1 |
| DPP62 | 3.31E-130 | 0.89058598 | 0.987 | 0.917 | 1.26E-125 | InN6 | DPP6 |
| SPHKAP3 | 2.72E-129 | 1.07524308 | 0.474 | 0.162 | 1.04E-124 | InN6 | SPHKAP |
| KCNT23 | 1.23E-128 | 0.97008276 | 0.854 | 0.544 | 4.71E-124 | InN6 | KCNT2 |
| GABRG33 | 2.63E-128 | 0.8508108 | 0.95 | 0.8 | 1.01E-123 | InN6 | GABRG3 |
| ERBB44 | 3.38E-128 | 0.49849777 | 0.986 | 0.572 | 1.29E-123 | InN6 | ERBB4 |
| CUBN | 5.74E-128 | 0.59824595 | 0.327 | 0.085 | 2.19E-123 | InN6 | CUBN |
| ADAMTS62 | 1.07E-126 | 0.91059013 | 0.413 | 0.127 | 4.09E-122 | InN6 | ADAMTS6 |
| CCK2 | 2.39E-126 | 1.93911163 | 0.706 | 0.453 | 9.15E-122 | InN6 | CCK |
| VWC23 | 3.52E-125 | 0.98379003 | 0.719 | 0.353 | 1.34E-120 | InN6 | VWC2 |
| GRIP25 | 3.33E-124 | 0.6174956 | 0.53 | 0.187 | 1.27E-119 | InN6 | GRIP2 |
| NR3C22 | 4.40E-124 | 0.83264594 | 0.93 | 0.804 | 1.68E-119 | InN6 | NR3C2 |
| RIMS22 | 3.55E-122 | 0.6866875 | 0.991 | 0.968 | 1.36E-117 | InN6 | RIMS2 |
| DENND2A3 | 3.86E-122 | 0.85799329 | 0.598 | 0.265 | 1.47E-117 | InN6 | DENND2A |
| GREB1L2 | 4.46E-122 | 1.02789282 | 0.653 | 0.312 | 1.70E-117 | InN6 | GREB1L |
| GRID23 | 9.10E-119 | 0.96648922 | 0.961 | 0.733 | 3.48E-114 | InN6 | GRID2 |
| SLCO5A12 | 4.37E-118 | 0.59633441 | 0.378 | 0.114 | 1.67E-113 | InN6 | SLCO5A1 |
| CHL13 | 2.15E-114 | 0.81972232 | 0.926 | 0.805 | 8.23E-110 | InN6 | CHL1 |
| SCG22 | 4.69E-114 | 0.89097379 | 0.581 | 0.259 | 1.79E-109 | InN6 | SCG2 |
| EEPD12 | 6.52E-114 | 0.93089141 | 0.678 | 0.382 | 2.49E-109 | InN6 | EEPD1 |
| PLPPR15 | 1.81E-112 | 0.82772193 | 0.635 | 0.294 | 6.93E-108 | InN6 | PLPPR1 |
| FGF133 | 1.99E-112 | 0.89083484 | 0.711 | 0.416 | 7.60E-108 | InN6 | FGF13 |
| TP53I111 | 7.82E-112 | 0.67252782 | 0.398 | 0.137 | 2.99E-107 | InN6 | TP53I11 |
| LINC01470 | 1.56E-111 | 0.99943748 | 0.301 | 0.081 | 5.96E-107 | InN6 | LINC01470 |
| TCF45 | 1.79E-111 | 0.59371046 | 0.986 | 0.957 | 6.82E-107 | InN6 | TCF4 |
| TMEM132C2 | 4.34E-111 | 0.6549177 | 0.616 | 0.261 | 1.66E-106 | InN6 | TMEM132C |
| BCL11B5 | 2.00E-110 | 0.82157281 | 0.582 | 0.255 | 7.63E-106 | InN6 | BCL11B |
| SH3D191 | 2.42E-110 | 0.99208534 | 0.725 | 0.457 | 9.25E-106 | InN6 | SH3D19 |
| DLX13 | 8.93E-108 | 0.42989765 | 0.342 | 0.097 | 3.41E-103 | InN6 | DLX1 |
| GAD24 | 7.93E-107 | 0.49928528 | 0.649 | 0.256 | 3.03E-102 | InN6 | GAD2 |
| PALLD4 | 2.95E-106 | 0.94451276 | 0.695 | 0.414 | 1.13E-101 | InN6 | PALLD |
| DNER4 | 4.47E-106 | 0.75527732 | 0.82 | 0.526 | 1.71E-101 | InN6 | DNER |
| DSCAML15 | 2.31E-105 | 0.75576931 | 0.93 | 0.757 | 8.84E-101 | InN6 | DSCAML1 |
| KCNJ33 | 3.40E-105 | 0.74161872 | 0.942 | 0.868 | 1.30E-100 | InN6 | KCNJ3 |
| KIRREL32 | 7.01E-103 | 0.94034532 | 0.934 | 0.792 | 2.68E-98 | InN6 | KIRREL3 |
| TMTC23 | 6.99E-102 | 0.94940377 | 0.815 | 0.538 | 2.67E-97 | InN6 | TMTC2 |
| GCNT23 | 7.45E-101 | 0.35881627 | 0.284 | 0.074 | 2.85E-96 | InN6 | GCNT2 |
| IGF15 | 7.27E-100 | 0.63278054 | 0.426 | 0.15 | 2.78E-95 | InN6 | IGF1 |
| LRRC3B2 | 4.00E-99 | 0.64261951 | 0.572 | 0.252 | 1.53E-94 | InN6 | LRRC3B |
| AL391832.42 | 1.62E-98 | 0.53359807 | 0.261 | 0.066 | 6.21E-94 | InN6 | AL391832.4 |

| | | | | | | | |
|---|---|---|---|---|---|---|---|
| NTRK23 | 3.80E-98 | 0.68195795 | 0.963 | 0.944 | 1.45E-93 | InN6 | NTRK2 |
| SPOCK14 | 5.00E-98 | 0.71533262 | 0.946 | 0.748 | 1.91E-93 | InN6 | SPOCK1 |
| NHS5 | 3.18E-97 | 0.76015707 | 0.762 | 0.448 | 1.22E-92 | InN6 | NHS |
| ANO44 | 4.73E-97 | 0.67929656 | 0.87 | 0.597 | 1.81E-92 | InN6 | ANO4 |
| ZNF804B5 | 5.14E-96 | 1.14935992 | 0.648 | 0.356 | 1.96E-91 | InN6 | ZNF804B |
| B3GAT22 | 1.20E-93 | 0.51156884 | 0.365 | 0.124 | 4.58E-89 | InN6 | B3GAT2 |
| RASSF84 | 1.38E-92 | 0.63749266 | 0.444 | 0.176 | 5.28E-88 | InN6 | RASSF8 |
| UNC5B1 | 3.01E-92 | 0.57690765 | 0.401 | 0.149 | 1.15E-87 | InN6 | UNC5B |
| SYT173 | 8.69E-92 | 0.66244123 | 0.593 | 0.297 | 3.32E-87 | InN6 | SYT17 |
| SVEP11 | 3.47E-91 | 0.67742296 | 0.476 | 0.199 | 1.33E-86 | InN6 | SVEP1 |
| CACNG43 | 1.32E-90 | 0.5014594 | 0.426 | 0.158 | 5.04E-86 | InN6 | CACNG4 |
| DNAH2 | 5.69E-90 | 0.47681283 | 0.269 | 0.077 | 2.18E-85 | InN6 | DNAH2 |
| KLHL292 | 1.10E-89 | 1.07171905 | 0.819 | 0.699 | 4.20E-85 | InN6 | KLHL29 |
| PRKD13 | 1.28E-89 | 0.88217021 | 0.77 | 0.509 | 4.89E-85 | InN6 | PRKD1 |
| SLC24A34 | 4.30E-89 | 0.36151322 | 0.693 | 0.309 | 1.64E-84 | InN6 | SLC24A3 |
| ELAVL23 | 1.32E-88 | 0.7075258 | 0.796 | 0.519 | 5.05E-84 | InN6 | ELAVL2 |
| FOXO12 | 3.29E-88 | 0.63817657 | 0.361 | 0.127 | 1.26E-83 | InN6 | FOXO1 |
| GABRA12 | 1.29E-87 | 0.79146545 | 0.8 | 0.641 | 4.94E-83 | InN6 | GABRA1 |
| PDE4B3 | 1.32E-87 | 0.81531899 | 0.838 | 0.611 | 5.05E-83 | InN6 | PDE4B |
| FGF142 | 3.67E-87 | 0.45264083 | 0.996 | 0.989 | 1.40E-82 | InN6 | FGF14 |
| TLE22 | 6.60E-87 | 0.64275052 | 0.457 | 0.198 | 2.52E-82 | InN6 | TLE2 |
| PLCB42 | 1.71E-86 | 0.85260906 | 0.77 | 0.562 | 6.52E-82 | InN6 | PLCB4 |
| TIMP24 | 5.12E-86 | 0.68502036 | 0.589 | 0.302 | 1.96E-81 | InN6 | TIMP2 |
| MARCH113 | 2.62E-85 | 0.78686809 | 0.597 | 0.34 | 1.00E-80 | InN6 | 11-Mar |
| INHBA-AS12 | 2.38E-84 | 0.50504344 | 0.399 | 0.15 | 9.10E-80 | InN6 | INHBA-AS1 |
| COL16A11 | 2.79E-84 | 0.74864984 | 0.422 | 0.183 | 1.07E-79 | InN6 | COL16A1 |
| SHISAL1 | 3.87E-84 | 0.54299007 | 0.38 | 0.146 | 1.48E-79 | InN6 | SHISAL1 |
| RERG3 | 3.29E-83 | 0.84703194 | 0.545 | 0.268 | 1.26E-78 | InN6 | RERG |
| DOCK91 | 4.54E-83 | 0.721869 | 0.857 | 0.752 | 1.73E-78 | InN6 | DOCK9 |
| ADRA1A4 | 4.54E-83 | 0.78914679 | 0.488 | 0.211 | 1.74E-78 | InN6 | ADRA1A |
| RIPOR24 | 1.20E-82 | 0.69985425 | 0.724 | 0.466 | 4.57E-78 | InN6 | RIPOR2 |
| DCX5 | 1.34E-82 | 0.75290716 | 0.649 | 0.395 | 5.11E-78 | InN6 | DCX |
| CHRM24 | 1.91E-82 | 0.65914875 | 0.498 | 0.215 | 7.29E-78 | InN6 | CHRM2 |
| HIVEP13 | 2.05E-82 | 0.76321889 | 0.691 | 0.46 | 7.83E-78 | InN6 | HIVEP1 |
| SEZ6L4 | 2.96E-82 | 0.77556775 | 0.865 | 0.727 | 1.13E-77 | InN6 | SEZ6L |
| MACROD23 | 8.67E-82 | 0.8493569 | 0.976 | 0.951 | 3.31E-77 | InN6 | MACROD2 |
| INSYN2A1 | 8.93E-82 | 0.48615503 | 0.302 | 0.097 | 3.41E-77 | InN6 | INSYN2A |
| SCN9A1 | 2.32E-81 | 0.628465 | 0.449 | 0.199 | 8.86E-77 | InN6 | SCN9A |
| NINL4 | 3.18E-80 | 0.49897679 | 0.451 | 0.186 | 1.22E-75 | InN6 | NINL |
| AMOTL14 | 3.38E-80 | 0.51863302 | 0.474 | 0.202 | 1.29E-75 | InN6 | AMOTL1 |
| RGS81 | 7.46E-80 | 0.33477676 | 0.261 | 0.076 | 2.85E-75 | InN6 | RGS8 |
| TCF126 | 1.07E-79 | 0.68925645 | 0.85 | 0.704 | 4.08E-75 | InN6 | TCF12 |
| LHFPL33 | 9.46E-79 | 0.68154676 | 0.568 | 0.273 | 3.62E-74 | InN6 | LHFPL3 |
| GABRG11 | 3.30E-78 | 0.50357159 | 0.364 | 0.134 | 1.26E-73 | InN6 | GABRG1 |
| PIK3R33 | 5.75E-78 | 0.76561106 | 0.573 | 0.316 | 2.20E-73 | InN6 | PIK3R3 |
| PLCB12 | 6.55E-78 | 0.55108371 | 0.996 | 0.978 | 2.51E-73 | InN6 | PLCB1 |
| GLYATL2 | 3.81E-77 | 0.47818815 | 0.369 | 0.143 | 1.46E-72 | InN6 | GLYATL2 |
| ZNF4623 | 8.09E-77 | 0.64359048 | 0.548 | 0.286 | 3.09E-72 | InN6 | ZNF462 |
| SYNE12 | 7.00E-76 | 0.54628388 | 0.978 | 0.952 | 2.67E-71 | InN6 | SYNE1 |
| FNBP1L5 | 7.84E-75 | 0.64925474 | 0.654 | 0.388 | 3.00E-70 | InN6 | FNBP1L |
| AC087564.14 | 3.03E-74 | 0.52765145 | 0.377 | 0.14 | 1.16E-69 | InN6 | AC087564.1 |
| GAD15 | 5.34E-74 | 0.45579601 | 0.627 | 0.286 | 2.04E-69 | InN6 | GAD1 |
| CACNA1B | 5.90E-74 | 0.52686743 | 0.957 | 0.911 | 2.26E-69 | InN6 | CACNA1B |
| MGAT54 | 6.91E-74 | 0.82139135 | 0.788 | 0.66 | 2.64E-69 | InN6 | MGAT5 |
| ITGB83 | 3.44E-73 | 0.87240974 | 0.528 | 0.286 | 1.31E-68 | InN6 | ITGB8 |
| SLC29A4 | 4.37E-73 | 0.44354363 | 0.382 | 0.155 | 1.67E-68 | InN6 | SLC29A4 |
| GPR1582 | 7.63E-73 | 0.68276059 | 0.878 | 0.75 | 2.92E-68 | InN6 | GPR158 |
| ARHGAP283 | 1.94E-72 | 0.56325006 | 0.465 | 0.207 | 7.40E-68 | InN6 | ARHGAP28 |
| REV3L4 | 5.37E-72 | 0.57586092 | 0.876 | 0.777 | 2.05E-67 | InN6 | REV3L |
| GRAMD1B | 1.49E-71 | 0.68799535 | 0.807 | 0.668 | 5.68E-67 | InN6 | GRAMD1B |
| ANKRD30BL3 | 2.46E-71 | 0.66537015 | 0.854 | 0.707 | 9.40E-67 | InN6 | ANKRD30BL |
| SPATA131 | 3.29E-69 | 0.57720504 | 0.355 | 0.143 | 1.26E-64 | InN6 | SPATA13 |
| NTM2 | 2.04E-68 | 0.69073917 | 0.954 | 0.85 | 7.78E-64 | InN6 | NTM |

| | | | | | | | |
|---|---|---|---|---|---|---|---|
| LINC012372 | 5.97E-68 | 0.51356256 | 0.384 | 0.161 | 2.28E-63 | InN6 | LINC01237 |
| GNG42 | 6.35E-68 | 0.4266062 | 0.369 | 0.145 | 2.43E-63 | InN6 | GNG4 |
| CAMK2D2 | 1.89E-67 | 0.78763723 | 0.829 | 0.669 | 7.23E-63 | InN6 | CAMK2D |
| ANKRD502 | 7.51E-67 | 0.52745163 | 0.424 | 0.197 | 2.87E-62 | InN6 | ANKRD50 |
| PYGO1 | 2.07E-66 | 0.31682944 | 0.265 | 0.086 | 7.93E-62 | InN6 | PYGO1 |
| AP000721.2 | 6.72E-66 | 0.40905673 | 0.304 | 0.111 | 2.57E-61 | InN6 | AP000721.2 |
| SLC8A14 | 7.20E-66 | 0.64295887 | 0.963 | 0.862 | 2.75E-61 | InN6 | SLC8A1 |
| ABCA11 | 8.56E-66 | 0.4279617 | 0.335 | 0.128 | 3.27E-61 | InN6 | ABCA1 |
| PDE3A2 | 1.93E-65 | 0.74581637 | 0.342 | 0.135 | 7.38E-61 | InN6 | PDE3A |
| LRRC393 | 1.18E-64 | 0.5129435 | 0.377 | 0.163 | 4.51E-60 | InN6 | LRRC39 |
| NCOA11 | 4.79E-64 | 0.58979526 | 0.907 | 0.875 | 1.83E-59 | InN6 | NCOA1 |
| SOX2-OT3 | 5.95E-64 | 0.90340675 | 0.715 | 0.49 | 2.28E-59 | InN6 | SOX2-OT |
| ZNF6084 | 8.73E-64 | 0.54824249 | 0.616 | 0.356 | 3.34E-59 | InN6 | ZNF608 |
| U91319.11 | 1.17E-63 | 0.78562921 | 0.636 | 0.435 | 4.47E-59 | InN6 | U91319.1 |
| TRDMT1 | 1.77E-63 | 0.5287768 | 0.388 | 0.175 | 6.75E-59 | InN6 | TRDMT1 |
| PELI23 | 2.44E-63 | 0.68647941 | 0.519 | 0.284 | 9.32E-59 | InN6 | PELI2 |
| OSBPL83 | 3.17E-63 | 0.64382943 | 0.837 | 0.751 | 1.21E-58 | InN6 | OSBPL8 |
| KCNQ53 | 5.63E-63 | 0.77529718 | 0.988 | 0.904 | 2.15E-58 | InN6 | KCNQ5 |
| GLUL1 | 7.21E-63 | 0.42982163 | 0.343 | 0.139 | 2.75E-58 | InN6 | GLUL |
| FREM13 | 1.68E-62 | 0.83719968 | 0.438 | 0.221 | 6.43E-58 | InN6 | FREM1 |
| ARID1B1 | 2.56E-62 | 0.46903218 | 0.953 | 0.919 | 9.80E-58 | InN6 | ARID1B |
| EGLN3 | 7.67E-61 | 0.42139614 | 0.314 | 0.122 | 2.93E-56 | InN6 | EGLN3 |
| SLC2A132 | 7.90E-61 | 0.54923206 | 0.907 | 0.856 | 3.02E-56 | InN6 | SLC2A13 |
| SH3RF35 | 1.54E-60 | 0.58330061 | 0.836 | 0.68 | 5.90E-56 | InN6 | SH3RF3 |
| ADCY83 | 3.26E-60 | 0.50109285 | 0.552 | 0.287 | 1.25E-55 | InN6 | ADCY8 |
| APP1 | 2.46E-59 | 0.53097261 | 0.942 | 0.911 | 9.40E-55 | InN6 | APP |
| BICD14 | 7.13E-59 | 0.49544509 | 0.875 | 0.793 | 2.72E-54 | InN6 | BICD1 |
| SPOCK23 | 7.17E-59 | 0.64435547 | 0.636 | 0.434 | 2.74E-54 | InN6 | SPOCK2 |
| PBX4 | 8.90E-59 | 0.50022194 | 0.36 | 0.162 | 3.40E-54 | InN6 | PBX4 |
| SGK13 | 1.42E-58 | 0.64874572 | 0.696 | 0.507 | 5.42E-54 | InN6 | SGK1 |
| ARHGAP211 | 3.57E-58 | 0.51188914 | 0.883 | 0.839 | 1.36E-53 | InN6 | ARHGAP21 |
| VWA5A | 4.69E-58 | 0.29933634 | 0.268 | 0.095 | 1.79E-53 | InN6 | VWA5A |
| SLCO3A14 | 6.07E-58 | 0.64614673 | 0.83 | 0.735 | 2.32E-53 | InN6 | SLCO3A1 |
| PTN | 1.26E-57 | 0.45209327 | 0.298 | 0.118 | 4.81E-53 | InN6 | PTN |
| TRMT9B3 | 2.76E-57 | 0.57212672 | 0.653 | 0.417 | 1.06E-52 | InN6 | TRMT9B |
| NCAM12 | 2.82E-57 | 0.42497883 | 0.962 | 0.937 | 1.08E-52 | InN6 | NCAM1 |
| RAB37 | 3.57E-57 | 0.72399829 | 0.439 | 0.236 | 1.36E-52 | InN6 | RAB37 |
| LIMA12 | 3.60E-57 | 0.52254252 | 0.415 | 0.202 | 1.38E-52 | InN6 | LIMA1 |
| SLC1A22 | 7.81E-57 | 0.58034278 | 0.661 | 0.46 | 2.98E-52 | InN6 | SLC1A2 |
| PCBP32 | 9.37E-56 | 0.44944584 | 0.393 | 0.182 | 3.58E-51 | InN6 | PCBP3 |
| MGAT4C3 | 6.84E-55 | 0.48466725 | 0.848 | 0.577 | 2.62E-50 | InN6 | MGAT4C |
| FRMPD32 | 2.75E-53 | 0.35427916 | 0.283 | 0.109 | 1.05E-48 | InN6 | FRMPD3 |
| ATP1B24 | 5.30E-53 | 0.43947826 | 0.459 | 0.231 | 2.02E-48 | InN6 | ATP1B2 |
| MOB3B1 | 9.08E-53 | 0.78138266 | 0.577 | 0.391 | 3.47E-48 | InN6 | MOB3B |
| GRK33 | 1.70E-52 | 0.56940638 | 0.816 | 0.707 | 6.48E-48 | InN6 | GRK3 |
| DPF32 | 3.95E-52 | 0.56633056 | 0.507 | 0.283 | 1.51E-47 | InN6 | DPF3 |
| ANKRD291 | 8.96E-52 | 0.46507158 | 0.403 | 0.198 | 3.42E-47 | InN6 | ANKRD29 |
| ECE23 | 1.69E-51 | 0.49022508 | 0.439 | 0.229 | 6.44E-47 | InN6 | ECE2 |
| NECAB22 | 1.73E-51 | 0.42787416 | 0.369 | 0.176 | 6.61E-47 | InN6 | NECAB2 |
| FOXN34 | 3.47E-51 | 0.53314315 | 0.84 | 0.707 | 1.33E-46 | InN6 | FOXN3 |
| UBE2QL1 | 6.00E-51 | 0.58049401 | 0.549 | 0.356 | 2.29E-46 | InN6 | UBE2QL1 |
| PBX11 | 6.97E-51 | 0.73544586 | 0.892 | 0.89 | 2.66E-46 | InN6 | PBX1 |
| TRIM361 | 7.40E-51 | 0.47365445 | 0.414 | 0.213 | 2.83E-46 | InN6 | TRIM36 |
| AF279873.32 | 1.08E-50 | 0.50332906 | 0.335 | 0.148 | 4.11E-46 | InN6 | AF279873.3 |
| MITF4 | 4.10E-50 | 0.46292158 | 0.41 | 0.196 | 1.57E-45 | InN6 | MITF |
| EPHA64 | 7.49E-50 | 0.49377616 | 0.891 | 0.659 | 2.86E-45 | InN6 | EPHA6 |
| INPP4B5 | 3.29E-49 | 0.47882724 | 0.907 | 0.754 | 1.26E-44 | InN6 | INPP4B |
| DGKD4 | 7.22E-49 | 0.36343751 | 0.557 | 0.311 | 2.76E-44 | InN6 | DGKD |
| FLNB1 | 1.11E-48 | 0.53867771 | 0.625 | 0.452 | 4.25E-44 | InN6 | FLNB |
| ZNF7043 | 1.26E-48 | 0.63421434 | 0.794 | 0.671 | 4.83E-44 | InN6 | ZNF704 |
| WWOX4 | 1.81E-48 | 0.56132123 | 0.903 | 0.817 | 6.91E-44 | InN6 | WWOX |
| ADCYAP1R15 | 2.16E-48 | 0.4426859 | 0.531 | 0.301 | 8.25E-44 | InN6 | ADCYAP1R1 |
| HES4 | 5.20E-48 | 0.39572761 | 0.314 | 0.14 | 1.99E-43 | InN6 | HES4 |

| | | | | | | | |
|---|---|---|---|---|---|---|---|
| MAN1A12 | 1.49E-47 | 0.65563083 | 0.683 | 0.489 | 5.70E-43 | InN6 | MAN1A1 |
| RXRA3 | 1.90E-47 | 0.38252368 | 0.411 | 0.208 | 7.27E-43 | InN6 | RXRA |
| PODXL24 | 2.07E-47 | 0.50062553 | 0.587 | 0.391 | 7.91E-43 | InN6 | PODXL2 |
| SNCAIP3 | 3.54E-47 | 0.49706527 | 0.343 | 0.161 | 1.35E-42 | InN6 | SNCAIP |
| TOX24 | 3.96E-47 | 0.4226092 | 0.413 | 0.205 | 1.51E-42 | InN6 | TOX2 |
| AL445623.23 | 5.47E-47 | 0.54873108 | 0.589 | 0.376 | 2.09E-42 | InN6 | AL445623.2 |
| GRIA13 | 2.43E-46 | 0.45045543 | 0.954 | 0.922 | 9.29E-42 | InN6 | GRIA1 |
| CNTLN1 | 3.09E-46 | 0.4938507 | 0.426 | 0.234 | 1.18E-41 | InN6 | CNTLN |
| RAB8B1 | 8.03E-46 | 0.48628844 | 0.484 | 0.29 | 3.07E-41 | InN6 | RAB8B |
| GABRG2 | 1.31E-45 | 0.54424595 | 0.821 | 0.744 | 5.02E-41 | InN6 | GABRG2 |
| GPR137C3 | 1.93E-45 | 0.54758127 | 0.735 | 0.606 | 7.38E-41 | InN6 | GPR137C |
| GNAS3 | 3.57E-45 | 0.54824841 | 0.883 | 0.824 | 1.36E-40 | InN6 | GNAS |
| NKAIN25 | 4.25E-45 | 0.66157382 | 0.949 | 0.839 | 1.62E-40 | InN6 | NKAIN2 |
| BMERB12 | 2.57E-44 | 0.58956999 | 0.804 | 0.724 | 9.82E-40 | InN6 | BMERB1 |
| FRMD33 | 3.38E-44 | 0.50147351 | 0.548 | 0.342 | 1.29E-39 | InN6 | FRMD3 |
| GRID14 | 6.17E-44 | 0.49375817 | 0.892 | 0.852 | 2.36E-39 | InN6 | GRID1 |
| SGO1-AS12 | 6.36E-44 | 0.76772129 | 0.3 | 0.136 | 2.43E-39 | InN6 | SGO1-AS1 |
| LARP11 | 8.86E-44 | 0.50070519 | 0.769 | 0.669 | 3.39E-39 | InN6 | LARP1 |
| AKAP62 | 3.63E-43 | 0.33760344 | 0.97 | 0.944 | 1.39E-38 | InN6 | AKAP6 |
| LAMA31 | 4.94E-43 | 0.3998787 | 0.302 | 0.132 | 1.89E-38 | InN6 | LAMA3 |
| HIVEP34 | 5.81E-43 | 0.41034659 | 0.898 | 0.862 | 2.22E-38 | InN6 | HIVEP3 |
| GPR1552 | 6.17E-43 | 0.56363381 | 0.716 | 0.6 | 2.36E-38 | InN6 | GPR155 |
| NKAIN33 | 7.90E-43 | 0.57077951 | 0.485 | 0.277 | 3.02E-38 | InN6 | NKAIN3 |
| CCNH3 | 1.12E-42 | 0.50940667 | 0.844 | 0.768 | 4.29E-38 | InN6 | CCNH |
| PREX23 | 1.60E-42 | 0.37329106 | 0.375 | 0.178 | 6.13E-38 | InN6 | PREX2 |
| C8orf343 | 2.13E-42 | 0.66948287 | 0.594 | 0.391 | 8.14E-38 | InN6 | C8orf34 |
| RNF1301 | 2.26E-42 | 0.49706213 | 0.8 | 0.728 | 8.63E-38 | InN6 | RNF130 |
| GRIA21 | 2.33E-42 | 0.41594192 | 0.963 | 0.95 | 8.91E-38 | InN6 | GRIA2 |
| ARHGAP63 | 2.41E-42 | 0.84768223 | 0.339 | 0.175 | 9.20E-38 | InN6 | ARHGAP6 |
| KCNQ1OT11 | 3.29E-42 | 0.60531358 | 0.65 | 0.507 | 1.26E-37 | InN6 | KCNQ1OT1 |
| ANK32 | 7.71E-42 | 0.29532631 | 0.986 | 0.976 | 2.95E-37 | InN6 | ANK3 |
| NR2F12 | 8.47E-42 | 0.52309872 | 0.578 | 0.404 | 3.24E-37 | InN6 | NR2F1 |
| GRIN3A3 | 1.01E-41 | 0.34698907 | 0.459 | 0.237 | 3.85E-37 | InN6 | GRIN3A |
| NACC21 | 2.27E-41 | 0.25681959 | 0.268 | 0.113 | 8.67E-37 | InN6 | NACC2 |
| GRAMD1C | 3.63E-41 | 0.46632926 | 0.293 | 0.141 | 1.39E-36 | InN6 | GRAMD1C |
| MAF5 | 1.59E-40 | 0.28042135 | 0.388 | 0.186 | 6.07E-36 | InN6 | MAF |
| LRRC4C5 | 6.94E-40 | 0.25912426 | 0.967 | 0.889 | 2.65E-35 | InN6 | LRRC4C |
| ADD32 | 1.13E-39 | 0.47608079 | 0.711 | 0.581 | 4.31E-35 | InN6 | ADD3 |
| MTUS15 | 1.59E-39 | 0.39977518 | 0.509 | 0.298 | 6.10E-35 | InN6 | MTUS1 |
| EPHA55 | 4.37E-39 | 0.77202262 | 0.72 | 0.599 | 1.67E-34 | InN6 | EPHA5 |
| GAS72 | 7.96E-39 | 0.47637237 | 0.846 | 0.78 | 3.04E-34 | InN6 | GAS7 |
| NAAA2 | 2.84E-38 | 0.40378377 | 0.384 | 0.21 | 1.09E-33 | InN6 | NAAA |
| DSCAM4 | 2.95E-38 | 0.34702038 | 0.98 | 0.904 | 1.13E-33 | InN6 | DSCAM |
| TOX35 | 3.04E-38 | 0.38024176 | 0.381 | 0.189 | 1.16E-33 | InN6 | TOX3 |
| SLC7A142 | 5.50E-38 | 0.52492792 | 0.644 | 0.514 | 2.10E-33 | InN6 | SLC7A14 |
| MAP3K12 | 7.55E-38 | 0.35825724 | 0.327 | 0.163 | 2.89E-33 | InN6 | MAP3K1 |
| LPIN24 | 8.15E-38 | 0.4826787 | 0.534 | 0.363 | 3.11E-33 | InN6 | LPIN2 |
| POT1-AS12 | 8.65E-38 | 0.57953201 | 0.573 | 0.394 | 3.31E-33 | InN6 | POT1-AS1 |
| CRTAC14 | 1.07E-37 | 0.4030947 | 0.426 | 0.238 | 4.08E-33 | InN6 | CRTAC1 |
| HIP12 | 2.11E-37 | 0.31573147 | 0.367 | 0.188 | 8.06E-33 | InN6 | HIP1 |
| TGFBR33 | 2.35E-37 | 0.39785387 | 0.363 | 0.19 | 8.98E-33 | InN6 | TGFBR3 |
| AC079296.14 | 9.68E-37 | 0.30507786 | 0.363 | 0.184 | 3.70E-32 | InN6 | AC079296.1 |
| NYAP23 | 1.22E-36 | 0.46323473 | 0.673 | 0.509 | 4.68E-32 | InN6 | NYAP2 |
| SAMD4A3 | 1.44E-36 | 0.53158306 | 0.628 | 0.469 | 5.51E-32 | InN6 | SAMD4A |
| HTRA13 | 2.49E-36 | 0.29638174 | 0.336 | 0.165 | 9.54E-32 | InN6 | HTRA1 |
| INTS7 | 3.04E-36 | 0.45839683 | 0.393 | 0.227 | 1.16E-31 | InN6 | INTS7 |
| FAM13C2 | 3.28E-36 | 0.49874982 | 0.645 | 0.511 | 1.25E-31 | InN6 | FAM13C |
| UBASH3B5 | 3.88E-36 | 0.30800537 | 0.556 | 0.34 | 1.48E-31 | InN6 | UBASH3B |
| FXYD64 | 5.60E-36 | 0.54464343 | 0.704 | 0.619 | 2.14E-31 | InN6 | FXYD6 |
| AC092640.11 | 9.07E-36 | 0.39030049 | 0.269 | 0.123 | 3.47E-31 | InN6 | AC092640.1 |
| SNCA4 | 1.55E-35 | 0.54749506 | 0.748 | 0.612 | 5.92E-31 | InN6 | SNCA |
| TTC282 | 1.89E-35 | 0.40564383 | 0.858 | 0.788 | 7.23E-31 | InN6 | TTC28 |
| LRCH21 | 2.42E-35 | 0.47237981 | 0.507 | 0.339 | 9.26E-31 | InN6 | LRCH2 |

| | | | | | | | |
|---|---|---|---|---|---|---|---|
| FCHSD26 | 3.06E-35 | 0.43384878 | 0.74 | 0.614 | 1.17E-30 | InN6 | FCHSD2 |
| RASGRF11 | 3.52E-35 | 0.43310999 | 0.648 | 0.512 | 1.34E-30 | InN6 | RASGRF1 |
| SYN33 | 3.86E-35 | 0.39196209 | 0.967 | 0.865 | 1.47E-30 | InN6 | SYN3 |
| ARL4C4 | 4.17E-35 | 0.38166759 | 0.457 | 0.27 | 1.59E-30 | InN6 | ARL4C |
| DHRSX | 4.68E-35 | 0.49666443 | 0.574 | 0.416 | 1.79E-30 | InN6 | DHRSX |
| FAM107A | 1.29E-34 | 0.34442999 | 0.272 | 0.128 | 4.93E-30 | InN6 | FAM107A |
| KCNC24 | 1.50E-34 | 0.26049583 | 0.925 | 0.835 | 5.75E-30 | InN6 | KCNC2 |
| NSG22 | 2.43E-34 | 0.49174334 | 0.69 | 0.556 | 9.28E-30 | InN6 | NSG2 |
| NR2F1-AS12 | 2.64E-34 | 0.46375638 | 0.859 | 0.788 | 1.01E-29 | InN6 | NR2F1-AS1 |
| GPR161 | 3.41E-34 | 0.26919246 | 0.252 | 0.114 | 1.30E-29 | InN6 | GPR161 |
| IRF2BP21 | 4.15E-34 | 0.40536342 | 0.322 | 0.17 | 1.59E-29 | InN6 | IRF2BP2 |
| SIPA1L22 | 5.23E-34 | 0.520545 | 0.572 | 0.427 | 2.00E-29 | InN6 | SIPA1L2 |
| KCND22 | 5.77E-34 | 0.4559898 | 0.946 | 0.872 | 2.20E-29 | InN6 | KCND2 |
| CFAP1614 | 1.26E-33 | 0.4995433 | 0.491 | 0.316 | 4.83E-29 | InN6 | CFAP161 |
| MACROD11 | 1.54E-33 | 0.38123031 | 0.377 | 0.217 | 5.88E-29 | InN6 | MACROD1 |
| LIFR1 | 2.19E-33 | 0.45694502 | 0.472 | 0.315 | 8.36E-29 | InN6 | LIFR |
| CNTFR | 2.50E-33 | 0.31269377 | 0.307 | 0.157 | 9.55E-29 | InN6 | CNTFR |
| AFDN2 | 2.64E-33 | 0.39719489 | 0.752 | 0.65 | 1.01E-28 | InN6 | AFDN |
| CPEB22 | 3.51E-33 | 0.47204533 | 0.58 | 0.44 | 1.34E-28 | InN6 | CPEB2 |
| LYPD6B5 | 5.49E-33 | 0.3791354 | 0.347 | 0.18 | 2.10E-28 | InN6 | LYPD6B |
| WASF34 | 9.78E-33 | 0.4416475 | 0.541 | 0.383 | 3.74E-28 | InN6 | WASF3 |
| LPGAT11 | 1.14E-32 | 0.60106498 | 0.647 | 0.546 | 4.37E-28 | InN6 | LPGAT1 |
| ERBIN4 | 1.22E-32 | 0.42108981 | 0.675 | 0.538 | 4.68E-28 | InN6 | ERBIN |
| USP35 | 1.41E-32 | 0.31381882 | 0.336 | 0.175 | 5.40E-28 | InN6 | USP3 |
| HS6ST21 | 1.55E-32 | 0.45728686 | 0.375 | 0.218 | 5.91E-28 | InN6 | HS6ST2 |
| KCND31 | 2.03E-32 | 0.40075562 | 0.895 | 0.827 | 7.76E-28 | InN6 | KCND3 |
| OSBPL103 | 2.68E-32 | 0.47706203 | 0.628 | 0.484 | 1.02E-27 | InN6 | OSBPL10 |
| KIDINS2202 | 4.62E-32 | 0.41365214 | 0.777 | 0.739 | 1.77E-27 | InN6 | KIDINS220 |
| KIAA19584 | 5.74E-32 | 0.45020727 | 0.595 | 0.431 | 2.19E-27 | InN6 | KIAA1958 |
| GABRA21 | 8.08E-32 | 0.44284784 | 0.744 | 0.638 | 3.09E-27 | InN6 | GABRA2 |
| KCNQ36 | 8.15E-32 | 0.45534059 | 0.941 | 0.902 | 3.11E-27 | InN6 | KCNQ3 |
| DPYSL31 | 8.22E-32 | 0.38325829 | 0.351 | 0.196 | 3.14E-27 | InN6 | DPYSL3 |
| LTBP11 | 1.36E-31 | 0.30323065 | 0.264 | 0.124 | 5.22E-27 | InN6 | LTBP1 |
| RGMB1 | 1.78E-31 | 0.40361168 | 0.413 | 0.255 | 6.82E-27 | InN6 | RGMB |
| TENM34 | 4.39E-31 | 0.43604882 | 0.821 | 0.673 | 1.68E-26 | InN6 | TENM3 |
| HIPK22 | 1.04E-30 | 0.44764579 | 0.519 | 0.364 | 3.96E-26 | InN6 | HIPK2 |
| SLC9A93 | 1.17E-30 | 0.30615189 | 0.532 | 0.353 | 4.49E-26 | InN6 | SLC9A9 |
| CPEB32 | 2.08E-30 | 0.36991548 | 0.871 | 0.847 | 7.96E-26 | InN6 | CPEB3 |
| GPRC5B2 | 3.00E-30 | 0.29629801 | 0.325 | 0.171 | 1.15E-25 | InN6 | GPRC5B |
| GTF2I3 | 3.76E-30 | 0.3629678 | 0.871 | 0.834 | 1.44E-25 | InN6 | GTF2I |
| GALNT23 | 4.67E-30 | 0.48409885 | 0.606 | 0.481 | 1.78E-25 | InN6 | GALNT2 |
| AC106795.32 | 8.92E-30 | 0.3122484 | 0.382 | 0.22 | 3.41E-25 | InN6 | AC106795.3 |
| SNRPN5 | 1.01E-29 | 0.35876585 | 0.845 | 0.762 | 3.88E-25 | InN6 | SNRPN |
| ABCC13 | 1.40E-29 | 0.41172053 | 0.502 | 0.356 | 5.36E-25 | InN6 | ABCC1 |
| GLRA23 | 3.35E-29 | 0.33154245 | 0.381 | 0.211 | 1.28E-24 | InN6 | GLRA2 |
| PCSK1N2 | 4.28E-29 | 0.57037242 | 0.586 | 0.458 | 1.64E-24 | InN6 | PCSK1N |
| AC087457.12 | 4.77E-29 | 0.25679104 | 0.26 | 0.124 | 1.82E-24 | InN6 | AC087457.1 |
| SLC8A33 | 6.06E-29 | 0.43440194 | 0.631 | 0.487 | 2.32E-24 | InN6 | SLC8A3 |
| MYT1L3 | 8.66E-29 | 0.25021275 | 0.975 | 0.971 | 3.31E-24 | InN6 | MYT1L |
| NRSN12 | 1.05E-28 | 0.41484716 | 0.619 | 0.489 | 4.03E-24 | InN6 | NRSN1 |
| MXD43 | 1.13E-28 | 0.32907323 | 0.413 | 0.256 | 4.32E-24 | InN6 | MXD4 |
| DLG51 | 2.06E-28 | 0.42221957 | 0.461 | 0.312 | 7.89E-24 | InN6 | DLG5 |
| VSTM2A3 | 4.46E-28 | 0.41339396 | 0.548 | 0.394 | 1.70E-23 | InN6 | VSTM2A |
| SYT113 | 6.75E-28 | 0.43261958 | 0.731 | 0.646 | 2.58E-23 | InN6 | SYT11 |
| SYT91 | 9.05E-28 | 0.31913527 | 0.276 | 0.145 | 3.46E-23 | InN6 | SYT9 |
| AC007364.13 | 1.28E-27 | 0.42766954 | 0.435 | 0.281 | 4.89E-23 | InN6 | AC007364.1 |
| SESN31 | 2.45E-27 | 0.46192338 | 0.715 | 0.631 | 9.36E-23 | InN6 | SESN3 |
| PHYHIPL3 | 4.06E-27 | 0.43281087 | 0.683 | 0.593 | 1.55E-22 | InN6 | PHYHIPL |
| ADCY24 | 5.28E-27 | 0.38027705 | 0.887 | 0.854 | 2.02E-22 | InN6 | ADCY2 |
| CA31 | 6.54E-27 | 0.38085353 | 0.35 | 0.208 | 2.50E-22 | InN6 | CA3 |
| HDAC2-AS22 | 1.16E-26 | 0.35528115 | 0.422 | 0.269 | 4.44E-22 | InN6 | HDAC2-AS2 |
| ULK2 | 1.20E-26 | 0.43141762 | 0.635 | 0.547 | 4.59E-22 | InN6 | ULK2 |
| SERINC12 | 1.28E-26 | 0.50403327 | 0.686 | 0.62 | 4.91E-22 | InN6 | SERINC1 |

| | | | | | | | |
|---|---|---|---|---|---|---|---|
| SLC38A13 | 2.83E-26 | 0.46625558 | 0.678 | 0.598 | 1.08E-21 | InN6 | SLC38A1 |
| CPLX21 | 3.02E-26 | 0.42809147 | 0.664 | 0.573 | 1.15E-21 | InN6 | CPLX2 |
| PNPLA71 | 3.38E-26 | 0.39056696 | 0.548 | 0.414 | 1.29E-21 | InN6 | PNPLA7 |
| DAAM14 | 4.19E-26 | 0.38098495 | 0.79 | 0.751 | 1.60E-21 | InN6 | DAAM1 |
| SLC4A73 | 7.81E-26 | 0.38051275 | 0.58 | 0.447 | 2.98E-21 | InN6 | SLC4A7 |
| ATP1B14 | 1.08E-25 | 0.49420199 | 0.79 | 0.78 | 4.13E-21 | InN6 | ATP1B1 |
| PALM2-AKAF | 2.08E-25 | 0.42182748 | 0.763 | 0.704 | 7.93E-21 | InN6 | PALM2-AKAP |
| PDE8B2 | 2.22E-25 | 0.33173543 | 0.821 | 0.663 | 8.49E-21 | InN6 | PDE8B |
| ARHGAP53 | 7.63E-25 | 0.43526143 | 0.668 | 0.592 | 2.92E-20 | InN6 | ARHGAP5 |
| ADAM222 | 1.05E-24 | 0.31826999 | 0.848 | 0.826 | 4.00E-20 | InN6 | ADAM22 |
| LINC004612 | 1.21E-24 | 0.38546327 | 0.827 | 0.785 | 4.62E-20 | InN6 | LINC00461 |
| REEP12 | 2.29E-24 | 0.38896711 | 0.694 | 0.625 | 8.74E-20 | InN6 | REEP1 |
| TSPAN52 | 2.34E-24 | 0.3700905 | 0.892 | 0.844 | 8.93E-20 | InN6 | TSPAN5 |
| APC5 | 2.46E-24 | 0.35823603 | 0.777 | 0.741 | 9.40E-20 | InN6 | APC |
| RAVER21 | 3.58E-24 | 0.38886574 | 0.426 | 0.292 | 1.37E-19 | InN6 | RAVER2 |
| SUSD43 | 3.83E-24 | 0.41162159 | 0.691 | 0.611 | 1.46E-19 | InN6 | SUSD4 |
| RERE1 | 4.07E-24 | 0.25351767 | 0.945 | 0.925 | 1.55E-19 | InN6 | RERE |
| C1orf611 | 4.83E-24 | 0.36223429 | 0.457 | 0.329 | 1.85E-19 | InN6 | C1orf61 |
| GTF2IRD13 | 6.66E-24 | 0.36129839 | 0.583 | 0.455 | 2.55E-19 | InN6 | GTF2IRD1 |
| BACH16 | 6.73E-24 | 0.3420771 | 0.385 | 0.234 | 2.57E-19 | InN6 | BACH1 |
| FYN1 | 7.97E-24 | 0.36968629 | 0.779 | 0.748 | 3.05E-19 | InN6 | FYN |
| TCF202 | 1.61E-23 | 0.38997874 | 0.707 | 0.632 | 6.17E-19 | InN6 | TCF20 |
| JAK13 | 3.58E-23 | 0.40144354 | 0.787 | 0.744 | 1.37E-18 | InN6 | JAK1 |
| NNMT | 4.20E-23 | 0.33700162 | 0.25 | 0.134 | 1.60E-18 | InN6 | NNMT |
| AFAP14 | 6.33E-23 | 0.3312014 | 0.547 | 0.398 | 2.42E-18 | InN6 | AFAP1 |
| SRGAP25 | 9.67E-23 | 0.37143385 | 0.598 | 0.484 | 3.70E-18 | InN6 | SRGAP2 |
| TBC1D301 | 1.05E-22 | 0.39100827 | 0.426 | 0.3 | 4.03E-18 | InN6 | TBC1D30 |
| NRXN22 | 1.10E-22 | 0.38947484 | 0.699 | 0.636 | 4.21E-18 | InN6 | NRXN2 |
| ELFN21 | 1.23E-22 | 0.32792088 | 0.33 | 0.203 | 4.69E-18 | InN6 | ELFN2 |
| PCSK62 | 1.65E-22 | 0.42904846 | 0.342 | 0.218 | 6.30E-18 | InN6 | PCSK6 |
| NMNAT23 | 2.28E-22 | 0.30693108 | 0.88 | 0.853 | 8.72E-18 | InN6 | NMNAT2 |
| FOXO3 | 3.19E-22 | 0.37670572 | 0.746 | 0.691 | 1.22E-17 | InN6 | FOXO3 |
| MT-ND43 | 3.98E-22 | 0.45188097 | 0.879 | 0.856 | 1.52E-17 | InN6 | MT-ND4 |
| AC009093.22 | 4.57E-22 | 0.25118988 | 0.252 | 0.135 | 1.75E-17 | InN6 | AC009093.2 |
| COBL4 | 8.25E-22 | 0.3147212 | 0.857 | 0.75 | 3.15E-17 | InN6 | COBL |
| EPHA74 | 1.03E-21 | 0.52863582 | 0.641 | 0.545 | 3.95E-17 | InN6 | EPHA7 |
| B3GALNT2 | 1.36E-21 | 0.38031445 | 0.439 | 0.321 | 5.21E-17 | InN6 | B3GALNT2 |
| ABLIM16 | 2.13E-21 | 0.2887745 | 0.899 | 0.833 | 8.15E-17 | InN6 | ABLIM1 |
| ARL6IP12 | 2.36E-21 | 0.4424383 | 0.547 | 0.446 | 9.03E-17 | InN6 | ARL6IP1 |
| GABRB23 | 3.61E-21 | 0.26304304 | 0.941 | 0.922 | 1.38E-16 | InN6 | GABRB2 |
| GRK51 | 4.17E-21 | 0.34396987 | 0.41 | 0.281 | 1.60E-16 | InN6 | GRK5 |
| CCND2 | 5.56E-21 | 0.40943963 | 0.598 | 0.507 | 2.13E-16 | InN6 | CCND2 |
| MT-ND34 | 6.92E-21 | 0.50296174 | 0.891 | 0.87 | 2.65E-16 | InN6 | MT-ND3 |
| LAPTM4B | 7.82E-21 | 0.31460752 | 0.317 | 0.195 | 2.99E-16 | InN6 | LAPTM4B |
| SBF22 | 1.83E-20 | 0.27910814 | 0.841 | 0.801 | 7.01E-16 | InN6 | SBF2 |
| DNM21 | 2.10E-20 | 0.39274923 | 0.552 | 0.447 | 8.04E-16 | InN6 | DNM2 |
| DISC12 | 6.65E-20 | 0.27321759 | 0.251 | 0.139 | 2.54E-15 | InN6 | DISC1 |
| MT-CO24 | 8.32E-20 | 0.51165716 | 0.908 | 0.899 | 3.18E-15 | InN6 | MT-CO2 |
| ENAH2 | 8.70E-20 | 0.32925133 | 0.628 | 0.541 | 3.33E-15 | InN6 | ENAH |
| TAFA52 | 9.54E-20 | 0.41350109 | 0.674 | 0.606 | 3.65E-15 | InN6 | TAFA5 |
| N4BP23 | 1.65E-19 | 0.32321546 | 0.495 | 0.366 | 6.30E-15 | InN6 | N4BP2 |
| FRYL2 | 2.86E-19 | 0.32531669 | 0.756 | 0.704 | 1.09E-14 | InN6 | FRYL |
| NXN1 | 3.53E-19 | 0.29684178 | 0.263 | 0.151 | 1.35E-14 | InN6 | NXN |
| SHANK21 | 3.80E-19 | 0.30249263 | 0.862 | 0.815 | 1.45E-14 | InN6 | SHANK2 |
| MAST22 | 4.24E-19 | 0.34381054 | 0.507 | 0.408 | 1.62E-14 | InN6 | MAST2 |
| CAMK43 | 9.01E-19 | 0.38258919 | 0.781 | 0.721 | 3.44E-14 | InN6 | CAMK4 |
| CADM11 | 1.67E-18 | 0.30488302 | 0.896 | 0.86 | 6.37E-14 | InN6 | CADM1 |
| NAP1L32 | 1.83E-18 | 0.38019875 | 0.423 | 0.305 | 7.01E-14 | InN6 | NAP1L3 |
| RYBP1 | 1.88E-18 | 0.37349774 | 0.541 | 0.438 | 7.19E-14 | InN6 | RYBP |
| ZMIZ1 | 1.95E-18 | 0.3426258 | 0.495 | 0.384 | 7.46E-14 | InN6 | ZMIZ1 |
| PARM13 | 2.65E-18 | 0.41077702 | 0.511 | 0.399 | 1.01E-13 | InN6 | PARM1 |
| RAB3IP3 | 2.78E-18 | 0.27344331 | 0.406 | 0.276 | 1.06E-13 | InN6 | RAB3IP |
| ARNT23 | 3.00E-18 | 0.28049607 | 0.865 | 0.839 | 1.15E-13 | InN6 | ARNT2 |

| | | | | | | | |
|---|---|---|---|---|---|---|---|
| SEMA4B1 | 3.28E-18 | 0.35027468 | 0.459 | 0.353 | 1.25E-13 | InN6 | SEMA4B |
| ZNF4831 | 3.49E-18 | 0.33718576 | 0.779 | 0.75 | 1.34E-13 | InN6 | ZNF483 |
| COL12A14 | 3.75E-18 | 0.39056822 | 0.314 | 0.199 | 1.43E-13 | InN6 | COL12A1 |
| GPR1762 | 6.88E-18 | 0.3363166 | 0.448 | 0.334 | 2.63E-13 | InN6 | GPR176 |
| SMAD5 | 8.30E-18 | 0.34797196 | 0.402 | 0.296 | 3.17E-13 | InN6 | SMAD5 |
| MGLL4 | 8.60E-18 | 0.31456639 | 0.557 | 0.439 | 3.29E-13 | InN6 | MGLL |
| OSTM12 | 1.02E-17 | 0.31980257 | 0.375 | 0.263 | 3.91E-13 | InN6 | OSTM1 |
| MGST21 | 2.18E-17 | 0.25203163 | 0.314 | 0.202 | 8.35E-13 | InN6 | MGST2 |
| KMT2A | 5.67E-17 | 0.31359548 | 0.836 | 0.832 | 2.17E-12 | InN6 | KMT2A |
| LINC011974 | 8.13E-17 | 0.2889706 | 0.485 | 0.352 | 3.11E-12 | InN6 | LINC01197 |
| FNBP12 | 9.97E-17 | 0.33536754 | 0.689 | 0.636 | 3.81E-12 | InN6 | FNBP1 |
| PCDH155 | 1.67E-16 | 0.32257856 | 0.459 | 0.327 | 6.39E-12 | InN6 | PCDH15 |
| SLITRK41 | 1.74E-16 | 0.38718266 | 0.469 | 0.372 | 6.64E-12 | InN6 | SLITRK4 |
| ZNF5162 | 1.81E-16 | 0.33343379 | 0.447 | 0.33 | 6.91E-12 | InN6 | ZNF516 |
| WSB12 | 2.04E-16 | 0.33417392 | 0.668 | 0.625 | 7.80E-12 | InN6 | WSB1 |
| ANKRD64 | 2.26E-16 | 0.31161048 | 0.532 | 0.419 | 8.66E-12 | InN6 | ANKRD6 |
| NEAT13 | 2.55E-16 | 0.57171279 | 0.501 | 0.383 | 9.75E-12 | InN6 | NEAT1 |
| STAT5B2 | 2.92E-16 | 0.30586499 | 0.457 | 0.353 | 1.11E-11 | InN6 | STAT5B |
| GSE13 | 3.60E-16 | 0.33149145 | 0.608 | 0.502 | 1.38E-11 | InN6 | GSE1 |
| HSP90AB12 | 3.71E-16 | 0.37479883 | 0.756 | 0.727 | 1.42E-11 | InN6 | HSP90AB1 |
| TNKS1 | 3.85E-16 | 0.28278586 | 0.819 | 0.816 | 1.47E-11 | InN6 | TNKS |
| TRAF3IP2-AS | 4.27E-16 | 0.31235674 | 0.782 | 0.75 | 1.63E-11 | InN6 | TRAF3IP2-AS |
| MT-CO13 | 4.36E-16 | 0.45741822 | 0.899 | 0.868 | 1.67E-11 | InN6 | MT-CO1 |
| HOMER22 | 4.42E-16 | 0.33395455 | 0.443 | 0.345 | 1.69E-11 | InN6 | HOMER2 |
| SEMA5A3 | 4.44E-16 | 0.43173666 | 0.284 | 0.176 | 1.70E-11 | InN6 | SEMA5A |
| CSAD | 4.51E-16 | 0.33945111 | 0.52 | 0.433 | 1.73E-11 | InN6 | CSAD |
| C14orf1321 | 4.58E-16 | 0.3254059 | 0.451 | 0.341 | 1.75E-11 | InN6 | C14orf132 |
| PGRMC11 | 4.95E-16 | 0.30314377 | 0.442 | 0.339 | 1.89E-11 | InN6 | PGRMC1 |
| SEM13 | 5.40E-16 | 0.37475632 | 0.57 | 0.482 | 2.06E-11 | InN6 | SEM1 |
| ANKH2 | 5.97E-16 | 0.25601345 | 0.817 | 0.788 | 2.28E-11 | InN6 | ANKH |
| HACD32 | 7.40E-16 | 0.30341198 | 0.586 | 0.509 | 2.83E-11 | InN6 | HACD3 |
| DIP2B2 | 7.61E-16 | 0.30365765 | 0.74 | 0.691 | 2.91E-11 | InN6 | DIP2B |
| AL589740.12 | 9.59E-16 | 0.44201183 | 0.536 | 0.439 | 3.67E-11 | InN6 | AL589740.1 |
| DGKH2 | 1.05E-15 | 0.36845594 | 0.481 | 0.384 | 4.00E-11 | InN6 | DGKH |
| ARSB | 1.08E-15 | 0.29244337 | 0.455 | 0.349 | 4.13E-11 | InN6 | ARSB |
| CANX2 | 1.09E-15 | 0.3262284 | 0.573 | 0.493 | 4.18E-11 | InN6 | CANX |
| ABAT3 | 1.16E-15 | 0.27462113 | 0.594 | 0.503 | 4.42E-11 | InN6 | ABAT |
| AGPAT42 | 1.43E-15 | 0.30597601 | 0.741 | 0.705 | 5.46E-11 | InN6 | AGPAT4 |
| CDH203 | 1.53E-15 | 0.2517745 | 0.524 | 0.383 | 5.84E-11 | InN6 | CDH20 |
| PPFIBP14 | 2.52E-15 | 0.33380973 | 0.752 | 0.663 | 9.64E-11 | InN6 | PPFIBP1 |
| TBL1XR1 | 5.56E-15 | 0.27292938 | 0.795 | 0.762 | 2.12E-10 | InN6 | TBL1XR1 |
| APBB23 | 6.66E-15 | 0.28334343 | 0.733 | 0.691 | 2.54E-10 | InN6 | APBB2 |
| TACC24 | 8.76E-15 | 0.30074956 | 0.506 | 0.413 | 3.35E-10 | InN6 | TACC2 |
| AK32 | 8.82E-15 | 0.25381803 | 0.389 | 0.281 | 3.37E-10 | InN6 | AK3 |
| ELAVL31 | 1.08E-14 | 0.29479636 | 0.798 | 0.775 | 4.12E-10 | InN6 | ELAVL3 |
| PRKG22 | 1.14E-14 | 0.29072488 | 0.275 | 0.178 | 4.34E-10 | InN6 | PRKG2 |
| RRAS21 | 1.41E-14 | 0.29011817 | 0.398 | 0.296 | 5.41E-10 | InN6 | RRAS2 |
| FOXG12 | 2.07E-14 | 0.29219522 | 0.386 | 0.288 | 7.90E-10 | InN6 | FOXG1 |
| PIK3C2A1 | 2.77E-14 | 0.27168117 | 0.555 | 0.47 | 1.06E-09 | InN6 | PIK3C2A |
| CA131 | 3.07E-14 | 0.25410616 | 0.259 | 0.167 | 1.17E-09 | InN6 | CA13 |
| NXPE3 | 3.63E-14 | 0.2594943 | 0.377 | 0.28 | 1.39E-09 | InN6 | NXPE3 |
| PAG13 | 4.08E-14 | 0.29781044 | 0.473 | 0.375 | 1.56E-09 | InN6 | PAG1 |
| PGAP12 | 4.67E-14 | 0.34694068 | 0.532 | 0.456 | 1.78E-09 | InN6 | PGAP1 |
| ANP32A1 | 5.22E-14 | 0.33068877 | 0.405 | 0.306 | 2.00E-09 | InN6 | ANP32A |
| WWP21 | 7.96E-14 | 0.30908828 | 0.407 | 0.311 | 3.04E-09 | InN6 | WWP2 |
| RPS6KA31 | 8.77E-14 | 0.32115092 | 0.643 | 0.595 | 3.35E-09 | InN6 | RPS6KA3 |
| GLDN4 | 9.36E-14 | 0.26394726 | 0.339 | 0.237 | 3.58E-09 | InN6 | GLDN |
| TXNDC162 | 1.01E-13 | 0.28533349 | 0.603 | 0.545 | 3.87E-09 | InN6 | TXNDC16 |
| PHF141 | 1.54E-13 | 0.27774668 | 0.788 | 0.78 | 5.89E-09 | InN6 | PHF14 |
| RANBP173 | 1.66E-13 | 0.2657314 | 0.583 | 0.472 | 6.33E-09 | InN6 | RANBP17 |
| WDR11-AS1: | 1.76E-13 | 0.29597086 | 0.39 | 0.293 | 6.73E-09 | InN6 | WDR11-AS1 |
| RAB30 | 1.80E-13 | 0.32656328 | 0.432 | 0.35 | 6.88E-09 | InN6 | RAB30 |
| MT-ND13 | 2.45E-13 | 0.38246382 | 0.821 | 0.782 | 9.38E-09 | InN6 | MT-ND1 |

| | | | | | | | |
|---|---|---|---|---|---|---|---|
| EPDR1 | 3.39E-13 | 0.25560721 | 0.311 | 0.223 | 1.30E-08 | InN6 | EPDR1 |
| LUZP24 | 3.80E-13 | 0.31591247 | 0.438 | 0.311 | 1.45E-08 | InN6 | LUZP2 |
| TMOD11 | 4.05E-13 | 0.31208276 | 0.562 | 0.486 | 1.55E-08 | InN6 | TMOD1 |
| CAPZB2 | 4.28E-13 | 0.32510409 | 0.658 | 0.637 | 1.64E-08 | InN6 | CAPZB |
| JAM22 | 4.53E-13 | 0.26450292 | 0.256 | 0.169 | 1.73E-08 | InN6 | JAM2 |
| NFE2L1 | 5.74E-13 | 0.29652303 | 0.43 | 0.348 | 2.19E-08 | InN6 | NFE2L1 |
| RAP1GDS12 | 6.58E-13 | 0.28444052 | 0.664 | 0.622 | 2.51E-08 | InN6 | RAP1GDS1 |
| DNAH145 | 6.95E-13 | 0.28596151 | 0.403 | 0.299 | 2.66E-08 | InN6 | DNAH14 |
| TET24 | 7.07E-13 | 0.25247371 | 0.526 | 0.438 | 2.70E-08 | InN6 | TET2 |
| HSP90AA14 | 7.55E-13 | 0.47711113 | 0.735 | 0.712 | 2.89E-08 | InN6 | HSP90AA1 |
| SEC14L11 | 1.44E-12 | 0.30852946 | 0.47 | 0.389 | 5.51E-08 | InN6 | SEC14L1 |
| TBC1D9 | 1.71E-12 | 0.26404869 | 0.499 | 0.417 | 6.54E-08 | InN6 | TBC1D9 |
| DKK32 | 1.83E-12 | 0.26934632 | 0.677 | 0.635 | 6.98E-08 | InN6 | DKK3 |
| PRNP1 | 1.83E-12 | 0.32109623 | 0.685 | 0.661 | 6.98E-08 | InN6 | PRNP |
| ERO1A | 1.83E-12 | 0.29901862 | 0.435 | 0.352 | 7.01E-08 | InN6 | ERO1A |
| MTRNR2L12 | 2.43E-12 | 0.50937354 | 0.787 | 0.748 | 9.29E-08 | InN6 | MTRNR2L12 |
| FAM49A1 | 5.61E-12 | 0.3113158 | 0.728 | 0.717 | 2.14E-07 | InN6 | FAM49A |
| FAM214A2 | 5.63E-12 | 0.30592441 | 0.593 | 0.541 | 2.15E-07 | InN6 | FAM214A |
| ELP4 | 5.77E-12 | 0.3312528 | 0.587 | 0.542 | 2.21E-07 | InN6 | ELP4 |
| PCLO5 | 7.72E-12 | 0.32149815 | 0.915 | 0.924 | 2.95E-07 | InN6 | PCLO |
| UBA6-AS12 | 1.03E-11 | 0.31458576 | 0.661 | 0.605 | 3.94E-07 | InN6 | UBA6-AS1 |
| CD813 | 1.13E-11 | 0.25129057 | 0.334 | 0.243 | 4.33E-07 | InN6 | CD81 |
| MTMR71 | 1.25E-11 | 0.29151772 | 0.648 | 0.616 | 4.78E-07 | InN6 | MTMR7 |
| ATP1A33 | 1.29E-11 | 0.34486434 | 0.578 | 0.538 | 4.92E-07 | InN6 | ATP1A3 |
| AC098829.11 | 1.29E-11 | 0.26166613 | 0.336 | 0.241 | 4.94E-07 | InN6 | AC098829.1 |
| KLC11 | 1.31E-11 | 0.2812124 | 0.694 | 0.659 | 5.00E-07 | InN6 | KLC1 |
| TMEM50B1 | 2.09E-11 | 0.25007677 | 0.393 | 0.303 | 7.99E-07 | InN6 | TMEM50B |
| MT-CO33 | 2.34E-11 | 0.46775646 | 0.838 | 0.85 | 8.95E-07 | InN6 | MT-CO3 |
| SPATA6L | 2.35E-11 | 0.26023056 | 0.485 | 0.402 | 8.99E-07 | InN6 | SPATA6L |
| ZEB26 | 2.41E-11 | 0.2826799 | 0.812 | 0.823 | 9.21E-07 | InN6 | ZEB2 |
| RPL283 | 2.64E-11 | 0.33306727 | 0.445 | 0.368 | 1.01E-06 | InN6 | RPL28 |
| HIPK11 | 2.66E-11 | 0.28153355 | 0.368 | 0.283 | 1.02E-06 | InN6 | HIPK1 |
| TUBA1A1 | 2.99E-11 | 0.35091664 | 0.401 | 0.323 | 1.14E-06 | InN6 | TUBA1A |
| MT-ND4L3 | 3.66E-11 | 0.34426925 | 0.611 | 0.546 | 1.40E-06 | InN6 | MT-ND4L |
| RALGPS13 | 3.84E-11 | 0.27102948 | 0.716 | 0.708 | 1.47E-06 | InN6 | RALGPS1 |
| TMEFF26 | 3.88E-11 | 0.38027498 | 0.794 | 0.765 | 1.48E-06 | InN6 | TMEFF2 |
| TRIM242 | 3.97E-11 | 0.28095982 | 0.464 | 0.389 | 1.52E-06 | InN6 | TRIM24 |
| SARAF1 | 4.72E-11 | 0.29572462 | 0.614 | 0.575 | 1.80E-06 | InN6 | SARAF |
| ATCAY1 | 5.53E-11 | 0.28350373 | 0.639 | 0.599 | 2.11E-06 | InN6 | ATCAY |
| SPANXA2-OT | 5.56E-11 | 0.45331644 | 0.344 | 0.265 | 2.13E-06 | InN6 | SPANXA2-OT |
| KIF1A2 | 6.53E-11 | 0.27409478 | 0.664 | 0.637 | 2.50E-06 | InN6 | KIF1A |
| PLXNA21 | 6.80E-11 | 0.26924466 | 0.717 | 0.674 | 2.60E-06 | InN6 | PLXNA2 |
| RIMBP22 | 6.90E-11 | 0.25283435 | 0.786 | 0.76 | 2.64E-06 | InN6 | RIMBP2 |
| LRRC8A | 7.53E-11 | 0.25558206 | 0.264 | 0.186 | 2.88E-06 | InN6 | LRRC8A |
| SV2A2 | 9.56E-11 | 0.28360513 | 0.619 | 0.592 | 3.65E-06 | InN6 | SV2A |
| ARRB1 | 1.06E-10 | 0.25222822 | 0.505 | 0.432 | 4.04E-06 | InN6 | ARRB1 |
| AC002460.23 | 1.31E-10 | 0.39387289 | 0.506 | 0.425 | 5.00E-06 | InN6 | AC002460.2 |
| GFRA2 | 1.64E-10 | 0.26819151 | 0.327 | 0.247 | 6.28E-06 | InN6 | GFRA2 |
| CASK1 | 1.69E-10 | 0.26036535 | 0.674 | 0.633 | 6.46E-06 | InN6 | CASK |
| NBL11 | 1.70E-10 | 0.34904951 | 0.28 | 0.204 | 6.50E-06 | InN6 | NBL1 |
| MIR646HG3 | 2.03E-10 | 0.78559973 | 0.338 | 0.279 | 7.78E-06 | InN6 | MIR646HG |
| TNRC181 | 2.56E-10 | 0.25379176 | 0.629 | 0.589 | 9.80E-06 | InN6 | TNRC18 |
| TLE5 | 2.87E-10 | 0.30226017 | 0.482 | 0.421 | 1.10E-05 | InN6 | TLE5 |
| EPB41L31 | 2.94E-10 | 0.25857838 | 0.82 | 0.782 | 1.12E-05 | InN6 | EPB41L3 |
| CDR22 | 3.36E-10 | 0.30024896 | 0.612 | 0.557 | 1.29E-05 | InN6 | CDR2 |
| SPINT21 | 3.48E-10 | 0.28484681 | 0.351 | 0.279 | 1.33E-05 | InN6 | SPINT2 |
| SLC1A12 | 3.48E-10 | 0.29073382 | 0.46 | 0.394 | 1.33E-05 | InN6 | SLC1A1 |
| CRADD3 | 4.59E-10 | 0.33656175 | 0.511 | 0.435 | 1.76E-05 | InN6 | CRADD |
| VLDLR-AS11 | 6.18E-10 | 0.31077542 | 0.436 | 0.356 | 2.36E-05 | InN6 | VLDLR-AS1 |
| PVT11 | 8.00E-10 | 0.30224571 | 0.527 | 0.444 | 3.06E-05 | InN6 | PVT1 |
| RDX3 | 9.28E-10 | 0.26662114 | 0.472 | 0.405 | 3.55E-05 | InN6 | RDX |
| DCBLD2 | 9.97E-10 | 0.25818525 | 0.443 | 0.378 | 3.81E-05 | InN6 | DCBLD2 |
| EPN22 | 1.01E-09 | 0.26019248 | 0.603 | 0.556 | 3.87E-05 | InN6 | EPN2 |

| | | | | | | | |
|---|---|---|---|---|---|---|---|
| PLPPR51 | 1.11E-09 | 0.2728856 | 0.558 | 0.504 | 4.25E-05 | InN6 | PLPPR5 |
| ST8SIA52 | 1.19E-09 | 0.27325755 | 0.656 | 0.624 | 4.56E-05 | InN6 | ST8SIA5 |
| GALNT83 | 1.44E-09 | 0.25179941 | 0.38 | 0.303 | 5.50E-05 | InN6 | GALNT8 |
| EFNB21 | 1.91E-09 | 0.27495579 | 0.355 | 0.284 | 7.31E-05 | InN6 | EFNB2 |
| NLGN4X2 | 2.48E-09 | 0.27015091 | 0.74 | 0.73 | 9.48E-05 | InN6 | NLGN4X |
| LGR41 | 4.66E-09 | 0.27457524 | 0.372 | 0.299 | 0.00017824 | InN6 | LGR4 |
| OIP5-AS13 | 5.29E-09 | 0.2832509 | 0.611 | 0.572 | 0.00020204 | InN6 | OIP5-AS1 |
| KITLG2 | 5.66E-09 | 0.31205366 | 0.367 | 0.294 | 0.00021623 | InN6 | KITLG |
| GUCY1B1 | 6.95E-09 | 0.29047689 | 0.56 | 0.518 | 0.00026559 | InN6 | GUCY1B1 |
| URI13 | 9.65E-09 | 0.25345302 | 0.523 | 0.469 | 0.00036876 | InN6 | URI1 |
| LMCD1-AS13 | 1.03E-08 | 0.30826558 | 0.367 | 0.291 | 0.00039315 | InN6 | LMCD1-AS1 |
| MT-ATP64 | 1.12E-08 | 0.2922407 | 0.861 | 0.88 | 0.00042751 | InN6 | MT-ATP6 |
| RNF1501 | 1.93E-08 | 0.26522768 | 0.685 | 0.661 | 0.00073807 | InN6 | RNF150 |
| CLVS22 | 1.96E-08 | 0.26549575 | 0.636 | 0.599 | 0.00074959 | InN6 | CLVS2 |
| DNAJB21 | 2.20E-08 | 0.27911064 | 0.486 | 0.43 | 0.00083998 | InN6 | DNAJB2 |
| ANKRD30B3 | 2.20E-08 | 0.33867888 | 0.347 | 0.28 | 0.00084002 | InN6 | ANKRD30B |
| HIBCH1 | 3.27E-08 | 0.25845157 | 0.654 | 0.619 | 0.00125038 | InN6 | HIBCH |
| STIM11 | 3.58E-08 | 0.25622147 | 0.553 | 0.512 | 0.00136906 | InN6 | STIM1 |
| APLP21 | 4.08E-08 | 0.29573237 | 0.607 | 0.596 | 0.00156079 | InN6 | APLP2 |
| MT-CYB4 | 4.16E-08 | 0.27152737 | 0.811 | 0.787 | 0.00159196 | InN6 | MT-CYB |
| GRB10 | 4.46E-08 | 0.25193509 | 0.618 | 0.578 | 0.00170589 | InN6 | GRB10 |
| USP112 | 4.47E-08 | 0.25430489 | 0.572 | 0.544 | 0.0017094 | InN6 | USP11 |
| UBB1 | 4.71E-08 | 0.33749745 | 0.406 | 0.339 | 0.00180046 | InN6 | UBB |
| GNAI11 | 6.59E-08 | 0.30988833 | 0.562 | 0.528 | 0.00252071 | InN6 | GNAI1 |
| PLPPR41 | 6.93E-08 | 0.30318642 | 0.595 | 0.588 | 0.00264911 | InN6 | PLPPR4 |
| RASGEF1B | 9.01E-08 | 0.46236713 | 0.272 | 0.214 | 0.00344311 | InN6 | RASGEF1B |
| ENTPD1-AS1 | 1.62E-07 | 0.27722571 | 0.453 | 0.402 | 0.00619693 | InN6 | ENTPD1-AS1 |
| PHF20L11 | 2.48E-07 | 0.32381338 | 0.62 | 0.635 | 0.00946341 | InN6 | PHF20L1 |
| CNTNAP44 | 8.54E-07 | 0.75653623 | 0.564 | 0.587 | 0.03265688 | InN6 | CNTNAP4 |
| MXI1 | 1.12E-06 | 0.25623233 | 0.54 | 0.513 | 0.04274436 | InN6 | MXI1 |
| HSPA81 | 5.02E-06 | 0.2666344 | 0.348 | 0.292 | 0.19170594 | InN6 | HSPA8 |
| NAALADL22 | 6.40E-06 | 0.33387113 | 0.544 | 0.523 | 0.24474255 | InN6 | NAALADL2 |
| CACNA1D2 | 7.57E-06 | 0.31872226 | 0.848 | 0.889 | 0.2894356 | InN6 | CACNA1D |
| ADNP | 8.57E-06 | 0.25005236 | 0.591 | 0.571 | 0.32739802 | InN6 | ADNP |
| AP1G2 | 1.55E-05 | 0.25841367 | 0.468 | 0.442 | 0.5931548 | InN6 | AP1G2 |
| LARGE12 | 0.00010469 | 0.26199786 | 0.878 | 0.912 | 1 | InN6 | LARGE1 |
| AL024495.13 | 0.00037983 | 0.39841595 | 0.323 | 0.306 | 1 | InN6 | AL024495.1 |

**Supplementary Tabe 5 Pseudotime Association**

| | waldStat | df | pvalue | meanLogFC |
|---|---|---|---|---|
| DGKB | 21401.9104 | 4 | 0 | 1.00181614 |
| TSHZ2 | 19795.448 | 4 | 0 | 2.07130829 |
| PDZD2 | 18260.5429 | 4 | 0 | 1.14361889 |
| HS6ST3 | 18228.569 | 4 | 0 | 0.80759053 |
| GRIA4 | 17214.5205 | 4 | 0 | 0.51250846 |
| DCC | 16371.8301 | 4 | 0 | 1.27104268 |
| PTPRT | 14718.8239 | 4 | 0 | 1.01295595 |
| RORB | 14333.6063 | 4 | 0 | 1.2339871 |
| IL1RAPL2 | 12290.1667 | 4 | 0 | 1.44414347 |
| POU6F2 | 12254.4271 | 4 | 0 | 1.4713195 |
| EPHA6 | 11663.749 | 4 | 0 | 1.11536445 |
| CNTNAP2 | 11563.1283 | 4 | 0 | 0.32871539 |
| RORA | 11500.2166 | 4 | 0 | 0.44275121 |
| SLC35F1 | 11381.0902 | 4 | 0 | 1.25662991 |
| CBLN2 | 11310.9912 | 4 | 0 | 1.14154363 |
| RGS6 | 11222.0237 | 4 | 0 | 0.902473 |
| TMEM132D | 10799.4392 | 4 | 0 | 0.62933147 |
| DAB1 | 10695.7416 | 4 | 0 | 0.53605928 |
| FAM19A1 | 10395.3841 | 4 | 0 | 1.18542228 |
| SNTG1 | 9844.11043 | 4 | 0 | 0.42638234 |
| ADGRL3 | 9587.99396 | 4 | 0 | 0.35673343 |
| FSTL5 | 9554.06909 | 4 | 0 | 1.87463727 |
| LRRC4C | 9468.04703 | 4 | 0 | 0.96292247 |
| SOX5 | 9451.59818 | 4 | 0 | 0.92497301 |
| KCNH5 | 9318.39395 | 4 | 0 | 1.06237686 |
| GRID2 | 9264.15234 | 4 | 0 | 0.86857553 |
| PLCH1 | 9155.00714 | 4 | 0 | 3.74591365 |
| VWC2L | 9076.63987 | 4 | 0 | 1.65454812 |
| CTNNA2 | 8966.04076 | 4 | 0 | 0.42301163 |
| CDH9 | 8963.39794 | 4 | 0 | 0.62808922 |
| TOX | 8913.35239 | 4 | 0 | 0.97912833 |
| PHACTR1 | 8762.22622 | 4 | 0 | 0.32618842 |
| KAZN | 8587.01787 | 4 | 0 | 0.42074794 |
| ROBO1 | 8561.22222 | 4 | 0 | 1.46502532 |
| FOXP2 | 8511.22816 | 4 | 0 | 1.87721201 |
| RASGRF2 | 8476.43513 | 4 | 0 | 0.9089749 |
| AC011287.1 | 8283.21028 | 4 | 0 | 0.87625189 |
| TMEM108 | 8179.32964 | 4 | 0 | 0.55341714 |
| KCNQ3 | 7973.72448 | 4 | 0 | 0.40766243 |
| MEIS2 | 7836.82597 | 4 | 0 | 1.46607053 |
| LINC01122 | 7805.73718 | 4 | 0 | 0.80204318 |

| | | | | |
|---|---|---|---|---|
| SPOCK1 | 7757.70753 | 4 | 0 | 0.60539835 |
| TANC2 | 7721.45927 | 4 | 0 | 0.41243214 |
| CUX2 | 7523.82369 | 4 | 0 | 1.09038083 |
| CHRM3 | 7521.09299 | 4 | 0 | 0.35408383 |
| DGKG | 7447.37698 | 4 | 0 | 0.77115103 |
| KCNH7 | 7334.01129 | 4 | 0 | 1.35352002 |
| GRM3 | 7168.94267 | 4 | 0 | 0.91612806 |
| GALNT13 | 7020.25883 | 4 | 0 | 1.07354232 |
| CACNB4 | 6999.35323 | 4 | 0 | 0.75103593 |
| RFX3 | 6973.91425 | 4 | 0 | 0.43983656 |
| CADPS | 6791.79239 | 4 | 0 | 0.45997081 |
| LSAMP | 6708.33888 | 4 | 0 | 0.46169401 |
| GRIK2 | 6612.79743 | 4 | 0 | 0.36178779 |
| RARB | 6469.96189 | 4 | 0 | 0.72845025 |
| TRHDE | 6434.67505 | 4 | 0 | 0.94566203 |
| ST6GALNAC5 | 6430.3679 | 4 | 0 | 0.51525696 |
| ROBO2 | 6430.05051 | 4 | 0 | 0.98582612 |
| LRRTM4 | 6342.18586 | 4 | 0 | 0.26706044 |
| NTNG1 | 6250.0199 | 4 | 0 | 1.24004109 |
| CACNA2D3 | 6239.82347 | 4 | 0 | 0.42306807 |
| RCAN2 | 6141.24038 | 4 | 0 | 0.56607951 |
| NDST3 | 6022.86823 | 4 | 0 | 0.61884338 |
| NKAIN2 | 5956.18906 | 4 | 0 | 0.50084314 |
| DIAPH2 | 5949.13916 | 4 | 0 | 0.59027377 |
| AC016687.2 | 5902.26707 | 4 | 0 | 1.97721525 |
| GABRG3 | 5781.76208 | 4 | 0 | 0.43786402 |
| ENC1 | 5765.37805 | 4 | 0 | 0.52374916 |
| PTPRR | 5737.21203 | 4 | 0 | 0.433383 |
| CA10 | 5733.54696 | 4 | 0 | 1.65266524 |
| KCNAB1 | 5721.02951 | 4 | 0 | 1.05003679 |
| LHFPL6 | 5623.47814 | 4 | 0 | 0.98314392 |
| CPNE4 | 5523.76943 | 4 | 0 | 0.91893804 |
| GLIS3 | 5459.67735 | 4 | 0 | 1.57236645 |
| SGCD | 5451.49551 | 4 | 0 | 0.72036786 |
| AK5 | 5380.39815 | 4 | 0 | 0.42816584 |
| GNAL | 5339.3465 | 4 | 0 | 1.3761478 |
| SLIT3 | 5306.15666 | 4 | 0 | 1.3191004 |
| AJ009632.2 | 5280.91219 | 4 | 0 | 2.21838928 |
| RIT2 | 5254.00786 | 4 | 0 | 1.420237 |
| PTPRD | 5217.67085 | 4 | 0 | 0.37173297 |
| RBFOX1 | 5128.54175 | 4 | 0 | 0.23223348 |
| PTCHD4 | 5125.42893 | 4 | 0 | 1.46957861 |
| SLC44A5 | 5114.24837 | 4 | 0 | 0.57156754 |

| | | | | |
|---|---|---|---|---|
| MAP3K5 | 5102.8434 | 4 | 0 | 0.40263226 |
| SIPA1L1 | 5050.52673 | 4 | 0 | 0.29485252 |
| PCLO | 5043.96651 | 4 | 0 | 0.27538145 |
| LINC01378 | 4944.36813 | 4 | 0 | 1.25786532 |
| ALCAM | 4898.43847 | 4 | 0 | 0.69550474 |
| GREB1L | 4890.63071 | 4 | 0 | 1.07315583 |
| DPY19L1 | 4885.54761 | 4 | 0 | 0.9488995 |
| MCTP1 | 4872.90429 | 4 | 0 | 0.66958511 |
| ERC2 | 4866.76518 | 4 | 0 | 0.31069103 |
| SYNPR | 4833.86312 | 4 | 0 | 1.58856221 |
| PLCXD3 | 4812.30219 | 4 | 0 | 1.45939211 |
| CNTNAP4 | 4809.32444 | 4 | 0 | 0.55984458 |
| PLEKHA5 | 4783.67646 | 4 | 0 | 0.5020194 |
| LINC01725 | 4781.20838 | 4 | 0 | 0.66266923 |
| AC109466.1 | 4742.65314 | 4 | 0 | 1.91622097 |
| CLSTN2 | 4732.3923 | 4 | 0 | 0.72471455 |
| PCDH7 | 4720.32867 | 4 | 0 | 0.52656829 |
| NRG1 | 4715.11961 | 4 | 0 | 0.81417917 |
| GPC6 | 4714.29485 | 4 | 0 | 0.90679222 |
| PRKCA | 4696.98055 | 4 | 0 | 0.72537311 |
| KCTD16 | 4675.6326 | 4 | 0 | 0.36029033 |
| ABLIM1 | 4634.71554 | 4 | 0 | 0.51115726 |
| MGAT4C | 4628.84783 | 4 | 0 | 1.87387178 |
| DPP10 | 4618.86985 | 4 | 0 | 0.49702479 |
| AC022639.1 | 4596.58102 | 4 | 0 | 0.7336135 |
| NECAB1 | 4595.29468 | 4 | 0 | 0.46910507 |
| AC098850.3 | 4563.09375 | 4 | 0 | 0.44436666 |
| THSD7A | 4562.53947 | 4 | 0 | 0.85215334 |
| ZNF804A | 4530.49202 | 4 | 0 | 0.75442935 |
| COL5A2 | 4485.41826 | 4 | 0 | 1.62941267 |
| MYRIP | 4445.66684 | 4 | 0 | 0.3662975 |
| GRIA3 | 4444.05684 | 4 | 0 | 0.49499439 |
| OXR1 | 4427.86514 | 4 | 0 | 0.39952986 |
| EDIL3 | 4411.03911 | 4 | 0 | 0.69599807 |
| TMTC2 | 4335.99314 | 4 | 0 | 0.96366463 |
| GRIN2B | 4324.18659 | 4 | 0 | 0.31208572 |
| PDE1C | 4290.41682 | 4 | 0 | 1.0537543 |
| SLC24A2 | 4261.03889 | 4 | 0 | 0.3017465 |
| AC110023.1 | 4236.96482 | 4 | 0 | 0.9755914 |
| KHDRBS2 | 4233.17215 | 4 | 0 | 0.22568445 |
| HS3ST4 | 4226.99909 | 4 | 0 | 1.12252605 |
| AC092422.1 | 4092.08671 | 4 | 0 | 0.96063115 |
| CNTN4 | 4036.34592 | 4 | 0 | 0.54389783 |

| | | | | |
|---|---|---|---|---|
| PTPRK | 4024.29742 | 4 | 0 | 0.25442812 |
| 1-Mar | 4014.07771 | 4 | 0 | 0.53685697 |
| FAM135B | 4006.00166 | 4 | 0 | 0.97629624 |
| GRM8 | 3993.38936 | 4 | 0 | 1.02603655 |
| CYP1B1-AS1 | 3980.84374 | 4 | 0 | 1.1820455 |
| RXFP1 | 3980.29808 | 4 | 0 | 1.8313282 |
| PTPRO | 3978.94655 | 4 | 0 | 0.81912029 |
| ARFGEF3 | 3967.8815 | 4 | 0 | 0.34472438 |
| TENM3 | 3960.87059 | 4 | 0 | 0.74998819 |
| LRP1B | 3950.3686 | 4 | 0 | 0.2784934 |
| LINC02232 | 3945.66182 | 4 | 0 | 1.441968 |
| STXBP6 | 3942.53931 | 4 | 0 | 0.49614435 |
| CNTN3 | 3937.81531 | 4 | 0 | 0.86120367 |
| KIAA1217 | 3926.45201 | 4 | 0 | 0.89142337 |
| AC092691.1 | 3897.33651 | 4 | 0 | 0.46636683 |
| PEX5L | 3894.39695 | 4 | 0 | 0.82273004 |
| ADAMTS19 | 3870.17237 | 4 | 0 | 0.91776865 |
| CDH13 | 3866.78941 | 4 | 0 | 1.33920716 |
| FMN1 | 3866.19783 | 4 | 0 | 0.41982447 |
| LRRK1 | 3858.36194 | 4 | 0 | 0.89457065 |
| BTBD9 | 3786.72169 | 4 | 0 | 0.24253907 |
| OPCML | 3750.54095 | 4 | 0 | 0.2839183 |
| PPFIA2 | 3732.30885 | 4 | 0 | 0.28195872 |
| TMEM132B | 3724.02373 | 4 | 0 | 0.26672875 |
| DLGAP1 | 3693.96133 | 4 | 0 | 0.19551183 |
| PRKG1 | 3675.53591 | 4 | 0 | 1.42990353 |
| DOK6 | 3668.96741 | 4 | 0 | 0.70551026 |
| ARNT2 | 3607.41005 | 4 | 0 | 0.23855327 |
| CDH8 | 3601.78971 | 4 | 0 | 0.72533024 |
| GRM7 | 3601.69341 | 4 | 0 | 0.32690676 |
| STXBP5L | 3596.85621 | 4 | 0 | 0.32112697 |
| PTK2B | 3569.63926 | 4 | 0 | 0.34923926 |
| SORBS2 | 3562.03018 | 4 | 0 | 0.3434105 |
| TLN2 | 3559.68601 | 4 | 0 | 0.49653509 |
| AC084816.1 | 3483.74957 | 4 | 0 | 1.30422056 |
| ARL15 | 3482.44965 | 4 | 0 | 0.47404232 |
| DTNA | 3477.92 | 4 | 0 | 0.43145244 |
| AC008415.1 | 3477.52709 | 4 | 0 | 2.1574514 |
| PDGFD | 3466.35129 | 4 | 0 | 1.31420204 |
| CALN1 | 3459.00404 | 4 | 0 | 0.78903486 |
| MIR99AHG | 3448.72832 | 4 | 0 | 0.40601248 |
| CDH12 | 3446.24841 | 4 | 0 | 0.51859269 |
| DYNC1I1 | 3441.89908 | 4 | 0 | 0.44468981 |

| | | | | |
|---|---|---|---|---|
| FNBP1L | 3424.08026 | 4 | 0 | 1.18062128 |
| ZFPM2 | 3422.46811 | 4 | 0 | 0.33407258 |
| GARNL3 | 3419.26526 | 4 | 0 | 0.49417624 |
| TESPA1 | 3398.84657 | 4 | 0 | 0.69631841 |
| LINC00276 | 3369.98894 | 4 | 0 | 0.91339647 |
| HDAC9 | 3364.1974 | 4 | 0 | 0.52864161 |
| TMTC1 | 3356.16177 | 4 | 0 | 1.23477751 |
| GALNTL6 | 3350.92078 | 4 | 0 | 0.47953291 |
| SYN3 | 3346.9961 | 4 | 0 | 0.51648482 |
| COBLL1 | 3332.50988 | 4 | 0 | 2.21515445 |
| PAM | 3318.21993 | 4 | 0 | 0.40173652 |
| SLC22A10 | 3304.70224 | 4 | 0 | 0.65290052 |
| HPCAL1 | 3296.67744 | 4 | 0 | 0.93727649 |
| KCND3 | 3295.16724 | 4 | 0 | 0.32451479 |
| PRUNE2 | 3292.28755 | 4 | 0 | 0.63242223 |
| DTNB | 3290.91927 | 4 | 0 | 0.40609934 |
| TCF4 | 3288.68526 | 4 | 0 | 0.33788774 |
| AC019211.1 | 3272.69075 | 4 | 0 | 0.69855799 |
| NFIB | 3261.50878 | 4 | 0 | 1.35096806 |
| RAPGEF5 | 3233.8797 | 4 | 0 | 0.26276557 |
| DPYD | 3222.51278 | 4 | 0 | 0.4163878 |
| GNG2 | 3213.5622 | 4 | 0 | 0.7216509 |
| SORBS1 | 3212.74321 | 4 | 0 | 0.28048381 |
| NEBL | 3176.4721 | 4 | 0 | 0.24465939 |
| CSMD1 | 3151.29324 | 4 | 0 | 0.19052845 |
| INPP4B | 3141.07388 | 4 | 0 | 0.44091393 |
| SERPINE2 | 3130.70686 | 4 | 0 | 1.75422167 |
| LINC00507 | 3118.5944 | 4 | 0 | 1.05421281 |
| CDH18 | 3115.35601 | 4 | 0 | 0.28401821 |
| KIF16B | 3104.73835 | 4 | 0 | 0.3644531 |
| GABRG1 | 3081.14172 | 4 | 0 | 1.42099324 |
| AC091078.1 | 3072.11721 | 4 | 0 | 0.92915647 |
| HS3ST2 | 3047.28484 | 4 | 0 | 0.7584543 |
| RPS6KA2 | 3036.83597 | 4 | 0 | 0.6226337 |
| TTC6 | 3022.70129 | 4 | 0 | 0.91004608 |
| IL1RAPL1 | 3021.41911 | 4 | 0 | 0.2948439 |
| PPP2R2B | 3013.99606 | 4 | 0 | 0.23512127 |
| TSHZ3 | 2996.77252 | 4 | 0 | 0.45996507 |
| AC246817.1 | 2979.23256 | 4 | 0 | 1.11241707 |
| MAN1A1 | 2977.29285 | 4 | 0 | 0.395048 |
| RETREG1 | 2967.94147 | 4 | 0 | 0.47714033 |
| AC005162.3 | 2966.08101 | 4 | 0 | 0.58238464 |
| BMPER | 2960.50604 | 4 | 0 | 0.33755595 |

| | | | | |
|---|---|---|---|---|
| ELAVL2 | 2959.63867 | 4 | 0 | 0.74266125 |
| CTDSPL | 2945.73912 | 4 | 0 | 0.33010691 |
| TBC1D1 | 2932.83777 | 4 | 0 | 0.53304658 |
| SLIT1 | 2920.55776 | 4 | 0 | 0.45994018 |
| OLFM3 | 2919.5923 | 4 | 0 | 0.31276115 |
| DLG2 | 2915.80431 | 4 | 0 | 0.19068565 |
| KIF13A | 2911.59599 | 4 | 0 | 0.45994621 |
| SPAG16 | 2906.82163 | 4 | 0 | 0.3675372 |
| MLLT3 | 2887.06202 | 4 | 0 | 0.45556422 |
| SLIT2 | 2878.84892 | 4 | 0 | 0.40041673 |
| SLC24A4 | 2857.67675 | 4 | 0 | 0.89737226 |
| OPRM1 | 2853.38694 | 4 | 0 | 1.52336681 |
| MSC-AS1 | 2850.6313 | 4 | 0 | 0.99194662 |
| ANKRD30BL | 2846.02002 | 4 | 0 | 0.38906887 |
| ITGA9 | 2839.78254 | 4 | 0 | 0.66453454 |
| FOXP1 | 2827.18688 | 4 | 0 | 0.30563936 |
| TLK1 | 2808.44781 | 4 | 0 | 0.40305927 |
| ARHGAP26 | 2783.74302 | 4 | 0 | 0.44152952 |
| DMD | 2783.68604 | 4 | 0 | 0.32406614 |
| NETO1 | 2779.81913 | 4 | 0 | 0.2853881 |
| AC067956.1 | 2768.6436 | 4 | 0 | 0.62995784 |
| DCAF6 | 2764.97217 | 4 | 0 | 0.41945268 |
| AC010127.1 | 2755.88003 | 4 | 0 | 0.7574306 |
| CEMIP | 2753.62444 | 4 | 0 | 1.09654099 |
| GULP1 | 2733.78715 | 4 | 0 | 1.10992629 |
| ZMAT4 | 2724.32267 | 4 | 0 | 0.46575743 |
| LINC01250 | 2711.61669 | 4 | 0 | 0.33689925 |
| KHDRBS3 | 2703.31168 | 4 | 0 | 0.21441648 |
| UNC5D | 2699.89195 | 4 | 0 | 0.27431947 |
| RANBP17 | 2667.00525 | 4 | 0 | 1.13075125 |
| FAM19A2 | 2662.98429 | 4 | 0 | 0.41073348 |
| DNAJC1 | 2662.69841 | 4 | 0 | 0.45097064 |
| SLC4A4 | 2660.04031 | 4 | 0 | 0.88697514 |
| CACNA1E | 2636.38879 | 4 | 0 | 0.19879588 |
| ADAMTS3 | 2633.90582 | 4 | 0 | 1.08794593 |
| CEMIP2 | 2625.28834 | 4 | 0 | 0.67071807 |
| KCNIP2 | 2622.75461 | 4 | 0 | 0.69107914 |
| AC073578.2 | 2622.23447 | 4 | 0 | 1.26469899 |
| COL24A1 | 2616.64018 | 4 | 0 | 1.66881048 |
| ARHGEF28 | 2611.32117 | 4 | 0 | 1.22934653 |
| FRAS1 | 2604.80926 | 4 | 0 | 0.87977643 |
| GRIP1 | 2601.43725 | 4 | 0 | 0.52476125 |
| GRIK4 | 2597.59084 | 4 | 0 | 0.80621013 |

| | | | | |
|---|---|---|---|---|
| SLC35F3 | 2597.27596 | 4 | 0 | 0.37011492 |
| CELF2 | 2593.97881 | 4 | 0 | 0.23788763 |
| MAGI2 | 2591.13836 | 4 | 0 | 0.2229393 |
| BRINP2 | 2588.73896 | 4 | 0 | 0.41469181 |
| XYLT1 | 2586.66079 | 4 | 0 | 0.49456644 |
| ANO3 | 2585.84838 | 4 | 0 | 0.5816289 |
| PTPRN2 | 2580.27653 | 4 | 0 | 0.26571557 |
| ITPK1 | 2565.36881 | 4 | 0 | 0.36637315 |
| TMEM178B | 2549.0195 | 4 | 0 | 0.19524698 |
| FAT3 | 2520.34886 | 4 | 0 | 0.36552176 |
| IL1RAP | 2515.15356 | 4 | 0 | 0.87814846 |
| KIAA0319 | 2514.20899 | 4 | 0 | 0.35851768 |
| PTPN13 | 2481.57281 | 4 | 0 | 0.93862035 |
| BRINP3 | 2474.6769 | 4 | 0 | 0.53524514 |
| ITPR1 | 2472.33441 | 4 | 0 | 0.17882176 |
| SORCS3 | 2469.40836 | 4 | 0 | 0.86910496 |
| ALDH1A1 | 2456.95747 | 4 | 0 | 1.42496628 |
| PPP4R4 | 2456.48022 | 4 | 0 | 0.61048747 |
| CACNG3 | 2451.64539 | 4 | 0 | 0.34004859 |
| BACH2 | 2448.49733 | 4 | 0 | 0.55015318 |
| P2RX7 | 2446.04591 | 4 | 0 | 0.326765 |
| PTK2 | 2433.98679 | 4 | 0 | 0.20684244 |
| DNAH14 | 2428.98783 | 4 | 0 | 0.51733757 |
| MIR181A1HG | 2412.65404 | 4 | 0 | 0.52198627 |
| RPS6KC1 | 2412.12527 | 4 | 0 | 0.31017213 |
| COL6A1 | 2404.38311 | 4 | 0 | 0.75964549 |
| LIMCH1 | 2403.82588 | 4 | 0 | 0.29026758 |
| PDZRN4 | 2398.74383 | 4 | 0 | 1.14790208 |
| PAG1 | 2389.14054 | 4 | 0 | 0.5801969 |
| PARM1 | 2368.56287 | 4 | 0 | 0.85070954 |
| PDE4B | 2368.14506 | 4 | 0 | 0.69707009 |
| CNTN5 | 2366.57881 | 4 | 0 | 0.64437954 |
| PDE7B | 2359.46677 | 4 | 0 | 0.31866757 |
| LAMB1 | 2358.83212 | 4 | 0 | 0.58340374 |
| KCTD1 | 2348.00993 | 4 | 0 | 0.39304966 |
| TCERG1L | 2347.71492 | 4 | 0 | 0.89828724 |
| PVT1 | 2334.03915 | 4 | 0 | 0.97674336 |
| PCBP3 | 2324.02764 | 4 | 0 | 1.98795209 |
| L3MBTL4 | 2310.48569 | 4 | 0 | 1.80675301 |
| MAPK4 | 2305.96573 | 4 | 0 | 0.41132593 |
| CDHR3 | 2304.84862 | 4 | 0 | 0.47065088 |
| DCLK2 | 2299.70686 | 4 | 0 | 0.3442617 |
| ASIC2 | 2299.10289 | 4 | 0 | 0.50165081 |

| | | | | |
|---|---|---|---|---|
| FARP1 | 2285.95983 | 4 | 0 | 0.52918125 |
| LINC01202 | 2280.1002 | 4 | 0 | 0.74704287 |
| LINC02389 | 2275.02965 | 4 | 0 | 0.48985559 |
| ARHGAP10 | 2253.31346 | 4 | 0 | 0.68572893 |
| GAREM1 | 2249.35669 | 4 | 0 | 0.64261716 |
| PTPRB | 2242.283 | 4 | 0 | 0.49697722 |
| AC008574.1 | 2234.23127 | 4 | 0 | 1.68153432 |
| USP25 | 2231.12413 | 4 | 0 | 0.28410425 |
| KCNB2 | 2216.81597 | 4 | 0 | 0.39965687 |
| ST6GAL2 | 2214.41176 | 4 | 0 | 0.7671439 |
| NPAS3 | 2212.49642 | 4 | 0 | 0.65940717 |
| KCNN2 | 2212.23531 | 4 | 0 | 0.64323769 |
| NLK | 2187.5208 | 4 | 0 | 0.28919772 |
| NCAM2 | 2167.8006 | 4 | 0 | 0.59073018 |
| MBNL1 | 2166.73155 | 4 | 0 | 0.22202774 |
| PDZRN3 | 2162.97491 | 4 | 0 | 0.38848469 |
| PARD3 | 2154.08628 | 4 | 0 | 1.48242136 |
| SLC5A4-AS1 | 2147.29679 | 4 | 0 | 0.39624641 |
| MED12L | 2145.13018 | 4 | 0 | 0.34944276 |
| EFNA5 | 2142.34298 | 4 | 0 | 0.32785306 |
| KIAA1958 | 2133.15571 | 4 | 0 | 0.49910712 |
| NEDD4L | 2127.1513 | 4 | 0 | 0.36761647 |
| LAMA2 | 2120.19706 | 4 | 0 | 1.56416226 |
| LINC01197 | 2115.32108 | 4 | 0 | 1.14012133 |
| CACNA1A | 2112.89429 | 4 | 0 | 0.19738671 |
| NTRK3 | 2103.8795 | 4 | 0 | 0.22283639 |
| PHYHIPL | 2101.15265 | 4 | 0 | 0.51500362 |
| AC090568.2 | 2098.37989 | 4 | 0 | 0.75258499 |
| NTM | 2083.33625 | 4 | 0 | 0.40559013 |
| CAMTA1 | 2081.48641 | 4 | 0 | 0.20907643 |
| MAN2A1 | 2076.78811 | 4 | 0 | 0.30559942 |
| LINC02306 | 2075.62328 | 4 | 0 | 0.9677997 |
| ATP6V1H | 2031.1632 | 4 | 0 | 0.35163825 |
| KIAA1211L | 2030.68724 | 4 | 0 | 0.22380339 |
| ATP2B1 | 2020.97877 | 4 | 0 | 0.16158821 |
| LYPD6B | 2016.69329 | 4 | 0 | 1.8547628 |
| OSBPL3 | 2015.9562 | 4 | 0 | 0.67879304 |
| LAMP5 | 2013.22633 | 4 | 0 | 1.73066294 |
| FAM189A1 | 2011.78116 | 4 | 0 | 0.51225637 |
| SIDT1 | 2009.86155 | 4 | 0 | 0.82986865 |
| GRIA2 | 2009.13054 | 4 | 0 | 0.13010521 |
| CDH22 | 2006.07907 | 4 | 0 | 0.382736 |
| DLGAP2 | 2001.97396 | 4 | 0 | 0.21946899 |

| | | | | |
|---|---|---|---|---|
| NGEF | 1998.07553 | 4 | 0 | 0.3466837 |
| FSHR | 1991.76559 | 4 | 0 | 1.27826004 |
| CADPS2 | 1976.43829 | 4 | 0 | 0.61322139 |
| MEG8 | 1966.12011 | 4 | 0 | 0.645191 |
| CNTNAP3B | 1962.56393 | 4 | 0 | 1.41570204 |
| CDH4 | 1953.09992 | 4 | 0 | 0.8122375 |
| PLCB1 | 1936.18785 | 4 | 0 | 0.38337659 |
| HHAT | 1932.83528 | 4 | 0 | 0.69663141 |
| RYR2 | 1930.78571 | 4 | 0 | 0.23187756 |
| DDAH1 | 1927.21165 | 4 | 0 | 0.47995691 |
| ATXN1 | 1917.65414 | 4 | 0 | 0.20209048 |
| ANK3 | 1914.76964 | 4 | 0 | 0.18266802 |
| SEMA3A | 1908.89683 | 4 | 0 | 0.8867237 |
| SPOCK3 | 1904.64729 | 4 | 0 | 1.1235165 |
| THRB | 1896.43397 | 4 | 0 | 0.17382535 |
| GLRA3 | 1895.60298 | 4 | 0 | 0.74712231 |
| VPS41 | 1889.31485 | 4 | 0 | 0.24619107 |
| AC120193.1 | 1886.56521 | 4 | 0 | 0.77274737 |
| DKK3 | 1884.88381 | 4 | 0 | 0.27072138 |
| STK39 | 1879.32933 | 4 | 0 | 0.53656054 |
| NRXN3 | 1872.96406 | 4 | 0 | 0.29166138 |
| MBOAT2 | 1863.23637 | 4 | 0 | 0.5932612 |
| NR3C2 | 1860.88864 | 4 | 0 | 0.28503421 |
| VLDLR-AS1 | 1855.55541 | 4 | 0 | 0.62015205 |
| RHOBTB1 | 1850.04164 | 4 | 0 | 0.8340473 |
| NKAIN3 | 1848.37714 | 4 | 0 | 1.20343795 |
| DSCAM | 1844.09787 | 4 | 0 | 0.18175969 |
| ADCY2 | 1834.22594 | 4 | 0 | 0.47187956 |
| LMO3 | 1834.19729 | 4 | 0 | 0.29160712 |
| STMN2 | 1824.43527 | 4 | 0 | 0.5198095 |
| CNGB1 | 1812.97779 | 4 | 0 | 1.90376821 |
| NREP | 1811.51392 | 4 | 0 | 0.87079747 |
| AGBL4 | 1800.2399 | 4 | 0 | 0.24836999 |
| PCDH9 | 1796.255 | 4 | 0 | 0.25306025 |
| GRIA1 | 1790.82369 | 4 | 0 | 0.35232822 |
| ADCY1 | 1786.81754 | 4 | 0 | 0.33506962 |
| TMEM65 | 1785.45838 | 4 | 0 | 0.59646007 |
| SMYD3 | 1784.2222 | 4 | 0 | 0.25861108 |
| CCBE1 | 1777.65527 | 4 | 0 | 1.49158891 |
| BICC1 | 1776.00724 | 4 | 0 | 0.64048753 |
| LARGE1 | 1770.82631 | 4 | 0 | 0.28374852 |
| EML6 | 1768.20981 | 4 | 0 | 0.39459395 |
| NIN | 1766.69982 | 4 | 0 | 0.24453027 |

| | | | | |
|---|---|---|---|---|
| TMEM117 | 1758.61915 | 4 | 0 | 0.46356833 |
| PTPRS | 1756.16715 | 4 | 0 | 0.31769636 |
| KCND2 | 1754.91315 | 4 | 0 | 0.37213326 |
| NUAK1 | 1738.73128 | 4 | 0 | 0.19950152 |
| CYP46A1 | 1738.62176 | 4 | 0 | 0.42828337 |
| CACNA2D1 | 1731.92283 | 4 | 0 | 0.22963411 |
| HCN1 | 1729.21724 | 4 | 0 | 0.33519776 |
| CEP126 | 1729.04298 | 4 | 0 | 0.49039172 |
| CNTNAP5 | 1727.79222 | 4 | 0 | 0.42258319 |
| AC093607.1 | 1717.23423 | 4 | 0 | 0.65525541 |
| HECW1 | 1707.73465 | 4 | 0 | 0.14600748 |
| DLEU2 | 1707.20251 | 4 | 0 | 0.28132539 |
| GSG1L | 1703.57591 | 4 | 0 | 0.87892542 |
| SMAD3 | 1697.35234 | 4 | 0 | 0.35277468 |
| MAST4 | 1696.96283 | 4 | 0 | 0.33698889 |
| SLC12A8 | 1693.6715 | 4 | 0 | 0.55060879 |
| NECTIN3 | 1672.77611 | 4 | 0 | 0.32158361 |
| VAV3 | 1671.82362 | 4 | 0 | 1.32503645 |
| RGS7 | 1670.67973 | 4 | 0 | 0.23057306 |
| SHISA6 | 1670.4801 | 4 | 0 | 0.83859208 |
| CERS6 | 1666.58986 | 4 | 0 | 0.29328401 |
| NRG3 | 1662.91578 | 4 | 0 | 0.1748597 |
| ASPH | 1651.95728 | 4 | 0 | 0.27015598 |
| ITGB8 | 1651.49636 | 4 | 0 | 0.98304926 |
| CDH10 | 1642.88718 | 4 | 0 | 0.21427063 |
| SEMA6D | 1634.94565 | 4 | 0 | 0.53546217 |
| GAB2 | 1633.99054 | 4 | 0 | 0.54813131 |
| GPR158 | 1626.42309 | 4 | 0 | 0.23726426 |
| FBXW7 | 1622.02149 | 4 | 0 | 0.18472893 |
| ALS2 | 1615.18421 | 4 | 0 | 0.28592217 |
| FIGN | 1612.1599 | 4 | 0 | 1.74090692 |
| LRRC8B | 1595.78815 | 4 | 0 | 0.22223004 |
| PDE10A | 1586.03169 | 4 | 0 | 0.52939473 |
| ZEB1 | 1583.99622 | 4 | 0 | 0.20031436 |
| ST3GAL6 | 1582.72149 | 4 | 0 | 0.43229074 |
| PALMD | 1581.21729 | 4 | 0 | 1.71844865 |
| AKAP13 | 1575.13895 | 4 | 0 | 0.4886147 |
| ELAVL4 | 1572.75406 | 4 | 0 | 0.41842607 |
| MAP7 | 1567.7116 | 4 | 0 | 0.2122274 |
| WWC2 | 1560.37134 | 4 | 0 | 0.77969724 |
| HSPA12A | 1559.10946 | 4 | 0 | 0.34470806 |
| SLC2A13 | 1548.94588 | 4 | 0 | 0.26810459 |
| UBE2E3 | 1545.30953 | 4 | 0 | 0.44218953 |

| | | | | |
|---|---|---|---|---|
| EVI5 | 1542.65077 | 4 | 0 | 0.37177353 |
| RIPOR2 | 1540.32468 | 4 | 0 | 0.64744259 |
| PCDH11Y | 1539.02598 | 4 | 0 | 1.26682589 |
| ATP2B4 | 1538.31086 | 4 | 0 | 0.32811194 |
| SFMBT2 | 1537.94949 | 4 | 0 | 0.77122382 |
| KLHL2 | 1536.80565 | 4 | 0 | 0.28360271 |
| RYR3 | 1536.05978 | 4 | 0 | 0.79563505 |
| CFAP221 | 1530.81992 | 4 | 0 | 0.38074823 |
| LINC00937 | 1530.47414 | 4 | 0 | 0.47879228 |
| KCNN3 | 1528.99528 | 4 | 0 | 0.63980385 |
| CNIH3 | 1525.961 | 4 | 0 | 0.46882352 |
| CPLX2 | 1521.77028 | 4 | 0 | 0.57965036 |
| KLHL32 | 1518.02198 | 4 | 0 | 0.28416056 |
| RTN4 | 1515.98361 | 4 | 0 | 0.19380044 |
| LINC01500 | 1514.17651 | 4 | 0 | 1.86527938 |
| KITLG | 1510.17874 | 4 | 0 | 0.78756925 |
| VWC2 | 1509.63915 | 4 | 0 | 0.91021628 |
| AC016831.7 | 1503.92564 | 4 | 0 | 0.23221194 |
| NEK11 | 1493.80961 | 4 | 0 | 0.44134783 |
| CIT | 1493.41214 | 4 | 0 | 0.32174179 |
| FHOD3 | 1482.84759 | 4 | 0 | 0.30017059 |
| FANCL | 1480.13737 | 4 | 0 | 0.28379725 |
| PDE4D | 1477.94584 | 4 | 0 | 0.22623786 |
| LINC01090 | 1477.59764 | 4 | 0 | 1.80004443 |
| SCD5 | 1473.22616 | 4 | 0 | 0.34784722 |
| B3GAT1 | 1467.73957 | 4 | 0 | 0.35993637 |
| LDLRAD4 | 1466.76326 | 4 | 0 | 0.32809325 |
| HTR4 | 1462.94348 | 4 | 0 | 1.66351283 |
| UPP2 | 1457.39596 | 4 | 0 | 0.31740016 |
| CSGALNACT1 | 1455.02697 | 4 | 0 | 0.33424839 |
| TMEFF2 | 1453.5438 | 4 | 0 | 0.45101797 |
| ADAMTS16 | 1447.07928 | 4 | 0 | 1.846665 |
| KCNT2 | 1443.81525 | 4 | 0 | 1.06760697 |
| MOXD1 | 1443.5747 | 4 | 0 | 0.96097315 |
| PKP4 | 1439.80024 | 4 | 0 | 0.21587627 |
| MTHFD2L | 1433.62732 | 4 | 0 | 0.31852889 |
| SCN1A | 1429.13303 | 4 | 0 | 0.21671828 |
| IL12A-AS1 | 1427.26795 | 4 | 0 | 1.02272402 |
| NETO2 | 1423.06356 | 4 | 0 | 0.76217708 |
| BAIAP2 | 1421.93562 | 4 | 0 | 0.24780452 |
| BAIAP3 | 1419.52364 | 4 | 0 | 1.03688988 |
| GAP43 | 1416.17685 | 4 | 0 | 0.35199538 |
| PTPRG | 1414.99219 | 4 | 0 | 0.28763687 |

| | | | | |
|---|---|---|---|---|
| SEZ6L | 1414.98048 | 4 | 0 | 0.3535607 |
| GPC5 | 1411.37293 | 4 | 0 | 0.80281043 |
| SLC38A11 | 1405.83311 | 4 | 0 | 0.34393694 |
| LPP | 1400.67996 | 4 | 0 | 0.65045382 |
| USP43 | 1399.42132 | 4 | 0 | 0.62260393 |
| NAPEPLD | 1398.98975 | 4 | 0 | 0.34046782 |
| PID1 | 1394.85351 | 4 | 0 | 0.74372931 |
| SGK3 | 1390.36323 | 4 | 0 | 0.26080517 |
| CARMIL1 | 1388.80005 | 4 | 0 | 0.25897786 |
| CCDC3 | 1385.88601 | 4 | 0 | 0.31236714 |
| TTC28 | 1385.75991 | 4 | 0 | 0.35378871 |
| SMAP1 | 1384.1607 | 4 | 0 | 0.26092368 |
| LINC01331 | 1383.26291 | 4 | 0 | 1.70938548 |
| GNAS | 1378.29003 | 4 | 0 | 0.35135853 |
| NLGN1 | 1373.77584 | 4 | 0 | 0.20997855 |
| ELL2 | 1372.66996 | 4 | 0 | 0.27061754 |
| OTOGL | 1371.93686 | 4 | 0 | 1.10726969 |
| PRKD1 | 1369.15499 | 4 | 0 | 1.04501537 |
| AC113383.1 | 1365.19624 | 4 | 0 | 0.5384104 |
| RHOBTB2 | 1365.05131 | 4 | 0 | 0.27130172 |
| CAMK2D | 1359.19596 | 4 | 0 | 0.3436877 |
| DPF3 | 1357.99674 | 4 | 0 | 0.59666217 |
| PHACTR2 | 1349.85063 | 4 | 0 | 0.40048457 |
| NOS1AP | 1349.41722 | 4 | 0 | 0.56843173 |
| TMEM163 | 1348.31723 | 4 | 0 | 0.85415246 |
| STXBP1 | 1346.58582 | 4 | 0 | 0.18709307 |
| CRYM | 1344.62769 | 4 | 0 | 0.70681101 |
| MARCKS | 1343.86523 | 4 | 0 | 0.57023636 |
| UNC13C | 1342.06987 | 4 | 0 | 0.66832341 |
| KIRREL3 | 1341.81843 | 4 | 0 | 0.35905248 |
| SCAPER | 1341.34176 | 4 | 0 | 0.29194375 |
| LIN7A | 1335.90179 | 4 | 0 | 0.30374512 |
| MIR4500HG | 1325.18771 | 4 | 0 | 0.70975667 |
| ZNF704 | 1323.54538 | 4 | 0 | 0.24366629 |
| DOCK10 | 1321.98238 | 4 | 0 | 0.79920968 |
| SOBP | 1313.4886 | 4 | 0 | 0.2487363 |
| DAPK1 | 1309.38923 | 4 | 0 | 0.26513291 |
| ESRRG | 1308.29935 | 4 | 0 | 0.80972379 |
| SLC8A1 | 1298.45695 | 4 | 0 | 0.19899564 |
| NUP93 | 1292.27663 | 4 | 0 | 0.42309875 |
| EML1 | 1291.64373 | 4 | 0 | 0.32806538 |
| FRMD6 | 1288.87272 | 4 | 0 | 0.65694242 |
| KCNIP4 | 1285.97381 | 4 | 0 | 0.13648183 |

| | | | | |
|---|---|---|---|---|
| ADGRB3 | 1281.36217 | 4 | 0 | 0.15096744 |
| RASAL2 | 1280.77567 | 4 | 0 | 0.2053686 |
| NHSL2 | 1276.23933 | 4 | 0 | 0.4071109 |
| FAM13A | 1271.0143 | 4 | 0 | 0.17534199 |
| NEO1 | 1266.57982 | 4 | 0 | 0.30385906 |
| TMOD1 | 1265.07377 | 4 | 0 | 0.34587657 |
| CTNNB1 | 1260.63407 | 4 | 0 | 0.36916708 |
| AGBL1 | 1259.92072 | 4 | 0 | 1.25729376 |
| AC139491.7 | 1259.44533 | 4 | 0 | 0.4616885 |
| DPP6 | 1259.36487 | 4 | 0 | 0.27227018 |
| GRM1 | 1256.63676 | 4 | 0 | 0.3320112 |
| TRPS1 | 1256.55638 | 4 | 0 | 0.74783511 |
| SHTN1 | 1256.36845 | 4 | 0 | 0.31225278 |
| SND1 | 1251.8394 | 4 | 0 | 0.168355 |
| HOPX | 1250.23737 | 4 | 0 | 0.32817171 |
| TRPC6 | 1247.08776 | 4 | 0 | 1.64493202 |
| STXBP5-AS1 | 1241.21517 | 4 | 0 | 0.34563284 |
| ANKRD28 | 1235.34374 | 4 | 0 | 0.23174067 |
| DAAM1 | 1234.2944 | 4 | 0 | 0.20558948 |
| ACAP2 | 1234.02582 | 4 | 0 | 0.22886283 |
| CACNB2 | 1232.30592 | 4 | 0 | 0.22818064 |
| PRICKLE2 | 1231.89974 | 4 | 0 | 0.28022742 |
| STXBP5 | 1231.253 | 4 | 0 | 0.16312238 |
| ABHD2 | 1230.31613 | 4 | 0 | 0.3958569 |
| SNTB2 | 1229.93658 | 4 | 0 | 0.30823374 |
| SYNJ2 | 1228.0349 | 4 | 0 | 0.43586703 |
| AC069133.1 | 1225.8013 | 4 | 0 | 0.59874001 |
| MACF1 | 1223.98778 | 4 | 0 | 0.14374765 |
| PLCL1 | 1215.38454 | 4 | 0 | 0.35399599 |
| IPCEF1 | 1214.3364 | 4 | 0 | 0.28465074 |
| RNF115 | 1210.39416 | 4 | 0 | 0.21323524 |
| PRAG1 | 1208.19746 | 4 | 0 | 0.65991721 |
| GHR | 1201.67286 | 4 | 0 | 0.58256431 |
| CTNND2 | 1195.31227 | 4 | 0 | 0.15002166 |
| CDK14 | 1177.51995 | 4 | 0 | 0.23435088 |
| BICD1 | 1174.08232 | 4 | 0 | 0.27695256 |
| SPECC1 | 1167.03788 | 4 | 0 | 0.31338623 |
| TTC3 | 1166.42464 | 4 | 0 | 0.14383658 |
| NRSN1 | 1165.40194 | 4 | 0 | 0.3790597 |
| PAK3 | 1162.31075 | 4 | 0 | 0.16505659 |
| MAPK1 | 1162.10213 | 4 | 0 | 0.25576259 |
| AC103855.3 | 1161.25708 | 4 | 0 | 0.49312059 |
| SCHLAP1 | 1158.12753 | 4 | 0 | 0.58318663 |

| | | | | |
|---|---|---|---|---|
| TTC39C | 1157.60531 | 4 | 0 | 0.33854424 |
| ZNF609 | 1157.10358 | 4 | 0 | 0.22338868 |
| LINC02296 | 1156.53727 | 4 | 0 | 0.84935329 |
| MMP16 | 1156.42024 | 4 | 0 | 0.39578706 |
| MPP6 | 1156.21798 | 4 | 0 | 0.30526031 |
| PLPPR4 | 1145.81549 | 4 | 0 | 0.37980847 |
| CHSY3 | 1143.03551 | 4 | 0 | 0.22516649 |
| RALYL | 1141.07559 | 4 | 0 | 0.09756474 |
| LMO4 | 1136.47996 | 4 | 0 | 0.1932411 |
| MIR4435-2H | 1135.81897 | 4 | 0 | 0.35876126 |
| CELF4 | 1129.71868 | 4 | 0 | 0.38895674 |
| CHKA | 1129.33359 | 4 | 0 | 0.43038876 |
| RAPH1 | 1126.58608 | 4 | 0 | 0.3155465 |
| PSD3 | 1125.88026 | 4 | 0 | 0.18863288 |
| FP700111.1 | 1122.48257 | 4 | 0 | 0.32371095 |
| CHN1 | 1122.40227 | 4 | 0 | 0.17547068 |
| CDK6 | 1120.93655 | 4 | 0 | 0.32794014 |
| MB21D2 | 1120.5985 | 4 | 0 | 0.37342805 |
| ZFHX3 | 1119.79569 | 4 | 0 | 0.45123176 |
| MAP2 | 1118.00875 | 4 | 0 | 0.12668206 |
| CCNH | 1116.20742 | 4 | 0 | 0.48334764 |
| EPHA5 | 1112.66791 | 4 | 0 | 0.44876257 |
| LINC01322 | 1112.57461 | 4 | 0 | 0.58012642 |
| MACROD2 | 1111.02599 | 4 | 0 | 0.16596662 |
| LY86-AS1 | 1109.56857 | 4 | 0 | 0.34867581 |
| AL445250.1 | 1107.90815 | 4 | 0 | 0.36646261 |
| NBAS | 1105.2142 | 4 | 0 | 0.22419947 |
| AC073365.1 | 1104.79024 | 4 | 0 | 0.33214627 |
| AP005328.1 | 1100.48223 | 4 | 0 | 1.0913544 |
| AC006504.5 | 1100.23837 | 4 | 0 | 0.3713246 |
| LCORL | 1096.54935 | 4 | 0 | 0.68120479 |
| NR2F1-AS1 | 1095.07886 | 4 | 0 | 0.37644872 |
| TRERF1 | 1093.47533 | 4 | 0 | 0.27070607 |
| AUTS2 | 1088.83141 | 4 | 0 | 0.20182951 |
| FCHSD2 | 1085.97871 | 4 | 0 | 0.47165405 |
| MOB1B | 1081.81298 | 4 | 0 | 0.40100354 |
| ADGRL2 | 1079.90778 | 4 | 0 | 0.33923966 |
| ANO5 | 1078.84383 | 4 | 0 | 0.27234152 |
| RTN1 | 1078.00181 | 4 | 0 | 0.18336563 |
| EPHA7 | 1077.07684 | 4 | 0 | 0.82856637 |
| MEIS3 | 1075.44955 | 4 | 0 | 0.64628763 |
| SHANK2 | 1074.79225 | 4 | 0 | 0.1952957 |
| AC007563.2 | 1074.34832 | 4 | 0 | 0.25908457 |

| | | | | |
|---|---|---|---|---|
| HERC1 | 1074.34606 | 4 | 0 | 0.13038095 |
| NBEA | 1073.94133 | 4 | 0 | 0.18385506 |
| AL392023.2 | 1072.50569 | 4 | 0 | 1.29546195 |
| TIAM1 | 1070.63627 | 4 | 0 | 0.27400073 |
| FAM155A | 1070.51776 | 4 | 0 | 0.1566113 |
| PRKCE | 1070.26434 | 4 | 0 | 0.30018914 |
| CACNA1C | 1061.99717 | 4 | 0 | 0.1456098 |
| RAB27B | 1060.12982 | 4 | 0 | 0.39801826 |
| SAMD5 | 1057.79901 | 4 | 0 | 0.62599975 |
| GLIS1 | 1051.92904 | 4 | 0 | 0.29413388 |
| COL26A1 | 1050.13829 | 4 | 0 | 0.40672699 |
| GUCY1A2 | 1048.06233 | 4 | 0 | 0.33062434 |
| DENND3 | 1045.68527 | 4 | 0 | 0.42117881 |
| ADAMTS9-AS | 1041.19649 | 4 | 0 | 1.50558499 |
| CASC15 | 1041.12062 | 4 | 0 | 0.23445925 |
| PDGFC | 1041.09585 | 4 | 0 | 0.53998686 |
| ASXL3 | 1039.67092 | 4 | 0 | 0.6027394 |
| PUM3 | 1037.67522 | 4 | 0 | 0.29719301 |
| SORCS1 | 1037.08428 | 4 | 0 | 0.51742429 |
| ANK2 | 1035.6794 | 4 | 0 | 0.13176425 |
| FUT8 | 1035.31438 | 4 | 0 | 0.30896215 |
| AL589740.1 | 1031.19874 | 4 | 0 | 1.12780061 |
| ASTN2 | 1027.14943 | 4 | 0 | 0.64680703 |
| PCSK6 | 1026.48844 | 4 | 0 | 0.93136487 |
| VSNL1 | 1023.62539 | 4 | 0 | 0.204155 |
| ENOX1 | 1023.43525 | 4 | 0 | 0.28348826 |
| PDE1A | 1023.31597 | 4 | 0 | 0.2911557 |
| NRXN1 | 1021.27745 | 4 | 0 | 0.11913549 |
| ZNF804B | 1018.18363 | 4 | 0 | 1.06873016 |
| AC109779.1 | 1018.13259 | 4 | 0 | 1.32463905 |
| FLRT2 | 1017.09023 | 4 | 0 | 0.19077519 |
| CACNA1D | 1016.9573 | 4 | 0 | 0.16557288 |
| SHROOM3 | 1016.93852 | 4 | 0 | 0.46709491 |
| BMPR1B | 1016.30253 | 4 | 0 | 0.90718544 |
| NAV2 | 1013.38449 | 4 | 0 | 0.18283554 |
| SYN2 | 1011.8765 | 4 | 0 | 0.19193092 |
| SPARCL1 | 1010.33529 | 4 | 0 | 0.21644674 |
| NFAT5 | 1008.24959 | 4 | 0 | 0.15002844 |
| PLCB4 | 1007.2575 | 4 | 0 | 0.3661876 |
| ST6GALNAC3 | 1007.18531 | 4 | 0 | 0.5219997 |
| WDR17 | 1005.42847 | 4 | 0 | 0.21496802 |
| ZNF827 | 996.559377 | 4 | 0 | 0.47361072 |
| FNDC3B | 996.325861 | 4 | 0 | 0.63642032 |

| | | | | |
|---|---|---|---|---|
| MAPK10 | 993.934914 | 4 | 0 | 0.15678184 |
| NCDN | 989.183367 | 4 | 0 | 0.45588217 |
| ZMYND8 | 988.210622 | 4 | 0 | 0.14606821 |
| SEC24D | 987.864682 | 4 | 0 | 0.5494669 |
| ACACA | 986.4009 | 4 | 0 | 0.26564754 |
| ACTN1 | 986.025116 | 4 | 0 | 0.23927431 |
| ATRNL1 | 984.461947 | 4 | 0 | 0.25954185 |
| IGSF11 | 982.748996 | 4 | 0 | 0.30464554 |
| KCNQ1OT1 | 981.144333 | 4 | 0 | 0.51183596 |
| TRPC5 | 979.850042 | 4 | 0 | 0.34927949 |
| ST8SIA1 | 977.12446 | 4 | 0 | 0.33617936 |
| C8orf34 | 977.119694 | 4 | 0 | 1.06655368 |
| FBXO32 | 975.977143 | 4 | 0 | 0.25523108 |
| SUSD4 | 975.453795 | 4 | 0 | 0.42609399 |
| IPO5 | 973.87058 | 4 | 0 | 0.24275418 |
| KCNK1 | 970.055125 | 4 | 0 | 0.1458829 |
| AGAP1 | 968.409318 | 4 | 0 | 0.13210961 |
| ZDHHC14 | 965.037177 | 4 | 0 | 0.21295298 |
| TJP1 | 962.978044 | 4 | 0 | 0.18522268 |
| DCLK1 | 961.920363 | 4 | 0 | 0.1789798 |
| SSBP3 | 960.626295 | 4 | 0 | 0.21438022 |
| DGKH | 960.576742 | 4 | 0 | 0.40214262 |
| ZNF385B | 960.095724 | 4 | 0 | 0.16157919 |
| PGBD5 | 957.388844 | 4 | 0 | 0.29295 |
| SESN1 | 955.823168 | 4 | 0 | 0.20643531 |
| MAML3 | 954.881696 | 4 | 0 | 0.61074665 |
| VWA3A | 950.155677 | 4 | 0 | 0.57919147 |
| SPATS2L | 945.254764 | 4 | 0 | 0.25667494 |
| FNDC3A | 941.818159 | 4 | 0 | 0.15533524 |
| SERPINI1 | 936.761152 | 4 | 0 | 0.30916723 |
| AC124312.1 | 932.948672 | 4 | 0 | 0.18382254 |
| DOCK9 | 930.812887 | 4 | 0 | 0.29088988 |
| LY6H | 923.316304 | 4 | 0 | 0.7202199 |
| RAP1GDS1 | 922.618181 | 4 | 0 | 0.32527416 |
| EPB41 | 922.380101 | 4 | 0 | 0.43118633 |
| PKN2-AS1 | 921.559496 | 4 | 0 | 0.50654777 |
| MYO6 | 920.286621 | 4 | 0 | 0.33024043 |
| CDH11 | 919.17245 | 4 | 0 | 0.42074219 |
| UTRN | 915.969611 | 4 | 0 | 1.77657709 |
| EML5 | 912.912905 | 4 | 0 | 0.17759027 |
| AC100818.1 | 912.419564 | 4 | 0 | 1.59909217 |
| PPP3CA | 911.366728 | 4 | 0 | 0.16460076 |
| CABP1 | 911.297482 | 4 | 0 | 0.36213981 |

| | | | | |
|---|---|---|---|---|
| AC025887.2 | 909.924342 | 4 | 0 | 0.81698331 |
| JPH1 | 908.196307 | 4 | 0 | 0.40839009 |
| STARD13 | 907.939871 | 4 | 0 | 0.6078338 |
| STMN1 | 905.941572 | 4 | 0 | 0.54252297 |
| MIR137HG | 903.903574 | 4 | 0 | 0.27266425 |
| LINGO2 | 903.801843 | 4 | 0 | 0.20716295 |
| BCL11A | 903.536765 | 4 | 0 | 0.35879414 |
| GOLGA8B | 900.719131 | 4 | 0 | 0.2688809 |
| KSR2 | 899.285747 | 4 | 0 | 0.22816603 |
| NIPAL2 | 899.272632 | 4 | 0 | 0.36667223 |
| KIAA1549 | 895.945153 | 4 | 0 | 0.32144152 |
| PRKCI | 894.423418 | 4 | 0 | 0.24182905 |
| SLF1 | 894.382986 | 4 | 0 | 0.1860115 |
| RBFOX3 | 893.217339 | 4 | 0 | 0.24038351 |
| CAMK2N1 | 891.204297 | 4 | 0 | 0.2768969 |
| KIAA2012 | 891.143015 | 4 | 0 | 0.71717999 |
| CRIM1 | 888.039697 | 4 | 0 | 0.35174562 |
| CDH20 | 883.167702 | 4 | 0 | 0.63686455 |
| PRKAR2B | 881.772424 | 4 | 0 | 0.15713256 |
| EPB41L2 | 881.140622 | 4 | 0 | 0.13627821 |
| PTPRF | 879.524813 | 4 | 0 | 0.5341145 |
| UBE3D | 879.466595 | 4 | 0 | 0.18644509 |
| SRPK1 | 878.011935 | 4 | 0 | 0.27054144 |
| RAB3C | 877.768738 | 4 | 0 | 0.44796694 |
| OGA | 877.629579 | 4 | 0 | 0.20935247 |
| JAZF1 | 875.9985 | 4 | 0 | 0.23475855 |
| IQCJ-SCHIP1 | 874.835722 | 4 | 0 | 0.16526126 |
| GDA | 874.69087 | 4 | 0 | 0.48720394 |
| NTRK2 | 873.509403 | 4 | 0 | 0.18837555 |
| VPS13D | 872.177635 | 4 | 0 | 0.13271552 |
| AC018697.1 | 871.744703 | 4 | 0 | 0.74347911 |
| QKI | 871.292729 | 4 | 0 | 0.27009948 |
| TENM2 | 868.380886 | 4 | 0 | 0.1100716 |
| RNF175 | 868.365596 | 4 | 0 | 0.33523468 |
| SLC25A29 | 868.130029 | 4 | 0 | 0.36731997 |
| RNF144A | 866.23281 | 4 | 0 | 0.27288365 |
| GRM5 | 864.933241 | 4 | 0 | 0.13451877 |
| GPM6A | 864.730554 | 4 | 0 | 0.17472651 |
| PTCHD1-AS | 863.987123 | 4 | 0 | 0.41840019 |
| TPK1 | 862.497778 | 4 | 0 | 0.20974219 |
| COBL | 857.820902 | 4 | 0 | 0.17875024 |
| KCTD8 | 852.275114 | 4 | 0 | 0.3601652 |
| ROCK2 | 851.949243 | 4 | 0 | 0.13355685 |

| | | | | |
|---|---|---|---|---|
| PRKDC | 851.890282 | 4 | 0 | 0.24099884 |
| KIAA1328 | 851.524793 | 4 | 0 | 0.20060919 |
| JAKMIP2 | 850.593306 | 4 | 0 | 0.11706066 |
| FAIM2 | 847.158096 | 4 | 0 | 0.2955701 |
| NEGR1 | 846.517296 | 4 | 0 | 0.14788628 |
| NR2F2-AS1 | 846.504459 | 4 | 0 | 1.19659269 |
| FHIT | 843.124455 | 4 | 0 | 0.2098433 |
| KALRN | 842.238236 | 4 | 0 | 0.09032029 |
| CPNE8 | 841.269674 | 4 | 0 | 0.44406934 |
| CTNNA3 | 841.200423 | 4 | 0 | 0.48897772 |
| TUBA1B | 832.359439 | 4 | 0 | 0.36168225 |
| TSPAN5 | 832.191971 | 4 | 0 | 0.21021845 |
| ATP11B | 832.154057 | 4 | 0 | 0.18861385 |
| CTNNAL1 | 831.177303 | 4 | 0 | 0.2705645 |
| PRKAG2 | 830.984909 | 4 | 0 | 0.2654344 |
| KLHL29 | 830.570216 | 4 | 0 | 0.28251772 |
| CAMK2A | 827.395236 | 4 | 0 | 0.22520715 |
| NCOA1 | 824.202464 | 4 | 0 | 0.15272455 |
| CHD9 | 819.996713 | 4 | 0 | 0.13259491 |
| PIAS1 | 817.846735 | 4 | 0 | 0.21011891 |
| ARPP19 | 816.329058 | 4 | 0 | 0.24545844 |
| AC092683.1 | 814.762432 | 4 | 0 | 0.45432136 |
| CCDC88A | 813.413113 | 4 | 0 | 0.21910383 |
| KCNMB4 | 812.88476 | 4 | 0 | 0.39799919 |
| MDGA2 | 810.764815 | 4 | 0 | 0.09608372 |
| ATP9B | 809.755268 | 4 | 0 | 0.15178215 |
| 11-Sep | 809.704643 | 4 | 0 | 0.24451401 |
| SRCIN1 | 808.824903 | 4 | 0 | 0.2616795 |
| EPB41L3 | 806.092202 | 4 | 0 | 0.14911661 |
| PARP8 | 802.285544 | 4 | 0 | 0.25367096 |
| PDE4DIP | 799.853753 | 4 | 0 | 0.13490503 |
| IMMP2L | 799.493263 | 4 | 0 | 0.17713924 |
| TSPAN7 | 796.501995 | 4 | 0 | 0.47987696 |
| WNK2 | 790.552067 | 4 | 0 | 0.23567728 |
| PCED1B | 790.394155 | 4 | 0 | 1.35437857 |
| TRPM3 | 789.58879 | 4 | 0 | 0.75280981 |
| HOMER1 | 789.266953 | 4 | 0 | 0.1918242 |
| TNIK | 786.964706 | 4 | 0 | 0.16770527 |
| MIR222HG | 786.630154 | 4 | 0 | 0.39151471 |
| PIP5K1B | 785.729839 | 4 | 0 | 0.50468187 |
| ANKRD6 | 784.951948 | 4 | 0 | 0.60333525 |
| KIF26B | 784.40728 | 4 | 0 | 0.37593451 |
| MYO1D | 783.063602 | 4 | 0 | 0.35118743 |

| | | | | |
|---|---|---|---|---|
| ITPR2 | 782.821743 | 4 | 0 | 0.88823295 |
| FKBP5 | 781.986962 | 4 | 0 | 1.47751182 |
| TRIM2 | 780.86259 | 4 | 0 | 0.18153595 |
| SLC39A11 | 779.953571 | 4 | 0 | 0.29518237 |
| CDKL1 | 779.730654 | 4 | 0 | 0.31143817 |
| TYW1B | 776.038188 | 4 | 0 | 0.41234554 |
| STIM1 | 775.548279 | 4 | 0 | 0.25806252 |
| CACNA1B | 774.467419 | 4 | 0 | 0.14026246 |
| ILDR2 | 773.982168 | 4 | 0 | 0.53891612 |
| APP | 773.617497 | 4 | 0 | 0.19547576 |
| ERC1 | 772.61158 | 4 | 0 | 0.11405763 |
| R3HDM1 | 771.841634 | 4 | 0 | 0.1616972 |
| SSH2 | 771.758608 | 4 | 0 | 0.19955067 |
| OSBPL10 | 771.67119 | 4 | 0 | 0.33902566 |
| ANO4 | 771.483162 | 4 | 0 | 0.99305327 |
| FAM153C | 767.826311 | 4 | 0 | 0.23827588 |
| MAP1B | 767.00956 | 4 | 0 | 0.14536498 |
| PRELID2 | 766.870249 | 4 | 0 | 0.39321497 |
| LPGAT1 | 765.909221 | 4 | 0 | 0.28222348 |
| PHACTR3 | 763.580955 | 4 | 0 | 0.23503615 |
| SH3BGRL | 763.170125 | 4 | 0 | 0.42788575 |
| SYNPO2 | 763.105698 | 4 | 0 | 0.42000839 |
| SNAP25 | 761.765283 | 4 | 0 | 0.1364817 |
| RHBDL3 | 758.998268 | 4 | 0 | 0.64512751 |
| SSBP2 | 758.874645 | 4 | 0 | 0.26253122 |
| PART1 | 758.59635 | 4 | 0 | 0.59470509 |
| FTX | 758.218674 | 4 | 0 | 0.13472294 |
| NOVA1 | 756.699387 | 4 | 0 | 0.25366952 |
| LONRF2 | 755.175192 | 4 | 0 | 0.12937054 |
| GLCE | 752.926908 | 4 | 0 | 0.40165409 |
| FGF14 | 752.372424 | 4 | 0 | 0.12645953 |
| NELL2 | 751.765172 | 4 | 0 | 0.17083233 |
| MEF2C | 750.728041 | 4 | 0 | 0.14911774 |
| STX1A | 748.568759 | 4 | 0 | 0.34091341 |
| MIR2052HG | 748.3655 | 4 | 0 | 0.67693303 |
| GPHN | 747.621615 | 4 | 0 | 0.14007659 |
| SAMD4A | 747.329241 | 4 | 0 | 0.5380629 |
| AL117329.1 | 746.117569 | 4 | 0 | 0.50886536 |
| BABAM2 | 744.312842 | 4 | 0 | 0.15973584 |
| E2F3 | 742.529932 | 4 | 0 | 0.2911007 |
| ZSWIM6 | 740.638587 | 4 | 0 | 0.22074583 |
| PRNP | 740.606677 | 4 | 0 | 0.32121387 |
| ZNF385D | 740.093073 | 4 | 0 | 0.39652584 |

| | | | | |
|---|---|---|---|---|
| MPDZ | 738.206955 | 4 | 0 | 0.23588499 |
| SLC44A1 | 737.473554 | 4 | 0 | 0.15986626 |
| SLCO3A1 | 735.184698 | 4 | 0 | 0.18828891 |
| DPY19L2 | 734.297538 | 4 | 0 | 0.22808246 |
| PPFIBP1 | 731.668902 | 4 | 0 | 0.3543878 |
| FOCAD | 727.463849 | 4 | 0 | 0.15906471 |
| STAMBPL1 | 727.316215 | 4 | 0 | 0.2110064 |
| FAM19A5 | 726.951586 | 4 | 0 | 0.31473136 |
| JUND | 726.859025 | 4 | 0 | 0.41570808 |
| KCNMA1 | 722.54817 | 4 | 0 | 0.09890615 |
| PHLPP1 | 718.826011 | 4 | 0 | 0.24816885 |
| HSP90AB1 | 716.177012 | 4 | 0 | 0.35030784 |
| SKAP2 | 716.058734 | 4 | 0 | 0.42180361 |
| ANKRD26 | 716.018706 | 4 | 0 | 0.11009767 |
| STOX2 | 711.897808 | 4 | 0 | 0.12378456 |
| NEK10 | 708.263994 | 4 | 0 | 0.1949594 |
| CPSF6 | 707.60844 | 4 | 0 | 0.29855331 |
| CALM1 | 705.958621 | 4 | 0 | 0.33873209 |
| PPARG | 703.61077 | 4 | 0 | 0.55488486 |
| PAK1 | 703.090954 | 4 | 0 | 0.19911887 |
| ATF7IP2 | 702.104125 | 4 | 0 | 0.27051499 |
| NMT2 | 701.49042 | 4 | 0 | 0.26469118 |
| RIMS1 | 700.87607 | 4 | 0 | 0.17426504 |
| ZBTB16 | 698.777446 | 4 | 0 | 0.22745163 |
| EPB41L4B | 697.650705 | 4 | 0 | 0.22558556 |
| CAMK2B | 697.557159 | 4 | 0 | 0.12656361 |
| FRMD4A | 691.249901 | 4 | 0 | 0.10541715 |
| CLU | 690.275172 | 4 | 0 | 0.23103969 |
| AC006148.1 | 690.093505 | 4 | 0 | 0.27412963 |
| NPTX1 | 689.820767 | 4 | 0 | 0.35867003 |
| LRRFIP1 | 688.107562 | 4 | 0 | 0.18093863 |
| CCNI | 687.622566 | 4 | 0 | 0.24007958 |
| DSCAML1 | 686.704823 | 4 | 0 | 0.49035377 |
| NAV1 | 682.485052 | 4 | 0 | 0.21699961 |
| CAMK2G | 679.31792 | 4 | 0 | 0.14995546 |
| NFIA | 678.657651 | 4 | 0 | 0.47996538 |
| PDK3 | 677.317477 | 4 | 0 | 0.35363266 |
| UNC13B | 677.246934 | 4 | 0 | 0.26391999 |
| LRRC28 | 677.024972 | 4 | 0 | 0.21290936 |
| SLC38A1 | 676.746199 | 4 | 0 | 0.26768999 |
| CDC40 | 675.551501 | 4 | 0 | 0.20132192 |
| KLHL4 | 675.018487 | 4 | 0 | 0.21187779 |
| ACTN4 | 674.266558 | 4 | 0 | 0.39141393 |

| | | | | |
|---|---|---|---|---|
| SESN3 | 671.659357 | 4 | 0 | 0.26562532 |
| COP1 | 671.513852 | 4 | 0 | 0.15611768 |
| CLASP1 | 668.03613 | 4 | 0 | 0.08307033 |
| FSTL4 | 666.557783 | 4 | 0 | 0.25541925 |
| AC024901.1 | 665.92875 | 4 | 0 | 0.47623563 |
| LRMDA | 664.44209 | 4 | 0 | 1.19791169 |
| LINC02552 | 664.151344 | 4 | 0 | 1.21940677 |
| PTMA | 664.002285 | 4 | 0 | 0.40710725 |
| INPP5F | 662.086178 | 4 | 0 | 0.25454038 |
| SYT14 | 659.552418 | 4 | 0 | 0.2605524 |
| ZEB2 | 659.377722 | 4 | 0 | 0.48437037 |
| HRH2 | 659.364712 | 4 | 0 | 0.40276217 |
| MAGI1 | 658.90526 | 4 | 0 | 0.11715145 |
| ATG7 | 658.475918 | 4 | 0 | 0.12377709 |
| RGL1 | 657.133648 | 4 | 0 | 0.23062417 |
| NRCAM | 657.092583 | 4 | 0 | 0.14523265 |
| CPNE5 | 656.896119 | 4 | 0 | 0.34969943 |
| ST8SIA5 | 655.961125 | 4 | 0 | 0.58409678 |
| CEP112 | 655.004188 | 4 | 0 | 0.16441994 |
| PLXDC2 | 654.816188 | 4 | 0 | 0.52460596 |
| PCDH11X | 654.385031 | 4 | 0 | 0.30168343 |
| NCAM1 | 652.783945 | 4 | 0 | 0.10293825 |
| ERICH3 | 652.369864 | 4 | 0 | 0.30564937 |
| UNC5C | 648.924735 | 4 | 0 | 0.27062259 |
| DLGAP4 | 648.77192 | 4 | 0 | 0.12412559 |
| ADAM23 | 647.382462 | 4 | 0 | 0.21020643 |
| PI4KA | 646.03653 | 4 | 0 | 0.12648741 |
| MARK1 | 645.88042 | 4 | 0 | 0.2035253 |
| LINC01830 | 644.914202 | 4 | 0 | 1.0348248 |
| VGLL4 | 644.724235 | 4 | 0 | 0.19836503 |
| SCN3A | 644.418218 | 4 | 0 | 0.3023102 |
| ME1 | 643.388568 | 4 | 0 | 0.27004163 |
| AL591501.1 | 642.617262 | 4 | 0 | 0.21741058 |
| MAN1C1 | 642.18041 | 4 | 0 | 0.18637743 |
| SLC25A27 | 641.856757 | 4 | 0 | 0.30789941 |
| CDK19 | 640.193071 | 4 | 0 | 0.12823654 |
| RASGRF1 | 638.496286 | 4 | 0 | 0.58375403 |
| TESK2 | 634.468561 | 4 | 0 | 0.71865615 |
| AC007091.1 | 633.810514 | 4 | 0 | 1.01067381 |
| ZNF483 | 633.375261 | 4 | 0 | 0.32151661 |
| PXYLP1 | 631.467267 | 4 | 0 | 0.30890703 |
| GABRA2 | 631.267449 | 4 | 0 | 0.21753804 |
| ACAP3 | 630.87311 | 4 | 0 | 0.18417176 |

| | | | | |
|---|---|---|---|---|
| KLF12 | 628.974948 | 4 | 0 | 0.77514066 |
| EPHA3 | 624.137653 | 4 | 0 | 0.37094793 |
| FOXN3 | 623.873875 | 4 | 0 | 0.30112856 |
| WASF1 | 623.564917 | 4 | 0 | 0.12338206 |
| GALNT17 | 623.081898 | 4 | 0 | 0.22729964 |
| GPR137C | 621.747669 | 4 | 0 | 0.37294112 |
| SLITRK5 | 619.653194 | 4 | 0 | 0.14731812 |
| LRFN2 | 619.335974 | 4 | 0 | 0.14687884 |
| TYRO3 | 617.76371 | 4 | 0 | 0.18919409 |
| GDAP1 | 617.713408 | 4 | 0 | 0.15307189 |
| AC138305.1 | 617.653105 | 4 | 0 | 0.70175049 |
| UBA6-AS1 | 616.900059 | 4 | 0 | 0.46040393 |
| RIMBP2 | 613.998298 | 4 | 0 | 0.13450802 |
| NPTXR | 612.429406 | 4 | 0 | 0.43484734 |
| CNTN6 | 610.826862 | 4 | 0 | 0.76182875 |
| ZFAND3 | 609.640599 | 4 | 0 | 0.09637919 |
| AGPAT4 | 609.542255 | 4 | 0 | 0.20139631 |
| SLC22A15 | 608.320711 | 4 | 0 | 0.2525411 |
| NOL4 | 608.061766 | 4 | 0 | 0.22717808 |
| FARS2 | 607.342902 | 4 | 0 | 0.18640589 |
| ARHGEF4 | 607.078034 | 4 | 0 | 0.3434117 |
| RPH3A | 606.012397 | 4 | 0 | 0.65023002 |
| LRRFIP2 | 605.451593 | 4 | 0 | 0.19807336 |
| AKAP6 | 603.230183 | 4 | 0 | 0.09004634 |
| CMSS1 | 601.566876 | 4 | 0 | 0.36348557 |
| WDFY2 | 600.872374 | 4 | 0 | 0.2160612 |
| ARHGEF7 | 600.050352 | 4 | 0 | 0.10828019 |
| LRRN1 | 599.803238 | 4 | 0 | 0.58418723 |
| RGS7BP | 596.936378 | 4 | 0 | 0.25613178 |
| AC060765.1 | 595.85793 | 4 | 0 | 1.54212281 |
| ABI1 | 593.30723 | 4 | 0 | 0.16040082 |
| HIVEP1 | 588.122567 | 4 | 0 | 0.37697236 |
| KCNQ5 | 586.787964 | 4 | 0 | 0.08297519 |
| DNM3 | 585.120773 | 4 | 0 | 0.22212554 |
| PDS5B | 585.097249 | 4 | 0 | 0.1963253 |
| KAT6B | 583.922031 | 4 | 0 | 0.18634937 |
| MT-CO3 | 583.89024 | 4 | 0 | 0.25290361 |
| ZFR | 583.884654 | 4 | 0 | 0.13549485 |
| ARHGAP32 | 583.541763 | 4 | 0 | 0.18430249 |
| ST8SIA3 | 582.38354 | 4 | 0 | 0.19893974 |
| MCF2L2 | 581.586019 | 4 | 0 | 0.19535979 |
| 7-Sep | 581.130348 | 4 | 0 | 0.24370183 |
| CENPC | 580.56882 | 4 | 0 | 0.27134938 |

| | | | | |
|---|---|---|---|---|
| SLC7A14 | 577.143473 | 4 | 0 | 0.50204788 |
| HNRNPLL | 576.839747 | 4 | 0 | 0.35386798 |
| RAP1GAP2 | 574.692289 | 4 | 0 | 0.17232267 |
| FAM13C | 574.362897 | 4 | 0 | 0.22492551 |
| WWOX | 573.580833 | 4 | 0 | 0.1412586 |
| HECW2 | 572.580367 | 4 | 0 | 0.16148177 |
| MAGI3 | 572.209265 | 4 | 0 | 0.15868631 |
| PLCL2 | 570.619439 | 4 | 0 | 0.15409706 |
| CADM2 | 569.799359 | 4 | 0 | 0.09596263 |
| TSPYL1 | 568.383163 | 4 | 0 | 0.35376749 |
| CERS4 | 568.286058 | 4 | 0 | 0.33514799 |
| GRIN2A | 568.067798 | 4 | 0 | 0.12960792 |
| EXOC6B | 566.980016 | 4 | 0 | 0.08142375 |
| MT-CO1 | 564.85517 | 4 | 0 | 0.29626296 |
| HECTD2 | 563.487936 | 4 | 0 | 0.26366522 |
| EXOC4 | 563.272361 | 4 | 0 | 0.14353541 |
| CAMK4 | 562.118298 | 4 | 0 | 0.26112012 |
| FYN | 562.073016 | 4 | 0 | 0.12076792 |
| OPA1 | 561.022836 | 4 | 0 | 0.18343621 |
| KIAA0930 | 560.165595 | 4 | 0 | 0.13229194 |
| TRIO | 559.957477 | 4 | 0 | 0.1183035 |
| XKR4 | 559.634948 | 4 | 0 | 0.27930909 |
| MYH10 | 557.486784 | 4 | 0 | 0.20506621 |
| MKLN1 | 553.568861 | 4 | 0 | 0.142167 |
| PTPRE | 552.372692 | 4 | 0 | 0.3291979 |
| HDAC4 | 551.39568 | 4 | 0 | 0.24317722 |
| OSBPL8 | 549.282178 | 4 | 0 | 0.13966075 |
| FRY | 548.352908 | 4 | 0 | 0.1290191 |
| COX7A2L | 548.055626 | 4 | 0 | 0.28826161 |
| CDKL5 | 547.29173 | 4 | 0 | 0.1833186 |
| PTPRM | 547.285641 | 4 | 0 | 0.37394949 |
| GALNT18 | 544.536434 | 4 | 0 | 0.17641963 |
| SLAIN1 | 541.364041 | 4 | 0 | 0.19832979 |
| AC079385.1 | 539.221266 | 4 | 0 | 0.37932047 |
| RAP1GAP | 538.427362 | 4 | 0 | 0.35908506 |
| LINC00894 | 536.496194 | 4 | 0 | 0.41468697 |
| TAOK1 | 536.18505 | 4 | 0 | 0.13784972 |
| BASP1 | 535.718019 | 4 | 0 | 0.22583898 |
| ENTPD1-AS1 | 535.056209 | 4 | 0 | 0.39491839 |
| PTH2R | 533.070889 | 4 | 0 | 0.52708637 |
| MXI1 | 532.468057 | 4 | 0 | 0.21387839 |
| MT-ND1 | 531.831318 | 4 | 0 | 0.31714839 |
| ENSA | 531.440579 | 4 | 0 | 0.29888104 |

| | | | | |
|---|---|---|---|---|
| C3orf67-AS1 | 530.563719 | 4 | 0 | 0.37362087 |
| DUBR | 530.536413 | 4 | 0 | 0.22908704 |
| FBXL2 | 527.370338 | 4 | 0 | 0.25441071 |
| SYNE1 | 526.268951 | 4 | 0 | 0.13356366 |
| SLC16A10 | 526.067233 | 4 | 0 | 0.42601192 |
| EPHA4 | 526.043409 | 4 | 0 | 0.12629128 |
| MKL2 | 525.689977 | 4 | 0 | 0.13588669 |
| AACS | 525.212548 | 4 | 0 | 0.30323761 |
| CADM1 | 525.045852 | 4 | 0 | 0.35825205 |
| IQCA1 | 524.60476 | 4 | 0 | 0.39917365 |
| HYDIN | 523.76272 | 4 | 0 | 0.13920338 |
| AFF1 | 521.961039 | 4 | 0 | 0.42830775 |
| NEK1 | 521.586223 | 4 | 0 | 0.29751122 |
| ST7 | 521.327028 | 4 | 0 | 0.15955255 |
| PHYHIP | 520.508751 | 4 | 0 | 0.26740187 |
| SV2B | 519.428324 | 4 | 0 | 0.10982153 |
| LINGO1 | 517.134108 | 4 | 0 | 0.12628701 |
| JAK2 | 516.649703 | 4 | 0 | 0.20106277 |
| LINC00923 | 516.331269 | 4 | 0 | 0.39044883 |
| CHL1 | 515.543223 | 4 | 0 | 0.14801728 |
| VWA8 | 514.352283 | 4 | 0 | 0.1075438 |
| GRB10 | 512.938105 | 4 | 0 | 0.20287962 |
| POLN | 512.36994 | 4 | 0 | 0.21537258 |
| CAMK1D | 510.748237 | 4 | 0 | 0.11918005 |
| KCNJ6 | 510.644195 | 4 | 0 | 0.23840012 |
| ETV6 | 510.196167 | 4 | 0 | 0.64936819 |
| DDX10 | 509.041648 | 4 | 0 | 0.16757068 |
| GPC1 | 508.959981 | 4 | 0 | 0.27974564 |
| CELF5 | 506.062658 | 4 | 0 | 0.16341131 |
| TIAM2 | 504.488229 | 4 | 0 | 0.18753315 |
| MBNL2 | 504.047281 | 4 | 0 | 0.16588859 |
| GPATCH8 | 503.933487 | 4 | 0 | 0.07563177 |
| ATP2B2 | 503.460588 | 4 | 0 | 0.20103308 |
| TLE1 | 501.60793 | 4 | 0 | 0.36206737 |
| TYW1 | 501.310587 | 4 | 0 | 0.17946309 |
| MICAL2 | 499.306862 | 4 | 0 | 0.12646864 |
| DTWD2 | 498.264397 | 4 | 0 | 0.28313414 |
| PGM2L1 | 497.739462 | 4 | 0 | 0.43334328 |
| GAB1 | 497.650249 | 4 | 0 | 0.12170218 |
| TENM4 | 497.135056 | 4 | 0 | 0.36130459 |
| SLC4A10 | 495.664274 | 4 | 0 | 0.14114731 |
| IDS | 493.361094 | 4 | 0 | 0.27789958 |
| APBB2 | 492.670284 | 4 | 0 | 0.10548686 |

| | | | | |
|---|---|---|---|---|
| LINC02398 | 491.865055 | 4 | 0 | 0.57824259 |
| NYAP2 | 490.982563 | 4 | 0 | 0.44170929 |
| DNAH6 | 489.322125 | 4 | 0 | 0.39792524 |
| GPATCH2 | 487.846125 | 4 | 0 | 0.27975606 |
| SCN8A | 483.569777 | 4 | 0 | 0.16616674 |
| ARID1B | 483.191306 | 4 | 0 | 0.15164423 |
| FGF12 | 481.077315 | 4 | 0 | 0.0936112 |
| VGF | 480.603256 | 4 | 0 | 0.61162815 |
| LRCH1 | 480.575558 | 4 | 0 | 0.44806164 |
| STX8 | 479.322298 | 4 | 0 | 0.13091534 |
| RPS6KA6 | 478.915765 | 4 | 0 | 0.15625124 |
| OLFM1 | 478.835124 | 4 | 0 | 0.24664548 |
| TOMM20 | 477.179485 | 4 | 0 | 0.35189715 |
| RECK | 476.386931 | 4 | 0 | 0.18500789 |
| TNPO1 | 475.324646 | 4 | 0 | 0.23750598 |
| PPP1R21 | 474.16305 | 4 | 0 | 0.27368133 |
| MLIP | 473.573551 | 4 | 0 | 0.19611549 |
| APLP1 | 473.398057 | 4 | 0 | 0.30833673 |
| MED13L | 472.749898 | 4 | 0 | 0.13702171 |
| MOB3B | 471.019082 | 4 | 0 | 0.35437919 |
| CUX1 | 470.997127 | 4 | 0 | 0.13325266 |
| BCL2 | 470.169471 | 4 | 0 | 0.29903702 |
| ZNF292 | 468.298583 | 4 | 0 | 0.11684285 |
| PCSK2 | 467.552689 | 4 | 0 | 0.28439587 |
| SNRPN | 466.956387 | 4 | 0 | 0.78671922 |
| NPTN | 464.905519 | 4 | 0 | 0.09631629 |
| AL139260.1 | 463.465941 | 4 | 0 | 0.20596458 |
| GRID1 | 463.049204 | 4 | 0 | 0.19115021 |
| ME3 | 462.341191 | 4 | 0 | 0.18354609 |
| TRIM9 | 462.198337 | 4 | 0 | 0.07312253 |
| PCSK1N | 460.772221 | 4 | 0 | 0.4266483 |
| TARBP1 | 460.559754 | 4 | 0 | 0.19463114 |
| CFAP299 | 459.435998 | 4 | 0 | 0.51131308 |
| PEBP1 | 458.265428 | 4 | 0 | 0.29428798 |
| MRPS30-DT | 458.207969 | 4 | 0 | 0.15549115 |
| MKNK1 | 458.095457 | 4 | 0 | 0.34335164 |
| SYN1 | 458.029305 | 4 | 0 | 0.3191028 |
| MT-ND2 | 457.962626 | 4 | 0 | 0.30208634 |
| LINC01252 | 456.63765 | 4 | 0 | 0.26685013 |
| LNX1 | 456.157998 | 4 | 0 | 0.14923294 |
| ATF2 | 455.337336 | 4 | 0 | 0.13068526 |
| RAPGEFL1 | 455.084475 | 4 | 0 | 0.20966878 |
| ITFG1 | 453.231802 | 4 | 0 | 0.090469 |

| | | | | |
|---|---|---|---|---|
| AGTPBP1 | 450.23031 | 4 | 0 | 0.11979868 |
| DLGAP1-AS4 | 449.77594 | 4 | 0 | 0.50445941 |
| AQP4-AS1 | 448.281564 | 4 | 0 | 0.29132577 |
| RNLS | 447.010246 | 4 | 0 | 0.1639509 |
| NRXN2 | 446.746458 | 4 | 0 | 0.14831821 |
| PCNX1 | 446.265507 | 4 | 0 | 0.1445641 |
| TCF12 | 445.991836 | 4 | 0 | 0.28973483 |
| SPEF2 | 444.744982 | 4 | 0 | 0.25411981 |
| ZBBX | 444.347489 | 4 | 0 | 0.47405307 |
| RPS6KA5 | 442.198271 | 4 | 0 | 0.15395808 |
| SYNDIG1 | 441.284506 | 4 | 0 | 0.19608558 |
| SERINC1 | 440.909961 | 4 | 0 | 0.36697703 |
| WAC | 440.431449 | 4 | 0 | 0.25960718 |
| GOLGA8A | 439.861907 | 4 | 0 | 0.21395679 |
| AC093523.1 | 439.753536 | 4 | 0 | 0.37856601 |
| FBXO34 | 439.085447 | 4 | 0 | 0.18278772 |
| ASAP1 | 438.57427 | 4 | 0 | 0.17765918 |
| EIF1 | 436.180086 | 4 | 0 | 0.38756762 |
| CCSER1 | 434.486891 | 4 | 0 | 0.14945312 |
| AL391807.1 | 434.205576 | 4 | 0 | 0.30927435 |
| RIMS2 | 434.102945 | 4 | 0 | 0.11966155 |
| MAST3 | 433.352944 | 4 | 0 | 0.11746363 |
| FAM153A | 432.503477 | 4 | 0 | 0.23332458 |
| NDRG4 | 431.939322 | 4 | 0 | 0.1527634 |
| MYO9A | 431.903121 | 4 | 0 | 0.10807121 |
| EFR3A | 431.546046 | 4 | 0 | 0.16714328 |
| PCDH19 | 430.352515 | 4 | 0 | 0.32194236 |
| ATXN7L1 | 430.247791 | 4 | 0 | 0.12962881 |
| ZNF721 | 429.702976 | 4 | 0 | 0.18244799 |
| MEF2A | 429.525411 | 4 | 0 | 0.16994101 |
| RFTN1 | 428.690493 | 4 | 0 | 0.22705699 |
| NAV3 | 428.21702 | 4 | 0 | 0.06899439 |
| TAPT1-AS1 | 425.308467 | 4 | 0 | 0.23560928 |
| SORT1 | 424.410922 | 4 | 0 | 0.09914705 |
| RUNX1T1 | 424.369768 | 4 | 0 | 0.24037271 |
| CTTNBP2 | 424.138802 | 4 | 0 | 0.11989891 |
| PITPNM2 | 423.534661 | 4 | 0 | 0.16104675 |
| SH3GL2 | 423.483997 | 4 | 0 | 0.16004175 |
| FUT9 | 423.276302 | 4 | 0 | 0.21434244 |
| CDH2 | 423.169662 | 4 | 0 | 0.23932764 |
| USP22 | 422.825573 | 4 | 0 | 0.24093756 |
| GAPDH | 422.625989 | 4 | 0 | 0.27335714 |
| PRMT8 | 421.761541 | 4 | 0 | 0.30206799 |

| | | | | |
|---|---|---|---|---|
| MEGF9 | 421.258334 | 4 | 0 | 0.25397352 |
| CNST | 420.639375 | 4 | 0 | 0.20178825 |
| MAP2K1 | 420.500028 | 4 | 0 | 0.17591936 |
| MFSD4A | 419.435916 | 4 | 0 | 0.23786509 |
| TMEM59L | 416.314042 | 4 | 0 | 0.16906355 |
| ZC3H8 | 415.904751 | 4 | 0 | 0.24377068 |
| CCDC85A | 415.632526 | 4 | 0 | 0.23993358 |
| GABRA3 | 414.77767 | 4 | 0 | 0.30654089 |
| AL662796.1 | 414.032645 | 4 | 0 | 0.34098316 |
| SYT11 | 413.592578 | 4 | 0 | 0.3207691 |
| MYO9B | 413.276969 | 4 | 0 | 0.20566981 |
| KCNC1 | 412.354827 | 4 | 0 | 0.20593669 |
| XKR6 | 412.304608 | 4 | 0 | 0.08226657 |
| MBP | 410.637281 | 4 | 0 | 0.55837767 |
| EFR3B | 409.636234 | 4 | 0 | 0.15554865 |
| MGAT5B | 409.251664 | 4 | 0 | 0.15587817 |
| PKIG | 409.111407 | 4 | 0 | 0.1095791 |
| TM7SF3 | 408.953271 | 4 | 0 | 0.15663447 |
| NWD2 | 408.566441 | 4 | 0 | 0.37240163 |
| LINC00632 | 408.546289 | 4 | 0 | 0.18780419 |
| BRINP1 | 405.971126 | 4 | 0 | 0.17407446 |
| AKT3 | 405.335724 | 4 | 0 | 0.09832989 |
| PACRG | 405.267233 | 4 | 0 | 0.1853092 |
| ACTB | 400.852591 | 4 | 0 | 0.22859481 |
| PIK3R1 | 400.184327 | 4 | 0 | 0.15245839 |
| GABRB1 | 397.000126 | 4 | 0 | 0.12346199 |
| PTPN4 | 396.514271 | 4 | 0 | 0.18242217 |
| MAP2K5 | 394.549593 | 4 | 0 | 0.13234021 |
| MSI2 | 394.42836 | 4 | 0 | 0.31827029 |
| ZBTB7C | 394.259868 | 4 | 0 | 0.60140974 |
| SAMD12 | 394.183335 | 4 | 0 | 0.13510234 |
| MPP7 | 393.468552 | 4 | 0 | 0.24478273 |
| CADM3 | 393.220656 | 4 | 0 | 0.18562019 |
| AC245297.3 | 392.268852 | 4 | 0 | 0.2126171 |
| CLIP2 | 391.54923 | 4 | 0 | 0.20101367 |
| PDE3B | 391.483094 | 4 | 0 | 0.2684258 |
| GPM6B | 388.536284 | 4 | 0 | 0.17497618 |
| FAM171A1 | 388.336776 | 4 | 0 | 0.13518114 |
| NFASC | 387.16992 | 4 | 0 | 0.12891845 |
| TSPYL2 | 385.690379 | 4 | 0 | 0.20007149 |
| MIR124-2HG | 384.850412 | 4 | 0 | 0.16227674 |
| ABCA10 | 384.344697 | 4 | 0 | 0.23433552 |
| NCALD | 382.295527 | 4 | 0 | 0.107312 |

| | | | | |
|---|---|---|---|---|
| FMN2 | 382.205509 | 4 | 0 | 0.12732626 |
| AP001347.1 | 380.8239 | 4 | 0 | 0.33589973 |
| SYP | 380.314822 | 4 | 0 | 0.19247993 |
| PKNOX2 | 380.24912 | 4 | 0 | 0.13746157 |
| UNC5A | 379.387398 | 4 | 0 | 0.14232061 |
| LIPA | 378.663232 | 4 | 0 | 0.27316067 |
| PEA15 | 378.421186 | 4 | 0 | 0.45016335 |
| BICDL1 | 378.325531 | 4 | 0 | 0.23211928 |
| SDCCAG8 | 377.708623 | 4 | 0 | 0.08463829 |
| GMDS | 377.477977 | 4 | 0 | 0.22920355 |
| AEBP2 | 377.114222 | 4 | 0 | 0.20301733 |
| FP236383.1 | 376.801144 | 4 | 0 | 0.21883015 |
| FP671120.1 | 375.06898 | 4 | 0 | 0.23533123 |
| LANCL2 | 373.721941 | 4 | 0 | 0.22372029 |
| VSTM2A | 372.937215 | 4 | 0 | 0.26501224 |
| PRKACB | 372.874064 | 4 | 0 | 0.12228085 |
| CASC4 | 372.203482 | 4 | 0 | 0.12553195 |
| BASP1-AS1 | 370.989326 | 4 | 0 | 0.21686075 |
| USP32 | 370.761829 | 4 | 0 | 0.17619209 |
| SDK1 | 370.28171 | 4 | 0 | 0.53392336 |
| TTLL5 | 370.196061 | 4 | 0 | 0.19622996 |
| ROCK1 | 370.011139 | 4 | 0 | 0.13534785 |
| SHISA9 | 369.735059 | 4 | 0 | 0.24934586 |
| AC008014.1 | 369.607838 | 4 | 0 | 0.35061272 |
| ACADVL | 369.58137 | 4 | 0 | 0.24443413 |
| NUCKS1 | 368.484783 | 4 | 0 | 0.2145975 |
| PEAK1 | 367.223296 | 4 | 0 | 0.098116 |
| CAPS2 | 366.757838 | 4 | 0 | 0.28034118 |
| PRB2 | 365.437746 | 4 | 0 | 0.1581067 |
| NR3C1 | 364.91307 | 4 | 0 | 0.26333361 |
| RNF165 | 364.383958 | 4 | 0 | 0.27300892 |
| SYBU | 363.8477 | 4 | 0 | 0.09552481 |
| PIK3CA | 362.600649 | 4 | 0 | 0.10696812 |
| SRRM2 | 362.370179 | 4 | 0 | 0.10992964 |
| SGIP1 | 361.822499 | 4 | 0 | 0.12083814 |
| PKD1 | 361.774711 | 4 | 0 | 0.11824769 |
| PWAR6 | 360.803073 | 4 | 0 | 0.20363829 |
| CORO6 | 358.204572 | 4 | 0 | 0.44529356 |
| ATP2C1 | 357.811229 | 4 | 0 | 0.18648724 |
| BAALC | 357.147159 | 4 | 0 | 0.30175082 |
| DENND1A | 356.746882 | 4 | 0 | 0.11022185 |
| TEAD1 | 356.676285 | 4 | 0 | 0.39482982 |
| RNF19A | 356.578433 | 4 | 0 | 0.19737745 |

| | | | | |
|---|---|---|---|---|
| PCDH15 | 356.34683 | 4 | 0 | 0.38154326 |
| CCDC91 | 355.962657 | 4 | 0 | 0.20422447 |
| IQSEC3 | 354.981564 | 4 | 0 | 0.11726104 |
| MIR29B2CH( | 354.324295 | 4 | 0 | 0.11807988 |
| ZFYVE9 | 354.073152 | 4 | 0 | 0.09170649 |
| REEP5 | 353.972092 | 4 | 0 | 0.28740968 |
| MT-CYB | 352.852872 | 4 | 0 | 0.24349498 |
| IQSEC1 | 352.801176 | 4 | 0 | 0.13299649 |
| GABRB3 | 352.415543 | 4 | 0 | 0.1809458 |
| HERC4 | 351.967067 | 4 | 0 | 0.1755738 |
| CNTN1 | 351.775661 | 4 | 0 | 0.09344244 |
| MT-ND3 | 350.459899 | 4 | 0 | 0.2178271 |
| FRMPD4 | 349.952675 | 4 | 0 | 0.10867886 |
| AC073050.1 | 349.43371 | 4 | 0 | 0.22328288 |
| MT-ATP6 | 347.800531 | 4 | 0 | 0.20923002 |
| MKL1 | 347.492445 | 4 | 0 | 0.11352971 |
| PREP | 346.457744 | 4 | 0 | 0.15967056 |
| ARF1 | 346.451602 | 4 | 0 | 0.31859499 |
| BTBD8 | 346.442629 | 4 | 0 | 0.19827002 |
| TRPM7 | 346.316517 | 4 | 0 | 0.10870163 |
| TLE4 | 345.592199 | 4 | 0 | 0.21244246 |
| MEG3 | 345.298434 | 4 | 0 | 0.27331059 |
| AHCYL2 | 344.53446 | 4 | 0 | 0.14673915 |
| SETBP1 | 344.48975 | 4 | 0 | 0.13678479 |
| CEP192 | 344.148019 | 4 | 0 | 0.09175319 |
| CCSER2 | 343.379822 | 4 | 0 | 0.16815106 |
| RABGAP1L | 342.688447 | 4 | 0 | 0.1927613 |
| NCKAP1 | 342.577965 | 4 | 0 | 0.08807162 |
| CALM3 | 342.039136 | 4 | 0 | 0.30181639 |
| PCNT | 341.021616 | 4 | 0 | 0.1550691 |
| AFF3 | 340.294668 | 4 | 0 | 0.1433298 |
| KIFC2 | 339.889729 | 4 | 0 | 0.2109386 |
| USP11 | 339.680324 | 4 | 0 | 0.34395432 |
| NME7 | 339.276467 | 4 | 0 | 0.14747971 |
| AKAP12 | 338.213802 | 4 | 0 | 0.32106302 |
| PSAP | 337.474196 | 4 | 0 | 0.22472368 |
| OSBPL1A | 335.856597 | 4 | 0 | 0.18362776 |
| AC124854.1 | 335.601907 | 4 | 0 | 0.37762474 |
| SNCB | 335.231057 | 4 | 0 | 0.2367123 |
| MBD5 | 335.069089 | 4 | 0 | 0.0973412 |
| MCHR2 | 335.057473 | 4 | 0 | 0.2800959 |
| HTR2A | 334.904758 | 4 | 0 | 0.26660847 |
| MTURN | 334.630632 | 4 | 0 | 0.20422831 |

| | | | | |
|---|---|---|---|---|
| TSGA10 | 334.054312 | 4 | 0 | 0.26744467 |
| SPATA6L | 333.825813 | 4 | 0 | 0.37430483 |
| TMSB4X | 333.19836 | 4 | 0 | 0.31273495 |
| EIF4G3 | 333.191798 | 4 | 0 | 0.10888319 |
| MT-ND5 | 332.977909 | 4 | 0 | 0.30818576 |
| ABCC5 | 332.518248 | 4 | 0 | 0.17171239 |
| PLXNA2 | 331.196276 | 4 | 0 | 0.10979152 |
| HLCS | 329.499987 | 4 | 0 | 0.20148776 |
| ENAH | 329.250192 | 4 | 0 | 0.29018824 |
| ACVR2A | 328.381242 | 4 | 0 | 0.34153841 |
| MT-CO2 | 328.359105 | 4 | 0 | 0.18897867 |
| CREB5 | 327.468104 | 4 | 0 | 0.29578193 |
| ASTN1 | 327.058199 | 4 | 0 | 0.11430312 |
| KIAA1324 | 326.827632 | 4 | 0 | 0.251661 |
| ARAP2 | 326.667158 | 4 | 0 | 0.17824505 |
| AC008591.1 | 326.259773 | 4 | 0 | 0.55566833 |
| COG6 | 325.930956 | 4 | 0 | 0.17361578 |
| DST | 325.680417 | 4 | 0 | 0.07840749 |
| CEP85L | 325.43529 | 4 | 0 | 0.14492914 |
| NRGN | 325.176971 | 4 | 0 | 0.26916296 |
| DIRC3 | 325.125115 | 4 | 0 | 0.23443461 |
| TMEM135 | 325.048597 | 4 | 0 | 0.15669955 |
| FAM81A | 324.895657 | 4 | 0 | 0.16837569 |
| AP3S1 | 324.722084 | 4 | 0 | 0.16987246 |
| ZNRF3 | 324.125411 | 4 | 0 | 0.1486796 |
| MAP3K4 | 323.355221 | 4 | 0 | 0.16437694 |
| YWHAB | 323.333046 | 4 | 0 | 0.33208725 |
| MT-ND4L | 322.533945 | 4 | 0 | 0.30246592 |
| MT-ND4 | 322.138766 | 4 | 0 | 0.22436637 |
| PLPPR5 | 321.515024 | 4 | 0 | 0.20838862 |
| IGF1R | 320.777168 | 4 | 0 | 0.2001824 |
| SIK3 | 320.62815 | 4 | 0 | 0.08589145 |
| GRK3 | 320.46663 | 4 | 0 | 0.23508726 |
| PNMA2 | 320.386602 | 4 | 0 | 0.24465958 |
| UNC80 | 319.984123 | 4 | 0 | 0.17125178 |
| NPAS2 | 319.203283 | 4 | 0 | 0.18737909 |
| YWHAG | 318.390595 | 4 | 0 | 0.20943251 |
| ARHGAP21 | 317.572045 | 4 | 0 | 0.08386847 |
| SYT1 | 317.047296 | 4 | 0 | 0.04272075 |
| STAT4 | 317.035742 | 4 | 0 | 0.45266091 |
| MAPK8 | 316.855375 | 4 | 0 | 0.13771489 |
| TACC1 | 316.279532 | 4 | 0 | 0.22564413 |
| APBA2 | 315.660107 | 4 | 0 | 0.09870326 |

| | | | | |
|---|---|---|---|---|
| PPIA | 314.750608 | 4 | 0 | 0.22368037 |
| CTIF | 314.136432 | 4 | 0 | 0.11075349 |
| ANKRD33B | 314.050537 | 4 | 0 | 0.14413698 |
| SCN3B | 313.794014 | 4 | 0 | 0.32573513 |
| ANKRD10 | 311.6289 | 4 | 0 | 0.18265143 |
| EIF4A2 | 311.502169 | 4 | 0 | 0.25018069 |
| WIPF3 | 311.493114 | 4 | 0 | 0.25871489 |
| LRRC49 | 311.137825 | 4 | 0 | 0.09816776 |
| ABI2 | 310.738955 | 4 | 0 | 0.10729627 |
| AC025159.1 | 310.306007 | 4 | 0 | 0.37270654 |
| 6-Mar | 310.209952 | 4 | 0 | 0.20232876 |
| PLCG2 | 310.181226 | 4 | 0 | 0.14413621 |
| TNFRSF25 | 310.060408 | 4 | 0 | 0.18921278 |
| TRAPPC9 | 309.28922 | 4 | 0 | 0.16337418 |
| ANKH | 308.788563 | 4 | 0 | 0.07765105 |
| AHI1 | 308.675493 | 4 | 0 | 0.15696797 |
| REPS2 | 307.944623 | 4 | 0 | 0.11814213 |
| ARHGEF3 | 307.901203 | 4 | 0 | 0.20154624 |
| CAMKK1 | 307.52573 | 4 | 0 | 0.16672112 |
| C11orf80 | 306.704201 | 4 | 0 | 0.17865744 |
| NORAD | 306.534367 | 4 | 0 | 0.31883853 |
| MAP4K5 | 306.425896 | 4 | 0 | 0.14662643 |
| OSBPL6 | 305.283019 | 4 | 0 | 0.06717183 |
| CXXC4-AS1 | 304.285207 | 4 | 0 | 0.16816501 |
| ANKRD11 | 304.045916 | 4 | 0 | 0.09243611 |
| MYT1L | 303.674378 | 4 | 0 | 0.06332471 |
| AC008537.1 | 303.104324 | 4 | 0 | 0.27136745 |
| ANAPC10 | 302.646132 | 4 | 0 | 0.19600994 |
| SLC12A2 | 301.953401 | 4 | 0 | 0.34237791 |
| CDYL2 | 301.75142 | 4 | 0 | 0.20024559 |
| RAPGEF2 | 301.504964 | 4 | 0 | 0.15216386 |
| LUZP1 | 301.389812 | 4 | 0 | 0.36591731 |
| HIVEP2 | 300.5319 | 4 | 0 | 0.11395344 |
| TSPAN3 | 299.839352 | 4 | 0 | 0.10076505 |
| C14orf132 | 298.820201 | 4 | 0 | 0.27208062 |
| PDE2A | 298.707095 | 4 | 0 | 0.21984644 |
| LINC01811 | 298.552035 | 4 | 0 | 0.45058127 |
| SYT7 | 296.305157 | 4 | 0 | 0.17766693 |
| EPHB1 | 295.626513 | 4 | 0 | 0.27784182 |
| FRMD5 | 295.616498 | 4 | 0 | 0.12816867 |
| ADAM22 | 294.791104 | 4 | 0 | 0.08179589 |
| SH3GL3 | 293.370864 | 4 | 0 | 0.22366248 |
| RNF130 | 292.979967 | 4 | 0 | 0.15246692 |

| | | | | |
|---|---|---|---|---|
| MNAT1 | 292.028383 | 4 | 0 | 0.13742905 |
| EHBP1 | 291.602844 | 4 | 0 | 0.12858637 |
| RAB11FIP3 | 291.288012 | 4 | 0 | 0.15996877 |
| LRRN2 | 290.789035 | 4 | 0 | 0.26019529 |
| CREG2 | 290.467002 | 4 | 0 | 0.20406569 |
| PURA | 289.869723 | 4 | 0 | 0.20745355 |
| CAMKMT | 289.755944 | 4 | 0 | 0.47541512 |
| LINC02223 | 288.812568 | 4 | 0 | 0.39519356 |
| AAK1 | 288.594567 | 4 | 0 | 0.0973358 |
| FNIP2 | 288.555508 | 4 | 0 | 0.11682493 |
| PPM1E | 288.353181 | 4 | 0 | 0.16750846 |
| SH3RF3 | 287.931121 | 4 | 0 | 0.17249443 |
| AKAP8L | 287.841682 | 4 | 0 | 0.11338614 |
| DOCK4 | 287.362824 | 4 | 0 | 0.16720977 |
| PAPSS1 | 286.282697 | 4 | 0 | 0.19026814 |
| NELL1 | 286.254518 | 4 | 0 | 0.15472252 |
| CHD6 | 285.905136 | 4 | 0 | 0.13615937 |
| KCNAB2 | 285.675352 | 4 | 0 | 0.17162329 |
| PAK5 | 284.434008 | 4 | 0 | 0.23667449 |
| ATP8A2 | 283.50049 | 4 | 0 | 0.17395931 |
| STRBP | 282.829296 | 4 | 0 | 0.22195276 |
| DIP2B | 282.631857 | 4 | 0 | 0.22007737 |
| SGK1 | 282.50183 | 4 | 0 | 0.34086599 |
| MAP4K4 | 282.402856 | 4 | 0 | 0.12577076 |
| GALNT1 | 282.081454 | 4 | 0 | 0.26186726 |
| ZDHHC17 | 281.516774 | 4 | 0 | 0.09486721 |
| ADD3 | 281.107194 | 4 | 0 | 0.19550183 |
| PKN2 | 280.703741 | 4 | 0 | 0.09121676 |
| KIAA1468 | 280.601641 | 4 | 0 | 0.12839911 |
| SRPK2 | 280.580096 | 4 | 0 | 0.12016088 |
| MAST1 | 280.241857 | 4 | 0 | 0.10839745 |
| AC073332.1 | 278.929801 | 4 | 0 | 0.53394963 |
| BBS9 | 278.855174 | 4 | 0 | 0.1091653 |
| TNR | 278.853386 | 4 | 0 | 0.16060071 |
| DGKI | 278.19448 | 4 | 0 | 0.09865014 |
| KIAA1549L | 277.090807 | 4 | 0 | 0.12588133 |
| MTRNR2L12 | 276.636406 | 4 | 0 | 0.26620568 |
| UNC13A | 275.944613 | 4 | 0 | 0.12400274 |
| CMTM4 | 275.360195 | 4 | 0 | 0.1272866 |
| UPF3A | 275.188282 | 4 | 0 | 0.15115628 |
| CKB | 274.898539 | 4 | 0 | 0.1218435 |
| SLC17A7 | 274.027702 | 4 | 0 | 0.27871135 |
| BCAS3 | 272.492857 | 4 | 0 | 0.12259481 |

| | | | | |
|---|---|---|---|---|
| AF117829.1 | 271.807755 | 4 | 0 | 0.3893428 |
| ATP1B1 | 271.550595 | 4 | 0 | 0.16134151 |
| LUC7L3 | 271.523578 | 4 | 0 | 0.21610118 |
| TBL1XR1 | 271.14027 | 4 | 0 | 0.15083046 |
| LYST | 270.770872 | 4 | 0 | 0.14403397 |
| LRRC7 | 270.180369 | 4 | 0 | 0.14786816 |
| VEZT | 269.282925 | 4 | 0 | 0.16271618 |
| NALCN | 269.08606 | 4 | 0 | 0.07610237 |
| CDC14B | 268.552843 | 4 | 0 | 0.34212978 |
| PABPN1 | 267.667355 | 4 | 0 | 0.13790656 |
| PCNX4 | 266.41357 | 4 | 0 | 0.1047331 |
| DTWD1 | 266.30649 | 4 | 0 | 0.11140324 |
| ICA1 | 265.951292 | 4 | 0 | 0.07505561 |
| MMP17 | 265.483945 | 4 | 0 | 0.39313309 |
| BTRC | 264.747615 | 4 | 0 | 0.06989504 |
| SRRM4 | 263.988759 | 4 | 0 | 0.12904664 |
| NMNAT2 | 262.77494 | 4 | 0 | 0.13838629 |
| LRRK2 | 262.700481 | 4 | 0 | 0.11489783 |
| ASCC3 | 262.627963 | 4 | 0 | 0.13538216 |
| ATRX | 262.592927 | 4 | 0 | 0.11388204 |
| HSD17B4 | 262.523149 | 4 | 0 | 0.158157 |
| SLC23A2 | 261.44175 | 4 | 0 | 0.14519515 |
| SNAP91 | 261.243817 | 4 | 0 | 0.12194012 |
| UBE4B | 260.83627 | 4 | 0 | 0.08770777 |
| RIF1 | 260.826319 | 4 | 0 | 0.17519405 |
| PBX1 | 260.799317 | 4 | 0 | 0.10223536 |
| ANKRD34C-A | 260.478495 | 4 | 0 | 0.12231961 |
| BICRAL | 259.450604 | 4 | 0 | 0.09218666 |
| FOXO3 | 259.40656 | 4 | 0 | 0.1926239 |
| R3HDM2 | 258.975646 | 4 | 0 | 0.13731143 |
| DPH6-DT | 258.930937 | 4 | 0 | 0.23486903 |
| COG5 | 258.88023 | 4 | 0 | 0.1914884 |
| AC239809.3 | 258.853879 | 4 | 0 | 0.12308094 |
| TUT4 | 258.852758 | 4 | 0 | 0.18284009 |
| INTS10 | 258.471748 | 4 | 0 | 0.14941279 |
| GAS2 | 257.938547 | 4 | 0 | 0.37275762 |
| ANKS1B | 257.639481 | 4 | 0 | 0.11154372 |
| PPP2R2C | 257.066893 | 4 | 0 | 0.11169071 |
| MAPRE2 | 256.907812 | 4 | 0 | 0.15598334 |
| LINC00461 | 256.569996 | 4 | 0 | 0.14595548 |
| OGT | 256.435442 | 4 | 0 | 0.25071612 |
| WNK1 | 256.311429 | 4 | 0 | 0.10785794 |
| MAP4K3 | 255.930399 | 4 | 0 | 0.19690624 |

| | | | | |
|---|---|---|---|---|
| CCNY | 255.823012 | 4 | 0 | 0.14913378 |
| TTC7B | 255.505684 | 4 | 0 | 0.14444849 |
| RIC8B | 255.490367 | 4 | 0 | 0.17364401 |
| GLCCI1 | 254.785681 | 4 | 0 | 0.22737282 |
| CPEB4 | 254.388987 | 4 | 0 | 0.08335988 |
| SPATS2 | 253.503291 | 4 | 0 | 0.16778652 |
| AC013652.1 | 253.417386 | 4 | 0 | 0.25785541 |
| CHCHD6 | 253.258506 | 4 | 0 | 0.22053231 |
| FMNL2 | 253.07258 | 4 | 0 | 0.16117916 |
| IQSEC2 | 252.582288 | 4 | 0 | 0.17983462 |
| BRWD1 | 252.123269 | 4 | 0 | 0.06682644 |
| THY1 | 251.849488 | 4 | 0 | 0.16057043 |
| GABRG2 | 251.408693 | 4 | 0 | 0.12191595 |
| TTC8 | 250.923557 | 4 | 0 | 0.19229002 |
| LINC00882 | 250.719055 | 4 | 0 | 0.06374735 |
| PPP6R2 | 249.81357 | 4 | 0 | 0.15267842 |
| MTERF1 | 249.524669 | 4 | 0 | 0.24715826 |
| ENO2 | 249.292953 | 4 | 0 | 0.1564161 |
| BSG | 249.208511 | 4 | 0 | 0.29027646 |
| KIF9-AS1 | 249.200186 | 4 | 0 | 0.20572789 |
| NUMB | 248.854784 | 4 | 0 | 0.18129111 |
| MTMR7 | 248.828486 | 4 | 0 | 0.17745988 |
| CALY | 248.474692 | 4 | 0 | 0.24314523 |
| SBF2 | 248.297694 | 4 | 0 | 0.11667719 |
| DHFR | 248.249085 | 4 | 0 | 0.1988958 |
| ACTR3B | 246.059067 | 4 | 0 | 0.09732473 |
| ZNF438 | 246.045694 | 4 | 0 | 0.23344901 |
| PPP2R5C | 246.037622 | 4 | 0 | 0.14382452 |
| SV2A | 245.47731 | 4 | 0 | 0.14960858 |
| YEATS2 | 244.473025 | 4 | 0 | 0.155331 |
| ATP8A1 | 244.410318 | 4 | 0 | 0.13528662 |
| ERCC6L2 | 242.798383 | 4 | 0 | 0.10669386 |
| SMARCA2 | 242.448844 | 4 | 0 | 0.16670219 |
| FCHO2 | 242.279755 | 4 | 0 | 0.21519989 |
| AMPH | 242.066805 | 4 | 0 | 0.11557613 |
| CD47 | 241.083517 | 4 | 0 | 0.23227915 |
| VPS13A | 241.073293 | 4 | 0 | 0.21259575 |
| TMSB10 | 240.733341 | 4 | 0 | 0.46765695 |
| ZBTB20 | 240.393436 | 4 | 0 | 0.19209111 |
| FAM69A | 240.233896 | 4 | 0 | 0.15774479 |
| LGI4 | 240.203505 | 4 | 0 | 0.21767743 |
| TMEM181 | 239.937468 | 4 | 0 | 0.14984535 |
| RASA1 | 239.791838 | 4 | 0 | 0.16197343 |

| | | | | |
|---|---|---|---|---|
| SPATA7 | 239.710353 | 4 | 0 | 0.21997585 |
| UBE2E2 | 239.655989 | 4 | 0 | 0.08688196 |
| BEX1 | 239.301609 | 4 | 0 | 0.31557829 |
| GLRB | 238.984033 | 4 | 0 | 0.08396421 |
| PTPN9 | 238.904337 | 4 | 0 | 0.24884577 |
| TSC22D1 | 238.7072 | 4 | 0 | 0.13374216 |
| HIVEP3 | 237.955843 | 4 | 0 | 0.1576594 |
| ARMC2 | 237.760578 | 4 | 0 | 0.14521441 |
| TBC1D32 | 237.285033 | 4 | 0 | 0.12240178 |
| FAM78B | 237.224711 | 4 | 0 | 0.18290844 |
| TRAF3 | 236.996768 | 4 | 0 | 0.25204641 |
| ADAMTSL1 | 236.742431 | 4 | 0 | 0.54610509 |
| RNF150 | 236.411181 | 4 | 0 | 0.12074043 |
| SVOP | 236.314786 | 4 | 0 | 0.17505897 |
| LARP1 | 236.275272 | 4 | 0 | 0.09609658 |
| HTT | 236.084398 | 4 | 0 | 0.07401401 |
| KCNH3 | 235.295531 | 4 | 0 | 0.17082663 |
| NELFA | 234.729103 | 4 | 0 | 0.20523345 |
| LINC02055 | 234.512805 | 4 | 0 | 0.85605124 |
| RBM6 | 234.344905 | 4 | 0 | 0.11294691 |
| PATJ | 234.137916 | 4 | 0 | 0.15144411 |
| ABCA3 | 233.242344 | 4 | 0 | 0.20922982 |
| AIG1 | 233.117726 | 4 | 0 | 0.13438306 |
| AC034195.1 | 233.108426 | 4 | 0 | 0.73124249 |
| SCFD2 | 232.869131 | 4 | 0 | 0.15550369 |
| PPM1H | 232.600325 | 4 | 0 | 0.142876 |
| FAM120A | 232.537662 | 4 | 0 | 0.15548468 |
| GALNT7 | 232.356081 | 4 | 0 | 0.1989259 |
| C16orf45 | 232.137467 | 4 | 0 | 0.14658641 |
| JAKMIP1 | 232.135135 | 4 | 0 | 0.1578334 |
| CAMKK2 | 232.039702 | 4 | 0 | 0.09043756 |
| SGCZ | 231.841355 | 4 | 0 | 0.7480693 |
| LINC01184 | 230.669556 | 4 | 0 | 0.21931227 |
| EXOC6 | 230.633787 | 4 | 0 | 0.15097879 |
| CDK5R1 | 230.590874 | 4 | 0 | 0.30990532 |
| PCNX2 | 230.46886 | 4 | 0 | 0.10691423 |
| MPPED2 | 230.414221 | 4 | 0 | 0.25714849 |
| MSH3 | 230.398503 | 4 | 0 | 0.14547374 |
| MBTPS1 | 230.346038 | 4 | 0 | 0.21799544 |
| TRIM37 | 229.001703 | 4 | 0 | 0.1111733 |
| CYTH3 | 228.394507 | 4 | 0 | 0.16643335 |
| TTBK2 | 228.137834 | 4 | 0 | 0.20165256 |
| SIPA1L3 | 227.388601 | 4 | 0 | 0.13921582 |

| | | | | |
|---|---|---|---|---|
| CAMKV | 227.222292 | 4 | 0 | 0.20965526 |
| MTUS2 | 227.208484 | 4 | 0 | 0.14737342 |
| RSRP1 | 227.168575 | 4 | 0 | 0.14529263 |
| ADD2 | 223.859178 | 4 | 0 | 0.14239513 |
| TRAF3IP2-AS | 223.457166 | 4 | 0 | 0.08869732 |
| KLHL7 | 223.336953 | 4 | 0 | 0.08766926 |
| ARID5B | 222.683595 | 4 | 0 | 0.12949331 |
| BCKDHB | 222.475115 | 4 | 0 | 0.23007144 |
| MED27 | 221.90192 | 4 | 0 | 0.153676 |
| ATP2A2 | 220.757388 | 4 | 0 | 0.09055059 |
| ADGRV1 | 220.320653 | 4 | 0 | 0.21721311 |
| SERINC3 | 220.266947 | 4 | 0 | 0.20614253 |
| SECISBP2L | 220.24703 | 4 | 0 | 0.21192561 |
| REV1 | 218.753144 | 4 | 0 | 0.16743996 |
| GNPTAB | 218.584062 | 4 | 0 | 0.11233294 |
| CUL3 | 218.31376 | 4 | 0 | 0.15547062 |
| ATR | 218.202117 | 4 | 0 | 0.15233087 |
| DDX24 | 218.153484 | 4 | 0 | 0.14809902 |
| ITM2B | 218.018086 | 4 | 0 | 0.1882916 |
| CACNG2 | 218.008998 | 4 | 0 | 0.09139048 |
| MAP3K13 | 217.068359 | 4 | 0 | 0.07612731 |
| CDC42SE2 | 216.822458 | 4 | 0 | 0.13851993 |
| SREBF2 | 216.548414 | 4 | 0 | 0.18502693 |
| CCDC171 | 216.116969 | 4 | 0 | 0.13009837 |
| SHC3 | 214.786057 | 4 | 0 | 0.20786332 |
| DNAH7 | 214.425212 | 4 | 0 | 0.20648659 |
| KMT2C | 214.269652 | 4 | 0 | 0.05592883 |
| USP47 | 214.189151 | 4 | 0 | 0.07542029 |
| SUPT3H | 213.48038 | 4 | 0 | 0.15169943 |
| YPEL3 | 213.369015 | 4 | 0 | 0.14976797 |
| HSP90AA1 | 213.126542 | 4 | 0 | 0.23390029 |
| SRRM3 | 211.744681 | 4 | 0 | 0.16485052 |
| UBAC2 | 211.619703 | 4 | 0 | 0.15220266 |
| ZC3H14 | 211.524532 | 4 | 0 | 0.153018 |
| PACS1 | 211.510839 | 4 | 0 | 0.14392307 |
| FAM126B | 211.02774 | 4 | 0 | 0.1013238 |
| SEC62 | 209.929973 | 4 | 0 | 0.27414325 |
| OXCT1 | 209.868823 | 4 | 0 | 0.1461087 |
| CRADD | 209.808725 | 4 | 0 | 0.19450417 |
| LINC00970 | 209.535326 | 4 | 0 | 0.16462426 |
| NSG2 | 209.292108 | 4 | 0 | 0.3157922 |
| PPM1L | 208.996058 | 4 | 0 | 0.07032375 |
| FRRS1L | 208.723378 | 4 | 0 | 0.14504913 |

| | | | | |
|---|---|---|---|---|
| ACTG1 | 208.239034 | 4 | 0 | 0.38686255 |
| SMG6 | 208.228814 | 4 | 0 | 0.09002166 |
| ARHGEF11 | 207.739452 | 4 | 0 | 0.1752621 |
| LINC01505 | 207.57768 | 4 | 0 | 0.47371335 |
| GABPB1-AS1 | 207.310607 | 4 | 0 | 0.22048449 |
| SLC8A2 | 207.028727 | 4 | 0 | 0.28044254 |
| REPS1 | 206.826902 | 4 | 0 | 0.15205559 |
| KCNT1 | 206.556769 | 4 | 0 | 0.30298786 |
| PRKAA2 | 206.109498 | 4 | 0 | 0.16234491 |
| TTLL11 | 206.102965 | 4 | 0 | 0.09777411 |
| GNAQ | 205.721753 | 4 | 0 | 0.09645314 |
| MPRIP | 205.52444 | 4 | 0 | 0.08331483 |
| RASA2 | 205.09256 | 4 | 0 | 0.16603029 |
| GALNT11 | 204.407296 | 4 | 0 | 0.12565623 |
| SH3BGRL2 | 203.965927 | 4 | 0 | 0.13692278 |
| LYRM4 | 203.749186 | 4 | 0 | 0.21789316 |
| UMAD1 | 203.579228 | 4 | 0 | 0.10408958 |
| NUBPL | 203.188761 | 4 | 0 | 0.2359365 |
| BCAP29 | 203.093621 | 4 | 0 | 0.21459581 |
| PTEN | 203.090347 | 4 | 0 | 0.11030043 |
| KLC1 | 203.084643 | 4 | 0 | 0.24736137 |
| NFIX | 202.834554 | 4 | 0 | 0.23267333 |
| YWHAZ | 202.742608 | 4 | 0 | 0.13733509 |
| EXT1 | 202.698613 | 4 | 0 | 0.10389774 |
| SNX29 | 202.410172 | 4 | 0 | 0.35462485 |
| AC096711.2 | 201.564335 | 4 | 0 | 0.1469209 |
| SFXN1 | 201.239474 | 4 | 0 | 0.10359851 |
| PHTF1 | 201.112293 | 4 | 0 | 0.15074607 |
| CHST11 | 200.458319 | 4 | 0 | 0.25988184 |
| HMGCLL1 | 199.515336 | 4 | 0 | 0.13500739 |
| SLC22A23 | 199.401078 | 4 | 0 | 0.12876388 |
| CCK | 199.381935 | 4 | 0 | 0.17539626 |
| FZD3 | 199.096872 | 4 | 0 | 0.16861995 |
| ELOVL6 | 199.034973 | 4 | 0 | 0.16732962 |
| MIAT | 198.731089 | 4 | 0 | 0.07418901 |
| SIRPA | 198.676759 | 4 | 0 | 0.15664969 |
| SOS1 | 198.418036 | 4 | 0 | 0.14438112 |
| NIM1K | 197.842837 | 4 | 0 | 0.12033471 |
| GRIN1 | 197.082783 | 4 | 0 | 0.12700022 |
| TMBIM6 | 196.752001 | 4 | 0 | 0.21189608 |
| CATSPER2 | 196.473416 | 4 | 0 | 0.08583365 |
| LMO7 | 196.367956 | 4 | 0 | 0.17677907 |
| TMEM161B-/ | 196.303729 | 4 | 0 | 0.10378397 |

| | | | | |
|---|---|---|---|---|
| SPRED1 | 196.086379 | 4 | 0 | 0.14429442 |
| NR6A1 | 196.011035 | 4 | 0 | 0.14282538 |
| CFL1 | 195.939087 | 4 | 0 | 0.30753071 |
| DZIP3 | 195.721873 | 4 | 0 | 0.12687341 |
| SGSM1 | 195.685892 | 4 | 0 | 0.13886728 |
| BBX | 195.585272 | 4 | 0 | 0.13427742 |
| RB1 | 195.429803 | 4 | 0 | 0.1828056 |
| G3BP2 | 195.123938 | 4 | 0 | 0.16448982 |
| FER | 195.046772 | 4 | 0 | 0.09969117 |
| TTC21B | 193.703089 | 4 | 0 | 0.0940459 |
| PPA2 | 193.524846 | 4 | 0 | 0.08183893 |
| TFDP2 | 193.291976 | 4 | 0 | 0.18511179 |
| NLGN4X | 192.275168 | 4 | 0 | 0.10117639 |
| FAM49B | 192.067779 | 4 | 0 | 0.14416441 |
| UNC79 | 191.834947 | 4 | 0 | 0.07120281 |
| CLIP3 | 190.521268 | 4 | 0 | 0.2152924 |
| CHCHD3 | 190.116395 | 4 | 0 | 0.06705895 |
| AC008691.1 | 190.099128 | 4 | 0 | 0.56469176 |
| LUZP2 | 190.010382 | 4 | 0 | 0.28640636 |
| SELENOW | 189.976235 | 4 | 0 | 0.23560863 |
| METTL15 | 189.810279 | 4 | 0 | 0.11831951 |
| ATP1B3 | 189.661524 | 4 | 0 | 0.215991 |
| XPR1 | 189.426655 | 4 | 0 | 0.0759231 |
| RALGPS1 | 189.381852 | 4 | 0 | 0.12314059 |
| PKM | 189.228534 | 4 | 0 | 0.16960821 |
| AC105402.3 | 188.766086 | 4 | 0 | 0.22001733 |
| ERBB4 | 188.425948 | 4 | 0 | 0.1597817 |
| SLC39A10 | 188.285518 | 4 | 0 | 0.2259615 |
| NAALADL2 | 187.481404 | 4 | 0 | 0.33977275 |
| DENND5B | 186.18742 | 4 | 0 | 0.11666686 |
| ATP6V1B2 | 185.042175 | 4 | 0 | 0.16396307 |
| ERBIN | 184.82887 | 4 | 0 | 0.20578412 |
| ZNF518A | 184.64946 | 4 | 0 | 0.18343513 |
| MAP1A | 184.544784 | 4 | 0 | 0.21459649 |
| SH3KBP1 | 184.495548 | 4 | 0 | 0.11799797 |
| PDXK | 184.463801 | 4 | 0 | 0.09068295 |
| NISCH | 184.060726 | 4 | 0 | 0.17324097 |
| UVRAG | 183.893558 | 4 | 0 | 0.12808326 |
| PRKAR1B | 183.703393 | 4 | 0 | 0.12382947 |
| KIAA0825 | 183.351382 | 4 | 0 | 0.0813654 |
| SLC16A7 | 182.558148 | 4 | 0 | 0.09597863 |
| SPTBN4 | 181.638113 | 4 | 0 | 0.08915787 |
| AGFG1 | 180.938871 | 4 | 0 | 0.14573162 |

| | | | | |
|---|---|---|---|---|
| ANO10 | 180.815286 | 4 | 0 | 0.18508451 |
| MEF2C-AS1 | 180.386742 | 4 | 0 | 0.19131709 |
| CDON | 180.247312 | 4 | 0 | 0.25033905 |
| CLVS2 | 180.104189 | 4 | 0 | 0.1545994 |
| MCF2L | 180.050859 | 4 | 0 | 0.08905081 |
| PKIA | 179.86967 | 4 | 0 | 0.11882145 |
| TTC14 | 179.482378 | 4 | 0 | 0.11333179 |
| NSD2 | 178.939785 | 4 | 0 | 0.13994647 |
| PJA2 | 178.841814 | 4 | 0 | 0.147908 |
| SH3PXD2A | 178.76848 | 4 | 0 | 0.14230374 |
| PREPL | 178.637686 | 4 | 0 | 0.11422635 |
| FAM172A | 178.382535 | 4 | 0 | 0.09665029 |
| TRIM33 | 178.292055 | 4 | 0 | 0.09130748 |
| ELMOD1 | 178.191596 | 4 | 0 | 0.07179054 |
| PPARGC1A | 178.064137 | 4 | 0 | 0.29698594 |
| CACNG8 | 177.613212 | 4 | 0 | 0.14417384 |
| SNED1 | 176.698145 | 4 | 0 | 0.16529686 |
| CDR2 | 176.217918 | 4 | 0 | 0.23220483 |
| SCAI | 175.712793 | 4 | 0 | 0.11118808 |
| AKAP11 | 175.662657 | 4 | 0 | 0.13592329 |
| PCMTD1 | 175.541125 | 4 | 0 | 0.10027991 |
| RNPC3 | 175.293466 | 4 | 0 | 0.20165646 |
| DLGAP3 | 175.214354 | 4 | 0 | 0.12684443 |
| KIF5C | 175.103908 | 4 | 0 | 0.07142709 |
| MROH1 | 174.994108 | 4 | 0 | 0.17647488 |
| C1orf21 | 174.495408 | 4 | 0 | 0.12249514 |
| STAG1 | 174.479021 | 4 | 0 | 0.12465894 |
| ADCY9 | 174.38623 | 4 | 0 | 0.17706835 |
| ULK2 | 173.837496 | 4 | 0 | 0.13227068 |
| TNKS | 173.650424 | 4 | 0 | 0.10848576 |
| RSPO2 | 173.524072 | 4 | 0 | 0.38582627 |
| ABR | 173.107808 | 4 | 0 | 0.07192309 |
| SATB2 | 172.221205 | 4 | 0 | 0.18635764 |
| ANKS1A | 172.164696 | 4 | 0 | 0.25927698 |
| CASK | 171.526518 | 4 | 0 | 0.13706129 |
| CRELD1 | 171.519501 | 4 | 0 | 0.08758262 |
| CHD2 | 170.555046 | 4 | 0 | 0.16009105 |
| DOCK7 | 170.380565 | 4 | 0 | 0.16498205 |
| NNT | 170.339624 | 4 | 0 | 0.12429501 |
| KCNB1 | 170.075923 | 4 | 0 | 0.10094015 |
| MPHOSPH8 | 169.938779 | 4 | 0 | 0.09061633 |
| HNRNPU | 169.484523 | 4 | 0 | 0.11013026 |
| ZDHHC11B | 169.44822 | 4 | 0 | 0.17039729 |

| | | | | |
|---|---|---|---|---|
| HGSNAT | 169.131655 | 4 | 0 | 0.12930593 |
| GRAMD1B | 169.087533 | 4 | 0 | 0.07993445 |
| GPR155 | 168.797622 | 4 | 0 | 0.12773026 |
| PPP1R9A | 168.154484 | 4 | 0 | 0.1429964 |
| ZNF365 | 168.106954 | 4 | 0 | 0.10944297 |
| TCF20 | 168.079287 | 4 | 0 | 0.15341241 |
| TPST1 | 168.015007 | 4 | 0 | 0.17343327 |
| REEP1 | 167.681056 | 4 | 0 | 0.1096628 |
| MTX2 | 167.126423 | 4 | 0 | 0.1262186 |
| NCOA2 | 166.634855 | 4 | 0 | 0.08656429 |
| PNISR | 166.293377 | 4 | 0 | 0.10472584 |
| ARFGEF2 | 166.21005 | 4 | 0 | 0.14603513 |
| NCEH1 | 165.929925 | 4 | 0 | 0.11852528 |
| SLC25A36 | 165.854309 | 4 | 0 | 0.20443351 |
| SLC38A6 | 164.937854 | 4 | 0 | 0.20283574 |
| SNX32 | 164.912771 | 4 | 0 | 0.12637008 |
| ARPP21 | 164.811095 | 4 | 0 | 0.08005108 |
| CLSTN3 | 164.080547 | 4 | 0 | 0.10633837 |
| BIN1 | 163.589863 | 4 | 0 | 0.13335877 |
| GTDC1 | 163.584515 | 4 | 0 | 0.14781586 |
| ATG5 | 163.407066 | 4 | 0 | 0.16527159 |
| SACS | 162.881377 | 4 | 0 | 0.11294242 |
| MAN1A2 | 162.875003 | 4 | 0 | 0.06076676 |
| NDUFS1 | 162.842679 | 4 | 0 | 0.13224588 |
| SORL1 | 162.639894 | 4 | 0 | 0.08155399 |
| MGAT5 | 161.919393 | 4 | 0 | 0.14854862 |
| PHTF2 | 161.91612 | 4 | 0 | 0.09546314 |
| ZNF407 | 161.852782 | 4 | 0 | 0.16177724 |
| HIC2 | 161.659363 | 4 | 0 | 0.13265347 |
| SNCA | 161.159269 | 4 | 0 | 0.17803457 |
| UGGT2 | 161.126349 | 4 | 0 | 0.05838155 |
| GFOD1 | 161.077208 | 4 | 0 | 0.05688536 |
| MIR646HG | 161.07038 | 4 | 0 | 0.36597582 |
| DDX17 | 161.067646 | 4 | 0 | 0.11077691 |
| OPTN | 160.605944 | 4 | 0 | 0.17652651 |
| FRYL | 160.40457 | 4 | 0 | 0.15080588 |
| SLC16A1-AS1 | 160.303093 | 4 | 0 | 0.13933168 |
| PLEKHA6 | 159.96251 | 4 | 0 | 0.11419206 |
| MAP3K7 | 159.758241 | 4 | 0 | 0.11937414 |
| CSMD2 | 159.306968 | 4 | 0 | 0.12045359 |
| PRR16 | 159.286626 | 4 | 0 | 0.14303308 |
| ECPAS | 159.228321 | 4 | 0 | 0.0982297 |
| FAM222B | 158.932576 | 4 | 0 | 0.15617563 |

| | | | | |
|---|---|---|---|---|
| BAZ2B | 158.636645 | 4 | 0 | 0.11115202 |
| SH3D19 | 158.502279 | 4 | 0 | 0.09611878 |
| ABHD18 | 158.450174 | 4 | 0 | 0.12558302 |
| PRKCB | 158.409673 | 4 | 0 | 0.05618568 |
| FGFR1OP2 | 158.002142 | 4 | 0 | 0.08688695 |
| FAM66D | 157.773636 | 4 | 0 | 0.13246289 |
| NSF | 157.344136 | 4 | 0 | 0.08460212 |
| RBM26 | 157.331612 | 4 | 0 | 0.06880805 |
| DLG1 | 157.314008 | 4 | 0 | 0.1356947 |
| BEX3 | 156.761466 | 4 | 0 | 0.20238624 |
| HSP90B1 | 156.658589 | 4 | 0 | 0.23219414 |
| FXYD7 | 156.573519 | 4 | 0 | 0.1795394 |
| PPP1R12B | 156.092008 | 4 | 0 | 0.13137959 |
| KDM6A | 155.718858 | 4 | 0 | 0.16522977 |
| NDFIP2 | 155.368474 | 4 | 0 | 0.20582948 |
| PRICKLE1 | 155.18196 | 4 | 0 | 0.07314232 |
| RPS6KA3 | 155.043782 | 4 | 0 | 0.15849497 |
| FBN1 | 155.007175 | 4 | 0 | 0.13491621 |
| ISPD | 154.926992 | 4 | 0 | 0.11747418 |
| GPI | 154.843526 | 4 | 0 | 0.16452715 |
| MTMR12 | 154.560876 | 4 | 0 | 0.13484306 |
| TNRC6C | 154.502127 | 4 | 0 | 0.08139888 |
| TCAF1 | 153.776583 | 4 | 0 | 0.11152151 |
| BRAF | 153.479867 | 4 | 0 | 0.06971414 |
| LMTK2 | 153.258218 | 4 | 0 | 0.14309958 |
| TTC37 | 153.118915 | 4 | 0 | 0.12178088 |
| AP2M1 | 152.983256 | 4 | 0 | 0.21653631 |
| SIL1 | 152.37669 | 4 | 0 | 0.13746478 |
| NLGN4Y | 152.296954 | 4 | 0 | 0.15701909 |
| AGAP3 | 151.758429 | 4 | 0 | 0.06800198 |
| FBXO16 | 151.705408 | 4 | 0 | 0.14591192 |
| CA11 | 151.551395 | 4 | 0 | 0.13418894 |
| CSNK1A1 | 151.031917 | 4 | 0 | 0.10609345 |
| SMG1 | 150.856003 | 4 | 0 | 0.11768168 |
| GABRB2 | 150.641969 | 4 | 0 | 0.06400776 |
| NLN | 150.240111 | 4 | 0 | 0.1642958 |
| MAPK14 | 150.127307 | 4 | 0 | 0.07501708 |
| LDB2 | 150.126188 | 4 | 0 | 0.08166316 |
| TUBB4A | 150.086064 | 4 | 0 | 0.2753724 |
| UBR3 | 149.949338 | 4 | 0 | 0.07420753 |
| FAM182B | 149.737253 | 4 | 0 | 0.08241357 |
| ARHGAP5 | 149.584205 | 4 | 0 | 0.08217659 |
| MTCH1 | 149.474441 | 4 | 0 | 0.31053965 |

| | | | | |
|---|---|---|---|---|
| GUCY1B1 | 149.089793 | 4 | 0 | 0.25311749 |
| PIK3CB | 148.690904 | 4 | 0 | 0.08832302 |
| PTPN2 | 148.56586 | 4 | 0 | 0.10555438 |
| RBM25 | 148.436403 | 4 | 0 | 0.0856401 |
| PHIP | 148.264425 | 4 | 0 | 0.16930354 |
| SFXN5 | 148.011901 | 4 | 0 | 0.08857688 |
| NBEAL1 | 147.856104 | 4 | 0 | 0.09371982 |
| LIPE-AS1 | 147.650688 | 4 | 0 | 0.1239524 |
| AC087280.2 | 147.436291 | 4 | 0 | 0.39626092 |
| 4-Mar | 147.071578 | 4 | 0 | 0.21982175 |
| SYT16 | 147.048223 | 4 | 0 | 0.0893864 |
| SARAF | 146.479749 | 4 | 0 | 0.25833591 |
| FTL | 146.295636 | 4 | 0 | 0.29412758 |
| C20orf194 | 145.774717 | 4 | 0 | 0.06973914 |
| C9orf3 | 145.635705 | 4 | 0 | 0.10670783 |
| GABRA4 | 145.599657 | 4 | 0 | 0.14875175 |
| ZNF532 | 145.501313 | 4 | 0 | 0.09179425 |
| PTBP2 | 145.321776 | 4 | 0 | 0.13897158 |
| CLIP1 | 144.788678 | 4 | 0 | 0.1187107 |
| DDHD1 | 144.739037 | 4 | 0 | 0.0937389 |
| CAP2 | 144.61867 | 4 | 0 | 0.12790978 |
| SCRN1 | 144.565254 | 4 | 0 | 0.09508841 |
| MVB12B | 144.486682 | 4 | 0 | 0.13989779 |
| ICE1 | 144.460816 | 4 | 0 | 0.13743065 |
| AMMECR1 | 144.295841 | 4 | 0 | 0.14743588 |
| SCN2A | 144.139283 | 4 | 0 | 0.09166817 |
| PHKB | 143.710525 | 4 | 0 | 0.20234222 |
| MYSM1 | 143.458569 | 4 | 0 | 0.10258513 |
| N4BP2L1 | 142.919706 | 4 | 0 | 0.20943151 |
| AFDN | 142.816476 | 4 | 0 | 0.14614017 |
| PHF21A | 142.672272 | 4 | 0 | 0.15816886 |
| EXOC2 | 142.520806 | 4 | 0 | 0.12768016 |
| MAP6 | 142.246382 | 4 | 0 | 0.1398138 |
| KLHL3 | 142.041643 | 4 | 0 | 0.11015629 |
| ADARB1 | 141.878027 | 4 | 0 | 0.23684275 |
| SLC4A8 | 141.726514 | 4 | 0 | 0.23964746 |
| CCDC92 | 140.848939 | 4 | 0 | 0.09418938 |
| BTAF1 | 140.328233 | 4 | 0 | 0.14247654 |
| TNK2 | 139.822085 | 4 | 0 | 0.18300836 |
| SCFD1 | 139.300499 | 4 | 0 | 0.11709573 |
| EPB41L1 | 138.765815 | 4 | 0 | 0.07143623 |
| FAM171B | 138.265385 | 4 | 0 | 0.21441429 |
| MSH2 | 138.206755 | 4 | 0 | 0.1764249 |

| | | | | |
|---|---|---|---|---|
| PRKCZ | 138.188143 | 4 | 0 | 0.079785 |
| ARID4A | 138.180914 | 4 | 0 | 0.14151881 |
| UBE2O | 137.667735 | 4 | 0 | 0.1927701 |
| FBXL17 | 137.592989 | 4 | 0 | 0.08215216 |
| RNF24 | 137.30448 | 4 | 0 | 0.12279598 |
| CNNM1 | 137.025959 | 4 | 0 | 0.08092706 |
| DMTF1 | 136.996166 | 4 | 0 | 0.12108539 |
| MINDY3 | 136.62185 | 4 | 0 | 0.09288179 |
| SPTBN1 | 136.422586 | 4 | 0 | 0.07351448 |
| USP24 | 135.895741 | 4 | 0 | 0.11383646 |
| JPH4 | 135.823602 | 4 | 0 | 0.12775913 |
| KATNAL1 | 135.636414 | 4 | 0 | 0.15853186 |
| ATXN10 | 135.233896 | 4 | 0 | 0.06669755 |
| ADAM10 | 134.997848 | 4 | 0 | 0.27303884 |
| ATP1A1 | 134.731713 | 4 | 0 | 0.11170526 |
| CCPG1 | 134.549728 | 4 | 0 | 0.08402941 |
| AL359710.1 | 134.270474 | 4 | 0 | 0.22995122 |
| ARGLU1 | 134.256651 | 4 | 0 | 0.15676004 |
| RAD9A | 133.380248 | 4 | 0 | 0.10366632 |
| SPIDR | 133.084977 | 4 | 0 | 0.09343409 |
| JPX | 132.923682 | 4 | 0 | 0.20936225 |
| DYNLL1 | 132.571758 | 4 | 0 | 0.2691725 |
| NDUFA4 | 132.360693 | 4 | 0 | 0.21156118 |
| ZRANB3 | 132.340981 | 4 | 0 | 0.14764456 |
| VPS53 | 132.32078 | 4 | 0 | 0.08634608 |
| GDI1 | 132.128746 | 4 | 0 | 0.14477977 |
| NOL10 | 131.799558 | 4 | 0 | 0.12732013 |
| LENG8 | 131.735476 | 4 | 0 | 0.13130393 |
| B4GALNT3 | 131.480964 | 4 | 0 | 0.17800724 |
| AC092378.1 | 131.354669 | 4 | 0 | 0.13267858 |
| WDPCP | 131.172242 | 4 | 0 | 0.14917053 |
| CASTOR3 | 131.095836 | 4 | 0 | 0.10128985 |
| SLC13A3 | 130.945248 | 4 | 0 | 0.15622369 |
| LINC00513 | 130.492223 | 4 | 0 | 0.27054092 |
| INTU | 130.404803 | 4 | 0 | 0.11372865 |
| UTY | 130.374875 | 4 | 0 | 0.22728612 |
| SLC12A5 | 130.336456 | 4 | 0 | 0.16148258 |
| CELF1 | 129.866193 | 4 | 0 | 0.04507136 |
| KMT2A | 129.840512 | 4 | 0 | 0.10628765 |
| MDH1 | 129.701716 | 4 | 0 | 0.17527493 |
| CENPP | 129.503832 | 4 | 0 | 0.12604296 |
| RGS4 | 129.336567 | 4 | 0 | 0.18260822 |
| C3orf58 | 129.016067 | 4 | 0 | 0.17115607 |

| | | | | |
|---|---|---|---|---|
| STPG2 | 128.627536 | 4 | 0 | 0.26541194 |
| RTN3 | 128.397699 | 4 | 0 | 0.10342349 |
| UBR1 | 127.887473 | 4 | 0 | 0.07635873 |
| ELMO1 | 127.84676 | 4 | 0 | 0.11516398 |
| MICU3 | 127.752878 | 4 | 0 | 0.05319453 |
| KIF5A | 127.407088 | 4 | 0 | 0.13981365 |
| ZNF91 | 127.352245 | 4 | 0 | 0.12337643 |
| CLASP2 | 127.069156 | 4 | 0 | 0.09160776 |
| JMJD1C | 126.922385 | 4 | 0 | 0.05820901 |
| GLS | 126.855684 | 4 | 0 | 0.0472454 |
| ARHGAP44 | 126.781743 | 4 | 0 | 0.09223449 |
| GABRA1 | 126.746129 | 4 | 0 | 0.11945778 |
| KCNC2 | 126.03444 | 4 | 0 | 0.1235339 |
| ABCC8 | 125.674627 | 4 | 0 | 0.165252 |
| C11orf49 | 125.550968 | 4 | 0 | 0.08795412 |
| SMAP2 | 125.009292 | 4 | 0 | 0.21315752 |
| OCIAD1 | 124.945749 | 4 | 0 | 0.13107655 |
| ATG2B | 124.716017 | 4 | 0 | 0.11696017 |
| SLC25A12 | 124.584998 | 4 | 0 | 0.0850159 |
| ZNF706 | 124.516658 | 4 | 0 | 0.17195992 |
| NUDCD3 | 124.310707 | 4 | 0 | 0.12966217 |
| AL357153.2 | 124.150633 | 4 | 0 | 0.14995844 |
| PTPN5 | 124.11938 | 4 | 0 | 0.15836021 |
| HOOK3 | 123.744278 | 4 | 0 | 0.10486193 |
| YLPM1 | 123.458759 | 4 | 0 | 0.10811617 |
| SNRNP70 | 123.254598 | 4 | 0 | 0.05660526 |
| OPHN1 | 123.19658 | 4 | 0 | 0.15946166 |
| ADK | 122.84533 | 4 | 0 | 0.07951774 |
| INPP4A | 122.460443 | 4 | 0 | 0.08543479 |
| CSPP1 | 122.356367 | 4 | 0 | 0.12589023 |
| DYNC1H1 | 122.243684 | 4 | 0 | 0.05331571 |
| LCOR | 121.881165 | 4 | 0 | 0.10033857 |
| INSR | 121.49064 | 4 | 0 | 0.09159444 |
| SGMS1 | 121.349574 | 4 | 0 | 0.07015485 |
| TOM1L2 | 121.161452 | 4 | 0 | 0.06123927 |
| ARMC9 | 120.998951 | 4 | 0 | 0.14001326 |
| LUC7L | 120.478279 | 4 | 0 | 0.09325547 |
| HSPH1 | 120.400759 | 4 | 0 | 0.17773705 |
| ZBTB38 | 120.375275 | 4 | 0 | 0.09729717 |
| TERF2IP | 119.889968 | 4 | 0 | 0.22737302 |
| ELP4 | 119.735898 | 4 | 0 | 0.10882809 |
| TRANK1 | 119.343958 | 4 | 0 | 0.2015688 |
| BCLAF1 | 119.29182 | 4 | 0 | 0.12547548 |

| | | | | |
|---|---|---|---|---|
| MTCL1 | 119.027681 | 4 | 0 | 0.13278411 |
| CSMD3 | 118.836577 | 4 | 0 | 0.09662215 |
| ZER1 | 118.734333 | 4 | 0 | 0.07184542 |
| ANTXR1 | 118.31789 | 4 | 0 | 0.16237331 |
| LPIN1 | 117.520757 | 4 | 0 | 0.13910796 |
| CBFA2T2 | 117.122301 | 4 | 0 | 0.09466034 |
| NAA25 | 116.852165 | 4 | 0 | 0.10048271 |
| SHPRH | 116.667001 | 4 | 0 | 0.09227146 |
| SPG7 | 116.530194 | 4 | 0 | 0.12339904 |
| VPS13C | 116.393193 | 4 | 0 | 0.07622228 |
| NT5C2 | 116.286122 | 4 | 0 | 0.11145991 |
| RTN4RL1 | 116.095578 | 4 | 0 | 0.13282927 |
| TNKS2 | 116.008577 | 4 | 0 | 0.15860179 |
| PHLPP2 | 115.916063 | 4 | 0 | 0.14805421 |
| CAPZB | 115.655218 | 4 | 0 | 0.09229033 |
| 2-Sep | 115.089263 | 4 | 0 | 0.11386894 |
| SESTD1 | 114.781237 | 4 | 0 | 0.122202 |
| CRMP1 | 114.5195 | 4 | 0 | 0.12224565 |
| FAM168A | 114.461008 | 4 | 0 | 0.08438364 |
| RAB28 | 114.455266 | 4 | 0 | 0.10796016 |
| TUSC3 | 114.381451 | 4 | 0 | 0.17092389 |
| SFSWAP | 114.005736 | 4 | 0 | 0.05770807 |
| RANBP9 | 113.789969 | 4 | 0 | 0.10815843 |
| CCDC102B | 113.680571 | 4 | 0 | 0.12090787 |
| RASGEF1B | 113.656563 | 4 | 0 | 0.06779271 |
| CNKSR2 | 113.633782 | 4 | 0 | 0.08377088 |
| ZBTB8OS | 113.141837 | 4 | 0 | 0.13212892 |
| FTH1 | 113.132072 | 4 | 0 | 0.23848714 |
| VSTM2B | 113.090662 | 4 | 0 | 0.25543474 |
| VTI1A | 113.055198 | 4 | 0 | 0.06772315 |
| TOP2B | 113.048368 | 4 | 0 | 0.21263627 |
| RBFOX2 | 112.907321 | 4 | 0 | 0.05159816 |
| BMPR1A | 112.872332 | 4 | 0 | 0.09553558 |
| KIDINS220 | 112.862822 | 4 | 0 | 0.10680227 |
| BTBD10 | 112.541717 | 4 | 0 | 0.07833117 |
| UBE2B | 112.052509 | 4 | 0 | 0.1539834 |
| TDRD3 | 111.723643 | 4 | 0 | 0.12841326 |
| ZDHHC21 | 111.559922 | 4 | 0 | 0.08563654 |
| DMTN | 110.566461 | 4 | 0 | 0.10327353 |
| SCG5 | 110.372758 | 4 | 0 | 0.19690381 |
| HLF | 110.215236 | 4 | 0 | 0.06369626 |
| YWHAH | 109.889981 | 4 | 0 | 0.24150991 |
| KLHL24 | 109.440333 | 4 | 0 | 0.14316207 |

| | | | | |
|---|---|---|---|---|
| CLEC16A | 108.779343 | 4 | 0 | 0.22285819 |
| UCHL1 | 108.659821 | 4 | 0 | 0.1379085 |
| CCDC146 | 108.470762 | 4 | 0 | 0.20739722 |
| DSTN | 108.378972 | 4 | 0 | 0.2027748 |
| JARID2 | 108.261063 | 4 | 0 | 0.14441777 |
| ZNF519 | 107.912126 | 4 | 0 | 0.15593536 |
| CLCN3 | 107.906078 | 4 | 0 | 0.07015085 |
| NUTM2A-AS | 107.606318 | 4 | 0 | 0.10628282 |
| RYK | 107.552404 | 4 | 0 | 0.22643266 |
| DNMT3A | 107.350157 | 4 | 0 | 0.0834859 |
| MTDH | 107.07619 | 4 | 0 | 0.10171612 |
| EEF1A2 | 107.07407 | 4 | 0 | 0.13851832 |
| HIBCH | 106.968415 | 4 | 0 | 0.10021549 |
| PDE8A | 106.855737 | 4 | 0 | 0.29793919 |
| DYNC2LI1 | 106.692027 | 4 | 0 | 0.13197866 |
| DPYSL2 | 106.670187 | 4 | 0 | 0.09342057 |
| TBC1D5 | 106.64793 | 4 | 0 | 0.06893458 |
| ESYT2 | 106.550788 | 4 | 0 | 0.11908236 |
| GMDS-DT | 106.371693 | 4 | 0 | 0.12644513 |
| ADGRB1 | 106.364123 | 4 | 0 | 0.13422042 |
| TMEM232 | 106.285213 | 4 | 0 | 0.08960644 |
| BMPR2 | 106.19133 | 4 | 0 | 0.10446013 |
| IGSF9B | 105.981496 | 4 | 0 | 0.07393445 |
| NEFL | 105.904552 | 4 | 0 | 0.20017849 |
| TNRC18 | 105.645533 | 4 | 0 | 0.19655365 |
| AFF4 | 105.62828 | 4 | 0 | 0.07668152 |
| PRRC2C | 105.332529 | 4 | 0 | 0.08916901 |
| TMEM130 | 105.010354 | 4 | 0 | 0.22262286 |
| POU2F1 | 104.807192 | 4 | 0 | 0.088655 |
| MCPH1 | 104.764331 | 4 | 0 | 0.1934111 |
| RNF13 | 104.412229 | 4 | 0 | 0.10901122 |
| CBR4 | 104.346137 | 4 | 0 | 0.14894934 |
| SH3YL1 | 104.256814 | 4 | 0 | 0.09391739 |
| FAM169A | 104.094235 | 4 | 0 | 0.116284 |
| GPALPP1 | 104.02928 | 4 | 0 | 0.07367815 |
| GABBR2 | 103.893325 | 4 | 0 | 0.05208168 |
| ZNF273 | 103.667913 | 4 | 0 | 0.13029406 |
| ZMYM2 | 103.319989 | 4 | 0 | 0.04841953 |
| FAR2 | 103.148537 | 4 | 0 | 0.22480365 |
| PLEKHM3 | 102.834913 | 4 | 0 | 0.08935367 |
| DYNC1I2 | 102.43438 | 4 | 0 | 0.1097486 |
| PNCK | 102.261479 | 4 | 0 | 0.19101332 |
| IGSF21 | 102.202714 | 4 | 0 | 0.25005156 |

| | | | | |
|---|---|---|---|---|
| CLSTN1 | 101.926609 | 4 | 0 | 0.09542673 |
| UBE2H | 101.747869 | 4 | 0 | 0.06103927 |
| COA1 | 101.588277 | 4 | 0 | 0.10557149 |
| INTS6 | 101.4626 | 4 | 0 | 0.1312834 |
| CDK17 | 101.053907 | 4 | 0 | 0.16056135 |
| NCOR2 | 101.041056 | 4 | 0 | 0.11103558 |
| ELF2 | 100.954005 | 4 | 0 | 0.09015842 |
| CLVS1 | 100.745252 | 4 | 0 | 0.14893736 |
| HNRNPC | 100.665373 | 4 | 0 | 0.14493723 |
| ANKRD36 | 100.62431 | 4 | 0 | 0.06299177 |
| LARP1B | 100.468027 | 4 | 0 | 0.09161669 |
| KCNH1 | 100.336731 | 4 | 0 | 0.09105837 |
| TMCC1 | 99.9938835 | 4 | 0 | 0.08351672 |
| KCNQ2 | 99.6336956 | 4 | 0 | 0.07246369 |
| MAPT | 99.1344561 | 4 | 0 | 0.05023818 |
| FUS | 98.2653939 | 4 | 0 | 0.12911017 |
| TMEM178A | 98.2607489 | 4 | 0 | 0.10534102 |
| MLLT10 | 98.2295914 | 4 | 0 | 0.06622711 |
| ABLIM2 | 97.4841903 | 4 | 0 | 0.080975 |
| MALAT1 | 97.4839676 | 4 | 0 | 0.07737141 |
| RABGAP1 | 97.3259125 | 4 | 0 | 0.07846626 |
| ICA1L | 97.2926125 | 4 | 0 | 0.08132412 |
| MGST3 | 97.2340163 | 4 | 0 | 0.1096522 |
| GNB5 | 96.4731725 | 4 | 0 | 0.08836503 |
| CTTN | 96.3349677 | 4 | 0 | 0.09831244 |
| GAS7 | 96.0614368 | 4 | 0 | 0.12362794 |
| LZTS3 | 96.0269663 | 4 | 0 | 0.14539465 |
| HTR1E | 95.9699383 | 4 | 0 | 0.1205958 |
| AC118549.1 | 95.9391579 | 4 | 0 | 0.11225035 |
| TRIM44 | 95.2352237 | 4 | 0 | 0.15498473 |
| MYCBP2 | 95.0560914 | 4 | 0 | 0.07767631 |
| ZNF148 | 94.8284491 | 4 | 0 | 0.05841207 |
| MAP2K4 | 94.1946482 | 4 | 0 | 0.04042002 |
| PLEKHA1 | 94.1644257 | 4 | 0 | 0.09018969 |
| ATL1 | 93.9674469 | 4 | 0 | 0.10880264 |
| CYFIP2 | 93.5146116 | 4 | 0 | 0.09613449 |
| FAM135A | 93.4906185 | 4 | 0 | 0.09526796 |
| BSN | 93.2180317 | 4 | 0 | 0.13829567 |
| PPFIA4 | 92.9585387 | 4 | 0 | 0.08172649 |
| FAR1 | 92.538195 | 4 | 0 | 0.10253851 |
| ZFYVE28 | 92.4667036 | 4 | 0 | 0.13444545 |
| SRGAP3 | 92.271364 | 4 | 0 | 0.0878236 |
| RALGAPA2 | 92.0757411 | 4 | 0 | 0.10129572 |

| | | | | |
|---|---|---|---|---|
| WDR7 | 91.9022422 | 4 | 0 | 0.05723846 |
| CDK8 | 91.5400917 | 4 | 0 | 0.12190966 |
| NQO2 | 91.3718362 | 4 | 0 | 0.1061132 |
| LRP4 | 91.2921627 | 4 | 0 | 0.24450947 |
| SPTAN1 | 91.2586247 | 4 | 0 | 0.04800711 |
| LINC00662 | 91.2038663 | 4 | 0 | 0.15871007 |
| PIK3C3 | 91.1567368 | 4 | 0 | 0.13595326 |
| OTUD7A | 91.0474794 | 4 | 0 | 0.03270653 |
| HNRNPA2B1 | 90.9905695 | 4 | 0 | 0.13033773 |
| SNX10 | 90.9844723 | 4 | 0 | 0.11353786 |
| ARNTL2 | 90.9481857 | 4 | 0 | 0.17211322 |
| ATCAY | 90.853591 | 4 | 0 | 0.09966896 |
| AJAP1 | 90.7833882 | 4 | 0 | 0.08471031 |
| RBM5 | 90.5760628 | 4 | 0 | 0.16872654 |
| BNIP3L | 90.2169067 | 4 | 0 | 0.11082435 |
| TBC1D19 | 90.2030884 | 4 | 0 | 0.11281741 |
| PPP1R12A | 90.0672917 | 4 | 0 | 0.07357803 |
| ZDBF2 | 90.0565812 | 4 | 0 | 0.09816156 |
| CHD5 | 90.0230559 | 4 | 0 | 0.07693139 |
| SCAMP5 | 90.0119534 | 4 | 0 | 0.10662182 |
| TTLL7 | 89.5694261 | 4 | 0 | 0.0716672 |
| DNM1 | 89.5263699 | 4 | 0 | 0.05556636 |
| MRPL41 | 89.3487879 | 4 | 0 | 0.07708664 |
| UBE2D3 | 89.3272134 | 4 | 0 | 0.08139888 |
| ZCCHC7 | 89.1326931 | 4 | 0 | 0.08139871 |
| N4BP2L2 | 89.0042025 | 4 | 0 | 0.13284428 |
| TAGLN3 | 88.9679345 | 4 | 0 | 0.15277303 |
| FNBP1 | 88.866001 | 4 | 0 | 0.07235114 |
| MOSMO | 88.5929989 | 4 | 0 | 0.09775965 |
| ZHX3 | 88.4635715 | 4 | 0 | 0.08507455 |
| RERE | 88.0750469 | 4 | 0 | 0.0542483 |
| SPRED2 | 88.0719684 | 4 | 0 | 0.11307498 |
| ZSWIM5 | 87.9486429 | 4 | 0 | 0.10966371 |
| SSX2IP | 87.9387385 | 4 | 0 | 0.10077387 |
| HMGB1 | 87.3491673 | 4 | 0 | 0.07906846 |
| IBTK | 87.1824083 | 4 | 0 | 0.09128096 |
| TTN-AS1 | 87.0201686 | 4 | 0 | 0.05917233 |
| RPTOR | 87.0169774 | 4 | 0 | 0.08831099 |
| SLC22A17 | 86.2939955 | 4 | 0 | 0.12217099 |
| KDM4B | 86.1579301 | 4 | 0 | 0.09545856 |
| CIRBP | 85.953664 | 4 | 0 | 0.18430107 |
| MTHFD1L | 85.7844168 | 4 | 0 | 0.16963327 |
| AC079447.1 | 85.3845728 | 4 | 0 | 0.09088535 |

| | | | | |
|---|---|---|---|---|
| TASP1 | 85.0567097 | 4 | 0 | 0.0890324 |
| IFT43 | 85.0199732 | 4 | 0 | 0.14118364 |
| RNF180 | 84.5508513 | 4 | 0 | 0.10170703 |
| RUFY3 | 84.5466634 | 4 | 0 | 0.05843847 |
| CCNT2-AS1 | 84.5042842 | 4 | 0 | 0.10950017 |
| AP1G2 | 84.2917558 | 4 | 0 | 0.10324818 |
| DIP2C | 83.9365449 | 4 | 0 | 0.03011144 |
| LRRC37A3 | 83.7664786 | 4 | 0 | 0.08483092 |
| THADA | 83.2939211 | 4 | 0 | 0.06826215 |
| ABL2 | 83.2142364 | 4 | 0 | 0.05621852 |
| FAM49A | 83.1772939 | 4 | 0 | 0.08126011 |
| TBRG1 | 83.0965507 | 4 | 0 | 0.06169963 |
| EVL | 82.6172813 | 4 | 0 | 0.05127896 |
| SGCE | 82.3069683 | 4 | 1.11E-16 | 0.15808916 |
| ETNK1 | 82.2598077 | 4 | 1.11E-16 | 0.08698769 |
| EPC2 | 81.6382469 | 4 | 1.11E-16 | 0.10095053 |
| CPE | 81.3856986 | 4 | 1.11E-16 | 0.09025323 |
| ZNF248 | 81.3658498 | 4 | 1.11E-16 | 0.12368801 |
| WDR33 | 81.3218629 | 4 | 1.11E-16 | 0.12431261 |
| KDM2A | 81.1571734 | 4 | 1.11E-16 | 0.04304696 |
| GNAO1 | 81.1360262 | 4 | 1.11E-16 | 0.10235472 |
| ARID4B | 80.9535092 | 4 | 1.11E-16 | 0.03577654 |
| VAPA | 80.8783191 | 4 | 1.11E-16 | 0.13098773 |
| CABIN1 | 80.8117605 | 4 | 1.11E-16 | 0.07202316 |
| NUTM2B-AS | 80.6382348 | 4 | 1.11E-16 | 0.06556042 |
| HDGFL3 | 80.4932552 | 4 | 1.11E-16 | 0.17343167 |
| HERC2 | 80.4415144 | 4 | 1.11E-16 | 0.07615033 |
| PCCA | 80.3211436 | 4 | 1.11E-16 | 0.15884941 |
| TBC1D22A | 79.9787761 | 4 | 2.22E-16 | 0.12149783 |
| TFCP2 | 79.5594126 | 4 | 2.22E-16 | 0.09494062 |
| ANKRD36C | 79.3520797 | 4 | 2.22E-16 | 0.05516571 |
| PTPRN | 79.1529724 | 4 | 2.22E-16 | 0.1012255 |
| SRSF11 | 78.9137132 | 4 | 3.33E-16 | 0.05874269 |
| ADGRB2 | 78.81027 | 4 | 3.33E-16 | 0.17118276 |
| NAP1L1 | 78.7162123 | 4 | 3.33E-16 | 0.05756247 |
| WIPF2 | 78.1783947 | 4 | 4.44E-16 | 0.07851617 |
| NRDC | 78.1392985 | 4 | 4.44E-16 | 0.09255774 |
| DZIP1 | 78.1020891 | 4 | 4.44E-16 | 0.04927695 |
| MICU1 | 77.8205888 | 4 | 5.55E-16 | 0.06229428 |
| FAM184A | 77.3181173 | 4 | 6.66E-16 | 0.24964901 |
| RNGTT | 76.9899744 | 4 | 7.77E-16 | 0.10775576 |
| C2CD5 | 76.911989 | 4 | 7.77E-16 | 0.11976015 |
| KIZ | 76.6257033 | 4 | 8.88E-16 | 0.17257638 |

| | | | | |
|---|---|---|---|---|
| XRCC5 | 76.4438292 | 4 | 9.99E-16 | 0.1764413 |
| NSMCE2 | 76.4354986 | 4 | 9.99E-16 | 0.10292533 |
| TRIP12 | 76.4317802 | 4 | 9.99E-16 | 0.07046389 |
| KMT2E | 76.1361988 | 4 | 1.11E-15 | 0.08825935 |
| TRA2A | 75.7544555 | 4 | 1.33E-15 | 0.07095467 |
| PUM1 | 75.637092 | 4 | 1.44E-15 | 0.14523157 |
| SRSF5 | 75.5226775 | 4 | 1.55E-15 | 0.08200816 |
| VPS13B | 75.5038784 | 4 | 1.55E-15 | 0.04537884 |
| SENP6 | 75.0808235 | 4 | 1.89E-15 | 0.12995664 |
| STAU2 | 74.9950108 | 4 | 2.00E-15 | 0.08743008 |
| MOK | 74.9467701 | 4 | 2.00E-15 | 0.11524135 |
| AP3B2 | 74.8796409 | 4 | 2.11E-15 | 0.09176792 |
| ZBTB18 | 74.5839085 | 4 | 2.44E-15 | 0.25348113 |
| RAB3GAP1 | 74.5534826 | 4 | 2.44E-15 | 0.08605035 |
| PHF3 | 74.3712619 | 4 | 2.66E-15 | 0.04729999 |
| ITSN2 | 74.1399806 | 4 | 3.00E-15 | 0.08426063 |
| TCF25 | 74.1393028 | 4 | 3.00E-15 | 0.04714527 |
| ZRANB2 | 74.0470261 | 4 | 3.22E-15 | 0.15368634 |
| TNRC6A | 74.0162793 | 4 | 3.22E-15 | 0.06300458 |
| ATRN | 73.7825543 | 4 | 3.55E-15 | 0.0689302 |
| WDR70 | 73.7703908 | 4 | 3.66E-15 | 0.06444794 |
| ARHGEF9 | 73.6890864 | 4 | 3.77E-15 | 0.14562514 |
| KDM5B | 73.5417895 | 4 | 4.11E-15 | 0.09861303 |
| FEZ2 | 73.5146422 | 4 | 4.11E-15 | 0.08122008 |
| TMEM245 | 73.4797541 | 4 | 4.22E-15 | 0.15031325 |
| PPM1B | 73.444248 | 4 | 4.22E-15 | 0.08839627 |
| PPHLN1 | 73.2281131 | 4 | 4.77E-15 | 0.09076289 |
| SCLT1 | 72.9627792 | 4 | 5.33E-15 | 0.06499502 |
| CAMSAP2 | 72.6456643 | 4 | 6.22E-15 | 0.05207809 |
| SPIRE1 | 72.6085749 | 4 | 6.33E-15 | 0.08050393 |
| TMOD2 | 72.51903 | 4 | 6.66E-15 | 0.04266749 |
| ABCA5 | 72.2933418 | 4 | 7.44E-15 | 0.05078913 |
| YWHAE | 71.9895727 | 4 | 8.66E-15 | 0.12776252 |
| SIMC1 | 71.965414 | 4 | 8.77E-15 | 0.08634377 |
| MAP3K2 | 71.9343722 | 4 | 8.88E-15 | 0.08639837 |
| ZC3H6 | 71.8701959 | 4 | 9.10E-15 | 0.12169241 |
| RSF1 | 71.8435641 | 4 | 9.21E-15 | 0.11921754 |
| PPP1R16B | 71.7140062 | 4 | 9.88E-15 | 0.05174555 |
| RUNDC3B | 71.6545013 | 4 | 1.01E-14 | 0.07852152 |
| BEND6 | 71.5035786 | 4 | 1.09E-14 | 0.26622585 |
| CC2D1A | 71.1303386 | 4 | 1.31E-14 | 0.11027484 |
| KPNA3 | 71.1124753 | 4 | 1.32E-14 | 0.11228308 |
| MICAL3 | 71.1013019 | 4 | 1.33E-14 | 0.03595076 |

| | | | | |
|---|---|---|---|---|
| CSTF3 | 70.9614123 | 4 | 1.42E-14 | 0.18294867 |
| SYTL2 | 70.7032774 | 4 | 1.61E-14 | 0.12514437 |
| SLC20A2 | 70.5954924 | 4 | 1.70E-14 | 0.08034648 |
| MYEF2 | 70.5505289 | 4 | 1.73E-14 | 0.11881023 |
| KPNB1 | 69.9194468 | 4 | 2.36E-14 | 0.09485434 |
| FAM13B | 69.8736192 | 4 | 2.41E-14 | 0.07091582 |
| DNAJC6 | 69.8505491 | 4 | 2.44E-14 | 0.06535706 |
| HUWE1 | 69.4007427 | 4 | 3.04E-14 | 0.13815409 |
| FAF1 | 69.2416812 | 4 | 3.29E-14 | 0.04805414 |
| TRAPPC12 | 69.1333858 | 4 | 3.46E-14 | 0.14790417 |
| CRTC1 | 69.093671 | 4 | 3.53E-14 | 0.08792594 |
| TENT2 | 69.0614119 | 4 | 3.59E-14 | 0.05788789 |
| PCSK7 | 68.7734173 | 4 | 4.12E-14 | 0.09772873 |
| AUH | 68.6211594 | 4 | 4.44E-14 | 0.1159101 |
| PACSIN1 | 67.4873541 | 4 | 7.69E-14 | 0.13011739 |
| COL19A1 | 67.4740944 | 4 | 7.75E-14 | 0.06839403 |
| CEP350 | 67.4508734 | 4 | 7.83E-14 | 0.06730709 |
| PPIP5K1 | 66.8806783 | 4 | 1.03E-13 | 0.05662433 |
| LRCH3 | 66.8091935 | 4 | 1.07E-13 | 0.09104464 |
| SFPQ | 66.7225552 | 4 | 1.12E-13 | 0.14290805 |
| MYO5A | 66.5302981 | 4 | 1.22E-13 | 0.06069779 |
| SCMH1 | 66.476247 | 4 | 1.26E-13 | 0.05316684 |
| HECTD4 | 66.3898819 | 4 | 1.31E-13 | 0.04546386 |
| MTA3 | 65.9683264 | 4 | 1.61E-13 | 0.10271858 |
| ITCH | 65.716904 | 4 | 1.82E-13 | 0.07465059 |
| NCOR1 | 65.6694579 | 4 | 1.86E-13 | 0.08376376 |
| DYNC2H1 | 65.6661264 | 4 | 1.86E-13 | 0.08962282 |
| LONP2 | 65.6512127 | 4 | 1.88E-13 | 0.13019632 |
| MAGED1 | 65.4996579 | 4 | 2.02E-13 | 0.0916016 |
| RB1CC1 | 65.4077925 | 4 | 2.11E-13 | 0.0674235 |
| FTO | 65.3049483 | 4 | 2.22E-13 | 0.05658851 |
| BDH1 | 65.2927373 | 4 | 2.23E-13 | 0.12513699 |
| NPEPPS | 65.2601146 | 4 | 2.27E-13 | 0.06205153 |
| ZNF131 | 65.1999836 | 4 | 2.33E-13 | 0.04755039 |
| LINC00599 | 65.1793569 | 4 | 2.36E-13 | 0.10075668 |
| ACSL3 | 65.1482911 | 4 | 2.39E-13 | 0.09047331 |
| HECTD1 | 65.096017 | 4 | 2.46E-13 | 0.08858547 |
| PRKRIP1 | 64.7570551 | 4 | 2.90E-13 | 0.08588912 |
| CHM | 64.6875687 | 4 | 2.99E-13 | 0.09175333 |
| PHC3 | 64.6163528 | 4 | 3.10E-13 | 0.10725368 |
| LINC01266 | 64.5265013 | 4 | 3.24E-13 | 0.48844352 |
| PCM1 | 64.4345092 | 4 | 3.39E-13 | 0.06441263 |
| KANSL1 | 64.4122246 | 4 | 3.42E-13 | 0.06687999 |

| | | | | |
|---|---|---|---|---|
| AKAP9 | 64.3066934 | 4 | 3.60E-13 | 0.06534122 |
| EP400 | 64.0503003 | 4 | 4.08E-13 | 0.11963141 |
| APBA1 | 64.0231718 | 4 | 4.13E-13 | 0.10458328 |
| RAB7A | 64.0204332 | 4 | 4.14E-13 | 0.10161013 |
| CLOCK | 63.9814857 | 4 | 4.22E-13 | 0.04713356 |
| MAPKAP1 | 63.9534413 | 4 | 4.27E-13 | 0.06750944 |
| FBXO33 | 63.9455758 | 4 | 4.29E-13 | 0.05218584 |
| AL445433.2 | 63.9002932 | 4 | 4.39E-13 | 0.12838904 |
| SMCHD1 | 63.8005611 | 4 | 4.60E-13 | 0.06568551 |
| CCDC6 | 63.4125208 | 4 | 5.56E-13 | 0.11395551 |
| MAPKBP1 | 63.1557477 | 4 | 6.29E-13 | 0.08578442 |
| KDM4C | 63.1332384 | 4 | 6.36E-13 | 0.0822881 |
| DPH6 | 63.0980758 | 4 | 6.47E-13 | 0.14794768 |
| RSRC1 | 62.862247 | 4 | 7.25E-13 | 0.07550106 |
| FAM66C | 62.6368064 | 4 | 8.09E-13 | 0.1166253 |
| KPNA1 | 62.5746507 | 4 | 8.34E-13 | 0.08175591 |
| TBCD | 62.3991935 | 4 | 9.08E-13 | 0.07658912 |
| PRKN | 62.3152603 | 4 | 9.46E-13 | 0.06326411 |
| GCC2 | 62.2630792 | 4 | 9.70E-13 | 0.05325998 |
| IRAK1BP1 | 62.1780075 | 4 | 1.01E-12 | 0.05199461 |
| ATXN2 | 61.9922021 | 4 | 1.11E-12 | 0.06302292 |
| PICALM | 61.599767 | 4 | 1.34E-12 | 0.06930517 |
| TAOK3 | 60.7615997 | 4 | 2.01E-12 | 0.0783888 |
| REV3L | 60.653985 | 4 | 2.11E-12 | 0.05323774 |
| JAM3 | 59.9887905 | 4 | 2.92E-12 | 0.08392288 |
| ATP6V0A1 | 59.6763787 | 4 | 3.39E-12 | 0.04107218 |
| DENND4A | 59.5342659 | 4 | 3.63E-12 | 0.04862044 |
| SLC41A2 | 59.4892093 | 4 | 3.71E-12 | 0.08153326 |
| KAT6A | 58.9819821 | 4 | 4.75E-12 | 0.07303182 |
| KIAA1109 | 58.7905212 | 4 | 5.21E-12 | 0.07637195 |
| MFSD6 | 58.6905127 | 4 | 5.47E-12 | 0.11757821 |
| KLHDC10 | 58.5059551 | 4 | 5.98E-12 | 0.13169986 |
| LRRTM3 | 58.4833873 | 4 | 6.04E-12 | 0.06531651 |
| TULP4 | 57.7757354 | 4 | 8.50E-12 | 0.06753605 |
| TET3 | 57.745113 | 4 | 8.63E-12 | 0.12427079 |
| HDAC8 | 57.4995763 | 4 | 9.72E-12 | 0.08859285 |
| ULK4 | 57.3533247 | 4 | 1.04E-11 | 0.09956246 |
| ELAVL3 | 57.2899104 | 4 | 1.08E-11 | 0.07486084 |
| NSMAF | 57.2015658 | 4 | 1.12E-11 | 0.0656267 |
| VPS45 | 57.1009032 | 4 | 1.18E-11 | 0.08743157 |
| PTPRA | 56.9840129 | 4 | 1.25E-11 | 0.09146394 |
| CMIP | 56.8883642 | 4 | 1.31E-11 | 0.06732426 |
| SMARCAD1 | 56.6825545 | 4 | 1.44E-11 | 0.04005871 |

| | | | | |
|---|---|---|---|---|
| MARK3 | 56.4477887 | 4 | 1.62E-11 | 0.065236 |
| RALGPS2 | 56.3108215 | 4 | 1.73E-11 | 0.10917612 |
| NAPB | 56.235095 | 4 | 1.79E-11 | 0.14377882 |
| SPRY4-AS1 | 56.1092973 | 4 | 1.90E-11 | 0.22619499 |
| WDR60 | 55.9015682 | 4 | 2.10E-11 | 0.0703344 |
| PLXNA4 | 55.5963088 | 4 | 2.44E-11 | 0.10793935 |
| SNX9 | 55.3834017 | 4 | 2.70E-11 | 0.11397165 |
| GGT7 | 55.2973239 | 4 | 2.81E-11 | 0.1396195 |
| RUNDC3A | 55.2966993 | 4 | 2.82E-11 | 0.0972645 |
| SUZ12 | 55.2019789 | 4 | 2.95E-11 | 0.0480179 |
| PPP6R3 | 55.0190387 | 4 | 3.22E-11 | 0.10153854 |
| ITSN1 | 54.70771 | 4 | 3.74E-11 | 0.07309486 |
| NFKB1 | 54.6478871 | 4 | 3.85E-11 | 0.10922643 |
| CSRNP3 | 54.5967571 | 4 | 3.95E-11 | 0.09614684 |
| TAX1BP1 | 54.4235592 | 4 | 4.29E-11 | 0.06599543 |
| USP15 | 54.3230034 | 4 | 4.50E-11 | 0.09872228 |
| PRRC2B | 54.3004187 | 4 | 4.55E-11 | 0.08590719 |
| KIFAP3 | 54.266289 | 4 | 4.63E-11 | 0.07122526 |
| FLG-AS1 | 54.2514544 | 4 | 4.66E-11 | 0.1584394 |
| DYRK1A | 54.0865351 | 4 | 5.05E-11 | 0.0912126 |
| ZNF207 | 54.0829152 | 4 | 5.06E-11 | 0.10117473 |
| FBXO21 | 53.9876799 | 4 | 5.29E-11 | 0.05917173 |
| ADGRL1 | 53.2191539 | 4 | 7.67E-11 | 0.03411087 |
| MICU2 | 52.9184917 | 4 | 8.86E-11 | 0.11214837 |
| MUC20-OT1 | 52.8982904 | 4 | 8.95E-11 | 0.0711525 |
| RICTOR | 52.7868207 | 4 | 9.44E-11 | 0.03148637 |
| ARHGEF12 | 52.7208464 | 4 | 9.75E-11 | 0.10196282 |
| NDFIP1 | 52.5980741 | 4 | 1.03E-10 | 0.1052653 |
| LMBRD1 | 52.5141244 | 4 | 1.08E-10 | 0.14694435 |
| ZNF106 | 52.0136884 | 4 | 1.37E-10 | 0.08832392 |
| PPP3CB | 51.9258577 | 4 | 1.43E-10 | 0.05783644 |
| COMMD10 | 51.8920643 | 4 | 1.45E-10 | 0.11048875 |
| UBE3C | 51.8728362 | 4 | 1.47E-10 | 0.05397326 |
| MGA | 51.6863814 | 4 | 1.60E-10 | 0.1389613 |
| NSD1 | 51.0945348 | 4 | 2.13E-10 | 0.04719815 |
| SYNJ1 | 50.81309 | 4 | 2.44E-10 | 0.05143412 |
| PSIP1 | 50.794057 | 4 | 2.46E-10 | 0.10134245 |
| USP9Y | 50.3522332 | 4 | 3.05E-10 | 0.18515849 |
| PUM2 | 50.1771382 | 4 | 3.32E-10 | 0.08472088 |
| TSC22D2 | 50.1234003 | 4 | 3.40E-10 | 0.07758666 |
| SLMAP | 49.9961822 | 4 | 3.62E-10 | 0.09654903 |
| MON2 | 49.9605577 | 4 | 3.68E-10 | 0.04881298 |
| RAB11FIP4 | 49.9152318 | 4 | 3.76E-10 | 0.09674963 |

| | | | | |
|---|---|---|---|---|
| CNNM2 | 49.8578632 | 4 | 3.87E-10 | 0.10069189 |
| AP2B1 | 49.3913753 | 4 | 4.84E-10 | 0.04242592 |
| ACTR2 | 49.3701953 | 4 | 4.89E-10 | 0.0625135 |
| TP53BP1 | 49.2321991 | 4 | 5.22E-10 | 0.09872951 |
| IGSF8 | 48.9927556 | 4 | 5.86E-10 | 0.19720751 |
| EFCAB2 | 48.6211997 | 4 | 7.00E-10 | 0.05951036 |
| NRG2 | 48.3470849 | 4 | 7.99E-10 | 0.04842121 |
| GPBP1 | 48.2949555 | 4 | 8.19E-10 | 0.09531281 |
| PCGF5 | 48.2800035 | 4 | 8.25E-10 | 0.08525356 |
| CDC42BPA | 48.1392896 | 4 | 8.83E-10 | 0.04451242 |
| PPM1K | 47.8768588 | 4 | 1.00E-09 | 0.08466644 |
| EHMT1 | 47.5524901 | 4 | 1.17E-09 | 0.06151004 |
| AP2A2 | 47.5511995 | 4 | 1.17E-09 | 0.08190527 |
| TRAK2 | 47.4969291 | 4 | 1.20E-09 | 0.05000083 |
| DGKZ | 46.9411136 | 4 | 1.57E-09 | 0.03358425 |
| ANKRD17 | 46.4953902 | 4 | 1.94E-09 | 0.04113548 |
| POGZ | 45.1819782 | 4 | 3.64E-09 | 0.05143035 |
| PBRM1 | 45.0919478 | 4 | 3.80E-09 | 0.07379578 |
| DANT2 | 45.0415322 | 4 | 3.90E-09 | 0.06489776 |
| AVL9 | 44.7155461 | 4 | 4.56E-09 | 0.06355642 |
| RC3H1 | 44.6610104 | 4 | 4.68E-09 | 0.07843093 |
| LMBR1 | 44.4658146 | 4 | 5.13E-09 | 0.11281665 |
| PTPRJ | 44.1368988 | 4 | 6.01E-09 | 0.05273016 |
| GOLGB1 | 43.8563701 | 4 | 6.87E-09 | 0.0682089 |
| RBPJ | 43.7633661 | 4 | 7.18E-09 | 0.13548115 |
| RANBP2 | 43.7135188 | 4 | 7.36E-09 | 0.09015883 |
| STX18-AS1 | 43.6650012 | 4 | 7.53E-09 | 0.04241231 |
| MAP4 | 43.587014 | 4 | 7.82E-09 | 0.05178167 |
| UBE2K | 43.5183398 | 4 | 8.08E-09 | 0.08756965 |
| AMBRA1 | 43.4064302 | 4 | 8.52E-09 | 0.03493303 |
| INO80D | 43.398021 | 4 | 8.56E-09 | 0.06118624 |
| MPHOSPH9 | 43.3252112 | 4 | 8.86E-09 | 0.08279227 |
| PHF20L1 | 43.2897519 | 4 | 9.01E-09 | 0.04791385 |
| LRFN5 | 42.7708549 | 4 | 1.15E-08 | 0.05077663 |
| TAF1D | 42.6891791 | 4 | 1.20E-08 | 0.0345168 |
| FAM85B | 42.4987355 | 4 | 1.31E-08 | 0.04612091 |
| TOP1 | 42.078019 | 4 | 1.61E-08 | 0.05704869 |
| AC097103.2 | 42.0107424 | 4 | 1.66E-08 | 0.10523034 |
| PHF14 | 41.9810486 | 4 | 1.68E-08 | 0.10069573 |
| PAFAH1B1 | 41.9457708 | 4 | 1.71E-08 | 0.06421298 |
| IPO9 | 41.9284081 | 4 | 1.73E-08 | 0.11169196 |
| CRTC3 | 41.6947937 | 4 | 1.93E-08 | 0.07914812 |
| WASHC1 | 41.5702464 | 4 | 2.05E-08 | 0.06238685 |

| | | | | |
|---|---|---|---|---|
| KIF21A | 41.5491056 | 4 | 2.07E-08 | 0.06385265 |
| PAN3 | 41.4533385 | 4 | 2.17E-08 | 0.07509204 |
| RAB6A | 41.3285009 | 4 | 2.30E-08 | 0.04416924 |
| RBM39 | 41.2218985 | 4 | 2.42E-08 | 0.09274614 |
| CDKAL1 | 40.6345174 | 4 | 3.20E-08 | 0.05995617 |
| U2SURP | 40.4624929 | 4 | 3.47E-08 | 0.11345611 |
| GATAD2B | 40.4100883 | 4 | 3.56E-08 | 0.05286785 |
| MSRA | 40.3373004 | 4 | 3.69E-08 | 0.03764088 |
| ANKRD12 | 40.3025779 | 4 | 3.75E-08 | 0.06205511 |
| GNB1 | 40.3018812 | 4 | 3.75E-08 | 0.05904258 |
| TMEM131 | 40.2778148 | 4 | 3.79E-08 | 0.04389697 |
| HMBOX1 | 40.2663928 | 4 | 3.81E-08 | 0.04429099 |
| APLP2 | 40.2634343 | 4 | 3.82E-08 | 0.05814824 |
| CNOT2 | 40.234719 | 4 | 3.87E-08 | 0.0765212 |
| ORC4 | 40.0032718 | 4 | 4.32E-08 | 0.09431994 |
| KIF1A | 39.9652174 | 4 | 4.40E-08 | 0.05848474 |
| SNAP25-AS1 | 39.7182119 | 4 | 4.95E-08 | 0.09577607 |
| HP1BP3 | 39.6435156 | 4 | 5.13E-08 | 0.10598599 |
| SERGEF | 39.3975788 | 4 | 5.77E-08 | 0.05644122 |
| MIB1 | 39.3408043 | 4 | 5.92E-08 | 0.04567061 |
| FOXJ3 | 39.3361631 | 4 | 5.94E-08 | 0.04412576 |
| CCDC66 | 39.3010534 | 4 | 6.04E-08 | 0.07086692 |
| CRYL1 | 38.6784807 | 4 | 8.12E-08 | 0.06243412 |
| MIPOL1 | 38.4373913 | 4 | 9.10E-08 | 0.07978807 |
| OLA1 | 38.4177779 | 4 | 9.19E-08 | 0.06335015 |
| DCAF5 | 38.2779523 | 4 | 9.82E-08 | 0.06774681 |
| PRDM2 | 38.1770272 | 4 | 1.03E-07 | 0.10343507 |
| FBXO42 | 38.1465683 | 4 | 1.05E-07 | 0.06925548 |
| KNDC1 | 38.0677529 | 4 | 1.09E-07 | 0.06842951 |
| GLG1 | 37.8981126 | 4 | 1.18E-07 | 0.05084999 |
| GTF2I | 37.2247868 | 4 | 1.62E-07 | 0.0378837 |
| ATP6V1A | 36.9051749 | 4 | 1.88E-07 | 0.04085425 |
| NRF1 | 36.8839319 | 4 | 1.90E-07 | 0.04519648 |
| ACBD6 | 36.6591121 | 4 | 2.12E-07 | 0.05224507 |
| RNF216 | 36.4141835 | 4 | 2.38E-07 | 0.03313054 |
| ADNP | 36.4069797 | 4 | 2.39E-07 | 0.08186274 |
| RCOR3 | 36.2149538 | 4 | 2.61E-07 | 0.10869757 |
| LARP4B | 35.9946759 | 4 | 2.90E-07 | 0.06014055 |
| NMD3 | 35.8576945 | 4 | 3.10E-07 | 0.13964895 |
| CYFIP1 | 35.7909098 | 4 | 3.19E-07 | 0.07477732 |
| TSPOAP1 | 35.6823222 | 4 | 3.36E-07 | 0.10874341 |
| SUGP2 | 35.5957081 | 4 | 3.50E-07 | 0.06367482 |
| LINC00486 | 35.5726695 | 4 | 3.54E-07 | 0.19431373 |

| | | | | |
|---|---|---|---|---|
| TMEM241 | 35.1437718 | 4 | 4.34E-07 | 0.06370163 |
| OSBP2 | 34.9409711 | 4 | 4.78E-07 | 0.05197187 |
| SON | 34.6002054 | 4 | 5.61E-07 | 0.06046989 |
| DMXL2 | 34.3484424 | 4 | 6.32E-07 | 0.03798294 |
| P4HTM | 33.796951 | 4 | 8.20E-07 | 0.07274989 |
| CORO2B | 33.4868197 | 4 | 9.49E-07 | 0.0302273 |
| HCG17 | 33.4674866 | 4 | 9.58E-07 | 0.06356714 |
| EPC1 | 33.3567563 | 4 | 1.01E-06 | 0.08560396 |
| PSPC1 | 33.2643928 | 4 | 1.05E-06 | 0.04692632 |
| ACYP2 | 33.2557528 | 4 | 1.06E-06 | 0.09051538 |
| TBCK | 33.0836653 | 4 | 1.15E-06 | 0.05099392 |
| WDFY3 | 33.0202508 | 4 | 1.18E-06 | 0.06804993 |
| NSD3 | 32.7967077 | 4 | 1.31E-06 | 0.04098346 |
| TRPC1 | 32.7053605 | 4 | 1.37E-06 | 0.02737996 |
| LINC02210-C | 32.6218367 | 4 | 1.43E-06 | 0.06244331 |
| ARNTL | 32.3231139 | 4 | 1.64E-06 | 0.11815881 |
| TAB2 | 32.1929465 | 4 | 1.75E-06 | 0.05129331 |
| HSD17B12 | 32.1127161 | 4 | 1.81E-06 | 0.05243777 |
| NFX1 | 32.0044837 | 4 | 1.91E-06 | 0.07042307 |
| TTTY14 | 31.9387264 | 4 | 1.97E-06 | 0.07031853 |
| MORF4L2 | 31.7424225 | 4 | 2.16E-06 | 0.09450313 |
| CPLANE1 | 31.5738215 | 4 | 2.34E-06 | 0.0618212 |
| PDXDC1 | 31.4876986 | 4 | 2.43E-06 | 0.07493993 |
| VMP1 | 31.4263207 | 4 | 2.51E-06 | 0.10657282 |
| GPATCH2L | 31.222284 | 4 | 2.76E-06 | 0.04213106 |
| GOLGA4 | 31.0119824 | 4 | 3.04E-06 | 0.06769246 |
| GALK2 | 30.9603411 | 4 | 3.12E-06 | 0.09516502 |
| SUN1 | 30.9538013 | 4 | 3.13E-06 | 0.09536363 |
| XIST | 30.9004776 | 4 | 3.21E-06 | 0.11435728 |
| BOD1L1 | 30.8471737 | 4 | 3.29E-06 | 0.03667641 |
| PKD2 | 30.8208636 | 4 | 3.33E-06 | 0.08103044 |
| CEP290 | 30.7010357 | 4 | 3.52E-06 | 0.06072338 |
| UBXN4 | 30.6398796 | 4 | 3.63E-06 | 0.05022719 |
| WSB1 | 30.6298838 | 4 | 3.64E-06 | 0.10063424 |
| KANTR | 30.5611226 | 4 | 3.76E-06 | 0.04182806 |
| DOPEY1 | 30.3910491 | 4 | 4.07E-06 | 0.04942773 |
| KIF1B | 30.3217868 | 4 | 4.21E-06 | 0.03545695 |
| ATP9A | 30.3021684 | 4 | 4.25E-06 | 0.04215071 |
| DNAJB14 | 30.2006607 | 4 | 4.45E-06 | 0.08281737 |
| STRN3 | 30.0826038 | 4 | 4.71E-06 | 0.06270042 |
| RPRD2 | 30.0324065 | 4 | 4.82E-06 | 0.04591082 |
| CAB39 | 30.0243939 | 4 | 4.84E-06 | 0.09669277 |
| UBE3A | 29.9444403 | 4 | 5.02E-06 | 0.04386326 |

| | | | | |
|---|---|---|---|---|
| RABEP1 | 29.7165189 | 4 | 5.59E-06 | 0.05308322 |
| RAB6B | 29.30961 | 4 | 6.76E-06 | 0.09899772 |
| FBXL20 | 29.1595432 | 4 | 7.26E-06 | 0.04991432 |
| HNRNPDL | 28.9239797 | 4 | 8.10E-06 | 0.09100901 |
| KIAA0232 | 28.799334 | 4 | 8.59E-06 | 0.05049529 |
| ANKIB1 | 28.7893592 | 4 | 8.63E-06 | 0.04645861 |
| HPRT1 | 28.6136947 | 4 | 9.36E-06 | 0.0576764 |
| DOCK3 | 28.5005018 | 4 | 9.87E-06 | 0.02012818 |
| CDC27 | 28.3680125 | 4 | 1.05E-05 | 0.05546252 |
| DYM | 28.2768061 | 4 | 1.10E-05 | 0.04645364 |
| TAF15 | 28.1919808 | 4 | 1.14E-05 | 0.02402258 |
| RFX7 | 28.0494286 | 4 | 1.22E-05 | 0.06796917 |
| PSMD1 | 28.0235103 | 4 | 1.23E-05 | 0.06122167 |
| RNF149 | 27.9371587 | 4 | 1.28E-05 | 0.04252037 |
| GIGYF2 | 27.1944558 | 4 | 1.82E-05 | 0.05171968 |
| FBXO11 | 26.8183136 | 4 | 2.16E-05 | 0.03430005 |
| ZC3H13 | 26.8141787 | 4 | 2.17E-05 | 0.07003717 |
| MDN1 | 26.6673734 | 4 | 2.32E-05 | 0.05818671 |
| SPAG9 | 26.6380142 | 4 | 2.35E-05 | 0.02760108 |
| ATF6 | 26.6350464 | 4 | 2.36E-05 | 0.06862034 |
| DDX5 | 26.4961613 | 4 | 2.51E-05 | 0.1173897 |
| CREBBP | 26.373995 | 4 | 2.66E-05 | 0.04721897 |
| FAM185A | 26.3006999 | 4 | 2.75E-05 | 0.0907846 |
| XRN1 | 26.0914224 | 4 | 3.03E-05 | 0.02582001 |
| UBR5 | 25.9978913 | 4 | 3.17E-05 | 0.02248424 |
| PDE7A | 25.8330908 | 4 | 3.42E-05 | 0.09480896 |
| BDP1 | 25.6786133 | 4 | 3.67E-05 | 0.03945135 |
| DNM1L | 25.6295384 | 4 | 3.76E-05 | 0.0400754 |
| 7-Mar | 25.5696177 | 4 | 3.86E-05 | 0.06680043 |
| KTN1 | 25.5141309 | 4 | 3.96E-05 | 0.09537153 |
| CCDC57 | 25.413189 | 4 | 4.15E-05 | 0.04968867 |
| RUFY2 | 25.3915802 | 4 | 4.20E-05 | 0.06034118 |
| USP34 | 25.335208 | 4 | 4.31E-05 | 0.03049461 |
| NME5 | 25.3125969 | 4 | 4.35E-05 | 0.08258605 |
| INVS | 25.273097 | 4 | 4.43E-05 | 0.02912704 |
| AGO3 | 25.0049544 | 4 | 5.02E-05 | 0.03227856 |
| TECPR2 | 24.927344 | 4 | 5.20E-05 | 0.02677268 |
| LRRC6 | 24.4329377 | 4 | 6.54E-05 | 0.04453288 |
| LHFPL4 | 24.3499023 | 4 | 6.80E-05 | 0.02842258 |
| ALMS1 | 24.3286412 | 4 | 6.86E-05 | 0.05792991 |
| DCUN1D4 | 24.0614927 | 4 | 7.76E-05 | 0.04881392 |
| APC | 23.5799484 | 4 | 9.70E-05 | 0.04958337 |
| AC098829.1 | 23.3225669 | 4 | 0.00010916 | 0.19428485 |

| | | | | |
|---|---|---|---|---|
| PPP1R13B | 23.2712901 | 4 | 0.00011176 | 0.02391669 |
| TOGARAM1 | 23.0556248 | 4 | 0.00012343 | 0.06522982 |
| RPRD1A | 22.9267675 | 4 | 0.00013096 | 0.06555209 |
| CCDC7 | 22.8617536 | 4 | 0.00013494 | 0.07055819 |
| DOC2A | 22.4249311 | 4 | 0.00016493 | 0.03213183 |
| MTR | 22.2734724 | 4 | 0.0001768 | 0.04776743 |
| TTC17 | 22.266541 | 4 | 0.00017736 | 0.05754829 |
| PPP2R2A | 22.2518939 | 4 | 0.00017856 | 0.04774383 |
| ADD1 | 22.1073006 | 4 | 0.0001908 | 0.06268263 |
| RALGAPA1 | 22.0459139 | 4 | 0.00019625 | 0.03664136 |
| ZNF638 | 22.0176617 | 4 | 0.0001988 | 0.04420265 |
| KDM5A | 21.9040818 | 4 | 0.00020943 | 0.03835816 |
| CDK13 | 21.8383896 | 4 | 0.00021582 | 0.03758373 |
| CLTC | 21.8166977 | 4 | 0.00021798 | 0.04565984 |
| DMXL1 | 21.67763 | 4 | 0.00023231 | 0.02385082 |
| GSK3B | 21.6403891 | 4 | 0.00023631 | 0.01869815 |
| DIS3L2 | 21.4726977 | 4 | 0.00025515 | 0.04078302 |
| PPIG | 21.4299452 | 4 | 0.00026019 | 0.05088088 |
| RAB40C | 21.428556 | 4 | 0.00026035 | 0.03630751 |
| NF1 | 20.6581213 | 4 | 0.00037012 | 0.04310934 |
| CHPT1 | 20.6293452 | 4 | 0.000375 | 0.05793365 |
| FAM208B | 20.6288945 | 4 | 0.00037508 | 0.07618147 |
| PLD3 | 20.5290521 | 4 | 0.00039254 | 0.02993718 |
| ZMYM4 | 20.4688592 | 4 | 0.00040345 | 0.02202318 |
| PIBF1 | 20.4230016 | 4 | 0.00041197 | 0.02490229 |
| SPIN1 | 20.3513655 | 4 | 0.00042563 | 0.03913297 |
| PIAS2 | 20.3285607 | 4 | 0.00043007 | 0.03057727 |
| PPIP5K2 | 20.1840201 | 4 | 0.00045931 | 0.04605256 |
| UBR2 | 19.9341483 | 4 | 0.00051457 | 0.03969934 |
| PDS5A | 19.8368818 | 4 | 0.00053782 | 0.0387082 |
| MAPK8IP3 | 19.8018841 | 4 | 0.00054644 | 0.03636962 |
| FAM193A | 19.7982022 | 4 | 0.00054735 | 0.04658342 |
| BRD4 | 19.6728942 | 4 | 0.00057939 | 0.05083343 |
| RAB10 | 19.4757567 | 4 | 0.00063359 | 0.04469336 |
| NFE2L2 | 19.2794873 | 4 | 0.00069254 | 0.10334154 |
| COMMD1 | 19.1752564 | 4 | 0.00072601 | 0.02778246 |
| CCDC30 | 19.1362394 | 4 | 0.00073895 | 0.03477823 |
| DICER1 | 19.0530501 | 4 | 0.0007673 | 0.06788807 |
| NEXMIF | 18.8047587 | 4 | 0.00085848 | 0.03854356 |
| LRBA | 18.3628321 | 4 | 0.00104802 | 0.01845667 |
| VPS35L | 18.1734581 | 4 | 0.00114139 | 0.05095225 |
| COPA | 18.0510307 | 4 | 0.00120608 | 0.06915554 |
| CPEB3 | 17.7380975 | 4 | 0.00138834 | 0.02910145 |

| | | | | |
|---|---|---|---|---|
| USP48 | 17.6581449 | 4 | 0.00143911 | 0.07803101 |
| NKTR | 17.3000794 | 4 | 0.00168991 | 0.01677864 |
| BPTF | 17.2476392 | 4 | 0.0017301 | 0.02699911 |
| ARL17B | 17.1261798 | 4 | 0.00182682 | 0.04997897 |
| TNRC6B | 17.0873349 | 4 | 0.00185886 | 0.02276028 |
| COL4A3BP | 16.8126367 | 4 | 0.00210185 | 0.02401336 |
| KATNBL1 | 16.7940347 | 4 | 0.00211939 | 0.07881115 |
| FSD1L | 16.5423043 | 4 | 0.00237147 | 0.04921336 |
| RNF111 | 16.5341096 | 4 | 0.00238016 | 0.06284154 |
| CUL5 | 16.3823653 | 4 | 0.00254675 | 0.03791368 |
| MACO1 | 16.3603781 | 4 | 0.00257182 | 0.02682971 |
| RAP2A | 16.2385412 | 4 | 0.00271522 | 0.07159818 |
| ARMC8 | 16.139706 | 4 | 0.00283732 | 0.02395588 |
| PLPP1 | 16.0478731 | 4 | 0.00295559 | 0.05177245 |
| UBE2R2 | 15.8122409 | 4 | 0.00328174 | 0.02870951 |
| SETD2 | 15.7535721 | 4 | 0.0033683 | 0.03026857 |
| SETD5 | 15.6750634 | 4 | 0.00348766 | 0.03209732 |
| SNX13 | 15.6462304 | 4 | 0.00353253 | 0.03080084 |
| LMBRD2 | 15.585745 | 4 | 0.00362852 | 0.04028227 |
| ASH1L | 15.5838657 | 4 | 0.00363154 | 0.01791105 |
| RIC3 | 15.3285155 | 4 | 0.00406618 | 0.05380754 |
| SENP5 | 15.287638 | 4 | 0.00414035 | 0.04271737 |
| CWC27 | 15.2876118 | 4 | 0.0041404 | 0.01929273 |
| WDR37 | 15.0223282 | 4 | 0.00465513 | 0.0330525 |
| MCCC1 | 14.8922807 | 4 | 0.00492993 | 0.05594369 |
| RBM33 | 14.7329538 | 4 | 0.00528838 | 0.05629186 |
| PIGN | 14.717339 | 4 | 0.00532485 | 0.05983077 |
| MED13 | 14.6849896 | 4 | 0.00540121 | 0.02522251 |
| YTHDC2 | 14.5162818 | 4 | 0.00581717 | 0.05828264 |
| TTC19 | 14.3098508 | 4 | 0.00636906 | 0.02796986 |
| SPPL3 | 13.7748283 | 4 | 0.0080495 | 0.02954091 |
| RNF145 | 13.7446155 | 4 | 0.00815637 | 0.02316244 |
| NDUFA10 | 13.2504491 | 4 | 0.01011473 | 0.03614496 |
| VPS8 | 13.2170081 | 4 | 0.01026272 | 0.0167236 |
| NIPBL | 13.1553047 | 4 | 0.01054136 | 0.05196091 |
| PGP | 13.1505106 | 4 | 0.01056331 | 0.02729324 |
| RHOT1 | 12.9567486 | 4 | 0.01148907 | 0.02696125 |
| BIRC6 | 12.9138697 | 4 | 0.0117044 | 0.02259269 |
| XPO1 | 12.8815157 | 4 | 0.01186948 | 0.04391306 |
| FXR1 | 12.8384526 | 4 | 0.01209272 | 0.07241417 |
| RAPGEF4 | 12.7780258 | 4 | 0.0124129 | 0.01551674 |
| HACE1 | 12.5473397 | 4 | 0.01371303 | 0.01946528 |
| MIGA1 | 12.44279 | 4 | 0.0143451 | 0.04557585 |

| | | | | |
|---|---|---|---|---|
| CUL1 | 12.4295418 | 4 | 0.01442719 | 0.03695493 |
| KCNJ3 | 12.3427031 | 4 | 0.01497673 | 0.02215375 |
| SCAF8 | 12.2127644 | 4 | 0.01583728 | 0.05711728 |
| VPS54 | 12.1779825 | 4 | 0.01607567 | 0.03086042 |
| FNBP4 | 11.9103773 | 4 | 0.01803032 | 0.02230798 |
| KIAA0319L | 11.4651536 | 4 | 0.02180494 | 0.01760916 |
| UBN2 | 11.1649869 | 4 | 0.02477104 | 0.01947859 |
| RLF | 11.0574612 | 4 | 0.02592576 | 0.03405876 |
| FBXW11 | 10.8425833 | 4 | 0.0283913 | 0.05304465 |
| YAF2 | 10.8225353 | 4 | 0.02863256 | 0.03460755 |
| ZDHHC20 | 10.4801243 | 4 | 0.03307188 | 0.02814763 |
| ZNF451 | 10.4759139 | 4 | 0.03313039 | 0.0404409 |
| ZMYND11 | 10.3752153 | 4 | 0.03455997 | 0.01474389 |
| VPS50 | 10.2993037 | 4 | 0.03567674 | 0.01782571 |
| ODF2L | 9.9726179 | 4 | 0.04089146 | 0.02278222 |
| USP33 | 9.62124909 | 4 | 0.04731459 | 0.0235172 |
| ATG10 | 9.19244395 | 4 | 0.05646523 | 0.04799361 |
| ARFGEF1 | 9.06998101 | 4 | 0.05937389 | 0.01517245 |
| USP9X | 8.94926853 | 4 | 0.06238011 | 0.04327121 |
| ELP2 | 8.92029087 | 4 | 0.063123 | 0.02885516 |
| SETX | 8.88272906 | 4 | 0.06409848 | 0.03051615 |
| NEMF | 8.47120145 | 4 | 0.07576498 | 0.01846262 |
| SHOC2 | 8.17140803 | 4 | 0.08549729 | 0.03165942 |
| ANKRD13C | 8.15055211 | 4 | 0.08621638 | 0.03690082 |
| RPAP2 | 7.94488992 | 4 | 0.09361794 | 0.02743845 |
| RAB2A | 7.90668156 | 4 | 0.095057 | 0.02220521 |
| AC007388.1 | 7.74425055 | 4 | 0.10140884 | 0.02300778 |
| ATE1 | 7.68663447 | 4 | 0.10375558 | 0.01683732 |
| FAM120B | 6.88070302 | 4 | 0.14232844 | 0.01655097 |
| EP300 | 6.66596872 | 4 | 0.15462881 | 0.01534476 |
| PARG | 6.56787047 | 4 | 0.16056391 | 0.0248664 |
| U91319.1 | 6.50719725 | 4 | 0.16433746 | 0.04350839 |
| CEP170 | 6.07768187 | 4 | 0.19342157 | 0.01115839 |
| LMTK3 | 5.94057783 | 4 | 0.20363012 | 0.01707483 |
| AC124319.2 | 5.8792905 | 4 | 0.20834611 | 0.02720959 |
| HK1 | 5.53955512 | 4 | 0.23627437 | 0.02230399 |
| GAPVD1 | 5.39709588 | 4 | 0.248924 | 0.01222347 |
| PIP4K2A | 5.32957248 | 4 | 0.25512128 | 0.03581375 |
| ZNF644 | 4.98078193 | 4 | 0.28927508 | 0.01626452 |
| HELZ | 3.67287415 | 4 | 0.45208351 | 0.02256515 |
| ARMH3 | 3.33713384 | 4 | 0.50307032 | 0.01045104 |
| LRRC37B | 2.89108751 | 4 | 0.57621394 | 0.00911932 |
| ARIH1 | 2.86918397 | 4 | 0.57995034 | 0.00745456 |

| | | | | |
|---|---|---|---|---|
| KIF3A | 2.2674921 | 4 | 0.68669372 | 0.00847615 |
| SIK2 | 2.25869884 | 4 | 0.68829832 | 0.01127979 |
| TCERG1 | 2.20455439 | 4 | 0.69819555 | 0.005782 |
| NCOA7 | 1.90521819 | 4 | 0.75318634 | 0.00653396 |
| SUMF1 | 1.88974353 | 4 | 0.75602887 | 0.00768132 |
| ZKSCAN1 | 1.79798937 | 4 | 0.77285019 | 0.01095419 |
| CNOT4 | 1.20487528 | 4 | 0.87729528 | 0.00603033 |
| SLC30A9 | 1.0936808 | 4 | 0.89527352 | 0.00643719 |
| PPP2R5E | 0.79805003 | 4 | 0.93870929 | 0.0042767 |
| DCAF10 | 0.44181034 | 4 | 0.97891287 | 0.00375612 |
| RAB3GAP2 | 0.38996177 | 4 | 0.98329048 | 0.00368875 |
| PAPOLA | 0.07722286 | 4 | 0.99927349 | 0.00157643 |
| AHCTF1 | 0.0060497 | 4 | 0.99999543 | 0.00037953 |
| KIAA2026 | 0.00567906 | 4 | 0.99999598 | 0.00033203 |
| AC007319.1 | 0.00246295 | 4 | 0.99999924 | 0.00021242 |

**Supplementary Table 6. Microgial Marker Genes**

| | p_val | avg_log2FC | pct.1 | pct.2 | p_val_adj | cluster | gene |
|---|---|---|---|---|---|---|---|
| HSPA1A | 2.86E-102 | 0.98947761 | 0.867 | 0.761 | 5.72E-99 | 0 | HSPA1A |
| HLA-DRA | 3.24E-86 | 0.77938399 | 0.949 | 0.876 | 6.49E-83 | 0 | HLA-DRA |
| GPR183 | 8.64E-84 | 0.76177894 | 0.933 | 0.876 | 1.73E-80 | 0 | GPR183 |
| SRGN | 3.53E-72 | 0.92642864 | 0.879 | 0.81 | 7.06E-69 | 0 | SRGN |
| CD74 | 1.41E-70 | 0.7026835 | 0.991 | 0.958 | 2.81E-67 | 0 | CD74 |
| RGS1 | 1.62E-67 | 0.90877961 | 0.797 | 0.7 | 3.24E-64 | 0 | RGS1 |
| APOE | 1.25E-65 | 0.70573436 | 0.939 | 0.878 | 2.50E-62 | 0 | APOE |
| HLA-DRB1 | 4.75E-64 | 0.57660668 | 0.931 | 0.887 | 9.50E-61 | 0 | HLA-DRB1 |
| HIST1H2AC | 1.94E-57 | 0.57171218 | 0.951 | 0.909 | 3.87E-54 | 0 | HIST1H2AC |
| DUSP1 | 1.07E-51 | 0.75304871 | 0.9 | 0.863 | 2.15E-48 | 0 | DUSP1 |
| AC120193.1 | 3.66E-44 | 0.51873775 | 0.914 | 0.875 | 7.31E-41 | 0 | AC120193.1 |
| C1QC | 5.10E-41 | 0.50199755 | 0.802 | 0.702 | 1.02E-37 | 0 | C1QC |
| AIF1 | 1.22E-40 | 0.46800743 | 0.912 | 0.879 | 2.44E-37 | 0 | AIF1 |
| FOS | 2.23E-40 | 0.8014955 | 0.904 | 0.876 | 4.47E-37 | 0 | FOS |
| CST3 | 1.41E-35 | 0.33016383 | 0.898 | 0.853 | 2.82E-32 | 0 | CST3 |
| ZFP36L2 | 7.00E-35 | 0.45942204 | 0.994 | 0.963 | 1.40E-31 | 0 | ZFP36L2 |
| RNASET2 | 1.25E-33 | 0.35243268 | 0.988 | 0.963 | 2.50E-30 | 0 | RNASET2 |
| CEBPD | 2.84E-32 | 0.63068656 | 0.8 | 0.762 | 5.68E-29 | 0 | CEBPD |
| CD14 | 4.28E-32 | 0.60112869 | 0.737 | 0.677 | 8.56E-29 | 0 | CD14 |
| ADAM28 | 2.81E-31 | 0.40051499 | 0.993 | 0.976 | 5.62E-28 | 0 | ADAM28 |
| PADI2 | 5.16E-30 | 0.60805998 | 0.858 | 0.794 | 1.03E-26 | 0 | PADI2 |
| CD69 | 5.26E-30 | 0.50584133 | 0.896 | 0.885 | 1.05E-26 | 0 | CD69 |
| SPP1 | 1.15E-29 | 0.35083893 | 0.838 | 0.737 | 2.31E-26 | 0 | SPP1 |
| ZFP36L1 | 9.84E-29 | 0.47220955 | 0.994 | 0.972 | 1.97E-25 | 0 | ZFP36L1 |
| SERPINB9 | 1.11E-28 | 0.65764478 | 0.819 | 0.793 | 2.22E-25 | 0 | SERPINB9 |
| C1QA | 2.71E-28 | 0.44063796 | 0.767 | 0.701 | 5.42E-25 | 0 | C1QA |
| C1QB | 2.82E-27 | 0.43458763 | 0.83 | 0.738 | 5.63E-24 | 0 | C1QB |
| ABCC4 | 2.06E-26 | 0.41072423 | 0.988 | 0.954 | 4.12E-23 | 0 | ABCC4 |
| TMSB10 | 5.71E-26 | 0.58406629 | 0.889 | 0.85 | 1.14E-22 | 0 | TMSB10 |
| IL18 | 1.25E-25 | 0.35508819 | 0.925 | 0.859 | 2.49E-22 | 0 | IL18 |
| OLR1 | 1.04E-24 | 0.54152908 | 0.856 | 0.853 | 2.07E-21 | 0 | OLR1 |
| SAT1 | 2.01E-24 | 0.36184615 | 0.91 | 0.88 | 4.03E-21 | 0 | SAT1 |
| C3 | 2.18E-24 | 0.30229467 | 0.963 | 0.922 | 4.36E-21 | 0 | C3 |
| NR4A2 | 3.22E-24 | 0.25647733 | 0.729 | 0.645 | 6.43E-21 | 0 | NR4A2 |
| CD86 | 2.36E-23 | 0.30210494 | 0.955 | 0.913 | 4.71E-20 | 0 | CD86 |
| BLNK | 2.06E-22 | 0.35180837 | 0.985 | 0.932 | 4.11E-19 | 0 | BLNK |
| SLC2A3 | 1.54E-20 | 0.82333864 | 0.584 | 0.526 | 3.09E-17 | 0 | SLC2A3 |
| APOC1 | 3.57E-18 | 0.3261214 | 0.775 | 0.749 | 7.13E-15 | 0 | APOC1 |
| CYBB | 1.17E-16 | 0.35589208 | 0.879 | 0.81 | 2.33E-13 | 0 | CYBB |
| CSF3R | 2.00E-16 | 0.3464512 | 0.938 | 0.911 | 4.00E-13 | 0 | CSF3R |
| INPP5D | 7.57E-15 | 0.34310876 | 0.989 | 0.964 | 1.51E-11 | 0 | INPP5D |
| CSF1R | 8.57E-15 | 0.25017612 | 0.959 | 0.914 | 1.71E-11 | 0 | CSF1R |
| SRGAP2B | 1.21E-14 | 0.32549836 | 0.956 | 0.91 | 2.42E-11 | 0 | SRGAP2B |
| TNFRSF1B | 2.42E-14 | 0.32686452 | 0.801 | 0.775 | 4.84E-11 | 0 | TNFRSF1B |
| IRF8 | 2.72E-14 | 0.42068032 | 0.837 | 0.828 | 5.43E-11 | 0 | IRF8 |
| CCL4 | 3.32E-14 | 0.45770486 | 0.65 | 0.605 | 6.63E-11 | 0 | CCL4 |
| CCDC26 | 4.02E-14 | 0.33667383 | 0.867 | 0.832 | 8.04E-11 | 0 | CCDC26 |
| SLC2A5 | 1.34E-13 | 0.41146405 | 0.771 | 0.743 | 2.68E-10 | 0 | SLC2A5 |

| | | | | | | | |
|---|---|---|---|---|---|---|---|
| CSGALNACT1 | 1.85E-13 | 0.38119954 | 0.981 | 0.925 | 3.70E-10 | 0 | CSGALNACT1 |
| ATP8B4 | 2.99E-13 | 0.28654387 | 0.984 | 0.942 | 5.98E-10 | 0 | ATP8B4 |
| TMEM163 | 9.29E-13 | 0.30493115 | 0.502 | 0.38 | 1.86E-09 | 0 | TMEM163 |
| USP53 | 1.49E-12 | 0.30982307 | 0.944 | 0.902 | 2.98E-09 | 0 | USP53 |
| RGS16 | 3.39E-12 | 0.3912306 | 0.477 | 0.361 | 6.77E-09 | 0 | RGS16 |
| CCL3 | 4.00E-12 | 0.48525688 | 0.53 | 0.436 | 8.00E-09 | 0 | CCL3 |
| PALD1 | 8.57E-12 | 0.274098 | 0.955 | 0.906 | 1.71E-08 | 0 | PALD1 |
| PLA2G4A | 9.64E-12 | 0.27664821 | 0.875 | 0.85 | 1.93E-08 | 0 | PLA2G4A |
| CSF2RA | 1.32E-11 | 0.3371264 | 0.928 | 0.892 | 2.64E-08 | 0 | CSF2RA |
| GRID2 | 1.98E-11 | 0.38941876 | 0.775 | 0.729 | 3.97E-08 | 0 | GRID2 |
| LAPTM5 | 2.71E-11 | 0.25244545 | 0.864 | 0.803 | 5.42E-08 | 0 | LAPTM5 |
| HS3ST4 | 7.49E-11 | 0.25544518 | 0.972 | 0.927 | 1.50E-07 | 0 | HS3ST4 |
| JUNB | 1.29E-10 | 0.34846548 | 0.762 | 0.771 | 2.57E-07 | 0 | JUNB |
| ARHGAP15 | 7.85E-10 | 0.31407447 | 0.991 | 0.948 | 1.57E-06 | 0 | ARHGAP15 |
| ARHGAP24 | 5.84E-09 | 0.27186076 | 0.998 | 0.984 | 1.17E-05 | 0 | ARHGAP24 |
| DDIT4 | 8.79E-09 | 0.44573576 | 0.495 | 0.413 | 1.76E-05 | 0 | DDIT4 |
| FCGBP | 3.78E-08 | 0.33330247 | 0.604 | 0.534 | 7.56E-05 | 0 | FCGBP |
| SLC8A1 | 8.54E-08 | 0.29584497 | 0.998 | 0.979 | 0.00017073 | 0 | SLC8A1 |
| CD83 | 5.67E-07 | 0.50870053 | 0.59 | 0.56 | 0.00113423 | 0 | CD83 |
| AOAH | 5.90E-07 | 0.27159266 | 0.989 | 0.955 | 0.00117942 | 0 | AOAH |
| GLUL | 7.49E-07 | 0.44015194 | 0.712 | 0.701 | 0.00149892 | 0 | GLUL |
| AL034397.3 | 1.67E-06 | 0.28426937 | 0.693 | 0.685 | 0.00333803 | 0 | AL034397.3 |
| MAML3 | 5.02E-05 | 0.25889946 | 0.997 | 0.965 | 0.10049829 | 0 | MAML3 |
| ANKRD22 | 9.15E-05 | 0.25156018 | 0.434 | 0.373 | 0.1829805 | 0 | ANKRD22 |
| CDKN1A | 0.00028053 | 0.37395301 | 0.476 | 0.401 | 0.56106037 | 0 | CDKN1A |
| FCGR2A | 0.00049363 | 0.33171868 | 0.607 | 0.607 | 0.98726429 | 0 | FCGR2A |
| VEGFA | 0.00106569 | 0.29065885 | 0.59 | 0.602 | 1 | 0 | VEGFA |
| HAMP | 0.00164615 | 0.5122486 | 0.52 | 0.501 | 1 | 0 | HAMP |
| RASGEF1C | 1.72E-74 | 0.47818146 | 0.986 | 0.935 | 3.43E-71 | 1 | RASGEF1C |
| AC008691.1 | 1.36E-73 | 0.50412432 | 0.967 | 0.879 | 2.73E-70 | 1 | AC008691.1 |
| PLXDC2 | 3.33E-73 | 0.33047849 | 1 | 0.991 | 6.66E-70 | 1 | PLXDC2 |
| FRMD4A | 1.70E-68 | 0.37811021 | 0.999 | 0.995 | 3.41E-65 | 1 | FRMD4A |
| ST6GALNAC3 | 2.96E-65 | 0.55426079 | 0.988 | 0.92 | 5.93E-62 | 1 | ST6GALNAC3 |
| AC008014.1 | 2.38E-62 | 0.39332308 | 0.901 | 0.844 | 4.75E-59 | 1 | AC008014.1 |
| P2RY12 | 4.25E-60 | 0.47079125 | 0.988 | 0.905 | 8.50E-57 | 1 | P2RY12 |
| APBB1IP | 2.53E-59 | 0.43371483 | 0.996 | 0.969 | 5.07E-56 | 1 | APBB1IP |
| DOCK8 | 4.99E-55 | 0.35078625 | 1 | 0.972 | 9.97E-52 | 1 | DOCK8 |
| TENM4 | 7.79E-54 | 0.36520123 | 0.99 | 0.93 | 1.56E-50 | 1 | TENM4 |
| LINC02232 | 1.08E-46 | 0.39039353 | 0.994 | 0.965 | 2.16E-43 | 1 | LINC02232 |
| CACNB4 | 4.38E-46 | 0.37502355 | 0.984 | 0.94 | 8.75E-43 | 1 | CACNB4 |
| ITPR2 | 1.01E-44 | 0.31679734 | 0.999 | 0.987 | 2.01E-41 | 1 | ITPR2 |
| HS3ST41 | 1.12E-44 | 0.43173433 | 0.991 | 0.927 | 2.24E-41 | 1 | HS3ST4 |
| PIP5K1B | 1.18E-44 | 0.28001148 | 0.893 | 0.795 | 2.36E-41 | 1 | PIP5K1B |
| IPCEF1 | 1.70E-44 | 0.315455 | 0.979 | 0.917 | 3.41E-41 | 1 | IPCEF1 |
| BLNK1 | 1.72E-44 | 0.38335872 | 0.985 | 0.936 | 3.44E-41 | 1 | BLNK |
| FOXP2 | 2.87E-43 | 0.3815024 | 0.972 | 0.916 | 5.73E-40 | 1 | FOXP2 |
| KCNIP1 | 3.43E-40 | 0.34265009 | 0.953 | 0.882 | 6.85E-37 | 1 | KCNIP1 |
| SH3RF3 | 8.18E-39 | 0.37984905 | 0.967 | 0.875 | 1.64E-35 | 1 | SH3RF3 |
| SRGAP2 | 9.63E-39 | 0.27352428 | 0.999 | 0.978 | 1.93E-35 | 1 | SRGAP2 |
| LINC00578 | 1.10E-38 | 0.26922582 | 0.814 | 0.676 | 2.20E-35 | 1 | LINC00578 |

| | | | | | | | |
|---|---|---|---|---|---|---|---|
| RASSF8 | 3.72E-38 | 0.40785261 | 0.938 | 0.838 | 7.44E-35 | 1 | RASSF8 |
| ALOX5 | 6.77E-38 | 0.29276518 | 0.911 | 0.801 | 1.35E-34 | 1 | ALOX5 |
| IKZF1 | 1.31E-37 | 0.32356005 | 0.956 | 0.852 | 2.61E-34 | 1 | IKZF1 |
| CPED1 | 1.38E-37 | 0.31757928 | 0.971 | 0.916 | 2.75E-34 | 1 | CPED1 |
| CARD11 | 4.28E-36 | 0.269314 | 0.933 | 0.843 | 8.56E-33 | 1 | CARD11 |
| SDK1 | 1.14E-35 | 0.40147566 | 0.961 | 0.881 | 2.28E-32 | 1 | SDK1 |
| ABCC41 | 4.03E-35 | 0.28625357 | 0.994 | 0.955 | 8.07E-32 | 1 | ABCC4 |
| FYB1 | 2.70E-34 | 0.33315474 | 0.993 | 0.953 | 5.40E-31 | 1 | FYB1 |
| PTCHD4 | 2.85E-33 | 0.36257923 | 0.841 | 0.757 | 5.70E-30 | 1 | PTCHD4 |
| PALD11 | 4.10E-33 | 0.27620104 | 0.981 | 0.905 | 8.20E-30 | 1 | PALD1 |
| MARCH3 | 6.96E-33 | 0.2994801 | 0.948 | 0.887 | 1.39E-29 | 1 | 3-Mar |
| SRGAP2B1 | 1.25E-32 | 0.29583736 | 0.98 | 0.909 | 2.50E-29 | 1 | SRGAP2B |
| LRRK1 | 7.66E-31 | 0.32906593 | 0.982 | 0.909 | 1.53E-27 | 1 | LRRK1 |
| SFMBT2 | 2.46E-30 | 0.27353056 | 0.999 | 0.984 | 4.91E-27 | 1 | SFMBT2 |
| MIR646HG | 3.92E-30 | 0.28141061 | 0.984 | 0.948 | 7.85E-27 | 1 | MIR646HG |
| ST6GAL1 | 4.61E-30 | 0.29791239 | 0.998 | 0.975 | 9.23E-27 | 1 | ST6GAL1 |
| LHFPL2 | 7.56E-30 | 0.27913421 | 0.985 | 0.922 | 1.51E-26 | 1 | LHFPL2 |
| ARHGAP241 | 9.50E-30 | 0.28041181 | 0.999 | 0.985 | 1.90E-26 | 1 | ARHGAP24 |
| FLI1 | 2.76E-29 | 0.29417939 | 0.986 | 0.929 | 5.52E-26 | 1 | FLI1 |
| ENTPD1 | 9.63E-29 | 0.31841246 | 0.984 | 0.945 | 1.93E-25 | 1 | ENTPD1 |
| TNS3 | 2.18E-28 | 0.30290898 | 0.974 | 0.876 | 4.35E-25 | 1 | TNS3 |
| DIAPH2 | 9.67E-28 | 0.28677252 | 0.981 | 0.923 | 1.93E-24 | 1 | DIAPH2 |
| AC008050.1 | 5.06E-27 | 0.26935515 | 0.778 | 0.658 | 1.01E-23 | 1 | AC008050.1 |
| OXR1 | 5.97E-27 | 0.31171171 | 0.994 | 0.98 | 1.19E-23 | 1 | OXR1 |
| ADAM281 | 1.15E-26 | 0.25909404 | 0.998 | 0.976 | 2.29E-23 | 1 | ADAM28 |
| XACT | 3.68E-26 | 0.26952568 | 0.848 | 0.776 | 7.37E-23 | 1 | XACT |
| DSCAM | 2.25E-25 | 0.32103403 | 0.97 | 0.936 | 4.50E-22 | 1 | DSCAM |
| DISC1 | 1.20E-24 | 0.25592634 | 0.997 | 0.962 | 2.41E-21 | 1 | DISC1 |
| DISP1 | 6.85E-24 | 0.34959638 | 0.813 | 0.698 | 1.37E-20 | 1 | DISP1 |
| MAML2 | 1.12E-23 | 0.27882639 | 0.996 | 0.967 | 2.24E-20 | 1 | MAML2 |
| CCDC261 | 1.00E-15 | 0.2579485 | 0.9 | 0.828 | 2.00E-12 | 1 | CCDC26 |
| PLXDC21 | 6.45E-78 | 0.32759367 | 1 | 0.991 | 1.29E-74 | 2 | PLXDC2 |
| IPCEF11 | 1.94E-64 | 0.51288983 | 0.988 | 0.915 | 3.88E-61 | 2 | IPCEF1 |
| P2RY121 | 2.42E-57 | 0.28605864 | 0.991 | 0.904 | 4.84E-54 | 2 | P2RY12 |
| APBB1IP1 | 7.41E-49 | 0.34336906 | 0.999 | 0.969 | 1.48E-45 | 2 | APBB1IP |
| DOCK81 | 9.93E-32 | 0.25195005 | 0.998 | 0.973 | 1.99E-28 | 2 | DOCK8 |
| ST6GALNAC3 | 5.25E-30 | 0.25323337 | 0.994 | 0.919 | 1.05E-26 | 2 | ST6GALNAC3 |
| ANTXR2 | 2.30E-140 | 0.26139909 | 0.773 | 0.577 | 4.60E-137 | 3 | ANTXR2 |
| TMEM178B | 7.77E-129 | 0.27435896 | 0.572 | 0.833 | 1.55E-125 | 3 | TMEM178B |
| SYNPO2 | 9.07E-109 | 0.26249937 | 0.78 | 0.802 | 1.81E-105 | 3 | SYNPO2 |
| BASP1-AS1 | 7.63E-104 | 0.31728482 | 0.816 | 0.791 | 1.53E-100 | 3 | BASP1-AS1 |
| PAM | 2.32E-103 | 0.65785841 | 0.801 | 0.596 | 4.64E-100 | 3 | PAM |
| PDE7B | 8.76E-94 | 0.28286572 | 0.33 | 0.667 | 1.75E-90 | 3 | PDE7B |
| LUZP2 | 3.83E-90 | 0.39967003 | 0.824 | 0.788 | 7.66E-87 | 3 | LUZP2 |
| NWD2 | 1.03E-86 | 0.34778886 | 0.78 | 0.847 | 2.06E-83 | 3 | NWD2 |
| LINC01322 | 1.03E-83 | 0.34135379 | 0.777 | 0.727 | 2.06E-80 | 3 | LINC01322 |
| BANK1 | 1.61E-83 | 0.27399446 | 0.765 | 0.869 | 3.22E-80 | 3 | BANK1 |
| NKAIN3 | 7.71E-78 | 0.36395112 | 0.813 | 0.803 | 1.54E-74 | 3 | NKAIN3 |
| EFNA5 | 6.11E-76 | 0.2714783 | 0.689 | 0.877 | 1.22E-72 | 3 | EFNA5 |
| CDH10 | 3.28E-71 | 0.47851285 | 0.791 | 0.769 | 6.57E-68 | 3 | CDH10 |

| | | | | | | | |
|---|---|---|---|---|---|---|---|
| CCK | 6.23E-68 | 0.46392977 | 0.722 | 0.681 | 1.25E-64 | 3 | CCK |
| AQP4 | 8.99E-63 | 0.32451741 | 0.787 | 0.79 | 1.80E-59 | 3 | AQP4 |
| TNR | 4.89E-60 | 0.3876401 | 0.502 | 0.798 | 9.77E-57 | 3 | TNR |
| GPC6 | 3.03E-59 | 0.42484427 | 0.783 | 0.778 | 6.06E-56 | 3 | GPC6 |
| CLSTN2 | 6.31E-59 | 0.33218014 | 0.737 | 0.632 | 1.26E-55 | 3 | CLSTN2 |
| ELAVL2 | 1.41E-56 | 0.25283868 | 0.744 | 0.772 | 2.81E-53 | 3 | ELAVL2 |
| CBLN2 | 7.83E-53 | 0.32008531 | 0.716 | 0.614 | 1.57E-49 | 3 | CBLN2 |
| ASTN2 | 3.50E-52 | 0.30743438 | 0.44 | 0.672 | 7.00E-49 | 3 | ASTN2 |
| AL117329.1 | 4.24E-52 | 0.33729782 | 0.401 | 0.652 | 8.48E-49 | 3 | AL117329.1 |
| NR2F1-AS1 | 2.36E-51 | 0.32334116 | 0.788 | 0.752 | 4.73E-48 | 3 | NR2F1-AS1 |
| OLFM3 | 6.09E-47 | 0.2768378 | 0.575 | 0.799 | 1.22E-43 | 3 | OLFM3 |
| CDH4 | 1.24E-45 | 0.32707711 | 0.724 | 0.632 | 2.47E-42 | 3 | CDH4 |
| RORB | 1.98E-44 | 0.27156031 | 0.742 | 0.683 | 3.96E-41 | 3 | RORB |
| PCSK2 | 1.29E-41 | 0.34939199 | 0.814 | 0.829 | 2.58E-38 | 3 | PCSK2 |
| STXBP5L | 1.57E-41 | 0.88418196 | 0.863 | 0.887 | 3.14E-38 | 3 | STXBP5L |
| GALNT17 | 3.05E-40 | 0.26462286 | 0.693 | 0.82 | 6.10E-37 | 3 | GALNT17 |
| CCSER1 | 8.82E-40 | 0.69849043 | 0.84 | 0.908 | 1.76E-36 | 3 | CCSER1 |
| EBF1 | 2.41E-39 | 0.34831225 | 0.641 | 0.534 | 4.83E-36 | 3 | EBF1 |
| DAB1 | 4.17E-38 | 0.37523384 | 0.748 | 0.883 | 8.34E-35 | 3 | DAB1 |
| FAM155A | 6.50E-38 | 1.1309302 | 0.883 | 0.903 | 1.30E-34 | 3 | FAM155A |
| RBFOX3 | 8.75E-37 | 0.42455799 | 0.544 | 0.745 | 1.75E-33 | 3 | RBFOX3 |
| TRPM3 | 1.85E-36 | 0.25489825 | 0.74 | 0.829 | 3.71E-33 | 3 | TRPM3 |
| GRIP1 | 2.40E-36 | 0.50442493 | 0.78 | 0.748 | 4.81E-33 | 3 | GRIP1 |
| HS3ST2 | 1.63E-35 | 0.31418183 | 0.64 | 0.856 | 3.26E-32 | 3 | HS3ST2 |
| GRM7 | 3.48E-35 | 0.53957012 | 0.733 | 0.862 | 6.96E-32 | 3 | GRM7 |
| AC007563.2 | 6.78E-35 | 0.27173071 | 0.631 | 0.813 | 1.36E-31 | 3 | AC007563.2 |
| ADAM23 | 7.12E-35 | 0.3199857 | 0.689 | 0.653 | 1.42E-31 | 3 | ADAM23 |
| CHN1 | 1.52E-34 | 0.73200819 | 0.758 | 0.657 | 3.04E-31 | 3 | CHN1 |
| XACT1 | 3.75E-34 | 0.28636127 | 0.712 | 0.8 | 7.50E-31 | 3 | XACT |
| XKR4 | 4.77E-34 | 0.52307632 | 0.678 | 0.569 | 9.54E-31 | 3 | XKR4 |
| PTPRM | 1.05E-33 | 0.25901082 | 0.552 | 0.715 | 2.10E-30 | 3 | PTPRM |
| GRIK2 | 6.20E-33 | 0.6120934 | 0.872 | 0.923 | 1.24E-29 | 3 | GRIK2 |
| PMEPA1 | 8.09E-33 | 0.28340909 | 0.483 | 0.671 | 1.62E-29 | 3 | PMEPA1 |
| GALNTL6 | 1.39E-32 | 0.40113786 | 0.571 | 0.733 | 2.78E-29 | 3 | GALNTL6 |
| DAPK1 | 2.07E-32 | 0.25198064 | 0.763 | 0.728 | 4.15E-29 | 3 | DAPK1 |
| DGKB | 1.14E-31 | 0.5184178 | 0.805 | 0.854 | 2.28E-28 | 3 | DGKB |
| STAT4 | 1.59E-31 | 0.28454434 | 0.606 | 0.62 | 3.19E-28 | 3 | STAT4 |
| PARP14 | 4.32E-31 | 0.28847657 | 0.817 | 0.89 | 8.65E-28 | 3 | PARP14 |
| KCNK13 | 4.78E-31 | 0.30350484 | 0.626 | 0.793 | 9.56E-28 | 3 | KCNK13 |
| ARPP21 | 7.38E-31 | 0.37036377 | 0.757 | 0.836 | 1.48E-27 | 3 | ARPP21 |
| GRM1 | 1.30E-30 | 0.40569805 | 0.787 | 0.81 | 2.60E-27 | 3 | GRM1 |
| AQP4-AS1 | 8.55E-30 | 0.27295435 | 0.749 | 0.759 | 1.71E-26 | 3 | AQP4-AS1 |
| RYR2 | 1.05E-29 | 0.64129569 | 0.682 | 0.852 | 2.09E-26 | 3 | RYR2 |
| KCNH7 | 1.61E-28 | 0.37038934 | 0.688 | 0.803 | 3.21E-25 | 3 | KCNH7 |
| LMO3 | 2.39E-28 | 0.27766498 | 0.722 | 0.861 | 4.78E-25 | 3 | LMO3 |
| KCNIP4 | 2.84E-28 | 1.33003356 | 0.975 | 0.982 | 5.67E-25 | 3 | KCNIP4 |
| WDFY4 | 8.03E-28 | 0.28240019 | 0.813 | 0.881 | 1.61E-24 | 3 | WDFY4 |
| PDE8B | 1.75E-27 | 0.29661636 | 0.686 | 0.57 | 3.49E-24 | 3 | PDE8B |
| GRIN2B | 1.87E-27 | 0.33614589 | 0.713 | 0.861 | 3.74E-24 | 3 | GRIN2B |
| XYLT1 | 2.18E-27 | 0.34685698 | 0.759 | 0.831 | 4.35E-24 | 3 | XYLT1 |

| | | | | | | | |
|---|---|---|---|---|---|---|---|
| ADGRV1 | 1.63E-26 | 0.40352661 | 0.845 | 0.877 | 3.26E-23 | 3 | ADGRV1 |
| CDH13 | 2.15E-26 | 0.2990866 | 0.553 | 0.696 | 4.30E-23 | 3 | CDH13 |
| GRIA3 | 2.17E-26 | 0.5577237 | 0.803 | 0.783 | 4.33E-23 | 3 | GRIA3 |
| CNTN3 | 6.10E-26 | 0.36273993 | 0.693 | 0.864 | 1.22E-22 | 3 | CNTN3 |
| CDHR3 | 1.09E-25 | 0.27036753 | 0.701 | 0.588 | 2.17E-22 | 3 | CDHR3 |
| HCN1 | 1.39E-25 | 0.48342547 | 0.789 | 0.77 | 2.78E-22 | 3 | HCN1 |
| UNC13C | 2.09E-25 | 0.34358821 | 0.81 | 0.81 | 4.19E-22 | 3 | UNC13C |
| TMEM108 | 2.42E-25 | 0.41293433 | 0.696 | 0.844 | 4.84E-22 | 3 | TMEM108 |
| RAB3C | 4.10E-25 | 0.34075335 | 0.57 | 0.708 | 8.21E-22 | 3 | RAB3C |
| SYNPR | 4.99E-25 | 0.38429129 | 0.72 | 0.666 | 9.97E-22 | 3 | SYNPR |
| CNKSR2 | 6.91E-25 | 0.51206884 | 0.733 | 0.81 | 1.38E-21 | 3 | CNKSR2 |
| RIN3 | 7.32E-25 | 0.27108059 | 0.756 | 0.881 | 1.46E-21 | 3 | RIN3 |
| MTUS2 | 9.60E-25 | 0.31226401 | 0.78 | 0.842 | 1.92E-21 | 3 | MTUS2 |
| KCTD16 | 1.57E-24 | 0.43192333 | 0.619 | 0.825 | 3.14E-21 | 3 | KCTD16 |
| RGS7 | 2.47E-24 | 0.57089072 | 0.815 | 0.888 | 4.94E-21 | 3 | RGS7 |
| PHACTR1 | 4.25E-24 | 0.48009771 | 0.822 | 0.916 | 8.51E-21 | 3 | PHACTR1 |
| LRRTM4 | 4.70E-24 | 1.05186773 | 0.899 | 0.924 | 9.39E-21 | 3 | LRRTM4 |
| SORBS2 | 5.18E-24 | 0.54780194 | 0.826 | 0.839 | 1.04E-20 | 3 | SORBS2 |
| KIAA1217 | 7.06E-24 | 0.29363647 | 0.528 | 0.676 | 1.41E-20 | 3 | KIAA1217 |
| MGAT4C | 3.88E-23 | 0.41894961 | 0.65 | 0.8 | 7.77E-20 | 3 | MGAT4C |
| CD831 | 6.85E-23 | 0.37373303 | 0.419 | 0.59 | 1.37E-19 | 3 | CD83 |
| AC008050.11 | 4.05E-22 | 0.31475192 | 0.51 | 0.704 | 8.11E-19 | 3 | AC008050.1 |
| GUCY1A2 | 2.25E-21 | 0.41127641 | 0.722 | 0.732 | 4.50E-18 | 3 | GUCY1A2 |
| SOX5 | 3.75E-21 | 0.50385956 | 0.792 | 0.843 | 7.51E-18 | 3 | SOX5 |
| SNTG1 | 4.01E-21 | 0.62092924 | 0.725 | 0.838 | 8.02E-18 | 3 | SNTG1 |
| KCTD8 | 4.51E-21 | 0.3306618 | 0.591 | 0.515 | 9.02E-18 | 3 | KCTD8 |
| MIR646HG1 | 6.46E-21 | 0.30940934 | 0.9 | 0.963 | 1.29E-17 | 3 | MIR646HG |
| CUX2 | 7.07E-21 | 0.33372022 | 0.674 | 0.683 | 1.41E-17 | 3 | CUX2 |
| PTPRG | 3.06E-20 | 0.321893 | 0.415 | 0.614 | 6.12E-17 | 3 | PTPRG |
| SRRM4 | 5.16E-20 | 0.40158068 | 0.688 | 0.712 | 1.03E-16 | 3 | SRRM4 |
| KCTD1 | 8.37E-20 | 0.28136509 | 0.804 | 0.823 | 1.67E-16 | 3 | KCTD1 |
| ADGRL2 | 8.73E-20 | 0.54792665 | 0.787 | 0.753 | 1.75E-16 | 3 | ADGRL2 |
| LINC01378 | 1.11E-19 | 0.30166897 | 0.812 | 0.847 | 2.22E-16 | 3 | LINC01378 |
| CCDC3 | 1.48E-19 | 0.33650201 | 0.652 | 0.669 | 2.95E-16 | 3 | CCDC3 |
| RGS161 | 1.49E-19 | 0.25331858 | 0.226 | 0.413 | 2.98E-16 | 3 | RGS16 |
| CACNA1B | 1.61E-19 | 0.36092059 | 0.603 | 0.732 | 3.22E-16 | 3 | CACNA1B |
| ROBO2 | 1.94E-19 | 1.08752776 | 0.863 | 0.871 | 3.87E-16 | 3 | ROBO2 |
| RBFOX1 | 2.63E-19 | 1.21637226 | 0.936 | 0.946 | 5.26E-16 | 3 | RBFOX1 |
| CABP1 | 7.14E-19 | 0.3489595 | 0.777 | 0.845 | 1.43E-15 | 3 | CABP1 |
| KCNMB2 | 8.73E-19 | 0.30225228 | 0.749 | 0.844 | 1.75E-15 | 3 | KCNMB2 |
| GABRG3 | 1.54E-18 | 0.31664738 | 0.624 | 0.792 | 3.07E-15 | 3 | GABRG3 |
| MEG3 | 1.80E-18 | 1.15247603 | 0.937 | 0.963 | 3.61E-15 | 3 | MEG3 |
| PRKAG2 | 2.86E-18 | 0.37613907 | 0.742 | 0.863 | 5.72E-15 | 3 | PRKAG2 |
| ZNF385B | 2.89E-18 | 0.46534505 | 0.793 | 0.835 | 5.77E-15 | 3 | ZNF385B |
| SNAP25 | 3.65E-18 | 0.82086576 | 0.793 | 0.749 | 7.29E-15 | 3 | SNAP25 |
| ABLIM1 | 1.24E-17 | 0.46765388 | 0.732 | 0.809 | 2.48E-14 | 3 | ABLIM1 |
| CTNNA2 | 1.55E-17 | 0.73049475 | 0.819 | 0.907 | 3.11E-14 | 3 | CTNNA2 |
| CNTN4 | 2.08E-17 | 0.31652821 | 0.577 | 0.747 | 4.16E-14 | 3 | CNTN4 |
| CDH18 | 3.11E-17 | 0.69400458 | 0.755 | 0.846 | 6.23E-14 | 3 | CDH18 |
| NELL2 | 3.33E-17 | 0.76679774 | 0.832 | 0.798 | 6.66E-14 | 3 | NELL2 |

| | | | | | | | |
|---|---|---|---|---|---|---|---|
| AC084816.1 | 5.60E-17 | 0.29557362 | 0.398 | 0.304 | 1.12E-13 | 3 | AC084816.1 |
| KIAA1549L | 6.92E-17 | 0.48552919 | 0.742 | 0.788 | 1.38E-13 | 3 | KIAA1549L |
| RIPOR2 | 1.26E-16 | 0.34472966 | 0.702 | 0.665 | 2.52E-13 | 3 | RIPOR2 |
| GRIN2A | 1.34E-16 | 0.46257352 | 0.732 | 0.84 | 2.68E-13 | 3 | GRIN2A |
| MCTP1 | 4.01E-16 | 0.46339789 | 0.622 | 0.726 | 8.02E-13 | 3 | MCTP1 |
| GABRB3 | 4.32E-16 | 0.57188553 | 0.834 | 0.836 | 8.63E-13 | 3 | GABRB3 |
| GFAP | 5.37E-16 | 0.35845335 | 0.618 | 0.567 | 1.07E-12 | 3 | GFAP |
| PECAM1 | 6.85E-16 | 0.25638932 | 0.797 | 0.793 | 1.37E-12 | 3 | PECAM1 |
| PTPRZ1 | 1.26E-15 | 0.60189288 | 0.672 | 0.672 | 2.53E-12 | 3 | PTPRZ1 |
| NELL1 | 2.42E-15 | 0.50153256 | 0.79 | 0.799 | 4.85E-12 | 3 | NELL1 |
| VSNL1 | 5.67E-15 | 0.38559154 | 0.629 | 0.718 | 1.13E-11 | 3 | VSNL1 |
| CDH12 | 6.65E-15 | 0.60572667 | 0.812 | 0.854 | 1.33E-11 | 3 | CDH12 |
| SLIT3 | 6.99E-15 | 0.25562628 | 0.773 | 0.814 | 1.40E-11 | 3 | SLIT3 |
| FGFR1 | 7.78E-15 | 0.27782805 | 0.419 | 0.339 | 1.56E-11 | 3 | FGFR1 |
| SFXN5 | 4.73E-14 | 0.37473973 | 0.699 | 0.578 | 9.46E-11 | 3 | SFXN5 |
| ADCY2 | 7.77E-14 | 0.51131655 | 0.765 | 0.819 | 1.55E-10 | 3 | ADCY2 |
| GABRA1 | 1.34E-13 | 0.39419742 | 0.67 | 0.75 | 2.68E-10 | 3 | GABRA1 |
| MYRIP | 1.83E-13 | 0.2501363 | 0.552 | 0.682 | 3.67E-10 | 3 | MYRIP |
| ATRNL1 | 5.15E-13 | 0.95550624 | 0.889 | 0.9 | 1.03E-09 | 3 | ATRNL1 |
| GRIK1 | 8.47E-13 | 0.53233949 | 0.636 | 0.565 | 1.69E-09 | 3 | GRIK1 |
| PDZRN4 | 1.05E-12 | 0.30154255 | 0.542 | 0.749 | 2.11E-09 | 3 | PDZRN4 |
| PTGDS | 2.18E-12 | 0.54346568 | 0.693 | 0.641 | 4.36E-09 | 3 | PTGDS |
| AC011287.1 | 6.68E-12 | 0.36780184 | 0.694 | 0.816 | 1.34E-08 | 3 | AC011287.1 |
| OPCML | 7.90E-12 | 1.09488813 | 0.894 | 0.908 | 1.58E-08 | 3 | OPCML |
| TMEM1631 | 2.36E-11 | 0.26151757 | 0.23 | 0.435 | 4.73E-08 | 3 | TMEM163 |
| SYT1 | 1.28E-10 | 1.11864452 | 0.899 | 0.941 | 2.56E-07 | 3 | SYT1 |
| NECAB1 | 1.79E-10 | 0.25018929 | 0.751 | 0.737 | 3.59E-07 | 3 | NECAB1 |
| SYBU | 1.97E-10 | 0.27364916 | 0.631 | 0.756 | 3.95E-07 | 3 | SYBU |
| CPM | 2.40E-10 | 0.35252722 | 0.514 | 0.655 | 4.80E-07 | 3 | CPM |
| PRKG1 | 4.69E-10 | 0.43913588 | 0.768 | 0.871 | 9.37E-07 | 3 | PRKG1 |
| TMEM132D | 7.22E-10 | 0.66616035 | 0.828 | 0.834 | 1.44E-06 | 3 | TMEM132D |
| PDE1A | 1.16E-09 | 0.48724937 | 0.796 | 0.849 | 2.32E-06 | 3 | PDE1A |
| MICAL2 | 1.18E-09 | 0.38000434 | 0.654 | 0.606 | 2.35E-06 | 3 | MICAL2 |
| FAM135B | 1.21E-09 | 0.2571568 | 0.641 | 0.732 | 2.42E-06 | 3 | FAM135B |
| RGS6 | 1.27E-09 | 0.38234301 | 0.769 | 0.847 | 2.53E-06 | 3 | RGS6 |
| OPHN1 | 1.31E-09 | 0.46699348 | 0.595 | 0.731 | 2.62E-06 | 3 | OPHN1 |
| MEG8 | 1.81E-09 | 0.50388493 | 0.799 | 0.826 | 3.62E-06 | 3 | MEG8 |
| EYA2 | 2.32E-09 | 0.32108121 | 0.612 | 0.652 | 4.65E-06 | 3 | EYA2 |
| PVT1 | 2.50E-09 | 0.27748507 | 0.466 | 0.592 | 5.00E-06 | 3 | PVT1 |
| IQGAP2 | 4.29E-09 | 0.48979404 | 0.72 | 0.726 | 8.59E-06 | 3 | IQGAP2 |
| PCLO | 5.72E-09 | 0.37297614 | 0.755 | 0.815 | 1.14E-05 | 3 | PCLO |
| CACNA1E | 5.77E-09 | 0.38176989 | 0.679 | 0.701 | 1.15E-05 | 3 | CACNA1E |
| NCALD | 6.34E-09 | 0.50306312 | 0.53 | 0.668 | 1.27E-05 | 3 | NCALD |
| SYN2 | 6.72E-09 | 0.48731921 | 0.745 | 0.763 | 1.34E-05 | 3 | SYN2 |
| GPM6A | 6.94E-09 | 1.18211221 | 0.804 | 0.797 | 1.39E-05 | 3 | GPM6A |
| AC092691.1 | 1.36E-08 | 0.53067669 | 0.678 | 0.773 | 2.72E-05 | 3 | AC092691.1 |
| MARCKS | 2.12E-08 | 0.36838713 | 0.767 | 0.792 | 4.24E-05 | 3 | MARCKS |
| HSPB1 | 2.12E-08 | 0.36665469 | 0.656 | 0.606 | 4.24E-05 | 3 | HSPB1 |
| FAT3 | 5.95E-08 | 0.33997087 | 0.829 | 0.871 | 0.00011891 | 3 | FAT3 |
| RASGRF2 | 6.39E-08 | 0.35024464 | 0.742 | 0.775 | 0.00012776 | 3 | RASGRF2 |

| | | | | | | | |
|---|---|---|---|---|---|---|---|
| FGF12 | 9.47E-08 | 0.79061508 | 0.786 | 0.808 | 0.00018943 | 3 | FGF12 |
| PDE4D | 1.39E-07 | 0.63813275 | 0.922 | 0.969 | 0.00027826 | 3 | PDE4D |
| ANKRD30BL | 1.76E-07 | 0.3512601 | 0.581 | 0.77 | 0.00035119 | 3 | ANKRD30BL |
| PCDH7 | 1.86E-07 | 0.63900274 | 0.838 | 0.861 | 0.00037192 | 3 | PCDH7 |
| CNTNAP2 | 2.35E-07 | 0.95229498 | 0.887 | 0.905 | 0.00046926 | 3 | CNTNAP2 |
| KAZN | 3.07E-07 | 0.68722059 | 0.68 | 0.804 | 0.00061467 | 3 | KAZN |
| LDB2 | 3.94E-07 | 0.52050852 | 0.843 | 0.889 | 0.00078824 | 3 | LDB2 |
| CIT | 4.24E-07 | 0.30438328 | 0.694 | 0.79 | 0.00084872 | 3 | CIT |
| ATP8A2 | 6.46E-07 | 0.56781834 | 0.707 | 0.829 | 0.00129126 | 3 | ATP8A2 |
| DLGAP2 | 7.23E-07 | 0.69044807 | 0.823 | 0.905 | 0.00144671 | 3 | DLGAP2 |
| LHFPL3 | 9.39E-07 | 0.470367 | 0.712 | 0.675 | 0.00187834 | 3 | LHFPL3 |
| RIMS2 | 1.43E-06 | 0.86417744 | 0.734 | 0.817 | 0.00286189 | 3 | RIMS2 |
| MED12L | 1.44E-06 | 0.39953592 | 0.561 | 0.632 | 0.00288187 | 3 | MED12L |
| EPHA6 | 2.53E-06 | 0.58039437 | 0.595 | 0.776 | 0.00505633 | 3 | EPHA6 |
| ZNF385D | 2.72E-06 | 0.45532949 | 0.432 | 0.516 | 0.00544525 | 3 | ZNF385D |
| LRRC4C | 3.41E-06 | 0.43360218 | 0.794 | 0.819 | 0.00682328 | 3 | LRRC4C |
| ZMAT4 | 3.88E-06 | 0.38775339 | 0.72 | 0.736 | 0.00775769 | 3 | ZMAT4 |
| TOX | 4.12E-06 | 0.32095848 | 0.636 | 0.687 | 0.00823114 | 3 | TOX |
| PCDH15 | 6.56E-06 | 0.39848415 | 0.654 | 0.747 | 0.01311464 | 3 | PCDH15 |
| C31 | 7.38E-06 | 0.35979466 | 0.83 | 0.947 | 0.01476396 | 3 | C3 |
| SLC4A4 | 8.37E-06 | 0.30524975 | 0.625 | 0.661 | 0.0167471 | 3 | SLC4A4 |
| CHL1 | 9.15E-06 | 0.41142283 | 0.754 | 0.831 | 0.01830949 | 3 | CHL1 |
| AC073050.1 | 9.38E-06 | 0.41842775 | 0.803 | 0.832 | 0.01875945 | 3 | AC073050.1 |
| NIPAL2 | 1.52E-05 | 0.34722482 | 0.84 | 0.807 | 0.03041063 | 3 | NIPAL2 |
| SYN3 | 2.42E-05 | 0.56304397 | 0.725 | 0.8 | 0.04841915 | 3 | SYN3 |
| SCN1A | 2.74E-05 | 0.40944193 | 0.545 | 0.684 | 0.05487945 | 3 | SCN1A |
| MDGA2 | 4.04E-05 | 0.74897693 | 0.853 | 0.866 | 0.08075042 | 3 | MDGA2 |
| CLU | 5.10E-05 | 0.90007672 | 0.735 | 0.743 | 0.10204838 | 3 | CLU |
| ASIC2 | 5.84E-05 | 0.5303457 | 0.732 | 0.781 | 0.11689707 | 3 | ASIC2 |
| CDH9 | 6.72E-05 | 0.3343378 | 0.754 | 0.788 | 0.13438938 | 3 | CDH9 |
| EPSTI1 | 7.81E-05 | 0.26948576 | 0.772 | 0.734 | 0.15629091 | 3 | EPSTI1 |
| SV2B | 9.99E-05 | 0.26883339 | 0.765 | 0.834 | 0.19983392 | 3 | SV2B |
| HS6ST3 | 0.00010991 | 0.61596464 | 0.805 | 0.864 | 0.21981132 | 3 | HS6ST3 |
| GRIA1 | 0.00018643 | 0.35294055 | 0.75 | 0.817 | 0.37286901 | 3 | GRIA1 |
| CADPS | 0.00023751 | 0.52136501 | 0.839 | 0.884 | 0.47501069 | 3 | CADPS |
| NRCAM | 0.00024993 | 0.81085239 | 0.762 | 0.782 | 0.49985812 | 3 | NRCAM |
| TMEM132B | 0.00027172 | 0.45144495 | 0.787 | 0.787 | 0.54344731 | 3 | TMEM132B |
| GABBR2 | 0.00038459 | 0.46938879 | 0.692 | 0.748 | 0.76918403 | 3 | GABBR2 |
| MMP16 | 0.00045672 | 0.36545229 | 0.65 | 0.777 | 0.91344677 | 3 | MMP16 |
| BMPR1B | 0.00055884 | 0.25583195 | 0.504 | 0.672 | 1 | 3 | BMPR1B |
| CACNA2D3 | 0.00059895 | 0.51013888 | 0.785 | 0.826 | 1 | 3 | CACNA2D3 |
| ESRRG | 0.00063069 | 0.25607869 | 0.455 | 0.563 | 1 | 3 | ESRRG |
| CNTN5 | 0.00115784 | 0.63781239 | 0.821 | 0.83 | 1 | 3 | CNTN5 |
| DPP10 | 0.00145654 | 0.83412374 | 0.802 | 0.884 | 1 | 3 | DPP10 |
| AFF3 | 0.00149544 | 0.49005092 | 0.786 | 0.861 | 1 | 3 | AFF3 |
| BRINP3 | 0.00153103 | 0.28955486 | 0.733 | 0.775 | 1 | 3 | BRINP3 |
| PTCHD1-AS | 0.00191356 | 0.29233237 | 0.71 | 0.818 | 1 | 3 | PTCHD1-AS |
| ERC2 | 0.00261675 | 0.53823474 | 0.705 | 0.756 | 1 | 3 | ERC2 |
| PTPRT | 0.0027783 | 0.38473828 | 0.723 | 0.792 | 1 | 3 | PTPRT |
| RALYL | 0.00586741 | 0.97978982 | 0.851 | 0.888 | 1 | 3 | RALYL |

| | | | | | | | |
|---|---|---|---|---|---|---|---|
| CHRM3 | 0.00627934 | 0.61285419 | 0.73 | 0.755 | 1 | 3 | CHRM3 |
| CPE | 0.00668355 | 0.55812982 | 0.729 | 0.716 | 1 | 3 | CPE |
| ADARB2 | 0.00733858 | 0.27406305 | 0.592 | 0.626 | 1 | 3 | ADARB2 |
| SLC35F3 | 0.00801959 | 0.36749104 | 0.792 | 0.742 | 1 | 3 | SLC35F3 |
| LINC02306 | 0.00847136 | 0.26288405 | 0.697 | 0.797 | 1 | 3 | LINC02306 |
| IQCJ-SCHIP1 | 0.00910895 | 0.77582047 | 0.863 | 0.897 | 1 | 3 | IQCJ-SCHIP1 |
| NPL | 6.51E-54 | 0.74531813 | 0.961 | 0.915 | 1.30E-50 | 4 | NPL |
| NHSL1 | 1.17E-45 | 0.79025028 | 0.875 | 0.759 | 2.34E-42 | 4 | NHSL1 |
| CPM1 | 2.29E-44 | 0.92657685 | 0.755 | 0.618 | 4.57E-41 | 4 | CPM |
| MYO1E | 2.85E-43 | 0.9205184 | 0.741 | 0.587 | 5.71E-40 | 4 | MYO1E |
| MSR1 | 1.11E-40 | 0.69328636 | 0.969 | 0.968 | 2.21E-37 | 4 | MSR1 |
| IQGAP21 | 1.17E-40 | 0.48219333 | 0.763 | 0.72 | 2.34E-37 | 4 | IQGAP2 |
| PDGFC | 2.70E-36 | 0.49849595 | 0.913 | 0.855 | 5.41E-33 | 4 | PDGFC |
| ZNF804A | 1.87E-32 | 0.95831012 | 0.754 | 0.746 | 3.74E-29 | 4 | ZNF804A |
| FMN1 | 4.62E-32 | 0.44001084 | 0.84 | 0.723 | 9.23E-29 | 4 | FMN1 |
| RBM47 | 3.30E-31 | 0.44524387 | 0.936 | 0.857 | 6.59E-28 | 4 | RBM47 |
| HRH1 | 5.22E-30 | 0.2933259 | 0.715 | 0.636 | 1.04E-26 | 4 | HRH1 |
| NEAT1 | 6.75E-28 | 0.3088059 | 0.998 | 0.977 | 1.35E-24 | 4 | NEAT1 |
| CD163 | 6.48E-26 | 0.64797732 | 0.599 | 0.472 | 1.30E-22 | 4 | CD163 |
| EYA21 | 2.00E-25 | 0.59622233 | 0.686 | 0.641 | 3.99E-22 | 4 | EYA2 |
| SAMD4A | 1.97E-24 | 0.50098535 | 0.878 | 0.821 | 3.94E-21 | 4 | SAMD4A |
| GPNMB | 2.14E-24 | 0.8288887 | 0.711 | 0.657 | 4.29E-21 | 4 | GPNMB |
| FRMD4B | 2.23E-23 | 0.3758995 | 0.905 | 0.833 | 4.46E-20 | 4 | FRMD4B |
| MAML21 | 4.31E-23 | 0.34178628 | 0.996 | 0.968 | 8.63E-20 | 4 | MAML2 |
| PPARG | 5.80E-22 | 0.62557441 | 0.631 | 0.545 | 1.16E-18 | 4 | PPARG |
| ITGAX | 7.17E-22 | 0.47364065 | 0.958 | 0.913 | 1.43E-18 | 4 | ITGAX |
| PIK3R5 | 1.37E-21 | 0.39451616 | 0.869 | 0.796 | 2.73E-18 | 4 | PIK3R5 |
| TFEC | 7.87E-21 | 0.3083658 | 0.892 | 0.825 | 1.57E-17 | 4 | TFEC |
| PCSK5 | 1.03E-19 | 0.32900755 | 0.829 | 0.742 | 2.07E-16 | 4 | PCSK5 |
| SLC1A3 | 7.43E-19 | 0.25115127 | 0.986 | 0.961 | 1.49E-15 | 4 | SLC1A3 |
| HGF | 8.31E-19 | 0.26639449 | 0.799 | 0.768 | 1.66E-15 | 4 | HGF |
| BNC2 | 8.04E-18 | 0.3587581 | 0.943 | 0.877 | 1.61E-14 | 4 | BNC2 |
| DIRC3 | 1.07E-16 | 0.64708878 | 0.802 | 0.795 | 2.13E-13 | 4 | DIRC3 |
| RGS12 | 2.55E-16 | 0.37171862 | 0.957 | 0.927 | 5.09E-13 | 4 | RGS12 |
| APOE1 | 4.54E-16 | 0.38593947 | 0.921 | 0.887 | 9.09E-13 | 4 | APOE |
| SAT11 | 5.70E-16 | 0.3706051 | 0.917 | 0.883 | 1.14E-12 | 4 | SAT1 |
| MS4A7 | 7.30E-16 | 0.421481 | 0.719 | 0.649 | 1.46E-12 | 4 | MS4A7 |
| GNB4 | 2.34E-15 | 0.28334208 | 0.923 | 0.868 | 4.68E-12 | 4 | GNB4 |
| TPRG1 | 3.48E-15 | 0.97211648 | 0.52 | 0.395 | 6.96E-12 | 4 | TPRG1 |
| PDE3B | 1.81E-14 | 0.38103443 | 0.952 | 0.911 | 3.62E-11 | 4 | PDE3B |
| PTPRG1 | 2.87E-14 | 1.11886943 | 0.628 | 0.581 | 5.73E-11 | 4 | PTPRG |
| MERTK | 1.47E-13 | 0.31610369 | 0.836 | 0.762 | 2.95E-10 | 4 | MERTK |
| LINC01374 | 2.82E-12 | 0.36304484 | 0.963 | 0.927 | 5.63E-09 | 4 | LINC01374 |
| IFI44L | 7.06E-12 | 0.44821231 | 0.714 | 0.677 | 1.41E-08 | 4 | IFI44L |
| CD832 | 8.85E-12 | 0.28046091 | 0.639 | 0.556 | 1.77E-08 | 4 | CD83 |
| BMP2K | 4.94E-11 | 0.27230424 | 0.995 | 0.966 | 9.89E-08 | 4 | BMP2K |
| TNS31 | 9.46E-11 | 0.28785893 | 0.942 | 0.884 | 1.89E-07 | 4 | TNS3 |
| ARHGAP18 | 2.86E-10 | 0.36459583 | 0.658 | 0.64 | 5.72E-07 | 4 | ARHGAP18 |
| MCTP11 | 1.92E-09 | 0.28778094 | 0.725 | 0.709 | 3.83E-06 | 4 | MCTP1 |
| GLDN | 1.78E-08 | 0.56214643 | 0.953 | 0.929 | 3.56E-05 | 4 | GLDN |

| | | | | | | | |
|---|---|---|---|---|---|---|---|
| KLHL6 | 5.30E-08 | 0.32555723 | 0.695 | 0.66 | 0.00010608 | 4 | KLHL6 |
| RASGEF1B | 1.91E-07 | 0.27333192 | 0.709 | 0.664 | 0.00038144 | 4 | RASGEF1B |
| PBX3 | 3.00E-07 | 0.30858036 | 0.813 | 0.787 | 0.00059935 | 4 | PBX3 |
| OLFML2B | 7.69E-06 | 0.27393378 | 0.73 | 0.728 | 0.0153864 | 4 | OLFML2B |
| CPNE8 | 1.39E-05 | 0.26336342 | 0.589 | 0.548 | 0.02787498 | 4 | CPNE8 |
| CHSY3 | 2.66E-05 | 0.26608915 | 0.671 | 0.647 | 0.05314444 | 4 | CHSY3 |
| NAMPT | 5.90E-05 | 0.25285874 | 0.582 | 0.526 | 0.11797164 | 4 | NAMPT |
| WIPF3 | 6.98E-05 | 0.33480515 | 0.789 | 0.744 | 0.13969986 | 4 | WIPF3 |
| CCNG2 | 0.00057328 | 0.25250578 | 0.738 | 0.71 | 1 | 4 | CCNG2 |
| MS4A4A | 0.00175728 | 0.3024899 | 0.547 | 0.522 | 1 | 4 | MS4A4A |
| TENM2 | 3.05E-16 | 0.30608011 | 0.799 | 0.872 | 6.10E-13 | 5 | TENM2 |
| ADGRV11 | 2.24E-14 | 0.25027423 | 0.801 | 0.879 | 4.47E-11 | 5 | ADGRV1 |
| CHST11 | 1.20E-11 | 0.32897013 | 0.975 | 0.962 | 2.41E-08 | 5 | CHST11 |
| KCNQ3 | 1.74E-10 | 0.34534893 | 0.987 | 0.977 | 3.49E-07 | 5 | KCNQ3 |
| MAML31 | 3.34E-10 | 0.49827435 | 0.987 | 0.971 | 6.69E-07 | 5 | MAML3 |
| LRMDA | 1.45E-09 | 0.25557303 | 1 | 0.994 | 2.91E-06 | 5 | LRMDA |
| SFXN51 | 2.70E-09 | 0.26036029 | 0.439 | 0.61 | 5.40E-06 | 5 | SFXN5 |
| SDK11 | 1.58E-08 | 0.59067187 | 0.894 | 0.893 | 3.15E-05 | 5 | SDK1 |
| RUNX1 | 8.45E-07 | 0.3593352 | 0.878 | 0.906 | 0.00168967 | 5 | RUNX1 |
| TBXAS1 | 8.81E-06 | 0.28001616 | 0.964 | 0.967 | 0.01762478 | 5 | TBXAS1 |
| SRGAP21 | 8.84E-06 | 0.33267773 | 0.997 | 0.98 | 0.01767974 | 5 | SRGAP2 |
| RASGEF1C1 | 1.78E-05 | 0.57816263 | 0.93 | 0.944 | 0.03563294 | 5 | RASGEF1C |
| DOCK82 | 5.80E-05 | 0.25580075 | 0.987 | 0.976 | 0.11601263 | 5 | DOCK8 |
| LINC022321 | 9.01E-05 | 0.51595783 | 0.962 | 0.97 | 0.18011845 | 5 | LINC02232 |
| HS3ST42 | 0.0001294 | 0.26214697 | 0.925 | 0.938 | 0.25879017 | 5 | HS3ST4 |
| AOAH1 | 0.00016045 | 0.38815022 | 0.977 | 0.961 | 0.32089542 | 5 | AOAH |
| LINC02057 | 0.00023303 | 0.26958217 | 0.804 | 0.851 | 0.46606752 | 5 | LINC02057 |
| RBM471 | 0.00033953 | 0.29876326 | 0.828 | 0.87 | 0.67906337 | 5 | RBM47 |
| LHFPL21 | 0.00041367 | 0.25201659 | 0.945 | 0.931 | 0.82733682 | 5 | LHFPL2 |
| SFMBT21 | 0.00069641 | 0.25375303 | 1 | 0.985 | 1 | 5 | SFMBT2 |
| LINC013741 | 0.00090392 | 0.43758817 | 0.932 | 0.931 | 1 | 5 | LINC01374 |
| AC087564.1 | 0.00111741 | 0.27389455 | 0.846 | 0.878 | 1 | 5 | AC087564.1 |
| AC012150.1 | 0.00130844 | 0.44490279 | 0.759 | 0.751 | 1 | 5 | AC012150.1 |
| PALD12 | 0.00289142 | 0.31196844 | 0.925 | 0.916 | 1 | 5 | PALD1 |
| CARD111 | 0.00323756 | 0.55053412 | 0.823 | 0.86 | 1 | 5 | CARD11 |
| DOCK2 | 0.00436948 | 0.28936809 | 0.941 | 0.959 | 1 | 5 | DOCK2 |
| MSR11 | 0.00640008 | 0.39427416 | 0.958 | 0.969 | 1 | 5 | MSR1 |
| DISC11 | 0.00903939 | 0.26510922 | 0.964 | 0.967 | 1 | 5 | DISC1 |
| PDE3B1 | 0.00976594 | 0.41767927 | 0.912 | 0.917 | 1 | 5 | PDE3B |
| AC005699.1 | 0.00993097 | 0.28489565 | 0.5 | 0.589 | 1 | 5 | AC005699.1 |
| ST18 | 2.17E-195 | 3.493259 | 0.988 | 0.555 | 4.34E-192 | 6 | ST18 |
| CTNNA3 | 1.94E-191 | 3.37765764 | 0.991 | 0.743 | 3.89E-188 | 6 | CTNNA3 |
| MOBP | 8.96E-178 | 3.14853068 | 0.962 | 0.54 | 1.79E-174 | 6 | MOBP |
| PLP1 | 7.77E-174 | 3.37659052 | 0.971 | 0.471 | 1.55E-170 | 6 | PLP1 |
| RNF220 | 4.74E-173 | 3.20565849 | 0.956 | 0.479 | 9.49E-170 | 6 | RNF220 |
| C10orf90 | 6.00E-167 | 2.71914768 | 0.948 | 0.521 | 1.20E-163 | 6 | C10orf90 |
| TMEM144 | 6.37E-166 | 2.5439205 | 0.977 | 0.668 | 1.27E-162 | 6 | TMEM144 |
| CERCAM | 2.40E-164 | 2.53476318 | 0.953 | 0.582 | 4.81E-161 | 6 | CERCAM |
| BCAS1 | 6.54E-162 | 2.24095919 | 0.95 | 0.614 | 1.31E-158 | 6 | BCAS1 |
| SPOCK3 | 4.39E-161 | 2.62983942 | 0.942 | 0.372 | 8.78E-158 | 6 | SPOCK3 |

| | | | | | | | |
|---|---|---|---|---|---|---|---|
| LINC01608 | 4.92E-157 | 2.58369971 | 0.93 | 0.499 | 9.84E-154 | 6 | LINC01608 |
| PTPRK | 9.78E-152 | 2.58427166 | 0.956 | 0.65 | 1.96E-148 | 6 | PTPRK |
| AK5 | 7.82E-149 | 2.62381999 | 0.95 | 0.762 | 1.56E-145 | 6 | AK5 |
| EPB41L4A | 2.41E-147 | 0.49007517 | 0.866 | 0.185 | 4.81E-144 | 6 | EPB41L4A |
| PTGDS1 | 3.31E-146 | 2.31282557 | 0.962 | 0.634 | 6.62E-143 | 6 | PTGDS |
| GRM3 | 3.82E-143 | 2.46150919 | 0.921 | 0.63 | 7.65E-140 | 6 | GRM3 |
| TF | 4.93E-143 | 2.52608856 | 0.904 | 0.299 | 9.86E-140 | 6 | TF |
| SLCO1A2 | 1.33E-141 | 2.10290945 | 0.901 | 0.473 | 2.66E-138 | 6 | SLCO1A2 |
| ENPP2 | 1.69E-141 | 2.29846377 | 0.91 | 0.395 | 3.37E-138 | 6 | ENPP2 |
| COL4A5 | 1.69E-138 | 2.11146294 | 0.913 | 0.587 | 3.39E-135 | 6 | COL4A5 |
| KCNH8 | 2.88E-136 | 2.45127469 | 0.892 | 0.298 | 5.77E-133 | 6 | KCNH8 |
| CLMN | 1.77E-135 | 2.15023988 | 0.901 | 0.519 | 3.54E-132 | 6 | CLMN |
| MBP | 2.22E-133 | 1.76277379 | 0.985 | 0.792 | 4.43E-130 | 6 | MBP |
| LINC00639 | 1.75E-132 | 2.13012792 | 0.895 | 0.607 | 3.50E-129 | 6 | LINC00639 |
| AC012494.1 | 1.99E-129 | 2.33142072 | 0.869 | 0.394 | 3.98E-126 | 6 | AC012494.1 |
| GALNT13 | 2.32E-123 | 1.66810184 | 0.921 | 0.754 | 4.64E-120 | 6 | GALNT13 |
| ZNF536 | 1.51E-119 | 2.28242123 | 0.883 | 0.566 | 3.03E-116 | 6 | ZNF536 |
| DLC1 | 1.81E-119 | 2.10259805 | 0.892 | 0.562 | 3.63E-116 | 6 | DLC1 |
| PWRN1 | 1.85E-118 | 1.63535539 | 0.895 | 0.779 | 3.70E-115 | 6 | PWRN1 |
| KIF6 | 6.23E-118 | 1.88353363 | 0.869 | 0.58 | 1.25E-114 | 6 | KIF6 |
| APOD | 3.24E-117 | 1.1673208 | 0.918 | 0.62 | 6.48E-114 | 6 | APOD |
| PARD3 | 2.58E-115 | 0.80038752 | 0.915 | 0.661 | 5.16E-112 | 6 | PARD3 |
| AL359091.1 | 3.11E-111 | 1.8942692 | 0.866 | 0.649 | 6.22E-108 | 6 | AL359091.1 |
| UGT8 | 3.59E-111 | 1.94338899 | 0.866 | 0.563 | 7.19E-108 | 6 | UGT8 |
| SCD | 1.69E-109 | 1.55449026 | 0.86 | 0.453 | 3.38E-106 | 6 | SCD |
| CNTNAP4 | 4.08E-105 | 2.15477578 | 0.86 | 0.659 | 8.16E-102 | 6 | CNTNAP4 |
| HHIP | 1.23E-103 | 1.95747644 | 0.851 | 0.579 | 2.46E-100 | 6 | HHIP |
| LINC01170 | 9.67E-103 | 1.76579216 | 0.837 | 0.477 | 1.93E-99 | 6 | LINC01170 |
| LINC02073 | 4.19E-101 | 0.54664619 | 0.86 | 0.746 | 8.37E-98 | 6 | LINC02073 |
| SLC4A41 | 1.10E-98 | 0.36919385 | 0.913 | 0.644 | 2.21E-95 | 6 | SLC4A4 |
| CNP | 2.14E-97 | 1.71398857 | 0.834 | 0.33 | 4.28E-94 | 6 | CNP |
| TMEM132C | 4.72E-97 | 0.82909787 | 0.848 | 0.571 | 9.43E-94 | 6 | TMEM132C |
| ERBB4 | 2.51E-96 | 1.70632073 | 0.933 | 0.788 | 5.01E-93 | 6 | ERBB4 |
| PLPP3 | 5.51E-96 | 0.7487813 | 0.851 | 0.707 | 1.10E-92 | 6 | PLPP3 |
| PCSK6 | 1.28E-95 | 1.83335937 | 0.825 | 0.402 | 2.57E-92 | 6 | PCSK6 |
| ZNF462 | 2.59E-94 | 1.13892699 | 0.837 | 0.626 | 5.18E-91 | 6 | ZNF462 |
| FAM107B | 3.07E-94 | 1.55135869 | 0.86 | 0.591 | 6.15E-91 | 6 | FAM107B |
| GPC5 | 8.96E-93 | 0.9983406 | 0.915 | 0.72 | 1.79E-89 | 6 | GPC5 |
| SHROOM4 | 3.63E-91 | 1.83191045 | 0.822 | 0.543 | 7.25E-88 | 6 | SHROOM4 |
| ABCA8 | 3.96E-90 | 1.76642671 | 0.822 | 0.508 | 7.91E-87 | 6 | ABCA8 |
| MOG | 3.94E-87 | 1.54418175 | 0.799 | 0.366 | 7.88E-84 | 6 | MOG |
| LDLRAD3 | 1.89E-86 | 1.10403113 | 0.819 | 0.48 | 3.77E-83 | 6 | LDLRAD3 |
| ADCY21 | 8.69E-85 | 0.3386991 | 0.924 | 0.807 | 1.74E-81 | 6 | ADCY2 |
| FBXL7 | 1.13E-84 | 2.13490902 | 0.822 | 0.657 | 2.25E-81 | 6 | FBXL7 |
| LMCD1-AS1 | 6.57E-84 | 1.5699466 | 0.904 | 0.82 | 1.31E-80 | 6 | LMCD1-AS1 |
| ATP10B | 4.14E-82 | 1.57933026 | 0.81 | 0.641 | 8.27E-79 | 6 | ATP10B |
| LINC01122 | 7.45E-82 | 0.69161402 | 0.898 | 0.74 | 1.49E-78 | 6 | LINC01122 |
| SOX6 | 1.45E-81 | 0.35213194 | 0.872 | 0.693 | 2.89E-78 | 6 | SOX6 |
| HS3ST5 | 5.24E-81 | 1.74004499 | 0.831 | 0.68 | 1.05E-77 | 6 | HS3ST5 |
| ADARB21 | 1.67E-80 | 1.18951191 | 0.875 | 0.61 | 3.34E-77 | 6 | ADARB2 |

| | | | | | | | |
|---|---|---|---|---|---|---|---|
| DAAM2 | 1.83E-80 | 1.82026286 | 0.808 | 0.633 | 3.67E-77 | 6 | DAAM2 |
| CNDP1 | 6.11E-80 | 1.67806473 | 0.816 | 0.586 | 1.22E-76 | 6 | CNDP1 |
| NECAB11 | 3.48E-79 | 1.37449381 | 0.86 | 0.734 | 6.97E-76 | 6 | NECAB1 |
| LINC01630 | 3.60E-79 | 1.49899667 | 0.802 | 0.573 | 7.20E-76 | 6 | LINC01630 |
| LINC00320 | 1.17E-78 | 1.52111868 | 0.778 | 0.275 | 2.35E-75 | 6 | LINC00320 |
| AC113383.1 | 2.32E-76 | 0.32833181 | 0.828 | 0.692 | 4.63E-73 | 6 | AC113383.1 |
| OPALIN | 1.01E-74 | 1.32294412 | 0.773 | 0.318 | 2.02E-71 | 6 | OPALIN |
| PTPRT1 | 1.22E-72 | 0.31131772 | 0.904 | 0.777 | 2.44E-69 | 6 | PTPRT |
| ADAMTS19 | 3.34E-72 | 0.30963467 | 0.845 | 0.745 | 6.68E-69 | 6 | ADAMTS19 |
| GRIN2A1 | 7.37E-72 | 0.62439135 | 0.945 | 0.82 | 1.47E-68 | 6 | GRIN2A |
| CHODL | 4.43E-70 | 0.27465976 | 0.778 | 0.548 | 8.85E-67 | 6 | CHODL |
| TMEM1081 | 4.52E-70 | 0.30811701 | 0.918 | 0.819 | 9.04E-67 | 6 | TMEM108 |
| STXBP6 | 2.24E-63 | 1.26116723 | 0.764 | 0.537 | 4.49E-60 | 6 | STXBP6 |
| FRMD4B1 | 2.45E-62 | 0.97646197 | 0.971 | 0.837 | 4.91E-59 | 6 | FRMD4B |
| KCNH1 | 2.80E-60 | 0.48605082 | 0.927 | 0.826 | 5.61E-57 | 6 | KCNH1 |
| TENM3 | 2.75E-59 | 0.5489336 | 0.878 | 0.801 | 5.50E-56 | 6 | TENM3 |
| GREB1L | 6.03E-59 | 0.88808594 | 0.805 | 0.803 | 1.21E-55 | 6 | GREB1L |
| PIEZO2 | 4.12E-57 | 1.66188665 | 0.761 | 0.557 | 8.24E-54 | 6 | PIEZO2 |
| LPAR1 | 7.06E-56 | 1.1145249 | 0.942 | 0.834 | 1.41E-52 | 6 | LPAR1 |
| RBMS3 | 8.28E-56 | 0.30266642 | 0.848 | 0.497 | 1.66E-52 | 6 | RBMS3 |
| CA2 | 3.28E-55 | 1.16241683 | 0.72 | 0.379 | 6.56E-52 | 6 | CA2 |
| CRYAB | 5.91E-55 | 1.06006762 | 0.717 | 0.28 | 1.18E-51 | 6 | CRYAB |
| ROBO1 | 1.61E-54 | 1.32875346 | 0.813 | 0.657 | 3.22E-51 | 6 | ROBO1 |
| SLC6A1 | 8.28E-54 | 0.883512 | 0.796 | 0.682 | 1.66E-50 | 6 | SLC6A1 |
| TOX3 | 9.63E-54 | 0.30158443 | 0.752 | 0.58 | 1.93E-50 | 6 | TOX3 |
| PDE1A1 | 1.52E-53 | 1.24614733 | 0.848 | 0.842 | 3.03E-50 | 6 | PDE1A |
| LRP1B | 2.72E-52 | 0.99604883 | 0.953 | 0.909 | 5.43E-49 | 6 | LRP1B |
| PLPP4 | 5.62E-52 | 0.31458324 | 0.72 | 0.501 | 1.12E-48 | 6 | PLPP4 |
| CLDN11 | 7.22E-52 | 1.24282535 | 0.749 | 0.553 | 1.44E-48 | 6 | CLDN11 |
| RYR3 | 2.53E-51 | 0.3046895 | 0.886 | 0.728 | 5.05E-48 | 6 | RYR3 |
| ABTB2 | 7.36E-51 | 0.66941719 | 0.767 | 0.777 | 1.47E-47 | 6 | ABTB2 |
| MAG | 1.18E-50 | 1.15294781 | 0.726 | 0.493 | 2.36E-47 | 6 | MAG |
| ATP8A21 | 2.99E-50 | 0.33166238 | 0.883 | 0.809 | 5.98E-47 | 6 | ATP8A2 |
| AL354809.1 | 1.77E-49 | 1.09290971 | 0.767 | 0.683 | 3.53E-46 | 6 | AL354809.1 |
| AC007364.1 | 9.13E-49 | 0.78235177 | 0.764 | 0.6 | 1.83E-45 | 6 | AC007364.1 |
| AL589740.1 | 1.27E-48 | 0.87775976 | 0.784 | 0.741 | 2.55E-45 | 6 | AL589740.1 |
| ITGA2 | 3.24E-48 | 1.19749415 | 0.691 | 0.343 | 6.49E-45 | 6 | ITGA2 |
| AL133346.1 | 2.63E-47 | 0.3073646 | 0.831 | 0.823 | 5.26E-44 | 6 | AL133346.1 |
| FAT4 | 5.93E-47 | 0.58189642 | 0.711 | 0.466 | 1.19E-43 | 6 | FAT4 |
| CNTNAP21 | 2.42E-46 | 1.03511352 | 0.942 | 0.901 | 4.84E-43 | 6 | CNTNAP2 |
| IFNG-AS1 | 2.88E-46 | 0.33052555 | 0.758 | 0.79 | 5.75E-43 | 6 | IFNG-AS1 |
| SPANXA2-OT | 3.25E-46 | 0.35461338 | 0.726 | 0.523 | 6.51E-43 | 6 | SPANXA2-OT1 |
| SGCD | 5.84E-46 | 0.99697934 | 0.813 | 0.802 | 1.17E-42 | 6 | SGCD |
| LINC00609 | 1.20E-45 | 1.77993545 | 0.72 | 0.569 | 2.39E-42 | 6 | LINC00609 |
| EYA4 | 6.58E-45 | 0.33356901 | 0.697 | 0.416 | 1.32E-41 | 6 | EYA4 |
| PLPPR1 | 2.01E-44 | 1.34067218 | 0.708 | 0.5 | 4.02E-41 | 6 | PLPPR1 |
| POT1-AS1 | 2.20E-44 | 0.54773258 | 0.875 | 0.766 | 4.41E-41 | 6 | POT1-AS1 |
| LRRC4C1 | 1.52E-42 | 0.92644957 | 0.907 | 0.811 | 3.05E-39 | 6 | LRRC4C |
| FOLH1 | 1.89E-41 | 1.42224003 | 0.7 | 0.536 | 3.78E-38 | 6 | FOLH1 |
| SLIT2 | 2.41E-41 | 0.89681602 | 0.805 | 0.663 | 4.82E-38 | 6 | SLIT2 |

| | | | | | | | |
|---|---|---|---|---|---|---|---|
| ANLN | 3.18E-41 | 1.17947442 | 0.7 | 0.435 | 6.36E-38 | 6 | ANLN |
| RELN | 1.28E-40 | 0.32049566 | 0.741 | 0.721 | 2.56E-37 | 6 | RELN |
| LURAP1L-AS | 1.79E-40 | 0.87512863 | 0.743 | 0.811 | 3.59E-37 | 6 | LURAP1L-AS1 |
| AC233296.1 | 5.61E-40 | 0.76615557 | 0.706 | 0.581 | 1.12E-36 | 6 | AC233296.1 |
| SEMA3C | 1.23E-39 | 0.81267226 | 0.732 | 0.727 | 2.47E-36 | 6 | SEMA3C |
| ESRRG1 | 2.73E-39 | 1.04236212 | 0.729 | 0.54 | 5.46E-36 | 6 | ESRRG |
| ZCCHC24 | 4.57E-39 | 0.96881881 | 0.773 | 0.685 | 9.14E-36 | 6 | ZCCHC24 |
| LRRC63 | 4.42E-38 | 0.72613066 | 0.717 | 0.699 | 8.84E-35 | 6 | LRRC63 |
| LINC01182 | 7.00E-38 | 0.28713068 | 0.723 | 0.704 | 1.40E-34 | 6 | LINC01182 |
| NOX4 | 4.15E-37 | 0.32611363 | 0.729 | 0.867 | 8.31E-34 | 6 | NOX4 |
| SV2B1 | 4.70E-37 | 0.32176718 | 0.898 | 0.821 | 9.40E-34 | 6 | SV2B |
| ARPP211 | 3.31E-35 | 0.56052909 | 0.84 | 0.825 | 6.61E-32 | 6 | ARPP21 |
| LTBP1 | 9.40E-35 | 0.77993466 | 0.708 | 0.655 | 1.88E-31 | 6 | LTBP1 |
| EPN2 | 3.86E-34 | 0.86412292 | 0.773 | 0.627 | 7.71E-31 | 6 | EPN2 |
| ROR1 | 8.64E-34 | 0.73831105 | 0.711 | 0.791 | 1.73E-30 | 6 | ROR1 |
| SELENOP | 9.93E-34 | 0.69295791 | 0.77 | 0.658 | 1.99E-30 | 6 | SELENOP |
| AC025159.1 | 2.94E-33 | 0.53077559 | 0.851 | 0.689 | 5.87E-30 | 6 | AC025159.1 |
| OLIG1 | 6.61E-33 | 0.63178932 | 0.746 | 0.759 | 1.32E-29 | 6 | OLIG1 |
| SORCS2 | 8.07E-33 | 0.69280956 | 0.685 | 0.515 | 1.61E-29 | 6 | SORCS2 |
| RASGEF1B1 | 8.76E-33 | 0.30333472 | 0.863 | 0.66 | 1.75E-29 | 6 | RASGEF1B |
| LGR5 | 2.53E-32 | 0.74913213 | 0.671 | 0.454 | 5.06E-29 | 6 | LGR5 |
| VRK2 | 3.15E-32 | 0.8495536 | 0.822 | 0.702 | 6.30E-29 | 6 | VRK2 |
| DCC | 3.23E-31 | 0.62296916 | 0.787 | 0.741 | 6.46E-28 | 6 | DCC |
| KAZN1 | 7.36E-31 | 0.77829311 | 0.848 | 0.783 | 1.47E-27 | 6 | KAZN |
| NEGR1 | 2.05E-29 | 0.6678109 | 0.927 | 0.844 | 4.10E-26 | 6 | NEGR1 |
| DNER | 4.74E-29 | 0.92790047 | 0.708 | 0.607 | 9.48E-26 | 6 | DNER |
| PDZD2 | 8.55E-29 | 0.75135302 | 0.831 | 0.835 | 1.71E-25 | 6 | PDZD2 |
| AC092691.11 | 7.15E-28 | 0.56832242 | 0.831 | 0.757 | 1.43E-24 | 6 | AC092691.1 |
| PARD3B | 3.66E-27 | 0.38075913 | 0.898 | 0.834 | 7.33E-24 | 6 | PARD3B |
| MET | 5.80E-27 | 0.36169219 | 0.732 | 0.817 | 1.16E-23 | 6 | MET |
| LAMA2 | 4.60E-26 | 1.41104258 | 0.665 | 0.582 | 9.21E-23 | 6 | LAMA2 |
| SGCZ | 6.28E-26 | 0.3144224 | 0.796 | 0.693 | 1.26E-22 | 6 | SGCZ |
| SH3TC2 | 1.17E-23 | 1.05091977 | 0.65 | 0.596 | 2.34E-20 | 6 | SH3TC2 |
| GRAMD2B | 2.21E-22 | 0.64375755 | 0.743 | 0.74 | 4.42E-19 | 6 | GRAMD2B |
| WWTR1 | 2.85E-22 | 0.26407994 | 0.726 | 0.691 | 5.71E-19 | 6 | WWTR1 |
| RAB3IP | 1.22E-21 | 0.57552024 | 0.606 | 0.359 | 2.44E-18 | 6 | RAB3IP |
| LRP2 | 3.55E-21 | 1.18754951 | 0.612 | 0.278 | 7.09E-18 | 6 | LRP2 |
| LINC01505 | 7.48E-21 | 0.97734137 | 0.673 | 0.621 | 1.50E-17 | 6 | LINC01505 |
| RASGRF21 | 8.48E-21 | 0.84791032 | 0.723 | 0.773 | 1.70E-17 | 6 | RASGRF2 |
| CRB1 | 7.30E-20 | 0.51632552 | 0.72 | 0.8 | 1.46E-16 | 6 | CRB1 |
| RYR21 | 1.44E-19 | 0.38515154 | 0.825 | 0.828 | 2.89E-16 | 6 | RYR2 |
| MYO1E1 | 5.70E-18 | 0.3125873 | 0.776 | 0.599 | 1.14E-14 | 6 | MYO1E |
| MEIS2 | 7.37E-18 | 1.06056269 | 0.703 | 0.818 | 1.47E-14 | 6 | MEIS2 |
| ASTN21 | 7.90E-17 | 0.85445675 | 0.676 | 0.638 | 1.58E-13 | 6 | ASTN2 |
| GRIK4 | 8.94E-17 | 0.28636749 | 0.379 | 0.702 | 1.79E-13 | 6 | GRIK4 |
| IQCJ-SCHIP11 | 1.55E-16 | 0.41288715 | 0.948 | 0.89 | 3.10E-13 | 6 | IQCJ-SCHIP1 |
| SLC5A11 | 1.82E-16 | 1.15208726 | 0.618 | 0.516 | 3.64E-13 | 6 | SLC5A11 |
| TLE4 | 1.02E-15 | 0.28233104 | 0.845 | 0.814 | 2.03E-12 | 6 | TLE4 |
| KAT2B | 1.54E-15 | 0.54186541 | 0.755 | 0.723 | 3.08E-12 | 6 | KAT2B |
| ACACB | 1.90E-15 | 0.52112426 | 0.746 | 0.719 | 3.80E-12 | 6 | ACACB |

| | | | | | | | |
|---|---|---|---|---|---|---|---|
| GPM6A1 | 4.51E-15 | 0.26011669 | 0.904 | 0.793 | 9.02E-12 | 6 | GPM6A |
| LINC00598 | 5.08E-15 | 0.27441424 | 0.656 | 0.596 | 1.02E-11 | 6 | LINC00598 |
| GRIA4 | 2.93E-14 | 0.60414854 | 0.816 | 0.872 | 5.85E-11 | 6 | GRIA4 |
| PREX2 | 8.02E-14 | 0.62501683 | 0.72 | 0.726 | 1.60E-10 | 6 | PREX2 |
| CDH19 | 9.04E-14 | 0.94706288 | 0.592 | 0.454 | 1.81E-10 | 6 | CDH19 |
| SVEP1 | 3.94E-13 | 0.71392154 | 0.571 | 0.36 | 7.88E-10 | 6 | SVEP1 |
| LINC01445 | 7.33E-13 | 0.28779423 | 0.566 | 0.207 | 1.47E-09 | 6 | LINC01445 |
| LINC00511 | 1.50E-12 | 0.26010601 | 0.691 | 0.684 | 2.99E-09 | 6 | LINC00511 |
| EGLN3 | 8.76E-12 | 0.42352501 | 0.569 | 0.451 | 1.75E-08 | 6 | EGLN3 |
| CTNNA21 | 1.22E-11 | 0.73605321 | 0.91 | 0.894 | 2.44E-08 | 6 | CTNNA2 |
| DGKD | 8.19E-11 | 0.30807172 | 0.659 | 0.509 | 1.64E-07 | 6 | DGKD |
| SLC1A2 | 8.88E-11 | 0.39658244 | 0.848 | 0.785 | 1.78E-07 | 6 | SLC1A2 |
| CDH8 | 1.14E-10 | 0.40500556 | 0.7 | 0.743 | 2.27E-07 | 6 | CDH8 |
| PROX1 | 4.62E-10 | 0.54649037 | 0.662 | 0.691 | 9.25E-07 | 6 | PROX1 |
| PLEKHH2 | 1.27E-09 | 0.307951 | 0.671 | 0.751 | 2.54E-06 | 6 | PLEKHH2 |
| LHFPL6 | 1.79E-09 | 0.64666737 | 0.7 | 0.803 | 3.58E-06 | 6 | LHFPL6 |
| PTN | 2.68E-09 | 0.8761594 | 0.598 | 0.649 | 5.37E-06 | 6 | PTN |
| MID1 | 4.78E-09 | 0.25550284 | 0.778 | 0.78 | 9.55E-06 | 6 | MID1 |
| CALN1 | 6.47E-09 | 0.2530022 | 0.379 | 0.525 | 1.29E-05 | 6 | CALN1 |
| AL445250.1 | 8.82E-08 | 0.55136846 | 0.606 | 0.632 | 0.00017648 | 6 | AL445250.1 |
| AL078590.2 | 5.88E-07 | 0.41405931 | 0.735 | 0.723 | 0.00117659 | 6 | AL078590.2 |
| ITPKB | 7.07E-07 | 0.27081957 | 0.813 | 0.816 | 0.00141451 | 6 | ITPKB |
| ELOVL7 | 7.83E-07 | 0.47266853 | 0.563 | 0.584 | 0.00156563 | 6 | ELOVL7 |
| KCTD11 | 1.15E-06 | 0.28809581 | 0.828 | 0.82 | 0.00229294 | 6 | KCTD1 |
| KCTD81 | 2.72E-06 | 0.96479675 | 0.551 | 0.524 | 0.0054445 | 6 | KCTD8 |
| SMOC1 | 9.05E-06 | 0.28562317 | 0.499 | 0.732 | 0.01809435 | 6 | SMOC1 |
| LINC01099 | 1.49E-05 | 0.48360215 | 0.539 | 0.461 | 0.02987318 | 6 | LINC01099 |
| APOLD1 | 0.00090089 | 0.46267708 | 0.548 | 0.544 | 1 | 6 | APOLD1 |
| ERMN | 0.00091866 | 0.69945822 | 0.548 | 0.642 | 1 | 6 | ERMN |
| PLD5 | 0.00362079 | 0.54392898 | 0.443 | 0.63 | 1 | 6 | PLD5 |
| KCNK1 | 0.00442647 | 0.30781788 | 0.536 | 0.605 | 1 | 6 | KCNK1 |
| AL392023.2 | 0.00550521 | 0.62173234 | 0.571 | 0.734 | 1 | 6 | AL392023.2 |
| F13A1 | 4.62E-101 | 3.63833864 | 0.956 | 0.637 | 9.24E-98 | 7 | F13A1 |
| CD1631 | 1.25E-86 | 2.47414701 | 0.916 | 0.477 | 2.50E-83 | 7 | CD163 |
| SELENOP1 | 1.33E-69 | 2.76130121 | 0.901 | 0.657 | 2.65E-66 | 7 | SELENOP |
| ATP10A | 9.03E-66 | 0.56294628 | 0.887 | 0.786 | 1.81E-62 | 7 | ATP10A |
| MRC1 | 9.54E-62 | 2.47999794 | 0.926 | 0.767 | 1.91E-58 | 7 | MRC1 |
| VAV3 | 3.28E-58 | 2.05266585 | 0.852 | 0.679 | 6.57E-55 | 7 | VAV3 |
| IQGAP22 | 3.06E-54 | 1.97542763 | 0.887 | 0.721 | 6.12E-51 | 7 | IQGAP2 |
| FGF13 | 8.78E-50 | 1.87444686 | 0.842 | 0.668 | 1.76E-46 | 7 | FGF13 |
| MS4A4E | 5.03E-48 | 1.38800225 | 0.847 | 0.619 | 1.01E-44 | 7 | MS4A4E |
| DAPK11 | 4.16E-47 | 1.73914233 | 0.857 | 0.73 | 8.32E-44 | 7 | DAPK1 |
| HRH11 | 2.50E-44 | 1.03182221 | 0.808 | 0.642 | 5.01E-41 | 7 | HRH1 |
| MDFIC | 2.17E-43 | 0.84774103 | 0.828 | 0.779 | 4.33E-40 | 7 | MDFIC |
| COLEC12 | 1.33E-41 | 2.41303392 | 0.783 | 0.513 | 2.66E-38 | 7 | COLEC12 |
| STAB1 | 5.60E-40 | 1.77277827 | 0.847 | 0.601 | 1.12E-36 | 7 | STAB1 |
| MS4A4A1 | 3.71E-39 | 1.57487047 | 0.798 | 0.518 | 7.43E-36 | 7 | MS4A4A |
| DIRC31 | 9.47E-36 | 1.88584987 | 0.877 | 0.794 | 1.89E-32 | 7 | DIRC3 |
| FMN11 | 2.15E-35 | 1.08386972 | 0.916 | 0.733 | 4.30E-32 | 7 | FMN1 |
| MAN1A1 | 3.60E-34 | 1.8141697 | 0.906 | 0.773 | 7.20E-31 | 7 | MAN1A1 |

| | | | | | | | |
|---|---|---|---|---|---|---|---|
| ANXA1 | 8.86E-32 | 0.56033376 | 0.759 | 0.567 | 1.77E-28 | 7 | ANXA1 |
| LYVE1 | 2.90E-30 | 1.81220184 | 0.749 | 0.575 | 5.81E-27 | 7 | LYVE1 |
| ANTXR21 | 6.39E-29 | 0.77769706 | 0.749 | 0.6 | 1.28E-25 | 7 | ANTXR2 |
| TSHZ3 | 1.62E-28 | 1.20523089 | 0.749 | 0.564 | 3.25E-25 | 7 | TSHZ3 |
| EYA22 | 2.00E-27 | 1.35769406 | 0.749 | 0.644 | 4.01E-24 | 7 | EYA2 |
| ALDH1A1 | 1.09E-26 | 0.48308859 | 0.759 | 0.792 | 2.19E-23 | 7 | ALDH1A1 |
| COBLL1 | 6.64E-26 | 0.30483864 | 0.724 | 0.539 | 1.33E-22 | 7 | COBLL1 |
| CD36 | 1.45E-24 | 0.70130817 | 0.709 | 0.593 | 2.91E-21 | 7 | CD36 |
| CDYL2 | 5.56E-22 | 0.25461903 | 0.813 | 0.715 | 1.11E-18 | 7 | CDYL2 |
| ABCA9 | 6.10E-22 | 0.53336277 | 0.704 | 0.507 | 1.22E-18 | 7 | ABCA9 |
| ARHGAP181 | 8.09E-22 | 1.28702619 | 0.764 | 0.639 | 1.62E-18 | 7 | ARHGAP18 |
| MCTP12 | 8.38E-22 | 1.34338829 | 0.759 | 0.71 | 1.68E-18 | 7 | MCTP1 |
| GNG12-AS1 | 1.22E-21 | 0.37100837 | 0.182 | 0.449 | 2.44E-18 | 7 | GNG12-AS1 |
| CPM2 | 6.36E-20 | 0.99072284 | 0.798 | 0.631 | 1.27E-16 | 7 | CPM |
| NEAT11 | 2.33E-19 | 0.537513 | 0.995 | 0.979 | 4.66E-16 | 7 | NEAT1 |
| EDA | 3.14E-19 | 1.27732468 | 0.857 | 0.797 | 6.29E-16 | 7 | EDA |
| IL12RB2 | 5.72E-19 | 0.97441939 | 0.69 | 0.64 | 1.14E-15 | 7 | IL12RB2 |
| PID1 | 3.68E-17 | 1.29963127 | 0.842 | 0.866 | 7.35E-14 | 7 | PID1 |
| NAMPT1 | 8.11E-17 | 1.3459579 | 0.69 | 0.529 | 1.62E-13 | 7 | NAMPT |
| FAM198B-AS | 8.15E-17 | 0.49471992 | 0.739 | 0.617 | 1.63E-13 | 7 | FAM198B-AS1 |
| NRP1 | 8.70E-17 | 1.38787076 | 0.773 | 0.662 | 1.74E-13 | 7 | NRP1 |
| CPNE81 | 7.12E-16 | 0.95533664 | 0.69 | 0.55 | 1.42E-12 | 7 | CPNE8 |
| LDLRAD31 | 1.50E-15 | 0.30852825 | 0.685 | 0.489 | 3.01E-12 | 7 | LDLRAD3 |
| RBM472 | 2.30E-15 | 0.70248551 | 0.936 | 0.865 | 4.60E-12 | 7 | RBM47 |
| CXCR4 | 9.28E-15 | 0.35659975 | 0.65 | 0.427 | 1.86E-11 | 7 | CXCR4 |
| MS4A6A | 1.89E-14 | 0.83855357 | 0.887 | 0.826 | 3.77E-11 | 7 | MS4A6A |
| HS3ST21 | 2.87E-14 | 1.52518282 | 0.7 | 0.829 | 5.75E-11 | 7 | HS3ST2 |
| CEMIP2 | 2.00E-13 | 1.04975505 | 0.586 | 0.336 | 4.00E-10 | 7 | CEMIP2 |
| PAM1 | 3.02E-13 | 0.27160328 | 0.773 | 0.621 | 6.04E-10 | 7 | PAM |
| SPON1 | 3.23E-13 | 0.30494155 | 0.409 | 0.638 | 6.46E-10 | 7 | SPON1 |
| LINC00278 | 1.04E-12 | 0.30980759 | 0.606 | 0.386 | 2.09E-09 | 7 | LINC00278 |
| MS4A71 | 8.48E-12 | 0.81741573 | 0.778 | 0.654 | 1.70E-08 | 7 | MS4A7 |
| LRP1B1 | 4.20E-11 | 0.31240357 | 0.941 | 0.91 | 8.41E-08 | 7 | LRP1B |
| MPPED2 | 4.46E-11 | 0.81739605 | 0.67 | 0.593 | 8.91E-08 | 7 | MPPED2 |
| MAMDC2 | 6.11E-11 | 0.65182376 | 0.793 | 0.781 | 1.22E-07 | 7 | MAMDC2 |
| GNG2 | 4.05E-10 | 0.4162299 | 0.901 | 0.87 | 8.10E-07 | 7 | GNG2 |
| CABLES1 | 9.65E-10 | 0.30355127 | 0.892 | 0.862 | 1.93E-06 | 7 | CABLES1 |
| SCN9A | 1.18E-09 | 0.873708 | 0.665 | 0.709 | 2.36E-06 | 7 | SCN9A |
| ADGRG6 | 1.47E-09 | 0.96364492 | 0.685 | 0.732 | 2.93E-06 | 7 | ADGRG6 |
| SRGN1 | 2.02E-09 | 0.68402742 | 0.837 | 0.825 | 4.04E-06 | 7 | SRGN |
| ESR1 | 3.88E-09 | 0.68163448 | 0.788 | 0.785 | 7.75E-06 | 7 | ESR1 |
| FRMD4B2 | 6.19E-09 | 0.62584002 | 0.887 | 0.841 | 1.24E-05 | 7 | FRMD4B |
| MERTK1 | 6.79E-09 | 0.44184484 | 0.837 | 0.77 | 1.36E-05 | 7 | MERTK |
| PLEK | 8.69E-09 | 0.56031923 | 0.828 | 0.755 | 1.74E-05 | 7 | PLEK |
| AC021594.2 | 9.52E-09 | 0.25368929 | 0.635 | 0.51 | 1.90E-05 | 7 | AC021594.2 |
| RASGEF1B2 | 1.17E-08 | 0.69578078 | 0.719 | 0.668 | 2.35E-05 | 7 | RASGEF1B |
| MCUB | 1.58E-08 | 0.28862193 | 0.798 | 0.726 | 3.16E-05 | 7 | MCUB |
| PDGFC1 | 2.30E-08 | 0.75401784 | 0.837 | 0.863 | 4.60E-05 | 7 | PDGFC |
| ADGRA3 | 2.80E-08 | 0.73520902 | 0.695 | 0.76 | 5.60E-05 | 7 | ADGRA3 |
| EGFL7 | 7.19E-08 | 0.42258995 | 0.626 | 0.596 | 0.00014374 | 7 | EGFL7 |

| | | | | | | | |
|---|---|---|---|---|---|---|---|
| OPHN11 | 1.73E-07 | 0.59929291 | 0.759 | 0.711 | 0.00034661 | 7 | OPHN1 |
| AL356124.1 | 3.19E-07 | 0.49951207 | 0.635 | 0.63 | 0.00063711 | 7 | AL356124.1 |
| PARP8 | 4.74E-07 | 0.59136334 | 0.946 | 0.886 | 0.00094799 | 7 | PARP8 |
| AP001636.3 | 5.67E-07 | 0.39122177 | 0.645 | 0.626 | 0.00113363 | 7 | AP001636.3 |
| AC100849.1 | 8.80E-07 | 1.77472563 | 0.586 | 0.606 | 0.00176063 | 7 | AC100849.1 |
| KHDRBS2 | 1.32E-06 | 0.45917261 | 0.872 | 0.852 | 0.00264899 | 7 | KHDRBS2 |
| UTRN | 1.78E-06 | 0.63911055 | 0.916 | 0.917 | 0.00355313 | 7 | UTRN |
| IFI44L1 | 2.23E-06 | 0.62128612 | 0.734 | 0.68 | 0.00446722 | 7 | IFI44L |
| SLC12A7 | 3.19E-06 | 0.43968557 | 0.828 | 0.771 | 0.00637464 | 7 | SLC12A7 |
| PVT11 | 3.35E-06 | 0.70868003 | 0.65 | 0.573 | 0.00669096 | 7 | PVT1 |
| FCGR2A1 | 3.61E-06 | 0.65420523 | 0.66 | 0.606 | 0.00722351 | 7 | FCGR2A |
| PARD31 | 7.66E-06 | 0.37750435 | 0.69 | 0.671 | 0.01532107 | 7 | PARD3 |
| MAF | 8.66E-06 | 0.41847396 | 0.931 | 0.896 | 0.01731928 | 7 | MAF |
| TRHDE | 1.62E-05 | 0.35976201 | 0.621 | 0.675 | 0.0324053 | 7 | TRHDE |
| COL5A3 | 2.22E-05 | 0.26494271 | 0.803 | 0.781 | 0.04449948 | 7 | COL5A3 |
| GALNTL61 | 3.23E-05 | 1.05019769 | 0.695 | 0.711 | 0.06460553 | 7 | GALNTL6 |
| AL160272.1 | 3.24E-05 | 0.26381513 | 0.818 | 0.792 | 0.06481625 | 7 | AL160272.1 |
| ZNF804A1 | 3.44E-05 | 0.62803897 | 0.739 | 0.747 | 0.06871076 | 7 | ZNF804A |
| CEBPD1 | 3.93E-05 | 0.41872554 | 0.773 | 0.77 | 0.07867482 | 7 | CEBPD |
| AC083837.1 | 5.74E-05 | 0.46932428 | 0.557 | 0.467 | 0.1147546 | 7 | AC083837.1 |
| NHSL11 | 7.00E-05 | 0.25763797 | 0.833 | 0.772 | 0.13996084 | 7 | NHSL1 |
| AFF31 | 7.30E-05 | 0.52434049 | 0.842 | 0.851 | 0.14601656 | 7 | AFF3 |
| RNF144B | 7.55E-05 | 0.62286948 | 0.759 | 0.709 | 0.15093394 | 7 | RNF144B |
| WLS | 9.91E-05 | 0.74463566 | 0.788 | 0.827 | 0.19826976 | 7 | WLS |
| TFEC1 | 0.00014716 | 0.29524492 | 0.862 | 0.833 | 0.29432849 | 7 | TFEC |
| EPSTI11 | 0.00025909 | 0.34360099 | 0.837 | 0.737 | 0.51818457 | 7 | EPSTI1 |
| AHNAK | 0.00026163 | 0.68470474 | 0.616 | 0.609 | 0.52325154 | 7 | AHNAK |
| STON2 | 0.00027664 | 0.26923756 | 0.798 | 0.796 | 0.55328995 | 7 | STON2 |
| GAS2L3 | 0.00029686 | 0.73079606 | 0.626 | 0.676 | 0.59372586 | 7 | GAS2L3 |
| HTR7 | 0.00037932 | 0.283267 | 0.626 | 0.687 | 0.75864902 | 7 | HTR7 |
| PBX31 | 0.00135622 | 0.47042874 | 0.803 | 0.79 | 1 | 7 | PBX3 |
| EPS8 | 0.00188391 | 0.31259755 | 0.847 | 0.854 | 1 | 7 | EPS8 |
| FAM110B | 0.00206971 | 0.25513728 | 0.571 | 0.516 | 1 | 7 | FAM110B |
| TTN | 0.00905206 | 1.1557987 | 0.507 | 0.502 | 1 | 7 | TTN |
| SAMD4A1 | 0.00971189 | 0.4011484 | 0.798 | 0.829 | 1 | 7 | SAMD4A |
| FAM155A1 | 6.20E-68 | 2.25472809 | 0.949 | 0.899 | 1.24E-64 | 8 | FAM155A |
| OPCML1 | 1.80E-67 | 2.33572144 | 0.97 | 0.905 | 3.59E-64 | 8 | OPCML |
| CNTNAP22 | 2.20E-66 | 2.47465929 | 0.965 | 0.901 | 4.41E-63 | 8 | CNTNAP2 |
| RBFOX11 | 3.37E-64 | 2.39783274 | 0.98 | 0.944 | 6.73E-61 | 8 | RBFOX1 |
| DLGAP1 | 3.93E-59 | 2.2225479 | 0.955 | 0.903 | 7.86E-56 | 8 | DLGAP1 |
| SYT11 | 6.41E-55 | 1.87606395 | 0.96 | 0.935 | 1.28E-51 | 8 | SYT1 |
| FGF121 | 4.36E-51 | 1.87997253 | 0.894 | 0.803 | 8.73E-48 | 8 | FGF12 |
| MEG31 | 8.53E-51 | 1.42561638 | 0.975 | 0.959 | 1.71E-47 | 8 | MEG3 |
| LRP1B2 | 9.80E-50 | 2.03710679 | 0.939 | 0.911 | 1.96E-46 | 8 | LRP1B |
| CADPS1 | 2.44E-48 | 1.79392671 | 0.914 | 0.877 | 4.89E-45 | 8 | CADPS |
| STXBP5L1 | 4.22E-48 | 1.68265279 | 0.919 | 0.882 | 8.45E-45 | 8 | STXBP5L |
| SNTG11 | 5.81E-48 | 2.03965888 | 0.919 | 0.819 | 1.16E-44 | 8 | SNTG1 |
| RIMS21 | 8.12E-47 | 1.76333865 | 0.904 | 0.803 | 1.62E-43 | 8 | RIMS2 |
| KALRN | 1.17E-45 | 1.79076239 | 0.934 | 0.874 | 2.34E-42 | 8 | KALRN |
| NEGR11 | 1.95E-45 | 1.74258989 | 0.924 | 0.846 | 3.90E-42 | 8 | NEGR1 |

| | | | | | | | |
|---|---|---|---|---|---|---|---|
| CCSER11 | 2.16E-45 | 1.82639981 | 0.939 | 0.898 | 4.32E-42 | 8 | CCSER1 |
| PLCB1 | 2.33E-45 | 2.11567746 | 0.904 | 0.876 | 4.66E-42 | 8 | PLCB1 |
| CACNA1C | 8.08E-45 | 1.73923563 | 0.894 | 0.844 | 1.62E-41 | 8 | CACNA1C |
| KAZN2 | 2.33E-44 | 2.19975746 | 0.879 | 0.784 | 4.65E-41 | 8 | KAZN |
| LRRTM41 | 3.13E-43 | 1.68034094 | 0.944 | 0.92 | 6.26E-40 | 8 | LRRTM4 |
| CTNNA22 | 9.14E-42 | 1.82191183 | 0.929 | 0.894 | 1.83E-38 | 8 | CTNNA2 |
| GRIK21 | 1.40E-41 | 1.89120084 | 0.924 | 0.915 | 2.80E-38 | 8 | GRIK2 |
| ATP8A22 | 4.32E-38 | 1.67007992 | 0.843 | 0.811 | 8.64E-35 | 8 | ATP8A2 |
| CNTN51 | 9.88E-38 | 2.09014344 | 0.879 | 0.827 | 1.98E-34 | 8 | CNTN5 |
| RYR22 | 1.94E-37 | 1.69230495 | 0.889 | 0.826 | 3.89E-34 | 8 | RYR2 |
| ROBO21 | 5.92E-36 | 2.42467321 | 0.904 | 0.869 | 1.18E-32 | 8 | ROBO2 |
| CACNA2D31 | 5.94E-36 | 1.72679417 | 0.879 | 0.819 | 1.19E-32 | 8 | CACNA2D3 |
| KCNIP41 | 1.27E-35 | 1.7758096 | 0.995 | 0.981 | 2.53E-32 | 8 | KCNIP4 |
| RGS71 | 2.19E-35 | 1.58103388 | 0.904 | 0.877 | 4.38E-32 | 8 | RGS7 |
| MDGA21 | 3.91E-35 | 1.62449936 | 0.899 | 0.863 | 7.81E-32 | 8 | MDGA2 |
| GRM5 | 9.96E-35 | 1.51969518 | 0.869 | 0.86 | 1.99E-31 | 8 | GRM5 |
| DAB11 | 2.90E-34 | 1.53414248 | 0.909 | 0.863 | 5.81E-31 | 8 | DAB1 |
| DLGAP21 | 4.32E-34 | 1.48697256 | 0.914 | 0.893 | 8.64E-31 | 8 | DLGAP2 |
| GRIP11 | 9.03E-34 | 1.8182928 | 0.823 | 0.751 | 1.81E-30 | 8 | GRIP1 |
| GRIA11 | 2.54E-33 | 1.53951677 | 0.854 | 0.806 | 5.07E-30 | 8 | GRIA1 |
| TMEM178B1 | 2.66E-33 | 1.37366121 | 0.848 | 0.796 | 5.33E-30 | 8 | TMEM178B |
| DPP101 | 3.29E-33 | 2.31037883 | 0.934 | 0.871 | 6.57E-30 | 8 | DPP10 |
| MEG81 | 6.22E-33 | 1.20120593 | 0.869 | 0.821 | 1.24E-29 | 8 | MEG8 |
| GRIN2B1 | 1.77E-32 | 1.43405779 | 0.859 | 0.84 | 3.54E-29 | 8 | GRIN2B |
| FRMPD4 | 6.05E-31 | 1.64167965 | 0.889 | 0.813 | 1.21E-27 | 8 | FRMPD4 |
| AGBL4 | 1.57E-30 | 1.6249417 | 0.859 | 0.828 | 3.13E-27 | 8 | AGBL4 |
| TENM21 | 2.60E-30 | 1.97546877 | 0.859 | 0.866 | 5.20E-27 | 8 | TENM2 |
| XKR41 | 3.41E-30 | 1.77341223 | 0.732 | 0.581 | 6.82E-27 | 8 | XKR4 |
| SYN31 | 5.03E-30 | 1.68426421 | 0.823 | 0.789 | 1.01E-26 | 8 | SYN3 |
| AC092691.12 | 6.02E-30 | 1.54166625 | 0.813 | 0.759 | 1.20E-26 | 8 | AC092691.1 |
| SNAP251 | 1.26E-29 | 1.17382504 | 0.843 | 0.753 | 2.51E-26 | 8 | SNAP25 |
| LINGO2 | 4.90E-29 | 1.67031386 | 0.884 | 0.905 | 9.79E-26 | 8 | LINGO2 |
| PDE4D1 | 2.74E-28 | 1.16863832 | 0.975 | 0.962 | 5.48E-25 | 8 | PDE4D |
| GRIA41 | 6.44E-28 | 1.29091943 | 0.889 | 0.869 | 1.29E-24 | 8 | GRIA4 |
| MTUS21 | 7.39E-28 | 1.65359259 | 0.848 | 0.833 | 1.48E-24 | 8 | MTUS2 |
| TMEM132B1 | 1.24E-27 | 1.35409616 | 0.828 | 0.786 | 2.47E-24 | 8 | TMEM132B |
| KCNQ5 | 1.27E-27 | 1.74649071 | 0.859 | 0.84 | 2.54E-24 | 8 | KCNQ5 |
| PCLO1 | 1.72E-27 | 1.40480967 | 0.843 | 0.806 | 3.45E-24 | 8 | PCLO |
| CDH181 | 2.74E-27 | 1.2977126 | 0.869 | 0.833 | 5.47E-24 | 8 | CDH18 |
| AFF32 | 1.69E-26 | 1.22971855 | 0.874 | 0.85 | 3.37E-23 | 8 | AFF3 |
| GALNT171 | 2.76E-26 | 1.44472392 | 0.833 | 0.802 | 5.52E-23 | 8 | GALNT17 |
| MIAT | 5.53E-26 | 1.02082119 | 0.874 | 0.869 | 1.11E-22 | 8 | MIAT |
| ADGRL21 | 6.83E-26 | 1.25977797 | 0.808 | 0.756 | 1.37E-22 | 8 | ADGRL2 |
| GABBR21 | 1.65E-25 | 1.24065926 | 0.773 | 0.739 | 3.29E-22 | 8 | GABBR2 |
| CNTN41 | 2.35E-25 | 1.53521745 | 0.788 | 0.722 | 4.69E-22 | 8 | CNTN4 |
| GPM6A2 | 2.86E-25 | 1.08911624 | 0.859 | 0.796 | 5.71E-22 | 8 | GPM6A |
| PCDH71 | 3.70E-25 | 1.55034145 | 0.879 | 0.857 | 7.40E-22 | 8 | PCDH7 |
| HS6ST31 | 4.20E-25 | 1.35706439 | 0.899 | 0.855 | 8.41E-22 | 8 | HS6ST3 |
| RALYL1 | 5.05E-25 | 1.72937132 | 0.884 | 0.883 | 1.01E-21 | 8 | RALYL |
| LRFN5 | 5.36E-25 | 1.43068611 | 0.828 | 0.827 | 1.07E-21 | 8 | LRFN5 |

| | | | | | | | |
|---|---|---|---|---|---|---|---|
| ERC21 | 5.53E-25 | 1.28634592 | 0.859 | 0.746 | 1.11E-21 | 8 | ERC2 |
| ABLIM11 | 7.39E-25 | 0.97405098 | 0.818 | 0.798 | 1.48E-21 | 8 | ABLIM1 |
| ADCY22 | 9.54E-25 | 1.18898635 | 0.828 | 0.811 | 1.91E-21 | 8 | ADCY2 |
| ATRNL11 | 9.60E-25 | 1.3170498 | 0.889 | 0.899 | 1.92E-21 | 8 | ATRNL1 |
| SCN1A1 | 1.16E-24 | 1.05583608 | 0.773 | 0.662 | 2.33E-21 | 8 | SCN1A |
| KHDRBS21 | 3.33E-24 | 1.56271549 | 0.874 | 0.851 | 6.66E-21 | 8 | KHDRBS2 |
| DCLK1 | 4.75E-24 | 1.30308666 | 0.838 | 0.855 | 9.51E-21 | 8 | DCLK1 |
| LRRC4C2 | 6.79E-24 | 1.35765755 | 0.848 | 0.815 | 1.36E-20 | 8 | LRRC4C |
| SYNPR1 | 1.22E-23 | 1.42569479 | 0.773 | 0.671 | 2.44E-20 | 8 | SYNPR |
| PDZRN3 | 4.05E-23 | 1.32469104 | 0.778 | 0.71 | 8.11E-20 | 8 | PDZRN3 |
| ASIC21 | 5.56E-23 | 1.66757408 | 0.778 | 0.774 | 1.11E-19 | 8 | ASIC2 |
| TNR1 | 5.87E-23 | 1.06654354 | 0.818 | 0.755 | 1.17E-19 | 8 | TNR |
| SYN21 | 1.53E-22 | 1.31692751 | 0.788 | 0.76 | 3.07E-19 | 8 | SYN2 |
| CNTNAP5 | 1.73E-22 | 1.65735612 | 0.833 | 0.858 | 3.46E-19 | 8 | CNTNAP5 |
| CDH121 | 2.59E-22 | 1.47681765 | 0.854 | 0.848 | 5.17E-19 | 8 | CDH12 |
| SLC4A10 | 3.11E-22 | 1.22179199 | 0.859 | 0.813 | 6.22E-19 | 8 | SLC4A10 |
| CACNA1B1 | 3.14E-22 | 1.28303267 | 0.773 | 0.713 | 6.27E-19 | 8 | CACNA1B |
| UNC5D | 4.53E-22 | 1.78516508 | 0.823 | 0.807 | 9.05E-19 | 8 | UNC5D |
| GRM71 | 7.31E-22 | 1.65144833 | 0.848 | 0.844 | 1.46E-18 | 8 | GRM7 |
| CDH101 | 8.83E-22 | 1.1147519 | 0.793 | 0.771 | 1.77E-18 | 8 | CDH10 |
| ASTN22 | 9.89E-22 | 1.27113524 | 0.742 | 0.637 | 1.98E-18 | 8 | ASTN2 |
| MYRIP1 | 1.14E-21 | 1.32963589 | 0.737 | 0.662 | 2.29E-18 | 8 | MYRIP |
| NRCAM1 | 3.03E-21 | 1.02692343 | 0.803 | 0.779 | 6.06E-18 | 8 | NRCAM |
| INPP4B | 3.55E-21 | 1.20100487 | 0.828 | 0.741 | 7.10E-18 | 8 | INPP4B |
| PHACTR11 | 3.76E-21 | 0.94506695 | 0.909 | 0.903 | 7.52E-18 | 8 | PHACTR1 |
| GRIN2A2 | 1.37E-20 | 1.47759162 | 0.808 | 0.826 | 2.75E-17 | 8 | GRIN2A |
| PTPRG2 | 1.98E-20 | 0.592861 | 0.773 | 0.582 | 3.95E-17 | 8 | PTPRG |
| RIMS1 | 2.98E-20 | 1.17853155 | 0.848 | 0.837 | 5.95E-17 | 8 | RIMS1 |
| SORBS21 | 3.19E-20 | 1.16046694 | 0.848 | 0.837 | 6.38E-17 | 8 | SORBS2 |
| KCNJ3 | 3.38E-20 | 1.11126002 | 0.742 | 0.696 | 6.76E-17 | 8 | KCNJ3 |
| RBMS31 | 3.69E-20 | 1.33558729 | 0.677 | 0.508 | 7.38E-17 | 8 | RBMS3 |
| PTPRT2 | 5.08E-20 | 1.50999905 | 0.818 | 0.781 | 1.02E-16 | 8 | PTPRT |
| PDE10A | 6.89E-20 | 1.22073198 | 0.768 | 0.784 | 1.38E-16 | 8 | PDE10A |
| HECW1 | 1.18E-19 | 1.20966102 | 0.813 | 0.816 | 2.36E-16 | 8 | HECW1 |
| PDE1A2 | 1.51E-19 | 1.08808732 | 0.894 | 0.841 | 3.02E-16 | 8 | PDE1A |
| IQCJ-SCHIP12 | 1.69E-19 | 1.62536577 | 0.899 | 0.892 | 3.39E-16 | 8 | IQCJ-SCHIP1 |
| HCN11 | 1.85E-19 | 1.03391336 | 0.808 | 0.772 | 3.70E-16 | 8 | HCN1 |
| GABRB2 | 1.96E-19 | 1.29859789 | 0.737 | 0.691 | 3.92E-16 | 8 | GABRB2 |
| SRRM41 | 6.61E-19 | 1.09826997 | 0.747 | 0.708 | 1.32E-15 | 8 | SRRM4 |
| CHRM31 | 1.77E-18 | 1.47034777 | 0.783 | 0.751 | 3.55E-15 | 8 | CHRM3 |
| KCNC2 | 2.43E-18 | 1.36736193 | 0.768 | 0.794 | 4.87E-15 | 8 | KCNC2 |
| PCSK21 | 2.66E-18 | 1.0776293 | 0.803 | 0.827 | 5.32E-15 | 8 | PCSK2 |
| SGCZ1 | 3.56E-18 | 1.76714638 | 0.763 | 0.696 | 7.11E-15 | 8 | SGCZ |
| ZNF385B1 | 4.63E-18 | 1.33850085 | 0.808 | 0.829 | 9.26E-15 | 8 | ZNF385B |
| SLC35F1 | 5.11E-18 | 0.97489361 | 0.828 | 0.862 | 1.02E-14 | 8 | SLC35F1 |
| ERBB41 | 9.61E-18 | 1.99147937 | 0.828 | 0.794 | 1.92E-14 | 8 | ERBB4 |
| CHL11 | 1.09E-17 | 0.96116665 | 0.833 | 0.82 | 2.18E-14 | 8 | CHL1 |
| GPC61 | 1.34E-17 | 1.13472642 | 0.773 | 0.779 | 2.68E-14 | 8 | GPC6 |
| SLC44A5 | 3.18E-17 | 1.0615208 | 0.793 | 0.794 | 6.36E-14 | 8 | SLC44A5 |
| KIAA12171 | 5.09E-17 | 1.52896366 | 0.697 | 0.654 | 1.02E-13 | 8 | KIAA1217 |

| | | | | | | | |
|---|---|---|---|---|---|---|---|
| SHISA9 | 5.99E-17 | 1.2546856 | 0.793 | 0.79 | 1.20E-13 | 8 | SHISA9 |
| TMEM1082 | 8.77E-17 | 1.193664 | 0.823 | 0.824 | 1.75E-13 | 8 | TMEM108 |
| TMEM132D1 | 1.03E-16 | 1.1465685 | 0.818 | 0.834 | 2.06E-13 | 8 | TMEM132D |
| GRIA31 | 2.55E-16 | 1.18042876 | 0.753 | 0.786 | 5.10E-13 | 8 | GRIA3 |
| KIAA1549L1 | 2.86E-16 | 1.04118904 | 0.813 | 0.781 | 5.73E-13 | 8 | KIAA1549L |
| KCND2 | 3.15E-16 | 1.39880625 | 0.773 | 0.821 | 6.30E-13 | 8 | KCND2 |
| SEZ6L | 3.65E-16 | 1.00793947 | 0.783 | 0.795 | 7.31E-13 | 8 | SEZ6L |
| SLC35F31 | 4.43E-16 | 0.93358542 | 0.747 | 0.749 | 8.87E-13 | 8 | SLC35F3 |
| RGS61 | 5.43E-16 | 1.5548324 | 0.803 | 0.837 | 1.09E-12 | 8 | RGS6 |
| ROBO11 | 7.68E-16 | 1.41662842 | 0.727 | 0.662 | 1.54E-12 | 8 | ROBO1 |
| ADAMTS9-AS | 9.41E-16 | 1.06660103 | 0.722 | 0.716 | 1.88E-12 | 8 | ADAMTS9-AS2 |
| DCC1 | 9.45E-16 | 1.38226426 | 0.763 | 0.743 | 1.89E-12 | 8 | DCC |
| AC073050.11 | 1.23E-15 | 1.2533213 | 0.803 | 0.829 | 2.47E-12 | 8 | AC073050.1 |
| FSTL5 | 1.49E-15 | 1.17796675 | 0.677 | 0.594 | 2.99E-12 | 8 | FSTL5 |
| CALN11 | 1.70E-15 | 0.98867155 | 0.601 | 0.516 | 3.41E-12 | 8 | CALN1 |
| PAM2 | 1.82E-15 | 0.78005764 | 0.722 | 0.622 | 3.64E-12 | 8 | PAM |
| DAPK12 | 3.00E-15 | 0.8548534 | 0.778 | 0.732 | 5.99E-12 | 8 | DAPK1 |
| GALNT18 | 3.12E-15 | 0.76725348 | 0.783 | 0.819 | 6.24E-12 | 8 | GALNT18 |
| GPR158 | 4.66E-15 | 0.7512188 | 0.763 | 0.782 | 9.31E-12 | 8 | GPR158 |
| RBFOX31 | 4.90E-15 | 1.20264918 | 0.702 | 0.717 | 9.81E-12 | 8 | RBFOX3 |
| KCNH11 | 5.26E-15 | 1.06277212 | 0.813 | 0.831 | 1.05E-11 | 8 | KCNH1 |
| THSD7A | 1.05E-14 | 1.07864329 | 0.732 | 0.754 | 2.10E-11 | 8 | THSD7A |
| SPOCK31 | 1.56E-14 | 1.02478304 | 0.556 | 0.393 | 3.12E-11 | 8 | SPOCK3 |
| GABRB31 | 1.64E-14 | 0.96973917 | 0.793 | 0.837 | 3.28E-11 | 8 | GABRB3 |
| FLRT2 | 1.79E-14 | 1.42634188 | 0.763 | 0.798 | 3.57E-11 | 8 | FLRT2 |
| NCALD1 | 1.82E-14 | 0.90981934 | 0.722 | 0.647 | 3.64E-11 | 8 | NCALD |
| MCTP13 | 1.90E-14 | 0.81869489 | 0.788 | 0.709 | 3.80E-11 | 8 | MCTP1 |
| EPHA61 | 2.04E-14 | 1.45777034 | 0.763 | 0.751 | 4.08E-11 | 8 | EPHA6 |
| SNRPN | 2.32E-14 | 0.87428997 | 0.763 | 0.78 | 4.63E-11 | 8 | SNRPN |
| PLXNA4 | 2.51E-14 | 1.00728341 | 0.758 | 0.708 | 5.02E-11 | 8 | PLXNA4 |
| CAMK4 | 3.47E-14 | 0.77354956 | 0.737 | 0.762 | 6.93E-11 | 8 | CAMK4 |
| PDE8B1 | 4.18E-14 | 0.68862938 | 0.652 | 0.584 | 8.35E-11 | 8 | PDE8B |
| CACNB2 | 4.19E-14 | 1.16133356 | 0.813 | 0.839 | 8.38E-11 | 8 | CACNB2 |
| CAMK2A | 5.05E-14 | 0.85590731 | 0.753 | 0.749 | 1.01E-10 | 8 | CAMK2A |
| GUCY1A21 | 7.48E-14 | 0.58828432 | 0.768 | 0.73 | 1.50E-10 | 8 | GUCY1A2 |
| CNTNAP41 | 8.60E-14 | 0.85290195 | 0.747 | 0.666 | 1.72E-10 | 8 | CNTNAP4 |
| NR2F1-AS11 | 9.84E-14 | 0.9022251 | 0.763 | 0.757 | 1.97E-10 | 8 | NR2F1-AS1 |
| CNTN31 | 2.44E-13 | 0.83817411 | 0.838 | 0.84 | 4.88E-10 | 8 | CNTN3 |
| KCNH71 | 3.22E-13 | 1.28256364 | 0.783 | 0.787 | 6.44E-10 | 8 | KCNH7 |
| KCTD161 | 3.74E-13 | 1.10685041 | 0.783 | 0.796 | 7.48E-10 | 8 | KCTD16 |
| GRM11 | 3.82E-13 | 1.0108397 | 0.763 | 0.808 | 7.65E-10 | 8 | GRM1 |
| EML6 | 4.18E-13 | 0.86015018 | 0.843 | 0.845 | 8.35E-10 | 8 | EML6 |
| KCTD82 | 6.44E-13 | 0.76526165 | 0.626 | 0.523 | 1.29E-09 | 8 | KCTD8 |
| MPPED21 | 7.06E-13 | 0.59736749 | 0.641 | 0.594 | 1.41E-09 | 8 | MPPED2 |
| FSTL4 | 8.66E-13 | 0.89656609 | 0.753 | 0.777 | 1.73E-09 | 8 | FSTL4 |
| NR2F2-AS1 | 9.30E-13 | 0.26507041 | 0.515 | 0.383 | 1.86E-09 | 8 | NR2F2-AS1 |
| LDB21 | 1.06E-12 | 1.12044003 | 0.864 | 0.883 | 2.13E-09 | 8 | LDB2 |
| ZNF385D1 | 2.35E-12 | 1.21611308 | 0.601 | 0.502 | 4.70E-09 | 8 | ZNF385D |
| TENM31 | 4.41E-12 | 1.32292093 | 0.742 | 0.806 | 8.82E-09 | 8 | TENM3 |
| FAM135B1 | 7.92E-12 | 0.7521129 | 0.712 | 0.72 | 1.58E-08 | 8 | FAM135B |

| | | | | | | | |
|---|---|---|---|---|---|---|---|
| LINC011221 | 8.27E-12 | 1.06089667 | 0.768 | 0.746 | 1.65E-08 | 8 | LINC01122 |
| ST6GALNAC5 | 8.38E-12 | 1.18766777 | 0.778 | 0.829 | 1.68E-08 | 8 | ST6GALNAC5 |
| ESRRG2 | 8.88E-12 | 0.62035756 | 0.626 | 0.546 | 1.78E-08 | 8 | ESRRG |
| NELL21 | 9.19E-12 | 1.09409751 | 0.758 | 0.804 | 1.84E-08 | 8 | NELL2 |
| PACRG | 9.40E-12 | 0.7643397 | 0.621 | 0.566 | 1.88E-08 | 8 | PACRG |
| BRINP1 | 1.29E-11 | 0.8412064 | 0.702 | 0.711 | 2.58E-08 | 8 | BRINP1 |
| NDST3 | 2.15E-11 | 0.71052823 | 0.773 | 0.816 | 4.30E-08 | 8 | NDST3 |
| ANKRD30BL1 | 2.22E-11 | 0.81766056 | 0.697 | 0.744 | 4.44E-08 | 8 | ANKRD30BL |
| AMPH | 2.72E-11 | 0.73404041 | 0.732 | 0.69 | 5.44E-08 | 8 | AMPH |
| TMTC1 | 2.97E-11 | 0.75890732 | 0.682 | 0.684 | 5.93E-08 | 8 | TMTC1 |
| FAM189A1 | 4.36E-11 | 0.70631806 | 0.601 | 0.594 | 8.71E-08 | 8 | FAM189A1 |
| DNER1 | 4.45E-11 | 0.8148497 | 0.641 | 0.611 | 8.90E-08 | 8 | DNER |
| FHOD3 | 5.34E-11 | 0.58998002 | 0.742 | 0.814 | 1.07E-07 | 8 | FHOD3 |
| SORCS1 | 6.02E-11 | 1.16220091 | 0.747 | 0.728 | 1.20E-07 | 8 | SORCS1 |
| KLHL29 | 6.32E-11 | 0.87162754 | 0.525 | 0.401 | 1.26E-07 | 8 | KLHL29 |
| KCNB2 | 1.05E-10 | 1.02702588 | 0.763 | 0.836 | 2.10E-07 | 8 | KCNB2 |
| GRID21 | 1.06E-10 | 0.63111474 | 0.838 | 0.736 | 2.12E-07 | 8 | GRID2 |
| CCDC31 | 1.08E-10 | 0.73695813 | 0.687 | 0.666 | 2.16E-07 | 8 | CCDC3 |
| GABRG31 | 1.36E-10 | 1.15871181 | 0.707 | 0.77 | 2.73E-07 | 8 | GABRG3 |
| R3HDM1 | 1.68E-10 | 0.58506085 | 0.924 | 0.899 | 3.36E-07 | 8 | R3HDM1 |
| DGKB1 | 1.70E-10 | 1.07638595 | 0.793 | 0.849 | 3.39E-07 | 8 | DGKB |
| ZFPM2 | 1.87E-10 | 0.71635238 | 0.727 | 0.741 | 3.73E-07 | 8 | ZFPM2 |
| GRIK11 | 2.98E-10 | 1.75997993 | 0.626 | 0.574 | 5.96E-07 | 8 | GRIK1 |
| RUNX1T1 | 4.82E-10 | 0.83028249 | 0.702 | 0.721 | 9.65E-07 | 8 | RUNX1T1 |
| CHSY31 | 6.03E-10 | 0.53018207 | 0.737 | 0.648 | 1.21E-06 | 8 | CHSY3 |
| ZNF5361 | 6.94E-10 | 0.83140315 | 0.657 | 0.578 | 1.39E-06 | 8 | ZNF536 |
| GABRA2 | 7.28E-10 | 0.53054142 | 0.707 | 0.738 | 1.46E-06 | 8 | GABRA2 |
| KIAA1211 | 1.34E-09 | 0.83940773 | 0.53 | 0.458 | 2.67E-06 | 8 | KIAA1211 |
| KCNMB21 | 1.65E-09 | 1.01732187 | 0.808 | 0.831 | 3.31E-06 | 8 | KCNMB2 |
| BASP1-AS11 | 2.10E-09 | 0.82516944 | 0.732 | 0.796 | 4.19E-06 | 8 | BASP1-AS1 |
| RAB3C1 | 2.32E-09 | 0.7834079 | 0.687 | 0.689 | 4.63E-06 | 8 | RAB3C |
| DTNA | 2.37E-09 | 0.32898477 | 0.924 | 0.847 | 4.74E-06 | 8 | DTNA |
| OLFM31 | 4.52E-09 | 0.94470249 | 0.727 | 0.769 | 9.04E-06 | 8 | OLFM3 |
| SHISA6 | 4.72E-09 | 0.73934436 | 0.672 | 0.692 | 9.45E-06 | 8 | SHISA6 |
| OLFM1 | 5.09E-09 | 0.41005316 | 0.692 | 0.7 | 1.02E-05 | 8 | OLFM1 |
| CEP112 | 5.17E-09 | 0.63347507 | 0.672 | 0.66 | 1.03E-05 | 8 | CEP112 |
| PLD51 | 5.28E-09 | 0.78419756 | 0.662 | 0.621 | 1.06E-05 | 8 | PLD5 |
| SOX51 | 5.85E-09 | 0.93350427 | 0.833 | 0.836 | 1.17E-05 | 8 | SOX5 |
| GALNTL62 | 7.02E-09 | 1.58531729 | 0.672 | 0.711 | 1.40E-05 | 8 | GALNTL6 |
| ARPP212 | 9.00E-09 | 1.08777032 | 0.793 | 0.826 | 1.80E-05 | 8 | ARPP21 |
| CDH41 | 1.03E-08 | 0.58030798 | 0.641 | 0.645 | 2.05E-05 | 8 | CDH4 |
| PARD32 | 1.08E-08 | 0.82265413 | 0.717 | 0.671 | 2.16E-05 | 8 | PARD3 |
| GAD2 | 1.14E-08 | 0.59109249 | 0.545 | 0.492 | 2.27E-05 | 8 | GAD2 |
| SH3GL2 | 1.19E-08 | 0.75528483 | 0.702 | 0.742 | 2.37E-05 | 8 | SH3GL2 |
| GRM8 | 1.24E-08 | 0.92541901 | 0.616 | 0.664 | 2.49E-05 | 8 | GRM8 |
| PWRN11 | 1.36E-08 | 0.5772651 | 0.753 | 0.785 | 2.71E-05 | 8 | PWRN1 |
| COL19A1 | 1.46E-08 | 0.36931915 | 0.616 | 0.593 | 2.92E-05 | 8 | COL19A1 |
| CHN11 | 1.67E-08 | 0.5622007 | 0.727 | 0.67 | 3.33E-05 | 8 | CHN1 |
| PLPPR11 | 1.68E-08 | 0.42808417 | 0.551 | 0.508 | 3.37E-05 | 8 | PLPPR1 |
| EPHA7 | 1.94E-08 | 0.4978391 | 0.742 | 0.804 | 3.87E-05 | 8 | EPHA7 |

| | | | | | | | |
|---|---|---|---|---|---|---|---|
| NXPH2 | 2.26E-08 | 0.37259058 | 0.49 | 0.425 | 4.52E-05 | 8 | NXPH2 |
| SGCD1 | 2.65E-08 | 0.58223856 | 0.818 | 0.803 | 5.30E-05 | 8 | SGCD |
| LINC01725 | 3.01E-08 | 0.3953051 | 0.581 | 0.558 | 6.02E-05 | 8 | LINC01725 |
| CLSTN21 | 4.96E-08 | 1.22785993 | 0.657 | 0.647 | 9.93E-05 | 8 | CLSTN2 |
| SVEP11 | 5.41E-08 | 0.25716998 | 0.465 | 0.366 | 0.00010825 | 8 | SVEP1 |
| PDZD21 | 5.84E-08 | 0.73113936 | 0.843 | 0.835 | 0.00011672 | 8 | PDZD2 |
| NELL11 | 7.27E-08 | 1.03951768 | 0.737 | 0.799 | 0.00014538 | 8 | NELL1 |
| KCNT2 | 7.85E-08 | 0.71944773 | 0.692 | 0.717 | 0.00015692 | 8 | KCNT2 |
| CACNA1E1 | 8.55E-08 | 0.70792095 | 0.672 | 0.698 | 0.00017108 | 8 | CACNA1E |
| EFNA51 | 1.13E-07 | 1.07979493 | 0.758 | 0.854 | 0.00022514 | 8 | EFNA5 |
| CDH131 | 1.17E-07 | 1.06095849 | 0.682 | 0.676 | 0.00023332 | 8 | CDH13 |
| CIT1 | 1.40E-07 | 0.66245802 | 0.768 | 0.777 | 0.00027901 | 8 | CIT |
| NETO2 | 1.72E-07 | 0.41375758 | 0.424 | 0.316 | 0.00034324 | 8 | NETO2 |
| ADARB22 | 1.95E-07 | 2.34059207 | 0.631 | 0.621 | 0.00039086 | 8 | ADARB2 |
| PTPRR | 2.05E-07 | 0.69838252 | 0.687 | 0.778 | 0.00041015 | 8 | PTPRR |
| GALNT131 | 2.15E-07 | 0.65779328 | 0.722 | 0.763 | 0.00042945 | 8 | GALNT13 |
| MYO16 | 2.15E-07 | 0.62094693 | 0.646 | 0.672 | 0.00042977 | 8 | MYO16 |
| PTPRK1 | 2.77E-07 | 0.76354646 | 0.672 | 0.663 | 0.00055419 | 8 | PTPRK |
| STXBP61 | 2.89E-07 | 0.55630424 | 0.581 | 0.546 | 0.00057885 | 8 | STXBP6 |
| ADAM231 | 2.98E-07 | 0.60098043 | 0.652 | 0.658 | 0.00059559 | 8 | ADAM23 |
| C3orf67-AS1 | 3.16E-07 | 0.28385476 | 0.692 | 0.828 | 0.00063166 | 8 | C3orf67-AS1 |
| AC090578.1 | 3.43E-07 | 0.63146678 | 0.742 | 0.796 | 0.00068515 | 8 | AC090578.1 |
| CNR1 | 3.45E-07 | 0.3419239 | 0.596 | 0.608 | 0.00068919 | 8 | CNR1 |
| MMP161 | 4.29E-07 | 1.0092722 | 0.732 | 0.76 | 0.00085787 | 8 | MMP16 |
| POU6F2 | 6.88E-07 | 0.98504098 | 0.712 | 0.771 | 0.00137637 | 8 | POU6F2 |
| SLIT1 | 7.27E-07 | 0.52170886 | 0.657 | 0.636 | 0.00145433 | 8 | SLIT1 |
| PRICKLE1 | 7.41E-07 | 0.67963144 | 0.722 | 0.813 | 0.00148131 | 8 | PRICKLE1 |
| CUX21 | 9.27E-07 | 0.47518355 | 0.641 | 0.682 | 0.00185352 | 8 | CUX2 |
| MICAL21 | 1.08E-06 | 0.4919783 | 0.672 | 0.611 | 0.00215257 | 8 | MICAL2 |
| KLF12 | 1.09E-06 | 0.49841447 | 0.702 | 0.674 | 0.00217659 | 8 | KLF12 |
| ADCY8 | 1.49E-06 | 0.47979485 | 0.596 | 0.594 | 0.00298026 | 8 | ADCY8 |
| RANBP17 | 1.49E-06 | 0.43515232 | 0.505 | 0.48 | 0.00298733 | 8 | RANBP17 |
| LINC02398 | 1.58E-06 | 0.36390692 | 0.692 | 0.745 | 0.00316983 | 8 | LINC02398 |
| ELAVL21 | 1.66E-06 | 0.64907914 | 0.697 | 0.77 | 0.00332074 | 8 | ELAVL2 |
| DSCAM1 | 2.03E-06 | 0.39726006 | 0.944 | 0.941 | 0.0040686 | 8 | DSCAM |
| DOK6 | 2.18E-06 | 0.80477121 | 0.722 | 0.791 | 0.00436223 | 8 | DOK6 |
| ZMAT41 | 2.51E-06 | 0.89497729 | 0.667 | 0.735 | 0.00501147 | 8 | ZMAT4 |
| FAT31 | 2.63E-06 | 0.89829435 | 0.818 | 0.866 | 0.0052673 | 8 | FAT3 |
| SEMA5A | 2.93E-06 | 0.29477899 | 0.48 | 0.404 | 0.00585369 | 8 | SEMA5A |
| PRKG11 | 3.01E-06 | 1.11863644 | 0.778 | 0.859 | 0.00602004 | 8 | PRKG1 |
| DLX6-AS1 | 3.70E-06 | 0.39476437 | 0.672 | 0.687 | 0.00739557 | 8 | DLX6-AS1 |
| PRKCB | 3.89E-06 | 0.2967236 | 0.884 | 0.835 | 0.00778544 | 8 | PRKCB |
| UNC13C1 | 4.01E-06 | 0.78125892 | 0.778 | 0.811 | 0.00801972 | 8 | UNC13C |
| TMEM232 | 4.47E-06 | 0.65609541 | 0.778 | 0.83 | 0.0089306 | 8 | TMEM232 |
| CCDC85A | 4.93E-06 | 0.93562003 | 0.712 | 0.758 | 0.00986404 | 8 | CCDC85A |
| TMEM178A | 5.11E-06 | 0.47560191 | 0.727 | 0.814 | 0.01021311 | 8 | TMEM178A |
| MGAT4C1 | 5.63E-06 | 1.01102152 | 0.697 | 0.781 | 0.01126611 | 8 | MGAT4C |
| LINC013221 | 5.94E-06 | 0.8168682 | 0.641 | 0.736 | 0.0118733 | 8 | LINC01322 |
| NETO1 | 6.15E-06 | 0.83404341 | 0.646 | 0.724 | 0.01229629 | 8 | NETO1 |
| CPE1 | 7.48E-06 | 0.50364236 | 0.682 | 0.719 | 0.01496934 | 8 | CPE |

| | | | | | | |
|---|---|---|---|---|---|---|
| NRG1 | 8.00E-06 | 1.47521753 | 0.742 | 0.804 | 0.01600825 | 8 NRG1 |
| SYT7 | 8.04E-06 | 0.39609109 | 0.652 | 0.693 | 0.0160799 | 8 SYT7 |
| CACNB41 | 9.08E-06 | 0.26344254 | 0.929 | 0.947 | 0.0181614 | 8 CACNB4 |
| CDH91 | 9.09E-06 | 0.77541466 | 0.707 | 0.786 | 0.01817329 | 8 CDH9 |
| SLC6A11 | 9.36E-06 | 0.87278081 | 0.657 | 0.688 | 0.01871743 | 8 SLC6A1 |
| VSNL11 | 1.34E-05 | 0.51851699 | 0.702 | 0.706 | 0.02687929 | 8 VSNL1 |
| NYAP2 | 1.36E-05 | 0.61387695 | 0.677 | 0.794 | 0.02722405 | 8 NYAP2 |
| AC124854.1 | 1.48E-05 | 0.27079236 | 0.444 | 0.392 | 0.02967337 | 8 AC124854.1 |
| TSHZ31 | 1.82E-05 | 0.56082286 | 0.556 | 0.569 | 0.03636114 | 8 TSHZ3 |
| SYBU1 | 2.05E-05 | 0.60734664 | 0.657 | 0.74 | 0.04105972 | 8 SYBU |
| CA10 | 2.06E-05 | 0.5112777 | 0.808 | 0.842 | 0.04128692 | 8 CA10 |
| THSD7B | 2.40E-05 | 0.38594028 | 0.535 | 0.519 | 0.04800709 | 8 THSD7B |
| GDA | 2.76E-05 | 0.25726354 | 0.737 | 0.824 | 0.05521155 | 8 GDA |
| ATP2B1 | 2.92E-05 | 0.27609992 | 0.864 | 0.881 | 0.05836306 | 8 ATP2B1 |
| C1orf61 | 2.96E-05 | 0.31507651 | 0.692 | 0.68 | 0.05910602 | 8 C1orf61 |
| AK51 | 3.09E-05 | 0.4215121 | 0.742 | 0.771 | 0.06184807 | 8 AK5 |
| COBLL11 | 3.77E-05 | 0.27645806 | 0.586 | 0.543 | 0.07548399 | 8 COBLL1 |
| CNKSR21 | 4.43E-05 | 0.74445835 | 0.763 | 0.8 | 0.08862557 | 8 CNKSR2 |
| LINC01250 | 4.87E-05 | 0.50492193 | 0.747 | 0.807 | 0.09743377 | 8 LINC01250 |
| RPH3A | 4.88E-05 | 0.52919646 | 0.687 | 0.771 | 0.0976762 | 8 RPH3A |
| SLC17A7 | 5.17E-05 | 0.36981103 | 0.697 | 0.756 | 0.10345193 | 8 SLC17A7 |
| PDE7B1 | 5.21E-05 | 0.48325684 | 0.601 | 0.621 | 0.1041008 | 8 PDE7B |
| TMEM132C1 | 5.35E-05 | 0.51620434 | 0.561 | 0.583 | 0.10705745 | 8 TMEM132C |
| ANK1 | 5.58E-05 | 0.49044322 | 0.616 | 0.68 | 0.11157919 | 8 ANK1 |
| CACNG3 | 5.77E-05 | 0.68216927 | 0.662 | 0.776 | 0.11545536 | 8 CACNG3 |
| PCDH11X | 7.12E-05 | 0.92477265 | 0.621 | 0.712 | 0.14232222 | 8 PCDH11X |
| PTPRM1 | 7.30E-05 | 0.53490028 | 0.672 | 0.693 | 0.14601505 | 8 PTPRM |
| TRHDE1 | 8.56E-05 | 0.50485583 | 0.616 | 0.675 | 0.17119841 | 8 TRHDE |
| AC024901.1 | 9.85E-05 | 0.41092584 | 0.702 | 0.794 | 0.19707403 | 8 AC024901.1 |
| KIAA0319 | 0.00010289 | 0.35287012 | 0.662 | 0.733 | 0.20578599 | 8 KIAA0319 |
| GAD1 | 0.00013203 | 0.42922762 | 0.636 | 0.676 | 0.26405397 | 8 GAD1 |
| EPHA5 | 0.00013473 | 0.80419435 | 0.657 | 0.705 | 0.26945494 | 8 EPHA5 |
| PTPRO | 0.00013912 | 0.42927296 | 0.813 | 0.822 | 0.27824377 | 8 PTPRO |
| KCNN3 | 0.00014477 | 0.28375578 | 0.561 | 0.788 | 0.28954528 | 8 KCNN3 |
| BCL11B | 0.00015578 | 0.43854196 | 0.611 | 0.663 | 0.31156207 | 8 BCL11B |
| AC007563.21 | 0.00015692 | 0.54086585 | 0.707 | 0.79 | 0.31384672 | 8 AC007563.2 |
| GNG4 | 0.00015728 | 0.26108865 | 0.384 | 0.348 | 0.31455863 | 8 GNG4 |
| ADAMTS191 | 0.00017277 | 0.46753487 | 0.682 | 0.752 | 0.34554187 | 8 ADAMTS19 |
| RELN1 | 0.00018092 | 0.65230395 | 0.641 | 0.724 | 0.3618436 | 8 RELN |
| LNX1 | 0.00019997 | 0.40333378 | 0.672 | 0.726 | 0.39994995 | 8 LNX1 |
| VWC2 | 0.00020441 | 0.5503474 | 0.581 | 0.613 | 0.40881561 | 8 VWC2 |
| LMO31 | 0.00021595 | 0.3945797 | 0.778 | 0.843 | 0.4318974 | 8 LMO3 |
| BRINP2 | 0.00022024 | 0.46444899 | 0.601 | 0.639 | 0.44047524 | 8 BRINP2 |
| DGKG | 0.00024417 | 0.60377872 | 0.662 | 0.731 | 0.48834442 | 8 DGKG |
| HOMER1 | 0.00025518 | 0.2742008 | 0.747 | 0.75 | 0.51036047 | 8 HOMER1 |
| GABRG2 | 0.00026854 | 0.56524873 | 0.692 | 0.797 | 0.5370751 | 8 GABRG2 |
| GABRA11 | 0.00027529 | 0.43394511 | 0.672 | 0.74 | 0.55058582 | 8 GABRA1 |
| U91319.1 | 0.00030114 | 0.42664419 | 0.667 | 0.759 | 0.60228375 | 8 U91319.1 |
| MIR124-2HG | 0.0003332 | 0.47114437 | 0.641 | 0.705 | 0.66640329 | 8 MIR124-2HG |
| TOX1 | 0.00034898 | 1.04341952 | 0.606 | 0.682 | 0.69795286 | 8 TOX |

| | | | | | | | |
|---|---|---|---|---|---|---|---|
| STXBP5-AS1 | 0.00036836 | 0.62028138 | 0.571 | 0.579 | 0.73672239 | 8 | STXBP5-AS1 |
| RAB27B | 0.00039292 | 0.47668402 | 0.662 | 0.774 | 0.78583539 | 8 | RAB27B |
| AC025159.11 | 0.00040004 | 0.40998327 | 0.682 | 0.696 | 0.80008765 | 8 | AC025159.1 |
| ARHGEF28 | 0.00045618 | 0.27701092 | 0.737 | 0.819 | 0.91236636 | 8 | ARHGEF28 |
| EGFR | 0.00046802 | 0.59577358 | 0.662 | 0.693 | 0.93603403 | 8 | EGFR |
| SPARCL1 | 0.00048342 | 0.42562994 | 0.722 | 0.799 | 0.96684837 | 8 | SPARCL1 |
| GULP1 | 0.00050033 | 0.25934418 | 0.682 | 0.878 | 1 | 8 | GULP1 |
| CASC15 | 0.00050344 | 0.82490727 | 0.657 | 0.726 | 1 | 8 | CASC15 |
| MCC | 0.00050823 | 0.35673914 | 0.606 | 0.611 | 1 | 8 | MCC |
| BCL11A | 0.00051133 | 0.48095193 | 0.763 | 0.802 | 1 | 8 | BCL11A |
| LINC02389 | 0.00052958 | 0.44157436 | 0.54 | 0.535 | 1 | 8 | LINC02389 |
| LUZP21 | 0.00053003 | 0.40143233 | 0.722 | 0.795 | 1 | 8 | LUZP2 |
| FRAS1 | 0.00061259 | 0.74888722 | 0.672 | 0.756 | 1 | 8 | FRAS1 |
| DNAH6 | 0.00061918 | 0.42483584 | 0.667 | 0.754 | 1 | 8 | DNAH6 |
| BTBD11 | 0.00062325 | 0.53940532 | 0.551 | 0.603 | 1 | 8 | BTBD11 |
| NWD21 | 0.00062606 | 0.3286881 | 0.717 | 0.841 | 1 | 8 | NWD2 |
| GRIN3A | 0.00063995 | 0.32696604 | 0.48 | 0.499 | 1 | 8 | GRIN3A |
| TOX2 | 0.00066941 | 0.36698384 | 0.495 | 0.5 | 1 | 8 | TOX2 |
| SLC1A21 | 0.00067413 | 0.59127559 | 0.747 | 0.789 | 1 | 8 | SLC1A2 |
| AC233296.11 | 0.00067648 | 0.39845398 | 0.545 | 0.588 | 1 | 8 | AC233296.1 |
| CLVS2 | 0.00069952 | 0.39712491 | 0.455 | 0.414 | 1 | 8 | CLVS2 |
| RASGRF22 | 0.0007001 | 0.48767858 | 0.722 | 0.772 | 1 | 8 | RASGRF2 |
| AC132803.1 | 0.00076873 | 0.37674294 | 0.571 | 0.553 | 1 | 8 | AC132803.1 |
| LINC013781 | 0.00085766 | 0.252322 | 0.742 | 0.844 | 1 | 8 | LINC01378 |
| GNAL | 0.00086742 | 0.38093299 | 0.636 | 0.706 | 1 | 8 | GNAL |
| SLC24A3 | 0.00086803 | 0.85508629 | 0.53 | 0.601 | 1 | 8 | SLC24A3 |
| SEL1L3 | 0.00091667 | 0.39731091 | 0.535 | 0.569 | 1 | 8 | SEL1L3 |
| ARHGAP28 | 0.00096136 | 0.34420445 | 0.591 | 0.635 | 1 | 8 | ARHGAP28 |
| PRR16 | 0.00099413 | 0.51067221 | 0.591 | 0.629 | 1 | 8 | PRR16 |
| AC011995.2 | 0.00105329 | 0.34932228 | 0.692 | 0.804 | 1 | 8 | AC011995.2 |
| MLIP | 0.00118654 | 0.51078687 | 0.737 | 0.844 | 1 | 8 | MLIP |
| EPHA3 | 0.00135352 | 0.59956027 | 0.692 | 0.796 | 1 | 8 | EPHA3 |
| GHR | 0.00141112 | 0.59198606 | 0.722 | 0.825 | 1 | 8 | GHR |
| AL591501.1 | 0.00141815 | 0.38925543 | 0.732 | 0.847 | 1 | 8 | AL591501.1 |
| ZNRF3 | 0.00156905 | 0.28898191 | 0.571 | 0.549 | 1 | 8 | ZNRF3 |
| OSBPL10 | 0.00180161 | 0.49567384 | 0.636 | 0.738 | 1 | 8 | OSBPL10 |
| NHS | 0.00182913 | 0.4810144 | 0.672 | 0.746 | 1 | 8 | NHS |
| MSC-AS1 | 0.00184959 | 0.31384488 | 0.722 | 0.829 | 1 | 8 | MSC-AS1 |
| HTR1E | 0.0018525 | 0.42640855 | 0.475 | 0.476 | 1 | 8 | HTR1E |
| AL445623.2 | 0.00199385 | 0.34454487 | 0.611 | 0.648 | 1 | 8 | AL445623.2 |
| LINC005111 | 0.00201161 | 0.45545931 | 0.591 | 0.686 | 1 | 8 | LINC00511 |
| KCTD12 | 0.00205246 | 0.4607791 | 0.712 | 0.823 | 1 | 8 | KCTD1 |
| HS3ST51 | 0.00214 | 0.26977304 | 0.657 | 0.688 | 1 | 8 | HS3ST5 |
| BAALC-AS1 | 0.00234406 | 0.26940387 | 0.384 | 0.558 | 1 | 8 | BAALC-AS1 |
| SLIT21 | 0.00246767 | 0.73066175 | 0.626 | 0.67 | 1 | 8 | SLIT2 |
| PID11 | 0.00294753 | 0.50054423 | 0.818 | 0.866 | 1 | 8 | PID1 |
| XYLT11 | 0.00301299 | 0.37933572 | 0.783 | 0.822 | 1 | 8 | XYLT1 |
| LHFPL31 | 0.00307112 | 0.77310518 | 0.641 | 0.681 | 1 | 8 | LHFPL3 |
| NOS1AP | 0.00325657 | 0.38216672 | 0.843 | 0.88 | 1 | 8 | NOS1AP |
| LY86-AS1 | 0.00355826 | 0.44603297 | 0.758 | 0.764 | 1 | 8 | LY86-AS1 |

| | | | | | | | |
|---|---|---|---|---|---|---|---|
| LHFPL61 | 0.00377668 | 0.6426487 | 0.732 | 0.8 | 1 | 8 | LHFPL6 |
| RIT2 | 0.00378498 | 0.37351781 | 0.601 | 0.67 | 1 | 8 | RIT2 |
| CXXC4-AS1 | 0.00413879 | 0.34395066 | 0.636 | 0.721 | 1 | 8 | CXXC4-AS1 |
| CDHR31 | 0.00419555 | 0.46191091 | 0.566 | 0.605 | 1 | 8 | CDHR3 |
| DLC11 | 0.00456527 | 0.31321888 | 0.556 | 0.577 | 1 | 8 | DLC1 |
| SLC4A42 | 0.00473907 | 0.37057345 | 0.616 | 0.657 | 1 | 8 | SLC4A4 |
| CADPS2 | 0.00479379 | 0.67778082 | 0.722 | 0.749 | 1 | 8 | CADPS2 |
| WASF1 | 0.00516642 | 0.48055967 | 0.692 | 0.761 | 1 | 8 | WASF1 |
| TRPC4 | 0.00526114 | 0.31229187 | 0.667 | 0.742 | 1 | 8 | TRPC4 |
| CARMIL1 | 0.00527983 | 0.44804018 | 0.652 | 0.682 | 1 | 8 | CARMIL1 |
| AC093523.1 | 0.00551342 | 0.37439397 | 0.606 | 0.688 | 1 | 8 | AC093523.1 |
| MED12L1 | 0.00565817 | 0.28340135 | 0.631 | 0.622 | 1 | 8 | MED12L |
| ADAMTSL3 | 0.00607205 | 0.28597079 | 0.611 | 0.669 | 1 | 8 | ADAMTSL3 |
| ZNF804B | 0.00608851 | 0.49912617 | 0.47 | 0.727 | 1 | 8 | ZNF804B |
| MPP7 | 0.00676173 | 0.35027805 | 0.631 | 0.73 | 1 | 8 | MPP7 |
| PCDH151 | 0.00695882 | 1.01883509 | 0.646 | 0.736 | 1 | 8 | PCDH15 |
| DGCR9 | 0.00714472 | 0.52090765 | 0.697 | 0.796 | 1 | 8 | DGCR9 |
| AQP4-AS11 | 0.00766641 | 0.53480661 | 0.677 | 0.76 | 1 | 8 | AQP4-AS1 |
| AC016687.2 | 0.0079734 | 0.80960026 | 0.485 | 0.494 | 1 | 8 | AC016687.2 |
| RIPOR21 | 0.00827556 | 0.57043915 | 0.596 | 0.672 | 1 | 8 | RIPOR2 |
| FBXL71 | 0.00876818 | 0.37903094 | 0.626 | 0.665 | 1 | 8 | FBXL7 |
| OCA2 | 0.00886973 | 0.33178695 | 0.687 | 0.775 | 1 | 8 | OCA2 |
| SLC22A10 | 0.00929114 | 0.73963564 | 0.747 | 0.848 | 1 | 8 | SLC22A10 |
| CTNNA31 | 2.64E-32 | 1.31311546 | 0.934 | 0.751 | 5.27E-29 | 9 | CTNNA3 |
| RNF2201 | 1.93E-29 | 1.21178451 | 0.861 | 0.494 | 3.85E-26 | 9 | RNF220 |
| ST181 | 1.03E-28 | 1.36534833 | 0.869 | 0.569 | 2.05E-25 | 9 | ST18 |
| TF1 | 1.62E-17 | 0.87673719 | 0.705 | 0.319 | 3.24E-14 | 9 | TF |
| PTPRK2 | 1.02E-16 | 1.09885495 | 0.811 | 0.661 | 2.04E-13 | 9 | PTPRK |
| ZNF5362 | 2.58E-15 | 0.82050872 | 0.779 | 0.576 | 5.16E-12 | 9 | ZNF536 |
| MOBP1 | 1.40E-14 | 0.94084888 | 0.762 | 0.555 | 2.79E-11 | 9 | MOBP |
| TMEM1441 | 5.13E-14 | 0.74945395 | 0.852 | 0.679 | 1.03E-10 | 9 | TMEM144 |
| SPOCK32 | 2.57E-13 | 1.02831237 | 0.705 | 0.392 | 5.13E-10 | 9 | SPOCK3 |
| CLMN1 | 1.06E-12 | 1.17414587 | 0.713 | 0.533 | 2.11E-09 | 9 | CLMN |
| PLP11 | 2.69E-12 | 0.54139907 | 0.77 | 0.489 | 5.39E-09 | 9 | PLP1 |
| BCAS11 | 1.02E-11 | 0.78497809 | 0.762 | 0.627 | 2.04E-08 | 9 | BCAS1 |
| GRM31 | 1.79E-10 | 0.99002692 | 0.721 | 0.641 | 3.58E-07 | 9 | GRM3 |
| DAAM21 | 6.50E-10 | 0.81477585 | 0.73 | 0.639 | 1.30E-06 | 9 | DAAM2 |
| MBP1 | 4.70E-09 | 0.32549456 | 0.959 | 0.798 | 9.39E-06 | 9 | MBP |
| LINC020731 | 5.58E-09 | 0.28377205 | 0.861 | 0.749 | 1.12E-05 | 9 | LINC02073 |
| ENPP21 | 6.37E-09 | 0.78713792 | 0.639 | 0.414 | 1.27E-05 | 9 | ENPP2 |
| ERBB42 | 1.34E-08 | 0.43922899 | 0.885 | 0.793 | 2.68E-05 | 9 | ERBB4 |
| C10orf901 | 1.85E-08 | 0.76720878 | 0.689 | 0.537 | 3.71E-05 | 9 | C10orf90 |
| FBXL72 | 9.25E-08 | 0.9609174 | 0.73 | 0.663 | 0.00018505 | 9 | FBXL7 |
| KCNH81 | 1.75E-07 | 0.93201489 | 0.598 | 0.32 | 0.00034938 | 9 | KCNH8 |
| L3MBTL4 | 7.92E-07 | 0.31171441 | 0.934 | 0.855 | 0.00158311 | 9 | L3MBTL4 |
| LINC016081 | 1.72E-06 | 0.96915319 | 0.656 | 0.516 | 0.00344942 | 9 | LINC01608 |
| HHIP1 | 6.07E-06 | 0.73690772 | 0.689 | 0.59 | 0.01213573 | 9 | HHIP |
| ROR11 | 1.53E-05 | 0.27475477 | 0.549 | 0.792 | 0.03061419 | 9 | ROR1 |
| SHROOM41 | 4.51E-05 | 0.6540417 | 0.639 | 0.554 | 0.09019935 | 9 | SHROOM4 |
| PTGDS2 | 4.68E-05 | 0.39685448 | 0.73 | 0.647 | 0.09357727 | 9 | PTGDS |

| | | | | | | | |
|---|---|---|---|---|---|---|---|
| FAM107B1 | 6.05E-05 | 0.64980947 | 0.672 | 0.601 | 0.12103047 | 9 | FAM107B |
| CNTNAP23 | 8.77E-05 | 0.37065225 | 0.926 | 0.902 | 0.17533951 | 9 | CNTNAP2 |
| LPAR11 | 0.00012908 | 0.27332259 | 0.852 | 0.839 | 0.25815556 | 9 | LPAR1 |
| BNC21 | 0.00017652 | 0.49022038 | 0.975 | 0.884 | 0.35303756 | 9 | BNC2 |
| LMCD1-AS11 | 0.00077122 | 0.56162071 | 0.82 | 0.824 | 1 | 9 | LMCD1-AS1 |
| AK52 | 0.00120944 | 0.46986477 | 0.754 | 0.771 | 1 | 9 | AK5 |
| CSF2RA1 | 0.00207205 | 0.28342102 | 0.967 | 0.899 | 1 | 9 | CSF2RA |
| LINC022322 | 0.00215407 | 0.26058238 | 0.992 | 0.969 | 1 | 9 | LINC02232 |
| CERCAM1 | 0.00266145 | 0.55315384 | 0.631 | 0.597 | 1 | 9 | CERCAM |
| PTCHD41 | 0.00271462 | 0.26171717 | 0.885 | 0.768 | 1 | 9 | PTCHD4 |
| DLC12 | 0.00358313 | 0.59395441 | 0.615 | 0.576 | 1 | 9 | DLC1 |
| PIEZO21 | 0.00359927 | 0.70857763 | 0.607 | 0.565 | 1 | 9 | PIEZO2 |
| AC012494.11 | 0.00426089 | 0.96925703 | 0.549 | 0.413 | 1 | 9 | AC012494.1 |
| LGR51 | 0.0051206 | 0.29472219 | 0.393 | 0.465 | 1 | 9 | LGR5 |
| PLA2G4A1 | 0.00535946 | 0.27018879 | 0.91 | 0.855 | 1 | 9 | PLA2G4A |
| MYOF | 0.00547535 | 0.30304223 | 0.885 | 0.777 | 1 | 9 | MYOF |
| GRID22 | 0.00552655 | 0.68404648 | 0.828 | 0.737 | 1 | 9 | GRID2 |
| ENTPD11 | 0.00568955 | 0.27110916 | 0.992 | 0.951 | 1 | 9 | ENTPD1 |
